# SGLT2 inhibition mitigates perturbations in nephron segment-specific metabolic transcripts and mTOR pathway activity in kidneys of young persons with type 2 diabetes

**DOI:** 10.1101/2022.07.23.22277943

**Authors:** Jennifer A. Schaub, Fadhl M. AlAkwaa, Phillip J. McCown, Abhijit S. Naik, Viji Nair, Sean Eddy, Rajasree Menon, Edgar A. Otto, John Hartman, Damian Fermin, Christopher O’Connor, Markus Bitzer, Roger Harned, Patricia Ladd, Laura Pyle, Jeffrey B. Hodgin, Frank C. Brosius, Robert G. Nelson, Matthias Kretzler, Petter Bjornstad

## Abstract

The molecular mechanisms of SGLT2 inhibitors (SGLT2i) remain incompletely understood. Single-cell RNA sequencing and morphometrics data were collected from research kidney biopsies donated by participants with youth onset type 2 diabetes (T2D), aged 12-21 years of age, and healthy controls (HC) to study the effects of SGLT2i on kidney transcriptomics. Participants with T2D were more obese, had higher glomerular filtration rate, mesangial and glomerular volumes than HC. There were no clinically significant differences between participants prescribed SGLT2i (T2Di(+), n=10) and other T2D (T2Di(-), n=6). Transcriptional profiles showed SGLT2 expression exclusively in the proximal tubular (PT) cluster. Transcriptional alterations in T2Di(+) compared to T2Di(-) were seen across most nephron segments, most prominently in the distal nephron. SGLT2i treatment was associated with suppression of genes in the glycolysis, gluconeogenesis, tricarboxylic acid cycle pathways in PT, but enhanced expression in thick ascending limb. The energy sensitive mTOR signaling pathway transcripts were suppressed towards HC level in all nephron segments in T2Di(+). These transcriptional changes were confirmed in a diabetes mouse model treated with SGLT2i. Therefore, the beneficial effects of SGLT2i treatment to the kidneys might be from mitigating diabetes-induced metabolic perturbations via suppression of mTORC1 signaling across nephron segments, including those not expressing SGLT2.

## Introduction

Youth-onset type 2 diabetes (T2D) is an increasingly common cause of diabetes in children and adolescents worldwide(1). It is associated with a more severe clinical course than youth-onset type 1 diabetes (T1D) and is characterized by the frequent appearance of diabetic kidney disease (DKD) in adolescence and young adulthood(2). The Treatment Options for Type 2 Diabetes in Adolescents and Youth follow-up study (TODAY2) documented a 15-year cumulative incidence of albuminuria of greater than 50% in young adults with T2D(3, 4). Yet, the young age and the relatively short follow-up period limit robust data on kidney failure in the TODAY2 cohort. However, among Southwest American Indians, in whom youth-onset T2D was first noted in the 1960s(5), kidney failure is nearly five times as high in midlife (ages 25-54) in those with youth-onset T2D than in those diagnosed with T2D later in life(6). Furthermore, adult Southwest American Indians with youth-onset T2D were more likely to have severe kidney structural lesions on kidney biopsy than those with adult-onset T2D of similar duration(7). These studies underscore the burden of DKD in youth-onset T2D and their lifetime risk of kidney failure and premature death.

Sodium-glucose cotransporter-2 (SGLT2) inhibitors are highly effective therapies that have revolutionized the management of DKD in patients with T2D. (8–11) Although not currently approved by the Food and Drug Administration (FDA) for patients < 18 years of age, SGLT2 inhibitors are often prescribed off-label in patients with T2D due to their high risk of DKD. However, the molecular mechanisms underlying the kidney-protective effects of SGLT2 inhibitors in youth onset diabetes are yet to be elucidated.

Accordingly, the objective of this study was to characterize the effects of youth-onset T2D on kidney tissue and function and describe the impact of SGLT2 inhibition on the morphological and molecular features of early kidney dysfunction in these patients. To this end, protocol research kidney biopsies provided by study participants underwent histological analysis and molecular profiling studies using single-cell RNA sequencing technology. Transcriptomic signatures of perturbed renal energy expenditure and substrate metabolism associated with youth-onset T2D were identified, and the impact of SGLT2 inhibitor use on these transcripts was evaluated.

## Methods

### Study Design and Participants

Adolescents and young adults (N=16) with T2D (12-21 years of age, T2D onset <18 years of age, diabetes duration 1-10 years, and HbA1c <11%) from the Renal Hemodynamics, Energetics and Insulin Resistance in Youth Onset Type 2 Diabetes Study (Renal-HEIR, NCT03584217) and the Impact of Metabolic Surgery on Pancreatic, Renal and Cardiovascular Health in Youth with Type 2 Diabetes (IMPROVE-T2D, NCT03620773) who volunteered for a nested protocol research kidney biopsy were included in this analysis (**Figure 1**). The participants were recruited from the Type 2 Diabetes and Metabolic Bariatric Surgery clinics at the Children’s Hospital Colorado at the Anschutz Medical Campus in Aurora, Colorado. T2D was defined by American Diabetes Association criteria plus the absence of glutamic acid decarboxylase, islet cell, zinc transporter 8, and/or insulin autoantibodies. Exclusion criteria are detailed in **Supplemental Table 1**. The Renal-HEIR and IMPROVE-T2D cohorts have intentionally harmonized study protocols and were both approved by the Colorado Multiple Institutional Review Board (COMIRB). Participants and/or parents provided informed consent as appropriate for age. Participants who opted to undergo the optional kidney biopsy were specifically and additionally consented by the research and biopsy teams. Medication use was recorded for all participants, and T2D treatment, including SGLT2 inhibition, was at the discretion of their medical provider. Normative reference tissue research biopsies were provided by 6 healthy young adult participants in the Control of Renal Oxygen Consumption, Mitochondrial Dysfunction, and Insulin Resistance (CROCODILE) study (NCT04074668).

**Figure 1:**
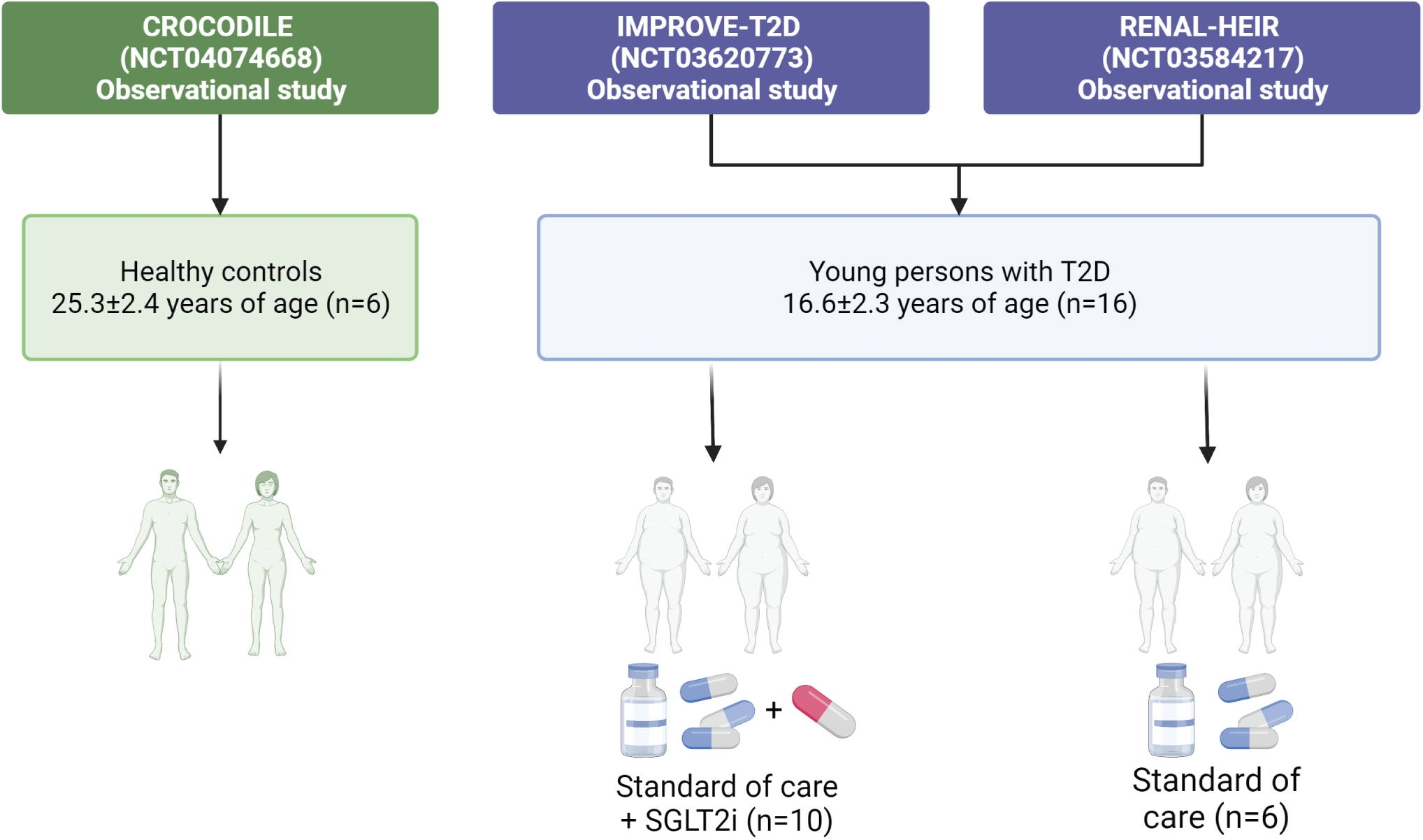
Participant data flow.

### Clinical measurements

In Renal-HEIR and IMPROVE-T2D, iohexol was administered as a bolus IV injection (5mL of 300 mg/ml [Omnipaque 300, GE Healthcare]). An equilibration period of 120 min was used, and blood collections for iohexol plasma disappearance were drawn at +120, +150, +180, +210, and +240 min (12). Because the Brøchner-Mortensen equation underestimates high values of GFR, the Jødal-Brøchner-Mortensen (JBM) equation was used to calculate the GFR(13) . Urine albumin to creatinine ratio (UACR) was measured from fasting untimed urine samples before and after renal clearance assessment and averaged.

All laboratory assays for the Renal-HEIR, IMPROVE-T2D, and CROCODILE cohorts were performed by the University of Colorado Clinical and Translational Research Centers (CTRC) Core Labs and the National Institute of Diabetes and Digestive and Kidney Diseases (NIDDK) laboratory in Phoenix, Arizona. As described previously, iohexol concentrations were measured in Phoenix by high-performance liquid chromatography (Waters, Milford, MA). (14–18) Other fasting laboratory evaluations included: total cholesterol, low-density lipoprotein cholesterol (LDL-C), high-density lipoprotein (HDL-C) cholesterol, triglycerides, glucose, and HbA1c (DCCT-calibrated); assays were performed by standard methods in the CTRC laboratory.

### Ultrasound-guided kidney biopsies and tissue processing

All 3 studies (i.e., Renal-HEIR, IMPROVE-T2D, and CROCODILE) use the same pathology protocol as KPMP. Briefly, an ultrasound-guided percutaneous kidney biopsy was performed by one of two highly experienced interventional radiologists (Drs. Patricia Ladd and Roger Harned). Per local protocol, up to 4 passages were performed to obtain 3 biopsy cores. Each core was immediately assessed for the presence of cortex by gross examination and digital imaging. Kidney tissue was placed in specific fixatives and shipped to the University of Michigan.(19) (https://dev.kpmp.org/wp-content/uploads/2019/06/KPMP_Pathology_Protocol_2019-06-07.pdf)

### Quantitative morphometrics

Light microscopy sections were assessed for pathologic diagnosis. For quantitative assessment of glomerular and mesangial volume and mesangial nuclear count, all glomeruli present in a 3 µm formalin-fixed paraffin-embedded section, stained with Periodic-Acid-Schiff, of each specimen, were assessed using quantitative morphometrics as previously described (20, 21). Mesangial index is % mesangial area per glomerular volume.

### Sample processing and Single-cell RNA sequencing

scRNAseq profiles were obtained using KPMP protocols.(22) Briefly, single cells were isolated from frozen tissues using Liberase TL at 37^0^C for 12 minutes. The single-cell suspension was immediately transferred to the University of Michigan Advanced Genomics Core facility for further processing. Sample demultiplexing, barcode processing, and gene expression quantifications were performed with the 10X Cell Ranger v6 pipeline using the hg38 GRCh38-2020-A reference genome(23–25). To remove ambient mRNA from the data, the cell ranger count matrices were processed using SoupX(26) using the default parameters. The resulting matrices were processed as previously described^22^, whereby cells were included only if gene counts were between 500 and 5000, with fewer than 50% mitochondrial genes. Individual matrices were then integrated using RunHarmony embedded in Seurat, version 4.0.0. Clusters were annotated based on previously established kidney cell markers (27, 28) (**Supplemental Figure 1A**).

### Murine Models

To validate our findings in the human data, we used RNA-sequencing data from the kidney tissue of recently published mouse models. In brief, female db/db mice BKS.Cg-Dock7m +/+ Leprdb/J from Jackson, BKS background, Lot 642) underwent left nephrectomy at 5 weeks of age and, at 12 weeks, infected with renin expressing adeno-associated virus (ReninAAV, 109 genomic copies), which accelerates disease progression (29). We compared ReninAAV Uninephrectomy db/db mice who received 2 weeks of SGLT2 inhibitor treatment with untreated ReninAAV Uninephrectomy db/db mice.

### Statistical Analysis

#### Groups

For our analyses, we stratified participants as follows: T2D without SGLT2i (T2Di(-)), T2D with SGLT2i (T2Di(+)), as well as normative reference tissue from healthy controls (HC). Additionally, the term T2D indicates all participants with T2D irrespective of SGLT2i use. Statistical power was limited to compare baseline clinical and morphometric comparisons between biopsy groups; thus, quantitative data are presented as mean and standard deviations.

#### Differential Expression Analysis

To identify the genes potentially influenced by SGLT2 inhibition, the Limma R package(30) was used to fit linear regression models with the Benjamini-Hochberg (BH) procedure to correct multiple testing. We calculated the fold change (Log_2_FC) between two comparisons: T2Di(-) *vs.* HC and T2Di(+) *vs.* T2Di(-). Transcripts were required to pass FDR-adjusted *p*-values <0.05 in T2Di(-) vs. HC and in T2Di(+) *vs.* T2Di(-) to be considered “reversed”. A T2D regulated transcript was considered to be: i) “suppressed” with SGLT2i if the log_2_FC of the transcript level in T2D relative to HC was >0 and log_2_FC T2Di(+) relative to T2Di(-) was <0 and ii) “enhanced” with SGLT2i if its log_2_FC in T2Di(-) relative to HC was <0 and log_2_FC T2Di(+) relative to T2Di(-) was >0 (**Supplemental Figure 2**).

#### Enrichment analysis

Enrichment for transcripts “reversed” by the SGLT2 inhibitor was determined using the enrichR package(31–33) and the Reactome database(34–36). A pathway was considered significant if its *p*-value <0.05 and included at least 5 reversal genes. Based on existing literature, genes and their Reactome terms were further categorized manually into smaller groupings.

#### Proteins Protein Interaction network

The Agilent Literature Search version 2.69 plugin(37) implemented in Cytoscape version 3.7.2(38) was used to generate the protein-protein interaction networks for “reversed” genes.

#### Pathway activity score

The single sample gene set enrichment analysis (ssGSEA) method implemented in GSVA R(39) package version 1.40.1 was used to compute the mTOR pathway activity score.

#### Analysis of Mouse Data

Mouse data were analyzed using the identical reversal analysis approach and definitions described for human gene expression studies to identify the genes suppressed/enhanced with SGLT2i. Limma R package(30) for DEGs identification, BH for multiple testing correction, and threshold FDR =0.05 were similarly applied.

Effects of SGLT2i on metabolic pathways were assessed in the murine data after applying the same analytical approach and criteria for defining reversed, suppressed, and enhanced genes as used in the human data.

## Results

### Clinical and Morphometric Characteristics of Cohort

Participants included in this study are summarized in **Figure 1**. When comparing all participants with T2D (both T2Di(-) and T2Di(+) groups) to HC, T2D were, on average, younger and had a higher BMI and percent body fat, greater dyslipidemia, and a higher GFR. The participants with T2D had an even distribution of males and females with an average age of 17±2 years. Only 18% of participants had elevated albuminuria at the time of screening. Participants with T2D, overall, had good glycemic control, and the T2Di(-) (n=6) and T2Di(+) (n=10) groups were well matched in terms of clinical, laboratory, and morphometric parameters (**Table 1**). The median time on SGLT2 inhibitors in the T2Di(+) group was 5 months.

**Table 1.**
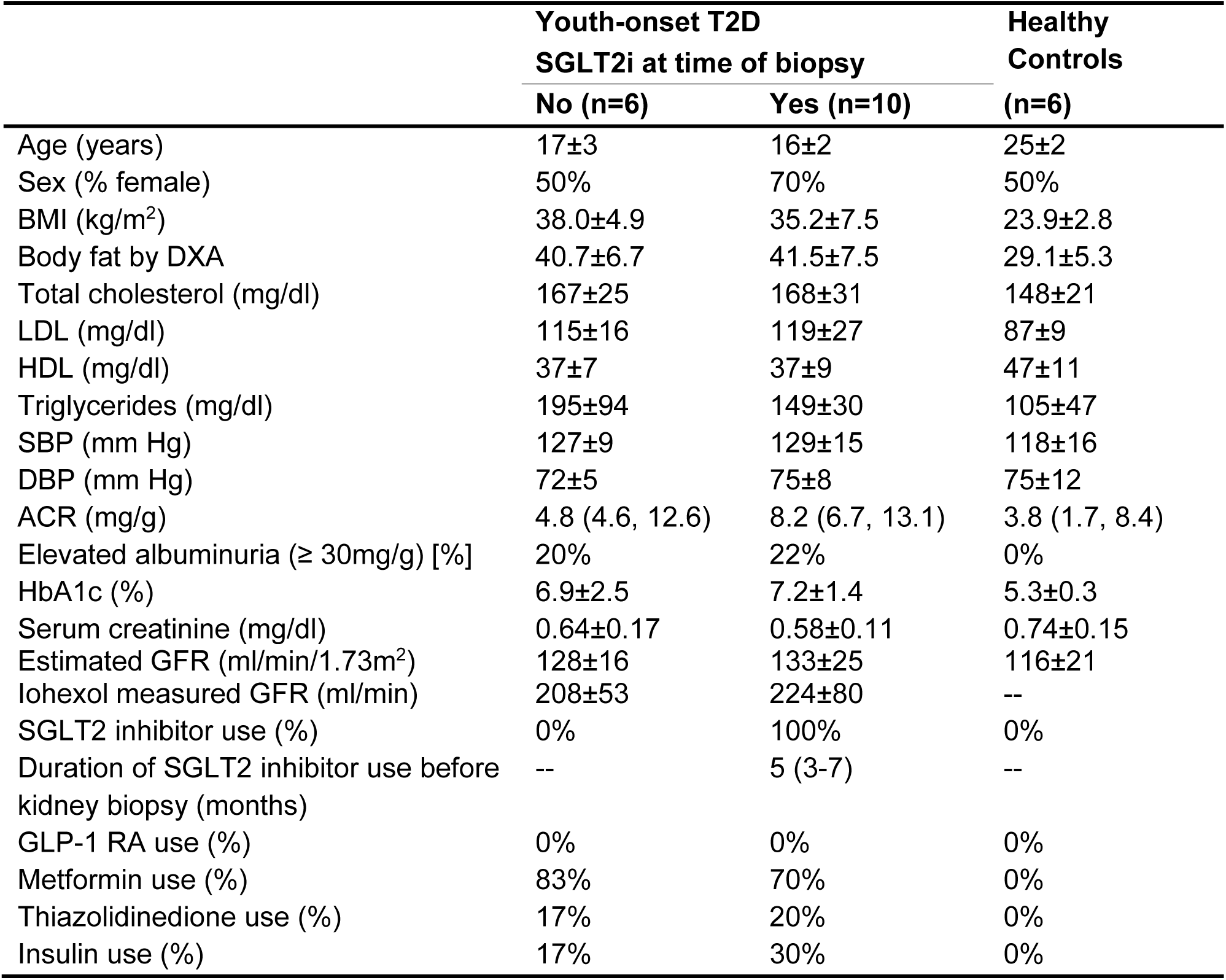
Participant Characteristics Stratified by T2D status and SGLT2 inhibitor use.

Morphometric assessment of the tissue demonstrated that mesangial matrix, mesangial nuclear count, mesangial volume, and glomerular volume were all quantitatively higher in participants with T2D than in HC. However, the fractional interstitial area was similar (**Table 2**). These data suggest that participants with T2D had findings consistent with early kidney dysfunction but no features consistent with advanced DKD.

**Table 2.**
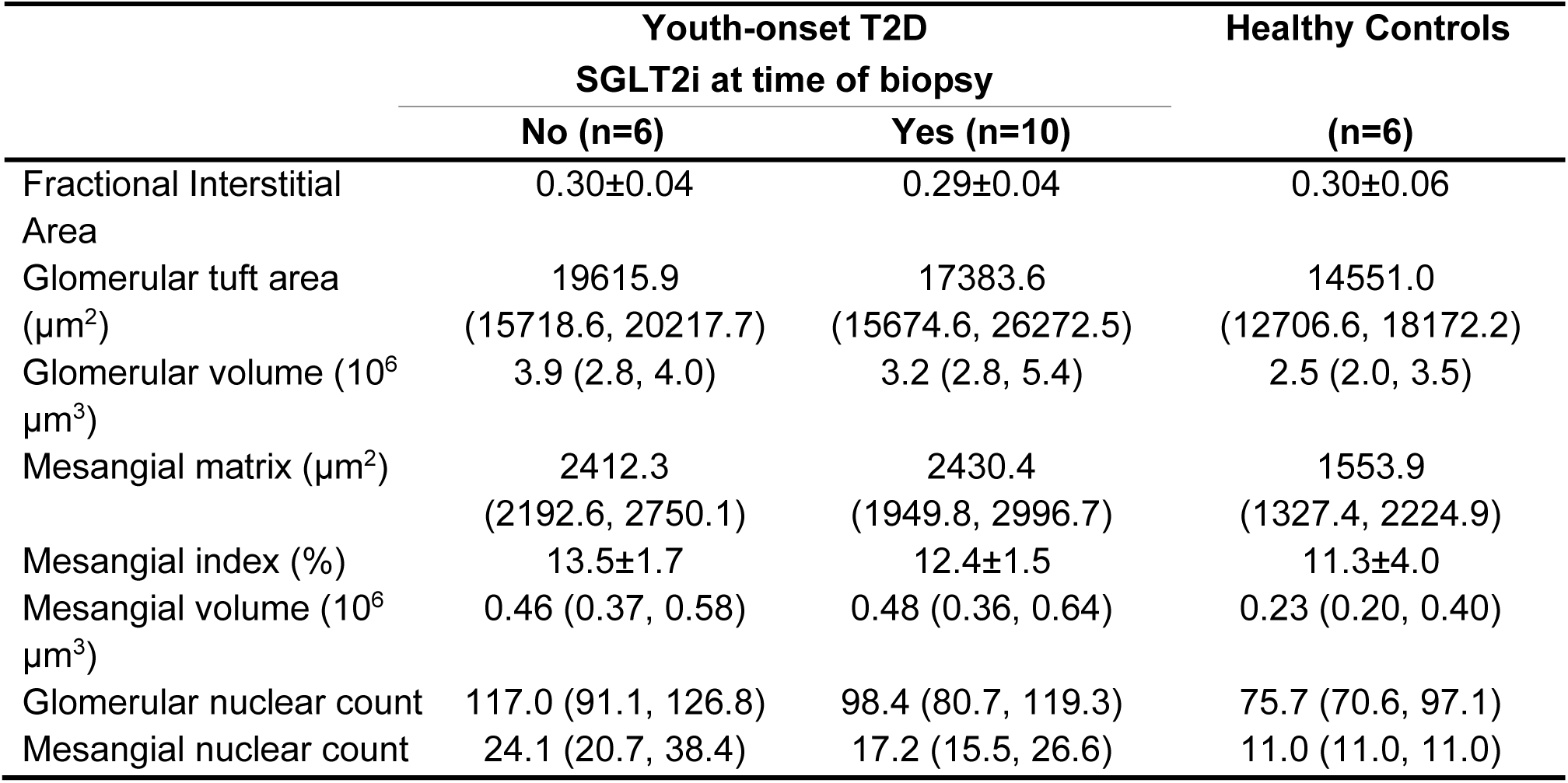
Morphometric Parameters Stratified by T2D status and SGLT2 inhibitor use.

|  | Youth-onset T2D |  | Healthy Controls |
| --- | --- | --- | --- |
|  | SGLT2i at time of biopsy |  | (n=6) |
|  | No (n=6) | Yes (n=10) |  |
| Fractional Interstitial Area | 0.30±0.04 | 0.29±0.04 | 0.30±0.06 |
| Glomerular tuft area (µm<sup>2</sup>) | 19615.9<br>(15718.6, 20217.7) | 17383.6<br>(15674.6, 26272.5) | 14551.0<br>(12706.6, 18172.2) |
| Glomerular volume (10<sup>6</sup> µm<sup>3</sup>) | 3.9 (2.8, 4.0) | 3.2 (2.8, 5.4) | 2.5 (2.0, 3.5) |
| Mesangial matrix (µm<sup>2</sup>) | 2412.3<br>(2192.6, 2750.1) | 2430.4<br>(1949.8, 2996.7) | 1553.9<br>(1327.4, 2224.9) |
| Mesangial index (%) | 13.5±1.7 | 12.4±1.5 | 11.3±4.0 |
| Mesangial volume (10<sup>6</sup> µm<sup>3</sup>) | 0.46 (0.37, 0.58) | 0.48 (0.36, 0.64) | 0.23 (0.20, 0.40) |
| Glomerular nuclear count | 117.0 (91.1, 126.8) | 98.4 (80.7, 119.3) | 75.7 (70.6, 97.1) |
| Mesangial nuclear count | 24.1 (20.7, 38.4) | 17.2 (15.5, 26.6) | 11.0 (11.0, 11.0) |

### scRNAseq Results

From 22 biopsies undergoing scRNAseq, 40,535 cells passed quality control requirements and were annotated to 18 clusters, representing all the major cell types in the nephron (**Figure 2A, Supplemental Figure 1A**). The expression of SGLT2 (*SLC5A2*) was enriched in the proximal tubular cell cluster (PT) (**Figure 2B**). Each cell cluster had a robust representation of the three biopsy groups **(Supplemental Figure 1B**).

**Figure 2:**
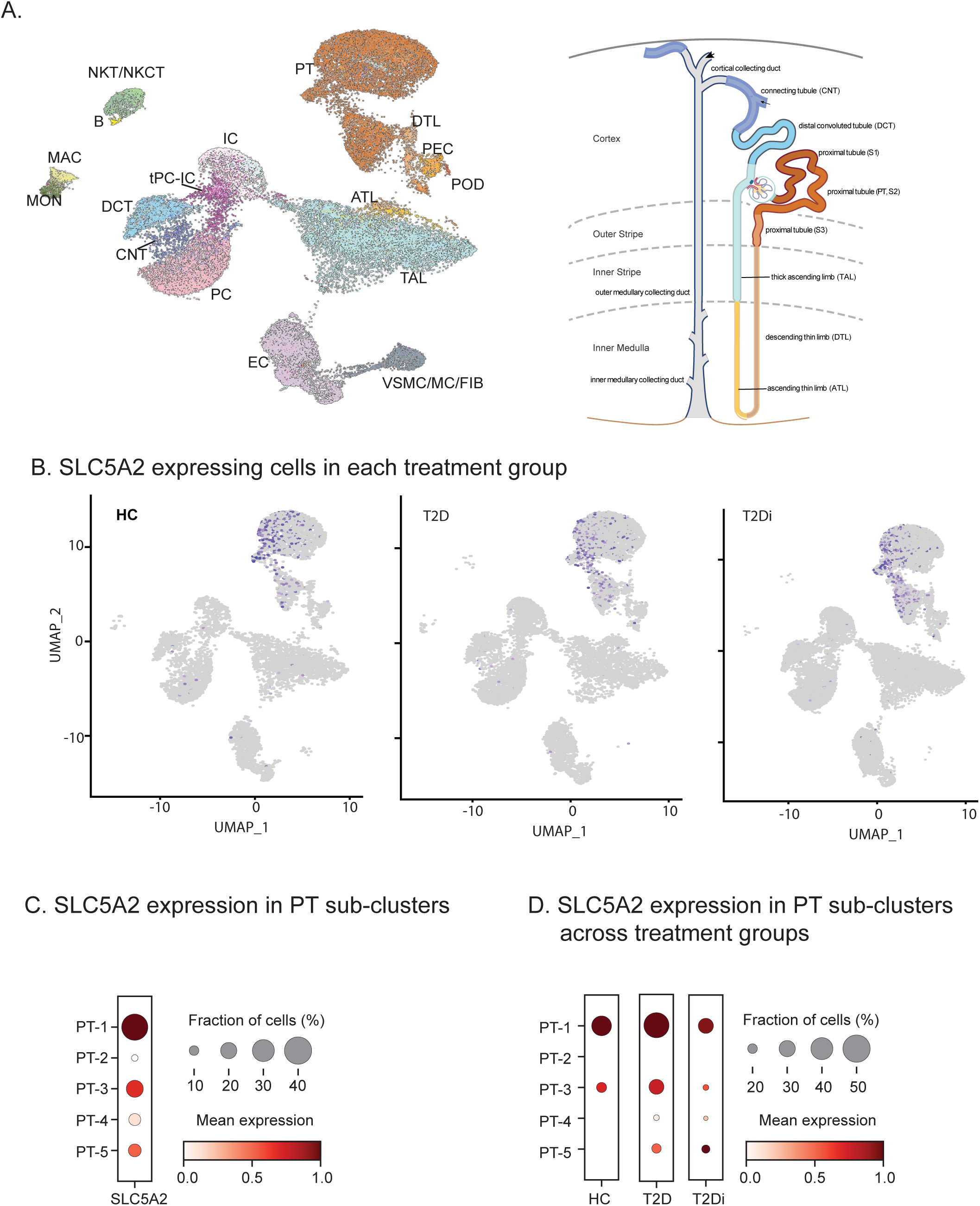
SLC5A2 expression is limited to PT clusters. **A)** UMAP projection of annotated cellular clusters from 3 groups: HC, T2Di (-), and T2Di(+) correspond to all major cell types in the nephron **B)** For all groups (HC, T2Di(-) and T2Di), SLC5A2 expression was limited to the PT cluster. **C)** Fraction of cells (%) expressing SLC5A2 varied in each PT subcluster with PT-1 having the highest expression and PT-2 and PT-4 having the lowest expression. **D)** Decreased percentage of cells expressed SLC5A2 in the T2Di(+) group. Abbreviations: PT - proximal tubule, DTL - descending thin limb, ATL - ascending thin limb, TAL - thick ascending limb, DCT - distal convoluted tubule, CNT - connecting tubule, IC - intercalated cells, PC - principal cells, tPC-IC transitioning intercalated/principal cells, EC - endothelial cells, vSMC/MC/Fib - vascular smooth muscle cells/mesangial cells/fibroblasts, PEC - parietal epithelial cells, POD - podocytes, MAC - macrophages, MON - monocytes, B - B cells, NKT/NKCT - Natural Killer T Cells/Natural Killer Cells with T cells

Within the PT cluster, five sub-clusters were identified using an unbiased approach, (PT-1 to PT-5). The expression of the SGLT2i target gene, *SLC5A2,* was highest in PT-1 followed by PT-3 and PT-5 sub-clusters (**Figure 2C**). PT-1 mapped back predominantly to S1 and S2 nephron segments using the Kidney Medicine Project (KPMP) segment-specific marker set.(40) Most of the other PT subclusters also mapped back to the S1 and S2 nephron segments, with PT-4 contributing minimally to the S3 nephron segment (**Supplemental Figure 3** A-B). PT-5 mostly mapped back to S1 and S2 nephron segments, but there were few cells in this sub-cluster (**Supplemental Figure 3C**). PT-4 had the highest expression of reactive cell markers (**Supplemental Figure 3D**), while cycling cell markers had low expression across and the subclusters (**Supplemental Figure 3E**) and degenerative markers were expressed relatively uniformly across the PT subclusters (**Supplemental Figure 3F**). The proportion of cells in PT-1 expressing *SLC5A2* (encoding SGLT2) was higher in T2DI(-) compared to those in HC and reverted to baseline level in T2DI(+) (**Figure 2D**).

### SGLT2 inhibitors are associated with transcriptional changes throughout the nephron

Despite *SLC5A2* being localized only to the PT clusters, the most significant number of genes reversed in the presence of SGLT2i were in the descending thin limb (DTL), followed by intercalated cells (IC), principal cells (PC), thick ascending limb (TAL), and then PT (**Figure 3A**).

**Figure 3:**
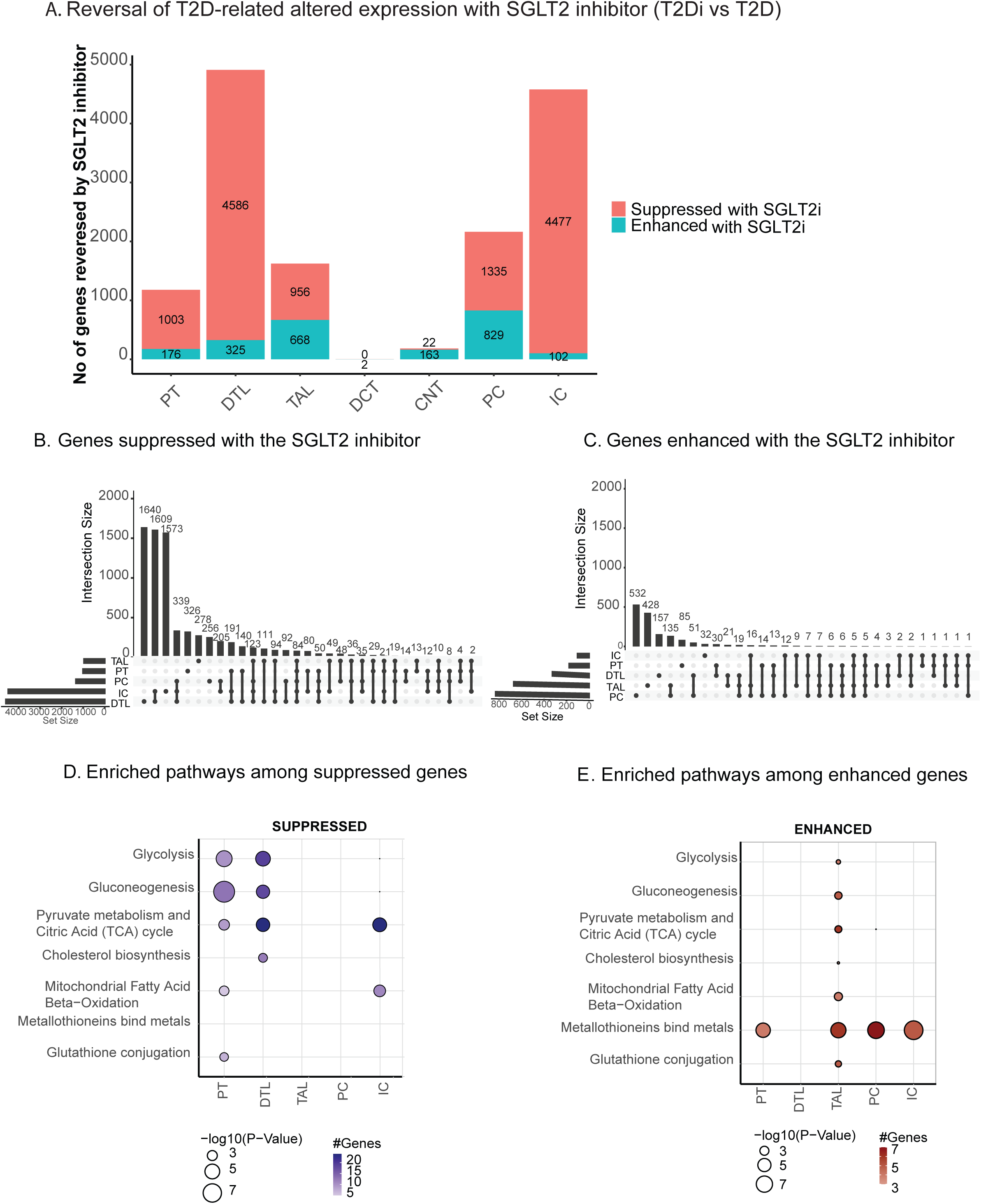
SGLT2 inhibition altered expression in the majority of tubular cell segments. **A)** The majority of genes altered with SGLT2 inhibition were in distal nephron segments. **B)** Most unique genes enhanced with SGLT2 inhibition were in PC, TAL, and DTL. PC and TAL had the greatest number of overlapping genes (135). **C)** Most genes suppressed with SGLT2 inhibition were in DTL, and DTL and IC shared the greatest number of genes. **D)** Using Reactome database, there was suppression of central metabolic pathways in PT, DTL, and IC. **E)** Using the Reactome database and Fisher’s exact test, metallothioneins were enhanced across all segments, except DTL. TAL had enhancement of all central metabolic processes.

Upset plots (**Figure 3B-C**) show the number of unique nephron segment-specific genes that were reversed (enhanced or suppressed) and changes shared between contiguous and non-contiguous tubular segments, and complete results of the reversal analysis are shown in **Supplemental Table 2**. PC and TAL had the greatest number of uniquely enhanced and mutually shared genes, consistent with their shared sodium reabsorption capacities. While DTL, IC, and PC had the greatest number of unique and shared suppressed genes. **Figures 3D and E** demonstrate pathways enriched within the SGLT2i regulated transcript sets. Key metabolic pathways such as glycolysis, gluconeogenesis, pyruvate metabolism, and citric acid cycle (TCA) were suppressed from PT to DTL segments. TCA cycle and fatty acid beta-oxidation were suppressed in IC, while PC did not show suppression of transcripts associated with any metabolic process (**Figure 3D**). However, these major metabolic pathways were enhanced in TAL (**Figure 3E**). The complete enrichment analysis results are available in **Supplemental Table 3**. Pathways involving metal-binding by metallothioneins, critical to mitigating damage from oxidative stress(41), were also enhanced by SGLT2i across all tubular segments except DTL.

#### Proximal Tubular Cells

An examination of the top twenty significantly altered (suppressed: **Figure 4A**, enhanced: **Figure 4B**) pathways confirm suppressed central metabolism-related pathways in PT (**Figure 4A**) with SGLTi exposure compared to T2DI(-). The transcriptional readouts of genes significantly reversed in each of the central metabolic pathways are presented in **Figure 4C**. Within glycolysis-specific enzymes, there was increased expression of key rate-limiting enzymes in T2Di(-) compared to HC, including hexokinase-2 (*HK2*) phosphofructokinase-liver (*PFKL*), and pyruvate kinase (*PKLR*). These genes were suppressed in the T2Di(+) group (relative to T2Di(-)). Similarly, other key glycolytic enzymes, such as aldolase (*ALDOC*), enolase (*ENO*), and glyceraldehyde 3-phosphate dehydrogenase (*GAPDH*), were also suppressed in T2Di. Furthermore, specific rate-limiting enzymes of the gluconeogenic pathways were suppressed, including phosphoenolpyruvate carboxykinase (*PCK1*) and fructose 1,6-bisphosphatase (*FBP1*). These data suggest that SGLT2 inhibitors suppress the elevated glycolysis and gluconeogenesis transcriptional profiles of PT cells in T2Di(-) towards HC.

**Figure 4:**
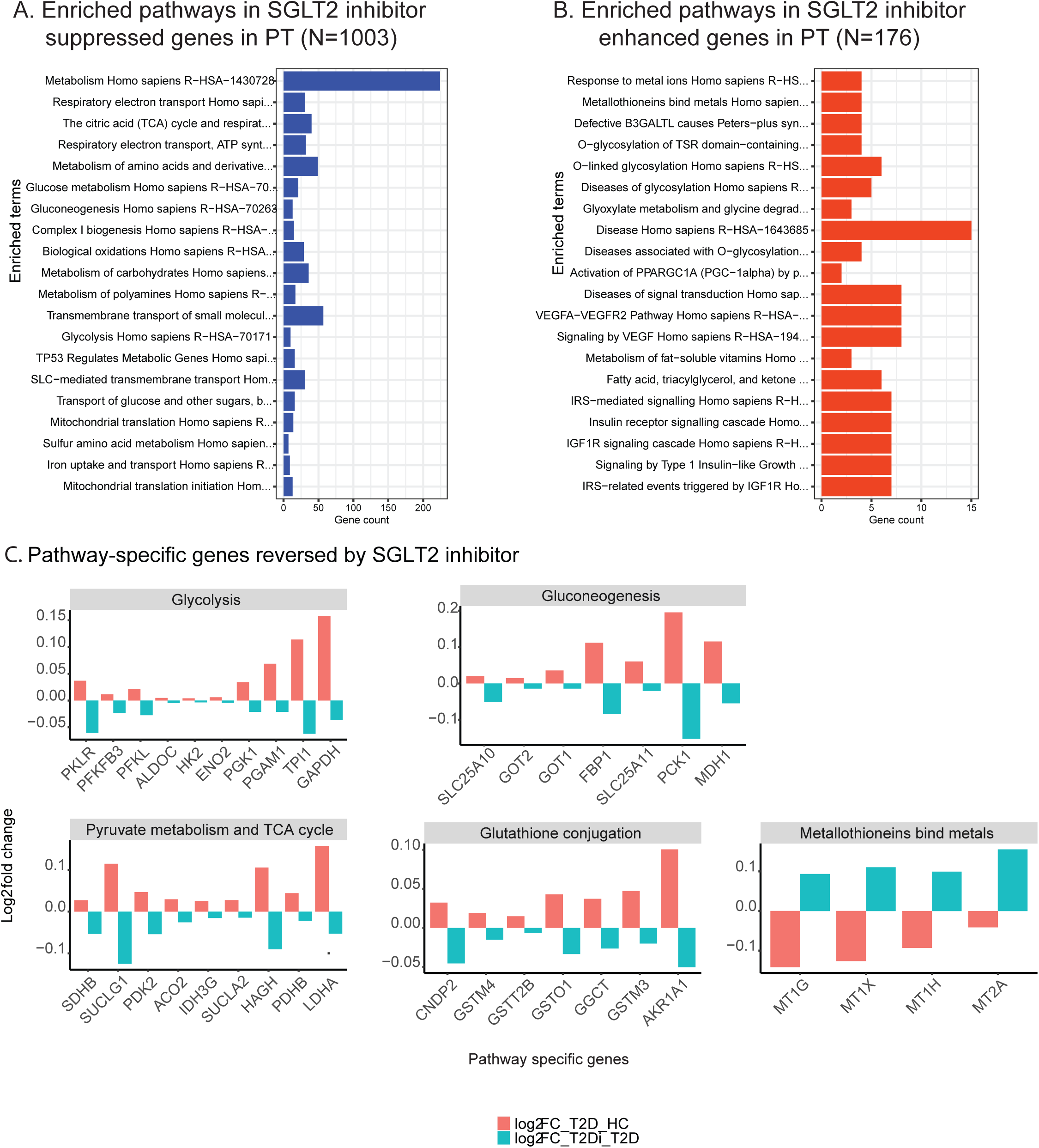
Suppression of central metabolic pathways with SGLT2 inhibition in PT. **A)** Pathway enrichment analysis using Reactome database of enhanced genes with SGLT2 inhibition. **B)** Enrichment analysis using Reactome data of suppressed genes with SGLT2 inhibition shows metabolism as the pathway with the greatest number of altered genes (n>200). **C)** Bar plots showing gene-level alterations (log_2_FC) when comparing T2Di(-) to HC (pink) and T2Di(+) to T2Di(-) (blue).

Downstream of glycolysis, SGLT2i was associated with the suppression of rate-limiting enzymes that allow pyruvate to enter the TCA cycle, such as pyruvate dehydrogenase complex (*PDHB*) and pyruvate dehydrogenase kinase (*PDK2*) (42). Moreover, aconitase (*ACO2*), isocitrate dehydrogenase (*IDH3G*), and components of succinyl Co-A synthase (*SUCLG1, SUCLA2*) were also suppressed. Genes critical to β-oxidation(43) like acyl-CoA dehydrogenase long-chain (*ACADL*) showed diminished expression in T2Di(-) (vs. HC) but were relatively enhanced in T2Di. Additionally, metallothionein, involved in mitigating oxidative stresses (41) was consistently enhanced by SGLT2i in PT (**Figure 4C**).

Differentially expressed gene (DEG) profiles in PT were visualized as physical networks using protein-protein interaction networks to demonstrate the effects of genes on central metabolism and oxidative stress. Although SGLT2 inhibitors reversed the direction of expression for many of these genes, they did not all completely revert to the HC state, as evidenced by the presence of significant logFC when comparing T2Di(+) to HC (**Supplemental Figure 4**).

#### Thick Ascending Limb

The TAL response differed from other nephron segments, with central carbon metabolism being the top pathway significantly enriched in genes enhanced by SGLT2i, whereas in PT, central carbon metabolism was one of the top suppressed pathways (**Figure 5A-B**). There were differences between the expression of key pathway genes for glycolysis in TAL compared to PT. For example, in PT, genes in the glycolysis pathway (*ALDOB, TPI1, PFKM, GAPDH*) had significantly elevated expression in T2Di(-), which were suppressed in T2Di(+) (**Figure 4C**). Whereas in TAL, these genes or corresponding isozymes (*ALDOC, TPI1, PFKL, GAPDH*) were suppressed in T2Di(-) and then enhanced with SGLT2 inhibition. Gluconeogenesis was among the pathways in TAL enhanced by SGLT2 inhibition (**Figure 2D**). However, the key gluconeogenic enzyme phosphoenolpyruvate carboxykinase (*PCK1*) was suppressed in T2Di(+) (**Figure 5C**). Overall, the findings of the transcriptional response in the distal nephron are consistent with the need to handle an increased glucose load as a consequence of the upstream inhibition of reabsorption in the proximal tubular segments.

**Figure 5:**
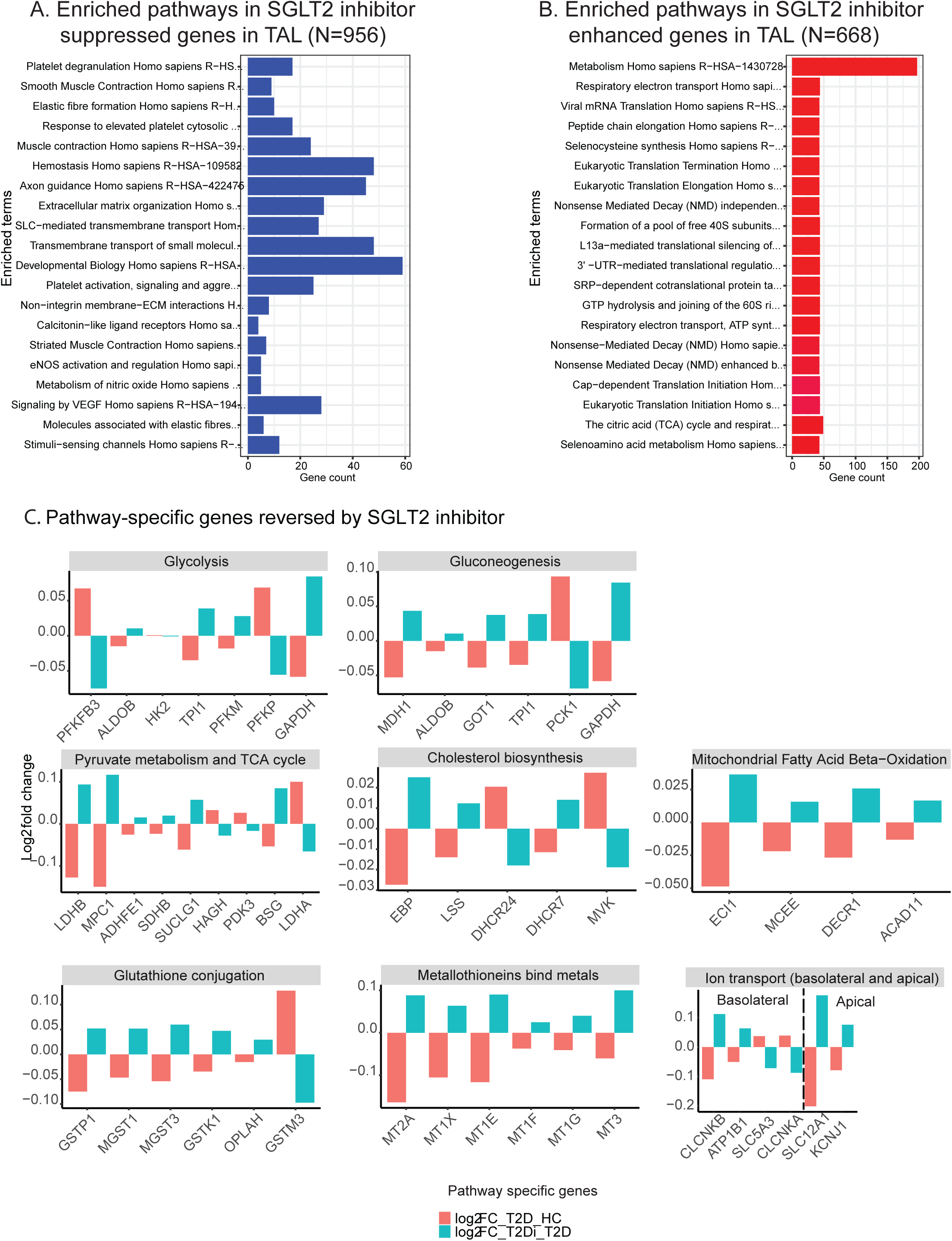
Enhancement of central metabolic pathways with SGLT2 inhibition in TAL. **A)** Enrichment analysis using Reactome database of enhanced genes with SGLT2 inhibition. **B)** Enrichment analysis using Reactome data of suppressed genes with SGLT2 inhibition showing metabolism as the pathway with the greatest number of altered genes (n>200). **C)** Bar plots showing gene level alterations when comparing T2Di(-) to HC (pink) and T2Di(+) to T2Di(-) (blue).

Similarly, TCA cycle genes were affected in opposite directions in TAL and PT with SGLT2i. In TAL, lactate dehydrogenase B, mitochondrial pyruvate carrier 1, and components of succinate synthase and the succinate dehydrogenase complex (*LDHB, MPC1, SUCLG1*, and *SDHB*, respectively) were enhanced with SGLT2i. In contrast, these TCA components were suppressed with SGLT2i in PT. Moreover, the reversal with SGLT2i of lanosterol synthase (*LSS*) and delta-7 sterol reductase (*DHCR7*) expression were unique to TAL, which code for enzymes that convert oxidosqualene into lanosterol and also convert 7-dehydrocholesterol into cholesterol, respectively. Several genes that oxidize branched and unsaturated fatty acids, including Enoyl-CoA Delta Isomerase 1 (*ECI1*), methylmalonyl CoA Epimerase (*MCEE*), and acyl CoA dehydrogenase family member 11 (*ACAD11*) were enhanced in T2Di(+).

Different pathway members involved in metal binding metallothioneins in PT and TAL were enhanced with SGLT2i in both nephron segments. The glutathione conjugation pathway was suppressed in PT but enhanced in TAL. The direction of expression changes of glutathione conjugation pathway members in PT and TAL was also opposite (**Figure 4C and 5C**), except for glutathione S-transferase mu 3 (*GSTM3*), which was suppressed in both segments with SGLT2i.

The expression of several genes related to ion transport, both basolateral and apical, was found to be reversed in TAL. Of these, the expression of electroneutral sodium/potassium/chloride transporter (NKCC2, *SLC12A1*), the apical tubular flow-mediated potassium rectifier channels (ROMK channels, *KCNJ1*), chloride voltage-gated channel B (*CLCNKB*), and the basolateral Na/potassium ATPase (*ATP1B1*) were enhanced by SGLT2i. Other ion transporters up-regulated in T2Di(-), like *SLC5A3* that codes for the Sodium-Myo-inositol Cotransporter critical for maintaining intracellular tonicity(44), were suppressed in T2Di(+). These data suggest that, with SGLT2 inhibition, TAL may have to compensate for increased sodium load resulting in transcriptional changes to support these increased energy demands.

Similar enrichment analyses are summarized for DTL, PC and IC in **Supplemental Figure 5** with the complete list for PT, TAL, DTL, PC and IC nephron segments provided in **Supplemental Table 3.**

### Validation of findings in murine models of diabetes treated with SGLT2 inhibitors

To validate the observed reversal by SGLT2i of metabolic pathways and genes in tubular segments impacted by T2Di(-), we compared this cohort to the transcriptional data from control and SGLT2i-treated ReninAAV db/db mice after uninephrectomy(29). The suppression of glycolysis, gluconeogenesis, TCA cycle, β-oxidation and glutathione conjugation with SGLT2 inhibition observed in the human PT were replicated in the murine kidney transcriptomic data (**Figure 6, Supplemental Table 4**). Given the preponderance of PT cells in the cortex (>80%), PT is a likely source of the transcriptional changes observed in the bulk murine cortical RNAseq data. Moreover, in the ReninAAV db/db mouse models, none of the metabolic pathways were enhanced at a transcriptional level, which also parallels the human PT data where metal-binding to metallothioneins was the only pathway enhanced in PT (**Figure 3E**). However, none of these genes were detected in the mouse data, precluding validation of this pathway (**Supplemental Figure 6** shows gene level alterations in human PT cells with SGLT2i and mouse cortex with SGLT2 inhibitors).

**Figure 6:**
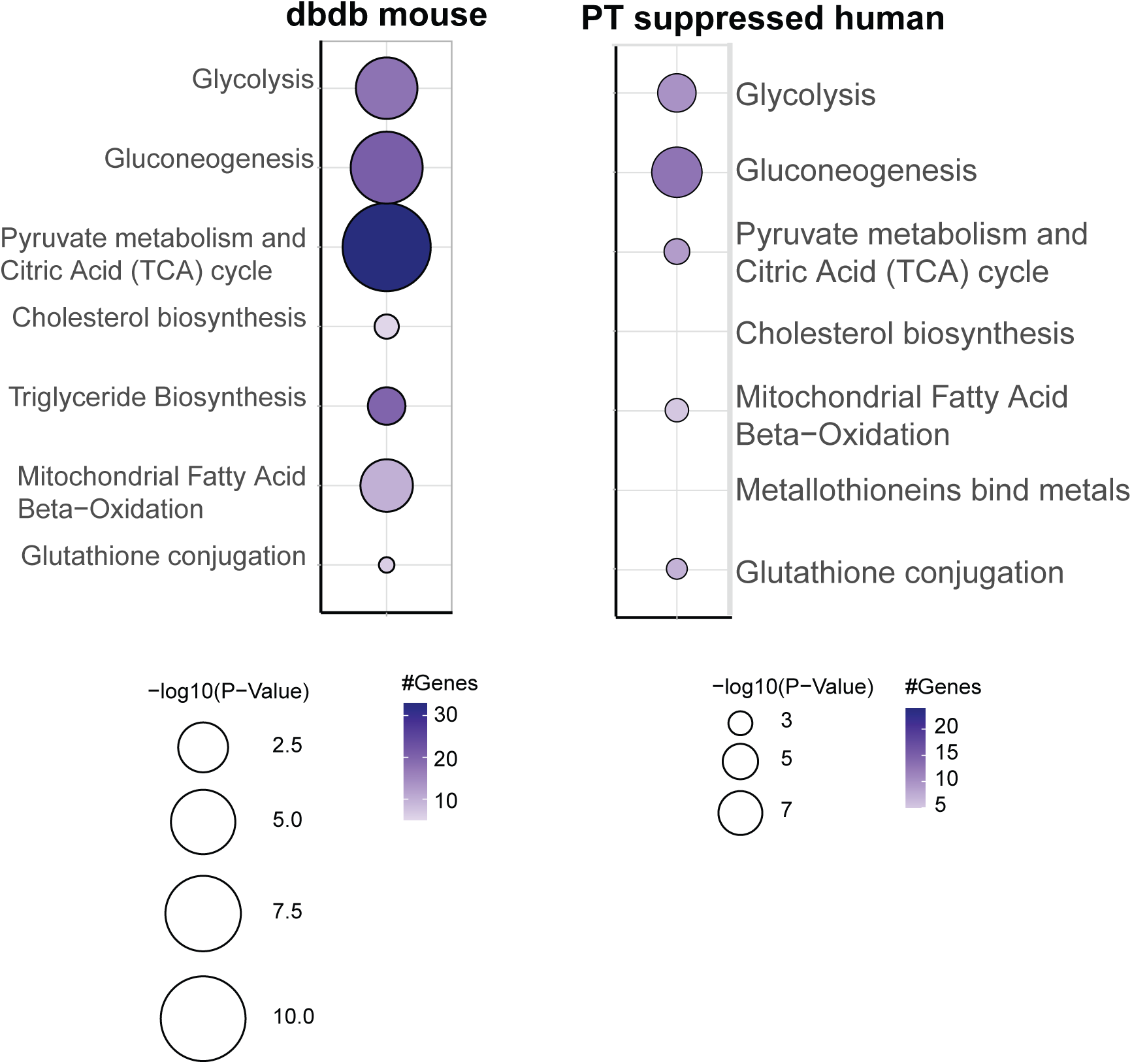
Transcriptomic alterations in Db/Db mouse model treated with SGLT2i validate alterations in central metabolic pathways in PT from humans. Using Reactome database, there was concordant regulation of central metabolic pathways between murine cortex and human PT in response to SGLT2 inhibition.

### mTOR Pathway as a Mediator of Effects of SGLT2 Inhibition

As most nephron segments, except TAL, had suppression of transcripts related to metabolic pathways, we hypothesized that decreased mTORC1 activity, which integrates signals regarding cellular energy state, was a candidate upstream regulator mediating the effects of SGLT2 inhibition. Decreased mTORC1 activity has previously been implicated in animal models(45) as mediating the effects of SGLT2 inhibitors. Using gene sets annotated for mTOR signaling, a delta mTOR pathway activity score for the T2Di(-) group was computed by subtracting the T2Di(-) mTOR score from the HC mTOR score. Similarly, for the T2Di(+) group, the delta mTOR pathway activity score was computed by subtracting the T2Di(+) mTOR score from the HC score. These analyses were performed across all tubular segments. The delta mTOR pathway activity score can be viewed as the departure of each of the group’s mTOR activity scores from the HC control score. SGLT2 inhibitor treatment was associated with lower mTOR activity scores compared to T2Di(-). The difference in the mTOR pathway activity score in all tubular segments in T2Di(+) versus T2Di(-) are shown in **Figure 7A**. These findings were reflected in the corresponding murine model data (**Figure 7B**).

**Figure 7:**
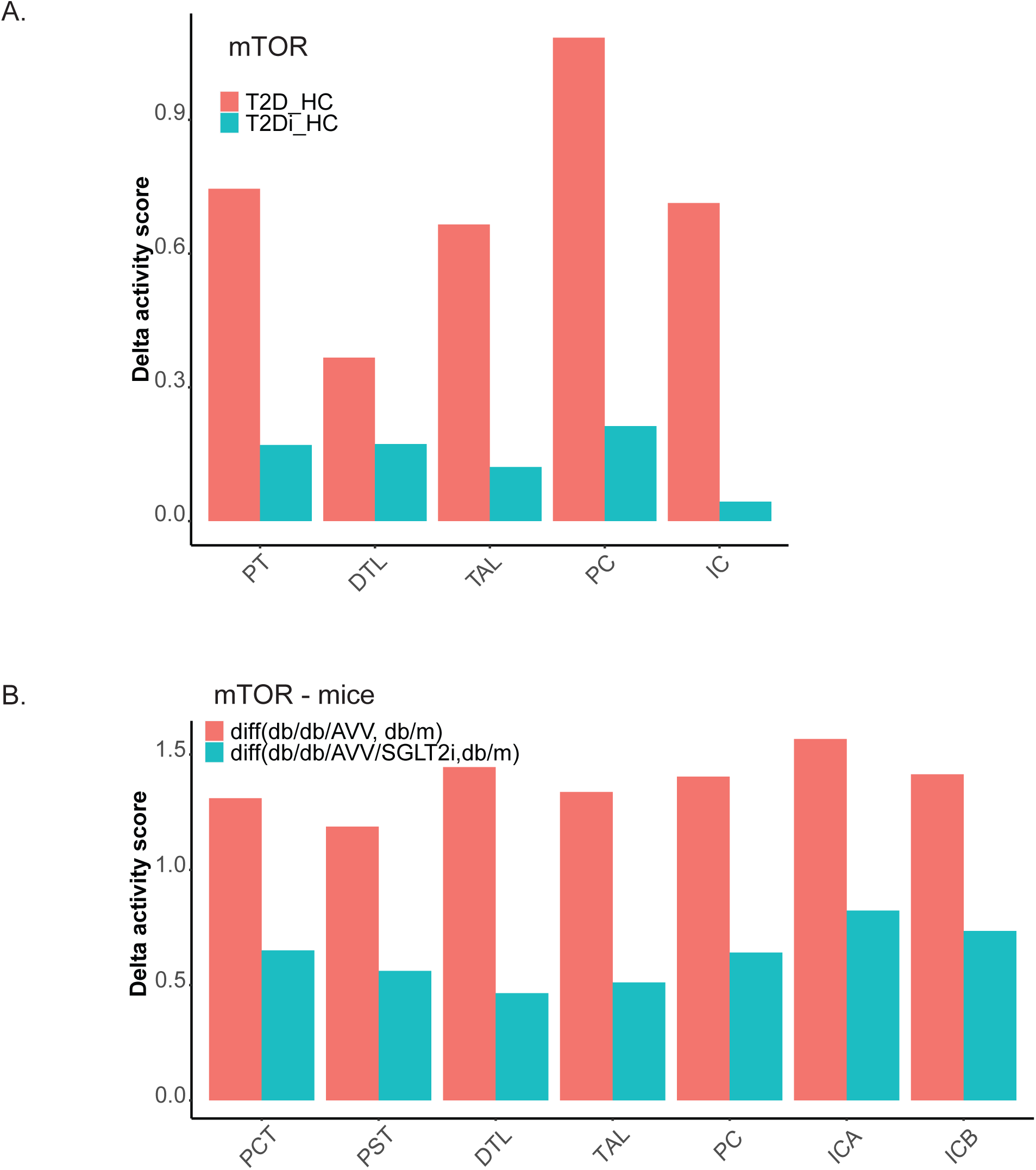
SGLT2i treatment associated mTor activity across nephron segments in human and mice data. Using Reactome database, 39 genes were associated with the mTOR pathway. A) Plot represents mTor pathway activity by nephron segment between T2Di(-) and HC (coral) and between T2Di(+) and HC (aqua). B) Similar comparison in the mouse model data where db/db/AVV is the diabetes model and mTOR activity in these mice was compared to the background db/m mice (coral) and the SGLT2i treated db/db/AVV with db/m mice (aqua).

## Discussion

In protocol research biopsies from a homogenous cohort of young persons with T2D and early kidney dysfunction, this study demonstrated that treatment with SGLT2 inhibitors associated with significant transcriptional changes across several tubular segments in the kidney despite localized SGLT2 expression in PT cells. Transcripts that reversed towards the healthy reference tissue state were enriched in metabolic pathways with subtle differences across nephron segments, with TAL most distinctly affected. The transcriptional regulation of metabolic pathways observed in PT in the human biopsies were validated in kidney cortical tissue from an SGLT2i-treated established diabetic kidney disease mouse model. This suggests that the transcriptional changes identified in our study may also be relevant in the later stages of diabetic kidney disease. In parallel, we observed a significant perturbation in mTOR signaling across all tubular cell types, despite SGLT2 being expressed only in the proximal tubule. Taken together, these data demonstrate that SGLT2 inhibition mitigates the perturbed cellular metabolic profiles induced by a hyperinsulinemic and insulin resistance state and is accompanied by reduction in mTORC1 signaling that is known to drive a DKD phenotype. (46–48)

Differences in morphology were also observed. Quantitatively glomerular volume and mesangial expansion were lower among diabetics treated with an inhibitor. However, sample sizes were limiting for this comparison. The morphology data is consistent with previous rodent models that noted a reduction in glomerular volume in animals treated with an SGLT2 inhibitor.(49) No differences in fractional interstitial area were observed between the three groups suggesting that transcriptomic differences in PTs are not due to structural differences. Another key observation was that multiple metal-binding to metallothionein pathway genes in both proximal and distal nephron segments were consistently increased with SGLT2 inhibitor treatment. Metallothioneins are intracellular metal-binding proteins that mitigate damage from oxidative stress(50, 51). In streptozocin-induced models of diabetes, metallothionein deficiency is associated with increased interstitial fibrosis and inflammatory interstitial infiltrates, which are prognostic markers of progressive DKD(52). While mice also express metallothioneins, they were not enriched in the ReninAAV db/db mouse model treated with SGLT2i. However, the metallothioneins enhanced with SGLT2i in the human data do not have exact orthologs in mice, so our data may highlight an additional pathway to mitigate oxidative stress in humans with SGLT2 inhibition. Murine studies have additionally shown that SGLT2 inhibitors may mitigate oxidative stress via the glutathione pathway in the kidney cortex, particularly glomeruli, but not in the medulla(53). The single-cell resolution of our data suggests that glutathione conjugation pathway activity may differ between PT and TAL, which further supports that there may be cell-specific differences in how the glutathione pathway mitigates oxidative stress with SGLT2 inhibition.

The role of metabolic derangements and perturbed renal energetics as drivers of DKD are well established (54–57). The transcriptional changes observed between our cohort’s diabetic and healthy control states align with prior evidence that diabetes increases glycolysis, β-oxidation, and TCA cycle activity in the kidney(55). Glomerular hyperfiltration in T2D is thought to exacerbate high tubular workload by increasing the filtered Na^+^ load, thereby augmenting SGLT2 activity and adenosine triphosphate (ATP) demand. (58–66) In parallel, T2D is associated with insulin resistance and hyperinsulinemia that increase renal tubular glucose and free fatty acid (FFA) uptake while impairing glucose and FFA oxidation resulting in a state of excess nutrients within the kidney.(56, 67–73) The kidney relies heavily on β-oxidation to meet its high metabolic demands, although there are nephron-segment-specific differences in metabolic processes and energy sources.(54, 74, 75) Notably, PT has a greater capacity for gluconeogenesis than glycolysis compared to the distal nephron, (76) which relies more on glycolysis and oxidative phosphorylation to meet its energy needs.(77)

This study provides evidence that SGLT2 inhibition might revert the transcriptional expression of many metabolic pathway genes closer to the healthy control state in this cohort of youth onset T2D and early renal dysfunction. These findings suggest that part of the beneficial effects of SGLT2 inhibition on the kidney may be through ameliorating some of the metabolic derangements present in T2D, which has been shown in animal studies (53, 55). Further, the scRNA-seq data in this study highlighted that the molecular changes in metabolism-related genes varied in each nephron segment, including anatomically contiguous segments. SGLT2 inhibitor treatment suppressed the up-regulated central metabolism pathway genes observed in T2D in PT, perhaps suggesting an attenuated state of “nutrient excess”. Meanwhile, with SGLT2 inhibitor treatment, TAL showed enhancements to genes involved in glycolysis, gluconeogenesis, TCA cycle, and β-oxidation. As TAL is the next nephron segment with sodium resorbing capabilities downstream of PT, our finding of increased expression of NKCC2 with SGLT2 inhibition could indicate several functional consequences of increased TAL workload. It is plausible that TAL has increased energy demands in response to SGLT2 inhibition to transport the increased tubular sodium content. Indeed, increased sodium transport and oxygen consumption has been predicted with SGLT2 inhibition in diabetic kidneys in rigorous mathematical models (78).

An advantage of transcriptional profiles is the opportunity to identify key transcriptional regulators of cellular responses to an external stimulus, using the mRNA signatures as an activity readout. mTORC1, a protein kinase, is a key signaling hub that integrates signals concerning nutrient and energy status, particularly glucose and branched-chain amino acids, to affect downstream pathways, including lipid metabolism, cell proliferation, cell growth and autophagy with a coordinated transcriptional response (79). Additionally, increased mTORC1 activity is a critical step in podocyte dysfunction in DKD (80), is implicated as a contributor to renal hypertrophy along the nephron and is known to induce a pro-inflammatory phenotype (81). In a recent study in the Akita mouse model of diabetes, SGLT2 inhibition decreased mTORC1 signaling in proximal tubular cells (82). Furthermore, increased mTORC1 activity was sufficient to abolish the beneficial effects of SGLT2 inhibitors (45) thereby suggesting that mTORC1 signaling may mediate the protective effects of SGLT2 inhibition. Using the differences in mTOR transcriptional scores between the diabetic groups and the healthy controls across tubular segments, we show evidence of reduced mTOR1 activation in proximal and distal tubular segments in diabetic patients on an SGLT2i.

SGLT2i were reno-protective in several high-quality clinical trials that enrolled older individuals with established DKD(83–86). The participants in this study had youth-onset T2D, an emerging epidemic with a high risk of early-onset DKD(87). At the time of kidney biopsy, some of these participants exhibited early signs of kidney dysfunction, including elevated GFR and albuminuria. While our sample size was too small for formal analysis, morphometric data also suggested early structural changes, including increased glomerular volume and mesangial expansion, which have been associated with future loss of kidney function(17). Our participants’ age and kidney function precludes extrapolating our results to older individuals with established DKD. However, these data provide important insights into the transcriptional alterations associated with SGLT2 inhibition in a high-risk population early in the disease process. Moreover, the effects of longstanding disease and associated comorbidities on structural and functional changes in kidneys, such as interstitial fibrosis, are minimized in these young participants.

To our knowledge, this is the first study to perform research on kidney biopsies in youth-onset T2D to examine the differences in intrarenal, cell-type-specific transcriptomic signatures associated with SGLT2 inhibitor treatment use. The single-cell resolution of this data provided insights into cell-type-specific responses and enabled the assessment of nephron segment-specific responses in a population of youth with T2D who are at uniquely high risk of DKD.

Our analyses also have important constraints, including the cross-sectional and observational examination, causal inferences of SGLT2 inhibitor treatment, and the relatively small sample size. For this reason, these data should be considered hypothesis-generating. While we do not currently have long-term outcomes data regarding the kidney health of these participants, such data are being collected. Additionally, our results show small absolute transcriptional changes at multiple steps in metabolic pathways, and we do not have metabolite flux data available to measure the changes in intra-renal metabolic flux through these pathways directly. However, we have previously shown that relatively modest transcriptional changes in metabolic pathways correspond to biologically meaningful changes in metabolic flux in DKD(55), and that transcript abundance in scRNAseq data correlates with protein and metabolite levels (88). Lastly, scRNAseq data may not capture genes that are expressed at a low level and in rare cell types, which may hinder our ability to classify cells, scarce cell types, such as juxtaglomerular cells and Type B intercalated cells. Although transcriptional changes may reflect the cell type-specific response to changes in metabolites(55), further investigations with metabolome measurements in this cohort will be needed to determine the ultimate impact of SGLT2 inhibition on the metabolic flux in the kidney. Spatial metabolomic techniques(89) could also provide more insights on the nephron segment-specific changes, particularly when integrating with other novel tissue interrogation technologies(90).

In conclusion, young persons with T2D with early kidney dysfunction exhibited transcriptional signatures of perturbed renal metabolism, which were attenuated in the presence of SGLT2 inhibition. Intriguingly, most transcriptional changes associated with SGLT2 inhibition were in the distal tubular segments where SLC5A2 is not expressed. While further investigations are needed to identify the mechanisms responsible for the changes in the distal nephron segments, our data suggest that alterations in the mTORC1 pathway may mediate some of these transcriptional changes. Future directions include designing a rigorous trial to examine the molecular and metabolic mechanisms by which SGLT2 inhibition mitigates the progression of DKD in T2D, as well as determining whether the transcriptional changes observed in this study predict long-term progression of structural injuries and kidney function decline.

## Data Availability

Data produced in the present study are available upon reasonable request to the authors and based on approval from respective study governing committees. Data produced as part of the present work are contained in the supplemental files.

## Acknowledgements

This work was supported in part by the University of Michigan O’Brien Kidney Translational Core Center grant (P30 DK081943) to M.K. The Renal-HEIR/IMPROVE-T2D and CROCODILE studies were supported by NIDDK (K23 DK116720, R01 DK132399, UC2 DK114886, P30 DK116073), JDRF (2-SRA-2019-845-S-B, 3-SRA-2022-1097-M-B), Boettcher Foundation and in part by the Intramural Research Program at NIDDK and the Centers for Disease Control and Prevention (CKD Initiative) under inter-Agency Agreement #21FED2100157DPG. P.B. receives salary and research support from NIDDK (R01 DK129211, R01 DK132399, R21 DK129720, K23 DK116720, UC2 DK114886), JDRF (3-SRA-2022-1097-M-B, 3-SRA-2022-1243-M-B, 3-SRA-2022-1230-M-B), Boettcher Foundation, American Heart Association (20IPA35260142), Ludeman Family Center for Women’s Health Research at the University of Colorado, the Department of Pediatrics, Section of Endocrinology and Barbara Davis Center for Diabetes at University of Colorado School of Medicine. J.A.S receives salary and research support from NIDDK (K08 DK124449). A.S.N. receives salary and research support from NIDDK (K23 DK125529). MB was supported by R01 DK100449.

## Supplementary Materials

### Supplemental Figures

**Supplemental Figure 1: Quality control for single-cell RNA seq.**

**Supplemental Figure 2: Bioinformatic Pipeline**

**Supplemental Figure 3: Anatomic and Cell State Annotations for Proximal Tubule Sub-clusters**

**Supplemental Figure 4: Protein-protein interaction networks of transcriptional alterations in PT with SGLT2 inhibition.**

**Supplemental Figure 5: Reactome Enrichment Analysis**

**Supplemental Figure 6: Gene level alterations in human PT cells and db/db mouse model treated with SGLT2 inhibitors.**

### Supplemental Tables

**Supplemental Table 1: Exclusion criteria for percutaneous kidney biopsy**

**Supplemental Table 2: List of reversed genes with SGLT2i across all nephron segments.**

**Supplemental Table 3: Pathways enriched by genes enhanced or suppressed with SGLT2 inhibitor across all nephron segments.**

**Supplemental Table 4: List of reversed genes in mouse model experiment with SGLT2i treatment.**

