## Supplemental materials for "SGLT2 inhibition mitigates perturbations in nephron segment-specific metabolic transcripts and mTOR pathway activity in kidneys of young persons with type 2 diabetes"

Supplemental Figure 1: Quality control for single-cell RNA seq. A) Cellular markers used to define cell clusters. B) HC, T2Di(-) and T2Di(+) contributed to each cell cluster.

A.

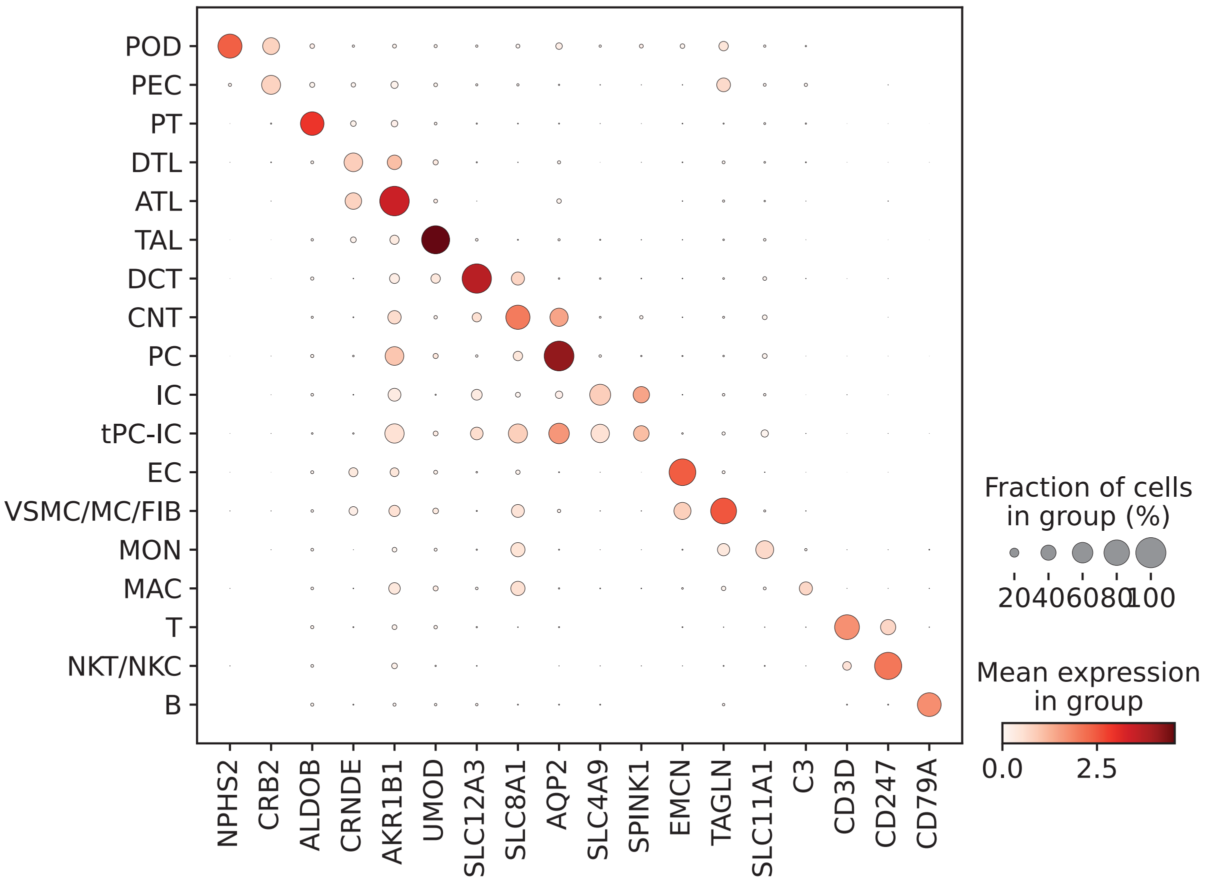


B.

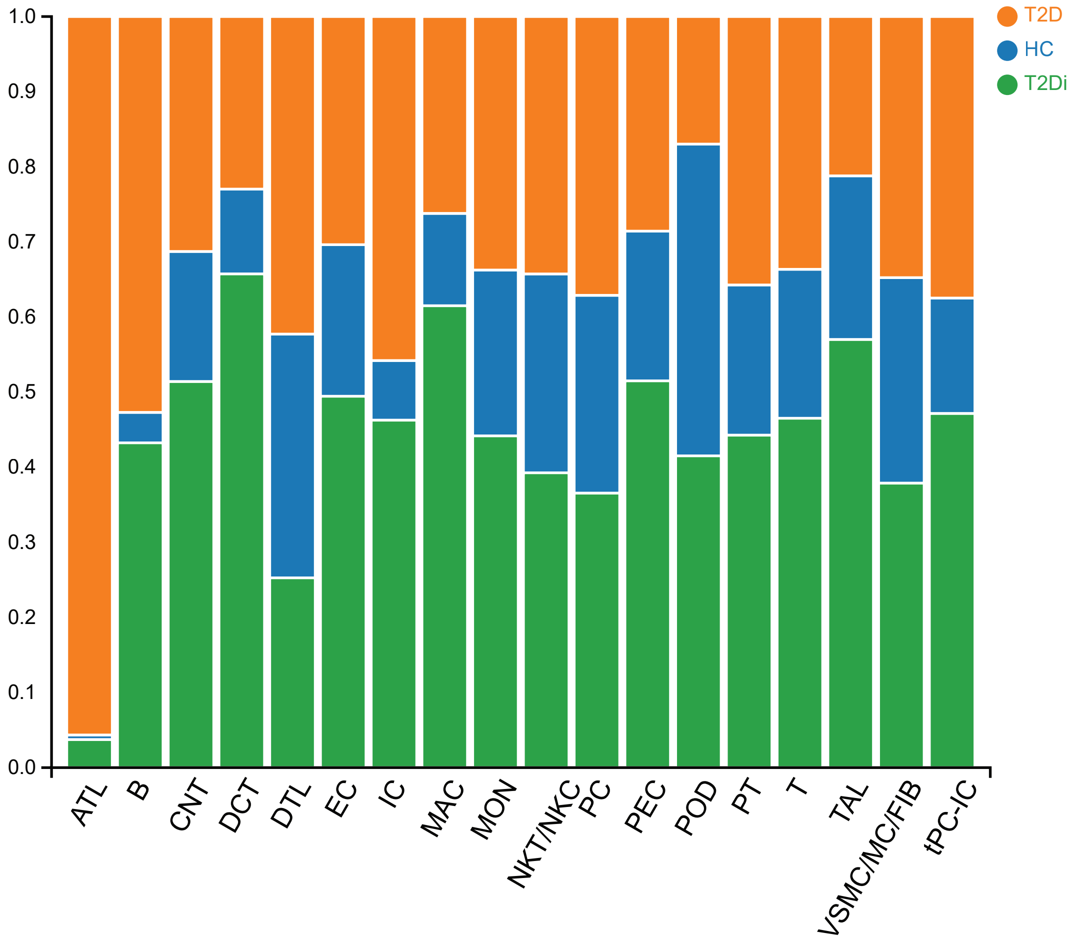

Supplementary Figure 2

Bioinformatic pipeline

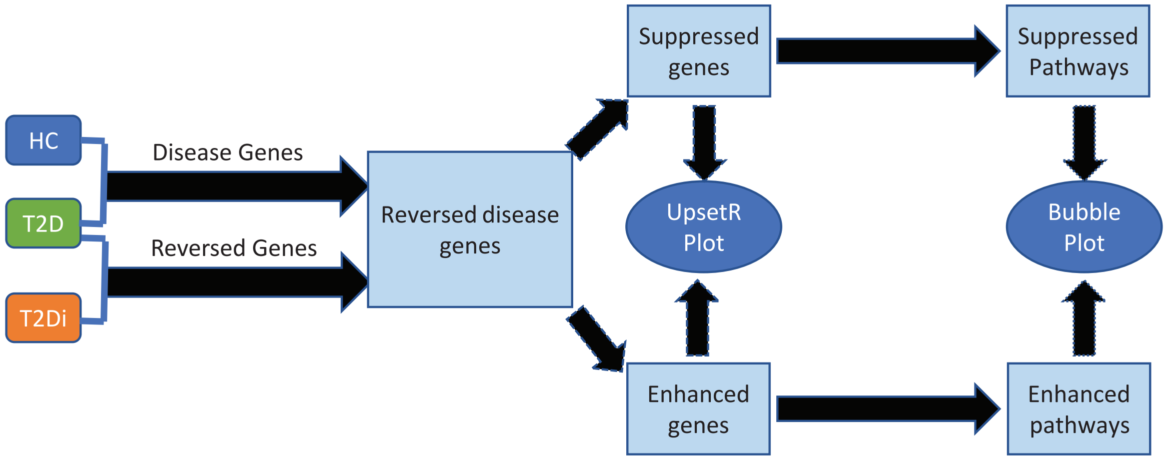

**Supplemental Figure 3. Anatomic and Cell State Annotations for Proximal Tubule Sub-clusters**

A-C) Transcriptional profiles from cells from HC, T2Di(-), or T2Di(+) were analyzed for proximal tubule (PT) biomarkers for compartments S1, S2, or S3, from the KPMP Consortium*. Compartment-specific transcripts were more abundant in clusters PT-1 through PT-3 (A) than PT-4 (B) or PT-5 (C). D) Adaptive/maladaptive state biomarkers defined by KPMP for PT-1 through PT-5. E) Cell cycling state biomarkers defined by KPMP for clusters PT-1 to PT-5. F) Degenerative state biomarkers from the KPMP for clusters PT-1 through PT-5.

*-Lake BB et al. bioRxiv. 2021:2021.07.28.454201.

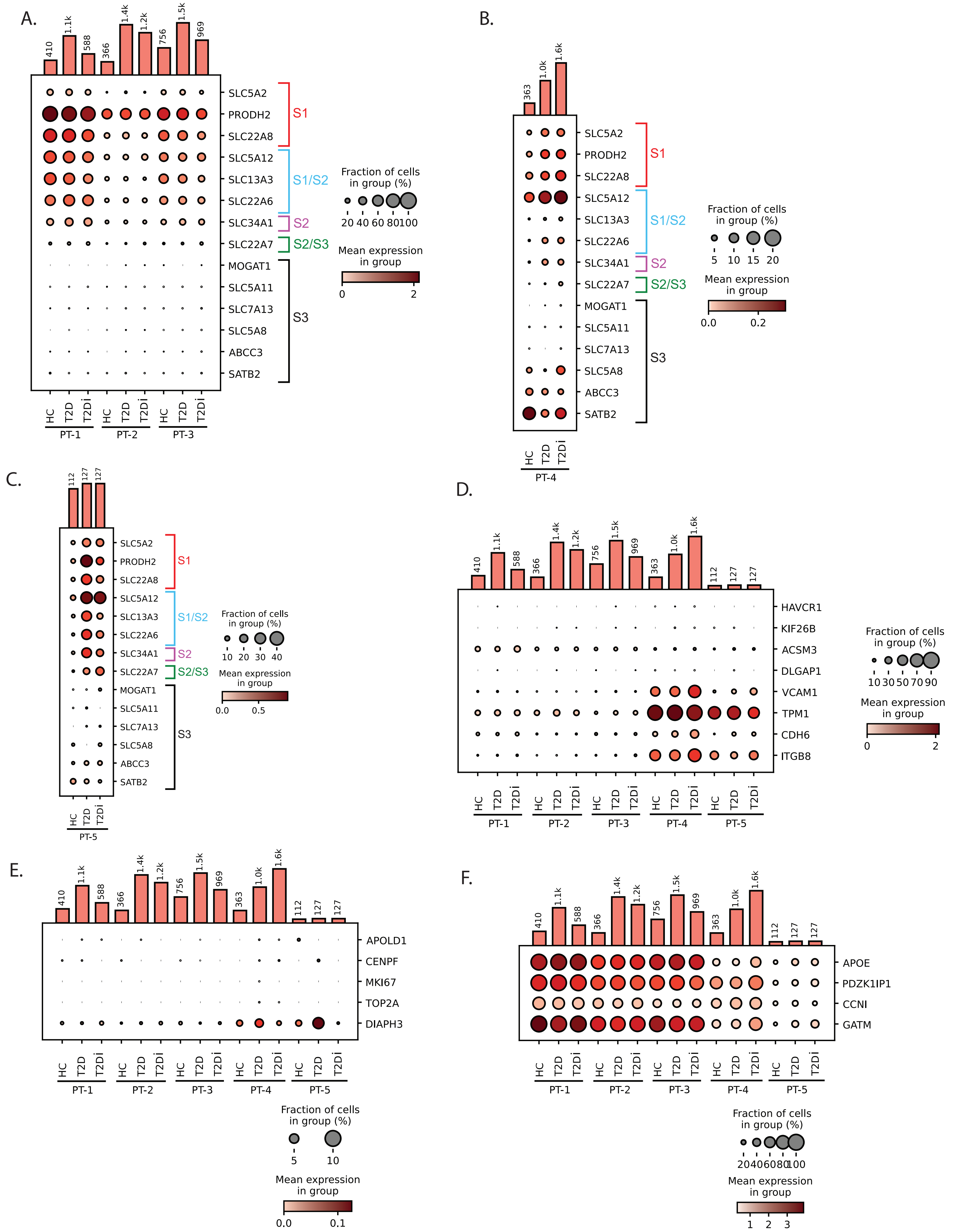

Supplemental Figure 4: Protein-protein interaction networks of transcriptional alterations in PT with SGLT2 inhibition. Protein-protein interaction networks in all PT clusters comparing A)T2Di(-) to HC, B) T2Di(+) to T2Di(-) and C) T2Di(+) to HC.
Each dot represents an individual gene and is color coded for any gene with a significant logFC ( red: 0 to 0.1 and blue: -0.1 to 0 logFC). Comparison between T2Di(+) to HC (panel C) shows that SGLT2 inhibition retains some differences from HC.

A. T2Di(-) to HC

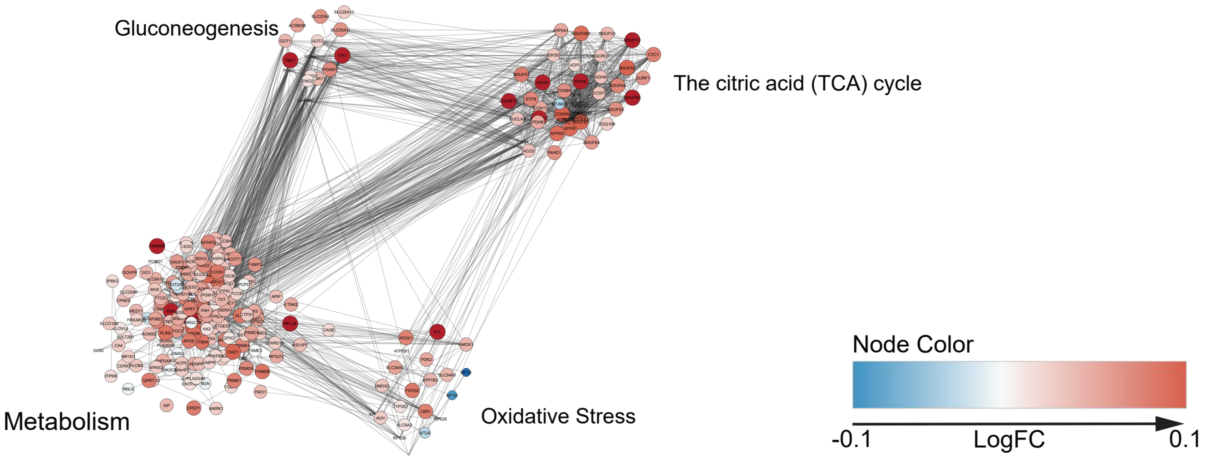


B. T2Di(+) to T2Di(-)

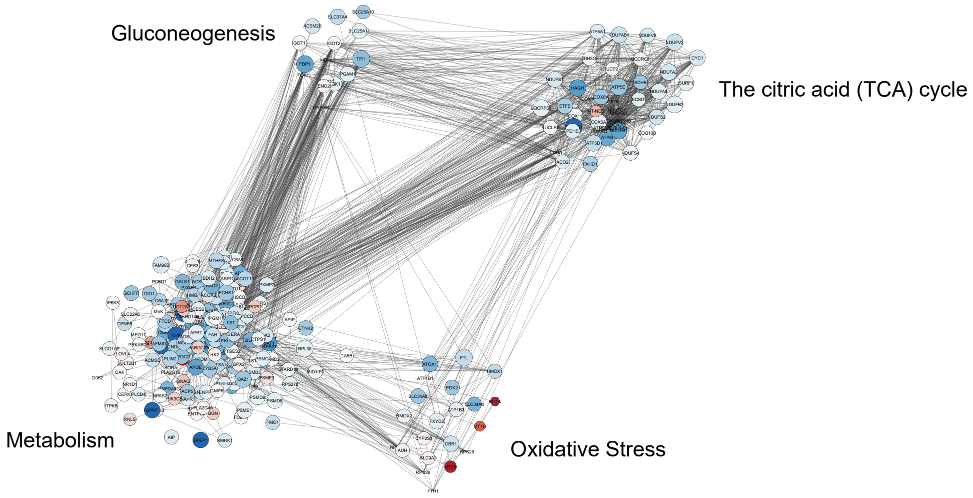


C. T2Di(+) to HC

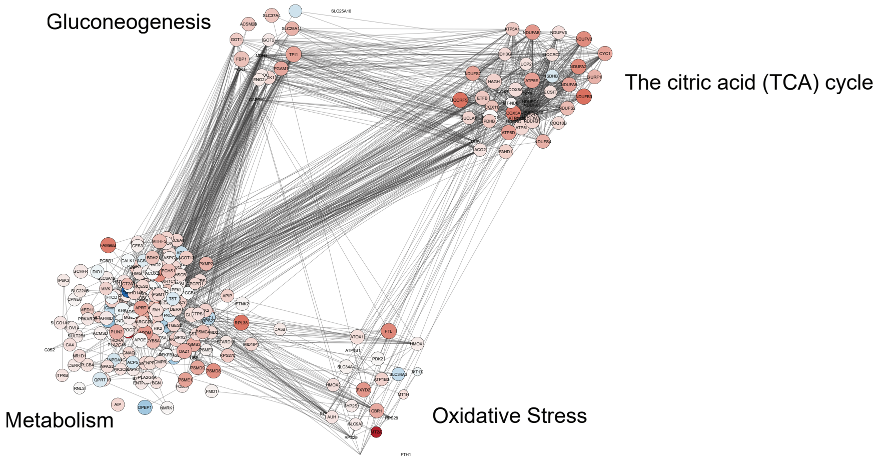

Supplemental Figure 5: Reactome Enrichment Analysis for A) Enhanced genes in DTL B) Suppressed genes in DTL C) Enhanced genes in PC D) Suppressed genes in PC E) Enhanced genes in IC F) Suppressed genes in IC

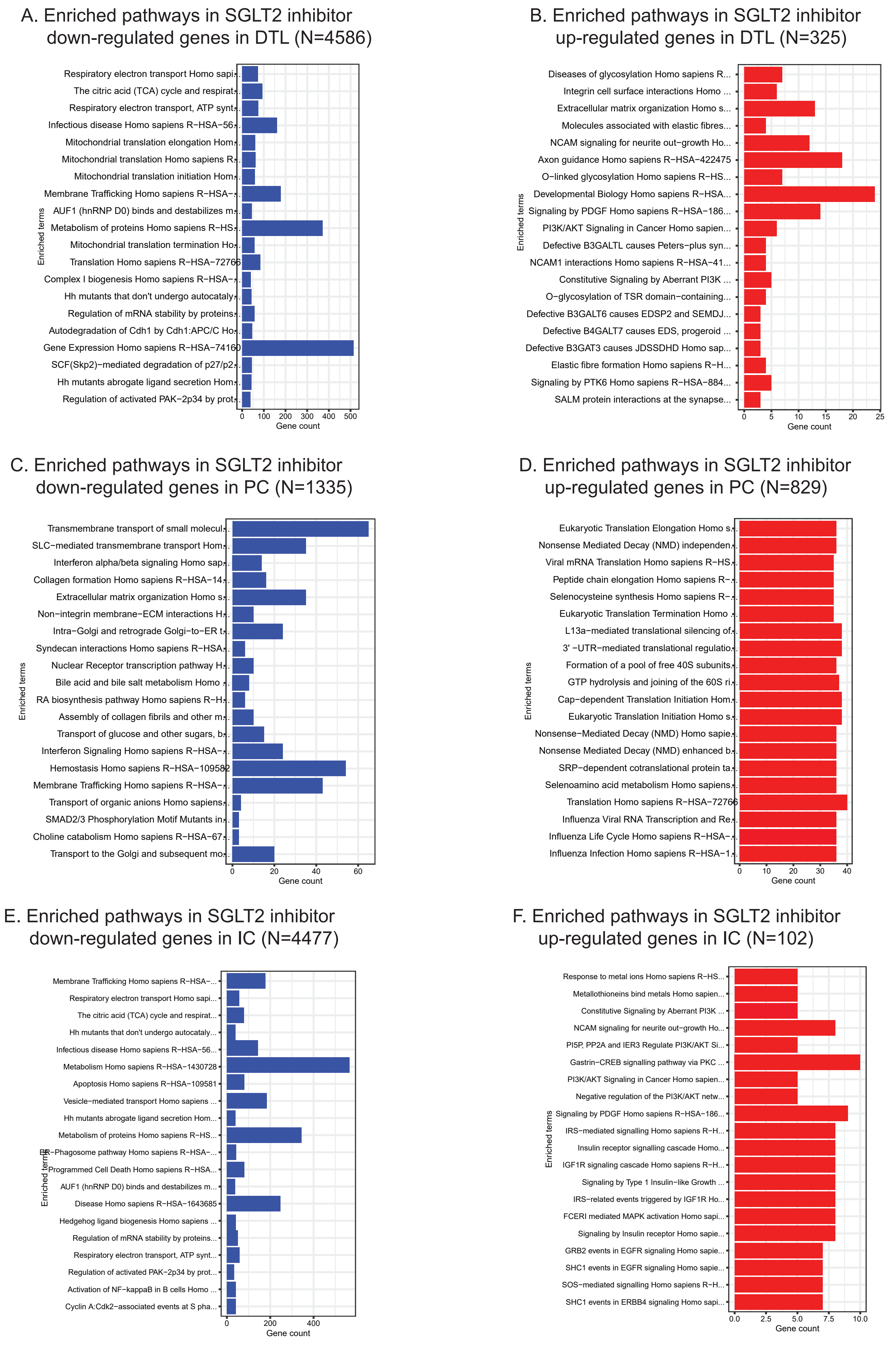

Supplemental Figure 6: Gene level alterations in human PT cells treated with SGLT2 inhibitors and db/db mouse model treated with SGLT2 inhibitors.

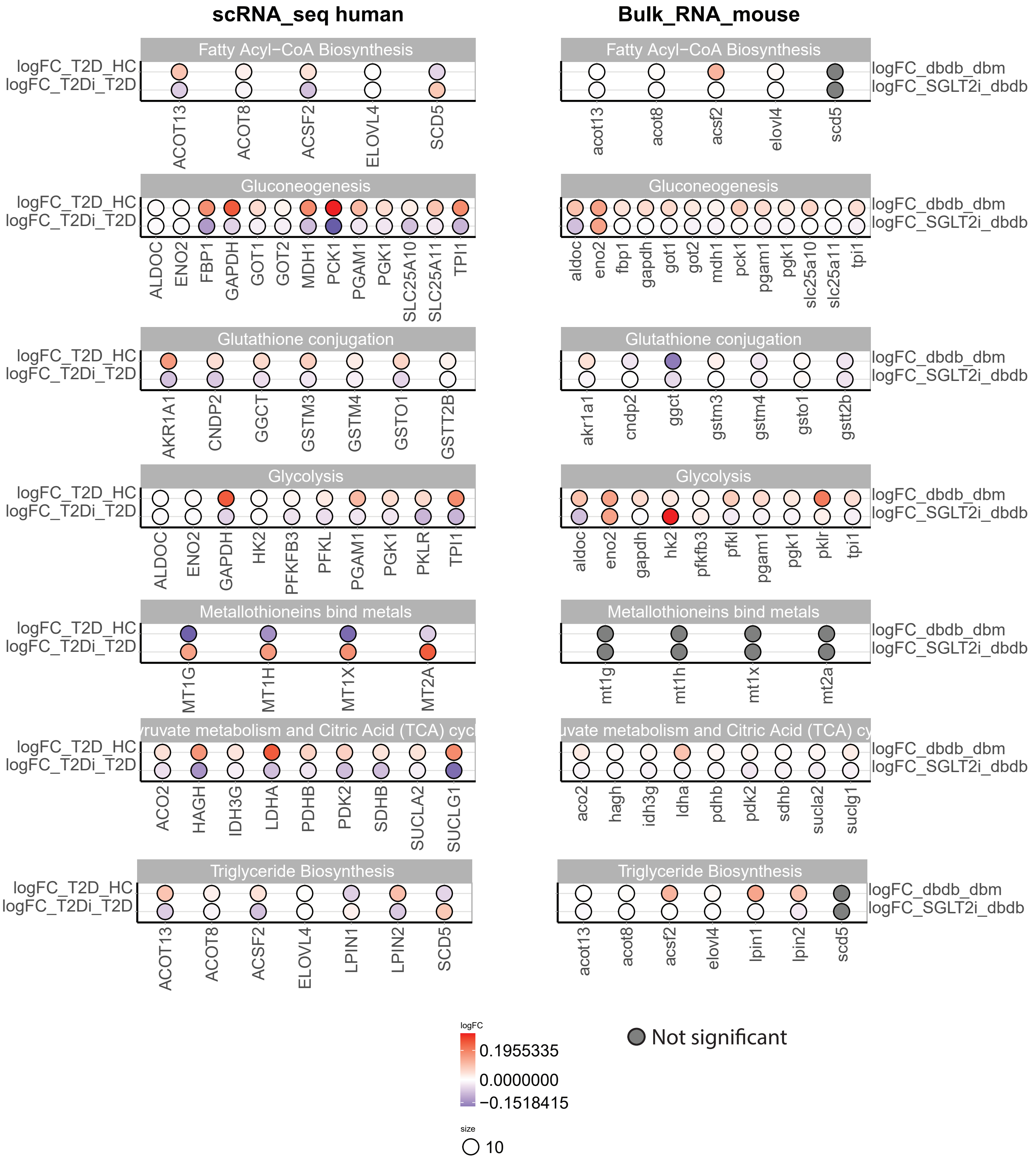

**Supplemental Table 1.** Exclusion criteria for percutaneous kidney biopsy

| Additional exclusion criteria for those undergoing kidney biopsy |
|---|
| Evidence of bleeding disorder or complications from bleeding |
| Use of aspirin, NSAIDS or other blood thinner that cannot be safely stopped for a sufficient time period before and after the biopsy so as to add no additional risk of bleeding |
| Blood urea nitrogen (BUN) > 80 gm/dL |
| INR > 1.4 |
| PTT > 35 seconds |
| Hemoglobin (Hb) < 10 mg/dL |
| Platelet count < 100,000 / µL |
| Uncontrolled or difficult to control hypertension (> 150/90 mmHg at the day of biopsy) |
| eGFR < 40 mL/min/1.73m$^2$ |
| Single kidney (either by history, documented by prior imaging or ultrasound performed prior to the biopsy) |
| > 2 cm discrepancy between left and right kidney sizes based on largest longitudinal diameter determined by US performed prior to the biopsy. |
| Kidney size: One or both kidneys < 9 cm |
| Hydronephrosis or other important renal ultrasound findings such as significant stone disease |
| Any evidence of a current urinary tract infection as indicated on day of biopsy |
| Clinical evidence of non-diabetic renal disease |
| Positive urine pregnancy test or pregnancy |

**Supplementary Table 2.** List of reversed genes with SGLT2i across all nephron segments.

**Proximal tubule cells**

| Genes | logFC_T2D_HC | logFC_T2Di_T2D | logFC_T2Di_HC |
|---|---|---|---|
| PPDPF | 0.200158443 | 0.105240043 | 0.305398486 |
| RPS10 | 0.194111658 | 0.026357534 | 0.220469192 |
| COX7C | 0.168970379 | -0.029850182 | 0.139120198 |
| RPL14 | 0.184299157 | 0.024096624 | 0.208395781 |
| NDUFA1 | 0.178598052 | -0.050617122 | 0.12798093 |
| RPS18 | 0.158088476 | 0.030926203 | 0.189014678 |
| RPS4X | 0.173209826 | 0.079811127 | 0.253020953 |
| CXCL14 | 0.211273345 | -0.032422785 | 0.17885056 |
| LDHA | 0.157481245 | -0.052618996 | 0.104862249 |
| NKAIN4 | 0.092885955 | -0.058462977 | 0.034422978 |
| RPS27 | 0.160541684 | -0.041229716 | 0.119311968 |
| RPL41 | 0.139371181 | 0.016576743 | 0.155947924 |
| RPL8 | 0.153917418 | 0.020251917 | 0.174169335 |
| GAPDH | 0.158302301 | -0.036743685 | 0.121558616 |
| PCK1 | 0.195533465 | -0.15184152 | 0.043691945 |
| RACK1 | 0.160077096 | 0.021929849 | 0.182006945 |
| RPS23 | 0.146340259 | 0.024618361 | 0.17095862 |
| NDUFB2 | 0.162022012 | -0.055765416 | 0.106256596 |
| RPS12 | 0.150370117 | 0.03503427 | 0.185404387 |
| COX7A2 | 0.161515191 | -0.023076197 | 0.138438994 |
| COL14A1 | 0.067940642 | -0.070194189 | -0.002253547 |
| RPL15 | 0.155484558 | 0.036299211 | 0.191783769 |
| RPS21 | 0.156789017 | -0.027279752 | 0.129509265 |
| EEF1A1 | 0.113489548 | 0.01530541 | 0.128794958 |
| SERF2 | 0.138137996 | -0.029127716 | 0.10901028 |
| RPL11 | 0.149494013 | 0.025671649 | 0.175165662 |
| RPS8 | 0.154848685 | 0.026672682 | 0.181521368 |
| RPS19 | 0.158357387 | 0.051118711 | 0.209476097 |
| CYBA | 0.169528376 | -0.037378933 | 0.132149443 |
| RPS3 | 0.16226434 | 0.022974579 | 0.185238919 |
| RPS28 | 0.129733677 | -0.01734016 | 0.112393517 |
| SMOC2 | 0.052025621 | -0.052168163 | -0.000142542 |
| FTH1 | 0.135783542 | -0.023511148 | 0.112272394 |
| PTGR1 | 0.144200756 | -0.050901698 | 0.093299057 |
| FABP1 | 0.201618986 | -0.235717071 | -0.034098085 |
| AKR1B1 | 0.093870511 | -0.062129497 | 0.031741014 |
| RPS9 | 0.136572473 | 0.020720706 | 0.15729318 |
| MRPS6 | 0.123083951 | -0.08271804 | 0.040365911 |
| UQCR11 | 0.148205651 | -0.044744991 | 0.10346066 |
| GATD3A | 0.068009739 | -0.014291971 | 0.053717768 |
| RPL29 | 0.138881047 | 0.025536325 | 0.164417372 |
| BX284668.5 | 0.075838487 | -0.077607893 | -0.001769406 |
| RPS6 | 0.132467177 | 0.035055591 | 0.167522767 |
| RPL10 | 0.134003064 | 0.035105677 | 0.16910874 |

| | | | |
|---|---|---|---|
| RANBP3L | 0.057094116 | -0.057459718 | -0.000365602 |
| ATP6V1F | 0.142213609 | -0.023104549 | 0.11910906 |
| RPS3A | 0.134670423 | 0.025203948 | 0.159874371 |
| G0S2 | 0.141675979 | -0.068273965 | 0.073402014 |
| COX6B1 | 0.140305352 | -0.027370006 | 0.112935346 |
| MGST3 | 0.144024085 | 0.037059285 | 0.18108337 |
| RPL28 | 0.134592144 | 0.048805368 | 0.183397512 |
| RPS27A | 0.123729907 | 0.018492877 | 0.142222784 |
| DBI | 0.146376254 | -0.050024016 | 0.096352237 |
| COL12A1 | 0.062370692 | -0.050806575 | 0.011564117 |
| SLIRP | 0.130659275 | -0.04100689 | 0.089652386 |
| ATP5IF1 | 0.140559434 | -0.036272848 | 0.104286585 |
| ACTG1 | 0.150307324 | 0.052281465 | 0.202588789 |
| RPL17 | 0.132618078 | 0.028311057 | 0.160929135 |
| RPS29 | 0.133938878 | -0.06407035 | 0.069868528 |
| RPL10A | 0.14363462 | 0.027756528 | 0.171391149 |
| TMEM176A | 0.140631308 | 0.030960209 | 0.171591517 |
| NDUFC1 | 0.13636666 | -0.035990612 | 0.100376048 |
| COX14 | 0.129832364 | -0.070543326 | 0.059289039 |
| CTSH | 0.120801217 | -0.067525758 | 0.053275459 |
| TMED6 | 0.050181807 | -0.040301005 | 0.009880801 |
| PLAAT4 | 0.094974787 | -0.036274407 | 0.05870038 |
| HIST1H4C | 0.13959432 | -0.037160709 | 0.102433611 |
| VEGFB | 0.105657828 | -0.042686227 | 0.062971601 |
| IER3 | 0.064081123 | -0.044602869 | 0.019478254 |
| PEBP1 | 0.106515344 | -0.06771444 | 0.038800904 |
| PLAAT3 | 0.124552359 | -0.064099143 | 0.060453216 |
| BTF3 | 0.136048383 | 0.025584222 | 0.161632606 |
| PPIA | 0.126633621 | 0.021619488 | 0.148253109 |
| UQCRQ | 0.133423601 | -0.052962853 | 0.080460749 |
| COPS9 | 0.125655959 | -0.049689121 | 0.075966838 |
| SNRPN | 0.115090911 | -0.033130397 | 0.081960513 |
| PCBD1 | 0.12898993 | -0.041558235 | 0.087431695 |
| NPM1 | 0.13408398 | 0.040917306 | 0.175001287 |
| RPS14 | 0.114011484 | 0.018264306 | 0.13227579 |
| CHCHD10 | 0.131495953 | 0.069715246 | 0.201211199 |
| RNF181 | 0.109910534 | -0.046607219 | 0.063303314 |
| RPL12 | 0.126459373 | 0.036230717 | 0.16269009 |
| SNHG5 | 0.106175409 | -0.029880396 | 0.076295014 |
| PCSK1N | 0.13666496 | -0.075664008 | 0.061000951 |
| AFP | -0.093733718 | -0.02973184 | -0.123465558 |
| KCNK5 | 0.067553402 | -0.082210839 | -0.014657437 |
| RPS2 | 0.126467647 | 0.035463887 | 0.161931534 |
| RPLP0 | 0.125897245 | 0.044711864 | 0.17060911 |
| RPL24 | 0.117242545 | 0.032196044 | 0.14943859 |
| C2 | 0.033262398 | -0.019620279 | 0.01364212 |
| LAMTOR5 | 0.125790621 | -0.037377655 | 0.088412965 |
| NEURL3 | 0.039394365 | -0.029693846 | 0.009700519 |
| NME2 | 0.127412357 | 0.031349201 | 0.158761558 |
| STK19 | 0.05937642 | -0.0312447 | 0.02813172 |

| | | | |
|---|---|---|---|
| TMSB10 | 0.153988889 | 0.154319477 | 0.308308366 |
| ELP5 | 0.065077394 | -0.040779015 | 0.024298379 |
| SNHG29 | 0.128333305 | 0.079470742 | 0.207804047 |
| UQCR10 | 0.123271477 | -0.028915742 | 0.094355734 |
| DYNLT1 | 0.116434281 | -0.020897819 | 0.095536462 |
| RPL32 | 0.108374215 | 0.023254743 | 0.131628958 |
| MRPL34 | 0.110872357 | -0.056987543 | 0.053884815 |
| NDUFA3 | 0.121345428 | -0.084385997 | 0.036959431 |
| ACAA1 | 0.127208979 | -0.075887775 | 0.051321204 |
| UBB | 0.117803542 | -0.028777255 | 0.089026286 |
| NAT8 | 0.138849239 | -0.044699551 | 0.094149688 |
| COX17 | 0.106347662 | -0.08760662 | 0.018741042 |
| SMDT1 | 0.114644124 | -0.029863986 | 0.084780137 |
| ZSWIM7 | 0.076009881 | -0.039489663 | 0.036520218 |
| COX7B | 0.120594421 | -0.032542862 | 0.088051559 |
| TMEM178A | 0.035216491 | -0.027851516 | 0.007364976 |
| TSTD1 | 0.112840819 | 0.02573272 | 0.13857354 |
| ATP5F1E | 0.110956359 | -0.056724242 | 0.054232118 |
| RNF24 | 0.04015205 | -0.039385163 | 0.000766887 |
| MDH1 | 0.115412405 | -0.054660061 | 0.060752343 |
| RPL18 | 0.116411437 | 0.028911783 | 0.14532322 |
| SLC4A11 | 0.061077186 | -0.076774947 | -0.01569776 |
| CALM1 | 0.117497613 | -0.08347711 | 0.034020503 |
| CENPX | 0.086235432 | -0.04514883 | 0.041086602 |
| TIMM10 | 0.086407995 | -0.034852536 | 0.051555459 |
| ATP5MF | 0.115869467 | -0.032701369 | 0.083168098 |
| RBP5 | 0.127423059 | -0.062247783 | 0.065175276 |
| AKR1C1 | 0.088406536 | -0.079143851 | 0.009262685 |
| ELF5 | 0.024997111 | -0.025001988 | -4.87694E-06 |
| FAM162A | 0.099022294 | -0.020773883 | 0.078248412 |
| TPI1 | 0.114173882 | -0.06212448 | 0.052049402 |
| H2AFZ | 0.104650727 | -0.034796985 | 0.069853743 |
| EEF1B2 | 0.11985968 | 0.02570334 | 0.14556302 |
| ATP1B1 | 0.13701635 | -0.044385481 | 0.092630869 |
| UQCRFS1 | 0.106302867 | -0.019937909 | 0.086364957 |
| CIAO2B | 0.105259803 | -0.025582287 | 0.079677516 |
| HAGH | 0.105796858 | -0.090350473 | 0.015446385 |
| EIF1 | 0.108353549 | 0.042361261 | 0.15071481 |
| PTMA | 0.120023678 | 0.051424136 | 0.171447814 |
| NDUFB3 | 0.108890132 | -0.020439529 | 0.088450603 |
| OCIAD2 | 0.119353638 | 0.047183008 | 0.166536646 |
| FXYD2 | 0.082182516 | -0.022078032 | 0.060104484 |
| LAMTOR1 | 0.09776873 | -0.041384852 | 0.056383878 |
| FAM107B | 0.092435438 | -0.040699285 | 0.051736153 |
| PLAAT2 | 0.039062639 | -0.037328135 | 0.001734505 |
| RPSA | 0.115627966 | 0.025139066 | 0.140767032 |
| S100A1 | 0.070675267 | -0.067583041 | 0.003092226 |
| DNMT3L | 0.051432955 | -0.050578987 | 0.000853968 |
| CLEC18C | 0.040766641 | -0.034699855 | 0.006066786 |
| SSR2 | 0.091266894 | 0.021813984 | 0.113080878 |

| | | | |
|---|---|---|---|
| TSPAN33 | 0.087782824 | -0.045433419 | 0.042349405 |
| RPL38 | 0.105976248 | -0.02034311 | 0.085633138 |
| DDT | 0.110846435 | -0.028848952 | 0.081997483 |
| ATOH8 | -0.022537668 | 0.005547495 | -0.016990172 |
| MUC13 | -0.026060903 | -0.007115012 | -0.033175915 |
| JAG1 | -0.04969998 | 0.048969866 | -0.000730114 |
| BRI3 | 0.10923079 | -0.03181065 | 0.07742014 |
| RPL18A | 0.105561714 | 0.031304457 | 0.13686617 |
| RPL39 | 0.104224041 | 0.027491603 | 0.131715644 |
| ARG2 | 0.070269223 | -0.07943105 | -0.009161827 |
| HIF1A | 0.072334231 | -0.040720467 | 0.031613765 |
| PRDX1 | 0.102775736 | -0.05537449 | 0.047401246 |
| DNPH1 | 0.108714083 | -0.035958622 | 0.072755461 |
| RGN | 0.081209282 | -0.06023695 | 0.020972333 |
| COX5A | 0.099655035 | -0.021674909 | 0.077980125 |
| SLPI | 0.038946321 | -0.040459009 | -0.001512689 |
| MIF | 0.099460816 | -0.032036998 | 0.067423818 |
| TOMM5 | 0.099277531 | -0.037633401 | 0.06164413 |
| LPIN1 | -0.038846314 | 0.015956414 | -0.0228899 |
| APOC2 | 0.056473253 | -0.059174847 | -0.002701594 |
| NDUFV2 | 0.100473269 | -0.03261189 | 0.067861379 |
| ATP5MD | 0.107932788 | -0.033394812 | 0.074537976 |
| PLCB4 | 0.025610486 | -0.020291754 | 0.005318731 |
| GLYATL1 | 0.104240301 | -0.077416099 | 0.026824202 |
| RPL13 | 0.084606706 | 0.039293381 | 0.123900088 |
| TMSB4X | 0.104832767 | 0.028247392 | 0.133080159 |
| PRAP1 | 0.111224261 | -0.113102069 | -0.001877808 |
| SLC5A3 | 0.076205428 | -0.086052927 | -0.009847499 |
| MT-ATP8 | -0.100768465 | -0.062636233 | -0.163404698 |
| TMEM14C | 0.096128637 | 0.020775835 | 0.116904472 |
| SLCO4A1 | 0.025845461 | -0.024159402 | 0.001686059 |
| CAMK2N1 | 0.09800744 | -0.047868325 | 0.050139116 |
| ELOB | 0.103787859 | -0.023986823 | 0.079801036 |
| PSMD8 | 0.085229715 | -0.020693131 | 0.064536584 |
| STXBP6 | 0.031367459 | -0.018692499 | 0.01267496 |
| SAT2 | 0.107524301 | -0.100716015 | 0.006808286 |
| MAL2 | 0.059353833 | -0.013008982 | 0.046344851 |
| FBP1 | 0.112043816 | -0.084226532 | 0.027817284 |
| LIX1 | 0.027224505 | -0.02803251 | -0.000808005 |
| REXO2 | 0.08952686 | -0.019059818 | 0.070467042 |
| SERPINF1 | 0.033330941 | -0.012823943 | 0.020506998 |
| COPE | 0.088713041 | -0.030666841 | 0.0580462 |
| B2M | 0.133186493 | 0.072690382 | 0.205876874 |
| SLC6A12 | 0.042678722 | -0.043226962 | -0.000548241 |
| NDUFA2 | 0.098565273 | -0.028367902 | 0.07019737 |
| MRPS15 | 0.077283037 | -0.039938916 | 0.037344121 |
| KHK | 0.110602997 | -0.127813964 | -0.017210967 |
| DDIT4L | 0.045370926 | -0.058833402 | -0.013462476 |
| RPS16 | 0.097728339 | 0.022603108 | 0.120331448 |
| PGGHG | 0.034901781 | -0.026701182 | 0.0082006 |

| | | | |
|---|---|---|---|
| RDH10 | 0.059658721 | -0.035087042 | 0.02457168 |
| PRDX6 | 0.099261123 | 0.081551181 | 0.180812305 |
| SUCLG1 | 0.114446507 | -0.124839552 | -0.010393045 |
| PPIB | 0.093685089 | 0.056407921 | 0.15009301 |
| CD82 | 0.029101075 | -0.021764169 | 0.007336907 |
| SLCO1A2 | 0.022120952 | -0.019430522 | 0.002690429 |
| NDUFA8 | 0.072893795 | -0.021019614 | 0.051874181 |
| MRPL57 | 0.093773532 | -0.038227 | 0.055546532 |
| HMGN3 | 0.100554513 | 0.030824369 | 0.131378882 |
| C4orf3 | 0.09781673 | -0.041532689 | 0.056284041 |
| LPIN2 | 0.0651367 | -0.044415801 | 0.020720899 |
| LGALS2 | 0.10624794 | -0.05575556 | 0.05049238 |
| HLA-DPB1 | 0.039484608 | -0.015325238 | 0.024159371 |
| CLU | 0.118928622 | 0.069904685 | 0.188833307 |
| S100A4 | 0.033918917 | -0.027330716 | 0.006588201 |
| ACAA2 | 0.095149122 | -0.030578184 | 0.064570938 |
| NOP10 | 0.088035605 | -0.01885366 | 0.069181945 |
| RPL4 | 0.096918949 | 0.022135545 | 0.119054493 |
| OAZ1 | 0.096707699 | -0.052954689 | 0.04375301 |
| CD63 | 0.112329995 | 0.088284313 | 0.200614308 |
| EDF1 | 0.097909814 | 0.049870183 | 0.147779997 |
| PLIN2 | 0.085360838 | -0.042075995 | 0.043284843 |
| ABRACL | 0.071392924 | -0.029742121 | 0.041650803 |
| APOM | 0.089026839 | -0.030131836 | 0.058895003 |
| FTL | 0.108384634 | -0.037273574 | 0.071111059 |
| NINJ1 | 0.075790892 | -0.036606307 | 0.039184586 |
| SIVA1 | 0.079804483 | -0.041914728 | 0.037889755 |
| NDUFB1 | 0.097950178 | -0.084940819 | 0.013009359 |
| DAD1 | 0.097549897 | 0.034117418 | 0.131667315 |
| AKR1A1 | 0.100536204 | -0.050227123 | 0.050309081 |
| TIMM13 | 0.089141805 | -0.036200747 | 0.052941058 |
| METTL27 | 0.035438115 | -0.014639099 | 0.020799016 |
| PET100 | 0.088146809 | -0.027780483 | 0.060366325 |
| CA2 | 0.096015258 | -0.087015836 | 0.008999422 |
| FKBP5 | 0.056035263 | -0.08019732 | -0.024162057 |
| CSRP2 | 0.063373617 | -0.022100996 | 0.041272621 |
| ECHDC2 | 0.08285488 | -0.019987927 | 0.062866953 |
| TMEM256 | 0.090571677 | -0.063849308 | 0.026722369 |
| THY1 | 0.097176912 | -0.046394902 | 0.05078201 |
| MIEN1 | 0.066282463 | -0.039737251 | 0.026545212 |
| SSNA1 | 0.073340453 | -0.036966601 | 0.036373851 |
| NDUFAB1 | 0.091000116 | -0.031996724 | 0.059003392 |
| ID1 | -0.058083091 | 0.09533506 | 0.037251969 |
| AL133351.1 | 0.030713822 | -0.041822494 | -0.011108672 |
| ARPC1B | 0.050686331 | -0.014877362 | 0.035808969 |
| FXYD4 | 0.029336667 | -0.025855476 | 0.003481192 |
| S100A11 | 0.105711137 | 0.100687791 | 0.206398928 |
| ATP5F1D | 0.090689898 | -0.03633382 | 0.054356079 |
| CYC1 | 0.075737527 | -0.025889231 | 0.049848296 |
| IGFBP5 | 0.046166113 | -0.056127049 | -0.009960936 |

| | | | |
|---|---|---|---|
| MT1X | -0.126436685 | 0.110565602 | -0.015871083 |
| ATP6V1G1 | 0.091752366 | -0.027207252 | 0.064545114 |
| MRPS26 | 0.073885344 | -0.024858905 | 0.049026439 |
| CLIC1 | 0.089586939 | 0.074054988 | 0.163641927 |
| AP2S1 | 0.078990056 | -0.035994303 | 0.042995754 |
| BAG1 | 0.078471675 | -0.041339077 | 0.037132598 |
| CBR1 | 0.076814347 | -0.031483256 | 0.045331091 |
| KRT18 | 0.086621415 | 0.074914337 | 0.161535752 |
| IGBP1 | 0.06201832 | -0.016147744 | 0.045870576 |
| ATP5ME | 0.092831237 | -0.081295768 | 0.011535469 |
| CLEC18A | 0.033780897 | -0.030956076 | 0.002824822 |
| HLA-DRB1 | 0.067084696 | 0.020404968 | 0.087489664 |
| MCCD1 | 0.06108891 | -0.052326708 | 0.008762202 |
| GAL3ST1 | 0.038364099 | -0.026337277 | 0.012026823 |
| MRC2 | -0.014604066 | 0.005089842 | -0.009514224 |
| LAMTOR2 | 0.078134428 | -0.025100352 | 0.053034076 |
| HSPA2 | 0.03716182 | -0.027052226 | 0.010109594 |
| PHB | 0.088014667 | -0.055327116 | 0.03268755 |
| USP2 | 0.056239603 | -0.068842472 | -0.012602869 |
| FCGBP | 0.027920238 | -0.025863312 | 0.002056926 |
| NDUFC2 | 0.090899586 | 0.024020439 | 0.114920025 |
| PSMD9 | 0.067918915 | -0.019384048 | 0.048534867 |
| HNRNPA1 | 0.092567136 | 0.051681886 | 0.144249022 |
| GET3 | 0.06625033 | -0.026085836 | 0.040164494 |
| CYB5A | 0.086049297 | -0.050872505 | 0.035176792 |
| MRPS36 | 0.081173127 | -0.032973919 | 0.048199208 |
| TXNDC17 | 0.087904443 | -0.051967941 | 0.035936502 |
| SEC11A | 0.079718301 | 0.031789379 | 0.11150768 |
| RNF186 | 0.048825359 | -0.059166461 | -0.010341102 |
| PIM3 | 0.047640813 | -0.011304229 | 0.036336584 |
| ROMO1 | 0.084086207 | -0.045302188 | 0.038784019 |
| JUND | 0.083823079 | 0.068751555 | 0.152574635 |
| PSME1 | 0.074133569 | -0.021134528 | 0.052999041 |
| PGAM1 | 0.068740313 | -0.021177011 | 0.047563302 |
| AP1S1 | 0.063132071 | 0.033904868 | 0.097036938 |
| PRSS23 | 0.034280995 | -0.015871201 | 0.018409795 |
| PFKP | 0.043982964 | 0.021107637 | 0.065090602 |
| TXNL4A | 0.0646525 | -0.018402097 | 0.046250403 |
| SLC25A5 | 0.092836676 | -0.052155883 | 0.040680793 |
| DBP | 0.046173771 | -0.011254723 | 0.034919048 |
| SNRPG | 0.084173304 | -0.024411012 | 0.059762292 |
| NAA38 | 0.078318123 | -0.03816583 | 0.040152293 |
| PAIP2 | 0.068653849 | -0.032768088 | 0.035885761 |
| HLA-C | 0.085459188 | 0.079394098 | 0.164853286 |
| ALDH1A3 | 0.021237176 | -0.016903105 | 0.004334072 |
| NDUFS7 | 0.078649685 | -0.026373581 | 0.052276104 |
| SASH1 | 0.036535165 | -0.02259498 | 0.013940185 |
| FIS1 | 0.085530222 | -0.026568006 | 0.058962215 |
| SSR4 | 0.078475087 | 0.045427506 | 0.123902592 |
| MT1G | -0.141865269 | 0.093561125 | -0.048304144 |

| | | | |
|---|---|---|---|
| DECR2 | 0.057821402 | -0.036476281 | 0.021345121 |
| TMEM208 | 0.061985723 | 0.015460288 | 0.077446011 |
| EEF1D | 0.08809226 | 0.036818646 | 0.124910907 |
| BORCS7 | 0.060706883 | -0.029136295 | 0.031570588 |
| ANAPC11 | 0.074165135 | -0.023303432 | 0.050861703 |
| ID4 | -0.057926884 | 0.090920705 | 0.032993821 |
| BIN1 | 0.071655537 | -0.048352755 | 0.023302781 |
| SMAD9 | -0.019632069 | 0.009877603 | -0.009754466 |
| SLC5A10 | 0.036461215 | -0.026085212 | 0.010376003 |
| PRXL2A | 0.067481934 | -0.028478467 | 0.039003467 |
| EIF3H | 0.074379766 | 0.020689652 | 0.095069418 |
| SLC37A4 | 0.062539302 | -0.036427114 | 0.026112188 |
| GRHPR | 0.080314452 | -0.023696014 | 0.056618439 |
| PKM | 0.073602091 | 0.036183337 | 0.109785429 |
| PNKD | 0.073033819 | -0.027124301 | 0.045909517 |
| EEF1G | 0.083267203 | 0.051360129 | 0.134627332 |
| NTM | 0.016633568 | -0.017281027 | -0.000647458 |
| ATP1B3 | 0.036209578 | -0.020842084 | 0.015367494 |
| LY6E | 0.067068451 | 0.035413981 | 0.102482432 |
| TMEM150A | 0.05003807 | -0.029685916 | 0.020352154 |
| LINC02754 | 0.057723879 | -0.066483518 | -0.00875964 |
| RAN | 0.07413637 | 0.021783925 | 0.095920295 |
| NDUFB9 | 0.078206387 | 0.026616897 | 0.104823284 |
| SCCPDH | 0.051618124 | -0.034487501 | 0.017130623 |
| DDIT4 | 0.029648721 | -0.029459022 | 0.000189699 |
| HIGD2A | 0.075437096 | -0.026759994 | 0.048677102 |
| SLC38A2 | 0.051316274 | -0.024638239 | 0.026678034 |
| POLR1D | 0.065207055 | 0.018495085 | 0.08370214 |
| RPS26 | 0.086003389 | 0.03598241 | 0.121985799 |
| CTSK | 0.016883075 | -0.014517549 | 0.002365527 |
| IER3IP1 | 0.068510612 | -0.031069088 | 0.037441523 |
| GLRX | 0.083435287 | -0.075107663 | 0.008327624 |
| F13A1 | -0.026560532 | -0.016991417 | -0.043551949 |
| SLC25A3 | 0.082577256 | -0.031763086 | 0.05081417 |
| PSMB3 | 0.06879348 | -0.023620792 | 0.045172687 |
| S100A13 | 0.081671904 | 0.022846507 | 0.104518411 |
| PSME2 | 0.06002364 | 0.024270513 | 0.084294153 |
| CPNE6 | 0.029345637 | -0.034805799 | -0.005460163 |
| SLC22A18AS | 0.03850476 | -0.034420234 | 0.004084526 |
| MTHFS | 0.071567332 | -0.03997782 | 0.031589512 |
| PPP1R14B | 0.065616193 | 0.042610222 | 0.108226415 |
| QPRT | 0.077983406 | -0.103852374 | -0.025868968 |
| RPS4Y1 | 0.068992085 | -0.17542378 | -0.106431694 |
| LINC01874 | 0.054624205 | -0.067848597 | -0.013224392 |
| GNG11 | 0.042740677 | -0.031277406 | 0.011463271 |
| MSRA | 0.080468098 | -0.070004014 | 0.010464083 |
| SLC25A11 | 0.060379483 | -0.02055862 | 0.039820863 |
| KDELR1 | 0.067457996 | 0.04085471 | 0.108312707 |
| CDKN1C | 0.053476364 | -0.019628681 | 0.033847683 |
| MRPS25 | 0.052242929 | -0.021432829 | 0.0308101 |

| | | | |
|---|---|---|---|
| SURF1 | 0.060929731 | -0.020926546 | 0.040003186 |
| CACNA2D1 | 0.027862527 | -0.022895859 | 0.004966668 |
| AEBP1 | -0.023880915 | 0.012425549 | -0.011455366 |
| ERI3 | 0.054777892 | -0.034650817 | 0.020127075 |
| DECR1 | 0.06439977 | -0.016381271 | 0.048018498 |
| NENF | 0.069717369 | 0.018060887 | 0.087778255 |
| KRT8 | 0.068787422 | 0.0801405 | 0.148927922 |
| FUNDC2 | 0.054912652 | -0.015234521 | 0.03967813 |
| RPL9 | 0.0705508 | 0.052149765 | 0.122700564 |
| ID3 | -0.033957843 | 0.040891642 | 0.006933798 |
| AF064858.1 | 0.010382215 | -0.009732808 | 0.000649407 |
| SLC39A5 | 0.050662736 | -0.025426893 | 0.025235843 |
| BNIP3 | 0.075706961 | -0.038341377 | 0.037365584 |
| HDHD3 | 0.058055945 | -0.036772035 | 0.02128391 |
| CAPN6 | 0.016426471 | -0.016099474 | 0.000326996 |
| GC | -0.013801666 | 0.005585628 | -0.008216038 |
| HINT2 | 0.071288678 | -0.03792824 | 0.033360438 |
| NDUFS2 | 0.063586976 | -0.030570061 | 0.033016915 |
| HOXB7 | 0.039097138 | -0.013955169 | 0.02514197 |
| APRT | 0.074572324 | -0.020994535 | 0.053577789 |
| TSPAN1 | 0.077186404 | 0.089309795 | 0.166496199 |
| HIGD1A | 0.060593374 | 0.01674303 | 0.077336404 |
| ZFAS1 | 0.071218223 | 0.058227576 | 0.129445798 |
| AKR7A3 | 0.074980931 | -0.053121229 | 0.021859702 |
| AFMID | 0.05319867 | -0.05289458 | 0.00030409 |
| CALML4 | 0.064288544 | -0.02187492 | 0.042413624 |
| VKORC1 | 0.058485191 | 0.022997157 | 0.081482348 |
| FAHD1 | 0.063767677 | -0.046796984 | 0.016970693 |
| SPARCL1 | 0.030540885 | -0.03208143 | -0.001540545 |
| MICOS13 | 0.070738972 | 0.025142809 | 0.095881781 |
| NDUFAF8 | 0.06545945 | -0.039657118 | 0.025802332 |
| PSMC4 | 0.05260754 | -0.02121178 | 0.03139576 |
| SEC61G | 0.07354754 | 0.024750108 | 0.098297649 |
| CNNM1 | 0.015640163 | -0.012427077 | 0.003213086 |
| ZNHIT1 | 0.074529716 | -0.025499754 | 0.049029962 |
| PDZK1 | 0.078457827 | 0.035153663 | 0.11361149 |
| ECHS1 | 0.08115625 | -0.050781824 | 0.030374426 |
| GSTM3 | 0.047241182 | -0.019948629 | 0.027292554 |
| UXT | 0.064357347 | -0.018030885 | 0.046326462 |
| H3F3B | 0.084903393 | 0.102756964 | 0.187660357 |
| MPST | 0.064435054 | -0.03230572 | 0.032129333 |
| AGFG2 | 0.018570144 | -0.020801869 | -0.002231725 |
| HAO2 | 0.070159883 | -0.073914197 | -0.003754314 |
| AIG1 | 0.062554139 | 0.021809556 | 0.084363696 |
| CHCHD5 | 0.059354878 | -0.018377283 | 0.040977595 |
| GALK1 | 0.058111129 | -0.070511829 | -0.0124007 |
| BDNF | 0.010135725 | -0.009472164 | 0.000663561 |
| UMOD | -0.052868674 | 0.055963317 | 0.003094643 |
| UROS | 0.05218628 | -0.03627147 | 0.015914809 |
| OCEL1 | 0.045133647 | -0.030885681 | 0.014247966 |

| | | | |
|---|---|---|---|
| CCND1 | -0.054658771 | 0.030097325 | -0.024561446 |
| HPCAL1 | 0.047592173 | -0.014643319 | 0.032948854 |
| TSFM | 0.043562906 | -0.014667833 | 0.028895073 |
| SLC51B | 0.038215186 | -0.020547766 | 0.01766742 |
| DPEP1 | 0.077795561 | -0.131096445 | -0.053300885 |
| AQP2 | 0.022837447 | -0.02271722 | 0.000120227 |
| NPC2 | 0.068608777 | 0.034319456 | 0.102928233 |
| KRTCAP2 | 0.06170806 | 0.038106219 | 0.099814279 |
| HOMER3 | 0.047011773 | -0.025011059 | 0.022000715 |
| CARHSP1 | 0.06136493 | 0.02131093 | 0.082675861 |
| AKR7A2 | 0.070175983 | -0.033411639 | 0.036764344 |
| BLOC1S1 | 0.073534434 | -0.035292733 | 0.038241701 |
| PCP4 | 0.026284313 | -0.020154658 | 0.006129656 |
| MTLN | 0.059032072 | -0.068065701 | -0.009033629 |
| MT-ND4 | 0.04795272 | 0.017268886 | 0.065221606 |
| ARRDC3 | -0.057792622 | 0.041782669 | -0.016009952 |
| RPS19BP1 | 0.06414638 | -0.020177036 | 0.043969345 |
| FAAP20 | 0.0551227 | -0.026539757 | 0.028582943 |
| AOC1 | 0.058192985 | -0.066281786 | -0.008088801 |
| RAB13 | 0.058606085 | 0.065224574 | 0.123830659 |
| LSM4 | 0.06283266 | -0.022253925 | 0.040578734 |
| CAPN3 | 0.044489121 | -0.075056404 | -0.030567283 |
| IMPA2 | 0.071122831 | 0.025234709 | 0.09635754 |
| NDUFS4 | 0.057176219 | -0.016229557 | 0.040946662 |
| RPA3 | 0.058450822 | -0.026780915 | 0.031669907 |
| WDR45 | 0.04006684 | -0.013650424 | 0.026416416 |
| SLC4A7 | 0.024483141 | -0.009096465 | 0.015386676 |
| CAPG | 0.052208227 | 0.020130301 | 0.072338528 |
| LMAN2 | 0.049234256 | 0.016767723 | 0.066001978 |
| PRELID1 | 0.071238295 | -0.049919359 | 0.021318937 |
| C9orf16 | 0.051979559 | -0.02836822 | 0.023611339 |
| FBXO17 | 0.04003588 | -0.011836836 | 0.028199044 |
| RNF187 | 0.049744233 | -0.014339841 | 0.035404392 |
| PFDN2 | 0.057875435 | 0.017396884 | 0.07527232 |
| NR2F6 | 0.044894403 | -0.013889278 | 0.031005125 |
| MGLL | 0.046089068 | -0.046473903 | -0.000384835 |
| PTMS | 0.071748979 | -0.028476249 | 0.043272729 |
| SPCS1 | 0.067841115 | 0.030536236 | 0.098377351 |
| ACOT13 | 0.05917998 | -0.040832901 | 0.018347079 |
| PSMB8 | 0.035371636 | 0.015528059 | 0.050899695 |
| PRKAR2B | 0.016126798 | -0.014176928 | 0.00194987 |
| MCUR1 | 0.054284669 | -0.034338073 | 0.019946596 |
| DYNLRB1 | 0.069960286 | -0.04458919 | 0.025371096 |
| ING2 | 0.027978255 | -0.018952775 | 0.00902548 |
| SCG5 | 0.00998102 | -0.007760357 | 0.002220663 |
| LINC02334 | 0.017906947 | -0.018796483 | -0.000889536 |
| SMIM4 | 0.049735642 | -0.054249064 | -0.004513422 |
| HIST1H1C | 0.048599765 | -0.018256329 | 0.030343435 |
| ETS2 | 0.028941488 | -0.020877812 | 0.008063676 |
| LGALS3 | 0.048096691 | -0.022032696 | 0.026063995 |

| | | | |
|---|---|---|---|
| ATOX1 | 0.074489254 | -0.068045698 | 0.006443556 |
| COA1 | 0.040370005 | -0.015332535 | 0.02503747 |
| MRPL12 | 0.051964211 | -0.023252687 | 0.028711525 |
| BANF1 | 0.055126856 | 0.02581586 | 0.080942716 |
| ABCA4 | 0.010485412 | -0.011054708 | -0.000569295 |
| SRI | 0.049747866 | 0.026091787 | 0.075839652 |
| ATP6V0E2 | 0.052553471 | -0.036988486 | 0.015564985 |
| ETFB | 0.071553667 | -0.048077452 | 0.023476215 |
| CREB5 | 0.03209158 | 0.025444044 | 0.057535624 |
| OSTC | 0.053268227 | 0.047655959 | 0.100924186 |
| CLTB | 0.054832731 | -0.035341872 | 0.019490859 |
| ARL2 | 0.064042175 | -0.02700056 | 0.037041615 |
| H2AFJ | 0.057147456 | 0.035996922 | 0.093144378 |
| SLC47A2 | 0.033064536 | -0.018108739 | 0.014955797 |
| RTCB | 0.042887451 | -0.012857934 | 0.030029517 |
| TOMM20 | 0.065503886 | 0.019636622 | 0.085140508 |
| PPP1R7 | 0.039180185 | -0.011526633 | 0.027653553 |
| SLC34A1 | 0.051645768 | -0.05563648 | -0.003990712 |
| NR1D1 | 0.017371669 | -0.007340853 | 0.010030817 |
| EHF | 0.019729719 | -0.019236439 | 0.00049328 |
| FKBP1A | 0.054633748 | 0.01661119 | 0.071244938 |
| EIF5A | 0.060670036 | -0.019245541 | 0.041424495 |
| C9orf78 | 0.044690803 | -0.019881804 | 0.024808999 |
| MT1H | -0.093214317 | 0.099441117 | 0.006226801 |
| AC004889.1 | -0.025354585 | 0.023238622 | -0.002115963 |
| DDOST | 0.038078592 | 0.02351253 | 0.061591122 |
| RPL27A | 0.066542767 | 0.049515539 | 0.116058306 |
| HSD17B14 | 0.055932026 | -0.051886432 | 0.004045594 |
| RGS3 | 0.024647662 | -0.015638062 | 0.0090096 |
| YPEL3 | 0.047187448 | -0.013874866 | 0.033312582 |
| NIPSNAP1 | 0.05020232 | -0.026325001 | 0.023877319 |
| SDHAF3 | 0.043962407 | -0.014553185 | 0.029409222 |
| PXMP2 | 0.060379917 | -0.022656889 | 0.037723028 |
| PLCXD2 | 0.041951128 | -0.029480296 | 0.012470832 |
| COX11 | 0.044226654 | -0.015997923 | 0.028228731 |
| CLEC18B | 0.026453425 | -0.017921805 | 0.008531619 |
| HEPHL1 | 0.00677639 | -0.00677639 | 0 |
| PRMT1 | 0.038904446 | 0.015247368 | 0.054151814 |
| QDPR | 0.066994187 | -0.071049772 | -0.004055585 |
| PDHB | 0.044157076 | -0.02185121 | 0.022305866 |
| BBOX1 | 0.075030592 | -0.086385086 | -0.011354494 |
| MRPS34 | 0.056111341 | -0.018510222 | 0.037601119 |
| DEFB1 | -0.071228699 | 0.230119748 | 0.158891049 |
| EPSTI1 | 0.025237829 | -0.01328338 | 0.011954449 |
| UNC13C | 0.008986347 | -0.008755927 | 0.00023042 |
| TNFRSF12A | 0.042656231 | 0.031032888 | 0.07368912 |
| RASSF8 | 0.034965541 | -0.024249839 | 0.010715702 |
| FAM174C | 0.046877243 | -0.016821913 | 0.030055329 |
| MED11 | 0.036621385 | -0.011378138 | 0.025243248 |
| PTCHD4 | 0.007981409 | -0.008195702 | -0.000214294 |

| | | | |
|---|---|---|---|
| RNF144B | -0.02969339 | 0.05077219 | 0.021078799 |
| CRYBB3 | 0.02382588 | -0.020700229 | 0.003125651 |
| AC005726.1 | 0.026846326 | -0.024345326 | 0.002501 |
| RER1 | 0.045426808 | 0.025718854 | 0.071145662 |
| SDF2 | 0.035778674 | 0.015723039 | 0.051501713 |
| CFI | 0.050024191 | 0.022698487 | 0.072722678 |
| POMC | -0.028030761 | 0.049956021 | 0.02192526 |
| MFSD10 | 0.034310765 | 0.018142318 | 0.052453083 |
| ABCG1 | 0.013479487 | -0.015167994 | -0.001688507 |
| FDX1 | 0.057392349 | -0.021431834 | 0.035960516 |
| AQP3 | 0.028729163 | -0.020688987 | 0.008040176 |
| TBCB | 0.043596308 | 0.015943561 | 0.059539869 |
| HMGN1 | 0.064990902 | 0.034636938 | 0.09962784 |
| PLXNA4 | 0.009199034 | -0.009552394 | -0.00035336 |
| C1orf43 | 0.055461572 | -0.026421731 | 0.029039842 |
| SHB | 0.027567144 | -0.013812622 | 0.013754522 |
| SYT10 | 0.009255398 | -0.009028262 | 0.000227136 |
| SMIM19 | 0.051127902 | -0.022166833 | 0.028961069 |
| NPAS2 | 0.019874629 | -0.013119433 | 0.006755196 |
| APOC1 | 0.012416189 | -0.011577146 | 0.000839044 |
| GRPEL1 | 0.043061711 | -0.029437446 | 0.013624265 |
| BCL6 | 0.016724721 | -0.013653648 | 0.003071073 |
| TP53I3 | 0.038376706 | 0.019730132 | 0.058106838 |
| SPCS2 | 0.054346602 | 0.025588287 | 0.079934889 |
| GGACT | -0.045901878 | -0.055132719 | -0.101034597 |
| HSBP1 | 0.061135371 | 0.031363056 | 0.092498427 |
| SCD5 | -0.033714608 | 0.05546244 | 0.021747832 |
| ARL6IP4 | 0.063190618 | -0.032014184 | 0.031176435 |
| PPIC | 0.03265355 | 0.022495964 | 0.055149515 |
| UCP2 | 0.0112549 | -0.006629524 | 0.004625375 |
| PERP | 0.048359933 | 0.025034921 | 0.073394854 |
| NOP53 | 0.058816387 | 0.030046003 | 0.08886239 |
| EMP1 | -0.015698342 | 0.01255094 | -0.003147402 |
| ARPC5L | 0.03310889 | -0.013663427 | 0.019445463 |
| SUGCT | 0.038248194 | -0.032416146 | 0.005832048 |
| CTPS1 | 0.021083357 | -0.015637149 | 0.005446207 |
| SLC2A13 | 0.028992305 | -0.019825057 | 0.009167248 |
| PIH1D1 | 0.035295158 | -0.015525824 | 0.019769334 |
| AGT | 0.026674964 | 0.011027228 | 0.037702192 |
| IGFBP7 | 0.090726359 | 0.033443694 | 0.124170052 |
| RNASEH2C | 0.053154464 | -0.020384257 | 0.032770208 |
| PACSIN2 | 0.034060305 | -0.035133255 | -0.00107295 |
| KRT10 | 0.057586695 | 0.030124454 | 0.087711149 |
| LAPTM4B | 0.038102621 | 0.032485324 | 0.070587945 |
| TSC22D4 | 0.026162693 | -0.016845807 | 0.009316886 |
| PRDX4 | 0.031414846 | 0.023088792 | 0.054503638 |
| CTTNBP2 | 0.025875232 | -0.015577447 | 0.010297786 |
| CALR | 0.060167481 | 0.047920478 | 0.108087959 |
| ATP5MPL | 0.06369764 | -0.025169201 | 0.038528439 |
| PDK2 | 0.046647775 | -0.054062405 | -0.00741463 |

| | | | |
|---|---|---|---|
| PPP1R16A | 0.061259908 | -0.033676417 | 0.027583491 |
| PAPPA | 0.020842384 | -0.007093598 | 0.013748786 |
| PEPD | 0.069567069 | -0.100569482 | -0.031002413 |
| SNRPE | 0.053005969 | 0.022260037 | 0.075266006 |
| HABP2 | 0.016228859 | -0.016811134 | -0.000582274 |
| COX8A | 0.06284911 | -0.054398136 | 0.008450974 |
| MID1IP1 | 0.029856879 | -0.016487131 | 0.013369748 |
| PKLR | 0.037031862 | -0.060736937 | -0.023705075 |
| GCHFR | 0.057086188 | -0.055089213 | 0.001996975 |
| ACYP2 | 0.025608287 | -0.014444992 | 0.011163294 |
| TMED9 | 0.040130911 | 0.023561289 | 0.0636922 |
| KLB | 0.008865093 | -0.008761475 | 0.000103618 |
| HMGCL | 0.047880058 | -0.041339246 | 0.006540812 |
| YBX3 | 0.039823285 | 0.02626987 | 0.066093155 |
| DAO | 0.04476621 | -0.048165681 | -0.003399471 |
| PDCD6 | 0.046396444 | 0.016627674 | 0.063024117 |
| RRBP1 | 0.040258953 | 0.041336694 | 0.081595647 |
| TRIM6 | -0.027392244 | 0.024040462 | -0.003351781 |
| CLDN4 | 0.046856653 | 0.049680858 | 0.09653751 |
| ERMP1 | 0.023620057 | -0.023489353 | 0.000130703 |
| TMEM238 | 0.041297463 | 0.016994181 | 0.058291644 |
| SLC12A1 | -0.042167323 | 0.066713972 | 0.024546649 |
| GRAMD2B | 0.023646 | -0.015825047 | 0.007820952 |
| SNRPD1 | 0.041068322 | 0.027105222 | 0.068173544 |
| SLC5A2 | 0.040367733 | -0.066050916 | -0.025683183 |
| CENPS | 0.033226622 | -0.01950149 | 0.013725132 |
| CLDN1 | 0.022074036 | -0.007878121 | 0.014195914 |
| SLC6A6 | 0.019867685 | -0.013242835 | 0.00662485 |
| PQBP1 | 0.031954776 | -0.011578249 | 0.020376527 |
| FOXC1 | -0.031667907 | 0.060612464 | 0.028944557 |
| ENPP1 | 0.029300323 | -0.020302076 | 0.008998247 |
| RNF212 | 0.015515822 | -0.008747837 | 0.006767985 |
| RGS5 | 0.018850838 | -0.012767173 | 0.006083666 |
| BTF3L4 | 0.038711918 | 0.012844802 | 0.05155672 |
| AL391883.1 | 0.018618561 | -0.010838716 | 0.007779845 |
| ABHD17A | 0.036548611 | -0.015795345 | 0.020753266 |
| ASPHD2 | 0.009541345 | -0.011505036 | -0.00196369 |
| SNRPB | 0.041680457 | 0.021636967 | 0.063317424 |
| SUB1 | 0.060190566 | 0.033546209 | 0.093736775 |
| GGCT | 0.037214214 | -0.026389365 | 0.010824849 |
| TCEAL4 | 0.047130126 | 0.03311446 | 0.080244586 |
| BGN | -0.017633348 | 0.022063788 | 0.004430439 |
| CIAO2A | 0.042295765 | -0.017661016 | 0.024634749 |
| EIF1AY | 0.021539582 | -0.037764077 | -0.016224495 |
| TPM1 | 0.069130518 | 0.073464889 | 0.142595407 |
| CCL15 | 0.024698881 | -0.009096341 | 0.01560254 |
| GNG4 | 0.007810324 | -0.006051095 | 0.001759229 |
| MRPL58 | 0.033247913 | -0.012931883 | 0.02031603 |
| STC1 | 0.009099745 | -0.007764829 | 0.001334916 |
| RASSF4 | 0.044240925 | 0.027993075 | 0.072234 |

| | | | |
|---|---|---|---|
| AQP1 | 0.072760088 | -0.02742662 | 0.045333468 |
| MAN1C1 | 0.023658243 | -0.040759552 | -0.017101308 |
| COMT | 0.040863609 | 0.039472759 | 0.080336368 |
| MAN1A1 | -0.032316719 | 0.015079188 | -0.017237532 |
| LINC02532 | 0.046511589 | 0.025056419 | 0.071568008 |
| RBP4 | -0.058055802 | 0.189871961 | 0.131816159 |
| SLC25A25 | 0.014349645 | -0.011461804 | 0.002887841 |
| ATRAID | 0.05109776 | 0.032465451 | 0.083563211 |
| NPR3 | -0.053593308 | 0.040017837 | -0.01357547 |
| TPCN1 | 0.033137937 | -0.027739358 | 0.005398579 |
| AFF3 | 0.011056234 | -0.006755315 | 0.004300919 |
| AP001059.1 | 0.017063116 | -0.012700528 | 0.004362588 |
| SH3YL1 | 0.057658064 | -0.100025677 | -0.042367613 |
| YWHAQ | 0.046052761 | 0.030442002 | 0.076494762 |
| IVNS1ABP | -0.037847456 | 0.020815178 | -0.017032278 |
| SMIM24 | 0.06487455 | -0.028534793 | 0.036339757 |
| SERPINA1 | 0.062222582 | -0.084698409 | -0.022475827 |
| BEX2 | 0.046592258 | 0.026754886 | 0.073347144 |
| ATP6V0D1 | 0.045077439 | -0.034315777 | 0.010761661 |
| CAMLG | 0.035762966 | -0.014488096 | 0.021274871 |
| CNN3 | 0.051635896 | -0.031003399 | 0.020632497 |
| GAS5 | 0.046849213 | 0.048914225 | 0.095763438 |
| DHRS4-AS1 | 0.042858686 | -0.056374122 | -0.013515437 |
| CIRBP | 0.058479732 | 0.046195276 | 0.104675008 |
| ATP13A3 | -0.036319649 | 0.037386115 | 0.001066466 |
| EBPL | 0.030701464 | 0.016253858 | 0.046955322 |
| UQCC2 | 0.043984863 | -0.015072929 | 0.028911934 |
| APOE | 0.078319189 | -0.085396476 | -0.007077287 |
| FN3K | 0.028143069 | -0.026723748 | 0.001419321 |
| ARF4 | 0.042260568 | 0.024338885 | 0.066599453 |
| LINC01589 | 0.006115681 | -0.006232429 | -0.000116748 |
| P4HB | 0.044180199 | 0.022242882 | 0.066423081 |
| MVK | 0.025674078 | -0.010283609 | 0.015390469 |
| MAGED2 | 0.044142758 | 0.049637113 | 0.09377987 |
| ENPEP | 0.048827579 | -0.052947437 | -0.004119858 |
| PLTP | 0.014316782 | -0.00594712 | 0.008369661 |
| SLC66A1L | 0.007173658 | -0.004820515 | 0.002353143 |
| UPB1 | 0.03610166 | -0.040233232 | -0.004131572 |
| SFXN4 | 0.028304576 | -0.014439595 | 0.01386498 |
| ITM2C | 0.039003254 | 0.036619999 | 0.075623254 |
| GSTT2B | 0.014904784 | -0.006414896 | 0.008489888 |
| SLC7A7 | 0.059852387 | -0.08723007 | -0.027377683 |
| POLR2I | 0.049599003 | -0.028055687 | 0.021543316 |
| TUBA1B | 0.041608917 | 0.028439088 | 0.070048005 |
| AK4 | -0.061278649 | -0.087477768 | -0.148756418 |
| C3orf85 | 0.03797239 | -0.038666334 | -0.000693944 |
| SLC22A12 | 0.048185546 | -0.064037995 | -0.015852448 |
| DNAJC8 | 0.041660487 | -0.01530223 | 0.026358258 |
| CA4 | 0.021388994 | -0.014230906 | 0.007158089 |
| MTIF3 | 0.045874178 | -0.053153659 | -0.007279481 |

| | | | |
|---|---|---|---|
| SNAPC1 | 0.017894 | -0.007882243 | 0.010011757 |
| LINC01139 | 0.028112794 | -0.012559159 | 0.015553636 |
| ETNK2 | 0.045215909 | -0.050519058 | -0.005303148 |
| PDE4DIP | 0.02948811 | -0.026691866 | 0.002796244 |
| FKBP2 | 0.021857284 | -0.008517512 | 0.013339773 |
| COX7A1 | 0.034680178 | 0.023534791 | 0.058214969 |
| MORN2 | 0.050111684 | 0.021066482 | 0.071178166 |
| CDC42 | 0.054580238 | 0.019370451 | 0.073950689 |
| TFF3 | 0.017862622 | -0.014112375 | 0.003750247 |
| FOXQ1 | 0.026570502 | 0.018959624 | 0.045530125 |
| CNTD2 | 0.008343928 | -0.005564575 | 0.002779353 |
| ZNRD1 | 0.029802459 | 0.015643965 | 0.045446424 |
| TMCO1 | 0.043457383 | 0.026661286 | 0.070118668 |
| DIO1 | 0.036373807 | -0.052958503 | -0.016584695 |
| BEX4 | 0.04592762 | 0.01721109 | 0.06313871 |
| MARCH2 | 0.024363437 | -0.018079079 | 0.006284359 |
| SNX29 | 0.034721588 | -0.020323329 | 0.014398259 |
| TGM2 | 0.012767908 | -0.010412216 | 0.002355692 |
| S100A6 | 0.072064989 | 0.07502926 | 0.147094249 |
| SKAP1 | 0.012483654 | -0.010925862 | 0.001557791 |
| RAB1B | 0.038128344 | -0.018748819 | 0.019379525 |
| LINC02294 | 0.030050062 | -0.010615597 | 0.019434464 |
| ECI2 | 0.054661031 | -0.043271194 | 0.011389837 |
| CRYL1 | 0.056981097 | -0.077707268 | -0.020726171 |
| GLS | 0.058028561 | -0.044227844 | 0.013800718 |
| LYAR | 0.013214959 | -0.006485214 | 0.006729745 |
| IFRD2 | 0.030248404 | -0.013705756 | 0.016542648 |
| S100A10 | 0.064156556 | 0.066673777 | 0.130830334 |
| DYNLL1 | 0.060155072 | 0.046801091 | 0.106956164 |
| AGXT | -0.026889439 | 0.009538854 | -0.017350584 |
| AIP | 0.034471462 | -0.02229293 | 0.012178533 |
| ZBTB16 | 0.026515277 | -0.026506156 | 9.12123E-06 |
| LRMDA | 0.02581583 | -0.029107815 | -0.003291985 |
| SLC22A2 | 0.053435718 | 0.098366496 | 0.151802214 |
| LINC01697 | 0.0365682 | 0.021867167 | 0.058435368 |
| TGIF1 | 0.030249988 | 0.013054712 | 0.0433047 |
| SAR1A | 0.035229899 | 0.013287209 | 0.048517108 |
| EGLN3 | 0.023487142 | -0.01648377 | 0.007003372 |
| GADD45B | 0.029802401 | -0.024668531 | 0.005133869 |
| CCDC25 | 0.032424695 | -0.020099596 | 0.012325099 |
| MARCH9 | 0.023689118 | -0.014331442 | 0.009357676 |
| ALDH1L1 | -0.048086148 | -0.028481677 | -0.076567826 |
| SLC34A3 | 0.034426197 | -0.071274413 | -0.036848215 |
| LINC02447 | 0.017550636 | -0.019788769 | -0.002238133 |
| TMEM179B | 0.03549484 | 0.023167243 | 0.058662083 |
| TRMT6 | 0.016142048 | -0.007081547 | 0.0090605 |
| DACH1 | 0.016682787 | -0.015110664 | 0.001572123 |
| PDIA6 | 0.044641204 | 0.043070261 | 0.087711465 |
| HLA-A | 0.065842162 | 0.160135317 | 0.22597748 |
| PPP1R35 | 0.027186761 | 0.011668898 | 0.038855658 |

| | | | |
|---|---|---|---|
| AC136475.3 | 0.006451586 | -0.006661802 | -0.000210216 |
| SPARC | 0.03547142 | 0.030698585 | 0.066170005 |
| FAH | 0.042649971 | -0.034213632 | 0.008436339 |
| ANP32B | 0.038703225 | 0.039016728 | 0.077719953 |
| COMMD8 | 0.039230271 | -0.021176558 | 0.018053713 |
| ATP6V1E1 | 0.046531908 | -0.063505042 | -0.016973134 |
| TLN2 | 0.039166799 | -0.060270751 | -0.021103952 |
| RASGEF1B | 0.02101033 | -0.018691054 | 0.002319276 |
| RAB17 | 0.042751884 | -0.024721432 | 0.018030451 |
| MAFG | 0.019773765 | -0.010536195 | 0.00923757 |
| NFE2L2 | 0.041281846 | 0.025791293 | 0.067073138 |
| USF2 | 0.029357473 | -0.011096526 | 0.018260947 |
| UBC | 0.057002741 | 0.063644424 | 0.120647165 |
| TRIL | 0.01413881 | -0.006907259 | 0.007231552 |
| HMOX1 | 0.037282957 | -0.038124137 | -0.000841181 |
| TM9SF1 | 0.029505122 | 0.019781287 | 0.049286409 |
| NME3 | 0.046919643 | 0.055999444 | 0.102919087 |
| LINC01578 | 0.041907752 | 0.038685757 | 0.080593509 |
| SELENOS | 0.032013766 | 0.028956401 | 0.060970167 |
| SLC14A2 | 0.006494158 | -0.00681157 | -0.000317412 |
| STRIP2 | 0.012120658 | -0.010595599 | 0.001525059 |
| RORA | 0.032627858 | -0.018232008 | 0.01439585 |
| AK2 | 0.0437627 | -0.02779109 | 0.01597161 |
| MRPL50 | 0.032190497 | 0.016262692 | 0.048453189 |
| SWI5 | 0.030000982 | -0.013371147 | 0.016629836 |
| RANBP1 | 0.037916633 | 0.021057773 | 0.058974406 |
| CMYA5 | 0.02841914 | 0.019888124 | 0.048307264 |
| RAB5C | 0.046346784 | -0.043488498 | 0.002858285 |
| EPCAM | 0.054793353 | 0.04118362 | 0.095976973 |
| GAMT | 0.050643384 | -0.022692864 | 0.02795052 |
| MRPL21 | 0.035518078 | 0.013250609 | 0.048768687 |
| SEPTIN1 | 0.021207836 | -0.024008396 | -0.00280056 |
| TRIM63 | 0.01505776 | -0.015953016 | -0.000895256 |
| BAX | 0.037530469 | 0.026467319 | 0.063997789 |
| MYO9A | -0.032589904 | 0.038921935 | 0.006332031 |
| SEC11C | 0.035052353 | 0.022595513 | 0.057647866 |
| CDC42EP3 | 0.013454837 | -0.009931823 | 0.003523014 |
| CLEC11A | 0.018518826 | 0.010059461 | 0.028578286 |
| MRPL46 | 0.03016764 | -0.015835649 | 0.014331991 |
| ACADM | 0.052256812 | -0.061498874 | -0.009242062 |
| OXR1 | -0.030560472 | 0.055953399 | 0.025392926 |
| AHNAK | -0.044123673 | 0.087100773 | 0.042977099 |
| SLC7A9 | 0.032985035 | -0.039568245 | -0.00658321 |
| AFAP1L2 | 0.007013046 | -0.006531838 | 0.000481208 |
| UFL1-AS1 | -0.007145525 | 0.004433982 | -0.002711544 |
| SLC7A8 | -0.057313527 | -0.130253413 | -0.18756694 |
| SDF2L1 | 0.031873569 | 0.029042315 | 0.060915884 |
| PON2 | 0.027552286 | 0.021289706 | 0.048841992 |
| NABP2 | 0.022725832 | -0.015378906 | 0.007346926 |
| MIOX | 0.072194741 | -0.134895668 | -0.062700927 |

| | | | |
|---|---|---|---|
| SLC25A39 | 0.045291688 | -0.052208998 | -0.00691731 |
| ELF3 | 0.033240409 | 0.034903273 | 0.068143682 |
| NEFH | 0.009886763 | -0.006141537 | 0.003745225 |
| CST3 | 0.056865044 | 0.056459066 | 0.11332411 |
| YWHAH | 0.028975153 | 0.020761435 | 0.049736588 |
| TM7SF2 | 0.032799486 | 0.022299027 | 0.055098513 |
| DERA | 0.039621036 | -0.031327058 | 0.008293978 |
| AHCY | 0.043159464 | -0.025521129 | 0.017638336 |
| LINC01060 | 0.018886461 | -0.00988941 | 0.008997051 |
| LINC00472 | -0.043078885 | 0.07559359 | 0.032514705 |
| IFITM2 | 0.022373573 | 0.016295858 | 0.038669431 |
| HAVCR2 | 0.021798606 | -0.012701864 | 0.009096742 |
| TMEM59 | 0.055179075 | 0.031098384 | 0.086277459 |
| GNPDA1 | 0.040277917 | -0.069502686 | -0.02922477 |
| PTTG1IP | 0.039267911 | 0.048013415 | 0.087281326 |
| MYDGF | 0.03155843 | 0.018554738 | 0.050113168 |
| IP6K3 | 0.009930435 | -0.009071136 | 0.000859299 |
| MEG3 | 0.026490209 | 0.026993895 | 0.053484104 |
| HDAC6 | 0.028011081 | -0.01619147 | 0.011819611 |
| ALPL | 0.027430368 | -0.029960272 | -0.002529904 |
| ITPKB | 0.01009252 | -0.00567524 | 0.00441728 |
| SAT1 | 0.055429998 | -0.094641767 | -0.039211769 |
| TIFA | 0.011295328 | -0.004474811 | 0.006820517 |
| PRSS35 | 0.011194902 | -0.011140123 | 5.47788E-05 |
| SREK1IP1 | 0.024776907 | -0.01278664 | 0.011990267 |
| COMMD9 | 0.026419835 | -0.010025143 | 0.016394692 |
| MPLKIP | 0.040564652 | 0.023084704 | 0.063649356 |
| DDIT3 | 0.027453238 | 0.02046199 | 0.047915228 |
| AC017000.1 | 0.008326252 | -0.004058177 | 0.004268075 |
| CRB3 | 0.024006431 | 0.009830207 | 0.033836637 |
| TSPO | 0.038989829 | 0.054156244 | 0.093146074 |
| FGF2 | 0.013162468 | -0.005985028 | 0.00717744 |
| APIP | 0.029537728 | -0.013064006 | 0.016473722 |
| MMP24OS | 0.044913185 | -0.023602429 | 0.021310757 |
| LINC01320 | -0.047817633 | 0.109680516 | 0.061862883 |
| YIF1A | 0.033470234 | 0.015880675 | 0.049350909 |
| SLC17A5 | 0.026552477 | -0.038984046 | -0.012431569 |
| KLF15 | 0.023304271 | -0.014102566 | 0.009201705 |
| GPRC5C | 0.035217924 | 0.02674614 | 0.061964064 |
| BHLHE40 | 0.027680901 | -0.010754695 | 0.016926205 |
| FOLR1 | 0.037743846 | 0.017163721 | 0.054907566 |
| ANXA2 | 0.039344865 | 0.045640032 | 0.084984897 |
| HLA-E | 0.035510452 | 0.038162528 | 0.07367298 |
| GCNT4 | -0.008173383 | 0.00457104 | -0.003602343 |
| TNIP2 | 0.021449886 | -0.009994994 | 0.011454892 |
| SLC25A33 | 0.018737386 | -0.01382269 | 0.004914696 |
| BNIP3L | 0.041900533 | -0.034734196 | 0.007166337 |
| DALRD3 | 0.024857332 | 0.010865765 | 0.035723097 |
| CDC26 | 0.036233224 | -0.020556688 | 0.015676536 |
| FUOM | 0.046354164 | -0.070412408 | -0.024058243 |

| | | | |
|---|---|---|---|
| ESRRA | 0.035637554 | -0.017698495 | 0.017939059 |
| GDPD3 | 0.025227117 | -0.016475352 | 0.008751765 |
| EIF3D | 0.036613596 | 0.0170326 | 0.053646196 |
| IL32 | 0.049236514 | 0.069423048 | 0.118659561 |
| EAPP | 0.032515967 | 0.017083484 | 0.049599451 |
| SH3GL3 | 0.004741133 | -0.004545473 | 0.00019566 |
| FKBP8 | 0.042009772 | -0.031670854 | 0.010338918 |
| AKAP12 | 0.030687706 | 0.047813105 | 0.078500811 |
| KCNH6 | 0.011523754 | -0.012077991 | -0.000554237 |
| SLC25A42 | 0.02874174 | -0.03720362 | -0.00846188 |
| HOXB8 | 0.018001779 | -0.010619444 | 0.007382335 |
| TP53I11 | 0.010179156 | -0.006199219 | 0.003979937 |
| AP005229.1 | 0.006695615 | -0.006489423 | 0.000206192 |
| RHOB | 0.040424587 | 0.019945425 | 0.060370012 |
| AC006441.4 | 0.016563094 | -0.018843603 | -0.002280509 |
| USP9Y | -0.017587769 | -0.016776301 | -0.03436407 |
| MRPL48 | 0.022269045 | -0.009993543 | 0.012275502 |
| SSB | 0.036534007 | 0.020521793 | 0.057055801 |
| BOLA3 | 0.03768317 | -0.023686106 | 0.013997064 |
| PSENEN | 0.03134096 | 0.015285327 | 0.046626288 |
| AARD | 0.01299496 | -0.005296508 | 0.007698452 |
| BDH2 | 0.051680206 | -0.023674632 | 0.028005574 |
| ACAD11 | 0.028963339 | -0.017782197 | 0.011181142 |
| HHLA2 | 0.012994498 | 0.006931217 | 0.019925715 |
| DDAH2 | 0.03756281 | 0.047180712 | 0.084743522 |
| THBS1 | -0.03635904 | 0.067094967 | 0.030735927 |
| SELENBP1 | 0.031249076 | -0.026868024 | 0.004381052 |
| STX8 | 0.027620147 | -0.011559128 | 0.016061019 |
| PDZK1IP1 | 0.054811495 | -0.035435991 | 0.019375504 |
| NR1H4 | 0.03275028 | -0.028892033 | 0.003858247 |
| ECSIT | 0.027277809 | -0.020888138 | 0.006389671 |
| MGAT4B | 0.032498469 | 0.032955397 | 0.065453866 |
| ANAPC13 | 0.037580438 | -0.0303481 | 0.007232339 |
| SEZ6L2 | 0.019848915 | 0.037589777 | 0.057438692 |
| VPS25 | 0.031953328 | 0.013981223 | 0.045934551 |
| MYH8 | 0.018869549 | -0.010021907 | 0.008847642 |
| GOT1 | 0.035738915 | -0.014298194 | 0.021440721 |
| MAL | 0.04728757 | 0.054116932 | 0.101404503 |
| LMAN1 | 0.040118031 | 0.018225761 | 0.058343792 |
| TMEM100 | 0.004624372 | -0.004375524 | 0.000248848 |
| C11orf52 | 0.026822925 | -0.012442563 | 0.014380363 |
| SLC6A8 | 0.028262434 | -0.014308736 | 0.013953698 |
| MGAM | 0.019656897 | -0.019326582 | 0.000330315 |
| FCRLB | 0.013078956 | -0.008835964 | 0.004242992 |
| C7orf50 | 0.040630892 | 0.0255863 | 0.066217192 |
| CYS1 | 0.042828071 | 0.033375438 | 0.076203509 |
| POLR2E | 0.034343497 | -0.024372558 | 0.009970939 |
| TRHDE-AS1 | -0.021116019 | 0.035108919 | 0.013992901 |
| SULT2B1 | 0.006466185 | -0.00606946 | 0.000396725 |
| SOD1 | 0.05065815 | -0.025956717 | 0.024701433 |

| | | | |
|---|---|---|---|
| TADA3 | 0.022934868 | -0.009597477 | 0.01333739 |
| AC008592.4 | 0.004296969 | -0.003698546 | 0.000598423 |
| TP53TG1 | 0.025754202 | -0.018484917 | 0.007269285 |
| ADGRG1 | 0.034299631 | 0.026938582 | 0.061238213 |
| AOPEP | 0.03194162 | 0.039946189 | 0.071887809 |
| REEP5 | 0.039505317 | 0.032694909 | 0.072200226 |
| CALCA | 0.004570044 | -0.0044926 | 7.7444E-05 |
| CD68 | 0.016233342 | -0.010435621 | 0.005797722 |
| TKT | 0.034965113 | -0.019935118 | 0.015029996 |
| IRX3 | -0.029424591 | 0.0793976 | 0.049973009 |
| RTN3 | 0.031580623 | 0.014356498 | 0.045937121 |
| CBLN2 | 0.006789839 | -0.006363969 | 0.00042587 |
| STRAP | 0.036507074 | -0.015404147 | 0.021102926 |
| RBM4 | 0.034699191 | 0.016268986 | 0.050968178 |
| MAP1LC3A | 0.029730908 | 0.019367474 | 0.049098382 |
| ACSM2B | 0.05109413 | -0.027710194 | 0.023383936 |
| LAP3 | 0.037205919 | -0.035225238 | 0.001980681 |
| PTH1R | 0.053140405 | 0.022620902 | 0.075761307 |
| HSP90B1 | 0.049933476 | 0.052673844 | 0.10260732 |
| OTUD6B-AS1 | 0.033661399 | 0.019522108 | 0.053183508 |
| PRKCSH | 0.031050603 | 0.036049471 | 0.067100074 |
| TAPBP | 0.029785596 | 0.038868693 | 0.068654289 |
| OGDHL | 0.038821599 | -0.026075173 | 0.012746426 |
| PLA2G4A | 0.004266406 | -0.003809019 | 0.000457386 |
| ABHD14B | 0.035431849 | -0.020806172 | 0.014625677 |
| GSTO1 | 0.042866998 | -0.033392725 | 0.009474273 |
| CLPTM1 | 0.030563861 | -0.017933467 | 0.012630394 |
| AIFM1 | 0.039195026 | -0.038078692 | 0.001116334 |
| HSPA1B | -0.035095264 | 0.034356273 | -0.000738991 |
| LINC01554 | 0.006597416 | -0.007139992 | -0.000542576 |
| OGG1 | 0.027100146 | -0.021622588 | 0.005477558 |
| EPN3 | 0.008278908 | -0.008063824 | 0.000215084 |
| DNAH17 | 0.004627828 | -0.005085931 | -0.000458103 |
| HCFC1R1 | 0.037914129 | 0.023583844 | 0.061497973 |
| SPON1 | -0.014004532 | 0.025232794 | 0.011228262 |
| RARA | 0.016682413 | -0.007791845 | 0.008890568 |
| SAMD4A | 0.016185531 | 0.009367012 | 0.025552543 |
| SMIM27 | 0.023499078 | -0.022258031 | 0.001241048 |
| F2R | 0.010054215 | -0.007412964 | 0.002641251 |
| RFC1 | 0.027733853 | -0.024859341 | 0.002874512 |
| BCL7C | 0.033130673 | 0.028796526 | 0.061927199 |
| EIF3M | 0.035299261 | 0.031573109 | 0.06687237 |
| MRPL15 | 0.030152871 | 0.017914088 | 0.048066959 |
| PACC1 | 0.012576332 | -0.005533663 | 0.007042669 |
| TNFSF10 | 0.048857003 | 0.100048865 | 0.148905868 |
| OPTN | 0.035789997 | -0.021794948 | 0.013995048 |
| SORBS2 | 0.026942821 | 0.026006584 | 0.052949405 |
| GALNT10 | -0.013058191 | 0.014592417 | 0.001534227 |
| SLC4A4 | -0.045611532 | -0.065110578 | -0.11072211 |
| UBAC2 | 0.020705714 | 0.010579899 | 0.031285613 |

| | | | |
|---|---|---|---|
| AC067930.3 | 0.004887746 | -0.003820771 | 0.001066975 |
| NAPEPLD | 0.026162226 | -0.02725485 | -0.001092624 |
| CASTOR1 | 0.020550089 | -0.029759761 | -0.009209673 |
| SMIM5 | 0.010476628 | -0.004651744 | 0.005824884 |
| AC006460.2 | 0.007209254 | -0.006840548 | 0.000368706 |
| LINC01510 | 0.030625581 | -0.021030289 | 0.009595292 |
| THOC7 | 0.032811227 | 0.013883531 | 0.046694758 |
| TINAGL1 | 0.035963638 | 0.05797787 | 0.093941508 |
| ATP5F1A | 0.046570509 | -0.025178836 | 0.021391672 |
| OCIAD1 | 0.03833244 | 0.028402524 | 0.066734965 |
| KIAA1191 | -0.038698285 | -0.01648802 | -0.055186305 |
| ITGB1 | 0.039031988 | 0.027459133 | 0.066491121 |
| DERL2 | 0.021864329 | 0.015821494 | 0.037685824 |
| MAP4K3-DT | 0.024863623 | -0.022858397 | 0.002005226 |
| ZNRF1 | 0.01548708 | -0.013667776 | 0.001819304 |
| TMED3 | 0.020095204 | 0.016746349 | 0.036841554 |
| SSBP4 | 0.01850751 | 0.007914919 | 0.026422429 |
| EAF2 | 0.03367444 | -0.016865018 | 0.016809421 |
| IFNGR2 | 0.022657842 | 0.017432548 | 0.040090391 |
| P4HA1 | 0.020910692 | -0.014766937 | 0.006143755 |
| TYMP | 0.032334838 | 0.057638833 | 0.08997367 |
| NUDT22 | 0.030301147 | -0.02710237 | 0.003198776 |
| PRR15 | 0.010155638 | -0.004697968 | 0.00545767 |
| TACC3 | 0.006581546 | -0.003304592 | 0.003276953 |
| AP001042.1 | 0.003510829 | -0.003367416 | 0.000143413 |
| AGMAT | 0.050958601 | -0.100532779 | -0.049574179 |
| COQ10B | 0.020871619 | -0.010324762 | 0.010546857 |
| CES3 | 0.014573133 | -0.017344157 | -0.002771024 |
| CPPED1 | 0.025179307 | -0.013948378 | 0.01123093 |
| SELENOF | 0.033733115 | 0.023417414 | 0.057150529 |
| AC087482.1 | 0.024772298 | -0.02833746 | -0.003565163 |
| HSCB | 0.018507794 | -0.010915022 | 0.007592772 |
| SLC13A2 | 0.024312788 | -0.034099167 | -0.009786379 |
| ANXA11 | 0.034378953 | 0.0322194 | 0.066598353 |
| MIDN | 0.024172507 | 0.01044759 | 0.034620096 |
| PLEKHH3 | 0.01184959 | -0.005748979 | 0.006100611 |
| PHPT1 | 0.046462512 | 0.058443826 | 0.104906338 |
| GABPB1-AS1 | -0.021564526 | 0.028873679 | 0.007309153 |
| CBX6 | 0.028557276 | 0.020294369 | 0.048851645 |
| NDFIP1 | 0.036608077 | 0.038436237 | 0.075044314 |
| MT-ND6 | -0.044417661 | 0.038145458 | -0.006272203 |
| MELTF | 0.008490172 | -0.004856829 | 0.003633343 |
| GRB7 | 0.019637023 | -0.015115658 | 0.004521365 |
| GABRE | -0.00797605 | 0.006809375 | -0.001166675 |
| C5orf24 | 0.033466535 | -0.029750686 | 0.003715849 |
| PTGES2 | 0.026507152 | -0.01195364 | 0.014553512 |
| GHITM | 0.043979905 | -0.035827067 | 0.008152838 |
| RAB25 | 0.017483915 | 0.013591016 | 0.031074931 |
| WBP1 | 0.028789898 | 0.020998414 | 0.049788312 |
| HMOX2 | 0.025525234 | -0.017870829 | 0.007654405 |

| | | | |
|---|---|---|---|
| LMO2 | 0.015670807 | -0.018624988 | -0.002954181 |
| AC022613.1 | 0.022684815 | 0.011821928 | 0.034506743 |
| UPK3BL1 | 0.022617018 | 0.02026738 | 0.042884398 |
| F2RL1 | 0.021110774 | 0.01157548 | 0.032686253 |
| CALD1 | 0.040729153 | 0.057949971 | 0.098679124 |
| TRIM47 | 0.026437815 | -0.011882255 | 0.01455556 |
| SELENON | 0.016302206 | -0.007430194 | 0.008872012 |
| SLC2A1 | 0.010521682 | -0.012938224 | -0.002416542 |
| CARD6 | 0.005866549 | -0.005122273 | 0.000744276 |
| DDX10 | 0.029261421 | -0.041924087 | -0.012662666 |
| LINC01965 | 0.004747775 | -0.005363517 | -0.000615742 |
| PPP1CB | 0.037164275 | 0.028554839 | 0.065719114 |
| ADM | 0.013951292 | -0.008959394 | 0.004991897 |
| DDX17 | -0.045237345 | 0.085641593 | 0.040404248 |
| HM13 | 0.02510054 | 0.011301032 | 0.036401571 |
| LAPTM4A | 0.054173732 | 0.059479137 | 0.113652869 |
| IRX5 | -0.017953747 | 0.038765791 | 0.020812044 |
| PPBP | 0.003751177 | -0.003751177 | 1.77636E-15 |
| SINHCAF | 0.018496154 | 0.008767598 | 0.027263753 |
| LSM8 | 0.029550926 | 0.018788472 | 0.048339398 |
| CAT | 0.043746149 | -0.092073692 | -0.048327543 |
| TTTY14 | 0.014325212 | -0.024107555 | -0.009782343 |
| STMP1 | 0.033477584 | -0.01749951 | 0.015978074 |
| DHRS12 | 0.022371924 | -0.009820391 | 0.012551533 |
| ZFAT | 0.012602369 | -0.008167308 | 0.004435062 |
| TCTEX1D2 | 0.018126852 | 0.008544086 | 0.026670938 |
| ELMOD1 | 0.009798752 | -0.010009747 | -0.000210995 |
| TIMP1 | 0.027580456 | 0.06640649 | 0.093986945 |
| PCMT1 | 0.024234882 | 0.019078626 | 0.043313508 |
| PIGF | 0.018917418 | 0.014168613 | 0.033086032 |
| OSTF1 | 0.020004118 | -0.015205416 | 0.004798702 |
| SNRNP70 | 0.030721771 | 0.026132198 | 0.05685397 |
| PTGER1 | 0.006256638 | -0.005828713 | 0.000427925 |
| DDX3Y | 0.014959187 | -0.030507883 | -0.015548696 |
| ARL6IP5 | 0.030168387 | 0.043017859 | 0.073186246 |
| TMEM165 | 0.023434378 | 0.015740486 | 0.039174864 |
| UNC119 | 0.01654259 | 0.010456647 | 0.026999237 |
| GUCD1 | 0.026375641 | -0.012600201 | 0.013775439 |
| NR1D2 | 0.020682713 | 0.009261624 | 0.029944336 |
| RNF152 | -0.040174041 | -0.061040135 | -0.101214176 |
| LPL | -0.007069211 | 0.004561141 | -0.002508069 |
| AL353751.1 | 0.013653973 | 0.008289314 | 0.021943287 |
| BTD | 0.018427003 | 0.009116158 | 0.027543162 |
| CA5B | 0.010939204 | -0.010180993 | 0.00075821 |
| MTPN | 0.026747293 | -0.014561406 | 0.012185887 |
| SESN2 | 0.02315302 | -0.012388856 | 0.010764164 |
| CYP2S1 | 0.004173514 | -0.004180663 | -7.14963E-06 |
| ISOC2 | 0.043083259 | -0.042698283 | 0.000384976 |
| XIST | 0.03432797 | 0.145119149 | 0.179447119 |
| CDK5RAP3 | 0.022114241 | 0.01056225 | 0.032676491 |

| | | | |
|---|---|---|---|
| SARS | 0.026862303 | -0.017597553 | 0.009264751 |
| TMEM205 | 0.036884848 | 0.063755398 | 0.100640246 |
| TUBA1C | 0.030026563 | 0.016660582 | 0.046687144 |
| MAGOH | 0.029580888 | 0.016393639 | 0.045974527 |
| HLA-DOA | -0.009616521 | 0.005748073 | -0.003868447 |
| EMC7 | 0.026941493 | 0.022375417 | 0.049316911 |
| KLHDC7A | -0.016398712 | 0.02021213 | 0.003813419 |
| BPI | -0.015614222 | 0.007950449 | -0.007663773 |
| PFDN1 | 0.026518703 | 0.012076383 | 0.038595086 |
| AC114316.1 | 0.002387911 | -0.002387911 | 1.77636E-15 |
| TWF2 | 0.017160452 | 0.008873479 | 0.026033931 |
| LYPD6B | 0.004200723 | -0.003183016 | 0.001017707 |
| FAM104B | 0.024299998 | -0.017791789 | 0.00650821 |
| CTSD | 0.046999779 | 0.1022422 | 0.149241979 |
| ZNF428 | 0.034611256 | 0.018415173 | 0.053026429 |
| HK2 | 0.004189344 | -0.003336891 | 0.000852454 |
| C1QTNF12 | 0.016483421 | -0.017568003 | -0.001084583 |
| BCAP31 | 0.034553436 | 0.019270142 | 0.053823579 |
| SCARA3 | 0.011446691 | -0.01505965 | -0.003612959 |
| FRY | 0.013706311 | -0.010897959 | 0.002808351 |
| AC016831.5 | 0.008022786 | -0.005485133 | 0.002537653 |
| AP1M1 | 0.018068511 | -0.013422167 | 0.004646345 |
| TINAG | 0.031896259 | 0.02366731 | 0.05556357 |
| NDUFV3 | 0.036339332 | -0.02824904 | 0.008090292 |
| MED10 | 0.022928995 | 0.011709868 | 0.034638863 |
| SERTAD4-AS1 | 0.015945495 | 0.019629914 | 0.035575409 |
| HNF4A | 0.035745938 | -0.047589746 | -0.011843808 |
| WASF2 | 0.026489102 | 0.012370131 | 0.038859233 |
| TMBIM4 | 0.034955966 | 0.03230831 | 0.067264276 |
| BX664615.2 | 0.018507939 | -0.010314928 | 0.00819301 |
| RGMB-AS1 | 0.006657263 | -0.005058145 | 0.001599119 |
| UBXN4 | 0.032300906 | 0.020388841 | 0.052689747 |
| GPCPD1 | -0.013050281 | 0.024721632 | 0.011671351 |
| WDR18 | 0.027676837 | -0.018254564 | 0.009422273 |
| PPP4R4 | 0.003593456 | -0.00242935 | 0.001164106 |
| TXNDC9 | 0.022132775 | 0.011801027 | 0.033933802 |
| ZSCAN16-AS1 | 0.032892925 | -0.048341911 | -0.015448986 |
| EIF2D | 0.017545619 | -0.011012138 | 0.006533481 |
| RPS27L | 0.044430358 | -0.021178289 | 0.023252068 |
| TMEM50A | 0.031665253 | 0.033783814 | 0.065449067 |
| PNOC | 0.003423748 | -0.003287878 | 0.00013587 |
| CISD3 | 0.036318397 | -0.035303397 | 0.001015 |
| PTPN1 | 0.017182084 | -0.012162337 | 0.005019746 |
| CD151 | 0.034088773 | 0.055733915 | 0.089822688 |
| PGM1 | 0.02163306 | -0.011561237 | 0.010071823 |
| SLC22A6 | 0.037718672 | -0.092060229 | -0.054341557 |
| BAIAP2 | 0.019535441 | -0.024869541 | -0.0053341 |
| ACYP1 | 0.01448595 | 0.007601051 | 0.022087 |
| ACBD4 | 0.017110406 | -0.01081674 | 0.006293666 |
| MMP7 | 0.029089929 | 0.051106777 | 0.080196706 |

| | | | |
|---|---|---|---|
| DDX6 | -0.027388075 | 0.019844565 | -0.00754351 |
| ZP3 | 0.005511197 | -0.005252302 | 0.000258896 |
| AC006460.1 | 0.004962581 | -0.005084867 | -0.000122286 |
| METRN | 0.032011723 | 0.037513251 | 0.069524974 |
| APEX1 | 0.031488879 | 0.015195566 | 0.046684444 |
| TMPRSS2 | 0.012159813 | -0.008301865 | 0.003857948 |
| PF4 | 0.003878351 | -0.003878351 | -1.77636E-15 |
| CASQ2 | -0.006395607 | 0.009597761 | 0.003202154 |
| PTGS1 | 0.00358523 | -0.003313021 | 0.000272208 |
| DNAJC1 | 0.021043391 | 0.026103281 | 0.047146672 |
| ERG28 | 0.019629564 | 0.014950603 | 0.034580167 |
| CD99 | 0.022109829 | 0.016061026 | 0.038170854 |
| TUSC2 | 0.030032186 | -0.01618549 | 0.013846696 |
| CREB3 | 0.020831901 | 0.009859605 | 0.030691507 |
| CAB39L | 0.010175321 | -0.006790911 | 0.00338441 |
| C16orf87 | 0.022677743 | -0.011520342 | 0.011157401 |
| HNRNPH3 | 0.028492993 | 0.030175685 | 0.058668677 |
| DNAJB11 | 0.016607239 | 0.008390007 | 0.024997246 |
| TMED10 | 0.028559489 | 0.02642344 | 0.054982929 |
| CCS | 0.030353214 | 0.024255676 | 0.05460889 |
| RHOBTB1 | 0.024196512 | -0.024785459 | -0.000588947 |
| FUZ | 0.025217177 | -0.031175026 | -0.005957849 |
| TUBG1 | 0.020675048 | -0.020305714 | 0.000369333 |
| SLC39A1 | 0.02202328 | 0.019210834 | 0.041234114 |
| ENO2 | 0.006005963 | -0.004345219 | 0.001660743 |
| BCAP29 | 0.015633572 | 0.008102667 | 0.02373624 |
| TMEM263 | 0.021320749 | 0.014457692 | 0.035778441 |
| SUCLA2 | 0.027677003 | -0.014295562 | 0.013381441 |
| SRM | 0.02106557 | 0.013545938 | 0.034611509 |
| ASPG | 0.0141458 | -0.015528512 | -0.001382712 |
| LINC00824 | 0.003864123 | -0.002398554 | 0.001465569 |
| RPL22L1 | 0.02624711 | 0.020500577 | 0.046747688 |
| SPON2 | 0.021513774 | 0.052013008 | 0.073526782 |
| SNTB2 | 0.022011284 | -0.021477673 | 0.000533611 |
| VAMP3 | 0.025244634 | -0.013845679 | 0.011398956 |
| PRRC2C | 0.029241603 | 0.02268982 | 0.051931423 |
| AC098483.1 | 0.008253512 | -0.006433161 | 0.001820351 |
| OSER1 | 0.019142473 | 0.01117263 | 0.030315103 |
| RGCC | 0.005998726 | -0.006087295 | -8.85693E-05 |
| SMIM1 | 0.028585799 | -0.046197448 | -0.017611648 |
| GRM5-AS1 | -0.005119428 | 0.002935068 | -0.00218436 |
| ARMCX6 | 0.014070434 | 0.010065507 | 0.024135941 |
| MTFR1 | 0.021371024 | -0.013364706 | 0.008006317 |
| ZDHHC12 | 0.020232603 | 0.017100219 | 0.037332823 |
| TSPAN6 | 0.022893744 | 0.022454661 | 0.045348404 |
| CCDC90B | 0.019815516 | 0.018190158 | 0.038005674 |
| PARG | 0.01730397 | -0.009351971 | 0.007951999 |
| ST14 | 0.019316325 | 0.018626097 | 0.037942422 |
| CES2 | 0.030412769 | -0.018845212 | 0.011567557 |
| TBC1D7 | 0.01509599 | -0.007832817 | 0.007263173 |

| | | | |
|---|---|---|---|
| SMIM3 | 0.00922616 | -0.006897335 | 0.002328824 |
| HLA-DQA1 | 0.009532781 | 0.005779759 | 0.01531254 |
| CYHR1 | 0.023235656 | 0.012951481 | 0.036187137 |
| SPSB3 | 0.024332182 | 0.012235225 | 0.036567407 |
| AGXT2 | 0.038947856 | -0.096775462 | -0.057827605 |
| TFCP2L1 | 0.025436579 | -0.031932812 | -0.006496233 |
| SBSPON | 0.014424156 | 0.011696517 | 0.026120673 |
| ANGPTL8 | 0.003931827 | -0.002697153 | 0.001234674 |
| CLIC5 | 0.011009636 | -0.009136685 | 0.001872951 |
| ANO3-AS1 | 0.00244661 | -0.00244661 | -1.77636E-15 |
| DPCD | 0.020415739 | 0.011373271 | 0.03178901 |
| RRP7A | 0.01879449 | -0.011847035 | 0.006947455 |
| OVOL1-AS1 | 0.01184889 | -0.007103215 | 0.004745676 |
| ATP6V0B | 0.035483341 | 0.048268665 | 0.083752006 |
| PDE7A | 0.0160549 | 0.01073906 | 0.026793961 |
| EMC3 | 0.021113331 | 0.014681483 | 0.035794813 |
| PLLP | 0.019704453 | 0.035241691 | 0.054946144 |
| SLC27A2 | -0.032851278 | 0.024099663 | -0.008751615 |
| CRYAB | 0.054851018 | 0.157099861 | 0.211950878 |
| TUBGCP2 | 0.025066013 | -0.012814166 | 0.012251847 |
| SCD | -0.009883171 | -0.005261597 | -0.015144768 |
| NMI | 0.013638999 | 0.008546103 | 0.022185101 |
| AP2A1 | 0.020911956 | -0.018251134 | 0.002660822 |
| IL6R | 0.015237129 | -0.012026695 | 0.003210435 |
| PRKCB | 0.00407271 | -0.004332716 | -0.000260005 |
| AMPH | 0.003742064 | -0.001939356 | 0.001802708 |
| FYN | 0.021165373 | -0.016464146 | 0.004701227 |
| CTSA | 0.031454394 | -0.038896012 | -0.007441618 |
| SLC38A11 | 0.007558199 | -0.006243266 | 0.001314933 |
| EFCAB14 | -0.022624546 | 0.021606545 | -0.001018001 |
| PLS1 | 0.020726062 | 0.019091881 | 0.039817943 |
| RPN2 | 0.027055389 | 0.014810145 | 0.041865534 |
| LSM12 | 0.022718602 | -0.012006073 | 0.010712529 |
| SPINT1-AS1 | 0.022046028 | 0.027493416 | 0.049539444 |
| ORMDL1 | 0.023450135 | 0.019055381 | 0.042505516 |
| RALYL | 0.014842508 | -0.008581905 | 0.006260602 |
| PRKCE | -0.017245483 | -0.020809966 | -0.038055449 |
| STK16 | 0.020580137 | -0.017627492 | 0.002952645 |
| RAB11FIP3 | 0.03961808 | -0.089829344 | -0.050211264 |
| TMEM9 | 0.024422334 | 0.021024311 | 0.045446645 |
| CLIP2 | 0.00876934 | -0.005736705 | 0.003032635 |
| NEU1 | 0.023378149 | 0.026782043 | 0.050160192 |
| NDRG1 | 0.035657251 | -0.038497211 | -0.00283996 |
| AC133552.5 | 0.013640357 | -0.009355649 | 0.004284708 |
| DCTPP1 | 0.019309433 | 0.014812052 | 0.034121485 |
| NAPA-AS1 | 0.009036862 | -0.004921666 | 0.004115196 |
| KRT7 | 0.029435357 | 0.076472949 | 0.105908306 |
| ARL1 | 0.029319192 | 0.02708637 | 0.056405562 |
| C2orf74 | -0.022171002 | 0.028045279 | 0.005874277 |
| CNIH4 | 0.021876149 | 0.01273335 | 0.0346095 |

| | | | |
|---|---|---|---|
| BTG1 | 0.034877955 | 0.030517362 | 0.065395317 |
| QPCT | 0.015402533 | 0.011106963 | 0.026509496 |
| WDR54 | 0.014275741 | 0.008442367 | 0.022718108 |
| ARL16 | 0.025300088 | 0.021774704 | 0.047074792 |
| SRSF10 | 0.025383082 | 0.017900895 | 0.043283977 |
| NRGN | 0.004972615 | -0.003899303 | 0.001073313 |
| PGRMC1 | 0.038955377 | 0.025006856 | 0.063962233 |
| IGFL4 | 0.002369569 | -0.002369569 | 0 |
| CCDC9 | 0.007693152 | -0.005061802 | 0.00263135 |
| AC009244.2 | 0.004392437 | -0.004018964 | 0.000373473 |
| SNHG32 | 0.026278554 | 0.034851446 | 0.06113 |
| ANP32A | 0.027616967 | 0.014915844 | 0.042532811 |
| GDF7 | -0.013057995 | 0.007553054 | -0.005504941 |
| SDF4 | 0.023611534 | 0.013767046 | 0.03737858 |
| GCDH | 0.019595992 | -0.013225158 | 0.006370833 |
| ASPSCR1 | 0.021672138 | -0.018396174 | 0.003275964 |
| SERPING1 | 0.020012163 | -0.013602868 | 0.006409295 |
| AC100810.1 | 0.028037414 | -0.021388805 | 0.006648609 |
| AC009509.1 | 0.002243716 | -0.002243716 | 0 |
| GCC2 | 0.027517737 | 0.037142487 | 0.064660224 |
| HLA-DQB1 | 0.012606829 | 0.013931931 | 0.02653876 |
| SMTNL2 | 0.019264643 | -0.010743204 | 0.008521439 |
| CYYR1 | 0.009913072 | -0.011359797 | -0.001446726 |
| TBX2-AS1 | 0.013229631 | -0.00780245 | 0.005427181 |
| AC023421.1 | 0.002739943 | -0.002739943 | 0 |
| PIGT | 0.017034748 | 0.014937353 | 0.031972101 |
| MPHOSPH8 | 0.022669369 | 0.01323623 | 0.035905599 |
| SERPINI1 | 0.010750763 | -0.007213726 | 0.003537038 |
| ACE2 | 0.024023544 | 0.025254749 | 0.049278292 |
| TUSC1 | 0.017569833 | 0.012038276 | 0.02960811 |
| DYNLT3 | 0.025636861 | -0.014605491 | 0.01103137 |
| PSMB10 | 0.016904552 | 0.021598371 | 0.038502923 |
| HAGHL | 0.008239266 | 0.004761778 | 0.013001043 |
| STOX1 | 0.007232141 | -0.004047772 | 0.003184369 |
| AVP | 0.019450025 | -0.034674113 | -0.015224088 |
| DMAC2 | 0.016791163 | -0.008896074 | 0.007895089 |
| PEX11G | 0.013193937 | -0.008099622 | 0.005094315 |
| FSCN1 | -0.008267187 | 0.009362131 | 0.001094945 |
| SSTR2 | 0.012501498 | -0.026742178 | -0.014240679 |
| DHRS7 | 0.02519483 | 0.021931424 | 0.047126254 |
| PIK3CD-AS2 | 0.013971029 | -0.012050659 | 0.001920369 |
| IAH1 | 0.023973709 | 0.014984185 | 0.038957895 |
| HES4 | -0.018064504 | 0.069959903 | 0.051895399 |
| MAGEF1 | 0.020588298 | 0.016416071 | 0.037004369 |
| DDX39B | 0.024456897 | 0.032049468 | 0.056506364 |
| LRRC28 | 0.018379464 | -0.011915012 | 0.006464451 |
| LRRC43 | 0.011016775 | -0.013980982 | -0.002964206 |
| GPR155 | 0.023918344 | -0.041562821 | -0.017644476 |
| GDF15 | 0.017132657 | 0.037852072 | 0.05498473 |
| RTP3 | 0.002121371 | -0.002121371 | 0 |

| | | | |
|---|---|---|---|
| GMPR | 0.013790428 | -0.007526817 | 0.006263611 |
| FGF9 | -0.007172356 | 0.006303834 | -0.000868522 |
| KMO | -0.020870904 | -0.018373188 | -0.039244092 |
| ZFPM1 | 0.015562906 | 0.023337041 | 0.038899948 |
| VIM | 0.042964837 | 0.136258603 | 0.17922344 |
| SURF2 | 0.021127884 | 0.012085368 | 0.033213252 |
| TRAF5 | -0.006179504 | 0.005591989 | -0.000587516 |
| COMMD2 | 0.019591187 | 0.021864962 | 0.041456149 |
| TMEM131 | 0.01480714 | -0.01043668 | 0.004370459 |
| ACSS3 | -0.017928729 | 0.010493986 | -0.007434743 |
| DLEU1 | 0.015719777 | -0.01161179 | 0.004107987 |
| AL662884.3 | 0.006125812 | -0.004378802 | 0.00174701 |
| GRAMD1B | 0.012465796 | -0.016342039 | -0.003876243 |
| TST | 0.037843335 | -0.063428975 | -0.02558564 |
| AFM | -0.018811462 | -0.012197829 | -0.031009291 |
| CALM2 | 0.038446782 | 0.043092534 | 0.081539317 |
| RASSF1 | 0.01034471 | -0.007095468 | 0.003249242 |
| PTGES3 | 0.036840268 | 0.041620305 | 0.078460572 |
| CD24 | 0.050506547 | 0.142665337 | 0.193171884 |
| SP100 | 0.017461323 | -0.009021849 | 0.008439473 |
| SET | 0.036679781 | 0.049021419 | 0.085701201 |
| RNLS | -0.015986563 | 0.008679052 | -0.007307512 |
| DPP3 | 0.013932645 | -0.00997641 | 0.003956235 |
| MON1A | 0.009453587 | -0.005840942 | 0.003612645 |
| SPINT2 | 0.041987997 | 0.088678891 | 0.130666887 |
| CDC25B | 0.008646866 | -0.007429254 | 0.001217612 |
| ZNF385B | 0.018306092 | 0.043326936 | 0.061633028 |
| LINC00504 | 0.002997867 | -0.003054595 | -5.67278E-05 |
| MED13L | -0.017206652 | 0.02061751 | 0.003410858 |
| ATAD1 | 0.016816019 | 0.023139598 | 0.039955617 |
| LPCAT3 | 0.016603342 | 0.008931309 | 0.025534651 |
| ADGRF4 | 0.003275768 | -0.004100408 | -0.000824641 |
| MT-ND2 | 0.028505251 | 0.030081667 | 0.058586918 |
| HEXIM1 | -0.022059024 | 0.031030526 | 0.008971502 |
| ADI1 | 0.04064561 | -0.075814943 | -0.035169333 |
| IQSEC2 | 0.01493568 | -0.012160351 | 0.002775329 |
| CDH6 | 0.025752599 | 0.056774416 | 0.082527015 |
| AATK | -0.006607081 | 0.009241411 | 0.00263433 |
| FAM167B | 0.00404453 | -0.002479063 | 0.001565467 |
| AC009107.2 | 0.010749342 | -0.011572573 | -0.000823231 |
| CNTF | 0.001993737 | -0.001993737 | 0 |
| SFT2D1 | 0.025505638 | 0.022927912 | 0.04843355 |
| THBD | -0.003545967 | 0.002711617 | -0.00083435 |
| LRRFIP2 | 0.016817305 | 0.012091099 | 0.028908404 |
| APMAP | 0.016095572 | 0.016508216 | 0.032603787 |
| SHANK2 | -0.013705827 | 0.011590685 | -0.002115141 |
| UBTD1 | 0.009908119 | -0.008161776 | 0.001746343 |
| RBM39 | 0.031385839 | 0.057643241 | 0.08902908 |
| ZEB2-AS1 | 0.006368995 | -0.005606376 | 0.000762619 |
| GMNN | 0.012335124 | -0.008825695 | 0.003509429 |

| | | | |
|---|---|---|---|
| AC103702.2 | -0.005350964 | 0.002815047 | -0.002535918 |
| AP003498.2 | -0.007978095 | 0.008585669 | 0.000607574 |
| SATB2 | -0.014165715 | 0.028937494 | 0.014771779 |
| LINC00623 | 0.016498794 | 0.009322989 | 0.025821783 |
| MACROD1 | 0.032870906 | -0.061006828 | -0.028135923 |
| ZNF266 | -0.006568128 | 0.007877587 | 0.001309459 |
| NIPAL3 | 0.013686643 | -0.00811122 | 0.005575423 |
| PLD3 | 0.020047108 | 0.012124481 | 0.032171589 |
| MYO10 | 0.010361977 | 0.005524806 | 0.015886783 |
| SLBP | 0.017844899 | 0.013692632 | 0.031537531 |
| ZNF350-AS1 | -0.004639635 | 0.005078577 | 0.000438942 |
| LINC02381 | -0.010523979 | 0.010331047 | -0.000192932 |
| ORMDL2 | 0.01439439 | 0.008882683 | 0.023277073 |
| TLN1 | 0.021839392 | 0.014534224 | 0.036373617 |
| FGF18 | 0.002402202 | -0.002278941 | 0.000123261 |
| GSN | 0.017822807 | 0.019326017 | 0.037148825 |
| PLEKHB1 | 0.006846711 | -0.00375542 | 0.003091291 |
| MRPL19 | 0.025162306 | -0.019327749 | 0.005834557 |
| RRAS | 0.014905048 | 0.016876447 | 0.031781496 |
| GPR146 | 0.006454453 | -0.003635044 | 0.002819409 |
| PLA2G12B | 0.009635495 | -0.005225904 | 0.004409591 |
| DRAM2 | 0.01565438 | 0.008650896 | 0.024305276 |
| AIMP1 | 0.024549842 | 0.023228799 | 0.047778641 |
| TPM4 | 0.015248723 | 0.017215012 | 0.032463735 |
| XBP1 | 0.022519521 | 0.030460773 | 0.052980294 |
| ACSF2 | 0.028637916 | -0.051355633 | -0.022717717 |
| PSMD2 | 0.022563431 | -0.012046875 | 0.010516556 |
| KLHL36 | 0.018184781 | 0.01424676 | 0.032431541 |
| MAPT | -0.023757384 | -0.036607722 | -0.060365105 |
| PLP2 | 0.012281659 | 0.015316156 | 0.027597815 |
| AP000688.2 | 0.004009227 | -0.002670884 | 0.001338342 |
| COMMD3 | 0.020643832 | 0.027332182 | 0.047976014 |
| GSTM4 | 0.019275681 | -0.015105696 | 0.004169984 |
| PRPF38B | 0.01615571 | 0.020210034 | 0.036365744 |
| FAM110A | 0.005169787 | -0.003029134 | 0.002140653 |
| ACMSD | 0.029962571 | -0.033282782 | -0.003320211 |
| NUDT9 | 0.017593811 | 0.013631784 | 0.031225595 |
| RGS18 | 0.002112801 | -0.002112801 | 1.77636E-15 |
| GLYAT | 0.038131847 | -0.025771641 | 0.012360206 |
| TNNC1 | 0.008608142 | -0.010403457 | -0.001795315 |
| SNW1 | 0.017125182 | 0.010997024 | 0.028122207 |
| IFITM1 | 0.003316461 | -0.002930506 | 0.000385955 |
| PGK1 | 0.034444099 | -0.021096941 | 0.013347157 |
| C16orf91 | 0.014049742 | 0.011234169 | 0.025283912 |
| LINC00240 | -0.005290282 | 0.004418197 | -0.000872085 |
| PAX2 | 0.024835757 | 0.0177436 | 0.042579356 |
| TUBB2A | 0.016727161 | 0.01312646 | 0.029853621 |
| CMBL | 0.040273492 | -0.078335831 | -0.038062339 |
| CTNNA1 | -0.029750971 | 0.021721869 | -0.008029102 |
| CBY1 | 0.017684899 | 0.014415014 | 0.032099912 |

| | | | |
|---|---|---|---|
| RSL1D1 | 0.027767151 | 0.027527419 | 0.05529457 |
| CD81 | 0.03633603 | 0.089036037 | 0.125372068 |
| SMARCE1 | 0.02245325 | 0.022929529 | 0.045382779 |
| THAP4 | 0.01931936 | -0.012880304 | 0.006439056 |
| CLCN7 | 0.017344721 | -0.010977129 | 0.006367592 |
| BLVRA | 0.015979392 | 0.010332947 | 0.026312339 |
| APOC3 | 0.021468758 | -0.039595492 | -0.018126734 |
| TMEM139 | 0.022163658 | 0.014758839 | 0.036922498 |
| PLPP3 | 0.01828395 | 0.019091715 | 0.037375665 |
| COLEC11 | -0.017832168 | -0.0143114 | -0.032143568 |
| KRTCAP3 | 0.010175479 | 0.006899737 | 0.017075216 |
| ERICH4 | 0.017486764 | -0.011709427 | 0.005777337 |
| TOMM40 | 0.015751007 | 0.008972146 | 0.024723153 |
| PAXX | 0.020328555 | 0.022124154 | 0.042452709 |
| SRGAP3 | 0.009362183 | -0.011612001 | -0.002249818 |
| B3GAT3 | 0.013791049 | 0.010105567 | 0.023896616 |
| TOGARAM1 | -0.013771223 | 0.011562324 | -0.0022089 |
| CITED2 | -0.027531191 | 0.077207812 | 0.049676621 |
| IDH3G | 0.025844491 | -0.015123666 | 0.010720825 |
| SDHA | 0.028421207 | 0.015937371 | 0.044358578 |
| MESD | 0.018063289 | 0.015490895 | 0.033554183 |
| MFSD12 | 0.020611022 | -0.024252244 | -0.003641222 |
| AC016924.1 | 0.009194099 | -0.007696164 | 0.001497935 |
| ITPA | 0.018953321 | 0.017335237 | 0.036288558 |
| LINC01831 | 0.001827591 | -0.001827591 | -1.77636E-15 |
| NUCB2 | 0.012821208 | 0.008965627 | 0.021786835 |
| NAP1L1 | 0.028404702 | 0.024585367 | 0.052990068 |
| FLVCR1-DT | 0.006084432 | -0.005809049 | 0.000275383 |
| ISYNA1 | 0.013396674 | 0.015453363 | 0.028850037 |
| STOX2 | -0.008539648 | 0.012726905 | 0.004187257 |
| COL18A1-AS1 | 0.005998789 | -0.007477756 | -0.001478967 |
| ACOT8 | 0.015919867 | -0.011037662 | 0.004882205 |
| PCGF2 | 0.019999807 | -0.017386725 | 0.002613082 |
| SHFL | 0.013275177 | 0.007933935 | 0.021209112 |
| MBNL3 | 0.011184376 | -0.006884944 | 0.004299432 |
| MVP | 0.024217488 | 0.017508156 | 0.041725644 |
| RARA-AS1 | 0.006678212 | -0.003921494 | 0.002756717 |
| DAB2 | 0.037288875 | -0.049911238 | -0.012622363 |
| GDA | -0.021844886 | -0.032635084 | -0.05447997 |
| MYL9 | 0.016004806 | 0.013379476 | 0.029384282 |
| PROZ | 0.007557595 | -0.008439203 | -0.000881608 |
| METRNL | 0.018194205 | 0.020136788 | 0.038330993 |
| HNRNPR | 0.022775639 | 0.015384235 | 0.038159874 |
| ANXA4 | 0.031509781 | 0.075947074 | 0.107456855 |
| CCT2 | 0.024429295 | 0.02346058 | 0.047889876 |
| ABHD14A | 0.023000419 | 0.027842234 | 0.050842653 |
| ANKRD44 | -0.006436423 | 0.005604148 | -0.000832274 |
| MMP14 | 0.012856363 | 0.016612606 | 0.029468969 |
| ZNF385D | 0.005767756 | -0.003336819 | 0.002430937 |
| THAP7 | 0.016931321 | -0.010814822 | 0.006116499 |

| | | | |
|---|---|---|---|
| AL031123.5 | 0.014542167 | -0.026813354 | -0.012271187 |
| HSPB8 | 0.013178579 | 0.016657725 | 0.029836304 |
| P4HTM | 0.015459611 | 0.017416946 | 0.032876556 |
| AC073174.1 | -0.013031413 | -0.01854089 | -0.031572303 |
| LINC00839 | -0.002617496 | 0.001934502 | -0.000682994 |
| HNRNPA2B1 | 0.035742894 | 0.03596405 | 0.071706944 |
| STAT5A | 0.011805705 | -0.008410888 | 0.003394816 |
| ELFN1 | -0.003174122 | 0.001882894 | -0.001291228 |
| SCOC-AS1 | 0.005214647 | -0.0029794 | 0.002235246 |
| SLCO4C1 | -0.022123982 | 0.017666311 | -0.004457672 |
| CHID1 | 0.021423758 | 0.029790925 | 0.051214683 |
| PALM2-AKAP2 | -0.016942726 | 0.015972657 | -0.000970069 |
| SRRM3 | 0.002080067 | -0.001760361 | 0.000319706 |
| SPPL2B | 0.014432504 | 0.011424238 | 0.025856742 |
| FMO1 | 0.031578547 | -0.035022607 | -0.00344406 |
| BCAR1 | 0.01819947 | 0.022405954 | 0.040605424 |
| NCL | 0.030824511 | 0.032770221 | 0.063594732 |
| FMO2 | 0.013201964 | 0.008614716 | 0.021816681 |
| TBC1D1 | 0.019784334 | -0.012986618 | 0.006797716 |
| ACO2 | 0.029560358 | -0.025548369 | 0.004011989 |
| WBP2 | 0.023628515 | -0.015433357 | 0.008195158 |
| NTPCR | 0.019602165 | 0.016922992 | 0.036525157 |
| CCL14 | 0.007104522 | -0.00486546 | 0.002239062 |
| IVD | 0.025463498 | -0.033516976 | -0.008053478 |
| AC020659.1 | -0.008084085 | 0.017453407 | 0.009369322 |
| CHD4 | 0.021637721 | 0.023752763 | 0.045390485 |
| TYMSOS | 0.016416007 | 0.010747303 | 0.02716331 |
| AL022726.1 | 0.003529123 | -0.003191204 | 0.000337919 |
| NEAT1 | 0.038457661 | 0.130257926 | 0.168715588 |
| GNB2 | 0.023998806 | 0.01760404 | 0.041602847 |
| ZNF576 | 0.012625858 | -0.007439172 | 0.005186686 |
| BMERB1 | 0.011354918 | -0.007154474 | 0.004200444 |
| OIP5-AS1 | 0.020103018 | 0.016001564 | 0.036104582 |
| ERGIC2 | 0.017651402 | 0.015600035 | 0.033251437 |
| SLC22A23 | 0.009645322 | -0.017559591 | -0.007914269 |
| AUH | 0.02179256 | -0.017728678 | 0.004063882 |
| PDIA3 | 0.02667804 | 0.034171211 | 0.06084925 |
| MPND | 0.014060633 | -0.013752767 | 0.000307866 |
| AXL | -0.004787592 | 0.004353644 | -0.000433947 |
| RITA1 | 0.015236178 | -0.014066672 | 0.001169506 |
| SLC2A9 | 0.012624458 | -0.00906414 | 0.003560318 |
| C2CD4C | 0.002293631 | -0.00197663 | 0.000317 |
| HDLBP | 0.026670043 | -0.016788157 | 0.009881886 |
| DAZAP2 | -0.028053544 | 0.025426513 | -0.002627031 |
| RASGRP1 | -0.009081909 | 0.009035144 | -4.67649E-05 |
| ENTPD3-AS1 | 0.01703933 | -0.02001338 | -0.00297405 |
| PINLYP | 0.007444822 | -0.004592028 | 0.002852793 |
| ACADSB | -0.025681062 | -0.026206332 | -0.051887394 |
| NOCT | 0.005100963 | -0.003337803 | 0.00176316 |
| H2AFY2 | 0.011110578 | 0.007448826 | 0.018559403 |

| | | | |
|---|---|---|---|
| AP000355.1 | 0.002015195 | -0.001507894 | 0.000507301 |
| WIPI2 | 0.020034271 | -0.013097963 | 0.006936308 |
| HRAS | 0.017348483 | 0.020182098 | 0.037530581 |
| PAPSS1 | 0.014307499 | 0.012088507 | 0.026396006 |
| VWA1 | 0.010399983 | 0.010575262 | 0.020975244 |
| ZNF22 | 0.015796196 | 0.020578103 | 0.036374299 |
| OLA1 | 0.020682464 | 0.015014448 | 0.035696912 |
| POLD3 | 0.015689537 | -0.019334051 | -0.003644514 |
| CHRDL1 | 0.006966078 | -0.011380017 | -0.004413939 |
| PLG | 0.028903805 | -0.076784168 | -0.047880363 |
| CTSF | 0.019187979 | 0.025379426 | 0.044567406 |
| LRRC4C | -0.006216256 | 0.007014938 | 0.000798681 |
| BSG | 0.035657643 | 0.086022521 | 0.121680164 |
| SPINT1 | 0.020797935 | 0.03374991 | 0.054547845 |
| PIPOX | 0.026235801 | -0.024275419 | 0.001960381 |
| AC099552.1 | 0.001519398 | -0.001519398 | 0 |
| SFTA2 | 0.008335918 | -0.00795825 | 0.000377668 |
| PLSCR1 | 0.016418832 | 0.018100808 | 0.03451964 |
| CKAP2 | 0.00832753 | -0.005495265 | 0.002832264 |
| PPT1 | 0.015327098 | 0.015865725 | 0.031192823 |
| UBA6 | -0.012294615 | 0.009585787 | -0.002708829 |
| AL807752.2 | 0.001991372 | -0.001631475 | 0.000359897 |
| MYLK4 | 0.007432072 | -0.009330223 | -0.001898152 |
| NRG1 | -0.012933371 | 0.029958191 | 0.017024819 |
| THUMPD3-AS1 | 0.017413594 | 0.024268027 | 0.041681621 |
| CBX5 | 0.021903232 | 0.021030728 | 0.042933961 |
| PRKCH | 0.003011951 | -0.002960333 | 5.16181E-05 |
| ZFYVE27 | 0.008086183 | 0.005650024 | 0.013736206 |
| SERINC2 | 0.019097075 | 0.034229677 | 0.053326751 |
| ACO1 | -0.029393853 | -0.084310068 | -0.113703921 |
| NOVA1 | 0.014529183 | 0.016696579 | 0.031225762 |
| SCIN | 0.020303818 | 0.027591324 | 0.047895143 |
| ZNF551 | -0.00420645 | 0.002474089 | -0.001732361 |
| FHOD3 | 0.013041108 | 0.015741191 | 0.028782299 |
| PSMB9 | 0.013312542 | 0.022072475 | 0.035385017 |
| CKLF | 0.012338569 | 0.010167094 | 0.022505663 |
| PMS2 | 0.013021556 | -0.008169787 | 0.00485177 |
| SDHB | 0.027394755 | -0.053493578 | -0.026098823 |
| RPL7 | 0.034824813 | 0.059109447 | 0.09393426 |
| AP1M2 | -0.026459434 | -0.017477773 | -0.043937207 |
| ERP44 | 0.015974601 | 0.01726727 | 0.033241871 |
| ACP5 | 0.022762445 | -0.051099094 | -0.028336649 |
| EFS | -0.004677648 | 0.006508048 | 0.0018304 |
| RCN2 | 0.019918161 | 0.028102071 | 0.048020232 |
| RPS11 | 0.033728877 | 0.020073652 | 0.053802529 |
| NEFL | -0.00431802 | 0.004903669 | 0.000585649 |
| RTCA | 0.01955275 | -0.01181686 | 0.00773589 |
| PTGER2 | -0.005701401 | 0.007542406 | 0.001841005 |
| CCR1 | -0.007338147 | -0.006979042 | -0.01431719 |
| TRIM38 | 0.016025841 | 0.024128447 | 0.040154288 |

| | | | |
|---|---|---|---|
| DNAJC21 | 0.019697236 | 0.017513891 | 0.037211126 |
| KIAA1522 | 0.015825842 | 0.00958759 | 0.025413432 |
| AC022424.1 | -0.009112619 | 0.024287464 | 0.015174845 |
| MACF1 | 0.023079295 | 0.033595567 | 0.056674863 |
| FTCD | 0.035106715 | -0.050223565 | -0.01511685 |
| ADAMTS15 | -0.00620581 | 0.00565841 | -0.0005474 |
| CD59 | 0.032246344 | 0.055938204 | 0.088184549 |
| ILVBL | 0.01873565 | 0.011848422 | 0.030584072 |
| RAB4B | 0.009173813 | -0.006916093 | 0.00225772 |
| LRRTM1 | 0.001815032 | -0.001619723 | 0.000195308 |
| SAMD5 | 0.012248331 | -0.010794579 | 0.001453752 |
| SIRPA | -0.014160659 | 0.012706793 | -0.001453866 |
| CBX2 | 0.002652526 | -0.002111225 | 0.0005413 |
| MTURN | 0.015614856 | 0.013006913 | 0.028621769 |
| PBX1 | -0.021199588 | 0.0150132 | -0.006186388 |
| MACC1 | -0.018930276 | 0.086135693 | 0.067205417 |
| ARHGAP1 | 0.015312149 | 0.0154127 | 0.030724849 |
| UBALD1 | 0.013063023 | -0.008159525 | 0.004903498 |
| PLBD1 | 0.012046784 | 0.024809611 | 0.036856395 |
| MEAF6 | 0.020096419 | 0.017137652 | 0.037234071 |
| EPS15L1 | 0.009698045 | -0.006774004 | 0.002924041 |
| CLDN7 | 0.024611308 | 0.089862555 | 0.114473863 |
| NEBL | -0.013224891 | 0.030907411 | 0.01768252 |
| CNDP2 | 0.032326187 | -0.045341851 | -0.013015664 |
| TATDN1 | 0.020070521 | 0.019342444 | 0.039412965 |
| TCL6 | 0.018853288 | -0.034104366 | -0.015251077 |
| AC004816.1 | 0.004943882 | -0.004497025 | 0.000446857 |
| MOB1A | 0.025672631 | -0.016119706 | 0.009552925 |
| NOX4 | -0.024733596 | -0.057080266 | -0.081813862 |
| THOC2 | 0.017392079 | 0.015732946 | 0.033125025 |
| TM2D1 | 0.016369988 | 0.011285821 | 0.027655809 |
| VEGFA | 0.020398999 | -0.017585196 | 0.002813804 |
| IMPDH2 | 0.015729015 | 0.012698525 | 0.02842754 |
| DCUN1D5 | 0.013553953 | 0.008774726 | 0.022328679 |
| ESPN | 0.0220242 | -0.030386903 | -0.008362702 |
| HLA-B | 0.033606758 | 0.139143082 | 0.17274984 |
| TBX2 | 0.017263197 | -0.017962386 | -0.00069919 |
| AC025171.2 | -0.004029303 | 0.003837736 | -0.000191567 |
| IL17RB | 0.014355863 | 0.011597986 | 0.025953849 |
| SPAG5 | 0.010069685 | -0.010374629 | -0.000304944 |
| SSC4D | 0.002817203 | -0.001893834 | 0.00092337 |
| C1orf56 | 0.01327897 | 0.009956908 | 0.023235878 |
| PTOV1 | 0.020623149 | -0.017209225 | 0.003413924 |
| KCNMB2 | -0.006416821 | 0.006406304 | -1.0516E-05 |
| ENPP6 | -0.01144529 | -0.009208552 | -0.020653842 |
| ALDOC | 0.004785238 | -0.004745782 | 3.94557E-05 |
| HIST2H4A | 0.001581144 | -0.001325396 | 0.000255748 |
| UBE2N | 0.020188044 | 0.013767867 | 0.033955911 |
| SLC50A1 | 0.011767661 | 0.007511398 | 0.019279059 |
| CCNL2 | 0.016677197 | 0.020796302 | 0.037473498 |

| | | | |
|---|---|---|---|
| AC009159.3 | 0.00438905 | -0.003530511 | 0.000858539 |
| SOX9-AS1 | 0.01236571 | -0.008108638 | 0.004257072 |
| MIR210HG | 0.005985336 | -0.006086972 | -0.000101636 |
| SLC44A4 | 0.018147578 | 0.023335284 | 0.041482861 |
| AC079848.2 | -0.003730011 | 0.00308293 | -0.000647081 |
| WFDC3 | 0.00396613 | -0.00248994 | 0.001476191 |
| GLT8D1 | 0.013159058 | 0.01213708 | 0.025296138 |
| BMP7 | -0.009760407 | 0.027403252 | 0.017642844 |
| KCNJ13 | 0.002748111 | -0.002072987 | 0.000675124 |
| AC010913.1 | 0.015202346 | -0.026533598 | -0.011331252 |
| LRRC29 | 0.007490217 | -0.004905205 | 0.002585012 |
| LEPROTL1 | 0.016877419 | 0.010559189 | 0.027436608 |
| AL451165.2 | 0.012192213 | -0.013436814 | -0.001244601 |
| GPX2 | 0.002277138 | -0.00196437 | 0.000312768 |
| APH1A | 0.019215523 | 0.014909323 | 0.034124847 |
| CD2AP | 0.022061015 | 0.014932539 | 0.036993554 |
| CERK | 0.011589894 | -0.009802643 | 0.00178725 |
| CASP8 | 0.011741658 | 0.009843122 | 0.02158478 |
| PFKFB3 | 0.011447129 | -0.023518043 | -0.012070914 |
| SON | 0.027994908 | 0.028190921 | 0.05618583 |
| HYAL2 | 0.011368809 | 0.014394959 | 0.025763768 |
| NASP | 0.015758029 | 0.010607648 | 0.026365677 |
| C9orf40 | 0.005443151 | -0.00373472 | 0.001708432 |
| TBX3 | 0.002760214 | -0.001775327 | 0.000984886 |
| RUBCNL | -0.002438317 | 0.001960483 | -0.000477834 |
| SRSF11 | 0.021756853 | 0.036001538 | 0.057758391 |
| TP53I13 | 0.016401601 | 0.03640085 | 0.052802451 |
| PLGRKT | 0.017236701 | 0.016979529 | 0.034216229 |
| WAPL | -0.015833871 | 0.017859193 | 0.002025322 |
| ACADL | -0.014443907 | 0.016859162 | 0.002415256 |
| IL18 | 0.010465487 | 0.014281545 | 0.024747033 |
| GOLGB1 | 0.024360011 | 0.042166321 | 0.066526331 |
| AL360169.3 | 0.010888888 | -0.028034143 | -0.017145255 |
| PCNP | 0.021221613 | 0.026640516 | 0.04786213 |
| SEMA5A | 0.018388359 | 0.046216262 | 0.064604621 |
| AC003984.1 | -0.006218328 | 0.013243081 | 0.007024753 |
| GRTP1 | 0.014528881 | 0.009954528 | 0.024483409 |
| SLC22A5 | 0.016008761 | -0.014326371 | 0.00168239 |
| MT1M | 0.012747069 | 0.021050409 | 0.033797477 |
| GRN | 0.025909893 | 0.026354559 | 0.052264452 |
| KDELR2 | 0.022354315 | 0.028569524 | 0.05092384 |
| ADGRF2 | 0.002454827 | -0.002990493 | -0.000535666 |
| LRATD2 | -0.018148603 | 0.015047046 | -0.003101558 |
| XAB2 | 0.018520017 | -0.024374741 | -0.005854724 |
| WNT7B | 0.006429095 | -0.013887141 | -0.007458046 |
| NFIB | 0.025943108 | 0.024329083 | 0.05027219 |
| TRIM45 | -0.005099906 | 0.00524256 | 0.000142654 |
| RBFA | 0.013560535 | 0.009260452 | 0.022820987 |
| SCOC | 0.024028474 | 0.020274653 | 0.044303127 |
| VMO1 | 0.003506977 | -0.002475405 | 0.001031572 |

| | | | |
|---|---|---|---|
| PAGR1 | 0.015041234 | 0.014008037 | 0.02904927 |
| TCHH | 0.001578459 | -0.001578459 | 0 |
| METTL7B | 0.010466215 | -0.007313017 | 0.003153198 |
| KCNAB2 | 0.013183624 | -0.026905547 | -0.013721923 |
| MEG8 | 0.007316 | 0.008352371 | 0.01566837 |
| LINC02062 | -0.003468188 | 0.002281584 | -0.001186604 |
| ZYX | 0.01181014 | 0.011614311 | 0.023424451 |
| OGA | -0.02081043 | 0.079070181 | 0.058259751 |
| SF3A2 | 0.009766592 | 0.007735964 | 0.017502556 |
| MSMO1 | 0.017204084 | 0.01288432 | 0.030088403 |
| KIF9 | 0.020303851 | 0.016332569 | 0.03663642 |
| SMIM15 | 0.018554588 | 0.012572276 | 0.031126864 |
| FNDC11 | 0.001603716 | -0.001496634 | 0.000107082 |
| MT2A | -0.041339372 | 0.1557626 | 0.114423228 |
| MITD1 | 0.012265407 | 0.009462925 | 0.021728333 |
| NRIP1 | -0.020075381 | 0.037135057 | 0.017059676 |
| GARS-DT | -0.009696907 | 0.014327787 | 0.00463088 |
| JUN | 0.024523647 | 0.04189727 | 0.066420916 |
| SNRNP200 | 0.017858915 | 0.016898327 | 0.034757242 |
| SEMA5B | 0.006575898 | 0.007557736 | 0.014133634 |
| BPHL | 0.027153997 | -0.03782339 | -0.010669393 |
| PSME3 | -0.011305282 | 0.007319709 | -0.003985573 |
| LMO3 | 0.002273961 | -0.001981475 | 0.000292486 |
| C12orf75 | 0.029388665 | 0.044571035 | 0.0739597 |
| RASSF7 | 0.017422606 | 0.021644804 | 0.03906741 |
| GTF2H2 | -0.009603756 | 0.014044031 | 0.004440275 |
| MAOA | 0.024849041 | -0.078069545 | -0.053220504 |
| BLOC1S2 | 0.015512604 | 0.010027419 | 0.025540023 |
| ZNF687-AS1 | 0.004083113 | -0.00309427 | 0.000988843 |
| MAP1S | 0.008385372 | 0.005647109 | 0.01403248 |
| CAPZA2 | 0.025321286 | 0.020376515 | 0.045697801 |
| CTAG2 | 0.001459851 | -0.001459851 | 0 |
| ACOX2 | 0.016857585 | -0.028009537 | -0.011151952 |
| DDR2 | 0.004707299 | -0.003928608 | 0.000778691 |
| FAIM | 0.013527327 | 0.023950037 | 0.037477363 |
| MAPKAPK5-AS1 | 0.010105624 | 0.009863754 | 0.019969377 |
| CD320 | 0.013386245 | 0.011167986 | 0.024554231 |
| SARS2 | 0.008251601 | -0.006269787 | 0.001981814 |
| ARID4B | 0.01476464 | 0.012025531 | 0.026790171 |
| TTC39B | 0.014265037 | -0.018564306 | -0.004299268 |
| HMGN4 | 0.016461315 | 0.023368664 | 0.039829979 |
| CTTN | 0.021366082 | 0.022985121 | 0.044351204 |
| GPAA1 | 0.020672729 | 0.02788586 | 0.048558589 |
| SLC52A2 | 0.009926306 | 0.008887947 | 0.018814253 |
| APCS | 0.001422156 | -0.001422156 | 0 |
| ILKAP | 0.013111436 | 0.009569006 | 0.022680442 |
| PREX2 | 0.002172533 | -0.001710583 | 0.00046195 |
| PCDH7 | 0.002096476 | -0.001857509 | 0.000238967 |
| TUBB | 0.024641752 | 0.049049474 | 0.073691226 |
| CDH2 | 0.020022109 | 0.023941398 | 0.043963507 |

| | | | |
|---|---|---|---|
| Z99572.1 | -0.003576883 | 0.003511679 | -6.52037E-05 |
| AL031726.2 | 0.001274267 | -0.001274267 | 1.77636E-15 |
| EYA3 | 0.011408259 | 0.011402976 | 0.022811235 |
| CCDC74A | 0.006220956 | 0.004519031 | 0.010739987 |
| ENC1 | -0.008013819 | 0.015288823 | 0.007275005 |
| VTI1B | 0.026007025 | -0.030548971 | -0.004541946 |
| NTS | 0.002203707 | -0.00230316 | -9.94532E-05 |
| CTSZ | 0.019142592 | 0.024020965 | 0.043163557 |
| B3GAT1 | 0.003767059 | -0.003055777 | 0.000711282 |
| GCA | 0.014245916 | 0.017457216 | 0.031703131 |
| TXNIP | 0.031773205 | 0.123536901 | 0.155310106 |
| NMRK1 | 0.019941861 | -0.026293516 | -0.006351655 |
| ZNRF3 | -0.012422651 | 0.018764522 | 0.006341871 |
| OGFRP1 | 0.00192422 | -0.001481368 | 0.000442852 |
| CNOT2 | 0.014194009 | 0.010108194 | 0.024302204 |
| TLE5 | 0.024946837 | 0.023395686 | 0.048342523 |
| PABPC4L | -0.003878244 | 0.005646441 | 0.001768197 |
| SHBG | 0.006185279 | -0.007976191 | -0.001790913 |
| AL136038.5 | 0.014588213 | -0.014132605 | 0.000455608 |
| ATP6AP2 | 0.023003445 | 0.047603805 | 0.07060725 |
| DICER1 | -0.016032552 | 0.022664425 | 0.006631873 |
| ICA1 | 0.006971624 | 0.008322681 | 0.015294305 |
| CRTAP | 0.019507958 | 0.013688252 | 0.03319621 |
| HPGD | 0.022782791 | -0.071399187 | -0.048616395 |
| SETBP1 | 0.0118195 | -0.008429862 | 0.003389638 |
| SCAMP2 | 0.01844989 | 0.017772013 | 0.036221904 |
| PRKAA2 | -0.020590649 | 0.032924082 | 0.012333433 |
| AP001269.1 | 0.002910896 | -0.002764213 | 0.000146684 |
| ALDH1B1 | 0.020610999 | -0.042556534 | -0.021945534 |
| TMEM35B | 0.018289893 | 0.048188846 | 0.066478739 |
| LINC00310 | 0.001783351 | -0.001345297 | 0.000438054 |
| MLLT3 | 0.008492289 | 0.006721198 | 0.015213487 |
| SGPP2 | 0.009376555 | 0.013254932 | 0.022631488 |
| CYP51A1 | 0.015025738 | 0.027769486 | 0.042795223 |
| CX3CL1 | 0.014029205 | 0.029299299 | 0.043328505 |
| CDNF | 0.004608977 | -0.003717448 | 0.000891529 |
| ENTPD4 | 0.007598558 | -0.005324199 | 0.002274359 |
| KIF5B | 0.019253238 | 0.015913218 | 0.035166456 |
| NPL | 0.023291236 | -0.068132592 | -0.044841356 |
| GNMT | -0.002843835 | 0.002315535 | -0.0005283 |
| CRELD1 | 0.009560024 | 0.012139583 | 0.021699606 |
| BST1 | 0.007968341 | 0.005746648 | 0.013714988 |
| SLC41A3 | 0.011032469 | 0.010415831 | 0.0214483 |
| RETREG1 | 0.017114427 | 0.037970525 | 0.055084952 |
| SLC2A3 | 0.001352447 | -0.001169206 | 0.000183241 |
| PCDH17 | 0.001800669 | -0.001711819 | 8.88494E-05 |
| TBC1D10A | 0.00992426 | -0.01164331 | -0.00171905 |
| CHPF | 0.011772447 | 0.029596936 | 0.041369383 |
| DOCK8-AS1 | 0.021573065 | -0.047552738 | -0.025979672 |
| GNAQ | -0.014960506 | 0.027705644 | 0.012745138 |

| | | | |
|---|---|---|---|
| FBXO48 | 0.005861716 | -0.00532504 | 0.000536675 |
| PPARGC1A | -0.013578159 | 0.019899265 | 0.006321106 |
| TMOD1 | 0.001497019 | -0.001283155 | 0.000213864 |
| RBM17 | 0.016656059 | 0.017649395 | 0.034305454 |
| TRA2B | -0.016474767 | 0.022727999 | 0.006253232 |
| B3GNT3 | 0.007088333 | 0.00779266 | 0.014880993 |
| KDR | 0.002681734 | -0.00209561 | 0.000586123 |
| PDLIM1 | 0.014715804 | 0.011103175 | 0.025818979 |
| MYOF | 0.010002512 | 0.008284934 | 0.018287446 |
| SLC39A13 | 0.012184682 | 0.010383231 | 0.022567914 |
| EFNB1 | 0.007932211 | 0.008795741 | 0.016727953 |
| PLEKHA2 | -0.010445313 | 0.013204389 | 0.002759077 |
| SEL1L3 | -0.0105573 | 0.031416279 | 0.020858979 |
| MINCR | 0.010995697 | -0.015338738 | -0.004343041 |
| ZNF90 | -0.00441403 | 0.00431113 | -0.000102899 |
| SLC22A11 | 0.016453213 | -0.01608516 | 0.000368053 |
| AP000345.2 | 0.002912709 | -0.002189309 | 0.0007234 |
| SF3B2 | 0.019094787 | 0.021535362 | 0.04063015 |
| ASS1 | 0.035087112 | -0.160030679 | -0.124943567 |
| SYNDIG1 | 0.007895577 | -0.008118694 | -0.000223117 |
| CALML3 | 0.020504878 | -0.075949888 | -0.055445009 |
| TGFBR3L | 0.006716428 | -0.005258944 | 0.001457484 |
| PRCP | 0.016112636 | 0.022036555 | 0.038149191 |
| CYP4F2 | 0.010388125 | -0.020607843 | -0.010219718 |
| SQSTM1 | 0.028651587 | 0.0516345 | 0.080286087 |
| MEIOB | 0.002068651 | -0.001557805 | 0.000510846 |
| CYP46A1 | 0.005791024 | -0.005822477 | -3.14525E-05 |
| ADAMTS9 | 0.010812269 | 0.019305551 | 0.03011782 |
| CSNK1A1 | 0.021350221 | 0.014475772 | 0.035825993 |
| WIPI1 | 0.013171021 | -0.014658917 | -0.001487896 |
| TFRC | -0.011978897 | 0.0287555 | 0.016776603 |
| MED30 | 0.009597148 | 0.007862621 | 0.017459769 |
| SLC9A3 | 0.003541399 | -0.003287454 | 0.000253945 |
| TMEM174 | 0.020074354 | -0.034398258 | -0.014323904 |
| GUCY1A1 | 0.009324833 | 0.011570441 | 0.020895274 |
| LTC4S | 0.011875174 | 0.017075337 | 0.028950511 |
| CDK18 | 0.014473841 | -0.020156325 | -0.005682483 |
| RBM3 | 0.022410428 | 0.063769742 | 0.08618017 |
| LHFPL3-AS2 | 0.016064055 | 0.018904536 | 0.034968591 |
| RESF1 | -0.011486951 | 0.022744899 | 0.011257947 |
| DM1-AS | 0.006469843 | -0.005932533 | 0.000537309 |
| BCAM | 0.027263787 | 0.038577198 | 0.065840985 |
| AC015802.6 | 0.005615922 | -0.004527614 | 0.001088308 |
| FAM120A | -0.02124333 | 0.01776355 | -0.00347978 |
| EMP3 | 0.010948134 | 0.009937947 | 0.020886082 |
| CLUAP1 | 0.012877579 | 0.011132626 | 0.024010204 |
| RNF130 | 0.017508521 | 0.027670119 | 0.04517864 |
| ATXN2L | 0.011938722 | 0.009881387 | 0.021820108 |
| C10orf143 | 0.006763058 | -0.004627251 | 0.002135807 |
| HACD3 | 0.017744591 | 0.019463007 | 0.037207598 |

| | | | |
|---|---|---|---|
| GLB1 | 0.013975366 | 0.018155588 | 0.032130954 |
| MARS | 0.008220417 | 0.006225082 | 0.014445498 |
| AC026471.3 | 0.00626914 | -0.006379921 | -0.000110781 |
| LINC01315 | 0.008394552 | 0.011165145 | 0.019559697 |
| ZKSCAN1 | 0.016480717 | 0.013184316 | 0.029665033 |
| YIPF3 | 0.018687561 | 0.02961079 | 0.048298351 |
| NAGLU | 0.014262281 | -0.017227438 | -0.002965157 |
| SYNE2 | 0.029597937 | 0.062503274 | 0.092101211 |
| GLDC | -0.014135652 | 0.024050632 | 0.00991498 |
| B4GALT1 | 0.017679687 | 0.046238403 | 0.06391809 |
| AC068987.1 | 0.001505648 | -0.001073693 | 0.000431955 |
| FOLH1 | 0.009675352 | -0.009051875 | 0.000623477 |
| NMB | 0.011855725 | 0.013226877 | 0.025082603 |
| UQCRC1 | 0.025199343 | -0.017603884 | 0.007595459 |
| RBP1 | 0.007382191 | 0.005339762 | 0.012721953 |
| LY86-AS1 | 0.001645822 | -0.001662095 | -1.62723E-05 |
| EMC10 | 0.018750305 | 0.022373 | 0.041123305 |
| AC068672.2 | 0.001293958 | -0.001293958 | 0 |
| NOL3 | 0.009627429 | 0.007604079 | 0.017231509 |
| FUCA2 | 0.017134984 | 0.020349716 | 0.0374847 |
| RNF43 | -0.00823299 | 0.011179605 | 0.002946616 |
| GRHL2 | -0.005898419 | 0.006269498 | 0.000371079 |
| STARD10 | 0.016803173 | -0.01442765 | 0.002375523 |
| ABCC2 | -0.011780996 | -0.014246508 | -0.026027504 |
| GOT2 | 0.014828623 | -0.014348467 | 0.000480156 |
| TPD52L2 | 0.013147932 | 0.010518892 | 0.023666824 |
| EHHADH | -0.025085925 | -0.033284093 | -0.058370017 |
| DERL1 | 0.012179955 | 0.013890082 | 0.026070037 |
| FRAS1 | 0.011915637 | 0.024056328 | 0.035971965 |
| TWSG1 | 0.009864212 | 0.008127396 | 0.017991609 |
| TMEM248 | 0.015973638 | 0.025575642 | 0.04154928 |
| MOGS | 0.010639258 | 0.008884842 | 0.019524101 |
| SRSF4 | 0.01390157 | 0.016605353 | 0.030506922 |
| KNG1 | -0.014105403 | 0.028464287 | 0.014358885 |
| TK1 | 0.006274092 | -0.008963116 | -0.002689024 |
| FAT1 | 0.02332518 | 0.038610977 | 0.061936157 |
| HSD17B12 | 0.016376493 | 0.011565315 | 0.027941808 |
| CARMIL1 | 0.011719445 | 0.018954287 | 0.030673731 |
| LMO7-AS1 | 0.004600979 | -0.003645222 | 0.000955757 |
| DCTD | 0.01365122 | 0.009837548 | 0.023488767 |
| HIPK2 | -0.023225521 | 0.017643747 | -0.005581774 |
| UGT2A3 | -0.027098863 | 0.061172612 | 0.034073749 |
| CCDC28B | 0.006100572 | 0.004944467 | 0.011045038 |
| ASB9 | 0.013103231 | -0.020329625 | -0.007226394 |
| SLC2A4RG | 0.026431891 | 0.077999123 | 0.104431014 |
| MCAT | 0.015463287 | -0.016476651 | -0.001013364 |
| SLC39A3 | 0.01124813 | 0.010132075 | 0.021380205 |
| UNC5CL | 0.009333251 | 0.008798792 | 0.018132044 |
| DMPK | 0.011212119 | -0.020859938 | -0.00964782 |
| RPH3AL | -0.005910153 | 0.007523794 | 0.00161364 |

| | | | |
|---|---|---|---|
| HNRNPA3 | 0.023748693 | 0.037131036 | 0.060879729 |
| DMGDH | 0.019837815 | -0.025875458 | -0.006037643 |
| MICU1 | 0.019414521 | -0.031830645 | -0.012416124 |
| PIK3CB | -0.014226121 | 0.019785885 | 0.005559763 |
| SCAP | 0.011237621 | 0.012640567 | 0.023878188 |
| AC096577.1 | 0.00749236 | 0.005347383 | 0.012839743 |
| ANG | 0.011210567 | 0.023660936 | 0.034871503 |
| ESCO1 | 0.01179199 | 0.011599152 | 0.023391142 |
| LIMS2 | -0.005838015 | 0.008464555 | 0.00262654 |
| TMEM106C | 0.016087169 | 0.016871363 | 0.032958531 |
| TMEM38B | 0.016837805 | 0.049114662 | 0.065952467 |
| CMTM8 | 0.009018559 | -0.006784095 | 0.002234464 |
| POLE3 | 0.012929653 | 0.009487422 | 0.022417075 |
| AC091729.3 | 0.007248452 | -0.008144029 | -0.000895577 |
| SLFN11 | 0.001255175 | -0.001069261 | 0.000185915 |
| GALNT4 | 0.008918375 | 0.006442839 | 0.015361214 |
| TNRC18 | -0.014136075 | 0.02477489 | 0.010638814 |
| C4orf48 | 0.009024903 | 0.019626362 | 0.028651265 |
| CCDC28A | 0.017215873 | -0.017239821 | -2.39472E-05 |
| ARMC7 | 0.006018238 | -0.004751411 | 0.001266827 |
| MAP9 | 0.012642908 | 0.013634381 | 0.026277289 |
| GUCY1B1 | 0.009044502 | 0.010750443 | 0.019794946 |
| NELFE | 0.015343056 | 0.017040702 | 0.032383757 |
| EEF1AKMT1 | 0.009457366 | 0.007345456 | 0.016802822 |
| MAP7 | 0.020012825 | -0.021683124 | -0.001670299 |
| MRPL37 | 0.016170058 | -0.012799193 | 0.003370866 |
| HOOK2 | 0.015552792 | 0.014752853 | 0.030305645 |
| SYCP2 | 0.006897494 | -0.01433904 | -0.007441546 |
| AC011377.1 | 0.000922456 | -0.000922456 | 0 |
| IKBIP | 0.008579084 | 0.010114738 | 0.018693822 |
| GPR173 | -0.002668112 | 0.002998237 | 0.000330125 |
| TEX41 | -0.005324012 | 0.01067369 | 0.005349678 |
| IL20RA | -0.002900251 | 0.003660628 | 0.000760378 |
| SLC25A10 | 0.020430339 | -0.051340362 | -0.030910023 |
| RNF13 | 0.016598669 | 0.027128677 | 0.043727347 |
| KIDINS220 | 0.01372088 | 0.016187548 | 0.029908428 |
| AL121839.2 | -0.003004681 | 0.00309488 | 9.01994E-05 |
| GNAS | 0.028601494 | 0.037433053 | 0.066034547 |
| PLAU | -0.007457556 | 0.011400879 | 0.003943323 |
| GTF2H2C | -0.006768225 | 0.005230116 | -0.001538109 |
| GLTPD2 | 0.016763454 | -0.014130518 | 0.002632936 |
| RNF135 | 0.012432741 | -0.015787132 | -0.003354391 |
| AC063919.2 | 0.001368544 | -0.001115579 | 0.000252965 |
| PCSK5 | 0.006221101 | -0.004993973 | 0.001227128 |
| NAPRT | 0.021044691 | -0.021225003 | -0.000180312 |
| XKR4 | -0.003237613 | 0.005209108 | 0.001971495 |
| MTDH | 0.018895378 | 0.019411593 | 0.038306971 |
| ELOVL4 | 0.00238868 | -0.002379794 | 8.8853E-06 |
| REEP6 | 0.014115157 | -0.023600414 | -0.009485257 |
| ASIP | 0.002183083 | -0.002691274 | -0.000508191 |

| | | | |
|---|---|---|---|
| BOC | 0.00548931 | -0.006543274 | -0.001053964 |
| RIN3 | 0.005370086 | -0.004147516 | 0.001222571 |
| LRRC8C | 0.002231367 | -0.002289856 | -5.84887E-05 |
| BTN3A2 | 0.005563544 | 0.004821394 | 0.010384938 |
| LGI2 | -0.010381826 | -0.010375035 | -0.02075686 |
| STAT1 | 0.015128327 | 0.045754097 | 0.060882424 |
| ARHGAP29 | 0.021335835 | 0.059101649 | 0.080437484 |
| SRSF7 | 0.019685901 | 0.044574579 | 0.064260481 |
| NKAIN3 | 0.001132634 | -0.00101933 | 0.000113304 |
| AL121999.1 | 0.001010034 | -0.001010034 | 0 |
| CA8 | 0.002017741 | -0.001718608 | 0.000299132 |
| STOML1 | 0.005582811 | 0.004664218 | 0.010247029 |
| RASGRF1 | 0.003089392 | -0.002798515 | 0.000290877 |
| P4HA2 | 0.013505617 | -0.019599132 | -0.006093514 |
| FASTKD5 | 0.008964982 | -0.009016316 | -5.13334E-05 |
| SH3BP2 | 0.012708179 | -0.022545933 | -0.009837754 |
| TKFC | 0.019700092 | -0.029389506 | -0.009689414 |
| SLC20A2 | 0.00864155 | 0.006383458 | 0.015025009 |
| AL109976.1 | 0.002310132 | -0.00224606 | 6.40718E-05 |
| ESR2 | 0.001320778 | -0.001009331 | 0.000311447 |
| SMARCA1 | 0.013642907 | 0.011707582 | 0.025350489 |
| TFG | 0.016939195 | 0.021551438 | 0.038490634 |
| AL021368.2 | -0.005473162 | 0.004056864 | -0.001416298 |
| WFDC2 | 0.025482692 | 0.163628752 | 0.189111444 |
| CDH15 | 0.001230891 | -0.000994133 | 0.000236758 |
| TMEM259 | 0.014010534 | 0.011160853 | 0.025171387 |
| ITGB6 | -0.004991246 | 0.006214607 | 0.001223361 |
| SFTA1P | 0.001114578 | -0.001114578 | 0 |
| MANF | 0.013566269 | 0.0301598 | 0.043726069 |
| AFDN | 0.01649735 | 0.019287544 | 0.035784894 |
| TBL1XR1 | 0.017299384 | 0.024637721 | 0.041937105 |
| ADSSL1 | 0.012546809 | -0.025637916 | -0.013091106 |
| VTN | 0.004517669 | -0.007206523 | -0.002688854 |
| RPL13A | 0.028131162 | 0.032575362 | 0.060706524 |
| C11orf98 | 0.005621584 | -0.00604123 | -0.000419646 |
| GOSR2 | 0.015488851 | -0.020899463 | -0.005410612 |
| LXN | 0.007804188 | 0.007948032 | 0.01575222 |
| FAT3 | 0.004229851 | -0.007171609 | -0.002941758 |
| KCNMB4 | 0.008920202 | 0.017478663 | 0.026398865 |
| HNF4A-AS1 | 0.00250781 | -0.001888479 | 0.000619331 |
| CCDC88C | 0.008085222 | -0.007282748 | 0.000802473 |
| BCL2L10 | -0.006108759 | 0.005348124 | -0.000760635 |
| GPT | -0.013847522 | 0.014330847 | 0.000483325 |
| ADAMTS16 | -0.006566647 | 0.018960588 | 0.012393941 |
| GJC1 | 0.001133927 | -0.001013184 | 0.000120743 |
| MYCBP | 0.010842801 | -0.009974526 | 0.000868275 |
| PFKL | 0.021492625 | -0.027459685 | -0.00596706 |
| HSPA8 | 0.027774916 | 0.028335489 | 0.056110405 |
| AL391117.1 | 0.001119835 | -0.000928828 | 0.000191007 |
| IRF2BP2 | -0.0167667 | 0.042706918 | 0.025940218 |

| | | | |
|---|---|---|---|
| ANKRD55 | 0.00121249 | -0.001041315 | 0.000171176 |
| SHANK3 | 0.007857502 | -0.007919104 | -6.16023E-05 |
| HEY1 | -0.003155931 | 0.00379286 | 0.000636929 |
| IER2 | 0.022092872 | 0.051494878 | 0.07358775 |
| MPPE1 | 0.010863987 | 0.019138618 | 0.030002605 |
| SPNS1 | 0.009690701 | 0.009019403 | 0.018710103 |
| ZBED2 | 0.002076219 | -0.00210441 | -2.81904E-05 |
| CMTM4 | -0.013879954 | 0.010243813 | -0.003636141 |
| CAPRIN1 | 0.016970763 | 0.01349146 | 0.030462224 |
| LMO7 | 0.015544925 | 0.0142249 | 0.029769825 |
| AC015845.2 | 0.003786504 | -0.00489697 | -0.001110466 |
| CCDC82 | 0.011489349 | 0.009967609 | 0.021456958 |
| EDIL3 | 0.010259392 | 0.007704586 | 0.017963978 |
| CFL2 | 0.015648676 | -0.015273102 | 0.000375574 |
| SYNJ2BP | 0.014815748 | 0.015639031 | 0.030454779 |
| TRABD | 0.01091327 | 0.008179198 | 0.019092467 |
| PMM2 | 0.007700205 | 0.005699946 | 0.013400152 |
| CDC42EP1 | 0.006876023 | 0.015304725 | 0.022180748 |
| MED12L | 0.001323946 | -0.000985339 | 0.000338607 |
| ABCA3 | 0.006483654 | 0.00761943 | 0.014103084 |
| MAD2L2 | 0.010047827 | 0.035002673 | 0.0450505 |
| CAPN12 | 0.011866162 | 0.03565531 | 0.047521472 |
| DONSON | -0.003780448 | 0.003068058 | -0.00071239 |
| CPVL | 0.019655394 | 0.042310255 | 0.061965648 |
| CCT4 | 0.019722551 | 0.017855798 | 0.037578348 |
| CLTRN | 0.026951505 | 0.032873002 | 0.059824507 |
| CEP63 | 0.009893869 | -0.00752331 | 0.002370559 |
| GTF2F2 | 0.009206219 | 0.008805595 | 0.018011814 |
| LOH12CR2 | 0.004716803 | -0.003941132 | 0.000775671 |
| FAM234A | 0.013128395 | 0.016903867 | 0.030032261 |
| ARHGAP18 | 0.011951759 | -0.009256208 | 0.002695551 |
| GRM5 | -0.002863962 | 0.004750659 | 0.001886697 |
| BICD1 | -0.006625978 | 0.009948242 | 0.003322264 |
| JDP2 | 0.009817529 | -0.009625457 | 0.000192072 |
| TONSL | 0.006070537 | -0.00815301 | -0.002082473 |
| LAMP1 | -0.022763841 | 0.028403338 | 0.005639497 |
| IMMP2L | 0.013451197 | 0.016792567 | 0.030243764 |
| FAM171A1 | -0.009575748 | 0.027530864 | 0.017955115 |
| PCCB | 0.013420966 | -0.022625439 | -0.009204473 |
| TCTN1 | 0.012690383 | 0.011277187 | 0.023967571 |
| AAK1 | 0.01886432 | 0.021329121 | 0.040193441 |
| LDHD | 0.014460924 | -0.018812524 | -0.004351599 |
| YWHAZ | 0.020461934 | 0.020366321 | 0.040828255 |
| DMXL1 | 0.016871308 | -0.024142604 | -0.007271296 |
| CALCRL | 0.008690807 | 0.009679127 | 0.018369934 |
| ANO10 | 0.011321585 | 0.016322984 | 0.027644569 |
| TTC17 | 0.011448252 | 0.018336642 | 0.029784894 |
| NFKBIE | 0.007080629 | 0.013096099 | 0.020176729 |
| TTC28 | 0.010820339 | 0.008186814 | 0.019007153 |
| TAP1 | 0.007625336 | 0.011902468 | 0.019527804 |

| | | | |
|---|---|---|---|
| TFCP2 | -0.007110964 | 0.010833403 | 0.003722439 |
| AC004556.3 | 0.006309636 | -0.028814367 | -0.022504731 |
| AC020656.2 | -0.00211978 | 0.002429333 | 0.000309553 |
| LYG1 | -0.008873751 | 0.022333127 | 0.013459376 |
| CCDC130 | 0.009244799 | 0.008138034 | 0.017382834 |
| UBE2Z | 0.013259095 | 0.01364816 | 0.026907255 |
| PRKDC | 0.015677388 | 0.01396488 | 0.029642268 |
| HOXD-AS2 | 0.014654523 | -0.021246701 | -0.006592178 |
| ZNF429 | -0.009457082 | 0.015567175 | 0.006110094 |
| TBCC | 0.009526118 | 0.009971381 | 0.019497499 |
| LINC00667 | 0.012930983 | 0.013360643 | 0.026291627 |
| CHD2 | 0.01446247 | 0.014923477 | 0.029385947 |
| AC005253.1 | -0.002012595 | 0.001565213 | -0.000447382 |
| AC098588.3 | 0.001215461 | -0.000982059 | 0.000233401 |
| DMKN | 0.016747899 | 0.056859541 | 0.073607439 |
| AC007906.2 | -0.009355592 | 0.018765659 | 0.009410067 |
| CALU | 0.011060199 | 0.021277216 | 0.032337415 |
| AR | 0.010887509 | 0.018815437 | 0.029702947 |
| GIPR | 0.001361613 | -0.001525925 | -0.000164313 |
| ABCA12 | 0.000798709 | -0.000798709 | -1.77636E-15 |
| PLEKHH2 | -0.012705179 | 0.026415002 | 0.013709823 |
| MAPK10 | 0.012692308 | 0.012651249 | 0.025343556 |
| TM2D2 | 0.013058284 | 0.013455977 | 0.026514261 |

**Descending thin limb cells**

| Genes | logFC_T2D_HC | logFC_T2Di_T2D | logFC_T2Di_HC |
|---|---|---|---|
| AKR1B1 | 0.923819973 | -0.902882636 | 0.020937338 |
| RARRES2 | 0.781197565 | -0.756013785 | 0.025183779 |
| SERPINA1 | 0.710199992 | -0.665894157 | 0.044305835 |
| AKR1C1 | 0.758736091 | -0.750654777 | 0.008081314 |
| HSPA2 | 0.703750658 | -0.711819791 | -0.008069134 |
| TMEM176B | 0.644576299 | -0.54196293 | 0.102613369 |
| RANBP3L | 0.570046184 | -0.595110613 | -0.025064429 |
| SCCPDH | 0.574900906 | -0.587828481 | -0.012927575 |
| NEURL3 | 0.479329746 | -0.467628318 | 0.011701428 |
| LDHA | 0.603480964 | -0.545595815 | 0.057885149 |
| GAS6 | 0.761476987 | -0.750544413 | 0.010932574 |
| ANK2 | 0.515177727 | -0.493086367 | 0.02209136 |
| ATP1B1 | 0.651236755 | -0.640338674 | 0.010898081 |
| ID1 | -0.687889518 | 0.553411354 | -0.134478164 |
| ARPC1B | 0.505293153 | -0.468890357 | 0.036402796 |
| TMEM176A | 0.537866229 | -0.458858753 | 0.079007476 |
| CLDN10 | 0.726106513 | -0.70956425 | 0.016542263 |
| ABCA4 | 0.405764388 | -0.40700386 | -0.001239471 |
| MAL2 | 0.556974048 | -0.561131472 | -0.004157424 |
| GAPDH | 0.616559402 | -0.373680148 | 0.242879254 |
| CALM1 | 0.605486671 | -0.520951193 | 0.084535478 |
| DDIT4L | 0.453167325 | -0.433946037 | 0.019221287 |
| ID4 | -0.611112892 | 0.458377517 | -0.152735376 |
| S100A11 | 0.618117278 | -0.393935627 | 0.224181651 |
| TPD52L1 | 0.42431602 | -0.374466796 | 0.049849224 |
| LPIN2 | 0.424849071 | -0.429943603 | -0.005094532 |
| CLCNKA | 0.503721114 | -0.493660109 | 0.010061006 |
| RNF24 | 0.403836823 | -0.398310686 | 0.005526137 |
| CXCL14 | 0.653889985 | -0.534496743 | 0.119393242 |
| JAG1 | -0.509253544 | 0.287554638 | -0.221698906 |
| COL4A2 | -0.497045795 | 0.309406623 | -0.187639172 |
| ID3 | -0.582830722 | 0.442606674 | -0.140224048 |
| COL4A1 | -0.456286122 | 0.221232554 | -0.235053568 |
| MRPS6 | 0.613397122 | -0.624949394 | -0.011552273 |
| ARG2 | 0.38460899 | -0.367381414 | 0.017227576 |
| SH3GL3 | 0.377503139 | -0.378494755 | -0.000991616 |
| LINC01320 | -0.490406012 | 0.171384208 | -0.319021804 |
| CYSTM1 | 0.554005053 | -0.37231032 | 0.181694733 |
| FXYD2 | 0.361379896 | -0.252081868 | 0.109298028 |
| TACSTD2 | 0.559307674 | -0.639862053 | -0.08055438 |
| RASSF8 | 0.362909362 | -0.3584373 | 0.004472062 |
| SULT2B1 | 0.335946651 | -0.341498103 | -0.005551453 |
| CD82 | 0.327456783 | -0.333337094 | -0.00588031 |
| FCGBP | 0.366364377 | -0.394093853 | -0.027729475 |
| S100A4 | 0.465293138 | -0.465323793 | -3.06552E-05 |

| | | | |
|---|---|---|---|
| CPE | 0.465239796 | -0.450347489 | 0.014892306 |
| PEBP1 | 0.460423498 | -0.414965864 | 0.045457634 |
| ATOH8 | -0.387763467 | 0.239622163 | -0.148141304 |
| GLS | 0.417407177 | -0.40091712 | 0.016490057 |
| ADIRF | -0.515834896 | 0.527875096 | 0.0120402 |
| CAT | 0.453784497 | -0.518199738 | -0.064415241 |
| HIF1A | 0.411381126 | -0.387104528 | 0.024276597 |
| MELTF | 0.298983667 | -0.29563558 | 0.003348087 |
| CLINT1 | 0.378362488 | -0.389021772 | -0.010659284 |
| PKM | 0.473101113 | -0.43500754 | 0.038093574 |
| S100A6 | 0.474314625 | -0.570035336 | -0.095720711 |
| BSND | 0.319847124 | -0.317654659 | 0.002192465 |
| H2AFZ | 0.431957016 | -0.380386569 | 0.051570447 |
| NAPEPLD | 0.330270741 | -0.332177574 | -0.001906833 |
| CA2 | 0.396857036 | -0.331357099 | 0.065499937 |
| GSTM3 | 0.481021663 | -0.490340096 | -0.009318433 |
| EEF1A1 | 0.296156661 | -0.157297416 | 0.138859245 |
| FBXO2 | 0.295051984 | -0.284081093 | 0.010970891 |
| CRYAB | 0.49693277 | -0.327621098 | 0.169311671 |
| MID1IP1 | 0.342489549 | -0.320402973 | 0.022086576 |
| GAL3ST1 | 0.318884747 | -0.320361781 | -0.001477034 |
| MFSD10 | 0.391247597 | -0.432898736 | -0.041651139 |
| SNRPN | 0.419694484 | -0.359679367 | 0.060015117 |
| DEFB1 | -0.484651533 | 0.231509513 | -0.25314202 |
| ISYNA1 | 0.306417941 | -0.278813856 | 0.027604085 |
| MIF | 0.448610497 | -0.216465334 | 0.232145163 |
| ECH1 | 0.366549649 | -0.236949763 | 0.129599886 |
| VAV3 | 0.320279035 | -0.283822243 | 0.036456792 |
| TLN2 | 0.357851523 | -0.384307219 | -0.026455696 |
| TMEM178A | 0.266323527 | -0.265498138 | 0.000825388 |
| GSTP1 | 0.42775281 | -0.318008584 | 0.109744226 |
| TFF3 | 0.304209642 | -0.312312312 | -0.00810267 |
| SLC4A7 | 0.296261764 | -0.320999718 | -0.024737954 |
| COL12A1 | 0.263789112 | -0.257323487 | 0.006465625 |
| CBR1 | 0.365092996 | -0.31258267 | 0.052510326 |
| SERPINA6 | 0.300725903 | -0.300725903 | -1.95399E-14 |
| CGNL1 | 0.273821216 | -0.256930046 | 0.016891171 |
| ATP1B3 | 0.323923973 | -0.329816988 | -0.005893014 |
| DANCR | 0.411271849 | -0.40252107 | 0.008750778 |
| SMAD9 | -0.342535633 | 0.159947284 | -0.182588348 |
| UBC | 0.435149323 | -0.284817367 | 0.150331956 |
| LIX1 | 0.351524725 | -0.298082611 | 0.053442114 |
| PPIA | 0.405191776 | -0.203544918 | 0.201646858 |
| MYL12A | 0.420062254 | -0.267169019 | 0.152893235 |
| GRAMD2B | 0.264385609 | -0.277662861 | -0.013277252 |
| MAL | 0.496079548 | -0.757857768 | -0.261778219 |
| C12orf75 | 0.379564336 | -0.292716384 | 0.086847952 |

| | | | |
|---|---|---|---|
| NME2 | 0.41384501 | -0.19353975 | 0.22030526 |
| CD24 | 0.383631911 | -0.492299085 | -0.108667174 |
| MYL12B | 0.418213803 | -0.249797715 | 0.168416088 |
| CLDN1 | 0.314793325 | -0.308262313 | 0.006531012 |
| EEF1E1 | 0.363593473 | -0.311942931 | 0.051650542 |
| GCHFR | 0.291894843 | -0.246619355 | 0.045275488 |
| CARHSP1 | 0.318860891 | -0.234188783 | 0.084672108 |
| BHMT2 | 0.285677439 | -0.20401662 | 0.081660819 |
| MIEN1 | 0.32292535 | -0.292558078 | 0.030367272 |
| COX5A | 0.353006772 | -0.231482906 | 0.121523867 |
| CA8 | 0.247238751 | -0.247238751 | -1.95399E-14 |
| S100A13 | 0.435303315 | -0.311788695 | 0.12351462 |
| PRKAR2B | 0.256935727 | -0.270040719 | -0.013104992 |
| GNAI1 | 0.346468488 | -0.374034115 | -0.027565627 |
| CAMK2N1 | 0.375067596 | -0.33745895 | 0.037608646 |
| TINAGL1 | 0.359007825 | -0.346773587 | 0.012234238 |
| NPR3 | -0.325364515 | 0.261911034 | -0.063453481 |
| SPOCK2 | -0.401232054 | 0.22363633 | -0.177595724 |
| CLCNKB | 0.288723472 | -0.290764087 | -0.002040615 |
| VDAC2 | 0.364274658 | -0.287278894 | 0.076995763 |
| SERF2 | 0.376074672 | -0.194691627 | 0.181383046 |
| NINJ1 | 0.300758951 | -0.283410983 | 0.017347968 |
| MYL6 | 0.373185825 | -0.223172469 | 0.150013356 |
| BAG1 | 0.362693881 | -0.363007835 | -0.000313954 |
| RPLP1 | 0.295619956 | -0.167759817 | 0.127860139 |
| VAMP8 | 0.398045595 | -0.254084323 | 0.143961272 |
| EVA1B | 0.265339984 | -0.243162057 | 0.022177927 |
| EXPH5 | 0.243618173 | -0.257987843 | -0.01436967 |
| BCL6 | 0.243735497 | -0.266607738 | -0.022872241 |
| RPL28 | 0.331437733 | -0.087333665 | 0.244104069 |
| YWHAH | 0.294283144 | -0.2474615 | 0.046821644 |
| NTRK2 | -0.283456681 | 0.141015915 | -0.142440766 |
| DBI | 0.378362839 | -0.245043836 | 0.133319003 |
| MXRA7 | 0.351077558 | -0.368725051 | -0.017647494 |
| GPNMB | -0.299551957 | 0.047367259 | -0.252184698 |
| CFL1 | 0.385574768 | -0.190603702 | 0.194971066 |
| GLUL | 0.339562132 | -0.332093011 | 0.007469121 |
| PCBD1 | 0.359105894 | -0.222991486 | 0.136114408 |
| PFN1 | 0.38754668 | -0.147136936 | 0.240409744 |
| S100A10 | 0.381153005 | -0.205669279 | 0.175483726 |
| PFKP | 0.256086753 | -0.223762055 | 0.032324698 |
| NDUFB2 | 0.38185852 | -0.269731218 | 0.112127302 |
| ATP5F1E | 0.356932581 | -0.167403809 | 0.189528772 |
| RNF144B | -0.318375217 | 0.181791318 | -0.136583898 |
| COMT | 0.363737652 | -0.373613634 | -0.009875982 |
| PDCD6 | 0.348735626 | -0.304312479 | 0.044423147 |
| TESC | 0.210786313 | -0.199289558 | 0.011496755 |

| | | | |
|---|---|---|---|
| CNN3 | 0.338813347 | -0.314782616 | 0.02403073 |
| HSPB7 | 0.220578469 | -0.217781889 | 0.00279658 |
| ARPC3 | 0.351795162 | -0.223623284 | 0.128171877 |
| ATP5F1C | 0.344137017 | -0.217416607 | 0.12672041 |
| RAN | 0.346969876 | -0.263288738 | 0.083681137 |
| ITPKB | 0.199324076 | -0.197544396 | 0.00177968 |
| COX7C | 0.353435893 | -0.137813781 | 0.215622112 |
| TPI1 | 0.362095222 | -0.225156098 | 0.136939124 |
| NDUFS5 | 0.362180599 | -0.139199304 | 0.222981296 |
| SLCO4A1 | 0.174024375 | -0.176805592 | -0.002781217 |
| PAK2 | 0.28722479 | -0.278736621 | 0.008488168 |
| CSRP2 | 0.276113493 | -0.241189831 | 0.034923662 |
| SH3BGRL3 | 0.30304046 | -0.190599945 | 0.112440515 |
| PDE4DIP | 0.263173945 | -0.29250542 | -0.029331475 |
| LDHB | 0.36667429 | -0.212689515 | 0.153984775 |
| F2R | 0.186397832 | -0.185651858 | 0.000745974 |
| MRPL34 | 0.297076357 | -0.246921607 | 0.05015475 |
| EHF | 0.266553663 | -0.305052502 | -0.038498839 |
| CDK2AP1 | 0.246463934 | -0.227864872 | 0.018599062 |
| ACADVL | 0.30661112 | -0.251051266 | 0.055559854 |
| RPS10 | 0.354620898 | -0.227422959 | 0.127197939 |
| KRT18 | 0.345270733 | -0.247626306 | 0.097644428 |
| GSPT1 | 0.293685769 | -0.302087342 | -0.008401573 |
| NME4 | 0.257921366 | -0.211509866 | 0.046411499 |
| CLU | 0.399723682 | -0.471613405 | -0.071889723 |
| C4orf48 | 0.239050992 | -0.225706311 | 0.013344681 |
| CAPS | 0.227264 | -0.21150422 | 0.01575978 |
| SLIT3 | -0.219797204 | 0.11116821 | -0.108628994 |
| SOD3 | 0.45938459 | -0.528749438 | -0.069364848 |
| CTSD | 0.358403461 | -0.336396372 | 0.022007089 |
| ACAA2 | 0.314833143 | -0.213900168 | 0.100932975 |
| STMP1 | 0.340148881 | -0.357246486 | -0.017097605 |
| RAB9A | 0.264308798 | -0.241256641 | 0.023052157 |
| GPX4 | 0.358711296 | -0.103307386 | 0.25540391 |
| HSBP1 | 0.3580346 | -0.236309364 | 0.121725235 |
| LARP6 | 0.224938608 | -0.219555363 | 0.005383246 |
| PGAM1 | 0.295429901 | -0.273843167 | 0.021586734 |
| RPL8 | 0.2978253 | -0.115474411 | 0.182350889 |
| APP | 0.366940207 | -0.554061851 | -0.187121644 |
| TSC22D4 | 0.250511646 | -0.274596703 | -0.024085056 |
| RHOA | 0.327020164 | -0.254472466 | 0.072547699 |
| SMOC2 | 0.207640105 | -0.217560391 | -0.009920286 |
| SPOCK1 | 0.168145171 | -0.167338063 | 0.000807109 |
| SLC25A5 | 0.310729782 | -0.184883278 | 0.125846504 |
| RACK1 | 0.327202661 | -0.144953772 | 0.182248889 |
| ATP5MC3 | 0.343326737 | -0.12531124 | 0.218015497 |
| TMEM37 | 0.280047072 | -0.281073691 | -0.001026619 |

| | | | |
|---|---|---|---|
| NPM1 | 0.348589199 | -0.204775274 | 0.143813925 |
| PRDX2 | 0.342638013 | -0.199966948 | 0.142671065 |
| ENO1 | 0.338896659 | -0.265226581 | 0.073670078 |
| RPS8 | 0.280647563 | -0.101557641 | 0.179089922 |
| PHLDA1 | 0.227387468 | -0.213072649 | 0.014314819 |
| PARK7 | 0.343868092 | -0.168255331 | 0.175612761 |
| ETS1 | 0.193286439 | -0.205224584 | -0.011938145 |
| OAZ1 | 0.346038497 | -0.211877389 | 0.134161108 |
| SAMD11 | -0.241567798 | 0.165410485 | -0.076157313 |
| GALNT10 | -0.214261358 | 0.091368269 | -0.122893088 |
| PRDX1 | 0.348691693 | -0.197387201 | 0.151304492 |
| HIGD1A | 0.293552576 | -0.202286762 | 0.091265814 |
| CSTB | 0.358508538 | -0.297488323 | 0.061020215 |
| ATP6V1F | 0.328835926 | -0.156045312 | 0.172790614 |
| CNP | 0.262882203 | -0.238948108 | 0.023934095 |
| SLCO3A1 | 0.189264178 | -0.191364126 | -0.002099947 |
| KRT7 | 0.322701036 | -0.239309409 | 0.083391627 |
| TUBB4B | 0.309950113 | -0.126865584 | 0.183084529 |
| CTNNAL1 | 0.197919875 | -0.193669224 | 0.004250651 |
| SLC25A3 | 0.339669535 | -0.197603032 | 0.142066503 |
| RPL7A | 0.286943042 | -0.123117637 | 0.163825405 |
| EIF2AK4 | 0.214120215 | -0.222695072 | -0.008574857 |
| IGFBP7 | 0.347118342 | -0.495148622 | -0.14803028 |
| CD59 | 0.34021869 | -0.365595499 | -0.025376809 |
| TACC1 | 0.229028804 | -0.237650355 | -0.008621551 |
| PTMA | 0.301851194 | -0.157609213 | 0.144241981 |
| SIPA1 | 0.188341538 | -0.194013315 | -0.005671777 |
| RRM2B | 0.236398159 | -0.260377215 | -0.023979057 |
| NAA38 | 0.29978296 | -0.174694431 | 0.125088529 |
| RPS13 | 0.29902511 | -0.125591442 | 0.173433668 |
| MTPN | 0.251183531 | -0.229621032 | 0.0215625 |
| SLC5A3 | 0.374938641 | -0.610478145 | -0.235539504 |
| COL4A3 | 0.267869154 | -0.308389015 | -0.040519861 |
| PRKX | 0.204312849 | -0.215426465 | -0.011113616 |
| SASH1 | 0.22174687 | -0.231724007 | -0.009977137 |
| ARPC2 | 0.273177591 | -0.218234506 | 0.054943085 |
| CPNE3 | 0.270041905 | -0.254536007 | 0.015505898 |
| EIF3K | 0.309267453 | -0.166570675 | 0.142696778 |
| GNG5 | 0.324494913 | -0.201121195 | 0.123373718 |
| PKN1 | 0.2032792 | -0.209433917 | -0.006154717 |
| HPS1 | 0.199781997 | -0.183756936 | 0.01602506 |
| DYNLRB1 | 0.334443097 | -0.202816694 | 0.131626403 |
| AFAP1L2 | 0.146489895 | -0.146429299 | 6.05959E-05 |
| PSMB6 | 0.286339288 | -0.187176561 | 0.099162727 |
| SLC6A12 | 0.252402304 | -0.307522773 | -0.055120469 |
| NDUFB3 | 0.300462254 | -0.168467778 | 0.131994475 |
| UQCRH | 0.329886395 | -0.170376457 | 0.159509937 |

| | | | |
|---|---|---|---|
| YPEL5 | 0.243565176 | -0.231295739 | 0.012269437 |
| SIM1 | 0.24815807 | -0.283223218 | -0.035065148 |
| HOMER3 | 0.186799429 | -0.148981551 | 0.037817877 |
| DNAJA1 | 0.279513902 | -0.250729622 | 0.02878428 |
| NR2F6 | 0.228561787 | -0.18691155 | 0.041650237 |
| ECHS1 | 0.276348767 | -0.166568335 | 0.109780431 |
| COPE | 0.277468891 | -0.218433999 | 0.059034892 |
| TNFRSF12A | 0.318423068 | -0.341259642 | -0.022836574 |
| LPIN1 | -0.26647666 | 0.08284807 | -0.18362859 |
| ATP5MF | 0.31822468 | -0.134286782 | 0.183937898 |
| C5orf38 | 0.205503214 | -0.21035712 | -0.004853907 |
| CHCHD2 | 0.329292986 | -0.166701705 | 0.162591281 |
| STXBP6 | 0.155916463 | -0.149666521 | 0.006249943 |
| PRDX5 | 0.312449246 | -0.154434029 | 0.158015217 |
| HAGH | 0.238413679 | -0.235740069 | 0.002673611 |
| TRHDE-AS1 | -0.176060514 | 0.048027089 | -0.128033425 |
| NUPR1 | -0.39574378 | 0.18156653 | -0.21417725 |
| S100A16 | 0.261110578 | -0.204716299 | 0.056394279 |
| YBX1 | 0.333602975 | -0.235355354 | 0.098247621 |
| BTG1 | 0.330324042 | -0.399467595 | -0.069143553 |
| DRAP1 | 0.257508946 | -0.172386529 | 0.085122417 |
| DCXR | 0.302835274 | -0.116151162 | 0.186684112 |
| COX5B | 0.319327841 | -0.156461665 | 0.162866176 |
| PROM1 | -0.287268504 | 0.115093667 | -0.172174836 |
| ASAH1 | 0.300089452 | -0.286303637 | 0.013785816 |
| SRGAP3 | 0.242424591 | -0.283922936 | -0.041498346 |
| STARD10 | 0.206053321 | -0.177836475 | 0.028216846 |
| FGFR3 | -0.22774365 | 0.096101348 | -0.131642301 |
| RPS23 | 0.253316882 | -0.058722423 | 0.194594459 |
| RHOD | 0.19976958 | -0.176064319 | 0.023705261 |
| FCHO2 | 0.216313495 | -0.24563819 | -0.029324694 |
| ETF1 | 0.217124283 | -0.202886563 | 0.01423772 |
| ST3GAL4 | 0.181265581 | -0.188408293 | -0.007142713 |
| SLC4A11 | 0.264594102 | -0.252358406 | 0.012235697 |
| GABARAP | 0.319666421 | -0.174558475 | 0.145107946 |
| RPL19 | 0.252707646 | -0.09778268 | 0.154924965 |
| PRSS23 | 0.291831004 | -0.310684501 | -0.018853497 |
| RPL3 | 0.222916979 | -0.11442157 | 0.108495409 |
| UBE2M | 0.250287812 | -0.159192513 | 0.091095299 |
| RPS9 | 0.26278111 | -0.080818547 | 0.181962563 |
| MRC2 | -0.20317333 | 0.144374353 | -0.058798977 |
| DPP4 | 0.210795428 | -0.226450464 | -0.015655036 |
| FAM200B | 0.261714093 | -0.180554016 | 0.081160077 |
| ILF2 | 0.252903625 | -0.214354301 | 0.038549324 |
| AOC1 | 0.166207643 | -0.1609461 | 0.005261543 |
| NDUFS2 | 0.224734689 | -0.160905253 | 0.063829436 |
| VAPA | 0.27979664 | -0.2588229 | 0.02097374 |

| | | | |
|---|---|---|---|
| SESN3 | -0.233369872 | 0.143164712 | -0.09020516 |
| RPS19BP1 | 0.269539198 | -0.23533983 | 0.034199368 |
| MRPL33 | 0.310170086 | -0.209379309 | 0.100790777 |
| KCNK15 | 0.152572573 | -0.150603165 | 0.001969408 |
| NDUFB4 | 0.291574931 | -0.131285545 | 0.160289386 |
| LAMTOR5 | 0.283909145 | -0.092004159 | 0.191904986 |
| RPL14 | 0.287072479 | -0.091766153 | 0.195306327 |
| NTAN1 | 0.178196426 | -0.151328312 | 0.026868114 |
| LHFPL3-AS2 | 0.199164044 | -0.173619822 | 0.025544222 |
| CLDN4 | 0.294298625 | -0.355775449 | -0.061476825 |
| HLA-C | 0.308647217 | -0.273603375 | 0.035043842 |
| PSMC4 | 0.21694577 | -0.179032343 | 0.037913427 |
| RNF5 | 0.238503842 | -0.158309057 | 0.080194785 |
| NDUFV2 | 0.266827656 | -0.16530777 | 0.101519885 |
| RPS12 | 0.234771703 | -0.083132654 | 0.151639049 |
| ARPC1A | 0.223753676 | -0.194138163 | 0.029615513 |
| WFS1 | 0.151653085 | -0.14696087 | 0.004692215 |
| LNPEP | 0.26561938 | -0.347434773 | -0.081815393 |
| ATXN10 | 0.239215023 | -0.22852772 | 0.010687303 |
| TSPO | 0.246455447 | -0.150424324 | 0.096031123 |
| DNAJC3 | 0.220453877 | -0.236776889 | -0.016323012 |
| ARRDC1 | 0.195277161 | -0.215482802 | -0.02020564 |
| COX14 | 0.263726175 | -0.216539984 | 0.047186192 |
| DBNDD2 | 0.179625539 | -0.162390006 | 0.017235533 |
| ELOB | 0.300308773 | -0.164801952 | 0.135506821 |
| DDX3Y | 0.195644076 | -0.233458094 | -0.037814019 |
| SQLE | 0.179714495 | -0.164016851 | 0.015697644 |
| SUMO2 | 0.313114879 | -0.197026541 | 0.116088337 |
| RPL36AL | 0.309801896 | -0.17791321 | 0.131888687 |
| PPA1 | 0.239956427 | -0.167821404 | 0.072135023 |
| RPS5 | 0.296020055 | -0.125840982 | 0.170179073 |
| SRSF9 | 0.286713889 | -0.215252645 | 0.071461244 |
| CCDC32 | 0.191743033 | -0.182357721 | 0.009385313 |
| NDUFB11 | 0.281691819 | -0.177118319 | 0.1045735 |
| ATP5F1D | 0.294954519 | -0.135775951 | 0.159178568 |
| YWHAB | 0.289026108 | -0.242764292 | 0.046261816 |
| PARD6A | 0.138959361 | -0.13341565 | 0.005543711 |
| ACTG1 | 0.286409595 | -0.131446721 | 0.154962875 |
| CNNM1 | 0.125875363 | -0.1236134 | 0.002261963 |
| ELF5 | 0.137807094 | -0.136877215 | 0.000929879 |
| DDX5 | 0.282414781 | -0.278781822 | 0.003632959 |
| RPS2 | 0.278956663 | -0.108419327 | 0.170537337 |
| ELF3 | 0.235547682 | -0.244739063 | -0.009191382 |
| PISD | 0.167434074 | -0.175500974 | -0.0080669 |
| VDAC1 | 0.244881118 | -0.144913945 | 0.099967173 |
| COX6B1 | 0.291845935 | -0.150513483 | 0.141332452 |
| RPS19 | 0.234761293 | -0.06795676 | 0.166804533 |

| | | | |
|---|---|---|---|
| RIPOR3 | 0.14306881 | -0.144216275 | -0.001147464 |
| PSMD8 | 0.259274086 | -0.188328709 | 0.070945377 |
| RAB2A | 0.265662744 | -0.222135233 | 0.043527511 |
| NDUFS8 | 0.251889386 | -0.140315908 | 0.111573478 |
| CYB5A | 0.299646218 | -0.141861967 | 0.157784251 |
| C1orf122 | 0.200023963 | -0.16959112 | 0.030432844 |
| SLC12A1 | -0.266684022 | 0.216107998 | -0.050576024 |
| ETS2 | 0.182154741 | -0.183253577 | -0.001098837 |
| AP2M1 | 0.272129043 | -0.224653831 | 0.047475213 |
| RPL29 | 0.259148941 | -0.061666527 | 0.197482414 |
| NAPSA | -0.224924163 | 0.202407653 | -0.02251651 |
| MAP2K2 | 0.224114339 | -0.163271426 | 0.060842912 |
| PSMB3 | 0.24243604 | -0.144619789 | 0.097816251 |
| ZBTB38 | 0.195410585 | -0.191386813 | 0.004023773 |
| METTL26 | 0.21823979 | -0.129738578 | 0.088501213 |
| PON2 | 0.225522441 | -0.227088376 | -0.001565935 |
| ATP5MG | 0.289811884 | -0.134500036 | 0.155311848 |
| CTSH | 0.221059626 | -0.236893534 | -0.015833908 |
| RPL10 | 0.22181846 | -0.05720703 | 0.16461143 |
| NEDD8 | 0.280462682 | -0.15059644 | 0.129866242 |
| CYP51A1 | 0.208930873 | -0.205942567 | 0.002988307 |
| HACD3 | 0.231332146 | -0.264887296 | -0.03355515 |
| CHMP5 | 0.257648271 | -0.204591153 | 0.053057118 |
| RAB17 | 0.210650227 | -0.196701976 | 0.013948251 |
| S100A1 | 0.218371816 | -0.154671996 | 0.063699819 |
| GRB14 | 0.210443172 | -0.232869381 | -0.022426209 |
| CNTD2 | 0.145771582 | -0.147307713 | -0.001536132 |
| SRP14 | 0.284923331 | -0.13926064 | 0.145662691 |
| TXNIP | -0.29185195 | 0.31669691 | 0.02484496 |
| MZT2B | 0.292129156 | -0.15264453 | 0.139484626 |
| EEF1D | 0.292178055 | -0.148410029 | 0.143768027 |
| EPCAM | 0.305140727 | -0.471977083 | -0.166836356 |
| REEP5 | 0.274964122 | -0.322848708 | -0.047884587 |
| LOXL2 | 0.122420315 | -0.123682487 | -0.001262172 |
| GCNT2 | 0.198497273 | -0.217895196 | -0.019397923 |
| AGFG2 | 0.1450559 | -0.147159479 | -0.002103579 |
| BRI3 | 0.284729286 | -0.181437304 | 0.103291982 |
| ANXA4 | 0.254964541 | -0.151792446 | 0.103172095 |
| PANK2 | 0.181640118 | -0.199706401 | -0.018066282 |
| RBX1 | 0.268227979 | -0.140416009 | 0.12781197 |
| CLTA | 0.281547607 | -0.149071641 | 0.132475967 |
| H3F3A | 0.279268087 | -0.173752521 | 0.105515566 |
| UQCRQ | 0.272405023 | -0.074106983 | 0.198298041 |
| PRRG4 | 0.214292629 | -0.248731905 | -0.034439275 |
| PDZK1IP1 | -0.253979519 | 0.285673525 | 0.031694006 |
| PSMA7 | 0.260589202 | -0.219621796 | 0.040967406 |
| SHB | 0.199875866 | -0.204230944 | -0.004355077 |

| | | | |
|---|---|---|---|
| TIMP1 | 0.226347385 | -0.160582605 | 0.06576478 |
| IVNS1ABP | -0.241718985 | 0.154995265 | -0.086723719 |
| UBXN1 | 0.256563817 | -0.211489145 | 0.045074672 |
| EFHD1 | 0.252208103 | -0.220837526 | 0.031370577 |
| SCP2 | 0.235513972 | -0.155492173 | 0.0800218 |
| MARVELD3 | 0.202432704 | -0.214479415 | -0.012046711 |
| TRIR | 0.273206919 | -0.183311494 | 0.089895425 |
| CIB1 | 0.236403819 | -0.157181924 | 0.079221895 |
| ZNF580 | 0.221662617 | -0.219860471 | 0.001802146 |
| DDRGK1 | 0.222027347 | -0.228853001 | -0.006825654 |
| GHITM | 0.22494568 | -0.164291125 | 0.060654555 |
| UQCR10 | 0.282325698 | -0.126881522 | 0.155444176 |
| TUBA1B | 0.248957464 | -0.189169401 | 0.059788063 |
| COX17 | 0.2488304 | -0.163073053 | 0.085757346 |
| RPL15 | 0.246445867 | -0.077290991 | 0.169154876 |
| QPRT | 0.178290111 | -0.161788655 | 0.016501457 |
| BRK1 | 0.246288751 | -0.149069994 | 0.097218758 |
| DYNLT1 | 0.258080892 | -0.146735742 | 0.111345149 |
| CDC42EP3 | 0.171339683 | -0.183364649 | -0.012024966 |
| RPL30 | 0.235545651 | -0.080187424 | 0.155358227 |
| ANXA11 | 0.238890494 | -0.187887793 | 0.051002701 |
| AK1 | 0.159871755 | -0.128652404 | 0.03121935 |
| SLC38A2 | 0.222080124 | -0.2424663 | -0.020386176 |
| ARL6IP4 | 0.269337155 | -0.209944738 | 0.059392417 |
| EMC6 | 0.202918052 | -0.187644778 | 0.015273274 |
| PFDN5 | 0.277492095 | -0.078080376 | 0.199411719 |
| LINC02532 | 0.186480993 | -0.17923408 | 0.007246912 |
| RPL6 | 0.2540595 | -0.091708758 | 0.162350742 |
| STARD13 | 0.131665276 | -0.1357692 | -0.004103924 |
| EIF4A1 | 0.244182962 | -0.112950512 | 0.131232449 |
| POLR2L | 0.27612583 | -0.086580101 | 0.189545728 |
| COX7A2 | 0.275323267 | -0.168960464 | 0.106362803 |
| AEBP1 | -0.167124197 | 0.07024916 | -0.096875038 |
| SFRP1 | -0.164115822 | 0.102684293 | -0.061431529 |
| TRMT112 | 0.272167927 | -0.124498169 | 0.147669757 |
| RPS15 | 0.233175208 | -0.076732218 | 0.15644299 |
| STK17A | 0.133259524 | -0.126407435 | 0.006852089 |
| ANXA2 | 0.276672528 | -0.245631083 | 0.031041445 |
| ARL5A | 0.216314916 | -0.18933652 | 0.026978396 |
| LINC01589 | 0.113239298 | -0.116347469 | -0.003108172 |
| CLTC | 0.227834359 | -0.187480829 | 0.04035353 |
| COX8A | 0.274448975 | -0.158532836 | 0.115916139 |
| SPSB3 | 0.218753442 | -0.232429162 | -0.01367572 |
| MVK | 0.15915505 | -0.150875067 | 0.008279983 |
| AP2S1 | 0.214465181 | -0.137532573 | 0.076932608 |
| STRAP | 0.203420727 | -0.141373469 | 0.062047257 |
| ATF7IP2 | 0.164438755 | -0.151832887 | 0.012605868 |

| | | | |
|---|---|---|---|
| ITGB1BP1 | 0.172308038 | -0.124320966 | 0.047987072 |
| PROX1 | 0.126630729 | -0.133595742 | -0.006965013 |
| CISD2 | 0.180837536 | -0.147748498 | 0.033089038 |
| CENPX | 0.210205496 | -0.209409251 | 0.000796245 |
| OLA1 | 0.179954343 | -0.131987622 | 0.047966721 |
| COX6A1 | 0.271341392 | -0.225671591 | 0.0456698 |
| TSTD1 | 0.27265547 | -0.184331747 | 0.088323723 |
| LAPTM4B | 0.20484019 | -0.246787465 | -0.041947275 |
| ALDH1A3 | 0.117517288 | -0.114748743 | 0.002768546 |
| CDKN1C | 0.171832406 | -0.123635934 | 0.048196472 |
| UQCR11 | 0.266415757 | -0.07762562 | 0.188790137 |
| EIF3G | 0.241753275 | -0.144063332 | 0.097689943 |
| IER3 | 0.209545055 | -0.180127225 | 0.02941783 |
| COX20 | 0.251143955 | -0.227792959 | 0.023350995 |
| VDR | 0.113525144 | -0.108003088 | 0.005522056 |
| EMP2 | 0.170181227 | -0.17791461 | -0.007733383 |
| LYAR | 0.131732214 | -0.133619918 | -0.001887704 |
| ITM2B | 0.259873127 | -0.375835674 | -0.115962547 |
| TMED3 | 0.16586656 | -0.165485524 | 0.000381035 |
| HINT1 | 0.263088173 | -0.077859439 | 0.185228734 |
| COX7B | 0.266509035 | -0.103965982 | 0.162543053 |
| NDUFA3 | 0.255387394 | -0.148949201 | 0.106438193 |
| EIF3H | 0.236797167 | -0.165168445 | 0.071628722 |
| DAP | 0.203412938 | -0.19754658 | 0.005866358 |
| COMTD1 | 0.165848491 | -0.159780919 | 0.006067572 |
| PEF1 | 0.16384301 | -0.13379177 | 0.030051239 |
| AC106795.2 | 0.108089544 | -0.108477635 | -0.000388091 |
| COPRS | 0.184123312 | -0.110274681 | 0.073848631 |
| SLBP | 0.174252366 | -0.128547256 | 0.04570511 |
| KIF21A | 0.195007406 | -0.214221317 | -0.019213911 |
| NOL7 | 0.241672179 | -0.22205362 | 0.019618559 |
| DOK5 | 0.109634357 | -0.10352701 | 0.006107347 |
| PRELID1 | 0.235494003 | -0.129260499 | 0.106233504 |
| NDFIP2 | 0.181911525 | -0.188055639 | -0.006144114 |
| MYL6B | 0.2154892 | -0.175491834 | 0.039997366 |
| UQCRFS1 | 0.228806676 | -0.137937746 | 0.09086893 |
| CHCHD1 | 0.20658362 | -0.118934732 | 0.087648888 |
| MIR22HG | 0.132060389 | -0.142564292 | -0.010503903 |
| MRPS24 | 0.237365377 | -0.174953408 | 0.062411969 |
| TMEM230 | 0.222150224 | -0.177878646 | 0.044271578 |
| NDUFS7 | 0.226442289 | -0.141316358 | 0.085125931 |
| MRPS26 | 0.232266259 | -0.168800765 | 0.063465494 |
| PSME1 | 0.239547128 | -0.173715128 | 0.065832 |
| ANXA5 | 0.250580804 | -0.172285866 | 0.078294938 |
| FLNA | -0.2153294 | 0.141551838 | -0.073777562 |
| GIPC1 | 0.181312278 | -0.191255747 | -0.00994347 |
| AURKAIP1 | 0.240671729 | -0.102543709 | 0.138128021 |

| | | | |
|---|---|---|---|
| CCNI | 0.268302738 | -0.208138514 | 0.060164225 |
| SIRPA | -0.188745861 | 0.089443353 | -0.099302508 |
| NDUFAB1 | 0.229804258 | -0.171555678 | 0.05824858 |
| PIM3 | 0.182200168 | -0.190016463 | -0.007816295 |
| NDUFA6 | 0.216134595 | -0.148967246 | 0.067167349 |
| SDCBP | 0.183464941 | -0.123613466 | 0.059851476 |
| MRPL51 | 0.245138331 | -0.149146732 | 0.095991599 |
| ARHGAP29 | 0.254759448 | -0.249764728 | 0.00499472 |
| OSTF1 | 0.140827126 | -0.129297761 | 0.011529365 |
| TMEM61 | 0.142924718 | -0.150194841 | -0.007270123 |
| ATP5PD | 0.235006252 | -0.113052468 | 0.121953784 |
| KRT19 | 0.245640141 | -0.082051866 | 0.163588275 |
| SLC39A13 | 0.179042341 | -0.181042522 | -0.002000181 |
| ERVMER34-1 | 0.129269645 | -0.145390297 | -0.016120652 |
| DVL3 | 0.156317903 | -0.171109222 | -0.014791319 |
| NDUFB10 | 0.243975315 | -0.120650218 | 0.123325098 |
| GUK1 | 0.230799085 | -0.121552448 | 0.109246637 |
| ABHD14B | 0.165039571 | -0.149698616 | 0.015340954 |
| RTN3 | 0.227760561 | -0.259280146 | -0.031519586 |
| PSMC5 | 0.217670545 | -0.185174962 | 0.032495583 |
| RDH10 | 0.161552317 | -0.159770021 | 0.001782296 |
| EID2 | 0.13258436 | -0.135370555 | -0.002786195 |
| PHB2 | 0.205192444 | -0.138955028 | 0.066237415 |
| CIAO2B | 0.230183754 | -0.146230092 | 0.083953662 |
| COX6C | 0.257989463 | -0.10911609 | 0.148873373 |
| MAP4K3 | 0.167939791 | -0.192030715 | -0.024090924 |
| RPL18 | 0.228808253 | -0.117201409 | 0.111606844 |
| MCRIP2 | 0.164943844 | -0.12882845 | 0.036115394 |
| ATP6V1G1 | 0.258202857 | -0.179676314 | 0.078526543 |
| SRP9 | 0.238739498 | -0.111644039 | 0.127095459 |
| ARMC10 | 0.159776465 | -0.133995333 | 0.025781132 |
| ARPC5L | 0.153752946 | -0.128488603 | 0.025264343 |
| RRAGA | 0.182059098 | -0.145158861 | 0.036900237 |
| SSU72 | 0.214682628 | -0.181853745 | 0.032828883 |
| HSP90AB1 | 0.264517672 | -0.216542529 | 0.047975143 |
| NDUFC1 | 0.258943069 | -0.096766141 | 0.162176928 |
| KIZ | 0.144066764 | -0.137648841 | 0.006417923 |
| ZDHHC2 | 0.176995185 | -0.192668549 | -0.015673364 |
| ATP6V0E2 | 0.15328064 | -0.138207621 | 0.015073019 |
| PSMB1 | 0.242610507 | -0.154013591 | 0.088596916 |
| EIF3I | 0.186073107 | -0.116303215 | 0.069769891 |
| SKP1 | 0.261160561 | -0.138201055 | 0.122959506 |
| CA5B | 0.116876687 | -0.117231228 | -0.000354541 |
| CRYL1 | 0.185780219 | -0.136046184 | 0.049734034 |
| KNG1 | 0.220070887 | -0.200557745 | 0.019513141 |
| EIF2S2 | 0.192352198 | -0.174708562 | 0.017643636 |
| BNIP3L | 0.202818264 | -0.184686067 | 0.018132197 |

| | | | |
|---|---|---|---|
| SEC11A | 0.236319338 | -0.171186602 | 0.065132736 |
| DNM1 | -0.130419402 | 0.051920335 | -0.078499067 |
| MRPL52 | 0.217887796 | -0.093370927 | 0.124516869 |
| BCL7C | 0.232464004 | -0.166766731 | 0.065697273 |
| AC009275.1 | 0.095076816 | -0.095076816 | -2.30926E-14 |
| TGFB2 | 0.108251361 | -0.113917127 | -0.005665766 |
| ATP5MC1 | 0.22465353 | -0.064836678 | 0.159816852 |
| TMEM160 | 0.218128169 | -0.210140353 | 0.007987816 |
| NDUFB8 | 0.240865629 | -0.154162699 | 0.08670293 |
| BTF3 | 0.266935832 | -0.142399996 | 0.124535836 |
| ARF4 | 0.191476022 | -0.133653654 | 0.057822368 |
| ADGRF1 | -0.161483251 | 0.060756327 | -0.100726924 |
| PACSIN2 | 0.156133266 | -0.166699249 | -0.010565984 |
| SPPL2A | 0.178819975 | -0.152307781 | 0.026512194 |
| ANXA7 | 0.204506261 | -0.167037833 | 0.037468428 |
| B3GNT3 | 0.105872961 | -0.106075119 | -0.000202158 |
| PLPP5 | 0.130766838 | -0.138401048 | -0.007634209 |
| TUBA1C | 0.16714693 | -0.11015824 | 0.05698869 |
| SLC47A2 | 0.10151175 | -0.101655686 | -0.000143936 |
| EEF1G | 0.245634879 | -0.095229481 | 0.150405398 |
| TECR | 0.221611654 | -0.229832714 | -0.008221061 |
| PSMB5 | 0.213428815 | -0.155230788 | 0.058198026 |
| ATP5PO | 0.22429055 | -0.100157516 | 0.124133034 |
| TP53I11 | 0.123887107 | -0.120268331 | 0.003618776 |
| COBLL1 | 0.259094997 | -0.344983512 | -0.085888515 |
| RAB13 | 0.238428161 | -0.192893828 | 0.045534332 |
| SARAF | 0.238766784 | -0.271398199 | -0.032631416 |
| NDUFA2 | 0.233466383 | -0.162187816 | 0.071278567 |
| HSBP1L1 | 0.172133559 | -0.15458193 | 0.01755163 |
| SERINC2 | 0.191460124 | -0.219883537 | -0.028423413 |
| RPS24 | 0.197680432 | -0.068939412 | 0.12874102 |
| MAP4K3-DT | 0.153881996 | -0.147078837 | 0.006803159 |
| MTCH1 | 0.214114146 | -0.224062206 | -0.00994806 |
| TMEM14C | 0.237915314 | -0.219251557 | 0.018663757 |
| ATP6V0B | 0.237515893 | -0.249102871 | -0.011586979 |
| C19orf53 | 0.235644931 | -0.133021648 | 0.102623283 |
| TOMM6 | 0.242618043 | -0.123611841 | 0.119006202 |
| SURF1 | 0.183504557 | -0.121946645 | 0.061557913 |
| SUCLG1 | 0.217363743 | -0.08695179 | 0.130411952 |
| PROSER2 | 0.150169545 | -0.153353596 | -0.003184051 |
| C1orf43 | 0.205029285 | -0.112707554 | 0.092321731 |
| OPTN | 0.173446094 | -0.149898115 | 0.023547979 |
| LSM7 | 0.213726721 | -0.175065891 | 0.038660829 |
| MLF2 | 0.208227734 | -0.168460055 | 0.039767679 |
| FAM49B | 0.148206398 | -0.13078647 | 0.017419928 |
| TUFM | 0.206671329 | -0.148031709 | 0.05863962 |
| FJX1 | 0.098199951 | -0.09485601 | 0.003343941 |

| | | | |
|---|---|---|---|
| PSMA2 | 0.219631973 | -0.141070577 | 0.078561396 |
| PSMB2 | 0.187197316 | -0.124076125 | 0.063121191 |
| PSME2 | 0.199615671 | -0.128731928 | 0.070883743 |
| TMEM165 | 0.176511407 | -0.188449401 | -0.011937994 |
| TCEA1 | 0.212919016 | -0.183022146 | 0.029896871 |
| PPP1R14B | 0.18766687 | -0.086254781 | 0.101412089 |
| LINC01116 | 0.116537753 | -0.113103869 | 0.003433884 |
| SLTM | 0.190658387 | -0.180798624 | 0.009859763 |
| NDUFA11 | 0.243365584 | -0.147972531 | 0.095393053 |
| ADRM1 | 0.169457291 | -0.117571779 | 0.051885512 |
| COL4A4 | 0.196170302 | -0.249240065 | -0.053069763 |
| C4orf3 | 0.230434722 | -0.178265722 | 0.052169 |
| ESAM | 0.187277406 | -0.214219857 | -0.026942451 |
| EMP1 | -0.176949477 | 0.072512987 | -0.10443649 |
| RPL18A | 0.203022872 | -0.063444129 | 0.139578743 |
| TMEM9B | 0.18898354 | -0.219582228 | -0.030598687 |
| EIF5 | 0.219410672 | -0.206470713 | 0.012939959 |
| SPINT1-AS1 | 0.181296061 | -0.137808647 | 0.043487415 |
| ARL2 | 0.22858748 | -0.122751118 | 0.105836361 |
| ATP5F1B | 0.236846486 | -0.135492347 | 0.101354139 |
| CALCA | 0.111815202 | -0.111815202 | -1.95399E-14 |
| RPL5 | 0.22844503 | -0.124506897 | 0.103938133 |
| SNHG7 | 0.21264495 | -0.170645767 | 0.041999184 |
| CBX3 | 0.202622226 | -0.179847485 | 0.022774741 |
| TXNL4A | 0.181831928 | -0.106497547 | 0.075334381 |
| PNRC2 | 0.210126107 | -0.229966338 | -0.019840231 |
| TMEM38B | 0.215313041 | -0.220918968 | -0.005605927 |
| FAU | 0.226723363 | -0.079500763 | 0.147222599 |
| PNKD | 0.230776252 | -0.216741713 | 0.014034539 |
| ADI1 | 0.19696988 | -0.08588567 | 0.11108421 |
| PGK1 | 0.215023369 | -0.112222828 | 0.102800541 |
| ATG101 | 0.136283169 | -0.109039787 | 0.027243383 |
| SLC9A3-AS1 | -0.163401824 | 0.060617296 | -0.102784528 |
| FAM241A | 0.096358988 | -0.092658398 | 0.00370059 |
| SCGB2A1 | 0.128727078 | -0.125082793 | 0.003644285 |
| ATP5PF | 0.243402141 | -0.102038725 | 0.141363416 |
| GAS6-DT | 0.089706933 | -0.087910461 | 0.001796473 |
| ERH | 0.232552964 | -0.121505234 | 0.11104773 |
| GPC3 | -0.17751241 | 0.166264596 | -0.011247814 |
| CCT5 | 0.177089919 | -0.131851835 | 0.045238084 |
| AMFR | 0.16281253 | -0.199819271 | -0.037006741 |
| NADK2 | 0.125598483 | -0.131223934 | -0.005625451 |
| GOLM1 | 0.181159549 | -0.213429362 | -0.032269813 |
| DHRS7 | 0.201323272 | -0.281617899 | -0.080294628 |
| MTLN | 0.179697279 | -0.138231618 | 0.041465661 |
| YWHAE | 0.252652887 | -0.143672928 | 0.108979959 |
| PPP2R3A | 0.150772381 | -0.166746381 | -0.015974 |

| | | | |
|---|---|---|---|
| UXT | 0.195774 | -0.142120232 | 0.053653768 |
| CLIC1 | 0.235275716 | -0.120876915 | 0.114398801 |
| BHLHE41 | 0.100602914 | -0.096520389 | 0.004082525 |
| NCOA7 | 0.170354795 | -0.183881937 | -0.013527141 |
| RFK | 0.142416421 | -0.132783655 | 0.009632766 |
| NDUFA13 | 0.226163907 | -0.152451661 | 0.073712247 |
| HSPA8 | 0.232664173 | -0.157682383 | 0.07498179 |
| RNF7 | 0.212936351 | -0.124809753 | 0.088126598 |
| SNX5 | 0.194797345 | -0.177626885 | 0.01717046 |
| RNH1 | 0.21176326 | -0.134666913 | 0.077096348 |
| TIFA | 0.111545323 | -0.101064224 | 0.0104811 |
| AC004889.1 | -0.167067186 | 0.076370748 | -0.090696437 |
| CPM | 0.115863996 | -0.113253237 | 0.002610759 |
| ATP5ME | 0.238087863 | -0.087933018 | 0.150154846 |
| SLC25A11 | 0.182021321 | -0.141199781 | 0.040821541 |
| RTN4 | 0.243991339 | -0.411151328 | -0.167159989 |
| BAD | 0.178619313 | -0.1248208 | 0.053798513 |
| CCT3 | 0.197774857 | -0.105486914 | 0.092287943 |
| NDUFB7 | 0.227299055 | -0.082830861 | 0.144468193 |
| SERP1 | 0.214572229 | -0.162989629 | 0.051582599 |
| DDX3X | 0.203573716 | -0.179144169 | 0.024429548 |
| IGBP1 | 0.162987784 | -0.125031679 | 0.037956105 |
| UQCC2 | 0.171987801 | -0.132829421 | 0.03915838 |
| C9orf16 | 0.18858296 | -0.148727365 | 0.039855595 |
| HSPH1 | 0.157850037 | -0.174795814 | -0.016945777 |
| PCDH9 | 0.119741634 | -0.113940984 | 0.00580065 |
| HSP90AA1 | 0.248987056 | -0.304583044 | -0.055595988 |
| MRPS34 | 0.196203056 | -0.161225966 | 0.034977089 |
| SH3BGRL | 0.18188246 | -0.157020719 | 0.024861741 |
| MRPL20 | 0.21298707 | -0.09960067 | 0.1133864 |
| PGP | 0.147437882 | -0.162917715 | -0.015479833 |
| RAD23A | 0.187136873 | -0.129860109 | 0.057276765 |
| UBE2E3 | 0.191296523 | -0.134827916 | 0.056468608 |
| ID2 | -0.143819933 | 0.136095055 | -0.007724879 |
| DCTN3 | 0.188265543 | -0.131640827 | 0.056624716 |
| PDHB | 0.160682963 | -0.128962496 | 0.031720467 |
| SNRPD2 | 0.199856073 | -0.10243764 | 0.097418432 |
| AIF1L | 0.248341577 | -0.296381066 | -0.048039489 |
| UACA | 0.162190762 | -0.184490382 | -0.02229962 |
| RPL23A | 0.232418281 | -0.089941247 | 0.142477034 |
| RAP1GAP | 0.159766856 | -0.158993133 | 0.000773723 |
| SMAD7 | -0.184132328 | 0.066724071 | -0.117408257 |
| PHF10 | 0.141297629 | -0.111685581 | 0.029612048 |
| FUNDC2 | 0.182901207 | -0.157417304 | 0.025483903 |
| TSG101 | 0.163755621 | -0.127852164 | 0.035903457 |
| PFDN2 | 0.197200959 | -0.123668054 | 0.073532905 |
| GADD45B | 0.229978692 | -0.265452394 | -0.035473702 |

| | | | |
|---|---|---|---|
| LAMB1 | -0.170264029 | 0.108106777 | -0.062157252 |
| MTSS1 | -0.163422497 | 0.087704018 | -0.075718479 |
| FEZ2 | 0.157906311 | -0.171207522 | -0.013301212 |
| VPS29 | 0.189878615 | -0.132483859 | 0.057394756 |
| GALNT9 | 0.087776359 | -0.089287719 | -0.001511361 |
| MRPL36 | 0.163691739 | -0.117437406 | 0.046254333 |
| LINC01426 | 0.080950893 | -0.080950893 | -2.30926E-14 |
| LIMA1 | 0.163098123 | -0.215451042 | -0.052352919 |
| CISD1 | 0.183585359 | -0.119109638 | 0.064475721 |
| EMC7 | 0.184233351 | -0.202163683 | -0.017930332 |
| SEC61B | 0.222930691 | -0.091538224 | 0.131392468 |
| OST4 | 0.23726167 | -0.112072991 | 0.125188679 |
| G3BP1 | 0.177547097 | -0.164846815 | 0.012700282 |
| ODC1 | -0.220107058 | 0.317021589 | 0.096914531 |
| SAP18 | 0.237433321 | -0.117916379 | 0.119516942 |
| APIP | 0.153692739 | -0.156949484 | -0.003256746 |
| PARD6B | 0.215601514 | -0.241663793 | -0.026062279 |
| SLC25A4 | 0.180859286 | -0.135484503 | 0.045374782 |
| TM2D1 | 0.167831561 | -0.200773178 | -0.032941617 |
| FBXO44 | 0.113791309 | -0.092188387 | 0.021602922 |
| BAMBI | 0.119512228 | -0.10549849 | 0.014013738 |
| CTSC | -0.191786521 | 0.105485682 | -0.086300839 |
| SMDT1 | 0.230755446 | -0.187844208 | 0.042911238 |
| CCDC47 | 0.18450284 | -0.236684029 | -0.052181189 |
| NGRN | 0.191690099 | -0.209903897 | -0.018213798 |
| UBL5 | 0.237116752 | -0.083618221 | 0.15349853 |
| DDX52 | 0.150512053 | -0.171164876 | -0.020652823 |
| HIST1H1E | 0.118831034 | -0.119308253 | -0.000477219 |
| REEP3 | 0.133040676 | -0.131125818 | 0.001914858 |
| CD9 | 0.246611974 | -0.405913322 | -0.159301349 |
| RPS27L | 0.227038125 | -0.074537497 | 0.152500629 |
| G3BP2 | 0.167252604 | -0.135268319 | 0.031984285 |
| PSMA5 | 0.183246614 | -0.13539875 | 0.047847864 |
| EEF1B2 | 0.239687937 | -0.157271382 | 0.082416555 |
| SHISA3 | 0.112989567 | -0.10265992 | 0.010329648 |
| TSPAN1 | 0.123400811 | -0.08434097 | 0.039059841 |
| CCT2 | 0.172215937 | -0.137107842 | 0.035108095 |
| CD2AP | 0.16340018 | -0.119616082 | 0.043784098 |
| PPP1R14C | -0.151114155 | 0.136933437 | -0.014180718 |
| GSDME | 0.081203902 | -0.080251224 | 0.000952678 |
| SELENOM | 0.220815085 | -0.216730461 | 0.004084624 |
| AL157373.2 | 0.085375116 | -0.084117329 | 0.001257788 |
| RPS27A | 0.157916801 | -0.06999672 | 0.087920081 |
| NFKBIA | 0.194686933 | -0.199102862 | -0.004415928 |
| CCDC107 | 0.144393117 | -0.104749515 | 0.039643602 |
| USP46 | 0.141214047 | -0.149335863 | -0.008121816 |
| CHMP2A | 0.205574951 | -0.121660411 | 0.083914541 |

| | | | |
|---|---|---|---|
| ZNHIT1 | 0.202993256 | -0.133155588 | 0.069837668 |
| MPP6 | 0.112567288 | -0.112789681 | -0.000222393 |
| WSB2 | 0.138116984 | -0.111245043 | 0.026871941 |
| NUTF2 | 0.164370448 | -0.073719823 | 0.090650625 |
| SEC14L1 | 0.156176381 | -0.165868601 | -0.00969222 |
| SSNA1 | 0.184898916 | -0.181848535 | 0.003050382 |
| LAMTOR4 | 0.214522937 | -0.14762345 | 0.066899487 |
| PHB | 0.201773849 | -0.145020464 | 0.056753385 |
| SOD2 | 0.179344094 | -0.084232662 | 0.095111432 |
| PDHA1 | 0.163599662 | -0.140152762 | 0.0234469 |
| NDUFA1 | 0.226841979 | -0.176888699 | 0.04995328 |
| SELENOH | 0.218135113 | -0.157703419 | 0.060431693 |
| KIF2A | 0.116031601 | -0.112782428 | 0.003249173 |
| RPSA | 0.230102672 | -0.08531156 | 0.144791112 |
| ILVBL | 0.142627622 | -0.128401039 | 0.014226583 |
| SORL1 | 0.186986189 | -0.30209188 | -0.115105691 |
| EIF5A | 0.196085513 | -0.107544476 | 0.088541038 |
| SRI | 0.201040102 | -0.146384572 | 0.05465553 |
| ISCU | 0.21863066 | -0.083039259 | 0.135591401 |
| SNX3 | 0.201460158 | -0.110259889 | 0.091200269 |
| NDUFS3 | 0.165074179 | -0.112234186 | 0.052839993 |
| CYCS | 0.193863024 | -0.095693051 | 0.098169974 |
| CRIM1-DT | 0.151748321 | -0.163098294 | -0.011349972 |
| SUMO1 | 0.210328522 | -0.144960367 | 0.065368155 |
| CREG1 | 0.199763874 | -0.241712081 | -0.041948207 |
| FKBP1A | 0.18342812 | -0.125340719 | 0.0580874 |
| C11orf58 | 0.212923446 | -0.173259777 | 0.039663668 |
| HTATSF1 | 0.174402952 | -0.217525804 | -0.043122852 |
| GRPEL1 | 0.124880916 | -0.103742924 | 0.021137992 |
| SMIM5 | 0.151789963 | -0.15414277 | -0.002352807 |
| MDH2 | 0.182881654 | -0.116961718 | 0.065919936 |
| JAK1 | 0.197638857 | -0.207346571 | -0.009707714 |
| GPC4 | -0.166222091 | 0.075123023 | -0.091099069 |
| RTF2 | 0.170718326 | -0.164368466 | 0.00634986 |
| SLIRP | 0.197515043 | -0.103075233 | 0.09443981 |
| GADD45GIP1 | 0.195370805 | -0.094273277 | 0.101097528 |
| ZMAT3 | 0.112710803 | -0.102774551 | 0.009936252 |
| GSTK1 | 0.176934051 | -0.183390758 | -0.006456707 |
| PANTR1 | 0.169452769 | -0.172391924 | -0.002939156 |
| XRCC5 | 0.204958109 | -0.209567101 | -0.004608992 |
| RAB5C | 0.171138189 | -0.116842849 | 0.05429534 |
| FAM162A | 0.178028429 | -0.108242783 | 0.069785646 |
| F2RL1 | 0.12823054 | -0.129039597 | -0.000809057 |
| TMEM106B | 0.185376884 | -0.245162874 | -0.05978599 |
| PRDX6 | 0.222531367 | -0.071643764 | 0.150887603 |
| SCIN | 0.207048767 | -0.272619691 | -0.065570924 |
| GRB7 | 0.148786688 | -0.16290428 | -0.014117591 |

| | | | |
|---|---|---|---|
| NDUFC2 | 0.218316942 | -0.19683819 | 0.021478752 |
| FRY | 0.151124934 | -0.183311107 | -0.032186173 |
| MPC2 | 0.232905619 | -0.204920112 | 0.027985506 |
| SUMO3 | 0.167990423 | -0.10354194 | 0.064448483 |
| SELENOT | 0.180563337 | -0.172283727 | 0.008279609 |
| DNAJC22 | -0.115578459 | 0.068706499 | -0.04687196 |
| WDR83OS | 0.219293883 | -0.200182702 | 0.019111181 |
| CD63 | 0.22599059 | -0.28080555 | -0.05481496 |
| RTRAF | 0.184056661 | -0.125196785 | 0.058859876 |
| KCNJ13 | 0.078268928 | -0.076518477 | 0.001750451 |
| TMEM45B | 0.082764361 | -0.083212693 | -0.000448333 |
| STUB1 | 0.197747977 | -0.111918374 | 0.085829603 |
| LRBA | 0.150937459 | -0.167098164 | -0.016160705 |
| ANXA3 | -0.165463101 | 0.164032017 | -0.001431083 |
| ATP5MC2 | 0.227435693 | -0.089732796 | 0.137702897 |
| CAB39L | 0.104819602 | -0.118695506 | -0.013875904 |
| GNAS | 0.230857881 | -0.248544425 | -0.017686545 |
| SINHCAF | 0.161953273 | -0.167542467 | -0.005589194 |
| HEIH | 0.141764556 | -0.114924976 | 0.026839581 |
| COX7A2L | 0.206906747 | -0.16364261 | 0.043264137 |
| PSMD9 | 0.156657926 | -0.11176025 | 0.044897676 |
| SUSD4 | 0.107229829 | -0.118728411 | -0.011498582 |
| NOP10 | 0.196680719 | -0.11111397 | 0.085566749 |
| C15orf61 | 0.128570951 | -0.095844207 | 0.032726744 |
| TXNL1 | 0.175768725 | -0.134398863 | 0.041369862 |
| FGF18 | 0.07621856 | -0.075109842 | 0.001108718 |
| LINC01965 | 0.083085664 | -0.089532666 | -0.006447002 |
| PPP2CB | 0.184756005 | -0.1680885 | 0.016667505 |
| GPCPD1 | -0.101769956 | 0.050241658 | -0.051528299 |
| RAC1 | 0.216572607 | -0.162103786 | 0.054468821 |
| NDUFS6 | 0.207018428 | -0.121904319 | 0.085114109 |
| FDX1 | 0.151830977 | -0.077521114 | 0.074309862 |
| ST13 | 0.210641123 | -0.12611052 | 0.084530603 |
| AIP | 0.150891522 | -0.135815595 | 0.015075927 |
| MZT2A | 0.179708238 | -0.150010617 | 0.029697621 |
| TNIP2 | 0.112562337 | -0.101675835 | 0.010886502 |
| NUDT16L1 | 0.155199868 | -0.137675045 | 0.017524823 |
| FOXC1 | -0.207457873 | 0.164453509 | -0.043004364 |
| TPM1 | 0.229231099 | -0.242197585 | -0.012966487 |
| COPZ1 | 0.16718242 | -0.135561522 | 0.031620898 |
| STAT3 | 0.164061665 | -0.195768376 | -0.031706711 |
| RAP1B | 0.154890735 | -0.134502613 | 0.020388122 |
| GOT1 | 0.139846057 | -0.112314607 | 0.02753145 |
| LSM2 | 0.15941567 | -0.099629135 | 0.059786535 |
| DCTPP1 | 0.131983972 | -0.093806777 | 0.038177195 |
| C22orf39 | 0.132827026 | -0.093676537 | 0.039150489 |
| MGST3 | 0.221544972 | -0.100298742 | 0.12124623 |

| | | | |
|---|---|---|---|
| CYP2S1 | 0.075885361 | -0.073699157 | 0.002186204 |
| GSN | 0.136039953 | -0.137793648 | -0.001753695 |
| CADM1 | -0.181630851 | 0.050000482 | -0.13163037 |
| PKD2 | 0.150375816 | -0.161637186 | -0.011261371 |
| ATP5IF1 | 0.219805823 | -0.085210391 | 0.134595432 |
| SLC25A48 | 0.080799388 | -0.074296194 | 0.006503194 |
| C11orf74 | 0.123353113 | -0.095051455 | 0.028301658 |
| SMCO4 | 0.143253675 | -0.147460049 | -0.004206374 |
| GAL | 0.078472117 | -0.078472117 | -2.30926E-14 |
| AIG1 | 0.166938344 | -0.157091193 | 0.009847151 |
| ATP6V0E1 | 0.220090839 | -0.259184668 | -0.03909383 |
| ANO6 | 0.126762019 | -0.14567909 | -0.01891707 |
| SERPINF2 | -0.116306637 | 0.059941666 | -0.05636497 |
| MORF4L1 | 0.21919631 | -0.142790337 | 0.076405973 |
| RPS4Y1 | 0.212819571 | -0.430695065 | -0.217875493 |
| TMED9 | 0.173116481 | -0.178416358 | -0.005299877 |
| VPS28 | 0.199994832 | -0.102687276 | 0.097307556 |
| TRAPPC4 | 0.158586051 | -0.120415072 | 0.038170979 |
| NARS | 0.160716678 | -0.142075255 | 0.018641423 |
| TAF10 | 0.163976802 | -0.148665316 | 0.015311485 |
| PPP1R7 | 0.137416614 | -0.104329753 | 0.033086861 |
| ITM2C | 0.167196031 | -0.124529149 | 0.042666881 |
| ERMP1 | 0.188217289 | -0.288152698 | -0.099935409 |
| DSTN | 0.218602496 | -0.130183542 | 0.088418954 |
| MAFA | 0.067633122 | -0.06609673 | 0.001536393 |
| PRSS35 | 0.133619281 | -0.123530884 | 0.010088397 |
| MDH1 | 0.183774274 | -0.080317824 | 0.10345645 |
| SNX6 | 0.167332193 | -0.151935901 | 0.015396292 |
| RPN2 | 0.169027109 | -0.186928019 | -0.01790091 |
| PEPD | 0.143648249 | -0.070140686 | 0.073507563 |
| GABARAPL2 | 0.216538169 | -0.183137061 | 0.033401108 |
| TMEM134 | 0.151380239 | -0.099256663 | 0.052123576 |
| TFAM | 0.125736907 | -0.110838278 | 0.014898629 |
| CCT4 | 0.160741942 | -0.091363707 | 0.069378235 |
| HNRNPF | 0.178640383 | -0.120742664 | 0.057897718 |
| GET3 | 0.148588224 | -0.117684129 | 0.030904095 |
| FBXW11 | 0.147056689 | -0.170241368 | -0.023184679 |
| JPT1 | 0.142225317 | -0.109576744 | 0.032648573 |
| GCNT4 | -0.098746976 | 0.067529851 | -0.031217126 |
| TRAPPC1 | 0.176978386 | -0.073303238 | 0.103675147 |
| ATP5F1A | 0.188694797 | -0.093741673 | 0.094953123 |
| LSM4 | 0.179962868 | -0.135606244 | 0.044356624 |
| MAP1LC3B | 0.159731465 | -0.128004373 | 0.031727092 |
| CREB3 | 0.124404406 | -0.119167423 | 0.005236983 |
| NAP1L1 | 0.19153049 | -0.194779259 | -0.003248769 |
| COPS9 | 0.18872924 | -0.100813424 | 0.087915816 |
| CKS1B | 0.127123838 | -0.114484009 | 0.012639829 |

| | | | |
|---|---|---|---|
| GSTO1 | 0.170725165 | -0.119667325 | 0.05105784 |
| MOB1A | 0.156921242 | -0.110728785 | 0.046192457 |
| HSPA4 | 0.154685082 | -0.14482207 | 0.009863012 |
| CCPG1 | 0.177665955 | -0.229094546 | -0.051428591 |
| PSMD11 | 0.139071359 | -0.123416816 | 0.015654544 |
| RTCB | 0.125288387 | -0.099930686 | 0.025357701 |
| SPINT2 | 0.218618034 | -0.317583204 | -0.09896517 |
| RABAC1 | 0.195974524 | -0.225235789 | -0.029261265 |
| UBE2A | 0.134522572 | -0.107818618 | 0.026703954 |
| NDUFAF4 | 0.109694041 | -0.095271747 | 0.014422294 |
| ATOX1 | 0.194354587 | -0.08789807 | 0.106456517 |
| SLCO1A2 | 0.080408503 | -0.079147878 | 0.001260625 |
| UBASH3B | 0.071397953 | -0.06799149 | 0.003406463 |
| SEC61G | 0.19802917 | -0.069143127 | 0.128886044 |
| HSPB1 | 0.184190656 | -0.051824369 | 0.132366287 |
| STAU1 | 0.148360534 | -0.135615739 | 0.012744795 |
| MAF1 | 0.159984506 | -0.143834135 | 0.01615037 |
| HMGN1 | 0.211082471 | -0.155575631 | 0.055506839 |
| RHEX | -0.101331526 | 0.058908383 | -0.042423143 |
| ENSA | 0.165342779 | -0.155235959 | 0.010106821 |
| LINC02672 | 0.068994958 | -0.065643864 | 0.003351094 |
| RNF181 | 0.162305723 | -0.078101003 | 0.08420472 |
| ATP6V0D1 | 0.146872756 | -0.079356995 | 0.067515761 |
| RAB11B | 0.157461139 | -0.118957996 | 0.038503143 |
| PSMG2 | 0.151702843 | -0.098077535 | 0.053625308 |
| TPMT | 0.121404213 | -0.084262275 | 0.037141937 |
| H1F0 | 0.172008282 | -0.14981722 | 0.022191062 |
| MRPL32 | 0.137711008 | -0.099352734 | 0.038358274 |
| MEA1 | 0.141947991 | -0.111562661 | 0.030385329 |
| REXO2 | 0.156064217 | -0.10174756 | 0.054316657 |
| B3GAT3 | 0.130683506 | -0.163969265 | -0.033285758 |
| RAB25 | 0.146136416 | -0.108231422 | 0.037904995 |
| MRFAP1 | 0.1962779 | -0.146916961 | 0.049360939 |
| LINC00472 | -0.22083954 | -0.077727494 | -0.298567035 |
| CCT7 | 0.152789462 | -0.102669716 | 0.050119746 |
| MARCKS | -0.138131374 | 0.136913534 | -0.00121784 |
| TSHZ2 | -0.102474601 | 0.056737339 | -0.045737262 |
| CERNA2 | -0.094566225 | 0.039306286 | -0.055259939 |
| AJAP1 | 0.066915828 | -0.061195501 | 0.005720327 |
| TCEAL4 | 0.184340715 | -0.121861311 | 0.062479404 |
| DTD1 | 0.136099934 | -0.122366732 | 0.013733202 |
| RHOC | 0.147051041 | -0.05003094 | 0.097020102 |
| UBE2D2 | 0.16145529 | -0.086801584 | 0.074653706 |
| PLOD2 | 0.156481337 | -0.170881842 | -0.014400506 |
| SNTB2 | 0.116747637 | -0.137040991 | -0.020293354 |
| PTTG1IP | 0.196274813 | -0.298376992 | -0.102102179 |
| UBE2H | 0.142107697 | -0.153539578 | -0.011431881 |

| | | | |
|---|---|---|---|
| DPP7 | 0.16351619 | -0.183888221 | -0.020372031 |
| NDUFB5 | 0.174809784 | -0.10880307 | 0.066006714 |
| KDELR1 | 0.199174875 | -0.214044838 | -0.014869963 |
| CCT8 | 0.161338821 | -0.125924712 | 0.03541411 |
| ALDH3A2 | 0.153788797 | -0.112877125 | 0.040911673 |
| SBDS | 0.188668102 | -0.154377831 | 0.034290271 |
| NDUFA12 | 0.184116956 | -0.133340457 | 0.050776499 |
| PLP2 | 0.136421689 | -0.133187462 | 0.003234227 |
| MLX | 0.113189681 | -0.095007854 | 0.018181827 |
| SPECC1L | 0.127295433 | -0.14653 | -0.019234566 |
| AKAP9 | 0.206002618 | -0.329912726 | -0.123910108 |
| MGAT1 | 0.121295309 | -0.121254783 | 4.05263E-05 |
| NCL | 0.202943484 | -0.18806024 | 0.014883244 |
| MKLN1 | 0.141555924 | -0.155547603 | -0.013991679 |
| TMEM171 | 0.099674866 | -0.109828701 | -0.010153835 |
| PTPN3 | 0.131961112 | -0.137796402 | -0.00583529 |
| RNASEK | 0.199028532 | -0.150944854 | 0.048083678 |
| ARID5B | -0.161850745 | 0.117820909 | -0.044029836 |
| WDR72 | 0.088033848 | -0.068297232 | 0.019736616 |
| CCDC186 | 0.145476507 | -0.165771476 | -0.020294969 |
| EIF6 | 0.169905107 | -0.063247942 | 0.106657165 |
| POLE4 | 0.122765923 | -0.061106819 | 0.061659104 |
| CFDP1 | 0.190520536 | -0.131774822 | 0.058745714 |
| FDFT1 | 0.109020943 | -0.088492123 | 0.02052882 |
| SLC25A6 | 0.205794929 | -0.131515146 | 0.074279783 |
| ENTPD8 | 0.063777115 | -0.061660128 | 0.002116987 |
| MTHFS | 0.134798649 | -0.095777613 | 0.039021036 |
| PHC2 | 0.129766428 | -0.101579044 | 0.028187383 |
| TCIM | 0.154784739 | -0.192429123 | -0.037644384 |
| VBP1 | 0.120758652 | -0.079822027 | 0.040936625 |
| PLS1 | 0.106312884 | -0.075936822 | 0.030376062 |
| KIAA2026 | 0.136776382 | -0.171840364 | -0.035063982 |
| EIF1 | 0.186235729 | -0.077513015 | 0.108722715 |
| PTMS | 0.206895401 | -0.114241788 | 0.092653613 |
| SPAG7 | 0.164101492 | -0.098056957 | 0.066044535 |
| LRRC2 | 0.071270309 | -0.07186018 | -0.000589871 |
| LITAF | 0.176729061 | -0.163298949 | 0.013430112 |
| ORMDL1 | 0.154342351 | -0.14904955 | 0.005292801 |
| LAMTOR1 | 0.181227361 | -0.13330595 | 0.047921411 |
| COA4 | 0.148702444 | -0.086010524 | 0.06269192 |
| SDHD | 0.183486908 | -0.145806779 | 0.037680129 |
| MRPL41 | 0.180188482 | -0.076766889 | 0.103421593 |
| SEPTIN11 | 0.142083136 | -0.100473961 | 0.041609175 |
| RBM4 | 0.17111499 | -0.176505079 | -0.005390089 |
| GALNT14 | -0.148117314 | 0.15277689 | 0.004659576 |
| SPECC1 | 0.061198762 | -0.061198762 | -2.13163E-14 |
| FBLN5 | 0.072341398 | -0.072437027 | -9.56292E-05 |

| | | | |
|---|---|---|---|
| MCUR1 | 0.141441948 | -0.116233899 | 0.025208049 |
| EHD4 | 0.0779095 | -0.072974846 | 0.004934653 |
| MPP7 | 0.104049937 | -0.099421278 | 0.004628659 |
| PAIP1 | 0.129157501 | -0.08117696 | 0.047980541 |
| COPS6 | 0.164554104 | -0.150805775 | 0.013748329 |
| MRPS15 | 0.124127119 | -0.066381997 | 0.057745122 |
| C11orf52 | 0.114579017 | -0.094709839 | 0.019869178 |
| LSM1 | 0.139270185 | -0.140645896 | -0.001375711 |
| SET | 0.207657147 | -0.174645139 | 0.033012008 |
| PTPN14 | 0.15434756 | -0.213649285 | -0.059301725 |
| FAM3B | 0.115468518 | -0.144328488 | -0.02885997 |
| VDAC3 | 0.155668267 | -0.113685582 | 0.041982685 |
| BAX | 0.140208096 | -0.10128698 | 0.038921115 |
| IRX2 | 0.139074132 | -0.162912222 | -0.02383809 |
| SNU13 | 0.186925189 | -0.15861119 | 0.028314 |
| NAXE | 0.134016625 | -0.09662698 | 0.037389645 |
| VEGFB | 0.158259971 | -0.091408222 | 0.066851749 |
| SEC14L6 | 0.071448191 | -0.068835838 | 0.002612353 |
| CYC1 | 0.161201453 | -0.118719828 | 0.042481626 |
| SSR4 | 0.197686198 | -0.179054762 | 0.018631436 |
| TMEM238 | 0.173341422 | -0.150766094 | 0.022575328 |
| GSTT2B | 0.080236463 | -0.058654832 | 0.021581631 |
| TIMM17B | 0.153970368 | -0.149006572 | 0.004963796 |
| POLR2K | 0.174806895 | -0.116248892 | 0.058558003 |
| PPP2R2B | 0.102182041 | -0.109368349 | -0.007186308 |
| PRKRA | 0.103344677 | -0.082639152 | 0.020705525 |
| COMMD2 | 0.143559314 | -0.127003694 | 0.01655562 |
| FAM50A | 0.134189008 | -0.109538481 | 0.024650528 |
| MRAS | 0.082507059 | -0.059303343 | 0.023203716 |
| OXR1 | -0.174658532 | 0.059897872 | -0.11476066 |
| COTL1 | 0.112007725 | -0.06964516 | 0.042362565 |
| FGF9 | -0.07852156 | 0.03230271 | -0.04621885 |
| RALA | 0.102875309 | -0.067403453 | 0.035471855 |
| CAPZB | 0.158680314 | -0.093020825 | 0.06565949 |
| DDIT4 | 0.077805861 | -0.074121817 | 0.003684044 |
| EXOSC7 | 0.12733712 | -0.116107957 | 0.011229162 |
| ADPRHL2 | 0.094690114 | -0.080115628 | 0.014574486 |
| LRATD2 | -0.156442633 | 0.099141487 | -0.057301146 |
| CDC25B | 0.074582928 | -0.062548273 | 0.012034655 |
| DEK | 0.177874215 | -0.175998294 | 0.001875921 |
| DNAJA2 | 0.148677846 | -0.136969208 | 0.011708638 |
| PTBP3 | 0.14100474 | -0.132068572 | 0.008936167 |
| CAPZA2 | 0.164965418 | -0.104549816 | 0.060415602 |
| CLTB | 0.171563077 | -0.160328027 | 0.01123505 |
| GDI2 | 0.137232196 | -0.092390662 | 0.044841534 |
| CAAP1 | 0.114186943 | -0.132115136 | -0.017928194 |
| NAA10 | 0.128774975 | -0.087912804 | 0.040862171 |

| | | | |
|---|---|---|---|
| CHMP2B | 0.123506397 | -0.101630891 | 0.021875507 |
| TAX1BP1 | 0.166874538 | -0.155604438 | 0.0112701 |
| CD151 | 0.180804974 | -0.22559327 | -0.044788296 |
| C1orf115 | 0.118857266 | -0.099982749 | 0.018874517 |
| CADPS2 | 0.099129058 | -0.100942233 | -0.001813175 |
| SLC39A1 | 0.148892229 | -0.154562212 | -0.005669983 |
| IFRD1 | 0.096952053 | -0.102792064 | -0.005840011 |
| TMEM219 | 0.167143432 | -0.125069712 | 0.04207372 |
| POP5 | 0.133381571 | -0.105814392 | 0.027567179 |
| CCND1 | -0.191595523 | 0.080834742 | -0.110760782 |
| ADK | 0.114303283 | -0.124536465 | -0.010233183 |
| TMA7 | 0.205345714 | -0.108156673 | 0.097189042 |
| SMIM22 | 0.070936437 | -0.063343272 | 0.007593164 |
| LMTK3 | 0.092501512 | -0.079043097 | 0.013458415 |
| EIF3F | 0.170588102 | -0.106679165 | 0.063908937 |
| KPNA2 | 0.076340628 | -0.062395108 | 0.01394552 |
| IRX1 | 0.141663748 | -0.146225993 | -0.004562244 |
| SELENOK | 0.170047224 | -0.122341701 | 0.047705523 |
| MRPL22 | 0.130353936 | -0.05546386 | 0.074890076 |
| PTS | 0.148518049 | -0.114897154 | 0.033620894 |
| THBS1 | -0.161305798 | 0.06735796 | -0.093947839 |
| NUP93 | 0.084726997 | -0.073342753 | 0.011384243 |
| AKIP1 | 0.094305814 | -0.088092156 | 0.006213658 |
| RAB18 | 0.143582077 | -0.134547665 | 0.009034411 |
| SNRPB | 0.164260767 | -0.088974612 | 0.075286156 |
| COQ10B | 0.101220747 | -0.094742615 | 0.006478132 |
| ZRSR2 | 0.086375592 | -0.071631269 | 0.014744323 |
| PAX2 | 0.162058351 | -0.208509162 | -0.04645081 |
| PIGH | 0.124099856 | -0.145871919 | -0.021772064 |
| RAB14 | 0.163582055 | -0.152421111 | 0.011160944 |
| AK2 | 0.137182765 | -0.085354904 | 0.051827861 |
| FXYD3 | 0.069673183 | -0.072135111 | -0.002461927 |
| MRPS35 | 0.12425538 | -0.106700241 | 0.017555139 |
| RRBP1 | 0.128264709 | -0.120608406 | 0.007656303 |
| STOM | -0.133177116 | 0.065271401 | -0.067905715 |
| IDH1 | 0.090046196 | -0.078000699 | 0.012045497 |
| GCA | 0.114091888 | -0.090303834 | 0.023788054 |
| ARL16 | 0.128556682 | -0.051018658 | 0.077538024 |
| MTCH2 | 0.11983299 | -0.040343078 | 0.079489912 |
| MRPL19 | 0.137059708 | -0.130068776 | 0.006990932 |
| YWHAQ | 0.16915866 | -0.125647259 | 0.043511402 |
| MTX2 | 0.125333026 | -0.096083571 | 0.029249456 |
| TOMM7 | 0.197878291 | -0.062061153 | 0.135817139 |
| UCHL3 | 0.097454795 | -0.07307339 | 0.024381405 |
| CDC123 | 0.113360437 | -0.093582277 | 0.01977816 |
| PLEKHB1 | 0.081480298 | -0.073374358 | 0.00810594 |
| BEX4 | 0.190413109 | -0.147379269 | 0.04303384 |

| | | | |
|---|---|---|---|
| CMPK1 | 0.141525552 | -0.095339653 | 0.046185899 |
| SPCS2 | 0.189228465 | -0.208800009 | -0.019571545 |
| TEX264 | 0.142838579 | -0.133888114 | 0.008950465 |
| PPM1G | 0.1494885 | -0.157434222 | -0.007945722 |
| HLA-DOA | -0.088769871 | 0.042313259 | -0.046456612 |
| WASL | 0.146704841 | -0.141931624 | 0.004773217 |
| TMPRSS4 | 0.068234994 | -0.068701682 | -0.000466688 |
| PACSIN3 | 0.108925581 | -0.07929154 | 0.029634041 |
| C14orf132 | 0.0796856 | -0.090688509 | -0.01100291 |
| THYN1 | 0.121127483 | -0.096350825 | 0.024776658 |
| CENPC | 0.128968581 | -0.156638308 | -0.027669727 |
| KLF5 | 0.100627435 | -0.091099148 | 0.009528287 |
| NDUFS4 | 0.147334416 | -0.086840807 | 0.06049361 |
| VSIG10 | 0.11519235 | -0.148870122 | -0.033677771 |
| MECOM | 0.126218029 | -0.176884567 | -0.050666538 |
| MRPL21 | 0.123041463 | -0.050915558 | 0.072125905 |
| CDV3 | 0.138978353 | -0.119009845 | 0.019968507 |
| COMMD6 | 0.193029638 | -0.059416952 | 0.133612686 |
| NDUFA4 | 0.176838911 | -0.168427218 | 0.008411694 |
| SSB | 0.159304692 | -0.126449526 | 0.032855166 |
| UBTD1 | 0.101660772 | -0.089447514 | 0.012213258 |
| ACBD5 | 0.119568459 | -0.110215035 | 0.009353424 |
| CYFIP2 | 0.157130015 | -0.185403329 | -0.028273314 |
| CALM2 | 0.204212208 | -0.155685547 | 0.048526661 |
| FAM136A | 0.110872702 | -0.088958591 | 0.021914111 |
| DDT | 0.185583213 | -0.05942335 | 0.126159863 |
| SNX2 | 0.158030667 | -0.148676291 | 0.009354376 |
| SSX2IP | 0.113003782 | -0.107268363 | 0.00573542 |
| CCT6A | 0.142782825 | -0.103581734 | 0.039201091 |
| C14orf119 | 0.111841563 | -0.079920617 | 0.031920946 |
| COA8 | 0.122822654 | -0.092884956 | 0.029937698 |
| SPCS3 | 0.123618197 | -0.132180564 | -0.008562366 |
| UCP2 | 0.075691285 | -0.063655325 | 0.01203596 |
| CCDC6 | 0.114679656 | -0.10777363 | 0.006906026 |
| MRPS12 | 0.131974834 | -0.071839396 | 0.060135437 |
| SCOC | 0.160963308 | -0.112828534 | 0.048134774 |
| ANAPC11 | 0.166329168 | -0.110151071 | 0.056178097 |
| FAM131C | 0.06137157 | -0.05591418 | 0.00545739 |
| PIN1 | 0.152891473 | -0.146920404 | 0.005971068 |
| DDX39A | 0.082283931 | -0.069083641 | 0.01320029 |
| PDIA3 | 0.167627347 | -0.209240636 | -0.041613289 |
| MICOS10 | 0.182162292 | -0.097089866 | 0.085072427 |
| UFC1 | 0.159524285 | -0.069699281 | 0.089825004 |
| MAB21L4 | 0.063053019 | -0.062333262 | 0.000719757 |
| SPINT1 | 0.148696481 | -0.183447208 | -0.034750728 |
| LAGE3 | 0.104749424 | -0.09097297 | 0.013776454 |
| ACP1 | 0.15278096 | -0.124728155 | 0.028052805 |

| | | | |
|---|---|---|---|
| ISCA2 | 0.126727092 | -0.079481302 | 0.04724579 |
| HLTF | 0.115674569 | -0.124689443 | -0.009014874 |
| TMEM223 | 0.10356366 | -0.0950756 | 0.00848806 |
| SREBF2 | 0.113409187 | -0.14081421 | -0.027405024 |
| ACYP1 | 0.104853367 | -0.09083901 | 0.014014358 |
| STOML2 | 0.140146568 | -0.109000775 | 0.031145793 |
| ANXA9 | 0.06552235 | -0.056002021 | 0.009520329 |
| SKAP1 | 0.106062183 | -0.124046221 | -0.017984037 |
| TUBB | 0.179623521 | -0.09087186 | 0.088751662 |
| DOCK1 | 0.137772382 | -0.151819975 | -0.014047593 |
| SSBP4 | 0.127685724 | -0.105178978 | 0.022506745 |
| SUDS3 | 0.125189826 | -0.131202087 | -0.006012261 |
| IGFBP6 | 0.073262009 | -0.062841519 | 0.01042049 |
| PHAX | 0.115180203 | -0.111001337 | 0.004178865 |
| CYYR1 | 0.059381034 | -0.059491232 | -0.000110198 |
| MRPS36 | 0.156867503 | -0.06158067 | 0.095286833 |
| SNRPG | 0.170877928 | -0.059785886 | 0.111092042 |
| MPG | 0.116400932 | -0.073384771 | 0.043016161 |
| SLC25A39 | 0.133229234 | -0.105345691 | 0.027883544 |
| LYPLA1 | 0.111250391 | -0.086154528 | 0.025095863 |
| DMKN | 0.182854214 | -0.149250942 | 0.033603272 |
| ARHGEF12 | 0.145942381 | -0.164829956 | -0.018887575 |
| SRSF4 | 0.129282628 | -0.132801266 | -0.003518638 |
| EIF4G3 | 0.110617112 | -0.116231407 | -0.005614295 |
| TMEM11 | 0.103458721 | -0.067788452 | 0.035670269 |
| LSM12 | 0.110437229 | -0.091741066 | 0.018696163 |
| PTPMT1 | 0.125350302 | -0.096141785 | 0.029208517 |
| DCPS | 0.090593905 | -0.09116602 | -0.000572115 |
| SLC41A2 | 0.070841494 | -0.072749652 | -0.001908158 |
| COX4I1 | 0.191876693 | -0.075406241 | 0.116470451 |
| TMBIM4 | 0.186333765 | -0.229464777 | -0.043131012 |
| MYO6 | 0.13059804 | -0.115739258 | 0.014858782 |
| CAP1 | 0.140217019 | -0.108205861 | 0.032011159 |
| TMBIM6 | 0.191976923 | -0.319714708 | -0.127737785 |
| SOD1 | 0.195439514 | -0.105697782 | 0.089741732 |
| HES4 | -0.148383276 | 0.102734385 | -0.045648891 |
| C18orf32 | 0.175237427 | -0.182309191 | -0.007071764 |
| MYDGF | 0.145683938 | -0.153616834 | -0.007932895 |
| PRMT1 | 0.14762499 | -0.098786945 | 0.048838044 |
| SMIM12 | 0.113450936 | -0.088612271 | 0.024838664 |
| YES1 | 0.099718921 | -0.073716115 | 0.026002806 |
| ATP6V1C1 | 0.11422511 | -0.105377489 | 0.008847621 |
| ERLEC1 | 0.12898837 | -0.164669698 | -0.035681328 |
| TMEM147 | 0.146581683 | -0.123645346 | 0.022936337 |
| SRRM4 | -0.071457291 | 0.03077033 | -0.040686961 |
| CLCN5 | -0.10232903 | 0.040011145 | -0.062317885 |
| RILPL2 | 0.112664525 | -0.110069373 | 0.002595152 |

| | | | |
|---|---|---|---|
| ATF4 | 0.134001754 | -0.122805355 | 0.011196399 |
| SYNE2 | 0.195794881 | -0.467686753 | -0.271891873 |
| MAP1LC3A | 0.131923126 | -0.089270699 | 0.042652427 |
| CAVIN1 | 0.147795247 | -0.188130068 | -0.040334821 |
| PDCD5 | 0.143575209 | -0.066383699 | 0.07719151 |
| SNAPC1 | 0.096939108 | -0.092823248 | 0.00411586 |
| TOP1MT | 0.064848127 | -0.06029413 | 0.004553997 |
| KCMF1 | 0.109752093 | -0.085035269 | 0.024716824 |
| CDC42 | 0.18081424 | -0.119457066 | 0.061357174 |
| PIGX | 0.097650996 | -0.084332511 | 0.013318485 |
| RAB5IF | 0.139921274 | -0.137211351 | 0.002709924 |
| MRPS21 | 0.172335771 | -0.063607361 | 0.108728409 |
| CHDH | 0.081466546 | -0.054701022 | 0.026765524 |
| FIBP | 0.127313629 | -0.073383652 | 0.053929977 |
| GTF3C6 | 0.144289341 | -0.084389572 | 0.059899769 |
| TUBA4A | 0.12045273 | -0.090104009 | 0.030348721 |
| DCTN2 | 0.125452333 | -0.111890257 | 0.013562075 |
| PCMTD1 | 0.152710785 | -0.188766228 | -0.036055443 |
| IP6K2 | 0.127182228 | -0.123578727 | 0.0036035 |
| STX12 | 0.106924004 | -0.100952183 | 0.005971821 |
| DGUOK | 0.128833869 | -0.108962866 | 0.019871003 |
| DCP2 | 0.11036401 | -0.116775098 | -0.006411088 |
| CTSZ | 0.151516262 | -0.1898754 | -0.038359138 |
| SNF8 | 0.134589993 | -0.079726306 | 0.054863688 |
| PPP1CB | 0.173984374 | -0.105504772 | 0.068479602 |
| SWAP70 | 0.115320946 | -0.111539499 | 0.003781447 |
| PUSL1 | 0.066060472 | -0.054760459 | 0.011300013 |
| TRHDE | -0.098443818 | 0.047065273 | -0.051378545 |
| PHF5A | 0.09990575 | -0.072287982 | 0.027617768 |
| UNC50 | 0.148024109 | -0.152111815 | -0.004087707 |
| SOCS3 | -0.098679391 | 0.037033148 | -0.061646243 |
| SYF2 | 0.173791623 | -0.167874707 | 0.005916916 |
| SSPN | 0.065292703 | -0.06098044 | 0.004312263 |
| MRPL14 | 0.135896466 | -0.127020619 | 0.008875847 |
| DUSP1 | 0.12869006 | -0.140762594 | -0.012072534 |
| KCNMB2 | -0.087383914 | 0.084430734 | -0.002953179 |
| SDHC | 0.136063867 | -0.060043946 | 0.07601992 |
| CHST1 | 0.054325881 | -0.054325881 | -2.13163E-14 |
| YWHAZ | 0.158109166 | -0.131913396 | 0.02619577 |
| DHCR24 | 0.129315193 | -0.182462503 | -0.05314731 |
| JUND | 0.195129816 | -0.091995479 | 0.103134337 |
| TMEM14B | 0.170314146 | -0.170419596 | -0.000105449 |
| ELOC | 0.148138217 | -0.074144629 | 0.073993589 |
| ACTR2 | 0.139012059 | -0.14653272 | -0.007520661 |
| PPP2R2A | 0.110126346 | -0.112794031 | -0.002667684 |
| RAB10 | 0.128984729 | -0.123583905 | 0.005400824 |
| BID | 0.085317515 | -0.066478997 | 0.018838518 |

| | | | |
|---|---|---|---|
| CAMLG | 0.118661181 | -0.107080729 | 0.011580453 |
| IDH3A | 0.08929254 | -0.089686997 | -0.000394457 |
| HNRNPAB | 0.126636715 | -0.111487163 | 0.015149552 |
| TMEM50A | 0.166038609 | -0.254134519 | -0.08809591 |
| CSRP1 | 0.110803829 | -0.065910677 | 0.044893152 |
| PSMD6 | 0.10764833 | -0.073176362 | 0.034471968 |
| ESYT2 | 0.114477894 | -0.139099236 | -0.024621341 |
| BEX3 | 0.191962002 | -0.180748949 | 0.011213053 |
| MRPL12 | 0.117376041 | -0.040717008 | 0.076659032 |
| FAM89B | 0.108573491 | -0.099159801 | 0.00941369 |
| LCMT1 | 0.102137348 | -0.069755358 | 0.032381991 |
| CTSK | 0.098401573 | -0.104288794 | -0.005887222 |
| TPT1 | 0.163281151 | 0.10450763 | 0.267788781 |
| CFAP298 | 0.131160164 | -0.135246332 | -0.004086167 |
| SHMT1 | 0.133222846 | -0.109594584 | 0.023628261 |
| EIF4A2 | 0.184184846 | -0.177241744 | 0.006943102 |
| PFN2 | 0.120737546 | -0.099325401 | 0.021412144 |
| FKBP8 | 0.161591499 | -0.117481171 | 0.044110328 |
| SARS | 0.129853025 | -0.124587075 | 0.00526595 |
| TRAPPC6A | 0.131898448 | -0.058835551 | 0.073062896 |
| TMEM59 | 0.191686426 | -0.295996317 | -0.104309892 |
| RPL10A | 0.179182348 | -0.061838027 | 0.117344321 |
| BX284668.5 | 0.084668433 | -0.057004977 | 0.027663456 |
| PSMA3 | 0.137125255 | -0.083053044 | 0.054072211 |
| RBCK1 | 0.112181332 | -0.073075043 | 0.03910629 |
| ELF1 | 0.127439901 | -0.129461836 | -0.002021934 |
| DNAJB6 | 0.142576146 | -0.147958448 | -0.005382302 |
| UBE2S | 0.092401894 | -0.074151781 | 0.018250112 |
| ZFP36L2 | -0.13269469 | 0.080743565 | -0.051951124 |
| PRXL2C | 0.083615398 | -0.088932143 | -0.005316745 |
| SNRPD3 | 0.162105087 | -0.105247894 | 0.056857193 |
| RNASEH2C | 0.143940589 | -0.13071638 | 0.01322421 |
| ELP5 | 0.104775942 | -0.099989153 | 0.004786789 |
| NUBP2 | 0.117432483 | -0.125448623 | -0.00801614 |
| HNRNPH3 | 0.159820359 | -0.163528913 | -0.003708554 |
| JTB | 0.162236009 | -0.163821184 | -0.001585175 |
| CCDC91 | 0.096498067 | -0.087680074 | 0.008817992 |
| SLC25A24 | 0.096652027 | -0.095547516 | 0.001104511 |
| EIF2A | 0.11525982 | -0.094858763 | 0.020401056 |
| RPAIN | 0.126124079 | -0.111889987 | 0.014234092 |
| ITPA | 0.117517109 | -0.068743532 | 0.048773577 |
| TPGS1 | 0.105465243 | -0.090515602 | 0.014949641 |
| MACC1 | -0.131058842 | 0.096292338 | -0.034766505 |
| SMTNL2 | 0.092177408 | -0.093190069 | -0.00101266 |
| LMAN2 | 0.146977082 | -0.144477937 | 0.002499144 |
| PSMB7 | 0.136916984 | -0.082754588 | 0.054162397 |
| PLEKHD1 | 0.054089461 | -0.054089461 | -1.95399E-14 |

| | | | |
|---|---|---|---|
| THY1 | 0.128290637 | -0.13028122 | -0.001990583 |
| PDE6D | 0.083428189 | -0.08149019 | 0.001937999 |
| CUTA | 0.177333868 | -0.147193072 | 0.030140796 |
| MBNL2 | 0.150426704 | -0.200747009 | -0.050320305 |
| NXT1 | 0.094867678 | -0.09434772 | 0.000519958 |
| SCD5 | -0.113318403 | 0.050480513 | -0.06283789 |
| POLR1D | 0.148401747 | -0.11134756 | 0.037054188 |
| SETD3 | 0.11972805 | -0.099162607 | 0.020565443 |
| CTSO | -0.102542739 | 0.040373574 | -0.062169166 |
| RPS21 | 0.157805412 | -0.076374161 | 0.081431251 |
| LNX1 | 0.071157803 | -0.072259087 | -0.001101284 |
| INPP1 | 0.065400414 | -0.061684125 | 0.003716289 |
| TERF2IP | 0.14783795 | -0.149533094 | -0.001695144 |
| SHTN1 | 0.121662865 | -0.128472351 | -0.006809486 |
| LMAN1 | 0.176369224 | -0.278366669 | -0.101997445 |
| NDUFA8 | 0.134926627 | -0.104228966 | 0.030697661 |
| RAB11A | 0.139424557 | -0.069087465 | 0.070337091 |
| CEBPD | 0.186233136 | -0.186076456 | 0.00015668 |
| PCNP | 0.137924078 | -0.093664072 | 0.044260006 |
| NDUFB9 | 0.165787162 | -0.078007834 | 0.087779327 |
| COPB1 | 0.125858186 | -0.099859065 | 0.025999121 |
| ARL8A | 0.112321991 | -0.098779209 | 0.013542782 |
| SLC50A1 | 0.090512786 | -0.081504629 | 0.009008157 |
| C1QBP | 0.145737024 | -0.05506185 | 0.090675174 |
| KAZN | 0.148371263 | -0.176833309 | -0.028462045 |
| GNAI3 | 0.114787254 | -0.069635238 | 0.045152016 |
| TPRN | 0.084508992 | -0.094740252 | -0.01023126 |
| JAGN1 | 0.117288351 | -0.104197696 | 0.013090655 |
| ERP29 | 0.167074569 | -0.194253548 | -0.027178979 |
| NDUFA7 | 0.16475986 | -0.075854728 | 0.088905132 |
| FAM177A1 | 0.124743417 | -0.087812554 | 0.036930863 |
| ZCRB1 | 0.131356561 | -0.110218903 | 0.021137658 |
| TMEM54 | 0.11296929 | -0.09610291 | 0.01686638 |
| TAGLN2 | 0.130989815 | -0.112606335 | 0.01838348 |
| PNOC | 0.083832922 | -0.084855956 | -0.001023034 |
| MBD2 | 0.1188808 | -0.086679723 | 0.032201077 |
| SMIM3 | 0.084623663 | -0.086621569 | -0.001997906 |
| SMIM26 | 0.158027962 | -0.095083635 | 0.062944327 |
| DZIP1 | 0.069660715 | -0.058483239 | 0.011177476 |
| ESD | 0.150320279 | -0.052083542 | 0.098236737 |
| WHAMM | 0.099748317 | -0.105264281 | -0.005515964 |
| SLC27A2 | -0.082379347 | 0.080008025 | -0.002371323 |
| OCIAD1 | 0.167388537 | -0.21670526 | -0.049316724 |
| SDHA | 0.119594955 | -0.077556951 | 0.042038004 |
| C1D | 0.137541459 | -0.090404583 | 0.047136876 |
| AASS | -0.081140841 | 0.033530317 | -0.047610524 |
| LIPH | 0.128888561 | -0.199356457 | -0.070467895 |

| | | | |
|---|---|---|---|
| ST3GAL6 | 0.101666065 | -0.132745957 | -0.031079892 |
| ARF1 | 0.170315034 | -0.161475958 | 0.008839075 |
| PPP4R2 | 0.11301124 | -0.108900206 | 0.004111034 |
| HK1 | 0.095238798 | -0.098069872 | -0.002831074 |
| DHPS | 0.109820449 | -0.128435437 | -0.018614987 |
| EMC8 | 0.080605123 | -0.051553816 | 0.029051307 |
| CUEDC2 | 0.120643105 | -0.093064228 | 0.027578877 |
| CLDN7 | 0.155705942 | -0.186028189 | -0.030322247 |
| CD99 | 0.083013196 | -0.081799156 | 0.00121404 |
| GORASP2 | 0.08745601 | -0.106186896 | -0.018730886 |
| SMAGP | 0.074941386 | -0.080747824 | -0.005806438 |
| HDDC2 | 0.141385929 | -0.114731301 | 0.026654628 |
| CDK4 | 0.113997104 | -0.068069372 | 0.045927732 |
| POLR3K | 0.090109144 | -0.069194173 | 0.020914972 |
| TWF1 | 0.099867813 | -0.085608547 | 0.014259266 |
| POLR2E | 0.129053216 | -0.0843848 | 0.044668416 |
| SPP1 | -0.171373417 | 0.119025434 | -0.052347983 |
| MOB2 | 0.091191393 | -0.077326819 | 0.013864573 |
| KRR1 | 0.126499913 | -0.126073194 | 0.00042672 |
| POLR2H | 0.127789237 | -0.084609098 | 0.043180139 |
| TOMM5 | 0.15133504 | -0.080000569 | 0.071334471 |
| RNF212 | 0.067201075 | -0.059310802 | 0.007890273 |
| OGFRL1 | 0.097549729 | -0.094979514 | 0.002570216 |
| IER3IP1 | 0.137395805 | -0.112019743 | 0.025376062 |
| TIMMDC1 | 0.122998292 | -0.104383211 | 0.018615081 |
| GTF2I | 0.171681467 | -0.224621671 | -0.052940204 |
| SS18L2 | 0.107718861 | -0.095375031 | 0.01234383 |
| SMIM19 | 0.133406585 | -0.130691346 | 0.002715239 |
| CFAP36 | 0.118609786 | -0.09416093 | 0.024448856 |
| PGLS | 0.125773351 | -0.057806498 | 0.067966854 |
| GLOD4 | 0.120966798 | -0.088127814 | 0.032838984 |
| MPV17 | 0.09901638 | -0.070740122 | 0.028276259 |
| SLC43A2 | 0.1192913 | -0.180461919 | -0.061170619 |
| P4HA1 | 0.091631548 | -0.095326023 | -0.003694476 |
| MXD4 | 0.126214861 | -0.12340488 | 0.002809981 |
| TACC3 | 0.05482251 | -0.057717104 | -0.002894594 |
| SLC14A2 | 0.088820321 | -0.088820321 | -1.95399E-14 |
| CORO1B | 0.108175614 | -0.092561493 | 0.015614121 |
| BCL7B | 0.09865125 | -0.073350391 | 0.025300859 |
| TBCA | 0.169521671 | -0.086825293 | 0.082696378 |
| PCMT1 | 0.128589749 | -0.153198839 | -0.024609091 |
| DRAM2 | 0.093986284 | -0.082929259 | 0.011057024 |
| AMPD2 | 0.075548068 | -0.071197767 | 0.004350301 |
| PIP4P2 | 0.077879384 | -0.07068478 | 0.007194603 |
| SNHG8 | 0.18130861 | -0.09211075 | 0.089197861 |
| AKTIP | 0.104736642 | -0.113713598 | -0.008976956 |
| PAICS | 0.094701839 | -0.072307624 | 0.022394216 |

| | | | |
|---|---|---|---|
| ZNF32 | 0.105402366 | -0.069343167 | 0.036059199 |
| FAM114A1 | 0.078297389 | -0.08669184 | -0.008394451 |
| DPYSL2 | 0.133306306 | -0.13161986 | 0.001686446 |
| PDAP1 | 0.137532224 | -0.145204602 | -0.007672378 |
| RNF187 | 0.126958756 | -0.084321334 | 0.042637422 |
| SELENOW | 0.155630847 | -0.104619379 | 0.051011468 |
| PMF1 | 0.128707954 | -0.110843473 | 0.017864482 |
| KIF5B | 0.15438207 | -0.188408658 | -0.034026589 |
| AP1M2 | 0.110448331 | -0.050895474 | 0.059552857 |
| IDH3G | 0.112921051 | -0.073682349 | 0.039238702 |
| IDI1 | 0.100394956 | -0.089466457 | 0.010928499 |
| SUCLA2 | 0.119482613 | -0.104631369 | 0.014851244 |
| FAM174C | 0.114191341 | -0.09136551 | 0.022825831 |
| MRPL4 | 0.115765584 | -0.078352179 | 0.037413404 |
| TIMM13 | 0.148833854 | -0.090398915 | 0.058434939 |
| XRCC6 | 0.138207495 | -0.102281893 | 0.035925602 |
| ISOC2 | 0.104675339 | -0.048976746 | 0.055698593 |
| EBNA1BP2 | 0.102836777 | -0.070443557 | 0.032393221 |
| HAUS1 | 0.070578418 | -0.045191359 | 0.02538706 |
| APMAP | 0.118939064 | -0.137557174 | -0.01861811 |
| PLPP1 | 0.08532733 | -0.083392633 | 0.001934697 |
| KCNJ16 | -0.132944719 | 0.090947926 | -0.041996793 |
| ZFP91 | 0.11250503 | -0.118978907 | -0.006473877 |
| NUDC | 0.150100529 | -0.117259173 | 0.032841356 |
| AFMID | 0.10610202 | -0.133710882 | -0.027608862 |
| ST6GALNAC6 | 0.072939449 | -0.0690324 | 0.003907049 |
| ANAPC13 | 0.138546967 | -0.081300521 | 0.057246446 |
| GTF3A | 0.123788246 | -0.099750227 | 0.024038019 |
| GLRX5 | 0.154407665 | -0.113071562 | 0.041336103 |
| CTSB | 0.123183799 | -0.107722029 | 0.01546177 |
| NAT14 | 0.113155261 | -0.138935547 | -0.025780285 |
| CRYM | -0.099284097 | 0.091218485 | -0.008065612 |
| UBXN4 | 0.160199933 | -0.174420771 | -0.014220838 |
| CXXC5 | 0.161005048 | -0.185327759 | -0.024322712 |
| ADD1 | 0.132861803 | -0.140259627 | -0.007397824 |
| CCDC25 | 0.106582153 | -0.093731162 | 0.012850991 |
| TXN2 | 0.134663587 | -0.095709983 | 0.038953604 |
| SHANK2 | -0.094773211 | 0.059228609 | -0.035544601 |
| NDUFA5 | 0.162615131 | -0.104821633 | 0.057793498 |
| TGM2 | 0.052313738 | -0.046112054 | 0.006201684 |
| DEGS1 | 0.103924618 | -0.107358746 | -0.003434128 |
| SLC39A7 | 0.107477721 | -0.119882255 | -0.012404534 |
| LINC-PINT | 0.1069682 | -0.124121828 | -0.017153628 |
| HS3ST1 | 0.052939142 | -0.05322948 | -0.000290338 |
| PLGRKT | 0.104751687 | -0.094220572 | 0.010531115 |
| MRPL38 | 0.112990027 | -0.071967124 | 0.041022902 |
| ATAD3C | 0.073426339 | -0.053087228 | 0.020339111 |

| | | | |
|---|---|---|---|
| ATP1A1 | 0.169190118 | -0.229881142 | -0.060691024 |
| PPP1R11 | 0.10406892 | -0.088524768 | 0.015544152 |
| OTUD6B-AS1 | 0.131786085 | -0.133368675 | -0.001582591 |
| PSMD2 | 0.110204708 | -0.08170608 | 0.028498629 |
| NABP1 | -0.055791386 | 0.020482988 | -0.035308399 |
| MARCH5 | 0.097676922 | -0.097458516 | 0.000218406 |
| MTOR | 0.074216388 | -0.0777469 | -0.003530513 |
| LAD1 | 0.088650539 | -0.066388505 | 0.022262034 |
| FKBP1B | 0.065633917 | -0.069938741 | -0.004304824 |
| SLC38A11 | 0.058694792 | -0.053741451 | 0.004953341 |
| HSPD1 | 0.15128284 | -0.062219282 | 0.089063558 |
| RHOQ | 0.084584887 | -0.067108454 | 0.017476433 |
| MRPS7 | 0.117641725 | -0.079745132 | 0.037896592 |
| PPP1CC | 0.109687098 | -0.071492417 | 0.038194681 |
| THNSL2 | -0.06841533 | 0.04770094 | -0.02071439 |
| MT-ND5 | -0.135474337 | 0.112278335 | -0.023196002 |
| SERBP1 | 0.165854399 | -0.124767295 | 0.041087104 |
| SNRPA1 | 0.09820439 | -0.085214135 | 0.012990255 |
| CNPY2 | 0.148843738 | -0.163351263 | -0.014507525 |
| REX1BD | 0.126854343 | -0.075817635 | 0.051036708 |
| DNAJC4 | 0.10097247 | -0.087292219 | 0.013680251 |
| C9orf78 | 0.12154604 | -0.127121927 | -0.005575886 |
| LMO7-AS1 | 0.070161585 | -0.071472767 | -0.001311182 |
| HSPA1A | 0.167551366 | -0.225818632 | -0.058267266 |
| BABAM1 | 0.10028016 | -0.099615292 | 0.000664868 |
| UBE2G1 | 0.080062569 | -0.066365591 | 0.013696977 |
| MSANTD3 | 0.079800418 | -0.080518647 | -0.000718228 |
| ARL3 | 0.14569748 | -0.121230755 | 0.024466725 |
| RASSF8-AS1 | 0.08188576 | -0.050501346 | 0.031384414 |
| WDR83 | 0.083282361 | -0.068373786 | 0.014908575 |
| KRTCAP2 | 0.160504604 | -0.137503716 | 0.023000887 |
| TSEN34 | 0.122329026 | -0.111747566 | 0.01058146 |
| NAA50 | 0.110064062 | -0.098148763 | 0.011915299 |
| RAB32 | 0.085051733 | -0.044529171 | 0.040522562 |
| QKI | 0.139910853 | -0.156565633 | -0.01665478 |
| ECE1 | 0.07828409 | -0.055345894 | 0.022938196 |
| GALNT3 | 0.11356734 | -0.137446524 | -0.023879184 |
| HLA-DMA | 0.071398461 | -0.04610298 | 0.025295481 |
| PSMD4 | 0.123632441 | -0.071021719 | 0.052610721 |
| MZT1 | 0.090118944 | -0.078558387 | 0.011560557 |
| LIMCH1 | 0.103384332 | -0.098901498 | 0.004482834 |
| PAXX | 0.120526543 | -0.076960303 | 0.043566241 |
| LAPTM4A | 0.179421167 | -0.292801481 | -0.113380314 |
| LACTB | 0.075004478 | -0.058756236 | 0.016248242 |
| PSMD13 | 0.113539562 | -0.073516169 | 0.040023393 |
| LIMS1 | 0.091511226 | -0.078316339 | 0.013194888 |
| FNIP2 | -0.093027238 | 0.064716648 | -0.02831059 |

| | | | |
|---|---|---|---|
| RP1 | 0.051389508 | -0.040679324 | 0.010710184 |
| KCNMA1 | -0.079238724 | 0.086618856 | 0.007380133 |
| SATB2 | -0.081710914 | 0.039398299 | -0.042312614 |
| GLO1 | 0.120066391 | -0.096896816 | 0.023169575 |
| RBM17 | 0.126794545 | -0.119237935 | 0.00755661 |
| PRR13 | 0.148690883 | -0.066216079 | 0.082474805 |
| UMOD | -0.129570977 | 0.102921452 | -0.026649525 |
| PDCD2 | 0.104049689 | -0.094907545 | 0.009142144 |
| GOLT1A | 0.054094587 | -0.046832389 | 0.007262198 |
| TAX1BP3 | 0.122516197 | -0.101171695 | 0.021344503 |
| TSC22D1 | -0.171368117 | 0.135949151 | -0.035418966 |
| TDGF1 | 0.055850597 | -0.064915528 | -0.009064931 |
| HEBP2 | 0.1283307 | -0.082439065 | 0.045891635 |
| MT-ND4L | -0.127822224 | 0.046823853 | -0.08099837 |
| SHISA4 | 0.069412961 | -0.068648981 | 0.00076398 |
| LFNG | -0.061158245 | 0.033941295 | -0.027216951 |
| PLEKHA5 | 0.092717572 | -0.100673813 | -0.007956242 |
| EIF4G2 | 0.156261705 | -0.149273871 | 0.006987834 |
| HSDL2 | 0.085353473 | -0.069224142 | 0.016129331 |
| TUSC3 | 0.120951155 | -0.126086891 | -0.005135736 |
| SLC2A6 | 0.048001976 | -0.044915146 | 0.00308683 |
| TRIM47 | 0.067133836 | -0.045938276 | 0.021195559 |
| MRPL17 | 0.111631532 | -0.090322985 | 0.021308547 |
| URM1 | 0.09256515 | -0.055527879 | 0.037037271 |
| NOVA1 | 0.07708752 | -0.057196001 | 0.019891519 |
| DMGDH | 0.064537554 | -0.057733537 | 0.006804017 |
| ENOPH1 | 0.083618661 | -0.052919481 | 0.03069918 |
| FBXO21 | 0.109804505 | -0.082462175 | 0.02734233 |
| BOLA3 | 0.108132369 | -0.061308114 | 0.046824256 |
| ZNF433-AS1 | 0.062860495 | -0.056705209 | 0.006155287 |
| TPM3 | 0.136592124 | -0.111365611 | 0.025226513 |
| SEL1L3 | -0.091934888 | 0.07051074 | -0.021424149 |
| MANF | 0.104409068 | -0.101814206 | 0.002594862 |
| SEPHS2 | 0.130978598 | -0.109243626 | 0.021734972 |
| ADH5 | 0.141761806 | -0.066885988 | 0.074875818 |
| WDSUB1 | 0.07792593 | -0.069544339 | 0.008381591 |
| PLS3 | 0.068472238 | -0.06334937 | 0.005122869 |
| HMGA1 | 0.083159366 | -0.083102356 | 5.701E-05 |
| DLGAP1-AS1 | 0.065189271 | -0.056980555 | 0.008208717 |
| ERI3 | 0.108141634 | -0.075002374 | 0.033139261 |
| DDX21 | 0.102904237 | -0.084450866 | 0.018453372 |
| FLRT3 | -0.079977916 | 0.085909367 | 0.00593145 |
| ACADM | 0.123242316 | -0.117416949 | 0.005825367 |
| FNDC3B | 0.11495672 | -0.10885642 | 0.0061003 |
| ARPC5 | 0.120647681 | -0.07076097 | 0.049886711 |
| HAT1 | 0.08800746 | -0.069614694 | 0.018392767 |
| UXS1 | 0.146254004 | -0.208686579 | -0.062432575 |

| | | | |
|---|---|---|---|
| PSMA4 | 0.123618333 | -0.094537385 | 0.029080948 |
| MRPL57 | 0.148695036 | -0.097292531 | 0.051402505 |
| GTF2A2 | 0.146920411 | -0.09218526 | 0.054735151 |
| SEPTIN7 | 0.153080524 | -0.136936392 | 0.016144132 |
| BUB3 | 0.091593796 | -0.069727417 | 0.021866379 |
| TXNDC9 | 0.100002363 | -0.046484954 | 0.053517409 |
| COPS5 | 0.109943736 | -0.085026689 | 0.024917047 |
| LINC02593 | -0.061870115 | 0.036981389 | -0.024888726 |
| SLC52A2 | 0.069923304 | -0.068418809 | 0.001504495 |
| HS6ST1 | -0.107960774 | 0.063330117 | -0.044630658 |
| WNT7B | 0.104905207 | -0.148284248 | -0.043379042 |
| FKBP4 | 0.132746636 | -0.118536676 | 0.01420996 |
| LURAP1 | 0.056516874 | -0.051775082 | 0.004741792 |
| TANK | 0.091521516 | -0.080233562 | 0.011287953 |
| PSMA6 | 0.13121315 | -0.077331945 | 0.053881205 |
| DGCR6L | 0.119187566 | -0.067774872 | 0.051412694 |
| CRYZ | 0.119596878 | -0.134628735 | -0.015031857 |
| LAMB2 | 0.114313084 | -0.12897974 | -0.014666656 |
| SEM1 | 0.154684389 | -0.05718294 | 0.097501449 |
| ERG28 | 0.108410046 | -0.104998915 | 0.003411131 |
| RAMAC | 0.074024816 | -0.056376588 | 0.017648228 |
| TAPT1 | 0.087151495 | -0.100643061 | -0.013491565 |
| TSSC4 | 0.097320736 | -0.084956059 | 0.012364678 |
| SRP19 | 0.10569158 | -0.11337565 | -0.00768407 |
| SLC35B3 | 0.078255245 | -0.076116283 | 0.002138962 |
| RALBP1 | 0.127741197 | -0.109729666 | 0.018011531 |
| PSMC2 | 0.099591439 | -0.113076408 | -0.013484969 |
| DNAJC7 | 0.121409539 | -0.128976426 | -0.007566887 |
| CREM | 0.080180133 | -0.052773382 | 0.027406751 |
| HEXA | 0.10450234 | -0.099444181 | 0.005058159 |
| ACAT2 | 0.071516383 | -0.053933356 | 0.017583027 |
| TSR2 | 0.104885166 | -0.098090197 | 0.006794969 |
| PPP4C | 0.124586033 | -0.091076012 | 0.033510021 |
| SSR2 | 0.148905014 | -0.099280633 | 0.049624381 |
| FAM3A | 0.095612409 | -0.077894762 | 0.017717647 |
| LINC00504 | 0.043235482 | -0.041361877 | 0.001873605 |
| TCF4 | 0.055885939 | -0.05646344 | -0.000577501 |
| APLP2 | 0.170076129 | -0.356510034 | -0.186433905 |
| GSR | 0.082716643 | -0.069376451 | 0.013340191 |
| IL1R1 | -0.083805833 | 0.050208952 | -0.033596881 |
| PTPN11 | 0.103915555 | -0.082609502 | 0.021306053 |
| ZBTB16 | -0.107853633 | 0.082080811 | -0.025772822 |
| COASY | 0.087423492 | -0.094681352 | -0.007257861 |
| HEBP1 | 0.137648663 | -0.114770672 | 0.022877991 |
| FREM2 | -0.0866254 | 0.03965056 | -0.046974839 |
| ITGB6 | -0.06789684 | 0.027987459 | -0.039909381 |
| ACBD6 | 0.087525048 | -0.086964296 | 0.000560752 |

| | | | |
|---|---|---|---|
| RWDD1 | 0.128722918 | -0.118499608 | 0.01022331 |
| RAB12 | 0.084546078 | -0.083885614 | 0.000660464 |
| HES6 | 0.0478103 | -0.047670128 | 0.000140172 |
| FUCA1 | 0.098454474 | -0.123267471 | -0.024812996 |
| JUP | 0.133498763 | -0.146105115 | -0.012606352 |
| RANBP1 | 0.134979927 | -0.0614119 | 0.073568027 |
| RNF144A | -0.058207935 | 0.027317978 | -0.030889957 |
| BOD1 | 0.094539609 | -0.080874197 | 0.013665412 |
| PRNP | 0.100363479 | -0.096450357 | 0.003913122 |
| AK6 | 0.103803582 | -0.051847349 | 0.051956234 |
| PCDH1 | 0.047539904 | -0.04374911 | 0.003790794 |
| GRHPR | 0.113184951 | -0.066765331 | 0.04641962 |
| ALS2CL | 0.061487107 | -0.062463904 | -0.000976797 |
| SCO2 | 0.065448614 | -0.035086168 | 0.030362446 |
| RBM8A | 0.140712279 | -0.110124508 | 0.030587772 |
| STK26 | 0.049705295 | -0.050838933 | -0.001133638 |
| FDPS | 0.109664622 | -0.092486392 | 0.01717823 |
| SURF2 | 0.101312658 | -0.063236012 | 0.038076647 |
| CALR | 0.160317153 | -0.212447939 | -0.052130785 |
| TPCN1 | 0.077658144 | -0.090506519 | -0.012848375 |
| PSMB8 | 0.114298071 | -0.067042905 | 0.047255167 |
| ESRRB | 0.042384571 | -0.041588163 | 0.000796408 |
| MYH8 | 0.05925042 | -0.043269647 | 0.015980773 |
| PLCB1 | 0.083802704 | -0.076021992 | 0.007780711 |
| MSMO1 | 0.097156286 | -0.116460137 | -0.019303851 |
| NT5C3B | 0.106496337 | -0.09308659 | 0.013409747 |
| PAFAH1B1 | 0.130685848 | -0.152513945 | -0.021828098 |
| AKR1C2 | 0.102823709 | -0.079090598 | 0.023733111 |
| CCNQ | 0.07002639 | -0.072871323 | -0.002844933 |
| NUDCD2 | 0.117794942 | -0.056503234 | 0.061291708 |
| DNAJC6 | 0.056893459 | -0.057604931 | -0.000711472 |
| SF3B6 | 0.156100766 | -0.064958538 | 0.091142227 |
| MFSD6 | 0.05455969 | -0.053411084 | 0.001148607 |
| C12orf57 | 0.139519472 | -0.068275564 | 0.071243908 |
| TRIP6 | 0.117059569 | -0.078767356 | 0.038292212 |
| DNAJC21 | 0.113838566 | -0.133193818 | -0.019355252 |
| NUDT22 | 0.096867819 | -0.064753471 | 0.032114348 |
| NHS | -0.061617319 | 0.02912541 | -0.032491909 |
| SHARPIN | 0.098289505 | -0.073410631 | 0.024878873 |
| AKAP8L | 0.105159511 | -0.101111959 | 0.004047552 |
| JADE1 | -0.104137284 | 0.054949432 | -0.049187851 |
| PFDN4 | 0.094460631 | -0.056705689 | 0.037754942 |
| RAB7A | 0.118552876 | -0.10111524 | 0.017437635 |
| HADHB | 0.124812649 | -0.055338654 | 0.069473995 |
| VASP | 0.076215713 | -0.071449328 | 0.004766385 |
| EIF2B5 | 0.086023013 | -0.085505848 | 0.000517165 |
| PELO | 0.083602229 | -0.053406942 | 0.030195287 |

| | | | |
|---|---|---|---|
| SDC1 | 0.077595346 | -0.069776963 | 0.007818383 |
| RITA1 | 0.073498564 | -0.063374022 | 0.010124542 |
| IGFBP3 | -0.211441417 | -0.115963621 | -0.327405039 |
| CBWD1 | 0.088449526 | -0.072110387 | 0.016339139 |
| SLC35B1 | 0.089875939 | -0.068003674 | 0.021872266 |
| NCBP2AS2 | 0.110144573 | -0.071444596 | 0.038699977 |
| KCNQ1 | 0.057966277 | -0.062423934 | -0.004457658 |
| WTAP | 0.124550552 | -0.097538912 | 0.027011641 |
| ARL1 | 0.130183825 | -0.12985903 | 0.000324795 |
| CDC26 | 0.108512615 | -0.087278975 | 0.021233641 |
| CYS1 | 0.157145317 | -0.190014214 | -0.032868897 |
| MEAF6 | 0.125046985 | -0.052852168 | 0.072194817 |
| PITHD1 | 0.096076131 | -0.089573357 | 0.006502774 |
| TUSC2 | 0.11028503 | -0.071970655 | 0.038314375 |
| NMRK1 | 0.081194511 | -0.07334509 | 0.007849421 |
| MED4 | 0.114214032 | -0.106452605 | 0.007761426 |
| POLD2 | 0.117544197 | -0.070697611 | 0.046846586 |
| ELOF1 | 0.082825375 | -0.044210087 | 0.038615289 |
| PRPF31 | 0.103486125 | -0.093585746 | 0.009900379 |
| GLTP | 0.11709325 | -0.143053125 | -0.025959875 |
| CDK11A | 0.053441741 | -0.051283668 | 0.002158073 |
| ABCB1 | -0.093560914 | 0.040651408 | -0.052909506 |
| ARSJ | 0.055220824 | -0.042747885 | 0.012472939 |
| SPNS2 | 0.124935526 | -0.135290938 | -0.010355412 |
| GGH | 0.057752722 | -0.045618626 | 0.012134096 |
| DNAJC19 | 0.131022682 | -0.095033203 | 0.035989478 |
| MED21 | 0.107570264 | -0.100921963 | 0.006648301 |
| PDXDC1 | 0.100394925 | -0.074213826 | 0.026181099 |
| ARFIP2 | 0.089441912 | -0.066214271 | 0.023227641 |
| ATP6AP2 | 0.147761486 | -0.232326375 | -0.084564889 |
| SSTR2 | 0.044264795 | -0.045449269 | -0.001184474 |
| ETFB | 0.138651463 | -0.07270373 | 0.065947733 |
| EEF2 | 0.158640023 | -0.090999174 | 0.067640849 |
| TSPAN7 | 0.05388603 | -0.054127682 | -0.000241652 |
| PDE4D | 0.073973358 | -0.067830702 | 0.006142656 |
| ASB13 | 0.073278787 | -0.068617892 | 0.004660895 |
| SDHB | 0.094101788 | -0.066727123 | 0.027374665 |
| ATRAID | 0.15609911 | -0.152807351 | 0.003291759 |
| FAM32A | 0.101762777 | -0.050697385 | 0.051065391 |
| HSPB2 | 0.058378511 | -0.044956355 | 0.013422156 |
| DPM3 | 0.14201147 | -0.149047848 | -0.007036377 |
| ZBTB8OS | 0.084482715 | -0.067787264 | 0.016695451 |
| PTGES3 | 0.156886489 | -0.0849508 | 0.071935689 |
| AFG3L2 | 0.105710845 | -0.113804451 | -0.008093605 |
| GLRX3 | 0.09769562 | -0.076256254 | 0.021439365 |
| ANP32B | 0.131850656 | -0.104306635 | 0.027544021 |
| KRTCAP3 | 0.084374376 | -0.087829138 | -0.003454762 |

| | | | |
|---|---|---|---|
| TBCB | 0.124617835 | -0.097135242 | 0.027482593 |
| CRMP1 | 0.039416552 | -0.039416552 | -1.95399E-14 |
| BRMS1 | 0.080636342 | -0.063551287 | 0.017085055 |
| NUDT14 | 0.10467274 | -0.112297962 | -0.007625222 |
| LYPD6B | 0.047400269 | -0.050098615 | -0.002698346 |
| CLDN8 | -0.104318508 | 0.097757615 | -0.006560893 |
| FAAP20 | 0.118427019 | -0.118936813 | -0.000509795 |
| CHCHD3 | 0.099443546 | -0.102401904 | -0.002958358 |
| CTDSPL | 0.125437645 | -0.155541845 | -0.0301042 |
| B4GALNT2 | 0.061193779 | -0.049825815 | 0.011367964 |
| HNRNPA3 | 0.152787888 | -0.166238248 | -0.01345036 |
| HNRNPDL | 0.153112907 | -0.173476302 | -0.020363395 |
| AAMP | 0.114304041 | -0.112927679 | 0.001376362 |
| PSENEN | 0.119373382 | -0.117054635 | 0.002318747 |
| APBB2 | 0.065614533 | -0.076045919 | -0.010431387 |
| DAD1 | 0.165296383 | -0.201643944 | -0.036347561 |
| HSD11B1L | 0.071393931 | -0.076826869 | -0.005432939 |
| SDF2 | 0.119178255 | -0.136481595 | -0.01730334 |
| GON7 | 0.090525693 | -0.082252726 | 0.008272967 |
| ST14 | 0.101073748 | -0.128838464 | -0.027764716 |
| PLIN3 | 0.093390067 | -0.083716796 | 0.009673271 |
| HNRNPR | 0.132950421 | -0.17903754 | -0.046087119 |
| RAB3B | -0.058634416 | 0.038588959 | -0.020045458 |
| MMP24OS | 0.13276782 | -0.098251991 | 0.034515829 |
| BLOC1S2 | 0.089849039 | -0.055344096 | 0.034504943 |
| RSL1D1 | 0.1391928 | -0.12156553 | 0.01762727 |
| HPRT1 | 0.091332266 | -0.068621629 | 0.022710636 |
| CLPP | 0.105524226 | -0.044176837 | 0.061347389 |
| AC148477.2 | 0.041621631 | -0.041293609 | 0.000328022 |
| NPEPL1 | 0.088784643 | -0.085499052 | 0.003285591 |
| CD302 | 0.045905775 | -0.043599685 | 0.00230609 |
| ROMO1 | 0.145506587 | -0.058509297 | 0.086997289 |
| STAM2 | 0.085872849 | -0.091246729 | -0.005373881 |
| NPEPPS | 0.104027247 | -0.098946728 | 0.005080519 |
| MTURN | 0.13378689 | -0.164038229 | -0.030251339 |
| HACL1 | 0.062673595 | -0.048145912 | 0.014527683 |
| SYAP1 | 0.099196114 | -0.084018616 | 0.015177497 |
| RBFA | 0.100545619 | -0.116893794 | -0.016348175 |
| MGAT4B | 0.109662438 | -0.099623557 | 0.010038881 |
| PHLDA3 | 0.072577508 | -0.07610869 | -0.003531182 |
| XBP1 | 0.119139488 | -0.107811288 | 0.0113282 |
| ETFA | 0.100160391 | -0.046953858 | 0.053206533 |
| PAIP2 | 0.1325484 | -0.075282477 | 0.057265923 |
| PINK1 | 0.089296424 | -0.054389273 | 0.034907151 |
| MAST4 | 0.073779856 | -0.070948453 | 0.002831403 |
| EPB41L5 | 0.08745674 | -0.092463754 | -0.005007014 |
| TOMM34 | 0.06731293 | -0.058032495 | 0.009280434 |

| | | | |
|---|---|---|---|
| MUC20 | 0.072571042 | -0.085924309 | -0.013353267 |
| PSMA1 | 0.121852613 | -0.082723252 | 0.039129361 |
| PSMD12 | 0.105400088 | -0.115576002 | -0.010175913 |
| EIF4A3 | 0.108845398 | -0.09833546 | 0.010509938 |
| MRGBP | 0.064189635 | -0.061699131 | 0.002490504 |
| THAP11 | 0.073289964 | -0.064327659 | 0.008962304 |
| DESI1 | 0.072403327 | -0.066359423 | 0.006043904 |
| ZNF511 | 0.10218994 | -0.073798034 | 0.028391906 |
| GMPR2 | 0.103887063 | -0.10353801 | 0.000349053 |
| TMX2 | 0.096214258 | -0.117873817 | -0.021659559 |
| APRT | 0.139166875 | -0.068187681 | 0.070979194 |
| PPIL1 | 0.068231776 | -0.063956179 | 0.004275597 |
| ASPSCR1 | 0.074741542 | -0.069644049 | 0.005097493 |
| SPTSSB | 0.079704809 | -0.059772344 | 0.019932465 |
| SAT2 | 0.132244409 | -0.113999132 | 0.018245276 |
| SYS1 | 0.095836889 | -0.101630974 | -0.005794085 |
| GRIN3B | 0.036049783 | -0.036049783 | -1.95399E-14 |
| ZNF44 | 0.097998782 | -0.115736775 | -0.017737992 |
| WDR46 | 0.070194878 | -0.066041313 | 0.004153566 |
| SKAP2 | 0.130154454 | -0.132315331 | -0.002160877 |
| ARFGEF1 | 0.094777776 | -0.10163697 | -0.006859194 |
| DERA | 0.091294833 | -0.067663148 | 0.023631685 |
| KLHL5 | 0.065085971 | -0.061169528 | 0.003916443 |
| VPS13C | 0.140302862 | -0.278893975 | -0.138591112 |
| NUAK1 | 0.06261389 | -0.054509474 | 0.008104416 |
| SPCS1 | 0.153664649 | -0.200294604 | -0.046629955 |
| SLC7A8 | 0.055598667 | -0.046548961 | 0.009049707 |
| EPN1 | 0.122133512 | -0.118475276 | 0.003658236 |
| RASD1 | -0.092056169 | 0.152163616 | 0.060107447 |
| LSM10 | 0.090209661 | -0.076954549 | 0.013255112 |
| AMD1 | 0.116261479 | -0.12863542 | -0.012373941 |
| SH3GLB1 | 0.116694918 | -0.115804212 | 0.000890706 |
| EIF4H | 0.119657136 | -0.111942373 | 0.007714764 |
| MKLN1-AS | 0.051847621 | -0.050364957 | 0.001482665 |
| ENC1 | -0.062874238 | 0.046427613 | -0.016446625 |
| PPME1 | 0.087216522 | -0.093075703 | -0.005859181 |
| SGPL1 | 0.08285701 | -0.083046794 | -0.000189784 |
| PQBP1 | 0.078442661 | -0.073193516 | 0.005249145 |
| RBM42 | 0.098067147 | -0.062503643 | 0.035563504 |
| TMEM47 | -0.066319649 | 0.049893112 | -0.016426537 |
| SIVA1 | 0.134765324 | -0.070107555 | 0.064657769 |
| DENR | 0.103593077 | -0.116471764 | -0.012878686 |
| GTF2B | 0.086135832 | -0.067731129 | 0.018404703 |
| PTPN18 | 0.081127748 | -0.079386039 | 0.001741708 |
| ME1 | 0.04826216 | -0.044508696 | 0.003753463 |
| PDGFB | 0.105407722 | -0.140706225 | -0.035298503 |
| AP4B1 | 0.046905654 | -0.053310482 | -0.006404829 |

| | | | |
|---|---|---|---|
| RUSC1 | 0.065731286 | -0.065400078 | 0.000331207 |
| TMEM205 | 0.146264433 | -0.167621084 | -0.021356651 |
| CDK2AP2 | 0.107263285 | -0.089692175 | 0.01757111 |
| NDUFA9 | 0.094761053 | -0.048630006 | 0.046131046 |
| HNRNPK | 0.156045984 | -0.128425775 | 0.027620209 |
| IDH2 | 0.09635146 | -0.052257259 | 0.0440942 |
| A4GALT | -0.066872429 | 0.058937255 | -0.007935174 |
| PBX1 | -0.088367801 | 0.075026398 | -0.013341403 |
| ZFAND2B | 0.085552601 | -0.057832501 | 0.0277201 |
| SSC4D | 0.048983361 | -0.05600567 | -0.007022309 |
| BCAR3 | 0.066794591 | -0.069855023 | -0.003060431 |
| EIF4E3 | 0.057157495 | -0.047422521 | 0.009734974 |
| LINC01503 | 0.0470537 | -0.048526771 | -0.001473071 |
| SEC11C | 0.120110894 | -0.105922145 | 0.014188749 |
| DDX24 | 0.13808616 | -0.193238689 | -0.055152528 |
| TCEAL3 | 0.107368757 | -0.126488885 | -0.019120128 |
| C1QL4 | -0.058245946 | 0.051507819 | -0.006738127 |
| TRAPPC10 | 0.068809324 | -0.07111407 | -0.002304746 |
| PEX3 | 0.075607488 | -0.0668 | 0.008807488 |
| LMBRD1 | 0.127158586 | -0.116494762 | 0.010663825 |
| EIF4E | 0.08772724 | -0.060875803 | 0.026851437 |
| FRG1 | 0.093978741 | -0.089890407 | 0.004088335 |
| HNRNPA0 | 0.118913361 | -0.107623072 | 0.011290289 |
| SELENBP1 | 0.057132272 | -0.03691976 | 0.020212511 |
| STK32C | 0.055548679 | -0.05606842 | -0.000519741 |
| EPHA7 | -0.08947119 | 0.060672371 | -0.028798819 |
| MSRA | 0.065739077 | -0.041988551 | 0.023750526 |
| AC004148.1 | 0.046596561 | -0.045579532 | 0.001017029 |
| RGS19 | 0.03952472 | -0.036707714 | 0.002817006 |
| ADGRG6 | -0.060692224 | 0.033162544 | -0.02752968 |
| MVB12A | 0.091188225 | -0.045717602 | 0.045470623 |
| UFD1 | 0.093186911 | -0.086622579 | 0.006564332 |
| ITGB1 | 0.151170528 | -0.270832705 | -0.119662177 |
| SLC2A13 | 0.099496511 | -0.133993186 | -0.034496676 |
| GNAQ | -0.093493056 | 0.046803348 | -0.046689708 |
| PUF60 | 0.103329156 | -0.064397032 | 0.038932124 |
| PSIP1 | 0.102811491 | -0.114007873 | -0.011196382 |
| TRPM7 | 0.095196725 | -0.108674057 | -0.013477331 |
| TSR3 | 0.103226666 | -0.049343285 | 0.053883381 |
| TLR5 | 0.056670551 | -0.052086503 | 0.004584049 |
| STARD7 | 0.104751328 | -0.084652979 | 0.020098349 |
| MRPS10 | 0.09271489 | -0.077645278 | 0.015069612 |
| EPHX1 | 0.059772395 | -0.037595014 | 0.022177381 |
| GLT8D2 | 0.03610386 | -0.035855896 | 0.000247964 |
| NDFIP1 | 0.144061471 | -0.18933734 | -0.045275869 |
| NTMT1 | 0.073532705 | -0.028283649 | 0.045249056 |
| RPP21 | 0.104922763 | -0.074280408 | 0.030642356 |

| | | | |
|---|---|---|---|
| LINC02471 | 0.053901553 | -0.045773393 | 0.00812816 |
| TCEA3 | 0.097100446 | -0.042836597 | 0.054263849 |
| SEPTIN1 | 0.064098366 | -0.065971852 | -0.001873485 |
| RCN2 | 0.125579163 | -0.127760483 | -0.002181321 |
| HSPA9 | 0.118510181 | -0.07869293 | 0.039817251 |
| CCDC115 | 0.104235953 | -0.080661962 | 0.023573992 |
| TXN | 0.147063008 | 0.113308951 | 0.260371959 |
| MRPL49 | 0.08694701 | -0.076196808 | 0.010750202 |
| LRPAP1 | 0.141574379 | -0.198366458 | -0.056792079 |
| CIAO2A | 0.092010514 | -0.095905771 | -0.003895256 |
| POLM | 0.075083844 | -0.078954999 | -0.003871155 |
| ATG3 | 0.089083178 | -0.079642064 | 0.009441114 |
| ABHD17A | 0.096480629 | -0.074370269 | 0.02211036 |
| POLR3GL | 0.085537044 | -0.08210814 | 0.003428904 |
| COMMD1 | 0.093400159 | -0.065307894 | 0.028092265 |
| OGT | 0.108760243 | -0.161662204 | -0.052901961 |
| FAM171A1 | -0.083162402 | 0.055465076 | -0.027697326 |
| TSPAN3 | 0.121252501 | -0.127064189 | -0.005811688 |
| ZNF618 | -0.080296886 | 0.051946783 | -0.028350103 |
| GNL1 | 0.077236425 | -0.085968686 | -0.008732262 |
| SEPTIN8 | 0.099034135 | -0.117546829 | -0.018512693 |
| SYBU | 0.046700106 | -0.037752708 | 0.008947398 |
| RFC1 | 0.110524181 | -0.12951742 | -0.018993239 |
| PPP2CA | 0.093453282 | -0.069704788 | 0.023748495 |
| CAPRIN1 | 0.10595249 | -0.104986417 | 0.000966074 |
| SDHAF2 | 0.083090772 | -0.052440579 | 0.030650193 |
| GFPT1 | 0.079258105 | -0.068463432 | 0.010794673 |
| WWC3 | 0.079237079 | -0.090313779 | -0.0110767 |
| TMPO | 0.111148057 | -0.137105089 | -0.025957032 |
| PHF23 | 0.0770226 | -0.060986767 | 0.016035833 |
| SORT1 | 0.110433731 | -0.152084594 | -0.041650863 |
| MRPL27 | 0.098518037 | -0.057239072 | 0.041278965 |
| NBDY | 0.112512083 | -0.069486819 | 0.043025263 |
| NOL4L | 0.052989193 | -0.038575947 | 0.014413246 |
| SMARCB1 | 0.091500773 | -0.076797801 | 0.014702972 |
| EID1 | 0.152982658 | -0.124697813 | 0.028284845 |
| ISOC1 | 0.068807767 | -0.04461906 | 0.024188707 |
| LMCD1 | 0.057327307 | -0.035551275 | 0.021776032 |
| DCTN6 | 0.0960086 | -0.071323356 | 0.024685244 |
| GPC1 | 0.068998608 | -0.071092247 | -0.002093639 |
| BORCS7 | 0.108258254 | -0.071839122 | 0.036419132 |
| TMED10 | 0.126497294 | -0.138350431 | -0.011853138 |
| HNRNPH2 | 0.095369103 | -0.070589602 | 0.024779501 |
| NT5C3A | 0.065440374 | -0.040971688 | 0.024468686 |
| SRPK2 | 0.072828522 | -0.077719182 | -0.00489066 |
| MISP3 | 0.077025991 | -0.066518668 | 0.010507323 |
| GDF15 | 0.054516406 | -0.04956981 | 0.004946595 |

| | | | |
|---|---|---|---|
| FBL | 0.108643238 | -0.141501907 | -0.032858669 |
| ATP12A | 0.044089643 | -0.044168537 | -7.88941E-05 |
| CAPN5 | 0.058828796 | -0.066334298 | -0.007505502 |
| DDX1 | 0.109282782 | -0.09049804 | 0.018784742 |
| CYP1B1-AS1 | 0.050690014 | -0.052896355 | -0.002206341 |
| GNG10 | 0.118721245 | -0.057416338 | 0.061304907 |
| METTL5 | 0.098365201 | -0.0663443 | 0.032020901 |
| VCP | 0.107011627 | -0.075431776 | 0.031579851 |
| SDHAF3 | 0.079807572 | -0.047866175 | 0.031941397 |
| FRMD3 | -0.078974229 | 0.095255653 | 0.016281424 |
| MRM2 | 0.065856513 | -0.047674241 | 0.018182272 |
| ARMCX3 | 0.097408311 | -0.113644006 | -0.016235695 |
| ATP2B1 | 0.10118239 | -0.122914314 | -0.021731925 |
| TCTA | 0.082844183 | -0.090894575 | -0.008050393 |
| TOMM20 | 0.151219689 | -0.17290794 | -0.021688251 |
| OSBPL8 | 0.087954137 | -0.079066291 | 0.008887846 |
| IRX3 | -0.138106504 | 0.088998431 | -0.049108072 |
| HMGB1 | 0.153217695 | -0.145497456 | 0.007720239 |
| ALDOA | 0.067412444 | -0.049790054 | 0.017622389 |
| CARD19 | 0.068561261 | -0.042654479 | 0.025906782 |
| ADAMTS1 | -0.063195851 | 0.043131153 | -0.020064698 |
| TMEM203 | 0.090290885 | -0.077527851 | 0.012763034 |
| NFU1 | 0.083254033 | -0.057501921 | 0.025752113 |
| CHST3 | -0.05490343 | 0.036280793 | -0.018622637 |
| HOXB7 | 0.109095191 | -0.126849022 | -0.017753831 |
| TMEM106C | 0.107372176 | -0.105628262 | 0.001743914 |
| TRAPPC5 | 0.13095362 | -0.082543462 | 0.048410158 |
| TMEM150A | 0.055173243 | -0.049766816 | 0.005406427 |
| AKAP5 | 0.079549677 | -0.088223389 | -0.008673712 |
| CTSL | 0.133225808 | -0.163656715 | -0.030430907 |
| HM13 | 0.110745855 | -0.10672448 | 0.004021374 |
| DNAJC8 | 0.119859464 | -0.117075523 | 0.002783941 |
| COMMD4 | 0.101817432 | -0.084882862 | 0.01693457 |
| SEMA3C | 0.042389165 | -0.040849876 | 0.00153929 |
| QDPR | 0.081823036 | -0.046409619 | 0.035413417 |
| MT-ATP8 | -0.148086528 | 0.07451006 | -0.073576468 |
| SERPINA5 | 0.047068698 | -0.034475622 | 0.012593076 |
| PRDX3 | 0.129694808 | -0.072541455 | 0.057153354 |
| NUDT5 | 0.085725295 | -0.071558113 | 0.014167182 |
| PTGES2 | 0.079544361 | -0.054194371 | 0.02534999 |
| SLC46A3 | 0.046640662 | -0.045953189 | 0.000687473 |
| FDX2 | 0.080149059 | -0.048140764 | 0.032008295 |
| TMEM60 | 0.071307362 | -0.061476373 | 0.009830989 |
| SEPTIN10 | 0.095838151 | -0.074664425 | 0.021173726 |
| AFDN | 0.120400253 | -0.164837581 | -0.044437327 |
| TMEM208 | 0.114407337 | -0.092786319 | 0.021621018 |
| ATF1 | 0.062539433 | -0.052131132 | 0.010408302 |

| | | | |
|---|---|---|---|
| GTF2H4 | 0.051874212 | -0.034536229 | 0.017337983 |
| ROGDI | 0.061204386 | -0.046391178 | 0.014813208 |
| SYPL1 | 0.130909742 | -0.13555453 | -0.004644788 |
| PHF1 | 0.088064856 | -0.093670114 | -0.005605257 |
| TMEM131 | 0.093401569 | -0.121011474 | -0.027609905 |
| SOGA1 | -0.100742437 | 0.045300741 | -0.055441696 |
| UBAC1 | 0.08081763 | -0.076676451 | 0.004141179 |
| EMC3 | 0.092056325 | -0.081142279 | 0.010914047 |
| ASCL2 | 0.033228701 | -0.032436493 | 0.000792209 |
| EMC4 | 0.104543113 | -0.076205274 | 0.028337839 |
| ZSCAN16-AS1 | 0.101688517 | -0.060205925 | 0.041482592 |
| CRACR2B | 0.053152753 | -0.050308591 | 0.002844163 |
| BUD31 | 0.117225457 | -0.086051521 | 0.031173937 |
| ABT1 | 0.066795824 | -0.039010512 | 0.027785312 |
| BAG5 | 0.0891052 | -0.106723407 | -0.017618207 |
| PPP1R35 | 0.093100474 | -0.108981266 | -0.015880792 |
| MRPL16 | 0.083843658 | -0.03357047 | 0.050273187 |
| SNX17 | 0.098500241 | -0.070395092 | 0.028105149 |
| SLC17A5 | 0.065784306 | -0.069164001 | -0.003379694 |
| TMEM126B | 0.094089348 | -0.072357906 | 0.021731442 |
| DHRS7B | 0.078368315 | -0.076675417 | 0.001692898 |
| SERTAD1 | 0.067900814 | -0.046728154 | 0.02117266 |
| TCF25 | 0.120145439 | -0.158924693 | -0.038779253 |
| PEX2 | 0.103425564 | -0.091150999 | 0.012274565 |
| TDG | 0.070020871 | -0.075462944 | -0.005442072 |
| STIP1 | 0.087382551 | -0.096433525 | -0.009050974 |
| SLC27A5 | 0.059606582 | -0.043303825 | 0.016302757 |
| OVOL2 | 0.051250136 | -0.05157515 | -0.000325014 |
| PRPF38B | 0.097340364 | -0.105727288 | -0.008386924 |
| MRPL48 | 0.069370328 | -0.058044787 | 0.011325541 |
| TNFSF12 | 0.078028673 | -0.064829428 | 0.013199246 |
| CNIH4 | 0.098899882 | -0.117251944 | -0.018352062 |
| MRPS18C | 0.103985562 | -0.053360711 | 0.050624851 |
| TSPAN8 | 0.078090527 | -0.080597857 | -0.00250733 |
| CDK6 | 0.05523914 | -0.050127819 | 0.005111321 |
| SMAP1 | 0.067448126 | -0.054692438 | 0.012755688 |
| HAX1 | 0.101799037 | -0.078884048 | 0.022914989 |
| ATP6V1D | 0.111980185 | -0.072721655 | 0.03925853 |
| SERTAD3 | 0.068564763 | -0.041056418 | 0.027508345 |
| EZR | 0.138442972 | -0.090034615 | 0.048408357 |
| CUL2 | 0.06132198 | -0.054239069 | 0.007082911 |
| IFNGR2 | 0.092652069 | -0.082524694 | 0.010127374 |
| THSD4 | -0.080632735 | 0.056168737 | -0.024463998 |
| ALDOC | 0.051730721 | -0.04297271 | 0.008758011 |
| ZBTB1 | 0.08030484 | -0.104386091 | -0.02408125 |
| MYCBP | 0.067297113 | -0.066675522 | 0.000621592 |
| PPDPF | 0.1560289 | 0.092261342 | 0.248290243 |

| | | | |
|---|---|---|---|
| KMT5A | 0.077211355 | -0.088538629 | -0.011327274 |
| DTYMK | 0.073526575 | -0.05327166 | 0.020254914 |
| PGM1 | 0.056704925 | -0.041655237 | 0.015049688 |
| MTA3 | 0.072551597 | -0.064633965 | 0.007917632 |
| TCP1 | 0.103015005 | -0.079903122 | 0.023111883 |
| PPFIBP1 | 0.13521444 | -0.132305321 | 0.002909119 |
| YY1 | 0.115690508 | -0.133482048 | -0.01779154 |
| COPS2 | 0.093284467 | -0.10662006 | -0.013335594 |
| RSBN1L | 0.104349079 | -0.130963611 | -0.026614531 |
| FBN2 | 0.038020695 | -0.038322521 | -0.000301826 |
| FBXL15 | 0.086204496 | -0.082251948 | 0.003952548 |
| NCOA3 | 0.080249792 | -0.093203793 | -0.012954001 |
| EGLN1 | 0.073646359 | -0.074227521 | -0.000581161 |
| SYNE4 | 0.06766547 | -0.067021477 | 0.000643993 |
| BCAS4 | 0.063665315 | -0.05620616 | 0.007459155 |
| SNX7 | 0.06905255 | -0.034076918 | 0.034975632 |
| ABCF1 | 0.088702311 | -0.082546923 | 0.006155388 |
| HNF1B | -0.10239871 | 0.045121167 | -0.057277544 |
| EIF4B | 0.136323197 | -0.11540549 | 0.020917707 |
| BIN3 | 0.051447207 | -0.049168256 | 0.002278951 |
| SPTSSA | 0.070150239 | -0.05580666 | 0.014343579 |
| PCYOX1 | 0.111309397 | -0.157036645 | -0.045727248 |
| C12orf10 | 0.092782998 | -0.055413966 | 0.037369032 |
| SAR1B | 0.087205327 | -0.121705235 | -0.034499909 |
| CPSF6 | 0.089174493 | -0.112825122 | -0.023650629 |
| ERP44 | 0.090309234 | -0.100465369 | -0.010156135 |
| DYNLT3 | 0.096737865 | -0.101140157 | -0.004402292 |
| INSIG1 | 0.05452116 | -0.060950599 | -0.006429439 |
| COL14A1 | 0.077659078 | -0.07523828 | 0.002420798 |
| MARCH2 | 0.069516325 | -0.074817267 | -0.005300942 |
| TMX1 | 0.085715576 | -0.087850369 | -0.002134793 |
| ALOX5 | 0.032891953 | -0.032891953 | -2.13163E-14 |
| TIMM10 | 0.088047099 | -0.055230718 | 0.032816382 |
| WASHC3 | 0.09205203 | -0.082377754 | 0.009674277 |
| IFITM2 | 0.115535 | -0.145211598 | -0.029676599 |
| CRLS1 | 0.077957705 | -0.045715415 | 0.03224229 |
| PCNA | 0.087309981 | -0.073889394 | 0.013420587 |
| SH3BP2 | 0.054263017 | -0.044402956 | 0.009860061 |
| VAPB | 0.069275256 | -0.060271188 | 0.009004068 |
| AC245297.3 | 0.091797434 | -0.117153851 | -0.025356417 |
| PCSK1N | 0.106295784 | -0.064918791 | 0.041376993 |
| ITGB2-AS1 | 0.033541931 | -0.03258372 | 0.00095821 |
| C8orf76 | 0.056073794 | -0.056166375 | -9.25814E-05 |
| SCPEP1 | 0.081849818 | -0.113460392 | -0.031610574 |
| MTMR12 | 0.074093956 | -0.077632486 | -0.003538531 |
| CHMP3 | 0.108295119 | -0.086162552 | 0.022132568 |
| TIMM10B | 0.070978604 | -0.04728703 | 0.023691574 |

| | | | |
|---|---|---|---|
| MPDU1 | 0.077637407 | -0.059836152 | 0.017801254 |
| FOS | -0.12130985 | 0.106705427 | -0.014604423 |
| HPN | 0.1013818 | -0.085462761 | 0.015919038 |
| DNAH17 | 0.035830337 | -0.0311781 | 0.004652237 |
| RRP36 | 0.073790015 | -0.052974481 | 0.020815534 |
| LINC02212 | 0.031642618 | -0.029816817 | 0.0018258 |
| MED10 | 0.101409762 | -0.087740743 | 0.013669019 |
| CARD6 | 0.034199098 | -0.032210062 | 0.001989036 |
| PARD3B | 0.070058303 | -0.071811884 | -0.001753581 |
| IGSF8 | 0.088089359 | -0.110722973 | -0.022633614 |
| NOP14 | 0.055727478 | -0.057057546 | -0.001330068 |
| CDC42EP4 | 0.052719583 | -0.040587974 | 0.012131608 |
| PRELID3B | 0.07123167 | -0.077439976 | -0.006208306 |
| NADK | 0.056582268 | -0.051599548 | 0.00498272 |
| DPM2 | 0.086037289 | -0.080599821 | 0.005437468 |
| COA3 | 0.113193085 | -0.070489866 | 0.042703219 |
| CERS2 | 0.113610664 | -0.11023455 | 0.003376113 |
| GALNT18 | 0.100444135 | -0.085329517 | 0.015114619 |
| PHYHD1 | 0.070298267 | -0.069187363 | 0.001110904 |
| RDH14 | 0.087233195 | -0.105602894 | -0.018369699 |
| TM9SF1 | 0.08860477 | -0.120043439 | -0.031438669 |
| ITPR2 | 0.066919168 | -0.078727253 | -0.011808085 |
| QPCT | 0.070257398 | -0.062810128 | 0.00744727 |
| AC010642.2 | 0.074961296 | -0.038378385 | 0.036582911 |
| CCNDBP1 | 0.094086717 | -0.112514076 | -0.018427359 |
| ZDHHC16 | 0.063416956 | -0.059161047 | 0.004255909 |
| CAST | 0.134829797 | -0.164887763 | -0.030057967 |
| PFKFB3 | 0.143483245 | -0.261394463 | -0.117911218 |
| STX4 | 0.075758916 | -0.06753412 | 0.008224796 |
| EIF1B | 0.130024193 | -0.072303839 | 0.057720354 |
| TRAPPC3 | 0.079468365 | -0.082522614 | -0.003054249 |
| NHP2 | 0.110295774 | -0.066088042 | 0.044207732 |
| LYPLAL1 | 0.104383705 | -0.070036601 | 0.034347104 |
| PRADC1 | 0.056180609 | -0.05048727 | 0.005693339 |
| SERPINB6 | 0.092019966 | -0.076419704 | 0.015600262 |
| KIF3A | 0.072384534 | -0.07895105 | -0.006566516 |
| RDH11 | 0.106140383 | -0.149419057 | -0.043278674 |
| UBL4A | 0.067766437 | -0.058259326 | 0.009507111 |
| UBE2E2 | 0.066150386 | -0.037733415 | 0.028416971 |
| MCF2L-AS1 | 0.029392504 | -0.029392504 | -2.30926E-14 |
| SNAPIN | 0.077829576 | -0.049567346 | 0.02826223 |
| UBE2I | 0.106427956 | -0.072753285 | 0.033674671 |
| HDAC11 | 0.066463462 | -0.04647882 | 0.019984642 |
| UBE2Q2 | 0.061754695 | -0.043285398 | 0.018469297 |
| SLC49A3 | 0.045375676 | -0.046125868 | -0.000750193 |
| MAPK13 | 0.063112959 | -0.061507212 | 0.001605747 |
| BORCS8 | 0.08196528 | -0.063211655 | 0.018753625 |

| | | | |
|---|---|---|---|
| DERL2 | 0.086857549 | -0.094947002 | -0.008089453 |
| HDGF | 0.108351827 | -0.122073119 | -0.013721292 |
| RPA3 | 0.112523585 | -0.052707815 | 0.05981577 |
| GGCT | 0.066730759 | -0.040901512 | 0.025829247 |
| CSNK2B | 0.108998125 | -0.068719593 | 0.040278532 |
| WDR18 | 0.070086202 | -0.030762724 | 0.039323479 |
| EPS8L2 | 0.137371525 | -0.306102201 | -0.168730676 |
| YPEL3 | 0.126785668 | -0.13567585 | -0.008890182 |
| TMED2 | 0.114280382 | -0.113340956 | 0.000939425 |
| ADGRL3 | -0.058131756 | 0.031681231 | -0.026450526 |
| HMGB2 | -0.100815518 | 0.107988831 | 0.007173313 |
| TMED5 | 0.098352771 | -0.115741388 | -0.017388617 |
| SPACA9 | 0.053127976 | -0.054310992 | -0.001183016 |
| SC5D | 0.079529249 | -0.097350178 | -0.017820929 |
| HOOK2 | 0.089741786 | -0.098744826 | -0.00900304 |
| CWC15 | 0.104354165 | -0.074406857 | 0.029947308 |
| SMIM14 | 0.098212143 | -0.101497641 | -0.003285498 |
| CDC37 | 0.109274211 | -0.092889772 | 0.016384439 |
| GALNT1 | 0.067129602 | -0.070016134 | -0.002886532 |
| MTFP1 | 0.050218986 | -0.041752038 | 0.008466947 |
| CBX5 | 0.126613267 | -0.184554438 | -0.057941171 |
| RAD23B | 0.083836801 | -0.072823651 | 0.01101315 |
| ZC3H15 | 0.101014395 | -0.09550615 | 0.005508244 |
| POLR2J | 0.115254366 | -0.070421382 | 0.044832985 |
| CD58 | 0.05771189 | -0.053068138 | 0.004643751 |
| ADIPOR1 | 0.084995102 | -0.098443504 | -0.013448402 |
| MMADHC | 0.080160582 | -0.039494319 | 0.040666263 |
| CSNK2A2 | 0.07220812 | -0.07567232 | -0.0034642 |
| CFAP20 | 0.057776368 | -0.043492583 | 0.014283785 |
| SDHAF1 | 0.068691024 | -0.072252149 | -0.003561125 |
| HSPG2 | 0.117567122 | -0.173498954 | -0.055931831 |
| GLRX2 | 0.063860942 | -0.044738235 | 0.019122707 |
| UNG | 0.05832383 | -0.054432996 | 0.003890834 |
| MAGEF1 | 0.091267388 | -0.083506492 | 0.007760896 |
| GSTM4 | 0.05704197 | -0.036076557 | 0.020965414 |
| SQSTM1 | 0.134349759 | -0.08185856 | 0.052491199 |
| C1orf52 | 0.068211755 | -0.065013241 | 0.003198514 |
| P4HTM | 0.09793487 | -0.123245843 | -0.025310973 |
| AKT2 | 0.094061131 | -0.114962695 | -0.020901564 |
| PRPF4B | 0.124328828 | -0.181668875 | -0.057340047 |
| MRPL43 | 0.112443189 | -0.120081529 | -0.00763834 |
| FBXL5 | 0.093171569 | -0.087165198 | 0.006006371 |
| ERGIC1 | 0.077997792 | -0.094194557 | -0.016196765 |
| TALDO1 | 0.128633757 | -0.054784895 | 0.073848862 |
| PLCXD3 | 0.036375499 | -0.034652255 | 0.001723244 |
| ATP6V1E1 | 0.098792779 | -0.049194377 | 0.049598402 |
| GRB2 | 0.090352827 | -0.071830163 | 0.018522663 |

| | | | |
|---|---|---|---|
| FBXO25 | 0.069175033 | -0.056062756 | 0.013112277 |
| NSFL1C | 0.079021469 | -0.080227062 | -0.001205593 |
| SNX27 | 0.07784014 | -0.07128311 | 0.00655703 |
| PWP1 | 0.074162646 | -0.069856613 | 0.004306033 |
| ACTR10 | 0.0956216 | -0.059883201 | 0.035738398 |
| RANBP6 | 0.069838346 | -0.074509825 | -0.004671479 |
| ECHDC1 | 0.080584101 | -0.051816438 | 0.028767663 |
| PSD3 | 0.037995125 | -0.036738895 | 0.00125623 |
| PET100 | 0.130816343 | -0.123780243 | 0.0070361 |
| DNAJB14 | 0.09836103 | -0.088971531 | 0.009389499 |
| PREPL | 0.102722245 | -0.145222689 | -0.042500443 |
| NEK7 | 0.070925148 | -0.078635765 | -0.007710617 |
| TMUB1 | 0.083613213 | -0.073368233 | 0.01024498 |
| GPC5 | 0.034009782 | -0.036407798 | -0.002398016 |
| STAMBP | 0.067172918 | -0.050336152 | 0.016836766 |
| DTWD1 | 0.08351613 | -0.085795432 | -0.002279302 |
| AL031123.4 | 0.060792222 | -0.060618024 | 0.000174198 |
| TXNDC12 | 0.095851348 | -0.10627332 | -0.010421973 |
| LRRFIP1 | 0.117491339 | -0.160361474 | -0.042870135 |
| SERTAD4 | 0.061676951 | -0.074158577 | -0.012481626 |
| TRIM2 | 0.099028619 | -0.123017834 | -0.023989215 |
| RHOF | 0.071506938 | -0.068300481 | 0.003206458 |
| TTC9 | 0.041474838 | -0.034379551 | 0.007095287 |
| AMIGO2 | 0.044480739 | -0.040307679 | 0.00417306 |
| FYTTD1 | 0.074758393 | -0.062648649 | 0.012109744 |
| HLA-DMB | 0.070797069 | -0.058592427 | 0.012204641 |
| VGLL4 | 0.091511008 | -0.073250835 | 0.018260172 |
| API5 | 0.078186393 | -0.094623833 | -0.01643744 |
| CACYBP | 0.090159921 | -0.076106357 | 0.014053563 |
| IQCD | 0.045870494 | -0.047633516 | -0.001763023 |
| LRRC32 | -0.041853187 | 0.024714463 | -0.017138724 |
| SMARCA1 | 0.081277286 | -0.082568379 | -0.001291093 |
| ILK | 0.091544647 | -0.051160005 | 0.040384642 |
| AHSA1 | 0.090410716 | -0.08235973 | 0.008050986 |
| LINC01315 | 0.054196076 | -0.043358109 | 0.010837967 |
| MOCS2 | 0.105725183 | -0.082068282 | 0.0236569 |
| CLYBL | 0.04881934 | -0.035020948 | 0.013798391 |
| MOB4 | 0.087956913 | -0.077054992 | 0.010901921 |
| STX8 | 0.078982465 | -0.044443481 | 0.034538984 |
| EIF2S1 | 0.082218464 | -0.074583207 | 0.007635257 |
| SPATC1L | 0.042224438 | -0.036900313 | 0.005324125 |
| BAG3 | 0.060727545 | -0.056541348 | 0.004186197 |
| ARFGAP3 | 0.072546537 | -0.054538904 | 0.018007634 |
| NSF | 0.058905905 | -0.044749316 | 0.014156589 |
| FERMT2 | 0.069119374 | -0.052186568 | 0.016932806 |
| CAPN1 | 0.096598277 | -0.085564154 | 0.011034123 |
| TMEM116 | 0.068121512 | -0.075168716 | -0.007047203 |

| | | | |
|---|---|---|---|
| XRN2 | 0.092318188 | -0.077875215 | 0.014442973 |
| MTX1 | 0.074457442 | -0.046746781 | 0.027710661 |
| TMSB10 | 0.144181637 | 0.209103768 | 0.353285405 |
| PRXL2A | 0.101481265 | -0.102301103 | -0.000819838 |
| CHP1 | 0.084773436 | -0.057722494 | 0.027050942 |
| ZNRD1 | 0.077266696 | -0.065282046 | 0.01198465 |
| GPAT3 | -0.050857659 | 0.027459232 | -0.023398427 |
| PICALM | 0.108066328 | -0.109331168 | -0.00126484 |
| MRPL28 | 0.087655772 | -0.05614581 | 0.031509962 |
| VPS37D | 0.061476101 | -0.046632914 | 0.014843187 |
| UBE2V1 | 0.10845818 | -0.110567385 | -0.002109206 |
| DPP3 | 0.0376559 | -0.036944669 | 0.000711231 |
| IFNAR1 | 0.09859616 | -0.122790764 | -0.024194603 |
| NAPA | 0.105056611 | -0.082355878 | 0.022700733 |
| SLC2A1 | 0.055216483 | -0.068778775 | -0.013562292 |
| UHMK1 | 0.08410019 | -0.101715182 | -0.017614992 |
| TMEM120A | 0.0599108 | -0.046148342 | 0.013762459 |
| SARNP | 0.096821196 | -0.072886137 | 0.023935059 |
| AC103702.2 | -0.072268379 | 0.047275346 | -0.024993034 |
| CBX1 | 0.09952062 | -0.098853699 | 0.000666921 |
| NELFCD | 0.081972927 | -0.079410899 | 0.002562028 |
| RUNX1 | 0.033616886 | -0.031884248 | 0.001732638 |
| NUDT8 | 0.066030817 | -0.055111503 | 0.010919314 |
| UBE2R2 | 0.086739032 | -0.088487972 | -0.00174894 |
| TRAM1 | 0.117955624 | -0.165519939 | -0.047564316 |
| MRPL2 | 0.084697199 | -0.073827758 | 0.010869441 |
| FLOT1 | 0.094879879 | -0.071894995 | 0.022984884 |
| SDF2L1 | 0.084817159 | -0.093116778 | -0.008299619 |
| ZNF770 | 0.074634218 | -0.082279673 | -0.007645455 |
| GSK3A | 0.068082155 | -0.035880057 | 0.032202098 |
| TMEM183A | 0.078018645 | -0.08576354 | -0.007744896 |
| KIF26B | 0.03597013 | -0.040978522 | -0.005008392 |
| PTPRN2 | 0.050464669 | -0.039833897 | 0.010630772 |
| MESD | 0.092412808 | -0.087453834 | 0.004958975 |
| PRMT2 | 0.093121168 | -0.092908046 | 0.000213122 |
| SIKE1 | 0.067701541 | -0.054956681 | 0.012744861 |
| YIPF3 | 0.111580773 | -0.120634143 | -0.00905337 |
| SRSF2 | 0.117669078 | -0.119248023 | -0.001578945 |
| SFT2D1 | 0.115595107 | -0.137217364 | -0.021622257 |
| MEGF9 | 0.066976171 | -0.070342523 | -0.003366353 |
| PDCL3 | 0.063764646 | -0.035332439 | 0.028432207 |
| TBK1 | 0.057597627 | -0.049899549 | 0.007698078 |
| TK1 | 0.045846938 | -0.057792692 | -0.011945754 |
| CHCHD10 | 0.139471193 | -0.079198238 | 0.060272955 |
| PPP1R2 | 0.085883277 | -0.050874869 | 0.035008408 |
| HLA-DQB1 | 0.034480694 | -0.02690121 | 0.007579483 |
| HOXA10-AS | 0.070688699 | -0.073019829 | -0.00233113 |

| | | | |
|---|---|---|---|
| PAFAH1B3 | 0.067039674 | -0.031283279 | 0.035756396 |
| MIR210HG | 0.05904223 | -0.047632888 | 0.011409341 |
| ERCC1 | 0.070775433 | -0.044636884 | 0.026138549 |
| NAPG | 0.066682845 | -0.06443638 | 0.002246465 |
| ELAVL1 | 0.082656455 | -0.089184294 | -0.00652784 |
| MAPRE3 | 0.054391573 | -0.049731391 | 0.004660182 |
| MED13L | -0.088238718 | 0.043253939 | -0.044984779 |
| GPN3 | 0.086727117 | -0.080937144 | 0.005789973 |
| WDR45B | 0.087739197 | -0.088818772 | -0.001079575 |
| STON2 | -0.054180153 | 0.023931693 | -0.03024846 |
| ACSL4 | 0.07040559 | -0.062684455 | 0.007721135 |
| ZFYVE16 | 0.09790832 | -0.11716182 | -0.0192535 |
| TMEM68 | 0.079729178 | -0.092830356 | -0.013101178 |
| SRP54 | 0.083389467 | -0.091989075 | -0.008599608 |
| PRAF2 | 0.049234368 | -0.046353317 | 0.00288105 |
| MAGOH | 0.100113262 | -0.069073083 | 0.031040179 |
| NAP1L4 | 0.097858351 | -0.09149791 | 0.006360441 |
| TCEAL8 | 0.093792948 | -0.043751293 | 0.050041655 |
| CHD5 | -0.039165404 | 0.018716666 | -0.020448738 |
| SIGIRR | 0.07968453 | -0.053911697 | 0.025772833 |
| TMEM126A | 0.069866607 | -0.053012965 | 0.016853642 |
| TOMM22 | 0.100252893 | -0.06929221 | 0.030960683 |
| NFAT5 | 0.10136205 | -0.142575792 | -0.041213742 |
| RTF1 | 0.096191305 | -0.077836809 | 0.018354496 |
| COMMD8 | 0.086930276 | -0.057921414 | 0.029008862 |
| ADGRG1 | 0.082850612 | -0.071417131 | 0.011433481 |
| NDUFA10 | 0.099324811 | -0.090214085 | 0.009110726 |
| UBE2F | 0.055664193 | -0.041009531 | 0.014654662 |
| MPP1 | 0.061974891 | -0.03295492 | 0.029019971 |
| EIF2B2 | 0.069772664 | -0.062610798 | 0.007161866 |
| TMEM8A | 0.055910072 | -0.047119509 | 0.008790563 |
| AMPH | 0.031440497 | -0.032765735 | -0.001325238 |
| HERPUD1 | 0.126603446 | -0.167032251 | -0.040428805 |
| SNRNP25 | 0.07465217 | -0.048694423 | 0.025957747 |
| DNM2 | 0.093431906 | -0.103740491 | -0.010308586 |
| DHRS4L2 | 0.077781708 | -0.054684436 | 0.023097272 |
| TMOD3 | 0.071341776 | -0.050043911 | 0.021297864 |
| SNRNP35 | 0.081465061 | -0.064943319 | 0.016521742 |
| STAP2 | 0.064020846 | -0.04334512 | 0.020675726 |
| MRPL44 | 0.060123533 | -0.027287724 | 0.032835809 |
| CFLAR | 0.108100994 | -0.170878612 | -0.062777618 |
| CDC42BPA | 0.084487873 | -0.11219828 | -0.027710407 |
| SLC25A38 | 0.055324428 | -0.068969119 | -0.013644691 |
| TPST1 | 0.065068381 | -0.063336662 | 0.001731719 |
| PDCD6IP | 0.094527274 | -0.106554382 | -0.012027108 |
| TBC1D16 | 0.081910422 | -0.090985461 | -0.009075039 |
| AC004130.2 | 0.053317859 | -0.041054133 | 0.012263726 |

| | | | |
|---|---|---|---|
| TRPS1 | 0.059084445 | -0.056901768 | 0.002182677 |
| B4GAT1 | 0.080085827 | -0.103837021 | -0.023751193 |
| AF064858.1 | 0.030445368 | -0.030445368 | -2.13163E-14 |
| ATIC | 0.058583197 | -0.040011934 | 0.018571263 |
| PLEKHB2 | 0.087639228 | -0.096461584 | -0.008822356 |
| ZC3H7A | 0.077840757 | -0.0686006 | 0.009240157 |
| RABGGTB | 0.073856307 | -0.07723663 | -0.003380324 |
| IDH3B | 0.077087679 | -0.070142236 | 0.006945442 |
| HADHA | 0.104929354 | -0.101962344 | 0.002967009 |
| OSBP | 0.091871136 | -0.13632972 | -0.044458584 |
| ECI2 | 0.092107217 | -0.05892175 | 0.033185467 |
| MAGED2 | 0.121099917 | -0.073915109 | 0.047184808 |
| BDKRB2 | 0.030701606 | -0.029145819 | 0.001555787 |
| UBE2L3 | 0.097025671 | -0.048144999 | 0.048880672 |
| R3HCC1 | 0.073051539 | -0.05292638 | 0.020125159 |
| SEC62 | 0.137434761 | -0.190954454 | -0.053519693 |
| HIKESHI | 0.07692904 | -0.041592742 | 0.035336298 |
| HIST1H2AC | 0.117296046 | -0.146359645 | -0.029063599 |
| LARP1B | 0.060693621 | -0.054535704 | 0.006157917 |
| BABAM2 | 0.071116977 | -0.070841969 | 0.000275008 |
| GLA | 0.048625076 | -0.04762302 | 0.001002056 |
| MLST8 | 0.048665625 | -0.033995157 | 0.014670468 |
| ITGB2 | 0.028495046 | -0.028495046 | -1.95399E-14 |
| BZW1 | 0.102432657 | -0.081338408 | 0.021094248 |
| THBD | -0.040636081 | 0.031990679 | -0.008645402 |
| HOXA9 | 0.09942413 | -0.119718049 | -0.020293919 |
| ARCN1 | 0.078884762 | -0.082597345 | -0.003712582 |
| TMEM242 | 0.067223222 | -0.063522922 | 0.0037003 |
| ATP6V1A | 0.093848945 | -0.085931196 | 0.007917749 |
| VTI1B | 0.107304057 | -0.063354782 | 0.043949275 |
| PGGHG | 0.070299636 | -0.086883386 | -0.01658375 |
| IGFBP2 | -0.151892957 | -0.135013405 | -0.286906362 |
| SSBP1 | 0.114562288 | -0.060352017 | 0.05421027 |
| SNRPD1 | 0.101954431 | -0.053760596 | 0.048193835 |
| KDELR2 | 0.096462098 | -0.098744214 | -0.002282116 |
| CTTN | 0.103805553 | -0.115653705 | -0.011848152 |
| ZSWIM7 | 0.081528472 | -0.056396303 | 0.025132169 |
| HSD17B10 | 0.099382722 | -0.048893358 | 0.050489364 |
| MGME1 | 0.065154269 | -0.07152493 | -0.006370661 |
| PRR15 | 0.030232314 | -0.025927664 | 0.00430465 |
| TMEM109 | 0.087484988 | -0.079015609 | 0.008469378 |
| LSM8 | 0.09990718 | -0.061344693 | 0.038562486 |
| RSAD1 | 0.049269893 | -0.041547861 | 0.007722032 |
| ACTR3 | 0.098517124 | -0.061725476 | 0.036791648 |
| ESRRA | 0.063090621 | -0.042786677 | 0.020303944 |
| VPS26A | 0.083631479 | -0.076695418 | 0.006936061 |
| STX5 | 0.065725732 | -0.037152305 | 0.028573427 |

| | | | |
|---|---|---|---|
| OSTC | 0.110808647 | -0.082143841 | 0.028664806 |
| BTF3L4 | 0.096944204 | -0.068022236 | 0.028921968 |
| BACE2 | 0.060214057 | -0.068669493 | -0.008455436 |
| ASAP1 | 0.038467071 | -0.04395746 | -0.005490389 |
| PPP6C | 0.070809502 | -0.053736746 | 0.017072756 |
| SSR3 | 0.103434441 | -0.073994184 | 0.029440257 |
| LRRFIP2 | 0.074743649 | -0.059832901 | 0.014910747 |
| IL17RB | 0.032913085 | -0.025967776 | 0.006945309 |
| PCCB | 0.049616851 | -0.0392546 | 0.010362251 |
| RASSF7 | 0.121581438 | -0.12378161 | -0.002200173 |
| COPS3 | 0.068442583 | -0.052001387 | 0.016441196 |
| IAH1 | 0.090893769 | -0.070258264 | 0.020635504 |
| CAPN6 | 0.037398883 | -0.03411586 | 0.003283023 |
| MIER1 | 0.086066289 | -0.102910569 | -0.01684428 |
| PPP1R36 | 0.027720851 | -0.02897592 | -0.001255069 |
| PPIF | 0.05211993 | -0.045436946 | 0.006682984 |
| TIAL1 | 0.094198981 | -0.104819273 | -0.010620292 |
| SUGT1 | 0.095363415 | -0.084176339 | 0.011187075 |
| TMEM130 | 0.036663229 | -0.036340288 | 0.000322941 |
| UBE2D3 | 0.113008587 | -0.104307995 | 0.008700592 |
| SLC3A1 | -0.068146779 | 0.088326606 | 0.020179827 |
| ARMCX1 | 0.064768023 | -0.056223643 | 0.00854438 |
| MAD2L1BP | 0.044737095 | -0.041040516 | 0.003696579 |
| TUNAR | 0.029150503 | -0.029150503 | -2.13163E-14 |
| RHEB | 0.112392527 | -0.069134315 | 0.043258212 |
| TUBG1 | 0.062473969 | -0.0445693 | 0.017904669 |
| LYPLA2 | 0.073159187 | -0.077878248 | -0.004719061 |
| URI1 | 0.087633384 | -0.101958552 | -0.014325168 |
| SPR | 0.050496608 | -0.043213914 | 0.007282694 |
| HRH1 | -0.050264662 | 0.049495048 | -0.000769614 |
| NCBP3 | 0.072354603 | -0.08475008 | -0.012395477 |
| FASTK | 0.081146757 | -0.079179283 | 0.001967475 |
| LINC01618 | 0.031895516 | -0.033528509 | -0.001632993 |
| PSMG1 | 0.063705398 | -0.032061878 | 0.03164352 |
| HIGD2A | 0.110521151 | -0.063872736 | 0.046648415 |
| FLII | 0.075812441 | -0.071184751 | 0.00462769 |
| ETFRF1 | 0.080708325 | -0.037224179 | 0.043484145 |
| ERRFI1 | -0.063092522 | 0.042034937 | -0.021057585 |
| NKAP | 0.060624325 | -0.049768251 | 0.010856073 |
| TOP1 | 0.094760217 | -0.092238229 | 0.002521988 |
| NR1H2 | 0.08380437 | -0.074458936 | 0.009345434 |
| AIMP1 | 0.101061337 | -0.107664152 | -0.006602814 |
| CCDC28A | 0.085766722 | -0.080435475 | 0.005331247 |
| ZNF581 | 0.051223596 | -0.035252391 | 0.015971204 |
| RNF115 | 0.076623202 | -0.089313633 | -0.012690431 |
| FH | 0.063790394 | -0.051225607 | 0.012564787 |
| AL662884.3 | 0.034855418 | -0.03774048 | -0.002885063 |

| | | | |
|---|---|---|---|
| PLPP4 | -0.043599997 | 0.043314929 | -0.000285068 |
| IVD | 0.075646312 | -0.059091132 | 0.016555179 |
| ECSIT | 0.059487585 | -0.035107963 | 0.024379622 |
| ATG5 | 0.066610912 | -0.056909647 | 0.009701265 |
| ABI3BP | 0.065572321 | -0.08443077 | -0.018858448 |
| PTGER2 | -0.059284793 | 0.042736776 | -0.016548017 |
| PSMC3 | 0.090551701 | -0.041278875 | 0.049272826 |
| FARSA | 0.06633623 | -0.041916972 | 0.024419258 |
| CASTOR3 | 0.044677857 | -0.055901818 | -0.011223961 |
| PAM16 | 0.083046297 | -0.067715878 | 0.015330419 |
| CPNE8 | 0.065089173 | -0.067554905 | -0.002465733 |
| HADH | 0.067312026 | -0.069887562 | -0.002575536 |
| WDR1 | 0.08127181 | -0.068148381 | 0.013123429 |
| ARL6IP6 | 0.064339536 | -0.068022164 | -0.003682628 |
| NRIP1 | -0.081416522 | 0.058516613 | -0.022899909 |
| IMP3 | 0.098075304 | -0.068722737 | 0.029352567 |
| C17orf75 | 0.057646969 | -0.044124494 | 0.013522475 |
| SLC25A33 | 0.065073369 | -0.060488279 | 0.00458509 |
| HSPB11 | 0.090327007 | -0.074783577 | 0.01554343 |
| FBXO3 | 0.068039581 | -0.081411973 | -0.013372393 |
| IRF2BP2 | -0.075402275 | 0.071384089 | -0.004018186 |
| UBA2 | 0.074567975 | -0.090352014 | -0.015784039 |
| OTULINL | 0.032024056 | -0.032356744 | -0.000332689 |
| CMC1 | 0.077530135 | -0.044135079 | 0.033395056 |
| SERP2 | 0.046028208 | -0.038074745 | 0.007953464 |
| NDUFB6 | 0.099027337 | -0.062734949 | 0.036292388 |
| MKRN1 | 0.087546358 | -0.09559268 | -0.008046322 |
| ZNF780A | 0.050712785 | -0.049388229 | 0.001324556 |
| RAB34 | 0.098773527 | -0.10444948 | -0.005675953 |
| CC2D2A | 0.059671094 | -0.048740868 | 0.010930226 |
| MAP7D1 | 0.053206027 | -0.030523578 | 0.022682449 |
| PTGES | 0.029107732 | -0.029005376 | 0.000102356 |
| ATRN | 0.069861011 | -0.083900421 | -0.014039409 |
| DHX9 | 0.096002901 | -0.094783925 | 0.001218977 |
| SF3B5 | 0.110681845 | -0.051891459 | 0.058790386 |
| AC245407.2 | 0.042068497 | -0.033352645 | 0.008715852 |
| CPTP | 0.050755558 | -0.034813895 | 0.015941663 |
| DNAJB12 | 0.071307947 | -0.062986714 | 0.008321234 |
| FAM153CP | 0.028878884 | -0.025241699 | 0.003637185 |
| BEND7 | 0.061650296 | -0.06337959 | -0.001729294 |
| PNMA1 | 0.069674782 | -0.047278044 | 0.022396737 |
| MIR200CHG | 0.031850634 | -0.029754744 | 0.00209589 |
| LRRC42 | 0.059032604 | -0.043198605 | 0.015834 |
| MUC15 | 0.116631036 | -0.242429456 | -0.12579842 |
| ASPH | 0.111598032 | -0.19895931 | -0.087361279 |
| HBP1 | 0.081374347 | -0.066443718 | 0.014930629 |
| GAS2L1 | 0.045017263 | -0.042301249 | 0.002716014 |

| | | | |
|---|---|---|---|
| CNPY3 | 0.06650417 | -0.07144045 | -0.00493628 |
| PTOV1 | 0.079488371 | -0.051390546 | 0.028097825 |
| SF3B2 | 0.096273965 | -0.071835299 | 0.024438666 |
| BUD23 | 0.094424576 | -0.117915238 | -0.023490662 |
| SH3YL1 | 0.099729387 | -0.119304172 | -0.019574785 |
| PSMC1 | 0.085802157 | -0.065708393 | 0.020093764 |
| CRYBB2 | 0.023495153 | -0.023495153 | -2.13163E-14 |
| SAC3D1 | 0.045579382 | -0.034993383 | 0.010585999 |
| SYTL2 | 0.084775818 | -0.115384494 | -0.030608676 |
| MAPK9 | 0.055715066 | -0.045406883 | 0.010308183 |
| HDGFL2 | 0.069637434 | -0.045757421 | 0.023880013 |
| COMMD7 | 0.08659546 | -0.066180079 | 0.02041538 |
| TCEA2 | 0.08489796 | -0.073501065 | 0.011396895 |
| N4BP2L2 | -0.130202694 | -0.068589184 | -0.198791878 |
| LAMP2 | 0.095229592 | -0.141007025 | -0.045777433 |
| CDKN2D | 0.038648313 | -0.040235763 | -0.00158745 |
| AL118522.1 | 0.025434112 | -0.025434112 | -2.13163E-14 |
| METTL23 | 0.078392142 | -0.068099185 | 0.010292956 |
| NTHL1 | 0.05286416 | -0.029374968 | 0.023489192 |
| PAGR1 | 0.078104466 | -0.03782212 | 0.040282346 |
| CCNC | 0.077514543 | -0.050465953 | 0.02704859 |
| ANO10 | 0.066936952 | -0.063322387 | 0.003614565 |
| R3HDM4 | 0.052815168 | -0.053461814 | -0.000646646 |
| PABPC4 | 0.076679622 | -0.084338841 | -0.007659219 |
| MGST2 | 0.078953943 | -0.086505602 | -0.007551659 |
| GAS7 | 0.024498773 | -0.024498773 | -2.30926E-14 |
| BSPRY | 0.051784597 | -0.048566564 | 0.003218033 |
| POLR2C | 0.082088467 | -0.056847805 | 0.025240662 |
| FAM117A | 0.039878625 | -0.038878759 | 0.000999866 |
| DDX49 | 0.054026843 | -0.042658762 | 0.01136808 |
| TSPAN15 | 0.058209328 | -0.070381656 | -0.012172328 |
| CIB4 | 0.02486592 | -0.02486592 | -2.30926E-14 |
| ZNF524 | 0.064740211 | -0.057248755 | 0.007491457 |
| RHPN1 | 0.061090528 | -0.05512064 | 0.005969888 |
| CLPTM1L | 0.078567604 | -0.082565172 | -0.003997568 |
| GYPC | 0.05091216 | -0.024022626 | 0.026889534 |
| KLHL13 | -0.0398382 | 0.026224712 | -0.013613488 |
| MRPS16 | 0.070578203 | -0.041458377 | 0.029119826 |
| TP53I3 | 0.072108326 | -0.034136625 | 0.037971701 |
| METRN | 0.058770829 | -0.036415009 | 0.02235582 |
| AC104461.1 | 0.02429832 | -0.02429832 | -2.30926E-14 |
| UBE2V2 | 0.079269955 | -0.049788543 | 0.029481412 |
| RERE | 0.098827309 | -0.121426515 | -0.022599206 |
| PSMD7 | 0.074269015 | -0.053961452 | 0.020307563 |
| SNRPB2 | 0.091736209 | -0.070186522 | 0.021549687 |
| TMCO1 | 0.11043442 | -0.156295692 | -0.045861272 |
| TMEM70 | 0.057551436 | -0.039287425 | 0.018264011 |

| | | | |
|---|---|---|---|
| SAMM50 | 0.059026748 | -0.044170364 | 0.014856383 |
| HOXA7 | 0.088036757 | -0.077408807 | 0.01062795 |
| CMAS | 0.064042386 | -0.048404386 | 0.015638 |
| EBP | 0.072433934 | -0.069642048 | 0.002791887 |
| TRMT6 | 0.048640474 | -0.039866687 | 0.008773788 |
| ADAM9 | 0.0768849 | -0.082127871 | -0.005242971 |
| SUZ12 | 0.079567025 | -0.079035817 | 0.000531208 |
| NENF | 0.115197231 | -0.08492617 | 0.030271061 |
| TTYH3 | 0.038584235 | -0.033270222 | 0.005314014 |
| STX2 | 0.072501647 | -0.070305029 | 0.002196618 |
| RASSF1 | 0.0492472 | -0.0391784 | 0.010068801 |
| SPAG16 | 0.086288759 | -0.044352506 | 0.041936253 |
| MRPL11 | 0.081179351 | -0.065557997 | 0.015621354 |
| MSRB2 | 0.101289302 | -0.05627926 | 0.045010042 |
| COBL | 0.063993137 | -0.074754741 | -0.010761604 |
| PIGF | 0.076176439 | -0.064797153 | 0.011379285 |
| OPHN1 | 0.049804204 | -0.054478315 | -0.004674111 |
| SH3GL1 | 0.056526268 | -0.045746223 | 0.010780045 |
| CACNA1C | 0.023643924 | -0.023643924 | -1.95399E-14 |
| ZMPSTE24 | 0.06242049 | -0.068163119 | -0.005742628 |
| RBMX | 0.104547377 | -0.095506867 | 0.00904051 |
| RPRD1A | 0.063383437 | -0.059647495 | 0.003735941 |
| RAB24 | 0.042152025 | -0.039716523 | 0.002435503 |
| NECAP2 | 0.057208174 | -0.051341415 | 0.005866759 |
| ZNF706 | 0.106770219 | -0.093831465 | 0.012938754 |
| TUBB6 | 0.064393441 | -0.07097227 | -0.006578829 |
| LRPPRC | 0.086146461 | -0.052092072 | 0.034054389 |
| TOMM40 | 0.065504581 | -0.048929552 | 0.016575029 |
| TMEM259 | 0.091242615 | -0.113483843 | -0.022241228 |
| USP8 | 0.078338134 | -0.065984419 | 0.012353715 |
| BOLA2 | 0.063440434 | -0.055302688 | 0.008137746 |
| SDC2 | 0.116477337 | -0.129089149 | -0.012611812 |
| NYAP1 | 0.036572051 | -0.043329869 | -0.006757818 |
| GID8 | 0.072718332 | -0.057785732 | 0.014932601 |
| FXYD6 | -0.058015993 | 0.099837466 | 0.041821473 |
| TRAFD1 | 0.046716588 | -0.040012058 | 0.00670453 |
| IFT80 | 0.072807997 | -0.099399428 | -0.026591431 |
| VAMP3 | 0.070654683 | -0.038140952 | 0.032513731 |
| PGS1 | 0.048937317 | -0.057848793 | -0.008911476 |
| HMG20B | 0.083209565 | -0.050508705 | 0.032700861 |
| GFM1 | 0.059409002 | -0.077645041 | -0.018236038 |
| SCTR | 0.024539301 | -0.024539301 | -1.95399E-14 |
| UTP18 | 0.063044991 | -0.058392656 | 0.004652335 |
| CRCP | 0.061111963 | -0.038512772 | 0.022599191 |
| MALSU1 | 0.071889711 | -0.05029266 | 0.021597051 |
| SAT1 | 0.131872842 | -0.16871486 | -0.036842018 |
| RPRD2 | 0.07797381 | -0.102927173 | -0.024953363 |

| | | | |
|---|---|---|---|
| STK24 | 0.071513071 | -0.042375287 | 0.029137784 |
| OCEL1 | 0.058121398 | -0.032938003 | 0.025183395 |
| ATP6V0C | 0.125563485 | -0.171605365 | -0.04604188 |
| REEP6 | 0.035146082 | -0.031128164 | 0.004017917 |
| SHLD2 | 0.073559756 | -0.065587092 | 0.007972663 |
| JKAMP | 0.076147754 | -0.09217109 | -0.016023336 |
| EIF3J | 0.091971742 | -0.07438883 | 0.017582911 |
| PSMD1 | 0.075857845 | -0.066670547 | 0.009187297 |
| SGSM3 | 0.066550417 | -0.06547436 | 0.001076057 |
| AKT1 | 0.070725228 | -0.059980481 | 0.010744747 |
| IFT22 | 0.090827725 | -0.044962881 | 0.045864844 |
| SRA1 | 0.066565919 | -0.040216052 | 0.026349867 |
| ZFR | 0.080218066 | -0.08979581 | -0.009577743 |
| RANGRF | 0.064371798 | -0.042373912 | 0.021997886 |
| RBKS | 0.053658213 | -0.037652571 | 0.016005642 |
| RPL26L1 | 0.077834037 | -0.048982548 | 0.028851489 |
| SYDE1 | 0.050371326 | -0.042418488 | 0.007952838 |
| THAP2 | 0.052514549 | -0.041510239 | 0.01100431 |
| FAM131A | 0.044909296 | -0.041783612 | 0.003125683 |
| YRDC | 0.047066066 | -0.037414946 | 0.00965112 |
| RAD54B | 0.031790778 | -0.034373816 | -0.002583038 |
| LMO4 | 0.067666973 | -0.069515823 | -0.001848849 |
| HSD17B4 | 0.072867304 | -0.097744423 | -0.024877119 |
| SHC2 | 0.035952248 | -0.021341548 | 0.014610701 |
| ANKRD33B | -0.062881045 | 0.032148091 | -0.030732954 |
| PBDC1 | 0.066431651 | -0.058403661 | 0.008027989 |
| CDKN2AIPNL | 0.067890902 | -0.075512303 | -0.007621401 |
| PACC1 | 0.029320497 | -0.022785271 | 0.006535227 |
| H2AFV | 0.102203445 | -0.11247631 | -0.010272865 |
| SUPT4H1 | 0.085769073 | -0.041409579 | 0.044359494 |
| TRABD | 0.065592172 | -0.066574552 | -0.000982379 |
| ORAI3 | 0.049333978 | -0.04968906 | -0.000355082 |
| MAPRE1 | 0.06905799 | -0.047379079 | 0.021678912 |
| PJA2 | 0.100755961 | -0.120942055 | -0.020186094 |
| AHDC1 | 0.044776489 | -0.0430062 | 0.001770289 |
| PPCS | 0.08640275 | -0.052762965 | 0.033639785 |
| DDX27 | 0.064673187 | -0.068603339 | -0.003930151 |
| DSG2 | -0.074322039 | 0.045125644 | -0.029196395 |
| HPS4 | 0.070892001 | -0.111787163 | -0.040895162 |
| AC006460.1 | 0.023700284 | -0.023700284 | -2.30926E-14 |
| WNT11 | 0.042458558 | -0.050659689 | -0.008201131 |
| TOR3A | 0.052594802 | -0.053721123 | -0.001126321 |
| NUCB2 | 0.067753923 | -0.070547869 | -0.002793946 |
| CCDC127 | 0.059059346 | -0.063848964 | -0.004789618 |
| MLEC | 0.093953733 | -0.13990516 | -0.045951427 |
| SMU1 | 0.07036787 | -0.072444599 | -0.002076729 |
| EIF3D | 0.101988048 | -0.061858657 | 0.040129391 |

| | | | |
|---|---|---|---|
| FAM98A | 0.071650495 | -0.093981587 | -0.022331092 |
| TLE1 | -0.057265017 | 0.065812072 | 0.008547055 |
| TENT5C | -0.043259535 | 0.033246421 | -0.010013115 |
| PTP4A2 | 0.098267542 | -0.122528588 | -0.024261045 |
| IFI27L1 | 0.046652476 | -0.052897633 | -0.006245156 |
| CCDC28B | 0.045051347 | -0.041508568 | 0.003542779 |
| LIN7C | 0.073335802 | -0.050808789 | 0.022527012 |
| AP1S2 | 0.044666762 | -0.03974417 | 0.004922592 |
| ANAPC16 | 0.127062161 | -0.083742323 | 0.043319838 |
| SNRNP27 | 0.068014397 | -0.070665545 | -0.002651148 |
| MRPL18 | 0.085088048 | -0.049642705 | 0.035445343 |
| MTG1 | 0.067450418 | -0.039496567 | 0.02795385 |
| PDIA6 | 0.117610833 | -0.189833895 | -0.072223062 |
| MRPL3 | 0.084221548 | -0.042704468 | 0.04151708 |
| DTNA | 0.027057258 | -0.024654875 | 0.002402383 |
| IGSF9 | -0.040188237 | 0.031945425 | -0.008242812 |
| KNL1 | 0.029081052 | -0.032858731 | -0.003777679 |
| EFNA4 | 0.03728554 | -0.029298798 | 0.007986742 |
| YTHDF2 | 0.075727577 | -0.089941707 | -0.014214131 |
| CCDC8 | -0.043282328 | 0.028102357 | -0.015179971 |
| ADGRE5 | 0.032451433 | -0.032434494 | 1.69388E-05 |
| ZMYND19 | 0.05243086 | -0.050433443 | 0.001997417 |
| PRPF18 | 0.058231871 | -0.056977598 | 0.001254273 |
| APOO | 0.056187771 | -0.037800757 | 0.018387014 |
| CSK | 0.041374771 | -0.032581836 | 0.008792935 |
| NDRG1 | -0.06766396 | 0.064703833 | -0.002960127 |
| UNC13C | 0.05455629 | -0.06717972 | -0.01262343 |
| TMEM267 | 0.062834974 | -0.056987746 | 0.005847228 |
| NECTIN2 | 0.094018427 | -0.114515197 | -0.020496771 |
| PIP4P1 | 0.057066538 | -0.045680867 | 0.011385671 |
| HMOX2 | 0.060289656 | -0.050467535 | 0.009822121 |
| DYNC1LI1 | 0.057717547 | -0.02801348 | 0.029704067 |
| FRGCA | 0.024495149 | -0.021562114 | 0.002933035 |
| EFNB2 | -0.045087723 | 0.034324901 | -0.010762822 |
| DHX29 | 0.082790176 | -0.054893983 | 0.027896194 |
| JUNB | -0.126591653 | 0.081547287 | -0.045044365 |
| IL1R2 | 0.029610005 | -0.023181681 | 0.006428324 |
| EI24 | 0.090881528 | -0.086911858 | 0.003969671 |
| CDA | 0.037439255 | -0.03921887 | -0.001779615 |
| CCNY | 0.069864584 | -0.072787219 | -0.002922636 |
| NSMCE4A | 0.060288882 | -0.047908104 | 0.012380778 |
| SPAG1 | 0.058627563 | -0.069008352 | -0.010380789 |
| HSF2 | 0.044669581 | -0.037912581 | 0.006757 |
| MIF4GD | 0.057230814 | -0.055741522 | 0.001489292 |
| TMEM141 | 0.1094398 | -0.058251179 | 0.051188621 |
| SLC25A26 | 0.056261885 | -0.050471844 | 0.005790042 |
| ERICH5 | 0.030277227 | -0.016252072 | 0.014025155 |

| | | | |
|---|---|---|---|
| RRP1 | 0.043373248 | -0.042506961 | 0.000866287 |
| BRD7 | 0.088049011 | -0.085182204 | 0.002866807 |
| UBE2D1 | 0.05401695 | -0.043083296 | 0.010933654 |
| USF2 | 0.075965635 | -0.083071221 | -0.007105585 |
| FURIN | 0.064676561 | -0.102509661 | -0.0378331 |
| VPS26B | 0.054549003 | -0.056800584 | -0.002251581 |
| IMMT | 0.075284597 | -0.089749456 | -0.014464859 |
| CYHR1 | 0.088433498 | -0.081739556 | 0.006693942 |
| SELENOS | 0.09017479 | -0.119614517 | -0.029439727 |
| HDLBP | 0.095704212 | -0.051644659 | 0.044059553 |
| CYP27A1 | -0.037527079 | 0.028326333 | -0.009200746 |
| SOSTDC1 | 0.060161694 | -0.071918124 | -0.01175643 |
| ABHD3 | 0.042343593 | -0.042370866 | -2.72732E-05 |
| CCDC149 | 0.051652353 | -0.054343285 | -0.002690932 |
| IMPACT | 0.053552001 | -0.036912558 | 0.016639443 |
| ZNF281 | 0.05271455 | -0.039854173 | 0.012860378 |
| PDCD10 | 0.083787147 | -0.059585775 | 0.024201372 |
| PPP4R4 | 0.023730747 | -0.023730747 | -2.13163E-14 |
| CGRRF1 | 0.054581215 | -0.048184846 | 0.006396369 |
| MRPS5 | 0.077925015 | -0.077785836 | 0.00013918 |
| THOC3 | 0.046335029 | -0.025210017 | 0.021125011 |
| HSD17B7 | 0.047331339 | -0.051423181 | -0.004091843 |
| WBP1 | 0.084996607 | -0.07888789 | 0.006108717 |
| C6orf120 | 0.053184626 | -0.049531012 | 0.003653614 |
| LRATD1 | -0.046408819 | 0.040283081 | -0.006125738 |
| SNRNP40 | 0.064166487 | -0.039769423 | 0.024397064 |
| TBRG1 | 0.064625835 | -0.040970501 | 0.023655334 |
| LARP4 | 0.067214112 | -0.089450231 | -0.022236119 |
| GPBAR1 | 0.026103246 | -0.025481864 | 0.000621382 |
| SLC35A3 | 0.067267181 | -0.072507204 | -0.005240023 |
| MEST | -0.040673467 | 0.038884893 | -0.001788574 |
| RMND5B | 0.051254167 | -0.037079729 | 0.014174437 |
| HSD17B12 | 0.087266619 | -0.151965771 | -0.064699152 |
| TAF6L | 0.044963264 | -0.032186105 | 0.012777159 |
| GMPR | 0.085803453 | -0.113254682 | -0.027451229 |
| PRSS3 | 0.022859118 | -0.022859118 | -1.95399E-14 |
| MAP2K3 | 0.052962128 | -0.045611643 | 0.007350485 |
| MDP1 | 0.062043171 | -0.048423244 | 0.013619927 |
| SEC22C | 0.054685948 | -0.047052186 | 0.007633762 |
| CRAT | 0.049465074 | -0.025368103 | 0.024096972 |
| SNW1 | 0.071267197 | -0.037756051 | 0.033511146 |
| PCGF5 | 0.067331281 | -0.056472809 | 0.010858472 |
| FBXO27 | 0.031153943 | -0.034401554 | -0.003247611 |
| SAFB | 0.073941173 | -0.072751184 | 0.001189989 |
| COPB2 | 0.075597388 | -0.041613456 | 0.033983932 |
| SPRYD7 | 0.059064838 | -0.06059037 | -0.001525531 |
| CCDC126 | 0.045880766 | -0.048068594 | -0.002187828 |

| | | | |
|---|---|---|---|
| GMPS | 0.06017277 | -0.072313952 | -0.012141183 |
| BLOC1S5 | 0.076802295 | -0.077188703 | -0.000386409 |
| ENDOG | 0.063140064 | -0.045159961 | 0.017980103 |
| SIAE | 0.054568566 | -0.07227348 | -0.017704914 |
| CBL | 0.043328086 | -0.027296914 | 0.016031172 |
| TM9SF3 | 0.108571886 | -0.200411042 | -0.091839156 |
| EEA1 | 0.068405955 | -0.086999016 | -0.01859306 |
| UBE2B | 0.102694763 | -0.084760848 | 0.017933915 |
| PA2G4 | 0.095296675 | -0.074609219 | 0.020687455 |
| TIMM50 | 0.045416144 | -0.030425812 | 0.014990333 |
| RSBN1 | 0.060879275 | -0.057763016 | 0.00311626 |
| RRP15 | 0.063745902 | -0.058250377 | 0.005495525 |
| MRPS9 | 0.06445903 | -0.057710283 | 0.006748747 |
| HIBADH | 0.071657489 | -0.03563978 | 0.036017709 |
| MAFG | 0.05176469 | -0.072057236 | -0.020292546 |
| TRADD | 0.051164507 | -0.028060774 | 0.023103733 |
| CDKL2 | 0.102927054 | -0.175786277 | -0.072859224 |
| ZNF667-AS1 | 0.077059247 | -0.059006719 | 0.018052528 |
| TMED1 | 0.077337356 | -0.087042792 | -0.009705435 |
| TSN | 0.070405862 | -0.068341572 | 0.00206429 |
| CALU | 0.06838932 | -0.083158303 | -0.014768984 |
| DDX46 | 0.092038052 | -0.149931555 | -0.057893502 |
| ARL2BP | 0.06963087 | -0.04300232 | 0.02662855 |
| NIPSNAP2 | 0.096225958 | -0.081066286 | 0.015159673 |
| C8orf82 | 0.068942953 | -0.034420842 | 0.034522111 |
| BMP4 | -0.037487513 | 0.031666717 | -0.005820797 |
| USP25 | 0.069502433 | -0.086351662 | -0.016849229 |
| EXOSC4 | 0.054317141 | -0.040563753 | 0.013753388 |
| ZYX | 0.058819755 | -0.035636928 | 0.023182827 |
| SERTAD4-AS1 | 0.072759964 | -0.063178251 | 0.009581713 |
| MACO1 | 0.064253509 | -0.056962336 | 0.007291172 |
| BGN | -0.052385288 | 0.038579209 | -0.013806079 |
| CCDC102A | 0.040051686 | -0.031957517 | 0.008094169 |
| SELENOF | 0.105873249 | -0.094839001 | 0.011034248 |
| FOXO3 | -0.060623975 | 0.036505637 | -0.024118339 |
| EGR1 | -0.056468636 | 0.044188314 | -0.012280322 |
| CLCN7 | 0.067256745 | -0.098153835 | -0.030897091 |
| DNMT1 | 0.055214779 | -0.050689614 | 0.004525165 |
| RNPS1 | 0.084967233 | -0.058208476 | 0.026758757 |
| RNF11 | 0.067659544 | -0.066041415 | 0.001618129 |
| CRIP2 | 0.102090628 | -0.087831085 | 0.014259543 |
| GEMIN8 | 0.035649596 | -0.025984798 | 0.009664798 |
| PSMF1 | 0.069137722 | -0.042051237 | 0.027086485 |
| MYO3B | 0.037034188 | -0.036098935 | 0.000935253 |
| Z98259.3 | 0.022375326 | -0.022375326 | -2.13163E-14 |
| GNG11 | -0.105032246 | 0.128740197 | 0.023707951 |
| APPL1 | 0.085376442 | -0.076068341 | 0.009308101 |

| | | | |
|---|---|---|---|
| TRMU | 0.047887929 | -0.033001457 | 0.014886471 |
| IFRD2 | 0.058864651 | -0.052828579 | 0.006036072 |
| RASL11B | 0.026741621 | -0.029954894 | -0.003213273 |
| BEX1 | -0.055693215 | 0.067867571 | 0.012174356 |
| STRADB | 0.045375012 | -0.03166818 | 0.013706832 |
| C6orf226 | 0.051842968 | -0.047848259 | 0.003994709 |
| NMNAT3 | 0.053946916 | -0.054450095 | -0.000503179 |
| PYROXD2 | 0.043639042 | -0.049657352 | -0.00601831 |
| HOXC10 | 0.084806677 | -0.091049011 | -0.006242334 |
| NSMCE3 | 0.060531523 | -0.063400724 | -0.002869201 |
| REL | 0.050538957 | -0.072804841 | -0.022265884 |
| TINF2 | 0.052331443 | -0.052227358 | 0.000104084 |
| GOLGB1 | 0.10631396 | -0.155020165 | -0.048706205 |
| ZNF69 | 0.049719867 | -0.053561592 | -0.003841725 |
| RAB27A | 0.047380628 | -0.065379915 | -0.017999287 |
| COL6A1 | -0.052892225 | 0.036298196 | -0.01659403 |
| SLC16A12 | 0.087063567 | -0.171459425 | -0.084395858 |
| RAB8A | 0.064870691 | -0.042135997 | 0.022734694 |
| TMEM80 | 0.074662956 | -0.087278833 | -0.012615877 |
| SLC25A1 | 0.066933073 | -0.04966308 | 0.017269993 |
| SAYSD1 | 0.05610084 | -0.055828063 | 0.000272777 |
| DTNBP1 | 0.051358094 | -0.049431517 | 0.001926576 |
| ARHGEF7 | 0.05497862 | -0.058980691 | -0.004002071 |
| CENPV | -0.052005366 | 0.082239523 | 0.030234158 |
| TRNT1 | 0.059648842 | -0.066800484 | -0.007151642 |
| HOXB-AS1 | 0.050902394 | -0.070704837 | -0.019802443 |
| LRRN2 | 0.031887568 | -0.026329329 | 0.005558238 |
| CFAP97 | 0.08824548 | -0.074184848 | 0.014060631 |
| RECQL5 | 0.063712245 | -0.102974992 | -0.039262747 |
| GNPDA1 | 0.051227132 | -0.044672673 | 0.006554459 |
| MMAB | 0.07427251 | -0.07268012 | 0.00159239 |
| FBXO22 | 0.064445852 | -0.074391242 | -0.009945389 |
| FST | 0.055578399 | -0.075736392 | -0.020157994 |
| RABEP2 | 0.043875435 | -0.036115598 | 0.007759837 |
| TMEM35B | 0.101376768 | -0.167489949 | -0.066113181 |
| TMED4 | 0.10544346 | -0.1199469 | -0.01450344 |
| BCAS1 | 0.083192317 | -0.18281908 | -0.099626763 |
| CDK5RAP3 | 0.073982875 | -0.087362674 | -0.013379798 |
| CYP39A1 | 0.030134834 | -0.027840644 | 0.00229419 |
| ZMAT2 | 0.075286634 | -0.076638507 | -0.001351873 |
| RCC1 | 0.033257381 | -0.036657701 | -0.00340032 |
| SPG21 | 0.066451886 | -0.059123585 | 0.007328301 |
| SPTAN1 | 0.094444632 | -0.138802566 | -0.044357934 |
| C8orf88 | 0.023969849 | -0.017514982 | 0.006454868 |
| AP1G2 | 0.051133896 | -0.06019234 | -0.009058444 |
| TWF2 | 0.057518796 | -0.041508988 | 0.016009807 |
| PDK2 | 0.060401724 | -0.047575438 | 0.012826286 |

| | | | |
|---|---|---|---|
| DHX15 | 0.075980719 | -0.082715771 | -0.006735053 |
| QTRT1 | 0.071845046 | -0.085197937 | -0.013352892 |
| CNBP | 0.105938038 | -0.130600381 | -0.024662343 |
| SLC6A8 | -0.056134596 | 0.029862986 | -0.026271609 |
| TRIP10 | 0.063609997 | -0.061315547 | 0.002294449 |
| AC110619.1 | 0.032243842 | -0.031548528 | 0.000695314 |
| SLC66A3 | 0.059941963 | -0.04516855 | 0.014773413 |
| CHMP4A | 0.080364298 | -0.059971899 | 0.020392399 |
| RALB | 0.054920289 | -0.046062168 | 0.008858121 |
| RASGRF2 | 0.030979167 | -0.036753226 | -0.005774059 |
| AKAP12 | 0.090262879 | -0.145784739 | -0.055521859 |
| KLF7 | -0.03977657 | 0.021082077 | -0.018694493 |
| MIA3 | 0.078868048 | -0.118706286 | -0.039838237 |
| TIMM17A | 0.052633459 | -0.059136367 | -0.006502909 |
| RPUSD2 | 0.039903831 | -0.031538981 | 0.008364851 |
| ORMDL2 | 0.057006008 | -0.03613737 | 0.020868638 |
| MLLT11 | 0.025796395 | -0.025114851 | 0.000681543 |
| RNF152 | 0.031921215 | -0.023467669 | 0.008453546 |
| HSD17B8 | 0.059320533 | -0.053716129 | 0.005604404 |
| CMTM6 | 0.073793137 | -0.056393958 | 0.017399179 |
| CSNK1A1 | 0.095188548 | -0.104706252 | -0.009517704 |
| ADAMTS15 | -0.036266992 | 0.023072806 | -0.013194186 |
| TIPRL | 0.063574748 | -0.040108976 | 0.023465772 |
| ADSL | 0.052249776 | -0.0333712 | 0.018878576 |
| CLIP2 | 0.039370568 | -0.047038981 | -0.007668413 |
| PELI2 | -0.037303104 | 0.026394241 | -0.010908864 |
| TSFM | 0.063009803 | -0.036009282 | 0.027000521 |
| TTC1 | 0.074991669 | -0.063046434 | 0.011945235 |
| KL | -0.05149702 | 0.055328917 | 0.003831897 |
| RBBP7 | 0.07947358 | -0.084799054 | -0.005325474 |
| PARP1 | 0.0599741 | -0.077817044 | -0.017842944 |
| CD74 | 0.071677526 | -0.048478988 | 0.023198538 |
| AL353152.1 | 0.023867908 | -0.023584879 | 0.000283029 |
| TIMP3 | 0.070702627 | -0.079047481 | -0.008344854 |
| AC097448.1 | 0.033575108 | -0.031888501 | 0.001686608 |
| KPNA4 | 0.071217801 | -0.079317628 | -0.008099827 |
| TTC33 | 0.052792546 | -0.04122446 | 0.011568086 |
| ANKRD46 | 0.048296894 | -0.039930445 | 0.008366449 |
| RMDN1 | 0.065566279 | -0.060982578 | 0.004583701 |
| HOXC-AS1 | 0.052814404 | -0.062035845 | -0.009221441 |
| SERGEF | 0.050607513 | -0.037995432 | 0.01261208 |
| TMEM139 | 0.091299394 | -0.066949772 | 0.024349622 |
| TMEM179B | 0.095388886 | -0.09558337 | -0.000194484 |
| TP53TG1 | 0.062593276 | -0.073982485 | -0.011389209 |
| EIF2B4 | 0.042240389 | -0.036543068 | 0.005697321 |
| RAPGEF3 | 0.091655992 | -0.116632244 | -0.024976253 |
| ZNF844 | -0.041359812 | 0.023453164 | -0.017906648 |

| | | | |
|---|---|---|---|
| ISCA1 | 0.072553016 | -0.050527571 | 0.022025446 |
| SMC3 | 0.083988089 | -0.093778978 | -0.009790889 |
| CCSER2 | 0.062235081 | -0.069945138 | -0.007710057 |
| RB1 | 0.065738498 | -0.097561842 | -0.031823343 |
| GCC2 | 0.099569366 | -0.190692595 | -0.091123229 |
| FAHD2A | 0.050673985 | -0.063212955 | -0.012538969 |
| UNC119 | 0.054244765 | -0.040140667 | 0.014104098 |
| ACYP2 | 0.04962272 | -0.047584303 | 0.002038417 |
| PELI1 | 0.051364925 | -0.075633677 | -0.024268752 |
| PNPLA2 | 0.087301659 | -0.112560722 | -0.025259064 |
| AP4E1 | 0.034126261 | -0.035557205 | -0.001430944 |
| FOXP1 | -0.066051533 | 0.041100535 | -0.024950999 |
| UTP11 | 0.047899034 | -0.046511676 | 0.001387359 |
| MFSD11 | 0.044664177 | -0.044609009 | 5.51687E-05 |
| SAP30L | 0.053564103 | -0.065312887 | -0.011748784 |
| SNHG18 | 0.057999163 | -0.031290978 | 0.026708186 |
| MPZL1 | 0.077412483 | -0.146485128 | -0.069072646 |
| CMTM8 | 0.033290491 | -0.029636258 | 0.003654233 |
| MANBAL | 0.071349911 | -0.063527058 | 0.007822853 |
| ETNK2 | 0.039119964 | -0.021816584 | 0.01730338 |
| POC1B | 0.029910303 | -0.020881939 | 0.009028364 |
| TPD52L2 | 0.059009665 | -0.032882425 | 0.02612724 |
| CKLF | 0.059423945 | -0.06758874 | -0.008164795 |
| NSD2 | 0.045545508 | -0.061010219 | -0.015464711 |
| TFG | 0.080142418 | -0.074165977 | 0.005976441 |
| CASTOR2 | 0.033718094 | -0.041254452 | -0.007536358 |
| TCEAL9 | 0.099009863 | -0.112832494 | -0.013822631 |
| EXOC6 | 0.0390215 | -0.039197548 | -0.000176048 |
| PIGZ | 0.019048003 | -0.019048003 | -1.95399E-14 |
| CCDC124 | 0.068985973 | -0.05609769 | 0.012888283 |
| PCSK5 | 0.021967165 | -0.020179515 | 0.00178765 |
| GFOD2 | 0.042176243 | -0.028272798 | 0.013903444 |
| B4GALT7 | 0.055537724 | -0.067228482 | -0.011690759 |
| DENND10 | 0.062237391 | -0.042281773 | 0.019955618 |
| RASA3 | -0.028885187 | 0.019366617 | -0.00951857 |
| UCHL5 | 0.059994608 | -0.060880085 | -0.000885478 |
| PLCB4 | 0.037369458 | -0.04165514 | -0.004285683 |
| COL5A1 | -0.033238228 | 0.021593259 | -0.011644969 |
| CTBS | 0.065158216 | -0.091434416 | -0.026276201 |
| TOR1AIP2 | 0.082356511 | -0.120575081 | -0.03821857 |
| MYLK | 0.048040884 | -0.052544297 | -0.004503413 |
| LRP10 | 0.088293058 | -0.144040556 | -0.055747499 |
| C15orf40 | 0.064362916 | -0.073860888 | -0.009497972 |
| PGAP2 | 0.049084615 | -0.063672696 | -0.014588081 |
| USP34 | 0.08977338 | -0.172421876 | -0.082648496 |
| NDUFAF8 | 0.084944661 | -0.046774356 | 0.038170305 |
| SEMA4B | 0.051090311 | -0.060734762 | -0.009644451 |

| | | | |
|---|---|---|---|
| ATE1 | 0.058585988 | -0.073511665 | -0.014925677 |
| HARS | 0.050824867 | -0.033230629 | 0.017594239 |
| TMEM265 | 0.069292088 | -0.048764526 | 0.020527562 |
| IFT20 | 0.069811757 | -0.065211839 | 0.004599918 |
| LTA4H | 0.060546553 | -0.048914034 | 0.011632518 |
| LANCL1 | 0.054205092 | -0.039970278 | 0.014234814 |
| TRIQK | 0.058507332 | -0.053771832 | 0.0047355 |
| SRP72 | 0.066779242 | -0.037027646 | 0.029751595 |
| LAMTOR3 | 0.070752589 | -0.046283234 | 0.024469355 |
| BTBD1 | 0.044944144 | -0.056836018 | -0.011891874 |
| CDR2L | 0.025861871 | -0.020037403 | 0.005824468 |
| TIMM22 | 0.049228404 | -0.062549044 | -0.01332064 |
| AC105942.1 | 0.029301607 | -0.027650124 | 0.001651483 |
| TMCO3 | 0.077429457 | -0.140016266 | -0.062586809 |
| KDM2A | 0.065680518 | -0.086954417 | -0.0212739 |
| FKBP3 | 0.079559446 | -0.058564223 | 0.020995223 |
| RAB28 | 0.050852766 | -0.05086437 | -1.16039E-05 |
| AGTRAP | 0.062916766 | -0.045052538 | 0.017864228 |
| ACTR1A | 0.053154332 | -0.055323729 | -0.002169397 |
| AP3S1 | 0.05150909 | -0.027658642 | 0.023850448 |
| USP2 | 0.052695772 | -0.054013551 | -0.001317779 |
| NRGN | 0.03197047 | -0.034181826 | -0.002211356 |
| CASP7 | 0.04477591 | -0.027791655 | 0.016984255 |
| PEX13 | 0.048644058 | -0.047296068 | 0.00134799 |
| REPIN1 | 0.06390753 | -0.041723809 | 0.022183721 |
| STXBP3 | 0.077439245 | -0.084726467 | -0.007287222 |
| CUL3 | 0.078492451 | -0.099336905 | -0.020844454 |
| ZNF277 | 0.051230165 | -0.042912084 | 0.008318081 |
| ABHD14A | 0.071060909 | -0.099366103 | -0.028305194 |
| KPTN | 0.029302352 | -0.029522981 | -0.000220629 |
| FAHD2B | 0.038984296 | -0.037947987 | 0.001036309 |
| C8orf58 | 0.027492928 | -0.026566349 | 0.000926579 |
| HDHD2 | 0.048835742 | -0.037769679 | 0.011066063 |
| KCTD17 | 0.027843962 | -0.021866078 | 0.005977884 |
| GABRP | 0.024515759 | -0.019421615 | 0.005094145 |
| DNAJC17 | 0.048573249 | -0.033101085 | 0.015472164 |
| LY75 | 0.024884523 | -0.021329041 | 0.003555482 |
| SF3A3 | 0.058668757 | -0.036333114 | 0.022335643 |
| AKIRIN1 | 0.078231797 | -0.116745926 | -0.038514129 |
| PIK3CD-AS2 | 0.030708794 | -0.026400326 | 0.004308468 |
| PDE4A | 0.028564325 | -0.018767634 | 0.009796691 |
| KLHL29 | 0.036229413 | -0.043842145 | -0.007612732 |
| MORC3 | 0.055865249 | -0.058357756 | -0.002492507 |
| DEF8 | 0.05166153 | -0.056021274 | -0.004359744 |
| TUSC1 | 0.065576157 | -0.066033798 | -0.000457641 |
| SEMA3F | -0.051605118 | 0.027919674 | -0.023685445 |
| PNRC1 | 0.106708668 | -0.113024643 | -0.006315975 |

| | | | |
|---|---|---|---|
| TAP1 | 0.045727824 | -0.037463168 | 0.008264656 |
| RTKN | 0.047433547 | -0.027286669 | 0.020146877 |
| MED11 | 0.060195278 | -0.039377563 | 0.020817715 |
| CD320 | 0.065818724 | -0.057201925 | 0.008616799 |
| CDC42SE1 | 0.074537472 | -0.095005254 | -0.020467782 |
| C2CD2L | 0.031619724 | -0.034312039 | -0.002692315 |
| IFT74 | 0.071227019 | -0.095257906 | -0.024030887 |
| MTDH | 0.098575497 | -0.156079621 | -0.057504124 |
| LARS | 0.081174746 | -0.086212571 | -0.005037825 |
| TM2D3 | 0.073096521 | -0.094457074 | -0.021360553 |
| MCFD2 | 0.070855502 | -0.080160372 | -0.00930487 |
| LZIC | 0.060588471 | -0.060077411 | 0.000511061 |
| QARS | 0.058182473 | -0.042082261 | 0.016100212 |
| LLPH | 0.053494016 | -0.050482951 | 0.003011065 |
| HBEGF | 0.035902879 | -0.0381249 | -0.002222021 |
| MT-ND6 | -0.087899844 | 0.145820364 | 0.057920521 |
| SURF4 | 0.065959805 | -0.071776342 | -0.005816536 |
| ANAPC5 | 0.073475842 | -0.09900956 | -0.025533718 |
| MRPS22 | 0.054792475 | -0.039456435 | 0.01533604 |
| IFI27L2 | 0.091306662 | -0.060226951 | 0.031079711 |
| PLXNA4 | 0.04666312 | -0.047840088 | -0.001176968 |
| SETD9 | 0.037618089 | -0.025733312 | 0.011884777 |
| ANTKMT | 0.077375061 | -0.0584229 | 0.018952161 |
| STK38 | 0.057821964 | -0.068774447 | -0.010952483 |
| MRM3 | 0.033744431 | -0.022985136 | 0.010759295 |
| CLSTN1 | 0.10425231 | -0.236598221 | -0.132345912 |
| MAZ | 0.081609318 | -0.066619958 | 0.01498936 |
| N4BP3 | 0.022734255 | -0.023625231 | -0.000890975 |
| TMEM25 | 0.052439103 | -0.054834083 | -0.002394979 |
| SREK1IP1 | 0.067627295 | -0.056197048 | 0.011430248 |
| BSCL2 | 0.083667561 | -0.079057107 | 0.004610454 |
| TM9SF2 | 0.092464929 | -0.138381515 | -0.045916586 |
| HOXD10 | 0.050716233 | -0.03342597 | 0.017290264 |
| NSD3 | 0.091637947 | -0.152400537 | -0.06076259 |
| ZMIZ1 | -0.067503615 | 0.036369052 | -0.031134563 |
| FAM204A | 0.075897201 | -0.089084248 | -0.013187047 |
| SNX1 | 0.063278612 | -0.048329909 | 0.014948703 |
| HAGLROS | 0.023908198 | -0.024231248 | -0.00032305 |
| VPS25 | 0.070329073 | -0.048281697 | 0.022047376 |
| PPM1D | 0.031890231 | -0.029795275 | 0.002094956 |
| UTRN | -0.106121555 | -0.093271489 | -0.199393044 |
| PPL | 0.066405027 | -0.121126312 | -0.054721285 |
| NRBF2 | 0.040606911 | -0.031879156 | 0.008727755 |
| SHKBP1 | 0.039008292 | -0.030201066 | 0.008807226 |
| TTC32 | 0.052358647 | -0.067186332 | -0.014827686 |
| PDE4B | -0.034772322 | 0.038805061 | 0.004032738 |
| HEXIM2 | 0.044117384 | -0.038628197 | 0.005489187 |

| | | | |
|---|---|---|---|
| FAM199X | 0.065327993 | -0.085020402 | -0.019692408 |
| AC108749.1 | 0.020952616 | -0.019211734 | 0.001740882 |
| RASGRP3 | 0.0335636 | -0.033229312 | 0.000334288 |
| CCL28 | 0.057263876 | -0.067116083 | -0.009852207 |
| SLC22A17 | -0.059718658 | 0.068239846 | 0.008521188 |
| NEDD4 | 0.030819204 | -0.022196933 | 0.008622271 |
| RTN2 | 0.02392397 | -0.022546772 | 0.001377199 |
| LINC01948 | 0.044817321 | -0.068561705 | -0.023744385 |
| CDC40 | 0.06274233 | -0.069857653 | -0.007115324 |
| MRPL46 | 0.053173596 | -0.043521615 | 0.009651982 |
| MTIF3 | 0.082996973 | -0.044217868 | 0.038779105 |
| RECQL | 0.05234624 | -0.055622665 | -0.003276425 |
| DUT | 0.095916615 | -0.096656197 | -0.000739582 |
| INSIG2 | 0.054991307 | -0.061677361 | -0.006686054 |
| RAPGEF1 | 0.047217113 | -0.044387006 | 0.002830107 |
| CBWD5 | 0.054640558 | -0.069622719 | -0.01498216 |
| VPS4A | 0.067336216 | -0.058321197 | 0.009015019 |
| KIAA0040 | 0.050840174 | -0.0797134 | -0.028873226 |
| GALNT6 | 0.035056328 | -0.040002699 | -0.004946371 |
| TXNDC15 | 0.057236571 | -0.075736756 | -0.018500185 |
| DHRS1 | 0.043792572 | -0.039142443 | 0.004650129 |
| ARHGAP12 | 0.053904034 | -0.053215006 | 0.000689028 |
| ACBD3 | 0.061580063 | -0.064675752 | -0.003095688 |
| LYRM9 | 0.07265121 | -0.066279382 | 0.006371827 |
| TPD52 | 0.072926718 | -0.080371069 | -0.007444351 |
| HOXD3 | 0.034363093 | -0.032078688 | 0.002284405 |
| AP1AR | 0.051764784 | -0.048397471 | 0.003367313 |
| STX10 | 0.06057209 | -0.060547208 | 2.48825E-05 |
| MRPS18B | 0.082287285 | -0.091926314 | -0.009639028 |
| CNIH1 | 0.056699948 | -0.034545939 | 0.022154009 |
| PIK3R3 | 0.027675802 | -0.029867318 | -0.002191516 |
| CYTOR | -0.042666736 | 0.023820891 | -0.018845846 |
| PPT2 | 0.025993915 | -0.02508085 | 0.000913065 |
| BLNK | -0.032705574 | 0.021278528 | -0.011427046 |
| PCBP4 | 0.040658831 | -0.029801254 | 0.010857577 |
| CHM | 0.057515291 | -0.049339672 | 0.00817562 |
| FXR1 | 0.080059615 | -0.106069382 | -0.026009767 |
| GABRA2 | -0.064994019 | 0.042968307 | -0.022025711 |
| ASPHD2 | 0.041671596 | -0.054141659 | -0.012470063 |
| MRPL58 | 0.05105058 | -0.036764233 | 0.014286347 |
| DEXI | 0.054044606 | -0.031893488 | 0.022151117 |
| CNOT2 | 0.067188169 | -0.058755538 | 0.008432631 |
| AP3S2 | 0.041826814 | -0.035052516 | 0.006774298 |
| WBP2 | 0.076595364 | -0.066281184 | 0.01031418 |
| TSPAN31 | 0.068681486 | -0.063619681 | 0.005061806 |
| SF3B4 | 0.063107061 | -0.066136613 | -0.003029552 |
| C2orf72 | 0.037930044 | -0.036165316 | 0.001764729 |

| | | | |
|---|---|---|---|
| FAM107A | 0.034304521 | -0.03043962 | 0.003864902 |
| KRAS | 0.062258842 | -0.070499191 | -0.008240348 |
| OSBPL3 | 0.051480532 | -0.066578795 | -0.015098263 |
| ATG4D | 0.042935813 | -0.043763821 | -0.000828008 |
| CSDE1 | 0.103035493 | -0.124297264 | -0.021261771 |
| ZNF143 | 0.031663324 | -0.02087965 | 0.010783674 |
| HTATIP2 | 0.047761007 | -0.036732165 | 0.011028842 |
| UBXN2A | 0.04726937 | -0.048622602 | -0.001353232 |
| CHIC2 | 0.056459244 | -0.050104885 | 0.00635436 |
| PROM2 | 0.046677679 | -0.057731673 | -0.011053994 |
| SMAP2 | 0.040055543 | -0.04304685 | -0.002991307 |
| ZFYVE19 | 0.03697512 | -0.02474302 | 0.0122321 |
| SSH2 | 0.052861502 | -0.058042478 | -0.005180975 |
| BOC | 0.055776641 | -0.0974292 | -0.041652559 |
| PAF1 | 0.052934327 | -0.053619794 | -0.000685467 |
| PCMTD2 | 0.068886359 | -0.10894389 | -0.040057531 |
| LCOR | 0.070158501 | -0.12553211 | -0.055373609 |
| SKIL | 0.074452789 | -0.097748583 | -0.023295793 |
| RSRC2 | 0.090129614 | -0.132250973 | -0.042121359 |
| TRIB1 | -0.031930706 | 0.024597612 | -0.007333095 |
| ADCY5 | 0.028754993 | -0.021635388 | 0.007119605 |
| CCDC74A | 0.044666299 | -0.03682359 | 0.00784271 |
| PPP4R1 | 0.055815527 | -0.050315411 | 0.005500115 |
| OSER1 | 0.058727878 | -0.033211673 | 0.025516205 |
| NASP | 0.073522501 | -0.077593924 | -0.004071423 |
| SPEF2 | -0.041119431 | 0.022895974 | -0.018223458 |
| WASF2 | 0.087416903 | -0.094577254 | -0.007160351 |
| LGALS3 | 0.102631143 | -0.123557345 | -0.020926202 |
| TIGAR | 0.02453153 | -0.024687298 | -0.000155768 |
| PPFIA1 | 0.063144512 | -0.049733513 | 0.013410999 |
| SMARCA5 | 0.076004166 | -0.069779461 | 0.006224705 |
| CELF2 | 0.022745143 | -0.023657331 | -0.000912188 |
| DENND1B | 0.053722994 | -0.071219398 | -0.017496404 |
| LHPP | 0.03471426 | -0.027756027 | 0.006958233 |
| GLB1 | 0.057369154 | -0.042720907 | 0.014648247 |
| MIR100HG | 0.029271689 | -0.034145674 | -0.004873986 |
| AHI1 | 0.071005183 | -0.116356712 | -0.045351529 |
| TMEM41B | 0.063873056 | -0.064707532 | -0.000834476 |
| BCAP29 | 0.056236118 | -0.065263032 | -0.009026914 |
| EDNRB | 0.024170205 | -0.025585557 | -0.001415352 |
| NDUFS1 | 0.072952319 | -0.073100173 | -0.000147854 |
| TVP23B | 0.043231812 | -0.037438929 | 0.005792883 |
| SLC44A3 | 0.063599075 | -0.06148401 | 0.002115065 |
| KLHDC8B | 0.03717105 | -0.030975147 | 0.006195903 |
| TSNAX | 0.06185109 | -0.041473946 | 0.020377144 |
| RNF6 | 0.046064669 | -0.043466684 | 0.002597986 |
| ELOVL6 | 0.032937443 | -0.029973903 | 0.00296354 |

| | | | |
|---|---|---|---|
| UCK1 | 0.051258898 | -0.054573108 | -0.00331421 |
| ILKAP | 0.058802937 | -0.048548594 | 0.010254342 |
| DLEU1 | 0.042993685 | -0.044733292 | -0.001739607 |
| FAM169A | 0.040871869 | -0.032041095 | 0.008830773 |
| EPHA2 | 0.057912568 | -0.063562052 | -0.005649484 |
| YIPF6 | 0.066020354 | -0.071283962 | -0.005263608 |
| CTNS | 0.038516871 | -0.047173405 | -0.008656534 |
| ITGAE | 0.056224509 | -0.03513112 | 0.021093389 |
| GALK1 | 0.050018786 | -0.045854249 | 0.004164537 |
| CLDN3 | 0.106898058 | -0.17884959 | -0.071951532 |
| ORAI1 | 0.038705222 | -0.03650331 | 0.002201913 |
| SRPK1 | 0.049382774 | -0.057934387 | -0.008551612 |
| PKDCC | 0.024004985 | -0.017250719 | 0.006754265 |
| DMAP1 | 0.057422435 | -0.033383606 | 0.024038829 |
| POMGNT1 | 0.048313318 | -0.047754204 | 0.000559114 |
| RARA-AS1 | 0.02457449 | -0.022067692 | 0.002506798 |
| IRF2 | 0.05957814 | -0.035641376 | 0.023936764 |
| PHKG2 | 0.049382406 | -0.043177443 | 0.006204963 |
| PRLR | -0.031522187 | 0.023726133 | -0.007796054 |
| PRKDC | 0.086262556 | -0.127445206 | -0.04118265 |
| LZTS3 | 0.058681597 | -0.10047247 | -0.041790873 |
| ACOT8 | 0.043975935 | -0.039080141 | 0.004895794 |
| AC013275.1 | 0.021763835 | -0.021635959 | 0.000127876 |
| PSRC1 | 0.022186585 | -0.01993581 | 0.002250775 |
| MBP | 0.063213374 | -0.052876221 | 0.010337153 |
| UBE2L6 | 0.052376153 | -0.040874121 | 0.011502032 |
| GADD45A | 0.060124815 | -0.047687083 | 0.012437733 |
| IRF3 | 0.058948142 | -0.064281753 | -0.005333611 |
| AC099552.3 | 0.016958986 | -0.016958986 | -1.95399E-14 |
| GNB5 | 0.047563338 | -0.049781258 | -0.002217919 |
| DNAH5 | 0.023459419 | -0.021970391 | 0.001489029 |
| MIR181A2HG | 0.021151289 | -0.019486592 | 0.001664698 |
| POLRMT | 0.04299091 | -0.035244109 | 0.007746802 |
| TMEM19 | 0.03888146 | -0.033859331 | 0.005022129 |
| CIAO1 | 0.064700869 | -0.068079217 | -0.003378348 |
| SELENOP | 0.098111789 | -0.164940498 | -0.066828709 |
| SRSF3 | -0.104709261 | 0.073772705 | -0.030936556 |
| NTN1 | 0.019509518 | -0.018377865 | 0.001131652 |
| SRRT | 0.057967626 | -0.062243452 | -0.004275826 |
| ZNF599 | 0.036500186 | -0.028865994 | 0.007634192 |
| SCG5 | 0.021026062 | -0.017397756 | 0.003628306 |
| ALG2 | 0.04696635 | -0.047379096 | -0.000412745 |
| SNHG15 | 0.048357751 | -0.034992458 | 0.013365293 |
| PEA15 | 0.04369271 | -0.035135622 | 0.008557088 |
| HDAC1 | 0.069366011 | -0.081438723 | -0.012072711 |
| DHX32 | 0.045707152 | -0.025659691 | 0.020047461 |
| PRSS22 | 0.06430064 | -0.096668088 | -0.032367448 |

| | | | |
|---|---|---|---|
| UBE3C | 0.048143729 | -0.0606059 | -0.012462171 |
| SYNGR1 | 0.100596584 | -0.154983285 | -0.054386701 |
| ZDHHC3 | 0.060731248 | -0.071373133 | -0.010641884 |
| C1orf50 | 0.049154831 | -0.030660317 | 0.018494515 |
| LIN37 | 0.034021542 | -0.028313175 | 0.005708367 |
| IFITM10 | 0.020357897 | -0.017425566 | 0.002932331 |
| FGFBP1 | 0.022444243 | -0.018356324 | 0.004087919 |
| CLPX | 0.052067514 | -0.033301328 | 0.018766186 |
| EBAG9 | 0.067164027 | -0.061357988 | 0.005806039 |
| NOS1AP | 0.019419072 | -0.020656627 | -0.001237555 |
| FOLR1 | -0.044617356 | 0.046120242 | 0.001502886 |
| DPP8 | 0.049086786 | -0.03474264 | 0.014344146 |
| RIC1 | 0.040270861 | -0.040263679 | 7.18206E-06 |
| FAAH | 0.047219309 | -0.050866502 | -0.003647193 |
| PTK2 | 0.064633861 | -0.037695791 | 0.02693807 |
| DUSP14 | 0.034536648 | -0.020474496 | 0.014062152 |
| GIGYF2 | 0.066562683 | -0.088163694 | -0.02160101 |
| KIFAP3 | 0.046879298 | -0.046101063 | 0.000778235 |
| DNAJB11 | 0.055140075 | -0.063740002 | -0.008599927 |
| UBAC2 | 0.062513579 | -0.055167359 | 0.00734622 |
| PABPC1 | 0.098228394 | -0.101266614 | -0.00303822 |
| ALAS1 | 0.041012764 | -0.030003897 | 0.011008867 |
| RAB26 | 0.019567827 | -0.016505646 | 0.003062181 |
| PLEKHJ1 | 0.071517498 | -0.045210557 | 0.026306941 |
| ELP6 | 0.052228107 | -0.045533043 | 0.006695063 |
| SAMD4A | 0.036973874 | -0.036350587 | 0.000623287 |
| AP003498.2 | -0.046684368 | 0.03404634 | -0.012638028 |
| CCDC82 | 0.057561848 | -0.067042418 | -0.00948057 |
| CCDC106 | 0.057853919 | -0.038905577 | 0.018948343 |
| DNAJB2 | 0.066897678 | -0.086634913 | -0.019737236 |
| NUDT9 | 0.055832322 | -0.064201057 | -0.008368735 |
| FADS3 | 0.042255049 | -0.052736377 | -0.010481329 |
| CTNNBL1 | 0.048247407 | -0.044306676 | 0.003940731 |
| KMO | 0.021704942 | -0.017905107 | 0.003799835 |
| CDK14 | 0.023824303 | -0.015087872 | 0.008736431 |
| DKC1 | 0.051411259 | -0.036689117 | 0.014722143 |
| LRCH1 | 0.032229805 | -0.028478227 | 0.003751578 |
| MRPL39 | 0.050435357 | -0.030561471 | 0.019873885 |
| TLN1 | 0.076896815 | -0.101844757 | -0.024947943 |
| NDUFV1 | 0.081944057 | -0.057265731 | 0.024678326 |
| P4HB | 0.082368108 | -0.130245318 | -0.04787721 |
| DNAJB9 | 0.060029973 | -0.052619705 | 0.007410268 |
| PYGO1 | 0.021621107 | -0.019084203 | 0.002536903 |
| RARB | 0.021611367 | -0.019091187 | 0.00252018 |
| RER1 | 0.079930163 | -0.089802005 | -0.009871842 |
| TXNRD2 | 0.040554951 | -0.03376086 | 0.006794091 |
| HIST1H1D | -0.027938816 | 0.015809075 | -0.01212974 |

| | | | |
|---|---|---|---|
| ESRP1 | 0.040662353 | -0.037570214 | 0.003092139 |
| RAB22A | 0.059011706 | -0.074709696 | -0.015697989 |
| CEBPG | 0.039284417 | -0.041366169 | -0.002081753 |
| ERO1B | 0.040655602 | -0.040664813 | -9.21067E-06 |
| KLHL7 | 0.037599717 | -0.028319005 | 0.009280712 |
| GDE1 | 0.046855529 | -0.050563579 | -0.00370805 |
| CD83 | 0.024364791 | -0.013843566 | 0.010521225 |
| IGSF22 | 0.022088071 | -0.022734116 | -0.000646045 |
| OXA1L | 0.057611438 | -0.035577225 | 0.022034213 |
| CASD1 | 0.047563578 | -0.050614484 | -0.003050906 |
| APCDD1 | 0.018848597 | -0.020244583 | -0.001395986 |
| ACACA | 0.039117016 | -0.051072308 | -0.011955292 |
| RSU1 | 0.04533239 | -0.026847152 | 0.018485238 |
| TFDP2 | 0.081457628 | -0.111551223 | -0.030093595 |
| LUC7L2 | 0.081623813 | -0.129197913 | -0.0475741 |
| MFAP1 | 0.066174088 | -0.057329006 | 0.008845082 |
| PRDX4 | 0.067840895 | -0.070156138 | -0.002315243 |
| SLC16A9 | -0.046040672 | 0.077970636 | 0.031929964 |
| COMMD10 | 0.053518427 | -0.041120706 | 0.012397722 |
| RNF114 | 0.067221608 | -0.048969187 | 0.018252421 |
| CSTF1 | 0.043710201 | -0.032854675 | 0.010855526 |
| HOOK3 | 0.064850458 | -0.080961959 | -0.016111501 |
| IFT57 | 0.067954085 | -0.039824616 | 0.028129469 |
| RGS11 | 0.016871936 | -0.016871936 | -1.95399E-14 |
| RPP25 | 0.024177614 | -0.024288795 | -0.000111181 |
| PEX1 | 0.054495068 | -0.080615899 | -0.02612083 |
| PLEKHA3 | 0.065129439 | -0.085154377 | -0.020024938 |
| RABEP1 | 0.053399991 | -0.064592713 | -0.011192722 |
| SLC12A2 | 0.037570011 | -0.033785246 | 0.003784764 |
| S100A14 | 0.091919523 | -0.081405622 | 0.010513901 |
| SH3GLB2 | 0.074183397 | -0.073636911 | 0.000546486 |
| H1FX | -0.087830271 | 0.124012824 | 0.036182553 |
| MCU | 0.035292703 | -0.031615818 | 0.003676885 |
| LRCH4 | 0.044882961 | -0.047576455 | -0.002693494 |
| TSPYL1 | 0.08618005 | -0.096619598 | -0.010439549 |
| APTR | 0.046241083 | -0.040863077 | 0.005378006 |
| USP48 | 0.06101428 | -0.070563113 | -0.009548833 |
| STXBP2 | 0.053514511 | -0.040503099 | 0.013011412 |
| RAB4A | 0.066857198 | -0.05216515 | 0.014692048 |
| CACNA1D | 0.018755559 | -0.016786519 | 0.00196904 |
| NUB1 | 0.06640715 | -0.078537842 | -0.012130691 |
| STK16 | 0.055393974 | -0.050381573 | 0.005012401 |
| REXO4 | 0.048696604 | -0.052988044 | -0.00429144 |
| C11orf49 | 0.044673704 | -0.044542266 | 0.000131439 |
| FNDC11 | 0.018049028 | -0.018049028 | -2.30926E-14 |
| GRIN2A | -0.030469431 | 0.034165898 | 0.003696467 |
| AC103740.1 | 0.024286553 | -0.025614161 | -0.001327608 |

| | | | |
|---|---|---|---|
| MAP3K1 | 0.074759723 | -0.112025799 | -0.037266076 |
| AGFG1 | 0.058231167 | -0.069425473 | -0.011194306 |
| DNAAF2 | 0.043697444 | -0.04458541 | -0.000887965 |
| GGNBP2 | 0.063697855 | -0.055234787 | 0.008463068 |
| FBXL17 | 0.039280142 | -0.031723458 | 0.007556683 |
| PTRH2 | 0.044144246 | -0.050198118 | -0.006053872 |
| PRRG2 | 0.03713586 | -0.04130559 | -0.00416973 |
| Z95331.1 | 0.037697033 | -0.048662073 | -0.01096504 |
| NOLC1 | 0.047347961 | -0.052530133 | -0.005182172 |
| SMIM10L1 | 0.057148583 | -0.063040277 | -0.005891694 |
| GRAMD1A | 0.032853386 | -0.021482039 | 0.011371347 |
| PLRG1 | 0.055394759 | -0.040240818 | 0.015153941 |
| PGM2L1 | 0.025060118 | -0.020816935 | 0.004243182 |
| LINC01671 | -0.02409993 | 0.017248558 | -0.006851372 |
| ANKRD37 | 0.038626238 | -0.035073224 | 0.003553014 |
| TXNDC11 | 0.046832135 | -0.040657914 | 0.006174221 |
| HNRNPA2B1 | 0.105007682 | -0.186599261 | -0.081591579 |
| KIAA1522 | 0.062161267 | -0.064999209 | -0.002837942 |
| LETMD1 | 0.061262932 | -0.076788041 | -0.015525109 |
| EPB41L4A | 0.043910985 | -0.055913328 | -0.012002343 |
| GOLGA2 | 0.058614961 | -0.086228175 | -0.027613214 |
| SETBP1 | 0.058138417 | -0.077989397 | -0.01985098 |
| PRPSAP1 | 0.050616166 | -0.062842117 | -0.012225951 |
| SMNDC1 | 0.052258246 | -0.043185671 | 0.009072576 |
| ZNF593 | 0.05809897 | -0.042452831 | 0.015646139 |
| TSPAN33 | 0.065774811 | -0.042865984 | 0.022908827 |
| GINM1 | 0.088171674 | -0.163601576 | -0.075429902 |
| TBC1D24 | 0.033846462 | -0.026858815 | 0.006987647 |
| VPS36 | 0.066028412 | -0.071669984 | -0.005641572 |
| THAP3 | 0.049324297 | -0.048746823 | 0.000577474 |
| HNRNPM | 0.093870946 | -0.097467541 | -0.003596595 |
| DHRS4 | 0.037008771 | -0.030632961 | 0.00637581 |
| PYM1 | 0.044445269 | -0.048897968 | -0.004452698 |
| CTPS1 | 0.024440651 | -0.02166079 | 0.002779861 |
| QSOX1 | 0.043403254 | -0.043285982 | 0.000117272 |
| RPAP3 | 0.049948789 | -0.057535373 | -0.007586585 |
| CHRDL1 | 0.019445618 | -0.020964574 | -0.001518956 |
| EIF2D | 0.048005721 | -0.04098774 | 0.007017981 |
| AC006230.1 | 0.021470199 | -0.021509856 | -3.96565E-05 |
| C12orf65 | 0.054481804 | -0.055043181 | -0.000561377 |
| GPR137 | 0.040574145 | -0.044700806 | -0.00412666 |
| ENPP4 | 0.057526058 | -0.084079623 | -0.026553565 |
| RAB1B | 0.076355371 | -0.064271389 | 0.012083983 |
| GNAI2 | 0.081657212 | -0.088151783 | -0.00649457 |
| IPMK | 0.028049393 | -0.029151142 | -0.001101748 |
| WDR33 | 0.069758961 | -0.05374195 | 0.016017011 |
| HIPK1 | 0.049298497 | -0.05022664 | -0.000928143 |

| | | | |
|---|---|---|---|
| TMEM132A | 0.021102537 | -0.01896187 | 0.002140667 |
| CD46 | 0.103198231 | -0.302169475 | -0.198971244 |
| DRG2 | 0.043565664 | -0.035850949 | 0.007714714 |
| PSMC6 | 0.067618023 | -0.039860123 | 0.0277579 |
| LINC02610 | 0.021854209 | -0.021475051 | 0.000379158 |
| EMG1 | 0.056108503 | -0.037677175 | 0.018431328 |
| TM7SF3 | 0.057230966 | -0.068778315 | -0.011547349 |
| CASC4 | 0.080462407 | -0.130232939 | -0.049770531 |
| PPP1CA | 0.072950667 | -0.071611788 | 0.001338879 |
| DALRD3 | 0.057958261 | -0.043391956 | 0.014566305 |
| YDJC | 0.053384699 | -0.037992104 | 0.015392596 |
| MRPL9 | 0.049774949 | -0.030391361 | 0.019383588 |
| PCID2 | 0.053187644 | -0.050104832 | 0.003082812 |
| NOTUM | 0.016143113 | -0.016143113 | -1.95399E-14 |
| TSC22D3 | -0.051792211 | 0.060206629 | 0.008414418 |
| MANSC1 | 0.033090457 | -0.032120846 | 0.000969611 |
| PPP3CB | 0.044361772 | -0.042725324 | 0.001636448 |
| MGMT | 0.07843228 | -0.051600655 | 0.026831625 |
| RGS12 | 0.048243464 | -0.046272408 | 0.001971056 |
| CCDC57 | 0.039429459 | -0.040614695 | -0.001185236 |
| SH3KBP1 | 0.024411477 | -0.025720507 | -0.001309031 |
| FGFR1OP2 | 0.060918424 | -0.069026029 | -0.008107606 |
| LRRC28 | 0.039715778 | -0.034877496 | 0.004838282 |
| RAB21 | 0.072456645 | -0.070334563 | 0.002122081 |
| VPS4B | 0.060865682 | -0.068081241 | -0.007215559 |
| SCYL1 | 0.056437209 | -0.036040923 | 0.020396286 |
| PRCC | 0.05039231 | -0.050356138 | 3.61719E-05 |
| KPNA3 | 0.039816727 | -0.040426516 | -0.00060979 |
| RNF10 | 0.059583195 | -0.076172459 | -0.016589264 |
| SH2D4A | 0.058838377 | -0.07703303 | -0.018194653 |
| TMEM86A | 0.024151981 | -0.024855042 | -0.000703061 |
| RPS6KB2 | 0.051141045 | -0.040804764 | 0.010336281 |
| GSDMD | 0.066393678 | -0.067207092 | -0.000813414 |
| TAF13 | 0.03774347 | -0.027666527 | 0.010076943 |
| NONO | 0.070802456 | -0.079173475 | -0.008371019 |
| HLA-E | 0.072841879 | -0.109154472 | -0.036312593 |
| KXD1 | 0.044394359 | -0.026878688 | 0.017515671 |
| TMCC3 | 0.0380452 | -0.034120509 | 0.00392469 |
| MICU2 | 0.067411362 | -0.077956426 | -0.010545064 |
| USB1 | 0.03646744 | -0.026704049 | 0.009763391 |
| CRELD2 | 0.042374546 | -0.052842451 | -0.010467905 |
| PDRG1 | 0.045925073 | -0.034402125 | 0.011522947 |
| CLDND1 | 0.062103502 | -0.063667361 | -0.001563859 |
| TULP4 | 0.060050412 | -0.073766141 | -0.013715729 |
| CXorf38 | 0.038107441 | -0.040905088 | -0.002797647 |
| CPQ | 0.065807314 | -0.068612545 | -0.002805231 |
| MCCC1 | 0.048342903 | -0.050764782 | -0.002421879 |

| | | | |
|---|---|---|---|
| AP000569.1 | 0.017766223 | -0.017616903 | 0.00014932 |
| GARS | 0.043658828 | -0.041902783 | 0.001756045 |
| TRMT11 | 0.047114826 | -0.054212178 | -0.007097352 |
| TTF2 | 0.030546095 | -0.031069463 | -0.000523368 |
| ARHGEF18 | 0.044107736 | -0.05326493 | -0.009157194 |
| FGD5-AS1 | 0.078410241 | -0.113839685 | -0.035429444 |
| NFKBIL1 | 0.048538418 | -0.047243838 | 0.00129458 |
| MAPKAPK5-AS1 | 0.045541214 | -0.044885919 | 0.000655295 |
| HEXIM1 | -0.063957674 | 0.052977268 | -0.010980406 |
| MRPL37 | 0.04424981 | -0.026115105 | 0.018134705 |
| TMEM121 | 0.034055167 | -0.045344273 | -0.011289106 |
| C16orf91 | 0.048801462 | -0.045762449 | 0.003039014 |
| CHMP4B | 0.063844958 | -0.051774636 | 0.012070322 |
| MAEA | 0.052598199 | -0.043091598 | 0.009506601 |
| GOT2 | 0.045151115 | -0.038262173 | 0.006888942 |
| SMYD3 | 0.037393861 | -0.044432852 | -0.007038992 |
| ANP32A | 0.077113754 | -0.09207925 | -0.014965496 |
| EPB41L4A-AS1 | 0.0828424 | -0.094745601 | -0.011903201 |
| PSMD3 | 0.054208209 | -0.033048116 | 0.021160094 |
| SLC7A6 | 0.037641915 | -0.046104635 | -0.00846272 |
| ING4 | 0.053901519 | -0.046594161 | 0.007307358 |
| TCF7L1 | 0.042150459 | -0.04261854 | -0.000468081 |
| DCAF6 | 0.05702095 | -0.079426448 | -0.022405498 |
| B3GNT2 | 0.052662726 | -0.096012519 | -0.043349793 |
| KLHDC3 | 0.052475496 | -0.062763652 | -0.010288156 |
| TJP1 | 0.07020287 | -0.102788058 | -0.032585188 |
| RNF126 | 0.047284135 | -0.053530275 | -0.00624614 |
| MYO5A | 0.021610588 | -0.021817412 | -0.000206825 |
| FUCA2 | 0.061464405 | -0.066748924 | -0.005284519 |
| FKBPL | 0.028985737 | -0.027117731 | 0.001868006 |
| NUDT15 | 0.042045694 | -0.036814737 | 0.005230957 |
| MLXIP | 0.055552144 | -0.073610232 | -0.018058088 |
| AP002748.4 | 0.020177725 | -0.017844214 | 0.002333511 |
| LLGL2 | 0.063252344 | -0.059765248 | 0.003487096 |
| DYNC1H1 | 0.081711472 | -0.113510344 | -0.031798872 |
| SNHG10 | 0.044076531 | -0.045965421 | -0.00188889 |
| CARS2 | 0.049516719 | -0.041344558 | 0.008172161 |
| DIMT1 | 0.040343118 | -0.049006439 | -0.008663321 |
| THEM6 | 0.042646967 | -0.035106536 | 0.007540431 |
| ILRUN | 0.042273892 | -0.04574096 | -0.003467068 |
| ERVK3-1 | 0.051145241 | -0.04358556 | 0.007559681 |
| RHPN2 | 0.037692026 | -0.033263139 | 0.004428887 |
| ACTL6A | 0.051640912 | -0.049794251 | 0.001846661 |
| RAB6A | 0.054648869 | -0.086463991 | -0.031815122 |
| MKKS | 0.036683672 | -0.038892803 | -0.002209131 |
| OAF | 0.031672216 | -0.041371192 | -0.009698976 |
| SLC25A28 | 0.040462561 | -0.028241762 | 0.012220798 |

| | | | |
|---|---|---|---|
| CDKN2AIP | 0.046094408 | -0.039312389 | 0.006782019 |
| NAT9 | 0.041738901 | -0.031943366 | 0.009795535 |
| ITFG1 | 0.069905822 | -0.111043613 | -0.041137791 |
| EEF1A2 | 0.019451445 | -0.015149974 | 0.004301471 |
| TFPT | 0.03613235 | -0.026149276 | 0.009983074 |
| NEIL1 | 0.030168479 | -0.029402705 | 0.000765774 |
| UBE2E1 | 0.051133896 | -0.041272355 | 0.009861542 |
| TMEM33 | 0.058669401 | -0.059701213 | -0.001031811 |
| PNN | 0.081724882 | -0.157304658 | -0.075579777 |
| RCAN1 | 0.043735422 | -0.028563441 | 0.015171982 |
| OCIAD2 | 0.088892929 | 0.081167277 | 0.170060206 |
| PIGT | 0.056795951 | -0.059682606 | -0.002886655 |
| UQCRC1 | 0.0703158 | -0.046407341 | 0.023908459 |
| STRIP1 | 0.031250453 | -0.036312739 | -0.005062286 |
| ZNF628 | 0.018807268 | -0.017174685 | 0.001632583 |
| NAE1 | 0.053912645 | -0.056362564 | -0.00244992 |
| ANKRD13A | 0.044807249 | -0.056457375 | -0.011650126 |
| RASAL1 | 0.033608722 | -0.041786007 | -0.008177284 |
| TBC1D25 | 0.022850701 | -0.02106408 | 0.00178662 |
| ZBED3 | 0.041528426 | -0.034868998 | 0.006659428 |
| SVIP | 0.064638328 | -0.049963776 | 0.014674552 |
| ENPP5 | 0.050297036 | -0.065164156 | -0.014867121 |
| TRABD2B | -0.043153126 | 0.096869469 | 0.053716344 |
| GRWD1 | 0.043911931 | -0.054605596 | -0.010693665 |
| MRPL45 | 0.046422862 | -0.038837235 | 0.007585627 |
| CYB561D2 | 0.04713661 | -0.042240985 | 0.004895626 |
| PROX1-AS1 | 0.015610084 | -0.014151115 | 0.001458969 |
| SS18 | 0.053152539 | -0.054490272 | -0.001337733 |
| FSBP | 0.027366088 | -0.033216575 | -0.005850487 |
| ARHGAP4 | 0.020844232 | -0.014566634 | 0.006277598 |
| C11orf68 | 0.050478532 | -0.052244875 | -0.001766344 |
| DMTN | 0.056421044 | -0.045063868 | 0.011357176 |
| CKAP2 | 0.032588032 | -0.035286883 | -0.002698852 |
| EPRS | 0.066041778 | -0.092063483 | -0.026021705 |
| BCAM | 0.105176447 | -0.276237724 | -0.171061277 |
| ERBB4 | -0.039700025 | 0.024625456 | -0.015074569 |
| ABHD8 | 0.040219215 | -0.027569713 | 0.012649502 |
| GRTP1 | 0.029780211 | -0.019378109 | 0.010402101 |
| ADGRV1 | 0.042861165 | -0.067070251 | -0.024209085 |
| PIP5K1B | 0.01733435 | -0.01733435 | -2.30926E-14 |
| MYPOP | 0.028747656 | -0.034259943 | -0.005512287 |
| NME3 | 0.093150402 | -0.127781587 | -0.034631184 |
| SRSF10 | 0.084506116 | -0.132376982 | -0.047870866 |
| GOPC | 0.061619942 | -0.082655396 | -0.021035454 |
| HIVEP2 | 0.034406078 | -0.03429725 | 0.000108828 |
| NFKB1 | 0.049860667 | -0.052075672 | -0.002215005 |
| HMGN2 | -0.097692382 | 0.168037741 | 0.070345359 |

| | | | |
|---|---|---|---|
| TMEM251 | 0.065385864 | -0.05732282 | 0.008063044 |
| EGLN2 | 0.049483534 | -0.054846802 | -0.005363268 |
| AC058791.1 | 0.038181377 | -0.063324093 | -0.025142716 |
| HDAC3 | 0.046301491 | -0.045749894 | 0.000551596 |
| VPS13B | 0.045377191 | -0.043184169 | 0.002193022 |
| STC1 | 0.033511576 | -0.043324687 | -0.009813112 |
| HNRNPC | 0.088738108 | -0.107948397 | -0.019210289 |
| VPS51 | 0.06365881 | -0.05274616 | 0.01091265 |
| HYAL2 | 0.048601522 | -0.039375281 | 0.009226241 |
| PDE1A | 0.03959215 | -0.032790295 | 0.006801856 |
| ABCC5 | 0.042684568 | -0.061879138 | -0.01919457 |
| ATP13A4 | -0.02443637 | 0.016246316 | -0.008190054 |
| TOB2 | 0.05044981 | -0.052569588 | -0.002119777 |
| FOLR3 | -0.035176711 | 0.024456639 | -0.010720072 |
| RCE1 | 0.038552581 | -0.034494757 | 0.004057825 |
| NMRAL1 | 0.045276641 | -0.038573626 | 0.006703015 |
| SLC40A1 | 0.055969809 | -0.042503123 | 0.013466686 |
| SLC43A3 | 0.014170998 | -0.013099618 | 0.001071381 |
| P4HA2 | 0.019061614 | -0.018029658 | 0.001031956 |
| MAP3K10 | 0.023507218 | -0.024511659 | -0.001004441 |
| WDR43 | 0.050816629 | -0.037961424 | 0.012855206 |
| MTHFD1 | 0.043748999 | -0.0356674 | 0.008081599 |
| NDUFAF5 | 0.032933837 | -0.027309002 | 0.005624835 |
| ZNF326 | 0.066482988 | -0.134648946 | -0.068165958 |
| ANKLE2 | 0.055943333 | -0.068234498 | -0.012291164 |
| MCRS1 | 0.05149432 | -0.035127428 | 0.016366892 |
| RFLNB | 0.03317242 | -0.036545428 | -0.003373008 |
| LTV1 | 0.038576274 | -0.033699136 | 0.004877138 |
| NRBP1 | 0.05865158 | -0.040812837 | 0.017838743 |
| KCTD1 | -0.040059644 | 0.032062645 | -0.007997 |
| FASTKD5 | 0.02830587 | -0.024568086 | 0.003737784 |
| SNX4 | 0.049938004 | -0.039354718 | 0.010583287 |
| CPSF3 | 0.029695898 | -0.032350509 | -0.002654611 |
| EIF4ENIF1 | 0.04128934 | -0.049175968 | -0.007886628 |
| ALDH1A1 | -0.086689684 | 0.122155209 | 0.035465525 |
| NCAPD3 | 0.034806815 | -0.04688622 | -0.012079406 |
| STK25 | 0.066146029 | -0.078100099 | -0.01195407 |
| TOPORS | 0.048098124 | -0.060240363 | -0.012142239 |
| DHX30 | 0.053536945 | -0.042345507 | 0.011191439 |
| ME3 | 0.030004287 | -0.024434926 | 0.005569361 |
| TMEM30A | 0.074897787 | -0.164122046 | -0.089224259 |
| FHDC1 | 0.022328128 | -0.027224799 | -0.004896671 |
| CDYL | 0.045777104 | -0.042706923 | 0.003070181 |
| HOXD1 | 0.021930832 | -0.020628822 | 0.00130201 |
| LINC01089 | 0.053168486 | -0.072559919 | -0.019391434 |
| SUPT6H | 0.051906242 | -0.053247672 | -0.00134143 |
| SCFD1 | 0.059550913 | -0.068091354 | -0.008540441 |

| | | | |
|---|---|---|---|
| GCK | 0.014513411 | -0.014513411 | -2.30926E-14 |
| AC016240.1 | 0.01460407 | -0.01460407 | -2.13163E-14 |
| PPP1R12C | 0.041818015 | -0.046275746 | -0.004457731 |
| CYB5R1 | 0.053783147 | -0.058179988 | -0.004396841 |
| CBY1 | 0.057483831 | -0.058399448 | -0.000915617 |
| OSTM1 | 0.041996725 | -0.049897085 | -0.007900359 |
| EIF2AK1 | 0.060036024 | -0.044492628 | 0.015543396 |
| BHMT | 0.052460072 | 0.078800796 | 0.131260868 |
| APAF1 | 0.03061928 | -0.028799927 | 0.001819353 |
| SLC35A1 | 0.039696404 | -0.025661098 | 0.014035305 |
| SPRYD4 | 0.036392101 | -0.022158087 | 0.014234014 |
| ZNF853 | 0.024431516 | -0.018025909 | 0.006405607 |
| AUXG01000058.1 | -0.024647138 | 0.024463942 | -0.000183196 |
| SEMA6D | -0.031170978 | 0.025330882 | -0.005840096 |
| GAN | 0.015913914 | -0.015398897 | 0.000515017 |
| PPP1R13B | 0.04283365 | -0.049945941 | -0.007112291 |
| CALY | 0.014956303 | -0.014956303 | -2.30926E-14 |
| EHMT1 | 0.047528123 | -0.049914251 | -0.002386129 |
| HIPK3 | 0.062303776 | -0.079292458 | -0.016988682 |
| TOR2A | 0.024362149 | -0.017223415 | 0.007138734 |
| FBXO33 | 0.035824025 | -0.035548816 | 0.000275209 |
| TMEM184B | 0.042744102 | -0.050172272 | -0.007428171 |
| NF2 | 0.045971813 | -0.044626729 | 0.001345084 |
| ARMCX2 | 0.045407753 | -0.031380488 | 0.014027266 |
| PKP3 | 0.01618302 | -0.016242735 | -5.97149E-05 |
| DR1 | 0.045125579 | -0.038253758 | 0.006871821 |
| CTBP2 | -0.054994137 | 0.047749405 | -0.007244732 |
| ZNRF1 | 0.029137309 | -0.027864337 | 0.001272972 |
| COQ8B | 0.031254416 | -0.032802355 | -0.001547939 |
| PAM | 0.058853101 | -0.079558939 | -0.020705838 |
| EPS8L1 | 0.052890368 | -0.054808561 | -0.001918193 |
| BROX | 0.061030411 | -0.04790804 | 0.013122371 |
| GET1 | 0.056066634 | -0.0564027 | -0.000336066 |
| CFL2 | 0.060781186 | -0.061113926 | -0.00033274 |
| ATXN1 | -0.037193446 | 0.023043411 | -0.014150035 |
| THOC2 | 0.064549625 | -0.076023573 | -0.011473948 |
| TESK1 | 0.033831318 | -0.035643994 | -0.001812676 |
| MBNL1 | 0.066946381 | -0.09900547 | -0.032059089 |
| AC015712.1 | 0.016984003 | -0.011242947 | 0.005741055 |
| DNASE1L1 | 0.030082265 | -0.034852779 | -0.004770515 |
| HDAC2 | 0.053360382 | -0.041736564 | 0.011623818 |
| ALG3 | 0.041261052 | -0.027770198 | 0.013490854 |
| FARP1 | -0.087409773 | 0.071631033 | -0.01577874 |
| PGGT1B | 0.043840373 | -0.031332989 | 0.012507383 |
| FAM92A | 0.040970676 | -0.041257597 | -0.000286922 |
| THUMPD2 | 0.053652013 | -0.068659089 | -0.015007076 |
| COL26A1 | 0.05922165 | -0.087869079 | -0.028647428 |

| | | | |
|---|---|---|---|
| SLC36A4 | 0.04632994 | -0.053677177 | -0.007347237 |
| MSANTD4 | 0.034187862 | -0.026405282 | 0.00778258 |
| WDR54 | 0.044694961 | -0.035439168 | 0.009255793 |
| LBH | -0.042709517 | 0.063465701 | 0.020756184 |
| SCAMP1 | 0.058788171 | -0.087223924 | -0.028435754 |
| ELOVL1 | 0.047965032 | -0.039068282 | 0.00889675 |
| ZNF250 | 0.029180734 | -0.023677016 | 0.005503718 |
| TCHP | 0.037779465 | -0.037815539 | -3.60747E-05 |
| RPP38 | 0.045570826 | -0.038082499 | 0.007488327 |
| AP001318.2 | 0.016841022 | -0.014529476 | 0.002311546 |
| MICU3 | 0.030947021 | -0.024545474 | 0.006401547 |
| TPR | 0.075350262 | -0.150156625 | -0.074806363 |
| VGLL1 | 0.047282581 | -0.040741401 | 0.006541181 |
| PAFAH1B2 | 0.059942363 | -0.055295835 | 0.004646528 |
| PRRG1 | 0.06383526 | -0.075195598 | -0.011360338 |
| WDR45 | 0.048768116 | -0.045112639 | 0.003655477 |
| DDOST | 0.070832853 | -0.10928124 | -0.038448387 |
| USP14 | 0.059809688 | -0.038129704 | 0.021679985 |
| AC106772.2 | 0.017379503 | -0.017493652 | -0.000114149 |
| USP3 | 0.041627474 | -0.045065579 | -0.003438106 |
| C16orf87 | 0.04024897 | -0.034319845 | 0.005929126 |
| HTT | 0.044203663 | -0.05897963 | -0.014775967 |
| NAA30 | 0.034667523 | -0.027675218 | 0.006992305 |
| TCTEX1D2 | 0.056959317 | -0.057691038 | -0.000731721 |
| TRMT9B | 0.032185333 | -0.030008318 | 0.002177015 |
| SRPRA | 0.05607121 | -0.084862351 | -0.028791141 |
| FAM117B | 0.028749538 | -0.030940869 | -0.002191332 |
| ZDHHC24 | 0.043779667 | -0.036310146 | 0.007469521 |
| SMAD1 | 0.0502095 | -0.073950247 | -0.023740746 |
| CREBL2 | 0.04824643 | -0.050535728 | -0.002289299 |
| MOB1B | 0.053985895 | -0.069549855 | -0.015563959 |
| TWISTNB | 0.046557812 | -0.039615112 | 0.0069427 |
| C6orf136 | 0.03847337 | -0.050059387 | -0.011586017 |
| DNASE2 | 0.048034975 | -0.061931479 | -0.013896504 |
| SUSD1 | 0.040028576 | -0.045734864 | -0.005706288 |
| ARIH2 | 0.048632129 | -0.054595876 | -0.005963747 |
| PDCL | 0.04378399 | -0.04345215 | 0.00033184 |
| BTN2A1 | 0.033756733 | -0.032810727 | 0.000946006 |
| LRP8 | 0.013586524 | -0.013586524 | -2.30926E-14 |
| PPP4R3A | 0.054882587 | -0.057961014 | -0.003078427 |
| SNRPA | 0.053701995 | -0.040318794 | 0.013383201 |
| TTC9C | 0.0409505 | -0.04398917 | -0.00303867 |
| BORCS6 | 0.030809121 | -0.024398244 | 0.006410877 |
| TIGD5 | 0.029392095 | -0.028187701 | 0.001204394 |
| SNHG12 | 0.039976342 | -0.059112266 | -0.019135924 |
| SFPQ | 0.087827874 | -0.132611302 | -0.044783428 |
| RND2 | 0.027524044 | -0.026129769 | 0.001394275 |

| | | | |
|---|---|---|---|
| CPEB2 | 0.038761265 | -0.048693816 | -0.009932551 |
| DPY19L4 | 0.051468082 | -0.067375932 | -0.01590785 |
| THRAP3 | 0.071595004 | -0.090010562 | -0.018415557 |
| NCKAP5L | 0.030300101 | -0.026777269 | 0.003522832 |
| EEFSEC | 0.022982478 | -0.01820705 | 0.004775428 |
| EPHX4 | 0.013603185 | -0.013603185 | -1.95399E-14 |
| AL160254.1 | 0.013787984 | -0.013787984 | -2.30926E-14 |
| AC006460.2 | 0.015056989 | -0.013546112 | 0.001510877 |
| ZFPL1 | 0.045652943 | -0.039872481 | 0.005780462 |
| JARID2 | 0.033141509 | -0.03809089 | -0.004949381 |
| C6orf47 | 0.043944801 | -0.03588096 | 0.008063841 |
| AGO2 | 0.045792696 | -0.065940958 | -0.020148262 |
| DCUN1D3 | 0.020878999 | -0.018976738 | 0.001902261 |
| AC006058.1 | -0.020184354 | 0.01504834 | -0.005136014 |
| TMTC4 | 0.024991419 | -0.016054516 | 0.008936903 |
| E2F4 | 0.041692424 | -0.036547119 | 0.005145305 |
| HID1 | 0.0220265 | -0.019287731 | 0.002738769 |
| LAMP1 | 0.084347749 | -0.132725404 | -0.048377655 |
| APOL6 | -0.030183386 | 0.031206211 | 0.001022825 |
| TRIM45 | -0.031277471 | 0.028873223 | -0.002404248 |
| UNC119B | 0.048493326 | -0.062706087 | -0.014212761 |
| EBPL | 0.062206675 | -0.089650995 | -0.02744432 |
| NCOR2 | 0.060964484 | -0.085882689 | -0.024918205 |
| ZDHHC6 | 0.043766775 | -0.042127397 | 0.001639378 |
| PFKL | 0.063694474 | -0.057486996 | 0.006207478 |
| PC | 0.022411194 | -0.023297378 | -0.000886185 |
| LIMK2 | 0.036746767 | -0.039521942 | -0.002775175 |
| BTC | 0.033997845 | -0.029077993 | 0.004919852 |
| WDFY3 | -0.067919493 | -0.044173196 | -0.112092689 |
| PFDN1 | 0.066047304 | -0.065132314 | 0.00091499 |
| NIP7 | 0.038782639 | -0.02472021 | 0.014062429 |
| LPAR2 | 0.025985387 | -0.030018317 | -0.00403293 |
| RNF113A | 0.033144648 | -0.04152299 | -0.008378342 |
| RHOG | 0.038487338 | -0.049940026 | -0.011452688 |
| TEN1 | 0.035550721 | -0.024228493 | 0.011322227 |
| COQ5 | 0.037923451 | -0.028719789 | 0.009203661 |
| LSM14A | 0.073896853 | -0.085693528 | -0.011796675 |
| ARHGEF16 | 0.043622786 | -0.069891235 | -0.026268449 |
| SIL1 | 0.052816464 | -0.073545843 | -0.020729379 |
| VANGL1 | 0.036079708 | -0.026693681 | 0.009386027 |
| SPAG9 | 0.070954974 | -0.053066073 | 0.017888902 |
| TBCEL | 0.031997587 | -0.027199147 | 0.00479844 |
| EXOSC3 | 0.043098099 | -0.042073284 | 0.001024815 |
| CCDC12 | 0.06430976 | -0.057727085 | 0.006582675 |
| ALDH7A1 | 0.053974706 | -0.044866109 | 0.009108597 |
| TPM4 | 0.054504004 | -0.042158346 | 0.012345658 |
| ARPP21 | 0.015258508 | -0.015258508 | -2.30926E-14 |

| | | | |
|---|---|---|---|
| AC009509.1 | 0.015033697 | -0.015033697 | -2.30926E-14 |
| COG2 | 0.044147754 | -0.047057686 | -0.002909932 |
| ABCA5 | 0.045907382 | -0.087553324 | -0.041645942 |
| RNF128 | 0.038554508 | -0.042111816 | -0.003557308 |
| WNK3 | 0.027015898 | -0.024023893 | 0.002992005 |
| STK32B | -0.026625331 | 0.017694282 | -0.008931049 |
| CDC42SE2 | 0.06182405 | -0.047854851 | 0.013969198 |
| DCAF13 | 0.046994667 | -0.03370443 | 0.013290237 |
| CCDC86 | 0.028674718 | -0.035355514 | -0.006680796 |
| CBWD2 | 0.037893959 | -0.036279971 | 0.001613988 |
| STK3 | 0.044561075 | -0.039120588 | 0.005440486 |
| MATN2 | 0.019015105 | -0.017256655 | 0.00175845 |
| CRTAP | 0.059286476 | -0.042154013 | 0.017132463 |
| RP9 | 0.038010016 | -0.02612309 | 0.011886925 |
| IQCA1 | 0.032493756 | -0.028700349 | 0.003793407 |
| MARS | 0.032566249 | -0.037034346 | -0.004468097 |
| ANKRD13D | 0.039218435 | -0.043303822 | -0.004085386 |
| CHPF | 0.044830635 | -0.031983871 | 0.012846764 |
| ZNF350-AS1 | -0.030254624 | 0.050906034 | 0.02065141 |
| AC009107.2 | 0.028956123 | -0.02501907 | 0.003937052 |
| PCOLCE | 0.025359933 | -0.028362963 | -0.00300303 |
| SPATS2 | 0.040678268 | -0.046895882 | -0.006217615 |
| RCOR1 | 0.03346341 | -0.050293982 | -0.016830572 |
| KAT8 | 0.046412092 | -0.031548664 | 0.014863428 |
| CLK3 | 0.039258735 | -0.040987324 | -0.001728589 |
| NGLY1 | 0.041964035 | -0.052235262 | -0.010271227 |
| TNNC1 | 0.056801167 | -0.074019615 | -0.017218448 |
| PERP | 0.093317701 | -0.143021212 | -0.049703511 |
| ZDHHC4 | 0.053689111 | -0.06878778 | -0.015098668 |
| PDK3 | 0.046196931 | -0.058814408 | -0.012617477 |
| FYN | 0.030120706 | -0.024530026 | 0.005590681 |
| CYP1B1 | 0.085151352 | -0.113837252 | -0.0286859 |
| ENTPD4 | 0.038267893 | -0.034610588 | 0.003657305 |
| KPNA1 | 0.042433286 | -0.046927299 | -0.004494014 |
| SKA2 | 0.064605194 | -0.054948357 | 0.009656837 |
| STK39 | 0.027434114 | -0.031477475 | -0.004043361 |
| NUFIP2 | 0.056119724 | -0.059026981 | -0.002907257 |
| MAPKAPK5 | 0.040400806 | -0.047654078 | -0.007253272 |
| LRRC47 | 0.04123092 | -0.028493005 | 0.012737914 |
| CYBA | 0.092429279 | -0.104984299 | -0.012555019 |
| HNRNPU | 0.093337451 | -0.176335387 | -0.082997936 |
| ABCF3 | 0.037817887 | -0.035694253 | 0.002123634 |
| RBM12B | -0.044892439 | 0.032283774 | -0.012608665 |
| SLCO2A1 | -0.027483158 | 0.020952641 | -0.006530517 |
| CYP20A1 | 0.051485072 | -0.048251696 | 0.003233377 |
| LYSMD3 | 0.046238502 | -0.05883852 | -0.012600018 |
| AL445524.1 | 0.021212026 | -0.020623138 | 0.000588888 |

| | | | |
|---|---|---|---|
| NOL4 | -0.023442176 | 0.027630829 | 0.004188653 |
| TEAD2 | 0.027888472 | -0.022851797 | 0.005036675 |
| SPATA2L | 0.029270736 | -0.023056476 | 0.006214261 |
| EXD3 | 0.03091217 | -0.041036788 | -0.010124618 |
| LUC7L | 0.04838454 | -0.057203592 | -0.008819052 |
| CAMSAP2 | 0.038951135 | -0.045664966 | -0.006713831 |
| TMEM243 | 0.067065209 | -0.093402248 | -0.026337039 |
| PPP2R5A | 0.053352704 | -0.046206161 | 0.007146542 |
| RINL | 0.02514396 | -0.022430044 | 0.002713916 |
| GIPR | 0.016724476 | -0.018924096 | -0.00219962 |
| AC011379.1 | 0.020231029 | -0.015231862 | 0.004999167 |
| EVC | 0.057345311 | -0.0824069 | -0.025061589 |
| GRSF1 | 0.050689636 | -0.034266736 | 0.0164229 |
| PAPSS1 | 0.04460205 | -0.029192273 | 0.015409777 |
| XPNPEP1 | 0.029819192 | -0.021559692 | 0.0082595 |
| HCCS | 0.0317836 | -0.034261242 | -0.002477641 |
| ARL6IP1 | 0.082660969 | -0.107058767 | -0.024397799 |
| DNAJA3 | 0.041784777 | -0.046074778 | -0.004290001 |
| DUSP3 | 0.05228596 | -0.058203362 | -0.005917402 |
| WBP11 | 0.058610037 | -0.060009827 | -0.00139979 |
| PCDH7 | 0.013170272 | -0.013170272 | -2.13163E-14 |
| ABCA3 | 0.029279954 | -0.026276571 | 0.003003383 |
| EVC2 | 0.027257709 | -0.026720688 | 0.000537021 |
| MCAT | 0.034445734 | -0.03494024 | -0.000494506 |
| UBE2J1 | 0.044943893 | -0.043918893 | 0.001025 |
| UQCC3 | 0.066663011 | -0.06191362 | 0.004749391 |
| C3orf38 | 0.035921973 | -0.023357196 | 0.012564777 |
| KBTBD2 | 0.038548874 | -0.038290755 | 0.000258118 |
| C1QTNF4 | 0.019206781 | -0.019194736 | 1.20447E-05 |
| CCDC90B | 0.047362902 | -0.037994348 | 0.009368554 |
| TNPO1 | 0.053439774 | -0.044242055 | 0.009197719 |
| AC080038.1 | -0.021597968 | 0.021001282 | -0.000596686 |
| MRPS27 | 0.041952795 | -0.049530139 | -0.007577343 |
| HOXC9 | 0.051379492 | -0.059155913 | -0.00777642 |
| DAB2IP | 0.036614843 | -0.036384436 | 0.000230407 |
| ERGIC2 | 0.062143694 | -0.063389034 | -0.00124534 |
| POLR2M | 0.041660069 | -0.044823729 | -0.003163659 |
| BUD13 | 0.028262012 | -0.031053031 | -0.002791019 |
| HDAC10 | 0.024849679 | -0.02175823 | 0.003091449 |
| RABGEF1 | 0.032328415 | -0.041927494 | -0.009599079 |
| PPIL4 | 0.054630516 | -0.053204768 | 0.001425748 |
| ACTR1B | 0.04296349 | -0.056876222 | -0.013912732 |
| FAM133B | 0.059522377 | -0.080527022 | -0.021004645 |
| KIAA1217 | 0.063357025 | -0.086953283 | -0.023596258 |
| SNAP47 | 0.040791356 | -0.027530276 | 0.013261079 |
| PMPCB | 0.053987417 | -0.043718654 | 0.010268764 |
| CDK5 | 0.028912637 | -0.024672922 | 0.004239715 |

| | | | |
|---|---|---|---|
| C1GALT1C1 | 0.052800071 | -0.069576311 | -0.01677624 |
| DEAF1 | 0.053884221 | -0.057750453 | -0.003866232 |
| RRP9 | 0.029454561 | -0.021391495 | 0.008063065 |
| HAGLR | 0.021701106 | -0.018112979 | 0.003588127 |
| UPF3A | 0.062919012 | -0.096114697 | -0.033195685 |
| HOXB2 | 0.07293602 | -0.101565494 | -0.028629474 |
| LINC01184 | 0.036333847 | -0.034014003 | 0.002319844 |
| DPH7 | 0.034841978 | -0.029393091 | 0.005448887 |
| SEMA4D | 0.065289541 | -0.113300658 | -0.048011117 |
| CHRNB1 | 0.03446061 | -0.029767772 | 0.004692838 |
| PPP2R5E | 0.049072911 | -0.046276084 | 0.002796827 |
| EPHA6 | 0.013831784 | -0.0120676 | 0.001764184 |
| ANKS3 | 0.041143646 | -0.038485732 | 0.002657915 |
| PCDH17 | 0.015914427 | -0.015914427 | -1.95399E-14 |
| C16orf86 | 0.031871882 | -0.028364358 | 0.003507523 |
| GATD3B | 0.049103032 | -0.034792877 | 0.014310155 |
| CNOT11 | 0.040669699 | -0.028353502 | 0.012316196 |
| CAPZA1 | 0.055473846 | -0.064293141 | -0.008819295 |
| HMGN4 | 0.060987988 | -0.046672081 | 0.014315907 |
| AP002387.2 | 0.024113395 | -0.021637283 | 0.002476112 |
| KLF16 | 0.022269824 | -0.014753529 | 0.007516295 |
| KCNK5 | 0.032813475 | -0.032777356 | 3.61186E-05 |
| PSMG4 | 0.04813017 | -0.034574903 | 0.013555267 |
| SOCS1 | 0.036826609 | -0.024019122 | 0.012807486 |
| GNL3 | 0.037754062 | -0.039617437 | -0.001863375 |
| PIGP | 0.066356167 | -0.093656252 | -0.027300084 |
| LINC00113 | -0.02126213 | 0.020697951 | -0.000564179 |
| SMAD4 | 0.06116673 | -0.085947531 | -0.024780801 |
| RRAGB | 0.043323449 | -0.033637708 | 0.009685742 |
| MNAT1 | 0.0451552 | -0.04020876 | 0.00494644 |
| HABP4 | 0.056525239 | -0.067997189 | -0.01147195 |
| SOX4 | -0.083191486 | 0.11179867 | 0.028607184 |
| USO1 | 0.052846372 | -0.056369451 | -0.003523078 |
| SLC25A46 | 0.035815278 | -0.029183891 | 0.006631388 |
| FAM174A | 0.048311597 | -0.047075309 | 0.001236288 |
| AC016831.5 | 0.029638335 | -0.043510171 | -0.013871837 |
| TRIM44 | 0.062797987 | -0.074441294 | -0.011643307 |
| NIFK | 0.051261284 | -0.036821787 | 0.014439497 |
| MRPS30 | 0.040969198 | -0.045459548 | -0.004490351 |
| MARC1 | 0.031916292 | -0.041887791 | -0.0099715 |
| PLEKHA1 | 0.059791711 | -0.104804567 | -0.045012856 |
| CABCOCO1 | -0.024463523 | 0.03051724 | 0.006053717 |
| AUP1 | 0.057017254 | -0.072936558 | -0.015919303 |
| AC025164.1 | 0.035738844 | -0.048087093 | -0.012348249 |
| GSTO2 | 0.043532909 | -0.033845378 | 0.009687531 |
| IDE | 0.026852358 | -0.029032285 | -0.002179927 |
| LONP1 | 0.037784296 | -0.040678344 | -0.002894048 |

| | | | |
|---|---|---|---|
| TGS1 | 0.046759418 | -0.070674245 | -0.023914827 |
| DHRS3 | -0.043319994 | 0.053698144 | 0.01037815 |
| TRIM5 | 0.032361682 | -0.022231665 | 0.010130017 |
| DCLK2 | 0.013977312 | -0.01200377 | 0.001973542 |
| SMARCD3 | 0.018382226 | -0.012649414 | 0.005732812 |
| SAMD12 | 0.046147242 | -0.067861831 | -0.021714589 |
| BDKRB1 | 0.012546727 | -0.012546727 | -1.95399E-14 |
| MINPP1 | 0.032905969 | -0.02834765 | 0.004558319 |
| CAMKMT | 0.037455915 | -0.031540234 | 0.005915681 |
| GMNN | 0.040347803 | -0.04089017 | -0.000542367 |
| PI4K2B | 0.033104503 | -0.028208473 | 0.004896031 |
| GRIPAP1 | 0.042735357 | -0.058726192 | -0.015990835 |
| ZNF24 | 0.056042299 | -0.064465341 | -0.008423041 |
| LTBP1 | 0.018756372 | -0.020934127 | -0.002177755 |
| NOSIP | 0.056081091 | -0.054729749 | 0.001351342 |
| NEMF | 0.057130746 | -0.090139821 | -0.033009076 |
| CUTC | 0.034135979 | -0.033242111 | 0.000893868 |
| LDLRAP1 | 0.024346173 | -0.021190509 | 0.003155665 |
| AZIN1-AS1 | 0.034094139 | -0.028102588 | 0.005991551 |
| PPIP5K2 | 0.054420686 | -0.068372923 | -0.013952237 |
| CDCA7L | 0.033616987 | -0.026693357 | 0.00692363 |
| GMPPB | 0.027019417 | -0.022782872 | 0.004236545 |
| RNPEPL1 | 0.037545684 | -0.039629646 | -0.002083962 |
| ATP6V1H | 0.049819676 | -0.037914179 | 0.011905496 |
| RAB43 | 0.019642024 | -0.017544705 | 0.002097319 |
| PHTF1 | 0.026275359 | -0.027719076 | -0.001443718 |
| STAM | 0.035427733 | -0.029789477 | 0.005638256 |
| DTWD2 | 0.026823888 | -0.031872299 | -0.005048411 |
| LMO7 | 0.07821074 | -0.163180099 | -0.084969359 |
| RAB4B | 0.03132654 | -0.029944075 | 0.001382466 |
| SLPI | 0.084448407 | -0.121061522 | -0.036613115 |
| RASSF2 | -0.02200827 | 0.027307142 | 0.005298871 |
| GPBP1 | 0.077232158 | -0.140024599 | -0.062792441 |
| TYW1 | 0.029811471 | -0.039126252 | -0.009314781 |
| MOGS | 0.04039345 | -0.032124202 | 0.008269248 |
| RAB8B | 0.033882466 | -0.031559717 | 0.002322749 |
| TMEM138 | 0.030760341 | -0.041636918 | -0.010876577 |
| SNRNP70 | 0.073111074 | -0.077523733 | -0.004412659 |
| RORA | 0.060589438 | -0.08513971 | -0.024550272 |
| PARL | 0.05354684 | -0.057702973 | -0.004156134 |
| KLF11 | 0.026417147 | -0.022945999 | 0.003471148 |
| DYM | 0.035557746 | -0.027089491 | 0.008468255 |
| GOLGA7 | 0.053389776 | -0.04897285 | 0.004416926 |
| SUPT16H | 0.060885555 | -0.047190859 | 0.013694696 |
| PATJ | 0.068330492 | -0.135445589 | -0.067115096 |
| ATF5 | 0.030683496 | -0.025394873 | 0.005288623 |
| SOCS4 | 0.038704515 | -0.043723811 | -0.005019295 |

| | | | |
|---|---|---|---|
| LINC01014 | -0.020583123 | 0.022939097 | 0.002355974 |
| C5orf51 | 0.037477551 | -0.0509847 | -0.013507149 |
| CDCP1 | 0.011715467 | -0.010841189 | 0.000874278 |
| PSMD14 | 0.047448963 | -0.047625725 | -0.000176762 |
| SGK2 | 0.024746514 | -0.019440915 | 0.005305599 |
| AR | -0.026413645 | 0.025625394 | -0.000788251 |
| HBS1L | 0.044209462 | -0.058766807 | -0.014557344 |
| P3H2 | 0.076539686 | -0.142269229 | -0.065729543 |
| DDX10 | 0.029290727 | -0.02151341 | 0.007777317 |
| ABRAXAS2 | 0.035242165 | -0.037042044 | -0.001799879 |
| WDR41 | 0.031571654 | -0.033298784 | -0.00172713 |
| AGT | 0.012897673 | -0.012897673 | -2.13163E-14 |
| DUSP12 | 0.036705716 | -0.036151265 | 0.000554451 |
| SLC45A4 | 0.029604998 | -0.042067915 | -0.012462917 |
| AATF | 0.043078899 | -0.031193751 | 0.011885148 |
| GPS2 | 0.065018041 | -0.084804948 | -0.019786907 |
| UBE2J2 | 0.048985217 | -0.034013052 | 0.014972165 |
| TBC1D17 | 0.040215384 | -0.044442458 | -0.004227074 |
| PRPF40A | 0.060460587 | -0.081826234 | -0.021365647 |
| NUP42 | 0.043045452 | -0.045960714 | -0.002915262 |
| SH2B2 | 0.016944659 | -0.015015435 | 0.001929223 |
| CEP170 | 0.032976771 | -0.047445829 | -0.014469059 |
| ARHGEF3 | 0.014081535 | -0.011213344 | 0.002868192 |
| EPPK1 | 0.02800399 | -0.026740143 | 0.001263847 |
| C19orf25 | 0.049477724 | -0.05648225 | -0.007004526 |
| MFSD12 | 0.029284675 | -0.031042636 | -0.001757961 |
| MDM2 | 0.041842999 | -0.037988694 | 0.003854305 |
| METTL27 | 0.034711484 | -0.040001729 | -0.005290245 |
| WAC | 0.068050915 | -0.089832167 | -0.021781252 |
| EXOSC1 | 0.039078169 | -0.027655233 | 0.011422936 |
| FMNL3 | 0.012391159 | -0.011279171 | 0.001111988 |
| TLCD5 | 0.03208778 | -0.031377107 | 0.000710673 |
| TRIP11 | 0.069417221 | -0.111393508 | -0.041976288 |
| PHYHIPL | -0.067769878 | 0.055099401 | -0.012670477 |
| GPRC5C | 0.06498355 | -0.046507365 | 0.018476184 |
| TRAPPC2B | 0.034744508 | -0.026991957 | 0.007752551 |
| NCEH1 | 0.04192893 | -0.067484208 | -0.025555278 |
| SERPINI1 | 0.023209851 | -0.022429894 | 0.000779957 |
| CHID1 | 0.064114862 | -0.069102955 | -0.004988093 |
| OGFOD1 | 0.030156933 | -0.04226274 | -0.012105807 |
| POLR3D | 0.024889052 | -0.021760625 | 0.003128427 |
| SLC25A51 | 0.022515862 | -0.015233613 | 0.007282248 |
| FAM222A | 0.014368333 | -0.009821031 | 0.004547302 |
| TBC1D2 | 0.018033777 | -0.015354838 | 0.002678939 |
| CDK11B | 0.034236967 | -0.040137291 | -0.005900324 |
| CCNB1 | 0.02361874 | -0.025525675 | -0.001906935 |
| ADCK2 | 0.032986718 | -0.024181344 | 0.008805374 |

| | | | |
|---|---|---|---|
| HSPA13 | 0.038233766 | -0.048420408 | -0.010186642 |
| FBXO32 | 0.021406948 | -0.019832397 | 0.001574552 |
| GNPNAT1 | 0.03024698 | -0.023155125 | 0.007091855 |
| PATL1 | 0.035051349 | -0.039728431 | -0.004677082 |
| UBE2K | 0.059505651 | -0.071401322 | -0.011895671 |
| POGK | 0.03934033 | -0.03421575 | 0.00512458 |
| RWDD4 | 0.045874795 | -0.032458361 | 0.013416434 |
| PTDSS2 | 0.034216727 | -0.032398563 | 0.001818164 |
| ZNF700 | 0.033092634 | -0.040699632 | -0.007606998 |
| LGALS3BP | 0.062277874 | -0.077102806 | -0.014824932 |
| AP2A2 | 0.056034039 | -0.083112926 | -0.027078887 |
| CD164 | 0.07911388 | -0.112539311 | -0.033425431 |
| RFNG | 0.043905116 | -0.053908497 | -0.010003381 |
| MIR34AHG | 0.011148935 | -0.011148935 | -1.95399E-14 |
| IRF2BPL | 0.059744798 | -0.112081548 | -0.05233675 |
| RBP1 | -0.055286201 | 0.081628811 | 0.026342609 |
| HMGXB4 | 0.036586648 | -0.037789458 | -0.00120281 |
| ESR2 | 0.012097469 | -0.012097469 | -2.30926E-14 |
| GMCL1 | 0.046004213 | -0.068265727 | -0.022261514 |
| ATXN7L3B | 0.065700473 | -0.086820976 | -0.021120503 |
| CHRNA5 | 0.011746835 | -0.011746835 | -2.13163E-14 |
| PIGK | 0.05106222 | -0.0743122 | -0.02324998 |
| ZNF124 | -0.029475651 | 0.026898231 | -0.00257742 |
| TYRO3 | -0.022857773 | 0.0188718 | -0.003985974 |
| CNOT7 | 0.060247082 | -0.047613711 | 0.012633371 |
| BICDL1 | 0.051995044 | -0.070346474 | -0.01835143 |
| MBOAT2 | 0.052341001 | -0.083192659 | -0.030851657 |
| FANCL | 0.031460358 | -0.038270662 | -0.006810304 |
| ZBTB33 | 0.021869406 | -0.020368029 | 0.001501377 |
| RBFOX3 | 0.011266686 | -0.011266686 | -1.95399E-14 |
| CCDC69 | 0.01441358 | -0.014185426 | 0.000228154 |
| DEDD | 0.031753913 | -0.032441173 | -0.000687261 |
| SBNO1 | 0.053559181 | -0.052465754 | 0.001093427 |
| THRA | 0.048480474 | -0.067974051 | -0.019493577 |
| FLYWCH2 | 0.050539904 | -0.045857499 | 0.004682404 |
| PLEKHA8 | 0.021788377 | -0.020324377 | 0.001464 |
| RPS29 | 0.077435127 | 0.073583276 | 0.151018403 |
| CHD1 | 0.045190469 | -0.049143496 | -0.003953027 |
| ANKRD49 | 0.044111201 | -0.042286327 | 0.001824874 |
| RAB11FIP4 | 0.032144047 | -0.02596628 | 0.006177766 |
| CCDC59 | 0.055507757 | -0.047817674 | 0.007690082 |
| PCBP3 | 0.011344646 | -0.011344646 | -1.77636E-14 |
| SYNRG | 0.053007642 | -0.094787536 | -0.041779893 |
| COX16 | 0.066293133 | -0.048057137 | 0.018235996 |
| NBR1 | 0.053715585 | -0.059121565 | -0.00540598 |
| MFN1 | 0.043289434 | -0.044623335 | -0.001333901 |
| NPPC | 0.0113377 | -0.0113377 | -1.95399E-14 |

| | | | |
|---|---|---|---|
| MBIP | 0.046626982 | -0.037172846 | 0.009454136 |
| TMEM44 | 0.011434816 | -0.011434816 | -2.13163E-14 |
| ACTR3C | 0.032578271 | -0.041241641 | -0.00866337 |
| NIPAL3 | 0.062474047 | -0.129288806 | -0.06681476 |
| CEBPZOS | 0.058667684 | -0.068096726 | -0.009429042 |
| CWH43 | -0.024139049 | 0.029675464 | 0.005536415 |
| LAMB3 | 0.018361447 | -0.018257739 | 0.000103708 |
| ENO2 | 0.030850971 | -0.028020673 | 0.002830298 |
| WBP4 | 0.048464592 | -0.052226858 | -0.003762266 |
| BCAR1 | 0.042520917 | -0.043810331 | -0.001289414 |
| PRKAR1A | 0.068791116 | -0.062968853 | 0.005822263 |
| PICK1 | 0.036347428 | -0.028180454 | 0.008166974 |
| SSBP2 | 0.038027858 | -0.047443002 | -0.009415143 |
| UBR2 | 0.047993647 | -0.049784965 | -0.001791318 |
| ZHX1 | 0.044308593 | -0.046574479 | -0.002265885 |
| ALDH9A1 | 0.057068304 | -0.064664051 | -0.007595747 |
| GTPBP8 | 0.033059261 | -0.029453437 | 0.003605824 |
| POGLUT3 | 0.023767087 | -0.021292809 | 0.002474278 |
| ARMC7 | 0.032676431 | -0.032479466 | 0.000196965 |
| ADGRF4 | 0.011943366 | -0.011943366 | -2.30926E-14 |
| DXO | 0.033317172 | -0.030699067 | 0.002618104 |
| DOK1 | 0.02153004 | -0.027036485 | -0.005506445 |
| VPS35 | 0.060575808 | -0.04572838 | 0.014847428 |
| MRPS25 | 0.051638801 | -0.044941972 | 0.006696829 |
| TMEM115 | 0.045958926 | -0.056911877 | -0.010952951 |
| LINC01341 | 0.014797441 | -0.013039923 | 0.001757518 |
| CSNK1G2 | 0.047999048 | -0.057753604 | -0.009754556 |
| ISG20 | 0.012418558 | -0.010469473 | 0.001949085 |
| SLC20A2 | 0.048376937 | -0.086012947 | -0.03763601 |
| RETREG2 | 0.054406806 | -0.072961184 | -0.018554377 |
| SECISBP2 | 0.056360614 | -0.090470992 | -0.034110379 |
| NIT2 | 0.057759415 | 0.045618146 | 0.10337756 |
| ICMT | 0.040914785 | -0.058343764 | -0.017428979 |
| EIF5B | 0.072729008 | -0.111557155 | -0.038828146 |
| AP001999.2 | 0.012825744 | -0.012825744 | -2.30926E-14 |
| COPS4 | 0.044617995 | -0.036103228 | 0.008514767 |
| ZBTB24 | 0.036664068 | -0.043988841 | -0.007324774 |
| ODR4 | 0.040922652 | -0.048048359 | -0.007125706 |
| SLITRK4 | 0.034754758 | -0.031822019 | 0.002932739 |
| PRDM2 | 0.044349696 | -0.063431868 | -0.019082172 |
| DDX18 | 0.069487935 | -0.078123818 | -0.008635882 |
| CTSA | 0.048710934 | -0.071585465 | -0.022874531 |
| TUBB2A | -0.036090667 | 0.073073972 | 0.036983305 |
| SOWAHD | 0.011405114 | -0.011405114 | -2.30926E-14 |
| GAA | 0.054321426 | -0.069708684 | -0.015387258 |
| MARCH9 | 0.043396325 | -0.061910941 | -0.018514616 |
| AVPR1A | -0.01766043 | 0.014728864 | -0.002931566 |

| | | | |
|---|---|---|---|
| SNX25 | 0.025295343 | -0.027851735 | -0.002556392 |
| LSR | 0.044157692 | -0.040599475 | 0.003558217 |
| UBQLN1 | 0.044135319 | -0.045956559 | -0.001821239 |
| RIOK3 | 0.053361763 | -0.045719135 | 0.007642627 |
| ANKRD40 | 0.040336258 | -0.031282065 | 0.009054193 |
| BTD | 0.028970675 | -0.029028232 | -5.75571E-05 |
| XRN1 | 0.050911381 | -0.069825643 | -0.018914262 |
| IL18R1 | 0.015867809 | -0.018090451 | -0.002222642 |
| RILPL1 | 0.035484818 | -0.055181306 | -0.019696487 |
| ICA1 | 0.03190136 | -0.024073724 | 0.007827636 |
| PCGF2 | 0.043221422 | -0.042009657 | 0.001211765 |
| ADPGK | 0.029594466 | -0.038730866 | -0.009136399 |
| DMPK | 0.01858517 | -0.018852406 | -0.000267236 |
| ADAP1 | 0.02581709 | -0.02253387 | 0.00328322 |
| JPT2 | 0.048129665 | -0.046358085 | 0.00177158 |
| CERKL | 0.014672743 | -0.016899216 | -0.002226473 |
| RIC8A | 0.045268667 | -0.036313805 | 0.008954861 |
| ERAL1 | 0.030641465 | -0.025187326 | 0.005454139 |
| PIP4K2C | 0.030077349 | -0.035426268 | -0.005348919 |
| RAD51B | 0.016568255 | -0.01626641 | 0.000301845 |
| ZFYVE27 | 0.026488822 | -0.033326484 | -0.006837662 |
| CEP19 | 0.023911101 | -0.025083211 | -0.001172111 |
| TMED7 | 0.058370018 | -0.109777201 | -0.051407184 |
| PAK1 | 0.030053575 | -0.022173271 | 0.007880304 |
| AL592183.1 | 0.014915407 | -0.016549805 | -0.001634398 |
| ZSCAN16 | 0.0201054 | -0.022971807 | -0.002866407 |
| SPA17 | 0.026799344 | -0.018764099 | 0.008035245 |
| LMTK2 | 0.033330502 | -0.027500966 | 0.005829536 |
| SLC22A15 | 0.019103 | -0.016811714 | 0.002291286 |
| ANGPT2 | -0.031182052 | 0.029588797 | -0.001593255 |
| SYNJ2BP | 0.052360456 | -0.063349054 | -0.010988598 |
| ADCK5 | 0.021622602 | -0.022080602 | -0.000458 |
| IGFBP4 | -0.040751347 | 0.096938172 | 0.056186824 |
| SMARCD2 | 0.034200137 | -0.042469816 | -0.008269679 |
| DOCK9 | 0.028737152 | -0.032868442 | -0.00413129 |
| FRAS1 | 0.044895337 | -0.060289774 | -0.015394437 |
| PPT1 | 0.058095434 | -0.083630921 | -0.025535487 |
| LINC00324 | 0.018317428 | -0.020983836 | -0.002666409 |
| ARRB1 | 0.022939279 | -0.024575103 | -0.001635824 |
| SATB2-AS1 | -0.042328568 | 0.051024334 | 0.008695767 |
| UBE2Q1 | 0.04192896 | -0.05302482 | -0.011095861 |
| DCAF8 | 0.047364485 | -0.046784642 | 0.000579843 |
| VOPP1 | 0.032997388 | -0.032192978 | 0.00080441 |
| LINC01607 | 0.015585138 | -0.01660647 | -0.001021332 |
| WIPI2 | 0.053464749 | -0.06095018 | -0.00748543 |
| GCH1 | 0.016726062 | -0.018699902 | -0.00197384 |
| CTNNBIP1 | 0.034131799 | -0.029462961 | 0.004668838 |

| | | | |
|---|---|---|---|
| SCAMP2 | 0.059679132 | -0.077447784 | -0.017768651 |
| EMC9 | 0.037587366 | -0.057872086 | -0.020284721 |
| FGF2 | 0.038767378 | -0.061225365 | -0.022457988 |
| IRAK1 | 0.029873155 | -0.023239861 | 0.006633293 |
| EDN1 | -0.023565593 | 0.020367421 | -0.003198172 |
| FAM210A | 0.032320413 | -0.03639422 | -0.004073806 |
| KAT2B | 0.023803341 | -0.020417717 | 0.003385623 |
| MUL1 | 0.032963512 | -0.032609464 | 0.000354048 |
| AKT1S1 | 0.035953126 | -0.025697097 | 0.010256029 |
| UBTD2 | 0.027278923 | -0.020299482 | 0.006979441 |
| DDX54 | 0.041626817 | -0.031838183 | 0.009788634 |
| MPPE1 | 0.042774112 | -0.050410498 | -0.007636386 |
| ACOT9 | 0.043914643 | -0.054577285 | -0.010662642 |
| ITGA2 | -0.058418693 | -0.070272286 | -0.128690979 |
| LINC01214 | 0.010596541 | -0.010596541 | -1.95399E-14 |
| ZNF358 | 0.036921093 | -0.036404008 | 0.000517086 |
| DRG1 | 0.036541755 | -0.031782776 | 0.004758979 |
| DMAC2L | 0.037060616 | -0.036373491 | 0.000687125 |
| MAGI2-AS3 | 0.048316097 | -0.071495406 | -0.023179309 |
| VMA21 | 0.049189978 | -0.037588369 | 0.011601609 |
| NAT2 | 0.011071461 | -0.011071461 | -2.30926E-14 |
| ME2 | 0.038387306 | -0.031018427 | 0.007368879 |
| ADCY3 | 0.011394021 | -0.010206432 | 0.00118759 |
| BMT2 | 0.028700334 | -0.036824508 | -0.008124175 |
| PMEL | 0.010517491 | -0.010517491 | -2.13163E-14 |
| PTPRF | -0.056073201 | 0.041064351 | -0.01500885 |
| TACC2 | 0.036311639 | -0.0477586 | -0.011446961 |
| BBC3 | 0.022168014 | -0.016327355 | 0.005840659 |
| TPBG | 0.030791524 | -0.025456709 | 0.005334815 |
| PREX2 | 0.015357558 | -0.014691794 | 0.000665764 |
| KIF5C | 0.015827531 | -0.019090915 | -0.003263384 |
| ENOX2 | 0.026213246 | -0.030676502 | -0.004463255 |
| ACTN1 | 0.036978045 | -0.035137681 | 0.001840364 |
| ACOT11 | 0.029693953 | -0.027133908 | 0.002560045 |
| MAPKAP1 | 0.0441156 | -0.054055864 | -0.009940264 |
| AL353194.1 | 0.023500855 | -0.026386884 | -0.002886029 |
| WDR34 | 0.05992295 | -0.075706888 | -0.015783938 |
| ACAD8 | 0.036117994 | -0.046628838 | -0.010510844 |
| SP100 | 0.048780338 | -0.052093742 | -0.003313404 |
| EDEM2 | 0.031211728 | -0.037427043 | -0.006215315 |
| SETDB2 | 0.029186532 | -0.035980058 | -0.006793526 |
| GPER1 | 0.02170053 | -0.018405558 | 0.003294972 |
| AP000688.2 | 0.012048374 | -0.010751012 | 0.001297362 |
| STAU2 | 0.036799528 | -0.028604038 | 0.008195491 |
| NCKAP5 | 0.012825488 | -0.014044218 | -0.00121873 |
| RNF19A | 0.050170016 | -0.058606961 | -0.008436945 |
| FBXO8 | 0.036576229 | -0.036479555 | 9.66735E-05 |

| | | | |
|---|---|---|---|
| ATG4A | 0.025864704 | -0.021420398 | 0.004444306 |
| SGCB | 0.055020728 | -0.079949858 | -0.024929131 |
| TGOLN2 | 0.073822728 | -0.154079023 | -0.080256295 |
| LMNA | 0.07024007 | -0.052675378 | 0.017564692 |
| PAX8 | 0.088862877 | -0.307801881 | -0.218939004 |
| KCNJ15 | 0.062179048 | -0.083982681 | -0.021803633 |
| HNRNPL | 0.050458323 | -0.046490906 | 0.003967418 |
| CBX4 | 0.021425014 | -0.018062551 | 0.003362463 |
| PPIE | 0.044104621 | -0.035499065 | 0.008605556 |
| SLC9A3 | 0.013165686 | -0.015114684 | -0.001948998 |
| ASB8 | 0.037571984 | -0.03324624 | 0.004325744 |
| SNN | 0.028254348 | -0.030338063 | -0.002083715 |
| MARK2 | 0.044207118 | -0.055395537 | -0.011188419 |
| AC004540.2 | 0.052736905 | -0.052809185 | -7.22798E-05 |
| ILF3 | 0.057968724 | -0.060321965 | -0.002353241 |
| CCN1 | 0.061014628 | -0.107697941 | -0.046683313 |
| TFAP2C | 0.028894639 | -0.040075992 | -0.011181353 |
| LCN12 | 0.043107589 | -0.031001702 | 0.012105887 |
| WDR74 | 0.04168057 | -0.047646353 | -0.005965783 |
| FAM227B | 0.029841525 | -0.043203935 | -0.01336241 |
| ARNT2 | 0.026636072 | -0.020875661 | 0.005760411 |
| LINC01571 | 0.027631873 | -0.026924853 | 0.00070702 |
| WDFY2 | 0.029649171 | -0.024102773 | 0.005546398 |
| CLDN19 | 0.060117764 | -0.104559672 | -0.044441908 |
| TWSG1 | 0.04252531 | -0.051609301 | -0.009083991 |
| VWA5A | 0.027510174 | -0.025299785 | 0.002210389 |
| LRRC59 | 0.037211987 | -0.035316711 | 0.001895276 |
| CPNE1 | 0.051859742 | -0.06078569 | -0.008925948 |
| PITPNB | 0.044960665 | -0.053454466 | -0.008493801 |
| LRRC8C-DT | 0.011388734 | -0.009843141 | 0.001545592 |
| NGDN | 0.035797339 | -0.029357942 | 0.006439397 |
| EIF2B3 | 0.02855673 | -0.032040832 | -0.003484102 |
| AFF3 | 0.016250158 | -0.013305433 | 0.002944725 |
| LEMD2 | 0.033733443 | -0.044961802 | -0.011228359 |
| NMT1 | 0.05562687 | -0.062987949 | -0.007361079 |
| ATP8B1 | 0.033948116 | -0.04629462 | -0.012346504 |
| MCPH1 | 0.032572945 | -0.041555713 | -0.008982768 |
| AL049629.1 | 0.014095189 | -0.01406441 | 3.07791E-05 |
| BMP2 | -0.015591758 | 0.013024897 | -0.002566862 |
| RABL6 | 0.050143348 | -0.050163531 | -2.01837E-05 |
| CROT | 0.030914405 | -0.024070114 | 0.006844291 |
| PLIN5 | -0.017937625 | 0.018409813 | 0.000472188 |
| EGFR | -0.064647626 | -0.053835035 | -0.118482661 |
| MYL3 | 0.026895894 | -0.023286163 | 0.003609731 |
| FOXJ1 | -0.027361846 | 0.020851299 | -0.006510547 |
| HRG | 0.014418544 | -0.011014797 | 0.003403746 |
| WIPI1 | 0.029382462 | -0.041872858 | -0.012490396 |

| | | | |
|---|---|---|---|
| DBNL | 0.044306724 | -0.037408735 | 0.006897989 |
| PRR5 | 0.034370416 | -0.02772585 | 0.006644566 |
| TNKS2 | 0.04160398 | -0.048481699 | -0.006877719 |
| SNHG30 | 0.035548147 | -0.029540484 | 0.006007663 |
| KRBOX1 | 0.02563989 | -0.027317633 | -0.001677743 |
| SERPING1 | -0.061961568 | -0.063846141 | -0.125807709 |
| JAG2 | 0.018640647 | -0.02497526 | -0.006334613 |
| TNFAIP8 | -0.032247855 | 0.038853785 | 0.00660593 |
| HIBCH | 0.047706108 | -0.040423504 | 0.007282604 |
| ITPK1 | 0.03318107 | -0.034507565 | -0.001326495 |
| BLOC1S6 | 0.037320262 | -0.041367649 | -0.004047387 |
| TPRG1L | 0.038252986 | -0.032371538 | 0.005881448 |
| SMURF2 | 0.041345394 | -0.054698805 | -0.013353411 |
| MFN2 | 0.034210227 | -0.035120228 | -0.00091 |
| AC020659.1 | 0.017201706 | -0.016056425 | 0.001145282 |
| ARNT | 0.043511815 | -0.061270348 | -0.017758533 |
| UNC93B1 | 0.030881061 | -0.027507556 | 0.003373505 |
| MANBA | 0.026957481 | -0.03303055 | -0.006073068 |
| RMDN3 | 0.037752241 | -0.045680082 | -0.00792784 |
| MAPK1 | 0.049324504 | -0.075842201 | -0.026517697 |
| KCNQ1OT1 | -0.053622323 | -0.057698378 | -0.111320701 |
| MSL3 | 0.029497588 | -0.04372706 | -0.014229472 |
| JMJD6 | 0.033088436 | -0.038655282 | -0.005566845 |
| LETM1 | 0.040513441 | -0.048070305 | -0.007556863 |
| GNL2 | 0.02830197 | -0.020985969 | 0.007316001 |
| USP9Y | -0.056557083 | -0.113514755 | -0.170071838 |
| TIMM29 | 0.02969253 | -0.028736056 | 0.000956474 |
| GTF2F2 | 0.034844329 | -0.027197154 | 0.007647175 |
| INPP5E | 0.021227529 | -0.019404399 | 0.00182313 |
| CLCN3 | 0.060056805 | -0.115355384 | -0.055298579 |
| CCM2 | 0.028079954 | -0.029611556 | -0.001531602 |
| AC036176.1 | 0.023599584 | -0.019528511 | 0.004071073 |
| CDHR1 | -0.015227208 | 0.011817203 | -0.003410005 |
| NIPA2 | 0.040705048 | -0.054335139 | -0.013630091 |
| OSBPL5 | 0.019823391 | -0.014868394 | 0.004954998 |
| MTUS1 | 0.027935728 | -0.021451376 | 0.006484352 |
| GOLPH3L | 0.034405035 | -0.027845527 | 0.006559509 |
| C10orf82 | -0.020893634 | 0.025620524 | 0.00472689 |
| TK2 | 0.034088943 | -0.029130761 | 0.004958182 |
| GOLT1B | 0.034296851 | -0.031300426 | 0.002996425 |
| POR | 0.049779098 | -0.068999587 | -0.01922049 |
| GUCY1A1 | -0.028942878 | 0.028595877 | -0.000347 |
| BRD4 | 0.051702186 | -0.044466858 | 0.007235327 |
| MCM8 | 0.026215571 | -0.032142734 | -0.005927163 |
| CDKL5 | 0.027883068 | -0.033169347 | -0.00528628 |
| DDB1 | 0.051734225 | -0.049116393 | 0.002617832 |
| SYCE1L | 0.047442999 | -0.039868476 | 0.007574523 |

| | | | |
|---|---|---|---|
| OLFM2 | 0.014468231 | -0.017194286 | -0.002726054 |
| RNASEH1 | 0.032213923 | -0.03867362 | -0.006459697 |
| MIB2 | 0.037954573 | -0.053531274 | -0.015576701 |
| PSMD10 | 0.028049746 | -0.031295407 | -0.003245661 |
| SGCG | 0.01060636 | -0.01060636 | -2.13163E-14 |
| RAD21 | 0.056249958 | -0.069372289 | -0.01312233 |
| C10orf95 | 0.016212037 | -0.017188958 | -0.000976921 |
| MFSD13A | 0.010676257 | -0.0094668 | 0.001209457 |
| TPRA1 | 0.041646213 | -0.038973877 | 0.002672336 |
| NOXA1 | 0.025033564 | -0.019117142 | 0.005916422 |
| SLC25A29 | 0.043332913 | -0.065917232 | -0.02258432 |
| PPFIBP2 | 0.023841047 | -0.034252659 | -0.010411612 |
| IRF5 | 0.01798911 | -0.01381506 | 0.00417405 |
| BCL10 | 0.043534758 | -0.052636767 | -0.009102009 |
| SERPINE2 | -0.025461068 | 0.051556411 | 0.026095342 |
| ZNF579 | 0.043339954 | -0.041900215 | 0.001439739 |
| INTS11 | 0.0414042 | -0.035300731 | 0.006103469 |
| USP39 | 0.029019294 | -0.024939509 | 0.004079785 |
| GPI | 0.051729437 | -0.04250554 | 0.009223897 |
| ZNF800 | 0.032582092 | -0.038302067 | -0.005719975 |
| ZBTB25 | 0.030831846 | -0.038918212 | -0.008086366 |
| BAIAP2L1 | 0.031426755 | -0.028744608 | 0.002682147 |
| PON3 | 0.011458128 | -0.009417794 | 0.002040334 |
| AP000866.2 | 0.035125722 | -0.030622423 | 0.004503299 |
| CARM1 | 0.027874281 | -0.023850552 | 0.00402373 |
| PYCARD | 0.026909267 | -0.020831149 | 0.006078118 |
| FAM83B | 0.019075048 | -0.018836691 | 0.000238357 |
| ALG5 | 0.05499499 | -0.050813409 | 0.004181582 |
| FAM218A | -0.017791648 | 0.013546858 | -0.00424479 |
| EIF3A | 0.064398715 | -0.098200897 | -0.033802182 |
| IARS2 | 0.050650028 | -0.071279481 | -0.020629453 |
| EEF1AKMT4 | 0.014804753 | -0.012064371 | 0.002740382 |
| DCUN1D4 | 0.036653043 | -0.047980308 | -0.011327265 |
| ZNF675 | 0.025118082 | -0.032953675 | -0.007835593 |
| PCP2 | 0.011230046 | -0.010433103 | 0.000796943 |
| PLA2G4A | 0.018960695 | -0.016917288 | 0.002043407 |
| WAPL | 0.047401589 | -0.043848817 | 0.003552773 |
| HOXB4 | 0.036800128 | -0.054574155 | -0.017774026 |
| GRINA | 0.047447979 | -0.05921851 | -0.011770531 |
| FOXA3 | 0.016402312 | -0.013484802 | 0.00291751 |
| WDR55 | 0.029135798 | -0.029620984 | -0.000485185 |
| GEMIN6 | 0.031901304 | -0.028930725 | 0.002970579 |
| SPTLC2 | -0.071491191 | -0.083381943 | -0.154873134 |
| SLC35E3 | 0.027546745 | -0.022366744 | 0.005180001 |
| C12orf76 | 0.036864874 | -0.039558489 | -0.002693616 |
| ARRDC1-AS1 | 0.028102488 | -0.031007899 | -0.002905411 |
| CHST15 | 0.01465232 | -0.013214369 | 0.001437951 |

| | | | |
|---|---|---|---|
| KIF1BP | 0.032604987 | -0.028037319 | 0.004567668 |
| RBM33 | 0.040300281 | -0.071146936 | -0.030846655 |
| AC073336.1 | 0.010999752 | -0.010999752 | -2.13163E-14 |
| CDC5L | 0.043806713 | -0.049137391 | -0.005330678 |
| TTC39B | 0.023006862 | -0.018177742 | 0.00482912 |
| RXYLT1 | 0.037044356 | -0.030077795 | 0.006966562 |
| LLGL1 | 0.01537736 | -0.015567826 | -0.000190466 |
| RAD17 | 0.037489015 | -0.033850457 | 0.003638558 |
| ULK1 | 0.029799892 | -0.039894317 | -0.010094426 |
| KHDRBS1 | 0.065258386 | -0.058802559 | 0.006455826 |
| CREBZF | 0.042893624 | -0.054579928 | -0.011686304 |
| WDR60 | 0.04557632 | -0.07122499 | -0.02564867 |
| CWC25 | 0.038401954 | -0.046624982 | -0.008223028 |
| SLC29A2 | 0.025887551 | -0.036087903 | -0.010200352 |
| GTF2E2 | 0.030765156 | -0.037299509 | -0.006534353 |
| FBXO9 | 0.051059337 | -0.089519393 | -0.038460056 |
| SNX14 | 0.042992853 | -0.070443802 | -0.027450949 |
| ENO3 | 0.022945137 | -0.022297135 | 0.000648002 |
| HP1BP3 | 0.066471009 | -0.105979535 | -0.039508526 |
| SERINC3 | 0.056263871 | -0.107930006 | -0.051666134 |
| FAM219B | 0.028931101 | -0.029366006 | -0.000434905 |
| BAG4 | 0.033765868 | -0.05745646 | -0.023690592 |
| ZNF567 | 0.030894157 | -0.041396491 | -0.010502334 |
| SAE1 | 0.023562779 | -0.030207708 | -0.006644929 |
| COPZ2 | 0.046421114 | -0.039682506 | 0.006738609 |
| COX19 | 0.035492596 | -0.042357844 | -0.006865248 |
| PANK3 | 0.035129733 | -0.031989309 | 0.003140423 |
| UNC45A | 0.034095622 | -0.038842439 | -0.004746817 |
| TRAPPC9 | 0.02498067 | -0.026257524 | -0.001276854 |
| MT1X | -0.071255394 | 0.181301696 | 0.110046301 |
| TANC1 | 0.029985706 | -0.038532256 | -0.008546549 |
| MATN3 | 0.015503812 | -0.017556537 | -0.002052725 |
| NCOR1 | 0.061296772 | -0.078113659 | -0.016816887 |
| AOPEP | 0.058065892 | -0.097274906 | -0.039209014 |
| HERC3 | 0.022632007 | -0.018960125 | 0.003671882 |
| WDR48 | 0.041493891 | -0.050742445 | -0.009248554 |
| MED8 | 0.03861709 | -0.030522386 | 0.008094705 |
| WDR47 | 0.019671455 | -0.017165195 | 0.00250626 |
| EPM2AIP1 | -0.050000992 | 0.039512141 | -0.010488851 |
| MED27 | 0.019080548 | -0.020619908 | -0.00153936 |
| CAMK2G | 0.02624465 | -0.021092171 | 0.005152479 |
| FIP1L1 | 0.042228879 | -0.03800173 | 0.004227149 |
| LINC01135 | 0.014297671 | -0.011278682 | 0.003018989 |
| LINC01137 | 0.018650817 | -0.020576953 | -0.001926136 |
| DCAF10 | 0.032957685 | -0.02592984 | 0.007027845 |
| SSH3 | 0.036463313 | -0.060173035 | -0.023709722 |
| EML2 | 0.030640398 | -0.032210039 | -0.00156964 |

| | | | |
|---|---|---|---|
| MOB3B | -0.025574798 | 0.024379391 | -0.001195407 |
| C1orf35 | 0.047649376 | -0.070640298 | -0.022990923 |
| MKRN2 | 0.027989865 | -0.027567802 | 0.000422063 |
| KCTD9 | 0.030396008 | -0.023572028 | 0.006823981 |
| IK | 0.058797755 | -0.06222731 | -0.003429554 |
| CUL1 | 0.036922021 | -0.031536921 | 0.0053851 |
| TMEM50B | 0.053519905 | -0.113281978 | -0.059762073 |
| KIDINS220 | 0.055331331 | -0.112322438 | -0.056991106 |
| TAL2 | 0.03244674 | -0.044525468 | -0.012078728 |
| AK3 | 0.05097978 | -0.042122929 | 0.008856852 |
| PLEKHH3 | 0.02605157 | -0.022483554 | 0.003568016 |
| WDR53 | 0.016077138 | -0.021091858 | -0.00501472 |
| ERP27 | -0.029240848 | 0.043244818 | 0.01400397 |
| FLVCR1 | 0.022209904 | -0.027412005 | -0.005202101 |
| XKR8 | 0.020455261 | -0.018922135 | 0.001533126 |
| DENND2A | 0.025315877 | -0.019885887 | 0.005429991 |
| ORAI2 | 0.049777481 | -0.062986943 | -0.013209462 |
| STOX1 | 0.015503739 | -0.013032728 | 0.002471011 |
| RASSF6 | -0.025517318 | 0.025366282 | -0.000151037 |
| MCOLN1 | 0.02846268 | -0.030343255 | -0.001880575 |
| PPP1R3F | 0.022833881 | -0.018180164 | 0.004653718 |
| NAXD | 0.027255941 | -0.032188897 | -0.004932956 |
| WDR20 | 0.038299405 | -0.041149 | -0.002849595 |
| NAAA | 0.041195053 | -0.082935636 | -0.041740583 |
| TMEM102 | 0.022605425 | -0.02996395 | -0.007358525 |
| MRPL10 | 0.039037363 | -0.033272691 | 0.005764672 |
| DHX40 | 0.039227068 | -0.043989729 | -0.004762661 |
| CLEC16A | 0.022975309 | -0.018481599 | 0.00449371 |
| TAF11 | 0.038584585 | -0.03520534 | 0.003379244 |
| PHACTR1 | -0.028811748 | 0.027543679 | -0.00126807 |
| AASDHPPT | 0.046199412 | -0.037417332 | 0.00878208 |
| NARF | 0.032801643 | -0.027795908 | 0.005005735 |
| CNOT6L | 0.045506477 | -0.039044555 | 0.006461922 |
| INO80B | 0.026088797 | -0.020280344 | 0.005808453 |
| AC016831.1 | 0.013638065 | -0.01729964 | -0.003661575 |
| ZUP1 | 0.021714908 | -0.017222377 | 0.004492531 |
| PRKCZ | 0.030671086 | -0.040461788 | -0.009790703 |
| CXADR | 0.052578832 | -0.083941343 | -0.031362511 |
| TMEM120B | 0.032652702 | -0.046463862 | -0.01381116 |
| MUS81 | 0.029456606 | -0.034979413 | -0.005522807 |
| HSPA5 | 0.064283197 | -0.116355639 | -0.052072442 |
| ANKRD39 | 0.028902179 | -0.034050817 | -0.005148638 |
| B4GALT3 | 0.03137517 | -0.044430601 | -0.013055431 |
| ACO1 | 0.034837235 | -0.027736035 | 0.0071012 |
| NHLRC2 | 0.035336359 | -0.044410768 | -0.009074409 |
| SERINC5 | -0.041540014 | -0.033195461 | -0.074735475 |
| DCLK1 | -0.016756036 | 0.016084932 | -0.000671104 |

| | | | |
|---|---|---|---|
| SMAD3 | -0.035419743 | 0.037553315 | 0.002133572 |
| C19orf12 | 0.025873481 | -0.033496908 | -0.007623427 |
| GLT8D1 | 0.047043037 | -0.060225346 | -0.013182308 |
| ATAD1 | 0.044277263 | -0.057303972 | -0.013026708 |
| TMEM177 | 0.026450978 | -0.028287621 | -0.001836643 |
| PECR | 0.045508246 | -0.035229681 | 0.010278565 |
| UBXN2B | 0.032263467 | -0.026866185 | 0.005397282 |
| BIRC6 | 0.052438572 | -0.080369551 | -0.02793098 |
| RNF43 | 0.041701347 | -0.061209158 | -0.019507812 |
| AP3B1 | 0.047413779 | -0.054788965 | -0.007375185 |
| TMEM234 | 0.032781998 | -0.039947605 | -0.007165607 |
| SEC24A | 0.025984005 | -0.025783769 | 0.000200237 |
| CLIP1 | 0.040467313 | -0.035973892 | 0.004493422 |
| DDX17 | -0.078389212 | -0.135763531 | -0.214152742 |
| LINC00642 | 0.009759814 | -0.009759814 | -1.95399E-14 |
| AC099552.1 | 0.009994701 | -0.009994701 | -2.13163E-14 |
| ZCCHC14 | -0.033390205 | 0.028274387 | -0.005115818 |
| CARMIL1 | 0.041909289 | -0.063114021 | -0.021204732 |
| ANO5 | 0.019974486 | -0.017661943 | 0.002312543 |
| TOR4A | 0.018654953 | -0.018411462 | 0.000243491 |
| TMEM237 | 0.031943755 | -0.044314471 | -0.012370715 |
| ZBTB44 | 0.060465497 | -0.117430354 | -0.056964857 |
| GTF2A1 | 0.033498598 | -0.037351665 | -0.003853066 |
| STAG2 | 0.058475795 | -0.089979506 | -0.031503711 |
| ANAPC15 | 0.045904786 | -0.038931488 | 0.006973298 |
| PPIB | 0.07613006 | -0.147399309 | -0.071269249 |
| PWWP2B | 0.030862838 | -0.033403291 | -0.002540453 |
| COQ10A | 0.026150644 | -0.034367977 | -0.008217333 |
| RBBP4 | 0.056316309 | -0.066352957 | -0.010036647 |
| PTPRD | -0.018463902 | 0.022269422 | 0.00380552 |
| REST | 0.05111781 | -0.065418837 | -0.014301027 |
| ING1 | 0.03066882 | -0.02796175 | 0.00270707 |
| M6PR | 0.056107874 | -0.098765303 | -0.042657428 |
| EPS15 | 0.036887816 | -0.037371699 | -0.000483883 |
| PGRMC2 | 0.052728928 | -0.095345291 | -0.042616363 |
| B3GALT4 | 0.025545952 | -0.034926319 | -0.009380367 |
| AC009403.1 | 0.022719191 | -0.020422807 | 0.002296384 |
| CD55 | 0.037132432 | -0.046512163 | -0.00937973 |
| DUSP8 | 0.014837499 | -0.01632535 | -0.001487851 |
| DCUN1D5 | 0.036221394 | -0.030206852 | 0.006014542 |
| SPAST | 0.02419699 | -0.029717651 | -0.005520661 |
| KCTD6 | 0.026634992 | -0.029717106 | -0.003082114 |
| LINC02361 | 0.012709239 | -0.015443902 | -0.002734663 |
| RAD50 | 0.052013971 | -0.063631454 | -0.011617484 |
| PLLP | 0.047069902 | -0.074809352 | -0.027739449 |
| HEXD | 0.039596199 | -0.039315219 | 0.000280979 |
| AC004846.1 | 0.011770262 | -0.011529033 | 0.000241228 |

| | | | |
|---|---|---|---|
| CDR2 | 0.03814663 | -0.031420857 | 0.006725773 |
| AC084740.1 | -0.013300467 | 0.010425021 | -0.002875447 |
| CRNDE | 0.073235127 | -0.155104466 | -0.08186934 |
| NACC2 | 0.021762421 | -0.020840458 | 0.000921962 |
| SMAD5 | 0.054440131 | -0.066112984 | -0.011672853 |
| TMEM168 | 0.038627951 | -0.042958707 | -0.004330755 |
| RANBP9 | 0.024486824 | -0.025985206 | -0.001498383 |
| AC016705.2 | 0.02071972 | -0.0268677 | -0.00614798 |
| COP1 | 0.023878867 | -0.020019467 | 0.003859401 |
| DOHH | 0.024284995 | -0.020067834 | 0.004217161 |
| PDLIM2 | -0.032916555 | 0.062076455 | 0.0291599 |
| CSE1L | 0.036120994 | -0.028675916 | 0.007445078 |
| DNAJC9 | 0.032618204 | -0.030838613 | 0.001779592 |
| MYZAP | 0.017243611 | -0.015307015 | 0.001936595 |
| CAMK1D | -0.016263022 | 0.014135713 | -0.002127309 |
| PPP6R2 | 0.049832131 | -0.075200731 | -0.0253686 |
| FUBP1 | 0.049578645 | -0.05665346 | -0.007074815 |
| DNM1L | 0.041684042 | -0.070548824 | -0.028864782 |
| NBAS | 0.035981722 | -0.045220888 | -0.009239166 |
| NUAK2 | 0.038272478 | -0.044360045 | -0.006087567 |
| FAM13B | 0.036483893 | -0.050321901 | -0.013838008 |
| PITPNA | 0.038170294 | -0.036252351 | 0.001917942 |
| TP53I13 | 0.045227328 | -0.047933393 | -0.002706065 |
| PIH1D1 | 0.04583761 | -0.050017177 | -0.004179566 |
| RHBDD2 | 0.040509321 | -0.034732516 | 0.005776804 |
| CNEP1R1 | 0.026269224 | -0.034125564 | -0.00785634 |
| KIFC2 | 0.022377233 | -0.01964573 | 0.002731504 |
| APH1A | 0.057348084 | -0.102755711 | -0.045407626 |
| ANKS6 | 0.032066037 | -0.04221484 | -0.010148803 |
| FAM126B | 0.036624092 | -0.047733999 | -0.011109907 |
| TRPV4 | 0.012186542 | -0.011492869 | 0.000693672 |
| ELF2 | 0.041575765 | -0.046102241 | -0.004526476 |
| RBM48 | 0.026316359 | -0.020723792 | 0.005592566 |
| RAB35 | 0.02624542 | -0.035217274 | -0.008971855 |
| CLDN12 | 0.025051807 | -0.02336217 | 0.001689637 |
| LYRM7 | 0.02610688 | -0.027507244 | -0.001400363 |
| KCP | 0.031741701 | -0.036305427 | -0.004563727 |
| ATP6AP1 | 0.060316128 | -0.087554436 | -0.027238308 |
| CCN2 | -0.050981501 | 0.052891517 | 0.001910016 |
| OSBPL11 | 0.029465197 | -0.024145806 | 0.00531939 |
| INPP5F | 0.029892696 | -0.036415436 | -0.006522741 |
| OGDHL | -0.028327278 | 0.046950057 | 0.018622779 |
| DNASE1 | 0.029019507 | -0.037701573 | -0.008682066 |
| NEK5 | -0.020326966 | 0.02336873 | 0.003041764 |
| MED24 | 0.025517051 | -0.020423319 | 0.005093733 |
| MBD4 | 0.045554657 | -0.076358302 | -0.030803645 |
| DPAGT1 | 0.027903584 | -0.023503875 | 0.004399709 |

| | | | |
|---|---|---|---|
| PRORP | 0.029582656 | -0.031469289 | -0.001886633 |
| ERFE | 0.008977427 | -0.007771433 | 0.001205994 |
| ICE1 | 0.042679454 | -0.067340142 | -0.024660689 |
| PPHLN1 | 0.040665779 | -0.046195544 | -0.005529765 |
| MMGT1 | 0.034983112 | -0.050008203 | -0.01502509 |
| BMS1 | 0.036513738 | -0.038023239 | -0.001509501 |
| MYO7A | 0.008335391 | -0.008335391 | -2.30926E-14 |
| BRCC3 | 0.030296737 | -0.035269172 | -0.004972435 |
| SEMA5B | 0.012181641 | -0.01205816 | 0.000123482 |
| AL022068.1 | -0.017237435 | 0.021826431 | 0.004588996 |
| FOXK2 | 0.027773823 | -0.022202062 | 0.005571761 |
| HNRNPD | 0.063756236 | -0.084413705 | -0.020657469 |
| AC025271.4 | 0.008466537 | -0.008466537 | -2.13163E-14 |
| TMEM189 | 0.025464199 | -0.031779175 | -0.006314976 |
| MFSD3 | 0.027747308 | -0.038867685 | -0.011120378 |
| SDR39U1 | 0.047027278 | -0.04952931 | -0.002502032 |
| DLGAP1-AS2 | 0.007881083 | -0.007881083 | -2.13163E-14 |
| NNT | 0.040892189 | -0.04317605 | -0.002283861 |
| AC114316.1 | 0.008614838 | -0.008614838 | -2.13163E-14 |
| ATXN2 | 0.040097599 | -0.064912109 | -0.024814511 |
| HOXB-AS3 | 0.041487968 | -0.047576785 | -0.006088817 |
| CHST14 | 0.032756137 | -0.037063237 | -0.004307101 |
| NKTR | -0.058147076 | -0.048762878 | -0.106909954 |
| SLC45A1 | 0.008545485 | -0.008545485 | -1.95399E-14 |
| VPS37B | 0.026192647 | -0.02499735 | 0.001195297 |
| SMC1A | 0.038889829 | -0.041829086 | -0.002939257 |
| PYGB | 0.035758607 | -0.038213792 | -0.002455185 |
| CCDC153 | 0.008237686 | -0.008237686 | -2.13163E-14 |
| TBC1D10A | 0.025208664 | -0.02596929 | -0.000760626 |
| TMEM129 | 0.035717511 | -0.029507801 | 0.00620971 |
| HEATR3 | 0.016834588 | -0.015926012 | 0.000908576 |
| MIR4435-2HG | -0.028715466 | 0.029649277 | 0.000933811 |
| MTMR6 | 0.028882939 | -0.023492737 | 0.005390202 |
| LPP | -0.051294016 | 0.046553011 | -0.004741005 |
| MED30 | 0.034691312 | -0.035138259 | -0.000446947 |
| FAM172A | 0.036615864 | -0.060912803 | -0.024296939 |
| NADSYN1 | 0.022897209 | -0.019030788 | 0.003866421 |
| TSPAN6 | 0.05244835 | -0.089450421 | -0.03700207 |
| EFCAB11 | 0.022890635 | -0.033622441 | -0.010731806 |
| UIMC1 | 0.027631298 | -0.033723607 | -0.006092308 |
| MARCH7 | 0.043147097 | -0.05207485 | -0.008927753 |
| GORAB | 0.017487991 | -0.023376093 | -0.005888102 |
| RARG | 0.027437503 | -0.036116167 | -0.008678664 |
| PCOTH | 0.010754264 | -0.010596834 | 0.00015743 |
| CNPY4 | 0.02330271 | -0.020093864 | 0.003208845 |
| SLC7A6OS | 0.026398248 | -0.022313417 | 0.004084831 |
| ATF6 | 0.046253312 | -0.075502837 | -0.029249524 |

| | | | |
|---|---|---|---|
| AC007036.3 | 0.008939401 | -0.008939401 | -2.13163E-14 |
| TADA3 | 0.037790405 | -0.039270558 | -0.001480152 |
| TMPRSS2 | 0.040028334 | -0.073789322 | -0.033760988 |
| MFAP3L | 0.042755345 | -0.087011442 | -0.044256096 |
| GGA1 | 0.033191011 | -0.041386714 | -0.008195703 |
| LNP1 | -0.016763533 | 0.018835879 | 0.002072346 |
| CCDC93 | 0.029973494 | -0.035755575 | -0.005782081 |
| AL161431.1 | 0.008195822 | -0.008195822 | -1.95399E-14 |
| LEPROT | 0.075019673 | -0.23885318 | -0.163833507 |
| FAM91A1 | 0.03117336 | -0.029772967 | 0.001400393 |
| MRPL20-AS1 | 0.026102915 | -0.034630451 | -0.008527536 |
| MSH6 | 0.033001965 | -0.049525482 | -0.016523517 |
| CNOT3 | 0.029492376 | -0.028364992 | 0.001127384 |
| AC092894.1 | -0.030086185 | 0.033001619 | 0.002915434 |
| PUS7 | 0.019675697 | -0.016946082 | 0.002729614 |
| FCHSD2 | 0.021315403 | -0.024313685 | -0.002998281 |
| ST7L | 0.031798195 | -0.029904556 | 0.001893639 |
| RAB5A | 0.03675084 | -0.046567874 | -0.009817035 |
| NR2C1 | 0.030275291 | -0.033761744 | -0.003486453 |
| AC011043.1 | 0.03591169 | -0.075868645 | -0.039956955 |
| DAB2 | -0.057447462 | 0.10288386 | 0.045436397 |
| ZBED6CL | 0.013540467 | -0.012662757 | 0.00087771 |
| ADAMTSL2 | 0.011078988 | -0.010627927 | 0.000451061 |
| SERAC1 | 0.02206564 | -0.027433573 | -0.005367933 |
| CETN2 | 0.05698662 | -0.051223504 | 0.005763116 |
| VMP1 | 0.04701509 | -0.069793413 | -0.022778323 |
| SPIRE1 | 0.022882787 | -0.020485268 | 0.002397519 |
| PHF6 | 0.045345651 | -0.070474158 | -0.025128508 |
| STMN3 | -0.020631381 | 0.019625873 | -0.001005508 |
| LMO2 | 0.013909609 | -0.013332162 | 0.000577448 |
| PRKAG1 | 0.032710106 | -0.029192487 | 0.003517619 |
| WTIP | 0.03412576 | -0.027953833 | 0.006171927 |
| FNBP1 | 0.03411003 | -0.035445365 | -0.001335335 |
| MFSD14A | 0.037985605 | -0.047821403 | -0.009835798 |
| LINC01687 | 0.00837256 | -0.00837256 | -2.13163E-14 |
| NAA16 | 0.0281779 | -0.036329253 | -0.008151353 |
| CRPPA | 0.020889674 | -0.017846893 | 0.003042782 |
| NCKAP1 | 0.052821099 | -0.090806316 | -0.037985217 |
| SETD4 | 0.028719751 | -0.023829494 | 0.004890257 |
| DNAJC11 | 0.031612767 | -0.046138076 | -0.014525309 |
| IBTK | 0.037633003 | -0.060941793 | -0.02330879 |
| TMF1 | 0.051049768 | -0.107972343 | -0.056922576 |
| GDI1 | 0.04165734 | -0.049181966 | -0.007524626 |
| USP12 | 0.034536782 | -0.042684214 | -0.008147432 |
| STN1 | 0.024228055 | -0.035076851 | -0.010848797 |
| AC092118.1 | 0.01238055 | -0.010431741 | 0.00194881 |
| TOB1 | 0.04908401 | -0.085917915 | -0.036833905 |

| | | | |
|---|---|---|---|
| RRAGD | -0.024559433 | 0.023918601 | -0.000640832 |
| USP7 | 0.044489816 | -0.061093175 | -0.01660336 |
| DNTTIP1 | 0.032402103 | -0.027700472 | 0.004701631 |
| C12orf43 | 0.028788658 | -0.027342363 | 0.001446295 |
| LINC00488 | 0.01285703 | -0.01623086 | -0.00337383 |
| RIN2 | -0.039673259 | 0.036290803 | -0.003382457 |
| LDOC1 | 0.034653437 | -0.037211687 | -0.00255825 |
| NR1D1 | 0.021373045 | -0.018976507 | 0.002396538 |
| RFX1 | 0.033125034 | -0.045106635 | -0.011981601 |
| WWC2 | 0.056216997 | -0.121276096 | -0.065059098 |
| CTTNBP2NL | 0.031618083 | -0.028102898 | 0.003515185 |
| CD200 | -0.029084671 | 0.02760005 | -0.001484621 |
| PRRC1 | 0.032481148 | -0.042436927 | -0.009955778 |
| PIP5KL1 | 0.01445884 | -0.016674118 | -0.002215278 |
| PLSCR1 | 0.041260929 | -0.039740617 | 0.001520313 |
| SLC37A1 | 0.018764741 | -0.020128735 | -0.001363994 |
| EDEM3 | 0.032715378 | -0.05326099 | -0.020545613 |
| MPHOSPH6 | 0.022994407 | -0.030087448 | -0.007093041 |
| ABCB7 | 0.023404473 | -0.025648867 | -0.002244395 |
| NOL10 | 0.027092937 | -0.030518293 | -0.003425356 |
| RAB11FIP2 | 0.043207752 | -0.044817675 | -0.001609923 |
| SEC22B | 0.048086278 | -0.040233578 | 0.0078527 |
| BECN1 | 0.034538171 | -0.028523875 | 0.006014295 |
| HACD4 | 0.023984162 | -0.034087949 | -0.010103786 |
| RPL7L1 | 0.050518798 | -0.069671458 | -0.01915266 |
| ARHGAP8 | -0.032266276 | 0.030898633 | -0.001367643 |
| ENTPD6 | 0.030984034 | -0.037362473 | -0.006378439 |
| SLC39A4 | 0.040661767 | -0.039396278 | 0.001265489 |
| RAD1 | 0.028206881 | -0.025398435 | 0.002808446 |
| GTPBP4 | 0.030479109 | -0.033415033 | -0.002935925 |
| AIDA | 0.033738979 | -0.031915251 | 0.001823729 |
| BRI3BP | 0.024699373 | -0.029672444 | -0.004973071 |
| OFD1 | 0.044444432 | -0.064959546 | -0.020515114 |
| SGTB | 0.013166995 | -0.011392799 | 0.001774196 |
| MSH2 | 0.029605542 | -0.038132016 | -0.008526473 |
| EVI5 | 0.033714346 | -0.03578753 | -0.002073183 |
| AMT | 0.031706648 | -0.031719126 | -1.24774E-05 |
| ELK4 | 0.03562909 | -0.031721991 | 0.003907099 |
| KIAA1549L | 0.010489411 | -0.011012744 | -0.000523333 |
| VSIR | 0.021832042 | -0.02084137 | 0.000990672 |
| ZNF322 | 0.035989395 | -0.055572098 | -0.019582703 |
| ORMDL3 | 0.025090805 | -0.035282139 | -0.010191334 |
| GRK2 | 0.03937551 | -0.056672608 | -0.017297098 |
| ACIN1 | 0.046224233 | -0.062631804 | -0.016407571 |
| ACSM3 | -0.025434254 | 0.037377364 | 0.01194311 |
| GSTM5 | 0.008963175 | -0.007930417 | 0.001032758 |
| FUT8 | 0.028816392 | -0.0263458 | 0.002470592 |

| | | | |
|---|---|---|---|
| HAUS8 | 0.01314457 | -0.014839496 | -0.001694926 |
| AP1G1 | 0.046031153 | -0.101102852 | -0.055071699 |
| CYBRD1 | 0.024491495 | -0.028614929 | -0.004123434 |
| TRPC4AP | 0.033717398 | -0.061669382 | -0.027951984 |
| COPS7B | 0.032244937 | -0.036432962 | -0.004188025 |
| HECTD1 | 0.051334021 | -0.055464251 | -0.004130231 |
| QRSL1 | 0.026746884 | -0.030187015 | -0.003440131 |
| MCF2L2 | 0.009552996 | -0.01073747 | -0.001184474 |
| S1PR3 | 0.010547078 | -0.010650114 | -0.000103036 |
| HLA-G | 0.007570182 | -0.007570182 | -1.95399E-14 |
| IER2 | -0.067155089 | 0.055934022 | -0.011221066 |
| AL162258.2 | 0.007395457 | -0.007395457 | -1.95399E-14 |
| PMPCA | 0.030005807 | -0.025219515 | 0.004786292 |
| ZNF92 | 0.021779031 | -0.024233488 | -0.002454457 |
| PPM1J | 0.011341952 | -0.01305428 | -0.001712328 |
| AL591848.4 | 0.007346839 | -0.007346839 | -1.95399E-14 |
| AC093690.1 | 0.00701321 | -0.00701321 | -2.13163E-14 |
| PTPN4 | 0.029395021 | -0.035342868 | -0.005947847 |
| GPSM3 | 0.010396777 | -0.008678241 | 0.001718536 |
| CSRNP1 | 0.017490736 | -0.016965321 | 0.000525415 |
| FAM240A | 0.007646957 | -0.007646957 | -2.13163E-14 |
| SGTA | 0.037041392 | -0.031550345 | 0.005491047 |
| SNTG2 | 0.007623563 | -0.007623563 | -2.13163E-14 |
| FBLIM1 | 0.028623618 | -0.030691833 | -0.002068215 |
| AC084759.2 | 0.008195626 | -0.008195626 | -2.13163E-14 |
| TGFB1 | 0.025528381 | -0.028047418 | -0.002519037 |
| KIAA0232 | 0.043929918 | -0.056195925 | -0.012266007 |
| LINC01991 | 0.008216615 | -0.008216615 | -2.13163E-14 |
| MICU1 | 0.035606537 | -0.04338708 | -0.007780543 |
| CDC27 | 0.030581014 | -0.035878435 | -0.005297421 |
| NPRL2 | 0.027238616 | -0.02420904 | 0.003029576 |
| PKNOX1 | 0.024538994 | -0.027931087 | -0.003392094 |
| ERBIN | 0.047669609 | -0.07866101 | -0.030991401 |
| AC136475.3 | 0.012654157 | -0.013098571 | -0.000444414 |
| AC100810.3 | 0.007855838 | -0.007855838 | -2.13163E-14 |
| SDF4 | 0.049363208 | -0.111266901 | -0.061903693 |
| SAFB2 | 0.043265955 | -0.057413261 | -0.014147306 |
| PYCR2 | 0.028719195 | -0.027082439 | 0.001636756 |
| TBC1D5 | 0.038621477 | -0.044416297 | -0.00579482 |
| ARMCX4 | 0.023979876 | -0.028135 | -0.004155124 |
| NIF3L1 | 0.024918871 | -0.022961689 | 0.001957182 |
| TMEM98 | -0.026353414 | 0.054478236 | 0.028124822 |
| FBP1 | 0.030846193 | 0.05994967 | 0.090795863 |
| FMN1 | 0.008687301 | -0.00963297 | -0.000945669 |
| FBXO48 | 0.017229915 | -0.015308786 | 0.001921129 |
| ZNHIT2 | 0.027182327 | -0.033416443 | -0.006234117 |
| AL031710.1 | 0.013759979 | -0.011622498 | 0.002137481 |

| | | | |
|---|---|---|---|
| LCORL | 0.028600793 | -0.03636646 | -0.007765667 |
| ACTR6 | 0.037056599 | -0.057106352 | -0.020049753 |
| BRD8 | 0.037875841 | -0.050603106 | -0.012727265 |
| SLC25A21-AS1 | -0.021428705 | 0.024162339 | 0.002733634 |
| SWSAP1 | 0.022535056 | -0.019588156 | 0.0029469 |
| RAP1A | 0.048797599 | -0.056958097 | -0.008160498 |
| EPB41 | 0.02681756 | -0.044129357 | -0.017311797 |
| L1CAM | 0.020829801 | -0.037094895 | -0.016265094 |
| PRICKLE3 | 0.012608646 | -0.011099058 | 0.001509589 |
| FOSL2 | 0.040421579 | -0.054251774 | -0.013830194 |
| UBE2T | 0.012484935 | -0.016681569 | -0.004196634 |
| YARS2 | 0.022106676 | -0.031858751 | -0.009752075 |
| RC3H2 | 0.03220634 | -0.035461854 | -0.003255514 |
| TFAP2B | 0.031070504 | -0.06424625 | -0.033175746 |
| TCEAL1 | 0.033153089 | -0.040647303 | -0.007494213 |
| NTN4 | 0.068844871 | -0.134446481 | -0.06560161 |
| KIAA0100 | 0.035372071 | -0.048793007 | -0.013420936 |
| LEPR | 0.017771119 | -0.016136957 | 0.001634161 |
| FUT1 | 0.009152768 | -0.010646578 | -0.00149381 |
| ZNF318 | 0.023614745 | -0.038048521 | -0.014433776 |
| BX255925.3 | 0.018246658 | -0.020626927 | -0.002380269 |
| HMGN5 | 0.029676556 | -0.033019384 | -0.003342828 |
| C20orf194 | 0.032104456 | -0.049049638 | -0.016945182 |
| NAMPT | 0.041163817 | -0.053364952 | -0.012201135 |
| ETV6 | 0.019925043 | -0.019399586 | 0.000525457 |
| TOR1A | 0.029966494 | -0.04433721 | -0.014370716 |
| HTRA2 | 0.029775142 | -0.038331701 | -0.00855656 |
| SART1 | 0.025714132 | -0.022913499 | 0.002800633 |
| SFTA1P | 0.007667271 | -0.007667271 | -1.95399E-14 |
| FSTL3 | -0.019040661 | 0.020582923 | 0.001542262 |
| LIX1L | 0.015758685 | -0.014951848 | 0.000806837 |
| RAB39B | 0.007852526 | -0.007041874 | 0.000810651 |
| NRDC | 0.042113459 | -0.075181206 | -0.033067748 |
| KBTBD4 | 0.020866365 | -0.024485159 | -0.003618794 |
| KLHDC2 | 0.048644666 | -0.044985559 | 0.003659107 |
| H2AFY | 0.05378013 | -0.095773686 | -0.041993556 |
| RIOK1 | 0.021124472 | -0.027008214 | -0.005883743 |
| TMEM170A | 0.041245351 | -0.063219553 | -0.021974202 |
| SEC23B | 0.032123145 | -0.03665405 | -0.004530905 |
| ZBTB17 | 0.019449425 | -0.020133549 | -0.000684124 |
| CACFD1 | 0.025237655 | -0.028277839 | -0.003040184 |
| TNK1 | 0.018367201 | -0.026839007 | -0.008471806 |
| MIDN | -0.035574227 | 0.036405242 | 0.000831016 |
| ALG8 | 0.025715521 | -0.028402046 | -0.002686526 |
| CXCL16 | 0.027310344 | -0.042554032 | -0.015243688 |
| ARL4C | -0.023640886 | 0.023969179 | 0.000328294 |
| ACTR8 | 0.021043686 | -0.030699293 | -0.009655608 |

| | | | |
|---|---|---|---|
| CITED4 | -0.034921286 | 0.066747034 | 0.031825748 |
| EDN2 | -0.019900662 | 0.034166277 | 0.014265615 |
| YWHAG | 0.035594001 | -0.031931541 | 0.00366246 |
| PNISR | -0.065927453 | -0.103158718 | -0.169086171 |
| CDC42BPB | 0.043072769 | -0.05954551 | -0.016472741 |
| IMPA1 | 0.028530068 | -0.035395382 | -0.006865314 |
| EOGT | 0.011454752 | -0.011068009 | 0.000386743 |
| SCRN2 | 0.033198241 | -0.02988273 | 0.003315511 |
| MON1A | 0.01471345 | -0.017398225 | -0.002684774 |
| CNOT4 | 0.042718524 | -0.075158923 | -0.032440399 |
| CSKMT | 0.030341068 | -0.051741925 | -0.021400857 |
| BAZ1A | 0.027001424 | -0.036305842 | -0.009304418 |
| VKORC1 | 0.059650453 | -0.08469367 | -0.025043218 |
| TFB1M | 0.023575425 | -0.029150481 | -0.005575056 |
| C5 | -0.020514249 | 0.03230781 | 0.011793561 |
| DLG1 | 0.041003185 | -0.067264362 | -0.026261178 |
| JAK2 | 0.013460187 | -0.012899852 | 0.000560335 |
| KBTBD3 | 0.025935055 | -0.030377513 | -0.004442458 |
| USP53 | 0.043455082 | -0.055770277 | -0.012315195 |
| CSRNP3 | 0.023682494 | -0.035695833 | -0.012013339 |
| PPTC7 | 0.021198013 | -0.024445928 | -0.003247916 |
| TYSND1 | 0.021823531 | -0.027396264 | -0.005572733 |
| USF3 | 0.033122728 | -0.045592892 | -0.012470165 |
| CDKN1B | 0.037988045 | -0.054587825 | -0.01659978 |
| ACAP3 | 0.019821418 | -0.026998487 | -0.007177069 |
| FAT3 | 0.032750493 | -0.071370913 | -0.03862042 |
| PCIF1 | 0.026720189 | -0.024301069 | 0.00241912 |
| STK17B | 0.020951828 | -0.026367555 | -0.005415727 |
| TSEN54 | 0.024596542 | -0.041354965 | -0.016758423 |
| C1orf56 | 0.044659034 | -0.084816573 | -0.040157539 |
| RABL3 | 0.025961103 | -0.02560218 | 0.000358923 |
| OSBPL9 | 0.045972551 | -0.072408196 | -0.026435644 |
| DDX56 | 0.029292742 | -0.027789638 | 0.001503104 |
| STX17-AS1 | 0.018861539 | -0.020380413 | -0.001518875 |
| ITGB8 | 0.059049563 | -0.136433933 | -0.07738437 |
| OTUD3 | 0.020978972 | -0.02702839 | -0.006049417 |
| ETNK1 | 0.045362639 | -0.084296351 | -0.038933711 |
| R3HDM1 | 0.022949423 | -0.030267153 | -0.007317731 |
| EMC2 | 0.044411421 | -0.043704219 | 0.000707201 |
| CYTH1 | 0.026402266 | -0.040115831 | -0.013713564 |
| CAV1 | -0.021123591 | 0.024523848 | 0.003400258 |
| EIF2AK2 | 0.043554854 | -0.052702358 | -0.009147504 |
| HOXD8 | 0.043073304 | -0.054283825 | -0.011210521 |
| MIR99AHG | 0.055008752 | -0.131073063 | -0.07606431 |
| PCBD2 | 0.022203648 | -0.019609876 | 0.002593773 |
| UBXN7 | 0.033600127 | -0.040099088 | -0.006498961 |
| L1TD1 | 0.006375019 | -0.006375019 | -2.30926E-14 |

| | | | |
|---|---|---|---|
| RASA1 | 0.025477888 | -0.027883847 | -0.002405959 |
| ERLIN2 | 0.038285288 | -0.073856606 | -0.035571318 |
| CAMTA2 | 0.026347187 | -0.039355275 | -0.013008088 |
| IRF2BP1 | 0.028822408 | -0.029360472 | -0.000538064 |
| AL023581.2 | 0.00595468 | -0.00595468 | -2.30926E-14 |
| CAHM | 0.014964269 | -0.021024829 | -0.00606056 |
| ABHD16A | 0.037938338 | -0.04558805 | -0.007649712 |
| RIPPLY3 | 0.008065584 | -0.008978869 | -0.000913286 |
| FKBP9 | 0.038373589 | -0.041003465 | -0.002629876 |
| SYT1 | 0.006563898 | -0.006563898 | -2.13163E-14 |
| CCNG2 | 0.045394496 | -0.051478945 | -0.006084449 |
| OS9 | 0.046559716 | -0.114542337 | -0.067982621 |
| AMOTL2 | 0.047002144 | -0.092093268 | -0.045091124 |
| CADM2 | 0.007112469 | -0.007112469 | -2.30926E-14 |
| MHENCR | 0.027914385 | -0.043037699 | -0.015123314 |
| AC093249.6 | 0.014407817 | -0.014677883 | -0.000270066 |
| HPCAL1 | 0.037783582 | -0.03780417 | -2.05872E-05 |
| ANKRD18B | 0.016115101 | -0.016178541 | -6.34404E-05 |
| TUBGCP2 | 0.046047336 | -0.061641637 | -0.015594301 |
| RRS1 | 0.016272941 | -0.017329447 | -0.001056506 |
| THAP12 | 0.03107872 | -0.042659492 | -0.011580773 |
| RNF26 | 0.025793458 | -0.033620257 | -0.007826799 |
| ZNF576 | 0.023174449 | -0.022546662 | 0.000627787 |
| SNAPC3 | 0.02466001 | -0.038832892 | -0.014172882 |
| MAVS | 0.03191766 | -0.045934806 | -0.014017147 |
| LINC01578 | 0.062713379 | -0.064287572 | -0.001574194 |
| LYRM1 | 0.037108761 | -0.033816235 | 0.003292526 |
| SMC4 | 0.024301819 | -0.038124409 | -0.01382259 |
| BCO2 | 0.012016432 | -0.011221635 | 0.000794797 |
| SPAG4 | 0.007791035 | -0.007791035 | -1.95399E-14 |
| TMEM238L | 0.006079995 | -0.006079995 | -1.95399E-14 |
| POGLUT1 | 0.028959955 | -0.032587461 | -0.003627507 |
| LINC02484 | 0.007317765 | -0.007317765 | -1.95399E-14 |
| AC099552.5 | 0.007023623 | -0.007023623 | -2.13163E-14 |
| USP45 | 0.019052173 | -0.023392472 | -0.0043403 |
| ZNF568 | 0.019945581 | -0.024166594 | -0.004221013 |
| TXLNA | 0.030802704 | -0.042995341 | -0.012192637 |
| CAPN14 | 0.007632398 | -0.007632398 | -2.13163E-14 |
| SLC6A13 | -0.035299778 | 0.112603382 | 0.077303604 |
| ZFP1 | 0.015268062 | -0.013678239 | 0.001589823 |
| ECD | 0.02456635 | -0.026576886 | -0.002010536 |
| ABTB1 | 0.027412831 | -0.033390054 | -0.005977224 |
| SAR1A | 0.04466216 | -0.046407 | -0.001744839 |
| CAPN15 | 0.022434643 | -0.023304075 | -0.000869431 |
| SMG5 | 0.026246764 | -0.025362958 | 0.000883806 |
| RILP | 0.018452551 | -0.021218365 | -0.002765814 |
| E2F6 | 0.019590927 | -0.029569153 | -0.009978226 |

| | | | |
|---|---|---|---|
| PTPRA | 0.050859311 | -0.09281189 | -0.041952579 |
| SPTLC1 | 0.039641284 | -0.067304111 | -0.027662827 |
| POLK | 0.033938602 | -0.050641737 | -0.016703134 |
| CIAO3 | 0.024363443 | -0.028821105 | -0.004457662 |
| AC023421.1 | 0.00576669 | -0.00576669 | -2.13163E-14 |
| CRIM1 | -0.068320507 | -0.163333513 | -0.23165402 |
| DNTTIP2 | 0.035823506 | -0.06328435 | -0.027460844 |
| SPTY2D1 | 0.031738733 | -0.056266166 | -0.024527432 |
| PDPK1 | 0.029791617 | -0.040819522 | -0.011027905 |
| ZC3H14 | 0.038223379 | -0.063906834 | -0.025683454 |
| C5orf24 | 0.043114277 | -0.048085614 | -0.004971336 |
| TAZ | 0.027495194 | -0.036544541 | -0.009049347 |
| RABGAP1L | 0.02749278 | -0.031602313 | -0.004109533 |
| RBPJ | 0.040267264 | -0.061162972 | -0.020895709 |
| SPATA6 | 0.02930052 | -0.038898676 | -0.009598155 |
| PLAA | 0.025510007 | -0.030069266 | -0.00455926 |
| AC006449.6 | 0.015692509 | -0.022996483 | -0.007303975 |
| IGFBP5 | 0.075655025 | -0.282536504 | -0.206881479 |
| CDH13 | 0.006289385 | -0.006289385 | -2.30926E-14 |
| LINC02298 | 0.019315783 | -0.020310354 | -0.000994571 |
| LEO1 | 0.024591311 | -0.03085511 | -0.006263798 |
| SLC4A1AP | 0.024980781 | -0.034247246 | -0.009266465 |
| THAP1 | 0.026156904 | -0.024625569 | 0.001531335 |
| DKK1 | 0.006954628 | -0.006954628 | -2.13163E-14 |
| ARID4B | 0.047939022 | -0.126817282 | -0.07887826 |
| PHKB | 0.035555502 | -0.043779532 | -0.00822403 |
| KIAA1324L | 0.00890942 | -0.010469204 | -0.001559784 |
| PRKAA1 | 0.035077364 | -0.044934158 | -0.009856794 |
| OGFR | 0.03430467 | -0.031808215 | 0.002496456 |
| USP24 | 0.031717797 | -0.036223204 | -0.004505407 |
| CU638689.4 | -0.013227107 | 0.018391488 | 0.005164381 |
| ATXN2L | 0.034787549 | -0.042158668 | -0.007371119 |
| AC006273.1 | 0.01860705 | -0.018469652 | 0.000137397 |
| KEAP1 | 0.024736323 | -0.033027094 | -0.008290771 |
| PAXBP1 | 0.036243021 | -0.052668213 | -0.016425192 |
| NPHP1 | 0.025361722 | -0.031284427 | -0.005922705 |
| THOC5 | 0.018046749 | -0.020193685 | -0.002146935 |
| C1GALT1 | 0.029869626 | -0.02784521 | 0.002024416 |
| FAAP100 | 0.017108291 | -0.016918039 | 0.000190252 |
| ZDHHC20 | 0.023145652 | -0.029245979 | -0.006100327 |
| ZNHIT6 | 0.028697618 | -0.029462043 | -0.000764425 |
| GOSR2 | 0.028345384 | -0.035263926 | -0.006918542 |
| PAXIP1 | 0.019697583 | -0.018750111 | 0.000947472 |
| RNASEH2B | 0.033077937 | -0.032472206 | 0.000605731 |
| UBE2G2 | 0.037753736 | -0.055606223 | -0.017852487 |
| CLN3 | 0.04187799 | -0.071807484 | -0.029929493 |
| MYMK | 0.007958478 | -0.007958478 | -2.13163E-14 |

| | | | |
|---|---|---|---|
| HEG1 | 0.031977602 | -0.067084428 | -0.035106826 |
| SLC16A3 | 0.009823765 | -0.011215851 | -0.001392086 |
| ANKRD11 | 0.047169712 | -0.079358438 | -0.032188726 |
| AJUBA | 0.03726838 | -0.049028353 | -0.011759973 |
| NME6 | 0.021170894 | -0.023068911 | -0.001898017 |
| PHF20 | 0.032151936 | -0.03486105 | -0.002709114 |
| MAN2A2 | 0.023326833 | -0.027324902 | -0.00399807 |
| SECISBP2L | 0.038643522 | -0.05642204 | -0.017778518 |
| MTIF2 | 0.024700029 | -0.023539163 | 0.001160866 |
| CNST | 0.023065406 | -0.031339231 | -0.008273825 |
| RHOT2 | 0.033487848 | -0.030976614 | 0.002511235 |
| PHRF1 | 0.025584628 | -0.031715827 | -0.006131199 |
| FOCAD | 0.023705754 | -0.024640162 | -0.000934408 |
| IRAK4 | 0.02366369 | -0.023536455 | 0.000127234 |
| PGAM2 | 0.013473049 | -0.014479289 | -0.00100624 |
| ABI2 | 0.042519195 | -0.072174154 | -0.02965496 |
| C2orf49 | 0.03550253 | -0.057364741 | -0.021862211 |
| NAGLU | 0.026565289 | -0.025726381 | 0.000838908 |
| MPP2 | 0.007630346 | -0.008719729 | -0.001089383 |
| RHOT1 | 0.028114244 | -0.037445466 | -0.009331222 |
| TMEM87B | 0.022140144 | -0.030037675 | -0.007897532 |
| RNF44 | 0.021840809 | -0.021901665 | -6.08562E-05 |
| MAPKBP1 | 0.012777803 | -0.014229637 | -0.001451834 |
| DDX50 | 0.033222382 | -0.038344318 | -0.005121936 |
| SLC30A9 | 0.033263199 | -0.045862544 | -0.012599345 |
| SUV39H2 | 0.011362385 | -0.012507358 | -0.001144973 |
| TMEM175 | 0.026158514 | -0.034506985 | -0.008348471 |
| AP2B1 | 0.045798087 | -0.06685187 | -0.021053783 |
| AC091488.1 | 0.00868487 | -0.010598553 | -0.001913683 |
| KLF3 | 0.033735708 | -0.034423316 | -0.000687607 |
| MCF2L | 0.029921322 | -0.040079502 | -0.010158181 |
| MAN1C1 | 0.013357698 | -0.015446137 | -0.002088439 |
| AC124852.1 | 0.008129534 | -0.009270384 | -0.00114085 |
| LRRC75B | -0.016367516 | 0.022391487 | 0.006023972 |
| CERT1 | 0.035131662 | -0.044884279 | -0.009752617 |
| BCL11B | -0.011989389 | 0.015752628 | 0.003763239 |
| BRIX1 | 0.032663779 | -0.042006004 | -0.009342225 |
| LIG4 | 0.018336738 | -0.030205148 | -0.01186841 |
| PHGDH | -0.047813239 | 0.086085749 | 0.03827251 |
| SAMD8 | 0.018957245 | -0.033014178 | -0.014056934 |
| SLC46A1 | 0.021226217 | -0.030263588 | -0.009037371 |
| EIF4G1 | 0.041183092 | -0.048358348 | -0.007175256 |
| ITGB3 | -0.020392713 | 0.018919361 | -0.001473352 |
| PIGR | -0.035229245 | 0.042474063 | 0.007244818 |
| FAM53C | 0.021689049 | -0.027188473 | -0.005499424 |
| AP005329.3 | 0.012225053 | -0.011767508 | 0.000457545 |
| CARS | 0.01711363 | -0.018240056 | -0.001126426 |

| | | | |
|---|---|---|---|
| CTCF | 0.036978102 | -0.055989124 | -0.019011021 |
| ITPRIPL2 | -0.021678118 | 0.025027859 | 0.003349741 |
| CRBN | 0.040692441 | -0.068200831 | -0.02750839 |
| VPS26C | 0.024492858 | -0.037552573 | -0.013059715 |
| TNF | 0.008552115 | -0.009804394 | -0.001252279 |
| IGF2R | -0.045117401 | -0.042993981 | -0.088111382 |
| MAP9 | 0.038233212 | -0.042682615 | -0.004449402 |
| PTGFRN | -0.016940258 | 0.021913149 | 0.00497289 |
| TXNRD1 | 0.029943607 | -0.038645156 | -0.008701549 |
| FAF2 | 0.032422197 | -0.044652108 | -0.012229912 |
| HERC4 | 0.031193902 | -0.048814814 | -0.017620913 |
| GNAO1 | 0.006028002 | -0.006028002 | -2.13163E-14 |
| IGSF10 | 0.006078814 | -0.006078814 | -2.30926E-14 |
| NR2C2 | 0.017225504 | -0.021053196 | -0.003827692 |
| BNIP2 | 0.035427643 | -0.049303701 | -0.013876058 |
| DHCR24-DT | 0.006123843 | -0.006123843 | -1.95399E-14 |
| EFCAB5 | 0.005983854 | -0.005983854 | -1.95399E-14 |
| MED12L | 0.005957727 | -0.005957727 | -2.13163E-14 |
| ISM2 | 0.005900163 | -0.005900163 | -2.30926E-14 |
| HPSE | 0.013192181 | -0.019205776 | -0.006013595 |
| TMIE | 0.005821904 | -0.005821904 | -2.30926E-14 |
| ARNTL | 0.013348589 | -0.0145911 | -0.001242511 |
| ARHGEF25 | 0.006163626 | -0.006163626 | -2.13163E-14 |
| BRMS1L | 0.020105875 | -0.022597548 | -0.002491673 |
| C2CD4C | 0.00616153 | -0.00616153 | -2.13163E-14 |
| VPS53 | 0.028185186 | -0.049150886 | -0.0209657 |
| SUCLG2 | 0.034530289 | -0.039208933 | -0.004678644 |
| FCHO1 | 0.005262531 | -0.005262531 | -2.30926E-14 |
| PDZD8 | 0.039398855 | -0.040768459 | -0.001369603 |
| NAF1 | 0.020751269 | -0.019433114 | 0.001318155 |
| HARBI1 | 0.00580301 | -0.00580301 | -2.13163E-14 |
| FTO | 0.027354814 | -0.025764005 | 0.001590809 |
| FRAT1 | 0.01291359 | -0.012863882 | 4.97073E-05 |
| QPCTL | 0.010818969 | -0.012285958 | -0.001466989 |
| KDSR | 0.038762309 | -0.073041591 | -0.034279281 |
| CEP57 | 0.032579134 | -0.043480511 | -0.010901377 |
| MAGIX | 0.021640422 | -0.022500772 | -0.00086035 |
| WFIKKN1 | 0.005246904 | -0.005246904 | -2.30926E-14 |
| GTPBP6 | 0.036978175 | -0.037565298 | -0.000587124 |
| ZMYM4 | 0.038167422 | -0.063043842 | -0.024876419 |
| CLP1 | 0.014624123 | -0.015455807 | -0.000831684 |
| TRMT61A | 0.021245116 | -0.025307766 | -0.004062651 |
| CLN5 | 0.043314993 | -0.083771445 | -0.040456452 |
| SH2D3A | 0.026554436 | -0.029277445 | -0.002723008 |
| STRN3 | 0.034796662 | -0.038917167 | -0.004120505 |
| NKX3-1 | 0.006465571 | -0.006465571 | -2.13163E-14 |
| ATL3 | 0.030563488 | -0.052254637 | -0.021691148 |

| | | | |
|---|---|---|---|
| OXSM | 0.021315026 | -0.023555289 | -0.002240263 |
| FOXN3 | 0.020586891 | -0.019775559 | 0.000811333 |
| HPCA | 0.006054376 | -0.006054376 | -2.13163E-14 |
| LINC01819 | 0.006128786 | -0.006128786 | -2.13163E-14 |
| ELK3 | 0.012815147 | -0.015314377 | -0.00249923 |
| STARD3 | 0.025840009 | -0.027725148 | -0.00188514 |
| SP3 | 0.037839077 | -0.058795434 | -0.020956357 |
| MNX1 | 0.005859584 | -0.005859584 | -2.13163E-14 |
| TAOK1 | 0.036862312 | -0.048132009 | -0.011269697 |
| IPO7 | 0.04237242 | -0.046538549 | -0.004166129 |
| SCGB2A2 | 0.0086609 | -0.0086609 | -1.95399E-14 |
| RPS26 | 0.062738139 | 0.132091002 | 0.194829141 |
| POLR2B | 0.036076942 | -0.046576805 | -0.010499863 |
| USP28 | 0.021168779 | -0.029340215 | -0.008171436 |
| RNF168 | 0.023510412 | -0.032132152 | -0.008621739 |
| AC093151.8 | 0.006225964 | -0.006225964 | -2.30926E-14 |
| EFNA3 | 0.008415009 | -0.008896141 | -0.000481132 |
| ZNF718 | 0.014655661 | -0.016516784 | -0.001861123 |
| SRPRB | 0.031449571 | -0.036530173 | -0.005080603 |
| IL10RB | 0.025275721 | -0.044476971 | -0.01920125 |
| AL596244.1 | 0.00601861 | -0.00601861 | -1.95399E-14 |
| HSD17B1 | 0.016395464 | -0.023822763 | -0.007427299 |
| PIGBOS1 | 0.026425772 | -0.035473919 | -0.009048147 |
| AC104667.2 | 0.010625806 | -0.01216944 | -0.001543634 |
| SENP5 | 0.025779608 | -0.048381533 | -0.022601925 |
| SRRD | 0.022408707 | -0.026935888 | -0.004527181 |
| MIR181A1HG | 0.013679163 | -0.018396242 | -0.004717079 |
| CD47 | 0.038449637 | -0.055562913 | -0.017113276 |
| IER5L | 0.030401545 | -0.035125651 | -0.004724106 |
| EAPP | 0.047559501 | -0.07693246 | -0.029372959 |
| ELMO3 | 0.019273525 | -0.022177078 | -0.002903553 |
| ADARB2 | 0.00671991 | -0.00671991 | -1.95399E-14 |
| PRKAR2A | 0.034333111 | -0.046069466 | -0.011736355 |
| VMAC | 0.016498263 | -0.016563102 | -6.48391E-05 |
| AC243964.3 | 0.01356757 | -0.014685657 | -0.001118087 |
| MUC1 | 0.040600722 | -0.06702576 | -0.026425038 |
| POLDIP3 | 0.033461804 | -0.035335118 | -0.001873314 |
| HRCT1 | -0.010919453 | 0.014398146 | 0.003478694 |
| YBEY | 0.026247533 | -0.036984781 | -0.010737248 |
| LINC00665 | -0.030199926 | 0.03961497 | 0.009415044 |
| EIF1AY | 0.03054737 | -0.081789369 | -0.051241999 |
| RPL27A | 0.058776867 | 0.104758567 | 0.163535433 |
| UTP25 | 0.020508362 | -0.021313882 | -0.000805521 |
| PRPSAP2 | 0.033375307 | -0.045473688 | -0.012098381 |
| RSF1 | 0.04219102 | -0.065391394 | -0.023200373 |
| ALKBH4 | 0.026225702 | -0.029396112 | -0.003170409 |
| OSBPL2 | 0.028235596 | -0.038171565 | -0.009935969 |

| | | | |
|---|---|---|---|
| VIM | 0.050959543 | 0.169239474 | 0.220199017 |
| FADD | 0.022026365 | -0.021320513 | 0.000705853 |
| FMNL1 | 0.011586112 | -0.013098509 | -0.001512397 |
| MYOM1 | 0.011760595 | -0.011487692 | 0.000272904 |
| MAGED1 | 0.031734819 | -0.038438187 | -0.006703368 |
| ELMO2 | 0.015779971 | -0.015456529 | 0.000323442 |
| HAUS7 | 0.021849634 | -0.026707273 | -0.00485764 |
| ADAM22 | 0.009833781 | -0.009598065 | 0.000235716 |
| IL20RA | -0.011466739 | 0.013606374 | 0.002139635 |
| COG6 | 0.019583262 | -0.025190698 | -0.005607437 |
| XAB2 | 0.030225955 | -0.034308692 | -0.004082736 |
| BOLA3-AS1 | 0.014460339 | -0.016600837 | -0.002140497 |
| STRN | 0.041299724 | -0.100007267 | -0.058707544 |
| HDHD5 | 0.018379044 | -0.026184251 | -0.007805207 |
| ASH2L | 0.023201523 | -0.022576145 | 0.000625378 |
| SIPA1L2 | -0.015147551 | 0.020909846 | 0.005762295 |
| AFF4 | 0.038269314 | -0.053803692 | -0.015534378 |
| PDZK1 | 0.054593817 | 0.11582574 | 0.170419557 |
| AFAP1 | -0.018688698 | 0.029619838 | 0.01093114 |
| TNFAIP1 | 0.023904378 | -0.02808058 | -0.004176202 |
| LINC02027 | -0.012657696 | 0.0197804 | 0.007122703 |
| LTBP3 | 0.046765443 | -0.063382949 | -0.016617506 |
| GMDS | -0.058374316 | 0.057097218 | -0.001277098 |
| TNFSF10 | -0.042465548 | 0.109241205 | 0.066775657 |
| NEU3 | -0.012878693 | 0.014908453 | 0.002029759 |
| CYREN | 0.032695655 | -0.034694358 | -0.001998702 |
| C15orf62 | 0.006423455 | -0.006423455 | -1.95399E-14 |
| HOXB6 | 0.038616437 | -0.086647494 | -0.048031057 |
| TIPARP | 0.014298059 | -0.020619736 | -0.006321677 |
| PPP1R10 | 0.03863765 | -0.053136875 | -0.014499225 |
| UGT2B7 | -0.052724353 | 0.098270218 | 0.045545865 |
| HINT3 | 0.028237171 | -0.031634872 | -0.003397701 |
| CRELD1 | 0.026406613 | -0.030118796 | -0.003712183 |
| ASCC3 | 0.029645892 | -0.033439536 | -0.003793644 |
| AP000553.2 | 0.007296568 | -0.008615937 | -0.001319369 |
| NUP50 | 0.028331838 | -0.035155245 | -0.006823407 |
| B3GLCT | 0.020010832 | -0.036616119 | -0.016605287 |
| ITCH | 0.033291255 | -0.049761917 | -0.016470663 |
| SCRN1 | 0.034289497 | -0.050419263 | -0.016129766 |
| ZNF354B | 0.018233088 | -0.02321421 | -0.004981121 |
| SLC25A19 | 0.013638767 | -0.015388488 | -0.001749721 |
| RNF13 | 0.045509967 | -0.073818603 | -0.028308636 |
| SLC2A9 | 0.016022194 | -0.023649953 | -0.007627758 |
| DPH3 | 0.020014149 | -0.025269054 | -0.005254905 |
| CHCHD6 | 0.02926131 | -0.033030458 | -0.003769148 |
| SMIM10 | 0.018777634 | -0.020947725 | -0.002170092 |
| CACNA2D3 | 0.020700007 | -0.026437364 | -0.005737357 |

| | | | |
|---|---|---|---|
| SNAP23 | 0.026235869 | -0.043137782 | -0.016901913 |
| APH1B | 0.025333213 | -0.028474501 | -0.003141289 |
| RBM18 | 0.021044718 | -0.020656993 | 0.000387725 |
| DIPK1B | -0.017476109 | 0.039132631 | 0.021656522 |
| MTO1 | 0.019043099 | -0.018867518 | 0.000175581 |
| CENPS | 0.028046572 | -0.045775513 | -0.017728941 |
| VCPIP1 | 0.023302576 | -0.024655695 | -0.001353119 |
| CACNB2 | -0.009197757 | 0.009244552 | 4.67944E-05 |
| CRKL | 0.020428235 | -0.020500101 | -7.18663E-05 |
| CAVIN2 | -0.014383511 | 0.017938357 | 0.003554846 |
| CTSF | 0.039522387 | -0.087251525 | -0.047729138 |
| CHST12 | 0.025467615 | -0.031871752 | -0.006404137 |
| CC2D1A | 0.020248716 | -0.022185428 | -0.001936712 |
| TMC4 | 0.026803652 | -0.03989656 | -0.013092909 |
| PRKAG2 | 0.028531926 | -0.047223644 | -0.018691718 |
| RNF216 | 0.02698314 | -0.037395677 | -0.010412538 |
| PRDM5 | 0.015832406 | -0.019443474 | -0.003611068 |
| TMEM72 | -0.015757481 | 0.016583182 | 0.000825702 |
| TNS2 | -0.018327328 | 0.01899828 | 0.000670951 |
| EML1 | 0.013438931 | -0.016898228 | -0.003459297 |
| ZNF667 | 0.017384738 | -0.024252261 | -0.006867523 |
| SNHG29 | 0.061694512 | 0.113863289 | 0.175557801 |
| PPM1K | 0.032842173 | -0.044947128 | -0.012104955 |
| ABCD3 | 0.031743204 | -0.042555831 | -0.010812627 |
| MAGI2 | 0.026279543 | -0.048225532 | -0.021945989 |
| HOXD9 | 0.029315364 | -0.043687018 | -0.014371654 |
| PALM3 | 0.036210155 | -0.058908842 | -0.022698686 |
| DNAJC15 | 0.040657995 | -0.041453188 | -0.000795193 |
| IMMP2L | 0.045214191 | -0.07383385 | -0.028619659 |
| DAGLA | 0.009119772 | -0.010299228 | -0.001179456 |
| KLHL36 | -0.028931894 | 0.032996313 | 0.004064419 |
| MRPL42 | 0.037942154 | -0.038249371 | -0.000307217 |
| MT-ND2 | -0.041161991 | 0.079384556 | 0.038222566 |
| GHDC | 0.023474689 | -0.026339105 | -0.002864416 |
| HDX | 0.013598731 | -0.01489543 | -0.001296699 |
| AC107953.2 | 0.005292044 | -0.005292044 | -2.30926E-14 |
| TCP11L2 | 0.016127145 | -0.025800711 | -0.009673565 |
| ARL6IP5 | 0.045548785 | -0.046507706 | -0.000958921 |
| PAQR5 | 0.028496314 | -0.031088559 | -0.002592246 |
| PCDH20 | -0.010477715 | 0.011008691 | 0.000530975 |
| TMEM218 | 0.030355373 | -0.034308741 | -0.003953368 |
| AL357033.1 | 0.005224547 | -0.005224547 | -2.30926E-14 |
| BEST3 | 0.005319617 | -0.005319617 | -2.13163E-14 |
| DACH2 | 0.005268737 | -0.005268737 | -1.95399E-14 |
| AL078587.2 | 0.005162486 | -0.005162486 | -1.95399E-14 |
| ARHGEF10 | -0.030106011 | -0.032315209 | -0.06242122 |
| TIAM1 | 0.005113667 | -0.005113667 | -2.30926E-14 |

| | | | |
|---|---|---|---|
| ARHGEF1 | 0.019041357 | -0.025821266 | -0.006779909 |
| LGR5 | 0.005709862 | -0.005709862 | -2.30926E-14 |
| FAM49A | 0.005395259 | -0.005395259 | -2.30926E-14 |
| TMEM255B | 0.004987716 | -0.004987716 | -2.13163E-14 |
| OXNAD1 | 0.014968835 | -0.015960726 | -0.00099189 |
| AL139383.1 | 0.005080019 | -0.005080019 | -2.13163E-14 |
| GPR143 | 0.016359976 | -0.020192677 | -0.003832701 |
| CHD4 | 0.041472782 | -0.086573993 | -0.045101211 |
| ARL13B | 0.026704132 | -0.038941261 | -0.012237129 |
| SDAD1 | 0.031277885 | -0.062304366 | -0.031026481 |
| DOLK | 0.015153689 | -0.016671468 | -0.001517779 |
| MORC2 | 0.021372271 | -0.032511777 | -0.011139506 |
| PTPN6 | 0.019255113 | -0.021545665 | -0.002290553 |
| ITSN2 | 0.033502233 | -0.07576255 | -0.042260317 |
| GATAD1 | 0.037217013 | -0.038045746 | -0.000828733 |
| CGN | 0.027419147 | -0.032138012 | -0.004718865 |
| KIAA0586 | 0.023837792 | -0.027380863 | -0.003543071 |
| IFNAR2 | 0.014753962 | -0.017050404 | -0.002296442 |
| ZFAND3 | 0.033104214 | -0.054304888 | -0.021200674 |
| VKORC1L1 | 0.027787947 | -0.048662721 | -0.020874774 |
| SDCBP2 | 0.010057871 | -0.010444046 | -0.000386176 |
| MSH3 | 0.028984609 | -0.037333736 | -0.008349127 |
| PLXNA2 | 0.018020395 | -0.027694983 | -0.009674588 |
| CUX1 | -0.039311151 | 0.046457936 | 0.007146785 |
| XPA | 0.032735421 | -0.035652878 | -0.002917458 |
| RBM28 | 0.028854703 | -0.051155154 | -0.022300451 |
| NEU1 | 0.035705137 | -0.046841103 | -0.011135966 |
| CYBC1 | 0.025177683 | -0.03403734 | -0.008859657 |
| MIPOL1 | 0.021187564 | -0.027138758 | -0.005951194 |
| TTC38 | 0.021048274 | 0.023084249 | 0.044132523 |
| EFHC1 | 0.03309359 | -0.053637218 | -0.020543628 |
| NUDT1 | 0.022890753 | -0.024070932 | -0.001180179 |
| ADGRL2 | -0.02059499 | 0.030216439 | 0.009621449 |
| FOXN2 | 0.016078285 | -0.016558955 | -0.00048067 |
| PRPF6 | 0.039219627 | -0.050853009 | -0.011633382 |
| NBPF1 | 0.019540983 | -0.028546627 | -0.009005643 |
| RO60 | 0.038654672 | -0.073851871 | -0.0351972 |
| ZDHHC23 | 0.015486803 | -0.027871923 | -0.01238512 |
| UBR7 | 0.021868231 | -0.022728083 | -0.000859852 |
| DENND4C | 0.025701959 | -0.029843814 | -0.004141854 |
| LNPK | 0.026242963 | -0.033574084 | -0.007331121 |
| RDH13 | 0.022112626 | -0.027446069 | -0.005333444 |
| MORC4 | 0.018723431 | -0.023050109 | -0.004326678 |
| CDK17 | 0.015621418 | -0.029027382 | -0.013405964 |
| SPOP | 0.034801865 | -0.062486721 | -0.027684857 |
| TARDBP | 0.035476664 | -0.067310252 | -0.031833589 |
| C1orf131 | 0.024336485 | -0.052180678 | -0.027844192 |

| | | | |
|---|---|---|---|
| FAM3C | 0.050675724 | -0.14788178 | -0.097206056 |
| TOR1AIP1 | 0.02826404 | -0.030703888 | -0.002439848 |
| BBX | 0.043585938 | -0.104884513 | -0.061298575 |
| GATC | 0.026393538 | -0.044465758 | -0.01807222 |
| MPHOSPH8 | 0.04374968 | -0.08510963 | -0.04135995 |
| CACNB4 | 0.008440236 | -0.009503246 | -0.00106301 |
| CD27-AS1 | 0.02575354 | -0.041920082 | -0.016166541 |
| KRI1 | 0.016866415 | -0.025745827 | -0.008879412 |
| ECPAS | 0.025124712 | -0.026804899 | -0.001680188 |
| AL109976.1 | 0.012367049 | -0.016841961 | -0.004474912 |
| PDS5B | 0.032887278 | -0.06500071 | -0.032113432 |
| ZNF467 | 0.01526816 | -0.016064021 | -0.000795861 |
| TAGLN | -0.018918553 | 0.027402425 | 0.008483872 |
| CFAP300 | 0.012315324 | -0.018493566 | -0.006178242 |
| ABHD13 | 0.023114003 | -0.024818879 | -0.001704875 |
| GPR108 | 0.035034362 | -0.0443293 | -0.009294939 |
| ARFIP1 | 0.027907254 | -0.038378954 | -0.0104717 |
| PGAP1 | 0.023843311 | -0.0397674 | -0.015924089 |
| APOOL | 0.025720562 | -0.038618935 | -0.012898373 |
| PAK6 | 0.012954503 | -0.019537094 | -0.006582591 |
| HERC2 | 0.034107346 | -0.069715644 | -0.035608298 |
| ZNF598 | 0.016179682 | -0.02299872 | -0.006819038 |
| FAM216A | 0.010891013 | -0.012942724 | -0.002051711 |
| DCAF12 | 0.019806091 | -0.023040145 | -0.003234055 |
| TMEM246 | -0.024636743 | 0.027186687 | 0.002549944 |
| CDK2 | 0.01585959 | -0.019553701 | -0.003694112 |
| GLYR1 | 0.039588106 | -0.059392798 | -0.019804692 |
| YAF2 | 0.02172878 | -0.031990425 | -0.010261644 |
| ZNF888 | 0.027919613 | -0.041456438 | -0.013536825 |
| RANBP3 | 0.022828455 | -0.036196577 | -0.013368122 |
| ZFP14 | 0.024707831 | -0.040652077 | -0.015944246 |
| URB1-AS1 | 0.021262049 | -0.042277662 | -0.021015613 |
| COX18 | 0.013393598 | -0.01490687 | -0.001513272 |
| TRAF5 | 0.023531518 | -0.044530007 | -0.020998489 |
| NFIC | 0.041995217 | -0.060975893 | -0.018980676 |
| HIST3H2A | -0.016943326 | 0.022158746 | 0.00521542 |
| LRRC23 | 0.024658562 | -0.032988355 | -0.008329793 |
| MYH10 | 0.03487071 | -0.049267573 | -0.014396863 |
| SIRT1 | 0.021253801 | -0.039490588 | -0.018236787 |
| PTPRM | 0.028869368 | -0.036774154 | -0.007904786 |
| FGFR4 | -0.015074995 | 0.017364368 | 0.002289373 |
| ST3GAL5 | -0.014677889 | 0.024902291 | 0.010224402 |
| PDCD7 | 0.030592649 | -0.054080485 | -0.023487836 |
| AC245140.2 | 0.008584091 | -0.009775934 | -0.001191843 |
| NCBP1 | 0.019146694 | -0.02358592 | -0.004439226 |
| STIMATE | 0.015869972 | -0.025925406 | -0.010055435 |
| MLLT3 | 0.02797922 | -0.062972829 | -0.03499361 |

| | | | |
|---|---|---|---|
| TOM1L1 | -0.031854246 | 0.05565085 | 0.023796605 |
| TRPM3 | -0.013012191 | 0.016009347 | 0.002997156 |
| CHPT1 | 0.030482052 | -0.03396888 | -0.003486828 |
| GULP1 | -0.025551361 | 0.041966592 | 0.016415231 |
| PINX1 | 0.015755731 | -0.021826533 | -0.006070802 |
| ZNF436-AS1 | 0.015955174 | -0.020129902 | -0.004174729 |
| SEPTIN2 | 0.049048818 | -0.06432254 | -0.015273722 |
| TLK2 | 0.022307824 | -0.025585247 | -0.003277423 |
| TUBGCP4 | 0.017699201 | -0.019724396 | -0.002025195 |
| RCOR3 | 0.034206386 | -0.065452208 | -0.031245822 |
| AC104794.5 | 0.013578804 | -0.016891781 | -0.003312977 |
| TRAPPC8 | 0.026816599 | -0.033601099 | -0.0067845 |
| TMSB4Y | 0.015390482 | -0.029425649 | -0.014035167 |
| CA3-AS1 | 0.010598108 | -0.016957728 | -0.00635962 |
| AL135925.1 | 0.022443463 | -0.030862341 | -0.008418877 |
| UPRT | 0.014087064 | -0.016378181 | -0.002291118 |
| ZNF652 | 0.033267815 | -0.062301377 | -0.029033562 |
| LYPD3 | 0.009076371 | -0.011941333 | -0.002864962 |
| WWOX | 0.019695199 | -0.022881681 | -0.003186482 |
| ERICH1 | 0.028221729 | -0.033678363 | -0.005456634 |
| TOLLIP-AS1 | 0.014842961 | -0.018691208 | -0.003848247 |
| TCOF1 | 0.023265281 | -0.032518095 | -0.009252814 |
| TMEM39B | 0.012021938 | -0.01466483 | -0.002642892 |
| SNRNP48 | 0.024205984 | -0.026506075 | -0.002300091 |
| ATP2A3 | 0.007202818 | -0.008688968 | -0.00148615 |
| NCOA2 | 0.027671603 | -0.043211506 | -0.015539903 |
| MMS19 | 0.022921025 | -0.031576895 | -0.008655871 |
| CYTH3 | 0.012584966 | -0.016311379 | -0.003726414 |
| SPTBN5 | 0.005579329 | -0.006437554 | -0.000858225 |
| SUCO | 0.031302627 | -0.048776024 | -0.017473396 |
| RUFY3 | 0.028378044 | -0.039219965 | -0.010841921 |
| AL161421.1 | -0.010183831 | 0.013667953 | 0.003484122 |
| CAPN7 | 0.031716374 | -0.041415041 | -0.009698667 |
| FKBP14 | 0.017805527 | -0.021192028 | -0.003386501 |
| CDK9 | 0.024446239 | -0.03345161 | -0.009005371 |
| LTN1 | 0.028768385 | -0.041093252 | -0.012324867 |
| TMEM150C | -0.01315839 | 0.014806518 | 0.001648129 |
| ARV1 | 0.027090851 | -0.043007484 | -0.015916633 |
| AOX1 | -0.012056792 | 0.014120779 | 0.002063988 |
| THAP5 | 0.032293688 | -0.037724397 | -0.005430709 |
| CREB1 | 0.029683712 | -0.045460897 | -0.015777185 |
| ZSWIM4 | 0.012933531 | -0.017977154 | -0.005043624 |
| SEL1L | 0.03713227 | -0.092719133 | -0.055586863 |
| SLC2A11 | 0.021028422 | -0.046438002 | -0.025409579 |
| ERCC5 | 0.02685168 | -0.05000316 | -0.02315148 |
| PKD1 | 0.01878908 | -0.020508238 | -0.001719158 |
| PLA2G15 | 0.024332882 | -0.038295593 | -0.013962711 |

| | | | |
|---|---|---|---|
| CHRAC1 | 0.025310473 | -0.037776571 | -0.012466098 |
| COIL | 0.025661444 | -0.043183939 | -0.017522495 |
| GSK3B | 0.027082431 | -0.031844818 | -0.004762387 |
| FAF1 | 0.025169763 | -0.030563894 | -0.005394131 |
| MFF | 0.042610124 | -0.0546968 | -0.012086676 |
| PDZD2 | 0.019758272 | -0.02434 | -0.004581727 |
| MARC2 | 0.02527344 | -0.028666095 | -0.003392655 |
| ARL17A | 0.016360507 | -0.019270645 | -0.002910138 |
| ROBO3 | 0.012446725 | -0.019870514 | -0.007423789 |
| VEGFA | 0.040339051 | -0.112840935 | -0.072501884 |
| HNRNPUL2 | 0.031365662 | -0.048075496 | -0.016709834 |
| KLC1 | 0.025679503 | -0.039652498 | -0.013972995 |

**Thick ascending limb cells**

| Genes | logFC_T2D_HC | logFC_T2Di_T2D | logFC_T2Di_HC |
|---|---|---|---|
| AKR1B1 | 0.261275424 | -0.266247556 | -0.004972131 |
| SERPINA1 | 0.150339254 | -0.150951196 | -0.000611941 |
| SERPINA6 | 0.106308414 | -0.106461433 | -0.000153019 |
| UMOD | -0.457005175 | 0.374794888 | -0.082210287 |
| MELTF | 0.158973143 | -0.161125568 | -0.002152425 |
| CPE | 0.084084816 | -0.083113159 | 0.000971657 |
| CKB | -0.239663366 | 0.197583728 | -0.042079638 |
| CALCA | 0.05444008 | -0.055039158 | -0.000599078 |
| DEFB1 | -0.248944426 | 0.248971221 | 2.67943E-05 |
| ARG2 | 0.064675503 | -0.061902632 | 0.002772871 |
| DST | -0.173856274 | 0.1017361 | -0.072120174 |
| ADIRF | -0.242534167 | 0.163720451 | -0.078813716 |
| FKBP5 | -0.131226783 | 0.084660604 | -0.046566179 |
| AKR1C1 | 0.047244405 | -0.048648908 | -0.001404503 |
| TACC3 | 0.040044319 | -0.040215535 | -0.000171217 |
| SLC6A12 | 0.058073556 | -0.052208594 | 0.005864962 |
| SLCO4A1 | 0.035319236 | -0.034925281 | 0.000393955 |
| PRKAR2B | 0.032795206 | -0.033331529 | -0.000536323 |
| PFKFB3 | 0.067511098 | -0.07499386 | -0.007482762 |
| DDIT4L | 0.08223276 | -0.079334417 | 0.002898342 |
| SERPINA5 | 0.196736556 | -0.10536687 | 0.091369685 |
| LTBP1 | 0.039435703 | -0.036627341 | 0.002808362 |
| RNF24 | 0.085006668 | -0.081387369 | 0.003619299 |
| GPC3 | -0.145339917 | 0.106062277 | -0.039277641 |
| SLC12A1 | -0.201477711 | 0.175968407 | -0.025509304 |
| SOCS2 | 0.066867442 | -0.047683155 | 0.019184286 |
| AC068631.1 | -0.108631518 | 0.085603444 | -0.023028073 |
| KNG1 | -0.177351133 | 0.132747568 | -0.044603565 |
| HSPB7 | 0.062011306 | -0.060594477 | 0.001416829 |
| RIPOR3 | 0.052163416 | -0.046615318 | 0.005548098 |
| CRYAB | 0.073675341 | -0.067663744 | 0.006011596 |
| TESC | 0.055625214 | -0.056212911 | -0.000587697 |
| SH3GL3 | 0.025636303 | -0.025023924 | 0.00061238 |
| CCDC71L | -0.086641566 | 0.031805168 | -0.054836398 |
| MT-ND2 | -0.137773254 | 0.072870505 | -0.064902749 |
| SPNS2 | 0.06003747 | -0.066631207 | -0.006593737 |
| TBC1D16 | 0.09403285 | -0.077139507 | 0.016893343 |
| MT-ND5 | -0.133060656 | 0.084389065 | -0.048671591 |
| RANBP3L | 0.105446592 | -0.096466248 | 0.008980344 |
| SOD3 | 0.098843167 | -0.088825367 | 0.0100178 |
| MT-ATP8 | -0.144846481 | 0.046192378 | -0.098654103 |
| NEK10 | 0.023301928 | -0.02335412 | -5.21913E-05 |
| MT1X | -0.105178306 | 0.063777696 | -0.041400609 |
| MT2A | -0.163689157 | 0.088393179 | -0.075295978 |
| MPC1 | -0.149601729 | 0.116928335 | -0.032673393 |

| | | | |
|---|---|---|---|
| LPL | -0.11835659 | 0.028341021 | -0.090015569 |
| LPIN2 | 0.066778778 | -0.071161362 | -0.004382585 |
| IGFBP2 | 0.125088325 | -0.039901736 | 0.085186589 |
| ARPC1B | 0.050205406 | -0.047311934 | 0.002893471 |
| SMIM24 | -0.062007629 | 0.018449466 | -0.043558163 |
| NEURL3 | 0.028720257 | -0.028395547 | 0.00032471 |
| MT-ND4L | -0.123974833 | 0.05468153 | -0.069293303 |
| TFF3 | 0.022016022 | -0.021532446 | 0.000483577 |
| BHLHE40 | -0.088240487 | 0.025789669 | -0.062450817 |
| FMN1 | 0.050019172 | -0.04997344 | 4.57318E-05 |
| LDHB | -0.127529949 | 0.093818594 | -0.033711355 |
| ID2 | -0.116005453 | 0.057541096 | -0.058464357 |
| AFAP1L2 | 0.025174194 | -0.025754972 | -0.000580779 |
| RNF128 | 0.023926146 | -0.025928641 | -0.002002494 |
| CNNM1 | 0.01624031 | -0.016119528 | 0.000120783 |
| MT-ND4 | -0.110805374 | 0.065659672 | -0.045145702 |
| MT-ND1 | -0.122177299 | 0.040745361 | -0.081431938 |
| SLC4A11 | 0.019679127 | -0.019679872 | -7.44896E-07 |
| BOC | 0.016477178 | -0.016054516 | 0.000422661 |
| UCP2 | 0.030422283 | -0.029864464 | 0.000557819 |
| RGCC | 0.022903837 | -0.02271479 | 0.000189047 |
| ASPHD2 | 0.037848538 | -0.0311489 | 0.006699638 |
| NEFL | -0.073097546 | 0.049596999 | -0.023500548 |
| GSTM3 | 0.128002165 | -0.096950488 | 0.031051677 |
| WNT7B | 0.032663762 | -0.039702038 | -0.007038276 |
| TPT1 | -0.13019749 | 0.056164963 | -0.074032527 |
| NTRK2 | -0.125063512 | 0.064629241 | -0.060434271 |
| CCND1 | -0.110890936 | 0.089763753 | -0.021127184 |
| ABI3 | -0.067641802 | 0.060020803 | -0.007620999 |
| RPL10 | -0.124072768 | 0.053736306 | -0.070336462 |
| KCNJ5 | 0.050214631 | -0.044929863 | 0.005284768 |
| RPS26 | -0.135190595 | 0.098239012 | -0.036951584 |
| MT1E | -0.116287118 | 0.090176278 | -0.02611084 |
| MT-CO3 | -0.112673998 | 0.066000106 | -0.046673892 |
| TDGF1 | -0.063568538 | 0.018907186 | -0.044661352 |
| DUSP9 | -0.127915411 | 0.070157025 | -0.057758386 |
| CAT | 0.105634015 | -0.116772427 | -0.011138412 |
| CYB5A | -0.130696232 | 0.057217609 | -0.073478623 |
| SNHG29 | -0.129885533 | 0.060360278 | -0.069525254 |
| CHDH | 0.032318252 | -0.030536665 | 0.001781587 |
| FOLR3 | -0.094634361 | 0.10256285 | 0.00792849 |
| PCSK1N | -0.132999588 | 0.101735625 | -0.031263963 |
| PPP1R16A | -0.076353398 | 0.043857445 | -0.032495952 |
| PCK1 | 0.093423795 | -0.069031133 | 0.024392662 |
| RSPO3 | 0.013694573 | -0.013555647 | 0.000138926 |
| TACSTD2 | 0.100267943 | -0.04520074 | 0.055067203 |
| ATP1B3 | 0.09301589 | -0.064587489 | 0.028428401 |

| | | | |
|---|---|---|---|
| LINC02381 | -0.097442923 | 0.050319956 | -0.047122967 |
| FOLR1 | -0.094003152 | 0.10803421 | 0.014031058 |
| FXYD2 | -0.127325599 | 0.033932358 | -0.093393241 |
| MUC1 | -0.120196571 | 0.110735765 | -0.009460806 |
| LINC01320 | -0.089799496 | 0.06324233 | -0.026557165 |
| LDHA | 0.100185044 | -0.065569858 | 0.034615186 |
| EGF | -0.134433472 | 0.088565905 | -0.045867567 |
| AC013275.1 | 0.022323224 | -0.020744035 | 0.001579189 |
| HIF1A | 0.083678219 | -0.066056316 | 0.017621903 |
| MT-ATP6 | -0.092638888 | 0.04901066 | -0.043628228 |
| AC009275.1 | 0.012631203 | -0.012608578 | 2.26254E-05 |
| FHL2 | 0.031316409 | -0.032934152 | -0.001617743 |
| LINC00379 | -0.06073201 | 0.035781667 | -0.024950344 |
| CDC25B | 0.021648607 | -0.021229737 | 0.00041887 |
| MT-CO1 | -0.099236133 | 0.050621643 | -0.04861449 |
| PFKP | 0.068640742 | -0.055554156 | 0.013086586 |
| SLC14A1 | 0.0180141 | -0.018141039 | -0.00012694 |
| TP53I11 | 0.013023887 | -0.013409328 | -0.000385441 |
| RPL24 | -0.1162233 | 0.049487319 | -0.066735981 |
| UQCRB | -0.114387982 | 0.095799926 | -0.018588056 |
| SLC3A1 | -0.104976926 | 0.064413834 | -0.040563092 |
| ID3 | -0.074021244 | 0.052312663 | -0.021708581 |
| AC108749.1 | 0.014584791 | -0.014467773 | 0.000117018 |
| SIM1 | 0.081977849 | -0.103114532 | -0.021136684 |
| MFSD10 | 0.054356263 | -0.040560904 | 0.013795359 |
| AIF1L | 0.104000891 | -0.089016202 | 0.014984689 |
| BEX2 | -0.099527087 | 0.042077891 | -0.057449196 |
| RPL39 | -0.116460199 | 0.057704635 | -0.058755564 |
| SDC1 | -0.103601671 | 0.104008469 | 0.000406798 |
| FAM131C | 0.051172059 | -0.044731344 | 0.006440716 |
| FABP3 | -0.104411674 | 0.109636839 | 0.005225165 |
| SLC22A2 | -0.093634471 | 0.087584977 | -0.006049495 |
| CNTD2 | 0.01494861 | -0.014563901 | 0.000384709 |
| SULT2B1 | 0.019233735 | -0.021441945 | -0.00220821 |
| MT-CYB | -0.095450983 | 0.052427047 | -0.043023937 |
| RPL41 | -0.100596244 | 0.052346377 | -0.048249867 |
| CLDN1 | 0.015713541 | -0.014930038 | 0.000783503 |
| PXMP2 | -0.08163154 | 0.075904415 | -0.005727125 |
| SSTR2 | 0.011840608 | -0.011842601 | -1.99263E-06 |
| PAX8 | 0.077003409 | -0.075522099 | 0.00148131 |
| CLCNKB | -0.109523274 | 0.112288426 | 0.002765152 |
| NDUFA4 | -0.103699231 | 0.07986665 | -0.023832581 |
| S100A4 | 0.072112285 | -0.074360081 | -0.002247797 |
| RPL26 | -0.106480859 | 0.044504976 | -0.061975883 |
| NFKBIA | 0.062646055 | -0.063024579 | -0.000378523 |
| SHB | 0.036914784 | -0.035640182 | 0.001274603 |
| RPS27A | -0.101819254 | 0.027809096 | -0.074010158 |

| | | | |
|---|---|---|---|
| FRY | 0.038887683 | -0.035561468 | 0.003326215 |
| LGR5 | 0.009980411 | -0.009606225 | 0.000374186 |
| CYFIP2 | -0.111355858 | 0.033389958 | -0.0779659 |
| MAL2 | 0.083783525 | -0.061387264 | 0.022396261 |
| HSPA8 | -0.098636926 | 0.064598383 | -0.034038543 |
| AC079447.1 | 0.039440687 | -0.045781379 | -0.006340692 |
| HMGN2 | -0.09341146 | 0.066509587 | -0.026901872 |
| SLC9A3R2 | 0.032289316 | -0.034975607 | -0.002686291 |
| RPL11 | -0.104136794 | 0.030399965 | -0.073736829 |
| GPER1 | 0.014998529 | -0.011908097 | 0.003090432 |
| MT-CO2 | -0.095935966 | 0.059039211 | -0.036896755 |
| MT-ND3 | -0.098058718 | 0.067083007 | -0.030975711 |
| FCGBP | 0.043998678 | -0.059549572 | -0.015550894 |
| RPS6 | -0.102207839 | 0.04653591 | -0.055671928 |
| RPS23 | -0.103718569 | 0.048933755 | -0.054784815 |
| RPS4Y1 | 0.073394015 | -0.188432678 | -0.115038663 |
| TMSB4X | 0.06349826 | -0.085159737 | -0.021661477 |
| OBSL1 | -0.086691091 | 0.083223696 | -0.003467395 |
| RPS4X | -0.104635303 | 0.089816812 | -0.01481849 |
| CMBL | -0.085733211 | 0.068023488 | -0.017709722 |
| COX7B | -0.10564745 | 0.102337498 | -0.003309953 |
| RPL13 | -0.093574733 | 0.039695286 | -0.053879447 |
| RPS14 | -0.097349943 | 0.02368576 | -0.073664183 |
| EIF1 | -0.100100647 | 0.053026588 | -0.047074059 |
| GMPR | -0.058956108 | 0.026315606 | -0.032640502 |
| PANK2 | 0.039891097 | -0.039109038 | 0.000782059 |
| GTF2H2 | -0.052903515 | 0.024036118 | -0.028867397 |
| LINC01687 | 0.007592291 | -0.007592291 | -3.55271E-15 |
| PLIN5 | -0.054292303 | 0.031576367 | -0.022715936 |
| FTL | -0.100963907 | 0.037212986 | -0.063750921 |
| GAS6 | 0.083100772 | -0.059877367 | 0.023223406 |
| MAOA | -0.056539834 | 0.026587426 | -0.029952408 |
| NTN4 | 0.06930767 | -0.049309388 | 0.019998281 |
| CARD6 | 0.012538445 | -0.012056995 | 0.00048145 |
| RPL34 | -0.100371608 | 0.043817414 | -0.056554194 |
| FBXO2 | 0.052065157 | -0.033834718 | 0.018230439 |
| SPOCK1 | 0.012357148 | -0.012799449 | -0.000442301 |
| PPM1K | -0.075996572 | 0.020824358 | -0.055172214 |
| SRSF3 | -0.083500251 | 0.03980627 | -0.043693981 |
| COX4I1 | -0.09397164 | 0.076635603 | -0.017336037 |
| CD83 | 0.030220133 | -0.020548622 | 0.009671511 |
| GRB14 | 0.059321424 | -0.049632343 | 0.00968908 |
| ALDH1L1 | -0.06738866 | 0.035691286 | -0.031697374 |
| PPDPF | -0.101835101 | 0.122740168 | 0.020905066 |
| EPHX2 | -0.079495785 | 0.079509852 | 1.40663E-05 |
| TMEM266 | 0.012214357 | -0.011713874 | 0.000500483 |
| VIM | 0.03177097 | -0.033719371 | -0.001948401 |

| | | | |
|---|---|---|---|
| STXBP6 | 0.010848518 | -0.010985971 | -0.000137453 |
| MRPL36 | -0.085407998 | 0.069825479 | -0.015582519 |
| SASH1 | 0.042205628 | -0.034124109 | 0.008081519 |
| KCNJ15 | 0.017753863 | -0.020873016 | -0.003119153 |
| NYNRIN | -0.033576843 | 0.009244245 | -0.024332598 |
| TPM1 | 0.060117349 | -0.052550347 | 0.007567002 |
| NELL1 | -0.059464108 | 0.016077138 | -0.043386969 |
| IFI27 | 0.017532992 | -0.01833187 | -0.000798878 |
| MXRA7 | 0.054926406 | -0.016524086 | 0.03840232 |
| ATP5MG | -0.094829424 | 0.087443895 | -0.007385529 |
| CALD1 | 0.024762567 | -0.020641314 | 0.004121253 |
| MFSD6 | 0.022886824 | -0.018762781 | 0.004124043 |
| SLC16A12 | -0.086875032 | 0.039387692 | -0.04748734 |
| ATP5IF1 | -0.092967515 | 0.067712526 | -0.02525499 |
| THNSL2 | -0.040248409 | 0.042133489 | 0.00188508 |
| CAVIN1 | 0.023519036 | -0.019983965 | 0.003535071 |
| RPS29 | -0.090401768 | 0.077802741 | -0.012599028 |
| RPL32 | -0.091841008 | 0.026948465 | -0.064892542 |
| CD82 | 0.037571326 | -0.038121753 | -0.000550427 |
| RHOBTB3 | -0.083848931 | 0.062577309 | -0.021271623 |
| PTGER3 | -0.095019993 | 0.045558791 | -0.049461202 |
| MACROD1 | -0.064081437 | 0.05680171 | -0.007279727 |
| SRGN | 0.014209747 | -0.014759317 | -0.00054957 |
| SLC25A3 | -0.088286594 | 0.053876442 | -0.034410152 |
| HSPB1 | -0.08371207 | 0.055051141 | -0.028660929 |
| DNAJC19 | -0.086839759 | 0.037938478 | -0.048901281 |
| ATP5F1D | -0.089268267 | 0.051209706 | -0.038058561 |
| CXCL12 | -0.098546111 | 0.07228795 | -0.026258161 |
| COX7C | -0.087775726 | 0.087592233 | -0.000183493 |
| QPRT | 0.026737522 | -0.029134807 | -0.002397285 |
| WFS1 | 0.030209098 | -0.031525108 | -0.00131601 |
| CALM2 | -0.087959709 | 0.058351743 | -0.029607966 |
| ETF1 | 0.049563924 | -0.060729928 | -0.011166004 |
| RPS28 | -0.086090425 | 0.033508796 | -0.052581629 |
| CBR1 | -0.085868913 | 0.071430057 | -0.014438856 |
| ID4 | -0.054721367 | 0.047813568 | -0.006907799 |
| ASS1 | -0.027764004 | 0.013158327 | -0.014605677 |
| TK1 | 0.014057086 | -0.014688812 | -0.000631727 |
| CAV1 | 0.011521902 | -0.009334499 | 0.002187403 |
| ATP5MC2 | -0.090552651 | 0.047899004 | -0.042653647 |
| SLC25A4 | -0.082991802 | 0.080943695 | -0.002048107 |
| IL17RB | 0.009135552 | -0.008925503 | 0.000210048 |
| SOX18 | 0.013493785 | -0.014060792 | -0.000567007 |
| NAPEPLD | 0.044553687 | -0.042171364 | 0.002382323 |
| MT3 | -0.060111535 | 0.099986591 | 0.039875056 |
| COX8A | -0.086362419 | 0.083653863 | -0.002708556 |
| SEPTIN11 | 0.041281391 | -0.047022651 | -0.00574126 |

| | | | |
|---|---|---|---|
| HSP90B1 | -0.092940126 | 0.060672452 | -0.032267674 |
| SEMA4D | 0.041913623 | -0.042273798 | -0.000360175 |
| SLC25A5 | -0.0904948 | 0.082638951 | -0.007855849 |
| CTSC | -0.076962343 | 0.058322735 | -0.018639607 |
| RAB17 | 0.059962461 | -0.053872396 | 0.006090065 |
| NDUFA1 | -0.087805519 | 0.098337168 | 0.01053165 |
| AFMID | 0.044241266 | -0.03694142 | 0.007299845 |
| RARRES2 | 0.080838271 | -0.07106919 | 0.009769081 |
| ALDH6A1 | -0.073692159 | 0.028515613 | -0.045176545 |
| ID1 | -0.056893084 | 0.051976759 | -0.004916325 |
| RPS18 | -0.081805592 | 0.031192075 | -0.050613517 |
| PHPT1 | -0.084991687 | 0.077601791 | -0.007389896 |
| LMO2 | 0.013260844 | -0.014203232 | -0.000942388 |
| PIEZO1 | 0.015593725 | -0.014935068 | 0.000658657 |
| FGF9 | -0.072711936 | 0.047655945 | -0.02505599 |
| AOX1 | -0.01603502 | 0.006833315 | -0.009201705 |
| LINC01589 | 0.009637459 | -0.009768093 | -0.000130634 |
| XIST | -0.08907157 | 0.257716647 | 0.168645076 |
| ITPKB | 0.0357744 | -0.024737507 | 0.011036894 |
| MARVELD3 | 0.020846725 | -0.022122594 | -0.001275869 |
| ALDH1A1 | -0.082302331 | 0.06967365 | -0.012628681 |
| ATP5F1A | -0.082019065 | 0.069777988 | -0.012241077 |
| EPS8L2 | 0.061638503 | -0.033491929 | 0.028146574 |
| GLRX | -0.076796003 | 0.046621453 | -0.03017455 |
| PIK3AP1 | -0.016754637 | 0.005452178 | -0.011302459 |
| MGP | 0.015363252 | -0.015002272 | 0.00036098 |
| GSTA1 | 0.062594701 | 0.028242653 | 0.090837353 |
| MYH8 | 0.006978603 | -0.006681482 | 0.000297122 |
| CYS1 | -0.082397083 | 0.04976922 | -0.032627863 |
| PECAM1 | 0.012460527 | -0.012245621 | 0.000214906 |
| ANXA2 | 0.035530741 | -0.02918346 | 0.006347281 |
| GAL3ST1 | 0.047715309 | -0.04321156 | 0.00450375 |
| CA2 | 0.065607256 | -0.045795496 | 0.01981176 |
| AC124017.1 | -0.064452561 | 0.038759999 | -0.025692561 |
| ATP5MC1 | -0.085272788 | 0.131439124 | 0.046166336 |
| LINC01197 | -0.025422805 | 0.024593462 | -0.000829342 |
| GAL | 0.00580355 | -0.00580355 | -3.55271E-15 |
| ERVMER34-1 | 0.009099906 | -0.008947167 | 0.000152739 |
| B4GALNT3 | 0.02469767 | -0.017033061 | 0.007664609 |
| CIRBP | -0.076021345 | 0.039320395 | -0.036700951 |
| KCNN3 | 0.029183558 | -0.035367054 | -0.006183495 |
| DDX3Y | 0.02456122 | -0.041923136 | -0.017361916 |
| CLIC4 | 0.017732534 | -0.016433318 | 0.001299216 |
| GGH | 0.028153837 | -0.015055699 | 0.013098138 |
| ITM2A | -0.039906627 | 0.021785299 | -0.018121327 |
| F2R | 0.008749103 | -0.009319263 | -0.00057016 |
| SDC4 | -0.070277163 | 0.080833255 | 0.010556092 |

| | | | |
|---|---|---|---|
| PTGDS | -0.039019223 | 0.026170351 | -0.012848872 |
| HES4 | -0.051090334 | 0.056251678 | 0.005161344 |
| EIF1B | -0.066353498 | 0.023330546 | -0.043022952 |
| KNL1 | 0.00865048 | -0.008564288 | 8.61928E-05 |
| RPL36A | -0.081710686 | 0.059437731 | -0.022272955 |
| COL12A1 | 0.011766908 | -0.011707031 | 5.98773E-05 |
| GP2 | -0.058992697 | 0.04542624 | -0.013566457 |
| RPL35A | -0.082168039 | 0.031327889 | -0.05084015 |
| NDUFB10 | -0.078600749 | 0.068890786 | -0.009709963 |
| COX6C | -0.079123341 | 0.066862328 | -0.012261013 |
| UQCRH | -0.082525698 | 0.061221808 | -0.021303889 |
| C11orf54 | -0.067406119 | 0.029556216 | -0.037849903 |
| ARID5B | -0.04426635 | 0.024915157 | -0.019351193 |
| GPT | -0.019709269 | 0.012691795 | -0.007017473 |
| FAM167B | 0.009039885 | -0.007657281 | 0.001382603 |
| RGS6 | -0.015778249 | 0.007500403 | -0.008277847 |
| ATP2A3 | 0.011856667 | -0.012964676 | -0.001108009 |
| PIK3R3 | 0.019194649 | -0.015413317 | 0.003781331 |
| CTAG2 | 0.006036702 | -0.006036702 | -3.55271E-15 |
| RPS15A | -0.082314723 | 0.033978038 | -0.048336686 |
| LINC02672 | 0.015804786 | -0.008978924 | 0.006825862 |
| USP53 | -0.053570386 | 0.034749585 | -0.018820801 |
| CALM1 | 0.072665502 | -0.088957145 | -0.016291643 |
| BMP4 | -0.026034467 | 0.009860328 | -0.016174139 |
| KCNJ1 | -0.078586292 | 0.07602682 | -0.002559471 |
| GSTP1 | -0.074681528 | 0.052056731 | -0.022624797 |
| TNFRSF1B | 0.007921684 | -0.008582303 | -0.000660619 |
| ERBB4 | -0.067728588 | 0.069083567 | 0.001354979 |
| HMGCS2 | 0.017705147 | -0.020134042 | -0.002428895 |
| LINC01205 | 0.015857284 | -0.016199347 | -0.000342063 |
| SCD5 | -0.068335201 | 0.060952391 | -0.007382809 |
| ITIH5 | 0.011100911 | -0.011604897 | -0.000503986 |
| GDF15 | 0.02963044 | -0.030658708 | -0.001028269 |
| CCL2 | 0.010875697 | -0.012889389 | -0.002013692 |
| DCXR | -0.069128077 | 0.061922905 | -0.007205172 |
| SNHG25 | -0.063686058 | 0.034561553 | -0.029124505 |
| LAMB2 | 0.044996528 | -0.023531133 | 0.021465395 |
| RPL3 | -0.070727811 | 0.033174229 | -0.037553582 |
| ATP5PF | -0.078709177 | 0.06517879 | -0.013530387 |
| FAU | -0.074701083 | 0.024697942 | -0.050003141 |
| B3GNT3 | 0.005654734 | -0.005416595 | 0.000238139 |
| SLC14A2 | 0.005792994 | -0.005549968 | 0.000243026 |
| FBXO21 | -0.054182612 | 0.027683356 | -0.026499256 |
| EHF | 0.030818748 | -0.02715707 | 0.003661678 |
| RPS8 | -0.075488856 | 0.027231479 | -0.048257377 |
| NDUFB1 | -0.078871642 | 0.097096782 | 0.01822514 |
| ATP5MC3 | -0.073432473 | 0.106389717 | 0.032957244 |

| | | | |
|---|---|---|---|
| RPL23A | -0.073307755 | 0.044538699 | -0.028769056 |
| ATP5ME | -0.077619876 | 0.075029623 | -0.002590253 |
| UACA | 0.033948492 | -0.03003837 | 0.003910122 |
| SOX17 | 0.006782137 | -0.006693279 | 8.88576E-05 |
| DPP10 | 0.004986473 | -0.004629334 | 0.000357139 |
| PLAU | -0.078832347 | 0.040160804 | -0.038671543 |
| FEZ1 | -0.025998734 | 0.023544969 | -0.002453765 |
| TNFRSF12A | 0.032870529 | -0.025494505 | 0.007376024 |
| PPP4R4 | 0.004880626 | -0.004715046 | 0.00016558 |
| PLIN4 | -0.017972093 | 0.009362372 | -0.008609721 |
| NDUFB7 | -0.073865032 | 0.049073817 | -0.024791216 |
| NAPRT | -0.035987174 | 0.025105337 | -0.010881837 |
| KLF2 | 0.012248634 | -0.010865181 | 0.001383453 |
| CST3 | -0.078175579 | 0.050890826 | -0.027284753 |
| TTYH3 | 0.013681603 | -0.012942238 | 0.000739365 |
| RECQL5 | 0.026062519 | -0.031352725 | -0.005290206 |
| TSC22D3 | -0.052879833 | 0.05729388 | 0.004414047 |
| HSPE1 | -0.074105761 | 0.04990139 | -0.02420437 |
| S100A14 | -0.034314281 | 0.023994461 | -0.01031982 |
| CYP2S1 | 0.005026967 | -0.005026967 | -3.55271E-15 |
| RPL37A | -0.069671009 | 0.076745315 | 0.007074307 |
| TPM2 | 0.014268917 | -0.009363301 | 0.004905616 |
| RAB15 | 0.013676427 | -0.008488241 | 0.005188185 |
| PGGHG | 0.010005735 | -0.0082013 | 0.001804435 |
| NDUFS6 | -0.07373283 | 0.060113582 | -0.013619248 |
| RP1 | -0.031500197 | 0.016920136 | -0.014580061 |
| NEB | -0.032005216 | 0.033868577 | 0.001863361 |
| GAS7 | 0.007503865 | -0.007297386 | 0.000206479 |
| LNPEP | 0.05218916 | -0.036647956 | 0.015541204 |
| RPL23 | -0.074160166 | 0.028184923 | -0.045975242 |
| TPM4 | 0.020281708 | -0.021918031 | -0.001636323 |
| AKAP12 | 0.009852999 | -0.008192595 | 0.001660405 |
| CHCHD10 | -0.073250502 | 0.093684462 | 0.02043396 |
| CLCN5 | -0.072170293 | 0.054241213 | -0.01792908 |
| HIST1H1C | 0.03583063 | -0.056321733 | -0.020491103 |
| NRAD1 | 0.018419897 | -0.019761341 | -0.001341444 |
| TMEM132A | 0.020224953 | -0.010401879 | 0.009823074 |
| RPS25 | -0.073366711 | 0.025329689 | -0.048037022 |
| F8A3 | 0.011360012 | -0.004650905 | 0.006709107 |
| ZBTB16 | -0.027530032 | 0.021826906 | -0.005703126 |
| NDUFB9 | -0.071264905 | 0.100815396 | 0.029550491 |
| HES1 | -0.054334288 | 0.056559055 | 0.002224766 |
| CNN3 | 0.056049124 | -0.039536282 | 0.016512842 |
| UNC13C | 0.01452065 | -0.018232199 | -0.003711549 |
| CDC42EP3 | 0.026469497 | -0.027103041 | -0.000633544 |
| GSN | 0.046246783 | -0.016553689 | 0.029693094 |
| ATP5MD | -0.076269088 | 0.077484958 | 0.00121587 |

| | | | |
|---|---|---|---|
| NUDT4 | -0.074293249 | 0.048023061 | -0.026270188 |
| KCNE4 | 0.004463675 | -0.004435716 | 2.79597E-05 |
| ACAT1 | -0.063972848 | 0.045550984 | -0.018421864 |
| SERPINE2 | -0.019006669 | 0.01531772 | -0.003688949 |
| SPECC1 | 0.013101109 | -0.013073747 | 2.73622E-05 |
| MLPH | -0.013861389 | 0.005831028 | -0.008030361 |
| BX890604.2 | -0.022826286 | 0.015015168 | -0.007811117 |
| IFIT1 | -0.038297857 | 0.025783846 | -0.012514011 |
| RNASE1 | 0.011461618 | -0.008456137 | 0.003005481 |
| HRG | -0.046142349 | 0.033324029 | -0.01281832 |
| ETS1 | 0.010494543 | -0.0113318 | -0.000837257 |
| EIF1AY | 0.023791896 | -0.050640059 | -0.026848162 |
| KCNJ12 | -0.020796763 | 0.012621133 | -0.008175631 |
| BCAT2 | -0.028775515 | 0.015773463 | -0.013002052 |
| PROM1 | 0.022199958 | -0.017112555 | 0.005087402 |
| HERPUD1 | -0.065245028 | 0.048850779 | -0.016394249 |
| ADGRF1 | 0.034204559 | -0.014995894 | 0.019208665 |
| CYYR1 | 0.016085588 | -0.017195109 | -0.00110952 |
| ATP5PB | -0.063616487 | 0.031380536 | -0.032235951 |
| FAM43B | 0.014873681 | -0.013468522 | 0.001405159 |
| C9orf24 | -0.028308836 | 0.022608458 | -0.005700378 |
| GPX4 | -0.069384814 | 0.030954568 | -0.038430246 |
| HIGD1B | 0.005547605 | -0.005457122 | 9.04825E-05 |
| HSPD1 | -0.064199658 | 0.041491508 | -0.02270815 |
| ATP5PO | -0.07024949 | 0.060440593 | -0.009808897 |
| RPL21 | -0.068390522 | 0.030879 | -0.037511522 |
| ECI2 | -0.057799142 | 0.032569417 | -0.025229725 |
| WFDC1 | 0.023199319 | -0.012081523 | 0.011117796 |
| MAT2A | -0.043062984 | 0.024904842 | -0.018158142 |
| NDUFB11 | -0.068069364 | 0.028424262 | -0.039645102 |
| RPL15 | -0.068625575 | 0.032801297 | -0.035824278 |
| ADGRL3 | -0.018204478 | 0.009188083 | -0.009016395 |
| RAB6B | -0.02404318 | 0.020033764 | -0.004009416 |
| VAMP5 | 0.017524629 | -0.019996808 | -0.00247218 |
| CALR | -0.071502573 | 0.077836909 | 0.006334336 |
| LINC01594 | -0.008927757 | 0.004617465 | -0.004310292 |
| CX3CL1 | 0.024322155 | -0.016898519 | 0.007423636 |
| TSPAN15 | -0.029418232 | 0.019328841 | -0.010089391 |
| PMM1 | -0.049757037 | 0.049143126 | -0.000613912 |
| SLC47A2 | 0.005153913 | -0.004903156 | 0.000250757 |
| MRPL41 | -0.069936219 | 0.064811748 | -0.005124471 |
| EPHX1 | 0.028635534 | -0.019399055 | 0.009236478 |
| GABRE | 0.004945304 | -0.005173249 | -0.000227945 |
| TMCC3 | 0.009745323 | -0.011265978 | -0.001520655 |
| METRN | -0.051504977 | 0.029209162 | -0.022295814 |
| HSPA5 | -0.06899854 | 0.064028816 | -0.004969724 |
| ZMAT3 | 0.033961731 | -0.039673949 | -0.005712218 |

| | | | |
|---|---|---|---|
| MICOS10 | -0.06888637 | 0.036803066 | -0.032083303 |
| TMEM54 | 0.016533494 | -0.0161124 | 0.000421093 |
| ERICH1 | -0.043475547 | 0.031375792 | -0.012099755 |
| RAMP2 | 0.013414558 | -0.012154381 | 0.001260177 |
| ATP1A1 | -0.072004154 | 0.090563727 | 0.018559573 |
| PTGES | 0.0067117 | -0.00712286 | -0.00041116 |
| HOXA9 | 0.042560364 | -0.022635607 | 0.019924757 |
| UBC | 0.06341824 | -0.028140694 | 0.035277546 |
| RPL8 | -0.064423386 | 0.025399331 | -0.039024055 |
| RGS5 | 0.013772209 | -0.012390665 | 0.001381544 |
| SUCLG1 | -0.061209539 | 0.057198175 | -0.004011364 |
| STK17A | 0.0267589 | -0.023598031 | 0.003160869 |
| CEBPD | 0.046307449 | -0.046921291 | -0.000613842 |
| HOMER3 | 0.023776475 | -0.024042257 | -0.000265781 |
| NANOS1 | 0.025401379 | -0.020629828 | 0.004771552 |
| TUBB2A | -0.047897645 | 0.066636102 | 0.018738457 |
| PKN1 | 0.024022908 | -0.025697282 | -0.001674374 |
| OTULINL | 0.02029608 | -0.020469627 | -0.000173546 |
| ATP5F1E | -0.066859326 | 0.050740167 | -0.016119159 |
| NDUFA13 | -0.065630074 | 0.054492761 | -0.011137313 |
| ADHFE1 | -0.025606528 | 0.015288194 | -0.010318334 |
| MT-ND6 | -0.063239502 | 0.047140143 | -0.01609936 |
| FTH1 | -0.062633441 | 0.024715148 | -0.037918294 |
| SHISA8 | 0.010984943 | -0.01258075 | -0.001595807 |
| SLC38A2 | 0.040163881 | -0.05162024 | -0.01145636 |
| ASIC2 | 0.003897699 | -0.003975693 | -7.79944E-05 |
| LIMA1 | 0.030295169 | -0.038759843 | -0.008464674 |
| FAM107A | 0.017334026 | -0.015575674 | 0.001758352 |
| PHGDH | -0.040170251 | 0.028185345 | -0.011984907 |
| ATP5F1C | -0.063975684 | 0.056838152 | -0.007137532 |
| H19 | 0.005598042 | -0.00523712 | 0.000360922 |
| THY1 | -0.065248366 | 0.049152846 | -0.01609552 |
| SCGB2A1 | 0.009059959 | -0.011799304 | -0.002739346 |
| MVK | 0.027257268 | -0.018847124 | 0.008410145 |
| AC009244.2 | 0.00452996 | -0.004763148 | -0.000233189 |
| ADGRE5 | 0.006702337 | -0.006496841 | 0.000205496 |
| CCDC69 | 0.008509223 | -0.009359587 | -0.000850364 |
| HSPG2 | 0.028237215 | -0.03243621 | -0.004198995 |
| GOLGA8B | -0.041090197 | 0.037400455 | -0.003689742 |
| TMA7 | -0.069869965 | 0.048717165 | -0.0211528 |
| CA12 | -0.073609691 | 0.073378084 | -0.000231607 |
| LGALS1 | 0.012168212 | -0.010405679 | 0.001762533 |
| CPT1A | 0.04356187 | -0.02402213 | 0.01953974 |
| DUSP1 | 0.027127106 | -0.03349228 | -0.006365174 |
| PDZK1 | 0.020489535 | -0.024105554 | -0.003616019 |
| COX5B | -0.064630211 | 0.094173293 | 0.029543081 |
| GCGR | -0.04957842 | 0.048789268 | -0.000789152 |

| | | | |
|---|---|---|---|
| RPL35 | -0.062557719 | 0.032431952 | -0.030125767 |
| FBRSL1 | -0.038966853 | 0.021173385 | -0.017793468 |
| KRT10 | -0.059934567 | 0.036547121 | -0.023387446 |
| CLCN7 | 0.036214063 | -0.038282745 | -0.002068683 |
| RNASEK | -0.063449666 | 0.026842363 | -0.036607303 |
| NUPR1 | 0.013006313 | -0.011107459 | 0.001898854 |
| RPL14 | -0.067061739 | 0.02550308 | -0.04155866 |
| ECH1 | 0.051427373 | -0.022687823 | 0.028739549 |
| SEC14L1 | 0.042876715 | -0.059279682 | -0.016402967 |
| RDH10 | 0.031528813 | -0.034341709 | -0.002812896 |
| BCAT1 | -0.009030811 | 0.003804518 | -0.005226293 |
| MICOS13 | -0.062181602 | 0.04649306 | -0.015688541 |
| TRNP1 | -0.030174503 | 0.022810324 | -0.007364179 |
| MAST4 | 0.038022391 | -0.032695833 | 0.005326558 |
| RGS3 | 0.023129327 | -0.015089883 | 0.008039444 |
| TMEM161B-AS1 | -0.052793984 | 0.042163751 | -0.010630233 |
| CHST6 | 0.003600622 | -0.003419957 | 0.000180665 |
| RGS19 | 0.00933613 | -0.008805395 | 0.000530735 |
| B4GALNT2 | 0.017848298 | -0.007768982 | 0.010079316 |
| NAAA | -0.041374145 | 0.028447402 | -0.012926743 |
| EIF3K | -0.063837155 | 0.037330325 | -0.02650683 |
| INPP1 | 0.017258485 | -0.01669607 | 0.000562415 |
| ELF5 | 0.00628173 | -0.00765694 | -0.00137521 |
| DOK7 | 0.00473805 | -0.00516549 | -0.000427439 |
| SPRY4 | 0.005226351 | -0.003804105 | 0.001422246 |
| NSA2 | -0.047921375 | 0.033445858 | -0.014475517 |
| DNAH17 | 0.003512744 | -0.003512744 | -3.55271E-15 |
| FBN2 | 0.003439813 | -0.003385153 | 5.46604E-05 |
| PIK3R1 | -0.054922361 | 0.045024819 | -0.009897542 |
| MYH9 | 0.047725231 | -0.025778097 | 0.021947134 |
| MARCKSL1 | 0.017811876 | -0.014979802 | 0.002832074 |
| ORAI1 | 0.01202374 | -0.013800975 | -0.001777235 |
| COASY | 0.033458368 | -0.022488145 | 0.010970223 |
| PRR13 | -0.059524026 | 0.053907125 | -0.005616901 |
| FAM241A | 0.01449997 | -0.015426619 | -0.000926648 |
| LIPG | 0.011972225 | -0.00978589 | 0.002186334 |
| ZNF275 | -0.018578623 | 0.010150876 | -0.008427747 |
| MGST1 | -0.04656256 | 0.051791948 | 0.005229388 |
| GLTP | 0.046911269 | -0.046835343 | 7.59256E-05 |
| CKS2 | 0.014002345 | -0.010976458 | 0.003025888 |
| COX7A2 | -0.063843301 | 0.060864197 | -0.002979104 |
| ALKBH7 | -0.053667949 | 0.028332138 | -0.025335811 |
| DTX1 | 0.022768254 | -0.022155138 | 0.000613115 |
| GALK1 | -0.02970709 | 0.027180833 | -0.002526258 |
| STK32B | -0.029233054 | 0.02147567 | -0.007757384 |
| SDSL | -0.032203437 | 0.01770955 | -0.014493887 |
| RPL19 | -0.060232103 | 0.027833848 | -0.032398255 |

| | | | |
|---|---|---|---|
| FJX1 | 0.008068833 | -0.008116349 | -4.7516E-05 |
| AKAP1 | -0.043339835 | 0.023411226 | -0.019928609 |
| TMEM204 | 0.005965577 | -0.005079275 | 0.000886302 |
| GLS | 0.04200255 | -0.0285995 | 0.01340305 |
| MRPL15 | -0.043372872 | 0.019703792 | -0.02366908 |
| CHPT1 | -0.035670236 | 0.031719595 | -0.00395064 |
| PRDX2 | -0.060950474 | 0.061637257 | 0.000686783 |
| GMDS | -0.043615818 | 0.03309198 | -0.010523838 |
| SAT1 | 0.057220124 | -0.062878254 | -0.00565813 |
| EPHA2 | 0.010879243 | -0.007673229 | 0.003206014 |
| PGM5 | 0.006699681 | -0.008935457 | -0.002235776 |
| HDDC2 | -0.049357246 | 0.045975057 | -0.003382189 |
| GUCY1A1 | -0.034323027 | 0.018930866 | -0.015392161 |
| FECH | -0.052847656 | 0.044715062 | -0.008132594 |
| ARHGAP24 | -0.059142683 | 0.052685182 | -0.006457501 |
| GIMAP7 | 0.006342614 | -0.007185459 | -0.000842845 |
| CHST1 | 0.004336934 | -0.004452031 | -0.000115096 |
| CTSO | -0.038592022 | 0.0199115 | -0.018680522 |
| TOMM7 | -0.062108265 | 0.045797202 | -0.016311063 |
| RPL36 | -0.060918793 | 0.031405099 | -0.029513694 |
| NETO2 | 0.006040489 | -0.004923468 | 0.001117021 |
| PROM2 | -0.051221628 | 0.047047161 | -0.004174468 |
| CDH4 | 0.002702378 | -0.002702378 | -3.55271E-15 |
| UTRN | 0.041224207 | -0.04121865 | 5.55671E-06 |
| A2M | 0.009819086 | -0.009936694 | -0.000117609 |
| AKR1C3 | 0.008327885 | -0.006301612 | 0.002026273 |
| AMPD2 | 0.022791291 | -0.016233919 | 0.006557372 |
| AMFR | 0.045494491 | -0.046258709 | -0.000764218 |
| MDH1 | -0.05288295 | 0.043532151 | -0.009350799 |
| CUTA | -0.056349937 | 0.022942688 | -0.033407249 |
| CXCL14 | -0.066383542 | 0.065206421 | -0.001177121 |
| AC009509.1 | 0.003108344 | -0.003108344 | -3.55271E-15 |
| COMMD6 | -0.061384178 | 0.042546065 | -0.018838113 |
| GAPDH | -0.058461257 | 0.084478127 | 0.02601687 |
| TUBB2B | -0.049644649 | 0.092942108 | 0.043297459 |
| GIMAP4 | 0.005832194 | -0.005240537 | 0.000591657 |
| PDGFB | 0.008964074 | -0.009660744 | -0.00069667 |
| H3F3B | -0.060222039 | 0.041230843 | -0.018991196 |
| RPL27A | -0.057669286 | 0.051814737 | -0.005854549 |
| NDUFAB1 | -0.057105754 | 0.03896805 | -0.018137703 |
| P4HA1 | 0.020983244 | -0.016587634 | 0.00439561 |
| NPR1 | 0.005978934 | -0.005990975 | -1.20406E-05 |
| DLC1 | 0.00920106 | -0.009750116 | -0.000549056 |
| FBLN2 | 0.006249182 | -0.006477364 | -0.000228182 |
| MRPS6 | 0.059954981 | -0.12373264 | -0.063777659 |
| TGFB2 | 0.004254671 | -0.00438812 | -0.000133449 |
| PDXK | -0.054826839 | 0.044566881 | -0.010259958 |

| | | | |
|---|---|---|---|
| ENTPD8 | 0.003556791 | -0.003227313 | 0.000329478 |
| PMEPA1 | 0.024417037 | -0.015477181 | 0.008939855 |
| ZNF208 | -0.017834176 | 0.01565983 | -0.002174346 |
| HEG1 | 0.00697994 | -0.007078398 | -9.84583E-05 |
| CKMT2 | -0.019366435 | 0.0168104 | -0.002556035 |
| SMIM5 | 0.021000527 | -0.021737885 | -0.000737358 |
| ORAI2 | 0.024933646 | -0.02120502 | 0.003728626 |
| TMEM53 | -0.022444944 | 0.014752324 | -0.00769262 |
| TMED3 | 0.032288596 | -0.019101637 | 0.013186959 |
| DENND1B | 0.031607203 | -0.021214223 | 0.01039298 |
| AGFG2 | 0.015029286 | -0.022027986 | -0.0069987 |
| AQP1 | 0.014026748 | -0.015224929 | -0.001198181 |
| HOXB5 | -0.029625012 | 0.019141853 | -0.010483158 |
| RHOJ | 0.003845986 | -0.003567692 | 0.000278295 |
| PCBP2 | -0.058584785 | 0.037424483 | -0.021160302 |
| TSC22D4 | 0.040578776 | -0.035047227 | 0.00553155 |
| GIMAP5 | 0.0044578 | -0.004066296 | 0.000391504 |
| SMOC2 | 0.006148726 | -0.007603994 | -0.001455268 |
| RAPGEF5 | 0.006594588 | -0.00670044 | -0.000105852 |
| EDIL3 | 0.006782839 | -0.005637585 | 0.001145255 |
| MT1F | -0.037158945 | 0.024884404 | -0.012274541 |
| MXD4 | 0.040789942 | -0.023837541 | 0.016952401 |
| ACSS1 | -0.045639923 | 0.040504873 | -0.00513505 |
| TSPAN7 | -0.037828969 | 0.030538013 | -0.007290956 |
| HOPX | 0.004751581 | -0.00421688 | 0.0005347 |
| AL353152.1 | 0.005159148 | -0.004193249 | 0.000965899 |
| WEE1 | -0.028076654 | 0.012740676 | -0.015335978 |
| ERG | 0.004912912 | -0.004768548 | 0.000144363 |
| BDH1 | -0.030867632 | 0.015956283 | -0.014911349 |
| IGFBP7 | -0.063343693 | 0.063102715 | -0.000240978 |
| PRRG4 | 0.028811947 | -0.03243801 | -0.003626063 |
| LIX1 | 0.006951834 | -0.007195224 | -0.000243389 |
| NT5DC2 | -0.029895862 | 0.035896604 | 0.006000741 |
| IFRD1 | 0.024424231 | -0.016777752 | 0.007646479 |
| FREM1 | -0.028733929 | 0.01619544 | -0.01253849 |
| MIR222HG | -0.0163852 | 0.007650894 | -0.008734306 |
| TBC1D3D | 0.00419905 | -0.005458484 | -0.001259434 |
| ECI1 | -0.04865591 | 0.036411515 | -0.012244395 |
| SLC2A4RG | -0.043620223 | 0.04693805 | 0.003317827 |
| FAT3 | 0.003473313 | -0.003318174 | 0.000155139 |
| MIR4458HG | -0.040982746 | 0.025923668 | -0.015059078 |
| LOXL2 | 0.005016005 | -0.004426381 | 0.000589624 |
| EPS8L1 | 0.025155669 | -0.012809291 | 0.012346378 |
| PCYT2 | -0.034734081 | 0.031190788 | -0.003543293 |
| FMC1 | -0.050470913 | 0.049325027 | -0.001145886 |
| EMP2 | 0.007554895 | -0.010198889 | -0.002643994 |
| AHI1 | 0.026319564 | -0.030590161 | -0.004270597 |

| | | | |
|---|---|---|---|
| COX6B1 | -0.058730648 | 0.056651802 | -0.002078846 |
| ALDH2 | -0.046800342 | 0.048513088 | 0.001712746 |
| UQCR11 | -0.059539787 | 0.071592652 | 0.012052865 |
| TIMP3 | 0.028604553 | -0.013620926 | 0.014983627 |
| HAGH | 0.032817872 | -0.027731363 | 0.005086509 |
| SESN3 | 0.019491125 | -0.014579936 | 0.004911188 |
| CHGB | 0.00765833 | -0.004702983 | 0.002955347 |
| SRSF5 | -0.051527985 | 0.057538093 | 0.006010108 |
| RPL31 | -0.051405322 | 0.034405671 | -0.016999651 |
| TAGLN | 0.011719402 | -0.008243121 | 0.003476281 |
| NDUFV3 | -0.044657658 | 0.03563721 | -0.009020448 |
| SLC35E2A | -0.02248275 | 0.010086132 | -0.012396618 |
| CLSTN2 | 0.00668396 | -0.005639219 | 0.001044741 |
| RUNX1 | 0.009738216 | -0.011620217 | -0.001882001 |
| TIMP1 | 0.008656308 | -0.008824692 | -0.000168384 |
| NDUFC2 | -0.057407419 | 0.028782962 | -0.028624456 |
| NDUFA3 | -0.056685901 | 0.07300995 | 0.016324049 |
| ABCB1 | 0.013788801 | -0.00656513 | 0.00722367 |
| NRGN | 0.008711863 | -0.005545062 | 0.003166801 |
| FERMT2 | 0.031624694 | -0.027077461 | 0.004547233 |
| FOXA3 | 0.002120344 | -0.002120344 | -5.32907E-15 |
| ITGB1 | 0.044726578 | -0.038216961 | 0.006509617 |
| TJP2 | -0.02868707 | 0.013428389 | -0.01525868 |
| ALDOB | -0.014842155 | 0.010534978 | -0.004307177 |
| TM4SF1 | 0.007527102 | -0.00902758 | -0.001500478 |
| FGD5 | 0.004309404 | -0.004352099 | -4.26945E-05 |
| RPS21 | -0.056029896 | 0.035727462 | -0.020302435 |
| PAK2 | 0.030846854 | -0.040742668 | -0.009895814 |
| FHOD3 | 0.027761971 | -0.02155828 | 0.00620369 |
| COTL1 | 0.015883042 | -0.021965526 | -0.006082484 |
| SGK1 | 0.011361403 | -0.009816038 | 0.001545366 |
| KCNMB2 | -0.017815795 | 0.010495769 | -0.007320026 |
| PGM1 | -0.033164905 | 0.032953561 | -0.000211343 |
| DMAC1 | -0.044139354 | 0.027280916 | -0.016858438 |
| KIF26A | -0.019897483 | 0.018077667 | -0.001819816 |
| COL4A4 | 0.037287388 | -0.029547891 | 0.007739497 |
| TRPS1 | 0.024956287 | -0.027065886 | -0.002109599 |
| ABCA4 | 0.013948467 | -0.018185102 | -0.004236635 |
| HPS4 | 0.025099767 | -0.026028818 | -0.000929051 |
| CACNA1C | 0.005649808 | -0.00464801 | 0.001001798 |
| METTL7A | -0.050317469 | 0.024134578 | -0.026182891 |
| HPN | -0.043991779 | 0.025139409 | -0.018852369 |
| STOX2 | -0.016832897 | 0.009075761 | -0.007757136 |
| KLHDC7A | -0.029424079 | 0.03561252 | 0.006188442 |
| ANXA1 | 0.006815877 | -0.007732091 | -0.000916214 |
| CHID1 | -0.040158443 | 0.035843896 | -0.004314547 |
| BDKRB2 | 0.023988772 | -0.018880863 | 0.005107909 |

| | | | |
|---|---|---|---|
| DPP10-AS1 | 0.003565844 | -0.003839095 | -0.000273251 |
| LDB2 | 0.005315204 | -0.005049199 | 0.000266005 |
| TLR5 | 0.02206643 | -0.023117576 | -0.001051146 |
| COL9A2 | 0.00850355 | -0.011054193 | -0.002550643 |
| IP6K2 | 0.031759401 | -0.02284703 | 0.00891237 |
| PRDX5 | -0.054791884 | 0.044745756 | -0.010046128 |
| NTAN1 | 0.022984362 | -0.019948497 | 0.003035865 |
| RAP1GAP | -0.051399058 | 0.029568491 | -0.021830567 |
| CD34 | 0.00638923 | -0.007170993 | -0.000781763 |
| BAZ1A | 0.010103064 | -0.009363178 | 0.000739886 |
| DEPP1 | -0.017935465 | 0.009423381 | -0.008512084 |
| ACTN1 | 0.015833652 | -0.015923873 | -9.02209E-05 |
| PRRX2 | 0.008911772 | -0.005326759 | 0.003585013 |
| NT5C3A | 0.020083568 | -0.017482563 | 0.002601005 |
| FZD2 | 0.003534404 | -0.003788978 | -0.000254574 |
| DOCK1 | 0.032648532 | -0.031816822 | 0.00083171 |
| EXPH5 | 0.020946668 | -0.022202699 | -0.001256031 |
| INPP4B | 0.030601649 | -0.030246783 | 0.000354867 |
| COMT | -0.049014349 | 0.023462866 | -0.025551482 |
| EIF5 | 0.045879406 | -0.046622292 | -0.000742886 |
| NRN1 | 0.004126971 | -0.004278296 | -0.000151326 |
| MISP3 | 0.025372942 | -0.028649096 | -0.003276154 |
| HLA-DPB1 | 0.008633554 | -0.007317995 | 0.001315559 |
| SEMA6D | -0.047396533 | 0.05899734 | 0.011600807 |
| ADGRL4 | 0.006025182 | -0.006869725 | -0.000844542 |
| KCNMB4 | 0.004627328 | -0.004073673 | 0.000553655 |
| DVL3 | 0.028344728 | -0.029150025 | -0.000805297 |
| FBXL16 | -0.015155944 | 0.01133682 | -0.003819124 |
| TMEM141 | -0.051774669 | 0.04649105 | -0.005283619 |
| UBL5 | -0.056596599 | 0.029284646 | -0.027311953 |
| LGI2 | 0.006794289 | -0.005275893 | 0.001518396 |
| RCAN2 | 0.0200213 | -0.021879362 | -0.001858062 |
| SCN1B | -0.013958899 | 0.007998083 | -0.005960816 |
| ZFYVE16 | 0.031999251 | -0.023841635 | 0.008157616 |
| NDUFB8 | -0.052461917 | 0.049251039 | -0.003210877 |
| CLIP2 | 0.016828669 | -0.014423246 | 0.002405423 |
| MKLN1-AS | 0.013825612 | -0.017098724 | -0.003273113 |
| AC099552.1 | 0.002267514 | -0.002267514 | -1.77636E-15 |
| SLC43A1 | -0.022538298 | 0.014047272 | -0.008491026 |
| ERC2 | 0.005061778 | -0.004387122 | 0.000674656 |
| AC004556.3 | 0.007160105 | -0.015829492 | -0.008669386 |
| MKNK2 | -0.030513645 | 0.014290647 | -0.016222998 |
| MAFA | 0.004056188 | -0.003112423 | 0.000943764 |
| MT1G | -0.040684271 | 0.039996534 | -0.000687738 |
| PRR34-AS1 | -0.038242393 | 0.035951327 | -0.002291066 |
| ACBD5 | 0.027478377 | -0.016252388 | 0.011225989 |
| IFITM2 | 0.034358514 | -0.024502752 | 0.009855763 |

| | | | |
|---|---|---|---|
| MGST3 | -0.053874193 | 0.05978666 | 0.005912467 |
| TMEM205 | -0.045647109 | 0.044006127 | -0.001640982 |
| KRT19 | 0.013255973 | -0.016513812 | -0.003257839 |
| IL2RG | 0.003155227 | -0.002940274 | 0.000214952 |
| STK26 | 0.011552304 | -0.015435851 | -0.003883548 |
| GATM | -0.02494453 | 0.014635409 | -0.01030912 |
| RPL38 | -0.051571224 | 0.039367418 | -0.012203806 |
| CAMSAP3 | -0.01875209 | 0.009787884 | -0.008964207 |
| AL031429.1 | -0.012936939 | 0.006833134 | -0.006103805 |
| FST | 0.003131121 | -0.00300745 | 0.000123671 |
| PAWR | 0.032406108 | -0.022501034 | 0.009905074 |
| APLP2 | -0.054249376 | 0.025867418 | -0.028381958 |
| BVES | 0.008450434 | -0.00697946 | 0.001470974 |
| JCAD | 0.005255521 | -0.005036482 | 0.000219039 |
| ATP5PD | -0.052410133 | 0.057073959 | 0.004663826 |
| CDYL2 | 0.011688203 | -0.010534947 | 0.001153256 |
| IGFBP5 | 0.015751601 | -0.025125772 | -0.009374172 |
| ARHGAP18 | -0.038617166 | 0.021998052 | -0.016619114 |
| DNER | 0.019819359 | -0.01341438 | 0.006404979 |
| ADGRF3 | -0.011917665 | 0.010530089 | -0.001387576 |
| PHLDA1 | 0.007065602 | -0.009239486 | -0.002173884 |
| ATP5MPL | -0.052643797 | 0.050905912 | -0.001737885 |
| NDRG2 | -0.04820508 | 0.05454711 | 0.00634203 |
| AC104461.1 | 0.004955279 | -0.004926588 | 2.86913E-05 |
| PRSS8 | -0.041985624 | 0.030309098 | -0.011676525 |
| SLC38A1 | 0.028663716 | -0.017269277 | 0.011394439 |
| ISG15 | 0.008277787 | -0.006199726 | 0.002078061 |
| AC104083.1 | -0.017808439 | 0.009541342 | -0.008267097 |
| TGM2 | 0.004276368 | -0.004273117 | 3.25094E-06 |
| HINT2 | -0.046752723 | 0.028609144 | -0.018143579 |
| DNASE1 | -0.048341653 | 0.039522694 | -0.008818959 |
| ALDH1A3 | 0.00340815 | -0.003535664 | -0.000127515 |
| BSG | -0.053425239 | 0.084757739 | 0.0313325 |
| FAT1 | 0.030171953 | -0.025769694 | 0.00440226 |
| THSD7A | -0.033728931 | 0.021828501 | -0.01190043 |
| KANSL1-AS1 | -0.018945791 | 0.015198083 | -0.003747708 |
| PNKD | -0.048012282 | 0.024704263 | -0.023308019 |
| ALCAM | -0.019726948 | 0.013118969 | -0.006607979 |
| PTMS | 0.046445215 | -0.031407318 | 0.015037897 |
| UQCR10 | -0.052767816 | 0.055381737 | 0.002613921 |
| BCAS4 | -0.01525145 | 0.009101182 | -0.006150268 |
| PDK3 | 0.026220799 | -0.016646934 | 0.009573865 |
| SLC5A8 | 0.004151988 | -0.004448322 | -0.000296334 |
| RASGRP3 | 0.009479372 | -0.005385771 | 0.004093601 |
| KAZN | 0.026272259 | -0.025315998 | 0.000956261 |
| TCIM | -0.040659868 | 0.025619515 | -0.015040352 |
| NDUFV1 | -0.045713323 | 0.0466198 | 0.000906477 |

| | | | |
|---|---|---|---|
| EDF1 | -0.050623289 | 0.048263777 | -0.002359513 |
| ABHD5 | 0.020564029 | -0.015236294 | 0.005327736 |
| KANTR | -0.017332373 | 0.013485843 | -0.003846531 |
| SLCO1A2 | 0.002496143 | -0.002496143 | -1.77636E-15 |
| NTS | 0.002498232 | -0.002588608 | -9.03761E-05 |
| CNPPD1 | -0.032407906 | 0.026252535 | -0.00615537 |
| PTGS1 | 0.003213984 | -0.003292183 | -7.81991E-05 |
| PTBP3 | 0.022701047 | -0.031235697 | -0.008534649 |
| SLC20A2 | -0.028626442 | 0.014921144 | -0.013705299 |
| AC012368.1 | -0.016149212 | 0.013592075 | -0.002557137 |
| UQCRC2 | -0.044087381 | 0.049852155 | 0.005764774 |
| JAM2 | 0.004467569 | -0.004281681 | 0.000185888 |
| PRR15 | 0.01504065 | -0.011646232 | 0.003394417 |
| LINC02432 | -0.028348306 | 0.027050668 | -0.001297639 |
| CADM1 | 0.039400496 | -0.020037907 | 0.019362589 |
| RASSF8 | 0.031674357 | -0.051124994 | -0.019450637 |
| SLC15A1 | 0.003716287 | -0.003428802 | 0.000287486 |
| SLC16A11 | -0.010050719 | 0.00491024 | -0.005140479 |
| TUBB | -0.040572529 | 0.023337024 | -0.017235505 |
| KLHL5 | 0.016453599 | -0.011700023 | 0.004753575 |
| AC111000.4 | 0.00371436 | -0.004392762 | -0.000678402 |
| PALM2-AKAP2 | 0.014038506 | -0.016544088 | -0.002505583 |
| BCL2 | -0.025885196 | 0.01619038 | -0.009694816 |
| LRRFIP1 | 0.037604754 | -0.057331672 | -0.019726918 |
| PRDX3 | -0.048243847 | 0.047012807 | -0.00123104 |
| TDRD12 | 0.003090122 | -0.002421507 | 0.000668615 |
| ECEL1 | 0.004691821 | -0.004179869 | 0.000511951 |
| SYNE2 | 0.04922283 | -0.061692951 | -0.012470122 |
| SNCAIP | 0.002564249 | -0.00243966 | 0.000124589 |
| LINC02716 | 0.002626626 | -0.002213494 | 0.000413132 |
| GLRX5 | -0.045824391 | 0.038700772 | -0.007123619 |
| CDC42BPA | 0.02619162 | -0.027898428 | -0.001706808 |
| OPTN | 0.01858848 | -0.017071179 | 0.001517301 |
| STARD13 | 0.016747733 | -0.020686464 | -0.003938731 |
| CKM | -0.008325806 | 0.007564537 | -0.000761269 |
| SPACA9 | 0.016082938 | -0.017286187 | -0.001203249 |
| MRPL33 | -0.050291054 | 0.027794915 | -0.022496139 |
| LINC02593 | -0.006252488 | 0.004524982 | -0.001727506 |
| CDHR2 | -0.007953162 | 0.004608558 | -0.003344604 |
| ATP1B1 | -0.050308984 | 0.063408549 | 0.013099564 |
| DPYSL2 | 0.022754451 | -0.038557139 | -0.015802689 |
| VWF | 0.004422735 | -0.003641875 | 0.000780859 |
| MALT1 | 0.012633882 | -0.010868165 | 0.001765717 |
| WLS | -0.039401138 | 0.030489576 | -0.008911561 |
| MAB21L4 | 0.008078123 | -0.010201047 | -0.002122924 |
| RPS2 | -0.046614765 | 0.039715992 | -0.006898772 |
| C7orf50 | -0.039659548 | 0.024860356 | -0.014799192 |

| | | | |
|---|---|---|---|
| CD81 | -0.048275148 | 0.025356469 | -0.022918679 |
| CLDN14 | -0.026029426 | 0.028799721 | 0.002770294 |
| CLIC1 | -0.044463156 | 0.041000479 | -0.003462677 |
| AC146944.4 | -0.012166013 | 0.006566839 | -0.005599175 |
| MRPL23 | -0.027940115 | 0.039702818 | 0.011762704 |
| AC007405.3 | -0.024500779 | 0.036117294 | 0.011616515 |
| ZFYVE28 | 0.012704789 | -0.009950999 | 0.00275379 |
| ETS2 | 0.014141955 | -0.019122176 | -0.004980221 |
| SCARA3 | 0.002969189 | -0.002779374 | 0.000189815 |
| GRINA | -0.033685451 | 0.020246844 | -0.013438607 |
| LINC01762 | -0.026737697 | 0.015181606 | -0.011556092 |
| CRTAC1 | 0.003797538 | -0.003898328 | -0.00010079 |
| FUS | -0.039312147 | 0.020177126 | -0.019135021 |
| SERPINA10 | 0.001773427 | -0.001773427 | -3.55271E-15 |
| CCDC3 | 0.003059171 | -0.002739602 | 0.000319569 |
| LINC01819 | 0.002607314 | -0.002819209 | -0.000211895 |
| C1QTNF7 | -0.013372648 | 0.018792751 | 0.005420104 |
| KCNK15 | 0.002747719 | -0.002735701 | 1.20187E-05 |
| CCNY | 0.028773541 | -0.037065822 | -0.008292281 |
| EIF2AK4 | 0.021583576 | -0.019764916 | 0.00181866 |
| PYGL | -0.006228479 | 0.003827508 | -0.00240097 |
| AP002360.1 | -0.029564306 | 0.025958192 | -0.003606114 |
| PHLDA2 | 0.003241892 | -0.003708947 | -0.000467055 |
| SYBU | -0.010353342 | 0.005368755 | -0.004984587 |
| GIMAP1 | 0.004700343 | -0.004710623 | -1.02796E-05 |
| ATP5F1B | -0.046870473 | 0.077816401 | 0.030945928 |
| NDUFS4 | -0.038918131 | 0.026757579 | -0.012160551 |
| PCBD1 | -0.041506038 | 0.024603799 | -0.016902239 |
| NARF | 0.018353191 | -0.020877034 | -0.002523843 |
| GOT1 | -0.038678736 | 0.037552096 | -0.001126639 |
| PREX2 | 0.003522883 | -0.002691629 | 0.000831255 |
| PDGFD | -0.010937898 | 0.006573223 | -0.004364675 |
| SREBF1 | -0.032774847 | 0.019078453 | -0.013696394 |
| CITED2 | -0.031859814 | 0.022202919 | -0.009656894 |
| PVALB | -0.043990401 | 0.087644105 | 0.043653705 |
| SLC6A8 | -0.03230217 | 0.01831375 | -0.013988419 |
| BOLA3 | -0.037128065 | 0.04347495 | 0.006346885 |
| BCL11B | -0.013584179 | 0.010103392 | -0.003480787 |
| NEDD4 | 0.003834616 | -0.003263679 | 0.000570937 |
| DENND11 | 0.016231023 | -0.017014561 | -0.000783538 |
| TRAPPC2L | -0.032047418 | 0.017000183 | -0.015047235 |
| ATOH8 | -0.006107529 | 0.003738628 | -0.002368901 |
| KDR | 0.003601167 | -0.002767181 | 0.000833985 |
| S100A1 | 0.005394959 | -0.007385964 | -0.001991005 |
| PGPEP1 | 0.017609104 | -0.015658354 | 0.001950751 |
| BX664615.2 | -0.019502286 | 0.013805331 | -0.005696955 |
| ETFB | -0.041851337 | 0.02557752 | -0.016273817 |

| | | | |
|---|---|---|---|
| INKA1 | 0.002317106 | -0.002116966 | 0.000200141 |
| WWC3 | 0.012919679 | -0.012941136 | -2.14575E-05 |
| ITM2C | -0.041093614 | 0.061357896 | 0.020264282 |
| STAT3 | 0.033045818 | -0.034271725 | -0.001225907 |
| HS6ST1 | 0.017395295 | -0.019520688 | -0.002125393 |
| MYO9A | -0.036976112 | 0.026392935 | -0.010583177 |
| PITHD1 | 0.021660788 | -0.018463325 | 0.003197462 |
| CASC15 | 0.018066504 | -0.013846614 | 0.004219891 |
| RPS10 | 0.047554443 | -0.037775517 | 0.009778926 |
| PALLD | 0.007460058 | -0.008015316 | -0.000555258 |
| PAM16 | -0.030436151 | 0.017481655 | -0.012954496 |
| EGFL6 | -0.012587987 | 0.018684052 | 0.006096066 |
| RPL37 | -0.044964932 | 0.050003843 | 0.005038911 |
| GALNT3 | 0.020702357 | -0.019841198 | 0.000861158 |
| RGN | -0.023392547 | 0.019279297 | -0.004113249 |
| COBLL1 | 0.043282591 | -0.023628083 | 0.019654508 |
| PNMA8A | -0.01960601 | 0.015428616 | -0.004177395 |
| PCAT19 | 0.002930558 | -0.002627794 | 0.000302764 |
| TPPP | -0.014743687 | 0.00803232 | -0.006711368 |
| NR1H4 | 0.016148772 | -0.012179674 | 0.003969097 |
| LINC02767 | 0.00568677 | -0.005584449 | 0.000102321 |
| TLE1 | -0.019803637 | 0.023902167 | 0.00409853 |
| GNPNAT1 | 0.019716639 | -0.017467638 | 0.002249001 |
| RPS27 | -0.044324403 | 0.033747632 | -0.010576771 |
| FXYD4 | 0.015351008 | -0.037855798 | -0.02250479 |
| PDLIM1 | 0.006602302 | -0.007352112 | -0.000749811 |
| CCPG1 | 0.02983867 | -0.029110632 | 0.000728039 |
| TOM1 | 0.021850226 | -0.016490832 | 0.005359394 |
| COQ8A | 0.020368014 | -0.024083335 | -0.003715322 |
| SAMD8 | 0.012109307 | -0.011531034 | 0.000578273 |
| SPARC | 0.006180593 | -0.005344204 | 0.000836389 |
| TACC1 | 0.040247906 | -0.09909075 | -0.058842844 |
| ANGPTL1 | -0.027474879 | 0.036227656 | 0.008752777 |
| SLC47A1 | -0.016021824 | 0.009044583 | -0.006977241 |
| PLA2G4F | 0.01478852 | -0.012186223 | 0.002602297 |
| AC073896.2 | -0.023784213 | 0.021105343 | -0.00267887 |
| GALNT16 | -0.011907777 | 0.008648577 | -0.0032592 |
| SNU13 | -0.040955101 | 0.030754568 | -0.010200534 |
| WNT11 | 0.008887586 | -0.01254429 | -0.003656705 |
| AC023421.1 | 0.002251991 | -0.001978974 | 0.000273018 |
| HIVEP2 | 0.012030801 | -0.009963568 | 0.002067234 |
| SOX5 | 0.002992102 | -0.002614838 | 0.000377264 |
| PTPN4 | -0.030887269 | 0.018042867 | -0.012844402 |
| MCTP1 | 0.002973185 | -0.003564198 | -0.000591012 |
| TMEM150A | 0.013802314 | -0.008537037 | 0.005265277 |
| ARL15 | 0.01262725 | -0.011350554 | 0.001276696 |
| CLDND2 | 0.009135223 | -0.004971964 | 0.004163259 |

| | | | |
|---|---|---|---|
| ZNF93 | 0.006475785 | -0.006016728 | 0.000459057 |
| RNF6 | 0.016560191 | -0.011028714 | 0.005531478 |
| KIF26B | 0.004786293 | -0.004754609 | 3.16846E-05 |
| PPARG | 0.005372118 | -0.005981039 | -0.00060892 |
| NXN | 0.006719266 | -0.006858336 | -0.000139071 |
| RDH11 | 0.029786801 | -0.025481237 | 0.004305565 |
| ECSCR | 0.003067591 | -0.00310006 | -3.2469E-05 |
| GDF11 | 0.007495228 | -0.007689544 | -0.000194316 |
| SH3BP5 | 0.004902451 | -0.004405113 | 0.000497338 |
| ZNF528-AS1 | -0.01829278 | 0.01221713 | -0.00607565 |
| UPK3BL1 | 0.024614162 | 0.03632398 | 0.060938142 |
| F11 | -0.01977166 | 0.013463595 | -0.006308065 |
| CAPS | 0.015388116 | -0.019388121 | -0.004000005 |
| RNF212 | 0.019514898 | -0.02139556 | -0.001880661 |
| MAFG | 0.01288193 | -0.012082526 | 0.000799404 |
| CLDN5 | 0.004538863 | -0.005979521 | -0.001440659 |
| RIMBP2 | 0.010722517 | -0.009966702 | 0.000755816 |
| BOLA2 | 0.020681594 | -0.018023328 | 0.002658266 |
| AGPAT5 | 0.016478616 | -0.016998099 | -0.000519482 |
| CA5B | 0.009592569 | -0.01227473 | -0.002682161 |
| EBP | -0.027361185 | 0.025050866 | -0.002310319 |
| ST5 | 0.017697369 | -0.016017401 | 0.001679967 |
| SPATS2L | 0.017363248 | -0.018449527 | -0.001086279 |
| HLA-A | 0.041104745 | -0.028578092 | 0.012526653 |
| KBTBD2 | 0.014242982 | -0.012514911 | 0.001728072 |
| RAB9A | 0.021734749 | -0.02756865 | -0.0058339 |
| C16orf87 | 0.012798224 | -0.010639271 | 0.002158953 |
| LPCAT2 | 0.005266718 | -0.0057363 | -0.000469582 |
| MZT2B | -0.042583027 | 0.027222423 | -0.015360604 |
| ERMP1 | 0.025036764 | -0.024578599 | 0.000458164 |
| FGF18 | 0.002283168 | -0.002322176 | -3.90078E-05 |
| EGR1 | -0.010444005 | 0.005827389 | -0.004616617 |
| PAIP2B | -0.016126998 | 0.015336015 | -0.000790983 |
| FOXA2 | 0.001892854 | -0.001745898 | 0.000146956 |
| ITGAV | 0.027912197 | -0.034242716 | -0.00633052 |
| MIF | -0.043709905 | 0.034253939 | -0.009455966 |
| CRMP1 | 0.002243279 | -0.002375514 | -0.000132235 |
| PTPRB | 0.004715369 | -0.004477731 | 0.000237638 |
| MYL12A | 0.036897865 | -0.043041547 | -0.006143682 |
| E2F3 | 0.005227935 | -0.004675888 | 0.000552047 |
| SPP1 | -0.051715909 | -0.042717548 | -0.094433458 |
| NUDT16 | -0.022257763 | 0.018274926 | -0.003982837 |
| MRPL13 | -0.0302996 | 0.023132193 | -0.007167408 |
| TNFRSF19 | 0.008389577 | -0.006565215 | 0.001824362 |
| IMMP1L | -0.024710272 | 0.014221351 | -0.01048892 |
| PSAP | -0.045616077 | 0.042949059 | -0.002667017 |
| NYAP1 | 0.00331267 | -0.002907461 | 0.000405209 |

| | | | |
|---|---|---|---|
| POLR2L | -0.0441956 | 0.064729663 | 0.020534063 |
| CEP170 | 0.011586782 | -0.007933571 | 0.003653212 |
| SLC7A6 | 0.014912982 | -0.01269004 | 0.002222942 |
| IFI16 | 0.004942829 | -0.004623264 | 0.000319565 |
| LMTK3 | 0.016959595 | -0.021611003 | -0.004651409 |
| B2M | -0.047199386 | 0.040057712 | -0.007141673 |
| UQCRQ | -0.045508294 | 0.086553864 | 0.04104557 |
| PSMB9 | 0.007582094 | -0.006891244 | 0.00069085 |
| TCF7L1 | 0.004483182 | -0.005095526 | -0.000612345 |
| AC004918.1 | 0.008357887 | -0.00818157 | 0.000176317 |
| LHFPL6 | 0.003973739 | -0.003083961 | 0.000889778 |
| SSR3 | -0.035258849 | 0.024366379 | -0.01089247 |
| PATJ | 0.028948442 | -0.016475481 | 0.012472961 |
| CPM | -0.03528519 | 0.031160548 | -0.004124642 |
| MRPS24 | -0.041570887 | 0.032945318 | -0.008625569 |
| NUAK1 | 0.012947272 | -0.00944775 | 0.003499522 |
| MAP1B | 0.003148703 | -0.003078535 | 7.01684E-05 |
| ANAPC11 | -0.037160876 | 0.023286772 | -0.013874104 |
| VLDLR | -0.02335584 | 0.031652274 | 0.008296433 |
| BCAS1 | 0.006388337 | -0.009256092 | -0.002867755 |
| ACSL6 | 0.0147816 | -0.014850914 | -6.93143E-05 |
| SIPA1 | 0.014588434 | -0.018632885 | -0.004044451 |
| RNF165 | -0.013985783 | 0.010578645 | -0.003407138 |
| RASGRF2 | 0.004590735 | -0.005253799 | -0.000663064 |
| UBASH3B | 0.003430577 | -0.003697772 | -0.000267195 |
| SNTB2 | 0.015310756 | -0.017023283 | -0.001712527 |
| AL121603.2 | 0.003122868 | -0.003473893 | -0.000351025 |
| CLSTN1 | 0.034960949 | -0.033015665 | 0.001945284 |
| BCL6 | 0.011266849 | -0.013275844 | -0.002008996 |
| ARID5A | 0.003735536 | -0.003532301 | 0.000203235 |
| NDUFB6 | -0.035686796 | 0.043757931 | 0.008071134 |
| DBT | -0.02949422 | 0.017985015 | -0.011509205 |
| CENPW | 0.005854848 | -0.005300117 | 0.000554732 |
| MTOR | 0.018838436 | -0.014425208 | 0.004413228 |
| PDE4D | 0.016965977 | -0.015342938 | 0.001623039 |
| CDIP1 | -0.020296674 | 0.01293723 | -0.007359444 |
| DHCR24 | 0.020464337 | -0.017920533 | 0.002543805 |
| AC016831.5 | 0.008819384 | -0.006290926 | 0.002528458 |
| CD53 | 0.003116237 | -0.003095694 | 2.05425E-05 |
| LOXL4 | 0.00247543 | -0.00281203 | -0.0003366 |
| CDCA7L | 0.009031265 | -0.00851279 | 0.000518475 |
| TFAP2A | 0.011444549 | -0.007977137 | 0.003467411 |
| ACADSB | -0.02724551 | 0.015932049 | -0.011313461 |
| ZSCAN16-AS1 | -0.032861273 | 0.022438699 | -0.010422574 |
| CRYBB2 | 0.005359578 | -0.007107453 | -0.001747875 |
| EQTN | 0.001771706 | -0.001571289 | 0.000200417 |
| CCND3 | -0.018628936 | 0.013070399 | -0.005558538 |

| | | | |
|---|---|---|---|
| PHB | -0.035991734 | 0.038953834 | 0.002962101 |
| NOSTRIN | 0.00478895 | -0.003172329 | 0.001616621 |
| PDZD2 | 0.003831052 | -0.004033063 | -0.000202011 |
| DYNC2H1 | -0.021819922 | 0.018922809 | -0.002897112 |
| YPEL5 | 0.02378283 | -0.030281866 | -0.006499037 |
| B4GALT6 | 0.012870206 | -0.010411467 | 0.00245874 |
| AC027031.2 | -0.019579475 | 0.012505299 | -0.007074176 |
| ANGPTL6 | 0.004426145 | -0.00401432 | 0.000411825 |
| PTPRM | 0.016060987 | -0.014233457 | 0.00182753 |
| TFCP2L1 | -0.037600426 | 0.065474456 | 0.02787403 |
| NDUFV2 | -0.03952877 | 0.046421242 | 0.006892472 |
| CCDC151 | -0.011789695 | 0.010709034 | -0.001080661 |
| MXRA5 | -0.012781129 | 0.011077987 | -0.001703142 |
| FUCA1 | 0.026778114 | -0.035134572 | -0.008356458 |
| GLYATL1 | -0.011708378 | 0.009553215 | -0.002155164 |
| NOTCH3 | -0.024030079 | 0.031255365 | 0.007225286 |
| FOXO3 | -0.023102115 | 0.015453335 | -0.007648781 |
| LAPTM4B | 0.02966529 | -0.029877251 | -0.000211961 |
| VPS13C | 0.028690123 | -0.033139681 | -0.004449558 |
| IMMP2L | -0.027470423 | 0.017757854 | -0.009712569 |
| SUMF2 | -0.027359222 | 0.02441971 | -0.002939512 |
| AL162424.1 | 0.001689256 | -0.001689256 | -1.77636E-15 |
| RRAD | 0.008302773 | -0.006162481 | 0.002140292 |
| MPP6 | 0.014300931 | -0.013678741 | 0.00062219 |
| GPR83 | 0.001519559 | -0.001462372 | 5.71865E-05 |
| ATP5MF | -0.041435577 | 0.066614841 | 0.025179264 |
| GBP2 | 0.002503009 | -0.002244437 | 0.000258572 |
| DIPK1B | -0.021362973 | 0.021589952 | 0.000226979 |
| NQO2 | -0.02364006 | 0.031195939 | 0.007555879 |
| PLXND1 | 0.003775223 | -0.003594813 | 0.00018041 |
| GSTK1 | -0.034470675 | 0.047141071 | 0.012670396 |
| PLEKHA1 | 0.019333614 | -0.024025947 | -0.004692333 |
| LSMEM1 | 0.005518658 | -0.004295766 | 0.001222892 |
| SH3BP2 | 0.006736014 | -0.007809745 | -0.001073731 |
| NOTCH4 | 0.005241567 | -0.004519215 | 0.000722352 |
| CAB39L | 0.01239078 | -0.020480499 | -0.00808972 |
| MCM8 | 0.009155911 | -0.008971022 | 0.000184889 |
| SLIT3 | 0.003870078 | -0.00417284 | -0.000302762 |
| KCNK3 | 0.001378496 | -0.001378496 | -3.55271E-15 |
| CTDSPL | 0.031135041 | -0.039340703 | -0.008205662 |
| KHDRBS3 | -0.023857603 | 0.015632705 | -0.008224899 |
| SULT1A1 | -0.023380934 | 0.040812827 | 0.017431893 |
| TMEM130 | 0.001965702 | -0.00174295 | 0.000222752 |
| TSPAN17 | 0.011855405 | -0.008385658 | 0.003469747 |
| FOSB | -0.007544218 | -0.005064029 | -0.012608248 |
| GALNT14 | -0.022605502 | 0.021679738 | -0.000925764 |
| HPRT1 | -0.028291656 | 0.021879587 | -0.006412069 |

| | | | |
|---|---|---|---|
| SMIM22 | 0.005417069 | -0.009439659 | -0.00402259 |
| CRACR2B | -0.01768592 | 0.013200382 | -0.004485537 |
| PBX4 | -0.01077306 | 0.009046368 | -0.001726692 |
| AKR1C2 | 0.002445551 | -0.002191258 | 0.000254293 |
| LDHD | -0.016738562 | 0.014589013 | -0.00214955 |
| AMT | -0.0196095 | 0.014357489 | -0.005252011 |
| FGF2 | 0.018147076 | -0.017419742 | 0.000727334 |
| MCOLN1 | 0.015472637 | -0.013283456 | 0.002189181 |
| ENDOG | -0.028185922 | 0.026995566 | -0.001190355 |
| AL831784.1 | 0.00331074 | -0.003082426 | 0.000228314 |
| HSD17B10 | -0.028139142 | 0.02034918 | -0.007789962 |
| SEMA5B | 0.002755255 | -0.001846262 | 0.000908993 |
| SLC36A4 | 0.018614281 | -0.020716189 | -0.002101908 |
| AC005616.1 | 0.001962118 | -0.001665947 | 0.000296171 |
| NADK2 | 0.01688795 | -0.013313314 | 0.003574635 |
| FAM162A | -0.036719993 | 0.025851969 | -0.010868024 |
| PLEKHG4B | 0.006435349 | -0.009882866 | -0.003447517 |
| CCM2L | 0.002430345 | -0.001979002 | 0.000451343 |
| CPA1 | 0.004878175 | -0.003017021 | 0.001861153 |
| NBEAL1 | -0.026083693 | 0.022773037 | -0.003310656 |
| PDE2A | 0.002668538 | -0.002497422 | 0.000171117 |
| NDUFS7 | -0.037223028 | 0.041769836 | 0.004546808 |
| CAPN14 | 0.001590016 | -0.001435994 | 0.000154022 |
| NDUFA12 | -0.037774267 | 0.024262556 | -0.013511711 |
| FOLH1 | -0.010244325 | 0.006448439 | -0.003795886 |
| ACSF2 | -0.010498893 | 0.009178381 | -0.001320512 |
| AC027237.2 | 0.001751268 | -0.001582147 | 0.000169121 |
| BNIP3L | 0.017856279 | -0.025661523 | -0.007805245 |
| TENM4 | -0.010387854 | 0.0072419 | -0.003145955 |
| GIPR | 0.00173526 | -0.001915323 | -0.000180063 |
| MCEE | -0.021915579 | 0.015722445 | -0.006193134 |
| CALCRL | 0.003689019 | -0.004215273 | -0.000526253 |
| KCNIP3 | -0.013850543 | 0.015543597 | 0.001693054 |
| DCAF12L1 | 0.002872525 | -0.002835826 | 3.66995E-05 |
| HOMER1 | 0.015087499 | -0.009979131 | 0.005108368 |
| KIF1C | 0.025491674 | -0.015809642 | 0.009682032 |
| FAR2 | 0.001867384 | -0.001722877 | 0.000144507 |
| SUSD1 | 0.00655849 | -0.006082318 | 0.000476172 |
| C15orf39 | 0.004164213 | -0.003050201 | 0.001114012 |
| MCCD1 | -0.024924237 | 0.023726941 | -0.001197296 |
| SLC16A7 | -0.039511636 | 0.033641339 | -0.005870297 |
| SP5 | -0.006902682 | 0.005223339 | -0.001679342 |
| TMPRSS12 | 0.002029346 | -0.001743453 | 0.000285893 |
| LSS | -0.013933041 | 0.012348516 | -0.001584525 |
| VLDLR-AS1 | -0.010774869 | 0.006730805 | -0.004044065 |
| NDUFB3 | -0.040362968 | 0.034159793 | -0.006203175 |
| LINC00551 | 0.001665269 | -0.001665269 | -3.55271E-15 |

| | | | |
|---|---|---|---|
| NDUFA4L2 | 0.003255567 | -0.003717131 | -0.000461564 |
| RNF182 | 0.001295729 | -0.00122935 | 6.63788E-05 |
| AC104211.2 | 0.001769804 | -0.001887451 | -0.000117647 |
| C15orf61 | -0.022913649 | 0.01677143 | -0.006142219 |
| RTN3 | 0.031191855 | -0.026040584 | 0.005151272 |
| SERGEF | 0.014694636 | -0.011520247 | 0.003174389 |
| MANF | -0.028565653 | 0.036343703 | 0.007778049 |
| NPC2 | -0.033597553 | 0.023679363 | -0.00991819 |
| HSPB3 | 0.001069451 | -0.001069451 | -3.55271E-15 |
| PCDHB5 | -0.010839807 | 0.006759905 | -0.004079903 |
| SLC26A7 | 0.011266674 | -0.017889946 | -0.006623271 |
| ARHGAP31 | 0.002878742 | -0.002104702 | 0.000774039 |
| TP53I3 | -0.025505703 | 0.032830713 | 0.00732501 |
| CCN3 | 0.004582289 | -0.004823038 | -0.000240749 |
| TMEM61 | -0.023794785 | 0.017026645 | -0.00676814 |
| NDUFB4 | -0.039851006 | 0.049870597 | 0.010019591 |
| CLDN10 | -0.046963803 | 0.040030115 | -0.006933688 |
| C10orf82 | -0.006193052 | 0.008222948 | 0.002029896 |
| TLCD3A | 0.004515314 | -0.004517816 | -2.50246E-06 |
| AL096854.1 | 0.000991316 | -0.000991316 | -3.55271E-15 |
| RBM39 | -0.035241526 | 0.021866409 | -0.013375118 |
| NOTUM | 0.001059817 | -0.001059817 | -3.55271E-15 |
| TMEM246 | -0.011798633 | 0.010299447 | -0.001499186 |
| FOXK2 | 0.016339336 | -0.02086374 | -0.004524404 |
| SESTD1 | 0.016589348 | -0.011107755 | 0.005481593 |
| OGA | -0.028246931 | 0.021605832 | -0.006641099 |
| REL | 0.009615433 | -0.010149285 | -0.000533852 |
| TIFA | 0.010745823 | -0.012633076 | -0.001887253 |
| CLCNKA | 0.038972256 | -0.087584324 | -0.048612068 |
| NLRP2 | -0.006154222 | 0.00484836 | -0.001305862 |
| HIST1H2AC | 0.012882268 | -0.01800303 | -0.005120762 |
| ZNF883 | 0.011641345 | -0.009844023 | 0.001797321 |
| PDLIM4 | -0.020517747 | 0.031543522 | 0.011025775 |
| SKIL | 0.013084662 | -0.011137544 | 0.001947117 |
| SMPD3 | 0.001267977 | -0.001267977 | -3.55271E-15 |
| CALCB | 0.00108218 | -0.00108218 | -3.55271E-15 |
| RASL11B | 0.002239791 | -0.00283587 | -0.000596079 |
| NUP210 | 0.014778773 | -0.017236894 | -0.002458121 |
| TBX2-AS1 | 0.004835302 | -0.004698892 | 0.00013641 |
| CFAP70 | -0.018928981 | 0.018527625 | -0.000401356 |
| SLC16A2 | -0.017044012 | 0.012481412 | -0.0045626 |
| C12orf75 | 0.013531258 | -0.021966967 | -0.008435709 |
| ACAT2 | -0.013984659 | 0.011811622 | -0.002173037 |
| P4HTM | -0.023874534 | 0.018275379 | -0.005599154 |
| CKAP2 | 0.008821455 | -0.007744714 | 0.001076741 |
| AC002451.1 | -0.009946414 | 0.007260644 | -0.00268577 |
| CREB3L2 | -0.030898989 | 0.041125792 | 0.010226803 |

| | | | |
|---|---|---|---|
| EIF4G3 | 0.018026605 | -0.017937416 | 8.91889E-05 |
| USP24 | 0.023573844 | -0.021337318 | 0.002236526 |
| TRAPPC5 | -0.033642342 | 0.028951122 | -0.00469122 |
| COX6A1 | -0.038721992 | 0.078637413 | 0.039915421 |
| ADGRB1 | -0.006400629 | 0.010024311 | 0.003623682 |
| SLC4A1 | -0.006701721 | -0.007663314 | -0.014365035 |
| ELOB | -0.040049207 | 0.027716204 | -0.012333003 |
| TMEM191B | -0.007639885 | 0.008636425 | 0.00099654 |
| PHC2 | 0.025541576 | -0.026164833 | -0.000623256 |
| GLP2R | 0.001097357 | -0.001097357 | -5.32907E-15 |
| EDARADD | 0.001888283 | -0.001378898 | 0.000509385 |
| ANKRD36B | 0.008602446 | -0.011131813 | -0.002529366 |
| TRANK1 | 0.005338012 | -0.005285023 | 5.29892E-05 |
| TBX3 | 0.002941111 | -0.003269004 | -0.000327893 |
| STK39 | -0.01334428 | 0.009683284 | -0.003660996 |
| PMF1 | 0.022503339 | -0.015167031 | 0.007336308 |
| MRPL24 | -0.020143493 | 0.015598598 | -0.004544896 |
| TECR | -0.031148592 | 0.028060472 | -0.00308812 |
| TMEM52B | -0.049947882 | 0.036435946 | -0.013511936 |
| S100A11 | 0.035559788 | -0.046380852 | -0.010821064 |
| DNAJC4 | -0.027570751 | 0.026768652 | -0.000802098 |
| SMAGP | 0.015919035 | -0.028295829 | -0.012376793 |
| CARHSP1 | 0.015625043 | -0.022575897 | -0.006950854 |
| JUND | 0.031543145 | -0.03579204 | -0.004248895 |
| SLC29A1 | 0.015714757 | -0.01155819 | 0.004156567 |
| SPTBN1 | 0.035187116 | -0.033798279 | 0.001388837 |
| LRPAP1 | 0.030548176 | -0.0266129 | 0.003935276 |
| NIPAL3 | 0.015548026 | -0.020361573 | -0.004813547 |
| WNK4 | -0.034407647 | 0.085327206 | 0.05091956 |
| MEIS2 | 0.004425588 | -0.00423729 | 0.000188298 |
| PPP1R36 | 0.005372341 | -0.005903742 | -0.000531401 |
| ITGA7 | -0.006851827 | 0.005095289 | -0.001756538 |
| DGLUCY | -0.020528187 | 0.028101304 | 0.007573117 |
| LINC01315 | 0.004706382 | -0.004932453 | -0.000226071 |
| RASGEF1B | 0.006505173 | -0.005324778 | 0.001180395 |
| RPN1 | 0.029347256 | -0.023395265 | 0.005951991 |
| HLA-DPA1 | 0.005878431 | -0.003984113 | 0.001894318 |
| KCNC4 | -0.013152787 | 0.009780394 | -0.003372394 |
| FBLIM1 | 0.003271936 | -0.003315405 | -4.34687E-05 |
| PLXNA4 | 0.006557114 | -0.007671282 | -0.001114168 |
| MRPL16 | -0.026780591 | 0.026057738 | -0.000722853 |
| HTRA1 | 0.005331813 | -0.004833282 | 0.00049853 |
| VWCE | 0.001619018 | -0.001247423 | 0.000371594 |
| OGFOD1 | 0.013079834 | -0.01027456 | 0.002805274 |
| THAP2 | 0.012779814 | -0.009336531 | 0.003443283 |
| SPNS1 | 0.015668392 | -0.014188512 | 0.00147988 |
| SLC2A9 | 0.004379027 | -0.003921809 | 0.000457219 |

| | | | |
|---|---|---|---|
| LINGO2 | 0.003986964 | -0.002729794 | 0.00125717 |
| NDUFA8 | -0.032452606 | 0.028812328 | -0.003640278 |
| ZSWIM7 | 0.02084472 | -0.019893863 | 0.000950858 |
| SLC25A25 | 0.008985231 | -0.007955663 | 0.001029568 |
| MTCH1 | -0.030972864 | 0.031935458 | 0.000962594 |
| SRGAP2 | 0.003475873 | -0.003191016 | 0.000284857 |
| NDUFA7 | -0.03482431 | 0.056286593 | 0.021462284 |
| SRGAP2B | 0.004235219 | -0.003762018 | 0.000473201 |
| USP9Y | 0.013239037 | -0.044625275 | -0.031386238 |
| LLGL1 | 0.005369114 | -0.005014584 | 0.00035453 |
| CU638689.4 | -0.006680881 | 0.009265479 | 0.002584597 |
| AL110505.1 | 0.001155346 | -0.001088202 | 6.71433E-05 |
| TM4SF18 | 0.006438837 | -0.007980519 | -0.001541682 |
| PPARGC1A | -0.030503555 | 0.052944586 | 0.022441031 |
| DCP2 | 0.017620589 | -0.015342082 | 0.002278507 |
| AC009996.1 | 0.002492434 | -0.00189312 | 0.000599314 |
| PWWP3A | -0.020161649 | 0.02209489 | 0.001933241 |
| ZNF703 | 0.004495644 | -0.004075248 | 0.000420396 |
| ASAP3 | -0.011954065 | 0.011217438 | -0.000736627 |
| NWD1 | -0.005215702 | 0.004287148 | -0.000928554 |
| CXADR | 0.017026065 | -0.013742621 | 0.003283444 |
| PRXL2C | 0.009054302 | -0.009976436 | -0.000922135 |
| PVR | 0.009630335 | -0.008278489 | 0.001351846 |
| TRIM47 | 0.007233664 | -0.005395735 | 0.001837929 |
| FLT1 | 0.002958036 | -0.00250012 | 0.000457916 |
| ZNF385B | -0.019614313 | 0.020711305 | 0.001096992 |
| MRPS18B | -0.027522993 | 0.019360105 | -0.008162888 |
| SMIM8 | -0.017946772 | 0.012593266 | -0.005353506 |
| SCCPDH | 0.015242854 | -0.034236272 | -0.018993418 |
| ANK2 | 0.030177956 | -0.062356433 | -0.032178477 |
| NPM1 | -0.036116869 | -0.025751686 | -0.061868555 |
| SULT4A1 | 0.002128891 | -0.00215003 | -2.11381E-05 |
| ADGRL1 | 0.010440633 | -0.007959739 | 0.002480894 |
| ARRB1 | 0.006723501 | -0.010306405 | -0.003582904 |
| PXDC1 | 0.005430685 | -0.003954421 | 0.001476264 |
| NDUFC1 | -0.038027638 | 0.053058637 | 0.015030999 |
| SNHG5 | 0.024441332 | -0.033466839 | -0.009025507 |
| MUC15 | -0.033952551 | 0.022719756 | -0.011232796 |
| LSAMP | -0.011118243 | 0.010390615 | -0.000727628 |
| JPX | -0.024696202 | 0.022236928 | -0.002459274 |
| CEP104 | 0.015586458 | -0.023984556 | -0.008398098 |
| AL512662.2 | 0.001734661 | -0.002092794 | -0.000358133 |
| SLC35E2B | 0.013405519 | -0.018435351 | -0.005029832 |
| ZNF439 | -0.010027213 | 0.007668297 | -0.002358916 |
| SLITRK4 | -0.021629586 | 0.032045425 | 0.010415839 |
| ACHE | 0.002635002 | -0.002382041 | 0.000252961 |
| ZNF844 | -0.016941389 | 0.014235981 | -0.002705408 |

| | | | |
|---|---|---|---|
| ANXA7 | 0.026467565 | -0.022616849 | 0.003850716 |
| SRGAP2C | 0.004928905 | -0.003866333 | 0.001062572 |
| GPR68 | 0.001341651 | -0.001341651 | -1.77636E-15 |
| HACD3 | 0.028421496 | -0.034463019 | -0.006041523 |
| SRSF2 | -0.030099924 | 0.020296136 | -0.009803788 |
| OTUB1 | -0.020652019 | 0.018048519 | -0.002603501 |
| RBFOX3 | 0.001260484 | -0.001260484 | -1.77636E-15 |
| NUDT8 | -0.020451943 | 0.015399577 | -0.005052365 |
| BSND | 0.021822622 | -0.015070631 | 0.006751991 |
| GRAMD2B | 0.015751968 | -0.015505767 | 0.000246201 |
| PISD | 0.012649954 | -0.011108156 | 0.001541798 |
| PTPRF | -0.023761203 | 0.018258384 | -0.005502819 |
| PPM1D | 0.007915006 | -0.008453762 | -0.000538756 |
| APMAP | 0.019831078 | -0.016261037 | 0.003570041 |
| PRKN | -0.015200795 | 0.014778327 | -0.000422467 |
| AC008443.6 | 0.0016154 | -0.001261306 | 0.000354094 |
| SERPING1 | -0.027174885 | 0.026342538 | -0.000832347 |
| PCDH7 | 0.004760958 | -0.004298138 | 0.00046282 |
| ADGRF4 | 0.001124152 | -0.001124152 | -3.55271E-15 |
| GYS1 | -0.01285058 | 0.013068826 | 0.000218246 |
| FOXD4 | 0.001337434 | -0.001054254 | 0.00028318 |
| AL133230.1 | 0.00101547 | -0.00101547 | -3.55271E-15 |
| RGS12 | 0.009187339 | -0.011654587 | -0.002467249 |
| GRAP | 0.001778248 | -0.001447641 | 0.000330607 |
| TMEM238L | 0.001150844 | -0.001150844 | -3.55271E-15 |
| PHLDB1 | 0.014977362 | -0.016906203 | -0.001928841 |
| TCF15 | 0.001715462 | -0.001461757 | 0.000253704 |
| GPSM2 | 0.004360628 | -0.004935847 | -0.000575219 |
| TTTY14 | 0.009180923 | -0.024603436 | -0.015422513 |
| TMEM8B | -0.016160955 | 0.012177034 | -0.003983921 |
| KIAA2026 | 0.019115831 | -0.025209468 | -0.006093637 |
| QPCT | 0.002483537 | -0.002950428 | -0.000466891 |
| LCN2 | 0.001405511 | -0.001245298 | 0.000160213 |
| PKP4 | -0.032687798 | 0.04703828 | 0.014350483 |
| ACAD11 | -0.013156372 | 0.016588267 | 0.003431895 |
| TMEM68 | 0.015370592 | -0.015904807 | -0.000534215 |
| MFAP2 | 0.002003094 | -0.001696847 | 0.000306247 |
| AC007344.1 | 0.00115347 | -0.00103411 | 0.000119359 |
| C2CD2L | 0.008566366 | -0.010263037 | -0.001696672 |
| MEF2C | -0.018000165 | 0.017675448 | -0.000324717 |
| NUDT18 | 0.005076175 | -0.003842356 | 0.001233819 |
| OSTM1 | 0.010389856 | -0.012244294 | -0.001854438 |
| PRR18 | -0.003150173 | 0.002492539 | -0.000657634 |
| DDX3X | 0.025500319 | -0.018637702 | 0.006862617 |
| UGT8 | -0.028459822 | 0.031386727 | 0.002926905 |
| RPS16 | -0.034511295 | 0.030453626 | -0.004057669 |
| YTHDF3-AS1 | 0.011485183 | -0.00882912 | 0.002656063 |

| | | | |
|---|---|---|---|
| PDE1A | -0.033340031 | 0.024630274 | -0.008709756 |
| GNS | 0.025625849 | -0.020043993 | 0.005581856 |
| GRM1 | -0.011018506 | 0.011884267 | 0.000865761 |
| PLVAP | 0.003102314 | -0.002953809 | 0.000148505 |
| ESYT2 | 0.0162436 | -0.015012219 | 0.001231381 |
| GJA4 | 0.002111206 | -0.002351779 | -0.000240573 |
| PF4V1 | -0.008701421 | 0.011396891 | 0.00269547 |
| UBE2T | 0.003640091 | -0.002642346 | 0.000997744 |
| TRPV4 | 0.005892636 | -0.004805028 | 0.001087607 |
| RCC1 | 0.005094649 | -0.004125095 | 0.000969554 |
| TPI1 | -0.034820238 | 0.038837509 | 0.004017271 |
| TPT1-AS1 | -0.015944561 | 0.024337753 | 0.008393192 |
| SEMA3G | 0.003007778 | -0.004073424 | -0.001065646 |
| SELENOI | -0.014568914 | 0.010449791 | -0.004119123 |
| PTH2R | -0.011325238 | 0.009599679 | -0.001725559 |
| IER5L | -0.018491183 | 0.022153768 | 0.003662585 |
| AL161669.3 | -0.008580055 | 0.00658835 | -0.001991706 |
| EHD2 | 0.00254784 | -0.002162704 | 0.000385137 |
| WHAMM | 0.010831509 | -0.012157113 | -0.001325604 |
| PHF11 | 0.014678466 | -0.012155305 | 0.002523161 |
| ATP9A | 0.013970412 | -0.017115485 | -0.003145073 |
| RBM14 | -0.016546776 | 0.019071379 | 0.002524603 |
| SLC5A3 | 0.037019953 | -0.071702202 | -0.034682249 |
| CHCHD5 | -0.024945356 | 0.018362176 | -0.00658318 |
| FRMD4A | -0.020832771 | 0.01619895 | -0.004633821 |
| ELK3 | 0.005975598 | -0.005481528 | 0.00049407 |
| C9orf16 | 0.025102924 | -0.049183615 | -0.024080691 |
| HEMK1 | -0.013486066 | 0.013008412 | -0.000477654 |
| TMEM178A | 0.008743157 | -0.008774566 | -3.14089E-05 |
| ROBO4 | 0.002257428 | -0.001883803 | 0.000373625 |
| ACACA | 0.011818697 | -0.014602194 | -0.002783497 |
| DHFR | 0.010284356 | -0.012869618 | -0.002585262 |
| ELP5 | 0.017157258 | -0.015245602 | 0.001911655 |
| CA8 | 0.004823427 | -0.008966681 | -0.004143254 |
| CDK2AP1 | 0.019121205 | -0.022208627 | -0.003087422 |
| SLC26A6 | 0.005069242 | -0.004309331 | 0.000759911 |
| PROX1 | -0.023034374 | -0.020966307 | -0.044000681 |
| GAL3ST3 | 0.012893773 | -0.010471848 | 0.002421925 |
| ZGPAT | 0.010263143 | -0.008368965 | 0.001894178 |
| ANGPT2 | 0.003780809 | -0.003170943 | 0.000609865 |
| PGM5-AS1 | 0.001347858 | -0.001103745 | 0.000244112 |
| MPV17 | -0.014903145 | 0.011550659 | -0.003352486 |
| LRRK2 | 0.015844884 | -0.012361497 | 0.003483387 |
| PACC1 | 0.005260763 | -0.005088844 | 0.000171919 |
| SLC45A4 | 0.013327885 | -0.013853985 | -0.000526101 |
| PRDX6 | -0.033873091 | 0.04245108 | 0.008577989 |
| TSTD1 | -0.033310297 | 0.030774089 | -0.002536208 |

| | | | |
|---|---|---|---|
| COA6-AS1 | -0.015658412 | 0.014039857 | -0.001618555 |
| TSPAN2 | 0.001690405 | -0.001582648 | 0.000107757 |
| GATA3-AS1 | 0.004313046 | -0.003167391 | 0.001145655 |
| BX284668.5 | 0.008891635 | -0.007603562 | 0.001288073 |
| MRPL12 | -0.026860643 | 0.020430871 | -0.006429772 |
| LINC01679 | 0.00184853 | -0.002303421 | -0.000454891 |
| ZDHHC2 | 0.021109966 | -0.03125114 | -0.010141174 |
| USP2 | -0.027525127 | 0.06131471 | 0.033789582 |
| SLC2A1 | 0.014673664 | -0.014432868 | 0.000240796 |
| ZCCHC24 | 0.003659382 | -0.003578109 | 8.12728E-05 |
| EPDR1 | 0.012357681 | -0.01362494 | -0.001267259 |
| GRIP1 | 0.002973595 | -0.002411058 | 0.000562537 |
| EGFR | 0.011507674 | -0.011761509 | -0.000253835 |
| CCN1 | 0.008775521 | -0.009544861 | -0.00076934 |
| S1PR3 | 0.001641593 | -0.001438461 | 0.000203131 |
| CAPZA2 | 0.026663424 | -0.030178432 | -0.003515008 |
| DOK1 | 0.001526663 | -0.00151546 | 1.12037E-05 |
| NFKBID | 0.003688321 | -0.002689011 | 0.00099931 |
| ZMAT1 | -0.020669738 | 0.018217355 | -0.002452383 |
| DECR1 | -0.026686924 | 0.025767584 | -0.00091934 |
| AC021321.2 | 0.001349247 | -0.001143457 | 0.00020579 |
| CHST15 | 0.001175112 | -0.001105724 | 6.93885E-05 |
| SLC52A2 | 0.012210707 | -0.008835965 | 0.003374742 |
| SHE | 0.002368753 | -0.002435515 | -6.67624E-05 |
| MTUS2 | -0.003011351 | 0.002609479 | -0.000401872 |
| RBP7 | 0.010186358 | -0.010423585 | -0.000237227 |
| AC011297.1 | 0.006188603 | -0.014176932 | -0.007988328 |
| TMEM177 | -0.015233117 | 0.024292458 | 0.009059341 |
| AL139280.1 | 0.002154554 | -0.001724671 | 0.000429882 |
| GSPT1 | 0.023525576 | -0.035985651 | -0.012460075 |
| TMOD3 | 0.019204792 | -0.01617768 | 0.003027113 |
| AL109976.1 | 0.001719409 | -0.001571892 | 0.000147516 |
| NDUFS8 | -0.032275392 | 0.033494609 | 0.001219217 |
| JAG1 | 0.021339019 | -0.023149251 | -0.001810232 |
| KLHL29 | 0.001393094 | -0.001341127 | 5.19667E-05 |
| TNNI1 | 0.00330127 | -0.002502391 | 0.000798879 |
| CH25H | 0.001145525 | -0.001145525 | -3.55271E-15 |
| AL121845.4 | 0.000987479 | -0.000928769 | 5.87104E-05 |
| CFAP73 | 0.001867943 | -0.001595152 | 0.000272792 |
| MSRB2 | -0.021943397 | 0.018318641 | -0.003624757 |
| AL627171.2 | -0.010363933 | 0.00817635 | -0.002187583 |
| THAP3 | 0.013010139 | -0.015736047 | -0.002725908 |
| FAHD2B | -0.013770211 | 0.010416645 | -0.003353566 |
| DHCR7 | -0.011487252 | 0.014089357 | 0.002602106 |
| AP1S2 | 0.008989885 | -0.008983722 | 6.16362E-06 |
| MAN2A2 | 0.011182456 | -0.00875382 | 0.002428637 |
| DDX58 | 0.006474079 | -0.005318466 | 0.001155613 |

| | | | |
|---|---|---|---|
| CDK14 | 0.011179722 | -0.008678549 | 0.002501173 |
| IFITM3 | 0.026484183 | -0.019267113 | 0.00721707 |
| CYBC1 | 0.012135561 | -0.008980062 | 0.003155499 |
| AL162411.1 | -0.006165793 | 0.006168 | 2.20763E-06 |
| TXNIP | -0.036045062 | 0.094934993 | 0.058889931 |
| FHIT | -0.017786342 | 0.01726684 | -0.000519503 |
| JADE2 | 0.011777911 | -0.011809983 | -3.20717E-05 |
| SH3GL1 | 0.009460166 | -0.007873059 | 0.001587107 |
| SLC2A6 | 0.002035382 | -0.002146406 | -0.000111023 |
| RANBP9 | 0.01258679 | -0.010676329 | 0.001910461 |
| NES | 0.002600326 | -0.002419166 | 0.00018116 |
| RBPMS2 | 0.005724518 | -0.005415497 | 0.000309021 |
| CRISP2 | 0.002392668 | -0.00224345 | 0.000149218 |
| RAP1B | 0.017719425 | -0.017043037 | 0.000676388 |
| TDRD6 | 0.006955999 | -0.005586954 | 0.001369045 |
| TIMM23B-AGAP6 | 0.001690956 | -0.001389598 | 0.000301359 |
| SLC29A4 | 0.006708141 | -0.007633148 | -0.000925007 |
| FAM167A-AS1 | -0.004290521 | 0.003143175 | -0.001147346 |
| RAB18 | 0.021515034 | -0.016649967 | 0.004865067 |
| PLEKHF1 | 0.003669821 | -0.003685636 | -1.58152E-05 |
| AVEN | 0.005478545 | -0.00558389 | -0.000105345 |
| KIAA1549 | 0.006752022 | -0.005710911 | 0.00104111 |
| ACTA2 | 0.002930222 | -0.00231798 | 0.000612242 |
| ZNRF1 | 0.008072524 | -0.007484088 | 0.000588436 |
| AGPAT2 | -0.0260018 | 0.050881703 | 0.024879903 |
| AC008735.1 | 0.001361392 | -0.001042395 | 0.000318996 |
| LGALS3BP | -0.027907156 | 0.033940962 | 0.006033805 |
| C19orf12 | 0.011018842 | -0.009835534 | 0.001183308 |
| ZEB1 | 0.003927134 | -0.003341823 | 0.000585311 |
| COX5A | -0.033748896 | 0.036018531 | 0.002269635 |
| SLC9A1 | -0.012050347 | 0.01230974 | 0.000259393 |
| SPATS2 | 0.008547108 | -0.006813107 | 0.001734001 |
| AC006230.1 | -0.006410667 | 0.005359723 | -0.001050944 |
| CLCN4 | 0.006262264 | -0.005935027 | 0.000327237 |
| NPAS2 | 0.003575746 | -0.003126373 | 0.000449373 |
| VPS26B | 0.012774017 | -0.013104113 | -0.000330095 |
| DUSP6 | 0.0077285 | -0.0058794 | 0.0018491 |
| SDHB | -0.023585015 | 0.019512512 | -0.004072503 |
| AL353751.1 | -0.007067913 | 0.006254517 | -0.000813396 |
| CHPF2 | 0.008364857 | -0.006453132 | 0.001911726 |
| CDK5RAP2 | 0.011888122 | -0.011102397 | 0.000785725 |
| RTTN | 0.008872219 | -0.008224019 | 0.0006482 |
| OST4 | -0.03407172 | 0.025728764 | -0.008342956 |
| SLCO4C1 | 0.01974179 | -0.015178292 | 0.004563498 |
| SNAPC1 | 0.010237312 | -0.009945517 | 0.000291795 |
| MCCC1 | -0.017618931 | 0.016876779 | -0.000742152 |
| SLC30A1 | -0.014641206 | 0.011047788 | -0.003593418 |

| | | | |
|---|---|---|---|
| PLOD2 | 0.021960771 | -0.027106672 | -0.005145901 |
| CDC42EP2 | 0.003621952 | -0.003800279 | -0.000178328 |
| ANGPTL4 | 0.001091901 | -0.001016153 | 7.57477E-05 |
| AMPH | 0.007124957 | -0.007576472 | -0.000451515 |
| SLPI | 0.003541846 | -0.004161637 | -0.000619791 |
| ATAD3C | 0.011612593 | -0.011635642 | -2.30487E-05 |
| SPX | -0.016908419 | 0.021714403 | 0.004805983 |
| DIPK2B | 0.001451781 | -0.001589145 | -0.000137364 |
| ZBTB38 | 0.020539262 | -0.022780838 | -0.002241576 |
| CNTN1 | -0.014161766 | 0.01308901 | -0.001072756 |
| COMMD9 | 0.018128597 | -0.017208106 | 0.00092049 |
| RAPGEF3 | 0.010825335 | -0.009169178 | 0.001656157 |
| TRAF3IP1 | 0.010503384 | -0.012061905 | -0.001558521 |
| SOHLH2 | 0.001344257 | -0.001274843 | 6.94137E-05 |
| PRLR | -0.01258619 | 0.011766467 | -0.000819723 |
| DENND5B | 0.007161953 | -0.008485006 | -0.001323053 |
| DOP1B | 0.004541962 | -0.003572524 | 0.000969438 |
| LMNTD1 | 0.000916711 | -0.000916711 | -3.55271E-15 |
| PTPRT | 0.00109277 | -0.00109277 | -3.55271E-15 |
| DPP3 | 0.007658685 | -0.006075198 | 0.001583488 |
| ACOT11 | -0.017807913 | 0.015975354 | -0.001832558 |
| KCNJ16 | -0.032519394 | 0.05150445 | 0.018985056 |
| MTRF1 | 0.00969759 | -0.007460467 | 0.002237122 |
| EZH2 | 0.004325027 | -0.003613784 | 0.000711243 |
| PRCP | -0.024559033 | 0.032221345 | 0.007662312 |
| TCHH | 0.000991762 | -0.000991762 | -3.55271E-15 |
| CST5 | -0.004998241 | 0.004682265 | -0.000315976 |
| GMFG | 0.003596181 | -0.003496335 | 9.98465E-05 |
| FAM245A | 0.004804291 | -0.005644203 | -0.000839912 |
| THORLNC | 0.001363686 | -0.001428398 | -6.47118E-05 |
| ZFP36L1 | -0.028768986 | -0.027820249 | -0.056589236 |
| DMGDH | 0.014908098 | -0.012216595 | 0.002691504 |
| NOX4 | -0.008099571 | 0.007076207 | -0.001023364 |
| NME2 | -0.033734119 | 0.041327315 | 0.007593196 |
| OLFM1 | 0.001757975 | -0.002354207 | -0.000596232 |
| ITGB2-AS1 | 0.0008298 | -0.0008298 | -3.55271E-15 |
| LYNX1 | 0.010657267 | -0.012057263 | -0.001399996 |
| AC069503.1 | 0.006762927 | -0.006248593 | 0.000514334 |
| CMC1 | -0.023600976 | 0.025022131 | 0.001421155 |
| KSR1 | 0.010600399 | -0.008480556 | 0.002119843 |
| C21orf62 | -0.019077018 | 0.025513833 | 0.006436815 |
| ACVRL1 | 0.001894952 | -0.001537695 | 0.000357258 |
| DLG4 | 0.002518315 | -0.003280527 | -0.000762212 |
| NTNG1 | 0.000997134 | -0.000997134 | -5.32907E-15 |
| ARSA | 0.008887201 | -0.00934635 | -0.000459149 |
| FHL3 | 0.004583591 | -0.003523724 | 0.001059866 |
| AC016240.1 | 0.001689472 | -0.001492532 | 0.00019694 |

| | | | |
|---|---|---|---|
| ARL4A | 0.008150124 | -0.009878975 | -0.001728851 |
| CABP1 | -0.023530997 | 0.079891425 | 0.056360428 |
| CASC18 | 0.000673926 | -0.000673926 | -1.77636E-15 |
| AL583827.1 | 0.000667501 | -0.000667501 | -3.55271E-15 |
| C1QBP | -0.025361475 | 0.035196808 | 0.009835333 |
| ABHD11 | -0.015560013 | 0.01196091 | -0.003599103 |
| GRIN3B | 0.0008489 | -0.0008489 | -3.55271E-15 |
| SRSF1 | -0.02435853 | 0.025717311 | 0.00135878 |
| NDUFS5 | -0.032360319 | 0.042482229 | 0.010121911 |
| DBF4 | 0.00532628 | -0.006309981 | -0.000983702 |
| FURIN | 0.014685616 | -0.014367542 | 0.000318074 |
| FXYD5 | 0.002957316 | -0.003058803 | -0.000101486 |
| DNAJC10 | 0.020064415 | -0.015728434 | 0.004335981 |
| PCBP3 | 0.003537957 | -0.005208003 | -0.001670045 |
| PXK | 0.012900308 | -0.014192765 | -0.001292457 |
| SOWAHC | 0.00932921 | -0.009887684 | -0.000558475 |
| HCN4 | 0.001136809 | -0.001094537 | 4.22723E-05 |
| HPS1 | 0.011984722 | -0.012162952 | -0.00017823 |
| NAA38 | -0.030791092 | 0.027992282 | -0.00279881 |
| PNRC2 | -0.023698807 | -0.021630435 | -0.045329241 |
| CLK1 | -0.021860297 | 0.020397566 | -0.001462731 |
| LINC02568 | 0.004368183 | -0.004777251 | -0.000409067 |
| CYP1A1 | 0.000748719 | -0.000748719 | -3.55271E-15 |
| UBR4 | 0.010968108 | -0.011521717 | -0.00055361 |
| MLLT11 | 0.00438684 | -0.003896461 | 0.000490379 |
| TBC1D1 | -0.019440528 | 0.015189086 | -0.004251442 |
| FBLN7 | 0.000896798 | -0.000896798 | -5.32907E-15 |
| PCDH17 | 0.002639584 | -0.0031575 | -0.000517917 |
| SGCG | 0.000810105 | -0.000810105 | -5.32907E-15 |
| BPHL | -0.013067432 | 0.011768728 | -0.001298704 |
| FAM110A | 0.0020512 | -0.001693236 | 0.000357964 |
| AC007993.3 | 0.001400356 | -0.001415272 | -1.49162E-05 |
| XBP1 | -0.022541628 | 0.023122877 | 0.00058125 |
| HK2 | 0.000808795 | -0.000808795 | -5.32907E-15 |
| ARNTL2 | 0.001006043 | -0.001006043 | -3.55271E-15 |
| ATP2B3 | -0.006610545 | 0.011483518 | 0.004872974 |
| CSRP2 | -0.027118211 | 0.0219654 | -0.00515281 |
| SAMD1 | 0.01426447 | -0.012602761 | 0.001661709 |
| POLN | -0.009383577 | 0.01538164 | 0.005998063 |
| ANXA5 | 0.028121829 | -0.02874465 | -0.000622821 |
| CADPS2 | 0.016198248 | -0.023450056 | -0.007251808 |
| TMED5 | 0.017261459 | -0.026011976 | -0.008750516 |
| PIK3C2B | 0.002629842 | -0.003265843 | -0.000636001 |
| PROSER2 | 0.00693913 | -0.00774052 | -0.00080139 |
| PLEKHB2 | 0.023571845 | -0.019725866 | 0.003845978 |
| SWAP70 | 0.012528148 | -0.017395553 | -0.004867405 |
| CHD2 | -0.020412461 | 0.02249355 | 0.002081089 |

| | | | |
|---|---|---|---|
| SRSF6 | -0.023866584 | 0.019934202 | -0.003932381 |
| BX470102.1 | 0.001079507 | -0.00094203 | 0.000137477 |
| NUPR2 | -0.020183184 | 0.023119122 | 0.002935938 |
| AL359636.2 | 0.000966549 | -0.000835161 | 0.000131389 |
| LTF | 0.001542289 | -0.001364976 | 0.000177312 |
| PCNX1 | -0.015351872 | 0.01217699 | -0.003174882 |
| TCAIM | -0.019022502 | 0.014865 | -0.004157503 |
| TRIM22 | -0.006776861 | 0.006069075 | -0.000707786 |
| PHAX | 0.01553615 | -0.018190329 | -0.002654179 |
| APOE | -0.018955892 | 0.030534847 | 0.011578955 |
| IL3RA | 0.001569544 | -0.001701991 | -0.000132447 |
| CLDN10-AS1 | -0.007403472 | 0.008781797 | 0.001378325 |
| RINL | 0.00329701 | -0.003601096 | -0.000304086 |
| FCHO2 | 0.014218248 | -0.012271936 | 0.001946311 |
| ATP8B2 | -0.00721067 | 0.006140891 | -0.001069778 |
| HSF2 | 0.008166585 | -0.009020797 | -0.000854212 |
| EIF4B | -0.027445006 | -0.028495551 | -0.055940557 |
| SHKBP1 | 0.009573019 | -0.007798892 | 0.001774127 |
| USP44 | 0.007141978 | -0.008995232 | -0.001853254 |
| SLC35A3 | 0.01600577 | -0.015230877 | 0.000774893 |
| SERPINB6 | 0.014964121 | -0.017360537 | -0.002396416 |
| KHK | -0.012622373 | 0.029960486 | 0.017338113 |
| TRO | -0.006074461 | 0.00867848 | 0.00260402 |
| RFTN1 | 0.002082466 | -0.002032414 | 5.00516E-05 |
| GRAMD1C | -0.017413338 | 0.014441238 | -0.0029721 |
| TXN2 | -0.025222571 | 0.024435144 | -0.000787428 |
| AC005696.1 | -0.004586765 | 0.003995394 | -0.00059137 |
| AL357033.1 | 0.001046436 | -0.000914501 | 0.000131936 |
| NUTM2A-AS1 | -0.016918858 | 0.01485169 | -0.002067168 |
| ERO1B | 0.009991861 | -0.009128094 | 0.000863767 |
| LINC002481 | 0.000965935 | -0.000829316 | 0.000136619 |
| CLTC | 0.02614919 | -0.023424647 | 0.002724543 |
| SLC6A9 | 0.003416449 | -0.002869745 | 0.000546704 |
| PFKM | -0.018006007 | 0.027977441 | 0.009971433 |
| MFSD9 | 0.005925564 | -0.008855154 | -0.00292959 |
| RPUSD1 | 0.008910547 | -0.007397257 | 0.00151329 |
| PHF13 | 0.008505759 | -0.009824408 | -0.001318649 |
| NFAT5 | 0.016353769 | -0.017146468 | -0.000792699 |
| GSE1 | 0.014862287 | -0.02557932 | -0.010717034 |
| UTY | 0.007007811 | -0.018484893 | -0.011477082 |
| FAM151B | 0.003039167 | -0.002464789 | 0.000574378 |
| SLC6A6 | 0.003209077 | -0.003259523 | -5.0446E-05 |
| CTSH | 0.020370256 | -0.035673198 | -0.015302942 |
| CBWD2 | -0.01076343 | 0.011655652 | 0.000892222 |
| IRAK2 | 0.001486143 | -0.001639526 | -0.000153383 |
| RILPL1 | -0.009778704 | 0.008131532 | -0.001647172 |
| GPR157 | -0.009041987 | 0.011790861 | 0.002748873 |

| | | | |
|---|---|---|---|
| SPR | 0.009208294 | -0.012694408 | -0.003486114 |
| MEDAG | 0.001819196 | -0.001832201 | -1.30051E-05 |
| ZNF281 | 0.01070847 | -0.011093125 | -0.000384655 |
| PPIF | -0.021720925 | 0.023980177 | 0.002259252 |
| LINC02471 | 0.001672989 | -0.001503581 | 0.000169408 |
| TTC9 | 0.002345251 | -0.002077551 | 0.0002677 |
| PGAP1 | 0.008723846 | -0.007554948 | 0.001168898 |
| PCAT6 | 0.003320128 | -0.002870172 | 0.000449956 |
| PKP3 | 0.001550928 | -0.00216019 | -0.000609262 |
| ABTB1 | 0.009759913 | -0.008125657 | 0.001634256 |
| HRC | 0.001956391 | -0.001820173 | 0.000136218 |
| PON3 | 0.001061139 | -0.001162881 | -0.000101741 |
| LINC02482 | -0.011561159 | 0.010738406 | -0.000822753 |
| AC007325.4 | 0.009344489 | -0.017869274 | -0.008524785 |
| RNPC3 | -0.021129451 | 0.024647629 | 0.003518178 |
| HRAS | -0.017760012 | 0.020605649 | 0.002845636 |
| FNDC3B | 0.016863742 | -0.023088862 | -0.006225119 |
| PALM3 | 0.009675566 | -0.012582022 | -0.002906456 |
| AC090709.1 | 0.002317574 | -0.002360548 | -4.29732E-05 |
| CPNE2 | 0.00312574 | -0.00294074 | 0.000184999 |
| NOL3 | -0.009787418 | 0.008897821 | -0.000889596 |
| AL021368.3 | -0.002891733 | 0.002655218 | -0.000236515 |
| AC073046.4 | 0.001908971 | -0.001583049 | 0.000325922 |
| GPR4 | 0.001557041 | -0.001867012 | -0.000309971 |
| FGF12 | 0.0015211 | -0.001365586 | 0.000155514 |
| CDO1 | 0.001098211 | -0.000957943 | 0.000140268 |
| RCN2 | 0.023142299 | -0.021073816 | 0.002068483 |
| GDF3 | 0.000882533 | -0.000825058 | 5.7475E-05 |
| MYT1 | 0.001037434 | -0.001245342 | -0.000207908 |
| ARAP1 | 0.012377513 | -0.010125016 | 0.002252497 |
| GRN | 0.021707235 | -0.03256714 | -0.010859905 |
| AC021016.3 | 0.003382347 | -0.003380328 | 2.0195E-06 |
| ARPC1A | 0.019409471 | -0.029774209 | -0.010364738 |
| ARHGEF37 | 0.007192707 | -0.007627532 | -0.000434824 |
| ITGB1BP2 | 0.001047826 | -0.000847822 | 0.000200004 |
| PRR15L | -0.018607651 | 0.028298314 | 0.009690662 |
| FUT11 | 0.005870656 | -0.004842122 | 0.001028534 |
| ZNF366 | 0.001517139 | -0.001488778 | 2.83604E-05 |
| SMC4 | 0.008263685 | -0.007767766 | 0.000495919 |
| LINC00629 | -0.005820166 | 0.005635666 | -0.0001845 |
| EFS | -0.006217943 | 0.005403583 | -0.00081436 |
| CUL9 | -0.008554469 | 0.00712209 | -0.001432379 |
| FZD9 | 0.001117516 | -0.001156945 | -3.94295E-05 |
| DUSP3 | 0.01604714 | -0.020467991 | -0.004420851 |
| GALNT6 | 0.009105293 | -0.007712961 | 0.001392332 |
| OSTF1 | 0.011234656 | -0.013285422 | -0.002050767 |
| MTMR7 | -0.005899123 | 0.005977178 | 7.80556E-05 |

| | | | |
|---|---|---|---|
| SEL1L3 | 0.003408262 | -0.00341145 | -3.18784E-06 |
| S100A13 | 0.025290097 | -0.056669707 | -0.03137961 |
| ANKRD13D | 0.009300289 | -0.007931862 | 0.001368427 |
| ITPR3 | 0.007065869 | -0.008866597 | -0.001800728 |
| TMEM258 | -0.027883202 | 0.04079239 | 0.012909188 |
| FRZB | 0.00241317 | -0.002362148 | 5.10215E-05 |
| SELP | 0.00094929 | -0.00080763 | 0.00014166 |
| PTPA | -0.019749694 | 0.018977818 | -0.000771875 |
| REPS2 | -0.004989061 | 0.004074615 | -0.000914446 |
| ZBED6CL | 0.003894488 | -0.004411886 | -0.000517398 |
| FRGCA | 0.000754125 | -0.000754125 | -3.55271E-15 |
| PPFIBP1 | 0.013179898 | -0.018262315 | -0.005082417 |
| HDLBP | 0.02464467 | -0.022273419 | 0.002371251 |
| ARPP21 | 0.00107699 | -0.001179897 | -0.000102907 |
| GIGYF2 | 0.018408597 | -0.020610129 | -0.002201532 |
| EMCN | 0.003478246 | -0.003147057 | 0.000331189 |
| EFNB2 | 0.002809765 | -0.002807531 | 2.23423E-06 |
| DSC2 | 0.010065751 | -0.008961243 | 0.001104507 |
| NNMT | 0.001811375 | -0.001702186 | 0.000109188 |
| TFAP2C | 0.001005044 | -0.000847783 | 0.000157261 |
| VOPP1 | 0.012944015 | -0.015223429 | -0.002279415 |
| AC005165.1 | 0.001178026 | -0.001325419 | -0.000147392 |
| LINC02352 | 0.001409563 | -0.001186851 | 0.000222712 |
| GADL1 | -0.00589322 | 0.011232651 | 0.005339431 |
| TIPARP | 0.008714358 | -0.010978209 | -0.00226385 |
| MRPS28 | -0.019483938 | 0.016333007 | -0.003150931 |
| CPVL | -0.025227785 | 0.028810884 | 0.003583099 |
| ARMC10 | 0.014330689 | -0.011931748 | 0.002398941 |
| KIZ | 0.010617908 | -0.011418195 | -0.000800288 |
| ENTPD1 | 0.002730126 | -0.002559316 | 0.00017081 |
| CELF6 | -0.014494466 | 0.013675586 | -0.00081888 |
| NXPE2 | 0.000914616 | -0.000799907 | 0.000114709 |
| QKI | 0.021254213 | -0.036109087 | -0.014854874 |
| STX3 | 0.011526588 | -0.010533319 | 0.000993269 |
| CLINT1 | 0.023321786 | -0.038312898 | -0.014991112 |
| SEPHS2 | -0.02336868 | -0.023797542 | -0.047166222 |
| OPLAH | -0.01528073 | 0.029459347 | 0.014178617 |
| ST6GALNAC2 | 0.00161947 | -0.001826133 | -0.000206663 |
| NLGN4Y | 0.003939016 | -0.00665102 | -0.002712004 |

**Principal cells**

| Genes | logFC_T2D_HC | logFC_T2Di_T2D | logFC_T2Di_HC |
|---|---|---|---|
| AKR1B1 | 0.529409115 | -0.424948487 | 0.104460629 |
| AKR1C1 | 0.277506164 | -0.280841783 | -0.003335619 |
| SLC5A3 | 0.423076321 | -0.430841928 | -0.007765607 |
| RANBP3L | 0.226831129 | -0.228491923 | -0.001660794 |
| ARG2 | 0.347063644 | -0.252903852 | 0.094159792 |
| NUPR1 | 0.280254947 | -0.285405126 | -0.005150178 |
| TFPI2 | 0.254691948 | -0.260035567 | -0.005343619 |
| PFKP | 0.29385365 | -0.274618083 | 0.019235567 |
| PTGS1 | 0.263021028 | -0.268036757 | -0.005015729 |
| SLC14A2 | 0.19123536 | -0.191285018 | -4.96578E-05 |
| MMP7 | 0.298637883 | -0.134548112 | 0.164089771 |
| MRPS6 | 0.357449898 | -0.368475558 | -0.011025661 |
| SLCO4A1 | 0.169209751 | -0.169740261 | -0.000530511 |
| FHL2 | 0.185697054 | -0.195029093 | -0.009332039 |
| CRYAB | 0.294075048 | -0.241544175 | 0.052530872 |
| RNF24 | 0.245801446 | -0.218805803 | 0.026995643 |
| ADIRF | -0.203933348 | 0.048433202 | -0.155500146 |
| RFC1 | 0.255946286 | -0.245376809 | 0.010569477 |
| ARPC1B | 0.267735629 | -0.200101091 | 0.067634538 |
| SLC6A12 | 0.240451176 | -0.266208484 | -0.025757307 |
| ANK2 | 0.175843473 | -0.190056632 | -0.014213159 |
| ST6GAL1 | 0.317737963 | -0.090206902 | 0.227531061 |
| SLC12A1 | -0.237445523 | 0.028723792 | -0.208721731 |
| NTS | 0.186753461 | -0.161694136 | 0.025059324 |
| PAPPA | 0.292759041 | -0.039201524 | 0.253557517 |
| AOC1 | 0.304904153 | -0.28372366 | 0.021180493 |
| SLC4A11 | 0.126778737 | -0.127362331 | -0.000583593 |
| PTGR1 | 0.298492988 | -0.13285497 | 0.165638018 |
| SLC4A7 | 0.157744309 | -0.154353846 | 0.003390462 |
| ID3 | -0.272281393 | 0.137461016 | -0.134820378 |
| HSPA2 | 0.260251591 | -0.225936777 | 0.034314813 |
| SLCO3A1 | 0.166591959 | -0.175886768 | -0.009294809 |
| ALDH8A1 | 0.116736849 | -0.117528451 | -0.000791602 |
| PGGHG | 0.20427304 | -0.252164265 | -0.047891225 |
| STXBP6 | 0.149020966 | -0.124995766 | 0.024025201 |
| PLXNA4 | 0.099141199 | -0.099609878 | -0.000468679 |
| AQP3 | 0.266001699 | -0.067305143 | 0.198696556 |
| TIMP3 | 0.204751933 | -0.110019032 | 0.094732901 |
| TPD52L1 | 0.197213427 | -0.164673274 | 0.032540154 |
| KNG1 | -0.182183071 | 0.040050672 | -0.142132399 |
| AC023421.1 | 0.096398098 | -0.095908726 | 0.000489372 |
| SASH1 | 0.112416312 | -0.112241283 | 0.000175028 |
| HIF1A | 0.222445493 | -0.133235533 | 0.089209961 |
| EGF | -0.121460242 | 0.018131294 | -0.103328948 |
| HLA-B | -0.24192327 | 0.148541143 | -0.093382127 |

| | | | |
|---|---|---|---|
| LDHA | 0.256378556 | -0.148742048 | 0.107636507 |
| SYNE2 | 0.259809141 | -0.243612363 | 0.016196778 |
| BAG1 | 0.242507175 | -0.178968196 | 0.063538979 |
| SSTR2 | 0.092960848 | -0.093079162 | -0.000118314 |
| CD82 | 0.210944286 | -0.197681286 | 0.013263 |
| NAPEPLD | 0.158137726 | -0.142823263 | 0.015314463 |
| DDIT4L | 0.095380537 | -0.093960898 | 0.001419638 |
| PRDX4 | 0.19604588 | -0.108104121 | 0.087941759 |
| ISYNA1 | 0.163285042 | -0.161344451 | 0.001940591 |
| TLN2 | 0.175020887 | -0.167160158 | 0.007860729 |
| GRAMD2B | 0.148953597 | -0.151026725 | -0.002073128 |
| B4GALNT2 | 0.158833734 | -0.142735201 | 0.016098533 |
| S100A10 | 0.252142724 | -0.107972578 | 0.144170146 |
| TMEM176B | 0.243559792 | -0.194741581 | 0.048818211 |
| PHACTR1 | 0.154589056 | -0.132316293 | 0.022272763 |
| GPER1 | 0.090074151 | -0.087001734 | 0.003072417 |
| DDIT4 | 0.161106897 | -0.111310427 | 0.04979647 |
| NTN4 | 0.223131551 | -0.230539759 | -0.007408208 |
| NFKBIA | 0.162450668 | -0.1365402 | 0.025910467 |
| FUCA1 | 0.208863376 | -0.130670337 | 0.078193039 |
| TGFBR1 | 0.144988675 | -0.094671163 | 0.050317512 |
| SOX4 | -0.183903457 | 0.112846122 | -0.071057335 |
| TMED5 | 0.212114002 | -0.125403918 | 0.086710084 |
| LOXL2 | 0.074081218 | -0.072830729 | 0.001250489 |
| HS3ST1 | 0.101031883 | -0.099583692 | 0.001448191 |
| ITM2B | 0.199540006 | -0.064970247 | 0.134569759 |
| TACSTD2 | 0.243057029 | -0.166763384 | 0.076293644 |
| FRY | 0.159142421 | -0.146097189 | 0.013045231 |
| HIP1 | 0.127910266 | -0.137703246 | -0.00979298 |
| EPHX1 | 0.132149264 | -0.0878653 | 0.044283963 |
| TMEM165 | 0.164487985 | -0.113916069 | 0.050571916 |
| RASSF8 | 0.125421057 | -0.114972631 | 0.010448425 |
| SAMD11 | -0.16119458 | 0.080262913 | -0.080931667 |
| GREB1L | 0.07772371 | -0.079614911 | -0.001891201 |
| SLC45A4 | 0.119764315 | -0.081849483 | 0.037914832 |
| FAM241A | 0.147700292 | -0.136529084 | 0.011171208 |
| NDRG1 | -0.211116974 | 0.280391518 | 0.069274544 |
| CCPG1 | 0.199116586 | -0.10727027 | 0.091846316 |
| TXNIP | -0.20297969 | 0.344124402 | 0.141144713 |
| OTULINL | 0.138677596 | -0.127329599 | 0.011347997 |
| SCCPDH | 0.188079957 | -0.146022509 | 0.042057448 |
| MGST1 | -0.15145716 | 0.155740922 | 0.004283762 |
| APOE | -0.09711739 | 0.025002333 | -0.072115057 |
| SLC44A4 | 0.109231799 | -0.103829687 | 0.005402112 |
| EHF | 0.14861759 | -0.100980212 | 0.047637378 |
| SMIM24 | -0.139095032 | 0.076439279 | -0.062655753 |
| PRRG4 | 0.128922303 | -0.113232393 | 0.01568991 |

| | | | |
|---|---|---|---|
| ANXA2 | 0.21421521 | -0.121175675 | 0.093039535 |
| UNC13C | 0.084378877 | -0.066191111 | 0.018187766 |
| HLA-DMB | 0.08135005 | -0.075315585 | 0.006034465 |
| ELF5 | 0.169834127 | -0.180750083 | -0.010915956 |
| GPT2 | 0.077757306 | -0.07279931 | 0.004957996 |
| SLC25A29 | 0.183849693 | -0.044064498 | 0.139785194 |
| SLC16A4 | 0.164375096 | -0.06787408 | 0.096501016 |
| PALM2-AKAP2 | -0.14377378 | 0.110432741 | -0.03334104 |
| ATP1A1 | 0.188832485 | -0.085742619 | 0.103089866 |
| TINAGL1 | 0.16343602 | -0.126301264 | 0.037134757 |
| FGF2 | 0.093991172 | -0.086997777 | 0.006993395 |
| MFSD10 | 0.142937103 | -0.092043606 | 0.050893496 |
| THSD7A | 0.188462971 | -0.090516023 | 0.097946948 |
| GAL3ST1 | 0.117803874 | -0.079286568 | 0.038517307 |
| ANGPTL4 | 0.056960638 | -0.052402841 | 0.004557797 |
| TMEM176A | 0.195028749 | -0.184350073 | 0.010678677 |
| TMEM50A | 0.167816891 | -0.070401215 | 0.097415676 |
| LPIN2 | 0.153327347 | -0.164129398 | -0.010802051 |
| CTSH | 0.193843214 | -0.114414309 | 0.079428904 |
| HS6ST1 | 0.170295673 | -0.087578924 | 0.082716748 |
| DST | 0.115570464 | -0.13998395 | -0.024413486 |
| KCNJ16 | 0.180800578 | -0.097786547 | 0.083014031 |
| AC009275.1 | 0.047200881 | -0.047573872 | -0.000372991 |
| RDH10 | 0.170181902 | -0.097126614 | 0.073055288 |
| ETF1 | 0.128905234 | -0.100696512 | 0.028208722 |
| ELF3 | 0.140605905 | -0.130466915 | 0.010138989 |
| KCNK3 | 0.051666937 | -0.051055874 | 0.000611064 |
| UCHL1 | -0.154801172 | 0.184874254 | 0.030073083 |
| MAP3K21 | -0.135872035 | 0.095504883 | -0.040367153 |
| SVIL | 0.093887127 | -0.106567464 | -0.012680337 |
| MAN1A1 | 0.178911129 | -0.144055563 | 0.034855565 |
| ORAI2 | 0.133078371 | -0.12082971 | 0.012248661 |
| VPS13C | 0.157582989 | -0.092457194 | 0.065125795 |
| MAL2 | 0.173414049 | -0.05135404 | 0.122060009 |
| PPIC | 0.144256069 | -0.027927583 | 0.116328486 |
| CLDN4 | 0.167169791 | -0.104757786 | 0.062412005 |
| LMO7 | 0.140378274 | -0.121489024 | 0.01888925 |
| CD24 | 0.154330667 | -0.126350197 | 0.027980469 |
| TSPAN1 | 0.189964933 | -0.107964256 | 0.082000677 |
| DDX3Y | 0.090568724 | -0.129300595 | -0.038731871 |
| PDE4DIP | 0.11082997 | -0.080112388 | 0.030717583 |
| FOS | -0.183310787 | 0.164090051 | -0.019220735 |
| DUSP9 | -0.067975921 | 0.017842471 | -0.05013345 |
| GCNT3 | 0.048528836 | -0.044634225 | 0.00389461 |
| FXYD4 | 0.240663704 | -0.176228397 | 0.064435307 |
| SLC44A1 | 0.142828424 | -0.079571805 | 0.063256618 |
| LIMA1 | 0.134306862 | -0.125391957 | 0.008914905 |

| | | | |
|---|---|---|---|
| AL162424.1 | 0.046883883 | -0.04700386 | -0.000119978 |
| SERTAD4-AS1 | 0.101450554 | -0.107095298 | -0.005644744 |
| TMEM68 | 0.10837413 | -0.08782255 | 0.02055158 |
| TMEM158 | -0.085260195 | 0.065896097 | -0.019364098 |
| S100A11 | 0.178389484 | -0.058463324 | 0.11992616 |
| LCK | 0.046837281 | -0.047841132 | -0.001003851 |
| EIF1AY | 0.083785666 | -0.117538341 | -0.033752675 |
| TSPO | 0.164652269 | -0.147888946 | 0.016763323 |
| PRDX1 | 0.176901454 | -0.0383785 | 0.138522953 |
| SHB | 0.106318133 | -0.066498572 | 0.039819561 |
| JPT1 | 0.138148475 | -0.091468815 | 0.04667966 |
| CNNM1 | 0.038881665 | -0.038881665 | 5.32907E-15 |
| ACKR4 | 0.054450949 | -0.050291066 | 0.004159883 |
| AC087379.2 | -0.103319735 | 0.108735204 | 0.005415469 |
| PAX8 | 0.168368972 | -0.096281939 | 0.072087034 |
| TMEM131 | 0.116778269 | -0.08898055 | 0.027797719 |
| GSPT1 | 0.1294804 | -0.10205247 | 0.027427929 |
| CD164 | 0.154211364 | -0.060713939 | 0.093497426 |
| ASPHD2 | 0.077053046 | -0.059991554 | 0.017061492 |
| TACC1 | 0.099871781 | -0.131722807 | -0.031851026 |
| PIM3 | 0.101467453 | -0.057441402 | 0.044026051 |
| AGFG2 | 0.079652671 | -0.084400695 | -0.004748024 |
| SLPI | 0.093043046 | -0.122009233 | -0.028966186 |
| TM7SF3 | 0.123782617 | -0.089481478 | 0.034301139 |
| BCL6 | 0.062095448 | -0.061457368 | 0.00063808 |
| CALM1 | 0.163418989 | -0.081994311 | 0.081424678 |
| PTBP3 | 0.14573606 | -0.057529612 | 0.088206448 |
| CLDN10 | -0.091211256 | 0.021895952 | -0.069315305 |
| OPHN1 | 0.088908031 | -0.065923972 | 0.022984059 |
| B3GNT3 | 0.04484175 | -0.05096058 | -0.00611883 |
| SLC66A3 | 0.140641574 | -0.069714822 | 0.070926751 |
| LGALS3 | -0.135622245 | 0.060133153 | -0.075489092 |
| UCP2 | 0.133465136 | -0.08103274 | 0.052432395 |
| PAK2 | 0.136745131 | -0.117600589 | 0.019144542 |
| CD151 | 0.150494078 | -0.098179509 | 0.05231457 |
| P4HA1 | 0.086542711 | -0.08135928 | 0.005183431 |
| PLPP4 | -0.068689144 | 0.024076446 | -0.044612698 |
| GOLGB1 | 0.148504566 | -0.073745974 | 0.074758592 |
| KCNMB4 | 0.050908225 | -0.048210779 | 0.002697446 |
| GRB14 | 0.068249751 | -0.06822825 | 2.15011E-05 |
| ZBTB38 | 0.116030596 | -0.093186169 | 0.022844427 |
| CHST6 | 0.038742216 | -0.038507638 | 0.000234578 |
| SLC17A5 | 0.082789166 | -0.077942999 | 0.004846167 |
| DEFB1 | -0.211424097 | 0.282778351 | 0.071354254 |
| AC136475.3 | 0.042257913 | -0.045827035 | -0.003569121 |
| JUN | -0.136182148 | 0.079472692 | -0.056709456 |
| PLAU | -0.04849656 | 0.009983933 | -0.038512626 |

| | | | |
|---|---|---|---|
| HIST1H2AC | 0.10768939 | -0.10825889 | -0.0005695 |
| APCDD1L | -0.099204953 | 0.062215164 | -0.036989788 |
| RTN3 | 0.157285569 | -0.092537541 | 0.064748028 |
| MPP7 | 0.088177814 | -0.084014961 | 0.004162854 |
| ZFYVE16 | 0.136379948 | -0.041254173 | 0.095125775 |
| BOC | 0.035570077 | -0.034573454 | 0.000996623 |
| CDK18 | 0.083092501 | -0.071790195 | 0.011302306 |
| DDRGK1 | 0.12288427 | -0.059542652 | 0.063341618 |
| LRBA | 0.11964276 | -0.068616122 | 0.051026638 |
| SERINC2 | 0.145572396 | -0.058929745 | 0.086642652 |
| KDELR1 | 0.143579477 | -0.077822855 | 0.065756622 |
| ID4 | -0.140126558 | 0.079255882 | -0.060870676 |
| COL4A4 | 0.060305317 | -0.050599356 | 0.009705962 |
| ACTN1 | 0.071154159 | -0.069567399 | 0.00158676 |
| TPM1 | 0.137084381 | -0.091959826 | 0.045124555 |
| RAB15 | 0.080064544 | -0.056994527 | 0.023070016 |
| JUNB | -0.147280458 | 0.091429556 | -0.055850903 |
| SEMA4D | 0.105019579 | -0.096849904 | 0.008169675 |
| ITGB5 | 0.097837133 | -0.083064329 | 0.014772804 |
| ITPR2 | 0.11947138 | -0.082123251 | 0.037348129 |
| S100A14 | -0.087291404 | 0.051744526 | -0.035546878 |
| S100A2 | -0.149841498 | 0.133579159 | -0.016262339 |
| CLN3 | 0.113229145 | -0.088414293 | 0.024814852 |
| CYSLTR2 | 0.067007113 | -0.031352951 | 0.035654162 |
| TP53I11 | 0.083620452 | -0.087529356 | -0.003908904 |
| HLA-DRA | 0.117222507 | -0.068544086 | 0.048678421 |
| QPCT | 0.05917907 | -0.048304054 | 0.010875015 |
| AIG1 | 0.122543419 | -0.043166389 | 0.079377029 |
| DCDC2 | -0.133209403 | 0.158788902 | 0.025579498 |
| LHFPL3-AS2 | 0.087266331 | -0.043384692 | 0.043881639 |
| NREP | 0.074768067 | -0.037408175 | 0.037359891 |
| ALDH1A3 | 0.037306303 | -0.037221462 | 8.48408E-05 |
| NEURL3 | 0.084608957 | -0.07173304 | 0.012875917 |
| NPAS2 | 0.086746924 | -0.072702486 | 0.014044438 |
| RCAN1 | 0.114244195 | -0.096936979 | 0.017307217 |
| CKB | -0.138572193 | 0.110950323 | -0.02762187 |
| HPN | -0.082065157 | 0.065696308 | -0.016368848 |
| GCNT4 | -0.088440127 | 0.063139438 | -0.025300689 |
| SCARA3 | 0.056433848 | -0.06012445 | -0.003690602 |
| TSC22D3 | 0.093648078 | -0.073405711 | 0.020242367 |
| HLA-DMA | 0.093429504 | -0.049894454 | 0.04353505 |
| FBP1 | 0.123305165 | -0.063713185 | 0.05959198 |
| ZNF503 | -0.123237083 | 0.15033455 | 0.027097467 |
| LTBP4 | -0.065112615 | 0.035286759 | -0.029825856 |
| GALNT3 | 0.112698913 | -0.101544437 | 0.011154476 |
| ANKS1B | 0.038101247 | -0.036421064 | 0.001680183 |
| ARFGEF1 | 0.117882677 | -0.091632019 | 0.026250658 |

| | | | |
|---|---|---|---|
| APCDD1L-DT | -0.103640628 | 0.093657121 | -0.009983506 |
| AC005165.1 | 0.032129693 | -0.031536421 | 0.000593272 |
| AC021517.1 | 0.034572292 | -0.034572292 | 1.06581E-14 |
| MBOAT2 | 0.110552435 | -0.075171403 | 0.035381031 |
| DNAJC3 | 0.111620825 | -0.056459518 | 0.055161307 |
| PRR15 | 0.074433102 | -0.03306312 | 0.041369982 |
| FMNL2 | 0.069928594 | -0.064077596 | 0.005850997 |
| ITGAV | 0.121300806 | -0.074367109 | 0.046933698 |
| SORT1 | 0.106927054 | -0.062799847 | 0.044127206 |
| FXYD2 | -0.205203794 | 0.151107387 | -0.054096407 |
| MTMR10 | -0.110074205 | 0.080651068 | -0.029423137 |
| UBAC2 | 0.109971217 | -0.030866691 | 0.079104526 |
| ETS2 | 0.049631122 | -0.049403273 | 0.000227849 |
| CDADC1 | 0.059167674 | -0.057181393 | 0.001986282 |
| EPS8L1 | 0.093344549 | -0.078594033 | 0.014750516 |
| OLFML3 | 0.034532799 | -0.035562773 | -0.001029974 |
| NIPAL3 | 0.096256463 | -0.086782536 | 0.009473927 |
| FDFT1 | 0.096226779 | -0.043872265 | 0.052354514 |
| LOXL4 | 0.041165506 | -0.040406116 | 0.00075939 |
| PRXL2A | 0.123820257 | -0.134810006 | -0.010989749 |
| VIM | 0.070800436 | -0.083237248 | -0.012436812 |
| TSC22D4 | 0.092510819 | -0.097520576 | -0.005009757 |
| TGFBR2 | 0.083063014 | -0.071314924 | 0.01174809 |
| UTRN | 0.111591118 | -0.094678374 | 0.016912745 |
| IFITM3 | 0.130210027 | -0.074719006 | 0.055491021 |
| TRPM3 | -0.125431689 | 0.177455691 | 0.052024002 |
| SEC62 | 0.138047516 | -0.082284194 | 0.055763323 |
| ABCA3 | 0.06532598 | -0.063387494 | 0.001938486 |
| LINC01320 | -0.067758367 | 0.030282287 | -0.03747608 |
| ADGRF1 | 0.114765724 | -0.063644276 | 0.051121449 |
| BTG1 | 0.131991843 | -0.130700918 | 0.001290925 |
| PLOD2 | 0.090219256 | -0.045178206 | 0.04504105 |
| SAT1 | -0.139052561 | 0.22561292 | 0.08656036 |
| GPR146 | 0.051142503 | -0.03829718 | 0.012845322 |
| CYBRD1 | -0.096321998 | 0.120296804 | 0.023974806 |
| PIK3IP1 | 0.097474281 | -0.051175028 | 0.046299253 |
| MGAT4B | 0.094963562 | -0.049479415 | 0.045484148 |
| GCHFR | 0.05553737 | -0.060212217 | -0.004674847 |
| PIGH | 0.081300208 | -0.065708857 | 0.015591351 |
| MTUS1 | 0.098322675 | -0.057977262 | 0.040345413 |
| KCNJ15 | -0.130422506 | 0.246346178 | 0.115923672 |
| TMEM192 | 0.085825704 | -0.08976866 | -0.003942956 |
| LYAR | 0.046637879 | -0.041495909 | 0.005141971 |
| S100A1 | 0.103838242 | -0.054514029 | 0.049324213 |
| PISD | 0.06827923 | -0.065276819 | 0.003002411 |
| TM9SF3 | 0.122707213 | -0.033706304 | 0.08900091 |
| INSIG2 | 0.08509712 | -0.083218901 | 0.00187822 |

| | | | |
|---|---|---|---|
| MT1G | -0.065240814 | 0.040409204 | -0.024831609 |
| WNT9B | -0.07550424 | 0.043815305 | -0.031688935 |
| SPP1 | -0.057077698 | 0.017649573 | -0.039428125 |
| LINC02381 | -0.071092365 | 0.033202843 | -0.037889522 |
| KCNK5 | 0.052155033 | -0.054376587 | -0.002221553 |
| KLF13 | 0.090249413 | -0.052750052 | 0.037499361 |
| CCDC107 | 0.08851642 | -0.047937334 | 0.040579086 |
| MARCKS | -0.092039007 | 0.104957808 | 0.012918801 |
| FAM20A | 0.087671433 | -0.082695985 | 0.004975448 |
| ID1 | -0.149715498 | 0.105292247 | -0.044423251 |
| ATP6V0A4 | 0.101721686 | -0.079521805 | 0.022199881 |
| HTATIP2 | 0.071710158 | -0.058023954 | 0.013686204 |
| KRTCAP3 | 0.064788656 | -0.074088645 | -0.009299989 |
| RAB13 | 0.117743188 | -0.028272755 | 0.089470433 |
| SLC52A3 | 0.091057752 | -0.080478273 | 0.01057948 |
| MACO1 | 0.071256203 | -0.059112371 | 0.012143832 |
| CAMK2N1 | 0.130932428 | -0.049316108 | 0.081616321 |
| ARL6IP5 | 0.118212776 | -0.08581481 | 0.032397966 |
| KIF26A | -0.038621481 | 0.020146951 | -0.018474531 |
| CTSC | -0.104076931 | 0.135052253 | 0.030975322 |
| NMB | 0.069469005 | -0.054788228 | 0.014680776 |
| RORA | 0.064592101 | -0.056417088 | 0.008175012 |
| FGF9 | -0.063409354 | 0.045960652 | -0.017448701 |
| HPS4 | 0.072587911 | -0.066784371 | 0.00580354 |
| BCAP29 | 0.082549097 | -0.056682071 | 0.025867026 |
| EGR1 | -0.070322549 | 0.052071812 | -0.018250737 |
| REEP3 | 0.097895392 | -0.064984543 | 0.032910848 |
| TIMP2 | -0.082796036 | 0.076466951 | -0.006329085 |
| PABPC4 | 0.08506589 | -0.084951769 | 0.000114121 |
| SDC4 | -0.11425299 | 0.138450878 | 0.024197888 |
| ZNF107 | 0.05113494 | -0.033589099 | 0.017545841 |
| ADAMTS9-AS1 | 0.03811008 | -0.044221659 | -0.006111579 |
| SLC38A2 | 0.117645378 | -0.032134843 | 0.085510535 |
| TBC1D7 | 0.06146348 | -0.041774942 | 0.019688538 |
| GOLGA8A | 0.083205532 | -0.022997577 | 0.060207955 |
| DNAJC1 | 0.089077517 | -0.049054512 | 0.040023004 |
| DRAM1 | 0.060110341 | -0.061694324 | -0.001583983 |
| ITGB1 | 0.124652698 | -0.067098402 | 0.057554296 |
| FGFR3 | -0.056558415 | 0.021026704 | -0.035531711 |
| HPGD | -0.092127969 | 0.097812209 | 0.00568424 |
| NID2 | 0.030553005 | -0.034025279 | -0.003472274 |
| H1FX | -0.091175058 | 0.062515579 | -0.028659478 |
| F2RL1 | 0.086342532 | -0.039162858 | 0.047179674 |
| DPYSL2 | 0.051976922 | -0.050647153 | 0.001329769 |
| CA12 | -0.133879452 | 0.258397116 | 0.124517663 |
| NEAT1 | 0.144136739 | -0.04915422 | 0.094982519 |
| MLLT1 | -0.095366138 | 0.099797391 | 0.004431253 |

| | | | |
|---|---|---|---|
| CPE | 0.032318473 | -0.029473247 | 0.002845226 |
| PPP1R10 | 0.084908008 | -0.056140724 | 0.028767284 |
| DOCK8 | 0.031783842 | -0.032665518 | -0.000881677 |
| FAM167B | 0.036117354 | -0.036234824 | -0.000117471 |
| H3F3B | -0.13078804 | 0.194432998 | 0.063644958 |
| SLC2A1 | 0.087046947 | -0.069181101 | 0.017865846 |
| CREG1 | 0.100209831 | -0.046499851 | 0.05370998 |
| LMO4 | -0.071665082 | 0.037598131 | -0.034066951 |
| BX284668.5 | 0.062538623 | -0.038863729 | 0.023674895 |
| PCSK1N | -0.13585367 | 0.218124135 | 0.082270465 |
| GRB7 | 0.076294199 | -0.061322654 | 0.014971545 |
| LAMB2 | 0.09387834 | -0.065756268 | 0.028122071 |
| SGPL1 | 0.08100397 | -0.04358835 | 0.03741562 |
| ALDOB | -0.047522461 | 0.016709605 | -0.030812856 |
| CAB39L | 0.06503942 | -0.057417129 | 0.007622291 |
| PANK2 | 0.066235442 | -0.028591041 | 0.037644401 |
| MELTF | 0.03324256 | -0.033917383 | -0.000674823 |
| BNIPL | -0.04307943 | 0.021117255 | -0.021962175 |
| ZNRF1 | 0.058692452 | -0.048209174 | 0.010483278 |
| CLIP2 | 0.04870574 | -0.05047197 | -0.001766231 |
| GPRC5B | -0.105630502 | 0.115995599 | 0.010365097 |
| MBP | 0.105457373 | -0.089904635 | 0.015552738 |
| LMO3 | 0.0655098 | -0.07641587 | -0.01090607 |
| FGD3 | -0.067279396 | 0.047669 | -0.019610397 |
| NR1H4 | 0.055853731 | -0.052077185 | 0.003776546 |
| C11orf52 | 0.071691844 | -0.047871131 | 0.023820713 |
| IER2 | -0.120795724 | 0.093339734 | -0.02745599 |
| ATP1B1 | 0.10220066 | -0.028986587 | 0.073214073 |
| FBXO25 | 0.06831633 | -0.054663047 | 0.013653283 |
| CLU | 0.127984259 | -0.065726136 | 0.062258124 |
| EMC6 | 0.092968329 | -0.054075025 | 0.038893304 |
| FABP3 | -0.040860532 | 0.01755267 | -0.023307862 |
| TFAP2C | 0.032986634 | -0.032083356 | 0.000903279 |
| SERTAD4 | 0.038425341 | -0.039502294 | -0.001076952 |
| ZFHX4 | 0.024746908 | -0.021792623 | 0.002954285 |
| PXYLP1 | 0.04274517 | -0.041730974 | 0.001014196 |
| CAT | 0.103414853 | -0.043135596 | 0.060279258 |
| CHD9 | 0.105002452 | -0.048772667 | 0.056229785 |
| MALT1 | 0.049915573 | -0.038240825 | 0.011674748 |
| PIGR | -0.118402004 | 0.218302781 | 0.099900777 |
| CXXC5 | 0.104443839 | -0.041590238 | 0.062853601 |
| OPTN | 0.062111509 | -0.049952326 | 0.012159182 |
| HACD3 | 0.114870508 | -0.041773994 | 0.073096513 |
| AL109976.1 | 0.020295415 | -0.020211297 | 8.41179E-05 |
| SLC3A1 | -0.068578447 | 0.040797315 | -0.027781132 |
| MYDGF | 0.091841549 | -0.046254132 | 0.045587417 |
| ID2 | -0.058830285 | 0.052613417 | -0.006216868 |

| | | | |
|---|---|---|---|
| CMYA5 | -0.085970959 | 0.118684391 | 0.032713433 |
| PHLDA2 | 0.063822454 | -0.054923621 | 0.008898834 |
| PACSIN2 | 0.080330554 | -0.058302802 | 0.022027752 |
| PIK3R3 | 0.05371746 | -0.043172365 | 0.010545095 |
| LRFN4 | 0.045920396 | -0.042172694 | 0.003747702 |
| CRIP1 | -0.103152854 | 0.033970412 | -0.069182443 |
| PLPP5 | 0.097177116 | -0.037841646 | 0.05933547 |
| FOSB | -0.050531893 | 0.019153854 | -0.031378039 |
| FGFR1 | 0.066127786 | -0.059734966 | 0.00639282 |
| GPR160 | 0.091487858 | -0.032902628 | 0.05858523 |
| SNAPC1 | 0.062698337 | -0.064121053 | -0.001422716 |
| GATM | -0.061045113 | 0.039207999 | -0.021837114 |
| HTATSF1 | 0.086754898 | -0.086147607 | 0.00060729 |
| DHRS7 | 0.113219889 | -0.126865874 | -0.013645984 |
| AMFR | 0.102508029 | -0.110354058 | -0.007846029 |
| GMNN | 0.092458721 | -0.038293879 | 0.054164842 |
| UNC50 | 0.089992843 | -0.030568007 | 0.059424836 |
| LGALS1 | -0.069312035 | 0.08458507 | 0.015273035 |
| PFKFB3 | 0.115249323 | -0.11362916 | 0.001620162 |
| TNFSF10 | 0.057603223 | -0.067029132 | -0.009425909 |
| XG | 0.020552256 | -0.01823853 | 0.002313726 |
| POR | 0.07843042 | -0.05176548 | 0.026664941 |
| ANXA3 | -0.071403136 | 0.070361836 | -0.0010413 |
| ECH1 | 0.10187234 | 0.030121276 | 0.131993616 |
| FGF12 | -0.042221875 | 0.026616257 | -0.015605617 |
| USP53 | 0.078316337 | -0.072070771 | 0.006245567 |
| MGLL | -0.07368102 | 0.096635171 | 0.022954151 |
| SUSD1 | 0.071142029 | -0.025453637 | 0.045688392 |
| SEC11C | 0.096771051 | -0.044231676 | 0.052539375 |
| RABAC1 | 0.109820769 | -0.043968988 | 0.065851781 |
| MYORG | 0.053406215 | -0.029628417 | 0.023777798 |
| CARHSP1 | 0.079500024 | -0.059945895 | 0.019554128 |
| ITLN1 | -0.10357491 | 0.242379613 | 0.138804702 |
| BMP4 | -0.040359913 | 0.025721421 | -0.014638492 |
| EDNRB | 0.082389914 | -0.03536117 | 0.047028744 |
| ATP11C | 0.055457692 | -0.062596119 | -0.007138428 |
| HK1 | 0.073166183 | -0.0636023 | 0.009563883 |
| SELENOS | 0.093996042 | -0.050577685 | 0.043418357 |
| PTGER3 | -0.084516782 | 0.107627101 | 0.02311032 |
| PRPS2 | -0.088012401 | 0.161312481 | 0.07330008 |
| CLCNKB | -0.094923619 | 0.182456441 | 0.087532823 |
| MARVELD3 | 0.047689193 | -0.04770526 | -1.60676E-05 |
| GCC2 | 0.099061405 | -0.041312427 | 0.057748978 |
| TMEM238L | 0.031886635 | -0.031902567 | -1.59322E-05 |
| AQP6 | 0.031488197 | -0.042147307 | -0.010659111 |
| SH2D4A | 0.058243596 | -0.068278642 | -0.010035046 |
| TMEM87B | 0.050097136 | -0.037377918 | 0.012719219 |

| | | | |
|---|---|---|---|
| MDGA1 | -0.032783092 | 0.023811144 | -0.008971949 |
| IER3IP1 | 0.093276196 | -0.026565155 | 0.066711041 |
| MIR210HG | 0.049345587 | -0.0524022 | -0.003056613 |
| SLC39A10 | 0.08174884 | -0.054083126 | 0.027665714 |
| ERLEC1 | 0.081872765 | -0.048834496 | 0.033038269 |
| HIP1R | -0.094785036 | 0.06046005 | -0.034324986 |
| PPIB | 0.108675722 | 0.054522918 | 0.16319864 |
| TGS1 | 0.074319004 | -0.051399875 | 0.022919129 |
| FUT8 | 0.03847543 | -0.024155819 | 0.014319611 |
| MT2A | -0.094180126 | 0.155708826 | 0.0615287 |
| HLA-DPA1 | 0.044340947 | -0.046743894 | -0.002402947 |
| LINC01503 | -0.069771844 | 0.076131784 | 0.00635994 |
| IGFBP2 | 0.128883435 | -0.100405852 | 0.028477583 |
| ZHX3 | 0.07502618 | -0.021924732 | 0.053101448 |
| CD9 | 0.118842279 | -0.060348796 | 0.058493484 |
| VPS53 | 0.059466008 | -0.060944865 | -0.001478857 |
| CIRBP | -0.100151551 | 0.151113708 | 0.050962157 |
| HGD | 0.040095979 | -0.023217883 | 0.016878096 |
| DSP | 0.100204638 | -0.070495869 | 0.029708769 |
| PRR13 | -0.087641501 | 0.085505616 | -0.002135885 |
| GSN | 0.073597985 | -0.060274548 | 0.013323437 |
| ERO1A | 0.05786749 | -0.042836759 | 0.015030732 |
| ACKR3 | 0.07133408 | -0.030219992 | 0.041114088 |
| MIA2 | 0.07724677 | -0.03910266 | 0.03814411 |
| CDH4 | -0.019082278 | 0.005824436 | -0.013257842 |
| SYNJ2BP | -0.094714258 | 0.071619544 | -0.023094714 |
| PDIA3 | 0.098583722 | 0.040513372 | 0.139097094 |
| FTL | -0.120143081 | 0.116583992 | -0.003559089 |
| PDK4 | 0.059206623 | -0.017241081 | 0.041965542 |
| SERINC3 | 0.076387829 | -0.056581204 | 0.019806626 |
| TXNDC12 | 0.0719085 | -0.042027838 | 0.029880662 |
| SMAP1 | 0.056453092 | -0.035534826 | 0.020918266 |
| CLIC3 | -0.030002566 | 0.019659882 | -0.010342684 |
| EIF4G3 | 0.069084166 | -0.0530737 | 0.016010466 |
| GAB1 | 0.053150918 | -0.050506167 | 0.002644751 |
| C1GALT1 | 0.068435744 | -0.026890405 | 0.041545338 |
| SRGN | 0.02841827 | -0.019874146 | 0.008544123 |
| PKD2 | 0.084882017 | -0.064729973 | 0.020152043 |
| FAM3A | 0.063276141 | -0.020765356 | 0.042510785 |
| CFB | 0.058897095 | -0.043305128 | 0.015591967 |
| FOLR3 | -0.046036469 | 0.0464459 | 0.000409431 |
| AL096854.1 | 0.017778597 | -0.015444566 | 0.002334031 |
| ADRA2C | 0.054920617 | -0.044688577 | 0.010232039 |
| RB1 | 0.062114904 | -0.031983267 | 0.030131637 |
| FAM110B | -0.050732363 | 0.031692955 | -0.019039408 |
| PHF6 | 0.06304483 | -0.041951302 | 0.021093528 |
| THEM4 | 0.074972802 | -0.055502455 | 0.019470347 |

| | | | |
|---|---|---|---|
| COL9A2 | -0.083322824 | 0.088552599 | 0.005229775 |
| MARCKSL1 | 0.055853093 | -0.079060798 | -0.023207705 |
| PHLDA1 | -0.062642752 | 0.063292737 | 0.000649984 |
| PIGQ | -0.10162844 | 0.127398355 | 0.025769915 |
| AVPI1 | 0.041959016 | -0.030635639 | 0.011323378 |
| MESD | 0.076168943 | -0.027255478 | 0.048913465 |
| SEMA3B | -0.060230712 | 0.036685454 | -0.023545258 |
| AC008397.1 | 0.017779324 | -0.017552679 | 0.000226645 |
| CD63 | 0.103289886 | 0.055423445 | 0.158713331 |
| PAX2 | 0.095907227 | -0.032352683 | 0.063554545 |
| SLC39A7 | 0.067054558 | -0.040425991 | 0.026628568 |
| KLF5 | 0.057311823 | -0.059948873 | -0.00263705 |
| MT1X | -0.061739042 | 0.083934803 | 0.022195761 |
| ACBD5 | 0.071789628 | -0.068352142 | 0.003437486 |
| NR1D1 | 0.028414745 | -0.026067867 | 0.002346878 |
| SPESP1 | 0.034588437 | -0.036669958 | -0.002081521 |
| IFI27L2 | -0.074771729 | 0.062803125 | -0.011968605 |
| FUT2 | 0.018872281 | -0.020343336 | -0.001471055 |
| ELMO3 | 0.049045267 | -0.031730363 | 0.017314903 |
| AGMO | 0.024128668 | -0.0243489 | -0.000220232 |
| GALNT18 | 0.078632033 | -0.083228971 | -0.004596938 |
| PLCL2 | -0.056312476 | 0.049291833 | -0.007020644 |
| SESN3 | 0.056334566 | -0.025758551 | 0.030576015 |
| RASSF8-AS1 | 0.047816467 | -0.04575451 | 0.002061957 |
| PPP2R3A | 0.086385952 | -0.034865733 | 0.051520218 |
| RRAS | 0.048387002 | -0.038691248 | 0.009695755 |
| MACF1 | 0.093242756 | -0.071058965 | 0.02218379 |
| PIGF | 0.066169298 | -0.023979453 | 0.042189845 |
| FOLH1 | 0.020961917 | -0.016863929 | 0.004097988 |
| SLC39A1 | 0.076251411 | -0.042046974 | 0.034204437 |
| SEMA4B | 0.067216038 | -0.03248979 | 0.034726248 |
| GOLIM4 | -0.086505888 | 0.051085125 | -0.035420763 |
| HOOK2 | 0.069175324 | -0.025139274 | 0.04403605 |
| PDZK1IP1 | -0.043221031 | 0.056699445 | 0.013478414 |
| C6orf132 | 0.041973803 | -0.050909273 | -0.00893547 |
| RRAD | 0.05083839 | -0.030121559 | 0.020716831 |
| ARMC7 | 0.04635789 | -0.043145472 | 0.003212418 |
| RNF213 | 0.070136168 | -0.058568685 | 0.011567483 |
| PLEKHA1 | 0.072777881 | -0.049399546 | 0.023378334 |
| LINC01205 | 0.014263672 | -0.014688831 | -0.000425159 |
| SPPL3 | 0.070071853 | -0.024237244 | 0.045834609 |
| LITAF | 0.087417604 | -0.105690643 | -0.018273038 |
| ZNF326 | 0.063805809 | -0.048269536 | 0.015536273 |
| SREK1 | 0.072118896 | -0.032063031 | 0.040055864 |
| PAPSS2 | -0.063356792 | 0.022718681 | -0.040638112 |
| TTYH3 | 0.031922654 | -0.02490755 | 0.007015104 |
| MAL | 0.094488459 | 0.043817244 | 0.138305703 |

| | | | |
|---|---|---|---|
| ITSN2 | 0.059962498 | -0.046972285 | 0.012990213 |
| ADIPOR1 | 0.068220959 | -0.035643831 | 0.032577128 |
| ADD3 | -0.080334569 | 0.091978581 | 0.011644012 |
| VPS13A | 0.069676149 | -0.0391571 | 0.03051905 |
| TNKS1BP1 | 0.05652098 | -0.041159771 | 0.015361209 |
| GPX8 | 0.043122527 | -0.046080873 | -0.002958346 |
| PNRC2 | 0.08471142 | -0.082932152 | 0.001779267 |
| HAGH | 0.067962164 | -0.031720249 | 0.036241915 |
| SLC35A3 | 0.077179628 | -0.035812414 | 0.041367214 |
| IMPA2 | -0.068918851 | 0.087597252 | 0.018678401 |
| SLC4A4 | 0.086864681 | -0.079016764 | 0.007847917 |
| DUSP2 | -0.028727885 | 0.023689211 | -0.005038673 |
| TUBA1C | 0.07488033 | -0.029290099 | 0.045590231 |
| PLEKHF1 | 0.020598217 | -0.017328173 | 0.003270044 |
| ERAP2 | 0.038656999 | -0.02024825 | 0.018408749 |
| ZNF69 | 0.034054829 | -0.033903588 | 0.000151241 |
| RPS26 | -0.106268275 | 0.190464158 | 0.084195883 |
| COL18A1 | -0.096849888 | 0.217453193 | 0.120603305 |
| CARD6 | 0.030961353 | -0.028767674 | 0.00219368 |
| TMTC2 | -0.064858158 | 0.046718764 | -0.018139395 |
| WDR41 | 0.041560502 | -0.031746756 | 0.009813746 |
| CRIM1 | -0.084531874 | 0.062260221 | -0.022271653 |
| RAP1B | 0.076267779 | -0.02480038 | 0.051467399 |
| SLTM | 0.07567388 | -0.046827292 | 0.028846589 |
| MACC1 | -0.082602539 | 0.099869374 | 0.017266835 |
| THNSL2 | -0.041988137 | 0.040128404 | -0.001859733 |
| GLUL | 0.074556491 | -0.03284928 | 0.041707211 |
| COL4A2 | -0.073327132 | 0.078881499 | 0.005554367 |
| TNFRSF11B | 0.066366664 | -0.089403831 | -0.023037168 |
| NEDD4L | -0.090336016 | 0.105769245 | 0.015433229 |
| CAPN8 | 0.013768944 | -0.013768944 | 1.77636E-15 |
| CRABP2 | -0.031222118 | 0.015432369 | -0.015789749 |
| MYH10 | 0.095658191 | -0.067601164 | 0.028057028 |
| HEG1 | 0.033616274 | -0.049405534 | -0.01578926 |
| SH3BP5 | 0.035056359 | -0.012174266 | 0.022882092 |
| AL157373.2 | 0.025456067 | -0.027276235 | -0.001820168 |
| C4orf48 | -0.089966221 | 0.176550303 | 0.086584082 |
| CNGA1 | 0.062391836 | -0.044604828 | 0.017787007 |
| KTN1 | 0.095984557 | -0.07329701 | 0.022687547 |
| ARHGAP24 | -0.083912106 | 0.127151961 | 0.043239855 |
| SBSPON | 0.072976698 | -0.032403843 | 0.040572855 |
| ZMAT3 | 0.05391191 | -0.030605658 | 0.023306252 |
| ADCY9 | 0.046231794 | -0.046667743 | -0.000435948 |
| RDH11 | 0.077630102 | -0.029075299 | 0.048554803 |
| AFDN | 0.081398333 | -0.065080447 | 0.016317885 |
| DUSP1 | 0.079037153 | -0.056793789 | 0.022243365 |
| HNRNPR | 0.076226066 | -0.042332223 | 0.033893844 |

| | | | |
|---|---|---|---|
| JKAMP | 0.06176014 | -0.028471779 | 0.033288361 |
| KIAA1109 | 0.086896884 | -0.033385036 | 0.053511848 |
| GSDMD | 0.056010133 | -0.027031751 | 0.028978382 |
| SLC7A6 | 0.038240086 | -0.015695676 | 0.022544409 |
| RPL39 | -0.10302288 | 0.110044891 | 0.007022011 |
| GATA2-AS1 | 0.035525076 | -0.022919411 | 0.012605664 |
| PEBP1 | 0.096633133 | -0.114873353 | -0.018240221 |
| GRP | 0.015122073 | -0.015122073 | 5.32907E-15 |
| LYPD6B | 0.060175501 | -0.06037502 | -0.000199519 |
| ENO1 | 0.092179504 | -0.033398618 | 0.058780886 |
| HSD17B11 | 0.047301768 | -0.028651406 | 0.018650362 |
| SMAD7 | -0.054851512 | 0.023843178 | -0.031008334 |
| SPTBN1 | 0.090499046 | -0.055027181 | 0.035471865 |
| FLNA | -0.075254331 | 0.103584191 | 0.02832986 |
| ASS1 | -0.040205073 | 0.027991912 | -0.012213161 |
| COASY | 0.059696422 | -0.025887139 | 0.033809283 |
| SNTB2 | 0.044827708 | -0.042145657 | 0.002682051 |
| MIA3 | 0.067282207 | -0.023789315 | 0.043492892 |
| ST6GALNAC6 | 0.048906083 | -0.031725937 | 0.017180145 |
| ELOVL1 | 0.056112379 | -0.037232338 | 0.01888004 |
| SCD5 | -0.035975837 | 0.028937012 | -0.007038825 |
| NR4A2 | 0.023820608 | -0.019225252 | 0.004595356 |
| ANKLE2 | 0.052955418 | -0.040730863 | 0.012224555 |
| SYS1 | 0.065475762 | -0.039270845 | 0.026204918 |
| PLA2G15 | 0.04802355 | -0.035711144 | 0.012312405 |
| CAPG | 0.09475434 | -0.076056106 | 0.018698234 |
| ENTPD8 | 0.028715789 | -0.022952446 | 0.005763343 |
| SRSF3 | -0.084684387 | 0.134268045 | 0.049583658 |
| GALNT2 | 0.043437193 | -0.036096652 | 0.007340541 |
| PAQR8 | -0.056496007 | 0.032935952 | -0.023560056 |
| JAG1 | -0.058834431 | 0.048023915 | -0.010810516 |
| RAB9A | 0.059953703 | -0.02269364 | 0.037260063 |
| CTIF | -0.037028563 | 0.027772144 | -0.009256418 |
| TCEAL2 | -0.055261545 | 0.078172509 | 0.022910964 |
| IGFBP7 | 0.090753797 | 0.054747453 | 0.14550125 |
| THRA | 0.065462363 | -0.056841491 | 0.008620872 |
| SRSF6 | -0.072435162 | 0.060314699 | -0.012120463 |
| MANSC1 | 0.057731745 | -0.039284363 | 0.018447382 |
| SPPL2A | 0.07267266 | -0.047574754 | 0.025097906 |
| ARRDC3 | -0.063687009 | 0.062531768 | -0.00115524 |
| CD74 | 0.084977544 | -0.071203689 | 0.013773855 |
| TAPT1 | 0.049581903 | -0.036973071 | 0.012608832 |
| TMBIM1 | 0.067974986 | -0.026870303 | 0.041104683 |
| IARS2 | 0.068786151 | -0.039625771 | 0.02916038 |
| REL | 0.042000905 | -0.045055888 | -0.003054983 |
| TRNP1 | -0.034026658 | 0.017934031 | -0.016092627 |
| TRIM38 | 0.056930658 | -0.028401802 | 0.028528856 |

| | | | |
|---|---|---|---|
| NFAT5 | 0.077006688 | -0.044538175 | 0.032468513 |
| SELENBP1 | 0.051816185 | -0.034570902 | 0.017245283 |
| SLC7A2 | 0.022227726 | -0.027590877 | -0.005363151 |
| DAP | 0.069479517 | -0.061915742 | 0.007563775 |
| ARPC2 | 0.083603728 | -0.057564938 | 0.026038789 |
| ARL6IP6 | 0.03970644 | -0.030271257 | 0.009435183 |
| CADM2 | 0.013529895 | -0.01363169 | -0.000101795 |
| PDE3B | 0.036031351 | 0.019160962 | 0.055192313 |
| SEPTIN7 | 0.080790209 | -0.026967241 | 0.053822968 |
| HLA-C | 0.093919619 | 0.045747509 | 0.139667128 |
| SLC2A6 | 0.01892366 | -0.019725003 | -0.000801343 |
| AC103740.1 | 0.014030662 | -0.01400015 | 3.05111E-05 |
| PPP1R9A | 0.071500079 | -0.055462316 | 0.016037763 |
| WNK1 | 0.10222303 | 0.091786324 | 0.194009354 |
| APMAP | 0.068082187 | -0.025220138 | 0.042862049 |
| AC022075.1 | 0.019559735 | -0.019060077 | 0.000499658 |
| LRP10 | 0.072913641 | -0.034308056 | 0.038605586 |
| TMF1 | 0.064613475 | -0.039009798 | 0.025603676 |
| TMEM80 | 0.062473576 | -0.052823785 | 0.009649792 |
| SERINC1 | 0.081056319 | -0.040628711 | 0.040427608 |
| GLRX | -0.069507548 | 0.055826244 | -0.013681304 |
| NAPRT | -0.06005171 | 0.045875481 | -0.014176229 |
| OGDHL | -0.074628102 | 0.148103521 | 0.073475419 |
| GABBR1 | 0.045531201 | -0.047882811 | -0.00235161 |
| NBL1 | -0.072081578 | 0.175080182 | 0.102998604 |
| AC004540.1 | -0.05400882 | 0.033057905 | -0.020950916 |
| FHL1 | 0.034210646 | -0.022860038 | 0.011350608 |
| BEX2 | -0.079005141 | 0.111078454 | 0.032073313 |
| EVA1B | 0.051425235 | -0.018350596 | 0.033074639 |
| PSME1 | 0.076012013 | -0.039544801 | 0.036467212 |
| CEBPD | 0.081873726 | -0.041068717 | 0.040805009 |
| KANSL1L | -0.066628043 | 0.092920213 | 0.02629217 |
| DYNC1H1 | 0.076639656 | -0.035670759 | 0.040968897 |
| NRDC | 0.058363168 | -0.026964027 | 0.031399141 |
| AKR1C3 | 0.049574733 | -0.044638285 | 0.004936448 |
| SETD3 | 0.057804037 | -0.03807027 | 0.019733767 |
| HVCN1 | 0.013893804 | -0.013713218 | 0.000180586 |
| SKIL | 0.051486146 | -0.044851719 | 0.006634427 |
| IRF2BP2 | -0.065293398 | 0.096767812 | 0.031474415 |
| NDUFV2 | 0.086276978 | 0.044935064 | 0.131212043 |
| CPNE7 | 0.013631527 | -0.013973912 | -0.000342385 |
| TMEM243 | 0.064033034 | -0.028640746 | 0.035392288 |
| IFNAR1 | 0.074510045 | -0.036604707 | 0.037905338 |
| SH3BP2 | 0.047018234 | -0.049924378 | -0.002906145 |
| AC025164.1 | 0.030032582 | -0.021064712 | 0.00896787 |
| TFCP2L1 | -0.084752109 | 0.11690083 | 0.032148721 |
| TMED10 | 0.0839301 | 0.03171893 | 0.11564903 |

| | | | |
|---|---|---|---|
| KCNS3 | 0.031252228 | -0.018072538 | 0.01317969 |
| C4orf54 | -0.036948993 | 0.021128261 | -0.015820732 |
| GPR155 | 0.041708913 | -0.028188205 | 0.013520708 |
| MLX | 0.045745679 | -0.03065651 | 0.015089169 |
| MAPK10 | 0.067399236 | -0.034242473 | 0.033156763 |
| STT3B | 0.0631993 | -0.023655251 | 0.039544049 |
| TK1 | 0.028477143 | -0.030232453 | -0.001755311 |
| HSPA1B | -0.077821283 | 0.073675323 | -0.00414596 |
| TRMT9B | -0.025425767 | 0.009754642 | -0.015671125 |
| SLC16A5 | 0.06036999 | -0.07986496 | -0.01949497 |
| C1orf210 | 0.04906964 | -0.026430568 | 0.022639071 |
| CCND1 | -0.063111091 | 0.044814341 | -0.01829675 |
| ARL4D | 0.076451984 | 0.036338901 | 0.112790885 |
| COL6A1 | -0.05727282 | 0.06251326 | 0.00524044 |
| KCTD12 | -0.070000923 | 0.059059132 | -0.010941791 |
| NOL7 | 0.072293819 | -0.037118611 | 0.035175208 |
| EGFR | 0.06850049 | -0.076776109 | -0.008275619 |
| PNMA1 | 0.051558486 | -0.025847443 | 0.025711043 |
| MUC1 | 0.085519618 | -0.031960079 | 0.053559539 |
| RPS17 | -0.082545561 | 0.066261754 | -0.016283807 |
| SPNS1 | 0.041027978 | -0.023278181 | 0.017749797 |
| S100A13 | 0.091049852 | -0.058894309 | 0.032155543 |
| GATA2 | -0.079101289 | 0.14602728 | 0.066925991 |
| GRIN2B | -0.019197813 | 0.011165575 | -0.008032238 |
| SLC15A1 | 0.012898881 | -0.01204781 | 0.000851071 |
| CTNNB1 | -0.073958738 | 0.041831993 | -0.032126744 |
| TFF1 | 0.015323262 | -0.015472846 | -0.000149584 |
| CALM2 | -0.08981588 | 0.149966778 | 0.060150898 |
| GBP4 | 0.01880945 | -0.020672202 | -0.001862753 |
| RPL27A | -0.084910812 | 0.09985492 | 0.014944109 |
| COX17 | 0.08838469 | -0.044593245 | 0.043791445 |
| PGP | 0.056436336 | -0.024801195 | 0.031635141 |
| NCEH1 | 0.035666207 | -0.027572256 | 0.008093951 |
| CLCN4 | 0.027182984 | -0.021699503 | 0.005483481 |
| MIF4GD | 0.044155092 | -0.034909511 | 0.009245581 |
| VSIG10 | 0.046025595 | -0.044548238 | 0.001477357 |
| TLCD5 | 0.03808792 | -0.024327706 | 0.013760213 |
| GRN | 0.084687701 | -0.035752862 | 0.048934839 |
| UBXN4 | 0.078841795 | -0.038435714 | 0.040406081 |
| HSPE1 | -0.089340305 | 0.159615874 | 0.070275569 |
| SLC22A23 | 0.031024982 | -0.030748568 | 0.000276414 |
| BNIP3L | 0.059346343 | -0.039142186 | 0.020204157 |
| AP1AR | 0.05445114 | -0.036096989 | 0.018354151 |
| PTGES | 0.027694905 | -0.02718569 | 0.000509215 |
| YES1 | 0.048736453 | -0.026314904 | 0.022421549 |
| JADE1 | -0.061263508 | 0.051060543 | -0.010202965 |
| MPHOSPH8 | 0.072474401 | -0.076756273 | -0.004281873 |

| | | | |
|---|---|---|---|
| SLC35B1 | 0.046884577 | -0.026801612 | 0.020082965 |
| TRPS1 | 0.052521643 | -0.039817225 | 0.012704418 |
| BMPR1A | 0.040076438 | -0.017523332 | 0.022553106 |
| MT1E | -0.068760103 | 0.144727187 | 0.075967084 |
| PPFIBP1 | 0.063255529 | -0.033855042 | 0.029400488 |
| GARS | 0.042615437 | -0.031377357 | 0.011238079 |
| PER1 | 0.033128283 | -0.021049348 | 0.012078935 |
| RALYL | -0.05435204 | 0.066346836 | 0.011994796 |
| MKLN1-AS | 0.023334813 | -0.023701156 | -0.000366343 |
| GXYLT1 | 0.047211221 | -0.028343049 | 0.018868172 |
| KIF5B | 0.075075313 | -0.03772131 | 0.037354003 |
| SLC49A4 | 0.040300883 | -0.018408288 | 0.021892595 |
| NECTIN1 | -0.034666287 | 0.025162443 | -0.009503844 |
| UQCRB | -0.090488821 | 0.153293691 | 0.06280487 |
| SOCS2 | -0.050220276 | 0.07599706 | 0.025776784 |
| DSG2 | -0.052127095 | 0.058324016 | 0.006196922 |
| CALD1 | 0.030465249 | -0.022770924 | 0.007694326 |
| PPP4R1 | 0.044064763 | -0.032485931 | 0.011578832 |
| GUCY1A1 | -0.025647712 | 0.019286933 | -0.006360778 |
| LY75 | 0.05551033 | -0.025256864 | 0.030253466 |
| NCS1 | -0.041344844 | 0.02898446 | -0.012360384 |
| ESYT2 | 0.064382838 | -0.041270135 | 0.023112703 |
| FAM50A | 0.050047336 | -0.029256022 | 0.020791314 |
| SRSF5 | -0.077710534 | 0.147348236 | 0.069637703 |
| OSBPL3 | 0.047551348 | -0.025861158 | 0.021690189 |
| CHST15 | 0.011809244 | -0.010956392 | 0.000852852 |
| TNIP2 | 0.039437455 | -0.020943076 | 0.018494379 |
| RPL7 | -0.086164902 | 0.092802674 | 0.006637772 |
| STIM2 | 0.067169142 | 0.085703545 | 0.152872687 |
| EIF2AK4 | 0.049633665 | -0.044030263 | 0.005603402 |
| SRCIN1 | 0.011909133 | -0.012232941 | -0.000323808 |
| CAPS | 0.079820534 | -0.052101283 | 0.027719251 |
| FMN1 | 0.017022645 | -0.015229359 | 0.001793286 |
| AC009244.2 | 0.015695693 | -0.014413489 | 0.001282204 |
| TBCEL | 0.036219202 | -0.028524348 | 0.007694855 |
| ADAMTS16 | -0.035977304 | 0.038205357 | 0.002228054 |
| POU2F3 | 0.02061749 | -0.019822477 | 0.000795014 |
| ORMDL3 | 0.045598095 | -0.020964211 | 0.024633884 |
| PLEKHH1 | 0.036404683 | -0.032903615 | 0.003501068 |
| SYCE1L | -0.025815903 | 0.012959797 | -0.012856107 |
| SPECC1L | 0.04649612 | -0.043501096 | 0.002995024 |
| FADS1 | -0.02711437 | 0.017539208 | -0.009575163 |
| TNS4 | 0.015972223 | -0.017905766 | -0.001933543 |
| TSPAN13 | 0.031811281 | -0.021177083 | 0.010634197 |
| TMCC3 | 0.035661107 | -0.020181974 | 0.015479133 |
| STOM | -0.042773606 | 0.037610115 | -0.00516349 |
| PLXNB2 | 0.077509803 | -0.032580952 | 0.044928851 |

| | | | |
|---|---|---|---|
| PARK7 | 0.081685044 | 0.041362632 | 0.123047676 |
| TOP1MT | 0.027815277 | -0.024430806 | 0.003384472 |
| MTDH | 0.074741771 | -0.068244451 | 0.00649732 |
| MRAP2 | 0.015467686 | -0.017476368 | -0.002008682 |
| MISP3 | 0.040552381 | -0.027075643 | 0.013476737 |
| ACTB | -0.07803605 | 0.161133536 | 0.083097486 |
| POPDC2 | 0.016231519 | -0.013722476 | 0.002509043 |
| PIEZO1 | 0.061215934 | -0.031610027 | 0.029605907 |
| NBPF9 | 0.017883096 | -0.014729121 | 0.003153974 |
| DCTN2 | 0.056493342 | -0.023064854 | 0.033428488 |
| JAM3 | -0.042882661 | 0.044993497 | 0.002110836 |
| WFS1 | 0.041286737 | -0.03384871 | 0.007438027 |
| DDX46 | 0.059863991 | -0.040628007 | 0.019235984 |
| LINC00882 | 0.048837584 | -0.032676259 | 0.016161324 |
| CPEB2 | 0.033039035 | -0.027347941 | 0.005691094 |
| TTF2 | 0.026923393 | -0.021616622 | 0.005306772 |
| IFRD1 | 0.0349331 | -0.017331202 | 0.017601899 |
| ARRDC1 | 0.046816703 | -0.039179763 | 0.00763694 |
| ZNF618 | -0.052665027 | 0.036471392 | -0.016193635 |
| TACC2 | 0.042641913 | -0.021515811 | 0.021126102 |
| ARPC5L | 0.055111973 | -0.036658259 | 0.018453713 |
| C9orf16 | 0.078563569 | -0.037493052 | 0.041070518 |
| KIF26B | 0.019055907 | -0.012544847 | 0.00651106 |
| SNHG29 | -0.084739399 | 0.068609079 | -0.01613032 |
| NBAS | 0.043101383 | -0.025726665 | 0.017374718 |
| WASIR2 | 0.010784306 | -0.010935643 | -0.000151337 |
| AMOTL2 | -0.053041483 | 0.028038379 | -0.025003104 |
| PALM3 | 0.051314545 | -0.047418122 | 0.003896423 |
| CSRP2 | -0.029235014 | 0.013912559 | -0.015322455 |
| NR4A3 | 0.01464184 | -0.014131986 | 0.000509853 |
| LCMT1 | 0.042565326 | -0.016667289 | 0.025898038 |
| KCNJ13 | 0.036489719 | -0.033683357 | 0.002806362 |
| ECT2 | 0.02817002 | -0.021184307 | 0.006985713 |
| LRP8 | 0.02050991 | -0.009950904 | 0.010559006 |
| ZFP36L1 | -0.078489531 | 0.072914221 | -0.00557531 |
| RAB5IF | 0.063406245 | -0.026626729 | 0.036779516 |
| RFLNB | 0.021609719 | -0.020109337 | 0.001500382 |
| TGM2 | 0.014096799 | -0.012535457 | 0.001561342 |
| CD99 | 0.05767232 | -0.056188283 | 0.001484038 |
| ERI1 | 0.025520304 | -0.0232318 | 0.002288504 |
| NSD3 | 0.064288642 | -0.048603108 | 0.015685533 |
| RBP5 | -0.019060883 | 0.008759941 | -0.010300942 |
| TMX1 | 0.057031181 | -0.023079132 | 0.033952049 |
| LEMD2 | 0.034920654 | -0.026495216 | 0.008425438 |
| MANBAL | 0.052524455 | -0.021612769 | 0.030911687 |
| BEX3 | -0.084745139 | 0.120562416 | 0.035817277 |
| ATP13A3 | 0.060781975 | -0.027109931 | 0.033672044 |

| | | | |
|---|---|---|---|
| CLDN19 | -0.018088682 | 0.007666568 | -0.010422114 |
| NADK2 | 0.037891948 | -0.024551583 | 0.013340364 |
| DUSP6 | 0.045673543 | 0.035090703 | 0.080764246 |
| TMEM59 | 0.080396073 | 0.051851599 | 0.132247672 |
| SNTB1 | 0.021280966 | -0.016385653 | 0.004895313 |
| AKAP9 | 0.075650848 | -0.046325125 | 0.029325723 |
| EFEMP2 | -0.049180396 | 0.06047731 | 0.011296914 |
| NYAP1 | 0.016127963 | -0.015253357 | 0.000874606 |
| FZD1 | -0.045444625 | 0.058586082 | 0.013141457 |
| BCL2L1 | -0.052673029 | 0.049385453 | -0.003287576 |
| MDFIC | 0.042760503 | -0.033656338 | 0.009104165 |
| TEX41 | -0.05943958 | 0.130160396 | 0.070720815 |
| VASP | 0.039158775 | -0.031029605 | 0.00812917 |
| TSPAN5 | -0.048787525 | 0.044897837 | -0.003889687 |
| HOTAIRM1 | -0.033463086 | 0.026171525 | -0.007291561 |
| CADM1 | -0.038772157 | 0.043240927 | 0.004468769 |
| CWH43 | 0.052734894 | -0.032400754 | 0.020334141 |
| SOCS1 | -0.040978939 | 0.058141467 | 0.017162529 |
| SPTLC2 | -0.067730265 | 0.107457229 | 0.039726964 |
| FAT2 | 0.026296039 | -0.019626211 | 0.006669828 |
| RPL24 | -0.082163341 | 0.112045416 | 0.029882075 |
| CC2D2A | 0.038597192 | -0.03107222 | 0.007524973 |
| SUCO | 0.050004624 | -0.025499064 | 0.02450556 |
| CCDC198 | -0.06891245 | 0.084098719 | 0.015186268 |
| PPARG | 0.041052419 | -0.020366623 | 0.020685796 |
| RPS20 | -0.067986534 | 0.059155143 | -0.008831391 |
| ISG15 | 0.027346545 | -0.024954806 | 0.002391739 |
| KLHDC7A | -0.045028017 | 0.047852463 | 0.002824446 |
| PRSS23 | -0.050094429 | 0.091976562 | 0.041882133 |
| MAN1C1 | 0.056785219 | -0.034206495 | 0.022578724 |
| HMGCS2 | 0.078964452 | 0.153917111 | 0.232881562 |
| HLA-A | 0.075645213 | 0.087517046 | 0.163162259 |
| PNP | 0.070307764 | 0.073534918 | 0.143842682 |
| LPIN1 | -0.038028634 | 0.027011229 | -0.011017405 |
| BTN2A1 | 0.029812588 | -0.022067232 | 0.007745356 |
| PCBP2 | -0.07200085 | 0.083773432 | 0.011772582 |
| PARP4 | 0.043542657 | -0.045661396 | -0.002118739 |
| CCDC47 | 0.063433168 | -0.038742762 | 0.024690406 |
| TCHH | 0.010165347 | -0.009890828 | 0.000274519 |
| TLCD3A | 0.018104804 | -0.01427301 | 0.003831794 |
| SLC16A11 | -0.020422433 | 0.017710214 | -0.002712219 |
| IDH2 | -0.063351363 | 0.124029177 | 0.060677814 |
| SMC4 | 0.041328954 | -0.030798958 | 0.010529996 |
| PRKDC | 0.064560964 | -0.03744086 | 0.027120103 |
| STK38 | 0.037784878 | -0.035843051 | 0.001941827 |
| CYS1 | -0.074491424 | 0.19289055 | 0.118399126 |
| IL10RB | 0.03475108 | -0.029343667 | 0.005407413 |

| | | | |
|---|---|---|---|
| IFITM2 | 0.063740652 | -0.036343328 | 0.027397324 |
| SPIDR | 0.033793826 | -0.01537942 | 0.018414405 |
| SRSF10 | 0.064939211 | -0.047187345 | 0.017751866 |
| AMPD2 | 0.034916968 | -0.025966214 | 0.008950754 |
| TMEM259 | 0.056674206 | -0.028060511 | 0.028613695 |
| NDUFB1 | -0.078142023 | 0.118179345 | 0.040037322 |
| MOB3B | -0.032253928 | 0.022898816 | -0.009355112 |
| GALM | 0.038902036 | -0.031647098 | 0.007254938 |
| KIAA2026 | 0.04608191 | -0.020654184 | 0.025427725 |
| PSMB3 | 0.06415237 | 0.027004416 | 0.091156786 |
| CCT5 | 0.052816261 | -0.028759425 | 0.024056836 |
| SST | 0.016631493 | -0.014103727 | 0.002527766 |
| RHOJ | 0.009228259 | -0.008750944 | 0.000477315 |
| NIPAL1 | -0.065185478 | 0.027084161 | -0.038101317 |
| LINC01224 | 0.015276527 | -0.014821952 | 0.000454574 |
| ADAMTS6 | 0.013781659 | -0.013295144 | 0.000486514 |
| SOWAHD | 0.016890071 | -0.011913767 | 0.004976304 |
| BICD1 | -0.042385446 | 0.020374564 | -0.022010882 |
| AC009509.1 | 0.008820636 | -0.008388312 | 0.000432324 |
| RPS4Y1 | 0.065458489 | -0.213294897 | -0.147836408 |
| COL11A1 | 0.008896132 | -0.008539491 | 0.000356641 |
| NUP93 | 0.028850188 | -0.019615401 | 0.009234787 |
| LINC01924 | 0.008574725 | -0.008574725 | 7.10543E-15 |
| EID2 | 0.031925425 | -0.022401043 | 0.009524381 |
| ZNF331 | 0.035998809 | -0.028308889 | 0.00768992 |
| RHEX | -0.011823668 | 0.00557729 | -0.006246378 |
| SUGT1 | 0.055612383 | -0.034072958 | 0.021539425 |
| LPCAT3 | 0.043994824 | -0.019738599 | 0.024256225 |
| TRIM2 | -0.057600888 | 0.077590098 | 0.019989209 |
| CLINT1 | 0.064043785 | -0.044029293 | 0.020014492 |
| CHL1 | -0.022695182 | 0.016399842 | -0.006295341 |
| TCTA | 0.044493935 | -0.027400148 | 0.017093787 |
| CYFIP2 | -0.073660292 | 0.110872733 | 0.03721244 |
| LMCD1 | -0.020676016 | 0.012244766 | -0.008431249 |
| NDNF | 0.009966936 | -0.010129903 | -0.000162967 |
| TMEM125 | 0.039080126 | -0.016148004 | 0.022932122 |
| HOXA7 | -0.026562101 | 0.01755054 | -0.009011561 |
| TRIM47 | 0.016431739 | -0.014323889 | 0.00210785 |
| LGI2 | 0.013983247 | -0.013895358 | 8.78889E-05 |
| CLCN7 | 0.043900099 | -0.037952136 | 0.005947963 |
| PDZRN3 | 0.023343839 | -0.021578855 | 0.001764984 |
| M6PR | 0.05696207 | -0.041152731 | 0.015809339 |
| FHOD3 | 0.05385756 | -0.022663959 | 0.031193601 |
| ITM2A | -0.015643874 | 0.007866081 | -0.007777793 |
| KRT19 | 0.079957719 | 0.097320761 | 0.177278481 |
| ABCC3 | 0.016370378 | -0.011143823 | 0.005226555 |
| STK24 | 0.045343261 | -0.039478097 | 0.005865164 |

| | | | |
|---|---|---|---|
| PDE4D | 0.062171766 | 0.09383908 | 0.156010846 |
| TFPI | 0.022562146 | -0.033906991 | -0.011344845 |
| MIF | 0.073371676 | 0.062297451 | 0.135669127 |
| TCF7L2 | -0.044680157 | 0.031417804 | -0.013262353 |
| TMUB2 | 0.041651753 | -0.020353912 | 0.021297841 |
| H2AFZ | 0.071008799 | -0.052227202 | 0.018781597 |
| ESAM | 0.02649263 | -0.027245358 | -0.000752728 |
| TPM4 | 0.079420296 | -0.068739196 | 0.0106811 |
| RPS29 | -0.075160746 | 0.045652216 | -0.02950853 |
| ATP5PB | -0.059671655 | 0.076033809 | 0.016362154 |
| FIRRE | 0.010078708 | -0.009961151 | 0.000117558 |
| RCN2 | 0.064049474 | 0.032255511 | 0.096304984 |
| ZDHHC3 | 0.057062957 | -0.033693448 | 0.023369508 |
| CD58 | 0.02756353 | -0.0242957 | 0.00326783 |
| HLA-E | 0.064388272 | -0.02926444 | 0.035123832 |
| C2CD2L | 0.024323183 | -0.018791231 | 0.005531952 |
| CDK2AP1 | 0.052047855 | -0.022178291 | 0.029869564 |
| LRRFIP2 | 0.042421964 | -0.032037559 | 0.010384405 |
| LCMT1-AS2 | 0.015382981 | -0.010182328 | 0.005200653 |
| ATE1 | 0.037206157 | -0.016512455 | 0.020693702 |
| SRD5A3 | 0.027555721 | -0.018834261 | 0.00872146 |
| MAFK | 0.039805718 | -0.028316701 | 0.011489017 |
| FGFR2 | -0.040040604 | 0.017143711 | -0.022896893 |
| PARP14 | 0.046582159 | -0.027844446 | 0.018737714 |
| DIAPH1 | 0.05145485 | -0.033018716 | 0.018436134 |
| WNT11 | 0.008646458 | -0.008366095 | 0.000280363 |
| PRPF38B | 0.056765645 | -0.049447183 | 0.007318463 |
| MAP3K3 | 0.029750486 | -0.022714869 | 0.007035617 |
| SORBS3 | -0.023170144 | 0.014387053 | -0.00878309 |
| CKMT2-AS1 | 0.02718786 | -0.017576386 | 0.009611474 |
| PNPLA6 | 0.044555734 | -0.024082989 | 0.020472745 |
| PTH1R | -0.03688336 | 0.030812218 | -0.006071142 |
| IFI44L | 0.018374355 | -0.01214896 | 0.006225395 |
| HERC2 | 0.047482095 | -0.043689382 | 0.003792713 |
| SGCZ | -0.018492849 | 0.01210422 | -0.006388628 |
| CMBL | -0.042165694 | 0.031238492 | -0.010927202 |
| RPL23 | -0.071819565 | 0.077925697 | 0.006106132 |
| TMEM61 | -0.023621391 | 0.013065192 | -0.010556198 |
| LINC01315 | 0.016693396 | -0.01365917 | 0.003034226 |
| CDA | -0.033099457 | 0.031686075 | -0.001413381 |
| FOXA3 | 0.016837887 | -0.014756367 | 0.00208152 |
| UBR4 | 0.036303673 | -0.023460627 | 0.012843046 |
| AC245297.3 | 0.043681165 | -0.038098304 | 0.005582861 |
| HAUS1 | 0.02332686 | -0.01601265 | 0.00731421 |
| PARM1 | -0.035041015 | 0.041978352 | 0.006937337 |
| MT-ND4L | -0.054143208 | 0.033412999 | -0.020730209 |
| RAD50 | 0.054006389 | -0.036838736 | 0.017167653 |

| | | | |
|---|---|---|---|
| FCGBP | 0.039618542 | -0.053497115 | -0.013878573 |
| LMTK2 | 0.032273189 | -0.030244636 | 0.002028553 |
| LINC01508 | -0.018755849 | 0.008338564 | -0.010417284 |
| AL031275.1 | 0.01284451 | -0.011225611 | 0.001618898 |
| TNFRSF1A | 0.03323218 | -0.017554917 | 0.015677263 |
| ENPP4 | 0.0422992 | -0.021939424 | 0.020359776 |
| RTL5 | -0.024021658 | 0.019828961 | -0.004192697 |
| ZNF138 | 0.034745751 | -0.024648206 | 0.010097546 |
| GABRE | 0.022415653 | -0.033713955 | -0.011298302 |
| GOLGA8B | -0.040585853 | 0.043355414 | 0.00276956 |
| TMEM144 | 0.017435299 | -0.01636243 | 0.001072868 |
| SKAP2 | -0.034186826 | 0.019424874 | -0.014761952 |
| AARD | 0.013295383 | -0.012454607 | 0.000840776 |
| HDAC1 | 0.053534797 | -0.028304811 | 0.025229986 |
| ARHGDIB | 0.031818319 | -0.020788941 | 0.011029378 |
| ADAM22 | 0.040573835 | -0.03688112 | 0.003692716 |
| NDUFA4 | -0.072572886 | 0.142679369 | 0.070106483 |
| KIDINS220 | 0.052906294 | -0.046554757 | 0.006351537 |
| AC004148.1 | 0.01755526 | -0.017506052 | 4.92085E-05 |
| EIF2S2 | 0.051369116 | -0.028023503 | 0.023345613 |
| RBM33 | 0.036998434 | -0.020477023 | 0.016521411 |
| LRWD1 | 0.022390485 | -0.011554227 | 0.010836258 |
| AFMID | 0.053503509 | -0.080147946 | -0.026644437 |
| AC006230.1 | 0.01733763 | -0.014892605 | 0.002445025 |
| SOX5 | 0.027615764 | -0.025910871 | 0.001704894 |
| ITPRID2 | 0.031898542 | -0.028202743 | 0.003695799 |
| MPC1 | -0.068894791 | 0.138291682 | 0.069396891 |
| NUP50 | 0.039463609 | -0.034787116 | 0.004676493 |
| CNBP | -0.059589602 | 0.062396591 | 0.002806989 |
| AL512662.2 | 0.023801145 | -0.021153198 | 0.002647947 |
| METAP2 | -0.05849319 | 0.055303036 | -0.003190155 |
| SLC25A37 | 0.049254842 | -0.059747532 | -0.01049269 |
| CGRRF1 | 0.035865739 | -0.022955617 | 0.012910122 |
| STON2 | -0.035736354 | 0.028243965 | -0.007492389 |
| SMTNL2 | 0.032025964 | -0.016059154 | 0.01596681 |
| UBTD1 | 0.025487526 | -0.022454199 | 0.003033327 |
| STC1 | -0.086160471 | 0.157865888 | 0.071705418 |
| AL033384.1 | 0.00971521 | -0.009659762 | 5.5448E-05 |
| TMEM41B | 0.041023788 | -0.023702322 | 0.017321466 |
| IQUB | -0.045288989 | 0.032384977 | -0.012904012 |
| MECOM | 0.069313171 | 0.055231268 | 0.124544438 |
| TCEA3 | -0.050237182 | 0.034005931 | -0.016231251 |
| SERINC5 | 0.033678874 | -0.027563734 | 0.00611514 |
| DAPK1 | 0.043071155 | -0.041116379 | 0.001954776 |
| TMCO4 | 0.035426619 | -0.031740737 | 0.003685882 |
| FAM234A | 0.048456051 | -0.022656607 | 0.025799444 |
| RMC1 | 0.020034606 | -0.010163637 | 0.009870969 |

| | | | |
|---|---|---|---|
| SLC33A1 | 0.035295795 | -0.017646003 | 0.017649792 |
| IFI27L1 | 0.035101333 | -0.016383027 | 0.018718306 |
| WEE1 | -0.030677629 | 0.026203184 | -0.004474445 |
| PDE9A | 0.029508702 | -0.020448311 | 0.009060391 |
| TACC3 | 0.010701969 | -0.008879216 | 0.001822753 |
| TCAIM | -0.037134362 | 0.020904788 | -0.016229574 |
| ATP7A | 0.033835296 | -0.024689494 | 0.009145802 |
| MET | 0.021451743 | -0.013475249 | 0.007976494 |
| DCP2 | 0.039228431 | -0.0355751 | 0.003653332 |
| L2HGDH | -0.02885355 | 0.01503703 | -0.013816521 |
| EBPL | 0.050714231 | -0.054852918 | -0.004138687 |
| PSMB10 | 0.032098579 | -0.021688476 | 0.010410103 |
| INPP5J | 0.057086536 | -0.027833883 | 0.029252653 |
| HSPA8 | -0.067434683 | 0.149762847 | 0.082328164 |
| COX7A1 | -0.03609344 | 0.045300151 | 0.009206711 |
| MIR22HG | 0.027135 | -0.022318108 | 0.004816892 |
| SLC39A6 | 0.067587053 | 0.093460748 | 0.161047801 |
| QSOX1 | -0.046493847 | 0.05857253 | 0.012078683 |
| CNKSR3 | -0.030872417 | 0.033934796 | 0.003062379 |
| REEP4 | 0.021123493 | -0.021571843 | -0.00044835 |
| PPARGC1A | -0.048235799 | 0.064494535 | 0.016258736 |
| RASSF7 | 0.062956729 | -0.034448118 | 0.028508611 |
| PHAX | 0.041171998 | -0.028420347 | 0.012751652 |
| WASHC1 | -0.043131547 | 0.020882244 | -0.022249303 |
| AGPAT2 | 0.052757941 | -0.037201194 | 0.015556747 |
| TNNC1 | -0.015179645 | 0.007698655 | -0.00748099 |
| MARK3 | 0.046662931 | -0.034236452 | 0.012426479 |
| ALB | -0.01077206 | 0.005595 | -0.00517706 |
| RRAGD | -0.032592948 | 0.025518139 | -0.007074808 |
| USP34 | 0.054003689 | -0.033821461 | 0.020182228 |
| C16orf87 | 0.037263833 | -0.02012946 | 0.017134372 |
| ACOX2 | 0.013570488 | -0.008567496 | 0.005002993 |
| ADH1C | -0.057743031 | 0.118446096 | 0.060703066 |
| HUNK | -0.028717303 | 0.02609656 | -0.002620743 |
| CLPTM1L | 0.043487213 | -0.022129927 | 0.021357286 |
| RNF14 | 0.032918075 | -0.018276212 | 0.014641863 |
| AC022424.1 | -0.02356718 | 0.021856225 | -0.001710956 |
| FAM124A | -0.01722168 | 0.010066814 | -0.007154866 |
| LINC-PINT | 0.033624998 | -0.039891625 | -0.006266627 |
| WWTR1 | 0.051916697 | -0.034225363 | 0.017691334 |
| ZBTB1 | 0.038451674 | -0.029961107 | 0.008490567 |
| PDK1 | 0.018079333 | -0.016643441 | 0.001435892 |
| OSMR | 0.021707388 | -0.015951096 | 0.005756293 |
| EPB41L1 | -0.048780367 | 0.052459803 | 0.003679435 |
| AUTS2 | 0.019344201 | -0.028842348 | -0.009498147 |
| WNK4 | -0.032380959 | 0.02494411 | -0.007436849 |
| SLC12A7 | 0.037581968 | -0.039640775 | -0.002058807 |

| | | | |
|---|---|---|---|
| MFSD4A | -0.031608774 | 0.021103203 | -0.010505571 |
| FUT11 | 0.028362016 | -0.027065805 | 0.001296211 |
| AKR1C2 | 0.006364906 | -0.005784363 | 0.000580543 |
| COQ8A | 0.031426106 | -0.020286436 | 0.01113967 |
| ILF3-DT | -0.042399224 | 0.032768065 | -0.009631159 |
| MUC19 | 0.007591041 | -0.00842733 | -0.000836289 |
| GHR | -0.028380921 | 0.03456768 | 0.006186759 |
| FAM149A | -0.037770456 | 0.033897358 | -0.003873098 |
| KIFAP3 | 0.035466249 | -0.016842331 | 0.018623918 |
| THY1 | -0.045080302 | 0.032948122 | -0.01213218 |
| OAS1 | 0.009983979 | -0.007717564 | 0.002266415 |
| DAPK1-IT1 | 0.017177976 | -0.02068296 | -0.003504984 |
| SPTLC1 | 0.046813134 | -0.029391037 | 0.017422098 |
| CHST9 | -0.019145812 | 0.022941382 | 0.00379557 |
| RRBP1 | 0.054298238 | -0.048661465 | 0.005636773 |
| LINC01983 | -0.048697961 | 0.063621707 | 0.014923746 |
| GFM1 | 0.038592011 | -0.018263793 | 0.020328218 |
| FMNL3 | 0.015513598 | -0.012371288 | 0.00314231 |
| NECTIN3 | 0.022102439 | -0.016279635 | 0.005822804 |
| NPDC1 | -0.019277308 | 0.01965542 | 0.000378112 |
| ANKRD37 | 0.023178815 | -0.022847418 | 0.000331396 |
| RER1 | 0.060816714 | 0.032228377 | 0.093045092 |
| TLE6 | 0.019518968 | -0.012614595 | 0.006904373 |
| C2CD2 | 0.02377121 | -0.019187119 | 0.00458409 |
| VPS26B | 0.033024475 | -0.021100984 | 0.011923491 |
| TLR5 | 0.02902813 | -0.031872627 | -0.002844497 |
| FAF2 | 0.036568654 | -0.018866552 | 0.017702102 |
| C6orf62 | -0.053654683 | 0.083031119 | 0.029376436 |
| PRRC2C | 0.059928885 | -0.031856156 | 0.028072729 |
| SPINK4 | 0.009105789 | -0.007525552 | 0.001580237 |
| MLLT3 | 0.030265172 | -0.014558327 | 0.015706845 |
| PIK3C2B | 0.017023829 | -0.016545573 | 0.000478256 |
| AC058791.1 | 0.017467353 | -0.016188152 | 0.001279201 |
| FABP1 | -0.013691468 | 0.009563334 | -0.004128134 |
| COL12A1 | 0.012599652 | -0.008949233 | 0.003650419 |
| IRF1 | 0.021752787 | -0.017049932 | 0.004702855 |
| CAV2 | -0.040426496 | 0.020768069 | -0.019658427 |
| CITED2 | -0.047195639 | 0.082782786 | 0.035587147 |
| BOD1L1 | 0.050541801 | -0.039251347 | 0.011290454 |
| PPM1D | 0.019585897 | -0.010421053 | 0.009164844 |
| MAFG | 0.023575211 | -0.017259615 | 0.006315596 |
| COL4A3 | 0.018684046 | -0.018418919 | 0.000265127 |
| RETREG1 | 0.031960965 | -0.022260848 | 0.009700117 |
| FARP1 | -0.048315568 | 0.025576746 | -0.022738822 |
| CEMIP2 | -0.037231701 | 0.044949505 | 0.007717804 |
| UGT3A1 | 0.055811667 | -0.030976649 | 0.024835017 |
| SIL1 | 0.047331961 | 0.02226874 | 0.069600701 |

| | | | |
|---|---|---|---|
| PPM1G | 0.047717752 | -0.029455127 | 0.018262625 |
| SLC16A3 | 0.032974977 | -0.029854173 | 0.003120804 |
| RUFY2 | 0.033923981 | -0.022793845 | 0.011130136 |
| AVIL | 0.014916622 | -0.009404389 | 0.005512232 |
| APOL3 | 0.007780545 | -0.006381483 | 0.001399062 |
| STX12 | 0.038384936 | -0.019562923 | 0.018822013 |
| CAPN2 | -0.050063051 | 0.066907415 | 0.016844364 |
| MAN2B2 | 0.032355161 | -0.027581419 | 0.004773742 |
| EYA3 | -0.051022389 | 0.115027191 | 0.064004802 |
| PLA1A | 0.010656929 | -0.011334272 | -0.000677343 |
| SNHG7 | 0.049570588 | -0.04961199 | -4.14025E-05 |
| NME3 | 0.060353287 | 0.05082516 | 0.111178447 |
| FAM89B | 0.033330669 | -0.023703605 | 0.009627064 |
| ABRACL | 0.01814838 | -0.011334457 | 0.006813923 |
| PNN | 0.051590715 | -0.027274386 | 0.024316329 |
| FECH | -0.029411703 | 0.029162361 | -0.000249342 |
| PC | 0.014708434 | -0.015185129 | -0.000476694 |
| NUDT3 | -0.047766173 | 0.056986228 | 0.009220055 |
| WDR75 | 0.025563422 | -0.015592527 | 0.009970896 |
| GIPR | 0.010390982 | -0.01308541 | -0.002694427 |
| MBD2 | 0.04012536 | -0.031104191 | 0.009021169 |
| SP140L | 0.024404915 | -0.018043772 | 0.006361143 |
| TIFA | 0.027051917 | -0.015118355 | 0.011933562 |
| RPL21 | -0.067227138 | 0.078801631 | 0.011574494 |
| POLM | 0.024555102 | -0.019230414 | 0.005324688 |
| SCN3B | -0.02024252 | 0.012619823 | -0.007622698 |
| TEX2 | 0.027086257 | -0.01862699 | 0.008459267 |
| WWC1 | -0.045717442 | 0.034187136 | -0.011530306 |
| RAB17 | 0.048288865 | -0.044735615 | 0.00355325 |
| LINC02593 | -0.027108173 | 0.013554363 | -0.01355381 |
| AL512274.1 | 0.008021977 | -0.008929178 | -0.000907202 |
| C11orf53 | 0.007710547 | -0.007328269 | 0.000382279 |
| DMGDH | 0.02717019 | -0.01828871 | 0.00888148 |
| NASP | 0.047533275 | 0.05110891 | 0.098642185 |
| NFIC | -0.053434119 | 0.046668277 | -0.006765842 |
| EFCAB5 | 0.010371496 | -0.007353087 | 0.00301841 |
| ARL17B | 0.014403172 | -0.009577487 | 0.004825685 |
| SSBP3 | -0.039179263 | 0.05317299 | 0.013993728 |
| MT1F | -0.03884292 | 0.066316515 | 0.027473594 |
| RAB27A | 0.024675481 | -0.023833062 | 0.000842419 |
| HLA-DRB1 | 0.049483733 | 0.069845469 | 0.119329202 |
| EPB41L2 | 0.014444407 | -0.012483524 | 0.001960883 |
| CKS2 | 0.018250064 | -0.009373521 | 0.008876543 |
| ASIC2 | 0.009590096 | -0.010349877 | -0.000759781 |
| BICC1 | -0.054797782 | 0.12232166 | 0.067523878 |
| CLIP1 | 0.038248451 | -0.030046544 | 0.008201908 |
| EEA1 | 0.036208461 | -0.029576866 | 0.006631596 |

| | | | |
|---|---|---|---|
| MAGED2 | 0.05638825 | 0.039427808 | 0.095816059 |
| GPX2 | 0.007980303 | -0.005912848 | 0.002067455 |
| RGS19 | 0.015803694 | -0.01125161 | 0.004552084 |
| PPP1R14C | -0.023055313 | 0.04361599 | 0.020560677 |
| TFF3 | 0.008047015 | -0.007310506 | 0.000736509 |
| RPL13A | -0.061657908 | 0.072601891 | 0.010943983 |
| ATP6V0A2 | 0.022277232 | -0.012309249 | 0.009967982 |
| AC091151.1 | 0.005874378 | -0.005874378 | 1.24345E-14 |
| NXPE4 | 0.00597788 | -0.00597788 | 7.10543E-15 |
| NEIL1 | 0.026381052 | -0.014525393 | 0.011855659 |
| LRRC59 | 0.028396219 | -0.014771913 | 0.013624306 |
| CDK6 | 0.04639362 | 0.022568215 | 0.068961834 |
| ERBB4 | -0.035239036 | 0.049319781 | 0.014080745 |
| HACL1 | 0.023684485 | -0.012944836 | 0.010739649 |
| SUB1 | -0.062943724 | 0.107602555 | 0.044658831 |
| PITPNM2 | 0.019698994 | -0.01958282 | 0.000116174 |
| NPNT | -0.046887877 | 0.069903437 | 0.023015561 |
| C9orf50 | -0.018733775 | 0.018237245 | -0.00049653 |
| PLIN5 | -0.019578069 | 0.019576493 | -1.57609E-06 |
| C20orf194 | 0.033062248 | -0.022187548 | 0.0108747 |
| DIPK1B | -0.023943595 | 0.028186863 | 0.004243269 |
| HOGA1 | -0.020968831 | 0.020737678 | -0.000231152 |
| KCNJ1 | -0.062181356 | 0.280308094 | 0.218126738 |
| ENPP5 | 0.031841853 | -0.017419507 | 0.014422346 |
| RNF152 | 0.02921273 | -0.046012028 | -0.016799298 |
| AL589935.1 | 0.01599432 | -0.009733037 | 0.006261283 |
| DEPTOR | -0.027566094 | 0.042681844 | 0.01511575 |
| BAZ1A | 0.019409761 | -0.019280895 | 0.000128866 |
| TRAK1 | -0.040425048 | 0.040370829 | -5.42189E-05 |
| AP001062.1 | 0.017529946 | -0.015667804 | 0.001862142 |
| KLK10 | 0.005774981 | -0.005774981 | 5.32907E-15 |
| EXOC4 | 0.02982306 | -0.019681371 | 0.010141689 |
| ZNF106 | -0.049978811 | 0.066023043 | 0.016044232 |
| AIF1L | 0.061591754 | 0.064390448 | 0.125982202 |
| CYYR1 | 0.024885156 | -0.015770184 | 0.009114972 |
| ATXN1 | -0.034768081 | 0.027382564 | -0.007385516 |
| NT5C3A | 0.028290429 | -0.018251881 | 0.010038549 |
| DLGAP1-AS2 | 0.009258608 | -0.009952417 | -0.000693809 |
| EFHD1 | -0.048719867 | 0.035461176 | -0.013258691 |
| PLEKHH3 | 0.024443117 | -0.016279184 | 0.008163933 |
| PSMB9 | 0.016631473 | -0.018111595 | -0.001480122 |
| SLC9A6 | 0.024330091 | -0.016047904 | 0.008282187 |
| MGST3 | 0.067686297 | 0.078961121 | 0.146647417 |
| TMEM189 | 0.03063572 | -0.024993628 | 0.005642092 |
| TPT1 | -0.065384246 | 0.071786152 | 0.006401906 |
| ADAMTS5 | -0.031628436 | 0.037422292 | 0.005793856 |
| PRRG1 | 0.032575764 | -0.02997372 | 0.002602044 |

| | | | |
|---|---|---|---|
| CDIPT | 0.044535605 | -0.023548755 | 0.020986849 |
| HS3ST3B1 | 0.013081223 | -0.014273813 | -0.00119259 |
| EPS8L2 | 0.056997711 | -0.080963448 | -0.023965736 |
| SREBF1 | -0.034100603 | 0.038969748 | 0.004869145 |
| MTPAP | -0.024165672 | 0.022244506 | -0.001921166 |
| DYNLL1 | -0.065789175 | 0.144978228 | 0.079189053 |
| MT-ND6 | -0.056570183 | 0.109916743 | 0.053346561 |
| DAPK2 | 0.013998953 | -0.020682519 | -0.006683566 |
| ST14 | 0.044242817 | -0.021793077 | 0.02244974 |
| AGMAT | -0.009202944 | 0.00544417 | -0.003758775 |
| SMARCA1 | 0.041007661 | -0.023599927 | 0.017407733 |
| RPL4 | -0.059287297 | 0.055421189 | -0.003866108 |
| ME1 | 0.008908545 | -0.008343566 | 0.000564979 |
| SYNPO | -0.013595543 | 0.00685326 | -0.006742283 |
| SPECC1 | 0.018144197 | -0.019285015 | -0.001140818 |
| ANO10 | 0.035585905 | -0.024728578 | 0.010857326 |
| CYP39A1 | 0.018351998 | -0.011775183 | 0.006576815 |
| ASPA | 0.040609847 | -0.020269288 | 0.020340559 |
| CAVIN3 | -0.020589915 | 0.023962765 | 0.003372851 |
| MAML2 | -0.042000706 | 0.030677202 | -0.011323504 |
| ZDHHC9 | 0.031466249 | -0.020872541 | 0.010593709 |
| COPZ2 | 0.024263833 | -0.0188907 | 0.005373133 |
| SYNGR2 | 0.049546797 | -0.048080947 | 0.00146585 |
| TRAPPC10 | 0.027379486 | -0.020377093 | 0.007002392 |
| TMUB1 | 0.038841971 | -0.019589992 | 0.019251979 |
| RNF5 | 0.04363766 | -0.030214033 | 0.013423626 |
| COL14A1 | 0.00880523 | -0.007667875 | 0.001137354 |
| AC093673.1 | 0.019248981 | -0.011062038 | 0.008186943 |
| UGT2B7 | -0.016496417 | 0.009405494 | -0.007090923 |
| PEX1 | 0.034701882 | -0.018550255 | 0.016151627 |
| MIS18BP1 | 0.026485104 | -0.018737357 | 0.007747747 |
| TPBG | 0.022378485 | -0.032224496 | -0.009846011 |
| TMEM229B | 0.01213622 | -0.012528872 | -0.000392652 |
| IP6K2 | 0.041473552 | -0.02058955 | 0.020884002 |
| ZNF318 | 0.023156559 | -0.014122724 | 0.009033835 |
| SLC39A11 | 0.023572265 | -0.017325114 | 0.006247152 |
| AMOTL1 | -0.020994414 | 0.022788239 | 0.001793825 |
| PBX1 | -0.032614956 | 0.025296122 | -0.007318835 |
| OBSCN | -0.027123504 | 0.019629673 | -0.007493831 |
| C1QL4 | 0.009481629 | -0.013096321 | -0.003614692 |
| KCNJ2 | 0.007608745 | -0.008573687 | -0.000964942 |
| GUCY1B1 | -0.023538325 | 0.029866343 | 0.006328018 |
| GATA3-AS1 | 0.055332099 | 0.079617128 | 0.134949227 |
| BST2 | 0.034384638 | -0.019211039 | 0.015173599 |
| GMPR | 0.02162839 | -0.031707421 | -0.010079031 |
| MCF2L | 0.02618521 | -0.031537784 | -0.005352574 |
| SOX13 | -0.029266286 | 0.033426972 | 0.004160686 |

| | | | |
|---|---|---|---|
| HOXA10 | -0.026689592 | 0.030458323 | 0.003768731 |
| NPR1 | 0.019534819 | -0.029362284 | -0.009827465 |
| EIF1 | -0.061020757 | 0.113712384 | 0.052691626 |
| BPGM | -0.029769321 | 0.036622296 | 0.006852975 |
| ZNF141 | 0.031376673 | -0.021573022 | 0.009803651 |
| CNTNAP3B | -0.024435826 | 0.016208594 | -0.008227232 |
| ELP5 | 0.034042282 | -0.030612455 | 0.003429827 |
| AL590550.1 | 0.005175737 | -0.005175737 | 5.32907E-15 |
| SP100 | 0.035964903 | -0.025485694 | 0.010479209 |
| TMEM64 | -0.034434932 | 0.048793189 | 0.014358257 |
| SRM | 0.030506672 | -0.017731577 | 0.012775095 |
| OTUD1 | -0.029220089 | 0.02073574 | -0.008484349 |
| CFAP69 | -0.018425323 | 0.014662729 | -0.003762594 |
| ARID5B | -0.027872798 | 0.039820529 | 0.011947731 |
| METTL9 | -0.057425442 | 0.15666573 | 0.099240288 |
| AC003681.1 | -0.013585876 | 0.008483183 | -0.005102693 |
| DNASE2 | 0.033597189 | -0.026488169 | 0.00710902 |
| MDN1 | 0.029315234 | -0.020314372 | 0.009000862 |
| ABCB1 | -0.057879404 | 0.161960968 | 0.104081563 |
| AC073336.1 | 0.005056758 | -0.005056758 | 7.10543E-15 |
| MBOAT1 | -0.034008601 | 0.02838 | -0.005628601 |
| POLR2A | -0.044974802 | 0.057992524 | 0.013017721 |
| SPCS1 | 0.059901368 | 0.043302657 | 0.103204026 |
| CA5B | 0.015962075 | -0.011130132 | 0.004831943 |
| PATJ | 0.050794728 | -0.031579026 | 0.019215702 |
| AATF | 0.030886142 | -0.022764828 | 0.008121314 |
| IFNAR2 | 0.019681131 | -0.012439178 | 0.007241953 |
| SPRY1 | -0.029908844 | 0.038055385 | 0.008146541 |
| RPL37A | -0.059279554 | 0.072152701 | 0.012873147 |
| ALDH1L1 | -0.024733269 | 0.024168853 | -0.000564415 |
| IQCK | -0.041508271 | 0.053676595 | 0.012168324 |
| PTGFR | -0.034725995 | 0.1152823 | 0.080556306 |
| GIGYF2 | 0.039798595 | -0.029277611 | 0.010520984 |
| ZNF853 | 0.013817386 | -0.013063244 | 0.000754141 |
| AC111000.4 | 0.005197948 | -0.004951637 | 0.000246311 |
| DNMT1 | 0.026151139 | -0.013482158 | 0.012668981 |
| NDUFA1 | -0.059795361 | 0.11436793 | 0.054572569 |
| IGDCC3 | -0.018536364 | 0.013669704 | -0.004866659 |
| MARCH2 | 0.027269224 | -0.01898858 | 0.008280644 |
| IRF3 | 0.033832843 | -0.021197966 | 0.012634877 |
| KIF5A | -0.014399699 | 0.008218525 | -0.006181174 |
| AC099552.5 | 0.00658126 | -0.007232793 | -0.000651533 |
| CENPC | 0.037067415 | -0.025255028 | 0.011812387 |
| ZFP36L2 | -0.047788369 | 0.065518722 | 0.017730352 |
| AC006460.1 | 0.005582115 | -0.005975312 | -0.000393196 |
| KCP | 0.033923965 | -0.030043763 | 0.003880201 |
| C2orf74 | -0.031652739 | 0.035886949 | 0.004234211 |

| | | | |
|---|---|---|---|
| HOXD9 | 0.041713896 | 0.036836976 | 0.078550872 |
| AC048341.1 | 0.013428888 | -0.007015382 | 0.006413507 |
| AC104806.2 | 0.005073731 | -0.005073731 | 8.88178E-15 |
| AC015712.1 | 0.015972634 | -0.010749552 | 0.005223082 |
| TES | 0.034469174 | -0.029417721 | 0.005051453 |
| ABCG1 | 0.01545207 | -0.025671032 | -0.010218962 |
| TFAP2B | -0.029424019 | 0.046126571 | 0.016702552 |
| GNS | 0.050988299 | 0.030147933 | 0.081136232 |
| TGFBR3 | 0.027871086 | -0.04171948 | -0.013848393 |
| UPK3BL1 | 0.036529786 | 0.050006051 | 0.086535837 |
| KLF3 | 0.031971573 | -0.038636754 | -0.006665181 |
| ADAMTSL3 | -0.029539981 | 0.029201685 | -0.000338296 |
| LINC00963 | 0.041693073 | -0.03105089 | 0.010642184 |
| GRM3 | -0.009281282 | 0.007455691 | -0.001825591 |
| HOXD8 | 0.053845875 | 0.101734499 | 0.155580374 |
| ICOSLG | 0.018578003 | -0.013634918 | 0.004943085 |
| DNPH1 | -0.049771824 | 0.098945923 | 0.049174099 |
| SLC9A3-AS1 | -0.019342156 | 0.010602497 | -0.008739659 |
| VPS25 | 0.043863231 | 0.046916592 | 0.090779824 |
| CNTRL | -0.021010847 | 0.015760277 | -0.00525057 |
| CYB561D2.1 | -0.027983528 | 0.029445621 | 0.001462093 |
| GLDC | -0.047482568 | 0.112117245 | 0.064634678 |
| LLGL1 | 0.012053193 | -0.007525176 | 0.004528017 |
| HACD4 | 0.020768505 | -0.016451494 | 0.004317011 |
| BDP1 | 0.040316737 | -0.037886161 | 0.002430576 |
| DOLK | 0.020258337 | -0.012204146 | 0.008054191 |
| BTG3 | 0.050210954 | 0.058552894 | 0.108763848 |
| FHOD1 | 0.014148553 | -0.008432496 | 0.005716056 |
| EPCAM | 0.060150715 | 0.072459837 | 0.132610552 |
| RERG | -0.014781894 | 0.010543665 | -0.004238229 |
| TCF25 | 0.048595927 | -0.047087148 | 0.001508779 |
| C19orf33 | -0.036866602 | -0.019847805 | -0.056714407 |
| DYNC1LI2 | 0.042721309 | -0.037733609 | 0.004987701 |
| STRN | 0.035774642 | -0.034185082 | 0.00158956 |
| PLCB2 | 0.010066789 | -0.014772743 | -0.004705954 |
| WHAMM | 0.022718599 | -0.025657046 | -0.002938447 |
| MCPH1 | 0.024971742 | -0.020802125 | 0.004169617 |
| UTP18 | 0.024907717 | -0.014241372 | 0.010666345 |
| PCDH9 | -0.025679361 | 0.037179857 | 0.011500496 |
| TOR4A | 0.026270077 | -0.019984219 | 0.006285858 |
| WDR60 | 0.035040945 | -0.028218563 | 0.006822382 |
| ALDH18A1 | 0.026747569 | -0.018117813 | 0.008629757 |
| VAT1 | -0.029171831 | 0.016703125 | -0.012468706 |
| PSD3 | 0.013283193 | -0.013263172 | 2.00217E-05 |
| SKAP1 | 0.03160949 | -0.018256655 | 0.013352835 |
| CNTN5 | 0.005300118 | -0.005617061 | -0.000316943 |
| NPC2 | -0.053220332 | 0.113828769 | 0.060608438 |

| | | | |
|---|---|---|---|
| ADGRL2 | -0.038671642 | 0.078537246 | 0.039865603 |
| MYOF | 0.029115657 | -0.017351189 | 0.011764469 |
| ARID3A | -0.024834872 | 0.032982566 | 0.008147694 |
| MYRFL | 0.008901732 | -0.007948742 | 0.00095299 |
| PRKY | 0.013642504 | -0.026237528 | -0.012595024 |
| FLRT3 | 0.019737769 | -0.016964422 | 0.002773347 |
| XPA | -0.031222772 | 0.023402205 | -0.007820567 |
| BX890604.2 | -0.015581888 | 0.014139244 | -0.001442644 |
| TRIP10 | 0.028810018 | -0.022575693 | 0.006234325 |
| LAPTM4A | 0.058226871 | 0.076089017 | 0.134315888 |
| OAT | -0.039777304 | 0.074763262 | 0.034985958 |
| TGFB1 | 0.032636988 | -0.04378166 | -0.011144672 |
| EIF1B | -0.047235829 | 0.092667019 | 0.04543119 |
| AC006460.2 | 0.006198798 | -0.007046348 | -0.00084755 |
| C1orf56 | -0.036812864 | 0.024033463 | -0.012779401 |
| MCF2L-AS1 | 0.014376293 | -0.01036877 | 0.004007523 |
| HMGN2 | -0.053247605 | 0.083938909 | 0.030691305 |
| ZNF519 | 0.012208944 | -0.008135624 | 0.004073321 |
| BACH1 | -0.0313208 | 0.021571367 | -0.009749433 |
| RPS4X | -0.061764362 | 0.151080111 | 0.089315748 |
| CTSZ | 0.036646465 | -0.021946392 | 0.014700073 |
| B4GALNT3 | 0.028438536 | -0.023540413 | 0.004898122 |
| MXI1 | 0.030058215 | -0.019326177 | 0.010732038 |
| SHTN1 | 0.025942185 | -0.027662048 | -0.001719863 |
| ARID1A | -0.037645063 | 0.041896123 | 0.004251059 |
| OSTM1 | 0.026725646 | -0.020802439 | 0.005923207 |
| SPIRE1 | -0.040734735 | 0.070702763 | 0.029968028 |
| PBLD | 0.0345263 | -0.036016648 | -0.001490348 |
| HES1 | 0.055739393 | 0.048812084 | 0.104551477 |
| SLC25A30 | -0.033910649 | 0.055174564 | 0.021263915 |
| MAN2A2 | 0.024575839 | -0.025045833 | -0.000469994 |
| IRX6 | -0.015328016 | 0.012395599 | -0.002932417 |
| EOGT | 0.016466044 | -0.011102647 | 0.005363397 |
| CGN | 0.03330525 | -0.0350637 | -0.00175845 |
| CASC8 | 0.004329368 | -0.004056998 | 0.000272371 |
| MINDY2 | -0.043541051 | 0.049340898 | 0.005799847 |
| FAM210A | 0.023077866 | -0.020218591 | 0.002859275 |
| TYW1 | 0.021673798 | -0.017984838 | 0.00368896 |
| FLVCR1 | 0.019361346 | -0.014252495 | 0.005108851 |
| PLXND1 | 0.009108825 | -0.010077334 | -0.000968508 |
| ACSS1 | -0.027875382 | 0.031963082 | 0.0040877 |
| ATP6V1F | 0.058803365 | 0.082679725 | 0.14148309 |
| RESF1 | -0.033414269 | 0.034443248 | 0.001028979 |
| IQGAP2 | 0.041934982 | 0.069824241 | 0.111759223 |
| BAIAP2 | 0.022089375 | -0.035395233 | -0.013305858 |
| EPHA2 | 0.020813185 | -0.033102524 | -0.01228934 |
| KCNK13 | -0.026677078 | 0.030704944 | 0.004027867 |

| | | | |
|---|---|---|---|
| SOD1 | -0.057381926 | 0.106155736 | 0.04877381 |
| EPB41L3 | 0.026447879 | 0.035639674 | 0.062087553 |
| ZNF217 | -0.028825963 | 0.024218509 | -0.004607454 |
| MT1H | -0.010985241 | 0.006170354 | -0.004814886 |
| ANKRD36B | 0.015226125 | -0.013664387 | 0.001561738 |
| RPS28 | -0.055455278 | 0.0557959 | 0.000340622 |
| PABPC1 | -0.050936711 | 0.044375526 | -0.006561185 |
| SLC35B4 | -0.034760242 | 0.058601486 | 0.023841243 |
| DCAF4L1 | 0.00612779 | -0.006344711 | -0.000216921 |
| CAPN15 | 0.021798702 | -0.016521287 | 0.005277415 |
| FAN1 | 0.032482469 | -0.023622788 | 0.008859681 |
| CGNL1 | 0.036886853 | -0.031435083 | 0.00545177 |
| AC004918.1 | 0.012650864 | -0.012986836 | -0.000335972 |
| LINC01687 | 0.004871191 | -0.005230806 | -0.000359616 |
| SLC22A15 | 0.018931681 | -0.017186564 | 0.001745116 |
| DENND5B | 0.012810402 | -0.008976407 | 0.003833995 |
| VANGL2 | -0.01520241 | 0.014028082 | -0.001174328 |
| COL4A1 | -0.035961171 | 0.062673739 | 0.026712567 |
| LRRK1 | 0.006420179 | -0.006972564 | -0.000552385 |
| HNF4A | -0.008940456 | 0.006279389 | -0.002661067 |
| SULT1A1 | -0.020319224 | 0.035547336 | 0.015228112 |
| PACSIN3 | -0.014000774 | 0.007921315 | -0.006079459 |
| IVNS1ABP | -0.048622073 | 0.136249079 | 0.087627006 |
| SLC25A48 | 0.005858453 | -0.005052112 | 0.000806342 |
| CENPV | -0.026771607 | 0.044082337 | 0.01731073 |
| C9orf78 | 0.034819433 | -0.019721007 | 0.015098427 |
| RGCC | 0.013679796 | -0.018821284 | -0.005141487 |
| SYNJ2 | 0.011382383 | -0.009141109 | 0.002241275 |
| ADNP2 | -0.020923423 | 0.018739413 | -0.00218401 |
| ARF6 | -0.041792151 | 0.060233916 | 0.018441765 |
| AGO3 | 0.030675487 | -0.022688969 | 0.007986518 |
| DAZAP2 | -0.048359667 | 0.066794501 | 0.018434835 |
| FKBP11 | 0.016756069 | -0.012797667 | 0.003958402 |
| LRCH4 | 0.020649066 | -0.018342996 | 0.00230607 |
| GTF2H2 | -0.022100252 | 0.01476783 | -0.007332422 |
| ATP13A2 | 0.024713125 | -0.022942459 | 0.001770666 |
| CLDND2 | 0.0171043 | -0.013963724 | 0.003140576 |
| NUB1 | 0.03511054 | -0.027970531 | 0.007140009 |
| GSTK1 | 0.050384045 | 0.062301345 | 0.11268539 |
| DDR1 | 0.049656137 | 0.049878147 | 0.099534284 |
| SGSM1 | -0.015134452 | 0.011462054 | -0.003672398 |
| CBWD5 | 0.026517473 | -0.018068055 | 0.008449418 |
| RANBP9 | 0.026576179 | -0.014958485 | 0.011617694 |
| ABCC4 | 0.009246938 | -0.008998006 | 0.000248932 |
| ARHGEF16 | 0.022030513 | -0.012681672 | 0.009348841 |
| METRNL | 0.047952914 | 0.063729838 | 0.111682752 |
| KIF23 | 0.009036625 | -0.006479169 | 0.002557456 |

| | | | |
|---|---|---|---|
| NR3C2 | -0.032459921 | 0.032365758 | -9.4163E-05 |
| CARD9 | 0.007004151 | -0.005199271 | 0.00180488 |
| LINC00324 | 0.013090539 | -0.011666896 | 0.001423643 |
| FANCC | 0.015077614 | -0.011938001 | 0.003139613 |
| EPHA6 | 0.004557776 | -0.004293106 | 0.00026467 |
| PRKAB1 | -0.02916343 | 0.047954052 | 0.018790621 |
| F2R | 0.009460389 | -0.007489314 | 0.001971075 |
| AL356740.3 | 0.005199428 | -0.004861826 | 0.000337602 |
| PDE1B | 0.005246246 | -0.00558379 | -0.000337545 |
| ARAP1 | 0.022204921 | -0.015653406 | 0.006551514 |
| CAPN6 | 0.010676492 | -0.007282208 | 0.003394284 |
| SPTAN1 | 0.046732108 | -0.039689453 | 0.007042655 |
| SLC35B3 | 0.022991037 | -0.019773947 | 0.00321709 |
| RPL34 | -0.057024149 | 0.059610179 | 0.00258603 |
| SUN2 | 0.025551766 | -0.019906071 | 0.005645696 |
| LINC00310 | 0.005616678 | -0.005261615 | 0.000355064 |
| HOXB6 | 0.04053066 | 0.023846042 | 0.064376703 |
| RAD17 | 0.028666803 | -0.017264305 | 0.011402498 |
| DES | 0.003729225 | -0.003729225 | 7.10543E-15 |
| LRP1 | -0.022091333 | 0.017063501 | -0.005027832 |
| SLC22A17 | -0.036385636 | 0.103348092 | 0.066962456 |
| INAVA | -0.014642801 | 0.00868192 | -0.005960881 |
| CCM2 | 0.020294331 | -0.017124428 | 0.003169904 |
| KDM1A | 0.033728197 | -0.026032811 | 0.007695386 |
| CHDH | 0.018915865 | -0.019468129 | -0.000552265 |
| JDP2 | 0.022583777 | -0.043652821 | -0.021069044 |
| FAM118A | -0.019661862 | 0.024356982 | 0.004695119 |
| CCDC8 | -0.026863022 | 0.032796825 | 0.005933803 |
| IL18R1 | 0.007393137 | -0.008537517 | -0.00114438 |
| ATP2A3 | 0.015123797 | -0.027396518 | -0.012272722 |
| ZNF680 | 0.026864169 | -0.021221678 | 0.005642492 |
| PAK4 | -0.027801648 | 0.022395933 | -0.005405715 |
| AL031123.5 | 0.022310101 | -0.016903018 | 0.005407083 |
| SCPEP1 | 0.05286724 | 0.090101518 | 0.142968758 |
| ZBED6CL | 0.009067088 | -0.011616783 | -0.002549695 |
| UAP1L1 | -0.011384358 | 0.00812995 | -0.003254408 |
| SGMS1 | -0.037948153 | 0.023292016 | -0.014656137 |
| SLC9A3R2 | 0.016626304 | -0.019547496 | -0.002921192 |
| PRR15L | -0.019878521 | 0.016926675 | -0.002951846 |
| MBD4 | 0.032181908 | -0.023647487 | 0.008534421 |
| GCGR | -0.013593031 | 0.012444315 | -0.001148716 |
| NOTCH1 | -0.016866943 | 0.017433962 | 0.000567019 |
| PLS3 | -0.050555644 | 0.135828912 | 0.085273267 |
| CLCN5 | -0.036833176 | 0.034531207 | -0.002301969 |
| TSC22D1 | 0.054544485 | 0.057300186 | 0.111844671 |
| HIST1H1C | 0.032720504 | -0.046407956 | -0.013687452 |
| BTN2A2 | 0.017012437 | -0.011282468 | 0.00572997 |

| | | | |
|---|---|---|---|
| PPM1M | -0.014349683 | 0.013892988 | -0.000456695 |
| METRN | -0.03478918 | 0.091518 | 0.05672882 |
| GTF2H5 | -0.044199827 | 0.079819181 | 0.035619353 |
| ZNF316 | 0.02097198 | -0.019770299 | 0.001201681 |
| SNX29 | 0.033246721 | -0.027015813 | 0.006230907 |
| HLF | -0.023728307 | 0.049576309 | 0.025848002 |
| CDO1 | -0.009892173 | 0.007237808 | -0.002654365 |
| SIPA1 | 0.02651482 | -0.021255202 | 0.005259618 |
| TMEM150C | -0.018025704 | 0.017286126 | -0.000739578 |
| BCAS3 | 0.022082632 | -0.01534511 | 0.006737522 |
| SELENOT | 0.046010311 | 0.036983301 | 0.082993613 |
| EIF3C | 0.012215985 | -0.010229247 | 0.001986737 |
| EYA4 | 0.035617439 | -0.051122234 | -0.015504795 |
| HOXA5 | -0.017133112 | 0.010574296 | -0.006558816 |
| CDC25B | 0.014940571 | -0.010356311 | 0.004584259 |
| PLCG2 | -0.032651848 | 0.03681389 | 0.004162042 |
| PELI2 | -0.020357072 | 0.01821958 | -0.002137492 |
| TPD52 | -0.03927186 | 0.063567631 | 0.024295771 |
| GJB1 | 0.020638839 | -0.012719377 | 0.007919462 |
| TSPAN18 | -0.010730805 | 0.006599936 | -0.004130869 |
| FAM3C | 0.038063165 | -0.030510624 | 0.007552542 |
| C1orf115 | 0.026331066 | -0.030517513 | -0.004186448 |
| GJA8 | 0.003834808 | -0.003834808 | 8.88178E-15 |
| CTSS | 0.01691624 | -0.019529179 | -0.00261294 |
| ALDH4A1 | -0.043206252 | 0.107797432 | 0.06459118 |
| ATL3 | 0.034225204 | -0.049165111 | -0.014939907 |
| AL441992.1 | 0.013320063 | -0.010065701 | 0.003254362 |
| KCTD9 | 0.02328154 | -0.017009687 | 0.006271853 |
| ZNF433 | 0.020000987 | -0.013977211 | 0.006023775 |
| EHMT1 | 0.026483455 | -0.020437457 | 0.006045998 |
| RBM3 | -0.036908519 | 0.054582481 | 0.017673962 |
| ALS2CL | 0.009512694 | -0.007044097 | 0.002468596 |
| MEGF9 | -0.035196042 | 0.028942115 | -0.006253927 |
| PDLIM4 | -0.014354533 | 0.011650607 | -0.002703926 |
| COA3 | -0.052165336 | 0.095746838 | 0.043581502 |
| GARS-DT | -0.024641303 | 0.019115943 | -0.00552536 |
| TSPAN12 | -0.034218875 | 0.04336807 | 0.009149195 |
| NORAD | -0.048359826 | 0.098192271 | 0.049832445 |
| PACRG | -0.017897191 | 0.01641799 | -0.001479201 |
| SEPTIN11 | 0.044555259 | 0.05883761 | 0.10339287 |
| HOXA9 | -0.025192026 | 0.034199451 | 0.009007425 |
| LIPH | 0.036085723 | -0.057201834 | -0.021116111 |
| SLC6A13 | 0.022897298 | -0.025602429 | -0.002705131 |
| ADIRF-AS1 | -0.008998252 | 0.006551547 | -0.002446705 |
| LPCAT2 | 0.045739409 | 0.046845414 | 0.092584822 |
| HOOK3 | 0.03167485 | -0.01956875 | 0.0121061 |
| ZFAND2B | 0.025372504 | -0.015633013 | 0.009739492 |

| | | | |
|---|---|---|---|
| AHCYL1 | -0.040814462 | 0.036909437 | -0.003905025 |
| DDR2 | 0.01142898 | -0.010372734 | 0.001056246 |
| ZNF846 | 0.013161579 | -0.013007629 | 0.000153951 |
| NDUFC2 | 0.05205949 | 0.037411697 | 0.089471187 |
| GREM1 | 0.004972781 | -0.004972781 | 1.24345E-14 |
| CU638689.4 | -0.012911413 | 0.018377531 | 0.005466118 |
| MAST4 | 0.011881722 | -0.018985661 | -0.00710394 |
| HNRNPH1 | -0.052289891 | 0.082768527 | 0.030478636 |
| TNRC6C | -0.021724668 | 0.022907875 | 0.001183206 |
| USP15 | 0.027792231 | -0.017391255 | 0.010400976 |
| RPL22L1 | -0.030582909 | 0.034703625 | 0.004120716 |
| AL591501.1 | -0.007386975 | 0.007044325 | -0.000342649 |
| ZNF836 | 0.015410381 | -0.014892995 | 0.000517386 |
| TDRD6 | 0.012564955 | -0.014115181 | -0.001550226 |
| MAT1A | 0.006461494 | -0.006911169 | -0.000449676 |
| AL139275.2 | -0.012357853 | 0.012275879 | -8.19734E-05 |
| TUFM | 0.046320164 | 0.054717863 | 0.101038027 |
| FTH1 | -0.053533271 | 0.093535754 | 0.040002483 |
| H1F0 | 0.037686573 | -0.072944896 | -0.035258323 |
| TMEM99 | 0.025831701 | -0.015486462 | 0.01034524 |
| PRDX3 | -0.044231358 | 0.088792238 | 0.04456088 |
| TTC22 | -0.015000009 | 0.017106403 | 0.002106394 |
| EFR3B | 0.010355423 | -0.011631674 | -0.001276251 |
| IRAK1BP1 | -0.02141703 | 0.014631179 | -0.006785851 |
| TMEM87A | 0.037665185 | -0.028905795 | 0.00875939 |
| TBCA | -0.048871582 | 0.08053671 | 0.031665128 |
| ZNF350-AS1 | -0.017808493 | 0.027007716 | 0.009199223 |
| LINC02477 | 0.003621185 | -0.003621185 | 3.55271E-15 |
| TMEM139 | 0.037047308 | -0.065589721 | -0.028542413 |
| CCN3 | 0.030679299 | -0.036359392 | -0.005680093 |
| CCNO | -0.027604259 | 0.055590445 | 0.027986186 |
| LINC00543 | -0.007929013 | 0.00798327 | 5.4257E-05 |
| CRYBG3 | 0.03189552 | -0.035833394 | -0.003937874 |
| LINC00642 | 0.00353977 | -0.00353977 | 5.32907E-15 |
| ODF2L | 0.034923703 | -0.022281406 | 0.012642297 |
| RBM47 | -0.042540281 | 0.035033121 | -0.00750716 |
| TMEM258 | 0.049704196 | 0.032799721 | 0.082503918 |
| PCCA-DT | -0.021756748 | 0.030708734 | 0.008951985 |
| EGLN3 | 0.022475658 | -0.032320802 | -0.009845143 |
| SAMD4A | 0.01311352 | -0.017698359 | -0.004584839 |
| TRPV4 | 0.033979799 | -0.024892786 | 0.009087013 |
| UFL1 | 0.032693863 | -0.02935103 | 0.003342833 |
| USP43 | 0.009699558 | -0.009492514 | 0.000207044 |
| MAP2K1 | -0.025136712 | 0.023669039 | -0.001467673 |
| TMPRSS3 | 0.00869071 | -0.010971674 | -0.002280963 |
| ADAL | -0.02318029 | 0.015334986 | -0.007845303 |
| TMEM17 | -0.013967992 | 0.009646301 | -0.004321691 |

| | | | |
|---|---|---|---|
| EPHX2 | -0.028206115 | 0.03123453 | 0.003028414 |
| CELSR2 | 0.041032044 | -0.03861528 | 0.002416764 |
| OS9 | 0.041631288 | 0.028825064 | 0.070456352 |
| PNLDC1 | -0.008772733 | 0.011865134 | 0.003092402 |
| PKP3 | 0.018859998 | -0.01636899 | 0.002491008 |
| REPS2 | 0.025208577 | -0.026857101 | -0.001648525 |
| TSGA10 | 0.02478555 | -0.020105319 | 0.004680232 |
| RSRP1 | 0.046872858 | 0.043549405 | 0.090422263 |
| SLC6A8 | -0.013508255 | 0.008324546 | -0.005183709 |
| LAD1 | 0.022708472 | -0.024969191 | -0.002260719 |
| ALDH6A1 | -0.037022816 | 0.031587127 | -0.005435689 |
| TBC1D1 | -0.037751742 | 0.081650374 | 0.043898632 |
| CRYBB2 | 0.008489809 | -0.009859118 | -0.00136931 |
| CEP295 | 0.019260564 | -0.015841431 | 0.003419133 |
| TMEM205 | 0.044459924 | 0.070725868 | 0.115185792 |
| RHBDD3 | 0.018098063 | -0.012175321 | 0.005922742 |
| RGN | -0.026035876 | 0.031325358 | 0.005289483 |
| NEFL | -0.006185798 | 0.004314878 | -0.00187092 |
| BEND7 | 0.025902908 | -0.023959151 | 0.001943757 |
| FSTL4 | -0.019562494 | 0.024240602 | 0.004678108 |
| RPS11 | -0.048108284 | 0.053155158 | 0.005046874 |
| MT-ND5 | -0.034442104 | 0.070083484 | 0.03564138 |
| KDM5B | 0.030996839 | -0.030853949 | 0.00014289 |
| COX7B | -0.054059872 | 0.178882108 | 0.124822236 |
| CEP131 | -0.016624429 | 0.012443387 | -0.004181043 |
| RASSF5 | 0.00494199 | -0.005682418 | -0.000740428 |
| PCSK7 | 0.022514008 | -0.018922713 | 0.003591295 |
| WNT5A | 0.00358055 | -0.003353604 | 0.000226946 |
| ERCC5 | 0.024350139 | -0.023761004 | 0.000589135 |
| PPFIA3 | -0.009466499 | 0.005884464 | -0.003582035 |
| KCNE3 | -0.016697104 | 0.031771478 | 0.015074374 |
| LONRF2 | -0.015924989 | 0.019153344 | 0.003228355 |
| RPL6 | -0.050363113 | 0.058262362 | 0.007899249 |
| PSENEN | 0.038315415 | 0.034548789 | 0.072864204 |
| SETD7 | -0.017116409 | 0.01213025 | -0.004986158 |
| PDGFRL | 0.003982688 | -0.003310075 | 0.000672613 |
| PPIF | -0.027231347 | 0.038079459 | 0.010848113 |
| CPVL | -0.049106755 | 0.150110832 | 0.101004077 |
| FOXO3 | -0.027447365 | 0.022587734 | -0.004859631 |
| PVALB | -0.038760376 | 0.131018317 | 0.092257941 |
| AC084759.2 | 0.005761484 | -0.004673977 | 0.001087507 |
| DDX43 | 0.005677044 | -0.005699582 | -2.25381E-05 |
| KIF13B | 0.03743373 | -0.026032814 | 0.011400915 |
| ZNF670 | 0.009329475 | -0.005875916 | 0.003453559 |
| RPL11 | -0.052183486 | 0.078206614 | 0.026023128 |
| MRPL33 | -0.050873911 | 0.086291168 | 0.035417258 |
| MIGA1 | 0.033666907 | -0.029884048 | 0.003782859 |

| | | | |
|---|---|---|---|
| PSAT1 | -0.007776709 | 0.005344912 | -0.002431797 |
| ERVMER34-1 | 0.028508064 | 0.018912058 | 0.047420122 |
| B3GALT4 | 0.019342103 | -0.013069672 | 0.006272431 |
| POC1B-AS1 | 0.013022477 | -0.008705525 | 0.004316952 |
| RPL9 | -0.049057395 | 0.070345579 | 0.021288184 |
| STX2 | 0.025574422 | -0.025139018 | 0.000435404 |
| ABTB2 | 0.022683928 | 0.021522296 | 0.044206224 |
| TSPYL1 | -0.042303338 | 0.079698351 | 0.037395013 |
| AC023908.3 | 0.009749597 | -0.006151359 | 0.003598238 |
| SNTG2-AS1 | 0.003330015 | -0.003330015 | 5.32907E-15 |
| MT-ND2 | -0.033646676 | 0.035545395 | 0.00189872 |
| FNDC10 | -0.008462167 | 0.010187026 | 0.001724859 |
| FAM181B | -0.015874258 | 0.016320738 | 0.000446479 |
| SMKR1 | -0.014929019 | 0.014297713 | -0.000631306 |
| CLSTN2 | 0.005751958 | -0.00521906 | 0.000532898 |
| CCDC74B | -0.013528676 | 0.013561218 | 3.25427E-05 |
| SLC44A3 | 0.026009437 | -0.029089079 | -0.003079642 |
| PTTG1IP | 0.043950846 | -0.04657308 | -0.002622234 |
| ANXA9 | -0.030676929 | 0.029494605 | -0.001182324 |
| ALDH1B1 | -0.014423792 | 0.016660883 | 0.002237091 |
| SCML1 | -0.028193349 | 0.044260274 | 0.016066925 |
| MCF2L2 | 0.005373008 | -0.005195287 | 0.00017772 |
| PLA2G4A | 0.004122564 | -0.004705325 | -0.000582762 |
| CYP27A1 | -0.010049481 | 0.006764562 | -0.003284919 |
| MEDAG | 0.003415741 | -0.003415741 | 7.10543E-15 |
| USP11 | -0.033769873 | 0.056418926 | 0.022649053 |
| NDUFA9 | 0.033865995 | 0.02940538 | 0.063271375 |
| RPL15 | -0.048239207 | 0.089665563 | 0.041426356 |
| HSH2D | 0.006035626 | -0.006745417 | -0.000709791 |
| NUDT16 | -0.02769747 | 0.046990599 | 0.01929313 |
| SEPSECS-AS1 | 0.011648328 | -0.012699448 | -0.00105112 |
| AC022613.1 | -0.020497656 | 0.032029863 | 0.011532207 |
| TTTY10 | 0.009270497 | -0.01479357 | -0.005523073 |
| ABCA6 | 0.00573592 | -0.005486837 | 0.000249082 |
| GPCPD1 | -0.023853314 | 0.040446595 | 0.01659328 |
| NPR3 | -0.013510014 | 0.009798865 | -0.003711149 |
| EVC2 | 0.010589662 | -0.008628035 | 0.001961627 |
| CTDSPL | 0.032098057 | -0.026435468 | 0.005662589 |
| SFT2D2 | 0.031324739 | -0.027643555 | 0.003681185 |
| AL109615.4 | 0.006805513 | -0.005547279 | 0.001258234 |
| AC092683.1 | 0.012104672 | -0.011205975 | 0.000898697 |
| ZNF44 | -0.036441874 | 0.059307894 | 0.02286602 |
| PLEKHA6 | -0.022028566 | 0.019106858 | -0.002921708 |
| MT3 | -0.008611757 | 0.009424864 | 0.000813106 |
| CSNK2A2 | 0.02225275 | -0.016698327 | 0.005554423 |
| MUC20 | -0.041973106 | 0.072104641 | 0.030131534 |
| DDC | -0.005704877 | 0.003668804 | -0.002036073 |

| | | | |
|---|---|---|---|
| KCNJ3 | 0.032385309 | -0.024381798 | 0.008003512 |
| RPS3A | -0.050645228 | 0.061808318 | 0.01116309 |
| FLRT1 | -0.013630846 | 0.022702231 | 0.009071385 |
| ENAM | -0.014282927 | 0.009552027 | -0.0047309 |
| ESR2 | 0.003499512 | -0.003254428 | 0.000245084 |
| IL17RD | -0.006895377 | 0.004924489 | -0.001970888 |
| TRIB1 | -0.019116736 | 0.02311703 | 0.004000294 |
| ZMYND11 | -0.034442 | 0.059294786 | 0.024852786 |
| PRNP | 0.021657291 | -0.020232094 | 0.001425197 |
| MYRF | -0.020188155 | 0.015569413 | -0.004618742 |
| SYNRG | 0.027451736 | -0.032218403 | -0.004766667 |
| LRRIQ1 | -0.021788207 | 0.039127202 | 0.017338995 |
| MACROD1 | -0.026459654 | 0.036894834 | 0.010435181 |
| ECHS1 | 0.041357871 | 0.043734665 | 0.085092536 |
| AC103702.1 | 0.019366104 | -0.012479537 | 0.006886568 |
| EFS | -0.011539323 | 0.013444221 | 0.001904898 |
| SRGAP3 | 0.037689583 | -0.046523287 | -0.008833704 |
| OAS2 | 0.004390669 | -0.00325028 | 0.001140389 |
| DTNB | -0.018883171 | 0.016029523 | -0.002853649 |
| TBL1XR1 | -0.036347842 | 0.070104228 | 0.033756386 |
| AC016705.2 | -0.011982723 | 0.012797609 | 0.000814886 |
| PEG10 | 0.021839146 | -0.024342388 | -0.002503242 |
| BAMBI | -0.021773044 | 0.034696878 | 0.012923834 |
| YWHAH | 0.034852284 | 0.02798628 | 0.062838564 |
| KLHL5 | -0.023195163 | 0.035758277 | 0.012563114 |
| VSIG10L | 0.006663611 | -0.004969164 | 0.001694448 |
| ADAMTS15 | -0.008176878 | 0.009342739 | 0.001165861 |
| RAP1GAP2 | -0.018644731 | 0.026990914 | 0.008346182 |
| MOG | 0.003093655 | -0.003093655 | 0 |
| RPS24 | -0.045793699 | 0.034151604 | -0.011642096 |
| ATP5ME | -0.049340129 | 0.113584372 | 0.064244243 |
| ATP5PO | -0.045994465 | 0.101735191 | 0.055740726 |
| TUBB | 0.042108199 | 0.031091992 | 0.073200191 |
| PBX4 | -0.008455394 | 0.006282028 | -0.002173366 |
| EFR3A | 0.034541919 | 0.036423819 | 0.070965738 |
| PROM2 | -0.037288969 | 0.101970571 | 0.064681602 |
| RPL32 | -0.049187111 | 0.074959584 | 0.025772474 |
| CNTN3 | 0.013046988 | -0.016718938 | -0.003671951 |
| EMP2 | 0.030966531 | -0.038340088 | -0.007373557 |
| STK17A | 0.014355411 | -0.016891736 | -0.002536325 |
| FBXL16 | -0.02244028 | 0.024614273 | 0.002173993 |
| CYCS | -0.045324805 | 0.123159338 | 0.077834533 |
| SYK | 0.035705426 | -0.053963996 | -0.01825857 |
| CTSA | 0.033002483 | 0.036749006 | 0.069751489 |
| AL445205.1 | -0.006749916 | 0.004692309 | -0.002057607 |
| FGL2 | 0.048367382 | 0.038075829 | 0.086443211 |
| KCNMA1 | -0.005654006 | 0.004020713 | -0.001633293 |

| | | | |
|---|---|---|---|
| IMMP1L | -0.027510443 | 0.039284927 | 0.011774484 |
| OLFM2 | 0.004174693 | -0.003400047 | 0.000774646 |
| YPEL2 | -0.030085225 | 0.048590937 | 0.018505712 |
| SPRY4 | 0.004989279 | -0.004012133 | 0.000977145 |
| CXorf38 | 0.018308277 | -0.019407168 | -0.00109889 |
| AP1M2 | 0.034306101 | 0.029762985 | 0.064069086 |
| DCN | 0.005576502 | -0.004395582 | 0.00118092 |
| RPS3 | -0.048241927 | 0.061162984 | 0.012921057 |
| RPP21 | 0.032996655 | 0.02517858 | 0.058175235 |
| AC140725.1 | -0.006593353 | 0.004560476 | -0.002032877 |
| TRIM73 | 0.015357078 | -0.017444417 | -0.002087339 |
| GJA4 | 0.005893756 | -0.007407776 | -0.00151402 |
| PCBP3 | 0.009701264 | -0.012253928 | -0.002552664 |
| GOLM1 | -0.025132742 | 0.051018948 | 0.025886206 |
| FAM27C | -0.019614925 | 0.020751548 | 0.001136623 |
| TDRD12 | 0.003595138 | -0.003143185 | 0.000451953 |
| SEC61G | -0.045999199 | 0.041888079 | -0.00411112 |
| RTP4 | 0.006262724 | -0.006750515 | -0.000487791 |
| C2orf49 | 0.023611309 | -0.02278012 | 0.000831189 |
| CHST10 | -0.010221094 | 0.008721723 | -0.001499371 |
| PACC1 | 0.013672939 | -0.009276302 | 0.004396637 |
| EFNA5 | -0.020312186 | 0.021242012 | 0.000929826 |
| SLC37A1 | 0.011179223 | -0.009443895 | 0.001735327 |
| DTX4 | 0.008518622 | -0.005625655 | 0.002892967 |
| DHX16 | 0.018105051 | -0.012671125 | 0.005433927 |
| CALCA | 0.007249665 | -0.011112195 | -0.00386253 |
| HLA-DQB1 | 0.020278623 | -0.015600985 | 0.004677639 |
| PCNX2 | 0.016180561 | -0.012422571 | 0.00375799 |
| CLN5 | 0.039923585 | 0.043405009 | 0.083328594 |
| ATP6V1D | 0.035633659 | 0.047741028 | 0.083374687 |
| FAM153B | 0.004658163 | -0.004027414 | 0.000630749 |
| CDYL2 | 0.009935857 | -0.006864044 | 0.003071813 |
| PRODH2 | -0.005973138 | 0.006376846 | 0.000403708 |
| RALGAPA1 | 0.030249006 | -0.021844417 | 0.00840459 |
| ZNF124 | -0.016874013 | 0.019888286 | 0.003014273 |
| AC116317.1 | 0.005949814 | -0.008732858 | -0.002783044 |
| PGK1 | 0.043402063 | 0.050019233 | 0.093421296 |
| SNHG16 | -0.025482167 | 0.031925895 | 0.006443729 |
| PSD4 | 0.019004786 | -0.021456403 | -0.002451617 |
| PSMA1 | 0.035484196 | 0.026357397 | 0.061841593 |
| FAT1 | 0.038275738 | 0.028280659 | 0.066556397 |
| NFKBIL1 | 0.019649737 | -0.018866059 | 0.000783678 |
| BRD3OS | -0.018414007 | 0.022228999 | 0.003814991 |
| AC016831.5 | 0.012206885 | -0.014051457 | -0.001844572 |
| ZNF423 | 0.004791688 | -0.004971099 | -0.000179411 |
| NDUFB2 | 0.047113314 | 0.074252602 | 0.121365916 |
| CHST2 | -0.011980432 | 0.014740538 | 0.002760106 |

| | | | |
|---|---|---|---|
| AC026356.1 | -0.008735654 | 0.006034892 | -0.002700762 |
| RDH13 | 0.022943883 | -0.020189563 | 0.00275432 |
| ARL17A | 0.015825247 | -0.01234313 | 0.003482117 |
| NPFF | 0.007653129 | -0.005710693 | 0.001942436 |
| ZNF615 | -0.016405767 | 0.015447076 | -0.000958691 |
| FCGRT | -0.040221198 | 0.089299187 | 0.049077989 |
| FMO5 | -0.019410682 | 0.017821917 | -0.001588765 |
| OBSL1 | -0.032959093 | 0.055615611 | 0.022656518 |
| EEF1D | -0.043464339 | 0.054716312 | 0.011251972 |
| EIF6 | 0.035882472 | 0.032644333 | 0.068526804 |
| AP000866.2 | 0.013592811 | -0.009538254 | 0.004054557 |
| NDRG2 | -0.029932574 | 0.05536962 | 0.025437045 |
| PNMA8A | -0.019246448 | 0.025866429 | 0.006619981 |
| FOXRED2 | -0.01038769 | 0.007952345 | -0.002435345 |
| TPCN1 | 0.029310917 | -0.02830432 | 0.001006597 |
| TMEM181 | 0.018691253 | -0.016756618 | 0.001934635 |
| C11orf16 | 0.007079496 | -0.005414249 | 0.001665247 |
| RPL36A | -0.045773722 | 0.075941529 | 0.030167807 |
| NT5DC2 | -0.020605471 | 0.037846162 | 0.017240692 |
| KLRG2 | 0.003550339 | -0.003874368 | -0.00032403 |
| HNRNPDL | 0.040728617 | 0.030984466 | 0.071713083 |
| PTP4A2 | 0.043534822 | 0.061790329 | 0.105325151 |
| MRPL22 | 0.029954127 | 0.023907702 | 0.05386183 |
| ATF4 | 0.029288667 | -0.02680883 | 0.002479836 |
| PLEC | 0.029276889 | -0.024649029 | 0.004627859 |
| KIAA1841 | 0.008968853 | -0.008085047 | 0.000883807 |
| AC073172.1 | -0.023455605 | 0.052006333 | 0.028550728 |
| CTXN1 | -0.017856775 | 0.021504546 | 0.003647771 |
| PLCL1 | -0.019022786 | 0.032371266 | 0.01334848 |
| B3GNT9 | -0.013141145 | 0.0167408 | 0.003599654 |
| MXRA5 | -0.040111485 | 0.069675676 | 0.029564191 |
| MIR503HG | -0.007905418 | 0.006508106 | -0.001397312 |
| PDE6G | -0.012727207 | 0.026820804 | 0.014093597 |
| AP001059.2 | 0.005466619 | -0.00374367 | 0.00172295 |
| CDKN2D | 0.011455346 | -0.00810569 | 0.003349656 |
| VEPH1 | -0.033092529 | 0.034040574 | 0.000948045 |
| NOP53 | -0.034771907 | 0.028949037 | -0.005822869 |
| AC129507.4 | -0.024542556 | 0.105517633 | 0.080975077 |
| PCDHA12 | 0.006610971 | -0.007232266 | -0.000621296 |
| DHCR24 | -0.023225907 | 0.035310128 | 0.012084221 |
| LDLRAD4 | 0.010198047 | -0.013480219 | -0.003282172 |
| ODAM | 0.014111168 | -0.010957755 | 0.003153413 |
| RPL12 | -0.048792695 | 0.075320953 | 0.026528258 |
| ANKRD13D | 0.017051403 | -0.015170666 | 0.001880737 |
| CEP41 | -0.015709805 | 0.01369996 | -0.002009845 |
| HADHB | 0.038505145 | 0.036634223 | 0.075139368 |
| POLR1D | 0.032329762 | 0.022625105 | 0.054954868 |

| | | | |
|---|---|---|---|
| MSH6 | 0.020662529 | -0.014697414 | 0.005965115 |
| TFF2 | 0.004242603 | -0.004611107 | -0.000368505 |
| IDNK | -0.014304013 | 0.015081978 | 0.000777965 |
| MAT2A | -0.028384992 | 0.043221788 | 0.014836796 |
| GAREM1 | -0.026588234 | 0.060799845 | 0.03421161 |
| TBX19 | 0.006093695 | -0.007090231 | -0.000996536 |
| HOXB1 | 0.002876067 | -0.002876067 | 7.10543E-15 |
| JUND | 0.046359354 | 0.056335447 | 0.102694801 |
| MALAT1 | 0.05686352 | 0.057509998 | 0.114373518 |
| MYOM1 | 0.004567669 | -0.00393837 | 0.000629299 |
| CCNB1IP1 | -0.028057783 | 0.041352047 | 0.013294264 |
| CDCP1 | 0.016266964 | 0.013389326 | 0.02965629 |
| MORC3 | 0.019603831 | -0.019740548 | -0.000136717 |
| AIDA | 0.023153471 | -0.021380348 | 0.001773123 |
| CFD | -0.010382277 | 0.008337522 | -0.002044755 |
| SIX5 | -0.014800179 | 0.011468197 | -0.003331983 |
| TAP1 | 0.014823863 | -0.01470802 | 0.000115843 |
| VAMP2 | -0.039818888 | 0.042963597 | 0.003144709 |
| ADGRE5 | 0.011925102 | -0.011301365 | 0.000623737 |
| LINC02229 | 0.002746021 | -0.002746021 | 8.88178E-15 |
| SYBU | 0.008998229 | -0.007693883 | 0.001304346 |
| EMX2 | -0.042392508 | 0.107948512 | 0.065556004 |
| FCHO2 | 0.022776617 | -0.02422794 | -0.001451323 |
| PPP1R15A | -0.017292978 | 0.013120085 | -0.004172893 |
| ATP5MG | -0.047269063 | 0.131150541 | 0.083881478 |
| GPC1 | -0.022408727 | 0.019593708 | -0.002815018 |
| HSBP1 | -0.043060597 | 0.102806345 | 0.059745748 |
| RPA1 | -0.018137688 | 0.019855509 | 0.001717821 |
| CHCHD3 | 0.023828615 | -0.016399028 | 0.007429587 |
| ST3GAL4 | -0.022595323 | 0.031259341 | 0.008664018 |
| GATD3A | 0.011935395 | 0.024699767 | 0.036635162 |
| FER | 0.021911527 | -0.019049453 | 0.002862075 |
| HSPA5 | 0.04133233 | 0.077177219 | 0.11850955 |
| COL16A1 | 0.013327068 | -0.020420046 | -0.007092977 |
| GOLT1B | 0.019943693 | -0.02081069 | -0.000866997 |
| PSMB6 | 0.03914848 | 0.028603813 | 0.067752293 |
| SPATA22 | 0.00307316 | -0.00307316 | 7.10543E-15 |
| ZNRF3 | -0.016529349 | 0.017675762 | 0.001146413 |
| SLC25A23 | -0.028515113 | 0.042357658 | 0.013842545 |
| GALNT9 | 0.012770541 | 0.011199126 | 0.023969667 |
| AC007614.1 | 0.003359441 | -0.002859813 | 0.000499628 |
| FBXL4 | -0.023154469 | 0.028894256 | 0.005739787 |
| CCDC181 | -0.02085531 | 0.021679989 | 0.00082468 |
| ANPEP | -0.007180092 | 0.007306922 | 0.00012683 |
| EML4 | -0.029268804 | 0.030829384 | 0.001560581 |
| LARGE2 | -0.019034378 | 0.025311033 | 0.006276655 |
| FBLN1 | 0.004840019 | -0.003636311 | 0.001203708 |

| | | | |
|---|---|---|---|
| BX470102.2 | -0.005588027 | 0.003954794 | -0.001633233 |
| CHST11 | 0.003630132 | -0.002651491 | 0.000978641 |
| B3GALNT1 | -0.01599338 | 0.028753833 | 0.012760452 |
| TMC4 | 0.03140601 | 0.028758661 | 0.060164671 |
| COA6-AS1 | -0.018040541 | 0.019581989 | 0.001541448 |
| SUMO1 | 0.041516501 | 0.058799181 | 0.100315682 |
| EHHADH | -0.021477639 | 0.025806115 | 0.004328475 |
| PHLDA3 | -0.014086283 | 0.011518758 | -0.002567525 |
| CDH3 | -0.012992845 | 0.01293034 | -6.25049E-05 |
| KIF21A | 0.022095913 | -0.017944691 | 0.004151223 |
| UFM1 | -0.031382291 | 0.047012544 | 0.015630254 |
| CACNA2D1 | -0.020456149 | 0.014775341 | -0.005680808 |
| ATF7IP | -0.029441917 | 0.025520351 | -0.003921567 |
| RIDA | -0.021108646 | 0.030518389 | 0.009409743 |
| COX16 | -0.036273187 | 0.048743456 | 0.012470269 |
| MAP3K13 | -0.033193157 | 0.072217709 | 0.039024552 |
| LAGE3 | -0.023955469 | 0.036625004 | 0.012669535 |
| KLHL14 | -0.017552223 | 0.036599241 | 0.019047018 |
| SDHA | 0.034220785 | 0.043256142 | 0.077476927 |
| LINC01226 | 0.008710853 | -0.01276593 | -0.004055078 |
| TMEM38B | 0.02132756 | 0.024857655 | 0.046185215 |
| TMEM9B-AS1 | 0.006687005 | -0.006036844 | 0.000650161 |
| ESYT1 | 0.022210037 | -0.016483083 | 0.005726954 |
| ENO3 | 0.008964788 | -0.006549612 | 0.002415176 |
| LRP3 | -0.01180718 | 0.008627441 | -0.003179739 |
| FAM171A1 | -0.030594642 | 0.06256744 | 0.031972797 |
| C17orf100 | -0.00803174 | 0.006259587 | -0.001772153 |
| NDUFB11 | 0.043210886 | 0.058704973 | 0.101915859 |
| AC005476.2 | 0.005465801 | -0.004737389 | 0.000728412 |
| NAT8L | -0.020420738 | 0.043082689 | 0.022661952 |
| AC069120.1 | 0.006624152 | -0.007713418 | -0.001089265 |
| EVA1A | -0.013464536 | 0.023879199 | 0.010414662 |
| CCDC151 | -0.015555775 | 0.020265071 | 0.004709296 |
| RHOB | -0.029814179 | 0.034542314 | 0.004728135 |
| TMEM98 | -0.018632108 | 0.032424943 | 0.013792835 |
| SLC20A1 | -0.012395756 | 0.012678097 | 0.000282342 |
| MAGI2-AS3 | -0.022548637 | 0.052635002 | 0.030086366 |
| C1D | 0.031867238 | 0.030999478 | 0.062866716 |
| DEAF1 | 0.021349702 | -0.015857834 | 0.005491868 |
| PHF13 | 0.013284187 | -0.010055083 | 0.003229104 |
| CCL2 | 0.003994066 | -0.003468508 | 0.000525558 |
| SNRPN | 0.043291578 | 0.031662464 | 0.074954043 |
| GPR27 | -0.01500646 | 0.03132437 | 0.01631791 |
| GAL3ST3 | -0.014283434 | 0.020995394 | 0.00671196 |
| AC008549.2 | 0.002654277 | -0.002654277 | 8.88178E-15 |
| GPC3 | 0.016001273 | -0.024264252 | -0.00826298 |
| CMTM7 | -0.010206707 | 0.013326995 | 0.003120288 |

| | | | |
|---|---|---|---|
| ADTRP | -0.007877764 | -0.006755047 | -0.014632811 |
| DIPK1A | 0.006152193 | -0.005197471 | 0.000954722 |
| COX20 | 0.041794748 | 0.060114515 | 0.101909263 |
| SERPINA5 | 0.028943707 | 0.032076076 | 0.061019783 |
| MGP | 0.026948875 | 0.030225382 | 0.057174257 |
| HOXC4 | -0.01068157 | 0.010040624 | -0.000640946 |
| AC025271.4 | 0.004085814 | -0.003349857 | 0.000735958 |
| HMGN3 | -0.043004355 | 0.104105627 | 0.061101272 |
| SMAD1 | -0.029697785 | 0.039520316 | 0.009822532 |
| FRAS1 | 0.035929286 | 0.079250214 | 0.1151795 |
| PDE3A | -0.005698985 | 0.004719354 | -0.000979631 |
| LINC02471 | 0.002830257 | -0.002583797 | 0.000246459 |
| AL133453.1 | 0.014575879 | -0.016490598 | -0.001914719 |
| DDX5 | 0.04022893 | 0.054449433 | 0.094678363 |
| PLCD1 | -0.013993563 | 0.012524038 | -0.001469525 |
| ZDHHC16 | 0.018445658 | -0.015973345 | 0.002472313 |
| C1orf116 | -0.014983532 | 0.014923051 | -6.04807E-05 |
| LIPC | 0.011491084 | -0.013011501 | -0.001520417 |
| KCNJ10 | -0.013954322 | 0.014156504 | 0.000202181 |
| TMEM132A | 0.004880769 | -0.004743079 | 0.000137691 |
| DYNLL2 | -0.027486057 | 0.0315795 | 0.004093443 |
| FDPS | 0.027372123 | 0.028270854 | 0.055642977 |
| RPL22 | -0.043092157 | 0.053273978 | 0.010181821 |
| DLG2 | 0.014827693 | -0.019863816 | -0.005036123 |
| AC004540.2 | -0.029017882 | 0.083484658 | 0.054466776 |
| GJB3 | 0.003246386 | -0.002968086 | 0.0002783 |
| ZNF426 | -0.017091701 | 0.022645143 | 0.005553442 |
| GAS1 | -0.014182819 | 0.022757364 | 0.008574546 |
| PRR5 | 0.012090107 | -0.012507062 | -0.000416955 |
| ABLIM1 | -0.028221329 | 0.038581257 | 0.010359928 |
| GIPC2 | -0.010553976 | 0.012586423 | 0.002032448 |
| RAB38 | -0.01307633 | 0.015630488 | 0.002554158 |
| FYB2 | 0.029477688 | -0.030840313 | -0.001362624 |
| CYHR1 | 0.033924264 | 0.027072153 | 0.060996417 |
| CCDC144A | -0.010964329 | 0.008296052 | -0.002668277 |
| HIPK2 | 0.038246278 | -0.038196938 | 4.93404E-05 |
| NFKBID | 0.006262209 | -0.005378384 | 0.000883825 |
| SMOC2 | 0.003573165 | -0.003959662 | -0.000386498 |
| AC093912.1 | 0.010183893 | -0.008070998 | 0.002112896 |
| MAPKAPK3 | -0.021382295 | 0.040222356 | 0.018840061 |
| FMOD | 0.010199402 | -0.016865286 | -0.006665885 |
| RAB1B | -0.031748799 | 0.036630562 | 0.004881762 |
| MTIF2 | 0.021953699 | -0.019814544 | 0.002139155 |
| MIR193BHG | 0.003120457 | -0.002508463 | 0.000611993 |
| BCL9 | -0.014088857 | 0.015158077 | 0.00106922 |
| MON2 | 0.027905375 | -0.024422813 | 0.003482562 |
| DAB2 | -0.033184132 | -0.046954675 | -0.080138806 |

| | | | |
|---|---|---|---|
| ARHGAP29 | -0.035954114 | 0.062561395 | 0.026607281 |
| BMT2 | 0.015013159 | -0.012458867 | 0.002554292 |
| AC127537.1 | 0.003068736 | -0.002530554 | 0.000538182 |
| EMD | -0.023799167 | 0.017952564 | -0.005846602 |
| KLB | 0.004011391 | -0.004732232 | -0.00072084 |
| PSMA2 | 0.035916319 | 0.031981294 | 0.067897614 |
| SLC19A2 | -0.016438297 | 0.021464248 | 0.005025951 |
| CDC42SE1 | -0.02852486 | 0.028320077 | -0.000204782 |
| ACSM2A | -0.007423257 | 0.007662264 | 0.000239006 |
| TRAF3IP2 | -0.016960848 | 0.024801488 | 0.00784064 |
| WDR82 | -0.024726615 | 0.034829953 | 0.010103338 |
| ITLN2 | -0.006589554 | 0.006734995 | 0.000145441 |
| BRPF1 | -0.011710075 | 0.008951349 | -0.002758726 |
| ADAMTS9-AS2 | -0.024203378 | 0.0292239 | 0.005020522 |
| ATP8B2 | -0.004983727 | 0.004048176 | -0.000935551 |
| DNAJB6 | 0.036514207 | 0.038106202 | 0.074620408 |
| ITGA10 | 0.003652569 | -0.004001257 | -0.000348688 |
| SAMD12-AS1 | 0.00386382 | -0.003681372 | 0.000182448 |
| APIP | 0.026006941 | -0.020005887 | 0.006001054 |
| PRSS8 | -0.0303137 | 0.040922059 | 0.010608358 |
| BAZ1B | 0.024311495 | -0.0231578 | 0.001153694 |
| FNDC1 | 0.002731267 | -0.002523389 | 0.000207878 |
| SPIRE2 | -0.012824197 | 0.015643065 | 0.002818867 |
| MAP3K10 | 0.01060892 | -0.008825068 | 0.001783852 |
| HPF1 | -0.022028621 | 0.03748669 | 0.015458069 |
| BCL7A | -0.027455795 | 0.030222836 | 0.002767041 |
| SCX | 0.008718372 | -0.009570698 | -0.000852326 |
| RREB1 | -0.024553349 | 0.020194596 | -0.004358753 |
| NCAPD3 | 0.016328036 | -0.015433484 | 0.000894552 |
| ENC1 | -0.009602341 | 0.010431158 | 0.000828817 |
| SLC41A2 | 0.00569738 | -0.006916509 | -0.001219129 |
| CARD10 | -0.012378918 | 0.010779801 | -0.001599117 |
| CHCHD10 | -0.043367114 | 0.158096995 | 0.114729881 |
| ATG101 | 0.021915023 | -0.023734117 | -0.001819094 |
| STK26 | 0.005064958 | -0.005213036 | -0.000148078 |
| LINC01341 | 0.00806123 | -0.007553447 | 0.000507783 |
| CYP4V2 | 0.024389424 | -0.020058414 | 0.00433101 |
| OGFR | 0.019039824 | -0.018124609 | 0.000915215 |
| WFDC2 | -0.046706798 | 0.121377185 | 0.074670387 |
| LRRC3 | 0.003870587 | -0.003234469 | 0.000636118 |
| TEKT2 | -0.005791292 | 0.004577051 | -0.001214241 |
| CDC42EP5 | 0.006350247 | -0.006509709 | -0.000159461 |
| ANK3 | 0.038376004 | -0.044271005 | -0.005895002 |
| SLC16A2 | -0.012120129 | 0.014445837 | 0.002325709 |
| TMOD3 | 0.026354896 | -0.025825896 | 0.000529 |
| MTFR1L | -0.027465804 | 0.050335371 | 0.022869568 |
| SEPHS2 | -0.030970297 | 0.040354172 | 0.009383875 |

| | | | |
|---|---|---|---|
| TBX3 | 0.026944792 | -0.028892024 | -0.001947232 |
| CDK5RAP2 | 0.018947686 | -0.020991875 | -0.002044189 |
| USP9Y | 0.019679799 | -0.076738041 | -0.057058242 |
| CABCOCO1 | -0.011591772 | 0.030591182 | 0.018999411 |
| C17orf97 | -0.020363012 | 0.036361649 | 0.015998637 |
| ATP5MPL | -0.041224237 | 0.093038457 | 0.05181422 |
| C8orf76 | 0.015695937 | -0.014262981 | 0.001432957 |
| CHST12 | 0.017576112 | -0.015802294 | 0.001773818 |
| TMEM231 | -0.016649624 | 0.018670303 | 0.002020679 |
| RPS23 | -0.042789845 | 0.041271372 | -0.001518473 |
| VDR | 0.008406973 | -0.015368004 | -0.006961031 |
| AC246817.1 | -0.00772791 | 0.006781557 | -0.000946353 |
| N4BP2L2 | 0.04011901 | 0.057211645 | 0.097330655 |
| FRY-AS1 | 0.00673006 | -0.009236568 | -0.002506508 |
| STAC | -0.006472305 | 0.006200955 | -0.000271351 |
| MBD6 | -0.022344508 | 0.025278453 | 0.002933945 |
| LINC02038 | 0.002207154 | -0.002207154 | 1.77636E-15 |
| AC108025.1 | 0.002302909 | -0.002302909 | 5.32907E-15 |
| RORC | -0.016257533 | 0.012623573 | -0.00363396 |
| SRPRB | 0.023609162 | 0.018354618 | 0.04196378 |
| FOXO1 | -0.017891953 | 0.029516895 | 0.011624943 |
| FAM162A | 0.035331458 | 0.030106231 | 0.065437689 |
| GCNA | 0.0069915 | -0.007528687 | -0.000537187 |
| RPS14 | -0.041111839 | 0.070621263 | 0.029509424 |
| GPSM1 | -0.006849752 | 0.005586921 | -0.001262831 |
| PLAGL1 | -0.016384762 | 0.033814551 | 0.017429789 |
| SMIM10 | 0.007241159 | -0.009478374 | -0.002237215 |
| COPS4 | 0.022345028 | 0.016764253 | 0.039109281 |
| CLBA1 | -0.014279789 | 0.020262376 | 0.005982587 |
| CNPY3 | 0.019941848 | -0.018253242 | 0.001688606 |
| PLEKHD1 | 0.002858663 | -0.002569883 | 0.00028878 |
| POLA2 | 0.007979734 | -0.007980345 | -6.10528E-07 |
| IFITM1 | 0.005587623 | -0.00484412 | 0.000743503 |
| RPL41 | -0.038506969 | 0.055056359 | 0.016549391 |
| ST3GAL5 | -0.008681676 | 0.02102296 | 0.012341284 |
| FAM171B | 0.005523998 | -0.005608857 | -8.48588E-05 |
| GADD45A | -0.021016312 | 0.04467088 | 0.023654568 |
| PWWP3A | -0.024096303 | 0.040790353 | 0.01669405 |
| NPTXR | 0.004849937 | -0.006401613 | -0.001551676 |
| MXRA7 | 0.027194966 | 0.023325408 | 0.050520374 |
| SSBP2 | 0.022305189 | -0.017051176 | 0.005254013 |
| PRKN | -0.017449416 | 0.033302263 | 0.015852847 |
| IGFBP5 | -0.024356508 | -0.021384581 | -0.045741089 |
| CDIP1 | -0.018213879 | 0.023608536 | 0.005394657 |
| EVC | 0.01473874 | -0.012264419 | 0.002474322 |
| KMT2C | 0.030638476 | -0.035769378 | -0.005130902 |
| SLCO4C1 | 0.019834485 | -0.015429003 | 0.004405483 |

| | | | |
|---|---|---|---|
| TMEM62 | 0.013175334 | -0.013749925 | -0.000574591 |
| UGT2B11 | 0.002608838 | -0.00295481 | -0.000345972 |
| KIFC2 | 0.0180152 | -0.015857518 | 0.002157683 |
| DECR1 | -0.033887086 | 0.089513589 | 0.055626503 |
| CDKN1B | -0.022541547 | 0.024314847 | 0.0017733 |
| ANGPT2 | 0.005869642 | -0.005582521 | 0.000287121 |
| ARHGAP6 | 0.017240812 | -0.020787952 | -0.00354714 |
| SPATA5 | 0.01103573 | -0.009690293 | 0.001345437 |
| LINC01836 | -0.013832011 | -0.017500211 | -0.031332222 |
| TBC1D8-AS1 | -0.005580749 | 0.004442731 | -0.001138018 |
| SP4 | -0.012955887 | 0.014099687 | 0.0011438 |
| REC8 | -0.01415739 | 0.015528122 | 0.001370732 |
| TRMT44 | 0.011207299 | -0.010206994 | 0.001000306 |
| APH1A | 0.032188523 | 0.027292109 | 0.059480632 |
| PIK3R2 | -0.019822459 | 0.022638809 | 0.00281635 |
| SLC46A1 | -0.013952618 | 0.016562348 | 0.002609731 |
| CLIC1 | 0.038401342 | 0.040910138 | 0.07931148 |
| PRTFDC1 | -0.014576249 | 0.030385255 | 0.015809006 |
| DRC3 | -0.010496778 | 0.008863446 | -0.001633332 |
| AF233439.1 | -0.007946014 | 0.008006991 | 6.09778E-05 |
| BLOC1S5 | -0.023809801 | 0.019572779 | -0.004237022 |
| ZNF710-AS1 | -0.016511594 | 0.028064494 | 0.011552901 |
| C9orf24 | -0.014481856 | 0.031230613 | 0.016748757 |
| ATP5PF | -0.042086078 | 0.124454807 | 0.082368729 |
| TFRC | 0.02264626 | 0.033951707 | 0.056597967 |
| TEAD1 | 0.027237812 | 0.024802791 | 0.052040603 |
| MEF2A | -0.027620525 | 0.028621066 | 0.001000541 |
| RPL10 | -0.04120625 | 0.04398933 | 0.00278308 |
| SNRPB2 | -0.028028444 | 0.050516125 | 0.022487681 |
| KLF9 | -0.017655652 | 0.023713007 | 0.006057355 |
| RGS5 | 0.007487009 | -0.007502841 | -1.58318E-05 |
| WSB1 | -0.041648172 | 0.172315601 | 0.130667429 |
| PALLD | -0.024592774 | 0.052118816 | 0.027526042 |
| PAG1 | -0.011476296 | 0.020083764 | 0.008607468 |
| BRD7 | 0.026676933 | -0.03122789 | -0.004550956 |
| PARD3B | 0.017196727 | -0.015281123 | 0.001915604 |
| ARC | 0.002380942 | -0.002160124 | 0.000220818 |
| CTSD | -0.038200711 | 0.091812337 | 0.053611626 |
| GALC | 0.016403007 | -0.013927259 | 0.002475748 |
| HYAL1 | -0.015373515 | 0.048231789 | 0.032858274 |
| PIK3C2G | 0.038855249 | 0.078871151 | 0.1177264 |
| PDGFA | -0.012129737 | 0.011507617 | -0.00062212 |
| GOT1 | 0.026942134 | 0.030997043 | 0.057939177 |
| AC103810.2 | 0.004696372 | -0.003904143 | 0.000792229 |
| FAH | -0.011476139 | 0.009153975 | -0.002322165 |
| SLC22A2 | -0.012278715 | 0.025658084 | 0.013379369 |
| TAB2 | -0.021203756 | 0.03195011 | 0.010746354 |

| | | | |
|---|---|---|---|
| UQCRH | -0.042217758 | 0.07761499 | 0.035397232 |
| PLEKHB2 | -0.029561243 | 0.046588476 | 0.017027232 |
| TAX1BP1 | 0.033916497 | 0.02719206 | 0.061108558 |
| PICALM | -0.027892933 | 0.043604455 | 0.015711522 |
| STYK1 | 0.004796346 | -0.003827296 | 0.00096905 |
| SREK1IP1 | 0.022847626 | -0.032745082 | -0.009897456 |
| AC015712.2 | 0.002111246 | -0.001865937 | 0.000245309 |
| ST13 | -0.034889125 | 0.043960771 | 0.009071646 |
| JCAD | 0.003143465 | -0.003168556 | -2.50907E-05 |
| ATP6AP2 | 0.035962847 | 0.042368908 | 0.078331755 |
| NAA38 | 0.036312949 | 0.034281471 | 0.07059442 |
| FAM87A | 0.002158886 | -0.002158886 | 8.88178E-15 |
| SERTAD3 | 0.021343358 | 0.020193265 | 0.041536622 |
| AC093898.1 | 0.002083896 | -0.002083896 | 8.88178E-15 |
| TLE3 | -0.02464936 | 0.046958552 | 0.022309192 |
| RPL27 | -0.038653882 | 0.083005712 | 0.044351831 |
| TMEM184C | 0.018959751 | 0.032342245 | 0.051301996 |
| NOX4 | -0.008940471 | 0.014564271 | 0.0056238 |
| LERFS | -0.008095294 | 0.011808657 | 0.003713363 |
| PIK3R1 | -0.020631479 | 0.023920683 | 0.003289204 |
| HMGB2 | -0.027412845 | 0.072218179 | 0.044805334 |
| COLCA1 | -0.025195514 | 0.053638192 | 0.028442679 |
| SPEF2 | -0.017741996 | 0.026910633 | 0.009168637 |
| UBXN10 | -0.009606857 | 0.014903592 | 0.005296735 |
| EMX1 | 0.037143036 | 0.101013386 | 0.138156422 |
| ATAD2 | 0.010284567 | -0.011148658 | -0.000864091 |
| NEXMIF | -0.013402152 | 0.024928138 | 0.011525985 |
| AL357033.1 | 0.00230797 | -0.002009829 | 0.000298142 |
| ACOT11 | -0.016812198 | 0.023521981 | 0.006709783 |
| LRRC34 | -0.011722534 | 0.014086209 | 0.002363674 |
| PPL | 0.022638285 | -0.034844266 | -0.012205981 |
| CTSF | -0.030304075 | 0.099034445 | 0.068730371 |
| RAF1 | 0.021873673 | -0.018038374 | 0.003835299 |
| SF3B1 | 0.03382108 | 0.041334461 | 0.075155541 |
| ATP6V0D1 | 0.030771351 | 0.026772647 | 0.057543998 |
| HMGA1 | -0.019801019 | 0.019363655 | -0.000437364 |
| PCBP1 | -0.033430717 | 0.066636525 | 0.033205807 |
| MFSD9 | 0.011819068 | -0.010337855 | 0.001481214 |
| PCDH17 | -0.01332202 | 0.02324625 | 0.00992423 |
| RIPOR3 | 0.005280777 | -0.004717912 | 0.000562865 |
| AL139002.1 | 0.002078723 | -0.002078723 | 5.32907E-15 |
| ZNF556 | -0.006005957 | 0.004913192 | -0.001092765 |
| LARP6 | 0.022340427 | 0.03091278 | 0.053253206 |
| WIZ | -0.015126262 | 0.01391575 | -0.001210512 |
| ZNF91 | 0.029967894 | -0.038191671 | -0.008223777 |
| PECAM1 | 0.006139027 | -0.006157647 | -1.86195E-05 |
| ATP5MF | 0.037985031 | 0.079757475 | 0.117742505 |

| | | | |
|---|---|---|---|
| ZNF800 | 0.018785566 | -0.018788956 | -3.38933E-06 |
| NOP14 | 0.013343049 | -0.011444539 | 0.00189851 |
| VRK2 | 0.011744856 | -0.009999432 | 0.001745424 |
| P3H3 | -0.009751013 | 0.011295412 | 0.001544399 |
| ZNF844 | -0.016569602 | 0.020361826 | 0.003792225 |
| FOXP1 | -0.023931278 | 0.025592837 | 0.001661559 |
| NMT1 | 0.023854174 | -0.022076388 | 0.001777786 |
| CYP4F11 | 0.005897502 | -0.006721992 | -0.00082449 |
| SOSTDC1 | -0.00610729 | 0.005770201 | -0.000337089 |
| CRCP | 0.016495836 | -0.016925467 | -0.000429631 |
| PCBP4 | -0.01278146 | 0.015410371 | 0.002628911 |
| AC231981.1 | 0.008894995 | -0.007364364 | 0.001530631 |
| KCTD1 | -0.017689511 | 0.027396636 | 0.009707125 |
| GPI | -0.030426455 | 0.087283922 | 0.056857467 |
| DCAF7 | -0.02485147 | 0.023753391 | -0.001098079 |
| PTGR2 | -0.020226103 | 0.035092511 | 0.014866408 |
| HERPUD1 | 0.029923682 | 0.024614637 | 0.054538319 |
| C10orf82 | -0.006666283 | 0.008203972 | 0.001537689 |
| GBP2 | 0.007078775 | -0.010697412 | -0.003618637 |
| NAAA | -0.016807376 | 0.0218705 | 0.005063124 |
| AC008467.1 | 0.004726159 | -0.006446735 | -0.001720576 |
| RHBDL2 | 0.005569175 | -0.005213984 | 0.000355191 |
| ASAP3 | -0.011772722 | 0.012973214 | 0.001200492 |
| ATP2B4 | -0.016585161 | 0.02961967 | 0.013034509 |
| ZNF549 | -0.009095866 | 0.007938989 | -0.001156877 |
| ANKRD36 | 0.013509169 | -0.013128527 | 0.000380641 |
| PWRN1 | 0.029475709 | -0.04191841 | -0.012442701 |
| NNT-AS1 | -0.018092475 | 0.021416014 | 0.003323538 |
| HNRNPAB | 0.023856434 | -0.026736955 | -0.002880522 |
| PLSCR2 | 0.008370923 | -0.007902623 | 0.0004683 |
| CCDC88C | -0.019785155 | 0.017479384 | -0.002305771 |
| LTBP3 | 0.031103778 | 0.030574291 | 0.061678069 |
| CSNK1A1 | 0.030872166 | 0.030904418 | 0.061776584 |
| KIAA0355 | -0.01982066 | 0.016475007 | -0.003345654 |
| CAPZB | 0.030114705 | 0.026862147 | 0.056976851 |
| AC010329.1 | -0.019102443 | 0.03557027 | 0.016467827 |
| DDX52 | 0.021012523 | -0.017758376 | 0.003254147 |
| TBL1X | 0.022889514 | -0.038771502 | -0.015881988 |
| UBASH3B | 0.002168404 | -0.002168404 | 8.88178E-15 |
| MYO1B | -0.030266961 | 0.12502043 | 0.094753468 |
| MCFD2 | 0.027054906 | 0.030687193 | 0.057742099 |
| LINC00504 | 0.00318402 | -0.002872411 | 0.000311609 |
| AC007566.1 | 0.008842729 | -0.009913046 | -0.001070317 |
| GLYAT | -0.004392527 | 0.004636729 | 0.000244202 |
| RASSF6 | -0.014691867 | 0.023158876 | 0.00846701 |
| CLDN1 | 0.005478813 | -0.008271395 | -0.002792582 |
| C4orf19 | -0.018343458 | 0.02037529 | 0.002031832 |

| | | | |
|---|---|---|---|
| EQTN | 0.002228469 | -0.001877967 | 0.000350502 |
| CTDSP2 | -0.025386324 | 0.046946152 | 0.021559828 |
| MFHAS1 | -0.019312924 | 0.026461917 | 0.007148993 |
| BLOC1S6 | -0.018542228 | 0.024463908 | 0.005921679 |
| ZDHHC2 | 0.027989235 | 0.027455922 | 0.055445157 |
| PRR35 | -0.032427064 | 0.12935461 | 0.096927546 |
| CEP350 | 0.025238078 | -0.022544084 | 0.002693994 |
| P4HA2 | 0.00965331 | -0.012900065 | -0.003246755 |
| PCDH11Y | 0.005730849 | -0.011950973 | -0.006220124 |
| CITED4 | -0.014217501 | 0.025823751 | 0.01160625 |
| ZNF772 | -0.010510656 | 0.014163741 | 0.003653086 |
| AC104461.1 | 0.002849382 | -0.002647996 | 0.000201386 |
| PRDM5 | 0.012606013 | -0.013619567 | -0.001013554 |
| MIB1 | -0.021023074 | 0.038483798 | 0.017460724 |
| CAMTA2 | 0.014974673 | -0.01462324 | 0.000351433 |
| RABGAP1L-IT1 | 0.002135025 | -0.001895479 | 0.000239546 |
| PARP6 | 0.013507367 | -0.011380911 | 0.002126456 |
| ZNF304 | -0.008928964 | 0.008645328 | -0.000283637 |
| PCAT1 | 0.007825389 | -0.009576408 | -0.001751019 |
| AC145207.5 | -0.006924136 | 0.00615936 | -0.000764775 |
| KIF22 | 0.017924085 | 0.015505436 | 0.033429521 |
| MTCH1 | -0.029856338 | 0.06260106 | 0.032744721 |
| FAT3 | 0.00241703 | -0.002594001 | -0.000176971 |
| DUSP15 | -0.019469522 | 0.045837975 | 0.026368453 |
| NRBF2 | 0.012351016 | -0.014301561 | -0.001950545 |
| LINC01547 | -0.005229014 | 0.005240136 | 1.11221E-05 |
| AL359397.2 | -0.005665032 | 0.005976668 | 0.000311636 |
| AC002070.1 | 0.003841772 | -0.003775768 | 6.60039E-05 |
| RAB11FIP5 | -0.017339677 | 0.032270837 | 0.01493116 |
| EVA1C | 0.008280483 | -0.008950779 | -0.000670296 |
| SNX27 | 0.021429937 | -0.022235762 | -0.000805825 |
| PLEKHA4 | -0.00720894 | 0.008338863 | 0.001129923 |
| RXRA | -0.017125827 | 0.02063201 | 0.003506183 |
| SMIM6 | -0.0201042 | 0.038949332 | 0.018845132 |
| AGRN | 0.032511872 | -0.029153583 | 0.003358288 |
| SLC25A24 | 0.018157042 | -0.018077795 | 7.92472E-05 |
| USP19 | -0.016700106 | 0.020885569 | 0.004185464 |
| SNHG5 | 0.022947875 | -0.022768551 | 0.000179324 |
| BAIAP2L1 | 0.011031691 | -0.015468457 | -0.004436766 |
| CWF19L2 | 0.022333587 | -0.032434345 | -0.010100758 |
| SALL1 | -0.009717652 | 0.013829651 | 0.004111999 |
| RNF10 | -0.027675933 | 0.058282784 | 0.030606851 |
| MARK1 | -0.01002819 | 0.00949258 | -0.00053561 |
| PABPC4L | -0.006545076 | 0.006846895 | 0.000301818 |
| MICOS10 | -0.037181174 | 0.112985781 | 0.075804606 |
| PRKAR1B | -0.012877745 | 0.012767789 | -0.000109956 |
| ESD | 0.029512855 | 0.040240511 | 0.069753366 |

| | | | |
|---|---|---|---|
| NUCKS1 | -0.034585412 | 0.072458162 | 0.03787275 |
| AC012358.3 | 0.007631007 | -0.008063814 | -0.000432807 |
| CABP4 | 0.003891527 | -0.004441296 | -0.000549769 |
| BLACAT1 | 0.001764095 | -0.001764095 | 7.10543E-15 |
| CISH | -0.012836369 | 0.027328893 | 0.014492524 |
| CINP | -0.015504067 | 0.026329489 | 0.010825423 |
| RPL38 | -0.035250076 | 0.05316965 | 0.017919574 |
| RBFA | 0.019750469 | -0.023185124 | -0.003434655 |
| AC092140.1 | 0.004790719 | -0.004417337 | 0.000373382 |
| HIF1A-AS3 | 0.002928674 | -0.00355624 | -0.000627567 |
| RPL35A | -0.037959791 | 0.065400672 | 0.02744088 |
| PLEKHN1 | 0.001902495 | -0.001902495 | 3.55271E-15 |
| PYROXD2 | 0.00795108 | -0.007680371 | 0.000270709 |
| PIGB | 0.009707777 | -0.009696463 | 1.13139E-05 |
| SMIM4 | -0.024475636 | 0.051052797 | 0.026577161 |
| ALKBH5 | -0.021262123 | 0.021007299 | -0.000254824 |
| FAT4 | -0.0146496 | 0.028005503 | 0.013355903 |
| NUS1 | -0.021719059 | 0.058285785 | 0.036566726 |
| CSNK2B | -0.027175026 | 0.033214731 | 0.006039705 |
| C12orf57 | -0.030568359 | 0.064217649 | 0.03364929 |
| CBX6 | -0.02577634 | 0.03867545 | 0.01289911 |
| PMM1 | -0.019954196 | 0.041158828 | 0.021204633 |
| ACTG1 | -0.032810796 | 0.09608702 | 0.063276224 |
| CCSER1 | -0.017782348 | 0.03928007 | 0.021497722 |
| SORD | -0.018005892 | 0.042371173 | 0.024365281 |
| LINC01138 | 0.007635035 | -0.008383312 | -0.000748276 |
| TRIM5 | 0.014446987 | -0.019530481 | -0.005083494 |
| C2orf88 | -0.005194923 | 0.005306202 | 0.000111279 |
| SLCO1A2 | 0.001696051 | -0.001696051 | 5.32907E-15 |
| SLCO2A1 | -0.006084847 | 0.006149601 | 6.47535E-05 |
| SMG1 | 0.025527061 | -0.032651332 | -0.007124271 |
| COBL | -0.015538023 | 0.014757315 | -0.000780708 |
| ATP5F1A | -0.03323369 | 0.096295613 | 0.063061923 |
| EIF1AX | -0.030092245 | 0.081045373 | 0.050953129 |
| AK3 | -0.025400179 | 0.040374772 | 0.014974593 |
| PAX8-AS1 | 0.015374496 | -0.022602912 | -0.007228416 |
| HSD17B8 | -0.020213335 | 0.044651707 | 0.024438373 |
| KANSL1-AS1 | -0.011640527 | 0.011496424 | -0.000144103 |
| DPY19L2 | 0.023592598 | 0.037525711 | 0.06111831 |
| PRTG | -0.015790736 | 0.040074211 | 0.024283475 |
| YBX1 | -0.035418099 | 0.055295669 | 0.01987757 |
| TMEM273 | 0.002703911 | -0.002716145 | -1.22336E-05 |
| AL513318.2 | -0.002946318 | 0.002928918 | -1.73994E-05 |
| LYRM2 | -0.023132729 | 0.025261252 | 0.002128524 |
| SNORC | 0.003051945 | -0.00298832 | 6.3625E-05 |
| COX8A | -0.035792844 | 0.088590935 | 0.052798091 |
| BHMT2 | 0.026272471 | 0.035879255 | 0.062151726 |

| | | | |
|---|---|---|---|
| SH3GL1 | 0.012840709 | -0.012544014 | 0.000296695 |
| AC107068.1 | -0.008164093 | 0.007993674 | -0.000170419 |
| GLYATL1 | -0.006938077 | 0.009260712 | 0.002322635 |
| LRRC4B | 0.004347004 | -0.008246405 | -0.003899401 |
| KLHDC10 | -0.020235194 | 0.02705324 | 0.006818046 |
| GPBP1 | 0.028779061 | 0.026319207 | 0.055098269 |
| SH3GLB2 | -0.02881723 | 0.063870784 | 0.035053553 |
| RHPN2 | 0.011716254 | -0.012098749 | -0.000382495 |
| MORN4 | -0.01289354 | 0.024250401 | 0.01135686 |
| AC099552.3 | 0.002513946 | -0.002304671 | 0.000209275 |
| ATP5MC2 | -0.03523099 | 0.083332657 | 0.048101668 |
| CERS4 | -0.014445256 | 0.016402096 | 0.00195684 |
| RIN2 | -0.020555452 | 0.032219279 | 0.011663827 |
| PCLO | 0.021924915 | -0.039030165 | -0.01710525 |
| ARHGAP44 | -0.012881131 | 0.016591127 | 0.003709996 |
| AC005083.1 | -0.016079306 | 0.045369396 | 0.029290091 |
| APOL1 | 0.0150186 | -0.023882205 | -0.008863605 |
| CAB39 | -0.026615767 | 0.07730953 | 0.050693762 |
| PXMP4 | -0.014164195 | 0.01751764 | 0.003353445 |
| COL26A1 | -0.01009257 | 0.01191121 | 0.00181864 |
| KCTD3 | -0.016033798 | 0.024231375 | 0.008197577 |
| NPEPPS | 0.024566368 | 0.030182476 | 0.054748843 |
| BCAR3 | -0.021198911 | 0.035456491 | 0.01425758 |
| ABHD17C | -0.010955778 | 0.014379854 | 0.003424076 |
| ZFR | 0.023749421 | -0.02236656 | 0.001382861 |
| SLC47A1 | -0.003479391 | 0.003438007 | -4.13835E-05 |
| ZMPSTE24 | 0.025379229 | 0.026699401 | 0.052078629 |
| OPLAH | -0.012368484 | 0.017127784 | 0.0047593 |
| PRDX6 | 0.033485139 | 0.046597319 | 0.080082459 |
| SNHG14 | 0.035121026 | 0.044508567 | 0.079629593 |
| METTL7B | 0.002456934 | -0.002432172 | 2.47613E-05 |
| LRP5 | -0.01449267 | 0.020495937 | 0.006003267 |
| ZNF396 | -0.008554195 | 0.009838242 | 0.001284047 |
| BANF1 | -0.029243468 | 0.055799626 | 0.026556159 |
| PPP1R36 | 0.010397848 | -0.015373438 | -0.00497559 |
| UBE3D | -0.007584567 | 0.007596561 | 1.19942E-05 |
| CPLX1 | -0.007554574 | 0.011029457 | 0.003474883 |
| PYGL | -0.00829779 | 0.009331848 | 0.001034058 |
| ZNF503-AS2 | -0.015056623 | 0.037754223 | 0.022697601 |
| GPAA1 | 0.026332788 | 0.051087357 | 0.077420144 |
| UFC1 | 0.029285085 | 0.03306468 | 0.062349764 |
| PRDM16-DT | 0.032480133 | 0.045456028 | 0.077936161 |
| FBXL3 | -0.020287043 | 0.022370069 | 0.002083026 |
| KCNG1 | 0.002999172 | -0.003135996 | -0.000136825 |
| NUDT21 | -0.02241566 | 0.056335312 | 0.033919651 |
| SNTA1 | -0.01076325 | 0.011210052 | 0.000446803 |
| BRINP1 | 0.002278915 | -0.002600026 | -0.000321112 |

| | | | |
|---|---|---|---|
| VPS28 | 0.030977243 | 0.040730862 | 0.071708105 |
| C1QBP | 0.027598065 | 0.044843623 | 0.072441688 |
| EIF2S3 | -0.025452069 | 0.05535126 | 0.029899192 |
| SAPCD2 | -0.005079478 | 0.005581125 | 0.000501647 |
| CCDC18 | 0.006726448 | -0.006935381 | -0.000208933 |
| MPC2 | -0.032345905 | 0.153575289 | 0.121229384 |
| C19orf12 | 0.013218883 | -0.015183932 | -0.00196505 |
| ACADM | 0.028179353 | 0.042678848 | 0.070858201 |
| SH3TC1 | 0.018944985 | 0.02362084 | 0.042565824 |
| ZBED5-AS1 | -0.014025182 | 0.018259721 | 0.004234539 |
| AC109347.1 | 0.005143524 | -0.004873135 | 0.000270388 |
| PRUNE1 | -0.015247448 | 0.022218119 | 0.006970671 |
| AC019197.1 | -0.028513475 | 0.027621284 | -0.000892191 |
| LINC01088 | 0.003380979 | -0.003619271 | -0.000238293 |
| LGALSL | -0.013261805 | 0.015756575 | 0.00249477 |
| AC093583.1 | 0.018060822 | 0.036529058 | 0.05458988 |
| NDUFA5 | -0.032847608 | 0.115469046 | 0.082621439 |
| CERT1 | -0.021199493 | 0.031124667 | 0.009925174 |
| RBM48 | 0.011685887 | -0.013615191 | -0.001929304 |
| RPAP1 | -0.00999927 | 0.009673369 | -0.000325901 |
| TMEM159 | 0.013422519 | -0.025561336 | -0.012138818 |
| PJA1 | -0.014117277 | 0.017664119 | 0.003546843 |
| MMAB | -0.019564274 | 0.039802827 | 0.020238553 |

**Intercalated cells**

| Genes | logFC_T2D_HC | logFC_T2Di_T2D | logFC_T2Di_HC |
|---|---|---|---|
| PTGR1 | 0.423134788 | -0.427395424 | -0.004260636 |
| AKR1C1 | 0.279070038 | -0.27410188 | 0.004968158 |
| UMOD | -0.348751151 | 0.123023065 | -0.225728086 |
| CALCA | 0.435070588 | -0.405842383 | 0.029228204 |
| PFKFB3 | 0.199700042 | -0.199919084 | -0.000219042 |
| HMGCS2 | 0.249889173 | -0.254646323 | -0.00475715 |
| ACADVL | 0.272103216 | -0.171583439 | 0.100519778 |
| CCPG1 | 0.292499526 | -0.271813675 | 0.020685851 |
| PTGES | 0.205242022 | -0.198859693 | 0.006382329 |
| CAT | 0.29015168 | -0.22483409 | 0.065317589 |
| FBP1 | 0.21918271 | -0.227888425 | -0.008705715 |
| SLC6A12 | 0.159624243 | -0.15993429 | -0.000310046 |
| SCCPDH | 0.248541836 | -0.229839647 | 0.018702189 |
| ELF5 | 0.151711652 | -0.155256899 | -0.003545246 |
| RNF24 | 0.163499799 | -0.167854596 | -0.004354798 |
| SLC12A1 | -0.243329292 | 0.102566834 | -0.140762458 |
| CLDN4 | 0.266353829 | -0.218801468 | 0.047552361 |
| TMEM106C | 0.270381999 | -0.211179944 | 0.059202055 |
| MRPS6 | 0.290352321 | -0.21760995 | 0.072742372 |
| CTSH | 0.271125153 | -0.267863234 | 0.003261919 |
| TACSTD2 | 0.274137201 | -0.296689187 | -0.022551986 |
| LINC01687 | 0.164727907 | -0.172142308 | -0.007414401 |
| HEPACAM2 | 0.265493479 | -0.162678937 | 0.102814541 |
| ANXA2 | 0.20215322 | -0.227413401 | -0.025260181 |
| HLA-DRA | 0.182880761 | -0.188996761 | -0.006116 |
| KNG1 | -0.189417348 | 0.078910451 | -0.110506897 |
| EPG5 | 0.160285 | -0.157624447 | 0.002660553 |
| ARL6IP5 | 0.220488369 | -0.1403249 | 0.080163469 |
| AQP6 | 0.289698083 | -0.288797046 | 0.000901037 |
| HLA-DMB | 0.151111291 | -0.145453412 | 0.005657879 |
| BAG1 | 0.23175009 | -0.17049157 | 0.06125852 |
| PRDX1 | 0.266597178 | -0.138933319 | 0.127663859 |
| FOLR1 | 0.215109326 | -0.228974643 | -0.013865317 |
| HLA-DMA | 0.152120746 | -0.146639075 | 0.005481671 |
| SLC66A3 | 0.231394056 | -0.172906591 | 0.058487465 |
| GALNT3 | 0.154123718 | -0.148491099 | 0.005632619 |
| SCIN | 0.249868067 | -0.186510004 | 0.063358064 |
| AIG1 | 0.200361587 | -0.160121598 | 0.040239989 |
| LDHA | 0.260905176 | -0.182163095 | 0.078742081 |
| LAPTM4B | 0.211575479 | -0.172106339 | 0.03946914 |
| DEFB1 | -0.381667765 | 0.30271584 | -0.078951925 |
| DRAM1 | 0.128705448 | -0.126360706 | 0.002344742 |
| FXYD4 | 0.228074721 | -0.269026043 | -0.040951323 |
| EHF | 0.111151471 | -0.107662042 | 0.003489429 |
| FAM107B | 0.247142225 | -0.184276041 | 0.062866185 |

| | | | |
|---|---|---|---|
| ZDHHC3 | 0.224492387 | -0.152391649 | 0.072100738 |
| CD74 | 0.210846658 | -0.207085554 | 0.003761103 |
| RHCG | 0.359711852 | -0.259626708 | 0.100085144 |
| RGCC | 0.192498553 | -0.230058728 | -0.037560175 |
| SLC38A2 | 0.184784908 | -0.203167468 | -0.01838256 |
| LIMA1 | 0.233868469 | -0.192609523 | 0.041258946 |
| EPHX1 | 0.124246545 | -0.118153691 | 0.006092855 |
| LPIN2 | 0.140742213 | -0.131898495 | 0.008843719 |
| TPD52L1 | 0.153739296 | -0.141439649 | 0.012299647 |
| TSPO | 0.232887275 | -0.139834289 | 0.093052987 |
| SLC14A2 | 0.104232109 | -0.10404874 | 0.000183368 |
| RTN3 | 0.205988421 | -0.176091065 | 0.029897356 |
| BTG1 | 0.234459782 | -0.188259737 | 0.046200046 |
| QPRT | 0.119908558 | -0.118082832 | 0.001825726 |
| STXBP6 | 0.095965712 | -0.094290512 | 0.001675201 |
| EPCAM | 0.252514739 | -0.191928699 | 0.06058604 |
| ITM2B | 0.245778566 | -0.201128667 | 0.044649899 |
| TMEM50A | 0.198006842 | -0.142189387 | 0.055817456 |
| IER3 | 0.1592041 | -0.170549515 | -0.011345415 |
| SYNE2 | 0.226790436 | -0.202205996 | 0.02458444 |
| TMED5 | 0.184733846 | -0.173273388 | 0.011460458 |
| SERPINA1 | 0.114166682 | -0.126272291 | -0.012105609 |
| ACAT1 | 0.230072771 | -0.222876681 | 0.007196089 |
| FXYD2 | -0.299812496 | 0.355306323 | 0.055493827 |
| SERPINA5 | 0.155695957 | -0.171343089 | -0.015647132 |
| HLA-DRB1 | 0.124753708 | -0.124722496 | 3.12119E-05 |
| NFKBIA | 0.141729296 | -0.145102931 | -0.003373635 |
| BTG3 | 0.162813028 | -0.139215143 | 0.023597885 |
| SLC5A3 | 0.226278908 | -0.206563471 | 0.019715437 |
| UBC | 0.216753602 | -0.150914204 | 0.065839398 |
| REEP5 | 0.20638965 | -0.112155932 | 0.094233718 |
| TSPAN1 | 0.193475764 | -0.193124409 | 0.000351355 |
| PGGHG | 0.090228158 | -0.085599243 | 0.004628914 |
| PLOD2 | 0.161856418 | -0.162297507 | -0.000441089 |
| PAX8 | 0.190489019 | -0.192780796 | -0.002291777 |
| ITPR2 | 0.209908515 | -0.14226262 | 0.067645895 |
| ATP7A | 0.144278216 | -0.127648294 | 0.016629922 |
| DDIT4L | 0.095661188 | -0.101092108 | -0.00543092 |
| TMEM116 | 0.158419416 | -0.098290813 | 0.060128603 |
| ARPC1B | 0.136366658 | -0.098033574 | 0.038333084 |
| SPINT2 | 0.210997293 | -0.154989822 | 0.056007471 |
| CD9 | 0.263840422 | -0.185463715 | 0.078376707 |
| ITGB1 | 0.179594474 | -0.144357285 | 0.035237189 |
| ACBD5 | 0.167157802 | -0.162849341 | 0.004308461 |
| CYB5A | 0.222331011 | -0.229003101 | -0.00667209 |
| AQP3 | 0.139175824 | -0.153853697 | -0.014677873 |
| TMPRSS2 | 0.193443543 | -0.101682157 | 0.091761387 |

| | | | |
|---|---|---|---|
| TMEM213 | 0.217013454 | -0.138867619 | 0.078145835 |
| MATN2 | 0.079703315 | -0.078391799 | 0.001311515 |
| KIT | 0.210238387 | -0.158564281 | 0.051674106 |
| LIN7A | 0.104064307 | -0.094941806 | 0.009122501 |
| TWF1 | 0.165222147 | -0.101032991 | 0.064189156 |
| EMP2 | 0.079410031 | -0.082248987 | -0.002838956 |
| HNRNPF | 0.185986372 | -0.068351203 | 0.117635169 |
| PPIC | 0.169565273 | -0.095027383 | 0.07453789 |
| KCNJ1 | -0.090915049 | 0.034096037 | -0.056819012 |
| C15orf65 | 0.11394358 | -0.090346128 | 0.023597452 |
| S100A1 | 0.082541276 | -0.078158996 | 0.00438228 |
| TPM1 | 0.192947076 | -0.112930519 | 0.080016557 |
| SETD3 | 0.136383817 | -0.112783124 | 0.023600692 |
| HLA-DPA1 | 0.076691886 | -0.074630531 | 0.002061355 |
| TMEM87A | 0.155021481 | -0.132536507 | 0.022484974 |
| CD164 | 0.179738448 | -0.141196893 | 0.038541556 |
| AC023421.1 | 0.071265478 | -0.070478253 | 0.000787225 |
| MAGED2 | 0.192846001 | -0.094306809 | 0.098539192 |
| TMEM238L | 0.072618301 | -0.074474052 | -0.001855752 |
| CPQ | 0.167769446 | -0.151274653 | 0.016494793 |
| ECH1 | 0.182100241 | -0.066910059 | 0.115190182 |
| FUCA1 | 0.14959765 | -0.118744593 | 0.030853057 |
| ARPC2 | 0.170722581 | -0.106795488 | 0.063927093 |
| BX284668.5 | 0.111310298 | -0.115623707 | -0.004313409 |
| TMBIM4 | 0.197328647 | -0.128366019 | 0.068962628 |
| NRCAM | 0.098462439 | -0.101984231 | -0.003521792 |
| PALM3 | 0.088178912 | -0.090515745 | -0.002336832 |
| CD24 | 0.236396292 | -0.239036724 | -0.002640432 |
| CLN3 | 0.120599885 | -0.112030257 | 0.008569628 |
| PCDH7 | 0.073472566 | -0.074806004 | -0.001333438 |
| UCP2 | 0.119551515 | -0.101364385 | 0.018187129 |
| SERPINA6 | 0.092383889 | -0.087721368 | 0.004662521 |
| THSD7A | 0.159534432 | -0.159619778 | -8.53458E-05 |
| ORAI1 | 0.123277811 | -0.088597598 | 0.034680213 |
| DHRS7 | 0.217041311 | -0.18785237 | 0.029188941 |
| ATP6V0A4 | 0.206306556 | -0.152647253 | 0.053659302 |
| CAPS | 0.179709533 | -0.126359334 | 0.0533502 |
| NAPEPLD | 0.107821365 | -0.118550953 | -0.010729588 |
| ELF3 | 0.158322426 | -0.110063062 | 0.048259363 |
| LY6K | 0.115412485 | -0.100296091 | 0.015116394 |
| HADHA | 0.181375585 | -0.101993324 | 0.079382261 |
| TLN2 | 0.088605049 | -0.100911149 | -0.0123061 |
| LMAN1 | 0.193385274 | -0.177699534 | 0.01568574 |
| KTN1 | 0.192078143 | -0.154342106 | 0.037736037 |
| CLDN8 | 0.195800893 | -0.162596205 | 0.033204688 |
| ATP6V0C | 0.209984363 | -0.102701716 | 0.107282647 |
| MGST3 | 0.201669099 | -0.164701709 | 0.03696739 |

| | | | |
|---|---|---|---|
| RANBP3L | 0.102443447 | -0.072893318 | 0.029550129 |
| MESD | 0.146519942 | -0.112115929 | 0.034404014 |
| KCNJ3 | 0.087975533 | -0.097964731 | -0.009989197 |
| AL031123.5 | 0.125167189 | -0.099549866 | 0.025617323 |
| HADHB | 0.205954836 | -0.060047516 | 0.14590732 |
| RAB17 | 0.123769461 | -0.104420975 | 0.019348485 |
| S100A11 | 0.212153189 | -0.128141983 | 0.084011207 |
| SERPINI1 | 0.087464727 | -0.084731255 | 0.002733472 |
| ERO1B | 0.103269335 | -0.095494221 | 0.007775114 |
| HTATIP2 | 0.110505836 | -0.074214881 | 0.036290955 |
| RAB9A | 0.131732538 | -0.084624047 | 0.047108491 |
| SLCO4A1 | 0.075452113 | -0.079203381 | -0.003751267 |
| RTN4 | 0.220692553 | -0.133130258 | 0.087562294 |
| CLINT1 | 0.146059426 | -0.126934091 | 0.019125335 |
| ECI1 | 0.184678551 | -0.058678324 | 0.126000226 |
| CD46 | 0.185827382 | -0.155603039 | 0.030224343 |
| PLIN2 | 0.097506924 | -0.09262489 | 0.004882034 |
| GMPR | 0.170450888 | -0.070628135 | 0.099822753 |
| RCN2 | 0.157411132 | -0.09658265 | 0.060828482 |
| LRRFIP1 | 0.137544777 | -0.117539461 | 0.020005316 |
| MRPS35 | 0.153045157 | -0.099406325 | 0.053638832 |
| TINAGL1 | 0.111525111 | -0.121689784 | -0.010164674 |
| ARHGAP18 | 0.194870833 | -0.102227067 | 0.092643766 |
| AFMID | 0.091767221 | -0.095120249 | -0.003353029 |
| CLDN16 | -0.063487251 | 0.021693347 | -0.041793904 |
| DIAPH2 | 0.071871671 | -0.07599679 | -0.004125118 |
| CD82 | 0.109244646 | -0.115658474 | -0.006413828 |
| DDX3Y | 0.08251911 | -0.091570443 | -0.009051333 |
| SH2D4A | 0.133783758 | -0.101278647 | 0.032505111 |
| SLTM | 0.141092901 | -0.110785923 | 0.030306977 |
| KDELR1 | 0.170499149 | -0.097310216 | 0.073188933 |
| KLF5 | 0.072655532 | -0.073253621 | -0.00059809 |
| KRTCAP3 | 0.117905972 | -0.112794937 | 0.005111035 |
| CDH16 | 0.187754314 | -0.209130377 | -0.021376063 |
| PGRMC1 | 0.17493347 | -0.141656953 | 0.033276517 |
| RNF152 | 0.170602772 | -0.167567017 | 0.003035756 |
| SCGB2A1 | 0.124047395 | -0.134148495 | -0.0101011 |
| PAK2 | 0.12842484 | -0.109605488 | 0.018819351 |
| EMP3 | 0.082743913 | -0.062428022 | 0.020315891 |
| EMX1 | 0.098523978 | -0.108942031 | -0.010418054 |
| SLC35B3 | 0.087179083 | -0.072225329 | 0.014953754 |
| UBAC2 | 0.122373674 | -0.081956908 | 0.040416765 |
| TMBIM6 | 0.175920349 | -0.141013985 | 0.034906364 |
| DCP2 | 0.110013362 | -0.090357903 | 0.019655459 |
| NDUFS2 | 0.165367789 | -0.088781385 | 0.076586405 |
| CAST | 0.156497955 | -0.103367145 | 0.05313081 |
| LMO7 | 0.148663149 | -0.116208337 | 0.032454812 |

| | | | |
|---|---|---|---|
| HGD | 0.115897699 | -0.097534139 | 0.01836356 |
| MUC20 | 0.20234646 | -0.140115197 | 0.062231263 |
| HSD17B4 | 0.130604011 | -0.112513781 | 0.01809023 |
| CAPG | 0.186098035 | -0.216581541 | -0.030483506 |
| ORMDL1 | 0.140905245 | -0.081837635 | 0.059067611 |
| PRDX4 | 0.133312195 | -0.099043139 | 0.034269057 |
| S100A6 | 0.158357597 | -0.114300903 | 0.044056694 |
| CLDN10 | -0.081095494 | 0.028374284 | -0.052721211 |
| SMIM22 | 0.115545272 | -0.142711437 | -0.027166165 |
| AMIGO2 | 0.081293258 | -0.080116517 | 0.001176741 |
| TSPAN3 | 0.139877311 | -0.124017136 | 0.015860175 |
| HAGH | 0.146555994 | -0.068658013 | 0.077897981 |
| ACAA2 | 0.14071842 | -0.097094295 | 0.043624125 |
| SERINC2 | 0.154806681 | -0.105933116 | 0.048873565 |
| ATRAID | 0.169198395 | -0.102629678 | 0.066568717 |
| PARP4 | 0.104998787 | -0.106782872 | -0.001784085 |
| CYP51A1 | 0.14236117 | -0.118039967 | 0.024321203 |
| GRN | 0.144946637 | -0.121947715 | 0.022998922 |
| DDIT4 | 0.068752508 | -0.072857842 | -0.004105334 |
| BMPR1B | 0.106798886 | -0.113240163 | -0.006441276 |
| MAL2 | 0.140545162 | -0.132365434 | 0.008179728 |
| TUBB4B | 0.16718793 | -0.086871572 | 0.080316359 |
| SYPL1 | 0.167773128 | -0.083988676 | 0.083784452 |
| PPIB | 0.175909925 | -0.096826429 | 0.079083496 |
| CUL5 | 0.136886519 | -0.109024949 | 0.027861569 |
| ADGRF1 | 0.18204942 | -0.154936051 | 0.027113369 |
| IFNAR1 | 0.129079556 | -0.120908248 | 0.008171308 |
| KIF5B | 0.146530512 | -0.121861314 | 0.024669198 |
| C9orf16 | 0.181434707 | -0.084217539 | 0.097217168 |
| CA2 | 0.189919472 | -0.134440996 | 0.055478475 |
| SDC2 | 0.157656238 | -0.08662194 | 0.071034298 |
| DRAM2 | 0.11712556 | -0.081459018 | 0.035666542 |
| SPTBN2 | 0.12233769 | -0.119071482 | 0.003266208 |
| TNFRSF11B | 0.070555552 | -0.070435851 | 0.000119701 |
| SLC16A5 | 0.119060428 | -0.091151972 | 0.027908456 |
| RAB15 | 0.066732849 | -0.058590686 | 0.008142164 |
| ATP6V0E1 | 0.186867225 | -0.107060059 | 0.079807167 |
| MRPS5 | 0.137303489 | -0.08642725 | 0.050876239 |
| CDH1 | 0.159782611 | -0.123334831 | 0.03644778 |
| THEM4 | 0.137054491 | -0.133020959 | 0.004033532 |
| SEC62 | 0.168284216 | -0.161529981 | 0.006754234 |
| PKM | 0.175528167 | -0.106780208 | 0.068747959 |
| UQCRFS1 | 0.174347798 | -0.044945952 | 0.129401846 |
| CORIN | 0.052876275 | -0.052876275 | -5.32907E-15 |
| GLTP | 0.162646998 | -0.090874301 | 0.071772697 |
| ATP5F1A | 0.168574324 | -0.077618762 | 0.090955562 |
| PLPP5 | 0.123398962 | -0.113806489 | 0.009592473 |

| | | | |
|---|---|---|---|
| SLC17A5 | 0.075812721 | -0.062802485 | 0.013010236 |
| AFDN | 0.130067471 | -0.105716551 | 0.024350921 |
| GHITM | 0.171142316 | -0.066623428 | 0.104518888 |
| ATP6AP1 | 0.161249757 | -0.136852715 | 0.024397041 |
| SCPEP1 | 0.144070632 | -0.11761753 | 0.026453102 |
| TMED9 | 0.140109027 | -0.088749216 | 0.051359811 |
| SHB | 0.082034502 | -0.074784303 | 0.007250199 |
| GLUL | 0.151062043 | -0.08499315 | 0.066068893 |
| TMEM165 | 0.129121995 | -0.094507161 | 0.034614834 |
| PLAU | -0.056584916 | 0.019720479 | -0.036864437 |
| GPRC5C | 0.144855579 | -0.124723057 | 0.020132522 |
| RFC1 | 0.117970519 | -0.087703865 | 0.030266654 |
| CFB | 0.047899353 | -0.047969773 | -7.04191E-05 |
| GSN | 0.1269585 | -0.071772975 | 0.055185525 |
| SPPL2A | 0.12773662 | -0.096170571 | 0.031566048 |
| DNAJA1 | 0.15283681 | -0.077336713 | 0.075500097 |
| PFKP | 0.078905657 | -0.072344772 | 0.006560885 |
| ERP29 | 0.161259278 | -0.0965537 | 0.064705578 |
| AKAP9 | 0.156951659 | -0.122627772 | 0.034323887 |
| DDRGK1 | 0.140433466 | -0.085831394 | 0.054602073 |
| RAPGEF3 | 0.07718407 | -0.068398447 | 0.008785622 |
| GOLGB1 | 0.14021324 | -0.11897364 | 0.0212396 |
| IER3IP1 | 0.143758822 | -0.051159462 | 0.09259936 |
| NRDC | 0.154177791 | -0.120170287 | 0.034007503 |
| UBE2M | 0.141365696 | -0.04824176 | 0.093123935 |
| MYO6 | 0.141289899 | -0.116156279 | 0.02513362 |
| ATP5F1C | 0.175171331 | -0.058935668 | 0.116235663 |
| EIF4G3 | 0.087800423 | -0.087321091 | 0.000479332 |
| CD81 | 0.181938094 | -0.101079897 | 0.080858196 |
| VPS13C | 0.126264805 | -0.115147547 | 0.011117258 |
| ZFYVE16 | 0.12331887 | -0.099968483 | 0.023350387 |
| NEBL | 0.10311756 | -0.09647294 | 0.006644621 |
| RDH11 | 0.116085631 | -0.098066261 | 0.018019371 |
| MFSD10 | 0.121659111 | -0.07997436 | 0.041684752 |
| COX6A1 | 0.163411833 | -0.091503212 | 0.071908621 |
| APIP | 0.122148527 | -0.095053551 | 0.027094975 |
| APMAP | 0.119008315 | -0.094604011 | 0.024404304 |
| LIPH | 0.093050037 | -0.084420846 | 0.008629191 |
| ACKR3 | 0.054234306 | -0.059163427 | -0.004929121 |
| KIAA1109 | 0.123288362 | -0.094312081 | 0.028976281 |
| PTPN13 | 0.092827372 | -0.093643417 | -0.000816045 |
| NCOR1 | 0.134369199 | -0.145117643 | -0.010748444 |
| CNDP2 | 0.126230869 | -0.094151874 | 0.032078995 |
| RAP1B | 0.128796192 | -0.050494097 | 0.078302095 |
| H2AFZ | 0.158839862 | -0.082167301 | 0.076672561 |
| CLIC1 | 0.161596217 | -0.077266256 | 0.084329961 |
| MYCBP2 | 0.108350461 | -0.100482292 | 0.007868169 |

| | | | |
|---|---|---|---|
| MYDGF | 0.133281712 | -0.092763148 | 0.040518564 |
| NDFIP1 | 0.157247996 | -0.076099069 | 0.081148927 |
| PDIA3 | 0.154064945 | -0.115561006 | 0.038503939 |
| APP | 0.167044627 | -0.186539269 | -0.019494641 |
| KIDINS220 | 0.126885982 | -0.099897812 | 0.026988171 |
| HTATSF1 | 0.1167349 | -0.106255154 | 0.010479747 |
| PCMT1 | 0.126792435 | -0.0741301 | 0.052662336 |
| AMFR | 0.150361273 | -0.105130912 | 0.045230361 |
| SDHA | 0.141607852 | -0.075885813 | 0.06572204 |
| PDHB | 0.142601532 | -0.071615507 | 0.070986025 |
| GRAMD2B | 0.09152394 | -0.076630852 | 0.014893088 |
| AKR1C3 | 0.06561223 | -0.0606527 | 0.00495953 |
| PPFIBP2 | 0.0943329 | -0.081216756 | 0.013116144 |
| KRT18 | 0.169575569 | -0.082953717 | 0.086621851 |
| MIR99AHG | 0.124905755 | -0.087386315 | 0.03751944 |
| CD63 | 0.177870803 | -0.098546028 | 0.079324775 |
| ENO1 | 0.152468032 | -0.120505812 | 0.03196222 |
| DSP | 0.156836478 | -0.105654803 | 0.051181675 |
| HLA-C | 0.181056439 | -0.051436879 | 0.129619559 |
| IL18 | 0.178246297 | -0.057175005 | 0.121071292 |
| PTPRJ | 0.124427074 | -0.113073298 | 0.011353776 |
| PPFIBP1 | 0.082417608 | -0.079013022 | 0.003404586 |
| TCF25 | 0.132988189 | -0.100950526 | 0.032037663 |
| PPP2R3A | 0.101490552 | -0.081171239 | 0.020319312 |
| FBXW11 | 0.1039062 | -0.09327126 | 0.01063494 |
| MELTF | 0.055585931 | -0.04932432 | 0.006261611 |
| NDUFA6 | 0.163058807 | -0.074353021 | 0.088705786 |
| SPCS2 | 0.156854686 | -0.082582738 | 0.074271948 |
| SCARB2 | 0.126997888 | -0.087638951 | 0.039358937 |
| ISYNA1 | 0.091450236 | -0.098028116 | -0.00657788 |
| DAB2 | 0.10465612 | -0.117275617 | -0.012619497 |
| P3H2 | 0.084231843 | -0.091797787 | -0.007565945 |
| RHOD | 0.087752354 | -0.063176687 | 0.024575667 |
| GINM1 | 0.134448874 | -0.098075944 | 0.03637293 |
| MTDH | 0.131839729 | -0.106565901 | 0.025273827 |
| CEBPD | 0.154903277 | -0.116010761 | 0.038892517 |
| DMKN | 0.151271449 | -0.086085632 | 0.065185817 |
| HIF1A | 0.113778939 | -0.085040049 | 0.02873889 |
| RALBP1 | 0.130696198 | -0.13988371 | -0.009187512 |
| RPL37 | 0.153138974 | -0.067253595 | 0.085885379 |
| CAMK1 | 0.09767633 | -0.095459964 | 0.002216366 |
| ELP5 | 0.108559651 | -0.07373524 | 0.034824411 |
| CLYBL | 0.111277788 | -0.086314652 | 0.024963135 |
| PSMC5 | 0.130656434 | -0.040873495 | 0.089782939 |
| RALGPS2 | 0.108666036 | -0.065038537 | 0.043627499 |
| PDHA1 | 0.134004394 | -0.08394724 | 0.050057154 |
| NDUFB9 | 0.154677298 | -0.046612554 | 0.108064744 |

| | | | |
|---|---|---|---|
| LAPTM4A | 0.160189913 | -0.095045884 | 0.065144029 |
| CALR | 0.164372378 | -0.107396382 | 0.056975997 |
| BORCS7 | 0.129713374 | -0.082417866 | 0.047295508 |
| SNRPN | 0.150190092 | -0.051513619 | 0.098676473 |
| ANXA3 | 0.110135076 | -0.082475197 | 0.027659878 |
| SLC9A3R1 | 0.118651583 | -0.053878259 | 0.064773324 |
| FNDC3A | 0.113542209 | -0.059771564 | 0.053770645 |
| CLTC | 0.133115652 | -0.06918488 | 0.063930772 |
| GOLGA8A | 0.092180289 | -0.067857526 | 0.024322763 |
| CRYAB | 0.092120841 | -0.083476414 | 0.008644426 |
| SNHG5 | 0.12209995 | -0.103390182 | 0.018709767 |
| HLA-E | 0.133611136 | -0.091210893 | 0.042400244 |
| MYZAP | 0.06557101 | -0.062694269 | 0.002876741 |
| NUDT9 | 0.098709874 | -0.060185442 | 0.038524432 |
| SIPA1 | 0.088600444 | -0.055434906 | 0.033165538 |
| MTR | 0.115725091 | -0.118282046 | -0.002556955 |
| ATP6V0B | 0.176259979 | -0.120685986 | 0.055573992 |
| DES | 0.060194977 | -0.063176818 | -0.002981841 |
| TM9SF3 | 0.150371144 | -0.123320782 | 0.027050363 |
| COBLL1 | 0.151739649 | -0.150260225 | 0.001479423 |
| S100A10 | 0.183956675 | -0.114329883 | 0.069626792 |
| CYC1 | 0.157606981 | -0.076979062 | 0.080627919 |
| PLEKHH3 | 0.060508096 | -0.050697614 | 0.009810482 |
| CPPED1 | 0.107713172 | -0.095857596 | 0.011855576 |
| INPP5J | 0.102466577 | -0.089558605 | 0.012907972 |
| FAM184B | 0.065903236 | -0.061954053 | 0.003949183 |
| CD151 | 0.123229206 | -0.096803804 | 0.026425402 |
| TMEM106B | 0.121609678 | -0.086045566 | 0.035564112 |
| MYL12A | 0.151253726 | -0.088224239 | 0.063029487 |
| RPS8 | 0.14440799 | -0.041742878 | 0.102665112 |
| B2M | 0.181951669 | -0.068880272 | 0.113071397 |
| ANK2 | 0.078674979 | -0.091591274 | -0.012916295 |
| EBPL | 0.124613632 | -0.061891746 | 0.062721885 |
| BOD1L1 | 0.110701643 | -0.100319926 | 0.010381717 |
| PLP2 | 0.103414674 | -0.067194344 | 0.03622033 |
| HS6ST1 | 0.077021509 | -0.063310754 | 0.013710755 |
| TMEM59 | 0.159742702 | -0.110971908 | 0.048770794 |
| TSTD1 | 0.150170909 | -0.030332138 | 0.119838771 |
| TALDO1 | 0.131032036 | -0.058190572 | 0.072841464 |
| RPS10 | 0.157235955 | -0.047931128 | 0.109304826 |
| DDX52 | 0.091733038 | -0.078179723 | 0.013553315 |
| PLBD1 | 0.08787127 | -0.082326731 | 0.00554454 |
| RDH10 | 0.093609104 | -0.118951405 | -0.025342301 |
| NIPAL3 | 0.084004407 | -0.075531377 | 0.00847303 |
| TUFM | 0.149370124 | -0.07187795 | 0.077492174 |
| SEPTIN10 | 0.083485726 | -0.068209662 | 0.015276063 |
| LRPAP1 | 0.132068847 | -0.09039917 | 0.041669677 |

| | | | |
|---|---|---|---|
| PIM3 | 0.107920274 | -0.066707235 | 0.041213039 |
| PRADC1 | 0.092269633 | -0.051669287 | 0.040600346 |
| HPS4 | 0.081991674 | -0.084548787 | -0.002557114 |
| GLS | 0.127618973 | -0.146381087 | -0.018762114 |
| TMED4 | 0.137649778 | -0.102887562 | 0.034762216 |
| RAB13 | 0.149959044 | -0.040262205 | 0.109696838 |
| GALM | 0.067417149 | -0.069849928 | -0.002432779 |
| EMC6 | 0.125179355 | -0.0540212 | 0.071158155 |
| LITAF | 0.15807664 | -0.033689732 | 0.124386909 |
| SSR4 | 0.15585637 | -0.055661088 | 0.100195281 |
| LAMP2 | 0.119073443 | -0.099700985 | 0.019372459 |
| GOLIM4 | 0.12720895 | -0.099877145 | 0.027331805 |
| HACD3 | 0.141446042 | -0.127520246 | 0.013925797 |
| RNH1 | 0.1383647 | -0.047888968 | 0.090475731 |
| SCAMP2 | 0.108560873 | -0.081814559 | 0.026746314 |
| PARK7 | 0.152243062 | -0.053131107 | 0.099111956 |
| TMX4 | 0.120536275 | -0.117878851 | 0.002657424 |
| SMAGP | 0.067404117 | -0.059784483 | 0.007619634 |
| CXCL12 | -0.049039224 | 0.020077212 | -0.028962012 |
| KCP | 0.044072609 | -0.04577061 | -0.001698001 |
| SELENBP1 | 0.112417608 | -0.072370704 | 0.040046904 |
| GRB14 | 0.152539941 | -0.092946161 | 0.059593781 |
| IFNGR2 | 0.096870081 | -0.05439711 | 0.042472971 |
| PRRG4 | 0.090239278 | -0.072536513 | 0.017702765 |
| CARHSP1 | 0.075999367 | -0.075207348 | 0.000792019 |
| TMEM139 | 0.074920997 | -0.061175798 | 0.0137452 |
| SRP54 | 0.092215031 | -0.058546324 | 0.033668708 |
| NDUFA10 | 0.135685654 | -0.065895803 | 0.06978985 |
| OS9 | 0.115170069 | -0.085854727 | 0.029315343 |
| SLC16A4 | 0.067548758 | -0.076896629 | -0.009347871 |
| NFAT5 | 0.102923316 | -0.10833838 | -0.005415064 |
| PGP | 0.106640017 | -0.040418776 | 0.06622124 |
| CNN3 | 0.138630978 | -0.05569729 | 0.082933689 |
| TECR | 0.145587483 | -0.103489276 | 0.042098207 |
| GAPDH | 0.156185581 | -0.04349246 | 0.112693121 |
| TRIP11 | 0.113674368 | -0.085494929 | 0.02817944 |
| GOT1 | 0.128806119 | -0.075615746 | 0.053190373 |
| ARL6IP1 | 0.132457958 | -0.128034294 | 0.004423664 |
| SON | 0.137311844 | -0.130798501 | 0.006513343 |
| GAL3ST1 | 0.065492089 | -0.067008021 | -0.001515932 |
| ERLEC1 | 0.106416823 | -0.092782762 | 0.013634062 |
| RERE | 0.106864238 | -0.086930872 | 0.019933366 |
| ELF1 | 0.090640709 | -0.07390048 | 0.016740228 |
| DDX5 | 0.142522023 | -0.112863985 | 0.029658038 |
| HLA-DPB1 | 0.041419468 | -0.037303065 | 0.004116404 |
| GSPT1 | 0.113985921 | -0.085351125 | 0.028634796 |
| SARAF | 0.144227185 | -0.081121601 | 0.063105584 |

| | | | |
|---|---|---|---|
| PXK | 0.123793291 | -0.056631911 | 0.06716138 |
| NR2F6 | 0.108722276 | -0.075379216 | 0.033343061 |
| AL031123.4 | 0.100883548 | -0.079621117 | 0.021262431 |
| HSPA9 | 0.126859043 | -0.076955206 | 0.049903838 |
| SPTBN1 | 0.129413103 | -0.127786395 | 0.001626708 |
| SLC8A1 | -0.067135583 | 0.015698452 | -0.051437131 |
| SRSF10 | 0.116663381 | -0.084448919 | 0.032214462 |
| STK24 | 0.100495816 | -0.070643121 | 0.029852695 |
| HERC2 | 0.081377063 | -0.070765871 | 0.010611192 |
| RPL19 | 0.140245096 | -0.046975135 | 0.093269961 |
| ATP6V1G1 | 0.144594117 | -0.053934528 | 0.090659588 |
| SLC50A1 | 0.09585863 | -0.078063661 | 0.017794969 |
| SPR | 0.076093174 | -0.056308101 | 0.019785073 |
| GATA3 | 0.081249733 | -0.083280908 | -0.002031175 |
| FH | 0.103242901 | -0.056200788 | 0.047042113 |
| TMEM243 | 0.101546361 | -0.092320445 | 0.009225916 |
| CAMK2N1 | 0.148999351 | -0.033739033 | 0.115260318 |
| UQCR10 | 0.145836213 | -0.053205993 | 0.09263022 |
| ARPC3 | 0.145113165 | -0.067809401 | 0.077303765 |
| UBXN4 | 0.126146984 | -0.10355935 | 0.022587634 |
| NDUFC2 | 0.148988184 | -0.073581705 | 0.075406479 |
| ARFGEF1 | 0.10023819 | -0.093401896 | 0.006836294 |
| PLLP | 0.140482675 | -0.035368381 | 0.105114294 |
| AFG3L2 | 0.117097751 | -0.109178364 | 0.007919387 |
| MAN1A1 | 0.110941737 | -0.102664579 | 0.008277158 |
| COASY | 0.079052562 | -0.069482577 | 0.009569986 |
| TGS1 | 0.072774128 | -0.06288279 | 0.009891339 |
| CTSD | 0.158154824 | -0.077645517 | 0.080509307 |
| EGFR | 0.093125901 | -0.067608388 | 0.025517513 |
| MYOF | 0.063526957 | -0.068686878 | -0.005159921 |
| ADSS | 0.085613129 | -0.048014625 | 0.037598504 |
| ILF2 | 0.111538507 | -0.062891159 | 0.048647349 |
| EXOSC7 | 0.15245286 | -0.040270714 | 0.112182146 |
| GBP3 | 0.05659544 | -0.044002797 | 0.012592643 |
| PIGF | 0.104751673 | -0.057419215 | 0.047332459 |
| STAP2 | 0.0683872 | -0.051631842 | 0.016755359 |
| NOL7 | 0.122614986 | -0.077522382 | 0.045092604 |
| P4HA1 | 0.067857116 | -0.069665617 | -0.001808501 |
| TMCO1 | 0.129988903 | -0.110506198 | 0.019482705 |
| IQGAP1 | 0.10156655 | -0.073618571 | 0.02794798 |
| CLN5 | 0.117232491 | -0.090088788 | 0.027143702 |
| REEP3 | 0.103164746 | -0.068262546 | 0.0349022 |
| PCMTD1 | 0.114019338 | -0.070697899 | 0.043321439 |
| EIF2AK4 | 0.077122848 | -0.058846524 | 0.018276324 |
| PIGH | 0.101740767 | -0.083957777 | 0.01778299 |
| QPCT | 0.044900003 | -0.046176361 | -0.001276357 |
| STARD10 | 0.120984725 | -0.02477329 | 0.096211434 |

| | | | |
|---|---|---|---|
| ETS2 | 0.056367719 | -0.046837799 | 0.00952992 |
| TMEM30A | 0.106897421 | -0.085001794 | 0.021895627 |
| SYS1 | 0.079644871 | -0.045820654 | 0.033824217 |
| CRYBG1 | 0.056358658 | -0.062450335 | -0.006091677 |
| TMEM35B | 0.100693467 | -0.078778234 | 0.021915233 |
| MBP | 0.100042117 | -0.067368664 | 0.032673453 |
| HSD17B11 | 0.069187111 | -0.070485825 | -0.001298714 |
| FAM177A1 | 0.107161381 | -0.072877284 | 0.034284097 |
| SPINT1 | 0.10895752 | -0.080162256 | 0.028795264 |
| ASPA | 0.038731042 | -0.041143034 | -0.002411993 |
| PLEK2 | 0.09197836 | -0.063818708 | 0.028159653 |
| IQCA1 | 0.042846381 | -0.040270966 | 0.002575415 |
| PYGO1 | 0.036124306 | -0.035607111 | 0.000517195 |
| MT-ND5 | -0.114317223 | 0.057531268 | -0.056785955 |
| CTSS | 0.053504201 | -0.055589627 | -0.002085426 |
| TEX264 | 0.130257549 | -0.08078024 | 0.049477309 |
| ATP5F1B | 0.149841232 | -0.060695552 | 0.08914568 |
| PSMA7 | 0.144859344 | -0.075587778 | 0.069271566 |
| MPDU1 | 0.104536571 | -0.067395448 | 0.037141123 |
| SNHG7 | 0.130710549 | -0.064213617 | 0.066496931 |
| FUCA2 | 0.104080148 | -0.088723537 | 0.015356611 |
| HSD17B12 | 0.11562114 | -0.054812607 | 0.060808533 |
| NDUFS5 | 0.143408616 | -0.081554035 | 0.061854581 |
| ATP5IF1 | 0.146835042 | -0.088867885 | 0.057967157 |
| TOM1L1 | 0.097420304 | -0.042581714 | 0.05483859 |
| AGPAT5 | 0.065800269 | -0.048094107 | 0.017706162 |
| PRPF4B | 0.109782587 | -0.099281494 | 0.010501093 |
| PNMT | 0.044656774 | -0.044842385 | -0.000185611 |
| SEPTIN11 | 0.105184198 | -0.095239988 | 0.00994421 |
| TMED10 | 0.1225078 | -0.094922159 | 0.027585641 |
| SLC25A4 | 0.142892345 | -0.034108567 | 0.108783777 |
| HEBP1 | 0.120210171 | -0.050495932 | 0.069714239 |
| AIFM1 | 0.100324119 | -0.069407329 | 0.03091679 |
| PDCD10 | 0.102884398 | -0.040796981 | 0.062087417 |
| ATP6V1A | 0.140527281 | -0.085379268 | 0.055148013 |
| MPP7 | 0.104093428 | -0.09562112 | 0.008472308 |
| SLC25A39 | 0.157705738 | -0.058549946 | 0.099155791 |
| AOC1 | 0.049771616 | -0.052333422 | -0.002561806 |
| ITFG1 | 0.100980115 | -0.060648389 | 0.040331726 |
| SPECC1L | 0.075222131 | -0.069572311 | 0.005649821 |
| TUBA1B | 0.130699678 | -0.057000265 | 0.073699413 |
| TGOLN2 | 0.12471417 | -0.107439622 | 0.017274549 |
| VDAC1 | 0.142583422 | -0.041241339 | 0.101342083 |
| ZBTB16 | 0.057802972 | -0.055265138 | 0.002537834 |
| KDM1A | 0.085743483 | -0.077153498 | 0.008589985 |
| CDC42EP2 | 0.044599304 | -0.045953094 | -0.00135379 |
| CRYL1 | 0.138634743 | -0.077541745 | 0.061092998 |

| | | | |
|---|---|---|---|
| GSTP1 | 0.144260353 | -0.075647163 | 0.06861319 |
| TM9SF2 | 0.122699716 | -0.113283477 | 0.00941624 |
| TMEM68 | 0.073181082 | -0.060009403 | 0.013171679 |
| TAPT1 | 0.067136736 | -0.045402951 | 0.021733785 |
| MTUS1 | 0.086714245 | -0.066019323 | 0.020694922 |
| TXNDC12 | 0.096272834 | -0.071376816 | 0.024896018 |
| FLOT1 | 0.100633116 | -0.073736656 | 0.02689646 |
| STX12 | 0.08237738 | -0.063794041 | 0.018583339 |
| DBI | 0.148259365 | -0.064209451 | 0.084049914 |
| INPP1 | 0.063796509 | -0.04084195 | 0.022954559 |
| IGSF8 | 0.081196233 | -0.052094645 | 0.029101589 |
| SORL1 | 0.120186995 | -0.10160305 | 0.018583945 |
| RPS4Y1 | 0.125339675 | -0.190630743 | -0.065291068 |
| LAMB1 | 0.10694455 | -0.092576922 | 0.014367628 |
| FXYD3 | 0.048376538 | -0.052187005 | -0.003810466 |
| HSPA5 | 0.127260753 | -0.070458121 | 0.056802632 |
| TUBA1C | 0.10280286 | -0.073163877 | 0.029638983 |
| TMEM179B | 0.119159395 | -0.057926266 | 0.061233128 |
| DNAJB11 | 0.078924815 | -0.026911243 | 0.052013572 |
| TMED2 | 0.112994903 | -0.048282349 | 0.064712554 |
| GNG5 | 0.138582977 | -0.065924782 | 0.072658196 |
| UQCRQ | 0.146178936 | -0.068867853 | 0.077311083 |
| NME2 | 0.137227373 | -0.039426779 | 0.097800594 |
| CHMP5 | 0.127966794 | -0.057770195 | 0.070196599 |
| TM2D1 | 0.113827709 | -0.057713643 | 0.056114067 |
| KIF13B | 0.080993777 | -0.076975252 | 0.004018525 |
| NTRK2 | -0.042346065 | 0.01779267 | -0.024553395 |
| SEC11C | 0.12449231 | -0.08148163 | 0.04301068 |
| ARMC7 | 0.07614836 | -0.04266493 | 0.033483431 |
| COPE | 0.139615316 | -0.039589099 | 0.100026217 |
| PHB | 0.131553875 | -0.033979598 | 0.097574277 |
| KIAA2026 | 0.081585825 | -0.074030074 | 0.007555751 |
| CTSL | 0.115277223 | -0.112503407 | 0.002773816 |
| SPSB3 | 0.09915347 | -0.083880352 | 0.015273118 |
| SEMA4B | 0.061234224 | -0.051891456 | 0.009342768 |
| H3F3A | 0.129294752 | -0.07507266 | 0.054222092 |
| EPS8L2 | 0.082625414 | -0.086313521 | -0.003688106 |
| STARD3 | 0.069564246 | -0.043099006 | 0.02646524 |
| PSMB6 | 0.131346102 | -0.046204839 | 0.085141263 |
| C18orf32 | 0.136739542 | -0.062793437 | 0.073946105 |
| TMEM54 | 0.073940434 | -0.049214932 | 0.024725501 |
| CALB1 | -0.086366305 | 0.020698668 | -0.065667637 |
| TFCP2L1 | 0.133833887 | -0.095936302 | 0.037897586 |
| TMEM9B | 0.112207597 | -0.08185099 | 0.030356606 |
| DPM3 | 0.129963058 | -0.099494405 | 0.030468653 |
| ANKRD22 | 0.088046298 | -0.072007736 | 0.016038562 |
| PLEKHJ1 | 0.124201587 | -0.041382068 | 0.082819518 |

| | | | |
|---|---|---|---|
| KMT2E | 0.106568418 | -0.074809667 | 0.031758751 |
| DNAJB6 | 0.115294465 | -0.061770535 | 0.05352393 |
| NMRK1 | 0.074998971 | -0.040007363 | 0.034991609 |
| TPM4 | 0.063985146 | -0.088182379 | -0.024197232 |
| YIPF4 | 0.100071557 | -0.075491396 | 0.024580161 |
| EMC7 | 0.111694219 | -0.070162469 | 0.04153175 |
| DHRS7B | 0.082690856 | -0.051750136 | 0.03094072 |
| SPINK7 | 0.04382566 | -0.04784208 | -0.00401642 |
| HNRNPH1 | 0.139309147 | -0.107662384 | 0.031646763 |
| TMEM147 | 0.118781704 | -0.057680793 | 0.061100912 |
| MCUR1 | 0.095710791 | -0.069743425 | 0.025967365 |
| FAM171A2 | 0.043697479 | -0.041892418 | 0.001805061 |
| SELENOT | 0.119479067 | -0.055185835 | 0.064293232 |
| RPL36 | 0.125397885 | -0.048687911 | 0.076709974 |
| PINK1 | 0.103017713 | -0.076804674 | 0.026213039 |
| FAM241A | 0.050748288 | -0.044120155 | 0.006628134 |
| ADRM1 | 0.09710809 | -0.047823551 | 0.049284539 |
| EGF | -0.082350911 | 0.033551014 | -0.048799897 |
| MTX2 | 0.114514938 | -0.04367329 | 0.070841649 |
| PRRG1 | 0.073452805 | -0.048360876 | 0.025091929 |
| SOD2 | 0.11363482 | -0.057754404 | 0.055880416 |
| GCC2 | 0.108018324 | -0.097422489 | 0.010595835 |
| ATP1B1 | 0.146356099 | -0.149578113 | -0.003222013 |
| PNN | 0.104428861 | -0.066612988 | 0.037815873 |
| GRB7 | 0.065163404 | -0.070676806 | -0.005513402 |
| COX14 | 0.125648027 | -0.068364638 | 0.057283389 |
| SLC38A1 | 0.108877021 | -0.075126455 | 0.033750567 |
| GGCT | 0.084480262 | -0.057044256 | 0.027436006 |
| OST4 | 0.137033434 | -0.053628322 | 0.083405112 |
| GOLGA4 | 0.10709594 | -0.074645898 | 0.032450041 |
| ALDH9A1 | 0.094094248 | -0.067243342 | 0.026850906 |
| SBDS | 0.109194157 | -0.059066075 | 0.050128082 |
| EIF5B | 0.105819904 | -0.111785317 | -0.005965413 |
| CLDN7 | 0.123001558 | -0.066058026 | 0.056943532 |
| DNAJB12 | 0.073048779 | -0.042518595 | 0.030530184 |
| PPP1R14B | 0.125659935 | -0.035235292 | 0.090424644 |
| STRN | 0.075671046 | -0.080669074 | -0.004998028 |
| SRGAP1 | 0.059505065 | -0.074375163 | -0.014870098 |
| LAP3 | 0.08757109 | -0.047235045 | 0.040336044 |
| MREG | 0.045446589 | -0.045409167 | 3.74215E-05 |
| SPINT1-AS1 | 0.118238438 | -0.045222093 | 0.073016345 |
| RPL37A | 0.124522799 | -0.037591097 | 0.086931702 |
| ANKRD12 | 0.100384611 | -0.089618959 | 0.010765653 |
| TMEM80 | 0.099755706 | -0.06556869 | 0.034187017 |
| EPRS | 0.083124386 | -0.077924696 | 0.00519969 |
| COPRS | 0.119485678 | -0.044465828 | 0.07501985 |
| SFRP1 | -0.047251116 | 0.020527923 | -0.026723194 |

| | | | |
|---|---|---|---|
| DPP7 | 0.117224724 | -0.050749075 | 0.066475648 |
| MICU2 | 0.096352372 | -0.031190267 | 0.065162105 |
| PSMB8 | 0.103708714 | -0.026605167 | 0.077103547 |
| SNRNP70 | 0.103429415 | -0.045907983 | 0.057521431 |
| TSPAN6 | 0.098625579 | -0.079121765 | 0.019503815 |
| SDF4 | 0.089425685 | -0.063727246 | 0.025698439 |
| MLX | 0.075033268 | -0.064283542 | 0.010749727 |
| AAK1 | 0.093263476 | -0.088912469 | 0.004351006 |
| PRRC2C | 0.107263971 | -0.079651351 | 0.02761262 |
| CNIH4 | 0.111973071 | -0.063660201 | 0.048312871 |
| DUSP9 | -0.052138802 | 0.026899956 | -0.025238846 |
| SUCLG2 | 0.100732441 | -0.065501092 | 0.035231349 |
| PCBD1 | 0.132656208 | -0.034637502 | 0.098018706 |
| CARD6 | 0.038272771 | -0.035126505 | 0.003146266 |
| PSMB5 | 0.117100529 | -0.04791254 | 0.069187989 |
| GPR155 | 0.060077979 | -0.055966091 | 0.004111888 |
| ATAD1 | 0.089735682 | -0.048347792 | 0.04138789 |
| LAMP1 | 0.108579893 | -0.095106409 | 0.013473485 |
| ERBB2 | 0.113037736 | -0.069872022 | 0.043165715 |
| LMAN2 | 0.113176537 | -0.063287078 | 0.049889459 |
| PLGRKT | 0.089477484 | -0.045490681 | 0.043986803 |
| DUT | 0.119146209 | -0.072613081 | 0.046533128 |
| CHCHD10 | 0.126796375 | -0.027818516 | 0.098977859 |
| TMX1 | 0.072431037 | -0.057110152 | 0.015320885 |
| ACSL5 | 0.046341583 | -0.037474523 | 0.00886706 |
| PSMB2 | 0.103969783 | -0.03588545 | 0.068084333 |
| TWSG1 | 0.068988868 | -0.054029678 | 0.01495919 |
| FBXO21 | 0.103063441 | -0.071115804 | 0.031947636 |
| CD302 | 0.038204145 | -0.040319451 | -0.002115306 |
| COX7A2 | 0.133469245 | -0.058327824 | 0.07514142 |
| EPS8L1 | 0.062893095 | -0.048860194 | 0.0140329 |
| TUBG1 | 0.063275221 | -0.03291277 | 0.030362451 |
| HSPA4 | 0.098321044 | -0.082988318 | 0.015332726 |
| PSMD8 | 0.122660202 | -0.054639381 | 0.06802082 |
| SNHG14 | 0.115975902 | -0.091660678 | 0.024315224 |
| MAF1 | 0.100211022 | -0.067681664 | 0.032529358 |
| OSTC | 0.107837004 | -0.043906446 | 0.063930558 |
| BCL6 | 0.054900453 | -0.063987997 | -0.009087544 |
| STAP1 | 0.104027037 | -0.032789576 | 0.071237462 |
| PLSCR1 | 0.08226335 | -0.065015011 | 0.017248339 |
| RPS15 | 0.123121834 | -0.051111222 | 0.072010612 |
| MRPL33 | 0.130940235 | -0.116210108 | 0.014730127 |
| FAM102A | 0.062523435 | -0.050319813 | 0.012203622 |
| NUB1 | 0.079893205 | -0.066107208 | 0.013785997 |
| M6PR | 0.093555256 | -0.06644424 | 0.027111016 |
| BCAM | 0.123476755 | -0.066944835 | 0.05653192 |
| SLC4A1 | 0.186957682 | -0.097502675 | 0.089455007 |

| | | | |
|---|---|---|---|
| TMEM14C | 0.126197032 | -0.067371426 | 0.058825607 |
| P4HTM | 0.090215037 | -0.023787846 | 0.066427191 |
| ST14 | 0.102558965 | -0.059145585 | 0.043413381 |
| FRG1 | 0.086000663 | -0.043348374 | 0.042652288 |
| NUPR1 | 0.044253291 | -0.044050005 | 0.000203287 |
| GALNT7 | 0.071298631 | -0.064777759 | 0.006520873 |
| LPCAT3 | 0.075633606 | -0.050960419 | 0.024673187 |
| RXYLT1 | 0.079344005 | -0.06207311 | 0.017270895 |
| FYB2 | 0.072769487 | -0.063540621 | 0.009228866 |
| HOOK2 | 0.078676375 | -0.06581786 | 0.012858515 |
| NDUFV2 | 0.133722544 | -0.037091221 | 0.096631323 |
| ATG101 | 0.085753914 | -0.036979515 | 0.048774399 |
| RAB5IF | 0.097854622 | -0.058352226 | 0.039502396 |
| MAP2K2 | 0.104573532 | -0.052873864 | 0.051699669 |
| GORASP2 | 0.072837698 | -0.073062279 | -0.000224581 |
| DDOST | 0.091984941 | -0.061826781 | 0.03015816 |
| CRTAP | 0.085329675 | -0.050792753 | 0.034536923 |
| YWHAE | 0.129396271 | -0.040585718 | 0.088810553 |
| P4HB | 0.099682656 | -0.078439618 | 0.021243038 |
| PCYOX1 | 0.100540065 | -0.068892628 | 0.031647437 |
| LYPLAL1 | 0.104937242 | -0.053488364 | 0.051448878 |
| PDCD5 | 0.111162536 | -0.058496131 | 0.052666405 |
| OCIAD1 | 0.1227984 | -0.054269133 | 0.068529267 |
| C19orf33 | 0.047984355 | -0.053247851 | -0.005263496 |
| PPP1R9A | 0.069759706 | -0.070605992 | -0.000846286 |
| TP53I11 | 0.076580258 | -0.057202131 | 0.019378127 |
| RACK1 | 0.13196328 | -0.041708478 | 0.090254803 |
| ITSN2 | 0.076798504 | -0.055874529 | 0.020923975 |
| RPL7A | 0.129460856 | -0.033394265 | 0.096066591 |
| MGAT4B | 0.088083843 | -0.040075402 | 0.048008441 |
| TBX2 | 0.079108993 | -0.080966782 | -0.00185779 |
| PRKDC | 0.104757095 | -0.09574091 | 0.009016185 |
| CASC4 | 0.092636943 | -0.067876757 | 0.024760186 |
| PDCD6 | 0.104816121 | -0.052274835 | 0.052541286 |
| RNF213 | 0.075849394 | -0.07824177 | -0.002392376 |
| CLIP1 | 0.097950362 | -0.047155243 | 0.050795119 |
| PIK3C2G | 0.062737838 | -0.081298918 | -0.01856108 |
| ITGA6 | 0.130832884 | -0.05506832 | 0.075764564 |
| KIF12 | 0.10024994 | -0.091349358 | 0.008900581 |
| SCAF11 | 0.104691692 | -0.070887797 | 0.033803895 |
| NDUFA8 | 0.121188408 | -0.035288734 | 0.085899675 |
| MYL6 | 0.121495158 | -0.069430618 | 0.052064541 |
| IMMT | 0.101157324 | -0.05869164 | 0.042465684 |
| RBPMS | 0.094001665 | -0.068047733 | 0.025953932 |
| SELENOM | 0.131262501 | -0.09251419 | 0.038748311 |
| IL27RA | 0.033949729 | -0.028606681 | 0.005343048 |
| SINHCAF | 0.084354462 | -0.076121027 | 0.008233435 |

| | | | |
|---|---|---|---|
| SYNGR1 | 0.060785252 | -0.039911151 | 0.020874101 |
| MARCH5 | 0.07811385 | -0.050402418 | 0.027711432 |
| IAH1 | 0.099692247 | -0.065933494 | 0.033758753 |
| SAR1B | 0.088111647 | -0.038339391 | 0.049772255 |
| OXGR1 | 0.081703089 | -0.042005087 | 0.039698002 |
| PCCB | 0.070602486 | -0.047021202 | 0.023581285 |
| HSBP1L1 | 0.081891078 | -0.064027636 | 0.017863442 |
| PTGES2 | 0.094905707 | -0.051119477 | 0.04378623 |
| SPATS2 | 0.049363978 | -0.037017617 | 0.012346361 |
| VAMP8 | 0.128720354 | -0.040013308 | 0.088707046 |
| NDUFA1 | 0.123638558 | -0.05875008 | 0.064888479 |
| ZBTB20 | 0.115857748 | -0.098201179 | 0.017656568 |
| DCTN2 | 0.092895027 | -0.049470248 | 0.043424779 |
| MICU1 | 0.069575451 | -0.052930728 | 0.016644723 |
| C1GALT1C1 | 0.078254141 | -0.041626772 | 0.03662737 |
| ELOVL1 | 0.07383447 | -0.058248244 | 0.015586225 |
| PPARG | 0.062148288 | -0.051047462 | 0.011100826 |
| SSR3 | 0.107456563 | -0.070170796 | 0.037285767 |
| SUCLG1 | 0.125539433 | -0.054300814 | 0.071238619 |
| DLG1 | 0.077700188 | -0.072944237 | 0.004755951 |
| TRAM1 | 0.109884143 | -0.098279173 | 0.01160497 |
| UBE2D2 | 0.103366603 | -0.050592076 | 0.052774527 |
| ADTRP | 0.115131652 | -0.08651455 | 0.028617102 |
| TAOK3 | 0.07934486 | -0.099604909 | -0.020260048 |
| SURF1 | 0.106657301 | -0.054659166 | 0.051998135 |
| DNAJC15 | 0.101292166 | -0.069858492 | 0.031433674 |
| ILVBL | 0.076051525 | -0.054685837 | 0.021365688 |
| TMOD3 | 0.081275838 | -0.050859488 | 0.030416351 |
| CAPN1 | 0.084961318 | -0.065602336 | 0.019358982 |
| ABCC3 | 0.04597223 | -0.048615868 | -0.002643638 |
| TMEM242 | 0.072687161 | -0.043412749 | 0.029274412 |
| SGPL1 | 0.06480719 | -0.050540212 | 0.014266978 |
| EIF5 | 0.107757318 | -0.083098735 | 0.024658584 |
| ERGIC2 | 0.092636266 | -0.043104852 | 0.049531414 |
| MYH10 | 0.066609213 | -0.065669315 | 0.000939898 |
| ATRX | 0.106306299 | -0.072059684 | 0.034246614 |
| LEPROT | 0.10199511 | -0.080043685 | 0.021951424 |
| TUBA4A | 0.097936278 | -0.022644819 | 0.075291459 |
| MPHOSPH8 | 0.096125531 | -0.09364969 | 0.002475841 |
| HSPA8 | 0.131535758 | -0.055348954 | 0.076186804 |
| MIR200CHG | 0.044729856 | -0.030002808 | 0.014727048 |
| PWRN1 | 0.036808452 | -0.038541796 | -0.001733344 |
| COL12A1 | 0.031599107 | -0.033408774 | -0.001809667 |
| INSIG2 | 0.078949432 | -0.0655183 | 0.013431132 |
| RARRES2 | 0.141756935 | -0.144996822 | -0.003239887 |
| LIMS1 | 0.103672436 | -0.06739994 | 0.036272496 |
| REL | 0.048616726 | -0.041235944 | 0.007380782 |

| | | | |
|---|---|---|---|
| NDUFA9 | 0.103493786 | -0.035974964 | 0.067518822 |
| SEL1L | 0.091999394 | -0.065894072 | 0.026105322 |
| APLP2 | 0.12677565 | -0.116176071 | 0.010599579 |
| XRN2 | 0.09286131 | -0.072564218 | 0.020297092 |
| LLGL2 | 0.093081567 | -0.060388106 | 0.032693461 |
| NDUFB11 | 0.127696444 | -0.054795901 | 0.072900543 |
| TMCO4 | 0.079127497 | -0.060792647 | 0.01833485 |
| PMF1 | 0.097640927 | -0.049393423 | 0.048247504 |
| DNAJC21 | 0.085990644 | -0.061419669 | 0.024570976 |
| MGLL | 0.088640307 | -0.078243878 | 0.01039643 |
| UQCR11 | 0.121566007 | -0.058284601 | 0.063281406 |
| SNAPC1 | 0.056519701 | -0.048498038 | 0.008021663 |
| AFAP1L2 | 0.036397476 | -0.041260992 | -0.004863516 |
| SNX2 | 0.091173965 | -0.040488706 | 0.050685259 |
| CHCHD3 | 0.079095889 | -0.043478939 | 0.03561695 |
| METTL5 | 0.094250249 | -0.035227988 | 0.059022261 |
| TSPAN15 | 0.058239863 | -0.057847603 | 0.00039226 |
| SDCBP | 0.085355813 | -0.065814698 | 0.019541116 |
| HNRNPDL | 0.11209061 | -0.070998633 | 0.041091977 |
| SMAD9 | 0.033627953 | -0.034945984 | -0.001318031 |
| OMA1 | 0.067850061 | -0.041871471 | 0.025978591 |
| CD55 | 0.074068245 | -0.069320901 | 0.004747344 |
| CASR | -0.029451878 | 0.010817541 | -0.018634338 |
| TXNL4A | 0.103388187 | -0.06457158 | 0.038816607 |
| PSMA5 | 0.099054244 | -0.038554067 | 0.060500177 |
| CCDC107 | 0.086148349 | -0.076657062 | 0.009491287 |
| SDF2L1 | 0.095667374 | -0.033224706 | 0.062442668 |
| PMEPA1 | 0.057341909 | -0.025121315 | 0.032220594 |
| ATP5MF | 0.124013526 | -0.046816976 | 0.077196551 |
| PDCD4 | 0.073955715 | -0.052066394 | 0.021889321 |
| TMEM14B | 0.122104555 | -0.046446675 | 0.07565788 |
| FAM162A | 0.123129922 | -0.051486758 | 0.071643164 |
| BUD23 | 0.097507466 | -0.063122138 | 0.034385328 |
| PIGK | 0.068844563 | -0.060314969 | 0.008529594 |
| RETREG1 | 0.085941196 | -0.081103526 | 0.00483767 |
| B3GAT3 | 0.085747043 | -0.061088232 | 0.024658811 |
| N4BP2L2 | 0.119064599 | -0.077646052 | 0.041418547 |
| CDK5RAP3 | 0.080109216 | -0.049583287 | 0.030525929 |
| MRPS26 | 0.107466735 | -0.057148728 | 0.050318007 |
| TMEM41B | 0.069572234 | -0.035697639 | 0.033874594 |
| ACOT2 | 0.084974162 | -0.037935622 | 0.047038539 |
| CDC42 | 0.118774549 | -0.041063674 | 0.077710876 |
| MUC1 | 0.122434974 | -0.107196934 | 0.01523804 |
| ACTG1 | 0.121361184 | -0.042956861 | 0.078404323 |
| NDUFB8 | 0.126374423 | -0.048431965 | 0.077942458 |
| MON2 | 0.06771574 | -0.063423709 | 0.004292031 |
| HNRNPA2B1 | 0.118148385 | -0.07504996 | 0.043098426 |

| | | | |
|---|---|---|---|
| RNF13 | 0.087184587 | -0.035268608 | 0.051915979 |
| RNF121 | 0.057411644 | -0.039438867 | 0.017972777 |
| SPG11 | 0.07134892 | -0.047907706 | 0.023441214 |
| ITIH5 | 0.112416863 | -0.026951445 | 0.085465417 |
| SLC35B1 | 0.077443121 | -0.043839949 | 0.033603172 |
| BAZ2B | 0.078439926 | -0.080021621 | -0.001581695 |
| STUB1 | 0.106414601 | -0.060545385 | 0.045869216 |
| UBAC1 | 0.085330125 | -0.065580813 | 0.019749312 |
| COMT | 0.103585469 | -0.075866323 | 0.027719146 |
| TMEM238 | 0.108491004 | -0.074525088 | 0.033965916 |
| UQCRC2 | 0.11618107 | -0.046799685 | 0.069381385 |
| BNIP3L | 0.065991036 | -0.059954322 | 0.006036714 |
| PNISR | 0.115915433 | -0.071746664 | 0.044168769 |
| GSDMD | 0.07631855 | -0.051838306 | 0.024480245 |
| FHL2 | 0.027062571 | -0.028989729 | -0.001927157 |
| ZSWIM7 | 0.083388068 | -0.057188878 | 0.02619919 |
| HIST1H1C | 0.079805854 | -0.078702849 | 0.001103006 |
| AC009244.2 | 0.025790212 | -0.026891727 | -0.001101515 |
| TMEM259 | 0.088496541 | -0.068163834 | 0.020332707 |
| THRAP3 | 0.090405282 | -0.054896464 | 0.035508818 |
| TMEM208 | 0.102635918 | -0.060353449 | 0.042282469 |
| SLC4A7 | 0.043602011 | -0.045339675 | -0.001737664 |
| RHOA | 0.11051813 | -0.075919309 | 0.034598821 |
| ARPC1A | 0.102913982 | -0.072758895 | 0.030155088 |
| SEC63 | 0.06983689 | -0.063458328 | 0.006378562 |
| TCIRG1 | 0.051621631 | -0.044098196 | 0.007523436 |
| ANKRD17 | 0.069467832 | -0.045218111 | 0.024249721 |
| PARD6B | 0.092310577 | -0.072054066 | 0.020256511 |
| CAB39L | 0.070264342 | -0.080806187 | -0.010541845 |
| TPI1 | 0.12717558 | -0.07323994 | 0.05393564 |
| RPS21 | 0.112574385 | -0.09001275 | 0.022561635 |
| KCNJ13 | 0.052205832 | -0.04106199 | 0.011143842 |
| RSRP1 | 0.109530153 | -0.059361061 | 0.050169092 |
| XPO1 | 0.076073627 | -0.049579957 | 0.02649367 |
| SUMO2 | 0.12410162 | -0.066701783 | 0.057399837 |
| NECTIN2 | 0.081309136 | -0.048711782 | 0.032597354 |
| IARS2 | 0.086053852 | -0.088027677 | -0.001973826 |
| DNAJB9 | 0.055994132 | -0.038845016 | 0.017149116 |
| PET100 | 0.117537585 | -0.087359145 | 0.03017844 |
| PSMD11 | 0.08199727 | -0.047041697 | 0.034955572 |
| PAFAH1B1 | 0.094737677 | -0.043479885 | 0.051257791 |
| CALM1 | 0.117283274 | -0.130639539 | -0.013356265 |
| ADAM10 | 0.068094536 | -0.045599903 | 0.022494633 |
| RCAN3 | 0.068869611 | -0.057106816 | 0.011762795 |
| RAB2A | 0.105262767 | -0.053784145 | 0.051478622 |
| PRR15 | 0.038444902 | -0.040030175 | -0.001585273 |
| CRYBG3 | 0.085523139 | -0.059977923 | 0.025545216 |

| | | | |
|---|---|---|---|
| CD2AP | 0.085504537 | -0.074622631 | 0.010881906 |
| TKT | 0.08598658 | -0.056100598 | 0.029885982 |
| RPS27 | 0.115205779 | -0.07302829 | 0.042177488 |
| ATP2A2 | 0.077378337 | -0.056716451 | 0.020661886 |
| RNASEK | 0.129127599 | -0.072374792 | 0.056752807 |
| LAMTOR1 | 0.10874979 | -0.06231558 | 0.04643421 |
| NUDT16L1 | 0.094405808 | -0.035179107 | 0.059226701 |
| AP1G1 | 0.075562883 | -0.05806308 | 0.017499802 |
| TPR | 0.093199665 | -0.078286075 | 0.01491359 |
| RPL30 | 0.119336254 | -0.060719357 | 0.058616897 |
| SEC11A | 0.122809854 | -0.050252339 | 0.072557514 |
| BCL7B | 0.069247053 | -0.051602646 | 0.017644408 |
| SUSD1 | 0.069425284 | -0.057037944 | 0.01238734 |
| RPLP2 | 0.116880025 | -0.065665872 | 0.051214153 |
| NAP1L4 | 0.09923458 | -0.061412779 | 0.037821801 |
| MDH2 | 0.118073622 | -0.052400101 | 0.065673521 |
| VPS35 | 0.085022782 | -0.062258634 | 0.022764148 |
| SYAP1 | 0.089585762 | -0.037533568 | 0.052052194 |
| NENF | 0.104435509 | -0.067252031 | 0.037183478 |
| DDX46 | 0.08335051 | -0.053225927 | 0.030124583 |
| VDAC3 | 0.101131427 | -0.049315477 | 0.051815949 |
| PSME1 | 0.10681259 | -0.070353688 | 0.036458901 |
| SEPTIN7 | 0.094714942 | -0.080452277 | 0.014262665 |
| CLK1 | 0.078915294 | -0.051288098 | 0.027627196 |
| SCP2 | 0.119636815 | -0.037631697 | 0.082005118 |
| CREBRF | 0.079433229 | -0.066807219 | 0.01262601 |
| LRBA | 0.081139577 | -0.06942232 | 0.011717256 |
| HDAC1 | 0.088008928 | -0.050838074 | 0.037170854 |
| PPP4R2 | 0.077183817 | -0.050558198 | 0.026625619 |
| NAA38 | 0.111458192 | -0.072692473 | 0.038765719 |
| NDUFA12 | 0.126202732 | -0.052067808 | 0.074134924 |
| DDX21 | 0.076501583 | -0.059168524 | 0.01733306 |
| MAEA | 0.074180422 | -0.043248475 | 0.030931947 |
| TM7SF3 | 0.066317684 | -0.065708074 | 0.00060961 |
| PIK3IP1 | 0.05220105 | -0.036077946 | 0.016123105 |
| LARP4 | 0.088909441 | -0.047997983 | 0.040911458 |
| ZC3H15 | 0.09018157 | -0.051082588 | 0.039098982 |
| ASAH1 | 0.103093336 | -0.106635484 | -0.003542148 |
| PSMD2 | 0.080093862 | -0.042989182 | 0.037104681 |
| GADD45G | 0.106695815 | -0.05306798 | 0.053627835 |
| TMEM258 | 0.118183842 | -0.059600997 | 0.058582844 |
| AKT1 | 0.073709642 | -0.045664563 | 0.02804508 |
| SLC35A3 | 0.06837307 | -0.052663997 | 0.015709074 |
| GCA | 0.071628056 | -0.025866074 | 0.045761982 |
| NPEPL1 | 0.068331385 | -0.039006959 | 0.029324426 |
| POR | 0.05284245 | -0.036454254 | 0.016388196 |
| MET | 0.072351206 | -0.045128418 | 0.027222788 |

| | | | |
|---|---|---|---|
| JUND | 0.120740202 | -0.077868236 | 0.042871966 |
| GNPTG | 0.078837546 | -0.047174499 | 0.031663047 |
| GNPTAB | 0.070893082 | -0.062187004 | 0.008706078 |
| DENND2D | 0.066229931 | -0.054504934 | 0.011724997 |
| DNAJC3 | 0.090250598 | -0.093996766 | -0.003746168 |
| SLC52A3 | 0.049250757 | -0.054093147 | -0.004842391 |
| AGTRAP | 0.054736038 | -0.029034501 | 0.025701536 |
| RDH14 | 0.074626443 | -0.045597858 | 0.029028585 |
| MTLN | 0.101442514 | -0.088092762 | 0.013349752 |
| LRPPRC | 0.092943089 | -0.048933767 | 0.044009321 |
| SCAMP1 | 0.083125008 | -0.050088852 | 0.033036156 |
| MYL12B | 0.127326535 | -0.065067355 | 0.06225918 |
| CMPK1 | 0.086470225 | -0.040530067 | 0.045940158 |
| PRKAG2 | 0.0773174 | -0.044251195 | 0.033066206 |
| CCT7 | 0.093875576 | -0.034821574 | 0.059054002 |
| CCT5 | 0.092122215 | -0.051349841 | 0.040772374 |
| SMARCB1 | 0.071459453 | -0.048876761 | 0.022582692 |
| METTL7A | 0.094229699 | -0.090007485 | 0.004222214 |
| BAD | 0.089430748 | -0.024342086 | 0.065088662 |
| PPP1R1A | -0.058480439 | 0.052536381 | -0.005944057 |
| FAM3C | 0.103341696 | -0.076908353 | 0.026433343 |
| PHB2 | 0.106598445 | -0.058831309 | 0.047767136 |
| G3BP2 | 0.080058899 | -0.040662017 | 0.039396882 |
| RPS19BP1 | 0.100562863 | -0.048893198 | 0.051669665 |
| PSME2 | 0.105778361 | -0.033944012 | 0.071834349 |
| MCCC1 | 0.068241418 | -0.070683615 | -0.002442197 |
| SFT2D1 | 0.103480796 | -0.058812917 | 0.044667879 |
| FAM131C | 0.061373079 | -0.053995751 | 0.007377328 |
| ZFYVE21 | 0.090238034 | -0.038857184 | 0.05138085 |
| B3GNT3 | 0.02433008 | -0.023919001 | 0.000411079 |
| PTPRF | 0.083359023 | -0.06397621 | 0.019382812 |
| HP1BP3 | 0.107042136 | -0.076164356 | 0.03087778 |
| NANS | 0.069534483 | -0.037512913 | 0.03202157 |
| MIGA1 | 0.073030833 | -0.055712962 | 0.017317871 |
| EIF3K | 0.123466619 | -0.034985092 | 0.088481527 |
| ADI1 | 0.115853238 | -0.037652963 | 0.078200275 |
| ANAPC11 | 0.108867023 | -0.070594066 | 0.038272958 |
| ETF1 | 0.076663029 | -0.063456289 | 0.013206741 |
| CAPN2 | 0.107542254 | -0.040937408 | 0.066604846 |
| TSPAN14 | 0.087158102 | -0.045562115 | 0.041595988 |
| MLEC | 0.095955233 | -0.054594201 | 0.041361032 |
| ABHD12 | 0.07801487 | -0.065386783 | 0.012628086 |
| PHAX | 0.067808149 | -0.051133999 | 0.01667415 |
| HNRNPU | 0.106307006 | -0.096990703 | 0.009316304 |
| HEXA | 0.099086384 | -0.025224108 | 0.073862276 |
| NDUFA11 | 0.116893107 | -0.052809224 | 0.064083884 |
| METTL27 | 0.044541609 | -0.032611762 | 0.011929847 |

| | | | |
|---|---|---|---|
| RPS7 | 0.115378644 | -0.038783982 | 0.076594663 |
| COX6B1 | 0.112367463 | -0.039852708 | 0.072514755 |
| ADIPOR2 | 0.076601716 | -0.052645294 | 0.023956422 |
| OPTN | 0.066654445 | -0.029148887 | 0.037505558 |
| SELENOP | 0.122427639 | -0.135164062 | -0.012736423 |
| SLC22A5 | 0.069562154 | -0.059713302 | 0.009848852 |
| ATP5F1E | 0.10952947 | -0.036340946 | 0.073188524 |
| RHBG | 0.114422424 | -0.068108361 | 0.046314063 |
| ARL3 | 0.0852654 | -0.058290766 | 0.026974634 |
| AKR1B1 | 0.083822249 | -0.081364643 | 0.002457606 |
| PRPF40A | 0.085506529 | -0.052118997 | 0.033387531 |
| DNASE2 | 0.075582865 | -0.044791006 | 0.030791859 |
| PAX2 | 0.079663541 | -0.088128237 | -0.008464696 |
| RBCK1 | 0.084232736 | -0.043364686 | 0.04086805 |
| LPP | 0.095477275 | -0.085679801 | 0.009797475 |
| TMTC1 | -0.051458355 | 0.021424935 | -0.030033419 |
| RPS19 | 0.115933637 | -0.03501712 | 0.080916517 |
| SDF2 | 0.080265649 | -0.042488874 | 0.037776775 |
| GGT6 | 0.063962235 | -0.064893831 | -0.000931596 |
| OAZ1 | 0.116921923 | -0.035737552 | 0.081184371 |
| RNF115 | 0.068284058 | -0.041181522 | 0.027102536 |
| RER1 | 0.10618287 | -0.047556758 | 0.058626112 |
| NGRN | 0.100381342 | -0.040292849 | 0.060088493 |
| ABHD16A | 0.067584055 | -0.059489826 | 0.008094229 |
| FAM200B | 0.099038776 | -0.060526003 | 0.038512773 |
| NEMF | 0.080552815 | -0.067007942 | 0.013544873 |
| CASP4 | 0.046679804 | -0.035044027 | 0.011635777 |
| DEK | 0.086886048 | -0.04858955 | 0.038296498 |
| CENPC | 0.077222922 | -0.067247119 | 0.009975803 |
| TDGF1 | -0.041610195 | 0.018119725 | -0.02349047 |
| EMC10 | 0.099512771 | -0.06788312 | 0.031629651 |
| SPATA13 | 0.063567113 | -0.057314022 | 0.00625309 |
| XRCC5 | 0.095554968 | -0.08834613 | 0.007208838 |
| FLII | 0.06205283 | -0.045295371 | 0.016757459 |
| COMMD1 | 0.077905139 | -0.036420948 | 0.04148419 |
| ARL1 | 0.096819843 | -0.043258307 | 0.053561536 |
| CDC42BPB | 0.077289502 | -0.043226657 | 0.034062845 |
| HNRNPA3 | 0.104556381 | -0.069948224 | 0.034608158 |
| TAX1BP1 | 0.102300068 | -0.046603258 | 0.05569681 |
| SEMA4D | 0.059151781 | -0.062550033 | -0.003398252 |
| USP15 | 0.059848298 | -0.050124433 | 0.009723864 |
| TAPBP | 0.078472769 | -0.055207198 | 0.023265571 |
| PSMG2 | 0.092671852 | -0.032808692 | 0.05986316 |
| VCP | 0.08339191 | -0.050796719 | 0.032595191 |
| MACF1 | 0.101116352 | -0.092864487 | 0.008251865 |
| CPEB4 | 0.097161122 | -0.04430305 | 0.052858072 |
| FAM3A | 0.069228807 | -0.043260418 | 0.025968389 |

| | | | |
|---|---|---|---|
| EMC4 | 0.082704472 | -0.052205333 | 0.030499139 |
| TRAPPC4 | 0.084150967 | -0.034682091 | 0.049468876 |
| SYNRG | 0.065840093 | -0.052917128 | 0.012922964 |
| ATG5 | 0.066544218 | -0.052142906 | 0.014401312 |
| TCEA1 | 0.091296872 | -0.061785166 | 0.029511706 |
| MAPK10 | 0.058736488 | -0.070012637 | -0.011276149 |
| CD99 | 0.064812514 | -0.054063543 | 0.010748971 |
| PRELID1 | 0.100283793 | -0.027087231 | 0.073196563 |
| POU3F4 | 0.037610085 | -0.044049752 | -0.006439668 |
| GTF3C6 | 0.093960694 | -0.071322104 | 0.022638591 |
| COX6C | 0.110959022 | -0.028725535 | 0.082233487 |
| PAPSS2 | 0.035936362 | -0.039777448 | -0.003841086 |
| NRAD1 | 0.031386694 | -0.030906443 | 0.000480252 |
| COA1 | 0.080563335 | -0.041979094 | 0.038584241 |
| AKAP8L | 0.06903866 | -0.044155489 | 0.02488317 |
| JMJD1C | 0.075567632 | -0.048776611 | 0.026791021 |
| NDUFC1 | 0.117854458 | -0.05910161 | 0.058752848 |
| GOPC | 0.064228087 | -0.050944382 | 0.013283705 |
| ARHGAP21 | 0.067545644 | -0.041452807 | 0.026092837 |
| VDAC2 | 0.113944856 | -0.0289681 | 0.084976757 |
| ACAD8 | 0.070756675 | -0.033670238 | 0.037086436 |
| FNBP1L | 0.093607567 | -0.065237263 | 0.028370304 |
| AHI1 | 0.067894695 | -0.060520953 | 0.007373742 |
| ADH5 | 0.093185343 | -0.050298469 | 0.042886874 |
| HECTD1 | 0.091353909 | -0.0697103 | 0.021643609 |
| FEZ2 | 0.069312156 | -0.033359282 | 0.035952874 |
| PERP | 0.098172417 | -0.073028777 | 0.025143639 |
| DERL2 | 0.079067908 | -0.041143634 | 0.037924274 |
| MAGI1 | 0.096116256 | -0.063302832 | 0.032813424 |
| TMPO | 0.066506774 | -0.051528384 | 0.01497839 |
| GLT8D1 | 0.060341568 | -0.039979851 | 0.020361717 |
| ITGB1BP1 | 0.090905946 | -0.066838575 | 0.024067371 |
| PGAM1 | 0.097924255 | -0.039645975 | 0.05827828 |
| SDC1 | 0.064467349 | -0.0526294 | 0.01183795 |
| HIBCH | 0.085903232 | -0.03040494 | 0.055498292 |
| ERMP1 | 0.071418152 | -0.075687509 | -0.004269357 |
| CNGA1 | 0.039397344 | -0.028622099 | 0.010775245 |
| PATJ | 0.079201178 | -0.087745461 | -0.008544283 |
| MTURN | 0.090075262 | -0.081383547 | 0.008691715 |
| ADIPOR1 | 0.076402001 | -0.032017437 | 0.044384564 |
| DNAJC1 | 0.064640352 | -0.047406992 | 0.017233359 |
| YWHAB | 0.109537113 | -0.055798575 | 0.053738538 |
| SSR1 | 0.081710043 | -0.06345431 | 0.018255733 |
| DHPS | 0.076517439 | -0.050448395 | 0.026069044 |
| PPM1G | 0.081124941 | -0.06286474 | 0.018260201 |
| RWDD1 | 0.090003172 | -0.056555168 | 0.033448003 |
| CD59 | 0.114153638 | -0.035701858 | 0.07845178 |

| | | | |
|---|---|---|---|
| ZDHHC9 | 0.053567854 | -0.036077319 | 0.017490535 |
| FBXO2 | 0.044752973 | -0.035589343 | 0.00916363 |
| MTG1 | 0.063778293 | -0.045791871 | 0.017986422 |
| ARHGEF28 | 0.076164863 | -0.066778659 | 0.009386204 |
| RAPH1 | 0.041842308 | -0.026543331 | 0.015298978 |
| PDE11A | 0.031444145 | -0.030761154 | 0.000682991 |
| C11orf80 | 0.061386202 | -0.061327565 | 5.86366E-05 |
| IPCEF1 | 0.046136837 | -0.027665045 | 0.018471791 |
| BST2 | 0.086714647 | -0.030071831 | 0.056642816 |
| SELENOS | 0.097418856 | -0.069389306 | 0.02802955 |
| RAN | 0.111040222 | -0.030604311 | 0.080435911 |
| PPP4R1 | 0.053811041 | -0.041608023 | 0.012203019 |
| ACO2 | 0.096233335 | -0.074724086 | 0.021509248 |
| LRRK2 | 0.039210664 | -0.046721086 | -0.007510422 |
| TJP3 | 0.047279595 | -0.04596961 | 0.001309985 |
| MTFP1 | 0.059694845 | -0.034224375 | 0.025470471 |
| RBM17 | 0.078939693 | -0.05992616 | 0.019013533 |
| SLC25A29 | 0.068301667 | -0.074216117 | -0.005914451 |
| FER1L6 | 0.024671581 | -0.025077791 | -0.00040621 |
| CHST15 | 0.023015267 | -0.022809658 | 0.000205608 |
| DTYMK | 0.061684156 | -0.040689079 | 0.020995077 |
| EIF4A3 | 0.071672066 | -0.049885152 | 0.021786914 |
| PSMA6 | 0.09232793 | -0.050206133 | 0.042121798 |
| PON2 | 0.067373768 | -0.051484545 | 0.015889223 |
| OSBPL8 | 0.069216999 | -0.042714519 | 0.02650248 |
| SDHD | 0.114676184 | -0.043347581 | 0.071328603 |
| JPT1 | 0.091690762 | -0.042063283 | 0.049627479 |
| CCT3 | 0.094628033 | -0.05393154 | 0.040696493 |
| HMGB1 | 0.112575112 | -0.043383459 | 0.069191653 |
| GTF3C3 | 0.050172347 | -0.025814017 | 0.02435833 |
| TMEM183A | 0.06662043 | -0.03333511 | 0.03328532 |
| PLEKHA1 | 0.069593774 | -0.043241724 | 0.02635205 |
| PMPCB | 0.097080222 | -0.031979469 | 0.065100752 |
| PKN2 | 0.08476206 | -0.076275885 | 0.008486174 |
| ZFHX3 | 0.064110081 | -0.061286442 | 0.002823639 |
| TMEM260 | 0.051381221 | -0.036625366 | 0.014755855 |
| PSMB1 | 0.110360105 | -0.037115672 | 0.073244433 |
| HK1 | 0.053484354 | -0.048759494 | 0.004724861 |
| ASPH | 0.080084563 | -0.099297493 | -0.01921293 |
| INPP5F | 0.04266622 | -0.039833229 | 0.002832991 |
| AGFG1 | 0.067904412 | -0.045645983 | 0.022258429 |
| MARCH6 | 0.086744694 | -0.052294027 | 0.034450667 |
| EIF2S2 | 0.085446777 | -0.058582502 | 0.026864275 |
| SYT10 | 0.067607804 | -0.068561566 | -0.000953763 |
| PNPLA8 | 0.093517042 | -0.026919396 | 0.066597646 |
| LMNA | 0.08085882 | -0.048101702 | 0.032757117 |
| CMTM6 | 0.07750431 | -0.053800025 | 0.023704285 |

| | | | |
|---|---|---|---|
| TBX3 | 0.04495719 | -0.062047985 | -0.017090795 |
| LACTB | 0.059872043 | -0.04923594 | 0.010636103 |
| DERA | 0.078866409 | -0.058956334 | 0.019910075 |
| KDELR2 | 0.079700603 | -0.040245341 | 0.039455262 |
| RBM4 | 0.091398008 | -0.040298437 | 0.051099572 |
| KDM2A | 0.056783092 | -0.05053363 | 0.006249462 |
| PFKM | 0.059730506 | -0.04761595 | 0.012114556 |
| ERP44 | 0.075813083 | -0.023336131 | 0.052476952 |
| NDUFB2 | 0.105561267 | -0.039237738 | 0.066323529 |
| RPLP1 | 0.096814561 | -0.060023735 | 0.036790826 |
| WASL | 0.089984863 | -0.073324128 | 0.016660735 |
| UFC1 | 0.100104273 | -0.057291162 | 0.04281311 |
| PHF1 | 0.068712961 | -0.049635199 | 0.019077762 |
| MIF4GD | 0.05394317 | -0.030357202 | 0.023585967 |
| ITGA9 | 0.021878016 | -0.022452418 | -0.000574403 |
| CREG1 | 0.075790126 | -0.055640437 | 0.02014969 |
| NEU1 | 0.067746079 | -0.041142334 | 0.026603745 |
| SPIRE1 | 0.065824753 | -0.05837697 | 0.007447783 |
| RAC1 | 0.104757315 | -0.056867426 | 0.047889889 |
| TMEM129 | 0.065044731 | -0.042408232 | 0.022636498 |
| FBLN5 | 0.030018251 | -0.032637679 | -0.002619428 |
| SRRM1 | 0.095862863 | -0.074044277 | 0.021818586 |
| POLR1D | 0.093201925 | -0.049476288 | 0.043725637 |
| PPIA | 0.105900463 | -0.033763583 | 0.072136879 |
| SLC25A11 | 0.092173389 | -0.038516819 | 0.05365657 |
| ZHX1 | 0.07260466 | -0.057852178 | 0.014752483 |
| LIMCH1 | 0.08052086 | -0.03874421 | 0.04177665 |
| COL24A1 | 0.037675362 | -0.028391674 | 0.009283688 |
| PTRHD1 | 0.081565009 | -0.052525813 | 0.029039195 |
| GMNN | 0.066127869 | -0.053531006 | 0.012596863 |
| ZNF326 | 0.062505097 | -0.061972334 | 0.000532763 |
| PSMA2 | 0.103941282 | -0.036568885 | 0.067372397 |
| CRB3 | 0.067114073 | -0.044068498 | 0.023045575 |
| MCC | 0.050620982 | -0.040201685 | 0.010419297 |
| EPB41 | 0.056509618 | -0.049000203 | 0.007509415 |
| COX5A | 0.119282131 | -0.046330223 | 0.072951908 |
| VMP1 | 0.069777577 | -0.043559355 | 0.026218222 |
| SLC30A9 | 0.077072878 | -0.055717881 | 0.021354997 |
| HMBOX1 | 0.059258916 | -0.040123624 | 0.019135292 |
| MRPL18 | 0.085010441 | -0.038456297 | 0.046554143 |
| SLC39A7 | 0.069326595 | -0.05512136 | 0.014205235 |
| COA8 | 0.084389924 | -0.02391049 | 0.060479433 |
| LRP10 | 0.081460914 | -0.061314026 | 0.020146888 |
| DHX29 | 0.074622387 | -0.062570963 | 0.012051423 |
| DZIP3 | 0.052730286 | -0.047496852 | 0.005233434 |
| NDUFB4 | 0.114984702 | -0.062236025 | 0.052748677 |
| SLC8B1 | 0.033954221 | -0.03456706 | -0.00061284 |

| | | | |
|---|---|---|---|
| TTF2 | 0.042206987 | -0.03277525 | 0.009431737 |
| PLEKHA8 | 0.044948326 | -0.026118709 | 0.018829617 |
| CWH43 | 0.032678732 | -0.032609678 | 6.90537E-05 |
| TMEM117 | 0.048423532 | -0.042825752 | 0.00559778 |
| EMC3 | 0.08094755 | -0.03632768 | 0.044619869 |
| FBXO25 | 0.061808997 | -0.033001596 | 0.028807401 |
| NCL | 0.100761935 | -0.072292358 | 0.028469577 |
| KIAA1522 | 0.070302928 | -0.061781308 | 0.00852162 |
| HEBP2 | 0.091938235 | -0.071783571 | 0.020154663 |
| RB1 | 0.058699382 | -0.047818585 | 0.010880796 |
| CTTN | 0.086433876 | -0.053101713 | 0.033332163 |
| HYAL2 | 0.062321181 | -0.031958881 | 0.0303623 |
| NAALADL2 | 0.040125576 | -0.040178926 | -5.33501E-05 |
| OGFRL1 | 0.062205003 | -0.056681514 | 0.005523489 |
| GLOD4 | 0.082089013 | -0.024186876 | 0.057902137 |
| ARGLU1 | 0.099564427 | -0.076224929 | 0.023339497 |
| APOO | 0.074433266 | -0.027238567 | 0.047194699 |
| SRSF4 | 0.072429239 | -0.053432975 | 0.018996265 |
| C1GALT1 | 0.073772926 | -0.041940873 | 0.031832054 |
| TMEM150C | 0.068612598 | -0.061753629 | 0.006858969 |
| ABHD14A | 0.070256005 | -0.048841988 | 0.021414017 |
| NNT | 0.099257694 | -0.04981213 | 0.049445564 |
| WDR1 | 0.092804064 | -0.052066743 | 0.040737322 |
| NDUFAB1 | 0.109654861 | -0.033871685 | 0.075783176 |
| ASH1L | 0.085543052 | -0.068180095 | 0.017362958 |
| CAMLG | 0.064103742 | -0.050209433 | 0.013894309 |
| SNX13 | 0.070582167 | -0.021107509 | 0.049474658 |
| SPCS1 | 0.110613298 | -0.065180514 | 0.045432785 |
| CD47 | 0.073578811 | -0.051060121 | 0.02251869 |
| POGLUT3 | 0.057361415 | -0.040225628 | 0.017135787 |
| HSCB | 0.050737204 | -0.022007655 | 0.028729549 |
| USP34 | 0.080685405 | -0.086477559 | -0.005792154 |
| REPS2 | 0.027868742 | -0.030922427 | -0.003053685 |
| CPT1A | 0.082191216 | -0.032406958 | 0.049784258 |
| GSR | 0.063177987 | -0.041777841 | 0.021400145 |
| SERINC1 | 0.09484468 | -0.079757272 | 0.015087408 |
| FOXJ3 | 0.051645991 | -0.051352159 | 0.000293832 |
| TIAL1 | 0.074558884 | -0.045922204 | 0.02863668 |
| DDX18 | 0.079447098 | -0.059869668 | 0.01957743 |
| GET3 | 0.078020113 | -0.03740261 | 0.040617504 |
| HSP90AB1 | 0.108563982 | -0.058743781 | 0.0498202 |
| GCOM1 | 0.027846438 | -0.029848593 | -0.002002155 |
| CARS2 | 0.066365244 | -0.045153367 | 0.021211877 |
| USP25 | 0.051636078 | -0.041030103 | 0.010605974 |
| NSD3 | 0.074588943 | -0.065354761 | 0.009234182 |
| SELENOH | 0.10758796 | -0.040280237 | 0.067307724 |
| PKN1 | 0.056665916 | -0.04146507 | 0.015200846 |

| | | | |
|---|---|---|---|
| SORT1 | 0.069448282 | -0.063607061 | 0.005841221 |
| LSM1 | 0.071778682 | -0.04409604 | 0.027682642 |
| CLCN7 | 0.054277742 | -0.040926955 | 0.013350787 |
| USP14 | 0.070416047 | -0.038632229 | 0.031783818 |
| MT2A | -0.101306333 | 0.10622035 | 0.004914017 |
| GIGYF2 | 0.076002516 | -0.059796443 | 0.016206073 |
| NDUFS3 | 0.096789977 | -0.04461707 | 0.052172907 |
| GPX3 | -0.087073059 | 0.063338343 | -0.023734717 |
| UPF2 | 0.054530305 | -0.04483782 | 0.009692485 |
| CD58 | 0.037229836 | -0.029059798 | 0.008170038 |
| CCT2 | 0.085435233 | -0.062237878 | 0.023197355 |
| EIF4A1 | 0.099044815 | -0.064049695 | 0.03499512 |
| CFLAR | 0.069294963 | -0.053152744 | 0.016142219 |
| GPR146 | 0.03638355 | -0.038345911 | -0.001962361 |
| BPTF | 0.082480032 | -0.056365757 | 0.026114275 |
| DYNC1H1 | 0.084426345 | -0.079424072 | 0.005002273 |
| MRPS36 | 0.102310653 | -0.044001185 | 0.058309468 |
| SYNGR2 | 0.067648267 | -0.042594932 | 0.025053335 |
| ANP32A | 0.105265177 | -0.037163537 | 0.06810164 |
| PIK3R3 | 0.034790098 | -0.037449038 | -0.00265894 |
| ACBD3 | 0.062636849 | -0.049671252 | 0.012965597 |
| TYW1 | 0.037904863 | -0.027658724 | 0.010246139 |
| TMEM33 | 0.084952101 | -0.04224241 | 0.042709691 |
| MXD4 | 0.078473999 | -0.04975741 | 0.028716589 |
| OTUD6B-AS1 | 0.092872067 | -0.034306735 | 0.058565331 |
| ACAA1 | 0.074274465 | -0.043622174 | 0.030652291 |
| MED10 | 0.077114988 | -0.046752709 | 0.030362279 |
| DGLUCY | 0.06357744 | -0.042905419 | 0.020672022 |
| DERL1 | 0.059851821 | -0.039377095 | 0.020474726 |
| BDP1 | 0.071644748 | -0.065073808 | 0.00657094 |
| PTGS1 | 0.018493984 | -0.018493984 | -1.77636E-15 |
| NR1D2 | 0.060499239 | -0.033138337 | 0.027360901 |
| NACA | 0.109849532 | -0.030022707 | 0.079826825 |
| SLK | 0.062014472 | -0.053626318 | 0.008388154 |
| RBBP7 | 0.077757773 | -0.055033305 | 0.022724468 |
| SCAMP5 | 0.03143226 | -0.03221331 | -0.00078105 |
| SMAP1 | 0.056528987 | -0.047132592 | 0.009396395 |
| KIAA0040 | 0.054845748 | -0.051737739 | 0.003108009 |
| BLVRB | 0.096905737 | -0.033994569 | 0.062911168 |
| RTCB | 0.067742799 | -0.048901109 | 0.01884169 |
| ABI2 | 0.060282959 | -0.033614484 | 0.026668476 |
| ARG2 | 0.063498454 | -0.058175281 | 0.005323173 |
| GLO1 | 0.088799404 | -0.036255405 | 0.052543999 |
| ATP5PD | 0.114135836 | -0.054262156 | 0.059873679 |
| PIP4P1 | 0.053375128 | -0.032385822 | 0.020989305 |
| EID1 | 0.101974324 | -0.074596934 | 0.027377391 |
| HAUS1 | 0.04960469 | -0.028270275 | 0.021334415 |

| | | | |
|---|---|---|---|
| UQCC2 | 0.092096306 | -0.034614914 | 0.057481392 |
| C9orf78 | 0.069032752 | -0.0430757 | 0.025957053 |
| HMGB3 | 0.06229678 | -0.048557052 | 0.013739728 |
| C1orf210 | 0.057158371 | -0.033672378 | 0.023485993 |
| UFL1 | 0.068895074 | -0.040874898 | 0.028020176 |
| PPIG | 0.087265042 | -0.04502289 | 0.042242152 |
| PTP4A2 | 0.092604486 | -0.074987174 | 0.017617312 |
| TP53I3 | 0.075717591 | -0.051780936 | 0.023936655 |
| SMS | 0.087912528 | -0.034254059 | 0.05365847 |
| JKAMP | 0.069414589 | -0.044848361 | 0.024566228 |
| HS6ST2 | -0.021380021 | 0.008560525 | -0.012819495 |
| TFB1M | 0.040945333 | -0.027895141 | 0.013050191 |
| LGALS3 | 0.129105496 | -0.076572247 | 0.052533249 |
| SCD5 | -0.048592652 | 0.043758627 | -0.004834025 |
| CDC37 | 0.080077457 | -0.039312935 | 0.040764521 |
| COX19 | 0.059482036 | -0.041522209 | 0.017959827 |
| PTBP3 | 0.097784925 | -0.083992464 | 0.013792461 |
| VEGFB | 0.096545964 | -0.040428081 | 0.056117883 |
| ADIRF | -0.068718544 | 0.025373807 | -0.043344737 |
| MIA3 | 0.065146046 | -0.054215634 | 0.010930412 |
| IVD | 0.074879484 | -0.031177005 | 0.043702478 |
| SREBF2 | 0.066440864 | -0.05694937 | 0.009491495 |
| SGSM3 | 0.057164877 | -0.047372727 | 0.00979215 |
| AIF1L | 0.090488513 | -0.073197134 | 0.017291379 |
| SPG7 | 0.070898903 | -0.063010675 | 0.007888227 |
| MDH1 | 0.10480583 | -0.041224491 | 0.063581339 |
| KIFC3 | 0.048456385 | -0.043067316 | 0.005389069 |
| TNFAIP8 | 0.056150365 | -0.043731762 | 0.012418603 |
| NASP | 0.075259844 | -0.037567782 | 0.037692062 |
| COX7C | 0.102161764 | -0.037648638 | 0.064513126 |
| HLA-F | 0.065749135 | -0.022814946 | 0.042934189 |
| PTPMT1 | 0.077300851 | -0.029117856 | 0.048182995 |
| VPS28 | 0.100284992 | -0.040092372 | 0.06019262 |
| SSR2 | 0.097466859 | -0.03793654 | 0.059530319 |
| JTB | 0.094065124 | -0.044566638 | 0.049498486 |
| EPN3 | 0.05481851 | -0.04854286 | 0.00627565 |
| CSPP1 | 0.043643284 | -0.022284982 | 0.021358303 |
| DECR1 | 0.093761649 | -0.037002796 | 0.056758852 |
| MOB1A | 0.084177991 | -0.054831252 | 0.029346739 |
| TBC1D9 | 0.060598619 | -0.048925163 | 0.011673456 |
| DDR1 | 0.083466181 | -0.059907615 | 0.023558566 |
| IFI6 | 0.061702823 | -0.040289238 | 0.021413585 |
| AC013472.2 | 0.032573536 | -0.022252157 | 0.010321379 |
| AP1M2 | 0.079760946 | -0.022229194 | 0.057531752 |
| NIPSNAP2 | 0.095549379 | -0.069369515 | 0.026179863 |
| MIER1 | 0.065525338 | -0.049890361 | 0.015634976 |
| MVB12A | 0.056604355 | -0.032365671 | 0.024238684 |

| | | | |
|---|---|---|---|
| HOMER3 | 0.044788053 | -0.034205191 | 0.010582861 |
| MANBAL | 0.065284908 | -0.046675291 | 0.018609617 |
| COLCA2 | 0.041754084 | -0.039917773 | 0.001836311 |
| DHX36 | 0.071908883 | -0.026131476 | 0.045777407 |
| STIM1 | 0.061216275 | -0.032106609 | 0.029109665 |
| SMIM14 | 0.068763271 | -0.028237437 | 0.040525834 |
| ROCK2 | 0.064850381 | -0.056357783 | 0.008492598 |
| CXADR | 0.067383704 | -0.057806869 | 0.009576835 |
| HLA-DQB1 | 0.030980606 | -0.037580175 | -0.006599569 |
| MARCH7 | 0.059141293 | -0.041228525 | 0.017912768 |
| PTGR2 | 0.056552192 | -0.037300762 | 0.01925143 |
| DNAJB14 | 0.071934405 | -0.050876128 | 0.021058276 |
| SSNA1 | 0.0834763 | -0.046207407 | 0.037268893 |
| EEF1B2 | 0.115499923 | -0.055231308 | 0.060268615 |
| MBOAT2 | 0.063957714 | -0.066701504 | -0.00274379 |
| RAD17 | 0.054880442 | -0.021347181 | 0.033533261 |
| MORF4L1 | 0.098923952 | -0.042338508 | 0.056585444 |
| IGSF22 | 0.03986867 | -0.022285406 | 0.017583264 |
| MISP3 | 0.049474452 | -0.038271609 | 0.011202843 |
| DHRS3 | 0.0774834 | -0.064237969 | 0.013245431 |
| U2SURP | 0.086376044 | -0.067908768 | 0.018467276 |
| SERTAD1 | 0.054717547 | -0.02983439 | 0.024883158 |
| HNRNPR | 0.08233706 | -0.06172545 | 0.02061161 |
| MCPH1 | 0.046268195 | -0.031651898 | 0.014616297 |
| PPP1R35 | 0.065364859 | -0.03054413 | 0.034820728 |
| METTL2A | 0.045634161 | -0.019323555 | 0.026310607 |
| SERBP1 | 0.093093239 | -0.074586833 | 0.018506406 |
| NPM1 | 0.108693065 | -0.059049119 | 0.049643946 |
| UQCRC1 | 0.094534478 | -0.056162985 | 0.038371493 |
| NPNT | 0.071928489 | -0.068423368 | 0.003505121 |
| SECISBP2 | 0.069531556 | -0.063135473 | 0.006396083 |
| RASSF7 | 0.072682722 | -0.075744957 | -0.003062234 |
| GFPT1 | 0.055960217 | -0.038698465 | 0.017261752 |
| ATF6 | 0.066055849 | -0.030411734 | 0.035644115 |
| UNC50 | 0.088581471 | -0.059002595 | 0.029578875 |
| CEP57 | 0.058298819 | -0.027035077 | 0.031263742 |
| MAP7 | 0.053504966 | -0.037577875 | 0.01592709 |
| MRPS22 | 0.060724415 | -0.025378692 | 0.035345723 |
| RNF149 | 0.050293414 | -0.045808634 | 0.004484779 |
| POMGNT1 | 0.06590779 | -0.039240196 | 0.026667594 |
| GSTK1 | 0.098168353 | -0.047592598 | 0.050575756 |
| POLR2C | 0.068623844 | -0.037683388 | 0.030940456 |
| TJP1 | 0.059378842 | -0.043874819 | 0.015504023 |
| PRKCI | 0.051366921 | -0.03637656 | 0.014990361 |
| SMCHD1 | 0.057836354 | -0.030075777 | 0.027760576 |
| ATP6V1C2 | 0.104004376 | -0.061663004 | 0.042341373 |
| CAMSAP2 | 0.061112835 | -0.037354998 | 0.023757836 |

| | | | |
|---|---|---|---|
| BRI3BP | 0.051671258 | -0.037826068 | 0.01384519 |
| ERBB3 | 0.066063061 | -0.04403512 | 0.022027941 |
| RAD50 | 0.073326362 | -0.056625219 | 0.016701143 |
| PHYH | 0.093784458 | 0.055505046 | 0.149289504 |
| SFXN1 | 0.052682308 | -0.047696265 | 0.004986043 |
| GNE | 0.042254005 | -0.048340821 | -0.006086816 |
| USP33 | 0.067276703 | -0.03458951 | 0.032687193 |
| RAB14 | 0.081912699 | -0.057937125 | 0.023975574 |
| MALAT1 | 0.166288339 | -0.205495533 | -0.039207194 |
| NDUFS4 | 0.092863863 | -0.042486197 | 0.050377666 |
| MIR210HG | 0.028839789 | -0.024531081 | 0.004308708 |
| MYPOP | 0.043938809 | -0.049786476 | -0.005847667 |
| NEDD9 | 0.033569295 | -0.037862679 | -0.004293385 |
| ACYP1 | 0.055885267 | -0.043246528 | 0.01263874 |
| CEBPZOS | 0.075471026 | -0.029958993 | 0.045512033 |
| RUFY3 | 0.067496719 | -0.073791398 | -0.006294679 |
| NT5C3A | 0.047206786 | -0.035046267 | 0.012160519 |
| NPC2 | 0.101304537 | -0.040733645 | 0.060570892 |
| IDH3G | 0.077086284 | -0.038266445 | 0.038819839 |
| ABO | 0.043559014 | -0.041313568 | 0.002245447 |
| PAIP1 | 0.073967755 | -0.050281076 | 0.023686679 |
| CENPV | 0.051549142 | -0.027663362 | 0.02388578 |
| MAN1C1 | 0.072207701 | -0.054820177 | 0.017387524 |
| SLC4A11 | 0.02177271 | -0.020887779 | 0.000884932 |
| PGLS | 0.087958547 | -0.032206267 | 0.05575228 |
| TBCB | 0.081413916 | -0.026589307 | 0.05482461 |
| RBFA | 0.052556292 | -0.050332698 | 0.002223594 |
| NUP50 | 0.052484987 | -0.031620484 | 0.020864503 |
| XRN1 | 0.065205877 | -0.066918166 | -0.00171229 |
| ZCRB1 | 0.078389974 | -0.046091947 | 0.032298027 |
| P2RX4 | 0.04250017 | -0.032229909 | 0.010270261 |
| RALA | 0.06261536 | -0.028796259 | 0.033819101 |
| MIA2 | 0.071537294 | -0.061352191 | 0.010185103 |
| TMEM52B | -0.092747782 | 0.060367517 | -0.032380265 |
| MR1 | 0.070337624 | -0.03066959 | 0.039668034 |
| SNX6 | 0.082648511 | -0.049467981 | 0.03318053 |
| SLC26A7 | 0.158022695 | -0.099673258 | 0.058349437 |
| BCAP31 | 0.089881988 | -0.063823371 | 0.026058617 |
| PYCARD | 0.043251466 | -0.038851414 | 0.004400052 |
| LPGAT1 | 0.060141393 | -0.053209849 | 0.006931544 |
| XBP1 | 0.083519487 | -0.067019565 | 0.016499922 |
| TM2D3 | 0.069655236 | -0.044343237 | 0.025311999 |
| GAS2L1 | 0.043094616 | -0.033265173 | 0.009829444 |
| ECI2 | 0.085646073 | -0.039973796 | 0.045672277 |
| OAF | 0.056699284 | -0.028696501 | 0.028002783 |
| SREK1 | 0.071017435 | -0.066915785 | 0.00410165 |
| RAP2B | 0.073305081 | -0.083551042 | -0.010245961 |

| | | | |
|---|---|---|---|
| DYNC1LI2 | 0.068754137 | -0.060726185 | 0.008027953 |
| RMND5B | 0.061692494 | -0.028391597 | 0.033300897 |
| ZMYND8 | 0.052963931 | -0.043695857 | 0.009268074 |
| GPER1 | 0.028488477 | -0.02800615 | 0.000482327 |
| C1orf43 | 0.091907455 | -0.033669586 | 0.058237869 |
| MT-ND2 | -0.077399996 | 0.027529056 | -0.04987094 |
| RAB4A | 0.067386892 | -0.057527027 | 0.009859865 |
| EIF2AK2 | 0.061402423 | -0.033715439 | 0.027686984 |
| SLC39A1 | 0.066234527 | -0.048858616 | 0.01737591 |
| CCDC174 | 0.051743021 | -0.039593366 | 0.012149654 |
| DAD1 | 0.101761096 | -0.066630708 | 0.035130388 |
| PABPC4 | 0.068401648 | -0.063727652 | 0.004673996 |
| DIAPH1 | 0.068227066 | -0.051828057 | 0.016399008 |
| MARCH9 | 0.047475432 | -0.029794197 | 0.017681235 |
| TOMM5 | 0.089857876 | -0.030616001 | 0.059241875 |
| EFNA1 | 0.055683397 | -0.042611807 | 0.01307159 |
| KCTD9 | 0.039915647 | -0.025134239 | 0.014781407 |
| ROMO1 | 0.101484694 | -0.042635019 | 0.058849675 |
| NIM1K | 0.017464291 | -0.017464291 | -5.32907E-15 |
| BAG5 | 0.064888751 | -0.024583625 | 0.040305126 |
| PCM1 | 0.077228443 | -0.07352277 | 0.003705673 |
| LY75 | 0.023658381 | -0.02599709 | -0.002338709 |
| PPP1R36 | 0.022949387 | -0.02475394 | -0.001804552 |
| SH3BP2 | 0.031765352 | -0.039217781 | -0.007452429 |
| PFDN5 | 0.103034309 | -0.033381479 | 0.06965283 |
| HNRNPAB | 0.069804871 | -0.037665183 | 0.032139687 |
| PPA2 | 0.068064489 | -0.019044567 | 0.049019922 |
| SNF8 | 0.079021973 | -0.038189111 | 0.040832862 |
| MRPL38 | 0.073976869 | -0.024610548 | 0.049366321 |
| C15orf61 | 0.068808647 | -0.039026071 | 0.029782576 |
| STAT1 | 0.055935992 | -0.026646583 | 0.029289409 |
| INSR | 0.075930795 | -0.087565755 | -0.011634959 |
| TMEM205 | 0.087993053 | -0.04312866 | 0.044864393 |
| MARCH1 | 0.021715671 | -0.023970805 | -0.002255134 |
| PRMT1 | 0.080509767 | -0.049044163 | 0.031465604 |
| SERPINB6 | 0.076962207 | -0.059439978 | 0.017522228 |
| STAT3 | 0.062200331 | -0.052849297 | 0.009351034 |
| ARF4 | 0.086471444 | -0.04098967 | 0.045481775 |
| CISD2 | 0.075453815 | -0.054755641 | 0.020698175 |
| ST7 | 0.047740716 | -0.019109671 | 0.028631045 |
| COPB2 | 0.064995117 | -0.041228807 | 0.02376631 |
| OXR1 | 0.06845442 | -0.035044759 | 0.033409661 |
| GALNT1 | 0.044043208 | -0.031118483 | 0.012924725 |
| DYNLRB1 | 0.099914733 | -0.076468019 | 0.023446715 |
| FCHO2 | 0.05571713 | -0.045607809 | 0.010109321 |
| MGST2 | 0.093387447 | -0.056108428 | 0.037279019 |
| ENOSF1 | 0.068192303 | -0.035137742 | 0.033054562 |

| | | | |
|---|---|---|---|
| SERINC3 | 0.072774201 | -0.063920843 | 0.008853358 |
| KCNK5 | 0.019488204 | -0.020052969 | -0.000564766 |
| NEDD4L | 0.098798733 | -0.039855423 | 0.05894331 |
| TSPAN31 | 0.067673734 | -0.040454479 | 0.027219255 |
| SLC25A24 | 0.051513017 | -0.046007696 | 0.005505321 |
| PPP1R7 | 0.07102572 | -0.033828911 | 0.037196809 |
| AUP1 | 0.070996844 | -0.060745946 | 0.010250897 |
| IRAK4 | 0.051842943 | -0.043363909 | 0.008479034 |
| ADCY6 | 0.048522409 | -0.04450433 | 0.004018079 |
| RCBTB2 | 0.043782903 | -0.036678379 | 0.007104524 |
| GAS5 | 0.09117148 | -0.028140037 | 0.063031443 |
| CCT4 | 0.086267254 | -0.037702546 | 0.048564708 |
| NFXL1 | 0.033800928 | -0.031430727 | 0.002370201 |
| SHTN1 | 0.057593768 | -0.044138783 | 0.013454985 |
| RSBN1L | 0.063305321 | -0.06387344 | -0.00056812 |
| MRPL14 | 0.083043999 | -0.033622713 | 0.049421286 |
| NPTN | 0.084137096 | -0.068831744 | 0.015305352 |
| PNKD | 0.096402151 | -0.045830421 | 0.05057173 |
| SUDS3 | 0.061820977 | -0.043242792 | 0.018578184 |
| GET1 | 0.060897612 | -0.031575766 | 0.029321846 |
| SPTAN1 | 0.082754194 | -0.101164399 | -0.018410205 |
| EFEMP1 | 0.069048327 | -0.122791645 | -0.053743319 |
| SLC39A4 | 0.068725031 | -0.079852993 | -0.011127961 |
| CMC2 | 0.081283558 | -0.041626153 | 0.039657405 |
| EBAG9 | 0.063483333 | -0.050090589 | 0.013392744 |
| STAU1 | 0.069814696 | -0.062022409 | 0.007792287 |
| RPL35A | 0.101787473 | -0.043348232 | 0.058439241 |
| NIPAL1 | 0.044737006 | -0.048683581 | -0.003946575 |
| SPTLC1 | 0.063478067 | -0.052167629 | 0.011310439 |
| HNRNPM | 0.08036672 | -0.036574788 | 0.043791931 |
| UGT8 | 0.076340648 | -0.02328771 | 0.053052938 |
| TMUB1 | 0.063985518 | -0.028401091 | 0.035584427 |
| GSTO1 | 0.092989259 | -0.036954849 | 0.05603441 |
| ARHGAP8 | 0.061846405 | -0.035609032 | 0.026237373 |
| LYAR | 0.034676259 | -0.04110609 | -0.006429831 |
| PNRC2 | 0.083016294 | -0.075016977 | 0.007999317 |
| TMED1 | 0.076972422 | -0.059027021 | 0.017945401 |
| ZFP91 | 0.055688791 | -0.033537228 | 0.022151563 |
| UQCC3 | 0.076163393 | -0.035935934 | 0.040227458 |
| TRIM2 | 0.086737499 | -0.051571587 | 0.035165911 |
| PSMD1 | 0.067925534 | -0.056500709 | 0.011424825 |
| EFNB2 | 0.036436887 | -0.042487012 | -0.006050125 |
| LRRC28 | 0.045894264 | -0.028889392 | 0.017004871 |
| NFIB | 0.047479312 | -0.036758846 | 0.010720466 |
| H1F0 | 0.08246574 | -0.094509273 | -0.012043534 |
| CDS1 | 0.051528978 | -0.023259369 | 0.028269609 |
| C11orf58 | 0.091765552 | -0.054549292 | 0.03721626 |

| | | | |
|---|---|---|---|
| MCFD2 | 0.0724142 | -0.035840589 | 0.036573611 |
| PISD | 0.049562974 | -0.039998113 | 0.009564861 |
| TBC1D16 | 0.046372062 | -0.034832511 | 0.011539551 |
| WBP1L | 0.05039421 | -0.045080149 | 0.005314061 |
| HSDL2 | 0.061267073 | -0.045436499 | 0.015830574 |
| ZNF69 | 0.027537166 | -0.027905735 | -0.000368569 |
| APH1A | 0.074533233 | -0.03011419 | 0.044419043 |
| TACC1 | 0.053102402 | -0.069416482 | -0.01631408 |
| RRAD | 0.028294316 | -0.030512966 | -0.00221865 |
| MBD4 | 0.063604649 | -0.039112309 | 0.024492341 |
| UBB | 0.102684072 | -0.033169339 | 0.069514732 |
| HEXB | 0.050194537 | -0.033879638 | 0.016314899 |
| MT-ND6 | -0.095024365 | 0.095686321 | 0.000661956 |
| EVA1B | 0.055461221 | -0.037880369 | 0.017580852 |
| RPS9 | 0.092071377 | -0.039393474 | 0.052677903 |
| RPL5 | 0.105811098 | -0.043023166 | 0.062787932 |
| DNMT1 | 0.046269266 | -0.042500786 | 0.003768481 |
| CDC123 | 0.058111302 | -0.035854654 | 0.022256648 |
| LYSMD3 | 0.0401486 | -0.024437219 | 0.015711381 |
| AGL | 0.065128183 | -0.047255215 | 0.017872968 |
| F2RL1 | 0.039924609 | -0.041457365 | -0.001532756 |
| PIGL | 0.045666896 | -0.028614722 | 0.017052175 |
| KLHDC2 | 0.077237515 | -0.049336536 | 0.027900979 |
| JAK1 | 0.071324755 | -0.079283119 | -0.007958364 |
| SAT1 | 0.104240123 | -0.098723702 | 0.005516421 |
| WDR83 | 0.042687605 | -0.036696233 | 0.005991372 |
| FUT2 | 0.018409795 | -0.019528541 | -0.001118745 |
| RDX | 0.071541365 | -0.043543341 | 0.027998024 |
| KNL1 | 0.025022189 | -0.025396613 | -0.000374424 |
| NDUFA3 | 0.103033458 | -0.073423221 | 0.029610236 |
| CCDC91 | 0.054382192 | -0.041387351 | 0.012994842 |
| FABP3 | -0.048873754 | 0.022017927 | -0.026855827 |
| NDFIP2 | 0.063324022 | -0.043347375 | 0.019976648 |
| SWI5 | 0.076181409 | -0.047375491 | 0.028805919 |
| AK2 | 0.081655271 | -0.068097057 | 0.013558214 |
| PDIA6 | 0.093929867 | -0.098209502 | -0.004279635 |
| POLR2J | 0.088015794 | -0.041110352 | 0.046905443 |
| ARID4B | 0.06515824 | -0.057303063 | 0.007855176 |
| UBE2E3 | 0.069703187 | -0.051634155 | 0.018069032 |
| HLA-B | 0.128806878 | 0.042306847 | 0.171113725 |
| LNPK | 0.053326235 | -0.039580361 | 0.013745874 |
| RASSF8 | 0.039119749 | -0.024341537 | 0.014778212 |
| DNPEP | 0.055951575 | -0.027967582 | 0.027983993 |
| PGK1 | 0.095183121 | -0.05561556 | 0.039567562 |
| HNRNPH3 | 0.079436548 | -0.065584154 | 0.013852394 |
| CSNK2A1 | 0.073101323 | -0.041934203 | 0.03116712 |
| HDLBP | 0.077999706 | -0.048394311 | 0.029605395 |

| | | | |
|---|---|---|---|
| ZDHHC2 | 0.064046897 | -0.028043925 | 0.036002972 |
| XIAP | 0.064053877 | -0.048824995 | 0.015228881 |
| CHD9 | 0.082411102 | -0.081581509 | 0.000829593 |
| SLCO4C1 | 0.032246515 | -0.036739352 | -0.004492837 |
| ABCD4 | 0.048051305 | -0.039605373 | 0.008445932 |
| TRIM16 | 0.025360288 | -0.016109591 | 0.009250697 |
| PDCD6IP | 0.072457627 | -0.045660786 | 0.026796841 |
| PEPD | 0.063892778 | -0.024618989 | 0.039273789 |
| COQ10B | 0.063241512 | -0.023953083 | 0.039288429 |
| VPS8 | 0.037494854 | -0.01651083 | 0.020984024 |
| TXNDC16 | 0.031180131 | -0.030790506 | 0.000389625 |
| BLVRA | 0.049517026 | -0.041709686 | 0.00780734 |
| WDR41 | 0.047551541 | -0.048085714 | -0.000534173 |
| C16orf58 | 0.051164885 | -0.01626182 | 0.034903066 |
| PPID | 0.06196026 | -0.021262949 | 0.040697311 |
| CABP1 | -0.016242123 | 0.00511714 | -0.011124984 |
| HERC3 | 0.036985972 | -0.029975358 | 0.007010614 |
| CHD3 | 0.05051983 | -0.048466805 | 0.002053025 |
| PTPRK | 0.046412405 | -0.049152679 | -0.002740274 |
| CUL3 | 0.065271726 | -0.038469796 | 0.02680193 |
| ESD | 0.091402346 | -0.03990472 | 0.051497626 |
| AKAP7 | 0.045766079 | -0.032628784 | 0.013137295 |
| IGFBP3 | -0.035051111 | 0.024794984 | -0.010256127 |
| EPC2 | 0.042915452 | -0.027680098 | 0.015235354 |
| USP43 | 0.023092908 | -0.020841012 | 0.002251896 |
| MEIOC | 0.039851836 | -0.036417218 | 0.003434617 |
| TRPM7 | 0.053173995 | -0.056755051 | -0.003581055 |
| CDV3 | 0.063831681 | -0.046515976 | 0.017315705 |
| ATP9A | 0.055495333 | -0.029491427 | 0.026003906 |
| CFAP70 | 0.030958713 | -0.028915835 | 0.002042877 |
| TPGS1 | 0.062486733 | -0.035676743 | 0.02680999 |
| BROX | 0.065059851 | -0.030215502 | 0.034844348 |
| UPF3A | 0.069420394 | -0.030892634 | 0.03852776 |
| RPL31 | 0.099018847 | -0.057164738 | 0.041854109 |
| MCRIP2 | 0.08855023 | -0.058570302 | 0.029979928 |
| BCAP29 | 0.083209671 | -0.064054783 | 0.019154888 |
| CWF19L2 | 0.056237937 | -0.043319974 | 0.012917963 |
| PHF11 | 0.061784384 | -0.035434527 | 0.026349857 |
| KIFAP3 | 0.047596481 | -0.023670379 | 0.023926103 |
| BAIAP2L1 | 0.05758104 | -0.045494253 | 0.012086787 |
| DPP8 | 0.052203344 | -0.032569601 | 0.019633743 |
| NDUFS8 | 0.099405001 | -0.037944966 | 0.061460035 |
| AHCTF1 | 0.047861569 | -0.03416965 | 0.013691919 |
| MRPL39 | 0.059312449 | -0.033518117 | 0.025794333 |
| MTIF3 | 0.074816879 | -0.057155324 | 0.017661555 |
| C22orf39 | 0.072320601 | -0.024303952 | 0.04801665 |
| TMF1 | 0.057910875 | -0.040342209 | 0.017568666 |

| | | | |
|---|---|---|---|
| POU2F3 | 0.018714824 | -0.018675849 | 3.89744E-05 |
| COIL | 0.045025756 | -0.038524832 | 0.006500923 |
| USP48 | 0.054563557 | -0.035661599 | 0.018901958 |
| PRKX | 0.042469023 | -0.053777097 | -0.011308074 |
| IFT57 | 0.060184595 | -0.030227304 | 0.029957291 |
| SNRPD2 | 0.090686876 | -0.029762483 | 0.060924393 |
| IFI27L1 | 0.058878357 | -0.035520663 | 0.023357694 |
| PSENEN | 0.068516284 | -0.031058565 | 0.037457719 |
| ZNF281 | 0.039255294 | -0.034904858 | 0.004350436 |
| KEAP1 | 0.040340577 | -0.022310294 | 0.018030282 |
| SARS | 0.067228232 | -0.044989814 | 0.022238418 |
| TMEM87B | 0.038553248 | -0.042102862 | -0.003549614 |
| SLC35F5 | 0.066233616 | -0.047125806 | 0.01910781 |
| FDX1 | 0.078300862 | -0.052714849 | 0.025586013 |
| PLS3 | 0.075195386 | -0.053327223 | 0.021868163 |
| TSFM | 0.063920259 | -0.03201856 | 0.031901699 |
| DUSP23 | 0.077961456 | -0.044669285 | 0.033292171 |
| LEMD2 | 0.033251799 | -0.020190415 | 0.013061384 |
| FNBP4 | 0.051877918 | -0.036841173 | 0.015036746 |
| SKAP1 | 0.065912174 | -0.028703523 | 0.037208651 |
| PRNP | 0.041314787 | -0.029021528 | 0.012293259 |
| NAPG | 0.058832863 | -0.040675003 | 0.01815786 |
| KIF1B | 0.062408104 | -0.046559795 | 0.015848308 |
| TRMT61B | 0.043757761 | -0.013308065 | 0.030449696 |
| RETREG2 | 0.071859589 | -0.049699262 | 0.022160327 |
| SPESP1 | 0.031467024 | -0.027471988 | 0.003995037 |
| MRPS24 | 0.092560268 | -0.03380151 | 0.058758758 |
| HCAR1 | 0.023125324 | -0.021400089 | 0.001725235 |
| MFAP3L | 0.036079233 | -0.03381062 | 0.002268613 |
| CDC42EP4 | 0.03632746 | -0.031649202 | 0.004678258 |
| SPATA20 | 0.060666484 | -0.045510711 | 0.015155773 |
| SYT17 | 0.07287559 | -0.02459279 | 0.0482828 |
| OCEL1 | 0.052982335 | -0.030483617 | 0.022498718 |
| TMLHE | 0.039981284 | -0.022070678 | 0.017910606 |
| ORMDL2 | 0.063596216 | -0.030353798 | 0.033242418 |
| AHSA1 | 0.073962911 | -0.033019187 | 0.040943724 |
| TLN1 | 0.061607416 | -0.042058711 | 0.019548705 |
| KRTCAP2 | 0.094956971 | -0.095177708 | -0.000220737 |
| DCUN1D4 | 0.039729134 | -0.023939669 | 0.015789465 |
| CHMP4B | 0.058893123 | -0.021310392 | 0.037582731 |
| PEBP1 | 0.09189451 | -0.075208234 | 0.016686276 |
| NAGK | 0.052516467 | -0.023604946 | 0.028911521 |
| POLE4 | 0.075456031 | -0.029112481 | 0.04634355 |
| RHBDL2 | 0.018578236 | -0.017878904 | 0.000699331 |
| MAN2A2 | 0.032357395 | -0.02981601 | 0.002541385 |
| HACD1 | 0.030784633 | -0.017411676 | 0.013372957 |
| SNTB2 | 0.045378436 | -0.042685895 | 0.00269254 |

| | | | |
|---|---|---|---|
| RIF1 | 0.056995354 | -0.038275309 | 0.018720045 |
| TMEM161A | 0.046220414 | -0.01891793 | 0.027302484 |
| GARS | 0.044722085 | -0.030332749 | 0.014389335 |
| ADAM17 | 0.046165531 | -0.031604468 | 0.014561063 |
| CA5B | 0.039189572 | -0.035499896 | 0.003689676 |
| EDDM3A | 0.015535678 | -0.015294077 | 0.0002416 |
| SRRM2 | 0.086033388 | -0.066538746 | 0.019494642 |
| USP9Y | 0.037142263 | -0.054539509 | -0.017397246 |
| FBXO44 | 0.042643136 | -0.035599924 | 0.007043211 |
| JDP2 | 0.024522887 | -0.019451418 | 0.005071469 |
| GPR160 | 0.060591766 | -0.043620249 | 0.016971517 |
| SLC52A2 | 0.050807939 | -0.032676281 | 0.018131658 |
| ACTN4 | 0.08280087 | -0.05025204 | 0.03254883 |
| LUC7L2 | 0.066728605 | -0.039040667 | 0.027687938 |
| SAP18 | 0.095005392 | -0.033671116 | 0.061334276 |
| VPS4B | 0.055304221 | -0.040142025 | 0.015162196 |
| INTS11 | 0.056604851 | -0.026949631 | 0.02965522 |
| ARHGDIB | 0.029920227 | -0.021982333 | 0.007937895 |
| FREM2 | 0.034343877 | -0.0290449 | 0.005298976 |
| HDDC2 | 0.077470156 | -0.041770449 | 0.035699707 |
| PTS | 0.078539814 | -0.032329361 | 0.046210453 |
| RPL29 | 0.10087776 | -0.041528797 | 0.059348963 |
| YPEL3 | 0.072162557 | -0.047895901 | 0.024266656 |
| MRPL57 | 0.08844267 | -0.053645107 | 0.034797563 |
| ARHGAP28 | 0.031767669 | -0.020105183 | 0.011662486 |
| UXT | 0.091468545 | -0.029939904 | 0.061528641 |
| ACADM | 0.08331458 | -0.08176366 | 0.001550919 |
| LEPROTL1 | 0.058791582 | -0.04851286 | 0.010278723 |
| CHMP2B | 0.058861809 | -0.041525464 | 0.017336345 |
| USF2 | 0.064518869 | -0.045004387 | 0.019514482 |
| MORC2 | 0.03213503 | -0.025265202 | 0.006869828 |
| SNX24 | 0.05434237 | -0.025162485 | 0.029179885 |
| HSPA1A | 0.115322182 | -0.095343243 | 0.019978939 |
| ECHDC3 | 0.075475897 | -0.029557721 | 0.045918176 |
| PRKAA2 | 0.083295483 | -0.031275989 | 0.052019494 |
| SMC3 | 0.062510449 | -0.051088241 | 0.011422208 |
| ADPRHL2 | 0.041838149 | -0.024534826 | 0.017303323 |
| NDUFB7 | 0.095454751 | -0.035121455 | 0.060333296 |
| SCYL1 | 0.05187907 | -0.017829133 | 0.034049937 |
| CYHR1 | 0.072352968 | -0.032829817 | 0.039523151 |
| CEBPZ | 0.062729418 | -0.041375872 | 0.021353546 |
| CFL1 | 0.098107744 | -0.037458476 | 0.060649269 |
| AC007993.3 | 0.017762568 | -0.018754456 | -0.000991888 |
| COX17 | 0.095856401 | -0.049020982 | 0.046835419 |
| RASSF9 | 0.023723735 | -0.027348216 | -0.003624482 |
| RPP21 | 0.071749231 | -0.045459963 | 0.026289268 |
| SSB | 0.079971853 | -0.038616556 | 0.041355296 |

| | | | |
|---|---|---|---|
| SRI | 0.096277036 | -0.03618398 | 0.060093055 |
| SNW1 | 0.061975131 | -0.046901189 | 0.015073942 |
| MAOA | 0.037471533 | -0.032417514 | 0.005054019 |
| PTCD3 | 0.058455352 | -0.043265561 | 0.015189791 |
| TOR1AIP2 | 0.063324164 | -0.041945658 | 0.021378506 |
| ANO6 | 0.047431652 | -0.05170603 | -0.004274378 |
| MOGS | 0.047855891 | -0.045298942 | 0.002556949 |
| PRKD3 | 0.03888225 | -0.030079905 | 0.008802345 |
| SMIM8 | 0.051173909 | -0.019571479 | 0.031602429 |
| UBL4A | 0.044558399 | -0.020227483 | 0.024330915 |
| SYNJ2BP | 0.07387426 | -0.032796098 | 0.041078162 |
| C6orf89 | 0.054086983 | -0.046188964 | 0.007898019 |
| AKR1A1 | 0.061336079 | -0.040930449 | 0.02040563 |
| TANC2 | 0.03765138 | -0.032765294 | 0.004886086 |
| RANBP2 | 0.070594404 | -0.048602381 | 0.021992022 |
| RBM25 | 0.069341521 | -0.053794808 | 0.015546713 |
| COL14A1 | 0.017370979 | -0.017370979 | -3.55271E-15 |
| NF2 | 0.061298739 | -0.044532927 | 0.016765812 |
| TAP1 | 0.034220335 | -0.016487367 | 0.017732968 |
| RAD23A | 0.074501666 | -0.044242055 | 0.030259611 |
| PGRMC2 | 0.058147312 | -0.043687924 | 0.014459387 |
| WDR60 | 0.041595643 | -0.036898455 | 0.004697187 |
| EIF2S1 | 0.05861384 | -0.031119772 | 0.027494068 |
| SRSF2 | 0.081156793 | -0.037112073 | 0.04404472 |
| MSMO1 | 0.057214288 | -0.039894877 | 0.017319411 |
| HSPD1 | 0.0903337 | -0.044813194 | 0.045520505 |
| FUT8 | 0.02522986 | -0.022095496 | 0.003134364 |
| PURA | 0.070133654 | -0.031170284 | 0.038963369 |
| SET | 0.089189409 | -0.047699619 | 0.04148979 |
| EIF3A | 0.075424504 | -0.057461262 | 0.017963243 |
| CFAP298 | 0.060014135 | -0.021023411 | 0.038990724 |
| PTPN11 | 0.059409392 | -0.022281911 | 0.037127481 |
| FAM174C | 0.066377542 | -0.024742183 | 0.04163536 |
| PLD3 | 0.068376 | -0.029638385 | 0.038737614 |
| CYTOR | 0.018486072 | -0.015248488 | 0.003237584 |
| SELENOF | 0.084860757 | -0.047918026 | 0.036942731 |
| PSMC4 | 0.065742925 | -0.032485293 | 0.033257632 |
| TBRG4 | 0.04721998 | -0.025932388 | 0.021287592 |
| LTN1 | 0.052499961 | -0.037472076 | 0.015027885 |
| RSPO3 | 0.019545799 | -0.021118781 | -0.001572982 |
| ARMC10 | 0.056817784 | -0.027272909 | 0.029544875 |
| CCDC47 | 0.071437478 | -0.086553411 | -0.015115932 |
| VAMP3 | 0.059119376 | -0.033359155 | 0.025760221 |
| RETREG3 | 0.048526418 | -0.039781472 | 0.008744945 |
| ARHGEF10L | 0.042750803 | -0.039261188 | 0.003489615 |
| NME3 | 0.08436981 | -0.065645236 | 0.018724574 |
| RPS5 | 0.105390962 | -0.047636196 | 0.057754766 |

| | | | |
|---|---|---|---|
| IGF2R | 0.05967362 | -0.051242535 | 0.008431085 |
| MARCKS | 0.091299491 | -0.060699331 | 0.03060016 |
| HOXB7 | 0.068703407 | -0.071661737 | -0.00295833 |
| LARS | 0.070415822 | -0.064832814 | 0.005583008 |
| HINT1 | 0.095336404 | -0.05125639 | 0.044080014 |
| HSP90AA1 | 0.093402289 | -0.090641822 | 0.002760467 |
| TXNL1 | 0.073632715 | -0.028194127 | 0.045438588 |
| MAPK13 | 0.055198886 | -0.028253042 | 0.026945844 |
| COMTD1 | 0.081376445 | -0.051121192 | 0.030255253 |
| RNF181 | 0.080734225 | -0.040551255 | 0.04018297 |
| PDE6D | 0.039913667 | -0.022719537 | 0.01719413 |
| SUCLA2 | 0.068493584 | -0.03372198 | 0.034771603 |
| MAX | 0.048227606 | -0.019162114 | 0.029065492 |
| YY1 | 0.06460031 | -0.054578401 | 0.010021909 |
| UBE2V1 | 0.075228909 | -0.046043716 | 0.029185193 |
| GNAI3 | 0.053123169 | -0.022924272 | 0.030198898 |
| TLE2 | 0.05376968 | -0.041118421 | 0.012651258 |
| ACO1 | 0.04798367 | -0.041360077 | 0.006623593 |
| MAP3K3 | 0.04012004 | -0.027040196 | 0.013079844 |
| ABHD17A | 0.061556168 | -0.026608305 | 0.034947863 |
| HM13 | 0.059668987 | -0.064895759 | -0.005226772 |
| TMEM164 | 0.031906451 | -0.032044753 | -0.000138302 |
| TLR5 | 0.024663971 | -0.029529328 | -0.004865357 |
| CCNI2 | 0.041642284 | -0.016685319 | 0.024956965 |
| MRPS14 | 0.058016982 | -0.019460696 | 0.038556287 |
| MANBA | 0.03549275 | -0.032352635 | 0.003140115 |
| KRR1 | 0.060552726 | -0.033731451 | 0.026821275 |
| TIMM22 | 0.045782346 | -0.024393986 | 0.02138836 |
| DHRS12 | 0.049595625 | -0.035231692 | 0.014363933 |
| SLBP | 0.070741716 | -0.02587374 | 0.044867976 |
| STT3B | 0.071045526 | -0.040256062 | 0.030789464 |
| TMEM18 | 0.056539945 | -0.028040136 | 0.028499809 |
| DLD | 0.074755018 | -0.037032251 | 0.037722767 |
| UBXN1 | 0.077902039 | -0.058620299 | 0.01928174 |
| FOSL2 | 0.048431756 | -0.03368368 | 0.014748076 |
| YPEL5 | 0.068952514 | -0.049014132 | 0.019938382 |
| SEPTIN6 | 0.062435538 | -0.04795285 | 0.014482687 |
| OPHN1 | 0.031650909 | -0.039563858 | -0.007912948 |
| DDX50 | 0.044544641 | -0.027505594 | 0.017039046 |
| DYNC2LI1 | 0.059323975 | -0.030811973 | 0.028512002 |
| EIF2D | 0.047973995 | -0.02468343 | 0.023290565 |
| TRIOBP | 0.062147284 | -0.068548489 | -0.006401205 |
| EFCAB11 | 0.032756525 | -0.019102157 | 0.013654368 |
| KIF2A | 0.037684174 | -0.022337957 | 0.015346217 |
| RECQL5 | 0.046068838 | -0.071783362 | -0.025714524 |
| APTR | 0.039647512 | -0.031266434 | 0.008381078 |
| NDUFV1 | 0.087915299 | -0.030355518 | 0.057559781 |

| | | | |
|---|---|---|---|
| ZBTB38 | 0.056076244 | -0.040860479 | 0.015215765 |
| AC019197.1 | 0.069141148 | -0.060359805 | 0.008781343 |
| ACADL | 0.049576787 | -0.05214761 | -0.002570823 |
| SVBP | 0.06152639 | -0.037782209 | 0.023744181 |
| YIPF3 | 0.071737692 | -0.034583892 | 0.0371538 |
| AASDH | 0.049802749 | -0.024558921 | 0.025243828 |
| CDK10 | 0.051233395 | -0.029429367 | 0.021804028 |
| BABAM1 | 0.059048473 | -0.023437566 | 0.035610907 |
| PSMD4 | 0.066491386 | -0.023219506 | 0.04327188 |
| RAB5C | 0.075639222 | -0.025206947 | 0.050432275 |
| ASPHD2 | 0.02135872 | -0.021909371 | -0.000550651 |
| TBL1XR1 | 0.061615071 | -0.04794141 | 0.013673661 |
| ATP5F1D | 0.091519465 | -0.041776673 | 0.049742792 |
| RABAC1 | 0.085573258 | -0.060625008 | 0.02494825 |
| PELO | 0.043377091 | -0.016232435 | 0.027144656 |
| ITGA2B | 0.018108841 | -0.014166465 | 0.003942376 |
| CYP1A1 | 0.016859275 | -0.016859275 | -5.32907E-15 |
| TM9SF4 | 0.049319158 | -0.015254508 | 0.03406465 |
| NMT1 | 0.059558694 | -0.04610559 | 0.013453103 |
| LINC01230 | 0.055023068 | -0.038272032 | 0.016751036 |
| SAFB2 | 0.055546285 | -0.040411183 | 0.015135102 |
| FUNDC2 | 0.076875144 | -0.043096891 | 0.033778252 |
| PRPF38B | 0.057666072 | -0.04637649 | 0.011289582 |
| CDKN1C | 0.091862013 | -0.029848339 | 0.062013674 |
| ACACA | 0.037975845 | -0.035141063 | 0.002834782 |
| PEX3 | 0.048286609 | -0.021695309 | 0.0265913 |
| ZNRF2 | 0.040821332 | -0.024511328 | 0.016310004 |
| ACOT13 | 0.055126126 | -0.027633007 | 0.027493119 |
| MKRN1 | 0.057574522 | -0.025975032 | 0.03159949 |
| THY1 | -0.034774251 | 0.016538779 | -0.018235472 |
| FBXO22 | 0.051328314 | -0.035676892 | 0.015651422 |
| NCAPD3 | 0.03612341 | -0.019962257 | 0.016161153 |
| H2AFJ | 0.082506415 | -0.063846286 | 0.01866013 |
| ACYP2 | 0.051904222 | -0.027768503 | 0.024135719 |
| SAYSD1 | 0.052155008 | -0.043116539 | 0.009038469 |
| YEATS4 | 0.041821911 | -0.022577995 | 0.019243916 |
| GUSB | 0.04948447 | -0.047177461 | 0.002307008 |
| TUBB2B | -0.026184435 | 0.012685179 | -0.013499256 |
| HMG20B | 0.060581744 | -0.039293792 | 0.021287952 |
| LSM14A | 0.055855195 | -0.033783342 | 0.022071853 |
| PHC2 | 0.058109511 | -0.02660062 | 0.031508891 |
| MRPS18B | 0.074819654 | -0.042524568 | 0.032295085 |
| QARS | 0.052933554 | -0.036773818 | 0.016159735 |
| CIZ1 | 0.04062132 | -0.02868888 | 0.01193244 |
| COPS6 | 0.07182487 | -0.050402208 | 0.021422662 |
| NUCB2 | 0.079265198 | -0.043417887 | 0.03584731 |
| IRF9 | 0.03774859 | -0.028795282 | 0.008953309 |

| | | | |
|---|---|---|---|
| SPCS3 | 0.078757876 | -0.027577232 | 0.051180644 |
| BHMT2 | 0.047689253 | -0.058425559 | -0.010736306 |
| IL1R1 | 0.037899948 | -0.037276779 | 0.000623169 |
| AL049629.1 | 0.017677994 | -0.01571241 | 0.001965584 |
| DENR | 0.063644821 | -0.046827753 | 0.016817068 |
| CAMTA1 | 0.08459801 | -0.036891953 | 0.047706056 |
| CLPP | 0.06304698 | -0.027990371 | 0.035056609 |
| OSBPL11 | 0.034835176 | -0.021960742 | 0.012874434 |
| AK9 | 0.033264797 | -0.026291779 | 0.006973018 |
| RPN2 | 0.083703291 | -0.08258631 | 0.001116981 |
| CLTB | 0.070532611 | -0.026033058 | 0.044499553 |
| MFF | 0.07026243 | -0.028751382 | 0.041511048 |
| PAQR8 | 0.033615069 | -0.033934658 | -0.000319588 |
| EIF3D | 0.077863328 | -0.031654123 | 0.046209205 |
| CYP17A1 | 0.015388699 | -0.014392163 | 0.000996537 |
| UNC93B1 | 0.030367359 | -0.026054928 | 0.004312431 |
| GPALPP1 | 0.041079304 | -0.02002914 | 0.021050164 |
| LGALS3BP | 0.075783123 | -0.032455325 | 0.043327798 |
| STX17 | 0.053478961 | -0.048332182 | 0.00514678 |
| LINC01341 | 0.018464852 | -0.019570725 | -0.001105874 |
| HNRNPK | 0.084216443 | -0.059206124 | 0.02501032 |
| LMO7-AS1 | 0.036083545 | -0.026807121 | 0.009276424 |
| SMAD2 | 0.056081774 | -0.035577202 | 0.020504572 |
| OCLN | 0.053777456 | -0.042063809 | 0.011713647 |
| UBA2 | 0.052946345 | -0.032706429 | 0.020239916 |
| PRXL2A | 0.070260367 | -0.118374274 | -0.048113907 |
| DRAP1 | 0.074548527 | -0.06439234 | 0.010156187 |
| LDHB | 0.093643843 | -0.032416143 | 0.0612277 |
| PTDSS2 | 0.04676942 | -0.021379107 | 0.025390313 |
| ULK1 | 0.046779905 | -0.042387209 | 0.004392695 |
| SAFB | 0.0526856 | -0.045703 | 0.0069826 |
| SLC46A3 | 0.034315398 | -0.022319907 | 0.011995492 |
| SLC9A1 | 0.028964332 | -0.025951513 | 0.003012819 |
| IRX1 | -0.01667581 | 0.006487169 | -0.010188641 |
| MSRA | 0.036582945 | -0.014523617 | 0.022059328 |
| EXPH5 | 0.044215468 | -0.046604352 | -0.002388884 |
| HOXB6 | 0.061259775 | -0.033460275 | 0.027799501 |
| RPL6 | 0.091485373 | -0.035077525 | 0.056407847 |
| MLH3 | 0.041916277 | -0.038743213 | 0.003173064 |
| LIFR | 0.055457294 | -0.051741746 | 0.003715548 |
| ZBTB7A | 0.058843145 | -0.042731441 | 0.016111704 |
| CYP39A1 | 0.038836654 | -0.022068354 | 0.0167683 |
| NIT2 | 0.056497278 | -0.036384399 | 0.020112879 |
| FAM50A | 0.065033724 | -0.042193338 | 0.022840386 |
| ATF6B | 0.049103236 | -0.034327009 | 0.014776227 |
| DDX3X | 0.070365854 | -0.070509185 | -0.000143331 |
| WDR83OS | 0.089611034 | -0.054248561 | 0.035362474 |

| | | | |
|---|---|---|---|
| NADK2 | 0.034800464 | -0.026737666 | 0.008062798 |
| TUBB | 0.077065006 | -0.042212924 | 0.034852082 |
| ATPAF1 | 0.057784476 | -0.021543684 | 0.036240792 |
| STK3 | 0.053359032 | -0.037407614 | 0.015951418 |
| KBTBD12 | 0.031945418 | -0.02992943 | 0.002015989 |
| CFAP97 | 0.061600967 | -0.043486467 | 0.0181145 |
| GTF2B | 0.0460279 | -0.018855177 | 0.027172722 |
| TXNRD2 | 0.054298466 | -0.036743724 | 0.017554742 |
| PELI1 | 0.017109019 | -0.017372427 | -0.000263408 |
| FIBP | 0.063137629 | -0.031340101 | 0.031797528 |
| LINC-PINT | 0.050425677 | -0.046990266 | 0.00343541 |
| SETD2 | 0.062124863 | -0.058304217 | 0.003820646 |
| ATXN10 | 0.059236997 | -0.027317825 | 0.031919172 |
| TSC22D4 | 0.056432921 | -0.066103692 | -0.009670771 |
| BOLA2 | 0.054003184 | -0.036253421 | 0.017749763 |
| MFAP1 | 0.052764102 | -0.029404435 | 0.023359667 |
| CHD2 | 0.066266443 | -0.058569363 | 0.00769708 |
| DNAJA2 | 0.068436703 | -0.025994259 | 0.042442444 |
| ZNF680 | 0.037565664 | -0.02187512 | 0.015690544 |
| TMEM159 | 0.03350004 | -0.026827594 | 0.006672446 |
| CHD6 | 0.048169629 | -0.027890216 | 0.020279413 |
| KMT2A | 0.060961729 | -0.054401866 | 0.006559863 |
| TIMM17B | 0.072654372 | -0.037761033 | 0.034893339 |
| ZC3H14 | 0.053262346 | -0.040367208 | 0.012895138 |
| HYAL4 | 0.016462631 | -0.017608306 | -0.001145674 |
| ATM | 0.047643642 | -0.042805028 | 0.004838614 |
| GMPR2 | 0.05170709 | -0.027700385 | 0.024006705 |
| CCNI | 0.091565019 | -0.042501178 | 0.04906384 |
| KCNG1 | 0.017494139 | -0.013844552 | 0.003649586 |
| MRPL19 | 0.067022908 | -0.048472822 | 0.018550086 |
| LGI2 | 0.018338922 | -0.020170903 | -0.001831981 |
| CRELD2 | 0.043585085 | -0.037271931 | 0.006313154 |
| MCF2L | 0.033052785 | -0.023330659 | 0.009722126 |
| L3MBTL4 | 0.029222948 | -0.020038423 | 0.009184525 |
| SLC48A1 | 0.053540397 | -0.048442783 | 0.005097613 |
| C5orf15 | 0.055079262 | -0.039550271 | 0.01552899 |
| SETD5 | 0.04343229 | -0.029386148 | 0.014046142 |
| ZNF207 | 0.062995146 | -0.051205077 | 0.011790069 |
| TRAPPC10 | 0.039803988 | -0.031788389 | 0.008015598 |
| VPS36 | 0.061400562 | -0.039569409 | 0.021831153 |
| FAM199X | 0.046794524 | -0.019816271 | 0.026978253 |
| CHST12 | 0.032574064 | -0.028571074 | 0.00400299 |
| RBM39 | 0.081396429 | -0.072893774 | 0.008502655 |
| MIR31HG | 0.014862569 | -0.016007087 | -0.001144518 |
| FAM204A | 0.05769506 | -0.045414378 | 0.012280681 |
| ECT2 | 0.02842967 | -0.018455387 | 0.009974283 |
| TLCD4 | 0.032800849 | -0.024718767 | 0.008082082 |

| | | | |
|---|---|---|---|
| PSMB3 | 0.083046591 | -0.035318894 | 0.047727697 |
| PCCA | 0.061962712 | -0.026559144 | 0.035403568 |
| ARL6IP6 | 0.040434638 | -0.039834478 | 0.00060016 |
| AKNA | 0.016701699 | -0.013526279 | 0.00317542 |
| PRPSAP2 | 0.048396438 | -0.046000496 | 0.002395942 |
| XRCC6 | 0.072094987 | -0.040939717 | 0.031155269 |
| CCNDBP1 | 0.069198749 | -0.039323472 | 0.029875277 |
| PTGER3 | -0.101790511 | 0.17632848 | 0.074537969 |
| THOC2 | 0.053815642 | -0.051675674 | 0.002139968 |
| DDHD2 | 0.042999059 | -0.025761642 | 0.017237417 |
| TRAP1 | 0.052103115 | -0.026760657 | 0.025342458 |
| COMMD6 | 0.092871348 | -0.031134282 | 0.061737065 |
| AQR | 0.045681407 | -0.032960929 | 0.012720478 |
| HIP1 | 0.036789018 | -0.055641889 | -0.01885287 |
| CEP78 | 0.056770025 | -0.061829785 | -0.005059761 |
| AL353572.4 | 0.025254912 | -0.009314483 | 0.015940429 |
| CCND3 | 0.026950681 | -0.019727904 | 0.007222777 |
| CYP2R1 | 0.025627436 | -0.023903784 | 0.001723651 |
| ELK4 | 0.048193703 | -0.031075362 | 0.017118341 |
| EPHB6 | 0.023026492 | -0.022968826 | 5.76656E-05 |
| ANAPC16 | 0.089165064 | -0.051787459 | 0.037377605 |
| ENPP4 | 0.045063506 | -0.037814866 | 0.00724864 |
| UBE3A | 0.06022632 | -0.032075995 | 0.028150324 |
| MED13L | 0.045717469 | -0.032401594 | 0.013315875 |
| MRPS34 | 0.075130285 | -0.038245239 | 0.036885047 |
| RBM6 | 0.056298299 | -0.056933629 | -0.000635331 |
| EPB41L5 | 0.050355528 | -0.048782998 | 0.00157253 |
| CXCL16 | 0.028601677 | -0.015045012 | 0.013556665 |
| MED21 | 0.058041255 | -0.020429251 | 0.037612004 |
| CHRDL2 | 0.018115546 | -0.022222535 | -0.004106989 |
| CASD1 | 0.044499771 | -0.032999692 | 0.011500079 |
| NDUFS7 | 0.088477037 | -0.04070917 | 0.047767868 |
| HIST1H4C | 0.095087048 | -0.101602485 | -0.006515437 |
| STRADB | 0.032304222 | -0.027082885 | 0.005221337 |
| SNRNP48 | 0.033665705 | -0.02017776 | 0.013487945 |
| MTREX | 0.047878734 | -0.024792494 | 0.02308624 |
| PAXBP1 | 0.039288004 | -0.028953701 | 0.010334303 |
| PHF5A | 0.047539908 | -0.021515022 | 0.026024886 |
| TRIM5 | 0.027706199 | -0.014916498 | 0.0127897 |
| DNAH10 | 0.013771888 | -0.013771888 | -5.32907E-15 |
| ZC3H6 | 0.042488924 | -0.030851702 | 0.011637222 |
| N4BP3 | 0.036951282 | -0.0269238 | 0.010027482 |
| SCAPER | 0.04353663 | -0.029737784 | 0.013798846 |
| CGRRF1 | 0.042471871 | -0.030063507 | 0.012408363 |
| TFG | 0.064739597 | -0.029006929 | 0.035732668 |
| HIGD2A | 0.087316947 | -0.03675343 | 0.050563517 |
| CAPN7 | 0.043695005 | -0.037840352 | 0.005854652 |

| | | | |
|---|---|---|---|
| CBY1 | 0.052563995 | -0.016834414 | 0.035729581 |
| KRT10 | 0.081131875 | -0.062864999 | 0.018266876 |
| CRYM | 0.021966958 | -0.02334432 | -0.001377362 |
| JAGN1 | 0.056413715 | -0.047084093 | 0.009329622 |
| WDR72 | 0.069151831 | -0.070640838 | -0.001489007 |
| FKBP8 | 0.074182242 | -0.025464322 | 0.04871792 |
| EMILIN2 | 0.017931462 | -0.016733287 | 0.001198175 |
| GTF2A2 | 0.078742807 | -0.031056553 | 0.047686253 |
| ANXA7 | 0.067636489 | -0.050430508 | 0.017205981 |
| FGD5-AS1 | 0.066471054 | -0.042928464 | 0.023542591 |
| TMEM123 | 0.076874613 | -0.043901989 | 0.032972625 |
| PRPF6 | 0.054539638 | -0.021025067 | 0.033514571 |
| ACSL3 | 0.071748789 | -0.043224068 | 0.028524721 |
| NUDCD2 | 0.061070975 | -0.029901301 | 0.031169674 |
| NOSIP | 0.056901737 | -0.022540305 | 0.034361432 |
| EIF3H | 0.083866464 | -0.035012186 | 0.048854278 |
| BABAM2 | 0.044894323 | -0.025295097 | 0.019599226 |
| C1orf115 | 0.032732808 | -0.034280233 | -0.001547424 |
| CCNC | 0.058261056 | -0.019164685 | 0.039096372 |
| FKBP3 | 0.066646322 | -0.028197972 | 0.03844835 |
| HSP90B1 | 0.09141134 | -0.050223378 | 0.041187962 |
| EPN2 | 0.043269514 | -0.032842561 | 0.010426953 |
| STK25 | 0.054480861 | -0.043147658 | 0.011333202 |
| SH3YL1 | 0.071641305 | -0.046316492 | 0.025324813 |
| LINC00472 | 0.074534663 | -0.090126684 | -0.015592021 |
| PLPP3 | 0.051505079 | -0.059936844 | -0.008431765 |
| KATNBL1 | 0.053852526 | -0.032906358 | 0.020946169 |
| RNF8 | 0.045145564 | -0.032808587 | 0.012336976 |
| MPRIP | 0.050916951 | -0.032363866 | 0.018553085 |
| ABCC5 | 0.062694022 | -0.032063674 | 0.030630348 |
| ALG5 | 0.066795796 | -0.037193741 | 0.029602055 |
| KIF25 | 0.015696513 | -0.016239681 | -0.000543168 |
| LRRFIP2 | 0.060071309 | -0.035184845 | 0.024886464 |
| SRC | 0.034816347 | -0.029966463 | 0.004849884 |
| TOR4A | 0.022007623 | -0.024047159 | -0.002039537 |
| SRSF11 | 0.070608028 | -0.061934818 | 0.00867321 |
| CERS5 | 0.032933504 | -0.01270748 | 0.020226024 |
| CCNO | 0.016543498 | -0.019341884 | -0.002798386 |
| ZMPSTE24 | 0.080557675 | -0.050036345 | 0.030521329 |
| B4GAT1 | 0.039567542 | -0.022442263 | 0.017125279 |
| MT1G | -0.047118579 | 0.028447414 | -0.018671165 |
| PDS5B | 0.045419613 | -0.029510416 | 0.015909197 |
| AIMP1 | 0.067709291 | -0.05203134 | 0.015677951 |
| CCT8 | 0.07242781 | -0.029979458 | 0.042448352 |
| PTK2 | 0.05665904 | -0.047793724 | 0.008865315 |
| SOWAHD | 0.014119935 | -0.013725771 | 0.000394164 |
| DNAJC19 | 0.077716775 | -0.041627924 | 0.036088851 |

| | | | |
|---|---|---|---|
| INO80E | 0.042851129 | -0.037418311 | 0.005432818 |
| HOXB2 | 0.06067375 | -0.035364145 | 0.025309605 |
| ATP2A3 | 0.018207724 | -0.017836462 | 0.000371262 |
| EML6 | 0.020325433 | -0.022918683 | -0.002593251 |
| CDK5 | 0.028675044 | -0.027686706 | 0.000988338 |
| ALG14 | 0.041337638 | -0.020025331 | 0.021312307 |
| TMEM70 | 0.051273912 | -0.021722784 | 0.029551129 |
| ENTPD8 | 0.013404207 | -0.013186862 | 0.000217345 |
| TUT4 | 0.058109506 | -0.050908766 | 0.007200741 |
| TMX2 | 0.063880676 | -0.032017343 | 0.031863333 |
| SQSTM1 | 0.074128186 | -0.03210126 | 0.042026926 |
| IRF2 | 0.043989687 | -0.031782186 | 0.012207501 |
| BTBD7 | 0.050973137 | -0.057174413 | -0.006201276 |
| BCKDHA | 0.067476803 | -0.033516596 | 0.033960207 |
| CES2 | 0.041400615 | -0.030471686 | 0.010928929 |
| SMIM19 | 0.06693471 | -0.041813677 | 0.025121032 |
| MT1E | -0.061810504 | 0.049997706 | -0.011812798 |
| R3HDM4 | 0.046367538 | -0.041073123 | 0.005294416 |
| SUMO3 | 0.060979204 | -0.044203032 | 0.016776172 |
| LPIN3 | 0.029788158 | -0.027516545 | 0.002271613 |
| POLI | 0.041403655 | -0.029926355 | 0.0114773 |
| PRPF8 | 0.064933871 | -0.047246098 | 0.017687773 |
| NOA1 | 0.032241162 | -0.014330735 | 0.017910426 |
| FCGRT | 0.07885864 | -0.057352741 | 0.021505899 |
| SETX | 0.054317111 | -0.047263449 | 0.007053663 |
| LSM12 | 0.061331695 | -0.026264972 | 0.035066722 |
| PACSIN2 | 0.04496441 | -0.041255113 | 0.003709297 |
| MORC3 | 0.038668388 | -0.034024827 | 0.004643561 |
| NR1H2 | 0.058393733 | -0.032456382 | 0.025937351 |
| TM9SF1 | 0.056308921 | -0.040570829 | 0.015738092 |
| MRPS9 | 0.045806752 | -0.022844416 | 0.022962336 |
| FDFT1 | 0.052443873 | -0.035896136 | 0.016547737 |
| INTS8 | 0.026702427 | -0.01363264 | 0.013069787 |
| TMEM256 | 0.073166597 | -0.04193088 | 0.031235717 |
| AL049828.1 | 0.021400504 | -0.022881622 | -0.001481117 |
| SLCO3A1 | 0.02705459 | -0.020966573 | 0.006088017 |
| PRXL2C | 0.037422328 | -0.021281684 | 0.016140644 |
| CCDC90B | 0.052659675 | -0.018264639 | 0.034395036 |
| FAM174A | 0.064777079 | -0.029038468 | 0.03573861 |
| SMARCA5 | 0.06239119 | -0.040643209 | 0.02174798 |
| UFSP2 | 0.056366839 | -0.055437728 | 0.000929111 |
| SMDT1 | 0.090451251 | -0.040442158 | 0.050009093 |
| C6orf132 | 0.024719552 | -0.027330355 | -0.002610803 |
| AP003068.2 | 0.023052755 | -0.014021123 | 0.009031632 |
| RTF1 | 0.061453098 | -0.047914423 | 0.013538675 |
| MACO1 | 0.048744846 | -0.037320016 | 0.01142483 |
| KCNQ1 | 0.049141178 | -0.040086921 | 0.009054258 |

| | | | |
|---|---|---|---|
| SLC12A3 | -0.064575139 | 0.114284533 | 0.049709394 |
| OSTF1 | 0.054851341 | -0.035615183 | 0.019236157 |
| CMYA5 | 0.047884347 | -0.065135122 | -0.017250775 |
| RGL3 | 0.055253729 | -0.031612317 | 0.023641411 |
| ZRANB2 | 0.065204087 | -0.066307958 | -0.001103871 |
| ARPC5L | 0.063251156 | -0.050779491 | 0.012471665 |
| FKBP9 | 0.047161991 | -0.039179383 | 0.007982608 |
| C5orf24 | 0.052780574 | -0.051232899 | 0.001547675 |
| CR1L | 0.028212084 | -0.01727058 | 0.010941505 |
| GPD2 | 0.037311961 | -0.019933419 | 0.017378543 |
| SLC27A4 | 0.043579681 | -0.021954165 | 0.021625516 |
| GRSF1 | 0.056514885 | -0.033635344 | 0.022879541 |
| OTULINL | 0.019902694 | -0.025402948 | -0.005500254 |
| MEAF6 | 0.064940268 | -0.021669354 | 0.043270915 |
| EIF4G2 | 0.077718482 | -0.064394233 | 0.013324249 |
| CHMP3 | 0.069437113 | -0.037070952 | 0.03236616 |
| KIF21A | 0.077045921 | -0.040812299 | 0.036233621 |
| TUBGCP2 | 0.051971412 | -0.036223506 | 0.015747906 |
| ARL8A | 0.051420349 | -0.023134664 | 0.028285685 |
| FTX | 0.055624992 | -0.032079686 | 0.023545306 |
| RNASET2 | 0.046416375 | -0.051499331 | -0.005082955 |
| AC024560.1 | 0.038158811 | -0.028053517 | 0.010105295 |
| LUZP1 | 0.046003905 | -0.035791138 | 0.010212767 |
| TAF1B | 0.037516631 | -0.032120237 | 0.005396394 |
| SLC12A7 | 0.042206368 | -0.030364801 | 0.011841567 |
| AC004148.1 | 0.027851694 | -0.026234696 | 0.001616997 |
| C8orf82 | 0.058296317 | -0.020528523 | 0.037767794 |
| VPS37D | 0.044054679 | -0.018078777 | 0.025975902 |
| CIT | 0.034381694 | -0.028036188 | 0.006345506 |
| POLR2E | 0.06162755 | -0.037693137 | 0.023934413 |
| ALDH5A1 | 0.035020579 | -0.012292163 | 0.022728416 |
| STK38 | 0.034891107 | -0.023591922 | 0.011299186 |
| RORA | 0.036697642 | -0.032245203 | 0.004452439 |
| VCL | 0.054035452 | -0.043573332 | 0.01046212 |
| PER3 | 0.043424586 | -0.037345288 | 0.006079298 |
| VKORC1L1 | 0.036647012 | -0.01446701 | 0.022180002 |
| FMN1 | 0.026945936 | -0.030781278 | -0.003835341 |
| RRAS | 0.035665474 | -0.025168473 | 0.010497 |
| GABBR1 | 0.026104895 | -0.02351279 | 0.002592105 |
| UBA6 | 0.045124358 | -0.034450003 | 0.010674355 |
| AIFM2 | 0.028392034 | -0.023448175 | 0.004943859 |
| CFDP1 | 0.060110523 | -0.038437075 | 0.021673448 |
| DEXI | 0.045706313 | -0.027395391 | 0.018310921 |
| MOB2 | 0.040946951 | -0.035669344 | 0.005277607 |
| ZBTB8OS | 0.055254747 | -0.019508536 | 0.035746211 |
| PPP1R12A | 0.046357946 | -0.02661568 | 0.019742265 |
| SLC22A23 | 0.043493676 | -0.0379061 | 0.005587576 |

| | | | |
|---|---|---|---|
| SP100 | 0.043458552 | -0.044665441 | -0.001206889 |
| FLNB | 0.062478475 | -0.025664768 | 0.036813708 |
| AL353699.1 | 0.022274612 | -0.026474995 | -0.004200383 |
| CD320 | 0.052145057 | -0.03799618 | 0.014148876 |
| CKS2 | 0.039412056 | -0.013448464 | 0.025963592 |
| SLC25A28 | 0.038992614 | -0.02126855 | 0.017724065 |
| RSL1D1 | 0.064697422 | -0.028415258 | 0.036282163 |
| HNRNPD | 0.068563346 | -0.028675374 | 0.039887972 |
| BAZ1A | 0.027696377 | -0.027813753 | -0.000117376 |
| LIPC | 0.018714556 | -0.017663238 | 0.001051317 |
| MEA1 | 0.056260325 | -0.0224703 | 0.033790025 |
| BLNK | 0.046300217 | -0.046682704 | -0.000382487 |
| CIAO2B | 0.078927931 | -0.032803223 | 0.046124708 |
| ACIN1 | 0.053013855 | -0.045042724 | 0.007971132 |
| NT5DC1 | 0.056431356 | -0.03470935 | 0.021722006 |
| BEX2 | -0.058531308 | 0.028655985 | -0.029875324 |
| ACOT9 | 0.035640847 | -0.025183732 | 0.010457115 |
| NUBP2 | 0.057239379 | -0.035297833 | 0.021941546 |
| PTPRN2 | 0.024569777 | -0.024241755 | 0.000328022 |
| GALNT4 | 0.028587144 | -0.022858615 | 0.005728529 |
| ARHGAP35 | 0.047996095 | -0.052747261 | -0.004751166 |
| GRINA | 0.055667415 | -0.046751498 | 0.008915917 |
| MIR22HG | 0.047877605 | -0.024839314 | 0.023038291 |
| CLPTM1L | 0.053512975 | -0.051571825 | 0.00194115 |
| ZNF107 | 0.02496434 | -0.023442544 | 0.001521796 |
| NF1 | 0.051030259 | -0.034243682 | 0.016786576 |
| OGT | 0.055956067 | -0.053615509 | 0.002340559 |
| CTDSPL | 0.062205915 | -0.034615666 | 0.027590249 |
| UBA3 | 0.040947143 | -0.030614648 | 0.010332495 |
| EML4 | 0.043595069 | -0.023357137 | 0.020237933 |
| WBP4 | 0.045351492 | -0.029181484 | 0.016170008 |
| KMT2C | 0.054672961 | -0.052555555 | 0.002117407 |
| SHMT1 | 0.040508771 | -0.049962595 | -0.009453825 |
| ZNF236 | 0.02839047 | -0.018612805 | 0.009777665 |
| PSMC2 | 0.05156738 | -0.021770205 | 0.029797175 |
| NUDT5 | 0.049694299 | -0.024481077 | 0.025213222 |
| SNHG6 | 0.081248506 | -0.058853636 | 0.02239487 |
| TFAM | 0.05722201 | -0.060116526 | -0.002894516 |
| MTOR | 0.038777337 | -0.035812283 | 0.002965054 |
| MRPL40 | 0.070575907 | -0.037057151 | 0.033518756 |
| MBTPS1 | 0.060022209 | -0.049072494 | 0.010949715 |
| SLC2A4RG | 0.060484034 | -0.039864576 | 0.020619458 |
| RWDD4 | 0.063607158 | -0.038256372 | 0.025350786 |
| PDCD2 | 0.057277136 | -0.036900979 | 0.020376157 |
| ANGPTL1 | 0.066172808 | -0.031512946 | 0.034659861 |
| SMIM7 | 0.069878793 | -0.036739685 | 0.033139108 |
| B4GALT1 | 0.053311731 | -0.047551339 | 0.005760392 |

| | | | |
|---|---|---|---|
| ABTB1 | 0.032501373 | -0.028691463 | 0.003809909 |
| PTTG1IP | 0.072052086 | -0.077200121 | -0.005148034 |
| ACTR6 | 0.044842728 | -0.018519667 | 0.026323061 |
| SUCO | 0.050680797 | -0.023733309 | 0.026947488 |
| ARHGEF16 | 0.03852022 | -0.0337897 | 0.004730521 |
| DAP | 0.06862578 | -0.056262661 | 0.01236312 |
| SSBP4 | 0.044618949 | -0.042467864 | 0.002151085 |
| ATP13A2 | 0.032438323 | -0.027560972 | 0.004877351 |
| ACOT4 | 0.031972344 | -0.025896372 | 0.006075972 |
| SUZ12 | 0.056006862 | -0.043037079 | 0.012969783 |
| IQSEC1 | 0.039709224 | -0.022161221 | 0.017548003 |
| STX16 | 0.052980691 | -0.027798164 | 0.025182526 |
| BEND7 | 0.044368772 | -0.035864323 | 0.008504449 |
| TRIM56 | 0.046200696 | -0.02735366 | 0.018847036 |
| TMEM161B | 0.041132984 | -0.029012151 | 0.012120833 |
| HERC5 | 0.022413791 | -0.02076146 | 0.001652331 |
| IRS2 | 0.030903447 | -0.030101 | 0.000802448 |
| ACTR10 | 0.064995186 | -0.022695264 | 0.042299922 |
| MKLN1-AS | 0.019902491 | -0.021428564 | -0.001526073 |
| WIPI2 | 0.054881486 | -0.03693268 | 0.017948806 |
| NARS | 0.068423267 | -0.03356673 | 0.034856537 |
| IST1 | 0.05368852 | -0.029688039 | 0.024000481 |
| PER2 | 0.028860875 | -0.017039626 | 0.011821249 |
| FBXW5 | 0.055849205 | -0.045765163 | 0.010084042 |
| LINC00324 | 0.022795215 | -0.019451507 | 0.003343708 |
| SIVA1 | 0.074549492 | -0.038966439 | 0.035583053 |
| HLTF | 0.05007024 | -0.045532366 | 0.004537874 |
| ELMO3 | 0.032812931 | -0.026901875 | 0.005911056 |
| CARMIL1 | 0.06284526 | -0.061863241 | 0.000982019 |
| TPRG1L | 0.040690924 | -0.017206874 | 0.02348405 |
| AP003498.2 | 0.034503776 | -0.025550416 | 0.00895336 |
| SSU72 | 0.074647287 | -0.043925791 | 0.030721496 |
| GATAD1 | 0.054511556 | -0.028684584 | 0.025826972 |
| ALDH3A2 | 0.04631646 | -0.021890617 | 0.024425843 |
| CRELD1 | 0.043968552 | -0.023297042 | 0.02067151 |
| TERF2IP | 0.063993333 | -0.022106195 | 0.041887138 |
| IDH3B | 0.054969542 | -0.028875224 | 0.026094318 |
| KIAA2013 | 0.04686321 | -0.025352705 | 0.021510506 |
| ECSIT | 0.048103706 | -0.033436282 | 0.014667424 |
| LACTB2 | 0.04983634 | -0.018648152 | 0.031188188 |
| C6orf136 | 0.048458304 | -0.025126455 | 0.023331849 |
| PSIP1 | 0.045785433 | -0.036979701 | 0.008805732 |
| TMCC3 | 0.027735263 | -0.026412053 | 0.00132321 |
| SLC2A1 | 0.031220835 | -0.026584382 | 0.004636453 |
| GPC3 | -0.051146438 | 0.046476108 | -0.004670329 |
| SH3GLB2 | 0.058385085 | -0.038681622 | 0.019703463 |
| PHACTR2 | 0.044162665 | -0.024970157 | 0.019192508 |

| | | | |
|---|---|---|---|
| SH3GLB1 | 0.059706022 | -0.032514205 | 0.027191817 |
| KRCC1 | 0.039235044 | -0.024185073 | 0.01504997 |
| UNG | 0.040456164 | -0.025609408 | 0.014846756 |
| PUF60 | 0.060797092 | -0.023989727 | 0.036807365 |
| SH3BP5-AS1 | 0.017865836 | -0.011582035 | 0.006283801 |
| ETNK1 | 0.055836152 | -0.028990849 | 0.026845304 |
| MRPS10 | 0.054023486 | -0.025347936 | 0.028675551 |
| SHARPIN | 0.057224426 | -0.020701289 | 0.036523136 |
| LUC7L3 | 0.075061429 | -0.048552738 | 0.026508691 |
| IRF7 | 0.028293079 | -0.028190918 | 0.000102161 |
| FAM32A | 0.049099515 | -0.026191702 | 0.022907813 |
| KLF6 | 0.051804732 | -0.067203809 | -0.015399076 |
| ZNF567 | 0.029835412 | -0.025057091 | 0.004778322 |
| PRKCD | 0.058335434 | -0.033423249 | 0.024912185 |
| AKIRIN1 | 0.058663618 | -0.030167111 | 0.028496507 |
| CRAT | 0.036618155 | -0.027024445 | 0.00959371 |
| LARP7 | 0.054457549 | -0.04465412 | 0.00980343 |
| OSTM1 | 0.033930656 | -0.031068188 | 0.002862468 |
| EXOC5 | 0.038828072 | -0.019160551 | 0.019667521 |
| KIF3A | 0.043715658 | -0.032094582 | 0.011621076 |
| ZNF292 | 0.054522154 | -0.041723302 | 0.012798852 |
| TCEAL4 | 0.067893236 | -0.031060963 | 0.036832273 |
| MCF2L-AS1 | 0.029093677 | -0.027127249 | 0.001966428 |
| MARVELD3 | 0.028307834 | -0.032784626 | -0.004476792 |
| LIMK2 | 0.020302302 | -0.019435965 | 0.000866337 |
| NDUFS1 | 0.067027849 | -0.043522131 | 0.023505718 |
| SLC37A4 | 0.035954906 | -0.029096571 | 0.006858335 |
| CTNS | 0.035120357 | -0.012157742 | 0.022962615 |
| UBR4 | 0.030412293 | -0.019127195 | 0.011285098 |
| PLEKHH1 | 0.037595437 | -0.043244298 | -0.005648861 |
| SMARCA2 | 0.051836749 | -0.038153173 | 0.013683576 |
| BRMS1 | 0.046259815 | -0.027155118 | 0.019104697 |
| ZDHHC4 | 0.054482081 | -0.035737041 | 0.018745041 |
| DIMT1 | 0.02904758 | -0.012258757 | 0.016788823 |
| KLHL36 | 0.046507463 | -0.028056729 | 0.018450733 |
| C1orf53 | 0.049407214 | -0.029253025 | 0.020154189 |
| PRDM16-DT | 0.067821301 | -0.080282387 | -0.012461086 |
| PKP2 | 0.029716413 | -0.026339569 | 0.003376844 |
| TSG101 | 0.065686447 | -0.033912435 | 0.031774012 |
| CAPZA2 | 0.071513249 | -0.038975042 | 0.032538207 |
| BEX3 | -0.085184318 | 0.068716813 | -0.016467505 |
| FCGBP | 0.023444852 | -0.032420467 | -0.008975615 |
| FARSA | 0.045782413 | -0.022580398 | 0.023202015 |
| AGRN | 0.040435919 | -0.034143734 | 0.006292185 |
| TTC19 | 0.058379304 | -0.046317753 | 0.012061551 |
| IK | 0.054551442 | -0.03584961 | 0.018701832 |
| CXXC1 | 0.037084195 | -0.024930829 | 0.012153365 |

| | | | |
|---|---|---|---|
| STX4 | 0.040816229 | -0.032815642 | 0.008000587 |
| MYSM1 | 0.044227145 | -0.037091942 | 0.007135203 |
| CELF1 | 0.055398223 | -0.036321596 | 0.019076627 |
| PLEKHA3 | 0.052787711 | -0.025269753 | 0.027517957 |
| CCDC134 | 0.024159727 | -0.017504555 | 0.006655173 |
| MAFG | 0.027698439 | -0.028534448 | -0.000836009 |
| SLC30A5 | 0.048583114 | -0.02303777 | 0.025545344 |
| CCL28 | 0.020617454 | -0.020864637 | -0.000247184 |
| MBNL1 | 0.053830028 | -0.047099179 | 0.006730849 |
| ARFGEF2 | 0.047262308 | -0.03349886 | 0.013763448 |
| GRB2 | 0.052056413 | -0.0304383 | 0.021618113 |
| PDHX | 0.043786982 | -0.018853525 | 0.024933457 |
| ERLIN1 | 0.0308727 | -0.026489194 | 0.004383506 |
| CCDC127 | 0.033069124 | -0.014586996 | 0.018482128 |
| CAAP1 | 0.047017283 | -0.043433337 | 0.003583946 |
| EIF3L | 0.075709475 | -0.042001528 | 0.033707947 |
| MOAP1 | 0.042905855 | -0.026685335 | 0.01622052 |
| VBP1 | 0.060761869 | -0.022134091 | 0.038627778 |
| TJP2 | 0.042845017 | -0.035775839 | 0.007069178 |
| ISG15 | 0.034378987 | -0.019315405 | 0.015063582 |
| SLC35F2 | 0.019413253 | -0.019114229 | 0.000299024 |
| ATG4D | 0.040882049 | -0.021998743 | 0.018883306 |
| TOP3B | 0.030014371 | -0.028276406 | 0.001737965 |
| MAP4K3-DT | 0.064231077 | -0.036024023 | 0.028207054 |
| LSR | 0.04773184 | -0.03953117 | 0.00820067 |
| ENDOG | 0.057394833 | -0.021587886 | 0.035806947 |
| TRIM8 | 0.069028766 | -0.036028076 | 0.03300069 |
| SECISBP2L | 0.055524058 | -0.034680228 | 0.02084383 |
| SYF2 | 0.070725999 | -0.048815864 | 0.021910136 |
| SF3B1 | 0.070519328 | -0.044832168 | 0.02568716 |
| STX5 | 0.040294639 | -0.024610118 | 0.015684521 |
| SLC44A4 | 0.019117999 | -0.025655269 | -0.00653727 |
| MCOLN1 | 0.035142521 | -0.026280798 | 0.008861723 |
| SMAD4 | 0.05009231 | -0.03031568 | 0.01977663 |
| MRPS25 | 0.064544302 | -0.030553343 | 0.033990959 |
| STX17-AS1 | 0.023686508 | -0.017638114 | 0.006048393 |
| KATNA1 | 0.033846533 | -0.019378927 | 0.014467606 |
| OSBP | 0.049486908 | -0.039455484 | 0.010031423 |
| MPHOSPH6 | 0.04557255 | -0.023188844 | 0.022383706 |
| ASB13 | 0.037129551 | -0.027047734 | 0.010081817 |
| GLUD1 | 0.071119351 | -0.050606652 | 0.020512699 |
| FAN1 | 0.042866196 | -0.026428788 | 0.016437408 |
| PDP1 | 0.042281273 | -0.028996257 | 0.013285016 |
| EFHD1 | 0.082114671 | -0.03970183 | 0.042412841 |
| RAB10 | 0.061923213 | -0.031607274 | 0.030315939 |
| ZNF726 | 0.027170565 | -0.021591766 | 0.0055788 |
| ACAP2 | 0.045211252 | -0.029963365 | 0.015247887 |

| | | | |
|---|---|---|---|
| C11orf54 | 0.064820978 | -0.048145652 | 0.016675326 |
| ANKLE2 | 0.039783963 | -0.027058548 | 0.012725415 |
| EIF3G | 0.075297737 | -0.035353607 | 0.03994413 |
| CDC73 | 0.045886454 | -0.019356827 | 0.026529626 |
| CCDC186 | 0.056018838 | -0.06310389 | -0.007085052 |
| EIF3I | 0.064262001 | -0.04676613 | 0.01749587 |
| B3GLCT | 0.02833759 | -0.015953146 | 0.012384444 |
| ZNF652 | 0.048474462 | -0.030648825 | 0.017825637 |
| AUH | 0.057541241 | -0.026769846 | 0.030771395 |
| CEP104 | 0.034896904 | -0.031572928 | 0.003323977 |
| DCAF13 | 0.045784229 | -0.019629934 | 0.026154295 |
| ZCCHC8 | 0.032996445 | -0.02088753 | 0.012108915 |
| SRGAP3 | 0.041395187 | -0.028688852 | 0.012706335 |
| ACTR2 | 0.063490058 | -0.043856687 | 0.019633371 |
| RCOR3 | 0.042962173 | -0.028617261 | 0.014344911 |
| CIAPIN1 | 0.043688618 | -0.029817949 | 0.013870669 |
| TES | 0.038781378 | -0.041163962 | -0.002382584 |
| LPL | -0.015842293 | 0.008263371 | -0.007578922 |
| CBWD1 | 0.049865216 | -0.02046697 | 0.029398246 |
| CTNNB1 | 0.064663499 | -0.038886971 | 0.025776528 |
| SSTR2 | 0.012707946 | -0.013539343 | -0.000831397 |
| CSTB | 0.082291997 | -0.067409084 | 0.014882914 |
| ARRDC1 | 0.048436679 | -0.03587088 | 0.012565799 |
| ADAM9 | 0.045316854 | -0.024659237 | 0.020657617 |
| EIF1 | 0.081936705 | -0.033306743 | 0.048629962 |
| GNG11 | 0.035794718 | -0.024040085 | 0.011754633 |
| GATA2-AS1 | 0.027253654 | -0.025160784 | 0.002092869 |
| RBBP4 | 0.059499695 | -0.033535769 | 0.025963926 |
| TBC1D23 | 0.031896912 | -0.013810144 | 0.018086767 |
| TNPO1 | 0.049099967 | -0.028882086 | 0.020217881 |
| RGL2 | 0.03045653 | -0.023541054 | 0.006915476 |
| LINC00623 | 0.044606202 | -0.022067228 | 0.022538974 |
| SOS1 | 0.038523496 | -0.029713151 | 0.008810344 |
| DPP3 | 0.029326684 | -0.018263883 | 0.011062801 |
| PHIP | 0.050304654 | -0.051418774 | -0.00111412 |
| NSUN6 | 0.039095505 | -0.03146024 | 0.007635266 |
| SMIM20 | 0.059139453 | -0.035373683 | 0.02376577 |
| CYB5B | 0.049513993 | -0.028680487 | 0.020833506 |
| SMARCA1 | 0.051825787 | -0.039789577 | 0.01203621 |
| CCDC82 | 0.039193253 | -0.036664229 | 0.002529024 |
| TMUB2 | 0.040131616 | -0.01847451 | 0.021657106 |
| CTBS | 0.051311659 | -0.031491528 | 0.019820131 |
| GTPBP6 | 0.049813926 | -0.021061468 | 0.028752457 |
| PRRG2 | 0.033983737 | -0.026466518 | 0.007517219 |
| LHFPL3-AS2 | 0.017768644 | -0.022971607 | -0.005202963 |
| THUMPD1 | 0.054553432 | -0.03226741 | 0.022286022 |
| IFNAR2 | 0.024049908 | -0.020053676 | 0.003996232 |

| | | | |
|---|---|---|---|
| TNFRSF14 | 0.036052412 | -0.028249971 | 0.007802441 |
| NSFL1C | 0.048840713 | -0.019593498 | 0.029247215 |
| BCL10 | 0.047593867 | -0.020143433 | 0.027450435 |
| SYT7 | 0.020990884 | -0.021412722 | -0.000421838 |
| SLC36A4 | 0.034335631 | -0.041697261 | -0.00736163 |
| RAB27A | 0.029225191 | -0.021990978 | 0.007234212 |
| EEA1 | 0.04538218 | -0.044295069 | 0.001087111 |
| MANF | 0.055512533 | -0.043755455 | 0.011757078 |
| IMPAD1 | 0.048000825 | -0.03260629 | 0.015394535 |
| AP2M1 | 0.067512382 | -0.026647946 | 0.040864436 |
| MTPN | 0.059605849 | -0.047130677 | 0.012475172 |
| FAM219B | 0.035479081 | -0.038479441 | -0.00300036 |
| PDIA4 | 0.046458477 | -0.023102347 | 0.02335613 |
| HSPA1B | 0.078835149 | -0.0504577 | 0.028377449 |
| VPS37B | 0.043126771 | -0.022072412 | 0.02105436 |
| RSBN1 | 0.041056801 | -0.025962567 | 0.015094234 |
| WSCD2 | 0.011290424 | -0.011001304 | 0.00028912 |
| CCS | 0.054702512 | -0.025681743 | 0.029020769 |
| RBM47 | 0.070139352 | -0.044660326 | 0.025479026 |
| THAP2 | 0.031070028 | -0.03095076 | 0.000119268 |
| UBIAD1 | 0.026364849 | -0.013150701 | 0.013214148 |
| WDR13 | 0.05473934 | -0.035269311 | 0.01947003 |
| RNF6 | 0.034894873 | -0.023043496 | 0.011851377 |
| PRR5L | 0.012649708 | -0.011819662 | 0.000830045 |
| B4GALT3 | 0.038454568 | -0.031913718 | 0.00654085 |
| ROGDI | 0.044665753 | -0.021160916 | 0.023504836 |
| STAMBP | 0.041555549 | -0.025349154 | 0.016206396 |
| CMBL | 0.049724628 | -0.039497951 | 0.010226677 |
| ANKRD49 | 0.040992788 | -0.029402651 | 0.011590136 |
| MIEN1 | 0.072600952 | -0.044437427 | 0.028163525 |
| RTKN | 0.034656211 | -0.02168519 | 0.012971022 |
| B4GALT7 | 0.042770027 | -0.021985528 | 0.020784499 |
| MTHFS | 0.055071592 | -0.022765314 | 0.032306279 |
| TRPV6 | 0.013554912 | -0.012551059 | 0.001003853 |
| SNX9 | 0.043295036 | -0.037310972 | 0.005984064 |
| EXOC4 | 0.035023805 | -0.019187089 | 0.015836717 |
| COPB1 | 0.056972555 | -0.050127128 | 0.006845426 |
| TVP23B | 0.029274118 | -0.025721021 | 0.003553097 |
| TADA3 | 0.045171852 | -0.034267737 | 0.010904115 |
| ACP1 | 0.06630146 | -0.049739401 | 0.01656206 |
| CSNK2A2 | 0.040272863 | -0.028983343 | 0.01128952 |
| RAB40B | 0.058839685 | -0.04459012 | 0.014249565 |
| KIF13A | 0.036338426 | -0.040918253 | -0.004579828 |
| NFX1 | 0.035019851 | -0.024430755 | 0.010589097 |
| AP1G2 | 0.039349463 | -0.036251969 | 0.003097494 |
| ATP13A1 | 0.034954782 | -0.029467823 | 0.005486959 |
| ZNF888 | 0.023971676 | -0.022697168 | 0.001274508 |

| | | | |
|---|---|---|---|
| RNMT | 0.054338982 | -0.029982292 | 0.02435669 |
| SUPT16H | 0.049843701 | -0.024110419 | 0.025733282 |
| GBA2 | 0.045560449 | -0.033094272 | 0.012466177 |
| POU5F1 | 0.019284732 | -0.020739085 | -0.001454353 |
| SPRY1 | 0.038024605 | -0.027697551 | 0.010327054 |
| TMBIM1 | 0.039618031 | -0.041255233 | -0.001637202 |
| ATE1 | 0.042291818 | -0.039155564 | 0.003136255 |
| SSH3 | 0.031968737 | -0.026905071 | 0.005063666 |
| PPP2R2A | 0.045723069 | -0.043225324 | 0.002497746 |
| CEP290 | 0.03782619 | -0.028426027 | 0.009400163 |
| PFN2 | 0.07572426 | -0.031491354 | 0.044232906 |
| TNFRSF12A | 0.02813382 | -0.023063392 | 0.005070429 |
| ENAH | 0.025249919 | -0.019207396 | 0.006042523 |
| GRPEL1 | 0.051291933 | -0.038267641 | 0.013024291 |
| YIPF6 | 0.054841794 | -0.029272201 | 0.025569593 |
| GMCL1 | 0.046038451 | -0.021534662 | 0.024503789 |
| ESYT2 | 0.040591383 | -0.035014595 | 0.005576788 |
| IP6K2 | 0.049568886 | -0.024195027 | 0.025373859 |
| DMAP1 | 0.043381705 | -0.016346945 | 0.02703476 |
| ELMOD1 | 0.019692572 | -0.015906291 | 0.003786282 |
| ELP2 | 0.041185451 | -0.022468387 | 0.018717064 |
| CRYZ | 0.060052111 | -0.051674283 | 0.008377829 |
| TMEM11 | 0.044647652 | -0.018996795 | 0.025650857 |
| RPS25 | 0.076401014 | -0.034724513 | 0.041676501 |
| UBR2 | 0.045366071 | -0.043314084 | 0.002051986 |
| TIFA | 0.026606102 | -0.01764166 | 0.008964441 |
| DNM1L | 0.052253766 | -0.044577123 | 0.007676643 |
| MST1 | 0.03663307 | -0.042294581 | -0.00566151 |
| GHDC | 0.029613901 | -0.019061108 | 0.010552794 |
| PRPSAP1 | 0.045543068 | -0.032803791 | 0.012739277 |
| PLS1 | 0.034287007 | -0.031440175 | 0.002846832 |
| USP40 | 0.037699007 | -0.038442286 | -0.000743278 |
| SGCE | 0.026545664 | -0.020366128 | 0.006179536 |
| RAB24 | 0.022429863 | -0.009523338 | 0.012906526 |
| TMEM63A | 0.031199419 | -0.036133587 | -0.004934168 |
| YES1 | 0.043279538 | -0.034153992 | 0.009125547 |
| IFNGR1 | 0.04731325 | -0.01848613 | 0.02882712 |
| CCDC25 | 0.050314484 | -0.038432686 | 0.011881798 |
| CHRNA3 | 0.010866001 | -0.010866001 | -3.55271E-15 |
| SPATS2L | 0.036798924 | -0.039708129 | -0.002909204 |
| AAMP | 0.052598455 | -0.055272582 | -0.002674127 |
| CDK5RAP2 | 0.035583995 | -0.033082803 | 0.002501192 |
| UBE2Z | 0.04512132 | -0.017699798 | 0.027421523 |
| MINPP1 | 0.029754553 | -0.01645663 | 0.013297923 |
| USP7 | 0.051549064 | -0.053388239 | -0.001839174 |
| KANSL1L | 0.046105291 | -0.038797396 | 0.007307894 |
| ITFG2 | 0.034174149 | -0.0294959 | 0.004678249 |

| | | | |
|---|---|---|---|
| SERGEF | 0.044874119 | -0.019213491 | 0.025660628 |
| PPP2R2D | 0.038999867 | -0.034241699 | 0.004758168 |
| MED13 | 0.048158003 | -0.023387771 | 0.024770231 |
| LRRC19 | 0.014592468 | -0.011299869 | 0.0032926 |
| MARCKSL1 | 0.08099778 | 0.067946618 | 0.148944398 |
| PSAP | 0.07586577 | -0.035410893 | 0.040454877 |
| KDM5A | 0.049716251 | -0.036753789 | 0.012962462 |
| PAWR | 0.044184513 | -0.018408891 | 0.025775622 |
| GNPDA1 | 0.031526539 | -0.015246282 | 0.016280257 |
| ITPK1 | 0.043153544 | -0.022171665 | 0.020981879 |
| ZDHHC21 | 0.040570119 | -0.039644965 | 0.000925154 |
| MALSU1 | 0.057002296 | -0.031672901 | 0.025329395 |
| SIRT6 | 0.027205853 | -0.011210491 | 0.015995362 |
| CISD3 | 0.054931546 | -0.047895346 | 0.0070362 |
| RASGRF2 | 0.021896198 | -0.012293681 | 0.009602517 |
| KDM4A | 0.026667532 | -0.010608008 | 0.016059524 |
| APPL1 | 0.051416302 | -0.057795909 | -0.006379607 |
| ACACB | 0.037027966 | -0.028809852 | 0.008218113 |
| BAZ1B | 0.040204925 | -0.034368868 | 0.005836057 |
| PPP4C | 0.060511525 | -0.030286196 | 0.030225329 |
| GTF2IRD2B | 0.029777564 | -0.020953092 | 0.008824472 |
| LTBP3 | 0.052023556 | -0.022666503 | 0.029357053 |
| TMTC3 | 0.034621163 | -0.031034134 | 0.003587029 |
| OPA1 | 0.046485945 | -0.024272011 | 0.022213934 |
| UCKL1 | 0.040960835 | -0.025745769 | 0.015215066 |
| CHURC1 | 0.067382036 | -0.051948723 | 0.015433313 |
| SETDB2 | 0.040971455 | -0.043320176 | -0.002348721 |
| CPNE3 | 0.05939855 | -0.029789587 | 0.029608963 |
| ABHD14B | 0.059664599 | -0.040370166 | 0.019294434 |
| NAGLU | 0.035660588 | -0.02755367 | 0.008106918 |
| SNIP1 | 0.02339006 | -0.012768995 | 0.010621065 |
| IPO5 | 0.050207125 | -0.023692578 | 0.026514547 |
| PHF20 | 0.039440989 | -0.023672966 | 0.015768023 |
| OFD1 | 0.050114778 | -0.039484044 | 0.010630734 |
| CYB561D2 | 0.046073036 | -0.030382397 | 0.01569064 |
| PAPOLA | 0.067400935 | -0.035632783 | 0.031768152 |
| CBX5 | 0.062455931 | -0.05135051 | 0.011105421 |
| DCUN1D1 | 0.040563609 | -0.025940874 | 0.014622735 |
| PPIL2 | 0.044332352 | -0.024954369 | 0.019377983 |
| C8orf88 | 0.010541855 | -0.010325333 | 0.000216522 |
| COQ9 | 0.062283995 | -0.031110954 | 0.031173041 |
| MHENCR | 0.036560713 | -0.039655115 | -0.003094402 |
| CAMSAP3 | 0.024721517 | -0.01813775 | 0.006583767 |
| ATRN | 0.035636471 | -0.030010825 | 0.005625647 |
| ICE1 | 0.043969039 | -0.026493146 | 0.017475893 |
| LMTK2 | 0.030585409 | -0.019181808 | 0.0114036 |
| KDM5D | 0.020171824 | -0.020154388 | 1.74359E-05 |

| | | | |
|---|---|---|---|
| DNMBP | 0.036923959 | -0.018552609 | 0.01837135 |
| TMEM50B | 0.062605819 | -0.05123255 | 0.01137327 |
| BANK1 | 0.016111592 | -0.01174247 | 0.004369122 |
| BRD7 | 0.05393333 | -0.034894154 | 0.019039175 |
| ANTKMT | 0.065214092 | -0.032921425 | 0.032292667 |
| ZNF486 | 0.01852045 | -0.017307094 | 0.001213356 |
| CPNE1 | 0.050913435 | -0.036970298 | 0.013943137 |
| AMPD2 | 0.02294908 | -0.021919318 | 0.001029762 |
| XPC | 0.034139562 | -0.024923504 | 0.009216058 |
| ALDH7A1 | 0.045181614 | -0.037008967 | 0.008172647 |
| MPPED2 | 0.055591016 | -0.061302374 | -0.005711357 |
| RRAGA | 0.052180271 | -0.028236358 | 0.023943913 |
| COPS5 | 0.056034657 | -0.044891275 | 0.011143381 |
| AGTR1 | 0.014285862 | -0.012749967 | 0.001535895 |
| PIGP | 0.063883296 | -0.024059996 | 0.039823299 |
| ANKRD44 | 0.027809864 | -0.02336637 | 0.004443494 |
| AK3 | 0.061566569 | -0.033648465 | 0.027918104 |
| IQSEC2 | 0.027159458 | -0.016528465 | 0.010630994 |
| MPZL1 | 0.043554621 | -0.037003331 | 0.00655129 |
| TBCEL | 0.023380693 | -0.024253229 | -0.000872536 |
| ATP1B3 | 0.047875734 | -0.058560618 | -0.010684883 |
| SENP7 | 0.03274837 | -0.022477461 | 0.010270909 |
| RPP38 | 0.033032685 | -0.021613611 | 0.011419074 |
| NFE2L2 | 0.072086819 | -0.037040386 | 0.035046432 |
| PSMB9 | 0.035987717 | -0.023838416 | 0.012149301 |
| PMPCA | 0.046751889 | -0.02402553 | 0.02272636 |
| SCX | 0.017150072 | -0.011056143 | 0.006093929 |
| PANK2 | 0.032508407 | -0.031267844 | 0.001240563 |
| SLC22A15 | 0.024434579 | -0.015062286 | 0.009372293 |
| AHNAK | 0.063891573 | -0.051389722 | 0.012501851 |
| ABCC1 | 0.020251882 | -0.016187504 | 0.004064377 |
| DALRD3 | 0.041416238 | -0.033638766 | 0.007777472 |
| DMTN | 0.047892215 | -0.041149082 | 0.006743132 |
| APEX1 | 0.05726568 | -0.024179446 | 0.033086234 |
| TOP2B | 0.05128575 | -0.028176229 | 0.023109521 |
| NIPA2 | 0.044692016 | -0.027494455 | 0.017197561 |
| RPL41 | 0.071060424 | -0.039395255 | 0.03166517 |
| SRPK2 | 0.035825112 | -0.024273976 | 0.011551137 |
| FAM126B | 0.034274371 | -0.023186052 | 0.011088319 |
| CCDC59 | 0.046532062 | -0.039088157 | 0.007443905 |
| ARCN1 | 0.045077799 | -0.027288904 | 0.017788895 |
| PSMD13 | 0.04792722 | -0.028552372 | 0.019374848 |
| TPCN1 | 0.030759105 | -0.035867171 | -0.005108066 |
| SPAG16 | 0.04581923 | -0.037824789 | 0.007994441 |
| SERINC5 | 0.037128423 | -0.021803002 | 0.01532542 |
| AKAP11 | 0.040888141 | -0.044816796 | -0.003928655 |
| TPT1-AS1 | 0.03553277 | -0.034018543 | 0.001514227 |

| | | | |
|---|---|---|---|
| ABCB7 | 0.034595596 | -0.019126045 | 0.015469551 |
| MYH9 | 0.05449472 | -0.031207063 | 0.023287657 |
| TCAP | 0.018053174 | -0.012149082 | 0.005904092 |
| POLR2H | 0.0543274 | -0.026067201 | 0.028260199 |
| CERS2 | 0.055896669 | -0.049089171 | 0.006807498 |
| ZMYM4 | 0.044287908 | -0.029712392 | 0.014575516 |
| CREBZF | 0.038583662 | -0.017981723 | 0.020601939 |
| TMEM38B | 0.039829781 | -0.0154878 | 0.024341981 |
| ITGAE | 0.04164547 | -0.021941186 | 0.019704283 |
| TMEM131 | 0.038778097 | -0.034430454 | 0.004347644 |
| CKLF | 0.032040531 | -0.034849118 | -0.002808588 |
| PPIP5K2 | 0.040955511 | -0.031800147 | 0.009155364 |
| RALGAPA2 | 0.039363368 | -0.032951208 | 0.00641216 |
| AC136475.3 | 0.009996747 | -0.009996747 | -5.32907E-15 |
| SLC9A6 | 0.019443472 | -0.01325454 | 0.006188931 |
| PKHD1 | 0.030264024 | -0.053359287 | -0.023095262 |
| ZNF322 | 0.038199226 | -0.040451388 | -0.002252162 |
| SPTSSA | 0.042203771 | -0.025957311 | 0.01624646 |
| GNS | 0.051621176 | -0.055413686 | -0.003792511 |
| IL10RB | 0.03368792 | -0.035262431 | -0.001574511 |
| MYO9A | 0.045439757 | -0.023375912 | 0.022063846 |
| TMEM115 | 0.041043431 | -0.0275978 | 0.013445631 |
| MAP3K21 | 0.033655236 | -0.024782543 | 0.008872693 |
| ITGB6 | 0.029433509 | -0.027317304 | 0.002116205 |
| WAC | 0.053621753 | -0.038080199 | 0.015541554 |
| SETBP1 | 0.039424586 | -0.023866908 | 0.015557678 |
| GPS2 | 0.056319531 | -0.024072905 | 0.032246625 |
| IGFBP5 | 0.106986031 | -0.106678815 | 0.000307215 |
| DOCK7 | 0.039993168 | -0.04034222 | -0.000349053 |
| TXNDC15 | 0.039044303 | -0.021367855 | 0.017676448 |
| NFYC | 0.042136768 | -0.026714906 | 0.015421862 |
| BBX | 0.059782253 | -0.029299202 | 0.030483051 |
| TMED7 | 0.05660868 | -0.025201515 | 0.031407165 |
| RMDN1 | 0.043898977 | -0.037647622 | 0.006251354 |
| CYP20A1 | 0.041026774 | -0.038087152 | 0.002939621 |
| SLC31A1 | 0.033882863 | -0.020346852 | 0.013536011 |
| TMED3 | 0.047330993 | -0.042022475 | 0.005308518 |
| CDC25B | 0.019421082 | -0.014913023 | 0.004508059 |
| FOXA3 | 0.012488006 | -0.01199798 | 0.000490025 |
| RSRC2 | 0.059312314 | -0.044332427 | 0.014979887 |
| YIPF2 | 0.042626474 | -0.022205398 | 0.020421076 |
| RPS24 | 0.070940638 | -0.030935358 | 0.040005279 |
| HCFC1R1 | 0.042419195 | -0.032416814 | 0.010002381 |
| PDK2 | 0.040253569 | -0.037700079 | 0.00255349 |
| SPPL3 | 0.045805835 | -0.036351525 | 0.00945431 |
| RAD21 | 0.05084468 | -0.03927012 | 0.01157456 |
| GFM1 | 0.044434355 | -0.029080179 | 0.015354176 |

| | | | |
|---|---|---|---|
| THRB | 0.039116899 | -0.018254294 | 0.020862605 |
| MRPL43 | 0.062750946 | -0.025690162 | 0.037060784 |
| MYO10 | 0.055300656 | -0.041225444 | 0.014075212 |
| AC006230.1 | 0.01692823 | -0.012058383 | 0.004869847 |
| DIP2A | 0.036048232 | -0.032932024 | 0.003116208 |
| DDX24 | 0.06578567 | -0.05709835 | 0.00868732 |
| KIF16B | 0.037775568 | -0.029016017 | 0.008759551 |
| DPM2 | 0.054493224 | -0.029890897 | 0.024602327 |
| SOX5 | 0.012659054 | -0.011969001 | 0.000690053 |
| SDCCAG8 | 0.046004845 | -0.055358348 | -0.009353504 |
| TRIM33 | 0.043236617 | -0.03360523 | 0.009631388 |
| KIAA1217 | 0.047898706 | -0.052298608 | -0.004399903 |
| APPL2 | 0.031563206 | -0.023874153 | 0.007689053 |
| PRSS22 | 0.023510923 | -0.023448012 | 6.29118E-05 |
| DCBLD1 | 0.018587706 | -0.011982263 | 0.006605444 |
| ARSA | 0.029148026 | -0.014030941 | 0.015117085 |
| MEGF9 | 0.048672488 | -0.034807062 | 0.013865426 |
| PAXX | 0.05893926 | -0.039389095 | 0.019550165 |
| TBC1D2 | 0.042281315 | -0.0198121 | 0.022469215 |
| BTBD6 | 0.03916337 | -0.033079552 | 0.006083818 |
| CTU1 | 0.03013703 | -0.012112261 | 0.018024768 |
| SENP6 | 0.054961649 | -0.07842754 | -0.02346589 |
| PIH1D1 | 0.047611518 | -0.042112569 | 0.005498949 |
| PARP14 | 0.042412369 | -0.045114098 | -0.002701729 |
| DHRSX | 0.035027559 | -0.020329545 | 0.014698014 |
| FAM133B | 0.047950856 | -0.049354698 | -0.001403841 |
| APOE | -0.134927879 | 0.288789932 | 0.153862052 |
| MCU | 0.030765862 | -0.019285999 | 0.011479863 |
| AC008443.1 | 0.024137246 | -0.016377761 | 0.007759485 |
| MRGBP | 0.02982247 | -0.018287299 | 0.011535171 |
| SMC6 | 0.028085049 | -0.015584434 | 0.012500615 |
| DARS | 0.053321775 | -0.042656666 | 0.010665109 |
| ST3GAL6 | 0.046124775 | -0.029584064 | 0.016540711 |
| FN3KRP | 0.031189375 | -0.027865715 | 0.00332366 |
| EP300 | 0.042438205 | -0.035657511 | 0.006780694 |
| MRPL27 | 0.063674973 | -0.032617053 | 0.03105792 |
| ARL15 | 0.037413313 | -0.026793117 | 0.010620195 |
| ZFX | 0.038935454 | -0.016004997 | 0.022930457 |
| BIRC6 | 0.050824269 | -0.047415033 | 0.003409236 |
| FAM234A | 0.046181815 | -0.025346967 | 0.020834848 |
| LMBRD1 | 0.058629374 | -0.040912089 | 0.017717286 |
| PLCG1 | 0.016462789 | -0.011869804 | 0.004592984 |
| PHYKPL | 0.036886255 | -0.03256179 | 0.004324465 |
| EMC2 | 0.053608062 | -0.035360764 | 0.018247298 |
| NT5C3B | 0.046696332 | -0.018830068 | 0.027866264 |
| PPL | 0.022164152 | -0.024834558 | -0.002670406 |
| MGAM | 0.014667434 | -0.015009911 | -0.000342478 |

| | | | |
|---|---|---|---|
| EXTL2 | 0.043477938 | -0.020137801 | 0.023340138 |
| CYSTM1 | 0.074431842 | -0.032582087 | 0.041849755 |
| ATOX1 | 0.075701903 | -0.037117004 | 0.038584899 |
| GTF2F2 | 0.041592205 | -0.023296317 | 0.018295888 |
| CKS1B | 0.045717342 | -0.01913623 | 0.026581112 |
| RANBP9 | 0.034505786 | -0.023958084 | 0.010547702 |
| NRBP1 | 0.038730105 | -0.026693013 | 0.012037092 |
| AC005821.1 | 0.013344059 | -0.011489927 | 0.001854133 |
| AP3B1 | 0.048424887 | -0.035980853 | 0.012444033 |
| SMC1A | 0.044080083 | -0.024681749 | 0.019398334 |
| FBH1 | 0.03530145 | -0.024799311 | 0.010502139 |
| MTAP | 0.030827828 | -0.016039665 | 0.014788163 |
| GLYR1 | 0.051408849 | -0.033678521 | 0.017730329 |
| TARSL2 | 0.031813881 | -0.020055408 | 0.011758473 |
| COA6 | 0.053997443 | -0.035782286 | 0.018215156 |
| UNC13C | 0.017665206 | -0.019876871 | -0.002211665 |
| GPX2 | 0.013424468 | -0.011128574 | 0.002295894 |
| TBC1D14 | 0.066190338 | -0.060236066 | 0.005954272 |
| RBM5 | 0.04833947 | -0.041788147 | 0.006551323 |
| ODR4 | 0.039164827 | -0.038158315 | 0.001006513 |
| MPHOSPH10 | 0.037921991 | -0.025464699 | 0.012457292 |
| KDM4B | 0.031547454 | -0.023200069 | 0.008347385 |
| CDADC1 | 0.031976288 | -0.033120548 | -0.00114426 |
| THUMPD3 | 0.037044952 | -0.025824197 | 0.011220755 |
| SMTNL2 | 0.026273696 | -0.032980132 | -0.006706435 |
| CCDC51 | 0.02485712 | -0.010518719 | 0.014338402 |
| CC2D1A | 0.026886678 | -0.012358997 | 0.014527681 |
| PSMD6 | 0.05032838 | -0.025252491 | 0.025075889 |
| RUFY1 | 0.035969584 | -0.031666113 | 0.004303471 |
| INTS10 | 0.041565422 | -0.027492834 | 0.014072588 |
| BICDL2 | 0.025725388 | -0.024238252 | 0.001487137 |
| LGMN | 0.048565747 | -0.035074372 | 0.013491375 |
| SLC16A10 | 0.033646526 | -0.029757836 | 0.003888689 |
| ARHGEF35 | 0.032879404 | -0.016432844 | 0.01644656 |
| EGLN2 | 0.03977159 | -0.021461344 | 0.018310246 |
| MIPOL1 | 0.022819335 | -0.012845262 | 0.009974074 |
| UROS | 0.045058926 | -0.022439481 | 0.022619444 |
| MUL1 | 0.036205165 | -0.025227286 | 0.010977879 |
| ANAPC5 | 0.055140967 | -0.039842369 | 0.015298597 |
| SMARCC2 | 0.044699664 | -0.028694458 | 0.016005206 |
| HSPB1 | 0.072479933 | -0.061202338 | 0.011277594 |
| RBMX | 0.058855698 | -0.055616477 | 0.003239221 |
| MAP3K1 | 0.032131983 | -0.031817798 | 0.000314185 |
| BSDC1 | 0.036644371 | -0.03214929 | 0.00449508 |
| QSOX1 | 0.045631822 | -0.048560864 | -0.002929042 |
| PPP1R3C | 0.029347018 | -0.018957346 | 0.010389672 |
| PHKB | 0.051054987 | -0.036343478 | 0.014711509 |

| | | | |
|---|---|---|---|
| AC099670.3 | 0.010613146 | -0.00930986 | 0.001303286 |
| ZHX3 | 0.03303536 | -0.036960337 | -0.003924977 |
| DNAJC4 | 0.060220595 | -0.025742909 | 0.034477686 |
| CCNA1 | 0.009685026 | -0.009449955 | 0.000235071 |
| ACTR3 | 0.058608588 | -0.028116332 | 0.030492256 |
| CCDC93 | 0.033501285 | -0.025971492 | 0.007529793 |
| MXD1 | 0.02221829 | -0.011086093 | 0.011132197 |
| PRKAR2A | 0.042311421 | -0.033143575 | 0.009167846 |
| RILP | 0.041930409 | -0.024908973 | 0.017021436 |
| LCA5 | 0.016695211 | -0.011182249 | 0.005512961 |
| TLK1 | 0.047706389 | -0.048003995 | -0.000297607 |
| CDK9 | 0.035364189 | -0.021083693 | 0.014280497 |
| POLD4 | 0.06429506 | -0.052964858 | 0.011330202 |
| VSIG10 | 0.029317439 | -0.03081679 | -0.001499351 |
| DNAJA4 | 0.030105207 | -0.013587173 | 0.016518033 |
| FAM89B | 0.035712283 | -0.037534276 | -0.001821993 |
| MAN2A1 | 0.053534772 | -0.039008729 | 0.014526043 |
| ITM2C | 0.060121363 | -0.074416629 | -0.014295266 |
| GOLGA2 | 0.044558366 | -0.028340889 | 0.016217477 |
| ETFB | 0.071881122 | -0.027568866 | 0.044312256 |
| UBR5 | 0.039092933 | -0.036206458 | 0.002886475 |
| SCO2 | 0.035179848 | -0.028987827 | 0.006192021 |
| ECPAS | 0.041467505 | -0.036336262 | 0.005131243 |
| SLC44A3 | 0.039232906 | -0.024903761 | 0.014329145 |
| LAMC1 | 0.036117354 | -0.024993251 | 0.011124102 |
| LRRC59 | 0.037449651 | -0.0174986 | 0.019951051 |
| PIR | 0.016472976 | -0.01393259 | 0.002540386 |
| MITF | 0.046409682 | -0.032779052 | 0.01363063 |
| SIGIRR | 0.05532049 | -0.045403904 | 0.009916586 |
| MICALL1 | 0.020354026 | -0.012505818 | 0.007848208 |
| SAMD9 | 0.021956133 | -0.013840951 | 0.008115182 |
| MFN1 | 0.037321458 | -0.048168034 | -0.010846577 |
| ADNP | 0.047829401 | -0.019568755 | 0.028260646 |
| ZNF780A | 0.026882701 | -0.011801002 | 0.0150817 |
| AL139280.1 | 0.010268246 | -0.009079035 | 0.001189211 |
| HOXB-AS1 | 0.020477573 | -0.012638426 | 0.007839147 |
| SBNO1 | 0.044044806 | -0.039584348 | 0.004460459 |
| ATXN7L3 | 0.02445221 | -0.013973166 | 0.010479044 |
| CD40 | 0.02122842 | -0.023963503 | -0.002735083 |
| HSPG2 | 0.033661846 | -0.035907294 | -0.002245448 |
| EIF4B | 0.069114875 | -0.045332108 | 0.023782767 |
| SNHG12 | 0.029296454 | -0.021235641 | 0.008060813 |
| JMJD6 | 0.030581997 | -0.016179841 | 0.014402156 |
| ARIH2 | 0.040560232 | -0.033420354 | 0.007139878 |
| DNM2 | 0.043251845 | -0.037656255 | 0.00559559 |
| FGGY | 0.024001857 | -0.019236535 | 0.004765321 |
| SEC61A1 | 0.045826463 | -0.025740028 | 0.020086435 |

| | | | |
|---|---|---|---|
| RAB8A | 0.046077637 | -0.036536275 | 0.009541361 |
| HIST1H1E | 0.025056413 | -0.022175872 | 0.002880541 |
| USP24 | 0.034379049 | -0.026061572 | 0.008317477 |
| LONP2 | 0.045563024 | -0.034940223 | 0.010622801 |
| SRPRA | 0.047747844 | -0.040721466 | 0.007026377 |
| MFSD4A | -0.036957972 | 0.034371821 | -0.002586151 |
| HOTAIRM1 | -0.024816611 | 0.010153194 | -0.014663417 |
| ZDHHC20 | 0.025446668 | -0.016644927 | 0.008801741 |
| TTLL3 | 0.025365208 | -0.025891449 | -0.000526241 |
| NCKAP1 | 0.05132954 | -0.047969637 | 0.003359903 |
| SLC29A4 | 0.035754714 | -0.047105283 | -0.011350568 |
| FAM3B | 0.062344811 | -0.05015716 | 0.012187651 |
| ZC3HAV1 | 0.034329278 | -0.030262369 | 0.004066909 |
| PGAP1 | 0.020840097 | -0.022470946 | -0.001630849 |
| HECTD4 | 0.031140715 | -0.03497953 | -0.003838815 |
| PPIL6 | 0.021387902 | -0.015853612 | 0.005534289 |
| TAF15 | 0.040577139 | -0.023880927 | 0.016696212 |
| CYS1 | -0.048104223 | 0.01989956 | -0.028204663 |
| GCLC | 0.022115471 | -0.015477868 | 0.006637603 |
| TMEM168 | 0.039521475 | -0.033684911 | 0.005836564 |
| AHCYL1 | 0.057985498 | -0.034384296 | 0.023601202 |
| LINC00930 | 0.009137794 | -0.009137794 | -3.55271E-15 |
| NUCB1 | 0.055001716 | -0.04490352 | 0.010098197 |
| ELP6 | 0.039717269 | -0.0252897 | 0.014427569 |
| RGS19 | 0.015766388 | -0.010124109 | 0.005642279 |
| BAX | 0.045743449 | -0.024681943 | 0.021061507 |
| ELMOD3 | 0.024371255 | -0.017223539 | 0.007147717 |
| ARFIP2 | 0.037641842 | -0.030253579 | 0.007388262 |
| VPS53 | 0.033068242 | -0.022032967 | 0.011035275 |
| ARID4A | 0.034710505 | -0.035485889 | -0.000775384 |
| COPA | 0.050487328 | -0.039778908 | 0.01070842 |
| HSPA2 | 0.042331786 | -0.065781079 | -0.023449293 |
| DVL3 | 0.034073006 | -0.031762126 | 0.002310881 |
| TOR2A | 0.030028068 | -0.026201223 | 0.003826845 |
| DHX15 | 0.042898735 | -0.028720763 | 0.014177972 |
| PEAK1 | 0.017214502 | -0.015989463 | 0.001225038 |
| SSH2 | 0.033446545 | -0.032734977 | 0.000711568 |
| TMEM160 | 0.06086635 | -0.046601542 | 0.014264807 |
| MMP7 | 0.015695674 | -0.013496725 | 0.00219895 |
| LCMT1 | 0.037748438 | -0.014992711 | 0.022755727 |
| APH1B | 0.03284898 | -0.028015028 | 0.004833952 |
| DCDC2 | 0.031507209 | -0.023543565 | 0.007963644 |
| KCNJ15 | 0.034037409 | -0.04882451 | -0.014787101 |
| HIST1H2AC | 0.03086174 | -0.034400613 | -0.003538872 |
| CDK11A | 0.026229526 | -0.017046797 | 0.009182729 |
| EDEM3 | 0.036464911 | -0.037851539 | -0.001386628 |
| DSC2 | 0.038933678 | -0.019126425 | 0.019807253 |

| | | | |
|---|---|---|---|
| MPG | 0.045316571 | -0.026425653 | 0.018890917 |
| SMARCD1 | 0.035631379 | -0.014293157 | 0.021338222 |
| UTRN | 0.032985468 | -0.035119754 | -0.002134287 |
| NCBP3 | 0.040507249 | -0.040428578 | 7.86708E-05 |
| CRCP | 0.03300814 | -0.013787354 | 0.019220786 |
| EIF4H | 0.056185559 | -0.026556104 | 0.029629455 |
| TNS3 | 0.045726149 | -0.045244302 | 0.000481848 |
| DDX59 | 0.029186072 | -0.013680264 | 0.015505808 |
| HUWE1 | 0.049555454 | -0.043124394 | 0.006431059 |
| CCNL1 | 0.045629616 | -0.034149275 | 0.011480341 |
| ALG6 | 0.022780172 | -0.013226672 | 0.0095535 |
| GMPPB | 0.025745683 | -0.011957797 | 0.013787886 |
| ABCC4 | 0.059308023 | -0.030275498 | 0.029032525 |
| SNRNP40 | 0.043750869 | -0.022523098 | 0.021227772 |
| WDR34 | 0.042419701 | -0.036187968 | 0.006231733 |
| YTHDC1 | 0.047202763 | -0.043089338 | 0.004113425 |
| PPIL4 | 0.044410807 | -0.025007314 | 0.019403492 |
| UNC119 | 0.037748902 | -0.020003567 | 0.017745334 |
| PLA2G15 | 0.036313672 | -0.033085576 | 0.003228096 |
| BAG6 | 0.044810324 | -0.046553283 | -0.001742958 |
| C19orf12 | 0.029121697 | -0.02143833 | 0.007683367 |
| ALDH16A1 | 0.0268718 | -0.014495802 | 0.012375998 |
| USP8 | 0.046952828 | -0.033405358 | 0.01354747 |
| EPHX3 | 0.016434497 | -0.01359518 | 0.002839317 |
| PITPNB | 0.038899268 | -0.019412277 | 0.019486991 |
| SIPA1L3 | 0.033525199 | -0.016870819 | 0.01665438 |
| PRDM2 | 0.03760985 | -0.037218247 | 0.000391603 |
| C10orf143 | 0.024659054 | -0.014591805 | 0.010067249 |
| CNOT4 | 0.0409748 | -0.033779539 | 0.00719526 |
| PSMD7 | 0.045191347 | -0.032286116 | 0.012905231 |
| MSL3 | 0.026166732 | -0.018847896 | 0.007318836 |
| MAN2C1 | 0.033829622 | -0.019533344 | 0.014296279 |
| GOLPH3 | 0.040003646 | -0.041062506 | -0.001058861 |
| ZNF787 | 0.02744321 | -0.01599925 | 0.01144396 |
| KPNA1 | 0.03392178 | -0.028322357 | 0.005599423 |
| ITPKB | 0.026354988 | -0.026267587 | 8.74015E-05 |
| TCP11L2 | 0.018589025 | -0.013887137 | 0.004701887 |
| DOCK6 | 0.023163369 | -0.010855574 | 0.012307795 |
| EXT2 | 0.029252799 | -0.017963231 | 0.011289568 |
| SSX2IP | 0.029194469 | -0.023685776 | 0.005508693 |
| XPR1 | 0.031913577 | -0.023642072 | 0.008271505 |
| MUC15 | 0.044661308 | -0.081238843 | -0.036577536 |
| STXBP5 | 0.017715916 | -0.016230129 | 0.001485787 |
| C5orf56 | 0.027510602 | -0.026069366 | 0.001441236 |
| BDH2 | 0.058074187 | -0.028391752 | 0.029682435 |
| BLCAP | 0.046310787 | -0.023608049 | 0.022702738 |
| NOP2 | 0.019886794 | -0.011757754 | 0.00812904 |

| | | | |
|---|---|---|---|
| SLC44A1 | 0.044372638 | -0.055212544 | -0.010839907 |
| CWC25 | 0.032356712 | -0.025935336 | 0.006421376 |
| KIAA0232 | 0.046983352 | -0.022133596 | 0.024849756 |
| SURF4 | 0.045650295 | -0.039641957 | 0.006008338 |
| HSF2 | 0.022884643 | -0.014234524 | 0.008650119 |
| NR1D1 | 0.01595741 | -0.012528388 | 0.003429022 |
| KIZ | 0.024950483 | -0.023946001 | 0.001004483 |
| FAM245A | 0.010168075 | -0.009075801 | 0.001092274 |
| DUS1L | 0.044878739 | -0.025313017 | 0.019565722 |
| KBTBD11 | -0.01432519 | 0.006830284 | -0.007494906 |
| TXNDC11 | 0.034472004 | -0.034638168 | -0.000166164 |
| U91319.1 | 0.011314741 | -0.013147927 | -0.001833186 |
| RCOR1 | 0.028951518 | -0.02184764 | 0.007103878 |
| PWP1 | 0.041253527 | -0.037924963 | 0.003328563 |
| TOP1 | 0.052607863 | -0.043221016 | 0.009386847 |
| ARPC4 | 0.052851817 | -0.033192236 | 0.01965958 |
| AASDHPPT | 0.041978523 | -0.019124552 | 0.022853971 |
| TFAP2C | 0.009728024 | -0.010606515 | -0.000878492 |
| LMBRD2 | 0.03185066 | -0.026273812 | 0.005576847 |
| CYB5R1 | 0.045055359 | -0.035465866 | 0.009589493 |
| ATL2 | 0.050283968 | -0.029001414 | 0.021282554 |
| DCAF12L1 | 0.009793215 | -0.009058876 | 0.000734339 |
| QRSL1 | 0.032331865 | -0.025808392 | 0.006523474 |
| PHLDB2 | 0.049271261 | -0.029963905 | 0.019307355 |
| CDYL | 0.031995696 | -0.01764234 | 0.014353356 |
| ACSM3 | 0.035006635 | -0.025294201 | 0.009712434 |
| ZMYND11 | 0.049188322 | -0.037499951 | 0.011688371 |
| TNS4 | 0.018206836 | -0.015691331 | 0.002515505 |
| TBK1 | 0.032553326 | -0.034778457 | -0.002225131 |
| ZNF638 | 0.051819919 | -0.04588046 | 0.005939459 |
| ZBTB1 | 0.035495778 | -0.036738565 | -0.001242787 |
| GJB1 | 0.017045322 | -0.019603831 | -0.002558509 |
| GIPR | 0.01277737 | -0.012582485 | 0.000194885 |
| ANKRD13D | 0.027054171 | -0.023838072 | 0.003216099 |
| TOR3A | 0.034112261 | -0.023658014 | 0.010454247 |
| FBXO34 | 0.038783886 | -0.023063083 | 0.015720802 |
| SLC35E2B | 0.02320102 | -0.014192372 | 0.009008648 |
| BMPR2 | 0.037145524 | -0.031977596 | 0.005167928 |
| CBLC | 0.01661801 | -0.013616654 | 0.003001356 |
| USP53 | 0.04066739 | -0.027278407 | 0.013388983 |
| PFKL | 0.052947501 | -0.047845101 | 0.0051024 |
| VPS26A | 0.044208252 | -0.028477348 | 0.015730904 |
| MVB12B | 0.030576063 | -0.041447934 | -0.010871871 |
| COPZ2 | 0.022075166 | -0.011886095 | 0.01018907 |
| LYST | 0.034080847 | -0.014437019 | 0.019643828 |
| LYRM9 | 0.035510053 | -0.020843612 | 0.014666441 |
| CUEDC1 | 0.036155089 | -0.022155737 | 0.013999352 |

| | | | |
|---|---|---|---|
| CDC37L1 | 0.033990127 | -0.014807432 | 0.019182694 |
| GPR108 | 0.041941209 | -0.037320558 | 0.004620651 |
| AMN | 0.039123812 | -0.054162594 | -0.015038781 |
| SNX7 | 0.037369438 | -0.04167318 | -0.004303742 |
| PLEKHF1 | 0.018399858 | -0.019256647 | -0.000856789 |
| CDKL1 | 0.026867806 | -0.029476197 | -0.002608391 |
| PLEKHH2 | 0.024273802 | -0.033088605 | -0.008814803 |
| NADK | 0.031902299 | -0.026529636 | 0.005372664 |
| PPP1R10 | 0.042725937 | -0.05132485 | -0.008598913 |
| HCFC2 | 0.025802969 | -0.014587838 | 0.01121513 |
| WWP1 | 0.054772132 | -0.041249957 | 0.013522175 |
| AL157373.2 | 0.021677227 | -0.017128725 | 0.004548502 |
| HARS | 0.033500125 | -0.014176638 | 0.019323487 |
| MAP4K5 | 0.03118559 | -0.025744102 | 0.005441488 |
| TESK1 | 0.026088029 | -0.013363999 | 0.01272403 |
| ERH | 0.069092464 | 0.02954725 | 0.098639714 |
| RHPN2 | 0.027728128 | -0.02985007 | -0.002121942 |
| LINC00923 | 0.013770673 | -0.015883276 | -0.002112603 |
| ABCA3 | 0.01905823 | -0.017599171 | 0.001459059 |
| MLF2 | 0.060797873 | -0.027962169 | 0.032835704 |
| AC097654.1 | 0.026650091 | -0.019581591 | 0.0070685 |
| ZFAND2B | 0.039191823 | -0.026560627 | 0.012631196 |
| CSNK1G2 | 0.036203357 | -0.017987197 | 0.01821616 |
| MLLT10 | 0.030564652 | -0.021422152 | 0.0091425 |
| THUMPD2 | 0.027727977 | -0.029554919 | -0.001826942 |
| VPS13A | 0.043818803 | -0.033730462 | 0.010088341 |
| COL6A2 | -0.025970223 | 0.025339941 | -0.000630282 |
| IFRD1 | 0.030737279 | -0.031462296 | -0.000725018 |
| SNRNP200 | 0.046474678 | -0.028878501 | 0.017596176 |
| TAZ | 0.029344154 | -0.025685188 | 0.003658967 |
| GPATCH11 | 0.023287534 | -0.010000915 | 0.013286619 |
| ACAT2 | 0.031604707 | -0.013959386 | 0.017645321 |
| FAHD1 | 0.03307938 | -0.020506784 | 0.012572596 |
| RHOQ | 0.037235259 | -0.021768178 | 0.015467081 |
| POLM | 0.026123234 | -0.01869191 | 0.007431325 |
| CRACR2B | 0.026717011 | -0.014489353 | 0.012227657 |
| ASMTL | 0.03339629 | -0.018284902 | 0.015111388 |
| ATP6V0D1 | 0.055256562 | -0.026750173 | 0.028506389 |
| EFNA4 | 0.026826349 | -0.015903152 | 0.010923196 |
| TSR3 | 0.047978317 | -0.031564443 | 0.016413874 |
| TPRN | 0.026023146 | -0.023309806 | 0.002713339 |
| SMG5 | 0.030737796 | -0.023721419 | 0.007016378 |
| DYNC1I2 | 0.055417677 | -0.097272242 | -0.041854565 |
| PAX8-AS1 | 0.01725623 | -0.018004888 | -0.000748658 |
| FAM131A | 0.022892892 | -0.013235428 | 0.009657463 |
| CHRAC1 | 0.035523757 | -0.023689487 | 0.01183427 |
| STXBP2 | 0.042538689 | -0.028086722 | 0.014451967 |

| | | | |
|---|---|---|---|
| HIF3A | 0.008941407 | -0.008545034 | 0.000396373 |
| TC2N | 0.026169482 | -0.028749563 | -0.00258008 |
| PTPN18 | 0.046101921 | -0.027429502 | 0.018672419 |
| TAF5 | 0.01158569 | -0.007058737 | 0.004526953 |
| KHDRBS1 | 0.056526716 | -0.031301453 | 0.025225263 |
| NEAT1 | 0.080612819 | -0.060710194 | 0.019902624 |
| CMAS | 0.034861019 | -0.031258025 | 0.003602994 |
| PPHLN1 | 0.035999891 | -0.01678511 | 0.01921478 |
| IQGAP2 | 0.054263847 | -0.047630941 | 0.006632907 |
| GSE1 | 0.0404461 | -0.019770536 | 0.020675564 |
| CNTN3 | 0.017748744 | -0.013981661 | 0.003767083 |
| ADD1 | 0.043254644 | -0.042063129 | 0.001191516 |
| OSER1 | 0.03603072 | -0.025572584 | 0.010458136 |
| RPA3 | 0.05847748 | -0.039579913 | 0.018897567 |
| COG2 | 0.033548604 | -0.02390468 | 0.009643924 |
| UBE3B | 0.033651818 | -0.013697065 | 0.019954754 |
| BSCL2 | 0.047342852 | -0.059272726 | -0.011929874 |
| FAIM | 0.037110584 | -0.032581557 | 0.004529027 |
| OGDH | 0.053697728 | -0.023803256 | 0.029894472 |
| TK2 | 0.027169869 | -0.015118325 | 0.012051544 |
| DLST | 0.038907704 | -0.033627243 | 0.005280461 |
| CDIPT | 0.041220583 | -0.034099805 | 0.007120777 |
| RUVBL1 | 0.03388167 | -0.02842695 | 0.00545472 |
| PPP4R3B | 0.044673586 | -0.021262811 | 0.023410775 |
| RRP1 | 0.028528947 | -0.01739155 | 0.011137397 |
| GTPBP4 | 0.031398173 | -0.01351646 | 0.017881713 |
| TXNRD1 | 0.025228714 | -0.024519262 | 0.000709453 |
| ZFAS1 | 0.067177651 | -0.044465088 | 0.022712563 |
| GANAB | 0.048616682 | -0.035215397 | 0.013401285 |
| MMEL1 | 0.008952974 | -0.008505653 | 0.000447322 |
| ATG14 | 0.024511852 | -0.020059277 | 0.004452574 |
| FAM160B2 | 0.024722566 | -0.017764709 | 0.006957857 |
| THRA | 0.039768533 | -0.033692322 | 0.006076212 |
| PARL | 0.043403848 | -0.023545888 | 0.01985796 |
| CCDC170 | 0.011463969 | -0.013676665 | -0.002212695 |
| LRWD1 | 0.020358811 | -0.015054605 | 0.005304206 |
| AP000569.1 | 0.015865022 | -0.012398184 | 0.003466838 |
| SEC14L1 | 0.046035961 | -0.018968742 | 0.027067219 |
| CYTH2 | 0.034886141 | -0.017336675 | 0.017549466 |
| METTL23 | 0.045949389 | -0.034849249 | 0.011100141 |
| PCP2 | 0.024490254 | -0.028689531 | -0.004199277 |
| FAM117A | 0.015487557 | -0.008552016 | 0.006935541 |
| TENM3 | 0.017858499 | -0.017358001 | 0.000500498 |
| DYRK1B | 0.02331653 | -0.014607482 | 0.008709048 |
| MFSD14A | 0.036037239 | -0.01644038 | 0.01959686 |
| RPRD1A | 0.036869847 | -0.016632113 | 0.020237734 |
| TUT7 | 0.026969771 | -0.022672312 | 0.004297458 |

| | | | |
|---|---|---|---|
| TMEM251 | 0.050643163 | -0.02837515 | 0.022268013 |
| DPY19L2 | 0.045374206 | -0.02726843 | 0.018105777 |
| NDUFV3 | 0.05897541 | -0.040572926 | 0.018402484 |
| ANXA5 | 0.068844534 | -0.063751562 | 0.005092972 |
| SUMF2 | 0.048243058 | -0.040858065 | 0.007384994 |
| SUMF1 | 0.036512397 | -0.026267467 | 0.01024493 |
| ATP11C | 0.021097847 | -0.029353033 | -0.008255186 |
| CLK4 | 0.03806928 | -0.038977255 | -0.000907975 |
| HEIH | 0.043957371 | -0.019642427 | 0.024314943 |
| CTR9 | 0.040300656 | -0.025858882 | 0.014441773 |
| TMEM150A | 0.032608532 | -0.017632856 | 0.014975676 |
| RILPL2 | 0.024467337 | -0.021992692 | 0.002474645 |
| POGZ | 0.043858056 | -0.046547879 | -0.002689823 |
| SHKBP1 | 0.030182546 | -0.01660773 | 0.013574816 |
| CYB5R4 | 0.025732527 | -0.019396208 | 0.006336319 |
| BORCS8 | 0.037352372 | -0.026560081 | 0.01079229 |
| SASH1 | 0.027112138 | -0.03280591 | -0.005693772 |
| TNFRSF1A | 0.03114161 | -0.020920076 | 0.010221534 |
| TDRD7 | 0.017791083 | -0.017920048 | -0.000128966 |
| LRFN4 | 0.022849584 | -0.026225775 | -0.003376191 |
| CHD4 | 0.049108975 | -0.042240833 | 0.006868142 |
| CHKB | 0.042523327 | -0.023870898 | 0.018652429 |
| DAAM1 | 0.042079856 | -0.034936547 | 0.007143309 |
| ING4 | 0.034006335 | -0.019177316 | 0.014829019 |
| C16orf91 | 0.036569718 | -0.018255835 | 0.018313882 |
| RARS | 0.034617012 | -0.020996671 | 0.013620341 |
| GSTO2 | 0.044684515 | -0.029869307 | 0.014815208 |
| ATXN2 | 0.038974366 | -0.031256765 | 0.007717601 |
| RBM48 | 0.024543455 | -0.019565633 | 0.004977822 |
| DGAT1 | 0.032884633 | -0.032002843 | 0.000881791 |
| COPZ1 | 0.054504339 | -0.029994146 | 0.024510193 |
| SMAD3 | 0.026502892 | -0.015478563 | 0.011024328 |
| STX10 | 0.034756098 | -0.032906807 | 0.001849291 |
| SLC49A4 | 0.025679955 | -0.021623444 | 0.00405651 |
| HOOK3 | 0.037344888 | -0.020752647 | 0.01659224 |
| ENTPD6 | 0.034104888 | -0.022603809 | 0.011501079 |
| SPAST | 0.022344319 | -0.018804572 | 0.003539747 |
| KAT8 | 0.033259467 | -0.029214211 | 0.004045256 |
| MT-ND4 | -0.05094264 | 0.028305942 | -0.022636699 |
| MTIF2 | 0.036524247 | -0.032528113 | 0.003996134 |
| SWAP70 | 0.025537535 | -0.022316304 | 0.003221231 |
| EFCAB14 | 0.05167812 | -0.036771656 | 0.014906464 |
| USP28 | 0.030609906 | -0.028314096 | 0.002295811 |
| PRCC | 0.032609295 | -0.025730676 | 0.006878619 |
| STXBP3 | 0.049836054 | -0.042540592 | 0.007295462 |
| ARL13B | 0.023117561 | -0.01198674 | 0.011130821 |
| TCERG1 | 0.031703186 | -0.019830402 | 0.011872784 |

| | | | |
|---|---|---|---|
| ATG2B | 0.025696308 | -0.019051185 | 0.006645123 |
| CALU | 0.041609083 | -0.028241672 | 0.013367411 |
| RIC8A | 0.030087215 | -0.016182362 | 0.013904852 |
| ZNF22 | 0.041311319 | -0.029223107 | 0.012088213 |
| UBE2Q2 | 0.030089757 | -0.023750761 | 0.006338995 |
| UBXN7 | 0.030639212 | -0.017093631 | 0.013545581 |
| FOXD1 | -0.021844316 | 0.02194564 | 0.000101323 |
| CAMKMT | 0.019613107 | -0.009560705 | 0.010052402 |
| RNF5 | 0.048860249 | -0.06508301 | -0.016222761 |
| PRMT2 | 0.045029069 | -0.046263085 | -0.001234016 |
| MANSC1 | 0.023114504 | -0.017849232 | 0.005265272 |
| PBDC1 | 0.045413514 | -0.037347478 | 0.008066035 |
| SLC6A6 | 0.015101174 | -0.014184738 | 0.000916435 |
| SLC66A1L | 0.009697202 | -0.009315215 | 0.000381986 |
| PPP6R3 | 0.044742603 | -0.039058637 | 0.005683966 |
| FAM234B | 0.035663586 | -0.031284745 | 0.004378841 |
| CCSER2 | 0.038955875 | -0.023946791 | 0.015009084 |
| PIGV | 0.024186119 | -0.019802059 | 0.00438406 |
| SPOP | 0.040043635 | -0.039583316 | 0.000460319 |
| TRAPPC1 | 0.060816237 | -0.035701177 | 0.02511506 |
| CASP2 | 0.021921237 | -0.01442596 | 0.007495277 |
| NSF | 0.029913185 | -0.023095145 | 0.00681804 |
| QTRT1 | 0.04770006 | -0.035598198 | 0.012101862 |
| GDE1 | 0.041346794 | -0.021695653 | 0.019651141 |
| CWC27 | 0.028893333 | -0.015013944 | 0.013879389 |
| CACNA2D1 | 0.018098014 | -0.028888039 | -0.010790025 |
| SPP1 | -0.043062018 | 0.040501779 | -0.00256024 |
| TTC17 | 0.038632757 | -0.037348336 | 0.001284422 |
| COA5 | 0.052214699 | -0.027775184 | 0.024439515 |
| THAP3 | 0.028389641 | -0.024330098 | 0.004059543 |
| ELAC2 | 0.03315148 | -0.031535435 | 0.001616045 |
| KIF23 | 0.015407207 | -0.01461461 | 0.000792597 |
| SLC9A4 | 0.019739881 | -0.018491097 | 0.001248784 |
| ARL10 | 0.010878929 | -0.007252846 | 0.003626083 |
| DAB2IP | 0.041322697 | -0.025284072 | 0.016038624 |
| NUP205 | 0.026925011 | -0.018227087 | 0.008697924 |
| MCEE | 0.042196621 | -0.034289428 | 0.007907193 |
| PIBF1 | 0.028706228 | -0.0258984 | 0.002807829 |
| GTF3C1 | 0.030330728 | -0.01873978 | 0.011590949 |
| RABGEF1 | 0.028042275 | -0.027096556 | 0.000945719 |
| TBL2 | 0.024055173 | -0.01096963 | 0.013085543 |
| ERI3 | 0.041170457 | -0.016967556 | 0.024202901 |
| RAD23B | 0.040360248 | -0.023507468 | 0.016852781 |
| SGK2 | 0.037143272 | -0.039665059 | -0.002521786 |
| TMEM30B | 0.034196638 | -0.032029095 | 0.002167543 |
| ARHGEF12 | 0.046468851 | -0.052064555 | -0.005595704 |
| TOP3A | 0.018944429 | -0.015540429 | 0.003404 |

| | | | |
|---|---|---|---|
| CPTP | 0.035000053 | -0.017600688 | 0.017399364 |
| DNAJA3 | 0.034365654 | -0.025216866 | 0.009148787 |
| GOT2 | 0.042816907 | -0.032395299 | 0.010421607 |
| WAPL | 0.03990991 | -0.033259648 | 0.006650262 |
| TNKS1BP1 | 0.019897211 | -0.022748115 | -0.002850903 |
| ANKRD11 | 0.043601319 | -0.04527077 | -0.001669451 |
| PKD1 | 0.031548524 | -0.015939895 | 0.015608629 |
| EPB41L4A-AS1 | 0.052292979 | -0.025196493 | 0.027096486 |
| SLC44A2 | 0.043090121 | -0.052127699 | -0.009037577 |
| PCYT2 | 0.036984278 | -0.019849638 | 0.01713464 |
| HIP1R | 0.043859708 | -0.076724345 | -0.032864637 |
| CCZ1B | 0.02486382 | -0.023266505 | 0.001597315 |
| CCM2 | 0.02693202 | -0.020786861 | 0.006145159 |
| KLHL22 | 0.024045437 | -0.013161622 | 0.010883815 |
| AL929091.1 | 0.019984628 | -0.018315066 | 0.001669562 |
| CNOT2 | 0.03886775 | -0.031201388 | 0.007666361 |
| PPFIA1 | 0.04038095 | -0.025232568 | 0.015148382 |
| SMARCAD1 | 0.029733672 | -0.023151612 | 0.00658206 |
| DDX39B | 0.053273736 | -0.037104245 | 0.016169491 |
| NUDT15 | 0.033541491 | -0.017396026 | 0.016145465 |
| RSAD1 | 0.030326408 | -0.025605706 | 0.004720702 |
| DDX1 | 0.050323041 | -0.042755934 | 0.007567108 |
| MIB2 | 0.035252053 | -0.02570631 | 0.009545743 |
| ERGIC1 | 0.042245671 | -0.031372434 | 0.010873237 |
| NFE2L1 | 0.056601375 | -0.0369627 | 0.019638675 |
| FZD6 | 0.036163132 | -0.0241426 | 0.012020533 |
| KIAA0319L | 0.039678772 | -0.038691717 | 0.000987055 |
| TMEM203 | 0.041224993 | -0.021602228 | 0.019622764 |
| MT1F | -0.031407297 | 0.023110758 | -0.00829654 |
| HDAC6 | 0.028902622 | -0.017962527 | 0.010940094 |
| PGAP2 | 0.023284786 | -0.018396546 | 0.00488824 |
| MT1H | -0.018605703 | 0.00950484 | -0.009100863 |
| LAMB4 | 0.017019425 | -0.017620036 | -0.000600612 |
| LAMA3 | 0.014952827 | -0.014514514 | 0.000438313 |
| USO1 | 0.045042431 | -0.035179891 | 0.00986254 |
| TARDBP | 0.040897728 | -0.032520557 | 0.008377171 |
| TERF1 | 0.033962874 | -0.024499201 | 0.009463673 |
| ELMOD2 | 0.032706131 | -0.028223237 | 0.004482894 |
| RASA2 | 0.024394711 | -0.017996683 | 0.006398028 |
| ZFYVE28 | 0.016219852 | -0.014697866 | 0.001521986 |
| FANCL | 0.022554872 | -0.016496229 | 0.006058644 |
| CEP170B | 0.024970322 | -0.012870544 | 0.012099778 |
| DNAJC18 | 0.017217288 | -0.00879416 | 0.008423128 |
| AKAP8 | 0.023188956 | -0.014101616 | 0.00908734 |
| JAK2 | 0.020493357 | -0.009188898 | 0.011304459 |
| TP53I13 | 0.043931355 | -0.033103663 | 0.010827692 |
| SUPT6H | 0.034888879 | -0.028000333 | 0.006888546 |

| | | | |
|---|---|---|---|
| TTC37 | 0.046836917 | -0.044736158 | 0.002100759 |
| SFXN4 | 0.037740887 | -0.031230142 | 0.006510744 |
| SRCAP | 0.028349627 | -0.024689938 | 0.003659689 |
| BIRC2 | 0.044314966 | -0.032518975 | 0.011795991 |
| EFR3A | 0.03655562 | -0.019014072 | 0.017541548 |
| HMGXB3 | 0.030352003 | -0.021272722 | 0.009079282 |
| PHF10 | 0.037584945 | -0.018480093 | 0.019104852 |
| CLPTM1 | 0.045890951 | -0.030876711 | 0.01501424 |
| SPATA5 | 0.016438721 | -0.010706525 | 0.005732197 |
| SNRNP35 | 0.039350905 | -0.018776123 | 0.020574782 |
| TCHP | 0.02868867 | -0.015619408 | 0.013069263 |
| TMC6 | 0.014789521 | -0.014326552 | 0.000462969 |
| MFSD8 | 0.027726091 | -0.022768088 | 0.004958003 |
| EBLN3P | 0.045350746 | -0.033598999 | 0.011751747 |
| AL353152.1 | 0.007768279 | -0.007579346 | 0.000188933 |
| R3HDM2 | 0.040518437 | -0.040680927 | -0.00016249 |
| CKMT2-AS1 | 0.018963926 | -0.013679553 | 0.005284373 |
| WDR11 | 0.037881647 | -0.037038861 | 0.000842786 |
| EHD1 | 0.024311154 | -0.022243442 | 0.002067712 |
| HEXD | 0.038372732 | -0.040328335 | -0.001955603 |
| TPP2 | 0.028903052 | -0.021960579 | 0.006942474 |
| BPNT1 | 0.024978611 | -0.013628327 | 0.011350284 |
| SUN1 | 0.040479517 | -0.031765659 | 0.008713859 |
| DLEU1 | 0.02482654 | -0.016075581 | 0.008750959 |
| GFOD2 | 0.019723326 | -0.013657747 | 0.006065579 |
| SNX5 | 0.043714859 | -0.042241191 | 0.001473667 |
| FUS | 0.05500915 | -0.064225621 | -0.009216471 |
| MATN3 | 0.012749833 | -0.013253036 | -0.000503204 |
| SMG9 | 0.018123072 | -0.013618625 | 0.004504447 |
| POM121C | 0.028510626 | -0.016573931 | 0.011936695 |
| ST7L | 0.033058827 | -0.027875239 | 0.005183588 |
| KDM5B | 0.033000447 | -0.019312733 | 0.013687714 |
| GGA1 | 0.033609308 | -0.018026496 | 0.015582811 |
| AIMP2 | 0.034118695 | -0.015170398 | 0.018948297 |
| MRPL34 | 0.06478788 | -0.051817632 | 0.012970249 |
| NDRG1 | -0.052878831 | 0.052964084 | 8.52531E-05 |
| GFM2 | 0.033202976 | -0.022120796 | 0.011082181 |
| RPAP3 | 0.031857358 | -0.03216429 | -0.000306932 |
| INTS3 | 0.030613897 | -0.028848779 | 0.001765118 |
| MSL1 | 0.032761564 | -0.031783906 | 0.000977658 |
| PWWP2A | 0.038256957 | -0.024674201 | 0.013582756 |
| KPNA4 | 0.039378823 | -0.018283801 | 0.021095022 |
| ERAP2 | 0.023546229 | -0.011660437 | 0.011885792 |
| AC105942.1 | 0.017930997 | -0.008384789 | 0.009546208 |
| CSDE1 | 0.060916291 | -0.040709899 | 0.020206392 |
| OGG1 | 0.022866721 | -0.015027373 | 0.007839348 |
| ZFR | 0.040691945 | -0.040731417 | -3.94725E-05 |

| | | | |
|---|---|---|---|
| PDCL3 | 0.042522794 | -0.027085009 | 0.015437785 |
| KHDC4 | 0.031683144 | -0.024720468 | 0.006962676 |
| DENND4C | 0.027200763 | -0.019271677 | 0.007929086 |
| CTTNBP2 | 0.033695711 | -0.020466484 | 0.013229227 |
| NBAS | 0.031839632 | -0.035480467 | -0.003640835 |
| DNTTIP2 | 0.034090828 | -0.019653082 | 0.014437746 |
| VPS16 | 0.024055785 | -0.016471915 | 0.00758387 |
| DEF8 | 0.032944893 | -0.025462121 | 0.007482773 |
| TCP1 | 0.051138189 | -0.039495905 | 0.011642284 |
| PCBP3 | 0.009066766 | -0.00718412 | 0.001882646 |
| CAND1 | 0.041809278 | -0.029828975 | 0.011980303 |
| CISD1 | 0.062680653 | -0.038950143 | 0.02373051 |
| LARP1B | 0.029592028 | -0.024234426 | 0.005357602 |
| GAA | 0.037806038 | -0.047154528 | -0.00934849 |
| MICAL1 | 0.009783609 | -0.009634273 | 0.000149336 |
| NSUN7 | 0.029004405 | -0.019734921 | 0.009269483 |
| SMU1 | 0.036554771 | -0.024823783 | 0.011730988 |
| DNAJC8 | 0.054252331 | -0.028194201 | 0.02605813 |
| POLR2B | 0.040802155 | -0.032741302 | 0.008060853 |
| TMEM248 | 0.044669975 | -0.024805097 | 0.019864878 |
| AFF4 | 0.039685604 | -0.031203814 | 0.008481791 |
| SFXN5 | 0.023085493 | -0.032907356 | -0.009821862 |
| HMGXB4 | 0.028498881 | -0.015741267 | 0.012757613 |
| TBC1D20 | 0.026480502 | -0.017582085 | 0.008898417 |
| ACTR1A | 0.035387183 | -0.021215573 | 0.01417161 |
| TLE3 | 0.02613217 | -0.023701425 | 0.002430745 |
| SHH | 0.010024297 | -0.012003259 | -0.001978962 |
| MPST | 0.037895064 | -0.022950069 | 0.014944995 |
| ACP2 | 0.028100512 | -0.017198347 | 0.010902166 |
| PPP1R3F | 0.018899524 | -0.009012266 | 0.009887258 |
| KCNQ1OT1 | 0.055006803 | -0.026838669 | 0.028168134 |
| ANKZF1 | 0.026536971 | -0.013485666 | 0.013051305 |
| METAP2 | 0.04478293 | -0.035996688 | 0.008786242 |
| EVC2 | 0.013551973 | -0.011720156 | 0.001831818 |
| RMDN3 | 0.033377751 | -0.023151276 | 0.010226475 |
| ADPGK | 0.027196207 | -0.033323771 | -0.006127564 |
| CDKN2AIP | 0.027815887 | -0.012201604 | 0.015614283 |
| DNAJC16 | 0.031357279 | -0.014718123 | 0.016639157 |
| VASP | 0.027059575 | -0.015926186 | 0.011133389 |
| HS3ST1 | 0.015126316 | -0.007189494 | 0.007936822 |
| NFKB1 | 0.028404611 | -0.031968114 | -0.003563503 |
| AP1AR | 0.029356473 | -0.029310754 | 4.57186E-05 |
| DCAF10 | 0.03361355 | -0.014414475 | 0.019199075 |
| SOX4 | -0.054946736 | 0.048726412 | -0.006220324 |
| PTOV1 | 0.045841421 | -0.029831514 | 0.016009907 |
| ATP2C1 | 0.038255378 | -0.026812156 | 0.011443222 |
| KRIT1 | 0.037119469 | -0.022311109 | 0.014808361 |

| | | | |
|---|---|---|---|
| TMEM245 | 0.049230224 | -0.043246275 | 0.005983949 |
| CPT2 | 0.032546763 | -0.014064223 | 0.018482541 |
| STRADA | 0.02310198 | -0.012208366 | 0.010893614 |
| MAP3K2 | 0.041434082 | -0.019597295 | 0.021836787 |
| GRHL3 | 0.007914383 | -0.007122121 | 0.000792262 |
| WHAMM | 0.022464317 | -0.022132818 | 0.000331499 |
| RPS28 | 0.061877285 | -0.038030322 | 0.023846963 |
| RAB11FIP1 | 0.033649897 | -0.029366811 | 0.004283086 |
| TATDN3 | 0.035024118 | -0.021886348 | 0.01313777 |
| DENND1B | 0.029514564 | -0.029342723 | 0.000171841 |
| CORO1B | 0.042509054 | -0.024277516 | 0.018231537 |
| ERO1A | 0.048050468 | -0.042851193 | 0.005199275 |
| CCDC32 | 0.032643061 | -0.021935191 | 0.01070787 |
| NUFIP2 | 0.039804402 | -0.027154382 | 0.01265002 |
| ARL6IP4 | 0.062064908 | -0.03262857 | 0.029436338 |
| ACOT12 | 0.007620335 | -0.007620335 | -7.10543E-15 |
| MARCH2 | 0.035322388 | -0.031802319 | 0.003520069 |
| MDN1 | 0.025924989 | -0.024859066 | 0.001065924 |
| MRPS7 | 0.052485438 | -0.024904796 | 0.027580642 |
| H2AFV | 0.057104916 | -0.02776106 | 0.029343856 |
| CHPT1 | 0.043378817 | -0.065550228 | -0.022171411 |
| IL11RA | 0.01986036 | -0.010669163 | 0.009191197 |
| FAF1 | 0.032858486 | -0.023745691 | 0.009112795 |
| ARSB | 0.013682027 | -0.006784981 | 0.006897046 |
| EBP | 0.047320115 | -0.02923 | 0.018090115 |
| MYO3B | 0.018478117 | -0.017286529 | 0.001191588 |
| OCIAD2 | 0.050117327 | -0.037431896 | 0.012685431 |
| MTMR6 | 0.028069446 | -0.016370874 | 0.011698572 |
| PER1 | 0.0225528 | -0.032060851 | -0.009508052 |
| NCALD | -0.023759869 | 0.01814728 | -0.005612589 |
| MRI1 | 0.024513081 | -0.012160147 | 0.012352935 |
| PNPLA2 | 0.034800976 | -0.050526285 | -0.015725308 |
| NAXE | 0.052809998 | -0.0392066 | 0.013603398 |
| F8A3 | 0.015221873 | -0.01221407 | 0.003007803 |
| PSMA1 | 0.056118692 | -0.030990618 | 0.025128074 |
| LRRC69 | 0.016781042 | -0.016759857 | 2.11855E-05 |
| SF3B3 | 0.033267311 | -0.019283827 | 0.013983484 |
| HACL1 | 0.026325144 | -0.019606467 | 0.006718677 |
| MALT1 | 0.019047377 | -0.019623717 | -0.00057634 |
| FANCC | 0.016707497 | -0.018570884 | -0.001863387 |
| SELENOO | 0.026963878 | -0.025816203 | 0.001147675 |
| CCDC74A | 0.013257341 | -0.00847846 | 0.004778881 |
| VPS11 | 0.026605849 | -0.015354705 | 0.011251145 |
| TNRC6A | 0.042656043 | -0.037155545 | 0.005500498 |
| HDHD3 | 0.033360221 | -0.026632646 | 0.006727575 |
| IGBP1 | 0.0437265 | -0.042524809 | 0.001201691 |
| AL161630.1 | -0.007039912 | 0.003359422 | -0.00368049 |

| | | | |
|---|---|---|---|
| RARG | 0.009868368 | -0.007166061 | 0.002702307 |
| HIPK1 | 0.038106649 | -0.017706736 | 0.020399913 |
| DDX10 | 0.023589018 | -0.014145128 | 0.009443889 |
| IDE | 0.024034395 | -0.024751084 | -0.000716688 |
| TSSC4 | 0.03735109 | -0.020800168 | 0.016550922 |
| G0S2 | 0.079019058 | 0.134399765 | 0.213418823 |
| MRPL32 | 0.047601516 | -0.024643813 | 0.022957703 |
| SIL1 | 0.040467606 | -0.041828785 | -0.001361179 |
| BCAR1 | 0.034147542 | -0.02769033 | 0.006457212 |
| TEAD2 | 0.020269328 | -0.017678586 | 0.002590742 |
| GPATCH4 | 0.026283101 | -0.011498886 | 0.014784215 |
| ING1 | 0.023994831 | -0.017764607 | 0.006230224 |
| UTY | 0.01605483 | -0.017915968 | -0.001861137 |
| ERBIN | 0.043749733 | -0.031546619 | 0.012203114 |
| TNIP2 | 0.03172006 | -0.03089021 | 0.00082985 |
| EDEM2 | 0.026918038 | -0.024123601 | 0.002794438 |
| CSNK1A1 | 0.05403519 | -0.038066499 | 0.01596869 |
| PPP1R2 | 0.043977328 | -0.020401821 | 0.023575507 |
| CBL | 0.020247911 | -0.009452612 | 0.0107953 |
| PNPLA6 | 0.029801155 | -0.021440279 | 0.008360876 |
| WDSUB1 | 0.027807577 | -0.025552593 | 0.002254984 |
| TCAF1 | 0.030175309 | -0.026351019 | 0.00382429 |
| LINC00237 | 0.007559388 | -0.007559388 | -7.10543E-15 |
| KLHL24 | 0.037558196 | -0.034283496 | 0.0032747 |
| ENGASE | 0.022456032 | -0.019105909 | 0.003350122 |
| SLC35E3 | 0.0267677 | -0.013800677 | 0.012967023 |
| NXPH2 | -0.050420357 | 0.089195724 | 0.038775368 |
| RELCH | 0.025266614 | -0.020805562 | 0.004461052 |
| TFAP2A | 0.030180204 | -0.01927796 | 0.010902244 |
| PLAAT4 | 0.075622816 | 0.035848016 | 0.111470831 |
| ENPP5 | 0.023147598 | -0.011182366 | 0.011965233 |
| TIMP3 | 0.055521013 | 0.051394011 | 0.106915025 |
| DTD1 | 0.037444055 | -0.025557795 | 0.01188626 |
| HAUS4 | 0.029604241 | -0.024715105 | 0.004889137 |
| HIVEP2 | 0.019479035 | -0.009652461 | 0.009826574 |
| ZFC3H1 | 0.03106727 | -0.020281767 | 0.010785503 |
| YIF1B | 0.027197463 | -0.016067703 | 0.011129759 |
| DOK1 | 0.012667205 | -0.006319133 | 0.006348072 |
| TMX3 | 0.031877888 | -0.021860732 | 0.010017156 |
| ZNF91 | 0.045839871 | -0.034809208 | 0.011030663 |
| ATP1A1 | 0.072223579 | -0.070206142 | 0.002017437 |
| DDX56 | 0.030249041 | -0.021958513 | 0.008290529 |
| GATA3-AS1 | 0.021122034 | -0.029543519 | -0.008421485 |
| TRIR | 0.060858268 | -0.042127301 | 0.018730967 |
| GATA2 | 0.047642716 | -0.028972044 | 0.018670672 |
| URI1 | 0.042484164 | -0.038824667 | 0.003659497 |
| IL6ST | 0.052037774 | -0.029015051 | 0.023022723 |

| | | | |
|---|---|---|---|
| CYB5D1 | -0.016804832 | 0.008795573 | -0.008009259 |
| NBN | 0.032937293 | -0.021985593 | 0.0109517 |
| API5 | 0.036600689 | -0.025410556 | 0.011190133 |
| LMF1 | 0.028895099 | -0.013785304 | 0.015109794 |
| RAB3GAP1 | 0.036027154 | -0.037236849 | -0.001209695 |
| SREK1IP1 | 0.036237228 | -0.04602985 | -0.009792622 |
| FURIN | 0.033002781 | -0.054330397 | -0.021327616 |
| SGMS1 | 0.040354038 | -0.037856449 | 0.00249759 |
| ETFDH | 0.034186982 | -0.025708764 | 0.008478218 |
| PSMC3IP | 0.01492591 | -0.010425974 | 0.004499936 |
| AAMDC | 0.042947887 | -0.043309482 | -0.000361595 |
| NBEAL2 | 0.032490868 | -0.040107361 | -0.007616493 |
| ASXL1 | 0.04663592 | -0.026076442 | 0.020559477 |
| AC104806.2 | 0.007369252 | -0.007032298 | 0.000336953 |
| WASHC4 | 0.035046471 | -0.024273789 | 0.010772682 |
| CCNJL | 0.019882128 | -0.017227155 | 0.002654973 |
| EIF4G1 | 0.043257977 | -0.039034111 | 0.004223867 |
| CDK5RAP1 | 0.026587623 | -0.014852031 | 0.011735592 |
| C1orf109 | 0.022217739 | -0.010428566 | 0.011789173 |
| RAB11FIP5 | 0.025209226 | -0.016577522 | 0.008631705 |
| ACOX3 | 0.013523955 | -0.008677286 | 0.004846669 |
| ARFGAP3 | 0.0390584 | -0.036160567 | 0.002897833 |
| NFATC2IP | 0.0318563 | -0.023978149 | 0.007878151 |
| AFF1 | 0.032364263 | -0.031683407 | 0.000680855 |
| PUSL1 | 0.022921787 | -0.014548483 | 0.008373305 |
| NQO1 | 0.0223511 | -0.026832884 | -0.004481784 |
| FMNL2 | 0.028155608 | -0.024496272 | 0.003659336 |
| SLC12A6 | 0.022030533 | -0.023298303 | -0.001267769 |
| CUL1 | 0.03037816 | -0.018227731 | 0.012150429 |
| ARHGAP6 | 0.027099151 | -0.02355769 | 0.003541461 |
| POGK | 0.028668292 | -0.014707701 | 0.013960591 |
| CCSER1 | 0.016309082 | -0.018815067 | -0.002505985 |
| TMEM192 | 0.031471952 | -0.036906306 | -0.005434354 |
| NDUFAF4 | 0.047325476 | -0.021465766 | 0.02585971 |
| CNP | 0.032845397 | -0.033200587 | -0.00035519 |
| CMSS1 | 0.032300903 | -0.015233745 | 0.017067158 |
| CALCOCO1 | 0.032710502 | -0.023337687 | 0.009372814 |
| ATG3 | 0.039894021 | -0.024787398 | 0.015106623 |
| SCFD1 | 0.039606595 | -0.022048791 | 0.017557804 |
| ZNF138 | 0.024902654 | -0.023019396 | 0.001883258 |
| DYNC1LI1 | 0.031283579 | -0.023625344 | 0.007658235 |
| ZBED3-AS1 | 0.016320022 | -0.008551133 | 0.007768889 |
| PUM1 | 0.043296992 | -0.045574534 | -0.002277543 |
| BID | 0.011517085 | -0.009855656 | 0.00166143 |
| RAI14 | 0.018959935 | -0.016223989 | 0.002735945 |
| RBM33 | 0.030347479 | -0.026096857 | 0.004250623 |
| COL5A2 | 0.008474488 | -0.006827727 | 0.001646761 |

| | | | |
|---|---|---|---|
| AARSD1 | 0.028135361 | -0.017619439 | 0.010515922 |
| TIA1 | 0.040116756 | -0.044450062 | -0.004333307 |
| PHF23 | 0.03140837 | -0.020198707 | 0.011209663 |
| DDX39A | 0.031930252 | -0.032184178 | -0.000253926 |
| NIPBL | 0.042233677 | -0.042589275 | -0.000355598 |
| SMAD6 | 0.017298915 | -0.011194711 | 0.006104204 |
| AC087741.1 | 0.01759903 | -0.013900124 | 0.003698906 |
| TRRAP | 0.02714438 | -0.030433384 | -0.003289004 |
| DOK4 | 0.018496405 | -0.018625085 | -0.000128681 |
| CASTOR1 | 0.019307995 | -0.011194588 | 0.008113407 |
| MAML3 | 0.016442567 | -0.013235316 | 0.003207251 |
| USP45 | 0.020052303 | -0.012568411 | 0.007483891 |
| ZNF552 | 0.017354682 | -0.014301538 | 0.003053144 |
| FAM20C | 0.021613813 | -0.010703407 | 0.010910405 |
| GCHFR | 0.040620152 | -0.024152152 | 0.016468 |
| NADSYN1 | 0.027679225 | -0.020820381 | 0.006858844 |
| ATR | 0.027584296 | -0.0162697 | 0.011314596 |
| ROCK1 | 0.040712076 | -0.04464804 | -0.003935964 |
| ITCH | 0.032526789 | -0.027234871 | 0.005291918 |
| CCDC180 | 0.014926603 | -0.011028392 | 0.003898211 |
| TAF3 | 0.024982764 | -0.013893936 | 0.011088828 |
| PTMS | 0.058564744 | -0.044713363 | 0.013851381 |
| CDK2AP1 | 0.045704618 | -0.037356335 | 0.008348284 |
| SIRT7 | 0.025846587 | -0.020301387 | 0.005545199 |
| LSM7 | 0.057731043 | -0.025666273 | 0.03206477 |
| STBD1 | 0.009809922 | -0.005375327 | 0.004434595 |
| MTERF4 | 0.030202327 | -0.015183031 | 0.015019295 |
| RITA1 | 0.024699546 | -0.021318309 | 0.003381237 |
| TRAPPC8 | 0.030017585 | -0.023326905 | 0.00669068 |
| C11orf49 | 0.02755064 | -0.019132879 | 0.008417761 |
| F11R | 0.039386728 | -0.03748855 | 0.001898177 |
| RBMS1 | -0.028201445 | 0.019440644 | -0.008760801 |
| WDR6 | 0.039151664 | -0.034814287 | 0.004337377 |
| HMGCR | 0.032694194 | -0.015060805 | 0.017633389 |
| TOR1A | 0.026008097 | -0.018500905 | 0.007507192 |
| PFDN1 | 0.042552919 | -0.024292801 | 0.018260119 |
| SPART | 0.038457224 | -0.021271487 | 0.017185737 |
| RIOX2 | 0.037389589 | -0.021847271 | 0.015542318 |
| CEP152 | 0.023503871 | -0.019437717 | 0.004066153 |
| CYP4F11 | 0.006870608 | -0.006645273 | 0.000225335 |
| CLCNKA | 0.050174863 | -0.050365674 | -0.000190811 |
| PEX11A | 0.019660887 | -0.012999286 | 0.0066616 |
| EPSTI1 | 0.009449776 | -0.0062545 | 0.003195276 |
| HIPK3 | 0.038681151 | -0.042625254 | -0.003944102 |
| VAV3 | 0.041112046 | -0.020444696 | 0.020667351 |
| EPHA1 | 0.013778613 | -0.00845601 | 0.005322603 |
| PRR15L | 0.025711487 | -0.015862744 | 0.009848743 |

| | | | |
|---|---|---|---|
| LYRM4 | 0.027966354 | -0.024699276 | 0.003267078 |
| LIPG | 0.010933919 | -0.009344328 | 0.001589591 |
| PSMF1 | 0.036792679 | -0.019710995 | 0.017081684 |
| SPATA7 | 0.018112403 | -0.012133561 | 0.005978842 |
| RFX1 | 0.025944876 | -0.022958334 | 0.002986542 |
| CARS | 0.027218699 | -0.015981393 | 0.011237306 |
| TSR2 | 0.04260302 | -0.023710067 | 0.018892953 |
| SCNN1A | 0.058237791 | -0.08379566 | -0.025557869 |
| EIF2B2 | 0.031033274 | -0.016206176 | 0.014827098 |
| ATPAF2 | 0.027007508 | -0.014426483 | 0.012581025 |
| POLE3 | 0.029931985 | -0.014811005 | 0.01512098 |
| SLC39A3 | 0.038770294 | -0.022221822 | 0.016548472 |
| FGFR1OP2 | 0.037780897 | -0.020314754 | 0.017466143 |
| RHOT2 | 0.036986275 | -0.030997266 | 0.005989009 |
| STAM2 | 0.034855167 | -0.024768822 | 0.010086345 |
| PSMB4 | 0.051851855 | -0.027245155 | 0.0246067 |
| PICK1 | 0.026344871 | -0.01266691 | 0.013677961 |
| NPM2 | 0.012608003 | -0.012032541 | 0.000575461 |
| LINC00278 | 0.009910714 | -0.009528482 | 0.000382232 |
| FTL | 0.063288187 | -0.069203527 | -0.005915339 |
| C21orf62 | -0.011810593 | 0.006192162 | -0.005618431 |
| LYPLA2 | 0.038013551 | -0.029531034 | 0.008482516 |
| RPS4X | 0.064597585 | 0.029128704 | 0.093726289 |
| BRD2 | 0.046980104 | -0.049742701 | -0.002762597 |
| ALG1 | 0.020557383 | -0.011654509 | 0.008902874 |
| AMN1 | 0.027284753 | -0.013921617 | 0.013363137 |
| MARS | 0.025276884 | -0.018398903 | 0.006877981 |
| C11orf24 | 0.018248643 | -0.010142746 | 0.008105897 |
| SERPINB1 | 0.050060441 | -0.038785746 | 0.011274695 |
| KLHDC3 | 0.029365357 | -0.018388382 | 0.010976975 |
| IFRD2 | 0.031104837 | -0.014607512 | 0.016497325 |
| SLIT3 | 0.009799187 | -0.011127472 | -0.001328285 |
| PLPP1 | 0.031902781 | -0.031029982 | 0.000872799 |
| MIOS | 0.026463467 | -0.018437788 | 0.008025679 |
| RO60 | 0.038265187 | -0.033704462 | 0.004560726 |
| SLC25A36 | 0.048458563 | -0.036045218 | 0.012413345 |
| AL390755.1 | 0.009303572 | -0.009419231 | -0.000115659 |
| FNTA | 0.038835772 | -0.023522872 | 0.015312899 |
| PBX3 | 0.025892445 | -0.020882979 | 0.005009466 |
| RAPGEF6 | 0.025182701 | -0.016466176 | 0.008716525 |
| FBXO3 | 0.034099512 | -0.023348566 | 0.010750947 |
| ZMYND12 | 0.009189693 | -0.00499058 | 0.004199113 |
| RIPK1 | 0.023155876 | -0.018893952 | 0.004261924 |
| MRM2 | 0.027182435 | -0.012597303 | 0.014585132 |
| TOR1B | 0.020478472 | -0.017061308 | 0.003417164 |
| SLC6A13 | 0.015643341 | -0.018775234 | -0.003131893 |
| TK1 | 0.009613977 | -0.008380888 | 0.001233089 |

| | | | |
|---|---|---|---|
| ARHGEF1 | 0.021936753 | -0.025073552 | -0.003136799 |
| ABCF1 | 0.036250094 | -0.028201378 | 0.008048716 |
| GPBP1L1 | 0.040858473 | -0.036564292 | 0.004294182 |
| ZNRF1 | 0.01920133 | -0.02645724 | -0.007255909 |
| MYEF2 | 0.035547966 | -0.032322045 | 0.003225921 |
| SAP30L | 0.027190522 | -0.022782292 | 0.00440823 |
| COPS9 | 0.060224936 | -0.031122343 | 0.029102593 |
| TFRC | 0.036213507 | -0.027826461 | 0.008387045 |
| CEP89 | 0.018904938 | -0.010866647 | 0.008038292 |
| CALCOCO2 | 0.043523756 | -0.031643406 | 0.01188035 |
| YARS | 0.020910222 | -0.011037445 | 0.009872776 |
| ALG2 | 0.033044713 | -0.020172729 | 0.012871984 |
| SPARCL1 | 0.069683518 | -0.120296325 | -0.050612807 |
| PPIE | 0.036204803 | -0.020127841 | 0.016076961 |
| RB1CC1 | 0.033231812 | -0.021040745 | 0.012191067 |
| NCOA1 | 0.033725715 | -0.026888685 | 0.00683703 |
| PTBP1 | 0.038836781 | -0.043616998 | -0.004780216 |
| WTAP | 0.045149892 | -0.040591553 | 0.004558339 |
| C11orf53 | 0.012955771 | -0.01552067 | -0.002564899 |
| ZBED6CL | 0.010154933 | -0.007412344 | 0.002742589 |
| BTN3A2 | 0.020571515 | -0.013958692 | 0.006612824 |
| TOPBP1 | 0.020600855 | -0.017745097 | 0.002855758 |
| UGP2 | 0.04899691 | -0.033054124 | 0.015942786 |
| SOCS4 | 0.029557464 | -0.018708051 | 0.010849413 |
| COMMD4 | 0.044021052 | -0.036177778 | 0.007843275 |
| NSUN2 | 0.026893752 | -0.013483249 | 0.013410502 |
| TSTD2 | 0.022032304 | -0.011571242 | 0.010461061 |
| LMNTD2 | 0.014477212 | -0.008706619 | 0.005770592 |
| NUTM2A-AS1 | 0.037499624 | -0.024482164 | 0.01301746 |
| TMPRSS3 | 0.020820701 | -0.014316658 | 0.006504044 |
| DUS4L | 0.022793694 | -0.01048241 | 0.012311283 |
| BTD | 0.027337218 | -0.031991211 | -0.004653993 |
| ESR1 | 0.009788096 | -0.011499243 | -0.001711147 |
| ACTR1B | 0.027223104 | -0.01781579 | 0.009407314 |
| PTPRA | 0.042416681 | -0.048260645 | -0.005843964 |
| CDK18 | 0.015611871 | -0.010220378 | 0.005391493 |
| ERCC5 | 0.026722713 | -0.018179698 | 0.008543015 |
| GTF2I | 0.053410105 | -0.060490583 | -0.007080477 |
| CDK7 | 0.021764594 | -0.012489768 | 0.009274827 |
| TPM3 | 0.047784889 | -0.030021173 | 0.017763716 |
| CNPY3 | 0.028807609 | -0.018191399 | 0.01061621 |
| DMXL1 | 0.03688048 | -0.04683191 | -0.009951431 |
| TFPI2 | 0.010865788 | -0.014770936 | -0.003905148 |
| RAF1 | 0.032931719 | -0.047223214 | -0.014291495 |
| RASSF1 | 0.017897229 | -0.013312869 | 0.004584359 |
| MUS81 | 0.023379371 | -0.015161196 | 0.008218175 |
| PRKAA1 | 0.034743824 | -0.03653642 | -0.001792596 |

| | | | |
|---|---|---|---|
| DPYSL2 | 0.019750418 | -0.022847335 | -0.003096916 |
| PARP12 | 0.02055749 | -0.019990563 | 0.000566926 |
| GGNBP2 | 0.037920136 | -0.02136817 | 0.016551966 |
| RRM2B | 0.02899912 | -0.014972767 | 0.014026353 |
| WBP11 | 0.039404349 | -0.023998232 | 0.015406117 |
| C8orf76 | 0.024552922 | -0.016748632 | 0.00780429 |
| FAM117B | 0.023746635 | -0.027400203 | -0.003653568 |
| GAPVD1 | 0.028557331 | -0.021560039 | 0.006997293 |
| HINFP | 0.015021225 | -0.00926181 | 0.005759415 |
| AL356124.1 | 0.015802319 | -0.017913604 | -0.002111285 |
| AQP2 | 0.044073045 | -0.126623012 | -0.082549968 |
| UBE2I | 0.04581204 | -0.024920207 | 0.020891834 |
| DDX60 | 0.017207433 | -0.009379725 | 0.007827708 |
| AC009509.1 | 0.00647225 | -0.00647225 | -8.88178E-15 |
| ESRRA | 0.045166391 | -0.033805934 | 0.011360457 |
| SERPINA4 | 0.011673745 | -0.014426172 | -0.002752427 |
| CTSB | 0.047856708 | -0.031067621 | 0.016789087 |
| ZNF511 | 0.04136106 | -0.025137887 | 0.016223172 |
| IGF2BP2 | 0.023145182 | -0.015510325 | 0.007634857 |
| NUP214 | 0.031946467 | -0.026110052 | 0.005836415 |
| MPP1 | 0.032036734 | -0.018320827 | 0.013715907 |
| CRBN | 0.034731638 | -0.022076271 | 0.012655367 |
| HDAC8 | 0.024650801 | -0.022318903 | 0.002331897 |
| U2AF2 | 0.031275547 | -0.026830106 | 0.004445441 |
| PQBP1 | 0.030998174 | -0.027353102 | 0.003645072 |
| RYK | 0.034828417 | -0.019839226 | 0.014989191 |
| ANKMY2 | 0.022583051 | -0.013471784 | 0.009111267 |
| TRAPPC12 | 0.030306508 | -0.016656497 | 0.013650011 |
| ARMT1 | 0.032937079 | -0.021154217 | 0.011782861 |
| DNAAF4 | 0.016439041 | -0.008401438 | 0.008037603 |
| POFUT2 | 0.021793464 | -0.020528011 | 0.001265453 |
| S100A2 | -0.061152822 | 0.04892526 | -0.012227562 |
| AKT2 | 0.030963955 | -0.01769039 | 0.013273565 |
| DPY19L4 | 0.033449664 | -0.039333053 | -0.00588339 |
| SMC2 | 0.02454526 | -0.019424174 | 0.005121086 |
| EPS15 | 0.035990599 | -0.025395126 | 0.010595473 |
| ULK3 | 0.029970022 | -0.028574895 | 0.001395127 |
| AC005674.2 | 0.010110514 | -0.005250608 | 0.004859905 |
| ZNF254 | 0.032169547 | -0.01889526 | 0.013274288 |
| DCBLD2 | 0.030500354 | -0.02826306 | 0.002237294 |
| ZNF563 | 0.011356831 | -0.00670995 | 0.004646881 |
| AC009275.1 | 0.007032638 | -0.006748208 | 0.00028443 |
| METTL4 | 0.020399613 | -0.01783043 | 0.002569183 |
| ZDHHC8 | 0.022955667 | -0.030088526 | -0.007132859 |
| UQCC1 | 0.026180208 | -0.01244293 | 0.013737278 |
| ALG12 | 0.023103358 | -0.023992399 | -0.000889041 |
| DAPK1 | 0.022178837 | -0.027828463 | -0.005649626 |

| | | | |
|---|---|---|---|
| FNBP1 | 0.025000529 | -0.021601611 | 0.003398918 |
| PLEKHM2 | 0.02340229 | -0.020597971 | 0.002804319 |
| GRAMD1A | 0.021394351 | -0.019322335 | 0.002072016 |
| UBN1 | 0.02516814 | -0.016527096 | 0.008641044 |
| SPTLC2 | 0.055432603 | -0.056602224 | -0.001169622 |
| CALCB | 0.006696526 | -0.006696526 | -3.55271E-15 |
| LARP1 | 0.042136508 | -0.031048689 | 0.011087819 |
| SIAE | 0.026741065 | -0.025615 | 0.001126065 |
| ZNF66 | 0.015396081 | -0.011724656 | 0.003671424 |
| WLS | 0.046769233 | -0.022704425 | 0.024064807 |
| EXOSC8 | 0.037390436 | -0.029148504 | 0.008241932 |
| NLN | 0.019230071 | -0.01319558 | 0.006034491 |
| PIGBOS1 | 0.02991764 | -0.026924639 | 0.002993001 |
| PSMB10 | 0.030649049 | -0.017847504 | 0.012801546 |
| SNX1 | 0.034833111 | -0.022511045 | 0.012322066 |
| IFIT2 | 0.021709397 | -0.024686113 | -0.002976716 |
| CEMIP2 | 0.014297703 | -0.01667434 | -0.002376637 |
| LPAR2 | 0.0129281 | -0.010371119 | 0.002556981 |
| ABHD2 | 0.052558548 | 0.025981851 | 0.078540399 |
| CCDC28B | 0.015037557 | -0.007947043 | 0.007090514 |
| EVL | 0.028422466 | -0.013379164 | 0.015043302 |
| LINC01315 | 0.007779464 | -0.005577075 | 0.002202388 |
| SIRT2 | 0.031718771 | -0.01881377 | 0.012905002 |
| NEFL | -0.010757802 | 0.006718618 | -0.004039184 |
| CYP4X1 | 0.007473449 | -0.006712628 | 0.000760821 |
| GOLT1B | 0.032012184 | -0.018223726 | 0.013788459 |
| RCC1L | 0.031244963 | -0.01721256 | 0.014032403 |
| ANKRD29 | 0.03146712 | -0.023931086 | 0.007536034 |
| PIAS1 | 0.033891893 | -0.027959563 | 0.00593233 |
| TMTC4 | 0.024957692 | -0.013291305 | 0.011666387 |
| DOLPP1 | 0.018886703 | -0.011199483 | 0.007687219 |
| HULC | 0.01316668 | -0.008758067 | 0.004408613 |
| CC2D2A | 0.019246374 | -0.020828421 | -0.001582047 |
| LINC02672 | 0.009403517 | -0.008019637 | 0.00138388 |
| RAP1GDS1 | 0.025737866 | -0.022854208 | 0.002883658 |
| GPR137B | 0.027819396 | -0.022780987 | 0.005038409 |
| AL133453.1 | 0.029939191 | -0.030366213 | -0.000427022 |
| GPATCH2L | 0.032490565 | -0.017736248 | 0.014754317 |
| PGM3 | 0.022447925 | -0.019692248 | 0.002755676 |
| ERCC6L2 | 0.028199627 | -0.022164292 | 0.006035335 |
| SAMM50 | 0.034332558 | -0.021241698 | 0.01309086 |
| RPA2 | 0.032736435 | -0.026713987 | 0.006022448 |
| PANX1 | 0.01369492 | -0.011091099 | 0.002603821 |
| QSOX2 | 0.014235036 | -0.009713722 | 0.004521314 |
| WRNIP1 | 0.027267324 | -0.015221146 | 0.012046178 |
| SRPK1 | 0.026622832 | -0.024400848 | 0.002221984 |
| M1AP | 0.005673681 | -0.005673681 | -5.32907E-15 |

| | | | |
|---|---|---|---|
| UBASH3B | 0.011228773 | -0.006138427 | 0.005090346 |
| RPP25 | 0.017188246 | -0.014842676 | 0.00234557 |
| BBS2 | 0.026219652 | -0.016806816 | 0.009412836 |
| VPS4A | 0.033591531 | -0.034024343 | -0.000432813 |
| EGLN3 | 0.01117424 | -0.011412508 | -0.000238268 |
| MCM3 | 0.02642587 | -0.023378936 | 0.003046935 |
| ADGRL1 | 0.019575853 | -0.013151082 | 0.006424771 |
| ZNF770 | 0.033237127 | -0.026860139 | 0.006376988 |
| RABGAP1 | 0.032299244 | -0.035258353 | -0.002959108 |
| TRMT11 | 0.026903873 | -0.02005213 | 0.006851743 |
| DNASE1L1 | 0.023136311 | -0.014894377 | 0.008241934 |
| ERBB4 | -0.028441379 | 0.023808737 | -0.004632642 |
| CCDC77 | 0.014078216 | -0.012242593 | 0.001835624 |
| HES6 | 0.016460736 | -0.009182775 | 0.007277962 |
| SHROOM3 | 0.037734383 | -0.020014882 | 0.0177195 |
| PARN | 0.026603009 | -0.013843815 | 0.012759194 |
| TIMM8B | 0.054028682 | 0.029314077 | 0.083342759 |
| GCNA | 0.008109499 | -0.006214615 | 0.001894883 |
| DCAF8 | 0.036478024 | -0.024267616 | 0.012210408 |
| MTSS1 | 0.031619619 | -0.019711862 | 0.011907757 |
| CAPNS1 | 0.052358447 | -0.033127677 | 0.01923077 |
| FAM210A | 0.025637523 | -0.017699005 | 0.007938518 |
| KCMF1 | 0.033118596 | -0.027620204 | 0.005498392 |
| IRF3 | 0.034341537 | -0.040497028 | -0.006155491 |
| ACTR8 | 0.0205223 | -0.015770626 | 0.004751673 |
| SLU7 | 0.02625109 | -0.01773581 | 0.00851528 |
| TSNAX | 0.039443972 | -0.022311782 | 0.017132189 |
| NUCKS1 | 0.057240384 | -0.040877825 | 0.016362559 |
| GLA | 0.022767452 | -0.018298994 | 0.004468458 |
| HMGCL | 0.038406905 | -0.027177299 | 0.011229606 |
| RBPJ | 0.041032499 | -0.030549795 | 0.010482704 |
| SCRN1 | 0.03580782 | -0.024335354 | 0.011472466 |
| ZBTB43 | 0.02575031 | -0.014231494 | 0.011518816 |
| ZNF160 | 0.023030513 | -0.011766559 | 0.011263955 |
| TRIM38 | 0.031532754 | -0.019385676 | 0.012147079 |
| FGFR3 | -0.023538387 | 0.030951473 | 0.007413085 |
| GIPC1 | 0.036346378 | -0.028850389 | 0.00749599 |
| KLF13 | 0.036282978 | -0.027165814 | 0.009117164 |
| MXI1 | 0.025329592 | -0.034141228 | -0.008811637 |
| RRNAD1 | 0.027227932 | -0.021118842 | 0.00610909 |
| SDHB | 0.045984267 | -0.027328813 | 0.018655454 |
| TKFC | 0.021802522 | -0.011573946 | 0.010228576 |
| RBM26 | 0.036559031 | -0.021591024 | 0.014968007 |
| RTCA | 0.032432244 | -0.027388539 | 0.005043705 |
| TRMT6 | 0.022881697 | -0.018702528 | 0.004179168 |
| ZC3H7A | 0.031680102 | -0.038670485 | -0.006990383 |
| ID2 | -0.030205857 | 0.020631595 | -0.009574263 |

| | | | |
|---|---|---|---|
| CASP7 | 0.025780291 | -0.024557072 | 0.001223218 |
| SVIL | 0.026427489 | -0.016721468 | 0.00970602 |
| PCID2 | 0.034522689 | -0.020814097 | 0.013708592 |
| TMEM143 | 0.018281807 | -0.010859002 | 0.007422806 |
| AC104031.1 | 0.007202459 | -0.006116729 | 0.00108573 |
| RSF1 | 0.039993006 | -0.029291083 | 0.010701923 |
| C3orf33 | 0.022056807 | -0.016434052 | 0.005622755 |
| URM1 | 0.035868362 | -0.019801171 | 0.016067191 |
| DTNBP1 | 0.025022524 | -0.014191812 | 0.010830713 |
| ATP5MD | 0.061487859 | -0.039790824 | 0.021697035 |
| SNAPC2 | 0.018967548 | -0.013296453 | 0.005671095 |
| TACC3 | 0.007767765 | -0.007861159 | -9.33945E-05 |
| PARP6 | 0.018883202 | -0.014446342 | 0.004436859 |
| RAB3GAP2 | 0.029293635 | -0.026551839 | 0.002741796 |
| SLC7A1 | 0.017769994 | -0.015317492 | 0.002452502 |
| CHMP4C | 0.022007211 | -0.015288673 | 0.006718538 |
| FBXO11 | 0.030765009 | -0.023192015 | 0.007572994 |
| LINC02343 | 0.0117837 | -0.011965632 | -0.000181932 |
| ZBTB41 | 0.028616953 | -0.015036607 | 0.013580346 |
| EIF3C | 0.013971858 | -0.010449987 | 0.003521871 |
| CYB561A3 | 0.022749858 | -0.016108407 | 0.006641452 |
| ENTR1 | 0.022350394 | -0.017442089 | 0.004908304 |
| SH3TC1 | 0.035440245 | -0.024286451 | 0.011153793 |
| STC1 | -0.014908029 | -0.009057572 | -0.023965601 |
| CHCHD5 | 0.049034811 | -0.027472703 | 0.021562108 |
| NAGA | 0.020896199 | -0.021838371 | -0.000942172 |
| CSK | 0.022955902 | -0.025274382 | -0.00231848 |
| NUDT7 | 0.032240134 | -0.031476229 | 0.000763905 |
| UBE2H | 0.041638162 | -0.024606966 | 0.017031196 |
| AL138756.1 | 0.026031634 | -0.0237288 | 0.002302834 |
| SMIM3 | 0.012908608 | -0.010492267 | 0.002416341 |
| RRP1B | 0.018751514 | -0.014126779 | 0.004624735 |
| CDK19 | 0.015198646 | -0.016261422 | -0.001062776 |
| GGA2 | 0.035501704 | -0.039670928 | -0.004169223 |
| PXN-AS1 | 0.017815232 | -0.013652603 | 0.004162629 |
| NUMB | 0.0407833 | -0.047161958 | -0.006378658 |
| ORMDL3 | 0.025924734 | -0.012521483 | 0.013403251 |
| TMEM181 | 0.027487857 | -0.02210322 | 0.005384637 |
| TAF1D | 0.043800175 | -0.035411598 | 0.008388577 |
| LINC00662 | 0.026450397 | -0.032028769 | -0.005578371 |
| PTRH2 | 0.024976435 | -0.014499054 | 0.010477381 |
| PTDSS1 | 0.020226089 | -0.018970366 | 0.001255723 |
| GON7 | 0.036711636 | -0.020995873 | 0.015715762 |
| FANCI | 0.008670146 | -0.005805069 | 0.002865076 |
| FAM91A1 | 0.02617491 | -0.020416807 | 0.005758103 |
| AL157392.3 | 0.013208613 | -0.010563263 | 0.00264535 |
| KIF1BP | 0.027544403 | -0.013816386 | 0.013728017 |

| | | | |
|---|---|---|---|
| MBOAT1 | 0.023425979 | -0.018890228 | 0.004535751 |
| TMEM161B-AS1 | 0.041651402 | -0.053512478 | -0.011861076 |
| CEP70 | 0.029519437 | -0.020576223 | 0.008943214 |
| IFIT3 | 0.021949195 | -0.015416472 | 0.006532723 |
| MMUT | 0.036484345 | -0.028077594 | 0.008406751 |
| NIFK-AS1 | 0.020241045 | -0.01030436 | 0.009936685 |
| OBSL1 | -0.030718453 | 0.020363623 | -0.01035483 |
| SLIT2 | -0.048967148 | 0.080179279 | 0.03121213 |
| SLC29A2 | 0.022148324 | -0.018089943 | 0.004058381 |
| PAFAH1B3 | 0.029444922 | -0.018856945 | 0.010587977 |
| FBXL19 | 0.015019136 | -0.012175914 | 0.002843222 |
| ZNF800 | 0.029081702 | -0.026219459 | 0.002862244 |
| MYL6B | 0.046096622 | -0.060495074 | -0.014398452 |
| LTA4H | 0.032285338 | -0.032136515 | 0.000148823 |
| MAGI2-AS3 | -0.024987173 | 0.018273072 | -0.006714101 |
| YTHDC2 | 0.026796427 | -0.022982928 | 0.003813499 |
| BMS1 | 0.028937291 | -0.02477657 | 0.004160721 |
| PELI3 | 0.01353955 | -0.007264951 | 0.0062746 |
| MBOAT7 | 0.028013466 | -0.029247361 | -0.001233895 |
| PDS5A | 0.029054633 | -0.026304789 | 0.002749843 |
| TMEM167A | 0.045230657 | -0.022548736 | 0.022681921 |
| UQCRH | 0.061228685 | -0.042426053 | 0.018802632 |
| COG6 | 0.021596096 | -0.012596483 | 0.008999614 |
| LRP6 | 0.031744256 | -0.017662149 | 0.014082107 |
| EHMT1 | 0.024995547 | -0.02171928 | 0.003276267 |
| COL4A4 | 0.038346474 | -0.035180287 | 0.003166187 |
| RSRC1 | 0.032588811 | -0.029473143 | 0.003115669 |
| TMEM220 | 0.030225284 | -0.022078053 | 0.008147231 |
| SNRPA1 | 0.036274804 | -0.020298231 | 0.015976573 |
| EIF2B4 | 0.023511019 | -0.018351109 | 0.005159911 |
| DIXDC1 | 0.015311978 | -0.010298841 | 0.005013137 |
| TNFSF12 | 0.021203341 | -0.012592145 | 0.008611196 |
| AL512274.1 | 0.007300519 | -0.006464826 | 0.000835693 |
| DDX23 | 0.026286434 | -0.023861827 | 0.002424607 |
| EWSR1 | 0.04040104 | -0.045444334 | -0.005043294 |
| PRRC2B | 0.043881121 | -0.023833848 | 0.020047274 |
| RASGEF1A | 0.005878993 | -0.005878993 | -3.55271E-15 |
| AC008467.1 | 0.006684804 | -0.006121237 | 0.000563567 |
| KLHL2 | 0.018673589 | -0.009191028 | 0.009482561 |
| CLIP2 | 0.01056011 | -0.011170057 | -0.000609947 |
| PJA2 | 0.044937695 | -0.035808501 | 0.009129195 |
| ASCC2 | 0.021635877 | -0.014076848 | 0.00755903 |
| UGGT2 | 0.023851463 | -0.017303204 | 0.006548259 |
| ALDOA | 0.03047947 | -0.020941512 | 0.009537958 |
| HCAR2 | 0.007911056 | -0.00969015 | -0.001779094 |
| SAMD9L | 0.014191533 | -0.016251363 | -0.002059831 |
| ZNF32 | 0.033265625 | -0.026924117 | 0.006341508 |

| | | | |
|---|---|---|---|
| SLC33A1 | 0.028345575 | -0.017277389 | 0.011068186 |
| DBNDD2 | 0.022415817 | -0.026186534 | -0.003770717 |
| BTG2 | 0.032201133 | -0.029371276 | 0.002829857 |
| NPAP1 | 0.008936294 | -0.011323227 | -0.002386933 |
| SESTD1 | 0.027638064 | -0.030750972 | -0.003112908 |
| EPN1 | 0.046317696 | -0.040597539 | 0.005720157 |
| CLMN | 0.042054538 | -0.051500116 | -0.009445577 |
| CGN | 0.022010902 | -0.018363019 | 0.003647883 |
| RBM28 | 0.029233626 | -0.024835256 | 0.00439837 |
| MRRF | 0.023778077 | -0.013188811 | 0.010589267 |
| SLC25A3 | 0.062641457 | -0.031919476 | 0.03072198 |
| PDXK | 0.045283611 | -0.034860632 | 0.01042298 |
| LNX1 | 0.025900671 | -0.019053792 | 0.006846879 |
| HLCS | 0.018703981 | -0.014760968 | 0.003943013 |
| ADAMTS1 | 0.009590834 | -0.006826447 | 0.002764387 |
| NOXA1 | 0.014814414 | -0.007730037 | 0.007084377 |
| WDR59 | 0.022564933 | -0.022870011 | -0.000305079 |
| TTC14 | 0.036414336 | -0.027140418 | 0.009273919 |
| TRIP10 | 0.021685147 | -0.019338994 | 0.002346153 |
| TMEM237 | 0.027847927 | -0.01554342 | 0.012304507 |
| LINC00963 | 0.027724257 | -0.015782478 | 0.01194178 |
| GALNT11 | 0.033896817 | -0.018295345 | 0.015601472 |
| CXorf40B | 0.017975566 | -0.01496106 | 0.003014507 |
| PSMG3 | 0.035182594 | -0.038276585 | -0.003093991 |
| LINC00958 | 0.025667291 | -0.022600499 | 0.003066792 |
| SNX14 | 0.037459076 | -0.02953763 | 0.007921446 |
| SMG1 | 0.036415958 | -0.048569937 | -0.012153979 |
| RDH13 | 0.026882588 | -0.021740799 | 0.005141789 |
| CNFN | 0.025265997 | -0.035567119 | -0.010301122 |
| BMP7 | -0.024552969 | 0.032075297 | 0.007522328 |
| SP3 | 0.02858106 | -0.015640926 | 0.012940134 |
| CAPN15 | 0.023988303 | -0.020205028 | 0.003783275 |
| MCRS1 | 0.033536633 | -0.028556515 | 0.004980119 |
| RRP15 | 0.027911504 | -0.017843829 | 0.010067674 |
| NMNAT1 | 0.019722466 | -0.015739347 | 0.00398312 |
| HSD17B7 | 0.023291944 | -0.023042128 | 0.000249816 |
| MLLT3 | 0.020640545 | -0.017717236 | 0.00292331 |
| SLC37A3 | 0.020989299 | -0.014500787 | 0.006488513 |
| AL355974.2 | 0.006375614 | -0.005932806 | 0.000442808 |
| LRRC42 | 0.021799257 | -0.022998099 | -0.001198843 |
| SMYD2 | 0.020976871 | -0.020954882 | 2.1989E-05 |
| LTBR | 0.026798582 | -0.036597669 | -0.009799086 |
| SNX15 | 0.022133224 | -0.012599369 | 0.009533854 |
| ARFIP1 | 0.031597153 | -0.027898217 | 0.003698936 |
| ARRB1 | 0.018474013 | -0.021431688 | -0.002957675 |
| LZTS3 | 0.021856916 | -0.024193074 | -0.002336158 |
| PAIP2B | 0.017370775 | -0.008793193 | 0.008577582 |

| | | | |
|---|---|---|---|
| NARS2 | 0.020696294 | -0.016884408 | 0.003811886 |
| ATAD2 | 0.013356501 | -0.01397183 | -0.000615329 |
| MBD2 | 0.031540343 | -0.029736018 | 0.001804325 |
| OCRL | 0.019759968 | -0.010452814 | 0.009307153 |
| ATL3 | 0.029301815 | -0.030574142 | -0.001272327 |
| HOXD3 | 0.021214888 | -0.015448049 | 0.005766838 |
| AC015845.2 | 0.005812776 | -0.005391513 | 0.000421263 |
| CCAR1 | 0.034360894 | -0.035625382 | -0.001264488 |
| NAE1 | 0.03460937 | -0.023502951 | 0.011106419 |
| PDZD2 | 0.029183849 | -0.027596686 | 0.001587163 |
| GCN1 | 0.023223837 | -0.01541739 | 0.007806447 |
| SLC25A19 | 0.015045323 | -0.011533294 | 0.003512029 |
| RNF212 | 0.027730841 | -0.035283675 | -0.007552834 |
| NAA10 | 0.0422829 | -0.028373575 | 0.013909325 |
| NUP98 | 0.024778871 | -0.020183805 | 0.004595066 |
| SSH1 | 0.019830895 | -0.021068817 | -0.001237922 |
| PPP2R3C | 0.027203176 | -0.018648529 | 0.008554647 |
| AP2A1 | 0.022627887 | -0.020663139 | 0.001964747 |
| LINC02814 | 0.006522666 | -0.005598999 | 0.000923667 |
| RUSC1 | 0.021692639 | -0.015701292 | 0.005991347 |
| COQ8A | 0.023521983 | -0.020950076 | 0.002571907 |
| EIF3J-DT | 0.027365924 | -0.02283884 | 0.004527085 |
| ACBD6 | 0.030607096 | -0.017941338 | 0.012665758 |
| KMT2B | 0.024946383 | -0.028430059 | -0.003483676 |
| LDLRAD4 | 0.010310814 | -0.01342229 | -0.003111475 |
| DDX17 | 0.057588119 | -0.050625415 | 0.006962704 |
| RMND1 | 0.022478664 | -0.018259748 | 0.004218916 |
| DENND4A | 0.023271297 | -0.014462582 | 0.008808716 |
| GRHL2 | 0.020353151 | -0.01997131 | 0.000381842 |
| ZNF141 | 0.02709641 | -0.016554534 | 0.010541876 |
| APOC1 | -0.063469844 | 0.127931348 | 0.064461504 |
| SLC39A11 | 0.018427279 | -0.022184426 | -0.003757147 |
| FBXO9 | 0.036134096 | -0.024243843 | 0.011890253 |
| C1QTNF4 | 0.018544209 | -0.020581333 | -0.002037124 |
| UBE2E1 | 0.03459574 | -0.027582571 | 0.00701317 |
| COX20 | 0.052602434 | -0.040707648 | 0.011894786 |
| ICE2 | 0.032082646 | -0.022434331 | 0.009648314 |
| ACVR1B | 0.032148407 | -0.0261898 | 0.005958607 |
| ABCC10 | 0.014243414 | -0.010569391 | 0.003674023 |
| PPP1R21 | 0.0228065 | -0.031241093 | -0.008434594 |
| SDAD1 | 0.027819842 | -0.01713171 | 0.010688132 |
| ADCY9 | 0.016058932 | -0.020931226 | -0.004872294 |
| BLOC1S5 | 0.024159639 | -0.022227774 | 0.001931865 |
| CDK11B | 0.019218992 | -0.011453957 | 0.007765035 |
| CLUAP1 | 0.026914872 | -0.023093426 | 0.003821446 |
| NAV2 | 0.029399168 | -0.024980728 | 0.00441844 |
| PSMA3 | 0.047611409 | -0.02729235 | 0.020319059 |

| | | | |
|---|---|---|---|
| UBALD1 | 0.021519177 | -0.021875074 | -0.000355897 |
| BRAF | 0.028810193 | -0.018387054 | 0.010423139 |
| SFT2D2 | 0.027939999 | -0.034523317 | -0.006583318 |
| KLC2 | 0.022705057 | -0.012802228 | 0.009902829 |
| TMEM128 | 0.034587096 | -0.03147873 | 0.003108366 |
| STAT2 | 0.024894195 | -0.01559644 | 0.009297755 |
| ZFPL1 | 0.032949248 | -0.027534399 | 0.005414849 |
| DBNL | 0.036819475 | -0.021606762 | 0.015212712 |
| CTPS2 | 0.014549934 | -0.008153355 | 0.00639658 |
| MRPL58 | 0.032442069 | -0.015876006 | 0.016566063 |
| APPBP2 | 0.027243143 | -0.02033099 | 0.006912153 |
| CREBL2 | 0.033281469 | -0.020750849 | 0.01253062 |
| SIRT1 | 0.019924776 | -0.011180598 | 0.008744178 |
| C2orf49 | 0.030074373 | -0.026872769 | 0.003201604 |
| C2CD5 | 0.025748853 | -0.025006868 | 0.000741985 |
| AP001107.9 | 0.008835226 | -0.006826637 | 0.002008589 |
| IFI44 | 0.013093671 | -0.007615213 | 0.005478459 |
| CYFIP1 | 0.031104798 | -0.030370931 | 0.000733867 |
| TRABD | 0.034782497 | -0.035067879 | -0.000285382 |
| AC016831.5 | 0.016186704 | -0.014872765 | 0.001313939 |
| CHMP1A | 0.03058247 | -0.018805414 | 0.011777056 |
| MFSD12 | 0.026536088 | -0.030379278 | -0.00384319 |
| TTC36 | 0.010608283 | -0.00674624 | 0.003862043 |
| DKC1 | 0.026988266 | -0.021597476 | 0.005390791 |
| COPS3 | 0.033848014 | -0.016612695 | 0.017235319 |
| RNF113A | 0.019638772 | -0.009910635 | 0.009728136 |
| HIRIP3 | 0.021479356 | -0.012943561 | 0.008535796 |
| SLFN12 | 0.012665364 | -0.012688784 | -2.34193E-05 |
| CCNB1IP1 | 0.043283795 | -0.026255771 | 0.017028023 |
| SPECC1 | 0.027493135 | -0.026166677 | 0.001326457 |
| CDR2 | 0.014410734 | -0.017485391 | -0.003074657 |
| ZNF518A | 0.033648007 | -0.019449424 | 0.014198583 |
| CUL4B | 0.025383109 | -0.014860557 | 0.010522552 |
| EPHA2 | 0.013493367 | -0.0123482 | 0.001145167 |
| B4GALNT3 | 0.022627157 | -0.01587886 | 0.006748297 |
| HADH | 0.033058647 | -0.020503488 | 0.012555158 |
| RAB3IP | 0.036819372 | -0.026899275 | 0.009920098 |
| CRYZL1 | 0.028985022 | -0.019913222 | 0.009071799 |
| ALDH4A1 | 0.024258524 | -0.022132401 | 0.002126123 |
| ICA1L | 0.01400727 | -0.018422311 | -0.004415041 |
| TRIP6 | 0.026061854 | -0.018220017 | 0.007841837 |
| MYH14 | 0.02681636 | -0.019646533 | 0.007169827 |
| RING1 | 0.029512533 | -0.017833793 | 0.01167874 |
| AC008610.1 | 0.008562012 | -0.00713232 | 0.001429692 |
| TTC21B | 0.021939425 | -0.023624876 | -0.001685451 |
| YDJC | 0.030990303 | -0.017465083 | 0.01352522 |
| ARSK | 0.017628055 | -0.021620821 | -0.003992766 |

| | | | |
|---|---|---|---|
| TRIM13 | 0.026179837 | -0.017709669 | 0.008470168 |
| ZCCHC9 | 0.028330203 | -0.029041289 | -0.000711086 |
| C11orf1 | 0.040965023 | -0.028922413 | 0.01204261 |
| SYVN1 | 0.027709535 | -0.016616952 | 0.011092583 |
| TTN-AS1 | 0.017590831 | -0.009174856 | 0.008415975 |
| GCNT2 | 0.026992321 | -0.022334008 | 0.004658313 |
| DACH1 | -0.01788693 | 0.015301993 | -0.002584937 |
| AGPAT3 | 0.030377335 | -0.020944505 | 0.009432831 |
| CDK12 | 0.027957041 | -0.02368423 | 0.004272812 |
| NELFB | 0.030050498 | -0.030050023 | 4.74614E-07 |
| C6orf226 | 0.026681341 | -0.015623203 | 0.011058138 |
| STX7 | 0.034637608 | -0.023592499 | 0.011045109 |
| CDC14B | 0.022965988 | -0.017506845 | 0.005459143 |
| SLFN13 | 0.013207804 | -0.018321018 | -0.005113214 |
| KCNMA1 | -0.017510448 | 0.020838654 | 0.003328206 |
| ZNF33B | 0.027857722 | -0.023859429 | 0.003998294 |
| IFI35 | 0.020429785 | -0.011356833 | 0.009072952 |
| KCNJ6 | -0.010933185 | 0.006439035 | -0.00449415 |
| SNUPN | 0.024165207 | -0.016387372 | 0.007777835 |
| DAP3 | 0.03465777 | -0.025142972 | 0.009514798 |
| HAX1 | 0.046616721 | -0.029611171 | 0.01700555 |
| CEP295 | 0.019935551 | -0.025766292 | -0.005830741 |
| NKAP | 0.02664586 | -0.018707896 | 0.007937964 |
| USP37 | 0.01815024 | -0.012629806 | 0.005520434 |
| MELTF-AS1 | 0.012588655 | -0.012401167 | 0.000187488 |
| EMX2OS | 0.034385038 | -0.029234152 | 0.005150886 |
| C9orf40 | 0.017226654 | -0.009461949 | 0.007764705 |
| TSBP1-AS1 | 0.009301471 | -0.01156337 | -0.002261899 |
| BCLAF1 | 0.041637302 | -0.033650338 | 0.007986964 |
| SNX25 | 0.017138371 | -0.011583362 | 0.005555009 |
| TP53BP1 | 0.026513906 | -0.015618474 | 0.010895432 |
| TGFB1 | 0.028891108 | -0.029904468 | -0.00101336 |
| KRBA2 | 0.01911285 | -0.014028199 | 0.00508465 |
| A1BG | 0.023424513 | -0.011794363 | 0.01163015 |
| AC002553.2 | 0.006715882 | -0.005270411 | 0.001445471 |
| FRRS1 | 0.012441208 | -0.012525002 | -8.37943E-05 |
| CREB1 | 0.025723589 | -0.015403162 | 0.010320427 |
| USP47 | 0.035685773 | -0.026753613 | 0.00893216 |
| NDUFAF6 | 0.025764091 | -0.015922783 | 0.009841308 |
| HPRT1 | 0.051415489 | 0.044308256 | 0.095723745 |
| AL589935.1 | 0.011082749 | -0.010618561 | 0.000464188 |
| MED19 | 0.031030665 | -0.017213251 | 0.013817414 |
| CCDC120 | 0.011846023 | -0.009023363 | 0.00282266 |
| AC008969.1 | 0.016141338 | -0.01186404 | 0.004277297 |
| VPS41 | 0.026609355 | -0.02448204 | 0.002127315 |
| PLEKHD1 | 0.008959573 | -0.007650113 | 0.001309459 |
| CUTA | 0.05597282 | -0.028180468 | 0.027792352 |

| | | | |
|---|---|---|---|
| QPCTL | 0.007555884 | -0.004303671 | 0.003252212 |
| COX8A | 0.056569042 | -0.06137371 | -0.004804667 |
| SMPD1 | 0.026890686 | -0.017451092 | 0.009439595 |
| NUDT8 | 0.02812272 | -0.031432565 | -0.003309846 |
| LRRC8B | 0.016230421 | -0.009362703 | 0.006867718 |
| NT5C2 | 0.027532866 | -0.023382814 | 0.004150053 |
| RELA | 0.026179868 | -0.017361065 | 0.008818803 |
| SHPRH | 0.032239872 | -0.038136273 | -0.005896401 |
| ABHD3 | 0.028351811 | -0.021990448 | 0.006361362 |
| LTBP2 | -0.016054765 | 0.008403092 | -0.007651673 |
| SPEN | 0.031553661 | -0.033965663 | -0.002412003 |
| GXYLT1 | 0.022119426 | -0.019088609 | 0.003030817 |
| CNNM4 | 0.014180519 | -0.011985021 | 0.002195498 |
| ASAP2 | 0.036742821 | -0.042617719 | -0.005874898 |
| EVPLL | 0.005858536 | -0.005501542 | 0.000356994 |
| EDA | 0.007132818 | -0.006780719 | 0.000352099 |
| UBE2K | 0.039034403 | -0.030479255 | 0.008555147 |
| RNF43 | 0.023430194 | -0.015664264 | 0.007765931 |
| INAFM1 | 0.021409289 | -0.015749613 | 0.005659676 |
| PHTF2 | 0.014587807 | -0.014965478 | -0.00037767 |
| UIMC1 | 0.018080459 | -0.014857084 | 0.003223375 |
| YBEY | 0.029744482 | -0.019731501 | 0.01001298 |
| ANKRD27 | 0.016564813 | -0.010962511 | 0.005602302 |
| ILRUN | 0.023422201 | -0.018167829 | 0.005254372 |
| CTBP2 | 0.034984535 | -0.02152724 | 0.013457294 |
| RABGGTA | 0.021552599 | -0.013776962 | 0.007775637 |
| DDAH2 | 0.042314082 | -0.033149626 | 0.009164456 |
| ORAI2 | 0.016490798 | -0.00964049 | 0.006850307 |
| AC104237.3 | 0.008594811 | -0.008671339 | -7.65279E-05 |
| BCL3 | 0.006676577 | -0.00499718 | 0.001679397 |
| RPL7L1 | 0.038467152 | -0.024859139 | 0.013608013 |
| ERC1 | 0.024386615 | -0.026954429 | -0.002567814 |
| TMEM187 | 0.020181604 | -0.013580346 | 0.006601259 |
| CENPS | 0.025604222 | -0.027325399 | -0.001721177 |
| AGA | 0.023897471 | -0.014001175 | 0.009896296 |
| AL035661.1 | 0.025066254 | -0.028932875 | -0.003866621 |
| DTWD2 | 0.018173655 | -0.018688707 | -0.000515052 |
| AC093627.5 | 0.013284843 | -0.007797601 | 0.005487242 |
| DBP | 0.022304673 | -0.021357919 | 0.000946754 |
| FZD5 | 0.013787161 | -0.01804847 | -0.004261309 |
| TUBB2A | 0.027627972 | -0.020576104 | 0.007051869 |
| ZNHIT3 | 0.035783729 | -0.021402402 | 0.014381327 |
| SPTBN5 | 0.007703365 | -0.008451358 | -0.000747993 |
| ICMT | 0.029601583 | -0.018557257 | 0.011044326 |
| AGK | 0.019371071 | -0.010849089 | 0.008521983 |
| IL3RA | 0.013640421 | -0.007804701 | 0.005835721 |
| NCOA2 | 0.028339229 | -0.021497065 | 0.006842164 |

| | | | |
|---|---|---|---|
| AC100801.1 | 0.010420787 | -0.005795693 | 0.004625095 |
| LY6L | 0.008428535 | -0.004681981 | 0.003746554 |
| ABRAXAS2 | 0.022459915 | -0.020326022 | 0.002133893 |
| NXT1 | 0.025088233 | -0.017031003 | 0.00805723 |
| ARAP1 | 0.038723997 | -0.022358271 | 0.016365726 |
| SEPTIN8 | 0.029880302 | -0.035128436 | -0.005248135 |
| PRMT7 | 0.016710986 | -0.012200087 | 0.004510899 |
| IFT74 | 0.022157663 | -0.025491364 | -0.003333701 |
| PLEKHA5 | 0.026538491 | -0.032772123 | -0.006233632 |
| GDI2 | 0.039879387 | -0.040942256 | -0.001062868 |
| SMAD7 | 0.023212182 | -0.011796821 | 0.01141536 |
| AL157938.3 | 0.012486377 | -0.006627101 | 0.005859276 |
| TIMM44 | 0.027707884 | -0.020017288 | 0.007690596 |
| POLR3GL | 0.034719224 | -0.021908126 | 0.012811098 |
| AC008147.2 | 0.009821724 | -0.007678248 | 0.002143476 |
| UBR1 | 0.023246612 | -0.025271133 | -0.002024521 |
| TMEM125 | 0.021780103 | -0.018963904 | 0.002816199 |
| EEF2 | 0.052601537 | -0.029589183 | 0.023012353 |
| EYA4 | 0.011919895 | -0.01681237 | -0.004892474 |
| LRCH3 | 0.020749856 | -0.013999377 | 0.006750479 |
| ATF4 | 0.03373116 | -0.024830185 | 0.008900975 |
| WFS1 | 0.017109638 | -0.021469204 | -0.004359567 |
| CRIM1 | -0.04487218 | 0.062541845 | 0.017669665 |
| SDR39U1 | 0.036779426 | -0.025143008 | 0.011636418 |
| RPS6KC1 | 0.014311596 | -0.009604642 | 0.004706954 |
| NSMCE4A | 0.029818135 | -0.02561448 | 0.004203655 |
| TSPYL5 | 0.024500937 | -0.017947493 | 0.006553444 |
| ARID2 | 0.025467452 | -0.018608365 | 0.006859087 |
| KIAA1143 | 0.02669266 | -0.020374611 | 0.006318049 |
| ZNF433-AS1 | 0.016930076 | -0.013880841 | 0.003049235 |
| FAM13A-AS1 | 0.010877384 | -0.009956349 | 0.000921035 |
| ZNF430 | 0.018080675 | -0.019519345 | -0.00143867 |
| LSAMP | 0.009161895 | -0.011305101 | -0.002143206 |
| FER | 0.024006093 | -0.020306441 | 0.003699652 |
| LINC01637 | 0.008923677 | -0.005096981 | 0.003826696 |
| ZDHHC7 | 0.024081792 | -0.015940913 | 0.008140879 |
| ITSN1 | 0.021930725 | -0.014123824 | 0.007806901 |
| MFSD4B | 0.020869208 | -0.011895644 | 0.008973564 |
| SLC16A7 | 0.047412275 | -0.030543005 | 0.01686927 |
| WDR43 | 0.027134431 | -0.016416127 | 0.010718305 |
| AVPR1A | -0.035445542 | 0.037356508 | 0.001910965 |
| BRWD1 | 0.037587359 | -0.030842266 | 0.006745093 |
| TSGA10 | 0.023749769 | -0.014620676 | 0.009129094 |
| FAM13A | 0.042308807 | -0.040428137 | 0.001880671 |
| TENT4B | 0.020249084 | -0.015228754 | 0.00502033 |
| STON1 | 0.01637602 | -0.015937535 | 0.000438484 |
| ZNF354A | 0.015737714 | -0.014942754 | 0.000794959 |

| | | | |
|---|---|---|---|
| FZR1 | 0.021092355 | -0.01274145 | 0.008350905 |
| TOR1AIP1 | 0.028800538 | -0.01890335 | 0.009897189 |
| TCTA | 0.029951317 | -0.023036373 | 0.006914943 |
| NCOA3 | 0.025400885 | -0.022103794 | 0.003297092 |
| WNT5A | 0.006594911 | -0.004967248 | 0.001627663 |
| SELENOI | 0.035315428 | -0.035425887 | -0.000110458 |
| TRIM73 | 0.02251639 | -0.028764702 | -0.006248312 |
| DDX43 | 0.00527878 | -0.004987737 | 0.000291044 |
| POLG | 0.01930847 | -0.01189177 | 0.007416701 |
| AC010969.2 | 0.013181458 | -0.006995517 | 0.006185941 |
| AP001042.1 | 0.005848614 | -0.005513511 | 0.000335103 |
| AC048341.1 | 0.010079486 | -0.011659862 | -0.001580376 |
| G3BP1 | 0.037845213 | -0.033067433 | 0.00477778 |
| FXR1 | 0.041291844 | -0.043585546 | -0.002293703 |
| GDF15 | 0.02853002 | -0.022422314 | 0.006107706 |
| MEN1 | 0.017516324 | -0.013881753 | 0.003634571 |
| MIR29B2CHG | 0.02488841 | -0.015728238 | 0.009160172 |
| ZNF205 | 0.017430801 | -0.009003768 | 0.008427033 |
| ANKRD13A | 0.028757877 | -0.016437303 | 0.012320573 |
| KIAA0100 | 0.030358252 | -0.031725099 | -0.001366847 |
| AC005532.1 | 0.005839086 | -0.005052434 | 0.000786652 |
| TMEM120A | 0.02438386 | -0.022803913 | 0.001579946 |
| SQOR | 0.027823301 | -0.019983918 | 0.007839384 |
| TBC1D31 | 0.010080225 | -0.006265596 | 0.003814629 |
| MAPK1 | 0.036426999 | -0.021853465 | 0.014573534 |
| MAPRE1 | 0.036498022 | -0.023124917 | 0.013373105 |
| BAK1 | 0.017022747 | -0.01140652 | 0.005616227 |
| AC073349.1 | 0.00788199 | -0.006344066 | 0.001537924 |
| MSN | 0.036348541 | -0.028013847 | 0.008334694 |
| WDFY3 | 0.023646504 | -0.019899508 | 0.003746996 |
| TMEM45B | 0.018511018 | -0.026676972 | -0.008165954 |
| PDGFA | -0.015231048 | 0.010058488 | -0.00517256 |
| AAAS | 0.027897917 | -0.023532538 | 0.004365379 |
| OSBPL3 | 0.024359945 | -0.024626724 | -0.000266779 |
| VOPP1 | 0.021651409 | -0.017968609 | 0.0036828 |
| ZFP14 | 0.023850688 | -0.02370562 | 0.000145068 |
| CCNT2 | 0.027966841 | -0.027628314 | 0.000338527 |
| BLZF1 | 0.021172114 | -0.013063927 | 0.008108188 |
| PIEZO1 | 0.022717502 | -0.018036289 | 0.004681213 |
| RPS6KB2 | 0.028495795 | -0.019475115 | 0.00902068 |
| RBBP6 | 0.032480381 | -0.019556067 | 0.012924315 |
| PRH1 | 0.026556027 | -0.019567777 | 0.00698825 |
| PPP6R2 | 0.03474894 | -0.036402914 | -0.001653974 |
| TMEM163 | 0.01434334 | -0.017783844 | -0.003440504 |
| SCT | 0.006183092 | -0.00548149 | 0.000701601 |
| ZNF316 | 0.019140167 | -0.011287634 | 0.007852533 |
| MIR3681HG | 0.013320462 | -0.011106384 | 0.002214078 |

| | | | |
|---|---|---|---|
| GPATCH2 | 0.022594089 | -0.017999307 | 0.004594782 |
| RNF14 | 0.029530924 | -0.026811518 | 0.002719406 |
| RPN1 | 0.040863226 | -0.042442574 | -0.001579348 |
| MARC1 | 0.01335477 | -0.018192298 | -0.004837528 |
| TBILA | 0.005282087 | -0.005282087 | -5.32907E-15 |
| ANK3 | 0.039299015 | -0.030359138 | 0.008939877 |
| PEX6 | 0.023496911 | -0.018907403 | 0.004589507 |
| ITPR3 | 0.011288697 | -0.012255547 | -0.00096685 |
| SND1 | 0.024738192 | -0.018093537 | 0.006644655 |
| CYP1A2 | 0.005155049 | -0.005155049 | -1.77636E-15 |
| ZNF836 | 0.012246382 | -0.007497299 | 0.004749084 |
| ADK | 0.025005889 | -0.03061737 | -0.005611481 |
| SMG6 | 0.022221088 | -0.021627263 | 0.000593826 |
| RPS6KA5 | 0.011706128 | -0.00986498 | 0.001841148 |
| TLE5 | 0.051470932 | -0.032928982 | 0.018541951 |
| TNKS2 | 0.028240866 | -0.030807849 | -0.002566983 |
| AIP | 0.033190734 | -0.023448382 | 0.009742351 |
| NEK7 | 0.02478443 | -0.017205624 | 0.007578806 |
| APOL6 | 0.019106561 | -0.011318584 | 0.007787977 |
| CENPT | 0.021917946 | -0.02089643 | 0.001021516 |
| GPSM3 | 0.009042473 | -0.008948029 | 9.44442E-05 |
| CHKA | 0.02518148 | -0.015909227 | 0.009272253 |
| GMPS | 0.029152232 | -0.016687834 | 0.012464398 |
| AL022323.1 | 0.005338505 | -0.004951847 | 0.000386658 |
| MTMR1 | 0.013432613 | -0.007250099 | 0.006182513 |
| SFPQ | 0.048713644 | -0.034384616 | 0.014329028 |
| UBE3C | 0.026609987 | -0.033699235 | -0.007089248 |
| TBC1D10B | 0.020218494 | -0.012419306 | 0.007799188 |
| DESI2 | 0.022910324 | -0.020675378 | 0.002234946 |
| AP4E1 | 0.015782258 | -0.01403332 | 0.001748938 |
| WDR75 | 0.020913959 | -0.018931425 | 0.001982534 |
| S100A16 | 0.022570039 | -0.02057743 | 0.001992609 |
| CU638689.4 | -0.010946543 | 0.014085394 | 0.003138851 |
| TMEM41A | 0.019826407 | -0.018872246 | 0.000954161 |
| HPS1 | 0.02215561 | -0.020647236 | 0.001508374 |
| NUMA1 | 0.039513386 | -0.043646247 | -0.004132861 |
| SKA2 | 0.031385879 | -0.017216057 | 0.014169823 |
| ERAL1 | 0.02544829 | -0.023423965 | 0.002024326 |
| RECQL | 0.024072796 | -0.01773598 | 0.006336816 |
| ZCCHC7 | 0.024802449 | -0.026554564 | -0.001752115 |
| LBR | 0.025794915 | -0.026011155 | -0.00021624 |
| EZH1 | 0.023541917 | -0.032060984 | -0.008519066 |
| C12orf65 | 0.027758482 | -0.016951479 | 0.010807003 |
| EIF3M | 0.041995613 | -0.024073068 | 0.017922545 |
| HSH2D | 0.006644962 | -0.007596893 | -0.000951931 |
| COG1 | 0.017776074 | -0.011578485 | 0.006197589 |
| ZNF581 | 0.018202283 | -0.010541676 | 0.007660607 |

| | | | |
|---|---|---|---|
| AC007114.1 | 0.014837585 | -0.016156854 | -0.001319269 |
| CNOT8 | 0.023285454 | -0.017454257 | 0.005831197 |
| OGFOD3 | 0.029016834 | -0.015588277 | 0.013428557 |
| LCORL | 0.021636138 | -0.017140742 | 0.004495395 |
| EMP1 | -0.009837607 | 0.007019064 | -0.002818544 |
| KMT5B | 0.03242053 | -0.021953945 | 0.010466585 |
| ANKS3 | 0.02339657 | -0.016693884 | 0.006702686 |
| MECOM | 0.044120485 | -0.078207615 | -0.03408713 |
| MYO5B | 0.019879243 | -0.011581896 | 0.008297347 |
| INSIG1 | 0.016701966 | -0.010041872 | 0.006660093 |
| DDX19B | 0.018822005 | -0.013612894 | 0.005209111 |
| METTL9 | 0.040203298 | -0.032261034 | 0.007942264 |
| UTP6 | 0.023164909 | -0.019677944 | 0.003486965 |
| ZFAND5 | 0.038335025 | -0.030154026 | 0.008180998 |
| FBXL2 | 0.017742888 | -0.011802107 | 0.005940781 |
| AP3D1 | 0.02875498 | -0.022107425 | 0.006647555 |
| TTC33 | 0.023598489 | -0.016343284 | 0.007255205 |
| PYROXD1 | 0.02474205 | -0.016458767 | 0.008283282 |
| AC007255.1 | 0.005691389 | -0.00467293 | 0.001018459 |
| ATG4B | 0.028392017 | -0.024600359 | 0.003791657 |
| NEURL1B | 0.008088126 | -0.006756706 | 0.001331419 |
| OSBPL9 | 0.037285492 | -0.039441816 | -0.002156324 |
| CTNNBL1 | 0.023515259 | -0.020494012 | 0.003021247 |
| TBRG1 | 0.026384649 | -0.01959891 | 0.006785739 |
| TRA2A | 0.036030995 | -0.020031366 | 0.015999629 |
| KDM1B | 0.013806964 | -0.009958307 | 0.003848657 |
| STX18 | 0.037620865 | -0.035376966 | 0.002243899 |
| ASCC3 | 0.024791378 | -0.014064097 | 0.010727281 |
| RFXANK | 0.02845241 | -0.020605869 | 0.007846541 |
| PALLD | -0.023366922 | 0.015942119 | -0.007424803 |
| ZFHX4 | 0.006648012 | -0.007663683 | -0.001015672 |
| ATF2 | 0.026589494 | -0.017499931 | 0.009089562 |
| AC139100.2 | 0.006181692 | -0.005124096 | 0.001057596 |
| HFE | 0.012452181 | -0.012158133 | 0.000294048 |
| KLHL8 | 0.021549552 | -0.023366206 | -0.001816654 |
| MAP2K1 | 0.03130752 | -0.017354758 | 0.013952763 |
| C9 | 0.014834093 | -0.01069786 | 0.004136233 |
| CFAP36 | 0.028028719 | -0.03035425 | -0.002325531 |
| GNL3 | 0.021217634 | -0.011591684 | 0.00962595 |
| LAMB2 | 0.027671447 | -0.040518998 | -0.012847551 |
| TMEM43 | 0.021786106 | -0.018845258 | 0.002940849 |
| BRWD3 | 0.017252678 | -0.015462656 | 0.001790022 |
| CENPJ | 0.010956203 | -0.009821442 | 0.001134761 |
| SHFL | 0.021830454 | -0.023441841 | -0.001611387 |
| BBIP1 | 0.035235904 | -0.025226265 | 0.010009639 |
| KIAA1211L | 0.005452794 | -0.004962388 | 0.000490406 |
| PHF14 | 0.030526929 | -0.020446621 | 0.010080308 |

| | | | |
|---|---|---|---|
| HOOK1 | 0.030000494 | -0.023916923 | 0.00608357 |
| EVI5 | 0.027350688 | -0.023711805 | 0.003638883 |
| TMEM8A | 0.035223569 | -0.02051612 | 0.01470745 |
| AL161772.1 | 0.006975365 | -0.003917254 | 0.003058111 |
| FAM98A | 0.023790678 | -0.026917789 | -0.003127111 |
| CEP19 | 0.009267151 | -0.010764684 | -0.001497533 |
| ATP8B1 | 0.021449394 | -0.031843963 | -0.01039457 |
| ADCK2 | 0.018797024 | -0.02122784 | -0.002430816 |
| AC002070.1 | 0.008634853 | -0.005781037 | 0.002853816 |
| MLH1 | 0.021157165 | -0.018287826 | 0.002869339 |
| USP9X | 0.038671901 | -0.025838762 | 0.01283314 |
| AC135507.1 | 0.013086542 | -0.009365176 | 0.003721367 |
| GIPC2 | -0.019778747 | 0.013916369 | -0.005862377 |
| TEAD1 | 0.0266043 | -0.025991398 | 0.000612902 |
| ZNF274 | 0.01530826 | -0.015345926 | -3.76662E-05 |
| AC091271.1 | -0.012381585 | 0.006638784 | -0.005742801 |
| TICAM1 | 0.012653118 | -0.009057923 | 0.003595195 |
| AKAP1 | 0.03263273 | -0.027893862 | 0.004738867 |
| FAS | 0.006564176 | -0.004099669 | 0.002464508 |
| TFF3 | 0.003581222 | -0.003581222 | -8.88178E-15 |
| PIGG | 0.019236199 | -0.012082274 | 0.007153925 |
| SLC22A18 | 0.019076056 | -0.020582373 | -0.001506317 |
| ARL4D | 0.050136698 | 0.042419047 | 0.092555744 |
| REXO4 | 0.023570351 | -0.027021202 | -0.003450851 |
| SLC18B1 | 0.023587 | -0.016535525 | 0.007051475 |
| PHF3 | 0.040543475 | -0.034794712 | 0.005748763 |
| GSK3B | 0.025043976 | -0.015581312 | 0.009462664 |
| HEATR5A | 0.021259792 | -0.020104109 | 0.001155683 |
| NOXO1 | 0.007098452 | -0.006144026 | 0.000954426 |
| RNF126 | 0.024630055 | -0.015573861 | 0.009056195 |
| INSC | 0.004918526 | -0.004918526 | -5.32907E-15 |
| DAZAP1 | 0.026566198 | -0.016085964 | 0.010480234 |
| LRSAM1 | 0.018232605 | -0.016470826 | 0.00176178 |
| STAG2 | 0.034176686 | -0.054490851 | -0.020314165 |
| WDR55 | 0.020027169 | -0.021594872 | -0.001567703 |
| CENPX | 0.039021445 | -0.021407365 | 0.01761408 |
| TPRA1 | 0.026935696 | -0.020307258 | 0.006628438 |
| RNPC3 | 0.029198939 | -0.024799326 | 0.004399613 |
| UBE2G1 | 0.027100397 | -0.021991587 | 0.00510881 |
| SLF2 | 0.021234164 | -0.0125451 | 0.008689063 |
| MAP3K11 | 0.019654601 | -0.02053637 | -0.000881769 |
| PNMA1 | 0.024222572 | -0.025859335 | -0.001636763 |
| N4BP2 | 0.019282552 | -0.012305591 | 0.006976961 |
| PPWD1 | 0.026054411 | -0.02921778 | -0.00316337 |
| SNRPC | 0.035682393 | -0.019493744 | 0.016188649 |
| SLC39A13 | 0.028353337 | -0.023365499 | 0.004987838 |
| SSRP1 | 0.025974225 | -0.014447746 | 0.011526479 |

| | | | |
|---|---|---|---|
| ZNF720 | 0.022624502 | -0.017689225 | 0.004935276 |
| ARHGDIA | 0.042755535 | -0.029696201 | 0.013059334 |
| FGF9 | -0.045949778 | 0.078459173 | 0.032509395 |
| INTS9 | 0.015920938 | -0.012968315 | 0.002952622 |
| SH3GL1 | 0.015528709 | -0.009164262 | 0.006364447 |
| GADD45B | 0.047284832 | -0.055767222 | -0.00848239 |
| RIOK3 | 0.027717092 | -0.018707971 | 0.009009121 |
| MAP2K5 | 0.013946392 | -0.011422888 | 0.002523504 |
| QKI | 0.034632435 | -0.020201728 | 0.014430708 |
| CDK13 | 0.028346668 | -0.026877345 | 0.001469323 |
| CCDC28A | 0.032549098 | -0.027651517 | 0.004897581 |
| IFT88 | 0.024735631 | -0.035306988 | -0.010571357 |
| PANK4 | 0.017512371 | -0.010968074 | 0.006544297 |
| PEX7 | 0.034463532 | 0.023172168 | 0.0576357 |
| UXS1 | 0.030382192 | -0.04572316 | -0.015340968 |
| NTS | 0.003881345 | -0.003881345 | -5.32907E-15 |
| PHLDA2 | 0.006378092 | -0.004972971 | 0.001405121 |
| SDHAF4 | 0.023564797 | -0.019747398 | 0.003817399 |
| STRBP | 0.028107902 | -0.02462772 | 0.003480182 |
| SLFN5 | 0.011189757 | -0.010341527 | 0.00084823 |
| ZBTB42 | 0.012391713 | -0.007733305 | 0.004658408 |
| MAD2L1 | 0.011755496 | -0.00769676 | 0.004058736 |
| PPM1A | 0.032335913 | -0.031367034 | 0.000968879 |
| AP006287.2 | 0.007916019 | -0.004484217 | 0.003431802 |
| DLG3 | 0.026122279 | -0.027890608 | -0.001768329 |
| PTPN3 | 0.028863834 | -0.035803871 | -0.006940038 |
| AL356740.3 | 0.006057357 | -0.00533246 | 0.000724896 |
| RCN1 | 0.028703803 | -0.02723021 | 0.001473593 |
| HIPK2 | 0.051771794 | 0.034235139 | 0.086006934 |
| TMEM184B | 0.019776157 | -0.017002924 | 0.002773233 |
| TRUB2 | 0.025659459 | -0.016869758 | 0.008789701 |
| MKLN1 | 0.033442174 | -0.022463195 | 0.010978979 |
| TRAF3IP1 | 0.017216731 | -0.011815191 | 0.00540154 |
| DDR2 | 0.009868336 | -0.009206327 | 0.000662009 |
| DDB1 | 0.039914386 | -0.027053339 | 0.012861047 |
| TAX1BP3 | 0.026529299 | -0.019014195 | 0.007515104 |
| INTU | 0.025414674 | -0.033119933 | -0.007705259 |
| ZNF33A | 0.03065431 | -0.034584307 | -0.003929997 |
| APC | 0.024915027 | -0.025139073 | -0.000224046 |
| SWT1 | 0.016156169 | -0.013098277 | 0.003057892 |
| ZNF793 | 0.015949392 | -0.010066229 | 0.005883164 |
| USP5 | 0.021905493 | -0.01219658 | 0.009708912 |
| HBP1 | 0.030126973 | -0.027521556 | 0.002605417 |
| MTO1 | 0.017026157 | -0.010022696 | 0.007003461 |
| TOX4 | 0.02909913 | -0.030187624 | -0.001088495 |
| DCAF4L1 | 0.006687088 | -0.00421653 | 0.002470558 |
| NAT9 | 0.023335307 | -0.015314703 | 0.008020604 |

| | | | |
|---|---|---|---|
| C4orf48 | 0.036477064 | 0.033047767 | 0.069524831 |
| AC018730.1 | 0.006543836 | -0.006062764 | 0.000481072 |
| GNAI2 | 0.0342962 | -0.01908001 | 0.01521619 |
| ITPRID2 | 0.020322229 | -0.015158907 | 0.005163322 |
| RPAP2 | 0.024048974 | -0.017024756 | 0.007024218 |
| HNRNPLL | 0.019056205 | -0.014248063 | 0.004808141 |
| DCLRE1B | 0.007335977 | -0.005169406 | 0.002166571 |
| ALG8 | 0.022328719 | -0.014565939 | 0.00776278 |
| B3GALT4 | 0.017977679 | -0.020435716 | -0.002458037 |
| TLCD5 | 0.014605551 | -0.009446585 | 0.005158966 |
| TMEM189 | 0.020607903 | -0.013794028 | 0.006813875 |
| ZBED3 | 0.023456665 | -0.029933689 | -0.006477024 |
| ZNF224 | 0.019005656 | -0.0147781 | 0.004227555 |
| REV1 | 0.032862858 | -0.025234759 | 0.007628099 |
| REEP4 | 0.014708805 | -0.01750696 | -0.002798154 |
| IDO1 | 0.005003855 | -0.004523514 | 0.000480341 |
| KPNA6 | 0.028860844 | -0.024190818 | 0.004670026 |
| UPF3B | 0.019695002 | -0.018031527 | 0.001663475 |
| BCL7A | 0.028904793 | -0.027846168 | 0.001058626 |
| LLGL1 | 0.009396493 | -0.011005202 | -0.00160871 |
| MRPL10 | 0.025635926 | -0.015969674 | 0.009666252 |
| TRAK2 | 0.027175541 | -0.020549211 | 0.00662633 |
| SIRT4 | 0.011621663 | -0.011652058 | -3.0395E-05 |
| TPST2 | 0.026501783 | -0.029299355 | -0.002797573 |
| HNRNPA0 | 0.040833275 | -0.023519416 | 0.017313859 |
| ZDHHC16 | 0.021992036 | -0.01224286 | 0.009749176 |
| TMEM170A | 0.031236285 | -0.020350725 | 0.010885559 |
| EXOC2 | 0.016630809 | -0.010535169 | 0.006095641 |
| TMEM99 | 0.022877694 | -0.021070941 | 0.001806753 |
| ARID1B | 0.035069812 | -0.037865966 | -0.002796155 |
| HCAR3 | 0.005451124 | -0.004548123 | 0.000903001 |
| FLOT2 | 0.026055745 | -0.020973425 | 0.00508232 |
| ANKRA2 | 0.020069887 | -0.016814873 | 0.003255014 |
| IFT20 | 0.027944977 | -0.026313697 | 0.00163128 |
| LINC01991 | 0.004430447 | -0.004430447 | -3.55271E-15 |
| DNAJC25 | 0.019994495 | -0.02030119 | -0.000306696 |
| CLDND1 | 0.029844563 | -0.01948857 | 0.010355994 |
| SLAIN1 | 0.006404025 | -0.005445386 | 0.000958639 |
| CTNNAL1 | 0.031432162 | -0.024274885 | 0.007157277 |
| ASCL2 | 0.017073142 | -0.027116179 | -0.010043036 |
| HDAC2 | 0.028874788 | -0.015853123 | 0.013021665 |
| PCED1B | 0.010149395 | -0.008265872 | 0.001883523 |
| SBNO1-AS1 | 0.017035472 | -0.012626901 | 0.004408572 |
| GK5 | 0.026392136 | -0.02645893 | -6.67942E-05 |
| CYYR1 | 0.008387142 | -0.012227102 | -0.003839959 |
| ANKRD26 | 0.025975617 | -0.04229292 | -0.016317303 |
| NFKBID | 0.007429753 | -0.004873626 | 0.002556127 |

| | | | |
|---|---|---|---|
| CRNKL1 | 0.022555309 | -0.014320948 | 0.008234361 |
| CUX1 | 0.032818573 | -0.026155704 | 0.006662868 |
| SMIM27 | 0.031086417 | -0.023476799 | 0.007609618 |
| EIF3J | 0.034529465 | -0.033943217 | 0.000586248 |
| SGTA | 0.029237003 | -0.022790185 | 0.006446818 |
| HDAC9 | 0.005258076 | -0.004453809 | 0.000804267 |
| ARF5 | 0.04009534 | -0.049041447 | -0.008946107 |
| ODF2L | 0.026033897 | -0.022174759 | 0.003859138 |
| S100A4 | 0.0352074 | -0.071486249 | -0.036278849 |
| SHC2 | 0.008254788 | -0.004728832 | 0.003525956 |
| TMEM39B | 0.014182044 | -0.010438616 | 0.003743428 |
| GIGYF1 | 0.026072863 | -0.024009715 | 0.002063148 |
| TSR1 | 0.018838706 | -0.011797646 | 0.00704106 |
| GRTP1 | 0.019014762 | -0.013685381 | 0.005329381 |
| ATRIP | 0.010246631 | -0.006061902 | 0.004184729 |
| CFAP20 | 0.021045873 | -0.013804483 | 0.00724139 |
| ZNF211 | 0.014536419 | -0.009094692 | 0.005441727 |
| NARF | 0.023440065 | -0.027112199 | -0.003672134 |
| HEY1 | 0.01597495 | -0.013398788 | 0.002576162 |
| ACAD9 | 0.020124385 | -0.013543616 | 0.006580769 |
| CCDC50 | 0.033557924 | -0.051238419 | -0.017680496 |
| DNAJC7 | 0.033011429 | -0.026210027 | 0.006801402 |
| SPATA6 | 0.023089956 | -0.027600357 | -0.004510402 |
| KLF3 | 0.023995407 | -0.022194842 | 0.001800565 |
| RIPOR1 | -0.023240315 | 0.028343423 | 0.005103108 |
| AL162424.1 | 0.003965277 | -0.003965277 | -7.10543E-15 |
| PTPN9 | 0.018279588 | -0.01411552 | 0.004164067 |
| TOMM70 | 0.024694531 | -0.02475472 | -6.01884E-05 |
| PPP1R13B | 0.018100311 | -0.01332187 | 0.004778441 |
| HOXD11 | 0.011493209 | -0.008546643 | 0.002946566 |
| MYRF | 0.008856891 | -0.006799511 | 0.00205738 |
| ACKR4 | 0.005643728 | -0.004552164 | 0.001091564 |
| ZNF131 | 0.025958365 | -0.016850531 | 0.009107834 |
| GOLGA5 | 0.020077468 | -0.015828699 | 0.004248769 |
| EXOC7 | 0.026522494 | -0.017665844 | 0.00885665 |
| DAPK3 | 0.016983663 | -0.010168562 | 0.006815101 |
| TMEM44 | 0.005370085 | -0.005905596 | -0.000535511 |
| NFIA | 0.030580719 | -0.017250408 | 0.013330311 |
| TGFBR1 | 0.023677314 | -0.023531219 | 0.000146095 |
| HLA-DQA2 | 0.004530841 | -0.004350015 | 0.000180826 |
| PDE4DIP | 0.020515154 | -0.018085855 | 0.0024293 |
| EID2 | 0.020815526 | -0.020526762 | 0.000288763 |
| DGKH | 0.010089603 | -0.007452967 | 0.002636635 |
| C20orf96 | 0.016784236 | -0.016740407 | 4.38291E-05 |
| PNPT1 | 0.023819094 | -0.022541084 | 0.00127801 |
| TAP2 | 0.014373432 | -0.008080522 | 0.00629291 |
| ZNF84 | 0.019350045 | -0.01194632 | 0.007403726 |

| | | | |
|---|---|---|---|
| ZNF737 | 0.021768033 | -0.014939427 | 0.006828606 |
| ACBD4 | 0.015269714 | -0.011718864 | 0.003550851 |
| SDC4 | 0.047069839 | -0.03052163 | 0.016548209 |
| XKR8 | 0.013866714 | -0.010753239 | 0.003113475 |
| IPP | 0.018590078 | -0.01392598 | 0.004664098 |
| MIR4435-2HG | 0.008687954 | -0.008703837 | -1.58827E-05 |
| ANKAR | 0.013489658 | -0.012651021 | 0.000838636 |
| SGF29 | 0.017448296 | -0.011746392 | 0.005701903 |
| CDKN1B | 0.026279185 | -0.023639483 | 0.002639702 |
| FAHD2A | 0.029479927 | -0.019561328 | 0.009918599 |
| TCEAL3 | 0.031030459 | -0.020207475 | 0.010822984 |
| CAV2 | -0.028974479 | 0.057371501 | 0.028397022 |
| AC104237.2 | 0.0071325 | -0.008343937 | -0.001211437 |
| ZNF704 | 0.025644802 | -0.020911459 | 0.004733344 |
| CCDC191 | 0.014092122 | -0.008698759 | 0.005393363 |
| STX6 | 0.022744765 | -0.013810767 | 0.008933999 |
| MIR3936HG | 0.021733739 | -0.022323549 | -0.000589809 |
| ABCB10 | 0.013181618 | -0.011362481 | 0.001819137 |
| MPPE1 | 0.028910091 | -0.027826876 | 0.001083215 |
| GOLM1 | -0.037599734 | 0.064837508 | 0.027237774 |
| AC091544.4 | 0.004272101 | -0.004272101 | -3.55271E-15 |
| DGCR8 | 0.015499153 | -0.011228445 | 0.004270708 |
| MKNK1 | 0.026367549 | -0.042778259 | -0.01641071 |
| PRELID3B | 0.02824009 | -0.017829531 | 0.010410559 |
| B3GNT2 | 0.032536618 | -0.038087889 | -0.005551272 |
| PLBD1-AS1 | 0.009389595 | -0.005556059 | 0.003833536 |
| SPIDR | 0.018854615 | -0.011450406 | 0.007404209 |
| AC003102.1 | 0.010810742 | -0.010819638 | -8.89583E-06 |
| PPM1D | 0.011614106 | -0.00813762 | 0.003476486 |
| NDUFAF7 | 0.018086955 | -0.015616169 | 0.002470787 |
| ADAMTS18 | 0.009784143 | -0.006206957 | 0.003577186 |
| MLST8 | 0.023652355 | -0.017419021 | 0.006233334 |
| KPNA2 | 0.016619635 | -0.014152841 | 0.002466794 |
| C3orf14 | 0.021239637 | -0.017236092 | 0.004003545 |
| PIK3C2B | 0.004808157 | -0.003829469 | 0.000978688 |
| PKP3 | 0.011279142 | -0.015151218 | -0.003872076 |
| CCDC62 | 0.004729953 | -0.003981616 | 0.000748337 |
| SLC23A3 | 0.0067769 | -0.007369141 | -0.000592241 |
| MSH3 | 0.022762274 | -0.031755027 | -0.008992752 |
| TXLNG | 0.018332778 | -0.012838136 | 0.005494643 |
| CSE1L | 0.023202763 | -0.017253773 | 0.00594899 |
| NPAS2 | 0.005144404 | -0.003724863 | 0.001419541 |
| KARS | 0.026976229 | -0.021363143 | 0.005613086 |
| RRAGD | -0.03345778 | 0.051772639 | 0.018314859 |
| MINDY4 | 0.010605491 | -0.009461638 | 0.001143853 |
| PRMT3 | 0.01497116 | -0.013315108 | 0.001656052 |
| METTL2B | 0.021620419 | -0.014832781 | 0.006787638 |

| | | | |
|---|---|---|---|
| DXO | 0.017036099 | -0.018317094 | -0.001280994 |
| TRMT10A | 0.016380587 | -0.00974145 | 0.006639137 |
| COPG2 | 0.014354002 | -0.015352557 | -0.000998556 |
| MAPK3 | 0.026065397 | -0.015777842 | 0.010287555 |
| BTBD2 | 0.021343173 | -0.014762181 | 0.006580992 |
| ANKS1A | 0.025411279 | -0.024591281 | 0.000819999 |
| EIF2B5 | 0.027472376 | -0.019978851 | 0.007493524 |
| IL13RA1 | 0.022124015 | -0.019065076 | 0.003058939 |
| PEX13 | 0.021299879 | -0.018055511 | 0.003244367 |
| AC103702.2 | 0.020194736 | -0.012812481 | 0.007382255 |
| ARHGEF2 | 0.018517818 | -0.015982607 | 0.002535211 |
| POGLUT1 | 0.017838844 | -0.020412935 | -0.002574091 |
| AGGF1 | 0.028740181 | -0.03168113 | -0.002940949 |
| TAF2 | 0.016989789 | -0.010238311 | 0.006751478 |
| ACVR2B | 0.012793063 | -0.008690702 | 0.004102361 |
| STK16 | 0.024655575 | -0.022085426 | 0.00257015 |
| G6PC3 | 0.03068621 | -0.021209392 | 0.009476818 |
| YPEL2 | 0.022579459 | -0.026557633 | -0.003978175 |
| RNF26 | 0.015946807 | -0.012105632 | 0.003841175 |
| UBE2T | 0.015847114 | -0.010606579 | 0.005240535 |
| ATP6V1C1 | 0.026557801 | -0.026635921 | -7.81198E-05 |
| APOL1 | 0.013598843 | -0.014508584 | -0.000909741 |
| STOX1 | 0.016458257 | -0.015072749 | 0.001385508 |
| DPAGT1 | 0.017166401 | -0.019591722 | -0.00242532 |
| TAF11 | 0.024551328 | -0.01972416 | 0.004827168 |
| BBS4 | 0.019291231 | -0.016485084 | 0.002806147 |
| USP1 | 0.024138523 | -0.019913926 | 0.004224597 |
| FAF2 | 0.022743284 | -0.027735573 | -0.004992289 |
| ITGA3 | 0.017132069 | -0.014990089 | 0.00214198 |
| CEP350 | 0.029009305 | -0.028659656 | 0.000349648 |
| ZNF331 | 0.018778477 | -0.017714861 | 0.001063616 |
| PIP4K2C | 0.024383707 | -0.016118344 | 0.008265363 |
| NPB | 0.005182537 | -0.0058967 | -0.000714162 |
| KPNA3 | 0.021928466 | -0.014868274 | 0.007060192 |
| TSTD3 | 0.019206659 | -0.013235578 | 0.005971081 |
| FARP1 | 0.035219521 | -0.032400198 | 0.002819323 |
| CNST | 0.017275495 | -0.020318903 | -0.003043408 |
| KYNU | 0.012394482 | -0.017306245 | -0.004911763 |
| BPHL | 0.026596177 | -0.020915399 | 0.005680778 |
| TASOR2 | 0.023756664 | -0.01621591 | 0.007540754 |
| FKBP15 | 0.015567103 | -0.014428216 | 0.001138887 |
| CBR1 | 0.044006418 | -0.030258844 | 0.013747573 |
| TIGD5 | 0.016988821 | -0.017441999 | -0.000453178 |
| IQCB1 | 0.01930981 | -0.020107207 | -0.000797397 |
| CLCN4 | 0.012509241 | -0.008564232 | 0.003945008 |
| FAAP24 | 0.006219028 | -0.003567399 | 0.00265163 |
| PHF6 | 0.025623464 | -0.015969245 | 0.009654219 |

| | | | |
|---|---|---|---|
| NCMAP-DT | 0.007726796 | -0.005813788 | 0.001913007 |
| FHL1 | 0.005954011 | -0.006444558 | -0.000490547 |
| PHACTR4 | 0.027543904 | -0.024661484 | 0.00288242 |
| MUSTN1 | 0.004621595 | -0.003897771 | 0.000723824 |
| TSC22D1 | 0.039018072 | -0.031483127 | 0.007534945 |
| VARS | 0.016651483 | -0.011700831 | 0.004950653 |
| YWHAH | 0.039917217 | -0.037793044 | 0.002124173 |
| TP53BP2 | 0.016883972 | -0.011947742 | 0.00493623 |
| UMAD1 | 0.022343439 | -0.017421938 | 0.004921501 |
| LRRN2 | 0.024290743 | -0.019201279 | 0.005089464 |
| INO80 | 0.021783468 | -0.034077484 | -0.012294016 |
| CX3CL1 | 0.019042793 | -0.015979217 | 0.003063577 |
| C11orf74 | 0.024931793 | -0.021769246 | 0.003162546 |
| UBR3 | 0.028610319 | -0.021432185 | 0.007178133 |
| SLC25A23 | 0.024979367 | -0.018465948 | 0.006513418 |
| ZNF24 | 0.031180756 | -0.02512146 | 0.006059296 |
| LRIT3 | 0.008707118 | -0.005116755 | 0.003590363 |
| CEPT1 | 0.026726612 | -0.032115602 | -0.00538899 |
| NHSL1 | 0.01444217 | -0.028994491 | -0.014552322 |
| MED8 | 0.023604461 | -0.015202317 | 0.008402144 |
| CNTRL | 0.012638454 | -0.009343492 | 0.003294962 |
| PDE1A | 0.048520361 | 0.037100715 | 0.085621075 |
| PHYHD1 | 0.028252743 | -0.022902739 | 0.005350003 |
| PRKACB | 0.037244054 | 0.02201375 | 0.059257804 |
| SLC25A26 | 0.020598614 | -0.023053095 | -0.002454481 |
| GCFC2 | 0.018487912 | -0.018160309 | 0.000327603 |
| OSBPL2 | 0.024713603 | -0.022165788 | 0.002547815 |
| LYPD6B | 0.017125939 | -0.010152112 | 0.006973827 |
| DHX9 | 0.033533611 | -0.021770724 | 0.011762887 |
| CIAO3 | 0.020700959 | -0.016964594 | 0.003736365 |
| KCNMB4 | 0.007521537 | -0.00766571 | -0.000144174 |
| DELE1 | 0.022646588 | -0.025581211 | -0.002934623 |
| IRX2 | -0.008237183 | 0.00565516 | -0.002582022 |
| SLC35A1 | 0.020589907 | -0.01618144 | 0.004408468 |
| WWTR1 | 0.020021101 | -0.014030082 | 0.005991019 |
| POLR3B | 0.013748187 | -0.015070562 | -0.001322375 |
| TBC1D5 | 0.0257419 | -0.023933229 | 0.001808671 |
| MSH6 | 0.018548628 | -0.024019937 | -0.005471309 |
| ZBTB18 | 0.0201607 | -0.03289775 | -0.012737051 |
| PSMA3-AS1 | 0.030573819 | -0.038138521 | -0.007564702 |
| SAC3D1 | 0.017548433 | -0.010776189 | 0.006772244 |
| LMLN | 0.011968761 | -0.007983808 | 0.003984953 |
| MTMR10 | 0.022299687 | -0.026216409 | -0.003916722 |
| GCAT | 0.021295239 | -0.013665224 | 0.007630015 |
| UBE4A | 0.025182343 | -0.016366005 | 0.008816338 |
| NFKBIL1 | 0.020858119 | -0.01801237 | 0.002845749 |
| ATXN7L3B | 0.032410284 | -0.023690488 | 0.008719796 |

| | | | |
|---|---|---|---|
| EGR1 | -0.034489317 | 0.028645923 | -0.005843394 |
| SAMD12 | 0.02316604 | -0.01855762 | 0.004608419 |
| BCL2L2 | 0.020952143 | -0.013006671 | 0.007945472 |
| NEIL1 | 0.015079642 | -0.021333712 | -0.00625407 |
| NTAN1 | 0.024139106 | -0.027352136 | -0.00321303 |
| PPP2R5B | 0.014156272 | -0.008790771 | 0.005365502 |
| PIGS | 0.024184577 | -0.027266381 | -0.003081804 |
| ZNF595 | 0.01353226 | -0.011629055 | 0.001903204 |
| ZMYM2 | 0.030683337 | -0.027613582 | 0.003069755 |
| TST | 0.033474361 | -0.024319629 | 0.009154731 |
| CCNY | 0.027622617 | -0.032360871 | -0.004738254 |
| EIF1AY | 0.021680851 | -0.04653413 | -0.024853279 |
| ZNF540 | 0.013313857 | -0.008319908 | 0.004993948 |
| PDLIM1 | 0.011449479 | -0.017331292 | -0.005881813 |
| SLC30A6 | 0.017153576 | -0.011514486 | 0.00563909 |
| CHD1 | 0.02312765 | -0.017656972 | 0.005470679 |
| MTRF1 | 0.0150085 | -0.012011539 | 0.002996961 |
| MYLK4 | 0.004129363 | -0.003662419 | 0.000466945 |
| NUDT16 | -0.03636276 | 0.048314828 | 0.011952068 |
| RRP9 | 0.017427647 | -0.012568176 | 0.004859472 |
| TFF2 | 0.006508392 | -0.004323168 | 0.002185225 |
| ZNF257 | 0.008438185 | -0.006139827 | 0.002298358 |
| RELL1 | 0.009221037 | -0.010521 | -0.001299963 |
| SLC22A2 | -0.015906139 | 0.021956337 | 0.006050197 |
| CD36 | 0.006809312 | -0.008509781 | -0.001700468 |
| LINC02591 | 0.015507751 | -0.012789228 | 0.002718523 |
| USF3 | 0.026089182 | -0.018650035 | 0.007439147 |
| DMGDH | 0.013739924 | -0.010649014 | 0.00309091 |
| VEZT | 0.025466464 | -0.019586585 | 0.005879879 |
| IFITM2 | 0.033372079 | 0.043209258 | 0.076581336 |
| ZC3H4 | 0.020833586 | -0.017212533 | 0.003621053 |
| PHKG2 | 0.023824168 | -0.018648491 | 0.005175677 |
| RINL | 0.016713109 | -0.012109787 | 0.004603322 |
| PPP4R3A | 0.026481291 | -0.027622168 | -0.001140877 |
| XYLT2 | 0.018869231 | -0.020571531 | -0.0017023 |
| SLC25A20 | 0.012561268 | -0.00752687 | 0.005034399 |
| MYRIP | 0.0122809 | -0.010854378 | 0.001426523 |
| CNOT1 | 0.029755472 | -0.034357746 | -0.004602274 |
| AL162231.1 | 0.016607426 | -0.015283948 | 0.001323478 |
| ZBTB17 | 0.014797538 | -0.010315605 | 0.004481933 |
| TRAFD1 | 0.013465183 | -0.00846314 | 0.005002043 |
| CALCR | 0.009212509 | -0.008969035 | 0.000243473 |
| KLHL23 | -0.020736865 | 0.014656244 | -0.006080621 |
| MX1 | 0.017377052 | -0.020643704 | -0.003266652 |
| ATG4C | 0.017216929 | -0.012415869 | 0.00480106 |
| LINC01863 | 0.004416783 | -0.003944088 | 0.000472694 |
| KAT2A | 0.019880272 | -0.011863235 | 0.008017038 |

| | | | |
|---|---|---|---|
| STIL | 0.004401245 | -0.003907253 | 0.000493991 |
| SNX10 | -0.019992498 | 0.012180131 | -0.007812366 |
| FBXL20 | 0.020234759 | -0.017397269 | 0.00283749 |
| C1RL | 0.011099991 | -0.011214834 | -0.000114843 |
| POU2F1 | 0.017262932 | -0.016728434 | 0.000534497 |
| AC006115.2 | 0.007510861 | -0.006744601 | 0.00076626 |
| RNF103 | 0.018609354 | -0.013564175 | 0.005045179 |
| EXOC6 | 0.020833143 | -0.015598996 | 0.005234147 |
| KDM7A | 0.014577145 | -0.009036203 | 0.005540942 |
| ZCWPW1 | 0.01026887 | -0.010651465 | -0.000382595 |
| RNF114 | 0.028747347 | -0.017814621 | 0.010932727 |
| PSMD12 | 0.030746063 | -0.035526787 | -0.004780724 |
| ZCCHC2 | 0.015217361 | -0.015431894 | -0.000214533 |
| HAUS3 | 0.015008501 | -0.0124775 | 0.002531001 |
| NEO1 | 0.021339267 | -0.023288837 | -0.00194957 |
| LINC01535 | 0.011694557 | -0.010279991 | 0.001414566 |
| SPEG | 0.004463134 | -0.004233145 | 0.000229988 |
| SUMO4 | 0.009631304 | -0.0065392 | 0.003092104 |
| ZC3H18 | 0.020362651 | -0.01848125 | 0.001881401 |
| CCDC137 | 0.021595712 | -0.019901347 | 0.001694365 |
| MARC2 | 0.02528269 | -0.019373477 | 0.005909213 |
| TPRG1-AS1 | 0.005110489 | -0.003926518 | 0.001183971 |
| ANKRD37 | 0.015198062 | -0.023349989 | -0.008151927 |
| SAT2 | -0.035288101 | 0.030561222 | -0.00472688 |
| PIP4P2 | 0.018890402 | -0.025232836 | -0.006342433 |
| MACROD2 | 0.017218912 | -0.01269188 | 0.004527032 |
| APTX | 0.02344644 | -0.014316406 | 0.009130034 |
| WDR70 | 0.019781721 | -0.023164443 | -0.003382721 |
| FAAH | 0.019962117 | -0.022459262 | -0.002497146 |
| TMEM222 | 0.024155403 | -0.017414977 | 0.006740426 |
| ODAM | 0.007476456 | -0.007056758 | 0.000419698 |
| ALS2CL | 0.021156965 | -0.018846485 | 0.00231048 |
| HES1 | 0.020328049 | -0.03833634 | -0.018008291 |
| LINC01725 | 0.008729706 | -0.005741421 | 0.002988285 |
| P3H4 | 0.014175341 | -0.008928754 | 0.005246587 |
| RPS29 | 0.046884698 | -0.047741747 | -0.000857049 |
| RALGAPB | 0.022616716 | -0.026113299 | -0.003496583 |
| PLXNA2 | -0.019590917 | 0.021754004 | 0.002163087 |
| NBPF1 | 0.013996935 | -0.01157876 | 0.002418174 |
| SLC35A5 | 0.019340847 | -0.012993424 | 0.006347423 |
| EPC1 | 0.027805935 | -0.026572263 | 0.001233672 |
| ARFGEF3 | 0.013821271 | -0.018060269 | -0.004238997 |
| BCR | 0.022427331 | -0.014813424 | 0.007613908 |
| STAT6 | 0.027607476 | -0.021841205 | 0.005766271 |
| C6orf163 | 0.005446393 | -0.003561704 | 0.001884689 |
| CFAP44 | 0.010133892 | -0.009131107 | 0.001002785 |
| COBL | -0.015194323 | 0.010674838 | -0.004519485 |

| | | | |
|---|---|---|---|
| EHBP1 | 0.016371795 | -0.010946981 | 0.005424814 |
| CTDSPL2 | 0.019824033 | -0.020141581 | -0.000317548 |
| DSG2 | 0.052639593 | 0.036545542 | 0.089185135 |
| MAST4 | -0.015393651 | 0.01356789 | -0.001825761 |
| MRPL2 | 0.033835173 | -0.020804227 | 0.013030946 |
| STXBP5L | 0.003951486 | -0.003951486 | -3.55271E-15 |
| CHM | 0.022869223 | -0.018538756 | 0.004330467 |
| RINT1 | 0.017328663 | -0.013855454 | 0.003473209 |
| TMPRSS12 | 0.004337359 | -0.003711576 | 0.000625783 |
| COX16 | 0.039004685 | -0.023979685 | 0.015025 |
| ILK | 0.027379877 | -0.023650659 | 0.003729218 |
| EIF2AK1 | 0.031646565 | -0.025330057 | 0.006316508 |
| AMT | 0.02505484 | -0.016477762 | 0.008577078 |
| ZGLP1 | 0.007514684 | -0.00448121 | 0.003033474 |
| ANG | 0.007154519 | -0.006520951 | 0.000633568 |
| RGS5 | 0.008994618 | -0.00970208 | -0.000707462 |
| CDKL5 | 0.012211128 | -0.009175936 | 0.003035192 |
| FOS | -0.065208382 | 0.061730156 | -0.003478226 |
| DHX30 | 0.030293125 | -0.018801511 | 0.011491615 |
| AL139824.1 | 0.00417037 | -0.00417037 | -5.32907E-15 |
| RNF19A | 0.025247908 | -0.025797641 | -0.000549733 |
| YAP1 | 0.025550471 | -0.033901803 | -0.008351331 |
| PABPC1L | 0.013733489 | -0.016978664 | -0.003245175 |
| MINCR | 0.021705001 | -0.014383771 | 0.007321231 |
| SBNO2 | 0.013445613 | -0.012849398 | 0.000596215 |
| ATXN3 | 0.017494955 | -0.01057044 | 0.006924515 |
| RESF1 | 0.017716107 | -0.017524724 | 0.000191383 |
| GTF2A1 | 0.022535341 | -0.017256532 | 0.005278808 |
| KCTD14 | 0.013253881 | -0.009200858 | 0.004053023 |
| MT-CO3 | -0.042969388 | 0.030641231 | -0.012328157 |
| TANK | 0.024182766 | -0.023820426 | 0.00036234 |
| MITD1 | 0.021764213 | -0.019640965 | 0.002123247 |
| ELP3 | 0.025038178 | -0.019423275 | 0.005614903 |
| SERTAD4 | 0.011369196 | -0.01118023 | 0.000188965 |
| TAPBPL | 0.019257148 | -0.017225957 | 0.00203119 |
| IRF2BP1 | 0.018158554 | -0.011506434 | 0.00665212 |
| DNAH10OS | 0.007655199 | -0.006168164 | 0.001487036 |
| C3orf52 | 0.008037066 | -0.008910585 | -0.000873519 |
| DNAJC13 | 0.02094433 | -0.012638528 | 0.008305803 |
| CEP126 | 0.006602623 | -0.006297132 | 0.000305491 |
| LAD1 | 0.016407631 | -0.015650864 | 0.000756767 |
| WDR53 | 0.010264447 | -0.007829022 | 0.002435425 |
| PPM1M | 0.011907667 | -0.010466386 | 0.001441282 |
| STAG1 | 0.022252064 | -0.024654357 | -0.002402293 |
| FOCAD | 0.015960301 | -0.019311838 | -0.003351538 |
| PSMD3 | 0.026110018 | -0.02436159 | 0.001748429 |
| RBM19 | 0.016050407 | -0.01247892 | 0.003571486 |

| | | | |
|---|---|---|---|
| FAM174B | 0.01616503 | -0.023984917 | -0.007819887 |
| LETM1 | 0.023707336 | -0.018893536 | 0.0048138 |
| OAS1 | 0.009968411 | -0.009874349 | 9.40617E-05 |
| FBXO38 | 0.015390055 | -0.017769644 | -0.002379589 |
| SESN3 | 0.013278903 | -0.014378252 | -0.001099349 |
| ETV6 | 0.013429973 | -0.012657461 | 0.000772512 |
| LRRC57 | 0.019937089 | -0.012322951 | 0.007614138 |
| AC083805.1 | 0.004370503 | -0.003596009 | 0.000774494 |
| SPINDOC | 0.010687162 | -0.007999872 | 0.00268729 |
| NMRAL1 | 0.024460237 | -0.014913963 | 0.009546274 |
| VIRMA | 0.019986598 | -0.021393545 | -0.001406947 |
| VPS26C | 0.01804274 | -0.012206484 | 0.005836255 |
| POLL | 0.015801846 | -0.010598046 | 0.0052038 |
| CLPB | 0.02272735 | -0.022420378 | 0.000306972 |
| PWWP2B | 0.020384708 | -0.016166201 | 0.004218507 |
| FAM160B1 | 0.013315441 | -0.01264551 | 0.000669931 |
| DDX27 | 0.023228133 | -0.019842403 | 0.00338573 |
| AGO3 | 0.021794976 | -0.017885756 | 0.00390922 |
| C12orf81 | 0.005328787 | -0.003870758 | 0.00145803 |
| SF1 | 0.038814291 | -0.051601711 | -0.01278742 |
| PAFAH2 | 0.015828644 | -0.014521862 | 0.001306782 |
| P4HA2 | 0.012329916 | -0.017745605 | -0.005415689 |
| TNFAIP1 | 0.015668013 | -0.011641966 | 0.004026046 |
| DDX41 | 0.019231183 | -0.012065483 | 0.0071657 |
| SLMAP | 0.020817322 | -0.017301571 | 0.003515751 |
| ANKRD30A | 0.0071061 | -0.006126494 | 0.000979605 |
| UFD1 | 0.029751618 | -0.023531313 | 0.006220305 |
| IPO7 | 0.03098487 | -0.022727189 | 0.008257681 |
| TBC1D9B | 0.021675603 | -0.014414243 | 0.007261361 |
| RPRD2 | 0.022733738 | -0.020756125 | 0.001977614 |
| SUGT1 | 0.031486101 | -0.031169316 | 0.000316784 |
| AC096642.1 | 0.004671611 | -0.003829347 | 0.000842264 |
| SCAP | 0.025553958 | -0.025602979 | -4.90203E-05 |
| SERHL2 | 0.009260199 | -0.009001491 | 0.000258708 |
| TFAP4 | 0.01315107 | -0.016199003 | -0.003047933 |
| VAMP1 | 0.007863267 | -0.005306391 | 0.002556877 |
| SHISA9 | 0.005392396 | -0.004082163 | 0.001310233 |
| DST | -0.032920106 | 0.023153444 | -0.009766663 |
| ZNF609 | 0.013022844 | -0.008397025 | 0.00462582 |
| SLC34A3 | 0.004603768 | -0.004105714 | 0.000498054 |
| PAQR5 | 0.028320595 | -0.017490902 | 0.010829693 |
| MFSD11 | 0.018414668 | -0.02051301 | -0.002098341 |

**Supplemental Table 3**: Pathways enriched by genes enhanced or suppressed with SGLT2 inhibitor across all nephron segments.

**Pathways enriched by genes suppressed with SGLT2 inhibitor in proximal tubular cells**

| | Term | P value | Adjusted P-value | Odds Ratio | Combined Score | Genes |
|---|---|---|---|---|---|---|
| 1 | Metabolism Homo sapiens R-HSA-1430728 | 1.97E-35 | 2.13E-32 | 2.96E+00 | 2.36E+02 | DMGDH;CERK;CPNE6;ECI2;ENO2;GLS;AQP1;PSMD8;PSMD9;PSMD2;RPL38;ACAD11;PDK2;GAMT;ACOT8;SLC6A12;CMBL;UPB1;SULT2B1;FDX1;PSME1;UQCRC1;SUCLG1;CES2;GCDH;MAOA;SLC5A2;RPS4Y1;SLC5A3;OAZ1;ACMSD;LDHA;APOM;PRKAR2B;AFMID;HMOX1;APOE;PCK1;DECR2;HAO2;GSTT2B;HMOX2;DECR1;GSTM4;CTSA;GSTM3;SURF1;GLYATL1;CYP4F2;MTHFS;NR1D1;GNG11;DERA;RPS28;SLC6A8;RPS27;ADI1;RPS29;CYP2S1;CALM1;RPS21;COX7B;QPRT;AKR1B1;ECSIT;COX7C;KHK;MED11;AUH;ACADM;TK1;COX8A;CBR1;ARG2;TPI1;ELOVL4;GSTO1;PGAM1;AKR1A1;PTGR1;APRT;GCHFR;ITPKB;BDH2;PLCB4;IVD;ADSSL1;PLIN2;ALDOC;TKT;MGLL;ECHS1;AHCY;UQCR11;GDPD3;UQCR10;COX5A;PTGS1;PSMB3;UCP2;AOC1;MDH1;PKLR;AKR1C1;NR1H4;BBOX1;G0S2;FMO1;FAH;AGMAT;FABP1;PSMC4;ETNK2;NDUFAB1;RORA;CYP46A1;SAT1;CNDP2;AKR7A2;AKR7A3;NAGLU;ENPP1;ACAA1;DIO1;SARS;PIPOX;ACOT13;SDHB;COX6B1;FOLH1;DPEP1;ABCG1;PFKFB3;AK2;ARL2;GMPR;GLYAT;NPAS2;UBB;PGK1;COX11;COX14;CYC1;NDUFV3;NDUFV2;PLTP;IP6K3;ESRRA;MGAM;GOT1;AGXT2;UROS;GOT2;ACSF2;GGCT;QDPR;GRHPR;DAO;UQCRQ;SLCO1A2;CRYL1;ACO2;GALK1;APIP;SLC2A1;SLC2A3;ETFB;ACSM2B;FT [illegible] |
| 2 | Respiratory electron transport Homo sapiens R-HSA-611105 | 1.16E-18 | 6.27E-16 | 1.06E+01 | 4.38E+02 | COX7B;NDUFB3;NDUFB2;NDUFB1;UQCR11;ETFB;UQCR10;ECSIT;COX7C;COX5A;UQCRFS1;COX11;COX14;NDUFV3;CYC1;NDUFV2;COX8A;NDUFA8;SURF1;NDUFA3;NDUFA2;NDUFA1;NDUFC1;SDHB;COX6B1;NDUFS7;UQCRQ;NDUFS4;NDUFAB1;UQCRC1;NDUFS2 |
| 3 | The citric acid (TCA) cycle and respiratory electron transport Homo sapiens R-HSA-1428517 | 2.29E-18 | 8.24E-16 | 6.94E+00 | 2.82E+02 | COX7B;NDUFB3;NDUFB2;NDUFB1;UQCR11;ETFB;UQCR10;PDHB;ECSIT;COX7C;COX5A;LDHA;UCP2;COX11;UQCRFS1;COX14;NDUFV3;CYC1;NDUFV2;PDK2;COX8A;NDUFA8;SURF1;IDH3G;NDUFA3;NDUFA2;NDUFA1;NDUFC1;SDHB;COX6B1;SUCLA2;NDUFS7;UQCRQ;NDUFS4;NDUFAB1;UQCRC1;NDUFS2;SUCLG1;ACO2;HAGH |

| | | | | | | |
|---|---|---|---|---|---|---|
| 4 | Respiratory electron transport, ATP synthesis by chemiosmotic coupling, and heat production by uncoupling proteins. Homo sapiens R-HSA-163200 | 1.46E-16 | 3.95E-14 | 8.10E+00 | 2.95E+02 | COX7B;NDUFB3;NDUFB2;NDUFB1;UQCR11;ETFB;UQCR10;ECSIT;COX7C;COX5A;UCP2;COX11;UQCRFS1;COX14;NDUFV3;CYC1;NDUFV2;COX8A;NDUFA8;SURF1;NDUFA3;NDUFA2;NDUFA1;NDUFC1;SDHB;COX6B1;NDUFS7;UQCRQ;NDUFS4;NDUFAB1;UQCRC1;NDUFS2 |
| 5 | Metabolism of amino acids and derivatives Homo sapiens R-HSA-71291 | 1.50E-11 | 3.25E-09 | 3.36E+00 | 8.37E+01 | APIP;SAT1;FTCD;GLS;CNDP2;PSMD8;PSMD9;AUH;PSMD2;RPL38;GAMT;ARG2;DIO1;SLC6A12;SARS;PIPOX;FOLH1;TST;IVD;ASPG;PSME1;SLC25A10;GCDH;AHCY;PDHB;RPS4Y1;OAZ1;ACMSD;PSMB3;AFMID;PCBD1;GOT1;AGXT2;GOT2;BBOX1;FAH;ASS1;AGMAT;QDPR;GRHPR;RPS28;RPS27;SLC6A8;DAO;RPS29;ADI1;PSMC4;NDUFAB1;RPS21 |
| 6 | Glucose metabolism Homo sapiens R-HSA-70326 | 1.92E-10 | 3.45E-08 | 6.98E+00 | 1.56E+02 | TPI1;PFKFB3;PKLR;GOT1;MDH1;PGAM1;GOT2;ENO2;HK2;PFKL;GNPDA1;UBB;PGK1;ALDOC;SLC25A10;PCK1;CALM1;FBP1;SLC25A11;GAPDH;PGM1 |
| 7 | Gluconeogenesis Homo sapiens R-HSA-70263 | 1.68E-09 | 2.59E-07 | 1.31E+01 | 2.65E+02 | TPI1;GOT1;MDH1;PGAM1;GOT2;ENO2;PGK1;ALDOC;SLC25A10;PCK1;SLC25A11;FBP1;GAPDH |
| 8 | Complex I biogenesis Homo sapiens R-HSA-6799198 | 9.10E-09 | 1.23E-06 | 8.47E+00 | 1.57E+02 | NDUFA8;NDUFA3;NDUFB3;NDUFA2;NDUFB2;NDUFA1;NDUFB1;NDUFC1;ECSIT;NDUFS7;NDUFS4;NDUFAB1;NDUFS2;NDUFV3;NDUFV2 |
| 9 | Biological oxidations Homo sapiens R-HSA-211859 | 2.33E-07 | 2.80E-05 | 3.30E+00 | 5.04E+01 | AHCY;MAOA;GLYAT;CYP46A1;BPHL;ACSM2B;PTGS1;CNDP2;AKR7A2;AKR7A3;ALDH1B1;GSTT2B;GSTM4;GSTM3;AOC1;GLYATL1;GSTO1;CYP4F2;AKR1A1;NR1H4;FMO1;ABHD14B;CMBL;SULT2B1;GGCT;CYP2S1;FDX1;DPEP1;CES2 |
| 10 | Metabolism of carbohydrates Homo sapiens R-HSA-71387 | 2.59E-07 | 2.80E-05 | 2.84E+00 | 4.30E+01 | PFKFB3;SLC2A1;AKR1B1;SLC2A3;ENO2;SLC5A2;SLC5A3;HK2;KHK;NAGLU;UBB;PGK1;PCK1;SLC37A4;PGM1;MGAM;TPI1;PKLR;MDH1;GOT1;PGAM1;GOT2;B3GAT1;AKR1A1;DERA;PFKL;GNPDA1;CRYL1;ALDOC;SLC25A10;CALM1;TKT;FBP1;SLC25A11;GAPDH;GALK1 |
| 11 | Metabolism of polyamines Homo sapiens R-HSA-351202 | 7.46E-07 | 7.33E-05 | 4.87E+00 | 6.87E+01 | GAMT;ARG2;GOT1;APIP;SLC6A12;SAT1;OAZ1;ASS1;AGMAT;PSMD8;PSMD9;SLC6A8;PSMB3;PSMC4;ADI1;PSMD2;PSME1 |

| | | | | | | |
|---|---|---|---|---|---|---|
| 12 | Transmembrane transport of small molecules Homo sapiens R-HSA-382551 | 2.08E-06 | 1.87E-04 | 2.07E+00 | 2.71E+01 | SLC2A1;SLC2A3;ABCA12;AQP2;AQP3;HK2;AQP1;SLC22A12;FTH1;NIPAL3;SLC39A5;SLC37A4;ATP6V1E1;SLC22A11;SLC34A3;SLC13A2;ATP6V1G1;SLC2A13;SLC34A1;SLC6A12;ATP1B3;ATP1B1;SLC9A3;SLC7A7;SLCO4A1;SLC7A9;SLC25A10;AVP;ATP6V0D1;ABCG1;FTL;SLC47A2;SLC22A6;SLC22A5;SLC5A2;SLC5A3;UBB;PRKAR2B;GNG4;SLC17A5;HMOX1;ATP6V0E2;SLC38A2;HMOX2;ATP6V1F;SLC14A2;ABCA4;GNG11;TPCN1;SLC2A9;CLCN7;SLC4A7;SLC6A6;FXYD4;FXYD2;SLCO1A2;CALM1 |
| 13 | Glycolysis Homo sapiens R-HSA-70171 | 2.26E-06 | 1.88E-04 | 8.69E+00 | 1.13E+02 | PFKL;PFKFB3;TPI1;PKLR;PGAM1;PGK1;ALDOC;ENO2;GAPDH;HK2 |
| 14 | TP53 Regulates Metabolic Genes Homo sapiens R-HSA-5628897 | 3.64E-06 | 2.81E-04 | 4.51E+00 | 5.65E+01 | COX8A;COX7B;GPX2;SURF1;COX7C;COX5A;GLS;COX6B1;PRDX1;SESN2;DDIT4;COX11;COX14;LAMTOR2;LAMTOR1;LAMTOR5 |
| 15 | SLC-mediated transmembrane transport Homo sapiens R-HSA-425407 | 1.34E-05 | 9.63E-04 | 2.52E+00 | 2.83E+01 | SLC47A2;SLC22A6;SLC22A5;SLC2A1;SLC2A3;SLC5A2;SLC5A3;HK2;SLC22A12;SLC17A5;SLC39A5;SLC37A4;SLC38A2;SLC22A11;SLC34A3;SLC13A2;SLC14A2;SLC34A1;SLC2A13;SLC6A12;SLC2A9;SLC4A7;SLC6A6;SLC9A3;SLC7A7;SLCO4A1;SLC7A9;SLCO1A2;SLC25A10;AVP;CALM1 |
| 16 | Transport of glucose and other sugars, bile salts and organic acids, metal ions and amine compounds Homo sapiens R-HSA-425366 | 4.10E-05 | 2.77E-03 | 3.61E+00 | 3.64E+01 | SLC47A2;SLC13A2;SLC22A6;SLC14A2;SLC22A5;SLC2A13;SLC2A1;SLC6A12;SLC2A3;SLC5A2;SLC5A3;SLC2A9;SLC22A12;SLC6A6;SLC39A5;SLC22A11 |
| 17 | Mitochondrial translation Homo sapiens R-HSA-5368287 | 1.49E-04 | 9.49E-03 | 3.52E+00 | 3.10E+01 | TSFM;MRPS15;MRPS26;MRPS25;MRPS36;MRPL19;MRPS34;MRPL37;MRPL48;MTIF3;MRPL34;MRPL12;MRPS6;MRPL46 |
| 18 | Sulfur amino acid metabolism Homo sapiens R-HSA-1614635 | 1.69E-04 | 1.02E-02 | 7.41E+00 | 6.43E+01 | AHCY;GOT1;TST;ADI1;APIP;SLC25A10;CNDP2 |
| 19 | Iron uptake and transport Homo sapiens R-HSA-917937 | 2.33E-04 | 1.33E-02 | 5.05E+00 | 4.22E+01 | ATP6V1G1;FTH1;HMOX1;ATP6V0E2;ATP6V1E1;ATP6V0D1;HMOX2;ATP6V1F;FTL |
| 20 | Mitochondrial translation initiation Homo sapiens R-HSA-5368286 | 2.68E-04 | 1.38E-02 | 3.50E+00 | 2.88E+01 | MRPS15;MRPS26;MRPS25;MRPS36;MRPL19;MRPS34;MRPL37;MRPL48;MTIF3;MRPL34;MRPL12;MRPS6;MRPL46 |

| | | | | | | |
|---|---|---|---|---|---|---|
| 21 | Mitochondrial translation elongation Homo sapiens R-HSA-5389840 | 2.68E-04 | 1.38E-02 | 3.50E+00 | 2.88E+01 | TSFM;MRPS15;MRPS26;MRPS25;MRPS36;MRPL19;MRPS34;MRPL37;MRPL48;MRPL34;MRPL12;MRPS6;MRPL46 |
| 22 | Mitochondrial protein import Homo sapiens R-HSA-1268020 | 3.07E-04 | 1.51E-02 | 4.34E+00 | 3.51E+01 | GRPEL1;IDH3G;COX17;LDHD;TIMM13;HSCB;TIMM10;CYC1;ACO2;TOMM5 |
| 23 | Vitamin C (ascorbate) metabolism Homo sapiens R-HSA-196836 | 3.74E-04 | 1.76E-02 | 1.90E+01 | 1.50E+02 | CYB5A;GSTO1;SLC2A1;SLC2A3 |
| 24 | Metabolism of lipids and lipoproteins Homo sapiens R-HSA-556833 | 4.89E-04 | 2.20E-02 | 1.69E+00 | 1.29E+01 | DMGDH;CERK;CPNE6;ECI2;AKR1B1;RORA;CYP46A1;MED11;ACADM;ACAD11;ACAA1;CBR1;ACOT8;GPX2;ELOVL4;ACOT13;PLA2G4A;PTGR1;BDH2;ACOX2;FDX1;PCCB;DPEP1;PLIN2;MGLL;ABCG1;ECHS1;MVK;HPGD;HSD17B14;GDPD3;NPAS2;PTGS1;HMGCL;APOE;DECR2;HAO2;PLTP;DECR1;ESRRA;CTSA;CYP4F2;AKR1C1;NR1H4;G0S2;APOC3;NR1D1;ACSF2;FABP1;ETNK2;APOC2;SLCO1A2;LPIN2 |
| 25 | Pyruvate metabolism and Citric Acid (TCA) cycle Homo sapiens R-HSA-71406 | 5.54E-04 | 2.40E-02 | 4.40E+00 | 3.30E+01 | LDHA;SUCLA2;IDH3G;SUCLG1;PDHB;HAGH;ACO2;SDHB;PDK2 |
| 26 | Phase 1 - Functionalization of compounds Homo sapiens R-HSA-211945 | 6.61E-04 | 2.75E-02 | 3.14E+00 | 2.30E+01 | AOC1;MAOA;CYP4F2;NR1H4;FMO1;CMBL;CYP46A1;BPHL;PTGS1;ALDH1B1;CYP2S1;FDX1;CES2 |
| 27 | Histidine, lysine, phenylalanine, tyrosine, proline and tryptophan catabolism Homo sapiens R-HSA-6788656 | 8.48E-04 | 3.40E-02 | 4.62E+00 | 3.27E+01 | QDPR;GCDH;AFMID;PIPOX;PCBD1;FAH;FTCD;ACMSD |
| 28 | Mitochondrial translation termination Homo sapiens R-HSA-5419276 | 9.54E-04 | 3.68E-02 | 3.18E+00 | 2.21E+01 | MRPS15;MRPS26;MRPS25;MRPS36;MRPL19;MRPS34;MRPL37;MRPL48;MRPL34;MRPL12;MRPS6;MRPL46 |
| 29 | Organic anion transport Homo sapiens R-HSA-561048 | 1.17E-03 | 4.22E-02 | 2.85E+01 | 1.92E+02 | SLC22A12;SLC22A6;SLC22A11 |
| 30 | Mitochondrial Fatty Acid Beta-Oxidation Homo sapiens R-HSA-77289 | 1.17E-03 | 4.22E-02 | 7.93E+00 | 5.35E+01 | ECHS1;PCCB;ACADM;ACAD11;DECR1 |

| | | | | | | |
|---|---|---|---|---|---|---|
| 31 | Fatty acid, triacylglycerol, and ketone body metabolism Homo sapiens R-HSA-535734 | 1.38E-03 | 4.82E-02 | 2.16E+00 | 1.42E+01 | ESRRA;ACOT8;ECHS1;ELOVL4;NR1H4;G0S2;ACOT13;RORA;NR1D1;NPAS2;ACSF2;FABP1;MED11;HMGCL;BDH2;PCCB;PLIN2;ACADM;ACAD11;DECR2;LPIN2;DECR1 |
| 32 | Phase II conjugation Homo sapiens R-HSA-156580 | 1.46E-03 | 4.92E-02 | 2.85E+00 | 1.86E+01 | GSTM4;GSTM3;GLYATL1;AHCY;GSTO1;AKR1A1;ABHD14B;GLYAT;ACSM2B;CNDP2;SULT2B1;GGCT;GSTT2B |
| 33 | HDL-mediated lipid transport Homo sapiens R-HSA-194223 | 2.03E-03 | 6.44E-02 | 6.79E+00 | 4.21E+01 | APOC2;APOC3;APOE;PLTP;ABCG1 |
| 34 | Citric acid cycle (TCA cycle) Homo sapiens R-HSA-71403 | 2.03E-03 | 6.44E-02 | 6.79E+00 | 4.21E+01 | SUCLA2;IDH3G;SUCLG1;ACO2;SDHB |
| 35 | Methionine salvage pathway Homo sapiens R-HSA-1237112 | 2.24E-03 | 6.74E-02 | 1.90E+01 | 1.16E+02 | GOT1;ADI1;APIP |
| 36 | Conjugation of benzoate with glycine Homo sapiens R-HSA-177135 | 2.24E-03 | 6.74E-02 | 1.90E+01 | 1.16E+02 | GLYATL1;GLYAT;ACSM2B |
| 37 | Glutathione conjugation Homo sapiens R-HSA-156590 | 2.52E-03 | 7.36E-02 | 4.30E+00 | 2.57E+01 | GSTM4;GGCT;GSTM3;GSTO1;AKR1A1;GSTT2B;CNDP2 |
| 38 | Autodegradation of Cdh1 by Cdh1:APC/C Homo sapiens R-HSA-174084 | 3.59E-03 | 1.01E-01 | 3.24E+00 | 1.82E+01 | PSMD8;PSMD9;UBB;PSMB3;PSMC4;CDC26;PSMD2;PSME1;ANAPC11 |
| 39 | Nuclear Receptor transcription pathway Homo sapiens R-HSA-383280 | 3.63E-03 | 1.01E-01 | 3.54E+00 | 1.99E+01 | ESRRA;HNF4A;RARA;NR1H4;RORA;NR1D1;ESR2;NR2F6 |
| 40 | Inositol transporters Homo sapiens R-HSA-429593 | 3.78E-03 | 1.02E-01 | 1.42E+01 | 7.95E+01 | SLC2A13;SLC5A2;SLC5A3 |
| 41 | Transport of inorganic cations/anions and amino acids/oligopeptides Homo sapiens R-HSA-425393 | 3.94E-03 | 1.04E-01 | 2.63E+00 | 1.46E+01 | SLC34A3;SLC4A7;SLC9A3;SLC6A6;SLC7A7;SLC7A9;SLC34A1;SLC17A5;SLC6A12;SLC25A10;CALM1;SLC38A2 |
| 42 | Organic cation/anion/zwitterion transport Homo sapiens R-HSA-549132 | 4.20E-03 | 1.06E-01 | 7.60E+00 | 4.16E+01 | SLC22A12;SLC22A6;SLC22A5;SLC22A11 |

| | | | | | | |
|---|---|---|---|---|---|---|
| 43 | Synthesis of Prostaglandins (PG) and Thromboxanes (TX) Homo sapiens R-HSA-2162123 | 4.20E-03 | 1.06E-01 | 7.60E+00 | 4.16E+01 | CBR1;HPGD;PTGR1;PTGS1 |
| 44 | BMAL1:CLOCK,NPAS2 activates circadian gene expression Homo sapiens R-HSA-1368108 | 4.52E-03 | 1.11E-01 | 3.81E+00 | 2.06E+01 | DBP;BHLHE40;RORA;NR1D1;AVP;HIF1A;NPAS2 |
| 45 | Arachidonic acid metabolism Homo sapiens R-HSA-2142753 | 4.63E-03 | 1.11E-01 | 3.39E+00 | 1.82E+01 | CBR1;GPX2;HPGD;CYP4F2;PLA2G4A;DPEP1;PTGR1;PTGS1 |
| 46 | Vasopressin regulates renal water homeostasis via Aquaporins Homo sapiens R-HSA-432040 | 5.18E-03 | 1.17E-01 | 3.70E+00 | 1.95E+01 | GNG4;PRKAR2B;AVP;AQP2;AQP3;GNG11;AQP1 |
| 47 | Detoxification of Reactive Oxygen Species Homo sapiens R-HSA-3299685 | 5.42E-03 | 1.17E-01 | 4.23E+00 | 2.21E+01 | GPX2;PRDX1;CAT;CYBA;ATOX1;SOD1 |
| 48 | APC/C:Cdc20 mediated degradation of Securin Homo sapiens R-HSA-174154 | 5.48E-03 | 1.17E-01 | 3.01E+00 | 1.57E+01 | PSMD8;PSMD9;UBB;PSMB3;PSMC4;CDC26;PSMD2;PSME1;ANAPC11 |
| 49 | WNT5A-dependent internalization of FZD4 Homo sapiens R-HSA-5099900 | 5.50E-03 | 1.17E-01 | 6.91E+00 | 3.60E+01 | PRKCB;CLTB;AP2S1;AP2A1 |
| 50 | PCP/CE pathway Homo sapiens R-HSA-4086400 | 5.76E-03 | 1.17E-01 | 2.62E+00 | 1.35E+01 | PSMD8;PSMD9;UBB;PRKCB;PSMB3;PSMC4;PSMD2;CLTB;PSME1;AP2S1;AP2A1 |
| 51 | Erythrocytes take up oxygen and release carbon dioxide Homo sapiens R-HSA-1247673 | 5.82E-03 | 1.17E-01 | 1.14E+01 | 5.86E+01 | CA2;CA4;AQP1 |
| 52 | VEGF ligand-receptor interactions Homo sapiens R-HSA-194313 | 5.82E-03 | 1.17E-01 | 1.14E+01 | 5.86E+01 | KDR;VEGFB;VEGFA |
| 53 | VEGF binds to VEGFR leading to receptor dimerization Homo sapiens R-HSA-195399 | 5.82E-03 | 1.17E-01 | 1.14E+01 | 5.86E+01 | KDR;VEGFB;VEGFA |
| 54 | Conjugation of salicylate with glycine Homo sapiens R-HSA-177128 | 5.82E-03 | 1.17E-01 | 1.14E+01 | 5.86E+01 | GLYATL1;GLYAT;ACSM2B |

| | | | | | | |
|---|---|---|---|---|---|---|
| 55 | ROS, RNS production in response to bacteria Homo sapiens R-HSA-1222556 | 6.31E-03 | 1.24E-01 | 4.08E+00 | 2.07E+01 | ATP6V1G1;CYBA;ATP6V0E2;ATP6V1E1;ATP6V0D1;ATP6V1F |
| 56 | Amino acid synthesis and interconversion (transamination) Homo sapiens R-HSA-70614 | 7.21E-03 | 1.36E-01 | 4.75E+00 | 2.34E+01 | FOLH1;GOT1;GOT2;ASPG;GLS |
| 57 | Glyoxylate metabolism and glycine degradation Homo sapiens R-HSA-389661 | 7.21E-03 | 1.36E-01 | 4.75E+00 | 2.34E+01 | GRHPR;DAO;AGXT2;NDUFAB1;PDHB |
| 58 | Bile acid and bile salt metabolism Homo sapiens R-HSA-194068 | 7.30E-03 | 1.36E-01 | 3.94E+00 | 1.94E+01 | ACOT8;ACOX2;AKR1C1;SLCO1A2;NR1H4;CYP46A1 |
| 59 | Metabolism of vitamins and cofactors Homo sapiens R-HSA-196854 | 7.50E-03 | 1.37E-01 | 2.15E+00 | 1.05E+01 | CYB5A;GSTO1;AKR1C1;QPRT;SLC2A1;APOC3;MTHFS;SLC2A3;APOM;NAPRT;APOC2;PCCB;ENPP1;ACP5;APOE |
| 60 | CDT1 association with the CDC6:ORC:origin complex Homo sapiens R-HSA-68827 | 8.05E-03 | 1.45E-01 | 3.05E+00 | 1.47E+01 | PSMD8;PSMD9;UBB;PSMB3;PSMC4;PSMD2;GMNN;PSME1 |
| 61 | Conjugation of carboxylic acids Homo sapiens R-HSA-159424 | 8.41E-03 | 1.47E-01 | 9.50E+00 | 4.54E+01 | GLYATL1;GLYAT;ACSM2B |
| 62 | Amino Acid conjugation Homo sapiens R-HSA-156587 | 8.41E-03 | 1.47E-01 | 9.50E+00 | 4.54E+01 | GLYATL1;GLYAT;ACSM2B |
| 63 | Insulin receptor recycling Homo sapiens R-HSA-77387 | 8.57E-03 | 1.47E-01 | 4.53E+00 | 2.15E+01 | ATP6V1G1;ATP6V0E2;ATP6V1E1;ATP6V0D1;ATP6V1F |
| 64 | APC/C:Cdh1 mediated degradation of Cdc20 and other APC/C:Cdh1 targeted proteins in late mitosis/early G1 Homo sapiens R-HSA-174178 | 8.84E-03 | 1.47E-01 | 2.77E+00 | 1.31E+01 | PSMD8;PSMD9;UBB;PSMB3;PSMC4;CDC26;PSMD2;PSME1;ANAPC11 |
| 65 | Cdc20:Phospho-APC/C mediated degradation of Cyclin A Homo sapiens R-HSA-174184 | 8.84E-03 | 1.47E-01 | 2.77E+00 | 1.31E+01 | PSMD8;PSMD9;UBB;PSMB3;PSMC4;CDC26;PSMD2;PSME1;ANAPC11 |

| | | | | | | |
|---|---|---|---|---|---|---|
| 66 | APC:Cdc20 mediated degradation of cell cycle proteins prior to satisfation of the cell cycle checkpoint Homo sapiens R-HSA-179419 | 9.66E-03 | 1.58E-01 | 2.72E+00 | 1.26E+01 | PSMD8;PSMD9;UBB;PSMB3;PSMC4;CDC26;PSMD2;PSME1;ANAPC11 |
| 67 | Peroxisomal lipid metabolism Homo sapiens R-HSA-390918 | 1.01E-02 | 1.60E-01 | 4.32E+00 | 1.99E+01 | ACOT8;ACOX2;ECI2;ACAA1;HAO2 |
| 68 | Synthesis of bile acids and bile salts Homo sapiens R-HSA-192105 | 1.01E-02 | 1.60E-01 | 4.32E+00 | 1.99E+01 | ACOT8;ACOX2;AKR1C1;NR1H4;CYP46A1 |
| 69 | Amino acid and oligopeptide SLC transporters Homo sapiens R-HSA-425374 | 1.06E-02 | 1.64E-01 | 3.17E+00 | 1.44E+01 | SLC6A6;SLC7A7;SLC7A9;SLC17A5;SLC6A12;SLC25A10;SLC38A2 |
| 70 | Regulation of activated PAK-2p34 by proteasome mediated degradation Homo sapiens R-HSA-211733 | 1.06E-02 | 1.64E-01 | 3.17E+00 | 1.44E+01 | PSMD8;PSMD9;UBB;PSMB3;PSMC4;PSMD2;PSME1 |
| 71 | APC/C:Cdc20 mediated degradation of mitotic proteins Homo sapiens R-HSA-176409 | 1.15E-02 | 1.64E-01 | 2.64E+00 | 1.18E+01 | PSMD8;PSMD9;UBB;PSMB3;PSMC4;CDC26;PSMD2;PSME1;ANAPC11 |
| 72 | Creatine metabolism Homo sapiens R-HSA-71288 | 1.16E-02 | 1.64E-01 | 8.14E+00 | 3.63E+01 | GAMT;SLC6A8;SLC6A12 |
| 73 | Metabolism of nucleotides Homo sapiens R-HSA-15869 | 1.17E-02 | 1.64E-01 | 2.47E+00 | 1.10E+01 | AGXT2;CAT;ADSSL1;AK2;GMPR;CTPS1;GLRX;TK1;UPB1;APRT |
| 74 | CDK-mediated phosphorylation and removal of Cdc6 Homo sapiens R-HSA-69017 | 1.19E-02 | 1.64E-01 | 3.10E+00 | 1.37E+01 | PSMD8;PSMD9;UBB;PSMB3;PSMC4;PSMD2;PSME1 |
| 75 | Ubiquitin-dependent degradation of Cyclin D1 Homo sapiens R-HSA-69229 | 1.19E-02 | 1.64E-01 | 3.10E+00 | 1.37E+01 | PSMD8;PSMD9;UBB;PSMB3;PSMC4;PSMD2;PSME1 |
| 76 | Ubiquitin-dependent degradation of Cyclin D Homo sapiens R-HSA-75815 | 1.19E-02 | 1.64E-01 | 3.10E+00 | 1.37E+01 | PSMD8;PSMD9;UBB;PSMB3;PSMC4;PSMD2;PSME1 |
| 77 | Regulation of Apoptosis Homo sapiens R-HSA-169911 | 1.19E-02 | 1.64E-01 | 3.10E+00 | 1.37E+01 | PSMD8;PSMD9;UBB;PSMB3;PSMC4;PSMD2;PSME1 |

| | | | | | | |
|---|---|---|---|---|---|---|
| 78 | Regulation of ornithine decarboxylase (ODC) Homo sapiens R-HSA-350562 | 1.19E-02 | 1.64E-01 | 3.10E+00 | 1.37E+01 | PSMD8;PSMD9;PSMB3;PSMC4;PSMD2;PSME1;OAZ1 |
| 79 | Activation of APC/C and APC/C:Cdc20 mediated degradation of mitotic proteins Homo sapiens R-HSA-176814 | 1.25E-02 | 1.71E-01 | 2.60E+00 | 1.14E+01 | PSMD8;PSMD9;UBB;PSMB3;PSMC4;CDC26;PSMD2;PSME1;ANAPC11 |
| 80 | Circadian Clock Homo sapiens R-HSA-400253 | 1.31E-02 | 1.72E-01 | 2.77E+00 | 1.20E+01 | DBP;UBB;BHLHE40;RORA;AVP;NR1D1;HIF1A;NPAS2 |
| 81 | Vpu mediated degradation of CD4 Homo sapiens R-HSA-180534 | 1.32E-02 | 1.72E-01 | 3.03E+00 | 1.31E+01 | PSMD8;PSMD9;UBB;PSMB3;PSMC4;PSMD2;PSME1 |
| 82 | Autodegradation of the E3 ubiquitin ligase COP1 Homo sapiens R-HSA-349425 | 1.32E-02 | 1.72E-01 | 3.03E+00 | 1.31E+01 | PSMD8;PSMD9;UBB;PSMB3;PSMC4;PSMD2;PSME1 |
| 83 | Aquaporin-mediated transport Homo sapiens R-HSA-445717 | 1.32E-02 | 1.72E-01 | 3.03E+00 | 1.31E+01 | GNG4;PRKAR2B;AVP;AQP2;AQP3;GNG11;AQP1 |
| 84 | Beta-catenin independent WNT signaling Homo sapiens R-HSA-3858494 | 1.35E-02 | 1.74E-01 | 2.05E+00 | 8.84E+00 | PRKCB;CLTB;AP2A1;GNG11;PSMD8;PSMD9;UBB;GNG4;PSMB3;PSMC4;PSMD2;PSME1;AP2S1;CALM1 |
| 85 | Transferrin endocytosis and recycling Homo sapiens R-HSA-917977 | 1.37E-02 | 1.74E-01 | 3.96E+00 | 1.70E+01 | ATP6V1G1;ATP6V0E2;ATP6V1E1;ATP6V0D1;ATP6V1F |
| 86 | p53-Independent DNA Damage Response Homo sapiens R-HSA-69610 | 1.46E-02 | 1.79E-01 | 2.96E+00 | 1.25E+01 | PSMD8;PSMD9;UBB;PSMB3;PSMC4;PSMD2;PSME1 |
| 87 | p53-Independent G1/S DNA damage checkpoint Homo sapiens R-HSA-69613 | 1.46E-02 | 1.79E-01 | 2.96E+00 | 1.25E+01 | PSMD8;PSMD9;UBB;PSMB3;PSMC4;PSMD2;PSME1 |
| 88 | Ubiquitin Mediated Degradation of Phosphorylated Cdc25A Homo sapiens R-HSA-69601 | 1.46E-02 | 1.79E-01 | 2.96E+00 | 1.25E+01 | PSMD8;PSMD9;UBB;PSMB3;PSMC4;PSMD2;PSME1 |
| 89 | Transport of organic anions Homo sapiens R-HSA-879518 | 1.53E-02 | 1.84E-01 | 7.12E+00 | 2.98E+01 | SLCO4A1;SLCO1A2;AVP |

| | | | | | | |
|---|---|---|---|---|---|---|
| 90 | Phenylalanine and tyrosine catabolism Homo sapiens R-HSA-71182 | 1.53E-02 | 1.84E-01 | 7.12E+00 | 2.98E+01 | QDPR;PCBD1;FAH |
| 91 | Vif-mediated degradation of APOBEC3G Homo sapiens R-HSA-180585 | 1.61E-02 | 1.90E-01 | 2.90E+00 | 1.20E+01 | PSMD8;PSMD9;UBB;PSMB3;PSMC4;PSMD2;PSME1 |
| 92 | DNA Replication Homo sapiens R-HSA-69306 | 1.62E-02 | 1.90E-01 | 2.23E+00 | 9.20E+00 | PSMD8;POLD3;PSMD9;UBB;RFC1;PSMB3;PSMC4;PSMD2;RPA3;GMNN;PSME1 |
| 93 | Transcriptional Regulation by TP53 Homo sapiens R-HSA-3700989 | 1.68E-02 | 1.93E-01 | 1.61E+00 | 6.58E+00 | COX7B;COX7C;COX5A;NDRG1;GLS;UBB;PRDX1;SESN2;POLR2E;PMS2;COX11;COX14;POLR2I;COX8A;BNIP3L;PRELID1;SURF1;GPX2;USP2;COX6B1;RGCC;BCL6;RPA3;DDIT4;LAMTOR2;LAMTOR1;LAMTOR5 |
| 94 | Activation of NF-kappaB in B cells Homo sapiens R-HSA-1169091 | 1.70E-02 | 1.93E-01 | 2.63E+00 | 1.07E+01 | PSMD8;PSMD9;UBB;PRKCB;PSMB3;PSMC4;PSMD2;PSME1 |
| 95 | Regulation of APC/C activators between G1/S and early anaphase Homo sapiens R-HSA-176408 | 1.72E-02 | 1.93E-01 | 2.45E+00 | 9.95E+00 | PSMD8;PSMD9;UBB;PSMB3;PSMC4;CDC26;PSMD2;PSME1;ANAPC11 |
| 96 | SCF-beta-TrCP mediated degradation of Emi1 Homo sapiens R-HSA-174113 | 1.77E-02 | 1.93E-01 | 2.83E+00 | 1.14E+01 | PSMD8;PSMD9;UBB;PSMB3;PSMC4;PSMD2;PSME1 |
| 97 | Stabilization of p53 Homo sapiens R-HSA-69541 | 1.77E-02 | 1.93E-01 | 2.83E+00 | 1.14E+01 | PSMD8;PSMD9;UBB;PSMB3;PSMC4;PSMD2;PSME1 |
| 98 | AUF1 (hnRNP D0) binds and destabilizes mRNA Homo sapiens R-HSA-450408 | 1.77E-02 | 1.93E-01 | 2.83E+00 | 1.14E+01 | PSMD8;PSMD9;UBB;PSMB3;PSMC4;PSMD2;PSME1 |
| 99 | Degradation of AXIN Homo sapiens R-HSA-4641257 | 1.77E-02 | 1.93E-01 | 2.83E+00 | 1.14E+01 | PSMD8;PSMD9;UBB;PSMB3;PSMC4;PSMD2;PSME1 |
| 100 | Amino acid transport across the plasma membrane Homo sapiens R-HSA-352230 | 1.80E-02 | 1.95E-01 | 3.66E+00 | 1.47E+01 | SLC6A6;SLC7A7;SLC7A9;SLC6A12;SLC38A2 |
| 101 | Assembly of the pre-replicative complex Homo sapiens R-HSA-68867 | 1.85E-02 | 1.98E-01 | 2.58E+00 | 1.03E+01 | PSMD8;PSMD9;UBB;PSMB3;PSMC4;PSMD2;GMNN;PSME1 |

| | | | | | | |
|---|---|---|---|---|---|---|
| 102 | Hh mutants that don't undergo autocatalytic processing are degraded by ERAD Homo sapiens R-HSA-5362768 | 1.94E-02 | 2.01E-01 | 2.77E+00 | 1.09E+01 | PSMD8;PSMD9;UBB;PSMB3;PSMC4;PSMD2;PSME1 |
| 103 | Erythrocytes take up carbon dioxide and release oxygen Homo sapiens R-HSA-1237044 | 1.97E-02 | 2.01E-01 | 6.33E+00 | 2.49E+01 | CA2;CA4;AQP1 |
| 104 | O2/CO2 exchange in erythrocytes Homo sapiens R-HSA-1480926 | 1.97E-02 | 2.01E-01 | 6.33E+00 | 2.49E+01 | CA2;CA4;AQP1 |
| 105 | Reversible hydration of carbon dioxide Homo sapiens R-HSA-1475029 | 1.97E-02 | 2.01E-01 | 6.33E+00 | 2.49E+01 | CA5B;CA2;CA4 |
| 106 | Passive transport by Aquaporins Homo sapiens R-HSA-432047 | 1.97E-02 | 2.01E-01 | 6.33E+00 | 2.49E+01 | AQP2;AQP3;AQP1 |
| 107 | Degradation of DVL Homo sapiens R-HSA-4641258 | 2.13E-02 | 2.05E-01 | 2.72E+00 | 1.05E+01 | PSMD8;PSMD9;UBB;PSMB3;PSMC4;PSMD2;PSME1 |
| 108 | DNA Replication Pre-Initiation Homo sapiens R-HSA-69002 | 2.14E-02 | 2.05E-01 | 2.35E+00 | 9.02E+00 | PSMD8;PSMD9;UBB;PSMB3;PSMC4;PSMD2;RPA3;GMNN;PSME1 |
| 109 | M/G1 Transition Homo sapiens R-HSA-68874 | 2.14E-02 | 2.05E-01 | 2.35E+00 | 9.02E+00 | PSMD8;PSMD9;UBB;PSMB3;PSMC4;PSMD2;RPA3;GMNN;PSME1 |
| 110 | Cyclin A:Cdk2-associated events at S phase entry Homo sapiens R-HSA-69656 | 2.17E-02 | 2.05E-01 | 2.50E+00 | 9.56E+00 | PSMD8;PSMD9;UBB;PSMB3;PSMC4;PSMD2;PSME1;CDC25B |
| 111 | Response to elevated platelet cytosolic Ca2+ Homo sapiens R-HSA-76005 | 2.22E-02 | 2.05E-01 | 2.12E+00 | 8.06E+00 | SERPINA1;PRKCB;PCDH7;VEGFB;SERPING1;PLG;PPBP;CALM1;VEGFA;SOD1;PF4 |
| 112 | Resolution of AP sites via the multiple-nucleotide patch replacement pathway Homo sapiens R-HSA-110373 | 2.23E-02 | 2.05E-01 | 4.22E+00 | 1.61E+01 | POLD3;PARG;RFC1;RPA3 |
| 113 | Na+-dependent glucose transporters Homo sapiens R-HSA-428808 | 2.27E-02 | 2.05E-01 | 1.27E+01 | 4.79E+01 | SLC5A2;SLC5A3 |

| | | | | | | |
|---|---|---|---|---|---|---|
| 114 | Sodium-coupled phosphate cotransporters Homo sapiens R-HSA-427652 | 2.27E-02 | 2.05E-01 | 1.27E+01 | 4.79E+01 | SLC34A3;SLC34A1 |
| 115 | Catabolism of glucuronate to xylulose-5-phosphate Homo sapiens R-HSA-5661270 | 2.27E-02 | 2.05E-01 | 1.27E+01 | 4.79E+01 | AKR1A1;CRYL1 |
| 116 | Beta oxidation of octanoyl-CoA to hexanoyl-CoA Homo sapiens R-HSA-77348 | 2.27E-02 | 2.05E-01 | 1.27E+01 | 4.79E+01 | ECHS1;ACADM |
| 117 | Beta oxidation of decanoyl-CoA to octanoyl-CoA-CoA Homo sapiens R-HSA-77346 | 2.27E-02 | 2.05E-01 | 1.27E+01 | 4.79E+01 | ECHS1;ACADM |
| 118 | Vitamin B2 (riboflavin) metabolism Homo sapiens R-HSA-196843 | 2.27E-02 | 2.05E-01 | 1.27E+01 | 4.79E+01 | ENPP1;ACP5 |
| 119 | Sulfide oxidation to sulfate Homo sapiens R-HSA-1614517 | 2.27E-02 | 2.05E-01 | 1.27E+01 | 4.79E+01 | TST;SLC25A10 |
| 120 | Galactose catabolism Homo sapiens R-HSA-70370 | 2.27E-02 | 2.05E-01 | 1.27E+01 | 4.79E+01 | PGM1;GALK1 |
| 121 | Hh mutants abrogate ligand secretion Homo sapiens R-HSA-5387390 | 2.33E-02 | 2.08E-01 | 2.66E+00 | 1.00E+01 | PSMD8;PSMD9;UBB;PSMB3;PSMC4;PSMD2;PSME1 |
| 122 | Synthesis of DNA Homo sapiens R-HSA-69239 | 2.35E-02 | 2.08E-01 | 2.19E+00 | 8.21E+00 | PSMD8;POLD3;PSMD9;UBB;RFC1;PSMB3;PSMC4;PSMD2;RPA3;PSME1 |
| 123 | Signaling by Insulin receptor Homo sapiens R-HSA-74752 | 2.45E-02 | 2.13E-01 | 1.60E+00 | 5.93E+00 | KLB;ATP6V1G1;CAB39L;RASGRF1;PEBP1;PHB;FGF2;PSMD8;PSMD9;UBB;PSMB3;PSMC4;PSMD2;FGF18;PSME1;LAMTOR2;FYN;LAMTOR1;ATP6V1E1;ATP6V0D1;CALM1;ATP6V0E2;LAMTOR5;ATP6V1F |
| 124 | Facilitative Na+-independent glucose transporters Homo sapiens R-HSA-428790 | 2.47E-02 | 2.13E-01 | 5.70E+00 | 2.11E+01 | SLC2A9;SLC2A1;SLC2A3 |
| 125 | WNT5A-dependent internalization of FZD2, FZD5 and ROR2 Homo sapiens R-HSA-5140745 | 2.47E-02 | 2.13E-01 | 5.70E+00 | 2.11E+01 | CLTB;AP2S1;AP2A1 |

| | | | | | | |
|---|---|---|---|---|---|---|
| 126 | Metabolism of water-soluble vitamins and cofactors Homo sapiens R-HSA-196849 | 2.51E-02 | 2.15E-01 | 2.16E+00 | 7.98E+00 | CYB5A;GSTO1;NAPRT;QPRT;PCCB;SLC2A1;ENPP1;ACP5;MTHFS;SLC2A3 |
| 127 | NIK-->noncanonical NF-kB signaling Homo sapiens R-HSA-5676590 | 2.53E-02 | 2.16E-01 | 2.61E+00 | 9.60E+00 | PSMD8;PSMD9;UBB;PSMB3;PSMC4;PSMD2;PSME1 |
| 128 | Gap-filling DNA repair synthesis and ligation in GG-NER Homo sapiens R-HSA-5696397 | 2.60E-02 | 2.17E-01 | 4.00E+00 | 1.46E+01 | POLD3;RFC1;UBB;RPA3 |
| 129 | Lipoprotein metabolism Homo sapiens R-HSA-174824 | 2.61E-02 | 2.17E-01 | 3.28E+00 | 1.19E+01 | APOC2;APOC3;APOE;PLTP;ABCG1 |
| 130 | APC/C-mediated degradation of cell cycle proteins Homo sapiens R-HSA-174143 | 2.64E-02 | 2.17E-01 | 2.25E+00 | 8.19E+00 | PSMD8;PSMD9;UBB;PSMB3;PSMC4;CDC26;PSMD2;PSME1;ANAPC11 |
| 131 | Regulation of mitotic cell cycle Homo sapiens R-HSA-453276 | 2.64E-02 | 2.17E-01 | 2.25E+00 | 8.19E+00 | PSMD8;PSMD9;UBB;PSMB3;PSMC4;CDC26;PSMD2;PSME1;ANAPC11 |
| 132 | Removal of licensing factors from origins Homo sapiens R-HSA-69300 | 2.73E-02 | 2.17E-01 | 2.38E+00 | 8.56E+00 | PSMD8;PSMD9;UBB;PSMB3;PSMC4;PSMD2;GMNN;PSME1 |
| 133 | Degradation of GLI1 by the proteasome Homo sapiens R-HSA-5610780 | 2.76E-02 | 2.17E-01 | 2.56E+00 | 9.20E+00 | PSMD8;PSMD9;UBB;PSMB3;PSMC4;PSMD2;PSME1 |
| 134 | GLI3 is processed to GLI3R by the proteasome Homo sapiens R-HSA-5610785 | 2.76E-02 | 2.17E-01 | 2.56E+00 | 9.20E+00 | PSMD8;PSMD9;UBB;PSMB3;PSMC4;PSMD2;PSME1 |
| 135 | Degradation of GLI2 by the proteasome Homo sapiens R-HSA-5610783 | 2.76E-02 | 2.17E-01 | 2.56E+00 | 9.20E+00 | PSMD8;PSMD9;UBB;PSMB3;PSMC4;PSMD2;PSME1 |
| 136 | The role of GTSE1 in G2/M progression after G2 checkpoint Homo sapiens R-HSA-8852276 | 2.76E-02 | 2.17E-01 | 2.56E+00 | 9.20E+00 | PSMD8;PSMD9;UBB;PSMB3;PSMC4;PSMD2;PSME1 |
| 137 | SCF(Skp2)-mediated degradation of p27/p21 Homo sapiens R-HSA-187577 | 2.76E-02 | 2.17E-01 | 2.56E+00 | 9.20E+00 | PSMD8;PSMD9;UBB;PSMB3;PSMC4;PSMD2;PSME1 |

| | | | | | | |
|---|---|---|---|---|---|---|
| 138 | Host Interactions of HIV factors Homo sapiens R-HSA-162909 | 2.88E-02 | 2.25E-01 | 1.95E+00 | 6.93E+00 | PSMD8;PSMD9;UBB;PSMB3;PSMC4;PSMD2;PSME1;AP2S1;AP2A1;FYN;SLC25A5;AP1M1 |
| 139 | Base Excision Repair Homo sapiens R-HSA-73884 | 2.92E-02 | 2.26E-01 | 3.17E+00 | 1.12E+01 | POLD3;PARG;RFC1;OGG1;RPA3 |
| 140 | Resolution of Abasic Sites (AP sites) Homo sapiens R-HSA-73933 | 2.92E-02 | 2.26E-01 | 3.17E+00 | 1.12E+01 | POLD3;PARG;RFC1;OGG1;RPA3 |
| 141 | Synthesis of bile acids and bile salts via 7alpha-hydroxycholesterol Homo sapiens R-HSA-193368 | 3.00E-02 | 2.27E-01 | 3.80E+00 | 1.33E+01 | ACOT8;ACOX2;AKR1C1;NR1H4 |
| 142 | Mismatch repair (MMR) directed by MSH2:MSH3 (MutSbeta) Homo sapiens R-HSA-5358606 | 3.02E-02 | 2.27E-01 | 5.18E+00 | 1.81E+01 | POLD3;RPA3;PMS2 |
| 143 | Mismatch repair (MMR) directed by MSH2:MSH6 (MutSalpha) Homo sapiens R-HSA-5358565 | 3.02E-02 | 2.27E-01 | 5.18E+00 | 1.81E+01 | POLD3;RPA3;PMS2 |
| 144 | FGFR4 ligand binding and activation Homo sapiens R-HSA-190322 | 3.02E-02 | 2.27E-01 | 5.18E+00 | 1.81E+01 | KLB;FGF18;FGF2 |
| 145 | Cellular responses to stress Homo sapiens R-HSA-2262752 | 3.05E-02 | 2.27E-01 | 1.52E+00 | 5.30E+00 | RPS19BP1;WDR45;HIF1A;ETS2;HDAC6;ANAPC11;UBB;CDC26;PRDX1;BAG1;HIST1H1C;EGLN3;GPX2;H2AFZ;CBX2;CYBA;WIPI1;HSPA2;WIPI2;ATOX1;VEGFA;SOD1;RPA3;CAT;LAMTOR2;LAMTOR1;LAMTOR5 |
| 146 | Hexose transport Homo sapiens R-HSA-189200 | 3.19E-02 | 2.34E-01 | 2.72E+00 | 9.35E+00 | SLC2A1;SLC2A3;SLC5A2;SLC37A4;SLC5A3;HK2 |
| 147 | Dectin-1 mediated noncanonical NF-kB signaling Homo sapiens R-HSA-5607761 | 3.24E-02 | 2.34E-01 | 2.47E+00 | 8.46E+00 | PSMD8;PSMD9;UBB;PSMB3;PSMC4;PSMD2;PSME1 |
| 148 | Fructose metabolism Homo sapiens R-HSA-5652084 | 3.29E-02 | 2.34E-01 | 9.49E+00 | 3.24E+01 | AKR1B1;KHK |

| | | | | | | |
|---|---|---|---|---|---|---|
| 149 | mitochondrial fatty acid beta-oxidation of unsaturated fatty acids Homo sapiens R-HSA-77288 | 3.29E-02 | 2.34E-01 | 9.49E+00 | 3.24E+01 | ACADM;DECR1 |
| 150 | Beta-oxidation of very long chain fatty acids Homo sapiens R-HSA-390247 | 3.29E-02 | 2.34E-01 | 9.49E+00 | 3.24E+01 | ACOT8;ACAA1 |
| 151 | Heme degradation Homo sapiens R-HSA-189483 | 3.29E-02 | 2.34E-01 | 9.49E+00 | 3.24E+01 | HMOX1;HMOX2 |
| 152 | Synthesis of Lipoxins (LX) Homo sapiens R-HSA-2142700 | 3.29E-02 | 2.34E-01 | 9.49E+00 | 3.24E+01 | HPGD;PTGR1 |
| 153 | Regulation of DNA replication Homo sapiens R-HSA-69304 | 3.39E-02 | 2.39E-01 | 2.27E+00 | 7.69E+00 | PSMD8;PSMD9;UBB;PSMB3;PSMC4;PSMD2;GMNN;PSME1 |
| 154 | Basigin interactions Homo sapiens R-HSA-210991 | 3.43E-02 | 2.41E-01 | 3.62E+00 | 1.22E+01 | SLC7A7;SLC7A9;ATP1B3;ATP1B1 |
| 155 | Cross-presentation of soluble exogenous antigens (endosomes) Homo sapiens R-HSA-1236978 | 3.49E-02 | 2.42E-01 | 2.65E+00 | 8.90E+00 | PSMD8;PSMD9;PSMB3;PSMC4;PSMD2;PSME1 |
| 156 | Gap-filling DNA repair synthesis and ligation in TC-NER Homo sapiens R-HSA-6782210 | 3.50E-02 | 2.42E-01 | 2.42E+00 | 8.12E+00 | POLD3;XAB2;RFC1;UBB;RPA3;POLR2E;POLR2I |
| 157 | Mismatch Repair Homo sapiens R-HSA-5358508 | 3.64E-02 | 2.51E-01 | 4.75E+00 | 1.57E+01 | POLD3;RPA3;PMS2 |
| 158 | Dual incision in TC-NER Homo sapiens R-HSA-6782135 | 3.77E-02 | 2.53E-01 | 2.38E+00 | 7.79E+00 | POLD3;XAB2;RFC1;UBB;RPA3;POLR2E;POLR2I |
| 159 | p53-Dependent G1/S DNA damage checkpoint Homo sapiens R-HSA-69580 | 3.77E-02 | 2.53E-01 | 2.38E+00 | 7.79E+00 | PSMD8;PSMD9;UBB;PSMB3;PSMC4;PSMD2;PSME1 |
| 160 | p53-Dependent G1 DNA Damage Response Homo sapiens R-HSA-69563 | 3.77E-02 | 2.53E-01 | 2.38E+00 | 7.79E+00 | PSMD8;PSMD9;UBB;PSMB3;PSMC4;PSMD2;PSME1 |
| 161 | Asymmetric localization of PCP proteins Homo sapiens R-HSA-4608870 | 3.77E-02 | 2.53E-01 | 2.38E+00 | 7.79E+00 | PSMD8;PSMD9;UBB;PSMB3;PSMC4;PSMD2;PSME1 |

| | | | | | | |
|---|---|---|---|---|---|---|
| 162 | Platelet degranulation Homo sapiens R-HSA-114608 | 3.79E-02 | 2.53E-01 | 2.00E+00 | 6.56E+00 | SERPINA1;PCDH7;VEGFB;SERPING1;PLG;PPBP;CALM1;VEGFA;SOD1;PF4 |
| 163 | Regulation of Hypoxia-inducible Factor (HIF) by oxygen Homo sapiens R-HSA-1234174 | 3.90E-02 | 2.57E-01 | 3.45E+00 | 1.12E+01 | EGLN3;UBB;HIF1A;VEGFA |
| 164 | Cellular response to hypoxia Homo sapiens R-HSA-2262749 | 3.90E-02 | 2.57E-01 | 3.45E+00 | 1.12E+01 | EGLN3;UBB;HIF1A;VEGFA |
| 165 | Hedgehog ligand biogenesis Homo sapiens R-HSA-5358346 | 4.06E-02 | 2.66E-01 | 2.34E+00 | 7.48E+00 | PSMD8;PSMD9;UBB;PSMB3;PSMC4;PSMD2;PSME1 |
| 166 | Ion homeostasis Homo sapiens R-HSA-5578775 | 4.14E-02 | 2.69E-01 | 2.53E+00 | 8.07E+00 | DMPK;FXYD4;FXYD2;ATP1B3;ATP1B1;CALM1 |
| 167 | Translesion synthesis by REV1 Homo sapiens R-HSA-110312 | 4.32E-02 | 2.75E-01 | 4.38E+00 | 1.38E+01 | RFC1;UBB;RPA3 |
| 168 | ER-Phagosome pathway Homo sapiens R-HSA-1236974 | 4.36E-02 | 2.75E-01 | 2.29E+00 | 7.19E+00 | PSMD8;PSMD9;UBB;PSMB3;PSMC4;PSMD2;PSME1 |
| 169 | G1/S DNA Damage Checkpoints Homo sapiens R-HSA-69615 | 4.36E-02 | 2.75E-01 | 2.29E+00 | 7.19E+00 | PSMD8;PSMD9;UBB;PSMB3;PSMC4;PSMD2;PSME1 |
| 170 | EGFR downregulation Homo sapiens R-HSA-182971 | 4.40E-02 | 2.75E-01 | 3.30E+00 | 1.03E+01 | UBB;AP2S1;AP2A1;EPS15L1 |
| 171 | Pyruvate metabolism Homo sapiens R-HSA-70268 | 4.40E-02 | 2.75E-01 | 3.30E+00 | 1.03E+01 | LDHA;PDHB;HAGH;PDK2 |
| 172 | VEGFR2 mediated cell proliferation Homo sapiens R-HSA-5218921 | 4.44E-02 | 2.75E-01 | 1.58E+00 | 4.93E+00 | KLB;PRKCB;RASGRF1;PEBP1;PHB;FGF2;VEGFA;PSMD8;PSMD9;UBB;PSMB3;PSMC4;PSMD2;FGF18;KDR;PSME1;LAMTOR2;FYN;CALM1 |
| 173 | Nef Mediated CD8 Down-regulation Homo sapiens R-HSA-182218 | 4.46E-02 | 2.75E-01 | 7.59E+00 | 2.36E+01 | AP2S1;AP2A1 |
| 174 | PTK6 promotes HIF1A stabilization Homo sapiens R-HSA-8857538 | 4.46E-02 | 2.75E-01 | 7.59E+00 | 2.36E+01 | LINC01139;HIF1A |
| 175 | Synthesis of Ketone Bodies Homo sapiens R-HSA-77111 | 4.46E-02 | 2.75E-01 | 7.59E+00 | 2.36E+01 | HMGCL;BDH2 |

| | | | | | | |
|---|---|---|---|---|---|---|
| 176 | S Phase Homo sapiens R-HSA-69242 | 4.72E-02 | 2.90E-01 | 1.85E+00 | 5.66E+00 | PSMD8;POLD3;PSMD9;UBB;RFC1;PSMB3;PSMC4;PSMD2;RPA3;PSME1;CDC25B |
| 177 | The role of Nef in HIV-1 replication and disease pathogenesis Homo sapiens R-HSA-164952 | 4.94E-02 | 2.95E-01 | 3.17E+00 | 9.52E+00 | AP2S1;AP2A1;FYN;AP1M1 |
| 178 | Energy dependent regulation of mTOR by LKB1-AMPK Homo sapiens R-HSA-380972 | 4.94E-02 | 2.95E-01 | 3.17E+00 | 9.52E+00 | CAB39L;LAMTOR2;LAMTOR1;LAMTOR5 |
| 179 | Recognition of DNA damage by PCNA-containing replication complex Homo sapiens R-HSA-110314 | 4.94E-02 | 2.95E-01 | 3.17E+00 | 9.52E+00 | POLD3;UBB;RFC1;RPA3 |
| 180 | Regulation of RAS by GAPs Homo sapiens R-HSA-5658442 | 5.00E-02 | 2.95E-01 | 2.22E+00 | 6.64E+00 | PSMD8;PSMD9;UBB;PSMB3;PSMC4;PSMD2;PSME1 |
| 181 | Trafficking of GluR2-containing AMPA receptors Homo sapiens R-HSA-416993 | 5.06E-02 | 2.95E-01 | 4.07E+00 | 1.21E+01 | PRKCB;AP2S1;AP2A1 |
| 182 | Translesion synthesis by POLK Homo sapiens R-HSA-5655862 | 5.06E-02 | 2.95E-01 | 4.07E+00 | 1.21E+01 | RFC1;UBB;RPA3 |
| 183 | Translesion synthesis by POLI Homo sapiens R-HSA-5656121 | 5.06E-02 | 2.95E-01 | 4.07E+00 | 1.21E+01 | RFC1;UBB;RPA3 |
| 184 | Chylomicron-mediated lipid transport Homo sapiens R-HSA-174800 | 5.06E-02 | 2.95E-01 | 4.07E+00 | 1.21E+01 | APOC2;APOC3;APOE |
| 185 | Metabolism of porphyrins Homo sapiens R-HSA-189445 | 5.06E-02 | 2.95E-01 | 4.07E+00 | 1.21E+01 | UROS;HMOX1;HMOX2 |
| 186 | Platelet activation, signaling and aggregation Homo sapiens R-HSA-76002 | 5.23E-02 | 3.04E-01 | 1.55E+00 | 4.57E+00 | PTPN1;PRKCH;SERPINA1;PRKCB;PCDH7;F2R;VEGFB;PLA2G4A;PLG;PPBP;GNG11;VEGFA;SOD1;GNG4;SERPING1;FYN;CALM1;MGLL;PF4 |
| 187 | Antigen processing-Cross presentation Homo sapiens R-HSA-1236975 | 5.30E-02 | 3.06E-01 | 2.06E+00 | 6.04E+00 | PSMD8;PSMD9;UBB;PSMB3;PSMC4;PSMD2;PSME1;CYBA |
| 188 | G2/M Transition Homo sapiens R-HSA-69275 | 5.31E-02 | 3.06E-01 | 1.68E+00 | 4.92E+00 | TUBGCP2;SSNA1;TUBG1;CDC25B;PSMD8;PSMD9;UBB;PRKAR2B;PSMB3;PSMC4;PSMD2;PSME1;CEP63;OPTN |

| | | | | | | |
|---|---|---|---|---|---|---|
| 189 | Cyclin E associated events during G1/S transition Homo sapiens R-HSA-69202 | 5.35E-02 | 3.06E-01 | 2.18E+00 | 6.39E+00 | PSMD8;PSMD9;UBB;PSMB3;PSMC4;PSMD2;PSME1 |
| 190 | Visual phototransduction Homo sapiens R-HSA-2187338 | 5.44E-02 | 3.09E-01 | 1.95E+00 | 5.67E+00 | APOM;RDH10;AKR1C1;APOC2;ABCA4;NAPEPLD;APOC3;APOE;CALM1 |
| 191 | Mitotic G2-G2/M phases Homo sapiens R-HSA-453274 | 5.74E-02 | 3.15E-01 | 1.66E+00 | 4.73E+00 | TUBGCP2;SSNA1;TUBG1;CDC25B;PSMD8;PSMD9;UBB;PRKAR2B;PSMB3;PSMC4;PSMD2;PSME1;CEP63;OPTN |
| 192 | PPARA activates gene expression Homo sapiens R-HSA-1989781 | 5.75E-02 | 3.15E-01 | 1.85E+00 | 5.28E+00 | ESRRA;FABP1;MED11;NR1H4;G0S2;RORA;PLIN2;NR1D1;ACADM;NPAS2 |
| 193 | Organic anion transporters Homo sapiens R-HSA-428643 | 5.75E-02 | 3.15E-01 | 6.32E+00 | 1.81E+01 | SLC17A5;SLC25A10 |
| 194 | mitochondrial fatty acid beta-oxidation of saturated fatty acids Homo sapiens R-HSA-77286 | 5.75E-02 | 3.15E-01 | 6.32E+00 | 1.81E+01 | ECHS1;ACADM |
| 195 | NR1D1 (REV-ERBA) represses gene expression Homo sapiens R-HSA-1368071 | 5.75E-02 | 3.15E-01 | 6.32E+00 | 1.81E+01 | NR1D1;NPAS2 |
| 196 | Ketone body metabolism Homo sapiens R-HSA-74182 | 5.75E-02 | 3.15E-01 | 6.32E+00 | 1.81E+01 | HMGCL;BDH2 |
| 197 | Signaling by Retinoic Acid Homo sapiens R-HSA-5362517 | 5.77E-02 | 3.15E-01 | 2.57E+00 | 7.32E+00 | ALDH1A3;RDH10;RARA;PDHB;PDK2 |
| 198 | Retinoid metabolism and transport Homo sapiens R-HSA-975634 | 5.77E-02 | 3.15E-01 | 2.57E+00 | 7.32E+00 | APOM;AKR1C1;APOC2;APOC3;APOE |
| 199 | Oxygen-dependent proline hydroxylation of Hypoxia-inducible Factor Alpha Homo sapiens R-HSA-1234176 | 5.85E-02 | 3.18E-01 | 3.80E+00 | 1.08E+01 | EGLN3;UBB;HIF1A |
| 200 | Hedgehog 'off' state Homo sapiens R-HSA-5610787 | 6.04E-02 | 3.25E-01 | 1.90E+00 | 5.34E+00 | PSMD8;PSMD9;UBB;PRKAR2B;PSMB3;PSMC4;PSMD2;PSME1;FUZ |
| 201 | Switching of origins to a post-replicative state Homo sapiens R-HSA-69052 | 6.08E-02 | 3.25E-01 | 2.11E+00 | 5.91E+00 | PSMD8;PSMD9;UBB;PSMB3;PSMC4;PSMD2;PSME1 |

| | | | | | | |
|---|---|---|---|---|---|---|
| 202 | Orc1 removal from chromatin Homo sapiens R-HSA-68949 | 6.08E-02 | 3.25E-01 | 2.11E+00 | 5.91E+00 | PSMD8;PSMD9;UBB;PSMB3;PSMC4;PSMD2;PSME1 |
| 203 | Termination of translesion DNA synthesis Homo sapiens R-HSA-5656169 | 6.12E-02 | 3.26E-01 | 2.92E+00 | 8.16E+00 | POLD3;UBB;RFC1;RPA3 |
| 204 | Hedgehog 'on' state Homo sapiens R-HSA-5632684 | 6.29E-02 | 3.33E-01 | 1.98E+00 | 5.46E+00 | PSMD8;PSMD9;UBB;PSMB3;PSMC4;PSMD2;BOC;PSME1 |
| 205 | Organelle biogenesis and maintenance Homo sapiens R-HSA-1852241 | 6.31E-02 | 3.33E-01 | 1.45E+00 | 4.00E+00 | ESRRA;TSFM;MRPS15;MRPS26;MRPS25;MRPS36;SSNA1;MRPL19;MRPS34;NR1D1;MRPL37;MRPL48;MTIF3;MRPL12;MRPL34;TUBG1;MRPS6;MRPL46;HDAC6;PRKAR2B;CEP63;DYNLRB1;RAB11FIP3 |
| 206 | Lipid digestion, mobilization, and transport Homo sapiens R-HSA-73923 | 6.47E-02 | 3.39E-01 | 2.08E+00 | 5.69E+00 | FABP1;APOC2;APOC3;APOE;PLTP;ABCG1;MGLL |
| 207 | Regulation of lipid metabolism by Peroxisome proliferator-activated receptor alpha (PPARalpha) Homo sapiens R-HSA-400206 | 6.63E-02 | 3.41E-01 | 1.79E+00 | 4.87E+00 | ESRRA;FABP1;MED11;NR1H4;G0S2;RORA;PLIN2;NR1D1;ACADM;NPAS2 |
| 208 | Signaling by VEGF Homo sapiens R-HSA-194138 | 6.66E-02 | 3.41E-01 | 1.44E+00 | 3.90E+00 | KLB;PRKCB;RASGRF1;VEGFB;PEBP1;CYBA;PHB;SHB;BAIAP2;FGF2;VEGFA;PSMD8;PSMD9;UBB;PSMB3;PSMC4;PSMD2;FGF18;KDR;PSME1;LAMTOR2;FYN;CALM1 |
| 209 | Signaling by FGFR2 IIIa TM Homo sapiens R-HSA-8851708 | 6.69E-02 | 3.41E-01 | 3.56E+00 | 9.62E+00 | POLR2E;POLR2I;FGF2 |
| 210 | Translesion Synthesis by POLH Homo sapiens R-HSA-110320 | 6.69E-02 | 3.41E-01 | 3.56E+00 | 9.62E+00 | RFC1;UBB;RPA3 |
| 211 | PCNA-Dependent Long Patch Base Excision Repair Homo sapiens R-HSA-5651801 | 6.69E-02 | 3.41E-01 | 3.56E+00 | 9.62E+00 | POLD3;RFC1;RPA3 |
| 212 | Scavenging by Class A Receptors Homo sapiens R-HSA-3000480 | 6.69E-02 | 3.41E-01 | 3.56E+00 | 9.62E+00 | FTH1;APOE;FTL |
| 213 | Negative regulation of FGFR4 signaling Homo sapiens R-HSA-5654733 | 6.76E-02 | 3.43E-01 | 2.81E+00 | 7.58E+00 | KLB;UBB;FGF18;FGF2 |

| | | | | | | |
|---|---|---|---|---|---|---|
| 214 | Formation of annular gap junctions Homo sapiens R-HSA-196025 | 7.16E-02 | 3.55E-01 | 5.42E+00 | 1.43E+01 | DAB2;CLTB |
| 215 | Nef Mediated CD4 Down-regulation Homo sapiens R-HSA-167590 | 7.16E-02 | 3.55E-01 | 5.42E+00 | 1.43E+01 | AP2S1;AP2A1 |
| 216 | Degradation of cysteine and homocysteine Homo sapiens R-HSA-1614558 | 7.16E-02 | 3.55E-01 | 5.42E+00 | 1.43E+01 | TST;SLC25A10 |
| 217 | Pregnenolone biosynthesis Homo sapiens R-HSA-196108 | 7.16E-02 | 3.55E-01 | 5.42E+00 | 1.43E+01 | FDX1;AKR1B1 |
| 218 | Pentose phosphate pathway (hexose monophosphate shunt) Homo sapiens R-HSA-71336 | 7.16E-02 | 3.55E-01 | 5.42E+00 | 1.43E+01 | TKT;DERA |
| 219 | Lagging Strand Synthesis Homo sapiens R-HSA-69186 | 7.59E-02 | 3.75E-01 | 3.35E+00 | 8.63E+00 | POLD3;RFC1;RPA3 |
| 220 | Phospholipase C-mediated cascade; FGFR4 Homo sapiens R-HSA-5654228 | 7.92E-02 | 3.89E-01 | 2.32E+00 | 5.87E+00 | KLB;PRKAR2B;FGF18;CALM1;FGF2 |
| 221 | FGFR2 mutant receptor activation Homo sapiens R-HSA-1839126 | 8.13E-02 | 3.98E-01 | 2.62E+00 | 6.57E+00 | FGF18;POLR2E;POLR2I;FGF2 |
| 222 | Interleukin-2 signaling Homo sapiens R-HSA-451927 | 8.39E-02 | 3.98E-01 | 1.47E+00 | 3.63E+00 | STAT5A;KLB;RASGRF1;PEBP1;PHB;FGF2;PSMD8;PSMD9;UBB;PSMB3;PSMC4;PSMD2;FGF18;PSME1;LAMTOR2;FYN;CALM1;HAVCR2 |
| 223 | VEGFA-VEGFR2 Pathway Homo sapiens R-HSA-4420097 | 8.41E-02 | 3.98E-01 | 1.41E+00 | 3.48E+00 | KLB;PRKCB;RASGRF1;PEBP1;CYBA;PHB;SHB;BAIAP2;FGF2;VEGFA;PSMD8;PSMD9;UBB;PSMB3;PSMC4;PSMD2;FGF18;KDR;PSME1;LAMTOR2;FYN;CALM1 |
| 224 | Nef-mediates down modulation of cell surface receptors by recruiting them to clathrin adapters Homo sapiens R-HSA-164938 | 8.54E-02 | 3.98E-01 | 3.16E+00 | 7.78E+00 | AP2S1;AP2A1;AP1M1 |

| | | | | | | |
|---|---|---|---|---|---|---|
| 225 | Regulation of Insulin-like Growth Factor (IGF) transport and uptake by Insulin-like Growth Factor Binding Proteins (IGFBPs) Homo sapiens R-HSA-381426 | 8.54E-02 | 3.98E-01 | 3.16E+00 | 7.78E+00 | IGFBP5;PAPPA;PLG |
| 226 | Synthesis of Leukotrienes (LT) and Eoxins (EX) Homo sapiens R-HSA-2142691 | 8.54E-02 | 3.98E-01 | 3.16E+00 | 7.78E+00 | CYP4F2;DPEP1;PTGR1 |
| 227 | Aflatoxin activation and detoxification Homo sapiens R-HSA-5423646 | 8.54E-02 | 3.98E-01 | 3.16E+00 | 7.78E+00 | AKR7A2;AKR7A3;DPEP1 |
| 228 | Transcription-Coupled Nucleotide Excision Repair (TC-NER) Homo sapiens R-HSA-6781827 | 8.62E-02 | 3.98E-01 | 1.93E+00 | 4.72E+00 | POLD3;XAB2;RFC1;UBB;RPA3;POLR2E;POLR2I |
| 229 | Gap junction degradation Homo sapiens R-HSA-190873 | 8.65E-02 | 3.98E-01 | 4.74E+00 | 1.16E+01 | DAB2;CLTB |
| 230 | Removal of the Flap Intermediate from the C-strand Homo sapiens R-HSA-174437 | 8.65E-02 | 3.98E-01 | 4.74E+00 | 1.16E+01 | POLD3;RPA3 |
| 231 | Regulation of gene expression by Hypoxia-inducible Factor Homo sapiens R-HSA-1234158 | 8.65E-02 | 3.98E-01 | 4.74E+00 | 1.16E+01 | HIF1A;VEGFA |
| 232 | Tetrahydrobiopterin (BH4) synthesis, recycling, salvage and regulation Homo sapiens R-HSA-1474151 | 8.65E-02 | 3.98E-01 | 4.74E+00 | 1.16E+01 | GCHFR;CALM1 |
| 233 | Beta-oxidation of pristanoyl-CoA Homo sapiens R-HSA-389887 | 8.65E-02 | 3.98E-01 | 4.74E+00 | 1.16E+01 | ACOT8;ACOX2 |
| 234 | Calcitonin-like ligand receptors Homo sapiens R-HSA-419812 | 8.65E-02 | 3.98E-01 | 4.74E+00 | 1.16E+01 | CALCA;ADM |
| 235 | Urea cycle Homo sapiens R-HSA-70635 | 8.65E-02 | 3.98E-01 | 4.74E+00 | 1.16E+01 | ARG2;ASS1 |

| | | | | | | |
|---|---|---|---|---|---|---|
| 236 | EPHA-mediated growth cone collapse Homo sapiens R-HSA-3928663 | 8.87E-02 | 4.05E-01 | 2.53E+00 | 6.13E+00 | KDR;FYN;SHB;VEGFA |
| 237 | Purine metabolism Homo sapiens R-HSA-73847 | 8.87E-02 | 4.05E-01 | 2.53E+00 | 6.13E+00 | CAT;ADSSL1;GMPR;APRT |
| 238 | MAPK6/MAPK4 signaling Homo sapiens R-HSA-5687128 | 9.04E-02 | 4.10E-01 | 1.81E+00 | 4.35E+00 | PSMD8;PSMD9;UBB;PSMB3;PSMC4;PSMD2;CDC42EP3;PSME1 |
| 239 | mTORC1-mediated signalling Homo sapiens R-HSA-166208 | 9.54E-02 | 4.30E-01 | 3.00E+00 | 7.04E+00 | LAMTOR2;LAMTOR1;LAMTOR5 |
| 240 | Telomere C-strand (Lagging Strand) Synthesis Homo sapiens R-HSA-174417 | 9.54E-02 | 4.30E-01 | 3.00E+00 | 7.04E+00 | POLD3;RFC1;RPA3 |
| 241 | MAPK1/MAPK3 signaling Homo sapiens R-HSA-5684996 | 9.91E-02 | 4.44E-01 | 1.44E+00 | 3.34E+00 | KLB;RASGRF1;PEBP1;PHB;FGF2;PSMD8;PSMD9;UBB;PSMB3;PSMC4;PSMD2;FGF18;PSME1;LAMTOR2;FYN;CALM1;IL6R |
| 242 | PI3K Cascade Homo sapiens R-HSA-109704 | 1.01E-01 | 4.51E-01 | 1.85E+00 | 4.24E+00 | KLB;CAB39L;FGF18;LAMTOR2;LAMTOR1;FGF2;LAMTOR5 |
| 243 | Processive synthesis on the C-strand of the telomere Homo sapiens R-HSA-174414 | 1.02E-01 | 4.52E-01 | 4.22E+00 | 9.61E+00 | POLD3;RPA3 |
| 244 | Recruitment of NuMA to mitotic centrosomes Homo sapiens R-HSA-380320 | 1.02E-01 | 4.52E-01 | 4.22E+00 | 9.61E+00 | TUBGCP2;TUBG1 |
| 245 | Tryptophan catabolism Homo sapiens R-HSA-71240 | 1.02E-01 | 4.52E-01 | 4.22E+00 | 9.61E+00 | AFMID;ACMSD |
| 246 | EPH-Ephrin signaling Homo sapiens R-HSA-2682334 | 1.04E-01 | 4.54E-01 | 1.75E+00 | 3.96E+00 | ARPC1B;CLTB;KDR;AP2S1;AP2A1;FYN;SHB;VEGFA |
| 247 | Formation of the ternary complex, and subsequently, the 43S complex Homo sapiens R-HSA-72695 | 1.04E-01 | 4.54E-01 | 2.11E+00 | 4.77E+00 | RPS28;RPS27;RPS29;RPS4Y1;RPS21 |
| 248 | Signaling by Leptin Homo sapiens R-HSA-2586552 | 1.05E-01 | 4.54E-01 | 1.43E+00 | 3.23E+00 | STAT5A;KLB;RASGRF1;PEBP1;PHB;FGF2;PSMD8;PSMD9;UBB;PSMB3;PSMC4;PSMD2;FGF18;PSME1;LAMTOR2;FYN;CALM1 |
| 249 | The canonical retinoid cycle in rods (twilight vision) Homo sapiens R-HSA-2453902 | 1.06E-01 | 4.54E-01 | 2.85E+00 | 6.39E+00 | RDH10;ABCA4;NAPEPLD |

| | | | | | | |
|---|---|---|---|---|---|---|
| 250 | SHC-mediated cascade:FGFR4 Homo sapiens R-HSA-5654719 | 1.06E-01 | 4.54E-01 | 2.85E+00 | 6.39E+00 | KLB;FGF18;FGF2 |
| 251 | APC/C:Cdc20 mediated degradation of Cyclin B Homo sapiens R-HSA-174048 | 1.06E-01 | 4.54E-01 | 2.85E+00 | 6.39E+00 | UBB;CDC26;ANAPC11 |
| 252 | Branched-chain amino acid catabolism Homo sapiens R-HSA-70895 | 1.06E-01 | 4.54E-01 | 2.85E+00 | 6.39E+00 | AUH;IVD;BBOX1 |
| 253 | Metabolism of fat-soluble vitamins Homo sapiens R-HSA-6806667 | 1.11E-01 | 4.75E-01 | 2.06E+00 | 4.53E+00 | APOM;AKR1C1;APOC2;APOC3;APOE |
| 254 | Translesion synthesis by Y family DNA polymerases bypasses lesions on DNA template Homo sapiens R-HSA-110313 | 1.13E-01 | 4.79E-01 | 2.30E+00 | 5.02E+00 | POLD3;UBB;RFC1;RPA3 |
| 255 | Endogenous sterols Homo sapiens R-HSA-211976 | 1.17E-01 | 4.89E-01 | 2.71E+00 | 5.83E+00 | FDX1;NR1H4;CYP46A1 |
| 256 | Pyrimidine metabolism Homo sapiens R-HSA-73848 | 1.17E-01 | 4.89E-01 | 2.71E+00 | 5.83E+00 | AGXT2;TK1;UPB1 |
| 257 | Degradation of beta-catenin by the destruction complex Homo sapiens R-HSA-195253 | 1.17E-01 | 4.89E-01 | 1.77E+00 | 3.81E+00 | PSMD8;PSMD9;UBB;PSMB3;PSMC4;PSMD2;PSME1 |
| 258 | Macroautophagy Homo sapiens R-HSA-1632852 | 1.19E-01 | 4.89E-01 | 1.87E+00 | 3.98E+00 | WIPI1;LAMTOR2;WIPI2;LAMTOR1;WDR45;LAMTOR5 |
| 259 | FGFR3 mutant receptor activation Homo sapiens R-HSA-2033514 | 1.19E-01 | 4.89E-01 | 3.79E+00 | 8.08E+00 | FGF18;FGF2 |
| 260 | Signaling by activated point mutants of FGFR3 Homo sapiens R-HSA-1839130 | 1.19E-01 | 4.89E-01 | 3.79E+00 | 8.08E+00 | FGF18;FGF2 |
| 261 | Miscellaneous substrates Homo sapiens R-HSA-211958 | 1.19E-01 | 4.89E-01 | 3.79E+00 | 8.08E+00 | CYP2S1;CYP4F2 |
| 262 | Pyrimidine catabolism Homo sapiens R-HSA-73621 | 1.19E-01 | 4.89E-01 | 3.79E+00 | 8.08E+00 | AGXT2;UPB1 |

| | | | | | | |
|---|---|---|---|---|---|---|
| 263 | Lysine catabolism Homo sapiens R-HSA-71064 | 1.19E-01 | 4.89E-01 | 3.79E+00 | 8.08E+00 | GCDH;PIPOX |
| 264 | CLEC7A (Dectin-1) signaling Homo sapiens R-HSA-5607764 | 1.24E-01 | 5.00E-01 | 1.67E+00 | 3.49E+00 | PSMD8;PSMD9;UBB;PSMB3;PSMC4;PSMD2;PSME1;CALM1 |
| 265 | Downstream TCR signaling Homo sapiens R-HSA-202424 | 1.24E-01 | 5.00E-01 | 1.67E+00 | 3.49E+00 | PSMD8;PSMD9;UBB;PSMB3;PSMC4;PSMD2;PSME1;HLA-DPB1 |
| 266 | IRS-mediated signalling Homo sapiens R-HSA-112399 | 1.24E-01 | 5.00E-01 | 1.36E+00 | 2.85E+00 | KLB;CAB39L;RASGRF1;PEBP1;PHB;FGF2;PSMD8;PSMD9;UBB;PSMB3;PSMC4;PSMD2;FGF18;PSME1;LAMTOR2;FYN;LAMTOR1;CALM1;LAMTOR5 |
| 267 | APC-Cdc20 mediated degradation of Nek2A Homo sapiens R-HSA-179409 | 1.28E-01 | 5.00E-01 | 2.59E+00 | 5.32E+00 | UBB;CDC26;ANAPC11 |
| 268 | ADP signalling through P2Y purinoceptor 1 Homo sapiens R-HSA-418592 | 1.28E-01 | 5.00E-01 | 2.59E+00 | 5.32E+00 | GNG4;PLA2G4A;GNG11 |
| 269 | Synthesis of IP3 and IP4 in the cytosol Homo sapiens R-HSA-1855204 | 1.28E-01 | 5.00E-01 | 2.59E+00 | 5.32E+00 | ITPKB;PLCB4;CALM1 |
| 270 | Synthesis and interconversion of nucleotide di- and triphosphates Homo sapiens R-HSA-499943 | 1.28E-01 | 5.00E-01 | 2.59E+00 | 5.32E+00 | AK2;GLRX;CTPS1 |
| 271 | mTOR signalling Homo sapiens R-HSA-165159 | 1.30E-01 | 5.00E-01 | 2.17E+00 | 4.43E+00 | CAB39L;LAMTOR2;LAMTOR1;LAMTOR5 |
| 272 | Dual Incision in GG-NER Homo sapiens R-HSA-5696400 | 1.30E-01 | 5.00E-01 | 2.17E+00 | 4.43E+00 | POLD3;UBB;RFC1;RPA3 |
| 273 | Golgi Associated Vesicle Biogenesis Homo sapiens R-HSA-432722 | 1.33E-01 | 5.00E-01 | 1.94E+00 | 3.91E+00 | RAB5C;BLOC1S1;FTH1;FTL;AP1M1 |
| 274 | Insulin receptor signalling cascade Homo sapiens R-HSA-74751 | 1.33E-01 | 5.00E-01 | 1.35E+00 | 2.72E+00 | KLB;CAB39L;RASGRF1;PEBP1;PHB;FGF2;PSMD8;PSMD9;UBB;PSMB3;PSMC4;PSMD2;FGF18;PSME1;LAMTOR2;FYN;LAMTOR1;CALM1;LAMTOR5 |
| 275 | GRB2 events in EGFR signaling Homo sapiens R-HSA-179812 | 1.33E-01 | 5.00E-01 | 1.39E+00 | 2.80E+00 | KLB;RASGRF1;PEBP1;PHB;FGF2;PSMD8;PSMD9;UBB;PSMB3;PSMC4;PSMD2;FGF18;PSME1;LAMTOR2;FYN;CALM1 |
| 276 | SHC1 events in EGFR signaling Homo sapiens R-HSA-180336 | 1.33E-01 | 5.00E-01 | 1.39E+00 | 2.80E+00 | KLB;RASGRF1;PEBP1;PHB;FGF2;PSMD8;PSMD9;UBB;PSMB3;PSMC4;PSMD2;FGF18;PSME1;LAMTOR2;FYN;CALM1 |

| | | | | | | |
|---|---|---|---|---|---|---|
| 277 | SOS-mediated signalling Homo sapiens R-HSA-112412 | 1.33E-01 | 5.00E-01 | 1.39E+00 | 2.80E+00 | KLB;RASGRF1;PEBP1;PHB;FGF2;PSMD8;PSMD9;UBB;PSMB3;PSMC4;PSMD2;FGF18;PSME1;LAMTOR2;FYN;CALM1 |
| 278 | SHC1 events in ERBB4 signaling Homo sapiens R-HSA-1250347 | 1.33E-01 | 5.00E-01 | 1.39E+00 | 2.80E+00 | KLB;RASGRF1;PEBP1;PHB;FGF2;PSMD8;PSMD9;UBB;PSMB3;PSMC4;PSMD2;FGF18;PSME1;LAMTOR2;FYN;CALM1 |
| 279 | RAF/MAP kinase cascade Homo sapiens R-HSA-5673001 | 1.33E-01 | 5.00E-01 | 1.39E+00 | 2.80E+00 | KLB;RASGRF1;PEBP1;PHB;FGF2;PSMD8;PSMD9;UBB;PSMB3;PSMC4;PSMD2;FGF18;PSME1;LAMTOR2;FYN;CALM1 |
| 280 | Cell surface interactions at the vascular wall Homo sapiens R-HSA-202733 | 1.34E-01 | 5.00E-01 | 1.63E+00 | 3.28E+00 | GRB7;SLC7A7;SLC7A9;ATP1B3;FYN;MIF;ATP1B1;PF4 |
| 281 | Retrograde neurotrophin signalling Homo sapiens R-HSA-177504 | 1.36E-01 | 5.00E-01 | 3.45E+00 | 6.88E+00 | AP2S1;AP2A1 |
| 282 | FGFRL1 modulation of FGFR1 signaling Homo sapiens R-HSA-5658623 | 1.36E-01 | 5.00E-01 | 3.45E+00 | 6.88E+00 | FGF18;FGF2 |
| 283 | FGFR2c ligand binding and activation Homo sapiens R-HSA-190375 | 1.36E-01 | 5.00E-01 | 3.45E+00 | 6.88E+00 | FGF18;FGF2 |
| 284 | FGFR3c ligand binding and activation Homo sapiens R-HSA-190372 | 1.36E-01 | 5.00E-01 | 3.45E+00 | 6.88E+00 | FGF18;FGF2 |
| 285 | FGFR3 ligand binding and activation Homo sapiens R-HSA-190239 | 1.36E-01 | 5.00E-01 | 3.45E+00 | 6.88E+00 | FGF18;FGF2 |
| 286 | Synthesis of PE Homo sapiens R-HSA-1483213 | 1.36E-01 | 5.00E-01 | 3.45E+00 | 6.88E+00 | ETNK2;LPIN2 |
| 287 | alpha-linolenic (omega3) and linoleic (omega6) acid metabolism Homo sapiens R-HSA-2046104 | 1.36E-01 | 5.00E-01 | 3.45E+00 | 6.88E+00 | ACOT8;ACAA1 |
| 288 | alpha-linolenic acid (ALA) metabolism Homo sapiens R-HSA-2046106 | 1.36E-01 | 5.00E-01 | 3.45E+00 | 6.88E+00 | ACOT8;ACAA1 |
| 289 | Purine salvage Homo sapiens R-HSA-74217 | 1.36E-01 | 5.00E-01 | 3.45E+00 | 6.88E+00 | GMPR;APRT |

| | | | | | | |
|---|---|---|---|---|---|---|
| 290 | IGF1R signaling cascade Homo sapiens R-HSA-2428924 | 1.36E-01 | 5.00E-01 | 1.34E+00 | 2.68E+00 | KLB;CAB39L;RASGRF1;PEBP1;PHB;FGF2;PSMD8;PSMD9;UBB;PSMB3;PSMC4;PSMD2;FGF18;PSME1;LAMTOR2;FYN;LAMTOR1;CALM1;LAMTOR5 |
| 291 | Signaling by Type 1 Insulin-like Growth Factor 1 Receptor (IGF1R) Homo sapiens R-HSA-2404192 | 1.36E-01 | 5.00E-01 | 1.34E+00 | 2.68E+00 | KLB;CAB39L;RASGRF1;PEBP1;PHB;FGF2;PSMD8;PSMD9;UBB;PSMB3;PSMC4;PSMD2;FGF18;PSME1;LAMTOR2;FYN;LAMTOR1;CALM1;LAMTOR5 |
| 292 | IRS-related events triggered by IGF1R Homo sapiens R-HSA-2428928 | 1.36E-01 | 5.00E-01 | 1.34E+00 | 2.68E+00 | KLB;CAB39L;RASGRF1;PEBP1;PHB;FGF2;PSMD8;PSMD9;UBB;PSMB3;PSMC4;PSMD2;FGF18;PSME1;LAMTOR2;FYN;LAMTOR1;CALM1;LAMTOR5 |
| 293 | FRS-mediated FGFR2 signaling Homo sapiens R-HSA-5654700 | 1.37E-01 | 5.00E-01 | 1.38E+00 | 2.75E+00 | KLB;RASGRF1;PEBP1;PHB;FGF2;PSMD8;PSMD9;UBB;PSMB3;PSMC4;PSMD2;FGF18;PSME1;LAMTOR2;FYN;CALM1 |
| 294 | FRS-mediated FGFR4 signaling Homo sapiens R-HSA-5654712 | 1.37E-01 | 5.00E-01 | 1.38E+00 | 2.75E+00 | KLB;RASGRF1;PEBP1;PHB;FGF2;PSMD8;PSMD9;UBB;PSMB3;PSMC4;PSMD2;FGF18;PSME1;LAMTOR2;FYN;CALM1 |
| 295 | FRS-mediated FGFR3 signaling Homo sapiens R-HSA-5654706 | 1.37E-01 | 5.00E-01 | 1.38E+00 | 2.75E+00 | KLB;RASGRF1;PEBP1;PHB;FGF2;PSMD8;PSMD9;UBB;PSMB3;PSMC4;PSMD2;FGF18;PSME1;LAMTOR2;FYN;CALM1 |
| 296 | FRS-mediated FGFR1 signaling Homo sapiens R-HSA-5654693 | 1.37E-01 | 5.00E-01 | 1.38E+00 | 2.75E+00 | KLB;RASGRF1;PEBP1;PHB;FGF2;PSMD8;PSMD9;UBB;PSMB3;PSMC4;PSMD2;FGF18;PSME1;LAMTOR2;FYN;CALM1 |
| 297 | trans-Golgi Network Vesicle Budding Homo sapiens R-HSA-199992 | 1.38E-01 | 5.02E-01 | 1.78E+00 | 3.52E+00 | RAB5C;BLOC1S1;FTH1;CLTB;FTL;AP1M1 |
| 298 | Clathrin derived vesicle budding Homo sapiens R-HSA-421837 | 1.38E-01 | 5.02E-01 | 1.78E+00 | 3.52E+00 | RAB5C;BLOC1S1;FTH1;CLTB;FTL;AP1M1 |
| 299 | PKB-mediated events Homo sapiens R-HSA-109703 | 1.39E-01 | 5.03E-01 | 2.11E+00 | 4.16E+00 | CAB39L;LAMTOR2;LAMTOR1;LAMTOR5 |
| 300 | Regulation of mRNA stability by proteins that bind AU-rich elements Homo sapiens R-HSA-450531 | 1.40E-01 | 5.05E-01 | 1.68E+00 | 3.31E+00 | PSMD8;PSMD9;UBB;PSMB3;PSMC4;PSMD2;PSME1 |
| 301 | Ion transport by P-type ATPases Homo sapiens R-HSA-936837 | 1.41E-01 | 5.05E-01 | 1.90E+00 | 3.72E+00 | FXYD4;FXYD2;ATP1B3;ATP1B1;CALM1 |

| | | | | | | |
|---|---|---|---|---|---|---|
| 302 | Gastrin-CREB signalling pathway via PKC and MAPK Homo sapiens R-HSA-881907 | 1.41E-01 | 5.06E-01 | 1.27E+00 | 2.49E+00 | KLB;RGS18;PTGER1;RASGRF1;PEBP1;PHB;NTS;FGF2;PSMD8;PSMD9;RGS3;UBB;PSMB3;GNG4;PSMD2;FYN;PRKCH;F2R;GNG11;PLCB4;PSMC4;FGF18;PSME1;LAMTOR2;AVP;CALM1;MGLL |
| 303 | Signaling by Hedgehog Homo sapiens R-HSA-5358351 | 1.46E-01 | 5.19E-01 | 1.51E+00 | 2.91E+00 | PSMD8;PSMD9;UBB;PRKAR2B;PSMB3;PSMC4;PSMD2;BOC;PSME1;FUZ |
| 304 | Selenocysteine synthesis Homo sapiens R-HSA-2408557 | 1.46E-01 | 5.19E-01 | 1.66E+00 | 3.20E+00 | RPS28;RPS27;RPS29;SARS;RPL38;RPS4Y1;RPS21 |
| 305 | ARMS-mediated activation Homo sapiens R-HSA-170984 | 1.48E-01 | 5.19E-01 | 1.36E+00 | 2.61E+00 | KLB;RASGRF1;PEBP1;PHB;FGF2;PSMD8;PSMD9;UBB;PSMB3;PSMC4;PSMD2;FGF18;PSME1;LAMTOR2;FYN;CALM1 |
| 306 | Signalling to p38 via RIT and RIN Homo sapiens R-HSA-187706 | 1.48E-01 | 5.19E-01 | 1.36E+00 | 2.61E+00 | KLB;RASGRF1;PEBP1;PHB;FGF2;PSMD8;PSMD9;UBB;PSMB3;PSMC4;PSMD2;FGF18;PSME1;LAMTOR2;FYN;CALM1 |
| 307 | Frs2-mediated activation Homo sapiens R-HSA-170968 | 1.51E-01 | 5.19E-01 | 1.36E+00 | 2.57E+00 | KLB;RASGRF1;PEBP1;PHB;FGF2;PSMD8;PSMD9;UBB;PSMB3;PSMC4;PSMD2;FGF18;PSME1;LAMTOR2;FYN;CALM1 |
| 308 | RORA activates gene expression Homo sapiens R-HSA-1368082 | 1.51E-01 | 5.19E-01 | 2.37E+00 | 4.48E+00 | RORA;NR1D1;NPAS2 |
| 309 | TP53 regulates transcription of several additional cell death genes whose specific roles in p53-dependent apoptosis remain uncertain Homo sapiens R-HSA-6803205 | 1.54E-01 | 5.19E-01 | 3.16E+00 | 5.92E+00 | BCL6;NDRG1 |
| 310 | Polymerase switching on the C-strand of the telomere Homo sapiens R-HSA-174411 | 1.54E-01 | 5.19E-01 | 3.16E+00 | 5.92E+00 | POLD3;RFC1 |
| 311 | Polymerase switching Homo sapiens R-HSA-69091 | 1.54E-01 | 5.19E-01 | 3.16E+00 | 5.92E+00 | POLD3;RFC1 |
| 312 | Leading Strand Synthesis Homo sapiens R-HSA-69109 | 1.54E-01 | 5.19E-01 | 3.16E+00 | 5.92E+00 | POLD3;RFC1 |
| 313 | Removal of the Flap Intermediate Homo sapiens R-HSA-69166 | 1.54E-01 | 5.19E-01 | 3.16E+00 | 5.92E+00 | POLD3;RPA3 |

| | | | | | | |
|---|---|---|---|---|---|---|
| 314 | Bile salt and organic anion SLC transporters Homo sapiens R-HSA-425471 | 1.54E-01 | 5.19E-01 | 3.16E+00 | 5.92E+00 | SLC47A2;SLC13A2 |
| 315 | SEMA3A-Plexin repulsion signaling by inhibiting Integrin adhesion Homo sapiens R-HSA-399955 | 1.54E-01 | 5.19E-01 | 3.16E+00 | 5.92E+00 | FYN;PLXNA4 |
| 316 | Methylation Homo sapiens R-HSA-156581 | 1.54E-01 | 5.19E-01 | 3.16E+00 | 5.92E+00 | AHCY;GSTO1 |
| 317 | Synthesis of bile acids and bile salts via 24-hydroxycholesterol Homo sapiens R-HSA-193775 | 1.54E-01 | 5.19E-01 | 3.16E+00 | 5.92E+00 | AKR1C1;CYP46A1 |
| 318 | Nicotinate metabolism Homo sapiens R-HSA-196807 | 1.54E-01 | 5.19E-01 | 3.16E+00 | 5.92E+00 | NAPRT;QPRT |
| 319 | Glutathione synthesis and recycling Homo sapiens R-HSA-174403 | 1.54E-01 | 5.19E-01 | 3.16E+00 | 5.92E+00 | GGCT;CNDP2 |
| 320 | Regulation of pyruvate dehydrogenase (PDH) complex Homo sapiens R-HSA-204174 | 1.54E-01 | 5.19E-01 | 3.16E+00 | 5.92E+00 | PDHB;PDK2 |
| 321 | Ribosomal scanning and start codon recognition Homo sapiens R-HSA-72702 | 1.57E-01 | 5.25E-01 | 1.83E+00 | 3.39E+00 | RPS28;RPS27;RPS29;RPS4Y1;RPS21 |
| 322 | Translation initiation complex formation Homo sapiens R-HSA-72649 | 1.57E-01 | 5.25E-01 | 1.83E+00 | 3.39E+00 | RPS28;RPS27;RPS29;RPS4Y1;RPS21 |
| 323 | Infectious disease Homo sapiens R-HSA-5663205 | 1.58E-01 | 5.25E-01 | 1.28E+00 | 2.37E+00 | AP2A1;RPS4Y1;PSMD8;PSMD9;MSRA;RPS28;RPS27;UBB;RPS29;PSMB3;PSMC4;PSMD2;PSME1;AP2S1;POLR2E;RPL38;FYN;CALM1;SLC25A5;POLR2I;RPS21;AP1M1 |
| 324 | Glucose transport Homo sapiens R-HSA-70153 | 1.58E-01 | 5.25E-01 | 2.00E+00 | 3.69E+00 | SLC2A1;SLC2A3;SLC37A4;HK2 |
| 325 | Fatty Acyl-CoA Biosynthesis Homo sapiens R-HSA-75105 | 1.58E-01 | 5.25E-01 | 2.00E+00 | 3.69E+00 | ACOT8;ELOVL4;ACOT13;ACSF2 |
| 326 | Prolonged ERK activation events Homo sapiens R-HSA-169893 | 1.59E-01 | 5.26E-01 | 1.35E+00 | 2.48E+00 | KLB;RASGRF1;PEBP1;PHB;FGF2;PSMD8;PSMD9;UBB;PSMB3;PSMC4;PSMD2;FGF18;PSME1;LAMTOR2;FYN;CALM1 |

| | | | | | | |
|---|---|---|---|---|---|---|
| 327 | Gap junction trafficking Homo sapiens R-HSA-190828 | 1.64E-01 | 5.37E-01 | 2.28E+00 | 4.12E+00 | GJC1;DAB2;CLTB |
| 328 | Extension of Telomeres Homo sapiens R-HSA-180786 | 1.64E-01 | 5.37E-01 | 2.28E+00 | 4.12E+00 | POLD3;RFC1;RPA3 |
| 329 | Ca-dependent events Homo sapiens R-HSA-111996 | 1.64E-01 | 5.37E-01 | 2.28E+00 | 4.12E+00 | PRKAR2B;PLA2G4A;CALM1 |
| 330 | Interleukin-3, 5 and GM-CSF signaling Homo sapiens R-HSA-512988 | 1.64E-01 | 5.37E-01 | 1.33E+00 | 2.40E+00 | STAT5A;KLB;RASGRF1;PEBP1;PHB;FGF2;PSMD8;PSMD9;UBB;PSMB3;PSMC4;PSMD2;FGF18;PSME1;LAMTOR2;FYN;CALM1 |
| 331 | Activation of the mRNA upon binding of the cap-binding complex and eIFs, and subsequent binding to 43S Homo sapiens R-HSA-72662 | 1.65E-01 | 5.37E-01 | 1.79E+00 | 3.23E+00 | RPS28;RPS27;RPS29;RPS4Y1;RPS21 |
| 332 | C-type lectin receptors (CLRs) Homo sapiens R-HSA-5621481 | 1.65E-01 | 5.37E-01 | 1.50E+00 | 2.70E+00 | PSMD8;PSMD9;UBB;PSMB3;PSMC4;PSMD2;PSME1;FYN;CALM1 |
| 333 | TP53 Regulates Transcription of Cell Death Genes Homo sapiens R-HSA-5633008 | 1.68E-01 | 5.41E-01 | 1.95E+00 | 3.48E+00 | BNIP3L;PRELID1;BCL6;NDRG1 |
| 334 | Signaling by FGFR2 in disease Homo sapiens R-HSA-5655253 | 1.68E-01 | 5.41E-01 | 1.95E+00 | 3.48E+00 | FGF18;POLR2E;POLR2I;FGF2 |
| 335 | Downstream signaling events of B Cell Receptor (BCR) Homo sapiens R-HSA-1168372 | 1.68E-01 | 5.41E-01 | 1.38E+00 | 2.46E+00 | KLB;PRKCB;FGF2;PSMD8;PSMD9;UBB;PSMB3;PSMC4;PSMD2;FGF18;PSME1;FYN;IER3 |
| 336 | Interleukin receptor SHC signaling Homo sapiens R-HSA-912526 | 1.70E-01 | 5.41E-01 | 1.33E+00 | 2.35E+00 | KLB;RASGRF1;PEBP1;PHB;FGF2;PSMD8;PSMD9;UBB;PSMB3;PSMC4;PSMD2;FGF18;PSME1;LAMTOR2;FYN;CALM1 |
| 337 | Depolymerisation of the Nuclear Lamina Homo sapiens R-HSA-4419969 | 1.72E-01 | 5.41E-01 | 2.92E+00 | 5.14E+00 | PRKCB;LPIN2 |
| 338 | Processive synthesis on the lagging strand Homo sapiens R-HSA-69183 | 1.72E-01 | 5.41E-01 | 2.92E+00 | 5.14E+00 | POLD3;RPA3 |

| | | | | | | |
|---|---|---|---|---|---|---|
| 339 | Association of licensing factors with the pre-replicative complex Homo sapiens R-HSA-69298 | 1.72E-01 | 5.41E-01 | 2.92E+00 | 5.14E+00 | UBB;GMNN |
| 340 | Glycogen synthesis Homo sapiens R-HSA-3322077 | 1.72E-01 | 5.41E-01 | 2.92E+00 | 5.14E+00 | UBB;PGM1 |
| 341 | Glycogen breakdown (glycogenolysis) Homo sapiens R-HSA-70221 | 1.72E-01 | 5.41E-01 | 2.92E+00 | 5.14E+00 | CALM1;PGM1 |
| 342 | Recycling of bile acids and salts Homo sapiens R-HSA-159418 | 1.72E-01 | 5.41E-01 | 2.92E+00 | 5.14E+00 | SLCO1A2;NR1H4 |
| 343 | Synthesis of bile acids and bile salts via 27-hydroxycholesterol Homo sapiens R-HSA-193807 | 1.72E-01 | 5.41E-01 | 2.92E+00 | 5.14E+00 | AKR1C1;NR1H4 |
| 344 | Signalling to RAS Homo sapiens R-HSA-167044 | 1.74E-01 | 5.48E-01 | 1.32E+00 | 2.31E+00 | KLB;RASGRF1;PEBP1;PHB;FGF2;PSMD8;PSMD9;UBB;PSMB3;PSMC4;PSMD2;FGF18;PSME1;LAMTOR2;FYN;CALM1 |
| 345 | HSF1 activation Homo sapiens R-HSA-3371511 | 1.76E-01 | 5.50E-01 | 2.19E+00 | 3.80E+00 | UBB;RPA3;HDAC6 |
| 346 | Negative regulation of FGFR3 signaling Homo sapiens R-HSA-5654732 | 1.76E-01 | 5.50E-01 | 2.19E+00 | 3.80E+00 | UBB;FGF18;FGF2 |
| 347 | Phospholipase C-mediated cascade; FGFR3 Homo sapiens R-HSA-5654227 | 1.78E-01 | 5.50E-01 | 1.90E+00 | 3.28E+00 | PRKAR2B;FGF18;CALM1;FGF2 |
| 348 | PLC beta mediated events Homo sapiens R-HSA-112043 | 1.78E-01 | 5.50E-01 | 1.90E+00 | 3.28E+00 | PLCB4;PRKAR2B;PLA2G4A;CALM1 |
| 349 | Inositol phosphate metabolism Homo sapiens R-HSA-1483249 | 1.78E-01 | 5.50E-01 | 1.90E+00 | 3.28E+00 | ITPKB;PLCB4;CALM1;IP6K3 |
| 350 | MAPK family signaling cascades Homo sapiens R-HSA-5683057 | 1.84E-01 | 5.67E-01 | 1.29E+00 | 2.18E+00 | KLB;RASGRF1;PEBP1;PHB;FGF2;PSMD8;PSMD9;UBB;PSMB3;PSMC4;PSMD2;CDC42EP3;FGF18;PSME1;LAMTOR2;FYN;CALM1;IL6R |
| 351 | Integration of energy metabolism Homo sapiens R-HSA-163685 | 1.87E-01 | 5.71E-01 | 1.49E+00 | 2.50E+00 | PKLR;PRKAR2B;GNG4;SLC2A1;ARL2;TKT;SLC25A5;GNG11 |
| 352 | G-protein mediated events Homo sapiens R-HSA-112040 | 1.88E-01 | 5.71E-01 | 1.85E+00 | 3.10E+00 | PLCB4;PRKAR2B;PLA2G4A;CALM1 |

| | | | | | | |
|---|---|---|---|---|---|---|
| 353 | Amine compound SLC transporters Homo sapiens R-HSA-425428 | 1.89E-01 | 5.71E-01 | 2.11E+00 | 3.51E+00 | SLC6A6;SLC14A2;SLC6A12 |
| 354 | Gap junction trafficking and regulation Homo sapiens R-HSA-157858 | 1.89E-01 | 5.71E-01 | 2.11E+00 | 3.51E+00 | GJC1;DAB2;CLTB |
| 355 | Activation of Kainate Receptors upon glutamate binding Homo sapiens R-HSA-451326 | 1.89E-01 | 5.71E-01 | 2.11E+00 | 3.51E+00 | GNG4;CALM1;GNG11 |
| 356 | Glucagon-type ligand receptors Homo sapiens R-HSA-420092 | 1.89E-01 | 5.71E-01 | 2.11E+00 | 3.51E+00 | GIPR;GNG4;GNG11 |
| 357 | Metabolism of steroid hormones Homo sapiens R-HSA-196071 | 1.89E-01 | 5.71E-01 | 2.11E+00 | 3.51E+00 | FDX1;AKR1B1;HSD17B14 |
| 358 | MAP3K8 (TPL2)-dependent MAPK1/3 activation Homo sapiens R-HSA-5684264 | 1.90E-01 | 5.71E-01 | 2.71E+00 | 4.50E+00 | UBB;TNIP2 |
| 359 | CRMPs in Sema3A signaling Homo sapiens R-HSA-399956 | 1.90E-01 | 5.71E-01 | 2.71E+00 | 4.50E+00 | FYN;PLXNA4 |
| 360 | Sema3A PAK dependent Axon repulsion Homo sapiens R-HSA-399954 | 1.90E-01 | 5.71E-01 | 2.71E+00 | 4.50E+00 | FYN;PLXNA4 |
| 361 | Selenoamino acid metabolism Homo sapiens R-HSA-2408522 | 1.94E-01 | 5.80E-01 | 1.47E+00 | 2.42E+00 | RPS28;RPS27;AHCY;RPS29;SARS;RPL38;RPS4Y1;RPS21 |
| 362 | DNA Damage Bypass Homo sapiens R-HSA-73893 | 1.98E-01 | 5.90E-01 | 1.81E+00 | 2.93E+00 | POLD3;UBB;RFC1;RPA3 |
| 363 | G alpha (z) signalling events Homo sapiens R-HSA-418597 | 1.98E-01 | 5.90E-01 | 1.81E+00 | 2.93E+00 | PRKCH;GNG4;PRKCB;GNG11 |
| 364 | Cytochrome P450 - arranged by substrate type Homo sapiens R-HSA-211897 | 1.99E-01 | 5.90E-01 | 1.66E+00 | 2.69E+00 | CYP4F2;CYP2S1;FDX1;NR1H4;CYP46A1 |
| 365 | TNFR2 non-canonical NF-kB pathway Homo sapiens R-HSA-5668541 | 1.99E-01 | 5.90E-01 | 1.51E+00 | 2.44E+00 | PSMD8;PSMD9;UBB;PSMB3;PSMC4;PSMD2;PSME1 |
| 366 | G1/S Transition Homo sapiens R-HSA-69206 | 2.00E-01 | 5.91E-01 | 1.46E+00 | 2.35E+00 | PSMD8;PSMD9;UBB;PSMB3;PSMC4;PSMD2;RPA3;PSME1 |

| | | | | | | |
|---|---|---|---|---|---|---|
| 367 | Trafficking of AMPA receptors Homo sapiens R-HSA-399719 | 2.02E-01 | 5.93E-01 | 2.03E+00 | 3.25E+00 | PRKCB;AP2S1;AP2A1 |
| 368 | Glutamate Binding, Activation of AMPA Receptors and Synaptic Plasticity Homo sapiens R-HSA-399721 | 2.02E-01 | 5.93E-01 | 2.03E+00 | 3.25E+00 | PRKCB;AP2S1;AP2A1 |
| 369 | Signalling to ERKs Homo sapiens R-HSA-187687 | 2.03E-01 | 5.95E-01 | 1.28E+00 | 2.05E+00 | KLB;RASGRF1;PEBP1;PHB;FGF2;PSMD8;PSMD9;UBB;PSMB3;PSMC4;PSMD2;FGF18;PSME1;LAMTOR2;FYN;CALM1 |
| 370 | Signaling by FGFR in disease Homo sapiens R-HSA-1226099 | 2.08E-01 | 6.05E-01 | 1.64E+00 | 2.57E+00 | STAT5A;FGF18;POLR2E;POLR2I;FGF2 |
| 371 | ABC transporters in lipid homeostasis Homo sapiens R-HSA-1369062 | 2.09E-01 | 6.05E-01 | 2.53E+00 | 3.96E+00 | ABCA12;ABCG1 |
| 372 | Activated point mutants of FGFR2 Homo sapiens R-HSA-2033519 | 2.09E-01 | 6.05E-01 | 2.53E+00 | 3.96E+00 | FGF18;FGF2 |
| 373 | Ras activation uopn Ca2+ infux through NMDA receptor Homo sapiens R-HSA-442982 | 2.09E-01 | 6.05E-01 | 2.53E+00 | 3.96E+00 | RASGRF1;CALM1 |
| 374 | Regulation of insulin secretion Homo sapiens R-HSA-422356 | 2.12E-01 | 6.13E-01 | 1.54E+00 | 2.38E+00 | GNG4;PRKAR2B;SLC2A1;ARL2;SLC25A5;GNG11 |
| 375 | DNA strand elongation Homo sapiens R-HSA-69190 | 2.15E-01 | 6.13E-01 | 1.96E+00 | 3.02E+00 | POLD3;RFC1;RPA3 |
| 376 | Signal amplification Homo sapiens R-HSA-392518 | 2.15E-01 | 6.13E-01 | 1.96E+00 | 3.02E+00 | GNG4;PLA2G4A;GNG11 |
| 377 | Thrombin signalling through proteinase activated receptors (PARs) Homo sapiens R-HSA-456926 | 2.15E-01 | 6.13E-01 | 1.96E+00 | 3.02E+00 | GNG4;F2R;GNG11 |
| 378 | EPH-ephrin mediated repulsion of cells Homo sapiens R-HSA-3928665 | 2.19E-01 | 6.13E-01 | 1.72E+00 | 2.62E+00 | CLTB;AP2S1;AP2A1;FYN |
| 379 | Ion channel transport Homo sapiens R-HSA-983712 | 2.20E-01 | 6.13E-01 | 1.30E+00 | 1.97E+00 | ATP6V1G1;ATP1B3;ATP1B1;TPCN1;CLCN7;FXYD4;UBB;FXYD2;ATP6V1E1;ATP6V0D1;CALM1;ATP6V0E2;ATP6V1F |

| | | | | | | |
|---|---|---|---|---|---|---|
| 380 | Opioid Signalling Homo sapiens R-HSA-111885 | 2.20E-01 | 6.13E-01 | 1.52E+00 | 2.30E+00 | PLCB4;GNG4;PRKAR2B;PLA2G4A;CALM1;GNG11 |
| 381 | HIV Infection Homo sapiens R-HSA-162906 | 2.26E-01 | 6.13E-01 | 1.28E+00 | 1.90E+00 | AP2A1;PSMD8;PSMD9;UBB;PSMB3;PSMC4;PSMD2;PSME1;AP2S1;POLR2E;FYN;SLC25A5;POLR2I;AP1M1 |
| 382 | Abacavir metabolism Homo sapiens R-HSA-2161541 | 2.27E-01 | 6.13E-01 | 4.74E+00 | 7.03E+00 | PCK1 |
| 383 | Negative regulation of TCF-dependent signaling by DVL-interacting proteins Homo sapiens R-HSA-5368598 | 2.27E-01 | 6.13E-01 | 4.74E+00 | 7.03E+00 | CCDC88C |
| 384 | Disinhibition of SNARE formation Homo sapiens R-HSA-114516 | 2.27E-01 | 6.13E-01 | 4.74E+00 | 7.03E+00 | PRKCB |
| 385 | Electric Transmission Across Gap Junctions Homo sapiens R-HSA-112303 | 2.27E-01 | 6.13E-01 | 4.74E+00 | 7.03E+00 | GJC1 |
| 386 | Transmission across Electrical Synapses Homo sapiens R-HSA-112307 | 2.27E-01 | 6.13E-01 | 4.74E+00 | 7.03E+00 | GJC1 |
| 387 | N-glycan trimming and elongation in the cis-Golgi Homo sapiens R-HSA-964739 | 2.27E-01 | 6.13E-01 | 4.74E+00 | 7.03E+00 | MAN1C1 |
| 388 | PTK6 Expression Homo sapiens R-HSA-8849473 | 2.27E-01 | 6.13E-01 | 4.74E+00 | 7.03E+00 | HIF1A |
| 389 | GRB7 events in ERBB2 signaling Homo sapiens R-HSA-1306955 | 2.27E-01 | 6.13E-01 | 4.74E+00 | 7.03E+00 | GRB7 |
| 390 | Cyclin B2 mediated events Homo sapiens R-HSA-157881 | 2.27E-01 | 6.13E-01 | 4.74E+00 | 7.03E+00 | CDC25B |
| 391 | Sodium-coupled sulphate, di- and tri-carboxylate transporters Homo sapiens R-HSA-433137 | 2.27E-01 | 6.13E-01 | 4.74E+00 | 7.03E+00 | SLC13A2 |
| 392 | Arachidonate production from DAG Homo sapiens R-HSA-426048 | 2.27E-01 | 6.13E-01 | 4.74E+00 | 7.03E+00 | MGLL |

| | | | | | | |
|---|---|---|---|---|---|---|
| 393 | Beta oxidation of hexanoyl-CoA to butanoyl-CoA Homo sapiens R-HSA-77350 | 2.27E-01 | 6.13E-01 | 4.74E+00 | 7.03E+00 | ECHS1 |
| 394 | Beta oxidation of lauroyl-CoA to decanoyl-CoA-CoA Homo sapiens R-HSA-77310 | 2.27E-01 | 6.13E-01 | 4.74E+00 | 7.03E+00 | ECHS1 |
| 395 | Propionyl-CoA catabolism Homo sapiens R-HSA-71032 | 2.27E-01 | 6.13E-01 | 4.74E+00 | 7.03E+00 | PCCB |
| 396 | Estrogen biosynthesis Homo sapiens R-HSA-193144 | 2.27E-01 | 6.13E-01 | 4.74E+00 | 7.03E+00 | HSD17B14 |
| 397 | The fatty acid cycling model Homo sapiens R-HSA-167826 | 2.27E-01 | 6.13E-01 | 4.74E+00 | 7.03E+00 | UCP2 |
| 398 | The proton buffering model Homo sapiens R-HSA-167827 | 2.27E-01 | 6.13E-01 | 4.74E+00 | 7.03E+00 | UCP2 |
| 399 | Mitochondrial Uncoupling Proteins Homo sapiens R-HSA-166187 | 2.27E-01 | 6.13E-01 | 4.74E+00 | 7.03E+00 | UCP2 |
| 400 | SHC-mediated cascade:FGFR3 Homo sapiens R-HSA-5654704 | 2.27E-01 | 6.13E-01 | 2.37E+00 | 3.51E+00 | FGF18;FGF2 |
| 401 | Cytosolic sulfonation of small molecules Homo sapiens R-HSA-156584 | 2.27E-01 | 6.13E-01 | 2.37E+00 | 3.51E+00 | SULT2B1;ABHD14B |
| 402 | Sialic acid metabolism Homo sapiens R-HSA-4085001 | 2.28E-01 | 6.13E-01 | 1.90E+00 | 2.80E+00 | CTSA;SLC17A5;NPL |
| 403 | Glucagon signaling in metabolic regulation Homo sapiens R-HSA-163359 | 2.28E-01 | 6.13E-01 | 1.90E+00 | 2.80E+00 | GNG4;PRKAR2B;GNG11 |
| 404 | Phospholipase C-mediated cascade; FGFR2 Homo sapiens R-HSA-5654221 | 2.30E-01 | 6.16E-01 | 1.69E+00 | 2.48E+00 | PRKAR2B;FGF18;CALM1;FGF2 |
| 405 | Triglyceride Biosynthesis Homo sapiens R-HSA-75109 | 2.36E-01 | 6.29E-01 | 1.56E+00 | 2.25E+00 | ACOT8;ELOVL4;ACOT13;LPIN2;ACSF2 |
| 406 | TCR signaling Homo sapiens R-HSA-202403 | 2.40E-01 | 6.40E-01 | 1.38E+00 | 1.97E+00 | PSMD8;PSMD9;UBB;PSMB3;PSMC4;PSMD2;PSME1;HLA-DPB1 |

| | | | | | | |
|---|---|---|---|---|---|---|
| 407 | Negative regulation of FGFR2 signaling Homo sapiens R-HSA-5654727 | 2.42E-01 | 6.40E-01 | 1.84E+00 | 2.60E+00 | UBB;FGF18;FGF2 |
| 408 | Striated Muscle Contraction Homo sapiens R-HSA-390522 | 2.42E-01 | 6.40E-01 | 1.84E+00 | 2.60E+00 | TMOD1;TNNC1;MYH8 |
| 409 | Viral mRNA Translation Homo sapiens R-HSA-192823 | 2.45E-01 | 6.40E-01 | 1.46E+00 | 2.05E+00 | RPS28;RPS27;RPS29;RPL38;RPS4Y1;RPS21 |
| 410 | Peptide chain elongation Homo sapiens R-HSA-156902 | 2.45E-01 | 6.40E-01 | 1.46E+00 | 2.05E+00 | RPS28;RPS27;RPS29;RPL38;RPS4Y1;RPS21 |
| 411 | Class B/2 (Secretin family receptors) Homo sapiens R-HSA-373080 | 2.45E-01 | 6.40E-01 | 1.46E+00 | 2.05E+00 | CALCA;GIPR;GNG4;WNT7B;ADM;GNG11 |
| 412 | Na+/Cl- dependent neurotransmitter transporters Homo sapiens R-HSA-442660 | 2.46E-01 | 6.40E-01 | 2.23E+00 | 3.13E+00 | SLC6A6;SLC6A12 |
| 413 | TP53 Regulates Transcription of Genes Involved in Cytochrome C Release Homo sapiens R-HSA-6803204 | 2.46E-01 | 6.40E-01 | 2.23E+00 | 3.13E+00 | BNIP3L;PRELID1 |
| 414 | Phosphorylation of the APC/C Homo sapiens R-HSA-176412 | 2.46E-01 | 6.40E-01 | 2.23E+00 | 3.13E+00 | CDC26;ANAPC11 |
| 415 | Conversion from APC/C:Cdc20 to APC/C:Cdh1 in late anaphase Homo sapiens R-HSA-176407 | 2.46E-01 | 6.40E-01 | 2.23E+00 | 3.13E+00 | CDC26;ANAPC11 |
| 416 | Prostacyclin signalling through prostacyclin receptor Homo sapiens R-HSA-392851 | 2.46E-01 | 6.40E-01 | 2.23E+00 | 3.13E+00 | GNG4;GNG11 |
| 417 | Signaling by EGFR Homo sapiens R-HSA-177929 | 2.48E-01 | 6.42E-01 | 1.19E+00 | 1.67E+00 | KLB;RASGRF1;PEBP1;AP2A1;EPS15L1;PHB;FGF2;PSMD8;PSMD9;UBB;PRKAR2B;PSMB3;PSMC4;PSMD2;FGF18;PSME1;AP2S1;LAMTOR2;FYN;CALM1;IER3 |
| 418 | Neurotransmitter Release Cycle Homo sapiens R-HSA-112310 | 2.52E-01 | 6.48E-01 | 1.61E+00 | 2.22E+00 | MAOA;SLC6A12;SLC38A2;GLS |
| 419 | Amyloid fiber formation Homo sapiens R-HSA-977225 | 2.55E-01 | 6.48E-01 | 1.51E+00 | 2.06E+00 | APCS;CALCA;NAT8;UBB;H2AFZ |

| | | | | | | |
|---|---|---|---|---|---|---|
| 420 | Lysosome Vesicle Biogenesis Homo sapiens R-HSA-432720 | 2.56E-01 | 6.48E-01 | 1.78E+00 | 2.43E+00 | BLOC1S1;CLTB;AP1M1 |
| 421 | MHC class II antigen presentation Homo sapiens R-HSA-2132295 | 2.59E-01 | 6.48E-01 | 1.38E+00 | 1.87E+00 | CTSA;CTSK;AP2S1;HLA-DPB1;CTSH;AP2A1;AP1M1 |
| 422 | NCAM signaling for neurite out-growth Homo sapiens R-HSA-375165 | 2.62E-01 | 6.48E-01 | 1.22E+00 | 1.63E+00 | KLB;RASGRF1;PEBP1;PHB;FGF2;PSMD8;PSMD9;UBB;PSMB3;PSMC4;PSMD2;FGF18;PSME1;LAMTOR2;FYN;CALM1 |
| 423 | mRNA Splicing - Minor Pathway Homo sapiens R-HSA-72165 | 2.63E-01 | 6.48E-01 | 1.58E+00 | 2.11E+00 | SNRPG;POLR2E;TXNL4A;POLR2I |
| 424 | Syndecan interactions Homo sapiens R-HSA-3000170 | 2.65E-01 | 6.48E-01 | 2.11E+00 | 2.80E+00 | VTN;FGF2 |
| 425 | Regulation of gene expression in beta cells Homo sapiens R-HSA-210745 | 2.65E-01 | 6.48E-01 | 2.11E+00 | 2.80E+00 | PKLR;HNF4A |
| 426 | FGFR2 ligand binding and activation Homo sapiens R-HSA-190241 | 2.65E-01 | 6.48E-01 | 2.11E+00 | 2.80E+00 | FGF18;FGF2 |
| 427 | Inhibition of the proteolytic activity of APC/C required for the onset of anaphase by mitotic spindle checkpoint components Homo sapiens R-HSA-141405 | 2.65E-01 | 6.48E-01 | 2.11E+00 | 2.80E+00 | CDC26;ANAPC11 |
| 428 | Inactivation of APC/C via direct inhibition of the APC/C complex Homo sapiens R-HSA-141430 | 2.65E-01 | 6.48E-01 | 2.11E+00 | 2.80E+00 | CDC26;ANAPC11 |
| 429 | G beta:gamma signalling through PLC beta Homo sapiens R-HSA-418217 | 2.65E-01 | 6.48E-01 | 2.11E+00 | 2.80E+00 | GNG4;GNG11 |
| 430 | eNOS activation and regulation Homo sapiens R-HSA-203765 | 2.65E-01 | 6.48E-01 | 2.11E+00 | 2.80E+00 | GCHFR;CALM1 |
| 431 | Metabolism of nitric oxide Homo sapiens R-HSA-202131 | 2.65E-01 | 6.48E-01 | 2.11E+00 | 2.80E+00 | GCHFR;CALM1 |

| | | | | | | |
|---|---|---|---|---|---|---|
| 432 | Hormone-sensitive lipase (HSL)-mediated triacylglycerol hydrolysis Homo sapiens R-HSA-163560 | 2.65E-01 | 6.48E-01 | 2.11E+00 | 2.80E+00 | FABP1;MGLL |
| 433 | Vasopressin-like receptors Homo sapiens R-HSA-388479 | 2.66E-01 | 6.48E-01 | 3.79E+00 | 5.03E+00 | AVP |
| 434 | GDP-fucose biosynthesis Homo sapiens R-HSA-6787639 | 2.66E-01 | 6.48E-01 | 3.79E+00 | 5.03E+00 | FUOM |
| 435 | FGFR1b ligand binding and activation Homo sapiens R-HSA-190370 | 2.66E-01 | 6.48E-01 | 3.79E+00 | 5.03E+00 | FGF2 |
| 436 | Activation of C3 and C5 Homo sapiens R-HSA-174577 | 2.66E-01 | 6.48E-01 | 3.79E+00 | 5.03E+00 | C2 |
| 437 | Choline catabolism Homo sapiens R-HSA-6798163 | 2.66E-01 | 6.48E-01 | 3.79E+00 | 5.03E+00 | DMGDH |
| 438 | Acyl chain remodeling of CL Homo sapiens R-HSA-1482798 | 2.66E-01 | 6.48E-01 | 3.79E+00 | 5.03E+00 | PLA2G4A |
| 439 | Metabolic disorders of biological oxidation enzymes Homo sapiens R-HSA-5579029 | 2.66E-01 | 6.48E-01 | 3.79E+00 | 5.03E+00 | FDX1 |
| 440 | Synthesis of 5-eicosatetraenoic acids Homo sapiens R-HSA-2142688 | 2.66E-01 | 6.48E-01 | 3.79E+00 | 5.03E+00 | GPX2 |
| 441 | Synthesis of 15-eicosatetraenoic acid derivatives Homo sapiens R-HSA-2142770 | 2.66E-01 | 6.48E-01 | 3.79E+00 | 5.03E+00 | GPX2 |
| 442 | Synthesis of 12-eicosatetraenoic acid derivatives Homo sapiens R-HSA-2142712 | 2.66E-01 | 6.48E-01 | 3.79E+00 | 5.03E+00 | GPX2 |
| 443 | Protein repair Homo sapiens R-HSA-5676934 | 2.66E-01 | 6.48E-01 | 3.79E+00 | 5.03E+00 | MSRA |
| 444 | Downstream signal transduction Homo sapiens R-HSA-186763 | 2.66E-01 | 6.49E-01 | 1.18E+00 | 1.57E+00 | STAT5A;KLB;GRB7;RASGRF1;PEBP1;PHB;FGF2;PSMD8;PSMD9;UBB;PRKAR2B;PSMB3;PSMC4;PSMD2;FGF18;PSME1;LAMTOR2;FYN;CALM1;IER3 |

| | | | | | | |
|---|---|---|---|---|---|---|
| 445 | Eukaryotic Translation Termination Homo sapiens R-HSA-72764 | 2.71E-01 | 6.56E-01 | 1.41E+00 | 1.84E+00 | RPS28;RPS27;RPS29;RPL38;RPS4Y1;RPS21 |
| 446 | Regulation of PLK1 Activity at G2/M Transition Homo sapiens R-HSA-2565942 | 2.71E-01 | 6.56E-01 | 1.41E+00 | 1.84E+00 | UBB;PRKAR2B;SSNA1;CEP63;TUBG1;OPTN |
| 447 | Formation of TC-NER Pre-Incision Complex Homo sapiens R-HSA-6781823 | 2.75E-01 | 6.64E-01 | 1.55E+00 | 2.00E+00 | XAB2;UBB;POLR2E;POLR2I |
| 448 | Signaling by SCF-KIT Homo sapiens R-HSA-1433557 | 2.78E-01 | 6.70E-01 | 1.18E+00 | 1.51E+00 | STAT5A;KLB;GRB7;RASGRF1;PEBP1;PHB;FGF2;PSMD8;PSMD9;UBB;PSMB3;PSMC4;PSMD2;FGF18;PSME1;LAMTOR2;FYN;CALM1;IER3 |
| 449 | Signaling by the B Cell Receptor (BCR) Homo sapiens R-HSA-983705 | 2.81E-01 | 6.77E-01 | 1.21E+00 | 1.54E+00 | KLB;PRKCB;FGF2;PSMD8;PSMD9;UBB;PSMB3;PSMC4;PSMD2;FGF18;PSME1;FYN;CALM1;IER3 |
| 450 | Steroid hormones Homo sapiens R-HSA-209943 | 2.83E-01 | 6.79E-01 | 1.67E+00 | 2.11E+00 | FDX1;AKR1B1;HSD17B14 |
| 451 | Mitotic Spindle Checkpoint Homo sapiens R-HSA-69618 | 2.84E-01 | 6.79E-01 | 2.00E+00 | 2.51E+00 | CDC26;ANAPC11 |
| 452 | Presynaptic function of Kainate receptors Homo sapiens R-HSA-500657 | 2.84E-01 | 6.79E-01 | 2.00E+00 | 2.51E+00 | GNG4;GNG11 |
| 453 | Signaling by PDGF Homo sapiens R-HSA-186797 | 2.85E-01 | 6.79E-01 | 1.16E+00 | 1.46E+00 | STAT5A;KLB;GRB7;RASGRF1;PEBP1;PLG;PHB;FGF2;PSMD8;PSMD9;UBB;PRKAR2B;PSMB3;PSMC4;PSMD2;FGF18;PSME1;LAMTOR2;FYN;CALM1;IER3 |
| 454 | Eukaryotic Translation Elongation Homo sapiens R-HSA-156842 | 2.88E-01 | 6.85E-01 | 1.37E+00 | 1.71E+00 | RPS28;RPS27;RPS29;RPL38;RPS4Y1;RPS21 |
| 455 | Nonsense Mediated Decay (NMD) independent of the Exon Junction Complex (EJC) Homo sapiens R-HSA-975956 | 2.88E-01 | 6.85E-01 | 1.37E+00 | 1.71E+00 | RPS28;RPS27;RPS29;RPL38;RPS4Y1;RPS21 |
| 456 | Separation of Sister Chromatids Homo sapiens R-HSA-2467813 | 2.96E-01 | 6.90E-01 | 1.25E+00 | 1.52E+00 | PSMD8;PSMD9;RPS27;UBB;PSMB3;PSMC4;CDC26;PSMD2;PSME1;ANAPC11 |

| | | | | | | |
|---|---|---|---|---|---|---|
| 457 | Transport of vitamins, nucleosides, and related molecules Homo sapiens R-HSA-425397 | 2.97E-01 | 6.90E-01 | 1.63E+00 | 1.97E+00 | SLCO4A1;SLCO1A2;AVP |
| 458 | Nucleotide Excision Repair Homo sapiens R-HSA-5696398 | 2.98E-01 | 6.90E-01 | 1.31E+00 | 1.59E+00 | POLD3;XAB2;UBB;RFC1;RPA3;POLR2E;POLR2I |
| 459 | SLC transporter disorders Homo sapiens R-HSA-5619102 | 3.02E-01 | 6.90E-01 | 3.16E+00 | 3.78E+00 | AVP |
| 460 | POLB-Dependent Long Patch Base Excision Repair Homo sapiens R-HSA-110362 | 3.02E-01 | 6.90E-01 | 3.16E+00 | 3.78E+00 | PARG |
| 461 | APEX1-Independent Resolution of AP Sites via the Single Nucleotide Replacement Pathway Homo sapiens R-HSA-5649702 | 3.02E-01 | 6.90E-01 | 3.16E+00 | 3.78E+00 | OGG1 |
| 462 | GLI proteins bind promoters of Hh responsive genes to promote transcription Homo sapiens R-HSA-5635851 | 3.02E-01 | 6.90E-01 | 3.16E+00 | 3.78E+00 | BOC |
| 463 | FGFR3b ligand binding and activation Homo sapiens R-HSA-190371 | 3.02E-01 | 6.90E-01 | 3.16E+00 | 3.78E+00 | FGF18 |
| 464 | Glycerophospholipid catabolism Homo sapiens R-HSA-6814848 | 3.02E-01 | 6.90E-01 | 3.16E+00 | 3.78E+00 | GDPD3 |
| 465 | Acyl chain remodeling of DAG and TAG Homo sapiens R-HSA-1482883 | 3.02E-01 | 6.90E-01 | 3.16E+00 | 3.78E+00 | MGLL |
| 466 | Defective HLCS causes multiple carboxylase deficiency Homo sapiens R-HSA-3371599 | 3.02E-01 | 6.90E-01 | 3.16E+00 | 3.78E+00 | PCCB |
| 467 | Defects in biotin (Btn) metabolism Homo sapiens R-HSA-3323169 | 3.02E-01 | 6.90E-01 | 3.16E+00 | 3.78E+00 | PCCB |

| | | | | | | |
|---|---|---|---|---|---|---|
| 468 | Histidine catabolism Homo sapiens R-HSA-70921 | 3.02E-01 | 6.90E-01 | 3.16E+00 | 3.78E+00 | FTCD |
| 469 | RA biosynthesis pathway Homo sapiens R-HSA-5365859 | 3.03E-01 | 6.90E-01 | 1.90E+00 | 2.26E+00 | ALDH1A3;RDH10 |
| 470 | Common Pathway of Fibrin Clot Formation Homo sapiens R-HSA-140875 | 3.03E-01 | 6.90E-01 | 1.90E+00 | 2.26E+00 | F2R;PF4 |
| 471 | Pink/Parkin Mediated Mitophagy Homo sapiens R-HSA-5205685 | 3.03E-01 | 6.90E-01 | 1.90E+00 | 2.26E+00 | UBB;TOMM5 |
| 472 | Mitophagy Homo sapiens R-HSA-5205647 | 3.03E-01 | 6.90E-01 | 1.90E+00 | 2.26E+00 | UBB;TOMM5 |
| 473 | Signaling by FGFR3 point mutants in cancer Homo sapiens R-HSA-8853338 | 3.03E-01 | 6.90E-01 | 1.90E+00 | 2.26E+00 | FGF18;FGF2 |
| 474 | Signaling by FGFR3 in disease Homo sapiens R-HSA-5655332 | 3.03E-01 | 6.90E-01 | 1.90E+00 | 2.26E+00 | FGF18;FGF2 |
| 475 | ADP signalling through P2Y purinoceptor 12 Homo sapiens R-HSA-392170 | 3.03E-01 | 6.90E-01 | 1.90E+00 | 2.26E+00 | GNG4;GNG11 |
| 476 | Collagen degradation Homo sapiens R-HSA-1442490 | 3.11E-01 | 7.03E-01 | 1.58E+00 | 1.85E+00 | COL14A1;CTSK;COL12A1 |
| 477 | Formation of Fibrin Clot (Clotting Cascade) Homo sapiens R-HSA-140877 | 3.11E-01 | 7.03E-01 | 1.58E+00 | 1.85E+00 | F2R;SERPING1;PF4 |
| 478 | Gamma carboxylation, hypusine formation and arylsulfatase activation Homo sapiens R-HSA-163841 | 3.11E-01 | 7.03E-01 | 1.58E+00 | 1.85E+00 | EIF5A;FN3K;PROZ |
| 479 | Signaling by Wnt Homo sapiens R-HSA-195721 | 3.12E-01 | 7.05E-01 | 1.16E+00 | 1.35E+00 | PRKCB;H2AFZ;WNT7B;CLTB;AP2A1;GNG11;PSMD8;PSMD9;UBB;PSMB3;PSMC4;GNG4;PSMD2;CCDC88C;PSME1;AP2S1;CALM1 |
| 480 | Influenza Viral RNA Transcription and Replication Homo sapiens R-HSA-168273 | 3.13E-01 | 7.05E-01 | 1.26E+00 | 1.47E+00 | RPS28;RPS27;RPS29;POLR2E;RPL38;RPS4Y1;POLR2I;RPS21 |

| | | | | | | |
|---|---|---|---|---|---|---|
| 481 | MicroRNA (miRNA) biogenesis Homo sapiens R-HSA-203927 | 3.22E-01 | 7.20E-01 | 1.81E+00 | 2.05E+00 | POLR2E;POLR2I |
| 482 | Abortive elongation of HIV-1 transcript in the absence of Tat Homo sapiens R-HSA-167242 | 3.22E-01 | 7.20E-01 | 1.81E+00 | 2.05E+00 | POLR2E;POLR2I |
| 483 | Thromboxane signalling through TP receptor Homo sapiens R-HSA-428930 | 3.22E-01 | 7.20E-01 | 1.81E+00 | 2.05E+00 | GNG4;GNG11 |
| 484 | FCERI mediated NF-kB activation Homo sapiens R-HSA-2871837 | 3.22E-01 | 7.20E-01 | 1.28E+00 | 1.45E+00 | PSMD8;PSMD9;UBB;PSMB3;PSMC4;PSMD2;PSME1 |
| 485 | Antigen processing: Ubiquitination & Proteasome degradation Homo sapiens R-HSA-983168 | 3.26E-01 | 7.26E-01 | 1.16E+00 | 1.30E+00 | ANAPC13;TRIM63;ZBTB16;FBXO17;ANAPC11;PSMD8;PSMD9;ZNRF1;UBB;PSMB3;PSMC4;CDC26;PSMD2;ASB9;PSME1 |
| 486 | Neurotransmitter Clearance In The Synaptic Cleft Homo sapiens R-HSA-112311 | 3.37E-01 | 7.33E-01 | 2.71E+00 | 2.94E+00 | MAOA |
| 487 | Cleavage of the damaged pyrimidine Homo sapiens R-HSA-110329 | 3.37E-01 | 7.33E-01 | 2.71E+00 | 2.94E+00 | OGG1 |
| 488 | Recognition and association of DNA glycosylase with site containing an affected pyrimidine Homo sapiens R-HSA-110328 | 3.37E-01 | 7.33E-01 | 2.71E+00 | 2.94E+00 | OGG1 |
| 489 | Depyrimidination Homo sapiens R-HSA-73928 | 3.37E-01 | 7.33E-01 | 2.71E+00 | 2.94E+00 | OGG1 |
| 490 | Ligand-receptor interactions Homo sapiens R-HSA-5632681 | 3.37E-01 | 7.33E-01 | 2.71E+00 | 2.94E+00 | BOC |
| 491 | Digestion of dietary carbohydrate Homo sapiens R-HSA-189085 | 3.37E-01 | 7.33E-01 | 2.71E+00 | 2.94E+00 | MGAM |
| 492 | Nef and signal transduction Homo sapiens R-HSA-164944 | 3.37E-01 | 7.33E-01 | 2.71E+00 | 2.94E+00 | FYN |

| | | | | | | |
|---|---|---|---|---|---|---|
| 493 | Downregulation of ERBB4 signaling Homo sapiens R-HSA-1253288 | 3.37E-01 | 7.33E-01 | 2.71E+00 | 2.94E+00 | UBB |
| 494 | Cross-presentation of particulate exogenous antigens (phagosomes) Homo sapiens R-HSA-1236973 | 3.37E-01 | 7.33E-01 | 2.71E+00 | 2.94E+00 | CYBA |
| 495 | Hydrolysis of LPC Homo sapiens R-HSA-1483115 | 3.37E-01 | 7.33E-01 | 2.71E+00 | 2.94E+00 | PLA2G4A |
| 496 | ChREBP activates metabolic gene expression Homo sapiens R-HSA-163765 | 3.37E-01 | 7.33E-01 | 2.71E+00 | 2.94E+00 | PKLR |
| 497 | Metabolism of ingested SeMet, Sec, MeSec into H2Se Homo sapiens R-HSA-2408508 | 3.37E-01 | 7.33E-01 | 2.71E+00 | 2.94E+00 | AHCY |
| 498 | Nicotinamide salvaging Homo sapiens R-HSA-197264 | 3.37E-01 | 7.33E-01 | 2.71E+00 | 2.94E+00 | NAPRT |
| 499 | Glutamate Neurotransmitter Release Cycle Homo sapiens R-HSA-210500 | 3.41E-01 | 7.35E-01 | 1.72E+00 | 1.86E+00 | SLC38A2;GLS |
| 500 | Growth hormone receptor signaling Homo sapiens R-HSA-982772 | 3.41E-01 | 7.35E-01 | 1.72E+00 | 1.86E+00 | STAT5A;PTPN1 |
| 501 | DARPP-32 events Homo sapiens R-HSA-180024 | 3.41E-01 | 7.35E-01 | 1.72E+00 | 1.86E+00 | PRKAR2B;CALM1 |
| 502 | Telomere Maintenance Homo sapiens R-HSA-157579 | 3.43E-01 | 7.39E-01 | 1.38E+00 | 1.47E+00 | POLD3;RFC1;H2AFZ;RPA3 |
| 503 | Cellular response to heat stress Homo sapiens R-HSA-3371556 | 3.51E-01 | 7.41E-01 | 1.26E+00 | 1.33E+00 | RPS19BP1;UBB;RPA3;BAG1;HSPA2;HDAC6 |
| 504 | Formation of a pool of free 40S subunits Homo sapiens R-HSA-72689 | 3.51E-01 | 7.41E-01 | 1.26E+00 | 1.33E+00 | RPS28;RPS27;RPS29;RPL38;RPS4Y1;RPS21 |

| | | | | | | |
|---|---|---|---|---|---|---|
| 505 | Non-integrin membrane-ECM interactions Homo sapiens R-HSA-3000171 | 3.53E-01 | 7.41E-01 | 1.46E+00 | 1.52E+00 | VTN;FGF2;DDR2 |
| 506 | Transcriptional regulation of pluripotent stem cells Homo sapiens R-HSA-452723 | 3.53E-01 | 7.41E-01 | 1.46E+00 | 1.52E+00 | POLR2E;POLR2I;FGF2 |
| 507 | Glucagon-like Peptide-1 (GLP1) regulates insulin secretion Homo sapiens R-HSA-381676 | 3.53E-01 | 7.41E-01 | 1.46E+00 | 1.52E+00 | GNG4;PRKAR2B;GNG11 |
| 508 | Translocation of GLUT4 to the plasma membrane Homo sapiens R-HSA-1445148 | 3.55E-01 | 7.41E-01 | 1.35E+00 | 1.40E+00 | TBC1D1;MYH8;ASPSCR1;CALM1 |
| 509 | Signaling by FGFR2 Homo sapiens R-HSA-5654738 | 3.55E-01 | 7.41E-01 | 1.11E+00 | 1.15E+00 | KLB;RASGRF1;PEBP1;PHB;FGF2;PSMD8;PSMD9;UBB;PRKAR2B;PSMB3;PSMC4;PSMD2;FGF18;PSME1;POLR2E;LAMTOR2;FYN;CALM1;POLR2I;IER3 |
| 510 | G alpha (q) signalling events Homo sapiens R-HSA-416476 | 3.63E-01 | 7.41E-01 | 1.16E+00 | 1.18E+00 | RGS18;RGS3;PRKCH;PLCB4;PTGER1;GNG4;F2R;AVP;NTS;GNG11;MGLL |
| 511 | Centrosome maturation Homo sapiens R-HSA-380287 | 3.63E-01 | 7.41E-01 | 1.28E+00 | 1.30E+00 | TUBGCP2;PRKAR2B;SSNA1;CEP63;TUBG1 |
| 512 | Recruitment of mitotic centrosome proteins and complexes Homo sapiens R-HSA-380270 | 3.63E-01 | 7.41E-01 | 1.28E+00 | 1.30E+00 | TUBGCP2;PRKAR2B;SSNA1;CEP63;TUBG1 |
| 513 | Cardiac conduction Homo sapiens R-HSA-5576891 | 3.66E-01 | 7.41E-01 | 1.19E+00 | 1.20E+00 | KCNK5;DMPK;FXYD4;CACNA2D1;FXYD2;ATP1B3;ATP1B1;CALM1 |
| 514 | Constitutive Signaling by Aberrant PI3K in Cancer Homo sapiens R-HSA-2219530 | 3.66E-01 | 7.41E-01 | 1.33E+00 | 1.34E+00 | KLB;FGF18;FYN;FGF2 |
| 515 | Mitotic Anaphase Homo sapiens R-HSA-68882 | 3.69E-01 | 7.41E-01 | 1.16E+00 | 1.16E+00 | PSMD8;PSMD9;RPS27;UBB;PSMB3;PSMC4;CDC26;PSMD2;PSME1;ANAPC11 |
| 516 | Transmission across Chemical Synapses Homo sapiens R-HSA-112315 | 3.69E-01 | 7.41E-01 | 1.14E+00 | 1.14E+00 | PRKCB;MAOA;GNG4;CACNA2D1;RASGRF1;AP2S1;SLC6A12;AP2A1;CALM1;GNG11;SLC38A2;GLS |

| | | | | | | |
|---|---|---|---|---|---|---|
| 517 | Bicarbonate transporters Homo sapiens R-HSA-425381 | 3.71E-01 | 7.41E-01 | 2.37E+00 | 2.35E+00 | SLC4A7 |
| 518 | Organic cation transport Homo sapiens R-HSA-549127 | 3.71E-01 | 7.41E-01 | 2.37E+00 | 2.35E+00 | SLC22A5 |
| 519 | Displacement of DNA glycosylase by APEX1 Homo sapiens R-HSA-110357 | 3.71E-01 | 7.41E-01 | 2.37E+00 | 2.35E+00 | OGG1 |
| 520 | Retinoid cycle disease events Homo sapiens R-HSA-2453864 | 3.71E-01 | 7.41E-01 | 2.37E+00 | 2.35E+00 | NAPEPLD |
| 521 | Diseases associated with visual transduction Homo sapiens R-HSA-2474795 | 3.71E-01 | 7.41E-01 | 2.37E+00 | 2.35E+00 | NAPEPLD |
| 522 | Sodium/Proton exchangers Homo sapiens R-HSA-425986 | 3.71E-01 | 7.41E-01 | 2.37E+00 | 2.35E+00 | SLC9A3 |
| 523 | Transport of gamma-carboxylated protein precursors from the endoplasmic reticulum to the Golgi apparatus Homo sapiens R-HSA-159763 | 3.71E-01 | 7.41E-01 | 2.37E+00 | 2.35E+00 | PROZ |
| 524 | MAPK1 (ERK2) activation Homo sapiens R-HSA-112411 | 3.71E-01 | 7.41E-01 | 2.37E+00 | 2.35E+00 | IL6R |
| 525 | PTK6 Regulates RTKs and Their Effectors AKT1 and DOK1 Homo sapiens R-HSA-8849469 | 3.71E-01 | 7.41E-01 | 2.37E+00 | 2.35E+00 | UBB |
| 526 | Inactivation of Cdc42 and Rac Homo sapiens R-HSA-428543 | 3.71E-01 | 7.41E-01 | 2.37E+00 | 2.35E+00 | SRGAP3 |
| 527 | Prostanoid ligand receptors Homo sapiens R-HSA-391908 | 3.71E-01 | 7.41E-01 | 2.37E+00 | 2.35E+00 | PTGER1 |
| 528 | Mitochondrial iron-sulfur cluster biogenesis Homo sapiens R-HSA-1362409 | 3.71E-01 | 7.41E-01 | 2.37E+00 | 2.35E+00 | FDX1 |

| | | | | | | |
|---|---|---|---|---|---|---|
| 529 | Synthesis of (16-20)-hydroxyeicosatetraenoic acids (HETE) Homo sapiens R-HSA-2142816 | 3.71E-01 | 7.41E-01 | 2.37E+00 | 2.35E+00 | CYP4F2 |
| 530 | Pyrimidine salvage reactions Homo sapiens R-HSA-73614 | 3.71E-01 | 7.41E-01 | 2.37E+00 | 2.35E+00 | TK1 |
| 531 | Diseases of signal transduction Homo sapiens R-HSA-5663202 | 3.73E-01 | 7.41E-01 | 1.12E+00 | 1.10E+00 | STAT5A;KLB;FGF2;HDAC6;PSMD8;PSMD9;UBB;PSMB3;PSMC4;PSMD2;FGF18;PSME1;NAPEPLD;POLR2E;FYN;POLR2I |
| 532 | Regulation of HSF1-mediated heat shock response Homo sapiens R-HSA-3371453 | 3.74E-01 | 7.41E-01 | 1.26E+00 | 1.24E+00 | RPS19BP1;UBB;RPA3;BAG1;HSPA2 |
| 533 | Influenza Life Cycle Homo sapiens R-HSA-168255 | 3.74E-01 | 7.41E-01 | 1.19E+00 | 1.17E+00 | RPS28;RPS27;RPS29;POLR2E;RPL38;RPS4Y1;POLR2I;RPS21 |
| 534 | Mitotic G1-G1/S phases Homo sapiens R-HSA-453279 | 3.74E-01 | 7.41E-01 | 1.19E+00 | 1.17E+00 | PSMD8;PSMD9;UBB;PSMB3;PSMC4;PSMD2;RPA3;PSME1 |
| 535 | Mitotic Metaphase and Anaphase Homo sapiens R-HSA-2555396 | 3.75E-01 | 7.41E-01 | 1.16E+00 | 1.13E+00 | PSMD8;PSMD9;RPS27;UBB;PSMB3;PSMC4;CDC26;PSMD2;PSME1;ANAPC11 |
| 536 | Cell Cycle, Mitotic Homo sapiens R-HSA-69278 | 3.76E-01 | 7.41E-01 | 1.09E+00 | 1.06E+00 | TUBGCP2;RAB1B;SSNA1;GMNN;ANAPC11;PSMD8;POLD3;PSMD9;UBB;PRKAR2B;PSMB3;PSMD2;CDC26;RFC1;PRKCB;H2AFZ;TUBG1;CDC25B;RPS27;PSMC4;RPA3;PSME1;CEP63;LPIN2;OPTN |
| 537 | Recycling pathway of L1 Homo sapiens R-HSA-437239 | 3.77E-01 | 7.41E-01 | 1.58E+00 | 1.54E+00 | AP2S1;AP2A1 |
| 538 | Attenuation phase Homo sapiens R-HSA-3371568 | 3.77E-01 | 7.41E-01 | 1.58E+00 | 1.54E+00 | UBB;HSPA2 |
| 539 | FGFR2 alternative splicing Homo sapiens R-HSA-6803529 | 3.77E-01 | 7.41E-01 | 1.58E+00 | 1.54E+00 | POLR2E;POLR2I |
| 540 | SHC-mediated cascade:FGFR2 Homo sapiens R-HSA-5654699 | 3.77E-01 | 7.41E-01 | 1.58E+00 | 1.54E+00 | FGF18;FGF2 |
| 541 | Regulation of Complement cascade Homo sapiens R-HSA-977606 | 3.77E-01 | 7.41E-01 | 1.58E+00 | 1.54E+00 | VTN;C2 |

| | | | | | | |
|---|---|---|---|---|---|---|
| 542 | CaM pathway Homo sapiens R-HSA-111997 | 3.77E-01 | 7.41E-01 | 1.58E+00 | 1.54E+00 | PRKAR2B;CALM1 |
| 543 | Calmodulin induced events Homo sapiens R-HSA-111933 | 3.77E-01 | 7.41E-01 | 1.58E+00 | 1.54E+00 | PRKAR2B;CALM1 |
| 544 | Downregulation of TGF-beta receptor signaling Homo sapiens R-HSA-2173788 | 3.77E-01 | 7.41E-01 | 1.58E+00 | 1.54E+00 | UBB;STRAP |
| 545 | FCERI mediated MAPK activation Homo sapiens R-HSA-2871796 | 3.78E-01 | 7.41E-01 | 1.11E+00 | 1.08E+00 | KLB;RASGRF1;PEBP1;PHB;FGF2;PSMD8;PSMD9;UBB;PSMB3;PSMC4;PSMD2;FGF18;PSME1;LAMTOR2;FYN;CALM1 |
| 546 | Signaling by FGFR Homo sapiens R-HSA-190236 | 3.79E-01 | 7.41E-01 | 1.10E+00 | 1.06E+00 | KLB;RASGRF1;PEBP1;PHB;FGF2;PSMD8;PSMD9;UBB;PRKAR2B;PSMB3;PSMC4;PSMD2;FGF18;PSME1;POLR2E;LAMTOR2;FYN;CALM1;POLR2I;IER3 |
| 547 | Downstream signaling of activated FGFR2 Homo sapiens R-HSA-5654696 | 3.86E-01 | 7.41E-01 | 1.10E+00 | 1.04E+00 | KLB;RASGRF1;PEBP1;PHB;FGF2;PSMD8;PSMD9;UBB;PRKAR2B;PSMB3;PSMC4;PSMD2;FGF18;PSME1;LAMTOR2;FYN;CALM1;IER3 |
| 548 | Downstream signaling of activated FGFR4 Homo sapiens R-HSA-5654716 | 3.86E-01 | 7.41E-01 | 1.10E+00 | 1.04E+00 | KLB;RASGRF1;PEBP1;PHB;FGF2;PSMD8;PSMD9;UBB;PRKAR2B;PSMB3;PSMC4;PSMD2;FGF18;PSME1;LAMTOR2;FYN;CALM1;IER3 |
| 549 | Downstream signaling of activated FGFR3 Homo sapiens R-HSA-5654708 | 3.86E-01 | 7.41E-01 | 1.10E+00 | 1.04E+00 | KLB;RASGRF1;PEBP1;PHB;FGF2;PSMD8;PSMD9;UBB;PRKAR2B;PSMB3;PSMC4;PSMD2;FGF18;PSME1;LAMTOR2;FYN;CALM1;IER3 |
| 550 | Collagen biosynthesis and modifying enzymes Homo sapiens R-HSA-1650814 | 3.89E-01 | 7.41E-01 | 1.29E+00 | 1.21E+00 | P4HA1;COL14A1;P4HA2;COL12A1 |
| 551 | Synthesis of substrates in N-glycan biosythesis Homo sapiens R-HSA-446219 | 3.89E-01 | 7.41E-01 | 1.29E+00 | 1.21E+00 | CTSA;SLC17A5;NPL;FUOM |
| 552 | Signaling by ERBB4 Homo sapiens R-HSA-1236394 | 3.91E-01 | 7.41E-01 | 1.09E+00 | 1.03E+00 | STAT5A;KLB;RASGRF1;PEBP1;PHB;FGF2;PSMD8;PSMD9;UBB;PSMB3;PSMC4;PSMD2;FGF18;PSME1;LAMTOR2;FYN;CALM1;IER3 |
| 553 | ABC-family proteins mediated transport Homo sapiens R-HSA-382556 | 3.94E-01 | 7.41E-01 | 1.35E+00 | 1.26E+00 | ABCA4;ABCA12;ABCG1 |
| 554 | Signaling by ERBB2 Homo sapiens R-HSA-1227986 | 3.94E-01 | 7.41E-01 | 1.35E+00 | 1.26E+00 | GRB7;UBB;FYN |

| | | | | | | |
|---|---|---|---|---|---|---|
| 555 | Diseases of metabolism Homo sapiens R-HSA-5668914 | 3.94E-01 | 7.41E-01 | 1.35E+00 | 1.26E+00 | UBB;PCCB;FDX1 |
| 556 | Muscle contraction Homo sapiens R-HSA-397014 | 3.95E-01 | 7.41E-01 | 1.13E+00 | 1.05E+00 | TMOD1;KCNK5;DMPK;FXYD4;CACNA2D1;FXYD2;TNNC1;MYH8;ATP1B3;CALM1;ATP1B1 |
| 557 | RNA Pol II CTD phosphorylation and interaction with CE Homo sapiens R-HSA-167160 | 3.95E-01 | 7.41E-01 | 1.52E+00 | 1.41E+00 | POLR2E;POLR2I |
| 558 | RNA Pol II CTD phosphorylation and interaction with CE Homo sapiens R-HSA-77075 | 3.95E-01 | 7.41E-01 | 1.52E+00 | 1.41E+00 | POLR2E;POLR2I |
| 559 | Interleukin-6 family signaling Homo sapiens R-HSA-6783589 | 3.95E-01 | 7.41E-01 | 1.52E+00 | 1.41E+00 | CNTF;IL6R |
| 560 | CREB phosphorylation through the activation of Ras Homo sapiens R-HSA-442742 | 3.95E-01 | 7.41E-01 | 1.52E+00 | 1.41E+00 | RASGRF1;CALM1 |
| 561 | Effects of PIP2 hydrolysis Homo sapiens R-HSA-114508 | 3.95E-01 | 7.41E-01 | 1.52E+00 | 1.41E+00 | PRKCH;MGLL |
| 562 | HDR through Homologous Recombination (HRR) Homo sapiens R-HSA-5685942 | 4.01E-01 | 7.41E-01 | 1.26E+00 | 1.16E+00 | POLD3;RFC1;UBB;RPA3 |
| 563 | Signaling by FGFR4 Homo sapiens R-HSA-5654743 | 4.01E-01 | 7.41E-01 | 1.09E+00 | 9.93E-01 | KLB;RASGRF1;PEBP1;PHB;FGF2;PSMD8;PSMD9;UBB;PRKAR2B;PSMB3;PSMC4;PSMD2;FGF18;PSME1;LAMTOR2;FYN;CALM1;IER3 |
| 564 | Downstream signaling of activated FGFR1 Homo sapiens R-HSA-5654687 | 4.01E-01 | 7.41E-01 | 1.09E+00 | 9.93E-01 | KLB;RASGRF1;PEBP1;PHB;FGF2;PSMD8;PSMD9;UBB;PRKAR2B;PSMB3;PSMC4;PSMD2;FGF18;PSME1;LAMTOR2;FYN;CALM1;IER3 |
| 565 | Interaction With The Zona Pellucida Homo sapiens R-HSA-1300644 | 4.02E-01 | 7.41E-01 | 2.11E+00 | 1.92E+00 | ZP3 |
| 566 | Zinc influx into cells by the SLC39 gene family Homo sapiens R-HSA-442380 | 4.02E-01 | 7.41E-01 | 2.11E+00 | 1.92E+00 | SLC39A5 |

| | | | | | | |
|---|---|---|---|---|---|---|
| 567 | Abacavir transport and metabolism Homo sapiens R-HSA-2161522 | 4.02E-01 | 7.41E-01 | 2.11E+00 | 1.92E+00 | PCK1 |
| 568 | Base-Excision Repair, AP Site Formation Homo sapiens R-HSA-73929 | 4.02E-01 | 7.41E-01 | 2.11E+00 | 1.92E+00 | OGG1 |
| 569 | Removal of aminoterminal propeptides from gamma-carboxylated proteins Homo sapiens R-HSA-159782 | 4.02E-01 | 7.41E-01 | 2.11E+00 | 1.92E+00 | PROZ |
| 570 | Gamma-carboxylation of protein precursors Homo sapiens R-HSA-159740 | 4.02E-01 | 7.41E-01 | 2.11E+00 | 1.92E+00 | PROZ |
| 571 | Nef mediated downregulation of MHC class I complex cell surface expression Homo sapiens R-HSA-164940 | 4.02E-01 | 7.41E-01 | 2.11E+00 | 1.92E+00 | AP1M1 |
| 572 | Activation of Ca-permeable Kainate Receptor Homo sapiens R-HSA-451308 | 4.02E-01 | 7.41E-01 | 2.11E+00 | 1.92E+00 | CALM1 |
| 573 | Ionotropic activity of Kainate Receptors Homo sapiens R-HSA-451306 | 4.02E-01 | 7.41E-01 | 2.11E+00 | 1.92E+00 | CALM1 |
| 574 | SLBP independent Processing of Histone Pre-mRNAs Homo sapiens R-HSA-111367 | 4.02E-01 | 7.41E-01 | 2.11E+00 | 1.92E+00 | SNRPG |
| 575 | MAPK3 (ERK1) activation Homo sapiens R-HSA-110056 | 4.02E-01 | 7.41E-01 | 2.11E+00 | 1.92E+00 | IL6R |
| 576 | IRAK2 mediated activation of TAK1 complex upon TLR7/8 or 9 stimulation Homo sapiens R-HSA-975163 | 4.02E-01 | 7.41E-01 | 2.11E+00 | 1.92E+00 | UBB |
| 577 | IRAK2 mediated activation of TAK1 complex Homo sapiens R-HSA-937042 | 4.02E-01 | 7.41E-01 | 2.11E+00 | 1.92E+00 | UBB |

| | | | | | | |
|---|---|---|---|---|---|---|
| 578 | Diseases of carbohydrate metabolism Homo sapiens R-HSA-5663084 | 4.02E-01 | 7.41E-01 | 2.11E+00 | 1.92E+00 | UBB |
| 579 | Myoclonic epilepsy of Lafora Homo sapiens R-HSA-3785653 | 4.02E-01 | 7.41E-01 | 2.11E+00 | 1.92E+00 | UBB |
| 580 | Glycogen storage diseases Homo sapiens R-HSA-3229121 | 4.02E-01 | 7.41E-01 | 2.11E+00 | 1.92E+00 | UBB |
| 581 | FGFR2b ligand binding and activation Homo sapiens R-HSA-190377 | 4.02E-01 | 7.41E-01 | 2.11E+00 | 1.92E+00 | FGF2 |
| 582 | Netrin mediated repulsion signals Homo sapiens R-HSA-418886 | 4.02E-01 | 7.41E-01 | 2.11E+00 | 1.92E+00 | FYN |
| 583 | Dectin-2 family Homo sapiens R-HSA-5621480 | 4.02E-01 | 7.41E-01 | 2.11E+00 | 1.92E+00 | FYN |
| 584 | eNOS activation Homo sapiens R-HSA-203615 | 4.02E-01 | 7.41E-01 | 2.11E+00 | 1.92E+00 | CALM1 |
| 585 | Synthesis of pyrophosphates in the cytosol Homo sapiens R-HSA-1855167 | 4.02E-01 | 7.41E-01 | 2.11E+00 | 1.92E+00 | IP6K3 |
| 586 | Thyroxine biosynthesis Homo sapiens R-HSA-209968 | 4.02E-01 | 7.41E-01 | 2.11E+00 | 1.92E+00 | DIO1 |
| 587 | Purine catabolism Homo sapiens R-HSA-74259 | 4.02E-01 | 7.41E-01 | 2.11E+00 | 1.92E+00 | CAT |
| 588 | PI5P, PP2A and IER3 Regulate PI3K/AKT Signaling Homo sapiens R-HSA-6811558 | 4.04E-01 | 7.42E-01 | 1.22E+00 | 1.10E+00 | KLB;FGF18;FYN;FGF2;IER3 |
| 589 | Glycerophospholipid biosynthesis Homo sapiens R-HSA-1483206 | 4.05E-01 | 7.43E-01 | 1.18E+00 | 1.07E+00 | DMGDH;ETNK2;CPNE6;PLA2G4A;LPIN2;MGLL |
| 590 | Signaling by FGFR3 Homo sapiens R-HSA-5654741 | 4.06E-01 | 7.44E-01 | 1.08E+00 | 9.77E-01 | KLB;RASGRF1;PEBP1;PHB;FGF2;PSMD8;PSMD9;UBB;PRKAR2B;PSMB3;PSMC4;PSMD2;FGF18;PSME1;LAMTOR2;FYN;CALM1;IER3 |

| | | | | | | |
|---|---|---|---|---|---|---|
| 591 | Signaling by Interleukins Homo sapiens R-HSA-449147 | 4.09E-01 | 7.49E-01 | 1.07E+00 | 9.59E-01 | STAT5A;KLB;CNTF;RASGRF1;PEBP1;PHB;FGF2;PSMD8;PSMD9;UBB;PSMB3;PSMC4;TNIP2;PSMD2;FGF18;PSME1;LAMTOR2;FYN;CALM1;IL6R;HAVCR2 |
| 592 | RNA Polymerase III Transcription Initiation From Type 3 Promoter Homo sapiens R-HSA-76071 | 4.13E-01 | 7.52E-01 | 1.46E+00 | 1.29E+00 | SNAPC1;POLR2E |
| 593 | G-protein activation Homo sapiens R-HSA-202040 | 4.13E-01 | 7.52E-01 | 1.46E+00 | 1.29E+00 | GNG4;GNG11 |
| 594 | Adrenaline,noradrenaline inhibits insulin secretion Homo sapiens R-HSA-400042 | 4.13E-01 | 7.52E-01 | 1.46E+00 | 1.29E+00 | GNG4;GNG11 |
| 595 | rRNA processing Homo sapiens R-HSA-72312 | 4.16E-01 | 7.54E-01 | 1.12E+00 | 9.78E-01 | RRP7A;RPS28;RPS27;RPS29;WDR18;RPS27L;RPL38;RPS4Y1;RPS21;NOP10 |
| 596 | NGF signalling via TRKA from the plasma membrane Homo sapiens R-HSA-187037 | 4.16E-01 | 7.54E-01 | 1.07E+00 | 9.39E-01 | KLB;RASGRF1;PEBP1;AP2A1;PHB;FGF2;PSMD8;PSMD9;UBB;PRKAR2B;PSMB3;PSMC4;PSMD2;FGF18;PSME1;AP2S1;LAMTOR2;FYN;CALM1;IER3 |
| 597 | Signaling by FGFR1 Homo sapiens R-HSA-5654736 | 4.21E-01 | 7.54E-01 | 1.07E+00 | 9.29E-01 | KLB;RASGRF1;PEBP1;PHB;FGF2;PSMD8;PSMD9;UBB;PRKAR2B;PSMB3;PSMC4;PSMD2;FGF18;PSME1;LAMTOR2;FYN;CALM1;IER3 |
| 598 | NOTCH1 Intracellular Domain Regulates Transcription Homo sapiens R-HSA-2122947 | 4.22E-01 | 7.54E-01 | 1.29E+00 | 1.12E+00 | UBB;HIF1A;HDAC6 |
| 599 | Phospholipase C-mediated cascade: FGFR1 Homo sapiens R-HSA-5654219 | 4.22E-01 | 7.54E-01 | 1.29E+00 | 1.12E+00 | PRKAR2B;CALM1;FGF2 |
| 600 | Cell Cycle Checkpoints Homo sapiens R-HSA-69620 | 4.30E-01 | 7.54E-01 | 1.10E+00 | 9.31E-01 | PSMD8;PSMD9;UBB;PSMB3;PSMC4;CDC26;PSMD2;RPA3;PSME1;ANAPC11 |
| 601 | Surfactant metabolism Homo sapiens R-HSA-5683826 | 4.31E-01 | 7.54E-01 | 1.40E+00 | 1.18E+00 | SLC34A1;CTSH |
| 602 | PIWI-interacting RNA (piRNA) biogenesis Homo sapiens R-HSA-5601884 | 4.31E-01 | 7.54E-01 | 1.40E+00 | 1.18E+00 | POLR2E;POLR2I |
| 603 | mRNA Capping Homo sapiens R-HSA-72086 | 4.31E-01 | 7.54E-01 | 1.40E+00 | 1.18E+00 | POLR2E;POLR2I |

| | | | | | | |
|---|---|---|---|---|---|---|
| 604 | Myogenesis Homo sapiens R-HSA-525793 | 4.31E-01 | 7.54E-01 | 1.40E+00 | 1.18E+00 | BOC;CDH15 |
| 605 | CDO in myogenesis Homo sapiens R-HSA-375170 | 4.31E-01 | 7.54E-01 | 1.40E+00 | 1.18E+00 | BOC;CDH15 |
| 606 | Uptake and function of anthrax toxins Homo sapiens R-HSA-5210891 | 4.32E-01 | 7.54E-01 | 1.89E+00 | 1.59E+00 | CALM1 |
| 607 | Gamma-carboxylation, transport, and amino-terminal cleavage of proteins Homo sapiens R-HSA-159854 | 4.32E-01 | 7.54E-01 | 1.89E+00 | 1.59E+00 | PROZ |
| 608 | SLBP Dependent Processing of Replication-Dependent Histone Pre-mRNAs Homo sapiens R-HSA-77588 | 4.32E-01 | 7.54E-01 | 1.89E+00 | 1.59E+00 | SNRPG |
| 609 | Interleukin-6 signaling Homo sapiens R-HSA-1059683 | 4.32E-01 | 7.54E-01 | 1.89E+00 | 1.59E+00 | IL6R |
| 610 | Membrane binding and targetting of GAG proteins Homo sapiens R-HSA-174490 | 4.32E-01 | 7.54E-01 | 1.89E+00 | 1.59E+00 | UBB |
| 611 | Synthesis And Processing Of GAG, GAGPOL Polyproteins Homo sapiens R-HSA-174495 | 4.32E-01 | 7.54E-01 | 1.89E+00 | 1.59E+00 | UBB |
| 612 | FGFR1c ligand binding and activation Homo sapiens R-HSA-190373 | 4.32E-01 | 7.54E-01 | 1.89E+00 | 1.59E+00 | FGF2 |
| 613 | Signaling by activated point mutants of FGFR1 Homo sapiens R-HSA-1839122 | 4.32E-01 | 7.54E-01 | 1.89E+00 | 1.59E+00 | FGF2 |
| 614 | RHO GTPases activate IQGAPs Homo sapiens R-HSA-5626467 | 4.32E-01 | 7.54E-01 | 1.89E+00 | 1.59E+00 | CALM1 |
| 615 | Interleukin-7 signaling Homo sapiens R-HSA-1266695 | 4.32E-01 | 7.54E-01 | 1.89E+00 | 1.59E+00 | STAT5A |
| 616 | Sodium/Calcium exchangers Homo sapiens R-HSA-425561 | 4.32E-01 | 7.54E-01 | 1.89E+00 | 1.59E+00 | CALM1 |

| | | | | | | |
|---|---|---|---|---|---|---|
| 617 | CLEC7A (Dectin-1) induces NFAT activation Homo sapiens R-HSA-5607763 | 4.32E-01 | 7.54E-01 | 1.89E+00 | 1.59E+00 | CALM1 |
| 618 | Biotin transport and metabolism Homo sapiens R-HSA-196780 | 4.32E-01 | 7.54E-01 | 1.89E+00 | 1.59E+00 | PCCB |
| 619 | CYP2E1 reactions Homo sapiens R-HSA-211999 | 4.32E-01 | 7.54E-01 | 1.89E+00 | 1.59E+00 | CYP2S1 |
| 620 | Heme biosynthesis Homo sapiens R-HSA-189451 | 4.32E-01 | 7.54E-01 | 1.89E+00 | 1.59E+00 | UROS |
| 621 | Signaling by PTK6 Homo sapiens R-HSA-8848021 | 4.35E-01 | 7.57E-01 | 1.20E+00 | 1.00E+00 | PTPN1;UBB;LINC01139;HIF1A |
| 622 | Degradation of the extracellular matrix Homo sapiens R-HSA-1474228 | 4.41E-01 | 7.61E-01 | 1.14E+00 | 9.32E-01 | CAPN6;COL14A1;CTSK;COL12A1;CAPN3;PLG |
| 623 | Nonsense-Mediated Decay (NMD) Homo sapiens R-HSA-927802 | 4.41E-01 | 7.61E-01 | 1.14E+00 | 9.32E-01 | RPS28;RPS27;RPS29;RPL38;RPS4Y1;RPS21 |
| 624 | Nonsense Mediated Decay (NMD) enhanced by the Exon Junction Complex (EJC) Homo sapiens R-HSA-975957 | 4.41E-01 | 7.61E-01 | 1.14E+00 | 9.32E-01 | RPS28;RPS27;RPS29;RPL38;RPS4Y1;RPS21 |
| 625 | L13a-mediated translational silencing of Ceruloplasmin expression Homo sapiens R-HSA-156827 | 4.41E-01 | 7.61E-01 | 1.14E+00 | 9.32E-01 | RPS28;RPS27;RPS29;RPL38;RPS4Y1;RPS21 |
| 626 | 3' -UTR-mediated translational regulation Homo sapiens R-HSA-157279 | 4.41E-01 | 7.61E-01 | 1.14E+00 | 9.32E-01 | RPS28;RPS27;RPS29;RPL38;RPS4Y1;RPS21 |
| 627 | SRP-dependent cotranslational protein targeting to membrane Homo sapiens R-HSA-1799339 | 4.50E-01 | 7.74E-01 | 1.13E+00 | 9.00E-01 | RPS28;RPS27;RPS29;RPL38;RPS4Y1;RPS21 |
| 628 | GTP hydrolysis and joining of the 60S ribosomal subunit Homo sapiens R-HSA-72706 | 4.50E-01 | 7.74E-01 | 1.13E+00 | 9.00E-01 | RPS28;RPS27;RPS29;RPL38;RPS4Y1;RPS21 |

| | | | | | | |
|---|---|---|---|---|---|---|
| 629 | Major pathway of rRNA processing in the nucleolus Homo sapiens R-HSA-6791226 | 4.54E-01 | 7.77E-01 | 1.09E+00 | 8.58E-01 | RRP7A;RPS28;RPS27;RPS29;WDR18;RPS27L;RPL38;RPS4Y1;RPS21 |
| 630 | Loss of Nlp from mitotic centrosomes Homo sapiens R-HSA-380259 | 4.58E-01 | 7.77E-01 | 1.17E+00 | 9.12E-01 | PRKAR2B;SSNA1;CEP63;TUBG1 |
| 631 | Loss of proteins required for interphase microtubule organization?from the centrosome Homo sapiens R-HSA-380284 | 4.58E-01 | 7.77E-01 | 1.17E+00 | 9.12E-01 | PRKAR2B;SSNA1;CEP63;TUBG1 |
| 632 | Influenza Infection Homo sapiens R-HSA-168254 | 4.58E-01 | 7.77E-01 | 1.09E+00 | 8.52E-01 | RPS28;RPS27;RPS29;POLR2E;RPL38;RPS4Y1;POLR2I;RPS21 |
| 633 | Ethanol oxidation Homo sapiens R-HSA-71384 | 4.61E-01 | 7.77E-01 | 1.72E+00 | 1.33E+00 | ALDH1B1 |
| 634 | CD28 dependent Vav1 pathway Homo sapiens R-HSA-389359 | 4.61E-01 | 7.77E-01 | 1.72E+00 | 1.33E+00 | FYN |
| 635 | NF-kB activation through FADD/RIP-1 pathway mediated by caspase-8 and -10 Homo sapiens R-HSA-933543 | 4.61E-01 | 7.77E-01 | 1.72E+00 | 1.33E+00 | RNF135 |
| 636 | PECAM1 interactions Homo sapiens R-HSA-210990 | 4.61E-01 | 7.77E-01 | 1.72E+00 | 1.33E+00 | FYN |
| 637 | Reduction of cytosolic Ca++ levels Homo sapiens R-HSA-418359 | 4.61E-01 | 7.77E-01 | 1.72E+00 | 1.33E+00 | CALM1 |
| 638 | Eicosanoids Homo sapiens R-HSA-211979 | 4.61E-01 | 7.77E-01 | 1.72E+00 | 1.33E+00 | CYP4F2 |
| 639 | Purine ribonucleoside monophosphate biosynthesis Homo sapiens R-HSA-73817 | 4.61E-01 | 7.77E-01 | 1.72E+00 | 1.33E+00 | ADSSL1 |
| 640 | DAP12 signaling Homo sapiens R-HSA-2424491 | 4.61E-01 | 7.77E-01 | 1.05E+00 | 8.11E-01 | KLB;RASGRF1;PEBP1;PHB;FGF2;PSMD8;PSMD9;UBB;PRKAR2B;PSMB3;PSMC4;PSMD2;FGF18;PSME1;LAMTOR2;FYN;CALM1;IER3 |

| | | | | | | |
|---|---|---|---|---|---|---|
| 641 | Class I MHC mediated antigen processing & presentation Homo sapiens R-HSA-983169 | 4.63E-01 | 7.77E-01 | 1.05E+00 | 8.08E-01 | ANAPC13;TRIM63;ZBTB16;FBXO17;CYBA;ANAPC11;PSMD8;PSMD9;ZNRF1;UBB;PSMB3;PSMC4;CDC26;PSMD2;ASB9;PSME1 |
| 642 | Activation of anterior HOX genes in hindbrain development during early embryogenesis Homo sapiens R-HSA-5617472 | 4.63E-01 | 7.77E-01 | 1.13E+00 | 8.68E-01 | H2AFZ;PCGF2;RARA;POLR2E;POLR2I |
| 643 | Activation of HOX genes during differentiation Homo sapiens R-HSA-5619507 | 4.63E-01 | 7.77E-01 | 1.13E+00 | 8.68E-01 | H2AFZ;PCGF2;RARA;POLR2E;POLR2I |
| 644 | Pausing and recovery of Tat-mediated HIV elongation Homo sapiens R-HSA-167238 | 4.65E-01 | 7.77E-01 | 1.31E+00 | 1.00E+00 | POLR2E;POLR2I |
| 645 | Tat-mediated HIV elongation arrest and recovery Homo sapiens R-HSA-167243 | 4.65E-01 | 7.77E-01 | 1.31E+00 | 1.00E+00 | POLR2E;POLR2I |
| 646 | FGFR1 mutant receptor activation Homo sapiens R-HSA-1839124 | 4.65E-01 | 7.77E-01 | 1.31E+00 | 1.00E+00 | STAT5A;FGF2 |
| 647 | Regulation of TNFR1 signaling Homo sapiens R-HSA-5357905 | 4.65E-01 | 7.77E-01 | 1.31E+00 | 1.00E+00 | UBB;USP2 |
| 648 | Negative regulation of the PI3K/AKT network Homo sapiens R-HSA-199418 | 4.73E-01 | 7.82E-01 | 1.11E+00 | 8.34E-01 | KLB;FGF18;FYN;FGF2;IER3 |
| 649 | M Phase Homo sapiens R-HSA-68886 | 4.77E-01 | 7.82E-01 | 1.04E+00 | 7.74E-01 | PRKCB;H2AFZ;RAB1B;ANAPC11;PSMD8;PSMD9;RPS27;UBB;PSMB3;PSMC4;CDC26;PSMD2;PSME1;LPIN2 |
| 650 | G2/M Checkpoints Homo sapiens R-HSA-69481 | 4.81E-01 | 7.82E-01 | 1.07E+00 | 7.82E-01 | PSMD8;PSMD9;UBB;PSMB3;PSMC4;PSMD2;RPA3;PSME1 |
| 651 | Activation of Matrix Metalloproteinases Homo sapiens R-HSA-1592389 | 4.82E-01 | 7.82E-01 | 1.26E+00 | 9.22E-01 | CTSK;PLG |
| 652 | Regulation of beta-cell development Homo sapiens R-HSA-186712 | 4.82E-01 | 7.82E-01 | 1.26E+00 | 9.22E-01 | PKLR;HNF4A |

| | | | | | | |
|---|---|---|---|---|---|---|
| 653 | DNA methylation Homo sapiens R-HSA-5334118 | 4.82E-01 | 7.82E-01 | 1.26E+00 | 9.22E-01 | DNMT3L;H2AFZ |
| 654 | HIV elongation arrest and recovery Homo sapiens R-HSA-167287 | 4.82E-01 | 7.82E-01 | 1.26E+00 | 9.22E-01 | POLR2E;POLR2I |
| 655 | Pausing and recovery of HIV elongation Homo sapiens R-HSA-167290 | 4.82E-01 | 7.82E-01 | 1.26E+00 | 9.22E-01 | POLR2E;POLR2I |
| 656 | Elongation arrest and recovery Homo sapiens R-HSA-112387 | 4.82E-01 | 7.82E-01 | 1.26E+00 | 9.22E-01 | POLR2E;POLR2I |
| 657 | Oncogene Induced Senescence Homo sapiens R-HSA-2559585 | 4.82E-01 | 7.82E-01 | 1.26E+00 | 9.22E-01 | UBB;ETS2 |
| 658 | Negative regulation of FGFR1 signaling Homo sapiens R-HSA-5654726 | 4.82E-01 | 7.82E-01 | 1.26E+00 | 9.22E-01 | UBB;FGF2 |
| 659 | DAG and IP3 signaling Homo sapiens R-HSA-1489509 | 4.82E-01 | 7.82E-01 | 1.26E+00 | 9.22E-01 | PRKAR2B;CALM1 |
| 660 | TGF-beta receptor signaling activates SMADs Homo sapiens R-HSA-2173789 | 4.82E-01 | 7.82E-01 | 1.26E+00 | 9.22E-01 | UBB;STRAP |
| 661 | Dissolution of Fibrin Clot Homo sapiens R-HSA-75205 | 4.88E-01 | 7.82E-01 | 1.58E+00 | 1.13E+00 | PLG |
| 662 | Activation of DNA fragmentation factor Homo sapiens R-HSA-211227 | 4.88E-01 | 7.82E-01 | 1.58E+00 | 1.13E+00 | HIST1H1C |
| 663 | Apoptosis induced DNA fragmentation Homo sapiens R-HSA-140342 | 4.88E-01 | 7.82E-01 | 1.58E+00 | 1.13E+00 | HIST1H1C |
| 664 | POU5F1 (OCT4), SOX2, NANOG activate genes related to proliferation Homo sapiens R-HSA-2892247 | 4.88E-01 | 7.82E-01 | 1.58E+00 | 1.13E+00 | FGF2 |
| 665 | p75NTR recruits signalling complexes Homo sapiens R-HSA-209543 | 4.88E-01 | 7.82E-01 | 1.58E+00 | 1.13E+00 | UBB |

| | | | | | | |
|---|---|---|---|---|---|---|
| 666 | NF-kB is activated and signals survival Homo sapiens R-HSA-209560 | 4.88E-01 | 7.82E-01 | 1.58E+00 | 1.13E+00 | UBB |
| 667 | Assembly Of The HIV Virion Homo sapiens R-HSA-175474 | 4.88E-01 | 7.82E-01 | 1.58E+00 | 1.13E+00 | UBB |
| 668 | Downregulation of ERBB2:ERBB3 signaling Homo sapiens R-HSA-1358803 | 4.88E-01 | 7.82E-01 | 1.58E+00 | 1.13E+00 | UBB |
| 669 | Advanced glycosylation endproduct receptor signaling Homo sapiens R-HSA-879415 | 4.88E-01 | 7.82E-01 | 1.58E+00 | 1.13E+00 | LGALS3 |
| 670 | Trafficking and processing of endosomal TLR Homo sapiens R-HSA-1679131 | 4.88E-01 | 7.82E-01 | 1.58E+00 | 1.13E+00 | CTSK |
| 671 | TNFR1-induced proapoptotic signaling Homo sapiens R-HSA-5357786 | 4.88E-01 | 7.82E-01 | 1.58E+00 | 1.13E+00 | USP2 |
| 672 | Platelet Adhesion to exposed collagen Homo sapiens R-HSA-75892 | 4.88E-01 | 7.82E-01 | 1.58E+00 | 1.13E+00 | FYN |
| 673 | Depolarization of the Presynaptic Terminal Triggers the Opening of Calcium Channels Homo sapiens R-HSA-112308 | 4.88E-01 | 7.82E-01 | 1.58E+00 | 1.13E+00 | CACNA2D1 |
| 674 | RHO GTPases Activate NADPH Oxidases Homo sapiens R-HSA-5668599 | 4.88E-01 | 7.82E-01 | 1.58E+00 | 1.13E+00 | CYBA |
| 675 | AURKA Activation by TPX2 Homo sapiens R-HSA-8854518 | 4.91E-01 | 7.86E-01 | 1.11E+00 | 7.94E-01 | PRKAR2B;SSNA1;CEP63;TUBG1 |
| 676 | Cargo concentration in the ER Homo sapiens R-HSA-5694530 | 4.98E-01 | 7.94E-01 | 1.22E+00 | 8.52E-01 | SERPINA1;GOSR2 |
| 677 | Formation of the Early Elongation Complex Homo sapiens R-HSA-113418 | 4.98E-01 | 7.94E-01 | 1.22E+00 | 8.52E-01 | POLR2E;POLR2I |

| | | | | | | |
|---|---|---|---|---|---|---|
| 678 | Formation of the HIV-1 Early Elongation Complex Homo sapiens R-HSA-167158 | 4.98E-01 | 7.94E-01 | 1.22E+00 | 8.52E-01 | POLR2E;POLR2I |
| 679 | XBP1(S) activates chaperone genes Homo sapiens R-HSA-381038 | 5.00E-01 | 7.97E-01 | 1.14E+00 | 7.87E-01 | GOSR2;WIPI1;ATP6V0D1 |
| 680 | Cap-dependent Translation Initiation Homo sapiens R-HSA-72737 | 5.11E-01 | 8.05E-01 | 1.05E+00 | 7.06E-01 | RPS28;RPS27;RPS29;RPL38;RPS4Y1;RPS21 |
| 681 | Eukaryotic Translation Initiation Homo sapiens R-HSA-72613 | 5.11E-01 | 8.05E-01 | 1.05E+00 | 7.06E-01 | RPS28;RPS27;RPS29;RPL38;RPS4Y1;RPS21 |
| 682 | Regulation of IFNG signaling Homo sapiens R-HSA-877312 | 5.14E-01 | 8.05E-01 | 1.46E+00 | 9.71E-01 | PTPN1 |
| 683 | Receptor-ligand binding initiates the second proteolytic cleavage of Notch receptor Homo sapiens R-HSA-156988 | 5.14E-01 | 8.05E-01 | 1.46E+00 | 9.71E-01 | UBB |
| 684 | TRAF3-dependent IRF activation pathway Homo sapiens R-HSA-918233 | 5.14E-01 | 8.05E-01 | 1.46E+00 | 9.71E-01 | RNF135 |
| 685 | Golgi Cisternae Pericentriolar Stack Reorganization Homo sapiens R-HSA-162658 | 5.14E-01 | 8.05E-01 | 1.46E+00 | 9.71E-01 | RAB1B |
| 686 | DCC mediated attractive signaling Homo sapiens R-HSA-418885 | 5.14E-01 | 8.05E-01 | 1.46E+00 | 9.71E-01 | FYN |
| 687 | HSF1-dependent transactivation Homo sapiens R-HSA-3371571 | 5.14E-01 | 8.05E-01 | 1.18E+00 | 7.88E-01 | UBB;HSPA2 |
| 688 | Negative regulators of RIG-I/MDA5 signaling Homo sapiens R-HSA-936440 | 5.14E-01 | 8.05E-01 | 1.18E+00 | 7.88E-01 | RNF135;UBB |
| 689 | EGFR interacts with phospholipase C-gamma Homo sapiens R-HSA-212718 | 5.14E-01 | 8.05E-01 | 1.18E+00 | 7.88E-01 | PRKAR2B;CALM1 |

| | | | | | | |
|---|---|---|---|---|---|---|
| 690 | PLC-gamma1 signalling Homo sapiens R-HSA-167021 | 5.14E-01 | 8.05E-01 | 1.18E+00 | 7.88E-01 | PRKAR2B;CALM1 |
| 691 | IRE1alpha activates chaperones Homo sapiens R-HSA-381070 | 5.25E-01 | 8.22E-01 | 1.09E+00 | 7.03E-01 | GOSR2;WIPI1;ATP6V0D1 |
| 692 | Regulation of TP53 Degradation Homo sapiens R-HSA-6804757 | 5.30E-01 | 8.22E-01 | 1.15E+00 | 7.29E-01 | UBB;USP2 |
| 693 | Fanconi Anemia Pathway Homo sapiens R-HSA-6783310 | 5.30E-01 | 8.22E-01 | 1.15E+00 | 7.29E-01 | UBB;RPA3 |
| 694 | Post NMDA receptor activation events Homo sapiens R-HSA-438064 | 5.30E-01 | 8.22E-01 | 1.15E+00 | 7.29E-01 | RASGRF1;CALM1 |
| 695 | DAP12 interactions Homo sapiens R-HSA-2172127 | 5.35E-01 | 8.22E-01 | 1.00E+00 | 6.25E-01 | KLB;RASGRF1;PEBP1;PHB;FGF2;PSMD8;PSMD9;UBB;PRKAR2B;PSMB3;PSMC4;PSMD2;FGF18;PSME1;LAMTOR2;FYN;CALM1;IER3 |
| 696 | Membrane Trafficking Homo sapiens R-HSA-199991 | 5.38E-01 | 8.22E-01 | 9.97E-01 | 6.18E-01 | SERPINA1;RAB5C;RAB1B;GOSR2;CLTB;ASPSCR1;PLA2G4A;GJC1;TBC1D1;DAB2;UBB;BLOC1S1;FTH1;MYH8;MAN1C1;NAA38;CALM1;COPE;VAMP3;FTL;AP1M1 |
| 697 | Disorders of transmembrane transporters Homo sapiens R-HSA-5619115 | 5.38E-01 | 8.22E-01 | 1.35E+00 | 8.39E-01 | AVP |
| 698 | mRNA decay by 5' to 3' exoribonuclease Homo sapiens R-HSA-430039 | 5.38E-01 | 8.22E-01 | 1.35E+00 | 8.39E-01 | LSM4 |
| 699 | Regulation of innate immune responses to cytosolic DNA Homo sapiens R-HSA-3134975 | 5.38E-01 | 8.22E-01 | 1.35E+00 | 8.39E-01 | UBB |
| 700 | Prolactin receptor signaling Homo sapiens R-HSA-1170546 | 5.38E-01 | 8.22E-01 | 1.35E+00 | 8.39E-01 | STAT5A |
| 701 | Constitutive Signaling by NOTCH1 HD Domain Mutants Homo sapiens R-HSA-2691232 | 5.38E-01 | 8.22E-01 | 1.35E+00 | 8.39E-01 | UBB |

| | | | | | | |
|---|---|---|---|---|---|---|
| 702 | Signaling by NOTCH1 HD Domain Mutants in Cancer Homo sapiens R-HSA-2691230 | 5.38E-01 | 8.22E-01 | 1.35E+00 | 8.39E-01 | UBB |
| 703 | NRIF signals cell death from the nucleus Homo sapiens R-HSA-205043 | 5.38E-01 | 8.22E-01 | 1.35E+00 | 8.39E-01 | UBB |
| 704 | FGFR1 ligand binding and activation Homo sapiens R-HSA-190242 | 5.38E-01 | 8.22E-01 | 1.35E+00 | 8.39E-01 | FGF2 |
| 705 | CREB phosphorylation through the activation of CaMKII Homo sapiens R-HSA-442729 | 5.38E-01 | 8.22E-01 | 1.35E+00 | 8.39E-01 | CALM1 |
| 706 | Eicosanoid ligand-binding receptors Homo sapiens R-HSA-391903 | 5.38E-01 | 8.22E-01 | 1.35E+00 | 8.39E-01 | PTGER1 |
| 707 | Fatty acids Homo sapiens R-HSA-211935 | 5.38E-01 | 8.22E-01 | 1.35E+00 | 8.39E-01 | CYP4F2 |
| 708 | Senescence-Associated Secretory Phenotype (SASP) Homo sapiens R-HSA-2559582 | 5.44E-01 | 8.28E-01 | 1.04E+00 | 6.32E-01 | UBB;H2AFZ;CDC26;ANAPC11 |
| 709 | RNA polymerase II transcribes snRNA genes Homo sapiens R-HSA-6807505 | 5.44E-01 | 8.28E-01 | 1.04E+00 | 6.32E-01 | SNAPC1;POLR2E;NABP2;POLR2I |
| 710 | RNA Polymerase III Transcription Initiation Homo sapiens R-HSA-76046 | 5.45E-01 | 8.28E-01 | 1.11E+00 | 6.76E-01 | SNAPC1;POLR2E |
| 711 | Regulation of TP53 Expression and Degradation Homo sapiens R-HSA-6806003 | 5.45E-01 | 8.28E-01 | 1.11E+00 | 6.76E-01 | UBB;USP2 |
| 712 | RHO GTPases Activate WASPs and WAVEs Homo sapiens R-HSA-5663213 | 5.45E-01 | 8.28E-01 | 1.11E+00 | 6.76E-01 | ARPC1B;BAIAP2 |
| 713 | Neuronal System Homo sapiens R-HSA-112316 | 5.48E-01 | 8.30E-01 | 9.93E-01 | 5.98E-01 | KCNH6;PRKCB;MAOA;CACNA2D1;RASGRF1;SLC6A12;AP2A1;KCNAB2;GNG11;GLS;GJC1;GNG4;AP2S1;CALM1;SLC38A2 |
| 714 | Meiotic synapsis Homo sapiens R-HSA-1221632 | 5.50E-01 | 8.32E-01 | 1.05E+00 | 6.29E-01 | SYCP2;H2AFZ;HSPA2 |

| | | | | | | |
|---|---|---|---|---|---|---|
| 715 | Biosynthesis of the N-glycan precursor (dolichol lipid-linked oligosaccharide, LLO) and transfer to a nascent protein Homo sapiens R-HSA-446193 | 5.54E-01 | 8.38E-01 | 1.02E+00 | 6.04E-01 | CTSA;SLC17A5;NPL;FUOM |
| 716 | p75NTR signals via NF-kB Homo sapiens R-HSA-193639 | 5.61E-01 | 8.38E-01 | 1.26E+00 | 7.30E-01 | UBB |
| 717 | TRAF6 mediated induction of TAK1 complex Homo sapiens R-HSA-937072 | 5.61E-01 | 8.38E-01 | 1.26E+00 | 7.30E-01 | UBB |
| 718 | Spry regulation of FGF signaling Homo sapiens R-HSA-1295596 | 5.61E-01 | 8.38E-01 | 1.26E+00 | 7.30E-01 | UBB |
| 719 | Regulation of KIT signaling Homo sapiens R-HSA-1433559 | 5.61E-01 | 8.38E-01 | 1.26E+00 | 7.30E-01 | FYN |
| 720 | Metabolism of Angiotensinogen to Angiotensins Homo sapiens R-HSA-2022377 | 5.61E-01 | 8.38E-01 | 1.26E+00 | 7.30E-01 | ENPEP |
| 721 | PKA activation Homo sapiens R-HSA-163615 | 5.61E-01 | 8.38E-01 | 1.26E+00 | 7.30E-01 | PRKAR2B |
| 722 | TGF-beta receptor signaling in EMT (epithelial to mesenchymal transition) Homo sapiens R-HSA-2173791 | 5.61E-01 | 8.38E-01 | 1.26E+00 | 7.30E-01 | UBB |
| 723 | Acyl chain remodelling of PI Homo sapiens R-HSA-1482922 | 5.61E-01 | 8.38E-01 | 1.26E+00 | 7.30E-01 | PLA2G4A |
| 724 | Metabolism of folate and pterines Homo sapiens R-HSA-196757 | 5.61E-01 | 8.38E-01 | 1.26E+00 | 7.30E-01 | MTHFS |
| 725 | Neurotransmitter Receptor Binding And Downstream Transmission In The Postsynaptic Cell Homo sapiens R-HSA-112314 | 5.73E-01 | 8.54E-01 | 9.82E-01 | 5.47E-01 | PRKCB;GNG4;RASGRF1;AP2S1;AP2A1;CALM1;GNG11 |

| | | | | | | |
|---|---|---|---|---|---|---|
| 726 | Signaling by FGFR1 in disease Homo sapiens R-HSA-5655302 | 5.75E-01 | 8.55E-01 | 1.05E+00 | 5.83E-01 | STAT5A;FGF2 |
| 727 | MAP2K and MAPK activation Homo sapiens R-HSA-5674135 | 5.75E-01 | 8.55E-01 | 1.05E+00 | 5.83E-01 | PEBP1;LAMTOR2 |
| 728 | Apoptosis Homo sapiens R-HSA-109581 | 5.76E-01 | 8.55E-01 | 9.77E-01 | 5.39E-01 | PSMD8;PSMD9;UBB;PSMB3;PSMC4;PSMD2;PSME1;HIST1H1C |
| 729 | Hemostasis Homo sapiens R-HSA-109582 | 5.82E-01 | 8.57E-01 | 9.73E-01 | 5.27E-01 | GRB7;SERPINA1;PLG;PRKAR2B;GNG4;FYN;PTPN1;PRKCH;PRKCB;PCDH7;F2R;VEGFB;PLA2G4A;ATP1B3;MIF;PPBP;ATP1B1;GNG11;VEGFA;SOD1;SLC7A7;SLC7A9;MAFG;SERPING1;CALM1;MGLL;PF4 |
| 730 | Zinc transporters Homo sapiens R-HSA-435354 | 5.83E-01 | 8.57E-01 | 1.18E+00 | 6.39E-01 | SLC39A5 |
| 731 | Formation of Senescence-Associated Heterochromatin Foci (SAHF) Homo sapiens R-HSA-2559584 | 5.83E-01 | 8.57E-01 | 1.18E+00 | 6.39E-01 | HIST1H1C |
| 732 | Platelet sensitization by LDL Homo sapiens R-HSA-432142 | 5.83E-01 | 8.57E-01 | 1.18E+00 | 6.39E-01 | PLA2G4A |
| 733 | PKA-mediated phosphorylation of CREB Homo sapiens R-HSA-111931 | 5.83E-01 | 8.57E-01 | 1.18E+00 | 6.39E-01 | PRKAR2B |
| 734 | PKA activation in glucagon signalling Homo sapiens R-HSA-164378 | 5.83E-01 | 8.57E-01 | 1.18E+00 | 6.39E-01 | PRKAR2B |
| 735 | Acyl chain remodelling of PG Homo sapiens R-HSA-1482925 | 5.83E-01 | 8.57E-01 | 1.18E+00 | 6.39E-01 | PLA2G4A |
| 736 | Acyl chain remodelling of PS Homo sapiens R-HSA-1482801 | 5.83E-01 | 8.57E-01 | 1.18E+00 | 6.39E-01 | PLA2G4A |
| 737 | Activation of NMDA receptor upon glutamate binding and postsynaptic events Homo sapiens R-HSA-442755 | 5.89E-01 | 8.64E-01 | 1.02E+00 | 5.42E-01 | RASGRF1;CALM1 |

| | | | | | | |
|---|---|---|---|---|---|---|
| 738 | Global Genome Nucleotide Excision Repair (GG-NER) Homo sapiens R-HSA-5696399 | 5.94E-01 | 8.66E-01 | 9.71E-01 | 5.06E-01 | POLD3;RFC1;UBB;RPA3 |
| 739 | TP53 Regulates Transcription of DNA Repair Genes Homo sapiens R-HSA-6796648 | 5.96E-01 | 8.66E-01 | 9.80E-01 | 5.07E-01 | POLR2E;PMS2;POLR2I |
| 740 | Ca2+ pathway Homo sapiens R-HSA-4086398 | 5.96E-01 | 8.66E-01 | 9.80E-01 | 5.07E-01 | GNG4;CALM1;GNG11 |
| 741 | Axon guidance Homo sapiens R-HSA-422475 | 5.96E-01 | 8.66E-01 | 9.65E-01 | 4.99E-01 | KLB;ARPC1B;RASGRF1;CLTB;PEBP1;AP2A1;PHB;SHB;FGF2;PSMD8;PSMD9;UBB;PSMB3;PSMD2;AP2S1;KDR;FYN;SRGAP3;PLXNA4;VEGFA;PSMC4;FGF18;PSME1;LAMTOR2;CALM1 |
| 742 | Programmed Cell Death Homo sapiens R-HSA-5357801 | 5.97E-01 | 8.66E-01 | 9.59E-01 | 4.95E-01 | PSMD8;PSMD9;UBB;PSMB3;PSMC4;PSMD2;PSME1;HIST1H1C |
| 743 | Negative regulation of MAPK pathway Homo sapiens R-HSA-5675221 | 6.03E-01 | 8.66E-01 | 9.97E-01 | 5.04E-01 | UBB;PEBP1 |
| 744 | Transcriptional activation of mitochondrial biogenesis Homo sapiens R-HSA-2151201 | 6.03E-01 | 8.66E-01 | 9.97E-01 | 5.04E-01 | ESRRA;NR1D1 |
| 745 | Miscellaneous transport and binding events Homo sapiens R-HSA-5223345 | 6.04E-01 | 8.66E-01 | 1.11E+00 | 5.62E-01 | NIPAL3 |
| 746 | Norepinephrine Neurotransmitter Release Cycle Homo sapiens R-HSA-181430 | 6.04E-01 | 8.66E-01 | 1.11E+00 | 5.62E-01 | MAOA |
| 747 | Activation of SMO Homo sapiens R-HSA-5635838 | 6.04E-01 | 8.66E-01 | 1.11E+00 | 5.62E-01 | BOC |
| 748 | RNA Polymerase III Chain Elongation Homo sapiens R-HSA-73780 | 6.04E-01 | 8.66E-01 | 1.11E+00 | 5.62E-01 | POLR2E |
| 749 | Signaling by cytosolic FGFR1 fusion mutants Homo sapiens R-HSA-1839117 | 6.04E-01 | 8.66E-01 | 1.11E+00 | 5.62E-01 | STAT5A |

| | | | | | | |
|---|---|---|---|---|---|---|
| 750 | Tie2 Signaling Homo sapiens R-HSA-210993 | 6.04E-01 | 8.66E-01 | 1.11E+00 | 5.62E-01 | GRB7 |
| 751 | Activation of IRF3/IRF7 mediated by TBK1/IKK epsilon Homo sapiens R-HSA-936964 | 6.04E-01 | 8.66E-01 | 1.11E+00 | 5.62E-01 | UBB |
| 752 | Regulation of signaling by CBL Homo sapiens R-HSA-912631 | 6.04E-01 | 8.66E-01 | 1.11E+00 | 5.62E-01 | FYN |
| 753 | Defects in vitamin and cofactor metabolism Homo sapiens R-HSA-3296482 | 6.04E-01 | 8.66E-01 | 1.11E+00 | 5.62E-01 | PCCB |
| 754 | Amine-derived hormones Homo sapiens R-HSA-209776 | 6.04E-01 | 8.66E-01 | 1.11E+00 | 5.62E-01 | DIO1 |
| 755 | Glycosphingolipid metabolism Homo sapiens R-HSA-1660662 | 6.16E-01 | 8.77E-01 | 9.71E-01 | 4.70E-01 | CTSA;CERK |
| 756 | RNA Polymerase III Abortive And Retractive Initiation Homo sapiens R-HSA-749476 | 6.16E-01 | 8.77E-01 | 9.71E-01 | 4.70E-01 | SNAPC1;POLR2E |
| 757 | RNA Polymerase III Transcription Homo sapiens R-HSA-74158 | 6.16E-01 | 8.77E-01 | 9.71E-01 | 4.70E-01 | SNAPC1;POLR2E |
| 758 | TNF signaling Homo sapiens R-HSA-75893 | 6.16E-01 | 8.77E-01 | 9.71E-01 | 4.70E-01 | UBB;USP2 |
| 759 | Collagen formation Homo sapiens R-HSA-1474290 | 6.23E-01 | 8.77E-01 | 9.35E-01 | 4.43E-01 | P4HA1;COL14A1;P4HA2;COL12A1 |
| 760 | Meiosis Homo sapiens R-HSA-1500620 | 6.23E-01 | 8.77E-01 | 9.35E-01 | 4.43E-01 | SYCP2;H2AFZ;RPA3;HSPA2 |
| 761 | GABA synthesis, release, reuptake and degradation Homo sapiens R-HSA-888590 | 6.24E-01 | 8.77E-01 | 1.05E+00 | 4.96E-01 | SLC6A12 |
| 762 | Gap junction assembly Homo sapiens R-HSA-190861 | 6.24E-01 | 8.77E-01 | 1.05E+00 | 4.96E-01 | GJC1 |
| 763 | Regulation of TP53 Activity through Methylation Homo sapiens R-HSA-6804760 | 6.24E-01 | 8.77E-01 | 1.05E+00 | 4.96E-01 | UBB |

| | | | | | | |
|---|---|---|---|---|---|---|
| 764 | Constitutive Signaling by Ligand-Responsive EGFR Cancer Variants Homo sapiens R-HSA-1236382 | 6.24E-01 | 8.77E-01 | 1.05E+00 | 4.96E-01 | UBB |
| 765 | Signaling by Ligand-Responsive EGFR Variants in Cancer Homo sapiens R-HSA-5637815 | 6.24E-01 | 8.77E-01 | 1.05E+00 | 4.96E-01 | UBB |
| 766 | Signaling by EGFR in Cancer Homo sapiens R-HSA-1643713 | 6.24E-01 | 8.77E-01 | 1.05E+00 | 4.96E-01 | UBB |
| 767 | Other semaphorin interactions Homo sapiens R-HSA-416700 | 6.24E-01 | 8.77E-01 | 1.05E+00 | 4.96E-01 | PLXNA4 |
| 768 | Ephrin signaling Homo sapiens R-HSA-3928664 | 6.24E-01 | 8.77E-01 | 1.05E+00 | 4.96E-01 | FYN |
| 769 | Phase 4 - resting membrane potential Homo sapiens R-HSA-5576886 | 6.24E-01 | 8.77E-01 | 1.05E+00 | 4.96E-01 | KCNK5 |
| 770 | Viral Messenger RNA Synthesis Homo sapiens R-HSA-168325 | 6.30E-01 | 8.80E-01 | 9.47E-01 | 4.38E-01 | POLR2E;POLR2I |
| 771 | EPHB-mediated forward signaling Homo sapiens R-HSA-3928662 | 6.30E-01 | 8.80E-01 | 9.47E-01 | 4.38E-01 | ARPC1B;FYN |
| 772 | Cooperation of PDCL (PhLP1) and TRiC/CCT in G-protein beta folding Homo sapiens R-HSA-6814122 | 6.30E-01 | 8.80E-01 | 9.47E-01 | 4.38E-01 | GNG4;GNG11 |
| 773 | tRNA Aminoacylation Homo sapiens R-HSA-379724 | 6.30E-01 | 8.80E-01 | 9.47E-01 | 4.38E-01 | SARS2;SARS |
| 774 | Chromosome Maintenance Homo sapiens R-HSA-73886 | 6.32E-01 | 8.81E-01 | 9.24E-01 | 4.24E-01 | POLD3;RFC1;H2AFZ;RPA3 |
| 775 | PI3K/AKT Signaling in Cancer Homo sapiens R-HSA-2219528 | 6.32E-01 | 8.81E-01 | 9.24E-01 | 4.24E-01 | KLB;FGF18;FYN;FGF2 |
| 776 | Cell Cycle Homo sapiens R-HSA-1640170 | 6.35E-01 | 8.83E-01 | 9.47E-01 | 4.31E-01 | SYCP2;TUBGCP2;RAB1B;SSNA1;GMNN;ANAPC11;PSMD8;POLD3;PSMD9;UBB;PRKAR2B;PSMB3;PSMD2;CDC26;RFC1;PRKCB;H2AFZ;HSPA2;TUBG1;CDC25B;RPS27;PSMC4;RPA3;PSME1;CEP63;LPIN2;OPTN |

| | | | | | | |
|---|---|---|---|---|---|---|
| 777 | Platelet homeostasis Homo sapiens R-HSA-418346 | 6.41E-01 | 8.83E-01 | 9.12E-01 | 4.06E-01 | GNG4;PLA2G4A;CALM1;GNG11 |
| 778 | HIV Transcription Elongation Homo sapiens R-HSA-167169 | 6.42E-01 | 8.83E-01 | 9.24E-01 | 4.09E-01 | POLR2E;POLR2I |
| 779 | Tat-mediated elongation of the HIV-1 transcript Homo sapiens R-HSA-167246 | 6.42E-01 | 8.83E-01 | 9.24E-01 | 4.09E-01 | POLR2E;POLR2I |
| 780 | Formation of HIV-1 elongation complex containing HIV-1 Tat Homo sapiens R-HSA-167200 | 6.42E-01 | 8.83E-01 | 9.24E-01 | 4.09E-01 | POLR2E;POLR2I |
| 781 | Formation of Incision Complex in GG-NER Homo sapiens R-HSA-5696395 | 6.42E-01 | 8.83E-01 | 9.24E-01 | 4.09E-01 | UBB;RPA3 |
| 782 | Voltage gated Potassium channels Homo sapiens R-HSA-1296072 | 6.42E-01 | 8.83E-01 | 9.24E-01 | 4.09E-01 | KCNH6;KCNAB2 |
| 783 | IL-6-type cytokine receptor ligand interactions Homo sapiens R-HSA-6788467 | 6.43E-01 | 8.83E-01 | 9.97E-01 | 4.40E-01 | CNTF |
| 784 | Signaling by Hippo Homo sapiens R-HSA-2028269 | 6.43E-01 | 8.83E-01 | 9.97E-01 | 4.40E-01 | MOB1A |
| 785 | Synthesis of PC Homo sapiens R-HSA-1483191 | 6.43E-01 | 8.83E-01 | 9.97E-01 | 4.40E-01 | LPIN2 |
| 786 | Xenobiotics Homo sapiens R-HSA-211981 | 6.43E-01 | 8.83E-01 | 9.97E-01 | 4.40E-01 | CYP2S1 |
| 787 | Intra-Golgi traffic Homo sapiens R-HSA-6811438 | 6.55E-01 | 8.83E-01 | 9.02E-01 | 3.82E-01 | GOSR2;MAN1C1 |
| 788 | Formation of HIV elongation complex in the absence of HIV Tat Homo sapiens R-HSA-167152 | 6.55E-01 | 8.83E-01 | 9.02E-01 | 3.82E-01 | POLR2E;POLR2I |
| 789 | RNA Polymerase II Transcription Elongation Homo sapiens R-HSA-75955 | 6.55E-01 | 8.83E-01 | 9.02E-01 | 3.82E-01 | POLR2E;POLR2I |

| | | | | | | |
|---|---|---|---|---|---|---|
| 790 | Formation of RNA Pol II elongation complex Homo sapiens R-HSA-112382 | 6.55E-01 | 8.83E-01 | 9.02E-01 | 3.82E-01 | POLR2E;POLR2I |
| 791 | RNA Polymerase II HIV Promoter Escape Homo sapiens R-HSA-167162 | 6.55E-01 | 8.83E-01 | 9.02E-01 | 3.82E-01 | POLR2E;POLR2I |
| 792 | RNA Polymerase II Promoter Escape Homo sapiens R-HSA-73776 | 6.55E-01 | 8.83E-01 | 9.02E-01 | 3.82E-01 | POLR2E;POLR2I |
| 793 | RNA Polymerase II Transcription Initiation And Promoter Clearance Homo sapiens R-HSA-76042 | 6.55E-01 | 8.83E-01 | 9.02E-01 | 3.82E-01 | POLR2E;POLR2I |
| 794 | RNA Polymerase II Transcription Initiation Homo sapiens R-HSA-75953 | 6.55E-01 | 8.83E-01 | 9.02E-01 | 3.82E-01 | POLR2E;POLR2I |
| 795 | RNA Polymerase II Transcription Pre-Initiation And Promoter Opening Homo sapiens R-HSA-73779 | 6.55E-01 | 8.83E-01 | 9.02E-01 | 3.82E-01 | POLR2E;POLR2I |
| 796 | HIV Transcription Initiation Homo sapiens R-HSA-167161 | 6.55E-01 | 8.83E-01 | 9.02E-01 | 3.82E-01 | POLR2E;POLR2I |
| 797 | Interleukin-1 signaling Homo sapiens R-HSA-446652 | 6.55E-01 | 8.83E-01 | 9.02E-01 | 3.82E-01 | UBB;TNIP2 |
| 798 | Phase 0 - rapid depolarisation Homo sapiens R-HSA-5576892 | 6.55E-01 | 8.83E-01 | 9.02E-01 | 3.82E-01 | CACNA2D1;CALM1 |
| 799 | HS-GAG degradation Homo sapiens R-HSA-2024096 | 6.61E-01 | 8.83E-01 | 9.47E-01 | 3.92E-01 | NAGLU |
| 800 | TP53 regulates transcription of additional cell cycle genes whose exact role in the p53 pathway remain uncertain Homo sapiens R-HSA-6804115 | 6.61E-01 | 8.83E-01 | 9.47E-01 | 3.92E-01 | RGCC |

| | | | | | | |
|---|---|---|---|---|---|---|
| 801 | Translocation of ZAP-70 to Immunological synapse Homo sapiens R-HSA-202430 | 6.61E-01 | 8.83E-01 | 9.47E-01 | 3.92E-01 | HLA-DPB1 |
| 802 | NOTCH2 Activation and Transmission of Signal to the Nucleus Homo sapiens R-HSA-2979096 | 6.61E-01 | 8.83E-01 | 9.47E-01 | 3.92E-01 | UBB |
| 803 | SHC-mediated cascade:FGFR1 Homo sapiens R-HSA-5654688 | 6.61E-01 | 8.83E-01 | 9.47E-01 | 3.92E-01 | FGF2 |
| 804 | Regulation of FZD by ubiquitination Homo sapiens R-HSA-4641263 | 6.61E-01 | 8.83E-01 | 9.47E-01 | 3.92E-01 | UBB |
| 805 | CD209 (DC-SIGN) signaling Homo sapiens R-HSA-5621575 | 6.61E-01 | 8.83E-01 | 9.47E-01 | 3.92E-01 | FYN |
| 806 | RHO GTPases activate PAKs Homo sapiens R-HSA-5627123 | 6.61E-01 | 8.83E-01 | 9.47E-01 | 3.92E-01 | CALM1 |
| 807 | VxPx cargo-targeting to cilium Homo sapiens R-HSA-5620916 | 6.61E-01 | 8.83E-01 | 9.47E-01 | 3.92E-01 | RAB11FIP3 |
| 808 | Synthesis of very long-chain fatty acyl-CoAs Homo sapiens R-HSA-75876 | 6.61E-01 | 8.83E-01 | 9.47E-01 | 3.92E-01 | ELOVL4 |
| 809 | Mitochondrial tRNA aminoacylation Homo sapiens R-HSA-379726 | 6.61E-01 | 8.83E-01 | 9.47E-01 | 3.92E-01 | SARS2 |
| 810 | Signalling by NGF Homo sapiens R-HSA-166520 | 6.65E-01 | 8.88E-01 | 9.26E-01 | 3.78E-01 | KLB;RASGRF1;PEBP1;AP2A1;PHB;FGF2;PCSK5;PSMD8;PSMD9;UBB;PRKAR2B;PSMB3;PSMC4;PSMD2;FGF18;PSME1;AP2S1;LAMTOR2;FYN;CALM1;IER3 |
| 811 | SUMOylation of RNA binding proteins Homo sapiens R-HSA-4570464 | 6.67E-01 | 8.89E-01 | 8.81E-01 | 3.57E-01 | CBX2;PCGF2 |
| 812 | mRNA Splicing - Major Pathway Homo sapiens R-HSA-72163 | 6.69E-01 | 8.90E-01 | 8.87E-01 | 3.57E-01 | SNRPG;POLR2E;DNAJC8;TXNL4A;POLR2I;LSM4 |
| 813 | COPII (Coat Protein 2) Mediated Vesicle Transport Homo sapiens R-HSA-204005 | 6.70E-01 | 8.90E-01 | 8.74E-01 | 3.50E-01 | SERPINA1;RAB1B;GOSR2 |

| | | | | | | |
|---|---|---|---|---|---|---|
| 814 | TCF dependent signaling in response to WNT Homo sapiens R-HSA-201681 | 6.72E-01 | 8.92E-01 | 8.96E-01 | 3.57E-01 | PSMD8;PSMD9;UBB;PSMB3;PSMC4;H2AFZ;PSMD2;CCDC88C;PSME1 |
| 815 | Intrinsic Pathway of Fibrin Clot Formation Homo sapiens R-HSA-140837 | 6.78E-01 | 8.94E-01 | 9.02E-01 | 3.51E-01 | SERPING1 |
| 816 | CD28 dependent PI3K/Akt signaling Homo sapiens R-HSA-389357 | 6.78E-01 | 8.94E-01 | 9.02E-01 | 3.51E-01 | FYN |
| 817 | CTLA4 inhibitory signaling Homo sapiens R-HSA-389513 | 6.78E-01 | 8.94E-01 | 9.02E-01 | 3.51E-01 | FYN |
| 818 | Cyclin A/B1 associated events during G2/M transition Homo sapiens R-HSA-69273 | 6.78E-01 | 8.94E-01 | 9.02E-01 | 3.51E-01 | CDC25B |
| 819 | Nephrin interactions Homo sapiens R-HSA-373753 | 6.78E-01 | 8.94E-01 | 9.02E-01 | 3.51E-01 | FYN |
| 820 | Post-chaperonin tubulin folding pathway Homo sapiens R-HSA-389977 | 6.78E-01 | 8.94E-01 | 9.02E-01 | 3.51E-01 | ARL2 |
| 821 | Antigen activates B Cell Receptor (BCR) leading to generation of second messengers Homo sapiens R-HSA-983695 | 6.90E-01 | 9.07E-01 | 8.41E-01 | 3.12E-01 | FYN;CALM1 |
| 822 | RNA Polymerase III Transcription Termination Homo sapiens R-HSA-73980 | 6.94E-01 | 9.07E-01 | 8.61E-01 | 3.14E-01 | POLR2E |
| 823 | RAF-independent MAPK1/3 activation Homo sapiens R-HSA-112409 | 6.94E-01 | 9.07E-01 | 8.61E-01 | 3.14E-01 | IL6R |
| 824 | Downregulation of SMAD2/3:SMAD4 transcriptional activity Homo sapiens R-HSA-2173795 | 6.94E-01 | 9.07E-01 | 8.61E-01 | 3.14E-01 | UBB |
| 825 | Signaling by BMP Homo sapiens R-HSA-201451 | 6.94E-01 | 9.07E-01 | 8.61E-01 | 3.14E-01 | CHRDL1 |

| | | | | | | |
|---|---|---|---|---|---|---|
| 826 | Acyl chain remodelling of PE Homo sapiens R-HSA-1482839 | 6.94E-01 | 9.07E-01 | 8.61E-01 | 3.14E-01 | PLA2G4A |
| 827 | Cholesterol biosynthesis Homo sapiens R-HSA-191273 | 6.94E-01 | 9.07E-01 | 8.61E-01 | 3.14E-01 | MVK |
| 828 | Death Receptor Signalling Homo sapiens R-HSA-73887 | 7.01E-01 | 9.14E-01 | 8.23E-01 | 2.92E-01 | UBB;USP2 |
| 829 | G beta:gamma signalling through PI3Kgamma Homo sapiens R-HSA-392451 | 7.01E-01 | 9.14E-01 | 8.23E-01 | 2.92E-01 | GNG4;GNG11 |
| 830 | Cellular Senescence Homo sapiens R-HSA-2559583 | 7.03E-01 | 9.14E-01 | 8.60E-01 | 3.03E-01 | UBB;H2AFZ;CDC26;CBX2;ETS2;ANAPC11;HIST1H1C |
| 831 | Phospholipid metabolism Homo sapiens R-HSA-1483257 | 7.03E-01 | 9.14E-01 | 8.60E-01 | 3.03E-01 | DMGDH;ETNK2;CPNE6;PLA2G4A;GDPD3;LPIN2;MGLL |
| 832 | Transcriptional regulation by small RNAs Homo sapiens R-HSA-5578749 | 7.07E-01 | 9.14E-01 | 8.23E-01 | 2.85E-01 | H2AFZ;POLR2E;POLR2I |
| 833 | Signaling by NOTCH1 Homo sapiens R-HSA-1980143 | 7.07E-01 | 9.14E-01 | 8.23E-01 | 2.85E-01 | UBB;HIF1A;HDAC6 |
| 834 | Phosphorylation of CD3 and TCR zeta chains Homo sapiens R-HSA-202427 | 7.09E-01 | 9.14E-01 | 8.23E-01 | 2.83E-01 | HLA-DPB1 |
| 835 | Post-Elongation Processing of Intronless pre-mRNA Homo sapiens R-HSA-112297 | 7.09E-01 | 9.14E-01 | 8.23E-01 | 2.83E-01 | SNRPG |
| 836 | Processing of Capped Intronless Pre-mRNA Homo sapiens R-HSA-75067 | 7.09E-01 | 9.14E-01 | 8.23E-01 | 2.83E-01 | SNRPG |
| 837 | TRAF6 mediated NF-kB activation Homo sapiens R-HSA-933542 | 7.09E-01 | 9.14E-01 | 8.23E-01 | 2.83E-01 | RNF135 |
| 838 | IKK complex recruitment mediated by RIP1 Homo sapiens R-HSA-937041 | 7.09E-01 | 9.14E-01 | 8.23E-01 | 2.83E-01 | UBB |
| 839 | Cytosolic tRNA aminoacylation Homo sapiens R-HSA-379716 | 7.09E-01 | 9.14E-01 | 8.23E-01 | 2.83E-01 | SARS |

| | | | | | | |
|---|---|---|---|---|---|---|
| 840 | Retrograde transport at the Trans-Golgi-Network Homo sapiens R-HSA-6811440 | 7.12E-01 | 9.15E-01 | 8.06E-01 | 2.74E-01 | NAA38;VAMP3 |
| 841 | Mitochondrial biogenesis Homo sapiens R-HSA-1592230 | 7.12E-01 | 9.15E-01 | 8.06E-01 | 2.74E-01 | ESRRA;NR1D1 |
| 842 | G alpha (s) signalling events Homo sapiens R-HSA-418555 | 7.22E-01 | 9.18E-01 | 8.35E-01 | 2.72E-01 | CALCA;GIPR;GNG4;ADM;AVP;GNG11 |
| 843 | Anchoring of the basal body to the plasma membrane Homo sapiens R-HSA-5620912 | 7.23E-01 | 9.18E-01 | 8.14E-01 | 2.64E-01 | PRKAR2B;SSNA1;CEP63;TUBG1 |
| 844 | Metal ion SLC transporters Homo sapiens R-HSA-425410 | 7.24E-01 | 9.18E-01 | 7.89E-01 | 2.55E-01 | SLC39A5 |
| 845 | Regulation of IFNA signaling Homo sapiens R-HSA-912694 | 7.24E-01 | 9.18E-01 | 7.89E-01 | 2.55E-01 | PTPN1 |
| 846 | Budding and maturation of HIV virion Homo sapiens R-HSA-162588 | 7.24E-01 | 9.18E-01 | 7.89E-01 | 2.55E-01 | UBB |
| 847 | RAF activation Homo sapiens R-HSA-5673000 | 7.24E-01 | 9.18E-01 | 7.89E-01 | 2.55E-01 | PHB |
| 848 | Activation of G protein gated Potassium channels Homo sapiens R-HSA-1296041 | 7.24E-01 | 9.18E-01 | 7.89E-01 | 2.55E-01 | GNG4 |
| 849 | Inhibition of voltage gated Ca2+ channels via Gbeta/gamma subunits Homo sapiens R-HSA-997272 | 7.24E-01 | 9.18E-01 | 7.89E-01 | 2.55E-01 | GNG4 |
| 850 | G protein gated Potassium channels Homo sapiens R-HSA-1296059 | 7.24E-01 | 9.18E-01 | 7.89E-01 | 2.55E-01 | GNG4 |
| 851 | Acyl chain remodelling of PC Homo sapiens R-HSA-1482788 | 7.24E-01 | 9.18E-01 | 7.89E-01 | 2.55E-01 | PLA2G4A |

| | | | | | | |
|---|---|---|---|---|---|---|
| 852 | Binding and Uptake of Ligands by Scavenger Receptors Homo sapiens R-HSA-2173782 | 7.24E-01 | 9.18E-01 | 8.00E-01 | 2.58E-01 | FTH1;APOE;FTL |
| 853 | Nuclear Envelope Breakdown Homo sapiens R-HSA-2980766 | 7.33E-01 | 9.18E-01 | 7.73E-01 | 2.40E-01 | PRKCB;LPIN2 |
| 854 | G-protein beta:gamma signalling Homo sapiens R-HSA-397795 | 7.33E-01 | 9.18E-01 | 7.73E-01 | 2.40E-01 | GNG4;GNG11 |
| 855 | SUMOylation of DNA damage response and repair proteins Homo sapiens R-HSA-3108214 | 7.33E-01 | 9.18E-01 | 7.89E-01 | 2.45E-01 | SP100;CBX2;PCGF2 |
| 856 | mRNA Splicing Homo sapiens R-HSA-72172 | 7.34E-01 | 9.18E-01 | 8.22E-01 | 2.54E-01 | SNRPG;POLR2E;DNAJC8;TXNL4A;POLR2I;LSM4 |
| 857 | Fertilization Homo sapiens R-HSA-1187000 | 7.38E-01 | 9.18E-01 | 7.57E-01 | 2.30E-01 | ZP3 |
| 858 | Reproduction Homo sapiens R-HSA-1474165 | 7.38E-01 | 9.18E-01 | 7.57E-01 | 2.30E-01 | ZP3 |
| 859 | A tetrasaccharide linker sequence is required for GAG synthesis Homo sapiens R-HSA-1971475 | 7.38E-01 | 9.18E-01 | 7.57E-01 | 2.30E-01 | B3GAT1 |
| 860 | PD-1 signaling Homo sapiens R-HSA-389948 | 7.38E-01 | 9.18E-01 | 7.57E-01 | 2.30E-01 | HLA-DPB1 |
| 861 | TNFR1-induced NFkappaB signaling pathway Homo sapiens R-HSA-5357956 | 7.38E-01 | 9.18E-01 | 7.57E-01 | 2.30E-01 | USP2 |
| 862 | WNT ligand biogenesis and trafficking Homo sapiens R-HSA-3238698 | 7.38E-01 | 9.18E-01 | 7.57E-01 | 2.30E-01 | WNT7B |
| 863 | Phase 2 - plateau phase Homo sapiens R-HSA-5576893 | 7.38E-01 | 9.18E-01 | 7.57E-01 | 2.30E-01 | CACNA2D1 |
| 864 | PI-3K cascade:FGFR1 Homo sapiens R-HSA-5654689 | 7.38E-01 | 9.18E-01 | 8.08E-01 | 2.45E-01 | KLB;FGF18;FYN;FGF2;IER3 |

| | | | | | | |
|---|---|---|---|---|---|---|
| 865 | PI-3K cascade:FGFR3 Homo sapiens R-HSA-5654710 | 7.38E-01 | 9.18E-01 | 8.08E-01 | 2.45E-01 | KLB;FGF18;FYN;FGF2;IER3 |
| 866 | PI3K events in ERBB4 signaling Homo sapiens R-HSA-1250342 | 7.38E-01 | 9.18E-01 | 8.08E-01 | 2.45E-01 | KLB;FGF18;FYN;FGF2;IER3 |
| 867 | PIP3 activates AKT signaling Homo sapiens R-HSA-1257604 | 7.38E-01 | 9.18E-01 | 8.08E-01 | 2.45E-01 | KLB;FGF18;FYN;FGF2;IER3 |
| 868 | PI-3K cascade:FGFR4 Homo sapiens R-HSA-5654720 | 7.38E-01 | 9.18E-01 | 8.08E-01 | 2.45E-01 | KLB;FGF18;FYN;FGF2;IER3 |
| 869 | PI-3K cascade:FGFR2 Homo sapiens R-HSA-5654695 | 7.38E-01 | 9.18E-01 | 8.08E-01 | 2.45E-01 | KLB;FGF18;FYN;FGF2;IER3 |
| 870 | Developmental Biology Homo sapiens R-HSA-1266738 | 7.39E-01 | 9.18E-01 | 9.06E-01 | 2.74E-01 | KLB;ARPC1B;RASGRF1;CLTB;PEBP1;AP2A1;PHB;SHB;FGF2;PSMD8;PSMD9;MED11;UBB;PSMB3;PSMD2;HNF4A;BOC;AP2S1;KDR;POLR2E;FYN;SRGAP3;PCK1;POLR2I;PLXNA4;PKLR;H2AFZ;PCGF2;VEGFA;PSMC4;FGF18;PSME1;RARA;LAMTOR2;CDH15;CALM1 |
| 871 | Vesicle-mediated transport Homo sapiens R-HSA-5653656 | 7.41E-01 | 9.20E-01 | 8.84E-01 | 2.65E-01 | SERPINA1;RAB5C;RAB1B;GOSR2;CLTB;ASPSCR1;PLA2G4A;GJC1;TBC1D1;DAB2;UBB;BLOC1S1;FTH1;MYH8;MAN1C1;NAA38;APOE;CALM1;COPE;VAMP3;FTL;AP1M1 |
| 872 | Stimuli-sensing channels Homo sapiens R-HSA-2672351 | 7.45E-01 | 9.24E-01 | 7.88E-01 | 2.32E-01 | CLCN7;UBB;CALM1;TPCN1 |
| 873 | Uptake and actions of bacterial toxins Homo sapiens R-HSA-5339562 | 7.51E-01 | 9.25E-01 | 7.28E-01 | 2.09E-01 | CALM1 |
| 874 | VEGFR2 mediated vascular permeability Homo sapiens R-HSA-5218920 | 7.51E-01 | 9.25E-01 | 7.28E-01 | 2.09E-01 | CALM1 |
| 875 | RNA Polymerase III Transcription Initiation From Type 2 Promoter Homo sapiens R-HSA-76066 | 7.51E-01 | 9.25E-01 | 7.28E-01 | 2.09E-01 | POLR2E |
| 876 | Integrin alphaIIb beta3 signaling Homo sapiens R-HSA-354192 | 7.51E-01 | 9.25E-01 | 7.28E-01 | 2.09E-01 | PTPN1 |

| | | | | | | |
|---|---|---|---|---|---|---|
| 877 | Platelet calcium homeostasis Homo sapiens R-HSA-418360 | 7.51E-01 | 9.25E-01 | 7.28E-01 | 2.09E-01 | CALM1 |
| 878 | Phase 1 - inactivation of fast Na+ channels Homo sapiens R-HSA-5576894 | 7.51E-01 | 9.25E-01 | 7.28E-01 | 2.09E-01 | CACNA2D1 |
| 879 | COPI-mediated anterograde transport Homo sapiens R-HSA-6807878 | 7.57E-01 | 9.29E-01 | 7.57E-01 | 2.11E-01 | RAB1B;GOSR2;COPE |
| 880 | GAB1 signalosome Homo sapiens R-HSA-180292 | 7.57E-01 | 9.29E-01 | 7.88E-01 | 2.19E-01 | KLB;FGF18;FYN;FGF2;IER3 |
| 881 | PI3K/AKT activation Homo sapiens R-HSA-198203 | 7.57E-01 | 9.29E-01 | 7.88E-01 | 2.19E-01 | KLB;FGF18;FYN;FGF2;IER3 |
| 882 | Heparan sulfate/heparin (HS-GAG) metabolism Homo sapiens R-HSA-1638091 | 7.61E-01 | 9.31E-01 | 7.28E-01 | 1.99E-01 | NAGLU;B3GAT1 |
| 883 | Meiotic recombination Homo sapiens R-HSA-912446 | 7.61E-01 | 9.31E-01 | 7.28E-01 | 1.99E-01 | H2AFZ;RPA3 |
| 884 | RNA Polymerase III Transcription Initiation From Type 1 Promoter Homo sapiens R-HSA-76061 | 7.63E-01 | 9.31E-01 | 7.01E-01 | 1.89E-01 | POLR2E |
| 885 | Endosomal Sorting Complex Required For Transport (ESCRT) Homo sapiens R-HSA-917729 | 7.63E-01 | 9.31E-01 | 7.01E-01 | 1.89E-01 | UBB |
| 886 | Synthesis of PA Homo sapiens R-HSA-1483166 | 7.63E-01 | 9.31E-01 | 7.01E-01 | 1.89E-01 | PLA2G4A |
| 887 | Extracellular matrix organization Homo sapiens R-HSA-1474244 | 7.64E-01 | 9.32E-01 | 8.37E-01 | 2.25E-01 | VTN;CAPN6;P4HA1;P4HA2;COL14A1;CTSK;COL12A1;KDR;CAPN3;PLG;FGF2;DDR2 |
| 888 | Translation Homo sapiens R-HSA-72766 | 7.74E-01 | 9.39E-01 | 7.82E-01 | 2.00E-01 | RPS28;RPS27;RPS29;RPL38;RPS4Y1;RPS21 |
| 889 | COPI-independent Golgi-to-ER retrograde traffic Homo sapiens R-HSA-6811436 | 7.75E-01 | 9.39E-01 | 6.76E-01 | 1.72E-01 | PLA2G4A |

| | | | | | | |
|---|---|---|---|---|---|---|
| 890 | Fc epsilon receptor (FCERI) signaling Homo sapiens R-HSA-2454202 | 7.75E-01 | 9.39E-01 | 8.49E-01 | 2.16E-01 | KLB;RASGRF1;PEBP1;PHB;FGF2;PSMD8;PSMD9;UBB;PSMB3;PSMC4;PSMD2;FGF18;PSME1;LAMTOR2;FYN;CALM1;IER3 |
| 891 | Chemokine receptors bind chemokines Homo sapiens R-HSA-380108 | 7.79E-01 | 9.39E-01 | 7.01E-01 | 1.75E-01 | PPBP;PF4 |
| 892 | Toll Like Receptor 10 (TLR10) Cascade Homo sapiens R-HSA-168142 | 7.86E-01 | 9.39E-01 | 7.18E-01 | 1.73E-01 | UBB;TNIP2;ECSIT |
| 893 | Toll Like Receptor 5 (TLR5) Cascade Homo sapiens R-HSA-168176 | 7.86E-01 | 9.39E-01 | 7.18E-01 | 1.73E-01 | UBB;TNIP2;ECSIT |
| 894 | MyD88 cascade initiated on plasma membrane Homo sapiens R-HSA-975871 | 7.86E-01 | 9.39E-01 | 7.18E-01 | 1.73E-01 | UBB;TNIP2;ECSIT |
| 895 | Molecules associated with elastic fibres Homo sapiens R-HSA-2129379 | 7.87E-01 | 9.39E-01 | 6.53E-01 | 1.57E-01 | VTN |
| 896 | RNA Polymerase I Promoter Opening Homo sapiens R-HSA-73728 | 7.87E-01 | 9.39E-01 | 6.53E-01 | 1.57E-01 | H2AFZ |
| 897 | RNA Polymerase I Promoter Escape Homo sapiens R-HSA-73772 | 7.87E-01 | 9.39E-01 | 6.53E-01 | 1.57E-01 | POLR2E |
| 898 | Activation of the pre-replicative complex Homo sapiens R-HSA-68962 | 7.87E-01 | 9.39E-01 | 6.53E-01 | 1.57E-01 | RPA3 |
| 899 | Activated NOTCH1 Transmits Signal to the Nucleus Homo sapiens R-HSA-2122948 | 7.87E-01 | 9.39E-01 | 6.53E-01 | 1.57E-01 | UBB |
| 900 | Tight junction interactions Homo sapiens R-HSA-420029 | 7.87E-01 | 9.39E-01 | 6.53E-01 | 1.57E-01 | CLDN1 |
| 901 | RNA Polymerase I Chain Elongation Homo sapiens R-HSA-73777 | 7.87E-01 | 9.39E-01 | 6.88E-01 | 1.65E-01 | H2AFZ;POLR2E |

| | | | | | | |
|---|---|---|---|---|---|---|
| 902 | Constitutive Signaling by NOTCH1 HD+PEST Domain Mutants Homo sapiens R-HSA-2894862 | 7.87E-01 | 9.39E-01 | 6.88E-01 | 1.65E-01 | UBB;HDAC6 |
| 903 | Signaling by NOTCH1 in Cancer Homo sapiens R-HSA-2644603 | 7.87E-01 | 9.39E-01 | 6.88E-01 | 1.65E-01 | UBB;HDAC6 |
| 904 | Signaling by NOTCH1 PEST Domain Mutants in Cancer Homo sapiens R-HSA-2644602 | 7.87E-01 | 9.39E-01 | 6.88E-01 | 1.65E-01 | UBB;HDAC6 |
| 905 | Constitutive Signaling by NOTCH1 PEST Domain Mutants Homo sapiens R-HSA-2644606 | 7.87E-01 | 9.39E-01 | 6.88E-01 | 1.65E-01 | UBB;HDAC6 |
| 906 | Signaling by NOTCH1 HD+PEST Domain Mutants in Cancer Homo sapiens R-HSA-2894858 | 7.87E-01 | 9.39E-01 | 6.88E-01 | 1.65E-01 | UBB;HDAC6 |
| 907 | Mitotic Prophase Homo sapiens R-HSA-68875 | 7.91E-01 | 9.42E-01 | 7.34E-01 | 1.72E-01 | PRKCB;H2AFZ;RAB1B;LPIN2 |
| 908 | TRAF6 mediated induction of NFkB and MAP kinases upon TLR7/8 or 9 activation Homo sapiens R-HSA-975138 | 7.93E-01 | 9.44E-01 | 7.09E-01 | 1.65E-01 | UBB;TNIP2;ECSIT |
| 909 | rRNA modification in the nucleus Homo sapiens R-HSA-6790901 | 7.95E-01 | 9.44E-01 | 6.76E-01 | 1.55E-01 | RRP7A;NOP10 |
| 910 | B-WICH complex positively regulates rRNA expression Homo sapiens R-HSA-5250924 | 7.95E-01 | 9.44E-01 | 6.76E-01 | 1.55E-01 | H2AFZ;POLR2E |
| 911 | Packaging Of Telomere Ends Homo sapiens R-HSA-171306 | 7.97E-01 | 9.44E-01 | 6.31E-01 | 1.43E-01 | H2AFZ |
| 912 | Adherens junctions interactions Homo sapiens R-HSA-418990 | 7.97E-01 | 9.44E-01 | 6.31E-01 | 1.43E-01 | CDH15 |

| | | | | | | |
|---|---|---|---|---|---|---|
| 913 | Inwardly rectifying K+ channels Homo sapiens R-HSA-1296065 | 7.97E-01 | 9.44E-01 | 6.31E-01 | 1.43E-01 | GNG4 |
| 914 | Intra-Golgi and retrograde Golgi-to-ER traffic Homo sapiens R-HSA-6811442 | 7.99E-01 | 9.45E-01 | 7.69E-01 | 1.72E-01 | RAB1B;GOSR2;MAN1C1;PLA2G4A;NAA38;COPE;VAMP3 |
| 915 | DNA Damage/Telomere Stress Induced Senescence Homo sapiens R-HSA-2559586 | 8.03E-01 | 9.47E-01 | 6.64E-01 | 1.46E-01 | H2AFZ;HIST1H1C |
| 916 | MyD88 dependent cascade initiated on endosome Homo sapiens R-HSA-975155 | 8.06E-01 | 9.47E-01 | 6.92E-01 | 1.49E-01 | UBB;TNIP2;ECSIT |
| 917 | Toll Like Receptor 7/8 (TLR7/8) Cascade Homo sapiens R-HSA-168181 | 8.06E-01 | 9.47E-01 | 6.92E-01 | 1.49E-01 | UBB;TNIP2;ECSIT |
| 918 | RNA Polymerase I Transcription Termination Homo sapiens R-HSA-73863 | 8.08E-01 | 9.47E-01 | 6.11E-01 | 1.31E-01 | POLR2E |
| 919 | Signaling by NOTCH2 Homo sapiens R-HSA-1980145 | 8.08E-01 | 9.47E-01 | 6.11E-01 | 1.31E-01 | UBB |
| 920 | SMAD2/SMAD3:SMAD4 heterotrimer regulates transcription Homo sapiens R-HSA-2173796 | 8.08E-01 | 9.47E-01 | 6.11E-01 | 1.31E-01 | UBB |
| 921 | TRAF6 mediated IRF7 activation Homo sapiens R-HSA-933541 | 8.08E-01 | 9.47E-01 | 6.11E-01 | 1.31E-01 | RNF135 |
| 922 | Signaling by Robo receptor Homo sapiens R-HSA-376176 | 8.08E-01 | 9.47E-01 | 6.11E-01 | 1.31E-01 | SRGAP3 |
| 923 | MAP kinase activation in TLR cascade Homo sapiens R-HSA-450294 | 8.10E-01 | 9.49E-01 | 6.52E-01 | 1.37E-01 | UBB;TNIP2 |
| 924 | Unfolded Protein Response (UPR) Homo sapiens R-HSA-381119 | 8.12E-01 | 9.50E-01 | 6.84E-01 | 1.42E-01 | GOSR2;WIPI1;ATP6V0D1 |
| 925 | CD28 co-stimulation Homo sapiens R-HSA-389356 | 8.17E-01 | 9.52E-01 | 5.91E-01 | 1.19E-01 | FYN |

| | | | | | | |
|---|---|---|---|---|---|---|
| 926 | Inactivation, recovery and regulation of the phototransduction cascade Homo sapiens R-HSA-2514859 | 8.17E-01 | 9.52E-01 | 5.91E-01 | 1.19E-01 | CALM1 |
| 927 | Smooth Muscle Contraction Homo sapiens R-HSA-445355 | 8.17E-01 | 9.52E-01 | 5.91E-01 | 1.19E-01 | CALM1 |
| 928 | Cell-cell junction organization Homo sapiens R-HSA-421270 | 8.17E-01 | 9.52E-01 | 6.41E-01 | 1.29E-01 | CDH15;CLDN1 |
| 929 | Metabolism of proteins Homo sapiens R-HSA-392499 | 8.19E-01 | 9.52E-01 | 8.80E-01 | 1.76E-01 | APCS;CALCA;ENPEP;SP100;SERPINA1;GOSR2;RAB1B;COX17;PROZ;TIMM13;ARL2;PLG;HSCB;TIMM10;NPL;FUOM;RPS4Y1;GRPEL1;UBB;GNG4;LDHD;MAN1C1;SLC17A5;CTSH;RPL38;CYC1;EIF5A;CTSA;FN3K;IGFBP5;IDH3G;H2AFZ;SLC34A1;PCGF2;CBX2;WIPI1;GNG11;MSRA;RPS28;NAT8;RPS27;RPS29;PAPPA;ACO2;ATP6V0D1;RPS21;TOMM5;COPE |
| 930 | HDR through Homologous Recombination (HR) or Single Strand Annealing (SSA) Homo sapiens R-HSA-5693567 | 8.19E-01 | 9.52E-01 | 7.00E-01 | 1.40E-01 | POLD3;UBB;RFC1;RPA3 |
| 931 | Toll Like Receptor 9 (TLR9) Cascade Homo sapiens R-HSA-168138 | 8.24E-01 | 9.57E-01 | 6.67E-01 | 1.29E-01 | UBB;TNIP2;ECSIT |
| 932 | Activated PKN1 stimulates transcription of AR (androgen receptor) regulated genes KLK2 and KLK3 Homo sapiens R-HSA-5625886 | 8.26E-01 | 9.57E-01 | 5.74E-01 | 1.09E-01 | H2AFZ |
| 933 | The phototransduction cascade Homo sapiens R-HSA-2514856 | 8.26E-01 | 9.57E-01 | 5.74E-01 | 1.09E-01 | CALM1 |
| 934 | Assembly of the primary cilium Homo sapiens R-HSA-5617833 | 8.34E-01 | 9.63E-01 | 7.35E-01 | 1.34E-01 | PRKAR2B;SSNA1;DYNLRB1;CEP63;TUBG1;RAB11FIP3;HDAC6 |

| | | | | | | |
|---|---|---|---|---|---|---|
| 935 | Generation of second messenger molecules Homo sapiens R-HSA-202433 | 8.35E-01 | 9.63E-01 | 5.57E-01 | 1.00E-01 | HLA-DPB1 |
| 936 | Interactions of Vpr with host cellular proteins Homo sapiens R-HSA-176033 | 8.35E-01 | 9.63E-01 | 5.57E-01 | 1.00E-01 | SLC25A5 |
| 937 | SIRT1 negatively regulates rRNA Expression Homo sapiens R-HSA-427359 | 8.35E-01 | 9.63E-01 | 5.57E-01 | 1.00E-01 | H2AFZ |
| 938 | RNA Polymerase II Pre-transcription Events Homo sapiens R-HSA-674695 | 8.38E-01 | 9.66E-01 | 6.10E-01 | 1.08E-01 | POLR2E;POLR2I |
| 939 | Oxidative Stress Induced Senescence Homo sapiens R-HSA-2559580 | 8.41E-01 | 9.68E-01 | 6.45E-01 | 1.11E-01 | UBB;H2AFZ;CBX2 |
| 940 | Disease Homo sapiens R-HSA-1643685 | 8.45E-01 | 9.68E-01 | 8.41E-01 | 1.41E-01 | KLB;AP2A1;RPS4Y1;FGF2;HDAC6;PSMD8;PSMD9;UBB;PSMB3;PSMD2;AP2S1;POLR2E;RPL38;FYN;POLR2I;AP1M1;STAT5A;MSRA;RPS28;RPS27;PSMC4;RPS29;FDX1;PCCB;FGF18;PSME1;NAPEPLD;AVP;SLC25A5;CALM1;RPS21 |
| 941 | MyD88:Mal cascade initiated on plasma membrane Homo sapiens R-HSA-166058 | 8.46E-01 | 9.68E-01 | 6.37E-01 | 1.06E-01 | UBB;TNIP2;ECSIT |
| 942 | Toll Like Receptor TLR1:TLR2 Cascade Homo sapiens R-HSA-168179 | 8.46E-01 | 9.68E-01 | 6.37E-01 | 1.06E-01 | UBB;TNIP2;ECSIT |
| 943 | Toll Like Receptor TLR6:TLR2 Cascade Homo sapiens R-HSA-168188 | 8.46E-01 | 9.68E-01 | 6.37E-01 | 1.06E-01 | UBB;TNIP2;ECSIT |
| 944 | Toll Like Receptor 2 (TLR2) Cascade Homo sapiens R-HSA-181438 | 8.46E-01 | 9.68E-01 | 6.37E-01 | 1.06E-01 | UBB;TNIP2;ECSIT |
| 945 | Generic Transcription Pathway Homo sapiens R-HSA-212436 | 8.47E-01 | 9.68E-01 | 8.48E-01 | 1.41E-01 | COX7B;RORA;COX7C;COX5A;NDRG1;GLS;UBB;HNF4A;PRDX1;SESN2;POLR2E;PMS2;COX11;COX14;POLR2I;COX8A;ESRRA;BNIP3L;SURF1;PRELID1;GPX2;USP2;NR1H4;NR1D1;ESR2;NR2F6;COX6B1;RGCC;BCL6;RPA3;DDIT4;RARA;LAMTOR2;LAMTOR1;LAMTOR5 |

| | | | | | | |
|---|---|---|---|---|---|---|
| 946 | Homology Directed Repair Homo sapiens R-HSA-5693538 | 8.49E-01 | 9.68E-01 | 6.63E-01 | 1.09E-01 | POLD3;UBB;RFC1;RPA3 |
| 947 | Cytosolic sensors of pathogen-associated DNA Homo sapiens R-HSA-1834949 | 8.50E-01 | 9.68E-01 | 5.91E-01 | 9.60E-02 | UBB;POLR2E |
| 948 | HDR through Single Strand Annealing (SSA) Homo sapiens R-HSA-5685938 | 8.51E-01 | 9.68E-01 | 5.26E-01 | 8.47E-02 | RPA3 |
| 949 | Activation of ATR in response to replication stress Homo sapiens R-HSA-176187 | 8.51E-01 | 9.68E-01 | 5.26E-01 | 8.47E-02 | RPA3 |
| 950 | Platelet Aggregation (Plug Formation) Homo sapiens R-HSA-76009 | 8.51E-01 | 9.68E-01 | 5.26E-01 | 8.47E-02 | PTPN1 |
| 951 | Interferon gamma signaling Homo sapiens R-HSA-877300 | 8.52E-01 | 9.68E-01 | 6.30E-01 | 1.01E-01 | PTPN1;SP100;HLA-DPB1 |
| 952 | Integrin cell surface interactions Homo sapiens R-HSA-216083 | 8.56E-01 | 9.69E-01 | 5.82E-01 | 9.05E-02 | VTN;KDR |
| 953 | Transcription of the HIV genome Homo sapiens R-HSA-167172 | 8.56E-01 | 9.69E-01 | 5.82E-01 | 9.05E-02 | POLR2E;POLR2I |
| 954 | Semaphorin interactions Homo sapiens R-HSA-373755 | 8.56E-01 | 9.69E-01 | 5.82E-01 | 9.05E-02 | FYN;PLXNA4 |
| 955 | Peptide ligand-binding receptors Homo sapiens R-HSA-375276 | 8.56E-01 | 9.69E-01 | 7.11E-01 | 1.10E-01 | PNOC;F2R;PPBP;AVP;NTS;SSTR2;PF4 |
| 956 | DNA Damage Recognition in GG-NER Homo sapiens R-HSA-5696394 | 8.59E-01 | 9.69E-01 | 5.11E-01 | 7.79E-02 | UBB |
| 957 | G1 Phase Homo sapiens R-HSA-69236 | 8.59E-01 | 9.69E-01 | 5.11E-01 | 7.79E-02 | UBB |
| 958 | Cyclin D associated events in G1 Homo sapiens R-HSA-69231 | 8.59E-01 | 9.69E-01 | 5.11E-01 | 7.79E-02 | UBB |
| 959 | Interferon alpha/beta signaling Homo sapiens R-HSA-909733 | 8.62E-01 | 9.70E-01 | 5.73E-01 | 8.53E-02 | PTPN1;IFITM1 |

| | | | | | | |
|---|---|---|---|---|---|---|
| 960 | Presynaptic phase of homologous DNA pairing and strand exchange Homo sapiens R-HSA-5693616 | 8.66E-01 | 9.70E-01 | 4.98E-01 | 7.17E-02 | RPA3 |
| 961 | Nuclear signaling by ERBB4 Homo sapiens R-HSA-1251985 | 8.66E-01 | 9.70E-01 | 4.98E-01 | 7.17E-02 | STAT5A |
| 962 | GABA B receptor activation Homo sapiens R-HSA-977444 | 8.66E-01 | 9.70E-01 | 4.98E-01 | 7.17E-02 | GNG4 |
| 963 | Activation of GABAB receptors Homo sapiens R-HSA-991365 | 8.66E-01 | 9.70E-01 | 4.98E-01 | 7.17E-02 | GNG4 |
| 964 | tRNA modification in the nucleus and cytosol Homo sapiens R-HSA-6782315 | 8.66E-01 | 9.70E-01 | 4.98E-01 | 7.17E-02 | TRMT6 |
| 965 | SUMO E3 ligases SUMOylate target proteins Homo sapiens R-HSA-3108232 | 8.66E-01 | 9.70E-01 | 6.10E-01 | 8.76E-02 | SP100;CBX2;PCGF2 |
| 966 | PRC2 methylates histones and DNA Homo sapiens R-HSA-212300 | 8.73E-01 | 9.76E-01 | 4.85E-01 | 6.61E-02 | H2AFZ |
| 967 | Elastic fibre formation Homo sapiens R-HSA-1566948 | 8.79E-01 | 9.77E-01 | 4.73E-01 | 6.10E-02 | VTN |
| 968 | Intraflagellar transport Homo sapiens R-HSA-5620924 | 8.79E-01 | 9.77E-01 | 4.73E-01 | 6.10E-02 | DYNLRB1 |
| 969 | Condensation of Prophase Chromosomes Homo sapiens R-HSA-2299718 | 8.79E-01 | 9.77E-01 | 4.73E-01 | 6.10E-02 | H2AFZ |
| 970 | Potassium Channels Homo sapiens R-HSA-1296071 | 8.79E-01 | 9.77E-01 | 5.91E-01 | 7.59E-02 | KCNH6;GNG4;KCNAB2 |
| 971 | Costimulation by the CD28 family Homo sapiens R-HSA-388841 | 8.82E-01 | 9.77E-01 | 5.40E-01 | 6.77E-02 | HLA-DPB1;FYN |

| | | | | | | |
|---|---|---|---|---|---|---|
| 972 | TRAF6 Mediated Induction of proinflammatory cytokines Homo sapiens R-HSA-168180 | 8.82E-01 | 9.77E-01 | 5.40E-01 | 6.77E-02 | UBB;TNIP2 |
| 973 | Homologous DNA Pairing and Strand Exchange Homo sapiens R-HSA-5693579 | 8.85E-01 | 9.77E-01 | 4.61E-01 | 5.63E-02 | RPA3 |
| 974 | Deactivation of the beta-catenin transactivating complex Homo sapiens R-HSA-3769402 | 8.85E-01 | 9.77E-01 | 4.61E-01 | 5.63E-02 | UBB |
| 975 | Netrin-1 signaling Homo sapiens R-HSA-373752 | 8.85E-01 | 9.77E-01 | 4.61E-01 | 5.63E-02 | FYN |
| 976 | Activation of gene expression by SREBF (SREBP) Homo sapiens R-HSA-2426168 | 8.85E-01 | 9.77E-01 | 4.61E-01 | 5.63E-02 | MVK |
| 977 | Positive epigenetic regulation of rRNA expression Homo sapiens R-HSA-5250913 | 8.87E-01 | 9.77E-01 | 5.33E-01 | 6.39E-02 | H2AFZ;POLR2E |
| 978 | NoRC negatively regulates rRNA expression Homo sapiens R-HSA-427413 | 8.87E-01 | 9.77E-01 | 5.33E-01 | 6.39E-02 | H2AFZ;POLR2E |
| 979 | Signaling by TGF-beta Receptor Complex Homo sapiens R-HSA-170834 | 8.87E-01 | 9.77E-01 | 5.33E-01 | 6.39E-02 | UBB;STRAP |
| 980 | SUMOylation Homo sapiens R-HSA-2990846 | 8.88E-01 | 9.77E-01 | 5.79E-01 | 6.90E-02 | SP100;CBX2;PCGF2 |
| 981 | Gene Silencing by RNA Homo sapiens R-HSA-211000 | 8.88E-01 | 9.77E-01 | 5.79E-01 | 6.90E-02 | H2AFZ;POLR2E;POLR2I |
| 982 | Protein folding Homo sapiens R-HSA-391251 | 8.88E-01 | 9.77E-01 | 5.79E-01 | 6.90E-02 | GNG4;ARL2;GNG11 |
| 983 | ERCC6 (CSB) and EHMT2 (G9a) positively regulate rRNA expression Homo sapiens R-HSA-427389 | 8.91E-01 | 9.79E-01 | 4.50E-01 | 5.21E-02 | H2AFZ |

| | | | | | | |
|---|---|---|---|---|---|---|
| 984 | Sphingolipid metabolism Homo sapiens R-HSA-428157 | 8.91E-01 | 9.79E-01 | 5.25E-01 | 6.03E-02 | CTSA;CERK |
| 985 | Cytokine Signaling in Immune system Homo sapiens R-HSA-1280215 | 8.94E-01 | 9.81E-01 | 7.91E-01 | 8.85E-02 | KLB;CNTF;IFITM1;SP100;RASGRF1;PEBP1;PHB;FGF2;PSMD8;PSMD9;UBB;PSMB3;PSMD2;FYN;IL6R;HAVCR2;STAT5A;PTPN1;PSMC4;TNIP2;FGF18;PSME1;HLA-DPB1;LAMTOR2;CALM1 |
| 986 | G alpha (12/13) signalling events Homo sapiens R-HSA-416482 | 8.96E-01 | 9.82E-01 | 5.18E-01 | 5.70E-02 | GNG4;GNG11 |
| 987 | Transcriptional activity of SMAD2/SMAD3:SMAD4 heterotrimer Homo sapiens R-HSA-2173793 | 8.96E-01 | 9.82E-01 | 4.40E-01 | 4.82E-02 | UBB |
| 988 | ER to Golgi Anterograde Transport Homo sapiens R-HSA-199977 | 9.00E-01 | 9.83E-01 | 5.95E-01 | 6.29E-02 | SERPINA1;RAB1B;GOSR2;COPE |
| 989 | Negative epigenetic regulation of rRNA expression Homo sapiens R-HSA-5250941 | 9.00E-01 | 9.83E-01 | 5.11E-01 | 5.38E-02 | H2AFZ;POLR2E |
| 990 | RNA Polymerase I Promoter Clearance Homo sapiens R-HSA-73854 | 9.00E-01 | 9.83E-01 | 5.11E-01 | 5.38E-02 | H2AFZ;POLR2E |
| 991 | RMTs methylate histone arginines Homo sapiens R-HSA-3214858 | 9.02E-01 | 9.83E-01 | 4.30E-01 | 4.46E-02 | H2AFZ |
| 992 | Asparagine N-linked glycosylation Homo sapiens R-HSA-446203 | 9.07E-01 | 9.86E-01 | 6.79E-01 | 6.63E-02 | CTSA;SERPINA1;RAB1B;GOSR2;SLC17A5;MAN1C1;NPL;FUOM;COPE |
| 993 | Adaptive Immune System Homo sapiens R-HSA-1280218 | 9.07E-01 | 9.86E-01 | 7.97E-01 | 7.75E-02 | KLB;ANAPC13;IFITM1;AP2A1;FGF2;ANAPC11;PSMD8;PSMD9;ZNRF1;UBB;PSMB3;PSMD2;CDC26;CTSK;AP2S1;CTSH;FYN;IER3;AP1M1;CTSA;TRIM63;PRKCB;ZBTB16;FBXO17;CYBA;PSMC4;FGF18;ASB9;PSME1;HLA-DPB1;CALM1 |
| 994 | DNA Repair Homo sapiens R-HSA-73894 | 9.11E-01 | 9.86E-01 | 6.86E-01 | 6.39E-02 | POLD3;XAB2;PARG;UBB;RFC1;OGG1;RPA3;POLR2E;PMS2;POLR2I |
| 995 | RNA Polymerase I Transcription Initiation Homo sapiens R-HSA-73762 | 9.11E-01 | 9.86E-01 | 4.11E-01 | 3.82E-02 | POLR2E |

| | | | | | | |
|---|---|---|---|---|---|---|
| 996 | RNA Polymerase I Transcription Homo sapiens R-HSA-73864 | 9.12E-01 | 9.86E-01 | 4.91E-01 | 4.54E-02 | H2AFZ;POLR2E |
| 997 | RIG-I/MDA5 mediated induction of IFN-alpha/beta pathways Homo sapiens R-HSA-168928 | 9.12E-01 | 9.86E-01 | 4.91E-01 | 4.54E-02 | RNF135;UBB |
| 998 | Transcriptional regulation of white adipocyte differentiation Homo sapiens R-HSA-381340 | 9.12E-01 | 9.86E-01 | 4.91E-01 | 4.54E-02 | MED11;PCK1 |
| 999 | Transport to the Golgi and subsequent modification Homo sapiens R-HSA-948021 | 9.14E-01 | 9.86E-01 | 6.01E-01 | 5.42E-02 | SERPINA1;RAB1B;GOSR2;MAN1C1;COPE |
| 1000 | Role of LAT2/NTAL/LAB on calcium mobilization Homo sapiens R-HSA-2730905 | 9.14E-01 | 9.86E-01 | 6.01E-01 | 5.42E-02 | KLB;FGF18;FYN;FGF2;IER3 |
| 1001 | Complement cascade Homo sapiens R-HSA-166658 | 9.15E-01 | 9.86E-01 | 4.85E-01 | 4.29E-02 | VTN;C2 |
| 1002 | TP53 Regulates Transcription of Cell Cycle Genes Homo sapiens R-HSA-6791312 | 9.16E-01 | 9.86E-01 | 4.02E-01 | 3.55E-02 | RGCC |
| 1003 | Golgi-to-ER retrograde transport Homo sapiens R-HSA-8856688 | 9.19E-01 | 9.86E-01 | 5.30E-01 | 4.49E-02 | RAB1B;PLA2G4A;COPE |
| 1004 | COPI-dependent Golgi-to-ER retrograde traffic Homo sapiens R-HSA-6811434 | 9.19E-01 | 9.86E-01 | 4.78E-01 | 4.06E-02 | RAB1B;COPE |
| 1005 | Processing of DNA double-strand break ends Homo sapiens R-HSA-5693607 | 9.19E-01 | 9.86E-01 | 4.78E-01 | 4.06E-02 | UBB;RPA3 |
| 1006 | G alpha (i) signalling events Homo sapiens R-HSA-418594 | 9.19E-01 | 9.86E-01 | 6.50E-01 | 5.51E-02 | RGS18;RGS5;GNG4;PNOC;PPBP;SSTR2;GNG11;PF4 |
| 1007 | FCGR activation Homo sapiens R-HSA-2029481 | 9.20E-01 | 9.86E-01 | 3.94E-01 | 3.29E-02 | FYN |

| | | | | | | |
|---|---|---|---|---|---|---|
| 1008 | Chondroitin sulfate/dermatan sulfate metabolism Homo sapiens R-HSA-1793185 | 9.24E-01 | 9.86E-01 | 3.86E-01 | 3.05E-02 | B3GAT1 |
| 1009 | Deadenylation-dependent mRNA decay Homo sapiens R-HSA-429914 | 9.24E-01 | 9.86E-01 | 3.86E-01 | 3.05E-02 | LSM4 |
| 1010 | Metabolism of non-coding RNA Homo sapiens R-HSA-194441 | 9.24E-01 | 9.86E-01 | 3.86E-01 | 3.05E-02 | SNRPG |
| 1011 | snRNP Assembly Homo sapiens R-HSA-191859 | 9.24E-01 | 9.86E-01 | 3.86E-01 | 3.05E-02 | SNRPG |
| 1012 | Activated TLR4 signalling Homo sapiens R-HSA-166054 | 9.24E-01 | 9.86E-01 | 5.20E-01 | 4.09E-02 | UBB;TNIP2;ECSIT |
| 1013 | Toll-Like Receptors Cascades Homo sapiens R-HSA-168898 | 9.25E-01 | 9.86E-01 | 5.55E-01 | 4.31E-02 | UBB;TNIP2;CTSK;ECSIT |
| 1014 | Processing of Capped Intron-Containing Pre-mRNA Homo sapiens R-HSA-72203 | 9.26E-01 | 9.86E-01 | 6.05E-01 | 4.66E-02 | SNRPG;POLR2E;DNAJC8;TXNL4A;POLR2I;LSM4 |
| 1015 | Cargo trafficking to the periciliary membrane Homo sapiens R-HSA-5620920 | 9.28E-01 | 9.86E-01 | 3.78E-01 | 2.84E-02 | RAB11FIP3 |
| 1016 | Apoptotic execution phase Homo sapiens R-HSA-75153 | 9.28E-01 | 9.86E-01 | 3.78E-01 | 2.84E-02 | HIST1H1C |
| 1017 | Deposition of new CENPA-containing nucleosomes at the centromere Homo sapiens R-HSA-606279 | 9.31E-01 | 9.86E-01 | 3.71E-01 | 2.64E-02 | H2AFZ |
| 1018 | Nucleosome assembly Homo sapiens R-HSA-774815 | 9.31E-01 | 9.86E-01 | 3.71E-01 | 2.64E-02 | H2AFZ |
| 1019 | Epigenetic regulation of gene expression Homo sapiens R-HSA-212165 | 9.32E-01 | 9.86E-01 | 5.06E-01 | 3.55E-02 | DNMT3L;H2AFZ;POLR2E |
| 1020 | Cell junction organization Homo sapiens R-HSA-446728 | 9.34E-01 | 9.86E-01 | 4.50E-01 | 3.07E-02 | CDH15;CLDN1 |

| | | | | | | |
|---|---|---|---|---|---|---|
| 1021 | RNA Polymerase I, RNA Polymerase III, and Mitochondrial Transcription Homo sapiens R-HSA-504046 | 9.35E-01 | 9.86E-01 | 5.01E-01 | 3.38E-02 | SNAPC1;H2AFZ;POLR2E |
| 1022 | GPVI-mediated activation cascade Homo sapiens R-HSA-114604 | 9.35E-01 | 9.86E-01 | 3.64E-01 | 2.45E-02 | FYN |
| 1023 | DNA Double-Strand Break Repair Homo sapiens R-HSA-5693532 | 9.37E-01 | 9.86E-01 | 5.35E-01 | 3.50E-02 | POLD3;UBB;RFC1;RPA3 |
| 1024 | ECM proteoglycans Homo sapiens R-HSA-3000178 | 9.41E-01 | 9.86E-01 | 3.50E-01 | 2.12E-02 | VTN |
| 1025 | GABA receptor activation Homo sapiens R-HSA-977443 | 9.41E-01 | 9.86E-01 | 3.50E-01 | 2.12E-02 | GNG4 |
| 1026 | Regulation of cholesterol biosynthesis by SREBP (SREBF) Homo sapiens R-HSA-1655829 | 9.41E-01 | 9.86E-01 | 3.50E-01 | 2.12E-02 | MVK |
| 1027 | Regulation of TP53 Activity through Phosphorylation Homo sapiens R-HSA-6804756 | 9.42E-01 | 9.86E-01 | 4.34E-01 | 2.60E-02 | UBB;RPA3 |
| 1028 | Fcgamma receptor (FCGR) dependent phagocytosis Homo sapiens R-HSA-2029480 | 9.44E-01 | 9.86E-01 | 4.84E-01 | 2.80E-02 | ARPC1B;FYN;BAIAP2 |
| 1029 | tRNA processing in the nucleus Homo sapiens R-HSA-6784531 | 9.47E-01 | 9.86E-01 | 3.38E-01 | 1.84E-02 | RTCB |
| 1030 | Signaling by NOTCH Homo sapiens R-HSA-157118 | 9.48E-01 | 9.86E-01 | 4.76E-01 | 2.55E-02 | UBB;HIF1A;HDAC6 |
| 1031 | Toll Like Receptor 4 (TLR4) Cascade Homo sapiens R-HSA-166016 | 9.48E-01 | 9.86E-01 | 4.76E-01 | 2.55E-02 | UBB;TNIP2;ECSIT |
| 1032 | Rho GTPase cycle Homo sapiens R-HSA-194840 | 9.48E-01 | 9.86E-01 | 4.76E-01 | 2.55E-02 | ARHGAP18;SRGAP3;RHOBTB1 |

| | | | | | | |
|---|---|---|---|---|---|---|
| 1033 | Formation of the beta-catenin:TCF transactivating complex Homo sapiens R-HSA-201722 | 9.50E-01 | 9.86E-01 | 3.32E-01 | 1.71E-02 | H2AFZ |
| 1034 | Recruitment and ATM-mediated phosphorylation of repair and signaling proteins at DNA double strand breaks Homo sapiens R-HSA-5693565 | 9.50E-01 | 9.86E-01 | 3.32E-01 | 1.71E-02 | UBB |
| 1035 | Initial triggering of complement Homo sapiens R-HSA-166663 | 9.50E-01 | 9.86E-01 | 3.32E-01 | 1.71E-02 | C2 |
| 1036 | RNA Polymerase II Transcription Homo sapiens R-HSA-73857 | 9.52E-01 | 9.86E-01 | 4.68E-01 | 2.32E-02 | SNRPG;POLR2E;POLR2I |
| 1037 | DNA Double Strand Break Response Homo sapiens R-HSA-5693606 | 9.52E-01 | 9.86E-01 | 3.26E-01 | 1.60E-02 | UBB |
| 1038 | Cleavage of Growing Transcript in the Termination Region Homo sapiens R-HSA-109688 | 9.55E-01 | 9.86E-01 | 3.20E-01 | 1.49E-02 | SNRPG |
| 1039 | RNA Polymerase II Transcription Termination Homo sapiens R-HSA-73856 | 9.55E-01 | 9.86E-01 | 3.20E-01 | 1.49E-02 | SNRPG |
| 1040 | Post-Elongation Processing of the Transcript Homo sapiens R-HSA-76044 | 9.55E-01 | 9.86E-01 | 3.20E-01 | 1.49E-02 | SNRPG |
| 1041 | RHO GTPases activate PKNs Homo sapiens R-HSA-5625740 | 9.55E-01 | 9.86E-01 | 3.20E-01 | 1.49E-02 | H2AFZ |
| 1042 | PI Metabolism Homo sapiens R-HSA-1483255 | 9.55E-01 | 9.86E-01 | 3.20E-01 | 1.49E-02 | GDPD3 |
| 1043 | Chaperonin-mediated protein folding Homo sapiens R-HSA-390466 | 9.55E-01 | 9.86E-01 | 4.06E-01 | 1.87E-02 | GNG4;GNG11 |
| 1044 | Cell death signalling via NRAGE, NRIF and NADE Homo sapiens R-HSA-204998 | 9.57E-01 | 9.86E-01 | 3.15E-01 | 1.39E-02 | UBB |

| | | | | | | |
|---|---|---|---|---|---|---|
| 1045 | L1CAM interactions Homo sapiens R-HSA-373760 | 9.57E-01 | 9.86E-01 | 4.02E-01 | 1.77E-02 | AP2S1;AP2A1 |
| 1046 | Late Phase of HIV Life Cycle Homo sapiens R-HSA-162599 | 9.58E-01 | 9.86E-01 | 4.53E-01 | 1.92E-02 | UBB;POLR2E;POLR2I |
| 1047 | MyD88-independent TLR3/TLR4 cascade Homo sapiens R-HSA-166166 | 9.59E-01 | 9.86E-01 | 3.98E-01 | 1.68E-02 | UBB;TNIP2 |
| 1048 | Toll Like Receptor 3 (TLR3) Cascade Homo sapiens R-HSA-168164 | 9.59E-01 | 9.86E-01 | 3.98E-01 | 1.68E-02 | UBB;TNIP2 |
| 1049 | TRIF-mediated TLR3/TLR4 signaling Homo sapiens R-HSA-937061 | 9.59E-01 | 9.86E-01 | 3.98E-01 | 1.68E-02 | UBB;TNIP2 |
| 1050 | Regulation of actin dynamics for phagocytic cup formation Homo sapiens R-HSA-2029482 | 9.59E-01 | 9.86E-01 | 3.98E-01 | 1.68E-02 | ARPC1B;BAIAP2 |
| 1051 | FCERI mediated Ca+2 mobilization Homo sapiens R-HSA-2871809 | 9.59E-01 | 9.86E-01 | 3.10E-01 | 1.30E-02 | CALM1 |
| 1052 | Cell-Cell communication Homo sapiens R-HSA-1500931 | 9.63E-01 | 9.90E-01 | 4.42E-01 | 1.67E-02 | FYN;CDH15;CLDN1 |
| 1053 | tRNA processing Homo sapiens R-HSA-72306 | 9.68E-01 | 9.94E-01 | 3.74E-01 | 1.21E-02 | RTCB;TRMT6 |
| 1054 | Interferon Signaling Homo sapiens R-HSA-913531 | 9.71E-01 | 9.96E-01 | 4.93E-01 | 1.45E-02 | PTPN1;IFITM1;SP100;UBB;HLA-DPB1 |
| 1055 | HATs acetylate histones Homo sapiens R-HSA-3214847 | 9.73E-01 | 9.97E-01 | 3.59E-01 | 9.75E-03 | TADA3;ELP5 |
| 1056 | HIV Life Cycle Homo sapiens R-HSA-162587 | 9.75E-01 | 9.97E-01 | 4.10E-01 | 1.04E-02 | UBB;POLR2E;POLR2I |
| 1057 | ISG15 antiviral mechanism Homo sapiens R-HSA-1169408 | 9.76E-01 | 9.97E-01 | 2.66E-01 | 6.59E-03 | UBB |

| | | | | | | |
|---|---|---|---|---|---|---|
| 1058 | Antiviral mechanism by IFN-stimulated genes Homo sapiens R-HSA-1169410 | 9.76E-01 | 9.97E-01 | 2.66E-01 | 6.59E-03 | UBB |
| 1059 | GPCR ligand binding Homo sapiens R-HSA-500792 | 9.81E-01 | 1.00E+00 | 6.07E-01 | 1.19E-02 | CALCA;GIPR;PTGER1;WNT7B;F2R;ADM;PPBP;NTS;SSTR2;GNG11;GNG4;PNOC;AVP;PF4 |
| 1060 | G2/M DNA damage checkpoint Homo sapiens R-HSA-69473 | 9.82E-01 | 1.00E+00 | 2.45E-01 | 4.44E-03 | RPA3 |
| 1061 | Regulation of TP53 Activity Homo sapiens R-HSA-5633007 | 9.83E-01 | 1.00E+00 | 3.82E-01 | 6.49E-03 | UBB;RPA3;USP2 |
| 1062 | Peptide hormone metabolism Homo sapiens R-HSA-2980736 | 9.85E-01 | 1.00E+00 | 2.36E-01 | 3.65E-03 | ENPEP |
| 1063 | Glycosaminoglycan metabolism Homo sapiens R-HSA-1630316 | 9.86E-01 | 1.00E+00 | 3.17E-01 | 4.60E-03 | NAGLU;B3GAT1 |
| 1064 | p75 NTR receptor-mediated signalling Homo sapiens R-HSA-193704 | 9.86E-01 | 1.00E+00 | 2.30E-01 | 3.21E-03 | UBB |
| 1065 | Innate Immune System Homo sapiens R-HSA-168249 | 9.87E-01 | 1.00E+00 | 6.72E-01 | 8.66E-03 | KLB;ARPC1B;RASGRF1;PEBP1;PHB;ECSIT;FGF2;C2;PSMD8;RNF135;PSMD9;LGALS3;VTN;UBB;PRKAR2B;PSMB3;PSMD2;CTSK;POLR2E;FYN;IER3;BAIAP2;PSMC4;TNIP2;FGF18;PSME1;LAMTOR2;CALM1 |
| 1066 | RHO GTPase Effectors Homo sapiens R-HSA-195258 | 9.90E-01 | 1.00E+00 | 4.53E-01 | 4.76E-03 | RPS27;H2AFZ;ARPC1B;CYBA;CALM1;BAIAP2 |
| 1067 | Gene Expression Homo sapiens R-HSA-74160 | 9.91E-01 | 1.00E+00 | 7.45E-01 | 6.94E-03 | COX7B;RTCB;RORA;COX7C;GLS;PSMD8;PSMD9;DNMT3L;SNAPC1;PSMD2;HNF4A;SESN2;RPL38;TXNL4A;COX8A;BNIP3L;GPX2;H2AFZ;USP2;SARS;COX6B1;SARS2;DDIT4;PSME1;RARA;SNRPG;DNAJC8;RPS27L;RPS4Y1;COX5A;NDRG1;RRP7A;UBB;PSMB3;PRDX1;POLR2E;PMS2;COX11;COX14;POLR2I;ESRRA;SURF1;PRELID1;WDR18;NR1H4;NR1D1;LSM4;ESR2;NR2F6;RPS28;RGCC;RPS27;BCL6;PSMC4;RPS29;RPA3;TRMT6;NABP2;LAMTOR2;LAMTOR1;RPS21;NOP10;LAMTOR5 |
| 1068 | Class A/1 (Rhodopsin-like receptors) Homo sapiens R-HSA-373076 | 9.93E-01 | 1.00E+00 | 4.77E-01 | 3.51E-03 | PTGER1;PNOC;F2R;PPBP;AVP;NTS;SSTR2;PF4 |

| | | | | | | |
|---|---|---|---|---|---|---|
| 1069 | Resolution of Sister Chromatid Cohesion Homo sapiens R-HSA-2500257 | 9.94E-01 | 1.00E+00 | 1.92E-01 | 1.17E-03 | RPS27 |
| 1070 | Factors involved in megakaryocyte development and platelet production Homo sapiens R-HSA-983231 | 9.94E-01 | 1.00E+00 | 2.71E-01 | 1.59E-03 | PRKAR2B;MAFG |
| 1071 | Post-translational protein modification Homo sapiens R-HSA-597592 | 9.94E-01 | 1.00E+00 | 5.55E-01 | 3.12E-03 | FN3K;EIF5A;CTSA;SP100;SERPINA1;PCGF2;RAB1B;CBX2;GOSR2;PROZ;NPL;FUOM;SLC17A5;MAN1C1;COPE |
| 1072 | Signaling by Rho GTPases Homo sapiens R-HSA-194315 | 9.96E-01 | 1.00E+00 | 4.71E-01 | 2.10E-03 | RPS27;H2AFZ;ARPC1B;ARHGAP18;CYBA;SRGAP3;CALM1;BAIAP2;RHOBTB1 |
| 1073 | Mitotic Prometaphase Homo sapiens R-HSA-68877 | 9.96E-01 | 1.00E+00 | 1.78E-01 | 7.14E-04 | RPS27 |
| 1074 | RHO GTPases Activate Formins Homo sapiens R-HSA-5663220 | 9.97E-01 | 1.00E+00 | 1.67E-01 | 4.66E-04 | RPS27 |
| 1075 | Immune System Homo sapiens R-HSA-168256 | 9.97E-01 | 1.00E+00 | 6.94E-01 | 1.85E-03 | KLB;CNTF;IFITM1;ARPC1B;RASGRF1;PEBP1;AP2A1;ECSIT;FGF2;PSMD8;PSMD9;LGALS3;PSMD2;CDC26;CTSK;CTSH;ATP6V1E1;IL6R;IER3;HAVCR2;ATP6V1G1;TRIM63;PRKCB;FBXO17;CYBA;PSME1;ATP6V0D1;ANAPC13;SP100;PHB;C2;ANAPC11;RNF135;ZNRF1;VTN;UBB;PRKAR2B;PSMB3;AP2S1;POLR2E;FYN;ATP6V0E2;AP1M1;ATP6V1F;CTSA;STAT5A;PTPN1;ZBTB16;BAIAP2;PSMC4;TNIP2;FGF18;ASB9;HLA-DPB1;LAMTOR2;CALM1 |

| | | | | | | |
|---|---|---|---|---|---|---|
| 1076 | Signal Transduction Homo sapiens R-HSA-162582 | 9.98E-01 | 1.00E+00 | 7.34E-01 | 1.13E-03 | KLB;CAB39L;ARPC1B;RASGRF1;CLTB;PEBP1;AP2A1;ADM;SHB;CHRDL1;PCSK5;FGF2;PSMD8;PSMD9;RGS5;RGS3;PSMD2;BOC;KDR;ATP6V1E1;IL6R;PDK2;IER3;ATP6V1G1;PRKCH;PRKCB;H2AFZ;USP2;F2R;PLA2G4A;CYBA;EPS15L1;SSTR2;ALDH1A3;PLCB4;CDC42EP3;PSME1;LINC01139;RARA;STRAP;AVP;ATP6V0D1;MGLL;RGS18;GRB7;CALCA;GIPR;PTGER1;ARHGAP18;PLG;PHB;PDHB;NTS;HIF1A;RHOBTB1;HDAC6;APOM;UBB;PRKAR2B;PSMB3;GNG4;RDH10;AP2S1;POLR2E;FYN;APOE;SRGAP3;POLR2I;ATP6V0E2;ATP6V1F;STAT5A;PTPN1;WNT7B;AKR1C1;ABCA4;VEGFB;APOC3;FUZ;PPBP;BAIAP2;GNG11;VEGFA;MOB1A;RPS27;PSMC4;APOC2;FGF18;PNOC;CCDC88C;NAPEPLD;LAMTOR2;LAMTOR1;CALM1;LAMTOR5;PF4 |
| 1077 | Chromatin organization Homo sapiens R-HSA-4839726 | 9.99E-01 | 1.00E+00 | 2.53E-01 | 1.81E-04 | H2AFZ;TADA3;ELP5 |
| 1078 | Chromatin modifying enzymes Homo sapiens R-HSA-3247509 | 9.99E-01 | 1.00E+00 | 2.53E-01 | 1.81E-04 | H2AFZ;TADA3;ELP5 |
| 1079 | Immunoregulatory interactions between a Lymphoid and a non-Lymphoid cell Homo sapiens R-HSA-198933 | 1.00E+00 | 1.00E+00 | 1.21E-01 | 3.66E-05 | IFITM1 |
| 1080 | Signaling by GPCR Homo sapiens R-HSA-372790 | 1.00E+00 | 1.00E+00 | 5.72E-01 | 6.36E-05 | KLB;RGS18;CALCA;GIPR;PTGER1;RASGRF1;PEBP1;ADM;PHB;NTS;FGF2;PSMD8;PSMD9;RGS5;RGS3;UBB;PRKAR2B;PSMB3;GNG4;PSMD2;FYN;PRKCH;PRKCB;WNT7B;F2R;PLA2G4A;PPBP;SSTR2;GNG11;PLCB4;PSMC4;FGF18;PNOC;PSME1;LAMTOR2;AVP;CALM1;MGLL;PF4 |
| 1081 | GPCR downstream signaling Homo sapiens R-HSA-388396 | 1.00E+00 | 1.00E+00 | 3.81E-01 | 2.19E-06 | RGS18;CALCA;PRKCH;GIPR;PRKCB;PTGER1;F2R;ADM;PPBP;SSTR2;NTS;GNG11;RGS5;RGS3;PLCB4;GNG4;PNOC;AVP;MGLL;PF4 |

**Pathways enriched by genes enhanced with SGLT2 inhibitor in proximal tubular cells**

| | Term | P value | Adjusted P value | Odds Ratio | Combined Score | Genes |
|---|---|---|---|---|---|---|
| 1 | Response to metal ions Homo sapiens R-HSA-5660526 | 1.82E-06 | 4.75E-04 | 6.58E+01 | 8.70E+02 | MT2A;MT1G;MT1X;MT1H |
| 2 | Metallothioneins bind metals Homo sapiens R-HSA-5661231 | 1.82E-06 | 4.75E-04 | 6.58E+01 | 8.70E+02 | MT2A;MT1G;MT1X;MT1H |
| 3 | Defective B3GALTL causes Peters-plus syndrome (PpS) Homo sapiens R-HSA-5083635 | 3.05E-04 | 4.41E-02 | 1.39E+01 | 1.13E+02 | ADAMTS16;ADAMTS15;SPON1;THBS1 |
| 4 | O-glycosylation of TSR domain-containing proteins Homo sapiens R-HSA-5173214 | 3.39E-04 | 4.41E-02 | 1.35E+01 | 1.08E+02 | ADAMTS16;ADAMTS15;SPON1;THBS1 |
| 5 | O-linked glycosylation Homo sapiens R-HSA-5173105 | 4.29E-04 | 4.47E-02 | 6.69E+00 | 5.19E+01 | ADAMTS16;ADAMTS15;SPON1;GCNT4;THBS1;GALNT10 |
| 6 | Diseases of glycosylation Homo sapiens R-HSA-3781865 | 1.08E-03 | 9.41E-02 | 6.95E+00 | 4.75E+01 | ADAMTS16;ADAMTS15;SPON1;BGN;THBS1 |
| 7 | Glyoxylate metabolism and glycine degradation Homo sapiens R-HSA-389661 | 1.34E-03 | 9.94E-02 | 1.56E+01 | 1.03E+02 | GLDC;AGXT;GNMT |
| 8 | Disease Homo sapiens R-HSA-1643685 | 1.92E-03 | 1.25E-01 | 2.51E+00 | 1.57E+01 | RNF43;SPON1;JAG1;BGN;NRG1;PIK3CB;GTF2H2;THBS1;ADAMTS16;POMC;ADAMTS15;RBP4;FGF9;HEY1;PSME3 |
| 9 | Diseases associated with O-glycosylation of proteins Homo sapiens R-HSA-3906995 | 2.17E-03 | 1.26E-01 | 7.93E+00 | 4.86E+01 | ADAMTS16;ADAMTS15;SPON1;THBS1 |
| 10 | Activation of PPARGC1A (PGC-1alpha) by phosphorylation Homo sapiens R-HSA-2151209 | 3.31E-03 | 1.72E-01 | 2.85E+01 | 1.63E+02 | PRKAA2;PPARGC1A |
| 11 | Diseases of signal transduction Homo sapiens R-HSA-5663202 | 4.07E-03 | 1.93E-01 | 3.32E+00 | 1.83E+01 | RNF43;RBP4;JAG1;FGF9;HEY1;PSME3;NRG1;PIK3CB |
| 12 | VEGFA-VEGFR2 Pathway Homo sapiens R-HSA-4420097 | 7.55E-03 | 3.28E-01 | 2.98E+00 | 1.46E+01 | FGF9;AXL;PSME3;CTNNA1;NEFL;NRG1;PIK3CB;RASGRP1 |
| 13 | Signaling by VEGF Homo sapiens R-HSA-194138 | 8.70E-03 | 3.43E-01 | 2.90E+00 | 1.38E+01 | FGF9;AXL;PSME3;CTNNA1;NEFL;NRG1;PIK3CB;RASGRP1 |
| 14 | Metabolism of fat-soluble vitamins Homo sapiens R-HSA-6806667 | 1.02E-02 | 3.43E-01 | 7.14E+00 | 3.27E+01 | RBP4;LPL;GC |
| 15 | Fatty acid, triacylglycerol, and ketone body metabolism Homo sapiens R-HSA-535734 | 1.26E-02 | 3.43E-01 | 3.28E+00 | 1.44E+01 | PRKAA2;ACADL;SCD5;LPIN1;PPARGC1A;MED13L |
| 16 | IRS-mediated signalling Homo sapiens R-HSA-112399 | 1.31E-02 | 3.43E-01 | 2.92E+00 | 1.27E+01 | PRKAA2;FGF9;PSME3;NEFL;NRG1;PIK3CB;RASGRP1 |
| 17 | Insulin receptor signalling cascade Homo sapiens R-HSA-74751 | 1.38E-02 | 3.43E-01 | 2.89E+00 | 1.24E+01 | PRKAA2;FGF9;PSME3;NEFL;NRG1;PIK3CB;RASGRP1 |
| 18 | IGF1R signaling cascade Homo sapiens R-HSA-2428924 | 1.40E-02 | 3.43E-01 | 2.88E+00 | 1.23E+01 | PRKAA2;FGF9;PSME3;NEFL;NRG1;PIK3CB;RASGRP1 |

| | | | | | | |
|---|---|---|---|---|---|---|
| 19 | Signaling by Type 1 Insulin-like Growth Factor 1 Receptor (IGF1R) Homo sapiens R-HSA-2404192 | 1.40E-02 | 3.43E-01 | 2.88E+00 | 1.23E+01 | PRKAA2;FGF9;PSME3;NEFL;NRG1;PIK3CB;RASGRP1 |
| 20 | IRS-related events triggered by IGF1R Homo sapiens R-HSA-2428928 | 1.40E-02 | 3.43E-01 | 2.88E+00 | 1.23E+01 | PRKAA2;FGF9;PSME3;NEFL;NRG1;PIK3CB;RASGRP1 |
| 21 | Regulation of FZD by ubiquitination Homo sapiens R-HSA-4641263 | 1.45E-02 | 3.43E-01 | 1.20E+01 | 5.07E+01 | RNF43;ZNRF3 |
| 22 | Constitutive Signaling by Aberrant PI3K in Cancer Homo sapiens R-HSA-2219530 | 1.66E-02 | 3.43E-01 | 5.91E+00 | 2.42E+01 | FGF9;NRG1;PIK3CB |
| 23 | Signaling by Insulin receptor Homo sapiens R-HSA-74752 | 2.04E-02 | 3.43E-01 | 2.66E+00 | 1.03E+01 | PRKAA2;FGF9;PSME3;NEFL;NRG1;PIK3CB;RASGRP1 |
| 24 | Interleukin receptor SHC signaling Homo sapiens R-HSA-912526 | 2.15E-02 | 3.43E-01 | 2.89E+00 | 1.11E+01 | FGF9;PSME3;NEFL;NRG1;PIK3CB;RASGRP1 |
| 25 | RORA activates gene expression Homo sapiens R-HSA-1368082 | 2.34E-02 | 3.43E-01 | 9.10E+00 | 3.42E+01 | NRIP1;PPARGC1A |
| 26 | Interleukin-2 signaling Homo sapiens R-HSA-451927 | 2.43E-02 | 3.43E-01 | 2.81E+00 | 1.04E+01 | FGF9;PSME3;NEFL;NRG1;PIK3CB;RASGRP1 |
| 27 | Downstream signaling events of B Cell Receptor (BCR) Homo sapiens R-HSA-1168372 | 2.76E-02 | 3.43E-01 | 3.07E+00 | 1.10E+01 | FGF9;PSME3;NRG1;PIK3CB;RASGRP1 |
| 28 | Interleukin-3, 5 and GM-CSF signaling Homo sapiens R-HSA-512988 | 2.82E-02 | 3.43E-01 | 2.71E+00 | 9.66E+00 | FGF9;PSME3;NEFL;NRG1;PIK3CB;RASGRP1 |
| 29 | Molecules associated with elastic fibres Homo sapiens R-HSA-2129379 | 2.85E-02 | 3.43E-01 | 8.13E+00 | 2.89E+01 | ITGB6;BMP7 |
| 30 | Cell-Cell communication Homo sapiens R-HSA-1500931 | 2.87E-02 | 3.43E-01 | 3.61E+00 | 1.28E+01 | CTNNA1;SIRPA;PIK3CB;LIMS2 |
| 31 | Metabolism of lipids and lipoproteins Homo sapiens R-HSA-556833 | 3.17E-02 | 3.43E-01 | 1.97E+00 | 6.81E+00 | POMC;PRKAA2;ACADL;SCD5;LPL;PIK3CB;GPCPD1;LPIN1;SLC27A2;PPARGC1A;MED13L |
| 32 | PI3K Cascade Homo sapiens R-HSA-109704 | 3.26E-02 | 3.43E-01 | 4.51E+00 | 1.54E+01 | PRKAA2;FGF9;PIK3CB |
| 33 | Transcriptional regulation of white adipocyte differentiation Homo sapiens R-HSA-381340 | 3.26E-02 | 3.43E-01 | 4.51E+00 | 1.54E+01 | LPL;PPARGC1A;MED13L |
| 34 | PI5P, PP2A and IER3 Regulate PI3K/AKT Signaling Homo sapiens R-HSA-6811558 | 3.69E-02 | 3.43E-01 | 4.28E+00 | 1.41E+01 | FGF9;NRG1;PIK3CB |
| 35 | Gastrin-CREB signalling pathway via PKC and MAPK Homo sapiens R-HSA-881907 | 3.77E-02 | 3.43E-01 | 2.18E+00 | 7.14E+00 | GRM5;FGF9;PSME3;GNAQ;NEFL;NRG1;RASGRP1;KNG1 |
| 36 | PI3K/AKT Signaling in Cancer Homo sapiens R-HSA-2219528 | 4.03E-02 | 3.43E-01 | 4.12E+00 | 1.32E+01 | FGF9;NRG1;PIK3CB |

| | | | | | | |
|---|---|---|---|---|---|---|
| 37 | Steroid hormones Homo sapiens R-HSA-209943 | 4.19E-02 | 3.43E-01 | 6.50E+00 | 2.06E+01 | POMC;GC |
| 38 | Signaling by PDGF Homo sapiens R-HSA-186797 | 4.25E-02 | 3.43E-01 | 2.26E+00 | 7.13E+00 | FGF9;PSME3;NEFL;NRG1;PIK3CB;RASGRP1;THBS1 |
| 39 | N-glycan trimming and elongation in the cis-Golgi Homo sapiens R-HSA-964739 | 4.32E-02 | 3.43E-01 | 2.83E+01 | 8.89E+01 | MAN1A1 |
| 40 | Beta oxidation of lauroyl-CoA to decanoyl-CoA-CoA Homo sapiens R-HSA-77310 | 4.32E-02 | 3.43E-01 | 2.83E+01 | 8.89E+01 | ACADL |
| 41 | Alpha-oxidation of phytanate Homo sapiens R-HSA-389599 | 4.32E-02 | 3.43E-01 | 2.83E+01 | 8.89E+01 | SLC27A2 |
| 42 | GRB7 events in ERBB2 signaling Homo sapiens R-HSA-1306955 | 4.32E-02 | 3.43E-01 | 2.83E+01 | 8.89E+01 | NRG1 |
| 43 | Activation of RAS in B cells Homo sapiens R-HSA-1169092 | 4.32E-02 | 3.43E-01 | 2.83E+01 | 8.89E+01 | RASGRP1 |
| 44 | Negative regulation of the PI3K/AKT network Homo sapiens R-HSA-199418 | 4.51E-02 | 3.43E-01 | 3.93E+00 | 1.22E+01 | FGF9;NRG1;PIK3CB |
| 45 | Formation of Fibrin Clot (Clotting Cascade) Homo sapiens R-HSA-140877 | 4.61E-02 | 3.43E-01 | 6.15E+00 | 1.89E+01 | THBD;KNG1 |
| 46 | Metabolism Homo sapiens R-HSA-1430728 | 4.71E-02 | 3.43E-01 | 1.50E+00 | 4.59E+00 | PRKAA2;GLDC;SCD5;BGN;LPL;MT1X;GPT;PIK3CB;GPCPD1;MED13L;GNMT;POMC;MT2A;RBP4;ACADL;PSME3;GNAQ;MT1G;MT1H;AGXT;GC;LPIN1;SLC27A2;PPARGC1A |
| 47 | Cytokine Signaling in Immune system Homo sapiens R-HSA-1280215 | 4.80E-02 | 3.43E-01 | 1.90E+00 | 5.76E+00 | MT2A;TRIM6;FGF9;PSME3;IL20RA;NEFL;NRG1;PIK3CB;RASGRP1;TRIM45 |
| 48 | Interferon gamma signaling Homo sapiens R-HSA-877300 | 4.89E-02 | 3.43E-01 | 3.80E+00 | 1.15E+01 | TRIM6;MT2A;TRIM45 |
| 49 | Elastic fibre formation Homo sapiens R-HSA-1566948 | 5.05E-02 | 3.43E-01 | 5.83E+00 | 1.74E+01 | ITGB6;BMP7 |
| 50 | mitochondrial fatty acid beta-oxidation of unsaturated fatty acids Homo sapiens R-HSA-77288 | 5.17E-02 | 3.43E-01 | 2.27E+01 | 6.71E+01 | ACADL |
| 51 | Metabolic disorders of biological oxidation enzymes Homo sapiens R-HSA-5579029 | 5.17E-02 | 3.43E-01 | 2.27E+01 | 6.71E+01 | POMC |
| 52 | PTK6 Regulates Cell Cycle Homo sapiens R-HSA-8849470 | 5.17E-02 | 3.43E-01 | 2.27E+01 | 6.71E+01 | CCND1 |
| 53 | Retinoid metabolism and transport Homo sapiens R-HSA-975634 | 5.27E-02 | 3.43E-01 | 5.69E+00 | 1.67E+01 | RBP4;LPL |
| 54 | BMAL1:CLOCK,NPAS2 activates circadian gene expression Homo sapiens R-HSA-1368108 | 5.27E-02 | 3.43E-01 | 5.69E+00 | 1.67E+01 | NRIP1;PPARGC1A |

| | | | | | | |
|---|---|---|---|---|---|---|
| 55 | Signaling by the B Cell Receptor (BCR) Homo sapiens R-HSA-983705 | 5.54E-02 | 3.43E-01 | 2.51E+00 | 7.27E+00 | FGF9;PSME3;NRG1;PIK3CB;RASGRP1 |
| 56 | Hemostasis Homo sapiens R-HSA-109582 | 5.58E-02 | 3.43E-01 | 1.91E+00 | 5.52E+00 | THBD;PLAU;GNAQ;KCNMB2;SIRPA;PIK3CB;RASGRP1;THBS1;KNG1 |
| 57 | GRB2 events in EGFR signaling Homo sapiens R-HSA-179812 | 5.71E-02 | 3.43E-01 | 2.49E+00 | 7.13E+00 | FGF9;PSME3;NEFL;NRG1;RASGRP1 |
| 58 | SHC1 events in EGFR signaling Homo sapiens R-HSA-180336 | 5.71E-02 | 3.43E-01 | 2.49E+00 | 7.13E+00 | FGF9;PSME3;NEFL;NRG1;RASGRP1 |
| 59 | SOS-mediated signalling Homo sapiens R-HSA-112412 | 5.71E-02 | 3.43E-01 | 2.49E+00 | 7.13E+00 | FGF9;PSME3;NEFL;NRG1;RASGRP1 |
| 60 | SHC1 events in ERBB4 signaling Homo sapiens R-HSA-1250347 | 5.71E-02 | 3.43E-01 | 2.49E+00 | 7.13E+00 | FGF9;PSME3;NEFL;NRG1;RASGRP1 |
| 61 | RAF/MAP kinase cascade Homo sapiens R-HSA-5673001 | 5.71E-02 | 3.43E-01 | 2.49E+00 | 7.13E+00 | FGF9;PSME3;NEFL;NRG1;RASGRP1 |
| 62 | FRS-mediated FGFR2 signaling Homo sapiens R-HSA-5654700 | 5.79E-02 | 3.43E-01 | 2.48E+00 | 7.07E+00 | FGF9;PSME3;NEFL;NRG1;RASGRP1 |
| 63 | FRS-mediated FGFR4 signaling Homo sapiens R-HSA-5654712 | 5.79E-02 | 3.43E-01 | 2.48E+00 | 7.07E+00 | FGF9;PSME3;NEFL;NRG1;RASGRP1 |
| 64 | FRS-mediated FGFR3 signaling Homo sapiens R-HSA-5654706 | 5.79E-02 | 3.43E-01 | 2.48E+00 | 7.07E+00 | FGF9;PSME3;NEFL;NRG1;RASGRP1 |
| 65 | FRS-mediated FGFR1 signaling Homo sapiens R-HSA-5654693 | 5.79E-02 | 3.43E-01 | 2.48E+00 | 7.07E+00 | FGF9;PSME3;NEFL;NRG1;RASGRP1 |
| 66 | Signaling by Interleukins Homo sapiens R-HSA-449147 | 5.88E-02 | 3.43E-01 | 2.09E+00 | 5.93E+00 | FGF9;PSME3;IL20RA;NEFL;NRG1;PIK3CB;RASGRP1 |
| 67 | Cell surface interactions at the vascular wall Homo sapiens R-HSA-202733 | 5.98E-02 | 3.43E-01 | 3.49E+00 | 9.83E+00 | THBD;SIRPA;PIK3CB |
| 68 | Cation-coupled Chloride cotransporters Homo sapiens R-HSA-426117 | 6.00E-02 | 3.43E-01 | 1.89E+01 | 5.31E+01 | SLC12A1 |
| 69 | Vitamin D (calciferol) metabolism Homo sapiens R-HSA-196791 | 6.00E-02 | 3.43E-01 | 1.89E+01 | 5.31E+01 | GC |
| 70 | Defective CHST3 causes SEDCJD Homo sapiens R-HSA-3595172 | 6.00E-02 | 3.43E-01 | 1.89E+01 | 5.31E+01 | BGN |
| 71 | Defective CHSY1 causes TPBS Homo sapiens R-HSA-3595177 | 6.00E-02 | 3.43E-01 | 1.89E+01 | 5.31E+01 | BGN |
| 72 | Defective CHST14 causes EDS, musculocontractural type Homo sapiens R-HSA-3595174 | 6.00E-02 | 3.43E-01 | 1.89E+01 | 5.31E+01 | BGN |
| 73 | Signaling by NOTCH1 t(7;9)(NOTCH1:M1580 K2555) Translocation Mutant Homo sapiens R-HSA-2660825 | 6.00E-02 | 3.43E-01 | 1.89E+01 | 5.31E+01 | JAG1 |

| | | | | | | |
|---|---|---|---|---|---|---|
| 74 | Constitutive Signaling by NOTCH1 t(7;9)(NOTCH1:M1580 K2555) Translocation Mutant Homo sapiens R-HSA-2660826 | 6.00E-02 | 3.43E-01 | 1.89E+01 | 5.31E+01 | JAG1 |
| 75 | FGFR3b ligand binding and activation Homo sapiens R-HSA-190371 | 6.00E-02 | 3.43E-01 | 1.89E+01 | 5.31E+01 | FGF9 |
| 76 | ARMS-mediated activation Homo sapiens R-HSA-170984 | 6.05E-02 | 3.43E-01 | 2.45E+00 | 6.87E+00 | FGF9;PSME3;NEFL;NRG1;RASGRP1 |
| 77 | Signalling to p38 via RIT and RIN Homo sapiens R-HSA-187706 | 6.05E-02 | 3.43E-01 | 2.45E+00 | 6.87E+00 | FGF9;PSME3;NEFL;NRG1;RASGRP1 |
| 78 | Frs2-mediated activation Homo sapiens R-HSA-170968 | 6.13E-02 | 3.43E-01 | 2.44E+00 | 6.80E+00 | FGF9;PSME3;NEFL;NRG1;RASGRP1 |
| 79 | MAPK1/MAPK3 signaling Homo sapiens R-HSA-5684996 | 6.22E-02 | 3.43E-01 | 2.43E+00 | 6.74E+00 | FGF9;PSME3;NEFL;NRG1;RASGRP1 |
| 80 | Prolonged ERK activation events Homo sapiens R-HSA-169893 | 6.31E-02 | 3.43E-01 | 2.42E+00 | 6.68E+00 | FGF9;PSME3;NEFL;NRG1;RASGRP1 |
| 81 | Signaling by Leptin Homo sapiens R-HSA-2586552 | 6.40E-02 | 3.43E-01 | 2.41E+00 | 6.61E+00 | FGF9;PSME3;NEFL;NRG1;RASGRP1 |
| 82 | Signalling to RAS Homo sapiens R-HSA-167044 | 6.67E-02 | 3.43E-01 | 2.38E+00 | 6.43E+00 | FGF9;PSME3;NEFL;NRG1;RASGRP1 |
| 83 | Signaling by SCF-KIT Homo sapiens R-HSA-1433557 | 6.80E-02 | 3.43E-01 | 2.16E+00 | 5.80E+00 | FGF9;PSME3;NEFL;NRG1;PIK3CB;RASGRP1 |
| 84 | RNF mutants show enhanced WNT signaling and proliferation Homo sapiens R-HSA-5340588 | 6.83E-02 | 3.43E-01 | 1.62E+01 | 4.34E+01 | RNF43 |
| 85 | Free fatty acids regulate insulin secretion Homo sapiens R-HSA-400451 | 6.83E-02 | 3.43E-01 | 1.62E+01 | 4.34E+01 | GNAQ |
| 86 | Fatty Acids bound to GPR40 (FFAR1) regulate insulin secretion Homo sapiens R-HSA-434316 | 6.83E-02 | 3.43E-01 | 1.62E+01 | 4.34E+01 | GNAQ |
| 87 | AMPK inhibits chREBP transcriptional activation activity Homo sapiens R-HSA-163680 | 6.83E-02 | 3.43E-01 | 1.62E+01 | 4.34E+01 | PRKAA2 |
| 88 | Hydrolysis of LPC Homo sapiens R-HSA-1483115 | 6.83E-02 | 3.43E-01 | 1.62E+01 | 4.34E+01 | GPCPD1 |
| 89 | mitochondrial fatty acid beta-oxidation of saturated fatty acids Homo sapiens R-HSA-77286 | 6.83E-02 | 3.43E-01 | 1.62E+01 | 4.34E+01 | ACADL |
| 90 | Metabolism of ingested SeMet, Sec, MeSec into H2Se Homo sapiens R-HSA-2408508 | 6.83E-02 | 3.43E-01 | 1.62E+01 | 4.34E+01 | GNMT |

| | | | | | | |
|---|---|---|---|---|---|---|
| 91 | NR1D1 (REV-ERBA) represses gene expression Homo sapiens R-HSA-1368071 | 6.83E-02 | 3.43E-01 | 1.62E+01 | 4.34E+01 | PPARGC1A |
| 92 | VEGFR2 mediated cell proliferation Homo sapiens R-HSA-5218921 | 6.85E-02 | 3.43E-01 | 2.36E+00 | 6.32E+00 | FGF9;PSME3;NEFL;NRG1;RASGRP1 |
| 93 | Mitochondrial biogenesis Homo sapiens R-HSA-1592230 | 6.92E-02 | 3.43E-01 | 4.84E+00 | 1.29E+01 | PRKAA2;PPARGC1A |
| 94 | Cross-presentation of soluble exogenous antigens (endosomes) Homo sapiens R-HSA-1236978 | 6.92E-02 | 3.43E-01 | 4.84E+00 | 1.29E+01 | MRC2;PSME3 |
| 95 | Downstream signaling of activated FGFR2 Homo sapiens R-HSA-5654696 | 7.12E-02 | 3.43E-01 | 2.13E+00 | 5.63E+00 | FGF9;PSME3;NEFL;NRG1;PIK3CB;RASGRP1 |
| 96 | Downstream signaling of activated FGFR4 Homo sapiens R-HSA-5654716 | 7.12E-02 | 3.43E-01 | 2.13E+00 | 5.63E+00 | FGF9;PSME3;NEFL;NRG1;PIK3CB;RASGRP1 |
| 97 | Downstream signaling of activated FGFR3 Homo sapiens R-HSA-5654708 | 7.12E-02 | 3.43E-01 | 2.13E+00 | 5.63E+00 | FGF9;PSME3;NEFL;NRG1;PIK3CB;RASGRP1 |
| 98 | Ubiquitin-dependent degradation of Cyclin D1 Homo sapiens R-HSA-69229 | 7.17E-02 | 3.43E-01 | 4.74E+00 | 1.25E+01 | CCND1;PSME3 |
| 99 | Ubiquitin-dependent degradation of Cyclin D Homo sapiens R-HSA-75815 | 7.17E-02 | 3.43E-01 | 4.74E+00 | 1.25E+01 | CCND1;PSME3 |
| 100 | Signaling by ERBB4 Homo sapiens R-HSA-1236394 | 7.20E-02 | 3.43E-01 | 2.12E+00 | 5.59E+00 | FGF9;PSME3;NEFL;NRG1;PIK3CB;RASGRP1 |
| 101 | Platelet activation, signaling and aggregation Homo sapiens R-HSA-76002 | 7.33E-02 | 3.43E-01 | 2.31E+00 | 6.03E+00 | GNAQ;PIK3CB;RASGRP1;THBS1;KNG1 |
| 102 | Signalling to ERKs Homo sapiens R-HSA-187687 | 7.33E-02 | 3.43E-01 | 2.31E+00 | 6.03E+00 | FGF9;PSME3;NEFL;NRG1;RASGRP1 |
| 103 | Signaling by FGFR4 Homo sapiens R-HSA-5654743 | 7.37E-02 | 3.43E-01 | 2.11E+00 | 5.51E+00 | FGF9;PSME3;NEFL;NRG1;PIK3CB;RASGRP1 |
| 104 | Downstream signaling of activated FGFR1 Homo sapiens R-HSA-5654687 | 7.37E-02 | 3.43E-01 | 2.11E+00 | 5.51E+00 | FGF9;PSME3;NEFL;NRG1;PIK3CB;RASGRP1 |
| 105 | Signaling by FGFR3 Homo sapiens R-HSA-5654741 | 7.45E-02 | 3.43E-01 | 2.10E+00 | 5.46E+00 | FGF9;PSME3;NEFL;NRG1;PIK3CB;RASGRP1 |
| 106 | Retinoid cycle disease events Homo sapiens R-HSA-2453864 | 7.65E-02 | 3.43E-01 | 1.42E+01 | 3.64E+01 | RBP4 |
| 107 | Diseases associated with visual transduction Homo sapiens R-HSA-2474795 | 7.65E-02 | 3.43E-01 | 1.42E+01 | 3.64E+01 | RBP4 |
| 108 | Ca2+ activated K+ channels Homo sapiens R-HSA-1296052 | 7.65E-02 | 3.43E-01 | 1.42E+01 | 3.64E+01 | KCNMB2 |
| 109 | Prostanoid ligand receptors Homo sapiens R-HSA-391908 | 7.65E-02 | 3.43E-01 | 1.42E+01 | 3.64E+01 | PTGER2 |

| | | | | | | |
|---|---|---|---|---|---|---|
| 110 | Glucocorticoid biosynthesis Homo sapiens R-HSA-194002 | 7.65E-02 | 3.43E-01 | 1.42E+01 | 3.64E+01 | POMC |
| 111 | Interleukin-19, 20, 22, 24 Homo sapiens R-HSA-8854691 | 7.65E-02 | 3.43E-01 | 1.42E+01 | 3.64E+01 | IL20RA |
| 112 | Small interfering RNA (siRNA) biogenesis Homo sapiens R-HSA-426486 | 7.65E-02 | 3.43E-01 | 1.42E+01 | 3.64E+01 | DICER1 |
| 113 | Signaling by FGFR1 Homo sapiens R-HSA-5654736 | 7.70E-02 | 3.43E-01 | 2.08E+00 | 5.34E+00 | FGF9;PSME3;NEFL;NRG1;PIK3CB;RASGRP1 |
| 114 | Downstream signal transduction Homo sapiens R-HSA-186763 | 8.14E-02 | 3.43E-01 | 2.05E+00 | 5.15E+00 | FGF9;PSME3;NEFL;NRG1;PIK3CB;RASGRP1 |
| 115 | DAP12 signaling Homo sapiens R-HSA-2424491 | 8.40E-02 | 3.43E-01 | 2.03E+00 | 5.04E+00 | FGF9;PSME3;NEFL;NRG1;PIK3CB;RASGRP1 |
| 116 | ECM proteoglycans Homo sapiens R-HSA-3000178 | 8.45E-02 | 3.43E-01 | 4.29E+00 | 1.06E+01 | BGN;ITGB6 |
| 117 | Acetylcholine regulates insulin secretion Homo sapiens R-HSA-399997 | 8.46E-02 | 3.43E-01 | 1.26E+01 | 3.11E+01 | GNAQ |
| 118 | Regulation of gene expression by Hypoxia-inducible Factor Homo sapiens R-HSA-1234158 | 8.46E-02 | 3.43E-01 | 1.26E+01 | 3.11E+01 | CITED2 |
| 119 | NCAM signaling for neurite out-growth Homo sapiens R-HSA-375165 | 8.64E-02 | 3.43E-01 | 2.19E+00 | 5.37E+00 | FGF9;PSME3;NEFL;NRG1;RASGRP1 |
| 120 | G alpha (q) signalling events Homo sapiens R-HSA-416476 | 8.85E-02 | 3.43E-01 | 2.44E+00 | 5.92E+00 | GRM5;GNAQ;RASGRP1;KNG1 |
| 121 | Constitutive Signaling by NOTCH1 HD+PEST Domain Mutants Homo sapiens R-HSA-2894862 | 8.98E-02 | 3.43E-01 | 4.13E+00 | 9.96E+00 | JAG1;HEY1 |
| 122 | Signaling by NOTCH1 in Cancer Homo sapiens R-HSA-2644603 | 8.98E-02 | 3.43E-01 | 4.13E+00 | 9.96E+00 | JAG1;HEY1 |
| 123 | Signaling by NOTCH1 PEST Domain Mutants in Cancer Homo sapiens R-HSA-2644602 | 8.98E-02 | 3.43E-01 | 4.13E+00 | 9.96E+00 | JAG1;HEY1 |
| 124 | Constitutive Signaling by NOTCH1 PEST Domain Mutants Homo sapiens R-HSA-2644606 | 8.98E-02 | 3.43E-01 | 4.13E+00 | 9.96E+00 | JAG1;HEY1 |
| 125 | Signaling by NOTCH1 HD+PEST Domain Mutants in Cancer Homo sapiens R-HSA-2894858 | 8.98E-02 | 3.43E-01 | 4.13E+00 | 9.96E+00 | JAG1;HEY1 |
| 126 | Transport of organic anions Homo sapiens R-HSA-879518 | 9.27E-02 | 3.43E-01 | 1.13E+01 | 2.69E+01 | SLCO4C1 |
| 127 | RHO GTPases activate IQGAPs Homo sapiens R-HSA-5626467 | 9.27E-02 | 3.43E-01 | 1.13E+01 | 2.69E+01 | CTNNA1 |
| 128 | Dermatan sulfate biosynthesis Homo sapiens R-HSA-2022923 | 9.27E-02 | 3.43E-01 | 1.13E+01 | 2.69E+01 | BGN |

| | | | | | | |
|---|---|---|---|---|---|---|
| 129 | Import of palmitoyl-CoA into the mitochondrial matrix Homo sapiens R-HSA-200425 | 9.27E-02 | 3.43E-01 | 1.13E+01 | 2.69E+01 | PRKAA2 |
| 130 | Androgen biosynthesis Homo sapiens R-HSA-193048 | 9.27E-02 | 3.43E-01 | 1.13E+01 | 2.69E+01 | POMC |
| 131 | Signaling by NOTCH3 Homo sapiens R-HSA-1980148 | 9.27E-02 | 3.43E-01 | 1.13E+01 | 2.69E+01 | JAG1 |
| 132 | Signaling by NOTCH4 Homo sapiens R-HSA-1980150 | 9.27E-02 | 3.43E-01 | 1.13E+01 | 2.69E+01 | JAG1 |
| 133 | FGFR1c ligand binding and activation Homo sapiens R-HSA-190373 | 9.27E-02 | 3.43E-01 | 1.13E+01 | 2.69E+01 | FGF9 |
| 134 | Signaling by activated point mutants of FGFR1 Homo sapiens R-HSA-1839122 | 9.27E-02 | 3.43E-01 | 1.13E+01 | 2.69E+01 | FGF9 |
| 135 | Signaling by NOTCH Homo sapiens R-HSA-157118 | 9.30E-02 | 3.43E-01 | 2.87E+00 | 6.82E+00 | JAG1;CCND1;HEY1 |
| 136 | PI-3K cascade:FGFR1 Homo sapiens R-HSA-5654689 | 9.30E-02 | 3.43E-01 | 2.87E+00 | 6.82E+00 | FGF9;NRG1;PIK3CB |
| 137 | PI-3K cascade:FGFR3 Homo sapiens R-HSA-5654710 | 9.30E-02 | 3.43E-01 | 2.87E+00 | 6.82E+00 | FGF9;NRG1;PIK3CB |
| 138 | PI3K events in ERBB4 signaling Homo sapiens R-HSA-1250342 | 9.30E-02 | 3.43E-01 | 2.87E+00 | 6.82E+00 | FGF9;NRG1;PIK3CB |
| 139 | PIP3 activates AKT signaling Homo sapiens R-HSA-1257604 | 9.30E-02 | 3.43E-01 | 2.87E+00 | 6.82E+00 | FGF9;NRG1;PIK3CB |
| 140 | PI-3K cascade:FGFR4 Homo sapiens R-HSA-5654720 | 9.30E-02 | 3.43E-01 | 2.87E+00 | 6.82E+00 | FGF9;NRG1;PIK3CB |
| 141 | PI-3K cascade:FGFR2 Homo sapiens R-HSA-5654695 | 9.30E-02 | 3.43E-01 | 2.87E+00 | 6.82E+00 | FGF9;NRG1;PIK3CB |
| 142 | Signaling by EGFR Homo sapiens R-HSA-177929 | 9.42E-02 | 3.46E-01 | 1.97E+00 | 4.65E+00 | FGF9;PSME3;NEFL;NRG1;PIK3CB;RASGRP1 |
| 143 | SCF(Skp2)-mediated degradation of p27/p21 Homo sapiens R-HSA-187577 | 9.52E-02 | 3.47E-01 | 3.99E+00 | 9.37E+00 | CCND1;PSME3 |
| 144 | DAP12 interactions Homo sapiens R-HSA-2172127 | 9.80E-02 | 3.47E-01 | 1.95E+00 | 4.52E+00 | FGF9;PSME3;NEFL;NRG1;PIK3CB;RASGRP1 |
| 145 | GAB1 signalosome Homo sapiens R-HSA-180292 | 9.82E-02 | 3.47E-01 | 2.80E+00 | 6.50E+00 | FGF9;NRG1;PIK3CB |
| 146 | PI3K/AKT activation Homo sapiens R-HSA-198203 | 9.82E-02 | 3.47E-01 | 2.80E+00 | 6.50E+00 | FGF9;NRG1;PIK3CB |
| 147 | Signaling by FGFR2 Homo sapiens R-HSA-5654738 | 1.00E-01 | 3.47E-01 | 1.94E+00 | 4.46E+00 | FGF9;PSME3;NEFL;NRG1;PIK3CB;RASGRP1 |
| 148 | Peptide hormone biosynthesis Homo sapiens R-HSA-209952 | 1.01E-01 | 3.47E-01 | 1.03E+01 | 2.36E+01 | POMC |
| 149 | Physiological factors Homo sapiens R-HSA-5578768 | 1.01E-01 | 3.47E-01 | 1.03E+01 | 2.36E+01 | HIPK2 |
| 150 | FGFR3 mutant receptor activation Homo sapiens R-HSA-2033514 | 1.01E-01 | 3.47E-01 | 1.03E+01 | 2.36E+01 | FGF9 |

| | | | | | | |
|---|---|---|---|---|---|---|
| 151 | Signaling by activated point mutants of FGFR3 Homo sapiens R-HSA-1839130 | 1.01E-01 | 3.47E-01 | 1.03E+01 | 2.36E+01 | FGF9 |
| 152 | Signaling by FGFR Homo sapiens R-HSA-190236 | 1.05E-01 | 3.47E-01 | 1.91E+00 | 4.30E+00 | FGF9;PSME3;NEFL;NRG1;PIK3CB;RASGRP1 |
| 153 | Extracellular matrix organization Homo sapiens R-HSA-1474244 | 1.05E-01 | 3.47E-01 | 2.06E+00 | 4.63E+00 | ADAMTS16;BGN;ITGB6;BMP7;THBS1 |
| 154 | Circadian Clock Homo sapiens R-HSA-400253 | 1.06E-01 | 3.47E-01 | 3.72E+00 | 8.35E+00 | NRIP1;PPARGC1A |
| 155 | MAPK family signaling cascades Homo sapiens R-HSA-5683057 | 1.06E-01 | 3.47E-01 | 2.05E+00 | 4.59E+00 | FGF9;PSME3;NEFL;NRG1;RASGRP1 |
| 156 | Signal regulatory protein (SIRP) family interactions Homo sapiens R-HSA-391160 | 1.09E-01 | 3.47E-01 | 9.43E+00 | 2.09E+01 | SIRPA |
| 157 | Dissolution of Fibrin Clot Homo sapiens R-HSA-75205 | 1.09E-01 | 3.47E-01 | 9.43E+00 | 2.09E+01 | PLAU |
| 158 | Synthesis of PE Homo sapiens R-HSA-1483213 | 1.09E-01 | 3.47E-01 | 9.43E+00 | 2.09E+01 | LPIN1 |
| 159 | FGFR2c ligand binding and activation Homo sapiens R-HSA-190375 | 1.09E-01 | 3.47E-01 | 9.43E+00 | 2.09E+01 | FGF9 |
| 160 | FGFR3c ligand binding and activation Homo sapiens R-HSA-190372 | 1.09E-01 | 3.47E-01 | 9.43E+00 | 2.09E+01 | FGF9 |
| 161 | FGFR3 ligand binding and activation Homo sapiens R-HSA-190239 | 1.09E-01 | 3.47E-01 | 9.43E+00 | 2.09E+01 | FGF9 |
| 162 | ERBB2 Activates PTK6 Signaling Homo sapiens R-HSA-8847993 | 1.09E-01 | 3.47E-01 | 9.43E+00 | 2.09E+01 | NRG1 |
| 163 | Downregulation of ERBB2:ERBB3 signaling Homo sapiens R-HSA-1358803 | 1.09E-01 | 3.47E-01 | 9.43E+00 | 2.09E+01 | NRG1 |
| 164 | FCERI mediated MAPK activation Homo sapiens R-HSA-2871796 | 1.12E-01 | 3.54E-01 | 2.01E+00 | 4.40E+00 | FGF9;PSME3;NEFL;NRG1;RASGRP1 |
| 165 | NGF signalling via TRKA from the plasma membrane Homo sapiens R-HSA-187037 | 1.13E-01 | 3.54E-01 | 1.87E+00 | 4.06E+00 | FGF9;PSME3;NEFL;NRG1;PIK3CB;RASGRP1 |
| 166 | O-linked glycosylation of mucins Homo sapiens R-HSA-913709 | 1.15E-01 | 3.54E-01 | 3.55E+00 | 7.68E+00 | GCNT4;GALNT10 |
| 167 | Triglyceride Biosynthesis Homo sapiens R-HSA-75109 | 1.15E-01 | 3.54E-01 | 3.55E+00 | 7.68E+00 | SCD5;LPIN1 |
| 168 | CS/DS degradation Homo sapiens R-HSA-2024101 | 1.16E-01 | 3.54E-01 | 8.71E+00 | 1.87E+01 | BGN |
| 169 | Synthesis of bile acids and bile salts via 24-hydroxycholesterol Homo sapiens R-HSA-193775 | 1.16E-01 | 3.54E-01 | 8.71E+00 | 1.87E+01 | SLC27A2 |

| | | | | | | |
|---|---|---|---|---|---|---|
| 170 | Receptor-ligand binding initiates the second proteolytic cleavage of Notch receptor Homo sapiens R-HSA-156988 | 1.16E-01 | 3.54E-01 | 8.71E+00 | 1.87E+01 | JAG1 |
| 171 | FGFR4 ligand binding and activation Homo sapiens R-HSA-190322 | 1.16E-01 | 3.54E-01 | 8.71E+00 | 1.87E+01 | FGF9 |
| 172 | Integrin cell surface interactions Homo sapiens R-HSA-216083 | 1.18E-01 | 3.54E-01 | 3.49E+00 | 7.48E+00 | ITGB6;THBS1 |
| 173 | Signaling by PTK6 Homo sapiens R-HSA-8848021 | 1.18E-01 | 3.54E-01 | 3.49E+00 | 7.48E+00 | CCND1;NRG1 |
| 174 | Cyclin E associated events during G1/S transition Homo sapiens R-HSA-69202 | 1.21E-01 | 3.54E-01 | 3.44E+00 | 7.28E+00 | CCND1;PSME3 |
| 175 | Cyclin A:Cdk2-associated events at S phase entry Homo sapiens R-HSA-69656 | 1.23E-01 | 3.54E-01 | 3.39E+00 | 7.09E+00 | CCND1;PSME3 |
| 176 | CREB phosphorylation through the activation of CaMKII Homo sapiens R-HSA-442729 | 1.24E-01 | 3.54E-01 | 8.09E+00 | 1.69E+01 | NEFL |
| 177 | Depolymerisation of the Nuclear Lamina Homo sapiens R-HSA-4419969 | 1.24E-01 | 3.54E-01 | 8.09E+00 | 1.69E+01 | LPIN1 |
| 178 | Eicosanoid ligand-binding receptors Homo sapiens R-HSA-391903 | 1.24E-01 | 3.54E-01 | 8.09E+00 | 1.69E+01 | PTGER2 |
| 179 | mRNA decay by 5' to 3' exoribonuclease Homo sapiens R-HSA-430039 | 1.24E-01 | 3.54E-01 | 8.09E+00 | 1.69E+01 | DDX6 |
| 180 | Constitutive Signaling by NOTCH1 HD Domain Mutants Homo sapiens R-HSA-2691232 | 1.24E-01 | 3.54E-01 | 8.09E+00 | 1.69E+01 | JAG1 |
| 181 | Signaling by NOTCH1 HD Domain Mutants in Cancer Homo sapiens R-HSA-2691230 | 1.24E-01 | 3.54E-01 | 8.09E+00 | 1.69E+01 | JAG1 |
| 182 | FGFR1 ligand binding and activation Homo sapiens R-HSA-190242 | 1.24E-01 | 3.54E-01 | 8.09E+00 | 1.69E+01 | FGF9 |
| 183 | ERBB2 Regulates Cell Motility Homo sapiens R-HSA-6785631 | 1.24E-01 | 3.54E-01 | 8.09E+00 | 1.69E+01 | NRG1 |
| 184 | Cell-extracellular matrix interactions Homo sapiens R-HSA-446353 | 1.32E-01 | 3.67E-01 | 7.55E+00 | 1.53E+01 | LIMS2 |
| 185 | Rap1 signalling Homo sapiens R-HSA-392517 | 1.32E-01 | 3.67E-01 | 7.55E+00 | 1.53E+01 | RASGRP1 |
| 186 | PI3K events in ERBB2 signaling Homo sapiens R-HSA-1963642 | 1.32E-01 | 3.67E-01 | 7.55E+00 | 1.53E+01 | NRG1 |
| 187 | GRB2 events in ERBB2 signaling Homo sapiens R-HSA-1963640 | 1.32E-01 | 3.67E-01 | 7.55E+00 | 1.53E+01 | NRG1 |

| | | | | | | |
|---|---|---|---|---|---|---|
| 188 | Signaling by NOTCH1 Homo sapiens R-HSA-1980143 | 1.32E-01 | 3.67E-01 | 3.24E+00 | 6.56E+00 | JAG1;HEY1 |
| 189 | Fc epsilon receptor (FCERI) signaling Homo sapiens R-HSA-2454202 | 1.36E-01 | 3.75E-01 | 1.76E+00 | 3.51E+00 | FGF9;PSME3;NEFL;NRG1;PIK3CB;RASGRP1 |
| 190 | Ras activation uopn Ca2+ infux through NMDA receptor Homo sapiens R-HSA-442982 | 1.40E-01 | 3.75E-01 | 7.07E+00 | 1.39E+01 | NEFL |
| 191 | Unblocking of NMDA receptor, glutamate binding and activation Homo sapiens R-HSA-438066 | 1.40E-01 | 3.75E-01 | 7.07E+00 | 1.39E+01 | NEFL |
| 192 | Chylomicron-mediated lipid transport Homo sapiens R-HSA-174800 | 1.40E-01 | 3.75E-01 | 7.07E+00 | 1.39E+01 | LPL |
| 193 | Mitochondrial Fatty Acid Beta-Oxidation Homo sapiens R-HSA-77289 | 1.40E-01 | 3.75E-01 | 7.07E+00 | 1.39E+01 | ACADL |
| 194 | Activated point mutants of FGFR2 Homo sapiens R-HSA-2033519 | 1.40E-01 | 3.75E-01 | 7.07E+00 | 1.39E+01 | FGF9 |
| 195 | Metabolism of vitamins and cofactors Homo sapiens R-HSA-196854 | 1.42E-01 | 3.80E-01 | 2.35E+00 | 4.59E+00 | RBP4;LPL;GC |
| 196 | Regulation of signaling by CBL Homo sapiens R-HSA-912631 | 1.47E-01 | 3.83E-01 | 6.66E+00 | 1.28E+01 | PIK3CB |
| 197 | cGMP effects Homo sapiens R-HSA-418457 | 1.47E-01 | 3.83E-01 | 6.66E+00 | 1.28E+01 | KCNMB2 |
| 198 | SHC-mediated cascade:FGFR3 Homo sapiens R-HSA-5654704 | 1.47E-01 | 3.83E-01 | 6.66E+00 | 1.28E+01 | FGF9 |
| 199 | SHC1 events in ERBB2 signaling Homo sapiens R-HSA-1250196 | 1.47E-01 | 3.83E-01 | 6.66E+00 | 1.28E+01 | NRG1 |
| 200 | Tie2 Signaling Homo sapiens R-HSA-210993 | 1.47E-01 | 3.83E-01 | 6.66E+00 | 1.28E+01 | PIK3CB |
| 201 | Defective B3GALT6 causes EDSP2 and SEMDJL1 Homo sapiens R-HSA-4420332 | 1.55E-01 | 3.97E-01 | 6.29E+00 | 1.17E+01 | BGN |
| 202 | Defective B4GALT7 causes EDS, progeroid type Homo sapiens R-HSA-3560783 | 1.55E-01 | 3.97E-01 | 6.29E+00 | 1.17E+01 | BGN |
| 203 | Defective B3GAT3 causes JDSSDHD Homo sapiens R-HSA-3560801 | 1.55E-01 | 3.97E-01 | 6.29E+00 | 1.17E+01 | BGN |
| 204 | Developmental Biology Homo sapiens R-HSA-1266738 | 1.56E-01 | 3.99E-01 | 1.48E+00 | 2.75E+00 | FGF9;PSME3;CTNNA1;LPL;NEFL;NRG1;RASGRP1;PPARGC1A;PBX1;MED13L |
| 205 | Chondroitin sulfate biosynthesis Homo sapiens R-HSA-2022870 | 1.62E-01 | 4.06E-01 | 5.96E+00 | 1.08E+01 | BGN |

| | | | | | | |
|---|---|---|---|---|---|---|
| 206 | Syndecan interactions Homo sapiens R-HSA-3000170 | 1.62E-01 | 4.06E-01 | 5.96E+00 | 1.08E+01 | THBS1 |
| 207 | Synthesis of PC Homo sapiens R-HSA-1483191 | 1.62E-01 | 4.06E-01 | 5.96E+00 | 1.08E+01 | LPIN1 |
| 208 | FGFR2 ligand binding and activation Homo sapiens R-HSA-190241 | 1.62E-01 | 4.06E-01 | 5.96E+00 | 1.08E+01 | FGF9 |
| 209 | Antigen processing-Cross presentation Homo sapiens R-HSA-1236975 | 1.63E-01 | 4.06E-01 | 2.84E+00 | 5.15E+00 | MRC2;PSME3 |
| 210 | Phospholipid metabolism Homo sapiens R-HSA-1483257 | 1.69E-01 | 4.17E-01 | 2.16E+00 | 3.83E+00 | PIK3CB;GPCPD1;LPIN1 |
| 211 | NOTCH2 Activation and Transmission of Signal to the Nucleus Homo sapiens R-HSA-2979096 | 1.69E-01 | 4.17E-01 | 5.66E+00 | 1.00E+01 | JAG1 |
| 212 | SHC-mediated cascade:FGFR1 Homo sapiens R-HSA-5654688 | 1.69E-01 | 4.17E-01 | 5.66E+00 | 1.00E+01 | FGF9 |
| 213 | Role of LAT2/NTAL/LAB on calcium mobilization Homo sapiens R-HSA-2730905 | 1.72E-01 | 4.17E-01 | 2.14E+00 | 3.78E+00 | FGF9;NRG1;PIK3CB |
| 214 | Metabolism of amino acids and derivatives Homo sapiens R-HSA-71291 | 1.74E-01 | 4.17E-01 | 1.73E+00 | 3.02E+00 | GLDC;PSME3;GPT;AGXT;GNMT |
| 215 | Cell junction organization Homo sapiens R-HSA-446728 | 1.75E-01 | 4.17E-01 | 2.70E+00 | 4.71E+00 | CTNNA1;LIMS2 |
| 216 | Post-translational protein modification Homo sapiens R-HSA-597592 | 1.77E-01 | 4.17E-01 | 1.56E+00 | 2.70E+00 | ADAMTS16;ADAMTS15;SPON1;MAN1A1;GCNT4;THBS1;GALNT10 |
| 217 | Nephrin interactions Homo sapiens R-HSA-373753 | 1.77E-01 | 4.17E-01 | 5.39E+00 | 9.34E+00 | PIK3CB |
| 218 | Common Pathway of Fibrin Clot Formation Homo sapiens R-HSA-140875 | 1.77E-01 | 4.17E-01 | 5.39E+00 | 9.34E+00 | THBD |
| 219 | Intrinsic Pathway of Fibrin Clot Formation Homo sapiens R-HSA-140837 | 1.77E-01 | 4.17E-01 | 5.39E+00 | 9.34E+00 | KNG1 |
| 220 | Signaling by FGFR3 point mutants in cancer Homo sapiens R-HSA-8853338 | 1.77E-01 | 4.17E-01 | 5.39E+00 | 9.34E+00 | FGF9 |
| 221 | Signaling by FGFR3 in disease Homo sapiens R-HSA-5655332 | 1.77E-01 | 4.17E-01 | 5.39E+00 | 9.34E+00 | FGF9 |
| 222 | The canonical retinoid cycle in rods (twilight vision) Homo sapiens R-HSA-2453902 | 1.84E-01 | 4.24E-01 | 5.14E+00 | 8.71E+00 | RBP4 |
| 223 | Thromboxane signalling through TP receptor Homo sapiens R-HSA-428930 | 1.84E-01 | 4.24E-01 | 5.14E+00 | 8.71E+00 | GNAQ |
| 224 | Signaling by BMP Homo sapiens R-HSA-201451 | 1.84E-01 | 4.24E-01 | 5.14E+00 | 8.71E+00 | SMAD9 |

| | | | | | | |
|---|---|---|---|---|---|---|
| 225 | MicroRNA (miRNA) biogenesis Homo sapiens R-HSA-203927 | 1.84E-01 | 4.24E-01 | 5.14E+00 | 8.71E+00 | DICER1 |
| 226 | SHC-mediated cascade:FGFR4 Homo sapiens R-HSA-5654719 | 1.84E-01 | 4.24E-01 | 5.14E+00 | 8.71E+00 | FGF9 |
| 227 | Regulation of TP53 Activity through Phosphorylation Homo sapiens R-HSA-6804756 | 1.85E-01 | 4.24E-01 | 2.61E+00 | 4.40E+00 | PRKAA2;HIPK2 |
| 228 | Synthesis of bile acids and bile salts via 7alpha-hydroxycholesterol Homo sapiens R-HSA-193368 | 1.91E-01 | 4.35E-01 | 4.92E+00 | 8.14E+00 | SLC27A2 |
| 229 | Endogenous sterols Homo sapiens R-HSA-211976 | 1.91E-01 | 4.35E-01 | 4.92E+00 | 8.14E+00 | POMC |
| 230 | Amino acid synthesis and interconversion (transamination) Homo sapiens R-HSA-70614 | 1.98E-01 | 4.45E-01 | 4.71E+00 | 7.63E+00 | GPT |
| 231 | ADP signalling through P2Y purinoceptor 1 Homo sapiens R-HSA-418592 | 1.98E-01 | 4.45E-01 | 4.71E+00 | 7.63E+00 | GNAQ |
| 232 | Nitric oxide stimulates guanylate cyclase Homo sapiens R-HSA-392154 | 1.98E-01 | 4.45E-01 | 4.71E+00 | 7.63E+00 | KCNMB2 |
| 233 | Diseases associated with glycosaminoglycan metabolism Homo sapiens R-HSA-3560782 | 2.05E-01 | 4.46E-01 | 4.53E+00 | 7.16E+00 | BGN |
| 234 | A tetrasaccharide linker sequence is required for GAG synthesis Homo sapiens R-HSA-1971475 | 2.05E-01 | 4.46E-01 | 4.53E+00 | 7.16E+00 | BGN |
| 235 | Attenuation phase Homo sapiens R-HSA-3371568 | 2.05E-01 | 4.46E-01 | 4.53E+00 | 7.16E+00 | HSPA1B |
| 236 | Regulation of Hypoxia-inducible Factor (HIF) by oxygen Homo sapiens R-HSA-1234174 | 2.05E-01 | 4.46E-01 | 4.53E+00 | 7.16E+00 | CITED2 |
| 237 | Cellular response to hypoxia Homo sapiens R-HSA-2262749 | 2.05E-01 | 4.46E-01 | 4.53E+00 | 7.16E+00 | CITED2 |
| 238 | SHC-mediated cascade:FGFR2 Homo sapiens R-HSA-5654699 | 2.05E-01 | 4.46E-01 | 4.53E+00 | 7.16E+00 | FGF9 |
| 239 | Signalling by NGF Homo sapiens R-HSA-166520 | 2.06E-01 | 4.46E-01 | 1.54E+00 | 2.43E+00 | FGF9;PSME3;NEFL;NRG1;PIK3CB;RASGRP1 |
| 240 | Visual phototransduction Homo sapiens R-HSA-2187338 | 2.10E-01 | 4.46E-01 | 2.39E+00 | 3.72E+00 | RBP4;LPL |
| 241 | Integrin alphaIIb beta3 signaling Homo sapiens R-HSA-354192 | 2.12E-01 | 4.46E-01 | 4.35E+00 | 6.74E+00 | RASGRP1 |
| 242 | CREB phosphorylation through the activation of Ras Homo sapiens R-HSA-442742 | 2.12E-01 | 4.46E-01 | 4.35E+00 | 6.74E+00 | NEFL |

| | | | | | | |
|---|---|---|---|---|---|---|
| 243 | Effects of PIP2 hydrolysis Homo sapiens R-HSA-114508 | 2.12E-01 | 4.46E-01 | 4.35E+00 | 6.74E+00 | RASGRP1 |
| 244 | VEGFR2 mediated vascular permeability Homo sapiens R-HSA-5218920 | 2.12E-01 | 4.46E-01 | 4.35E+00 | 6.74E+00 | CTNNA1 |
| 245 | Peroxisomal lipid metabolism Homo sapiens R-HSA-390918 | 2.12E-01 | 4.46E-01 | 4.35E+00 | 6.74E+00 | SLC27A2 |
| 246 | Synthesis of bile acids and bile salts Homo sapiens R-HSA-192105 | 2.12E-01 | 4.46E-01 | 4.35E+00 | 6.74E+00 | SLC27A2 |
| 247 | RNA Pol II CTD phosphorylation and interaction with CE Homo sapiens R-HSA-167160 | 2.12E-01 | 4.46E-01 | 4.35E+00 | 6.74E+00 | GTF2H2 |
| 248 | RNA Pol II CTD phosphorylation and interaction with CE Homo sapiens R-HSA-77075 | 2.12E-01 | 4.46E-01 | 4.35E+00 | 6.74E+00 | GTF2H2 |
| 249 | Downstream TCR signaling Homo sapiens R-HSA-202424 | 2.17E-01 | 4.53E-01 | 2.34E+00 | 3.58E+00 | PSME3;PIK3CB |
| 250 | G-protein activation Homo sapiens R-HSA-202040 | 2.19E-01 | 4.54E-01 | 4.19E+00 | 6.36E+00 | POMC |
| 251 | Energy dependent regulation of mTOR by LKB1-AMPK Homo sapiens R-HSA-380972 | 2.19E-01 | 4.54E-01 | 4.19E+00 | 6.36E+00 | PRKAA2 |
| 252 | Myogenesis Homo sapiens R-HSA-525793 | 2.26E-01 | 4.54E-01 | 4.04E+00 | 6.00E+00 | CTNNA1 |
| 253 | CDO in myogenesis Homo sapiens R-HSA-375170 | 2.26E-01 | 4.54E-01 | 4.04E+00 | 6.00E+00 | CTNNA1 |
| 254 | COPI-independent Golgi-to-ER retrograde traffic Homo sapiens R-HSA-6811436 | 2.26E-01 | 4.54E-01 | 4.04E+00 | 6.00E+00 | BICD1 |
| 255 | Transferrin endocytosis and recycling Homo sapiens R-HSA-917977 | 2.26E-01 | 4.54E-01 | 4.04E+00 | 6.00E+00 | TFRC |
| 256 | HSF1 activation Homo sapiens R-HSA-3371511 | 2.26E-01 | 4.54E-01 | 4.04E+00 | 6.00E+00 | HSPA1B |
| 257 | YAP1- and WWTR1 (TAZ)-stimulated gene expression Homo sapiens R-HSA-2032785 | 2.26E-01 | 4.54E-01 | 4.04E+00 | 6.00E+00 | HIPK2 |
| 258 | mRNA Capping Homo sapiens R-HSA-72086 | 2.26E-01 | 4.54E-01 | 4.04E+00 | 6.00E+00 | GTF2H2 |
| 259 | Negative regulation of FGFR3 signaling Homo sapiens R-HSA-5654732 | 2.26E-01 | 4.54E-01 | 4.04E+00 | 6.00E+00 | FGF9 |
| 260 | Glycerophospholipid biosynthesis Homo sapiens R-HSA-1483206 | 2.26E-01 | 4.54E-01 | 2.27E+00 | 3.37E+00 | GPCPD1;LPIN1 |

| | | | | | | |
|---|---|---|---|---|---|---|
| 261 | Adaptive Immune System Homo sapiens R-HSA-1280218 | 2.30E-01 | 4.58E-01 | 1.36E+00 | 2.01E+00 | MRC2;RNF144B;FGF9;UBA6;PSME3;NRG1;PIK3CB;HLA-DOA;RASGRP1 |
| 262 | Metabolism of steroid hormones Homo sapiens R-HSA-196071 | 2.33E-01 | 4.60E-01 | 3.90E+00 | 5.68E+00 | POMC |
| 263 | RNA Polymerase I Promoter Escape Homo sapiens R-HSA-73772 | 2.33E-01 | 4.60E-01 | 3.90E+00 | 5.68E+00 | GTF2H2 |
| 264 | Activated NOTCH1 Transmits Signal to the Nucleus Homo sapiens R-HSA-2122948 | 2.33E-01 | 4.60E-01 | 3.90E+00 | 5.68E+00 | JAG1 |
| 265 | Platelet degranulation Homo sapiens R-HSA-114608 | 2.36E-01 | 4.64E-01 | 2.20E+00 | 3.18E+00 | THBS1;KNG1 |
| 266 | Adherens junctions interactions Homo sapiens R-HSA-418990 | 2.40E-01 | 4.66E-01 | 3.77E+00 | 5.38E+00 | CTNNA1 |
| 267 | Negative regulation of FGFR4 signaling Homo sapiens R-HSA-5654733 | 2.40E-01 | 4.66E-01 | 3.77E+00 | 5.38E+00 | FGF9 |
| 268 | FGFR1 mutant receptor activation Homo sapiens R-HSA-1839124 | 2.40E-01 | 4.66E-01 | 3.77E+00 | 5.38E+00 | FGF9 |
| 269 | Signal amplification Homo sapiens R-HSA-392518 | 2.47E-01 | 4.69E-01 | 3.65E+00 | 5.11E+00 | GNAQ |
| 270 | Thrombin signalling through proteinase activated receptors (PARs) Homo sapiens R-HSA-456926 | 2.47E-01 | 4.69E-01 | 3.65E+00 | 5.11E+00 | GNAQ |
| 271 | RNA Polymerase I Transcription Termination Homo sapiens R-HSA-73863 | 2.47E-01 | 4.69E-01 | 3.65E+00 | 5.11E+00 | GTF2H2 |
| 272 | Signaling by NOTCH2 Homo sapiens R-HSA-1980145 | 2.47E-01 | 4.69E-01 | 3.65E+00 | 5.11E+00 | JAG1 |
| 273 | Oncogene Induced Senescence Homo sapiens R-HSA-2559585 | 2.47E-01 | 4.69E-01 | 3.65E+00 | 5.11E+00 | ID1 |
| 274 | Negative regulation of FGFR1 signaling Homo sapiens R-HSA-5654726 | 2.47E-01 | 4.69E-01 | 3.65E+00 | 5.11E+00 | FGF9 |
| 275 | Interferon Signaling Homo sapiens R-HSA-913531 | 2.49E-01 | 4.70E-01 | 1.76E+00 | 2.45E+00 | MT2A;TRIM6;TRIM45 |
| 276 | Response to elevated platelet cytosolic Ca2+ Homo sapiens R-HSA-76005 | 2.52E-01 | 4.70E-01 | 2.10E+00 | 2.89E+00 | THBS1;KNG1 |
| 277 | Integration of energy metabolism Homo sapiens R-HSA-163685 | 2.52E-01 | 4.70E-01 | 2.10E+00 | 2.89E+00 | PRKAA2;GNAQ |
| 278 | Formation of the Early Elongation Complex Homo sapiens R-HSA-113418 | 2.53E-01 | 4.70E-01 | 3.53E+00 | 4.86E+00 | GTF2H2 |

| | | | | | | |
|---|---|---|---|---|---|---|
| 279 | Formation of the HIV-1 Early Elongation Complex Homo sapiens R-HSA-167158 | 2.53E-01 | 4.70E-01 | 3.53E+00 | 4.86E+00 | GTF2H2 |
| 280 | FGFR2 mutant receptor activation Homo sapiens R-HSA-1839126 | 2.53E-01 | 4.70E-01 | 3.53E+00 | 4.86E+00 | FGF9 |
| 281 | FCERI mediated NF-kB activation Homo sapiens R-HSA-2871837 | 2.56E-01 | 4.70E-01 | 2.08E+00 | 2.84E+00 | PSME3;RASGRP1 |
| 282 | TCF dependent signaling in response to WNT Homo sapiens R-HSA-201681 | 2.56E-01 | 4.70E-01 | 1.74E+00 | 2.37E+00 | RNF43;ZNRF3;PSME3 |
| 283 | G1/S Transition Homo sapiens R-HSA-69206 | 2.59E-01 | 4.70E-01 | 2.06E+00 | 2.78E+00 | CCND1;PSME3 |
| 284 | Lipoprotein metabolism Homo sapiens R-HSA-174824 | 2.60E-01 | 4.70E-01 | 3.43E+00 | 4.62E+00 | LPL |
| 285 | Synthesis of PIPs at the plasma membrane Homo sapiens R-HSA-1660499 | 2.60E-01 | 4.70E-01 | 3.43E+00 | 4.62E+00 | PIK3CB |
| 286 | HSF1-dependent transactivation Homo sapiens R-HSA-3371571 | 2.60E-01 | 4.70E-01 | 3.43E+00 | 4.62E+00 | HSPA1B |
| 287 | Signaling by WNT in cancer Homo sapiens R-HSA-4791275 | 2.60E-01 | 4.70E-01 | 3.43E+00 | 4.62E+00 | RNF43 |
| 288 | Negative regulation of FGFR2 signaling Homo sapiens R-HSA-5654727 | 2.60E-01 | 4.70E-01 | 3.43E+00 | 4.62E+00 | FGF9 |
| 289 | PPARA activates gene expression Homo sapiens R-HSA-1989781 | 2.62E-01 | 4.73E-01 | 2.04E+00 | 2.73E+00 | PPARGC1A;MED13L |
| 290 | Post NMDA receptor activation events Homo sapiens R-HSA-438064 | 2.66E-01 | 4.77E-01 | 3.33E+00 | 4.40E+00 | NEFL |
| 291 | Bile acid and bile salt metabolism Homo sapiens R-HSA-194068 | 2.66E-01 | 4.77E-01 | 3.33E+00 | 4.40E+00 | SLC27A2 |
| 292 | Regulation of lipid metabolism by Peroxisome proliferator-activated receptor alpha (PPARalpha) Homo sapiens R-HSA-400206 | 2.72E-01 | 4.85E-01 | 1.99E+00 | 2.59E+00 | PPARGC1A;MED13L |
| 293 | TCR signaling Homo sapiens R-HSA-202403 | 2.78E-01 | 4.95E-01 | 1.95E+00 | 2.50E+00 | PSME3;PIK3CB |
| 294 | Platelet Aggregation (Plug Formation) Homo sapiens R-HSA-76009 | 2.79E-01 | 4.95E-01 | 3.14E+00 | 4.01E+00 | RASGRP1 |
| 295 | Class I MHC mediated antigen processing & presentation Homo sapiens R-HSA-983169 | 2.82E-01 | 4.96E-01 | 1.51E+00 | 1.91E+00 | MRC2;RNF144B;UBA6;PSME3 |

| | | | | | | |
|---|---|---|---|---|---|---|
| 296 | Transport of vitamins, nucleosides, and related molecules Homo sapiens R-HSA-425397 | 2.86E-01 | 4.96E-01 | 3.06E+00 | 3.83E+00 | SLCO4C1 |
| 297 | Beta defensins Homo sapiens R-HSA-1461957 | 2.86E-01 | 4.96E-01 | 3.06E+00 | 3.83E+00 | DEFB1 |
| 298 | G1 Phase Homo sapiens R-HSA-69236 | 2.86E-01 | 4.96E-01 | 3.06E+00 | 3.83E+00 | CCND1 |
| 299 | Cyclin D associated events in G1 Homo sapiens R-HSA-69231 | 2.86E-01 | 4.96E-01 | 3.06E+00 | 3.83E+00 | CCND1 |
| 300 | Signaling by FGFR1 in disease Homo sapiens R-HSA-5655302 | 2.86E-01 | 4.96E-01 | 3.06E+00 | 3.83E+00 | FGF9 |
| 301 | Immune System Homo sapiens R-HSA-168256 | 2.87E-01 | 4.97E-01 | 1.19E+00 | 1.49E+00 | UBA6;IL20RA;DEFB1;NRG1;PIK3CB;RASGRP1;MRC2;RNF144B;MT2A;TRIM6;FGF9;PSME3;NEFL;BPI;HLA-DOA;TRIM45 |
| 302 | Activation of NMDA receptor upon glutamate binding and postsynaptic events Homo sapiens R-HSA-442755 | 2.92E-01 | 4.98E-01 | 2.98E+00 | 3.66E+00 | NEFL |
| 303 | mTOR signalling Homo sapiens R-HSA-165159 | 2.92E-01 | 4.98E-01 | 2.98E+00 | 3.66E+00 | PRKAA2 |
| 304 | Dual Incision in GG-NER Homo sapiens R-HSA-5696400 | 2.92E-01 | 4.98E-01 | 2.98E+00 | 3.66E+00 | GTF2H2 |
| 305 | Nuclear signaling by ERBB4 Homo sapiens R-HSA-1251985 | 2.92E-01 | 4.98E-01 | 2.98E+00 | 3.66E+00 | NRG1 |
| 306 | S Phase Homo sapiens R-HSA-69242 | 2.98E-01 | 5.03E-01 | 1.86E+00 | 2.25E+00 | CCND1;PSME3 |
| 307 | Class C/3 (Metabotropic glutamate/pheromone receptors) Homo sapiens R-HSA-420499 | 2.98E-01 | 5.03E-01 | 2.90E+00 | 3.51E+00 | GRM5 |
| 308 | PKB-mediated events Homo sapiens R-HSA-109703 | 2.98E-01 | 5.03E-01 | 2.90E+00 | 3.51E+00 | PRKAA2 |
| 309 | Transcriptional activation of mitochondrial biogenesis Homo sapiens R-HSA-2151201 | 2.98E-01 | 5.03E-01 | 2.90E+00 | 3.51E+00 | PPARGC1A |
| 310 | Pre-NOTCH Transcription and Translation Homo sapiens R-HSA-1912408 | 3.04E-01 | 5.09E-01 | 2.83E+00 | 3.36E+00 | CCND1 |
| 311 | Non-integrin membrane-ECM interactions Homo sapiens R-HSA-3000171 | 3.10E-01 | 5.09E-01 | 2.76E+00 | 3.23E+00 | THBS1 |
| 312 | Fatty Acyl-CoA Biosynthesis Homo sapiens R-HSA-75105 | 3.10E-01 | 5.09E-01 | 2.76E+00 | 3.23E+00 | SCD5 |
| 313 | Transcriptional regulation of pluripotent stem cells Homo sapiens R-HSA-452723 | 3.10E-01 | 5.09E-01 | 2.76E+00 | 3.23E+00 | PBX1 |
| 314 | Iron uptake and transport Homo sapiens R-HSA-917937 | 3.16E-01 | 5.09E-01 | 2.69E+00 | 3.10E+00 | TFRC |

| | | | | | | |
|---|---|---|---|---|---|---|
| 315 | HIV Transcription Elongation Homo sapiens R-HSA-167169 | 3.16E-01 | 5.09E-01 | 2.69E+00 | 3.10E+00 | GTF2H2 |
| 316 | Tat-mediated elongation of the HIV-1 transcript Homo sapiens R-HSA-167246 | 3.16E-01 | 5.09E-01 | 2.69E+00 | 3.10E+00 | GTF2H2 |
| 317 | Formation of HIV-1 elongation complex containing HIV-1 Tat Homo sapiens R-HSA-167200 | 3.16E-01 | 5.09E-01 | 2.69E+00 | 3.10E+00 | GTF2H2 |
| 318 | Formation of Incision Complex in GG-NER Homo sapiens R-HSA-5696395 | 3.16E-01 | 5.09E-01 | 2.69E+00 | 3.10E+00 | GTF2H2 |
| 319 | Signaling by FGFR2 in disease Homo sapiens R-HSA-5655253 | 3.16E-01 | 5.09E-01 | 2.69E+00 | 3.10E+00 | FGF9 |
| 320 | Phospholipase C-mediated cascade; FGFR3 Homo sapiens R-HSA-5654227 | 3.22E-01 | 5.09E-01 | 2.63E+00 | 2.97E+00 | FGF9 |
| 321 | Intra-Golgi traffic Homo sapiens R-HSA-6811438 | 3.22E-01 | 5.09E-01 | 2.63E+00 | 2.97E+00 | MAN1A1 |
| 322 | Formation of HIV elongation complex in the absence of HIV Tat Homo sapiens R-HSA-167152 | 3.22E-01 | 5.09E-01 | 2.63E+00 | 2.97E+00 | GTF2H2 |
| 323 | RNA Polymerase II Transcription Elongation Homo sapiens R-HSA-75955 | 3.22E-01 | 5.09E-01 | 2.63E+00 | 2.97E+00 | GTF2H2 |
| 324 | Formation of RNA Pol II elongation complex Homo sapiens R-HSA-112382 | 3.22E-01 | 5.09E-01 | 2.63E+00 | 2.97E+00 | GTF2H2 |
| 325 | RNA Polymerase II HIV Promoter Escape Homo sapiens R-HSA-167162 | 3.22E-01 | 5.09E-01 | 2.63E+00 | 2.97E+00 | GTF2H2 |
| 326 | RNA Polymerase II Promoter Escape Homo sapiens R-HSA-73776 | 3.22E-01 | 5.09E-01 | 2.63E+00 | 2.97E+00 | GTF2H2 |
| 327 | RNA Polymerase II Transcription Initiation And Promoter Clearance Homo sapiens R-HSA-76042 | 3.22E-01 | 5.09E-01 | 2.63E+00 | 2.97E+00 | GTF2H2 |
| 328 | RNA Polymerase II Transcription Initiation Homo sapiens R-HSA-75953 | 3.22E-01 | 5.09E-01 | 2.63E+00 | 2.97E+00 | GTF2H2 |
| 329 | RNA Polymerase II Transcription Pre-Initiation And Promoter Opening Homo sapiens R-HSA-73779 | 3.22E-01 | 5.09E-01 | 2.63E+00 | 2.97E+00 | GTF2H2 |
| 330 | HIV Transcription Initiation Homo sapiens R-HSA-167161 | 3.22E-01 | 5.09E-01 | 2.63E+00 | 2.97E+00 | GTF2H2 |
| 331 | Diseases of metabolism Homo sapiens R-HSA-5668914 | 3.28E-01 | 5.14E-01 | 2.57E+00 | 2.86E+00 | POMC |

| | | | | | | |
|---|---|---|---|---|---|---|
| 332 | RMTs methylate histone arginines Homo sapiens R-HSA-3214858 | 3.28E-01 | 5.14E-01 | 2.57E+00 | 2.86E+00 | CCND1 |
| 333 | Signaling by ERBB2 Homo sapiens R-HSA-1227986 | 3.28E-01 | 5.14E-01 | 2.57E+00 | 2.86E+00 | NRG1 |
| 334 | Signal Transduction Homo sapiens R-HSA-162582 | 3.30E-01 | 5.15E-01 | 1.12E+00 | 1.25E+00 | RNF43;JAG1;PRKAA2;PTGER2;LPL;SMAD9;NRG1;PIK3CB;RASGRP1;THBS1;MYO9A;KNG1;POMC;RBP4;GRM5;ZNRF3;CCND1;FGF9;HEY1;AXL;PSME3;GNAQ;CTNNA1;NEFL |
| 335 | Phospholipase C-mediated cascade; FGFR4 Homo sapiens R-HSA-5654228 | 3.34E-01 | 5.20E-01 | 2.51E+00 | 2.75E+00 | FGF9 |
| 336 | Mitotic G1-G1/S phases Homo sapiens R-HSA-453279 | 3.37E-01 | 5.21E-01 | 1.69E+00 | 1.84E+00 | CCND1;PSME3 |
| 337 | Phospholipase C-mediated cascade: FGFR1 Homo sapiens R-HSA-5654219 | 3.40E-01 | 5.21E-01 | 2.46E+00 | 2.65E+00 | FGF9 |
| 338 | Defensins Homo sapiens R-HSA-1461973 | 3.40E-01 | 5.21E-01 | 2.46E+00 | 2.65E+00 | DEFB1 |
| 339 | RNA Polymerase I Transcription Initiation Homo sapiens R-HSA-73762 | 3.40E-01 | 5.21E-01 | 2.46E+00 | 2.65E+00 | GTF2H2 |
| 340 | NOTCH1 Intracellular Domain Regulates Transcription Homo sapiens R-HSA-2122947 | 3.40E-01 | 5.21E-01 | 2.46E+00 | 2.65E+00 | HEY1 |
| 341 | G beta:gamma signalling through PI3Kgamma Homo sapiens R-HSA-392451 | 3.46E-01 | 5.29E-01 | 2.40E+00 | 2.55E+00 | PIK3CB |
| 342 | Phospholipase C-mediated cascade; FGFR2 Homo sapiens R-HSA-5654221 | 3.52E-01 | 5.34E-01 | 2.35E+00 | 2.46E+00 | FGF9 |
| 343 | Regulation of activated PAK-2p34 by proteasome mediated degradation Homo sapiens R-HSA-211733 | 3.52E-01 | 5.34E-01 | 2.35E+00 | 2.46E+00 | PSME3 |
| 344 | G alpha (s) signalling events Homo sapiens R-HSA-418555 | 3.56E-01 | 5.34E-01 | 1.62E+00 | 1.67E+00 | POMC;PTGER2 |
| 345 | Chondroitin sulfate/dermatan sulfate metabolism Homo sapiens R-HSA-1793185 | 3.58E-01 | 5.34E-01 | 2.31E+00 | 2.37E+00 | BGN |
| 346 | Deadenylation-dependent mRNA decay Homo sapiens R-HSA-429914 | 3.58E-01 | 5.34E-01 | 2.31E+00 | 2.37E+00 | DDX6 |
| 347 | CDK-mediated phosphorylation and removal of Cdc6 Homo sapiens R-HSA-69017 | 3.58E-01 | 5.34E-01 | 2.31E+00 | 2.37E+00 | PSME3 |

| | | | | | | |
|---|---|---|---|---|---|---|
| 348 | Regulation of Apoptosis Homo sapiens R-HSA-169911 | 3.58E-01 | 5.34E-01 | 2.31E+00 | 2.37E+00 | PSME3 |
| 349 | Regulation of ornithine decarboxylase (ODC) Homo sapiens R-HSA-350562 | 3.58E-01 | 5.34E-01 | 2.31E+00 | 2.37E+00 | PSME3 |
| 350 | G-protein beta:gamma signalling Homo sapiens R-HSA-397795 | 3.63E-01 | 5.36E-01 | 2.26E+00 | 2.29E+00 | PIK3CB |
| 351 | Nuclear Envelope Breakdown Homo sapiens R-HSA-2980766 | 3.63E-01 | 5.36E-01 | 2.26E+00 | 2.29E+00 | LPIN1 |
| 352 | Vpu mediated degradation of CD4 Homo sapiens R-HSA-180534 | 3.63E-01 | 5.36E-01 | 2.26E+00 | 2.29E+00 | PSME3 |
| 353 | Autodegradation of the E3 ubiquitin ligase COP1 Homo sapiens R-HSA-349425 | 3.63E-01 | 5.36E-01 | 2.26E+00 | 2.29E+00 | PSME3 |
| 354 | p53-Independent DNA Damage Response Homo sapiens R-HSA-69610 | 3.69E-01 | 5.40E-01 | 2.22E+00 | 2.21E+00 | PSME3 |
| 355 | p53-Independent G1/S DNA damage checkpoint Homo sapiens R-HSA-69613 | 3.69E-01 | 5.40E-01 | 2.22E+00 | 2.21E+00 | PSME3 |
| 356 | Ubiquitin Mediated Degradation of Phosphorylated Cdc25A Homo sapiens R-HSA-69601 | 3.69E-01 | 5.40E-01 | 2.22E+00 | 2.21E+00 | PSME3 |
| 357 | GPVI-mediated activation cascade Homo sapiens R-HSA-114604 | 3.74E-01 | 5.42E-01 | 2.17E+00 | 2.13E+00 | PIK3CB |
| 358 | Formation of TC-NER Pre-Incision Complex Homo sapiens R-HSA-6781823 | 3.74E-01 | 5.42E-01 | 2.17E+00 | 2.13E+00 | GTF2H2 |
| 359 | Vif-mediated degradation of APOBEC3G Homo sapiens R-HSA-180585 | 3.74E-01 | 5.42E-01 | 2.17E+00 | 2.13E+00 | PSME3 |
| 360 | Heparan sulfate/heparin (HS-GAG) metabolism Homo sapiens R-HSA-1638091 | 3.80E-01 | 5.42E-01 | 2.13E+00 | 2.06E+00 | BGN |
| 361 | Golgi Associated Vesicle Biogenesis Homo sapiens R-HSA-432722 | 3.80E-01 | 5.42E-01 | 2.13E+00 | 2.06E+00 | TFRC |
| 362 | SCF-beta-TrCP mediated degradation of Emi1 Homo sapiens R-HSA-174113 | 3.80E-01 | 5.42E-01 | 2.13E+00 | 2.06E+00 | PSME3 |
| 363 | Stabilization of p53 Homo sapiens R-HSA-69541 | 3.80E-01 | 5.42E-01 | 2.13E+00 | 2.06E+00 | PSME3 |
| 364 | AUF1 (hnRNP D0) binds and destabilizes mRNA Homo sapiens R-HSA-450408 | 3.80E-01 | 5.42E-01 | 2.13E+00 | 2.06E+00 | PSME3 |

| | | | | | | |
|---|---|---|---|---|---|---|
| 365 | Degradation of AXIN Homo sapiens R-HSA-4641257 | 3.80E-01 | 5.42E-01 | 2.13E+00 | 2.06E+00 | PSME3 |
| 366 | Regulation of TP53 Activity Homo sapiens R-HSA-5633007 | 3.84E-01 | 5.47E-01 | 1.52E+00 | 1.45E+00 | PRKAA2;HIPK2 |
| 367 | Hh mutants that don't undergo autocatalytic processing are degraded by ERAD Homo sapiens R-HSA-5362768 | 3.85E-01 | 5.47E-01 | 2.09E+00 | 1.99E+00 | PSME3 |
| 368 | Degradation of DVL Homo sapiens R-HSA-4641258 | 3.91E-01 | 5.53E-01 | 2.05E+00 | 1.93E+00 | PSME3 |
| 369 | RNA Polymerase I Chain Elongation Homo sapiens R-HSA-73777 | 3.96E-01 | 5.56E-01 | 2.02E+00 | 1.87E+00 | GTF2H2 |
| 370 | Pre-NOTCH Expression and Processing Homo sapiens R-HSA-1912422 | 3.96E-01 | 5.56E-01 | 2.02E+00 | 1.87E+00 | CCND1 |
| 371 | Hh mutants abrogate ligand secretion Homo sapiens R-HSA-5387390 | 3.96E-01 | 5.56E-01 | 2.02E+00 | 1.87E+00 | PSME3 |
| 372 | NIK-->noncanonical NF-kB signaling Homo sapiens R-HSA-5676590 | 4.02E-01 | 5.59E-01 | 1.98E+00 | 1.81E+00 | PSME3 |
| 373 | CDT1 association with the CDC6:ORC:origin complex Homo sapiens R-HSA-68827 | 4.02E-01 | 5.59E-01 | 1.98E+00 | 1.81E+00 | PSME3 |
| 374 | Antigen processing: Ubiquitination & Proteasome degradation Homo sapiens R-HSA-983168 | 4.02E-01 | 5.59E-01 | 1.32E+00 | 1.20E+00 | RNF144B;UBA6;PSME3 |
| 375 | Cellular responses to stress Homo sapiens R-HSA-2262752 | 4.05E-01 | 5.59E-01 | 1.25E+00 | 1.13E+00 | PRKAA2;CITED2;ID1;HSPA1B |
| 376 | Degradation of GLI1 by the proteasome Homo sapiens R-HSA-5610780 | 4.07E-01 | 5.59E-01 | 1.95E+00 | 1.75E+00 | PSME3 |
| 377 | GLI3 is processed to GLI3R by the proteasome Homo sapiens R-HSA-5610785 | 4.07E-01 | 5.59E-01 | 1.95E+00 | 1.75E+00 | PSME3 |
| 378 | Degradation of GLI2 by the proteasome Homo sapiens R-HSA-5610783 | 4.07E-01 | 5.59E-01 | 1.95E+00 | 1.75E+00 | PSME3 |
| 379 | The role of GTSE1 in G2/M progression after G2 checkpoint Homo sapiens R-HSA-8852276 | 4.07E-01 | 5.59E-01 | 1.95E+00 | 1.75E+00 | PSME3 |
| 380 | Role of phospholipids in phagocytosis Homo sapiens R-HSA-2029485 | 4.12E-01 | 5.62E-01 | 1.91E+00 | 1.70E+00 | PIK3CB |
| 381 | PI Metabolism Homo sapiens R-HSA-1483255 | 4.12E-01 | 5.62E-01 | 1.91E+00 | 1.70E+00 | PIK3CB |

| | | | | | | |
|---|---|---|---|---|---|---|
| 382 | Translocation of GLUT4 to the plasma membrane Homo sapiens R-HSA-1445148 | 4.12E-01 | 5.62E-01 | 1.91E+00 | 1.70E+00 | PRKAA2 |
| 383 | Innate Immune System Homo sapiens R-HSA-168249 | 4.17E-01 | 5.63E-01 | 1.13E+00 | 9.92E-01 | FGF9;PSME3;NEFL;DEFB1;NRG1;BPI;PIK3CB;RASGRP1 |
| 384 | Cell-cell junction organization Homo sapiens R-HSA-421270 | 4.17E-01 | 5.63E-01 | 1.88E+00 | 1.65E+00 | CTNNA1 |
| 385 | TP53 Regulates Transcription of DNA Repair Genes Homo sapiens R-HSA-6796648 | 4.17E-01 | 5.63E-01 | 1.88E+00 | 1.65E+00 | GTF2H2 |
| 386 | Dectin-1 mediated noncanonical NF-kB signaling Homo sapiens R-HSA-5607761 | 4.17E-01 | 5.63E-01 | 1.88E+00 | 1.65E+00 | PSME3 |
| 387 | Cytochrome P450 - arranged by substrate type Homo sapiens R-HSA-211897 | 4.22E-01 | 5.65E-01 | 1.85E+00 | 1.60E+00 | POMC |
| 388 | Gap-filling DNA repair synthesis and ligation in TC-NER Homo sapiens R-HSA-6782210 | 4.22E-01 | 5.65E-01 | 1.85E+00 | 1.60E+00 | GTF2H2 |
| 389 | Autodegradation of Cdh1 by Cdh1:APC/C Homo sapiens R-HSA-174084 | 4.22E-01 | 5.65E-01 | 1.85E+00 | 1.60E+00 | PSME3 |
| 390 | Dual incision in TC-NER Homo sapiens R-HSA-6782135 | 4.27E-01 | 5.65E-01 | 1.82E+00 | 1.55E+00 | GTF2H2 |
| 391 | Signaling by FGFR in disease Homo sapiens R-HSA-1226099 | 4.27E-01 | 5.65E-01 | 1.82E+00 | 1.55E+00 | FGF9 |
| 392 | p53-Dependent G1/S DNA damage checkpoint Homo sapiens R-HSA-69580 | 4.27E-01 | 5.65E-01 | 1.82E+00 | 1.55E+00 | PSME3 |
| 393 | p53-Dependent G1 DNA Damage Response Homo sapiens R-HSA-69563 | 4.27E-01 | 5.65E-01 | 1.82E+00 | 1.55E+00 | PSME3 |
| 394 | Asymmetric localization of PCP proteins Homo sapiens R-HSA-4608870 | 4.27E-01 | 5.65E-01 | 1.82E+00 | 1.55E+00 | PSME3 |
| 395 | RNA Polymerase II Pre-transcription Events Homo sapiens R-HSA-674695 | 4.33E-01 | 5.69E-01 | 1.79E+00 | 1.50E+00 | GTF2H2 |
| 396 | Hedgehog ligand biogenesis Homo sapiens R-HSA-5358346 | 4.33E-01 | 5.69E-01 | 1.79E+00 | 1.50E+00 | PSME3 |
| 397 | ER-Phagosome pathway Homo sapiens R-HSA-1236974 | 4.38E-01 | 5.73E-01 | 1.76E+00 | 1.46E+00 | PSME3 |
| 398 | G1/S DNA Damage Checkpoints Homo sapiens R-HSA-69615 | 4.38E-01 | 5.73E-01 | 1.76E+00 | 1.46E+00 | PSME3 |
| 399 | APC/C:Cdc20 mediated degradation of Securin Homo sapiens R-HSA-174154 | 4.43E-01 | 5.76E-01 | 1.74E+00 | 1.42E+00 | PSME3 |

| | | | | | | |
|---|---|---|---|---|---|---|
| 400 | Activation of NF-kappaB in B cells Homo sapiens R-HSA-1169091 | 4.43E-01 | 5.76E-01 | 1.74E+00 | 1.42E+00 | PSME3 |
| 401 | Macroautophagy Homo sapiens R-HSA-1632852 | 4.47E-01 | 5.77E-01 | 1.71E+00 | 1.38E+00 | PRKAA2 |
| 402 | Transcription of the HIV genome Homo sapiens R-HSA-167172 | 4.47E-01 | 5.77E-01 | 1.71E+00 | 1.38E+00 | GTF2H2 |
| 403 | Regulation of RAS by GAPs Homo sapiens R-HSA-5658442 | 4.47E-01 | 5.77E-01 | 1.71E+00 | 1.38E+00 | PSME3 |
| 404 | Assembly of the pre-replicative complex Homo sapiens R-HSA-68867 | 4.47E-01 | 5.77E-01 | 1.71E+00 | 1.38E+00 | PSME3 |
| 405 | trans-Golgi Network Vesicle Budding Homo sapiens R-HSA-199992 | 4.62E-01 | 5.90E-01 | 1.64E+00 | 1.26E+00 | TFRC |
| 406 | Clathrin derived vesicle budding Homo sapiens R-HSA-421837 | 4.62E-01 | 5.90E-01 | 1.64E+00 | 1.26E+00 | TFRC |
| 407 | Switching of origins to a post-replicative state Homo sapiens R-HSA-69052 | 4.62E-01 | 5.90E-01 | 1.64E+00 | 1.26E+00 | PSME3 |
| 408 | Orc1 removal from chromatin Homo sapiens R-HSA-68949 | 4.62E-01 | 5.90E-01 | 1.64E+00 | 1.26E+00 | PSME3 |
| 409 | Lipid digestion, mobilization, and transport Homo sapiens R-HSA-73923 | 4.67E-01 | 5.92E-01 | 1.61E+00 | 1.23E+00 | LPL |
| 410 | APC/C:Cdh1 mediated degradation of Cdc20 and other APC/C:Cdh1 targeted proteins in late mitosis/early G1 Homo sapiens R-HSA-174178 | 4.67E-01 | 5.92E-01 | 1.61E+00 | 1.23E+00 | PSME3 |
| 411 | Cdc20:Phospho-APC/C mediated degradation of Cyclin A Homo sapiens R-HSA-174184 | 4.67E-01 | 5.92E-01 | 1.61E+00 | 1.23E+00 | PSME3 |
| 412 | Intra-Golgi and retrograde Golgi-to-ER traffic Homo sapiens R-HSA-6811442 | 4.69E-01 | 5.93E-01 | 1.28E+00 | 9.67E-01 | MAN1A1;BICD1 |
| 413 | APC:Cdc20 mediated degradation of cell cycle proteins prior to satisfation of the cell cycle checkpoint Homo sapiens R-HSA-179419 | 4.71E-01 | 5.93E-01 | 1.59E+00 | 1.20E+00 | PSME3 |
| 414 | Removal of licensing factors from origins Homo sapiens R-HSA-69300 | 4.71E-01 | 5.93E-01 | 1.59E+00 | 1.20E+00 | PSME3 |
| 415 | Axon guidance Homo sapiens R-HSA-422475 | 4.76E-01 | 5.96E-01 | 1.11E+00 | 8.23E-01 | FGF9;PSME3;NEFL;NRG1;RASGRP1 |

| | | | | | | |
|---|---|---|---|---|---|---|
| 416 | NoRC negatively regulates rRNA expression Homo sapiens R-HSA-427413 | 4.76E-01 | 5.96E-01 | 1.57E+00 | 1.16E+00 | GTF2H2 |
| 417 | APC/C:Cdc20 mediated degradation of mitotic proteins Homo sapiens R-HSA-176409 | 4.81E-01 | 6.01E-01 | 1.55E+00 | 1.13E+00 | PSME3 |
| 418 | Signaling by Wnt Homo sapiens R-HSA-195721 | 4.82E-01 | 6.01E-01 | 1.16E+00 | 8.46E-01 | RNF43;ZNRF3;PSME3 |
| 419 | Activation of APC/C and APC/C:Cdc20 mediated degradation of mitotic proteins Homo sapiens R-HSA-176814 | 4.85E-01 | 6.02E-01 | 1.53E+00 | 1.10E+00 | PSME3 |
| 420 | Regulation of DNA replication Homo sapiens R-HSA-69304 | 4.85E-01 | 6.02E-01 | 1.53E+00 | 1.10E+00 | PSME3 |
| 421 | Negative epigenetic regulation of rRNA expression Homo sapiens R-HSA-5250941 | 4.90E-01 | 6.03E-01 | 1.50E+00 | 1.07E+00 | GTF2H2 |
| 422 | RNA Polymerase I Promoter Clearance Homo sapiens R-HSA-73854 | 4.90E-01 | 6.03E-01 | 1.50E+00 | 1.07E+00 | GTF2H2 |
| 423 | Transcription-Coupled Nucleotide Excision Repair (TC-NER) Homo sapiens R-HSA-6781827 | 4.90E-01 | 6.03E-01 | 1.50E+00 | 1.07E+00 | GTF2H2 |
| 424 | RNA Polymerase I Transcription Homo sapiens R-HSA-73864 | 5.03E-01 | 6.17E-01 | 1.45E+00 | 9.93E-01 | GTF2H2 |
| 425 | Regulation of APC/C activators between G1/S and early anaphase Homo sapiens R-HSA-176408 | 5.03E-01 | 6.17E-01 | 1.45E+00 | 9.93E-01 | PSME3 |
| 426 | Membrane Trafficking Homo sapiens R-HSA-199991 | 5.07E-01 | 6.17E-01 | 1.08E+00 | 7.37E-01 | PRKAA2;TFRC;MAN1A1;BICD1 |
| 427 | Regulation of insulin secretion Homo sapiens R-HSA-422356 | 5.08E-01 | 6.17E-01 | 1.43E+00 | 9.68E-01 | GNAQ |
| 428 | Regulation of HSF1-mediated heat shock response Homo sapiens R-HSA-3371453 | 5.08E-01 | 6.17E-01 | 1.43E+00 | 9.68E-01 | HSPA1B |
| 429 | Peptide ligand-binding receptors Homo sapiens R-HSA-375276 | 5.08E-01 | 6.17E-01 | 1.18E+00 | 8.00E-01 | POMC;KNG1 |
| 430 | Peptide hormone metabolism Homo sapiens R-HSA-2980736 | 5.12E-01 | 6.18E-01 | 1.41E+00 | 9.44E-01 | POMC |
| 431 | Opioid Signalling Homo sapiens R-HSA-111885 | 5.12E-01 | 6.18E-01 | 1.41E+00 | 9.44E-01 | POMC |
| 432 | Global Genome Nucleotide Excision Repair (GG-NER) Homo sapiens R-HSA-5696399 | 5.16E-01 | 6.18E-01 | 1.39E+00 | 9.21E-01 | GTF2H2 |

| | | | | | | |
|---|---|---|---|---|---|---|
| 433 | Degradation of beta-catenin by the destruction complex Homo sapiens R-HSA-195253 | 5.16E-01 | 6.18E-01 | 1.39E+00 | 9.21E-01 | PSME3 |
| 434 | DNA Replication Pre-Initiation Homo sapiens R-HSA-69002 | 5.16E-01 | 6.18E-01 | 1.39E+00 | 9.21E-01 | PSME3 |
| 435 | M/G1 Transition Homo sapiens R-HSA-68874 | 5.16E-01 | 6.18E-01 | 1.39E+00 | 9.21E-01 | PSME3 |
| 436 | TP53 Regulates Metabolic Genes Homo sapiens R-HSA-5628897 | 5.25E-01 | 6.26E-01 | 1.36E+00 | 8.76E-01 | PRKAA2 |
| 437 | Metabolism of polyamines Homo sapiens R-HSA-351202 | 5.25E-01 | 6.26E-01 | 1.36E+00 | 8.76E-01 | PSME3 |
| 438 | Hedgehog 'on' state Homo sapiens R-HSA-5632684 | 5.29E-01 | 6.26E-01 | 1.34E+00 | 8.55E-01 | PSME3 |
| 439 | APC/C-mediated degradation of cell cycle proteins Homo sapiens R-HSA-174143 | 5.29E-01 | 6.26E-01 | 1.34E+00 | 8.55E-01 | PSME3 |
| 440 | Regulation of mitotic cell cycle Homo sapiens R-HSA-453276 | 5.29E-01 | 6.26E-01 | 1.34E+00 | 8.55E-01 | PSME3 |
| 441 | Regulation of mRNA stability by proteins that bind AU-rich elements Homo sapiens R-HSA-450531 | 5.33E-01 | 6.30E-01 | 1.33E+00 | 8.35E-01 | PSME3 |
| 442 | Platelet homeostasis Homo sapiens R-HSA-418346 | 5.37E-01 | 6.33E-01 | 1.31E+00 | 8.15E-01 | KCNMB2 |
| 443 | Class A/1 (Rhodopsin-like receptors) Homo sapiens R-HSA-373076 | 5.43E-01 | 6.39E-01 | 1.06E+00 | 6.45E-01 | POMC;PTGER2;KNG1 |
| 444 | Activation of anterior HOX genes in hindbrain development during early embryogenesis Homo sapiens R-HSA-5617472 | 5.45E-01 | 6.39E-01 | 1.28E+00 | 7.77E-01 | PBX1 |
| 445 | Activation of HOX genes during differentiation Homo sapiens R-HSA-5619507 | 5.45E-01 | 6.39E-01 | 1.28E+00 | 7.77E-01 | PBX1 |
| 446 | PCP/CE pathway Homo sapiens R-HSA-4086400 | 5.53E-01 | 6.46E-01 | 1.25E+00 | 7.41E-01 | PSME3 |
| 447 | GPCR ligand binding Homo sapiens R-HSA-500792 | 5.56E-01 | 6.47E-01 | 1.02E+00 | 5.97E-01 | POMC;GRM5;PTGER2;KNG1 |
| 448 | Phase 1 - Functionalization of compounds Homo sapiens R-HSA-211945 | 5.57E-01 | 6.47E-01 | 1.24E+00 | 7.24E-01 | POMC |
| 449 | MAPK6/MAPK4 signaling Homo sapiens R-HSA-5687128 | 5.57E-01 | 6.47E-01 | 1.24E+00 | 7.24E-01 | PSME3 |
| 450 | TNFR2 non-canonical NF-kB pathway Homo sapiens R-HSA-5668541 | 5.69E-01 | 6.59E-01 | 1.20E+00 | 6.76E-01 | PSME3 |

| | | | | | | |
|---|---|---|---|---|---|---|
| 451 | Cellular response to heat stress Homo sapiens R-HSA-3371556 | 5.73E-01 | 6.62E-01 | 1.19E+00 | 6.61E-01 | HSPA1B |
| 452 | Synthesis of DNA Homo sapiens R-HSA-69239 | 5.77E-01 | 6.65E-01 | 1.17E+00 | 6.47E-01 | PSME3 |
| 453 | HIV Infection Homo sapiens R-HSA-162906 | 5.84E-01 | 6.66E-01 | 1.02E+00 | 5.51E-01 | PSME3;GTF2H2 |
| 454 | Transport of inorganic cations/anions and amino acids/oligopeptides Homo sapiens R-HSA-425393 | 5.84E-01 | 6.66E-01 | 1.15E+00 | 6.19E-01 | SLC12A1 |
| 455 | Potassium Channels Homo sapiens R-HSA-1296071 | 5.84E-01 | 6.66E-01 | 1.15E+00 | 6.19E-01 | KCNMB2 |
| 456 | CLEC7A (Dectin-1) signaling Homo sapiens R-HSA-5607764 | 5.84E-01 | 6.66E-01 | 1.15E+00 | 6.19E-01 | PSME3 |
| 457 | Hedgehog 'off' state Homo sapiens R-HSA-5610787 | 5.84E-01 | 6.66E-01 | 1.15E+00 | 6.19E-01 | PSME3 |
| 458 | Signaling by GPCR Homo sapiens R-HSA-372790 | 5.90E-01 | 6.71E-01 | 9.64E-01 | 5.08E-01 | POMC;GRM5;FGF9;PSME3;GNAQ;PTGER2;NEFL;NRG1;PIK3CB;RASGRP1;KNG1 |
| 459 | Gene Silencing by RNA Homo sapiens R-HSA-211000 | 5.91E-01 | 6.71E-01 | 1.13E+00 | 5.92E-01 | DICER1 |
| 460 | Transcriptional Regulation by TP53 Homo sapiens R-HSA-3700989 | 5.94E-01 | 6.72E-01 | 9.79E-01 | 5.10E-01 | PRKAA2;GTF2H2;HIPK2 |
| 461 | MHC class II antigen presentation Homo sapiens R-HSA-2132295 | 5.99E-01 | 6.76E-01 | 1.10E+00 | 5.67E-01 | HLA-DOA |
| 462 | DNA Replication Homo sapiens R-HSA-69306 | 6.06E-01 | 6.83E-01 | 1.08E+00 | 5.43E-01 | PSME3 |
| 463 | Degradation of the extracellular matrix Homo sapiens R-HSA-1474228 | 6.09E-01 | 6.85E-01 | 1.07E+00 | 5.32E-01 | ADAMTS16 |
| 464 | Mitotic Prophase Homo sapiens R-HSA-68875 | 6.13E-01 | 6.88E-01 | 1.06E+00 | 5.21E-01 | LPIN1 |
| 465 | Nucleotide Excision Repair Homo sapiens R-HSA-5696398 | 6.16E-01 | 6.90E-01 | 1.05E+00 | 5.10E-01 | GTF2H2 |
| 466 | Golgi-to-ER retrograde transport Homo sapiens R-HSA-8856688 | 6.23E-01 | 6.96E-01 | 1.03E+00 | 4.89E-01 | BICD1 |
| 467 | Selenoamino acid metabolism Homo sapiens R-HSA-2408522 | 6.26E-01 | 6.97E-01 | 1.02E+00 | 4.79E-01 | GNMT |
| 468 | G alpha (i) signalling events Homo sapiens R-HSA-418594 | 6.26E-01 | 6.97E-01 | 9.46E-01 | 4.42E-01 | POMC;KNG1 |
| 469 | Vesicle-mediated transport Homo sapiens R-HSA-5653656 | 6.32E-01 | 7.02E-01 | 9.21E-01 | 4.23E-01 | PRKAA2;TFRC;MAN1A1;BICD1 |
| 470 | Epigenetic regulation of gene expression Homo sapiens R-HSA-212165 | 6.39E-01 | 7.09E-01 | 9.88E-01 | 4.42E-01 | GTF2H2 |

| | | | | | | |
|---|---|---|---|---|---|---|
| 471 | RNA Polymerase I, RNA Polymerase III, and Mitochondrial Transcription Homo sapiens R-HSA-504046 | 6.42E-01 | 7.11E-01 | 9.79E-01 | 4.33E-01 | GTF2H2 |
| 472 | Fcgamma receptor (FCGR) dependent phagocytosis Homo sapiens R-HSA-2029480 | 6.55E-01 | 7.23E-01 | 9.46E-01 | 4.01E-01 | PIK3CB |
| 473 | Glycosaminoglycan metabolism Homo sapiens R-HSA-1630316 | 6.58E-01 | 7.25E-01 | 9.38E-01 | 3.93E-01 | BGN |
| 474 | Rho GTPase cycle Homo sapiens R-HSA-194840 | 6.61E-01 | 7.25E-01 | 9.30E-01 | 3.85E-01 | MYO9A |
| 475 | Toll Like Receptor 4 (TLR4) Cascade Homo sapiens R-HSA-166016 | 6.61E-01 | 7.25E-01 | 9.30E-01 | 3.85E-01 | BPI |
| 476 | C-type lectin receptors (CLRs) Homo sapiens R-HSA-5621481 | 6.64E-01 | 7.27E-01 | 9.23E-01 | 3.78E-01 | PSME3 |
| 477 | RNA Polymerase II Transcription Homo sapiens R-HSA-73857 | 6.67E-01 | 7.28E-01 | 9.15E-01 | 3.71E-01 | GTF2H2 |
| 478 | Late Phase of HIV Life Cycle Homo sapiens R-HSA-162599 | 6.79E-01 | 7.40E-01 | 8.86E-01 | 3.44E-01 | GTF2H2 |
| 479 | Host Interactions of HIV factors Homo sapiens R-HSA-162909 | 6.81E-01 | 7.41E-01 | 8.79E-01 | 3.37E-01 | PSME3 |
| 480 | SLC-mediated transmembrane transport Homo sapiens R-HSA-425407 | 6.86E-01 | 7.43E-01 | 8.45E-01 | 3.19E-01 | SLC12A1;SLCO4C1 |
| 481 | M Phase Homo sapiens R-HSA-68886 | 6.86E-01 | 7.43E-01 | 8.45E-01 | 3.19E-01 | PSME3;LPIN1 |
| 482 | mRNA Splicing - Major Pathway Homo sapiens R-HSA-72163 | 6.95E-01 | 7.52E-01 | 8.46E-01 | 3.07E-01 | TRA2B |
| 483 | Cardiac conduction Homo sapiens R-HSA-5576891 | 6.98E-01 | 7.53E-01 | 8.40E-01 | 3.02E-01 | HIPK2 |
| 484 | Signaling by Hedgehog Homo sapiens R-HSA-5358351 | 7.01E-01 | 7.54E-01 | 8.33E-01 | 2.96E-01 | PSME3 |
| 485 | Toll-Like Receptors Cascades Homo sapiens R-HSA-168898 | 7.11E-01 | 7.64E-01 | 8.09E-01 | 2.76E-01 | BPI |
| 486 | HIV Life Cycle Homo sapiens R-HSA-162587 | 7.14E-01 | 7.65E-01 | 8.03E-01 | 2.71E-01 | GTF2H2 |
| 487 | Neurotransmitter Receptor Binding And Downstream Transmission In The Postsynaptic Cell Homo sapiens R-HSA-112314 | 7.16E-01 | 7.66E-01 | 7.98E-01 | 2.66E-01 | NEFL |
| 488 | mRNA Splicing Homo sapiens R-HSA-72172 | 7.21E-01 | 7.68E-01 | 7.86E-01 | 2.57E-01 | TRA2B |
| 489 | Beta-catenin independent WNT signaling Homo sapiens R-HSA-3858494 | 7.21E-01 | 7.68E-01 | 7.86E-01 | 2.57E-01 | PSME3 |

| | | | | | | |
|---|---|---|---|---|---|---|
| 490 | Generic Transcription Pathway Homo sapiens R-HSA-212436 | 7.24E-01 | 7.70E-01 | 8.33E-01 | 2.69E-01 | ZNF551;PRKAA2;ZNF429;GTF2H2;ZNF266;HIPK2 |
| 491 | Metabolism of proteins Homo sapiens R-HSA-392499 | 7.34E-01 | 7.79E-01 | 8.38E-01 | 2.59E-01 | ADAMTS16;POMC;ADAMTS15;SPON1;MAN1A1;GCNT4;THBS1;GALNT10 |
| 492 | G2/M Checkpoints Homo sapiens R-HSA-69481 | 7.36E-01 | 7.79E-01 | 7.55E-01 | 2.32E-01 | PSME3 |
| 493 | Neuronal System Homo sapiens R-HSA-112316 | 7.46E-01 | 7.88E-01 | 7.51E-01 | 2.20E-01 | KCNMB2;NEFL |
| 494 | Cellular Senescence Homo sapiens R-HSA-2559583 | 7.60E-01 | 8.01E-01 | 7.02E-01 | 1.92E-01 | ID1 |
| 495 | Transport to the Golgi and subsequent modification Homo sapiens R-HSA-948021 | 7.63E-01 | 8.01E-01 | 6.98E-01 | 1.89E-01 | MAN1A1 |
| 496 | Separation of Sister Chromatids Homo sapiens R-HSA-2467813 | 7.63E-01 | 8.01E-01 | 6.98E-01 | 1.89E-01 | PSME3 |
| 497 | Apoptosis Homo sapiens R-HSA-109581 | 7.65E-01 | 8.02E-01 | 6.94E-01 | 1.86E-01 | PSME3 |
| 498 | GPCR downstream signaling Homo sapiens R-HSA-388396 | 7.67E-01 | 8.03E-01 | 8.00E-01 | 2.12E-01 | POMC;GRM5;GNAQ;PTGER2;PIK3CB;RASGRP1;KNG1 |
| 499 | Programmed Cell Death Homo sapiens R-HSA-5357801 | 7.71E-01 | 8.05E-01 | 6.81E-01 | 1.77E-01 | PSME3 |
| 500 | Cell Cycle, Mitotic Homo sapiens R-HSA-69278 | 7.76E-01 | 8.09E-01 | 7.32E-01 | 1.85E-01 | CCND1;PSME3;LPIN1 |
| 501 | Organelle biogenesis and maintenance Homo sapiens R-HSA-1852241 | 7.84E-01 | 8.13E-01 | 6.92E-01 | 1.68E-01 | PRKAA2;PPARGC1A |
| 502 | Mitotic Anaphase Homo sapiens R-HSA-68882 | 7.85E-01 | 8.13E-01 | 6.53E-01 | 1.58E-01 | PSME3 |
| 503 | G2/M Transition Homo sapiens R-HSA-69275 | 7.85E-01 | 8.13E-01 | 6.53E-01 | 1.58E-01 | PSME3 |
| 504 | Mitotic Metaphase and Anaphase Homo sapiens R-HSA-2555396 | 7.87E-01 | 8.13E-01 | 6.49E-01 | 1.56E-01 | PSME3 |
| 505 | Mitotic G2-G2/M phases Homo sapiens R-HSA-453274 | 7.89E-01 | 8.13E-01 | 6.45E-01 | 1.53E-01 | PSME3 |
| 506 | Cell Cycle Checkpoints Homo sapiens R-HSA-69620 | 8.01E-01 | 8.25E-01 | 6.20E-01 | 1.37E-01 | PSME3 |
| 507 | Infectious disease Homo sapiens R-HSA-5663205 | 8.14E-01 | 8.36E-01 | 6.47E-01 | 1.33E-01 | PSME3;GTF2H2 |
| 508 | Processing of Capped Intron-Containing Pre-mRNA Homo sapiens R-HSA-72203 | 8.20E-01 | 8.41E-01 | 5.84E-01 | 1.16E-01 | TRA2B |
| 509 | Muscle contraction Homo sapiens R-HSA-397014 | 8.25E-01 | 8.44E-01 | 5.75E-01 | 1.11E-01 | HIPK2 |
| 510 | Biological oxidations Homo sapiens R-HSA-211859 | 8.29E-01 | 8.47E-01 | 5.66E-01 | 1.06E-01 | POMC |
| 511 | Signaling by Rho GTPases Homo sapiens R-HSA-194315 | 8.36E-01 | 8.53E-01 | 6.13E-01 | 1.09E-01 | CTNNA1;MYO9A |

| | | | | | | |
|---|---|---|---|---|---|---|
| 512 | Transmission across Chemical Synapses Homo sapiens R-HSA-112315 | 8.47E-01 | 8.62E-01 | 5.34E-01 | 8.89E-02 | NEFL |
| 513 | Chromatin organization Homo sapiens R-HSA-4839726 | 8.66E-01 | 8.78E-01 | 4.98E-01 | 7.17E-02 | CCND1 |
| 514 | Chromatin modifying enzymes Homo sapiens R-HSA-3247509 | 8.66E-01 | 8.78E-01 | 4.98E-01 | 7.17E-02 | CCND1 |
| 515 | Cell Cycle Homo sapiens R-HSA-1640170 | 8.78E-01 | 8.89E-01 | 5.93E-01 | 7.69E-02 | CCND1;PSME3;LPIN1 |
| 516 | RHO GTPase Effectors Homo sapiens R-HSA-195258 | 8.97E-01 | 9.05E-01 | 4.40E-01 | 4.81E-02 | CTNNA1 |
| 517 | Transmembrane transport of small molecules Homo sapiens R-HSA-382551 | 8.98E-01 | 9.05E-01 | 5.64E-01 | 6.08E-02 | TFRC;SLC12A1;SLCO4C1 |
| 518 | Asparagine N-linked glycosylation Homo sapiens R-HSA-446203 | 9.00E-01 | 9.05E-01 | 4.33E-01 | 4.56E-02 | MAN1A1 |
| 519 | Gene Expression Homo sapiens R-HSA-74160 | 9.16E-01 | 9.20E-01 | 6.76E-01 | 5.90E-02 | ZNF551;DDX6;PRKAA2;PSME3;ZNF429;TRA2B;DICER1;GTF2H2;ZNF266;HIPK2 |
| 520 | Metabolism of carbohydrates Homo sapiens R-HSA-71387 | 9.19E-01 | 9.21E-01 | 3.97E-01 | 3.37E-02 | BGN |
| 521 | DNA Repair Homo sapiens R-HSA-73894 | 9.21E-01 | 9.21E-01 | 3.93E-01 | 3.24E-02 | GTF2H2 |

**Pathways enriched by genes suppressed with SGLT2 inhibitor in descending thin limb cells**

| | Term | P value | Adjusted P value | Odds Ratio | Combined Score | Genes |
|---|---|---|---|---|---|---|
| 1 | Respiratory electron transport Homo sapiens R-HSA-611105 | 9.33E-33 | 1.33E-29 | 1.66E+01 | 1.22E+03 | COX7B;NDUFA13;NDUFA11;NDUFA12;COX4I1;NDUFA10;ETFA;ETFB;ECSIT;COX6A1;COX7C;UQCRFS1;COX8A;NDUFC2;NDUFC1;SDHC;SDHD;SDHA;SDHB;COX6B1;COX7A2L;NDUFS8;NDUFS7;SCO2;NDUFS6;NDUFS5;NDUFS4;NDUFS3;UQCRC1;NDUFS2;NDUFS1;TMEM126B;NDUFB9;COX19;NDUFB8;NDUFB7;NDUFB6;NDUFB10;COX16;NDUFB11;NDUFB5;NDUFB4;COX18;NDUFB3;NDUFB2;UQCR11;UQCR10;COX5B;COX5A;UQCRH;COX14;CYC1;NDUFV2;NDUFV1;NDUFA9;NDUFA8;SURF1;NDUFA7;TIMMDC1;NDUFA6;NDUFA5;NDUFA4;NDUFA3;NDUFA2;NDUFA1;COX6C;LRPPRC;UQCRQ;NDUFAF4;NDUFAB1;NDUFAF5;CYCS;COX20 |
| 2 | The citric acid (TCA) cycle and respiratory electron transport Homo sapiens R-HSA-1428517 | 1.59E-24 | 1.14E-21 | 5.45E+00 | 2.98E+02 | NDUFA13;COX7B;NDUFA11;NDUFA12;COX4I1;NDUFA10;ETFA;ETFB;COX6A1;ECSIT;COX7C;NNT;MPC2;IDH3B;PDK3;UQCRFS1;SLC16A3;PDK2;IDH3A;COX8A;NDUFC2;SDHC;NDUFC1;SDHD;SDHA;SDHB;COX6B1;COX7A2L;NDUFS8;NDUFS7;SCO2;NDUFS6;NDUFS5;NDUFS4;UQCRC1;NDUFS3;SUCLG2;NDUFS2;SUCLG1;NDUFS1;TMEM126B;HAGH;COX19;NDUFB9;NDUFB8;FH;NDUFB7;NDUFB10;COX16;NDUFB6;NDUFB11;NDUFB5;COX18;GLO1;NDUFB4;NDUFB3;NDUFB2;UQCR11;UQCR10;PDHB;COX5B;COX5A;UQCRH;LDHB;LDHA;UCP2;COX14;CYC1;NDUFV2;NDUFV1;NDUFA9;NDUFA8;SURF1;NDUFA7;PDHA1;TIMMDC1;NDUFA6;NDUFA5;MDH2;IDH3G;NDUFA4;NDUFA3;IDH2;NDUFA2;NDUFA1;COX6C;LRPPRC;SUCLA2;UQCRQ;NDUFAF4;NDUFAF5;NDUFAB1;CYCS;COX20 |
| 3 | Respiratory electron transport, ATP synthesis by chemiosmotic coupling, and heat production by uncoupling proteins. Homo sapiens R-HSA-163200 | 1.79E-23 | 8.55E-21 | 7.21E+00 | 3.77E+02 | NDUFA13;COX7B;NDUFA11;NDUFA12;COX4I1;NDUFA10;ETFA;ETFB;COX6A1;ECSIT;COX7C;UQCRFS1;COX8A;NDUFC2;SDHC;NDUFC1;SDHD;SDHA;SDHB;COX6B1;COX7A2L;NDUFS8;NDUFS7;SCO2;NDUFS6;NDUFS5;NDUFS4;UQCRC1;NDUFS3;NDUFS2;NDUFS1;TMEM126B;COX19;NDUFB9;NDUFB8;NDUFB7;NDUFB10;COX16;NDUFB6;NDUFB11;NDUFB5;COX18;NDUFB4;NDUFB3;NDUFB2;UQCR11;UQCR10;COX5B;COX5A;UQCRH;UCP2;COX14;CYC1;NDUFV2;NDUFV1;NDUFA9;NDUFA8;SURF1;NDUFA7;TIMMDC1;NDUFA6;NDUFA5;NDUFA4;NDUFA3;NDUFA2;NDUFA1;COX6C;LRPPRC;UQCRQ;NDUFAF4;NDUFAF5;NDUFAB1;CYCS;COX20 |

| | | | | | | |
|---|---|---|---|---|---|---|
| 4 | Infectious disease Homo sapiens R-HSA-5663205 | 4.07E-22 | 1.45E-19 | 2.96E+00 | 1.46E+02 | RPL5;RPL30;RPL3;GTF2B;RPL8;RPL10A;RPL6;PSMD8;RPS15;PSMD9;PSMD6;PSMD7;PSMD4;RPS19;RPL18A;PSMD2;PSMD3;AP1S2;PSMD1;RCC1;KPNA4;KPNA2;RPS10;KPNA3;RPS13;AP2M1;RPS12;SKP1;KPNA1;MAP2K3;SUPT16H;RPS9;MAP2K2;RPS8;RPS5;LIG4;RPSA;GTF2F2;RBX1;NUP93;PSME1;PSME2;CHMP4B;CHMP4A;RPL29;RPL28;PPIA;VPS28;PSMD10;PSMD12;TSG101;PSMD11;PSMD14;PSMD13;SUPT4H1;RPS4Y1;GTF2E2;TPR;UBC;ATP6V1H;PSMF1;AP1M2;XRCC6;TGFB1;SYT1;XRCC5;HMGA1;EIF2AK2;EEF2;CDK9;MSRA;CHMP2B;NMT1;CHMP2A;CALR;CALM1;MNAT1;GRSF1;RPS21;RPS24;RPS23;ARF1;HSP90AB1;RPLP1;CLTA;PSIP1;AP2A2;RPL7A;AP1G1;NELFCD;RAC1;HSP90AA1;PDCD6IP;NCBP1;VPS37D;VPS37B;PSMA5;DNAJC3;PSMA6;PSMA3;PSMA4;PSMA1;PSMA2;NUP50;CHMP3;SLC25A5;SLC25A4;HBEGF;CHMP5;SLC25A6;GTF2A1;GTF2A2;RPL10;VPS4B;VPS4A;FURIN;PSMA7;PSMB6;PSMB7;PSMB5;PSMB2;PSMB3;POLR2B;POLR2C;PSMB1;AP2S1;RPL14;POLR2E;FYN;RPL15;RPS2;POLR2H;RPL18;RPS27A;PAK2;POLR2J;POLR2K;RPL19;POLR2L;RANBP1;NPM1;TAF13;TXNRD1;TAF10;TAF11;RPL23A;AP2B1;GTF2H4;PSMB8;PSMC5;PSMC6;PSMC3;PSMC4;PSMC1;PSMC2;TCEA1;CD9;FAU;RAN;RPL26L1;HSPA1A |
| 5 | Mitochondrial translation elongation Homo sapiens R-HSA-5389840 | 4.67E-21 | 1.34E-18 | 8.50E+00 | 3.98E+02 | MRPS15;GFM1;MRPS16;MRPS12;MRPL38;MRPL39;MRPS10;MRPL36;MRPL37;MRPL34;MRPL32;MRPL33;MRPL41;MRPL4;MRPL42;MRPL3;MRPL2;MRPL9;CHCHD1;TSFM;MRPS26;MRPS27;MRPS24;MRPS25;MRPS22;MRPS18B;MRPL49;MRPS21;MRPL48;MRPS7;MRPL45;MRPS6;MRPL46;MRPS5;MRPL43;TUFM;MRPS18C;MRPL44;MRPL52;MRPS9;MRPL51;AURKAIP1;MRPS35;MRPS36;MRPL18;MRPL19;MRPS34;MRPL16;MRPL17;MRPL14;MRPS30;MRPL12;MRPL10;MRPL11;MRPL20;GADD45GIP1;MRPL27;MRPL28;MRPL21;MRPL22 |
| 6 | Mitochondrial translation Homo sapiens R-HSA-5368287 | 2.11E-20 | 5.03E-18 | 7.53E+00 | 3.41E+02 | MRPS15;GFM1;MRPS16;MRPS12;MRPL38;MRPL39;MRPS10;MRPL36;MRPL37;MRPL34;MRPL32;MRPL33;MRPL41;MRPL4;MRPL42;MRPL3;MRPL2;MRPL9;CHCHD1;TSFM;MRPS26;MRPS27;MRPS24;MRPS25;MRPS22;MRPS18B;MRPL49;MRPS21;MRPL48;MRPS7;MRPL45;MRPS6;MRPL46;MRPS5;MRPL43;TUFM;MRPS18C;MRPL44;MRPL52;MRPS9;MRPL51;AURKAIP1;MRPS35;MRPS36;MRPL18;MRPL19;MRPS34;MRPL16;MRPL17;MTIF2;MRPL14;MTIF3;MRPS30;MRPL12;MRPL10;MRPL11;MRPL20;GADD45GIP1;MRPL27;MRPL28;MRPL21;MRPL22 |

| | | | | | | |
|---|---|---|---|---|---|---|
| 7 | Mitochondrial translation initiation Homo sapiens R-HSA-5368286 | 3.84E-20 | 7.85E-18 | 8.02E+00 | 3.59E+02 | MRPS15;MRPS16;MRPS12;MRPL38;MRPL39;MRPS10;MRPL36;MRPL37;MRPL34;MRPL32;MRPL33;MRPL41;MRPL4;MRPL42;MRPL3;MRPL2;MRPL9;CHCHD1;MRPS26;MRPS27;MRPS24;MRPS25;MRPS22;MRPS18B;MRPL49;MRPS21;MRPL48;MRPS7;MRPL45;MRPS6;MRPL46;MRPS5;MRPL43;MRPS18C;MRPL44;MRPL52;MRPS9;MRPL51;AURKAIP1;MRPS35;MRPS36;MRPL18;MRPL19;MRPS34;MRPL16;MRPL17;MTIF2;MRPL14;MTIF3;MRPS30;MRPL12;MRPL10;MRPL11;MRPL20;GADD45GIP1;MRPL27;MRPL28;MRPL21;MRPL22 |
| 8 | Membrane Trafficking Homo sapiens R-HSA-199991 | 1.18E-19 | 1.89E-17 | 2.56E+00 | 1.11E+02 | APP;SCOC;SYS1;KDELR1;GCC2;RAB43;GOLGA2;TFG;KIF5C;AKT2;KIF5B;AP1S2;AKT1;ARFIP2;KIF21A;KIFAP3;VPS36;AP2M1;TPD52;NSF;COG6;COG2;ANK2;CHMP4B;KDELR2;CHMP4A;VPS25;STAM2;VPS28;COPB2;SPTBN5;RAB5C;TMED10;TSG101;STXBP3;SAR1B;COPB1;ARL1;KLC1;TGOLN2;BLOC1S6;SCFD1;SNX2;MAN2A2;KIF3A;MYO6;UBC;RAB6A;AP1M2;SNX5;TRAPPC3;HSPA8;RAB4A;SNAPIN;TRAPPC1;TRAPPC4;TRAPPC5;SURF4;M6PR;ASPSCR1;MYO5A;LNPEP;SNF8;TRAPPC9;STAM;AP4B1;CNIH1;TMEM115;GOLGB1;CHMP2B;CHMP2A;CALM1;COPE;PICALM;YWHAE;ARF4;ARF1;MCFD2;SERPINA1;YWHAB;CLTC;SNAP23;CTSZ;CLTB;PRKAG1;AP4E1;CLTA;PRKAG2;ARRB1;MIA3;CLINT1;PPP6C;AP1G2;YWHAQ;AP1G1;TMED3;TMED2;AP3S1;RAC1;TMED7;SPTAN1;RAB8A;YWHAG;YWHAH;TMED9;NAA30;COPZ2;VPS37D;PLA2G4A;AP3B1;VPS37B;ARFGAP3;YWHAZ;COPZ1;DNM2;VAMP8;TRAPPC6A;KIFC2;KIF2A;DNAJC6;MYH8;PLIN3;CHMP3;EXOC6;NAA38;DNASE2;SEC22B;PAFAH1B3;PAFAH1B2;RHOQ;VAMP3;PAFAH1B1;SEC22C;CHMP5;NAPA;DCTN6;NBAS;RALA;TMF1;DCTN2;RAB1B;VPS4B;GOSR2;USO1;VPS4A;DCTN3;NAPG;STX10;CYTH3;LMAN1;VPS51;LMAN2;VPS53;MAN1C1;STX5;STX4;CD59;RPS27A;CD55;CYTH1;DYNC1H1;RIC1;SEC24A;SORT1;TRAPPC10;GALNT1;DTNBP1;TPD52L1;RAB11A;ARCN1;RAB10;TJP1;RAB14;KIF26B;RAB13;RAB18;YIPF6;TRIP11;RAB9A |
| 9 | AUF1 (hnRNP D0) binds and destabilizes mRNA Homo sapiens R-HSA-450408 | 1.19E-19 | 1.89E-17 | 1.49E+01 | 6.50E+02 | PSMD10;PSMD12;PSMD11;PSMD14;PSMD13;HSPB1;PSMA7;PSMD8;PSMB6;PSMD9;PSMB7;PSMD6;PSMD7;PSMB5;PSMD4;PSMB2;PSMB3;PSMD2;PSMD3;UBC;PSMB1;PSMD1;PSMF1;RPS27A;HSPA8;PSMB8;PSMA5;PSMA6;PSMA3;PSMC5;PSMA4;PSMC6;PSMA1;PSMC3;PSMA2;PSMC4;PSMC1;PSMC2;PSME1;HNRNPD;PSME2;PABPC1;HSPA1A;EIF4G1 |

| | | | | | |
|---|---|---|---|---|---|
| 10 | Metabolism of proteins Homo sapiens R-HSA-392499 | 3.84E-19 | 5.50E-17 | 1.84E+00 | 7.81E+01 | EIF4A2;EIF4A1;RPL5;APP;RPL30;PGAP1;RPL3;CMAS;KDELR1;UBE2L6;RPL8;SMC3;ZDHHC2;RPL6;RPS15;GOLGA2;PCMT1;C1GALT1C1;HERC2;TFG;RPS19;RPL18A;SEC62;RPS10;RPS13;RPS12;ALG8;COG6;CSNK2A2;ALG5;ALG2;COG2;ALG3;MOGS;ANK2;HSPG2;NUP93;CSNK2B;XRN2;KDELR2;VDAC1;RPL29;TLN1;ATP6V0D1;ATF6;RPL28;ATF4;SPTBN5;RPN2;SAR1B;RPS4Y1;GNG10;SCFD1;GRPEL1;ARSJ;RAD21;NEU1;DPH3;ST3GAL4;MLEC;ST3GAL6;DPH7;EDEM3;CTSA;XBP1;PHC2;ICMT;ABCA3;FUCA1;EDEM2;H3F3A;MYO5A;RAB27A;CCT6A;MSRA;EIF5;HNRNPK;STAG2;GOLGB1;CALR;HNRNPC;FBXL5;RPS21;ITM2B;RPS24;RPS23;ARF4;ARF1;TRAM1;PIGT;SERPINA1;FKBP14;RPLP1;CETN2;PIGP;CTSZ;SRP54;MIA3;PIGX;HERPUD1;TRMT112;TUBA1C;TUBA1B;SERP1;SEC61G;ADAMTSL2;TMED3;CTSH;GNPNAT1;TMED2;MUC15;SEC61B;TMED7;LMCD1;CTSD;SPTAN1;CCT3;CCT2;TMED9;SSR4;PARP1;SSR2;SSR3;COPZ2;TIMM22;SRP9;COPZ1;RNF168;TCP1;PIGK;B3GNT3;B3GNT2;EXOC6;PABPC1;GAS6;PIGF;DCP2;PIGH;NAPA;SP100;PCNA;TOMM40;RPL10;USO1;GNAI3;TIMM13;DERL2;TIMM10;GNAI1;NAPG;GNAI2;EXOSC7;GGA1;LMAN1;EXOSC4;SAMM50;OS9;SRP72;LMAN2;LMNA;RPL14;SLC17A5;STX5;CD59;MGAT1;CCT8;RPL15;RPS2;RPS27A;EXOSC3;CCT7;RPL18;EXOSC1;CD55;CCT5;CCT4;RPL19;DYNC1H1;GSN;MTX1;MTX2;TRAPPC10;PCGF2;STAT3;C1GALT1;DOHH;TUBB4B;AGT;ARCN1;SPCS3;SPCS2;SPCS1;MSRB2;TOMM7;HDGF;FAU;TOMM5;TOMM6;DPAGT1;SRP19;SEMA5B;TUSC3;GALNT18;SRP14;RPL10A;TO [illegible] |
| 11 | Mitochondrial translation termination Homo sapiens R-HSA-5419276 | 2.20E-18 | 2.86E-16 | 7.17E+00 | 2.92E+02 | MRPS15;MRPS16;MRPS12;MRPL38;MRPL39;MRPS10;MRPL36;MRPL37;MRPL34;MRPL32;MRPL33;MRPL41;MRPL4;MRPL42;MRPL3;MRPL2;MRPL9;CHCHD1;MRPS26;MRPS27;MRPS24;MRPS25;MRPS22;MRPS18B;MRPL49;MRPS21;MRPL48;MRPS7;MRPL45;MRPS6;MRPL46;MRPS5;MRPL43;MRPS18C;MRPL44;MRPL52;MRPS9;MRPL51;AURKAIP1;MRPS35;MRPS36;MRPL18;MRPL19;MRPS34;MRPL16;MRPL17;MRPL14;MRPS30;MRPL12;MRPL10;MRPL11;MRPL20;GADD45GIP1;MRPL27;MRPL28;MRPL21;MRPL22 |
| 12 | Translation Homo sapiens R-HSA-72766 | 3.26E-18 | 3.89E-16 | 4.27E+00 | 1.72E+02 | EIF4A2;RPL5;SRP19;EIF4A1;RPL30;TRAM1;RPL3;RPLP1;SRP54;SRP14;RPL8;RPL10A;RPL6;EEF1B2;RPS15;TRMT112;RPL7A;RPS19;RPL18A;SEC61G;SEC61B;RPS10;RPS13;RPS12;EIF5B;RPS9;SSR4;RPS8;RPS5;SSR2;SSR3;RPSA;DDOST;SRP9;EEF1A1;EEF1G;EEF1D;EEF1A2;ETF1;PABPC1;RPL29;RPL28;RPN2;RPL10;RPS4Y1;SRP72;RPL14;EIF4H;RPL15;RPS2;RPL18;RPS27A;EIF4E;SEC11A;EIF4B;RPL19;SEC11C;EIF2B5;EIF2B4;EIF2B3;EIF2B2;RPL23A;EEF2;EIF2S2;EIF2S1;SPCS3;SPCS2;EIF5;SPCS1;EIF3K;EIF3I;EIF3J;EIF3G;SRPRB;EIF3H;FAU;EIF3F;RPS21;EIF3D;RPS24;RPL26L1;EIF3A;EIF4G1;RPS23 |

| | | | | | | |
|---|---|---|---|---|---|---|
| 13 | Complex I biogenesis Homo sapiens R-HSA-6799198 | 4.90E-18 | 5.39E-16 | 1.51E+01 | 6.00E+02 | NDUFB9;NDUFA13;NDUFB8;NDUFB7;NDUFA11;NDUFB6;NDUFB10;NDUFA12;NDUFB11;NDUFB5;NDUFB4;NDUFA10;NDUFB3;NDUFB2;ECSIT;NDUFV2;NDUFV1;NDUFA9;NDUFA8;NDUFA7;TIMMDC1;NDUFA6;NDUFA5;NDUFA3;NDUFA2;NDUFA1;NDUFC2;NDUFC1;NDUFS8;NDUFS7;NDUFS6;NDUFS5;NDUFAF4;NDUFS4;NDUFAB1;NDUFAF5;NDUFS3;NDUFS2;NDUFS1;TMEM126B |
| 14 | Hh mutants that don't undergo autocatalytic processing are degraded by ERAD Homo sapiens R-HSA-5362768 | 5.79E-18 | 5.92E-16 | 1.21E+01 | 4.82E+02 | PSMD10;PSMD12;VCP;PSMD11;PSMD14;PSMD13;SEL1L;DERL2;PSMA7;PSMD8;PSMB6;PSMD9;ERLEC1;PSMB7;PSMD6;PSMD7;PSMB5;PSMD4;OS9;PSMB2;PSMB3;PSMD2;PSMD3;UBC;PSMB1;PSMD1;PSMF1;RPS27A;PSMB8;PSMA5;PSMA6;PSMA3;PSMC5;PSMA4;PSMC6;PSMA1;PSMC3;PSMA2;PSMC4;PSMC1;PSMC2;PSME1;PSME2 |
| 15 | Regulation of mRNA stability by proteins that bind AU-rich elements Homo sapiens R-HSA-450531 | 1.19E-17 | 1.14E-15 | 6.68E+00 | 2.60E+02 | YWHAB;HSPB1;ELAVL1;PSMD8;PSMD9;PSMD6;PSMD7;PSMD4;PSMD2;PSMD3;AKT1;PSMD1;TNPO1;YWHAZ;PSMA5;PSMA6;PSMA3;PSMA4;XRN1;PSMA1;PSMA2;PSME1;PSME2;PABPC1;DCP2;PSMD10;PSMD12;PSMD11;SET;PSMD14;ANP32A;PSMD13;PSMA7;EXOSC7;PSMB6;PSMB7;EXOSC4;PSMB5;PSMB2;PSMB3;UBC;PSMB1;PSMF1;EXOSC3;RPS27A;EXOSC1;HSPA8;PSMB8;PSMC5;PSMC6;PSMC3;PSMC4;PSMC1;PSMC2;HNRNPD;HSPA1A;EIF4G1 |
| 16 | Autodegradation of Cdh1 by Cdh1:APC/C Homo sapiens R-HSA-174084 | 1.55E-17 | 1.39E-15 | 9.75E+00 | 3.77E+02 | PSMD10;PSMD12;PSMD11;ANAPC15;PSMD14;ANAPC16;PSMD13;UBE2D1;PSMA7;ANAPC11;PSMD8;PSMB6;PSMD9;PSMB7;PSMD6;PSMD7;PSMB5;PSMD4;PSMB2;PSMB3;CDC26;PSMD2;PSMD3;CDC27;UBC;PSMB1;PSMD1;PSMF1;RPS27A;UBE2E1;PSMB8;PSMA5;PSMA6;PSMA3;PSMC5;PSMA4;PSMC6;PSMA1;PSMC3;PSMA2;PSMC4;PSMC1;PSMC2;PSME1;PSME2;ANAPC5 |

| | | | | | | |
|---|---|---|---|---|---|---|
| 17 | Gene Expression Homo sapiens R-HSA-74160 | 3.41E-17 | 2.87E-15 | 1.62E+00 | 6.15E+01 | POP5;RPL5;RPL30;RPL3;HNRNPU;EHMT1;HNRNPR;PHAX;EPRS;RBPJ;RPL8;TSEN54;RPL6;GLS;RRP9;LSM10;PSMD8;PSMD9;PSMD6;PSMD7;PSMD4;CLP1;AKT2;PSMD2;PSMD3;AKT1;PSMD1;CSNK2A2;MED8;MED4;NUP93;DDX39A;SCO2;SARNP;DDIT4;PSME1;PSME2;RPL29;RPL28;SLBP;SET;MEAF6;RPN2;MAPKAP1;RPS4Y1;PRDX2;PRDX5;PRDX1;NRBF2;PSMF1;ZNF143;PRELID1;H3F3A;UPF3A;NR1D1;NR2F6;HNRNPL;HNRNPM;HNRNPK;EIF5;QARS;HNRNPF;AGO2;HNRNPD;RAD17;HNRNPC;FARSA;GPI;TRMU;RARG;CDKN1B;ZNF250;PHF20;SETD9;PHF1;RPLP1;PRKAG1;SRP54;PRKAG2;CHM;WDR46;WDR43;SMG5;BBC3;TRMT112;MED10;NELFCD;RIOK1;JARID2;TEAD2;CNOT6L;SSR4;SSR2;SSR3;MED27;SIRT1;SRP9;NCOR2;PSMA5;KAT2B;PSMA6;PSMA3;MED24;NCOR1;PSMA4;PSMA1;PSMA2;RARB;GARS;ZNF599;SF3B4;SF3B5;SF3B2;RPL10;URM1;SF3B6;SRRT;RPS27L;PSMA7;PSMB6;PSMB7;PSMB5;PSMB2;SRP72;PSMB3;PSMB1;RPL14;RRP36;HARS;RPL15;RPS2;BID;RPS27A;RPL18;BRD7;PAIP1;RPL19;SF3A3;RIOK3;WDR18;NR1H2;MBD2;GSR;HIPK1;GTF2H4;PSMB8;LRPPRC;PSMC5;PHF5A;PSMC6;PSMC3;PSMC4;NFIC;PSMC1;PSMC2;TRMT6;LAMTOR1;LAMTOR4;LAMTOR3;HSPA1A;LAMTOR5;CRCP;PDCD7;GTF2B;UBE2D3;UBE2D1;RORA;RPL10A;ZNF568;ZNF567;TXNL4A;SNRNP35;BNIP3L;SUPT16H;USP7;RPS9;RPS8;RPS5;USP2;GTF3A;SARS;THOC3;RPSA;ZCRB1;THOC2;POLRMT;THOC5;GTF2F2;NRBP1;MSH2;SRSF2;SNRPG;NHP2;SNRNP27;SRSF4;MAPKAPK5;SNRNP25;ZNF675;SNRPA;SRSF9;SNRPB;DNMT1;GTF2[illegible] |
| 18 | SCF(Skp2)-mediated degradation of p27/p21 Homo sapiens R-HSA-187577 | 5.63E-17 | 4.48E-15 | 9.95E+00 | 3.72E+02 | PSMD10;PSMD12;CDKN1B;PSMD11;PSMD14;PSMD13;CUL1;PSMA7;CKS1B;PSMD8;PSMB6;PSMD9;PSMB7;PSMD6;PSMD7;PSMB5;PSMD4;PSMB2;PSMB3;PSMD2;PSMD3;UBC;PSMB1;PSMD1;PSMF1;RPS27A;SKP1;PSMB8;PSMA5;PSMA6;PSMA3;PSMC5;PSMA4;PSMC6;PSMA1;PSMC3;PSMA2;PSMC4;CDK4;PSMC1;PSMC2;CDK2;PSME1;PSME2 |
| 19 | Hh mutants abrogate ligand secretion Homo sapiens R-HSA-5387390 | 6.14E-17 | 4.63E-15 | 1.04E+01 | 3.89E+02 | PSMD10;PSMD12;VCP;PSMD11;PSMD14;PSMD13;SEL1L;DERL2;PSMA7;PSMD8;PSMB6;PSMD9;ERLEC1;PSMB7;PSMD6;PSMD7;PSMB5;PSMD4;OS9;PSMB2;PSMB3;PSMD2;PSMD3;UBC;PSMB1;PSMD1;PSMF1;RPS27A;PSMB8;PSMA5;PSMA6;PSMA3;PSMC5;PSMA4;PSMC6;PSMA1;PSMC3;PSMA2;PSMC4;PSMC1;PSMC2;PSME1;PSME2 |
| 20 | Regulation of activated PAK-2p34 by proteasome mediated degradation Homo sapiens R-HSA-211733 | 6.70E-17 | 4.79E-15 | 1.32E+01 | 4.92E+02 | PSMD10;PSMD12;PSMD11;PSMD14;PSMD13;PSMA7;PSMD8;PSMB6;PSMD9;PSMB7;PSMD6;PSMD7;PSMB5;PSMD4;PSMB2;PSMB3;PSMD2;PSMD3;UBC;PSMB1;PSMD1;PSMF1;RPS27A;PAK2;PSMB8;PSMA5;PSMA6;PSMA3;PSMC5;PSMA4;PSMC6;PSMA1;PSMC3;PSMA2;PSMC4;PSMC1;PSMC2;PSME1;PSME2 |

| | | | | | | |
|---|---|---|---|---|---|---|
| 21 | HIV Infection Homo sapiens R-HSA-162906 | 1.04E-16 | 7.11E-15 | 3.18E+00 | 1.17E+02 | GTF2B;PSMD8;PSMD9;PSMD6;PSMD7;PSMD4;PSMD2;PSMD3;AP1S2;PSMD1;RCC1;AP2M1;SKP1;KPNA1;SUPT16H;LIG4;GTF2F2;RBX1;NUP93;PSME1;PSME2;CHMP4B;CHMP4A;PPIA;VPS28;PSMD10;PSMD12;TSG101;PSMD11;PSMD14;PSMD13;SUPT4H1;GTF2E2;TPR;UBC;ATP6V1H;PSMF1;AP1M2;XRCC6;XRCC5;HMGA1;CDK9;CHMP2B;NMT1;CHMP2A;MNAT1;ARF1;PSIP1;AP2A2;AP1G1;NELFCD;RAC1;PDCD6IP;NCBP1;VPS37D;VPS37B;PSMA5;PSMA6;PSMA3;PSMA4;PSMA1;PSMA2;NUP50;CHMP3;SLC25A5;SLC25A4;CHMP5;SLC25A6;GTF2A1;GTF2A2;VPS4B;VPS4A;FURIN;PSMA7;PSMB6;PSMB7;PSMB5;PSMB2;PSMB3;POLR2B;POLR2C;PSMB1;AP2S1;POLR2E;FYN;POLR2H;RPS27A;PAK2;POLR2J;POLR2K;POLR2L;RANBP1;NPM1;TAF13;TAF10;TAF11;AP2B1;GTF2H4;PSMB8;PSMC5;PSMC6;PSMC3;PSMC4;PSMC1;PSMC2;TCEA1;RAN |
| 22 | Hedgehog ligand biogenesis Homo sapiens R-HSA-5358346 | 1.23E-16 | 8.03E-15 | 8.67E+00 | 3.17E+02 | PSMD10;PSMD12;VCP;PSMD11;PSMD14;PSMD13;SEL1L;DERL2;PSMA7;PSMD8;PSMB6;PSMD9;ERLEC1;PSMB7;PSMD6;PSMD7;PSMB5;PSMD4;OS9;PSMB2;PSMB3;PSMD2;PSMD3;UBC;PSMB1;PSMD1;GPC5;PSMF1;RPS27A;PSMB8;PSMA5;PSMA6;PSMA3;PSMC5;PSMA4;PSMC6;PSMA1;PSMC3;PSMA2;PSMC4;PSMC1;PSMC2;PSME1;PSME2;P4HB;NOTUM |
| 23 | CDK-mediated phosphorylation and removal of Cdc6 Homo sapiens R-HSA-69017 | 2.37E-16 | 1.30E-14 | 1.20E+01 | 4.32E+02 | PSMD10;PSMD12;PSMD11;PSMD14;PSMD13;PSMA7;PSMD8;PSMB6;PSMD9;PSMB7;PSMD6;PSMD7;PSMB5;PSMD4;PSMB2;PSMB3;PSMD2;PSMD3;UBC;PSMB1;PSMD1;PSMF1;RPS27A;PSMB8;PSMA5;PSMA6;PSMA3;PSMC5;PSMA4;PSMC6;PSMA1;PSMC3;PSMA2;PSMC4;PSMC1;PSMC2;CDK2;PSME1;PSME2 |
| 24 | Ubiquitin-dependent degradation of Cyclin D1 Homo sapiens R-HSA-69229 | 2.37E-16 | 1.30E-14 | 1.20E+01 | 4.32E+02 | PSMD10;PSMD12;PSMD11;PSMD14;PSMD13;PSMA7;PSMD8;PSMB6;PSMD9;PSMB7;PSMD6;PSMD7;PSMB5;PSMD4;PSMB2;PSMB3;PSMD2;PSMD3;UBC;PSMB1;PSMD1;PSMF1;RPS27A;PSMB8;PSMA5;PSMA6;PSMA3;PSMC5;PSMA4;PSMC6;PSMA1;PSMC3;PSMA2;PSMC4;CDK4;PSMC1;PSMC2;PSME1;PSME2 |
| 25 | Ubiquitin-dependent degradation of Cyclin D Homo sapiens R-HSA-75815 | 2.37E-16 | 1.30E-14 | 1.20E+01 | 4.32E+02 | PSMD10;PSMD12;PSMD11;PSMD14;PSMD13;PSMA7;PSMD8;PSMB6;PSMD9;PSMB7;PSMD6;PSMD7;PSMB5;PSMD4;PSMB2;PSMB3;PSMD2;PSMD3;UBC;PSMB1;PSMD1;PSMF1;RPS27A;PSMB8;PSMA5;PSMA6;PSMA3;PSMC5;PSMA4;PSMC6;PSMA1;PSMC3;PSMA2;PSMC4;CDK4;PSMC1;PSMC2;PSME1;PSME2 |

| | | | | | | |
|---|---|---|---|---|---|---|
| 26 | Regulation of Apoptosis Homo sapiens R-HSA-169911 | 2.37E-16 | 1.30E-14 | 1.20E+01 | 4.32E+02 | PSMD10;PSMD12;PSMD11;PSMD14;PSMD13;PSMA7;PSMD8;PSMB6;PSMD9;PSMB7;PSMD6;PSMD7;PSMB5;PSMD4;PSMB2;PSMB3;PSMD2;PSMD3;UBC;PSMB1;PSMD1;PSMF1;RPS27A;PAK2;PSMB8;PSMA5;PSMA6;PSMA3;PSMC5;PSMA4;PSMC6;PSMA1;PSMC3;PSMA2;PSMC4;PSMC1;PSMC2;PSME1;PSME2 |
| 27 | ER-Phagosome pathway Homo sapiens R-HSA-1236974 | 3.28E-16 | 1.74E-14 | 8.21E+00 | 2.93E+02 | PSMD10;PSMD12;PSMD11;PSMD14;PSMD13;PSMA7;PSMD8;PSMB6;PSMD9;PSMB7;PSMD6;PSMD7;PSMB5;PSMD4;PSMB2;PSMB3;SEC61G;PSMD2;PSMD3;UBC;PSMB1;PSMD1;SEC61B;PSMF1;RPS27A;PDIA3;TAP1;HLA-C;HLA-G;PSMB8;HLA-E;PSMA5;PSMA6;PSMA3;PSMC5;PSMA4;PSMC6;PSMA1;PSMC3;PSMA2;PSMC4;PSMC1;PSMC2;PSME1;PSME2;CALR |
| 28 | GLI3 is processed to GLI3R by the proteasome Homo sapiens R-HSA-5610785 | 5.30E-16 | 2.53E-14 | 9.11E+00 | 3.20E+02 | PSMD10;GSK3B;PSMD12;PSMD11;PSMD14;PSMD13;CUL1;PSMA7;PSMD8;PSMB6;PSMD9;PSMB7;PSMD6;PSMD7;PSMB5;PSMD4;PSMB2;PSMB3;PSMD2;PSMD3;UBC;PSMB1;PSMD1;PSMF1;RPS27A;SKP1;CSNK1A1;PSMB8;RBX1;PSMA5;PSMA6;PSMA3;PSMC5;PSMA4;PSMC6;PSMA1;PSMC3;PSMA2;PSMC4;PSMC1;PSMC2;PSME1;PSME2 |
| 29 | Degradation of GLI2 by the proteasome Homo sapiens R-HSA-5610783 | 5.30E-16 | 2.53E-14 | 9.11E+00 | 3.20E+02 | PSMD10;GSK3B;PSMD12;PSMD11;PSMD14;PSMD13;CUL1;PSMA7;PSMD8;PSMB6;PSMD9;PSMB7;PSMD6;PSMD7;PSMB5;PSMD4;PSMB2;PSMB3;PSMD2;PSMD3;UBC;PSMB1;PSMD1;PSMF1;RPS27A;SKP1;CSNK1A1;PSMB8;RBX1;PSMA5;PSMA6;PSMA3;PSMC5;PSMA4;PSMC6;PSMA1;PSMC3;PSMA2;PSMC4;PSMC1;PSMC2;PSME1;PSME2 |
| 30 | The role of GTSE1 in G2/M progression after G2 checkpoint Homo sapiens R-HSA-8852276 | 5.30E-16 | 2.53E-14 | 9.11E+00 | 3.20E+02 | PSMD10;PSMD12;HSP90AB1;PSMD11;PSMD14;PSMD13;PSMA7;PSMD8;PSMB6;PSMD9;PSMB7;PSMD6;CCNB1;PSMD7;PSMB5;PSMD4;PSMB2;PSMB3;PSMD2;PSMD3;UBC;PSMB1;PSMD1;PSMF1;RPS27A;HSP90AA1;FKBPL;PSMB8;PSMA5;PSMA6;PSMA3;PSMC5;PSMA4;PSMC6;PSMA1;PSMC3;PSMA2;PSMC4;PSMC1;PSMC2;PSME1;PSME2;MAPRE1 |
| 31 | Vpu mediated degradation of CD4 Homo sapiens R-HSA-180534 | 7.84E-16 | 3.62E-14 | 1.10E+01 | 3.83E+02 | PSMD10;PSMD12;PSMD11;PSMD14;PSMD13;PSMA7;PSMD8;PSMB6;PSMD9;PSMB7;PSMD6;PSMD7;PSMB5;PSMD4;PSMB2;PSMB3;PSMD2;PSMD3;UBC;PSMB1;PSMD1;PSMF1;RPS27A;SKP1;PSMB8;PSMA5;PSMA6;PSMA3;PSMC5;PSMA4;PSMC6;PSMA1;PSMC3;PSMA2;PSMC4;PSMC1;PSMC2;PSME1;PSME2 |

| | | | | | | |
|---|---|---|---|---|---|---|
| 32 | APC/C:Cdc20 mediated degradation of Securin Homo sapiens R-HSA-174154 | 8.43E-16 | 3.77E-14 | 7.80E+00 | 2.71E+02 | PSMD10;PSMD12;PSMD11;ANAPC15;PSMD14;ANAPC16;PSMD13;UBE2D1;PSMA7;ANAPC11;PSMD8;PSMB6;PSMD9;PSMB7;PSMD6;PSMD7;PSMB5;PSMD4;PSMB2;PSMB3;CDC26;PSMD2;PSMD3;CDC27;UBC;PSMB1;PSMD1;PSMF1;RPS27A;UBE2E1;PSMB8;PSMA5;PSMA6;PSMA3;PSMC5;PSMA4;PSMC6;PSMA1;PSMC3;PSMA2;PSMC4;PSMC1;PSMC2;PSME1;PSME2;ANAPC5 |
| 33 | Cyclin A:Cdk2-associated events at S phase entry Homo sapiens R-HSA-69656 | 1.68E-15 | 7.16E-14 | 7.24E+00 | 2.46E+02 | RB1;PSMD10;PSMD12;CDKN1B;PSMD11;PSMD14;PSMD13;CUL1;PSMA7;CKS1B;PSMD8;PSMB6;PSMD9;PSMB7;PSMD6;PSMD7;PSMB5;PSMD4;PSMB2;PSMB3;PSMD2;PSMD3;UBC;PSMB1;PSMD1;PSMF1;RPS27A;SKP1;PSMB8;CDC25B;PSMA5;PSMA6;PSMA3;PSMC5;PSMA4;PSMC6;PSMA1;PSMC3;PSMA2;PSMC4;CDK4;PSMC1;PSMC2;CDK2;PSME1;PSME2;MNAT1 |
| 34 | NIK-->noncanonical NF-kB signaling Homo sapiens R-HSA-5676590 | 1.70E-15 | 7.16E-14 | 8.90E+00 | 3.03E+02 | PSMD10;PSMD12;PSMD11;PSMD14;PSMD13;CUL1;PSMA7;PSMD8;PSMB6;PSMD9;PSMB7;PSMD6;PSMD7;PSMB5;PSMD4;PSMB2;PSMB3;PSMD2;PSMD3;UBC;PSMB1;PSMD1;PSMF1;RPS27A;SKP1;FBXW11;PSMB8;PSMA5;PSMA6;PSMA3;PSMC5;PSMA4;PSMC6;PSMA1;PSMC3;PSMA2;PSMC4;PSMC1;PSMC2;PSME1;PSME2;UBE2M |
| 35 | Degradation of DVL Homo sapiens R-HSA-4641258 | 1.95E-15 | 7.99E-14 | 9.26E+00 | 3.14E+02 | PSMD10;PSMD12;PSMD11;PSMD14;PSMD13;CUL3;PSMA7;PSMD8;PSMB6;PSMD9;PSMB7;PSMD6;PSMD7;PSMB5;PSMD4;PSMB2;PSMB3;PSMD2;PSMD3;UBC;PSMB1;PSMD1;DVL3;PSMF1;RPS27A;PSMB8;RBX1;PSMA5;PSMA6;PSMA3;PSMC5;PSMA4;PSMC6;PSMA1;PSMC3;PSMA2;PSMC4;PSMC1;PSMC2;PSME1;PSME2 |
| 36 | SCF-beta-TrCP mediated degradation of Emi1 Homo sapiens R-HSA-174113 | 2.21E-15 | 8.53E-14 | 9.68E+00 | 3.27E+02 | PSMD10;PSMD12;PSMD11;PSMD14;PSMD13;CUL1;PSMA7;PSMD8;PSMB6;PSMD9;PSMB7;PSMD6;PSMD7;PSMB5;PSMD4;PSMB2;PSMB3;PSMD2;PSMD3;UBC;PSMB1;PSMD1;PSMF1;RPS27A;SKP1;PSMB8;PSMA5;PSMA6;PSMA3;PSMC5;PSMA4;PSMC6;PSMA1;PSMC3;PSMA2;PSMC4;PSMC1;PSMC2;PSME1;PSME2 |
| 37 | Degradation of AXIN Homo sapiens R-HSA-4641257 | 2.21E-15 | 8.53E-14 | 9.68E+00 | 3.27E+02 | PSMD10;PSMD12;PSMD11;PSMD14;PSMD13;PSMA7;PSMD8;PSMB6;PSMD9;PSMB7;PSMD6;PSMD7;PSMB5;PSMD4;TNKS2;PSMB2;PSMB3;PSMD2;PSMD3;UBC;PSMB1;PSMD1;PSMF1;RPS27A;SMURF2;PSMB8;PSMA5;PSMA6;PSMA3;PSMC5;PSMA4;PSMC6;PSMA1;PSMC3;PSMA2;PSMC4;PSMC1;PSMC2;PSME1;PSME2 |

| | | | | | | |
|---|---|---|---|---|---|---|
| 38 | Regulation of APC/C activators between G1/S and early anaphase Homo sapiens R-HSA-176408 | 3.16E-15 | 1.19E-13 | 6.18E+00 | 2.06E+02 | UBE2D1;PSMD8;PSMD9;PSMD6;PSMD7;PSMD4;CDC26;PSMD2;PSMD3;CDC27;PSMD1;SKP1;UBE2E1;PSMA5;PSMA6;PSMA3;PSMA4;PSMA1;PSMA2;PSME1;PSME2;ANAPC5;PSMD10;PSMD12;PSMD11;ANAPC15;PSMD14;ANAPC16;PSMD13;CUL1;PSMA7;ANAPC11;PSMB6;PSMB7;CCNB1;PSMB5;PSMB2;PSMB3;UBC;PSMB1;PSMF1;BUB3;RPS27A;PSMB8;PSMC5;PSMC6;PSMC3;PSMC4;PSMC1;PSMC2;CDK2 |
| 39 | Organelle biogenesis and maintenance Homo sapiens R-HSA-1852241 | 3.59E-15 | 1.32E-13 | 2.51E+00 | 8.36E+01 | KIFAP3;CHCHD1;MRPS18B;IFT80;CYS1;POLRMT;TUBA4A;MRPS18C;TUFM;AKAP9;TFAM;AURKAIP1;UNC119B;MRPL18;ARL3;MRPL19;MRPL16;MRPL17;MRPL14;MRPL12;MRPL10;MRPL11;MRPL20;PRKAR2B;KIF3A;INPP5E;NPHP1;TCTEX1D2;ESRRA;MRPL27;MRPL28;NR1D1;MRPL21;MRPL22;IFT20;AHI1;IFT22;WDR60;CYCS;DYNLRB1;CEP57;YWHAE;ARF4;MRPS15;GFM1;MRPS16;ALAS1;CETN2;MRPS12;MRPL38;PRKAG1;MRPL39;MRPS10;PRKAG2;MRPL36;MRPL37;TFB1M;MRPL34;PKD2;PKD1;MRPL32;HSPB11;MRPL33;MRPL41;MRPL4;MRPL42;MRPL3;ACTR1A;MRPL2;MRPL9;TNPO1;IFT57;CC2D2A;RAB8A;YWHAG;OFD1;CCT3;NCOA2;CCT2;TSFM;MRPS26;MKKS;HSP90AA1;MRPS27;MRPS24;MRPS25;TUBB;MRPS22;MRPL49;MRPS21;MRPL48;TUBG1;MRPS7;WDR34;MRPL45;MRPS6;MRPL46;MRPS5;MRPL43;MRPL44;HAUS1;MRPL52;MRPS9;NCOR1;MRPL51;CREB1;TCP1;EXOC6;MAPRE1;PAFAH1B1;SMARCD3;HDAC3;MRPS35;DCTN2;MRPS36;SSNA1;MRPS34;DCTN3;ASAP1;MTIF2;MRPS30;MTIF3;IFT74;HAUS8;ARL13B;HAUS7;TGS1;PDE6D;CCT8;CCT5;CCT4;GADD45GIP1;DYNC1H1;TUBB4B;RAB11A;CARM1;TRIP11;SSBP1 |
| 40 | Degradation of GLI1 by the proteasome Homo sapiens R-HSA-5610780 | 4.59E-15 | 1.64E-13 | 8.37E+00 | 2.76E+02 | PSMD10;PSMD12;PSMD11;PSMD14;PSMD13;CUL1;PSMA7;PSMD8;PSMB6;PSMD9;PSMB7;PSMD6;PSMD7;PSMB5;PSMD4;PSMB2;PSMB3;PSMD2;PSMD3;UBC;PSMB1;PSMD1;PSMF1;RPS27A;SKP1;PSMB8;RBX1;PSMA5;PSMA6;PSMA3;PSMC5;ITCH;PSMA4;PSMC6;PSMA1;PSMC3;PSMA2;PSMC4;PSMC1;PSMC2;PSME1;PSME2 |
| 41 | Cyclin E associated events during G1/S transition Homo sapiens R-HSA-69202 | 5.03E-15 | 1.76E-13 | 7.09E+00 | 2.33E+02 | RB1;PSMD10;PSMD12;CDKN1B;PSMD11;PSMD14;PSMD13;CUL1;PSMA7;CKS1B;PSMD8;PSMB6;PSMD9;PSMB7;PSMD6;PSMD7;PSMB5;PSMD4;PSMB2;PSMB3;PSMD2;PSMD3;UBC;PSMB1;PSMD1;PSMF1;RPS27A;SKP1;PSMB8;PSMA5;PSMA6;PSMA3;PSMC5;PSMA4;PSMC6;PSMA1;PSMC3;PSMA2;PSMC4;CDK4;PSMC1;PSMC2;CDK2;PSME1;PSME2;MNAT1 |

| | | | | | | |
|---|---|---|---|---|---|---|
| 42 | mRNA Splicing Homo sapiens R-HSA-72172 | 6.17E-15 | 2.10E-13 | 3.80E+00 | 1.24E+02 | PDCD7;EIF4A3;HNRNPU;HNRNPR;YBX1;USP39;ELAVL1;SART1;SNRPD2;CLP1;SNRPD1;SNRNP70;MAGOH;SNRPD3;TXNL4A;SNRNP35;CTNNBL1;NCBP1;ZCRB1;PLRG1;GTF2F2;CDC40;PRPF6;SNRPG;SRSF2;SNRNP27;SRSF4;HNRNPH2;SNRPA1;SNRNP25;DNAJC8;SNRPA;SNRPB;SRSF9;SF3B4;SF3B5;DDX5;SF3B2;RBM8A;DHX9;SF3B6;SRRT;FIP1L1;POLR2B;POLR2C;CSTF1;POLR2E;SNRPB2;POLR2H;POLR2J;POLR2K;POLR2L;HNRNPA0;SF3A3;HSPA8;HNRNPA3;CPSF3;CDC5L;CWC15;LSM4;LSM2;HNRNPL;HNRNPM;LSM8;LSM7;SNRNP40;HNRNPK;PHF5A;HNRNPF;HNRNPA2B1;HNRNPD;RNPS1;PRPF31;SNRNP48;HNRNPC;RBMX |
| 43 | Activation of NF-kappaB in B cells Homo sapiens R-HSA-1169091 | 6.32E-15 | 2.10E-13 | 7.26E+00 | 2.37E+02 | PSMD10;PSMD12;PSMD11;PSMD14;PSMD13;CUL1;BCL10;PSMA7;PSMD8;PSMB6;PSMD9;PSMB7;PSMD6;PSMD7;PSMB5;PSMD4;PSMB2;PSMB3;PSMD2;PSMD3;UBC;PSMB1;PSMD1;PSMF1;RPS27A;SKP1;FBXW11;NFKB1;PSMB8;PSMA5;NFKBIA;PSMA6;PSMA3;PSMC5;PSMA4;PSMC6;PSMA1;PSMC3;PSMA2;PSMC4;PSMC1;PSMC2;PSME1;REL;PSME2 |
| 44 | Vif-mediated degradation of APOBEC3G Homo sapiens R-HSA-180585 | 7.19E-15 | 2.34E-13 | 9.43E+00 | 3.07E+02 | PSMD10;PSMD12;PSMD11;PSMD14;PSMD13;PSMA7;PSMD8;PSMB6;PSMD9;PSMB7;PSMD6;PSMD7;PSMB5;PSMD4;PSMB2;PSMB3;PSMD2;PSMD3;UBC;PSMB1;PSMD1;PSMF1;RPS27A;PSMB8;RBX1;PSMA5;PSMA6;PSMA3;PSMC5;PSMA4;PSMC6;PSMA1;PSMC3;PSMA2;PSMC4;PSMC1;PSMC2;PSME1;PSME2 |
| 45 | Autodegradation of the E3 ubiquitin ligase COP1 Homo sapiens R-HSA-349425 | 8.05E-15 | 2.56E-13 | 9.90E+00 | 3.21E+02 | PSMD10;PSMD12;PSMD11;PSMD14;PSMD13;PSMA7;PSMD8;PSMB6;PSMD9;PSMB7;PSMD6;PSMD7;PSMB5;PSMD4;PSMB2;PSMB3;PSMD2;PSMD3;UBC;PSMB1;PSMD1;PSMF1;RPS27A;PSMB8;PSMA5;PSMA6;PSMA3;PSMC5;PSMA4;PSMC6;PSMA1;PSMC3;PSMA2;PSMC4;PSMC1;PSMC2;PSME1;PSME2 |
| 46 | Cdc20:Phospho-APC/C mediated degradation of Cyclin A Homo sapiens R-HSA-174184 | 9.13E-15 | 2.84E-13 | 6.64E+00 | 2.15E+02 | PSMD10;PSMD12;PSMD11;ANAPC15;PSMD14;ANAPC16;PSMD13;UBE2D1;PSMA7;ANAPC11;PSMD8;PSMB6;PSMD9;PSMB7;PSMD6;PSMD7;PSMB5;PSMD4;PSMB2;PSMB3;CDC26;PSMD2;PSMD3;CDC27;UBC;PSMB1;PSMD1;PSMF1;BUB3;RPS27A;UBE2E1;PSMB8;PSMA5;PSMA6;PSMA3;PSMC5;PSMA4;PSMC6;PSMA1;PSMC3;PSMA2;PSMC4;PSMC1;PSMC2;PSME1;PSME2;ANAPC5 |
| 47 | APC/C:Cdc20 mediated degradation of mitotic proteins Homo sapiens R-HSA-176409 | 1.56E-14 | 4.76E-13 | 6.26E+00 | 1.99E+02 | PSMD10;PSMD12;PSMD11;ANAPC15;PSMD14;ANAPC16;PSMD13;UBE2D1;PSMA7;ANAPC11;PSMD8;PSMB6;PSMD9;PSMB7;PSMD6;CCNB1;PSMD7;PSMB5;PSMD4;PSMB2;PSMB3;CDC26;PSMD2;PSMD3;CDC27;UBC;PSMB1;PSMD1;PSMF1;BUB3;RPS27A;UBE2E1;PSMB8;PSMA5;PSMA6;PSMA3;PSMC5;PSMA4;PSMC6;PSMA1;PSMC3;PSMA2;PSMC4;PSMC1;PSMC2;PSME1;PSME2;ANAPC5 |

| | | | | | | |
|---|---|---|---|---|---|---|
| 48 | Stabilization of p53 Homo sapiens R-HSA-69541 | 2.01E-14 | 5.97E-13 | 8.81E+00 | 2.78E+02 | PSMD10;PSMD12;PSMD11;PSMD14;PSMD13;PSMA7;PSMD8;PSMB6;PSMD9;PSMB7;PSMD6;PSMD7;PSMB5;PSMD4;PSMB2;PSMB3;PSMD2;PSMD3;UBC;PSMB1;PSMD1;PSMF1;RPS27A;PSMB8;PSMA5;PSMA6;PSMA3;PSMC5;PSMA4;PSMC6;PSMA1;PSMC3;PSMA2;PSMC4;PSMC1;PSMC2;PSME1;MDM2;PSME2 |
| 49 | APC:Cdc20 mediated degradation of cell cycle proteins prior to satisfation of the cell cycle checkpoint Homo sapiens R-HSA-179419 | 2.04E-14 | 5.97E-13 | 6.37E+00 | 2.01E+02 | PSMD10;PSMD12;PSMD11;ANAPC15;PSMD14;ANAPC16;PSMD13;UBE2D1;PSMA7;ANAPC11;PSMD8;PSMB6;PSMD9;PSMB7;PSMD6;PSMD7;PSMB5;PSMD4;PSMB2;PSMB3;CDC26;PSMD2;PSMD3;CDC27;UBC;PSMB1;PSMD1;PSMF1;BUB3;RPS27A;UBE2E1;PSMB8;PSMA5;PSMA6;PSMA3;PSMC5;PSMA4;PSMC6;PSMA1;PSMC3;PSMA2;PSMC4;PSMC1;PSMC2;PSME1;PSME2;ANAPC5 |
| 50 | p53-Independent DNA Damage Response Homo sapiens R-HSA-69610 | 2.33E-14 | 6.40E-13 | 9.19E+00 | 2.89E+02 | PSMD10;PSMD12;PSMD11;PSMD14;PSMD13;PSMA7;PSMD8;PSMB6;PSMD9;PSMB7;PSMD6;PSMD7;PSMB5;PSMD4;PSMB2;PSMB3;PSMD2;PSMD3;UBC;PSMB1;PSMD1;PSMF1;RPS27A;PSMB8;PSMA5;PSMA6;PSMA3;PSMC5;PSMA4;PSMC6;PSMA1;PSMC3;PSMA2;PSMC4;PSMC1;PSMC2;PSME1;PSME2 |
| 51 | p53-Independent G1/S DNA damage checkpoint Homo sapiens R-HSA-69613 | 2.33E-14 | 6.40E-13 | 9.19E+00 | 2.89E+02 | PSMD10;PSMD12;PSMD11;PSMD14;PSMD13;PSMA7;PSMD8;PSMB6;PSMD9;PSMB7;PSMD6;PSMD7;PSMB5;PSMD4;PSMB2;PSMB3;PSMD2;PSMD3;UBC;PSMB1;PSMD1;PSMF1;RPS27A;PSMB8;PSMA5;PSMA6;PSMA3;PSMC5;PSMA4;PSMC6;PSMA1;PSMC3;PSMA2;PSMC4;PSMC1;PSMC2;PSME1;PSME2 |
| 52 | Ubiquitin Mediated Degradation of Phosphorylated Cdc25A Homo sapiens R-HSA-69601 | 2.33E-14 | 6.40E-13 | 9.19E+00 | 2.89E+02 | PSMD10;PSMD12;PSMD11;PSMD14;PSMD13;PSMA7;PSMD8;PSMB6;PSMD9;PSMB7;PSMD6;PSMD7;PSMB5;PSMD4;PSMB2;PSMB3;PSMD2;PSMD3;UBC;PSMB1;PSMD1;PSMF1;RPS27A;PSMB8;PSMA5;PSMA6;PSMA3;PSMC5;PSMA4;PSMC6;PSMA1;PSMC3;PSMA2;PSMC4;PSMC1;PSMC2;PSME1;PSME2 |
| 53 | Regulation of ornithine decarboxylase (ODC) Homo sapiens R-HSA-350562 | 2.64E-14 | 7.12E-13 | 9.64E+00 | 3.01E+02 | PSMD10;PSMD12;PSMD11;PSMD14;PSMD13;OAZ1;PSMA7;PSMD8;PSMB6;PSMD9;PSMB7;PSMD6;PSMD7;PSMB5;PSMD4;PSMB2;PSMB3;PSMD2;PSMD3;PSMB1;PSMD1;PSMF1;PSMB8;PSMA5;PSMA6;PSMA3;PSMC5;PSMA4;PSMC6;PSMA1;PSMC3;PSMA2;PSMC4;PSMC1;PSMC2;PSME1;PSME2 |

| | | | | | | |
|---|---|---|---|---|---|---|
| 54 | Dectin-1 mediated noncanonical NF-kB signaling Homo sapiens R-HSA-5607761 | 2.97E-14 | 7.88E-13 | 7.49E+00 | 2.33E+02 | PSMD10;PSMD12;PSMD11;PSMD14;PSMD13;CUL1;PSMA7;PSMD8;PSMB6;PSMD9;PSMB7;PSMD6;PSMD7;PSMB5;PSMD4;PSMB2;PSMB3;PSMD2;PSMD3;UBC;PSMB1;PSMD1;PSMF1;RPS27A;SKP1;FBXW11;PSMB8;PSMA5;PSMA6;PSMA3;PSMC5;PSMA4;PSMC6;PSMA1;PSMC3;PSMA2;PSMC4;PSMC1;PSMC2;PSME1;PSME2;UBE2M |
| 55 | Activation of APC/C and APC/C:Cdc20 mediated degradation of mitotic proteins Homo sapiens R-HSA-176814 | 3.37E-14 | 8.78E-13 | 6.03E+00 | 1.87E+02 | PSMD10;PSMD12;PSMD11;ANAPC15;PSMD14;ANAPC16;PSMD13;UBE2D1;PSMA7;ANAPC11;PSMD8;PSMB6;PSMD9;PSMB7;PSMD6;CCNB1;PSMD7;PSMB5;PSMD4;PSMB2;PSMB3;CDC26;PSMD2;PSMD3;CDC27;UBC;PSMB1;PSMD1;PSMF1;BUB3;RPS27A;UBE2E1;PSMB8;PSMA5;PSMA6;PSMA3;PSMC5;PSMA4;PSMC6;PSMA1;PSMC3;PSMA2;PSMC4;PSMC1;PSMC2;PSME1;PSME2;ANAPC5 |
| 56 | mRNA Splicing - Major Pathway Homo sapiens R-HSA-72163 | 3.67E-14 | 9.38E-13 | 3.83E+00 | 1.19E+02 | EIF4A3;HNRNPU;HNRNPR;YBX1;USP39;ELAVL1;SART1;SNRPD2;CLP1;SNRPD1;SNRNP70;MAGOH;SNRPD3;TXNL4A;CTNNBL1;NCBP1;PLRG1;GTF2F2;CDC40;PRPF6;SNRPG;SRSF2;SNRNP27;SRSF4;HNRNPH2;SNRPA1;DNAJC8;SNRPA;SNRPB;SRSF9;SF3B4;SF3B5;DDX5;SF3B2;RBM8A;DHX9;SF3B6;SRRT;FIP1L1;POLR2B;POLR2C;CSTF1;POLR2E;SNRPB2;POLR2H;POLR2J;POLR2K;POLR2L;HNRNPA0;SF3A3;HSPA8;HNRNPA3;CPSF3;CDC5L;CWC15;LSM4;LSM2;HNRNPL;HNRNPM;LSM8;LSM7;SNRNP40;HNRNPK;PHF5A;HNRNPF;HNRNPA2B1;HNRNPD;RNPS1;PRPF31;HNRNPC;RBMX |
| 57 | APC/C:Cdh1 mediated degradation of Cdc20 and other APC/C:Cdh1 targeted proteins in late mitosis/early G1 Homo sapiens R-HSA-174178 | 5.88E-14 | 1.48E-12 | 6.24E+00 | 1.90E+02 | PSMD10;PSMD12;PSMD11;ANAPC15;PSMD14;ANAPC16;PSMD13;UBE2D1;PSMA7;ANAPC11;PSMD8;PSMB6;PSMD9;PSMB7;PSMD6;PSMD7;PSMB5;PSMD4;PSMB2;PSMB3;CDC26;PSMD2;PSMD3;CDC27;UBC;PSMB1;PSMD1;PSMF1;RPS27A;UBE2E1;PSMB8;PSMA5;PSMA6;PSMA3;PSMC5;PSMA4;PSMC6;PSMA1;PSMC3;PSMA2;PSMC4;PSMC1;PSMC2;PSME1;PSME2;ANAPC5 |
| 58 | Cross-presentation of soluble exogenous antigens (endosomes) Homo sapiens R-HSA-1236978 | 8.58E-14 | 2.12E-12 | 9.37E+00 | 2.82E+02 | PSMD10;PSMD12;PSMD11;PSMD14;PSMD13;PSMA7;PSMD8;PSMB6;PSMD9;PSMB7;PSMD6;PSMD7;PSMB5;PSMD4;PSMB2;PSMB3;PSMD2;PSMD3;PSMB1;PSMD1;PSMF1;PSMB8;PSMA5;PSMA6;PSMA3;PSMC5;PSMA4;PSMC6;PSMA1;PSMC3;PSMA2;PSMC4;PSMC1;PSMC2;PSME1;PSME2 |

| | | | | | | |
|---|---|---|---|---|---|---|
| 59 | Regulation of RAS by GAPs Homo sapiens R-HSA-5658442 | 1.00E-13 | 2.44E-12 | 6.48E+00 | 1.94E+02 | PSMD10;PSMD12;PSMD11;PSMD14;PSMD13;CUL3;RASAL1;PSMA7;PSMD8;PSMB6;PSMD9;PSMB7;PSMD6;PSMD7;PSMB5;PSMD4;PSMB2;PSMB3;PSMD2;PSMD3;UBC;PSMB1;PSMD1;PSMF1;RPS27A;DAB2IP;PSMB8;RBX1;PSMA5;PSMA6;PSMA3;PSMC5;PSMA4;PSMC6;PSMA1;PSMC3;PSMA2;PSMC4;RASA1;PSMC1;PSMC2;PSME1;PSME2;KRAS |
| 60 | CDT1 association with the CDC6:ORC:origin complex Homo sapiens R-HSA-68827 | 1.12E-13 | 2.68E-12 | 7.53E+00 | 2.24E+02 | PSMD10;PSMD12;PSMD11;PSMD14;MCM8;PSMD13;GMNN;PSMA7;PSMD8;PSMB6;PSMD9;PSMB7;PSMD6;PSMD7;PSMB5;PSMD4;PSMB2;PSMB3;PSMD2;PSMD3;UBC;PSMB1;PSMD1;PSMF1;RPS27A;PSMB8;PSMA5;PSMA6;PSMA3;PSMC5;PSMA4;PSMC6;PSMA1;PSMC3;PSMA2;PSMC4;PSMC1;PSMC2;PSME1;PSME2 |
| 61 | Degradation of beta-catenin by the destruction complex Homo sapiens R-HSA-195253 | 1.60E-13 | 3.75E-12 | 5.30E+00 | 1.56E+02 | GSK3B;PSMD8;PSMD9;FRAT1;PSMD6;PSMD7;PSMD4;PPP2R5E;PSMD2;PSMD3;PSMD1;SKP1;PPP2R5A;RBX1;PSMA5;PSMA6;PSMA3;PSMA4;PSMA1;PSMA2;PSME1;PSME2;PSMD10;PSMD12;PSMD11;PSMD14;PSMD13;HDAC1;CUL1;PSMA7;PPP2CA;PSMB6;PSMB7;PPP2CB;PSMB5;PSMB2;PSMB3;UBC;PSMB1;PSMF1;RPS27A;TCF7L1;CSNK1A1;PSMB8;PSMC5;PSMC6;PSMC3;PSMC4;PSMC1;PSMC2 |
| 62 | p53-Dependent G1/S DNA damage checkpoint Homo sapiens R-HSA-69580 | 1.68E-13 | 3.81E-12 | 6.78E+00 | 1.99E+02 | PSMD10;PSMD12;CDKN1B;PSMD11;PSMD14;PSMD13;PSMA7;PSMD8;PSMB6;PSMD9;PSMB7;PSMD6;PSMD7;PSMB5;PSMD4;PSMB2;PCBP4;PSMB3;PSMD2;PSMD3;UBC;PSMB1;PSMD1;PSMF1;RPS27A;PSMB8;PSMA5;PSMA6;PSMA3;PSMC5;PSMA4;PSMC6;PSMA1;PSMC3;PSMA2;PSMC4;PSMC1;PSMC2;CDK2;PSME1;MDM2;PSME2 |
| 63 | p53-Dependent G1 DNA Damage Response Homo sapiens R-HSA-69563 | 1.68E-13 | 3.81E-12 | 6.78E+00 | 1.99E+02 | PSMD10;PSMD12;CDKN1B;PSMD11;PSMD14;PSMD13;PSMA7;PSMD8;PSMB6;PSMD9;PSMB7;PSMD6;PSMD7;PSMB5;PSMD4;PSMB2;PCBP4;PSMB3;PSMD2;PSMD3;UBC;PSMB1;PSMD1;PSMF1;RPS27A;PSMB8;PSMA5;PSMA6;PSMA3;PSMC5;PSMA4;PSMC6;PSMA1;PSMC3;PSMA2;PSMC4;PSMC1;PSMC2;CDK2;PSME1;MDM2;PSME2 |

| | | | | | | |
|---|---|---|---|---|---|---|
| 64 | Disease Homo sapiens R-HSA-1643685 | 2.17E-13 | 4.85E-12 | 1.82E+00 | 5.32E+01 | RPL5;RPL30;RPL3;HDAC10;HDAC11;CCNC;RBPJ;RPL8;RPL6;PSMD8;RPS15;PSMD9;PSMD6;C1GALT1C1;PSMD7;PSMD4;RPS19;RPL18A;AKT2;PSMD2;PSMD3;FGFR1OP2;AP1S2;AKT1;PSMD1;RCC1;RPS10;RPS13;RPS12;SKP1;MCCC1;DKK1;HSPG2;NUP93;BTC;CDC37;FDX1;PSME1;PSME2;RPL29;RPL28;VPS28;MMADHC;MAPKAP1;SDC2;PDGFB;SUPT4H1;PIK3R3;RPS4Y1;APH1A;APH1B;PSMF1;JAG2;XRCC6;XRCC5;EIF2AK2;ERLIN2;MSRA;SNW1;NMT1;NAPEPLD;SDC1;GRB2;CALR;CALM1;RPS21;RPS24;CD320;RPS23;ARF1;CDKN1B;RPLP1;CLTA;AP2A2;ADAMTSL2;NELFCD;RAC1;MUC15;PSENEN;NCOR2;PSMA5;KAT2B;PSMA6;PSMA3;NCOR1;PSMA4;PSMA1;PSMA2;TLR5;HBEGF;RPL10;DERL2;PSMA7;PSMB6;PPP2CA;PSMB7;PPP2CB;PSMB5;PSMB2;OS9;PSMB3;GPC1;PSMB1;AP2S1;RPL14;GPC5;RPL15;RPS2;RPL18;RPS27A;PAK2;RPL19;RANBP1;NPM1;CSNK1A1;STAT3;C1GALT1;MIB2;AP2B1;GTF2H4;PSMB8;GCK;FKBP1A;PSMC5;PSMC6;PSMC3;PSMC4;PSMC1;PSMC2;TCEA1;FAU;HSPA1A;SEMA5B;GTF2B;RPL10A;FGF2;KPNA4;KPNA2;KPNA3;AP2M1;KPNA1;MAP2K3;RNF43;SUPT16H;RPS9;MAP2K2;RPS8;RPS5;LIG4;RPSA;GTF2F2;RBX1;CHMP4B;CHMP4A;PPIA;PSMD10;VCP;PSMD12;TSG101;PSMD11;PSMD14;PSMD13;SEL1L;CUL1;GTF2E2;ACACA;BAG4;TPR;UBC;ATP6V1H;AP1M2;SMAD4;TGFB1;CPSF6;SYT1;IDH1;HMGA1;EEF2;NFKB1;CDK9;NFKBIA;FGF18;CHMP2B;MDM2;CHMP2A;MNAT1;GRSF1;MUC20;PRR5;GSK3B;GSK3A;HSP90AB1;PSIP1;RPL7A;AP1G1;PPP2R5E;MLST8;LRRFIP1;HSP90AA1;PDCD6IP;NCBP[illegible] |
| 65 | APC/C-mediated degradation of cell cycle proteins Homo sapiens R-HSA-174143 | 2.27E-13 | 4.92E-12 | 5.09E+00 | 1.48E+02 | UBE2D1;PSMD8;PSMD9;PSMD6;PSMD7;PSMD4;CDC26;PSMD2;PSMD3;CDC27;PSMD1;SKP1;UBE2E1;PSMA5;PSMA6;PSMA3;PSMA4;PSMA1;PSMA2;PSME1;PSME2;ANAPC5;PSMD10;PSMD12;PSMD11;ANAPC15;PSMD14;ANAPC16;PSMD13;CUL1;PSMA7;ANAPC11;PSMB6;PSMB7;CCNB1;PSMB5;PSMB2;PSMB3;UBC;PSMB1;PSMF1;BUB3;RPS27A;PSMB8;PSMC5;PSMC6;PSMC3;PSMC4;PSMC1;PSMC2;CDK2 |
| 66 | Regulation of mitotic cell cycle Homo sapiens R-HSA-453276 | 2.27E-13 | 4.92E-12 | 5.09E+00 | 1.48E+02 | UBE2D1;PSMD8;PSMD9;PSMD6;PSMD7;PSMD4;CDC26;PSMD2;PSMD3;CDC27;PSMD1;SKP1;UBE2E1;PSMA5;PSMA6;PSMA3;PSMA4;PSMA1;PSMA2;PSME1;PSME2;ANAPC5;PSMD10;PSMD12;PSMD11;ANAPC15;PSMD14;ANAPC16;PSMD13;CUL1;PSMA7;ANAPC11;PSMB6;PSMB7;CCNB1;PSMB5;PSMB2;PSMB3;UBC;PSMB1;PSMF1;BUB3;RPS27A;PSMB8;PSMC5;PSMC6;PSMC3;PSMC4;PSMC1;PSMC2;CDK2 |

| | | | | | | |
|---|---|---|---|---|---|---|
| 67 | Processing of Capped Intron-Containing Pre-mRNA Homo sapiens R-HSA-72203 | 3.16E-13 | 6.76E-12 | 2.98E+00 | 8.56E+01 | NXT1;PDCD7;EIF4A3;HNRNPU;HNRNPR;YBX1;USP39;ELAVL1;SART1;SNRPD2;CLP1;SNRPD1;SNRNP70;MAGOH;SNRPD3;TXNL4A;SNRNP35;CTNNBL1;NCBP1;THOC3;THOC2;ZCRB1;PLRG1;THOC5;GTF2F2;CDC40;NUP93;DDX39A;PRPF6;SARNP;NUP50;SNRPG;SRSF2;SNRNP27;HNRNPH2;SRSF4;SNRPA1;SNRNP25;DNAJC8;SNRPA;SNRPB;SRSF9;SF3B4;FYTTD1;SLBP;SF3B5;SF3B2;DDX5;RBM8A;DHX9;POLDIP3;SF3B6;SRRT;FIP1L1;POLR2B;POLR2C;TPR;CSTF1;POLR2E;SNRPB2;POLR2H;EIF4E;POLR2J;POLR2K;POLR2L;HNRNPA0;SF3A3;HSPA8;HNRNPA3;WTAP;CPSF3;CDC5L;CWC15;LSM4;LSM2;HNRNPL;HNRNPM;LSM8;LSM7;SNRNP40;HNRNPK;PHF5A;HNRNPF;HNRNPA2B1;HNRNPD;RNPS1;PRPF31;SNRNP48;HNRNPC;RBMX |
| 68 | G2/M Transition Homo sapiens R-HSA-69275 | 3.81E-13 | 8.02E-12 | 3.14E+00 | 8.97E+01 | YWHAE;CEP57;HSP90AB1;CETN2;PPP2R2A;PPP1CB;PSMD8;PSMD9;PSMD6;ACTR1A;PSMD7;PSMD4;PSMD2;PSMD3;PSMD1;PHLDA1;YWHAG;RAB8A;SKP1;OFD1;HSP90AA1;FKBPL;FBXW11;TUBB;TUBG1;TUBA4A;CDC25B;HAUS1;PSMA5;PSMA6;PSMA3;PSMA4;PSMA1;PSMA2;AKAP9;PSME1;PSME2;MAPRE1;AJUBA;OPTN;PAFAH1B1;PSMD10;PSMD12;PSMD11;PSMD14;LIN37;DCTN2;TUBGCP2;PSMD13;SSNA1;DCTN3;CUL1;PSMA7;PPP2CA;PSMB6;PSMB7;PPP2CB;CCNB1;PSMB5;RBBP4;HAUS8;PSMB2;PRKAR2B;HAUS7;PSMB3;UBC;PSMB1;PSMF1;RPS27A;DYNC1H1;CDK11A;TUBB4B;CDK11B;PSMB8;PSMC5;PSMC6;PSMC3;PSMC4;PSMC1;CDK2;PSMC2;TUBGCP4;MNAT1 |
| 69 | TP53 Regulates Metabolic Genes Homo sapiens R-HSA-5628897 | 6.01E-13 | 1.25E-11 | 4.99E+00 | 1.40E+02 | YWHAE;GPI;COX7B;YWHAB;COX4I1;PRKAG1;PRKAG2;COX6A1;COX7C;GLS;YWHAQ;AKT2;MLST8;AKT1;YWHAG;YWHAH;COX8A;YWHAZ;COX6B1;RRAGA;COX7A2L;RRM2B;RRAGB;SCO2;DDIT4;COX19;PRKAA1;COX16;COX18;COX5B;COX5A;PRDX2;PRDX5;PRDX1;COX14;SURF1;NDUFA4;TXNRD1;GSR;COX6C;LRPPRC;MTOR;RHEB;AGO2;CYCS;LAMTOR1;LAMTOR4;LAMTOR3;COX20;LAMTOR5 |
| 70 | Host Interactions of HIV factors Homo sapiens R-HSA-162909 | 6.62E-13 | 1.35E-11 | 3.67E+00 | 1.03E+02 | ARF1;PSIP1;AP2A2;PSMD8;PSMD9;PSMD6;PSMD7;PSMD4;AP1G1;PSMD2;PSMD3;AP1S2;PSMD1;RCC1;RAC1;AP2M1;KPNA1;SKP1;RBX1;NUP93;PSMA5;PSMA6;PSMA3;PSMA4;PSMA1;PSMA2;NUP50;PSME1;PSME2;SLC25A5;PPIA;SLC25A4;SLC25A6;PSMD10;PSMD12;PSMD11;PSMD14;PSMD13;PSMA7;PSMB6;PSMB7;PSMB5;PSMB2;PSMB3;TPR;UBC;PSMB1;AP2S1;ATP6V1H;FYN;PSMF1;RPS27A;PAK2;AP1M2;RANBP1;NPM1;HMGA1;AP2B1;PSMB8;CDK9;PSMC5;PSMC6;PSMC3;PSMC4;PSMC1;PSMC2;RAN |

| | | | | | | |
|---|---|---|---|---|---|---|
| 71 | Vesicle-mediated transport Homo sapiens R-HSA-5653656 | 7.73E-13 | 1.56E-11 | 2.01E+00 | 5.62E+01 | APP;SCOC;SYS1;KDELR1;GCC2;RAB43;GOLGA2;TFG;KIF5C;AKT2;KIF5B;AP1S2;AKT1;ARFIP2;KIF21A;KIFAP3;VPS36;AP2M1;TPD52;NSF;COG6;COG2;ANK2;CHMP4B;KDELR2;CHMP4A;VPS25;STAM2;VPS28;COPB2;SPTBN5;RAB5C;TMED10;TSG101;STXBP3;SAR1B;COPB1;ARL1;KLC1;TGOLN2;BLOC1S6;SCFD1;HSPH1;SNX2;MAN2A2;KIF3A;MYO6;UBC;RAB6A;AP1M2;SNX5;TRAPPC3;HSPA8;RAB4A;SNAPIN;TRAPPC1;TRAPPC4;TRAPPC5;SURF4;M6PR;ASPSCR1;MYO5A;LNPEP;SNF8;TRAPPC9;STAM;AP4B1;CNIH1;TMEM115;GOLGB1;CHMP2B;CHMP2A;CALR;CALM1;COPE;PICALM;YWHAE;ARF4;ARF1;MCFD2;SERPINA1;YWHAB;CLTC;SNAP23;CTSZ;CLTB;PRKAG1;AP4E1;CLTA;PRKAG2;ARRB1;MIA3;CLINT1;PPP6C;AP1G2;YWHAQ;AP1G1;TMED3;TMED2;AP3S1;RAC1;TMED7;SPTAN1;RAB8A;YWHAG;YWHAH;HSP90AA1;TMED9;NAA30;COPZ2;VPS37D;PLA2G4A;AP3B1;VPS37B;ARFGAP3;YWHAZ;COPZ1;DNM2;VAMP8;TRAPPC6A;KIFC2;KIF2A;DNAJC6;MYH8;PLIN3;CHMP3;EXOC6;NAA38;DNASE2;SEC22B;PAFAH1B3;PAFAH1B2;RHOQ;VAMP3;PAFAH1B1;SEC22C;CHMP5;NAPA;DCTN6;NBAS;RALA;TMF1;DCTN2;RAB1B;VPS4B;GOSR2;USO1;VPS4A;DCTN3;NAPG;STX10;CYTH3;LMAN1;VPS51;LMAN2;VPS53;MAN1C1;STX5;STX4;CD59;RPS27A;CD55;CYTH1;DYNC1H1;RIC1;SEC24A;SORT1;TRAPPC10;GALNT1;DTNBP1;TPD52L1;RAB11A;ARCN1;RAB10;TJP1;RAB14;KIF26B;RAB13;RAB18;YIPF6;TRIP11;RAB9A |
| 72 | Antigen processing-Cross presentation Homo sapiens R-HSA-1236975 | 8.31E-13 | 1.62E-11 | 5.03E+00 | 1.40E+02 | PSMD8;PSMD9;PSMD6;PSMD7;PSMD4;CTSL;SEC61G;PSMD2;PSMD3;PSMD1;SEC61B;PDIA3;TAP1;HLA-C;CYBA;HLA-G;HLA-E;PSMA5;PSMA6;PSMA3;PSMA4;PSMA1;PSMA2;PSME1;PSME2;PSMD10;PSMD12;PSMD11;PSMD14;PSMD13;PSMA7;PSMB6;PSMB7;PSMB5;PSMB2;PSMB3;UBC;PSMB1;PSMF1;RPS27A;LNPEP;PSMB8;PSMC5;PSMC6;PSMC3;PSMC4;PSMC1;PSMC2;CALR |
| 73 | G1/S DNA Damage Checkpoints Homo sapiens R-HSA-69615 | 8.34E-13 | 1.62E-11 | 6.19E+00 | 1.72E+02 | PSMD10;PSMD12;CDKN1B;PSMD11;PSMD14;PSMD13;PSMA7;PSMD8;PSMB6;PSMD9;PSMB7;PSMD6;PSMD7;PSMB5;PSMD4;PSMB2;PCBP4;PSMB3;PSMD2;PSMD3;UBC;PSMB1;PSMD1;PSMF1;RPS27A;PSMB8;PSMA5;PSMA6;PSMA3;PSMC5;PSMA4;PSMC6;PSMA1;PSMC3;PSMA2;PSMC4;PSMC1;PSMC2;CDK2;PSME1;MDM2;PSME2 |

| | | | | | | |
|---|---|---|---|---|---|---|
| 74 | Mitotic G2-G2/M phases Homo sapiens R-HSA-453274 | 8.37E-13 | 1.62E-11 | 3.07E+00 | 8.54E+01 | YWHAE;CEP57;HSP90AB1;CETN2;PPP2R2A;PPP1CB;PSMD8;PSMD9;PSMD6;ACTR1A;PSMD7;PSMD4;PSMD2;PSMD3;PSMD1;PHLDA1;YWHAG;RAB8A;SKP1;OFD1;HSP90AA1;FKBPL;FBXW11;TUBB;TUBG1;TUBA4A;CDC25B;HAUS1;PSMA5;PSMA6;PSMA3;PSMA4;PSMA1;PSMA2;AKAP9;PSME1;PSME2;MAPRE1;AJUBA;OPTN;PAFAH1B1;PSMD10;PSMD12;PSMD11;PSMD14;LIN37;DCTN2;TUBGCP2;PSMD13;SSNA1;DCTN3;CUL1;PSMA7;PPP2CA;PSMB6;PSMB7;PPP2CB;CCNB1;PSMB5;RBBP4;HAUS8;PSMB2;PRKAR2B;HAUS7;PSMB3;UBC;PSMB1;PSMF1;RPS27A;DYNC1H1;CDK11A;TUBB4B;CDK11B;PSMB8;PSMC5;PSMC6;PSMC3;PSMC4;PSMC1;CDK2;PSMC2;TUBGCP4;MNAT1 |
| 75 | Hedgehog 'on' state Homo sapiens R-HSA-5632684 | 1.13E-12 | 2.17E-11 | 4.84E+00 | 1.33E+02 | ARRB1;PSMD8;PSMD9;PSMD6;PSMD7;PSMD4;PSMD2;BOC;PSMD3;PSMD1;RBX1;PSMA5;PSMA6;PSMA3;EVC;PSMA4;PSMA1;PSMA2;PSME1;PSME2;PSMD10;PSMD12;PSMD11;PSMD14;PSMD13;CUL3;PSMA7;PSMB6;PSMB7;PSMB5;PSMB2;PSMB3;KIF3A;UBC;PSMB1;SPOP;PSMF1;RPS27A;EVC2;SMURF2;CSNK1A1;PSMB8;PSMC5;ITCH;PSMC6;PSMC3;PSMC4;PSMC1;PSMC2;DZIP1 |
| 76 | Signaling by Hedgehog Homo sapiens R-HSA-5358351 | 1.30E-12 | 2.44E-11 | 3.50E+00 | 9.58E+01 | GSK3B;ARRB1;PSMD8;PSMD9;PSMD6;PSMD7;PSMD4;PSMD2;PSMD3;BOC;PSMD1;IFT57;SKP1;OFD1;RBX1;PSMA5;PSMA6;PSMA3;EVC;PSMA4;PSMA1;PRKAR1A;PSMA2;PSME1;PSME2;PSMD10;VCP;PSMD12;PSMD11;PSMD14;PSMD13;CUL3;SEL1L;CUL1;ADCY3;DERL2;PSMA7;ADCY5;PSMB6;ERLEC1;PSMB7;PSMB5;OS9;PSMB2;KIF3A;PRKAR2B;PSMB3;PRKAR2A;UBC;PSMB1;SPOP;GPC5;PSMF1;RPS27A;EVC2;SMURF2;CSNK1A1;PSMB8;PSMC5;ITCH;PSMC6;PSMC3;PSMC4;PSMC1;PSMC2;GNAS;P4HB;NOTUM;DZIP1 |
| 77 | PCP/CE pathway Homo sapiens R-HSA-4086400 | 1.95E-12 | 3.63E-11 | 4.52E+00 | 1.22E+02 | CLTC;CLTB;CLTA;AP2A2;PSMD8;PSMD9;PSMD6;PSMD7;PSMD4;PSMD2;PSMD3;PSMD1;RAC1;AP2M1;RHOA;PSMA5;PSMA6;PSMA3;PSMA4;PSMA1;PSMA2;PSME1;PSME2;PFN1;PSMD10;PSMD12;PSMD11;PSMD14;PSMD13;PSMA7;PSMB6;PSMB7;PARD6A;WNT11;PSMB5;PSMB2;PSMB3;UBC;PSMB1;AP2S1;DVL3;PSMF1;RPS27A;SMURF2;AP2B1;PSMB8;PSMC5;PSMC6;PSMC3;PSMC4;PSMC1;PSMC2 |

| | | | | | | |
|---|---|---|---|---|---|---|
| 78 | Antigen processing: Ubiquitination & Proteasome degradation Homo sapiens R-HSA-983168 | 5.81E-12 | 1.07E-10 | 2.46E+00 | 6.37E+01 | UBE3C;UBE2D2;FBXO27;UBE2D3;LTN1;KEAP1;UBE2D1;UBE2L6;UBE2L3;PSMD8;PSMD9;HERC4;NPEPPS;RNF115;HERC3;PSMD6;RNF114;PSMD7;HERC2;PSMD4;CDC26;PSMD2;RNF19A;PSMD3;CDC27;FBXO3;PSMD1;SKP1;FBXW11;FBXO2;UBE2E3;UBE2E1;UBE2E2;RBX1;RNF126;PSME1;UBE2V2;PSME2;ANAPC5;RBCK1;UBE2V1;PSMD10;ANAPC13;PSMD12;PSMD11;PSMD14;PSMD13;CUL3;CUL2;CUL1;UBR2;UBE2J2;FBXO44;UBE2J1;ANAPC11;SOCS1;BTBD1;UBC;PSMF1;UBE2F;UBE2H;UBE2B;SMURF2;LNPEP;UBE2A;UBAC1;UBE2S;MKRN1;UBE2K;UBE2M;UBE2Q1;UBE2Q2;ARIH2;PJA2;PSMA5;PSMA6;PSMA3;PSMA4;PSMA1;UBE2R2;GAN;PSMA2;ASB13;RNF6;PSMA7;PSMB6;PSMB7;ZNRF1;PSMB5;HECTD1;PSMB2;PSMB3;PSMB1;RPS27A;MIB2;UBE2G1;UBE2G2;PSMB8;PSMC5;ITCH;PSMC6;PSMC3;PSMC4;PSMC1;NEDD4;ASB8;PSMC2;LNX1;STUB1 |
| 79 | Asymmetric localization of PCP proteins Homo sapiens R-HSA-4608870 | 6.70E-12 | 1.21E-10 | 5.89E+00 | 1.51E+02 | PSMD10;PSMD12;PSMD11;PSMD14;PSMD13;PSMA7;PSMD8;PSMB6;PSMD9;PSMB7;PSMD6;PSMD7;PARD6A;PSMB5;PSMD4;PSMB2;PSMB3;PSMD2;PSMD3;UBC;PSMB1;PSMD1;PSMF1;RPS27A;SMURF2;PSMB8;PSMA5;PSMA6;PSMA3;PSMC5;PSMA4;PSMC6;PSMA1;PSMC3;PSMA2;PSMC4;PSMC1;PSMC2;PSME1;PSME2 |

| | | | | | |
|---|---|---|---|---|---|
| 80 | Metabolism Homo sapiens R-HSA-1430728 | 7.08E-12 | 1.27E-10 | 1.45E+00 | 3.72E+01 | CDA;PI4K2B;RPL5;RPL30;RPL3;ENO1;EPRS;ENO2;RPL8;ENO3;RPL6;GLS;PSMD8;PSMD9;PSMD6;PSMD7;SCP2;PSMD4;PSMD2;LIPH;NAMPT;PSMD3;AKT1;PSMD1;CYP1B1;PIP4K2C;PRSS3;GSTK1;ACOT9;ACOT8;SECISBP2;MED8;MED4;HSPG2;NUP93;SULT2B1;MTHFD1;SCO2;PRKAR1A;FDX1;UQCRC1;PSME1;PSME2;TMEM126B;HPRT1;RPL29;RPL28;SAR1B;ABCB7;SHMT1;RPS4Y1;SLC5A3;OAZ1;ADH5;MTMR6;CIAO1;LDHB;CYB5R1;LDHA;RDH11;PRKAR2B;CHST12;PRKAR2A;PTPMT1;AFMID;ST3GAL4;CHST15;PIP5K1B;ST3GAL6;LTA4H;PSMF1;CHST14;HMOX2;FDFT1;LHPP;GSTM4;GSTM3;MTHFS;NR1D1;SQLE;UCK1;QARS;ARL2BP;ADI1;PPT1;MMS19;PPT2;ITPA;GSTM5;GPI;PYGB;MOCS2;ARF1;RPLP1;HEXA;QPRT;ADK;PRKAG2;ECSIT;MED11;TRMT112;ATIC;IMPA1;MED10;RFK;TK2;TK1;LDLRAP1;ENOPH1;TEAD2;ELOVL1;LYPLA1;ARG2;TPI1;GSTO2;GSTO1;PGAM1;PGAM2;ITPK1;ELOVL6;MED27;NCOR2;HADHB;GCHFR;PSMA5;ITPKB;HADHA;PSMA6;PSMA3;MED24;NCOR1;PSMA4;PLCB4;PSMA1;PSMA2;MED21;IVD;CHST1;PLIN3;PLCB1;RAPGEF3;FH;ASAH1;ECHS1;RPL10;GSTP1;ADCY3;PSMA7;ADCY5;PAPSS1;PSMB6;PSMB7;PSMB5;PSMB2;PSMB3;ALOX5;PSMB1;UCP2;RPL14;SC5D;RPL15;RPS2;RPS27A;RPL18;RPL19;AOC1;TIMMDC1;MDH1;MDH2;GSR;NMNAT3;PSMB8;LRPPRC;AGT;PSMC5;PSMC6;PSMC3;ETNK2;PSMC4;ETNK1;PSMC1;PSMC2;CRAT;VKORC1L1;PECR;TPMT;CHPF;RORA;RPL10A;SAT1;PPP1CB;PPP1CC;NAGLU;MPC2;OSBP;CHP1;NUDT15;SLC16A3;RPS9;RPS8;RPS5;IPMK;SARS;RPSA;MINPP1;SLC27[illegible] |
| 81 | Diseases of signal transduction Homo sapiens R-HSA-5663202 | 1.17E-11 | 2.07E-10 | 2.33E+00 | 5.87E+01 | HDAC10;HDAC11;CCNC;RBPJ;FGF2;PSMD8;PSMD9;PSMD6;PSMD7;PSMD4;AKT2;PSMD2;PSMD3;FGFR1OP2;AKT1;PSMD1;SKP1;RNF43;DKK1;GTF2F2;RBX1;BTC;CDC37;PSME1;PSME2;PSMD10;VCP;PSMD12;PSMD11;PSMD14;PSMD13;MAPKAP1;SEL1L;PDGFB;CUL1;PIK3R3;APH1A;BAG4;APH1B;UBC;PSMF1;JAG2;SMAD4;TGFB1;CPSF6;ERLIN2;SNW1;FGF18;NAPEPLD;MDM2;GRB2;PRR5;GSK3B;GSK3A;CDKN1B;PPP2R5E;MLST8;LRRFIP1;PSENEN;HSP90AA1;NCBP1;PDPK1;PPP2R5A;NCOR2;PSMA5;KAT2B;PSMA6;PSMA3;NCOR1;PSMA4;CREB1;PSMA1;PSMA2;AKT1S1;HBEGF;HDAC2;HDAC3;HDAC1;DERL2;CBL;PSMA7;PPP2CA;PSMB6;PSMB7;PPP2CB;ERLEC1;PSMB5;OS9;PSMB2;TNKS2;PSMB3;POLR2B;POLR2C;PSMB1;POLR2E;FYN;POLR2H;RPS27A;POLR2J;POLR2K;POLR2L;CSNK1A1;BAD;STAT3;MIB2;PTPN11;PSMB8;MTOR;FKBP1A;PSMC5;PSMC6;PSMC3;PSMC4;PSMC1;PSMC2;RPS6KB2;KRAS |

| | | | | | | |
|---|---|---|---|---|---|---|
| 82 | Class I MHC mediated antigen processing & presentation Homo sapiens R-HSA-983169 | 1.46E-11 | 2.55E-10 | 2.27E+00 | 5.67E+01 | UBE3C;UBE2D2;FBXO27;UBE2D3;LTN1;KEAP1;UBE2D1;UBE2L6;UBE2L3;PSMD8;PSMD9;HERC4;NPEPPS;RNF115;HERC3;PSMD6;RNF114;PSMD7;HERC2;PSMD4;CDC26;PSMD2;RNF19A;PSMD3;CDC27;FBXO3;PSMD1;SKP1;PDIA3;FBXW11;FBXO2;UBE2E3;UBE2E1;HLA-C;UBE2E2;TAP1;CYBA;HLA-G;HLA-E;RBX1;RNF126;PSME1;UBE2V2;PSME2;ANAPC5;RBCK1;UBE2V1;PSMD10;ANAPC13;PSMD12;PSMD11;PSMD14;SAR1B;PSMD13;CUL3;CUL2;CUL1;UBR2;UBE2J2;FBXO44;UBE2J1;ANAPC11;SOCS1;BTBD1;UBC;PSMF1;UBE2F;UBE2H;UBE2B;HSPA5;SMURF2;LNPEP;UBE2A;UBAC1;UBE2S;MKRN1;CALR;UBE2K;UBE2M;UBE2Q1;CTSL;UBE2Q2;SEC61G;ARIH2;SEC61B;PJA2;PSMA5;PSMA6;PSMA3;PSMA4;PSMA1;UBE2R2;GAN;PSMA2;ASB13;RNF6;PSMA7;PSMB6;PSMB7;ZNRF1;PSMB5;HECTD1;PSMB2;PSMB3;PSMB1;RPS27A;SEC24A;MIB2;UBE2G1;UBE2G2;PSMB8;PSMC5;ITCH;PSMC6;PSMC3;PSMC4;PSMC1;NEDD4;ASB8;PSMC2;LNX1;STUB1 |
| 83 | Removal of licensing factors from origins Homo sapiens R-HSA-69300 | 2.12E-11 | 3.66E-10 | 5.02E+00 | 1.23E+02 | RB1;PSMD10;PSMD12;CDKN1B;PSMD11;PSMD14;MCM8;PSMD13;GMNN;PSMA7;PSMD8;PSMB6;PSMD9;PSMB7;PSMD6;PSMD7;PSMB5;PSMD4;PSMB2;PSMB3;PSMD2;PSMD3;UBC;PSMB1;PSMD1;PSMF1;RPS27A;PSMB8;PSMA5;PSMA6;PSMA3;PSMC5;PSMA4;PSMC6;PSMA1;PSMC3;PSMA2;PSMC4;PSMC1;PSMC2;CDK2;PSME1;PSME2 |
| 84 | Switching of origins to a post-replicative state Homo sapiens R-HSA-69052 | 2.94E-11 | 4.94E-10 | 5.08E+00 | 1.23E+02 | RB1;PSMD10;PSMD12;CDKN1B;PSMD11;PSMD14;MCM8;PSMD13;PSMA7;PSMD8;PSMB6;PSMD9;PSMB7;PSMD6;PSMD7;PSMB5;PSMD4;PSMB2;PSMB3;PSMD2;PSMD3;UBC;PSMB1;PSMD1;PSMF1;RPS27A;PSMB8;PSMA5;PSMA6;PSMA3;PSMC5;PSMA4;PSMC6;PSMA1;PSMC3;PSMA2;PSMC4;PSMC1;PSMC2;CDK2;PSME1;PSME2 |
| 85 | Orc1 removal from chromatin Homo sapiens R-HSA-68949 | 2.94E-11 | 4.94E-10 | 5.08E+00 | 1.23E+02 | RB1;PSMD10;PSMD12;CDKN1B;PSMD11;PSMD14;MCM8;PSMD13;PSMA7;PSMD8;PSMB6;PSMD9;PSMB7;PSMD6;PSMD7;PSMB5;PSMD4;PSMB2;PSMB3;PSMD2;PSMD3;UBC;PSMB1;PSMD1;PSMF1;RPS27A;PSMB8;PSMA5;PSMA6;PSMA3;PSMC5;PSMA4;PSMC6;PSMA1;PSMC3;PSMA2;PSMC4;PSMC1;PSMC2;CDK2;PSME1;PSME2 |
| 86 | CLEC7A (Dectin-1) signaling Homo sapiens R-HSA-5607764 | 3.68E-11 | 6.05E-10 | 3.91E+00 | 9.38E+01 | UBE2D2;UBE2D1;PYCARD;PSMD8;PSMD9;PSMD6;PPP3CB;PSMD7;PSMD4;PSMD2;PSMD3;PSMD1;SKP1;FBXW11;PDPK1;PSMA5;PSMA6;PSMA3;PSMA4;PSMA1;PSMA2;PSME1;PSME2;UBE2V1;PSMD10;PSMD12;PSMD11;PSMD14;PSMD13;CUL1;ITPR2;BCL10;PSMA7;PSMB6;PSMB7;PSMB5;PSMB2;PSMB3;UBC;PSMB1;PSMF1;RPS27A;NFKB1;PSMB8;NFKBIA;PSMC5;PSMC6;PSMC3;PSMC4;PSMC1;PSMC2;CALM1;UBE2M |

| | | | | | | |
|---|---|---|---|---|---|---|
| 87 | Hedgehog 'off' state Homo sapiens R-HSA-5610787 | 3.68E-11 | 6.05E-10 | 3.91E+00 | 9.38E+01 | GSK3B;PSMD8;PSMD9;PSMD6;PSMD7;PSMD4;PSMD2;PSMD3;PSMD1;IFT57;SKP1;OFD1;RBX1;PSMA5;PSMA6;PSMA3;PSMA4;PSMA1;PRKAR1A;PSMA2;PSME1;PSME2;PSMD10;PSMD12;PSMD11;PSMD14;PSMD13;CUL1;ADCY3;PSMA7;ADCY5;PSMB6;PSMB7;PSMB5;PSMB2;KIF3A;PRKAR2B;PSMB3;PRKAR2A;UBC;PSMB1;PSMF1;RPS27A;CSNK1A1;PSMB8;PSMC5;ITCH;PSMC6;PSMC3;PSMC4;PSMC1;PSMC2;GNAS |
| 88 | Apoptosis Homo sapiens R-HSA-109581 | 6.44E-11 | 1.05E-09 | 2.90E+00 | 6.79E+01 | YWHAE;PPP1R13B;YWHAB;TRADD;HMGB1;BBC3;PSMD8;PSMD9;PSMD6;CASP7;PSMD7;PSMD4;YWHAQ;AKT2;PSMD2;PSMD3;AKT1;PSMD1;HIST1H1E;DNM1L;SPTAN1;YWHAG;KPNA1;YWHAH;APPL1;DBNL;APAF1;YWHAZ;PSMA5;PSMA6;PSMA3;PSMA4;PSMA1;PSMA2;PSME1;PSME2;PSMD10;PSMD12;PSMD11;PSMD14;MAGED1;PSMD13;PSMA7;ADD1;PSMB6;PSMB7;PSMB5;PSMB2;PSMB3;UBC;LMNA;PSMB1;PSMF1;FADD;RPS27A;BID;PAK2;GSN;BAD;CFLAR;PTK2;PSMB8;TJP1;H1F0;PSMC5;PSMC6;STK24;PSMC3;PSMC4;PSMC1;PSMC2;NMT1;CYCS;BAX;ACIN1 |
| 89 | Assembly of the pre-replicative complex Homo sapiens R-HSA-68867 | 1.08E-10 | 1.73E-09 | 5.01E+00 | 1.15E+02 | PSMD10;PSMD12;PSMD11;PSMD14;MCM8;PSMD13;GMNN;PSMA7;PSMD8;PSMB6;PSMD9;PSMB7;PSMD6;PSMD7;PSMB5;PSMD4;PSMB2;PSMB3;PSMD2;PSMD3;UBC;PSMB1;PSMD1;PSMF1;RPS27A;PSMB8;PSMA5;PSMA6;PSMA3;PSMC5;PSMA4;PSMC6;PSMA1;PSMC3;PSMA2;PSMC4;PSMC1;PSMC2;PSME1;PSME2 |
| 90 | Regulation of DNA replication Homo sapiens R-HSA-69304 | 1.36E-10 | 2.17E-09 | 4.55E+00 | 1.03E+02 | RB1;PSMD10;PSMD12;CDKN1B;PSMD11;PSMD14;MCM8;PSMD13;GMNN;PSMA7;PSMD8;PSMB6;PSMD9;PSMB7;PSMD6;PSMD7;PSMB5;PSMD4;PSMB2;PSMB3;PSMD2;PSMD3;UBC;PSMB1;PSMD1;PSMF1;RPS27A;PSMB8;PSMA5;PSMA6;PSMA3;PSMC5;PSMA4;PSMC6;PSMA1;PSMC3;PSMA2;PSMC4;PSMC1;PSMC2;CDK2;PSME1;PSME2 |
| 91 | Programmed Cell Death Homo sapiens R-HSA-5357801 | 1.86E-10 | 2.93E-09 | 2.80E+00 | 6.27E+01 | YWHAE;PPP1R13B;YWHAB;TRADD;HMGB1;BBC3;PSMD8;PSMD9;PSMD6;CASP7;PSMD7;PSMD4;YWHAQ;AKT2;PSMD2;PSMD3;AKT1;PSMD1;HIST1H1E;DNM1L;SPTAN1;YWHAG;KPNA1;YWHAH;APPL1;DBNL;APAF1;YWHAZ;PSMA5;PSMA6;PSMA3;PSMA4;PSMA1;PSMA2;PSME1;PSME2;PSMD10;PSMD12;PSMD11;PSMD14;MAGED1;PSMD13;PSMA7;ADD1;PSMB6;PSMB7;PSMB5;PSMB2;PSMB3;UBC;LMNA;PSMB1;PSMF1;FADD;RPS27A;BID;PAK2;GSN;BAD;CFLAR;PTK2;PSMB8;TJP1;H1F0;PSMC5;PSMC6;STK24;PSMC3;PSMC4;PSMC1;PSMC2;NMT1;CYCS;BAX;ACIN1 |

| | | | | | |
|---|---|---|---|---|---|
| 92 | Cap-dependent Translation Initiation Homo sapiens R-HSA-72737 | 2.37E-10 | 3.65E-09 | 3.39E+00 | 7.51E+01 EIF4A2;RPL5;EIF4A1;RPL30;RPL3;RPLP1;RPL8;RPL10A;RPL6;RPS15;RPL7A;RPS19;RPL18A;RPS10;RPS13;RPS12;EIF5B;RPS9;RPS8;RPS5;RPSA;PABPC1;RPL29;RPL28;RPL10;RPS4Y1;RPL14;EIF4H;RPL15;RPS2;RPL18;RPS27A;EIF4E;EIF4B;RPL19;EIF2B5;EIF2B4;EIF2B3;EIF2B2;RPL23A;EIF2S2;EIF2S1;EIF5;EIF3K;EIF3I;EIF3J;EIF3G;EIF3H;FAU;EIF3F;RPS21;EIF3D;RPS24;RPL26L1;EIF3A;EIF4G1;RPS23 |
| 93 | Eukaryotic Translation Initiation Homo sapiens R-HSA-72613 | 2.37E-10 | 3.65E-09 | 3.39E+00 | 7.51E+01 EIF4A2;RPL5;EIF4A1;RPL30;RPL3;RPLP1;RPL8;RPL10A;RPL6;RPS15;RPL7A;RPS19;RPL18A;RPS10;RPS13;RPS12;EIF5B;RPS9;RPS8;RPS5;RPSA;PABPC1;RPL29;RPL28;RPL10;RPS4Y1;RPL14;EIF4H;RPL15;RPS2;RPL18;RPS27A;EIF4E;EIF4B;RPL19;EIF2B5;EIF2B4;EIF2B3;EIF2B2;RPL23A;EIF2S2;EIF2S1;EIF5;EIF3K;EIF3I;EIF3J;EIF3G;EIF3H;FAU;EIF3F;RPS21;EIF3D;RPS24;RPL26L1;EIF3A;EIF4G1;RPS23 |
| 94 | Metabolism of polyamines Homo sapiens R-HSA-351202 | 2.60E-10 | 3.96E-09 | 4.10E+00 | 9.05E+01 PSMD10;PSMD12;PSMD11;PSMD14;PSMD13;AMD1;APIP;SAT1;OAZ1;PSMA7;PSMD8;PSMB6;PSMD9;PSMB7;PSMD6;PSMD7;PSMB5;PSMD4;PSMB2;PSMB3;PSMD2;PSMD3;PSMB1;PSMD1;PSMF1;ENOPH1;ARG2;GOT1;SLC6A12;NMRAL1;PSMB8;PSMA5;PSMA6;PSMA3;PSMC5;PSMA4;PSMC6;PSMA1;PSMC3;PSMA2;PSMC4;ADI1;PSMC1;PSMC2;PSME1;PSME2 |
| 95 | Transcriptional Regulation by TP53 Homo sapiens R-HSA-3700989 | 7.01E-10 | 1.06E-08 | 2.03E+00 | 4.29E+01 EHMT1;COX6A1;GLS;AKT2;AKT1;BNIP3L;SUPT16H;USP7;CSNK2A2;USP2;GTF2F2;COX6B1;RRAGA;COX7A2L;MSH2;RRAGB;SCO2;CSNK2B;DDIT4;MAPKAPK5;COX19;MEAF6;COX16;MAPKAP1;COX18;SUPT4H1;PRDX2;NUAK1;PRDX5;CCNB1;PCBP4;PRDX1;UBC;COX14;PRELID1;SURF1;GADD45A;CNOT4;CDK9;CNOT7;TP53I3;CDK5;BCL6;CNOT11;RHEB;CNOT2;AGO2;CNOT3;CDK2;MDM2;RAD17;PIN1;CYCS;MNAT1;COX20;YWHAE;PRR5;GPI;COX7B;CDKN1B;PPP1R13B;PHF20;SETD9;YWHAB;COX4I1;PRKAG1;CHM;PRKAG2;CHD4;COX7C;BBC3;YWHAQ;MLST8;NELFCD;YWHAG;YWHAH;COX8A;CNOT6L;APAF1;PRMT1;PDPK1;RABGGTB;YWHAZ;TFDP2;RRM2B;TMEM219;PRKAA1;HDAC2;PCNA;HDAC1;COX5B;COX5A;PPP2CA;PPP2CB;RBBP4;POLR2B;PERP;POLR2C;POLR2E;E2F4;RBBP7;POLR2H;RPS27A;BID;BRD7;POLR2J;POLR2K;POLR2L;NPM1;TAF13;NDUFA4;TXNRD1;TAF10;GSR;TAF11;COX6C;HIPK1;GTF2H4;LRPPRC;MTOR;RAD50;RPA3;CARM1;TCEA1;BAX;RAD1;LAMTOR1;LAMTOR4;LAMTOR3;LAMTOR5 |

| | | | | | | |
|---|---|---|---|---|---|---|
| 96 | SRP-dependent cotranslational protein targeting to membrane Homo sapiens R-HSA-1799339 | 1.54E-09 | 2.30E-08 | 3.33E+00 | 6.75E+01 | RPL5;SRP19;RPL30;TRAM1;RPL3;RPLP1;SRP54;SRP14;RPL8;RPL10A;RPL6;RPS15;RPL7A;RPS19;RPL18A;SEC61G;SEC61B;RPS10;RPS13;RPS12;RPS9;SSR4;RPS8;RPS5;SSR2;SSR3;RPSA;DDOST;SRP9;RPL29;RPL28;RPN2;RPL10;RPS4Y1;SRP72;RPL14;RPL15;RPS2;RPL18;RPS27A;SEC11A;RPL19;SEC11C;RPL23A;SPCS3;SPCS2;SPCS1;SRPRB;FAU;RPS21;RPS24;RPL26L1;RPS23 |
| 97 | MAPK6/MAPK4 signaling Homo sapiens R-HSA-5687128 | 3.56E-09 | 5.25E-08 | 3.54E+00 | 6.88E+01 | PSMD10;PSMD12;PSMD11;PSMD14;PSMD13;HSPB1;PSMA7;PSMD8;CDC42;PSMB6;PSMD9;PSMB7;PSMD6;PAK1;PSMD7;PSMB5;PSMD4;PSMB2;PSMB3;PSMD2;PSMD3;UBC;PSMB1;PSMD1;RAC1;PSMF1;RPS27A;PAK2;NCOA3;PSMB8;PSMA5;PSMA6;PSMA3;PSMC5;PSMA4;PSMC6;PSMA1;PSMC3;PSMA2;PSMC4;PSMC1;CDC42EP3;PSMC2;AGO2;PSME1;PSME2;MAPKAPK5 |
| 98 | Cellular responses to stress Homo sapiens R-HSA-2262752 | 4.24E-09 | 6.19E-08 | 1.94E+00 | 3.74E+01 | RB1;UBE2D2;UBE2D3;EHMT1;UBE2D1;ETS1;ETS2;TXN2;CDC26;CDC27;HIST1H2AC;MAP2K3;ATG3;GABARAPL2;TINF2;UBE2E1;CYBA;RBX1;NUP93;EEF1A1;RRAGA;RRAGB;CHMP4B;ULK1;MAPKAPK5;CHMP4A;ANAPC5;FKBP4;VCP;ANAPC15;ANAPC16;RPS19BP1;MRPL18;CUL2;HIF1A;GABARAP;ANAPC11;PRDX3;PRDX2;PRDX5;BAG4;HSPH1;BAG5;ATG101;BAG3;PRDX1;TPR;UBC;BAG1;IGFBP7;ATG5;HSPA9;HSPA8;PHC2;CBX4;HSPA5;ST13;HSPA4;H3F3A;HMGA1;HSPA2;PRDX6;NFKB1;H1F0;CDK6;RHEB;CDK4;CHMP2B;CDK2;CHMP2A;MDM2;CYCS;YWHAE;GSK3B;CDKN1B;HSP90AB1;PRKAG1;PRKAG2;WDR45;MAP1LC3B;MAP1LC3A;MLST8;HIST1H1E;HSP90AA1;HSBP1;H2AFZ;ARNT;WIPI1;WIPI2;SIRT1;DNAJC7;NUP50;AKT1S1;CAT;CHMP3;TERF2IP;ATG4A;ATG4D;BECN1;PRKAA1;WDR45B;GSTP1;HSPA13;MAPK9;RBBP4;DNAJB6;MAPK1;RBBP7;RPS27A;CAMK2G;EGLN1;SUZ12;CDKN2D;EGLN2;TXNRD2;PTGES3;TXNRD1;STAT3;GSR;SOD2;SOD3;MTOR;ATOX1;VEGFA;SOD1;RAD50;RPA3;LAMTOR1;P4HB;LAMTOR4;LAMTOR3;HSPA1A;LAMTOR5 |
| 99 | Downstream signaling events of B Cell Receptor (BCR) Homo sapiens R-HSA-1168372 | 5.12E-09 | 7.33E-08 | 2.43E+00 | 4.63E+01 | PRR5;GSK3B;GSK3A;CDKN1B;FGF2;PSMD8;PSMD9;PSMD6;PSMD7;PSMD4;PPP2R5E;AKT2;PSMD2;PSMD3;MLST8;AKT1;PSMD1;PIP4K2C;IER3;SKP1;FBXW11;PDPK1;PPP2R5A;BTC;PSMA5;PSMA6;PSMA3;CREB1;PSMA4;PSMA1;PSMA2;AKT1S1;PSME1;PSME2;HBEGF;PSMD10;PSMD12;PSMD11;PSMD14;PSMD13;MAPKAP1;CUL1;PDGFB;PIK3R3;BCL10;PSMA7;RASGRP3;PPP2CA;PSMB6;PSMB7;PPP2CB;PSMB5;PSMB2;PSMB3;UBC;PSMB1;MAPK1;PIP5K1B;FYN;PSMF1;RPS27A;BAD;PTPN11;NFKB1;MTOR;PSMB8;NFKBIA;PSMC5;PSMC6;PSMC3;PSMC4;PSMC1;FGF18;AGO2;PSMC2;RPS6KB2;MDM2;REL;GRB2;KRAS |

| | | | | | | |
|---|---|---|---|---|---|---|
| 100 | GTP hydrolysis and joining of the 60S ribosomal subunit Homo sapiens R-HSA-72706 | 5.12E-09 | 7.33E-08 | 3.20E+00 | 6.11E+01 | EIF4A2;RPL5;EIF4A1;RPL30;RPL3;RPLP1;RPL8;RPL10A;RPL6;RPS15;RPL7A;RPS19;RPL18A;RPS10;RPS13;RPS12;EIF5B;RPS9;RPS8;RPS5;RPSA;RPL29;RPL28;RPL10;RPS4Y1;RPL14;EIF4H;RPL15;RPS2;RPL18;RPS27A;EIF4E;EIF4B;RPL19;RPL23A;EIF2S2;EIF2S1;EIF5;EIF3K;EIF3I;EIF3J;EIF3G;EIF3H;FAU;EIF3F;RPS21;EIF3D;RPS24;RPL26L1;EIF3A;EIF4G1;RPS23 |
| 101 | Signaling by Insulin receptor Homo sapiens R-HSA-74752 | 6.32E-09 | 8.95E-08 | 2.03E+00 | 3.82E+01 | PEBP1;WDR83;FGF2;PSMD8;PSMD9;PSMD6;PSMD7;PSMD4;AKT2;PSMD2;PSMD3;AKT1;PSMD1;ATP6V1E1;ATP6V1G1;ATP6V0E1;MAP2K2;RBX1;BTC;RRAGA;RRAGB;AKAP9;PSME1;PSME2;TLN1;ATP6V0D1;ATP6V1A;PSMD10;SPTBN5;PSMD12;SHC2;PSMD11;PSMD14;PSMD13;CUL3;PDGFB;RASAL1;PHB;RASGRP3;RAP1B;RAP1A;UBC;ATP6V1H;PSMF1;ATP6V1D;ATP6V1C1;ATP6V1F;PTK2;PTPRA;RHEB;FGF18;GRB2;CALM1;EIF4G1;CAB39L;YWHAB;RASGRF2;PRKAG1;PRKAG2;ARRB1;PPP2R5E;PEA15;MLST8;JAK2;SPTAN1;JAK1;ATP6V0B;ATP6AP1;DUSP1;PDPK1;STRADB;PPP2R5A;DUSP8;PSMA5;PSMA6;PSMA3;PSMA4;PSMA1;PSMA2;RASA1;AKT1S1;ATP6V0C;HBEGF;PRKAA1;PSMA7;PPP2CA;PSMB6;PSMB7;PPP2CB;PSMB5;PSMB2;PSMB3;PSMB1;MAPK1;CSK;FYN;RPS27A;ATP6V0E2;EIF4E;CAMK2G;EIF4B;DAB2IP;PTPN11;PSMB8;MTOR;PSMC5;PSMC6;PSMC3;PSMC4;PSMC1;PSMC2;KRAS;LAMTOR1;LAMTOR4;LAMTOR3;LAMTOR5 |
| 102 | Downstream TCR signaling Homo sapiens R-HSA-202424 | 6.53E-09 | 9.17E-08 | 3.32E+00 | 6.25E+01 | UBE2D2;UBE2D1;PSMD8;PSMD9;PSMD6;PSMD7;PSMD4;PSMD2;PSMD3;PSMD1;SKP1;FBXW11;PDPK1;PSMA5;PSMA6;PSMA3;PSMA4;PSMA1;PSMA2;PSME1;PSME2;UBE2V1;HLA-DQB1;PSMD10;PSMD12;PSMD11;PSMD14;PSMD13;CUL1;BCL10;PSMA7;PSMB6;PSMB7;PSMB5;PSMB2;PSMB3;UBC;PSMB1;PSMF1;RPS27A;NFKB1;PSMB8;NFKBIA;PSMC5;PSMC6;PSMC3;PSMC4;PSMC1;PSMC2 |
| 103 | C-type lectin receptors (CLRs) Homo sapiens R-HSA-5621481 | 8.78E-09 | 1.22E-07 | 2.93E+00 | 5.43E+01 | UBE2D2;UBE2D1;PYCARD;PSMD8;PSMD9;PSMD6;PPP3CB;PSMD7;PSMD4;PSMD2;PSMD3;PSMD1;SKP1;FBXW11;PDPK1;PSMA5;PSMA6;PSMA3;PSMA4;PSMA1;PSMA2;PSME1;PSME2;UBE2V1;PSMD10;PSMD12;PSMD11;PSMD14;PSMD13;CUL1;ITPR2;BCL10;PSMA7;PSMB6;PSMB7;PAK1;PSMB5;PSMB2;PSMB3;UBC;PSMB1;FYN;PSMF1;RPS27A;PAK2;NFKB1;PSMB8;NFKBIA;PSMC5;PSMC6;PSMC3;PSMC4;PSMC1;PSMC2;KRAS;CALM1;UBE2M |

| | | | | | | |
|---|---|---|---|---|---|---|
| 104 | Asparagine N-linked glycosylation Homo sapiens R-HSA-446203 | 8.92E-09 | 1.23E-07 | 2.14E+00 | 3.96E+01 | DPAGT1;CMAS;KDELR1;TUSC3;GOLGA2;TFG;NUDT14;PDIA3;NSF;ALG8;COG6;ALG5;COG2;ALG2;ALG3;MOGS;ANK2;DPM2;DPM3;KDELR2;COPB2;SPTBN5;TMED10;RPN2;SAR1B;COPB1;SEL1L;SCFD1;FUT8;MAN2A2;NEU1;ST3GAL4;MLEC;ST3GAL6;MPDU1;EDEM3;CTSA;TRAPPC3;TRAPPC1;FUCA1;TRAPPC4;EDEM2;TRAPPC5;GFPT1;DOLK;TRAPPC9;CNIH1;TMEM115;GOLGB1;CALR;COPE;ARF4;B4GALT3;ARF1;MCFD2;SERPINA1;CTSZ;MIA3;GMPPB;PPP6C;TMED3;GNPNAT1;TMED2;TMED7;SPTAN1;TMED9;COPZ2;ARFGAP3;COPZ1;DDOST;TRAPPC6A;DAD1;SEC22B;ST6GALNAC6;SEC22C;NAPA;DCTN6;DCTN2;RAB1B;GOSR2;USO1;DCTN3;DERL2;NAPG;LMAN1;OS9;LMAN2;SLC17A5;MAN1C1;STX5;CD59;MGAT1;CD55;DYNC1H1;SLC35A1;SEC24A;TRAPPC10;ARCN1;GLB1;MGAT4B |
| 105 | Mitochondrial protein import Homo sapiens R-HSA-1268020 | 1.02E-08 | 1.39E-07 | 4.92E+00 | 9.05E+01 | COX19;TOMM40;COX17;TIMM13;TIMM10;TAZ;TOMM20;TOMM22;TIMM50;HSPD1;CHCHD2;SAMM50;GRPEL1;TIMM17A;PMPCB;TIMM17B;PMPCA;CYC1;HSPA9;DNAJC19;MTX1;MTX2;IDH3G;PAM16;TIMM22;TIMM10B;TOMM7;VDAC1;SLC25A4;TOMM5;SLC25A6;TOMM6 |
| 106 | L13a-mediated translational silencing of Ceruloplasmin expression Homo sapiens R-HSA-156827 | 1.10E-08 | 1.47E-07 | 3.14E+00 | 5.75E+01 | EIF4A2;RPL5;EIF4A1;RPL30;RPL3;RPLP1;RPL8;RPL10A;RPL6;RPS15;RPL7A;RPS19;RPL18A;RPS10;RPS13;RPS12;RPS9;RPS8;RPS5;RPSA;PABPC1;RPL29;RPL28;RPL10;RPS4Y1;RPL14;EIF4H;RPL15;RPS2;RPL18;RPS27A;EIF4E;EIF4B;RPL19;RPL23A;EIF2S2;EIF2S1;EIF3K;EIF3I;EIF3J;EIF3G;EIF3H;FAU;EIF3F;RPS21;EIF3D;RPS24;RPL26L1;EIF3A;EIF4G1;RPS23 |
| 107 | 3' -UTR-mediated translational regulation Homo sapiens R-HSA-157279 | 1.10E-08 | 1.47E-07 | 3.14E+00 | 5.75E+01 | EIF4A2;RPL5;EIF4A1;RPL30;RPL3;RPLP1;RPL8;RPL10A;RPL6;RPS15;RPL7A;RPS19;RPL18A;RPS10;RPS13;RPS12;RPS9;RPS8;RPS5;RPSA;PABPC1;RPL29;RPL28;RPL10;RPS4Y1;RPL14;EIF4H;RPL15;RPS2;RPL18;RPS27A;EIF4E;EIF4B;RPL19;RPL23A;EIF2S2;EIF2S1;EIF3K;EIF3I;EIF3J;EIF3G;EIF3H;FAU;EIF3F;RPS21;EIF3D;RPS24;RPL26L1;EIF3A;EIF4G1;RPS23 |
| 108 | G1/S Transition Homo sapiens R-HSA-69206 | 1.18E-08 | 1.56E-07 | 3.04E+00 | 5.55E+01 | RB1;CDKN1B;MCM8;CKS1B;PSMD8;PSMD9;PSMD6;PSMD7;PSMD4;PSMD2;PSMD3;PSMD1;TK2;SKP1;PSMA5;PSMA6;PSMA3;PSMA4;PSMA1;PSMA2;PSME1;PSME2;PSMD10;PSMD12;PSMD11;PCNA;PSMD14;PSMD13;CUL1;PSMA7;PPP2CA;PSMB6;PSMB7;PPP2CB;CCNB1;PSMB5;PSMB2;PSMB3;UBC;PSMB1;PSMF1;RPS27A;PSMB8;PSMC5;PSMC6;PSMC3;CDK4;PSMC4;PSMC1;RPA3;CDK2;PSMC2;MNAT1 |

| | | | | | | |
|---|---|---|---|---|---|---|
| 109 | Mitotic G1-G1/S phases Homo sapiens R-HSA-453279 | 1.26E-08 | 1.63E-07 | 2.76E+00 | 5.02E+01 | RB1;CDKN1B;MCM8;PPP2R2A;CKS1B;PSMD8;PSMD9;PSMD6;PSMD7;PSMD4;PSMD2;PSMD3;PSMD1;TK2;SKP1;PSMA5;PSMA6;PSMA3;PSMA4;TFDP2;PSMA1;PSMA2;PSME1;PSME2;PSMD10;PSMD12;PSMD11;PCNA;PSMD14;LIN37;PSMD13;HDAC1;CUL1;PSMA7;PPP2CA;PSMB6;PSMB7;PPP2CB;CCNB1;PSMB5;RBBP4;PSMB2;PSMB3;UBC;PSMB1;PSMF1;E2F4;RPS27A;CDKN2D;PSMB8;PSMC5;CDK6;PSMC6;PSMC3;CDK4;PSMC4;PSMC1;RPA3;CDK2;PSMC2;MNAT1 |
| 110 | S Phase Homo sapiens R-HSA-69242 | 1.26E-08 | 1.63E-07 | 2.88E+00 | 5.24E+01 | RB1;CDKN1B;MCM8;SMC3;CKS1B;PSMD8;PSMD9;PSMD6;PSMD7;PSMD4;PSMD2;PSMD3;PSMD1;SKP1;RFC1;SMC1A;CDC25B;PSMA5;PSMA6;PSMA3;PSMA4;PSMA1;PSMA2;PSME1;PSME2;PSMD10;PSMD12;PSMD11;PCNA;PSMD14;PSMD13;CUL1;PDS5B;PSMA7;PSMB6;PSMB7;PSMB5;PSMB2;PSMB3;RAD21;UBC;PSMB1;POLD2;PSMF1;RPS27A;PSMB8;PSMC5;STAG2;PSMC6;PSMC3;CDK4;PSMC4;PSMC1;RPA3;CDK2;PSMC2;MNAT1 |
| 111 | trans-Golgi Network Vesicle Budding Homo sapiens R-HSA-199992 | 1.27E-08 | 1.63E-07 | 4.02E+00 | 7.30E+01 | NAPA;APP;ARF1;RAB5C;SNAP23;CTSZ;CLTC;CLTB;AP4E1;CLTA;ARRB1;CLINT1;TGOLN2;BLOC1S6;SNX2;AP1G2;AP1G1;AP1S2;STX4;AP3S1;SNX5;AP1M2;TPD52;HSPA8;SNAPIN;SORT1;DTNBP1;M6PR;AP3B1;TPD52L1;AP4B1;DNM2;VAMP8;DNAJC6;YIPF6;CHMP2A;DNASE2;PICALM |
| 112 | Clathrin derived vesicle budding Homo sapiens R-HSA-421837 | 1.27E-08 | 1.63E-07 | 4.02E+00 | 7.30E+01 | NAPA;APP;ARF1;RAB5C;SNAP23;CTSZ;CLTC;CLTB;AP4E1;CLTA;ARRB1;CLINT1;TGOLN2;BLOC1S6;SNX2;AP1G2;AP1G1;AP1S2;STX4;AP3S1;SNX5;AP1M2;TPD52;HSPA8;SNAPIN;SORT1;DTNBP1;M6PR;AP3B1;TPD52L1;AP4B1;DNM2;VAMP8;DNAJC6;YIPF6;CHMP2A;DNASE2;PICALM |
| 113 | Ribosomal scanning and start codon recognition Homo sapiens R-HSA-72702 | 1.35E-08 | 1.70E-07 | 4.65E+00 | 8.42E+01 | EIF4A2;EIF4A1;RPS4Y1;RPS15;RPS19;EIF4H;RPS2;RPS27A;RPS10;EIF4E;RPS13;EIF4B;RPS12;RPS9;RPS8;RPS5;RPSA;EIF2S2;EIF2S1;EIF5;EIF3K;EIF3I;EIF3J;EIF3G;EIF3H;FAU;EIF3F;RPS21;EIF3D;EIF3A;RPS24;EIF4G1;RPS23 |
| 114 | Translation initiation complex formation Homo sapiens R-HSA-72649 | 1.35E-08 | 1.70E-07 | 4.65E+00 | 8.42E+01 | EIF4A2;EIF4A1;RPS4Y1;RPS15;RPS19;EIF4H;RPS2;RPS27A;RPS10;EIF4E;RPS13;EIF4B;RPS12;RPS9;RPS8;RPS5;RPSA;EIF2S2;EIF2S1;EIF3K;EIF3I;EIF3J;EIF3G;EIF3H;PABPC1;FAU;EIF3F;RPS21;EIF3D;EIF3A;RPS24;EIF4G1;RPS23 |
| 115 | DNA Replication Pre-Initiation Homo sapiens R-HSA-69002 | 2.16E-08 | 2.67E-07 | 3.55E+00 | 6.27E+01 | PSMD10;PSMD12;PSMD11;PSMD14;MCM8;PSMD13;GMNN;PSMA7;PSMD8;PSMB6;PSMD9;PSMB7;PSMD6;PSMD7;PSMB5;PSMD4;PSMB2;PSMB3;PSMD2;PSMD3;UBC;PSMB1;PSMD1;PSMF1;RPS27A;PSMB8;PSMA5;PSMA6;PSMA3;PSMC5;PSMA4;PSMC6;PSMA1;PSMC3;PSMA2;PSMC4;PSMC1;RPA3;PSMC2;CDK2;PSME1;PSME2 |

| | | | | | | |
|---|---|---|---|---|---|---|
| 116 | M/G1 Transition Homo sapiens R-HSA-68874 | 2.16E-08 | 2.67E-07 | 3.55E+00 | 6.27E+01 | PSMD10;PSMD12;PSMD11;PSMD14;MCM8;PSMD13;GMNN;PSMA7;PSMD8;PSMB6;PSMD9;PSMB7;PSMD6;PSMD7;PSMB5;PSMD4;PSMB2;PSMB3;PSMD2;PSMD3;UBC;PSMB1;PSMD1;PSMF1;RPS27A;PSMB8;PSMA5;PSMA6;PSMA3;PSMC5;PSMA4;PSMC6;PSMA1;PSMC3;PSMA2;PSMC4;PSMC1;RPA3;PSMC2;CDK2;PSME1;PSME2 |
| 117 | Activation of the mRNA upon binding of the cap-binding complex and eIFs, and subsequent binding to 43S Homo sapiens R-HSA-72662 | 2.45E-08 | 3.00E-07 | 4.46E+00 | 7.82E+01 | EIF4A2;EIF4A1;RPS4Y1;RPS15;RPS19;EIF4H;RPS2;RPS27A;RPS10;EIF4E;RPS13;EIF4B;RPS12;RPS9;RPS8;RPS5;RPSA;EIF2S2;EIF2S1;EIF3K;EIF3I;EIF3J;EIF3G;EIF3H;PABPC1;FAU;EIF3F;RPS21;EIF3D;EIF3A;RPS24;EIF4G1;RPS23 |
| 118 | Signalling by NGF Homo sapiens R-HSA-166520 | 4.26E-08 | 5.17E-07 | 1.76E+00 | 2.98E+01 | ATF1;PEBP1;WDR83;FGF2;CRKL;PSMD8;PSMD9;PSMD6;PSMD7;PSMD4;AKT2;PSMD2;PSMD3;AKT1;PSMD1;PIP4K2C;AP2M1;IER3;MAP2K2;RBX1;BTC;TIAM1;PRKAR1A;AKAP9;PSME1;PSME2;TLN1;SQSTM1;PSMD10;SHC2;SPTBN5;PSMD12;PSMD11;PSMD14;MAGED1;PSMD13;MAPKAP1;CUL3;PDE1A;PDGFB;ITPR2;PIK3R3;RASAL1;PHB;RTN4;RASGRP3;RAP1B;APH1A;RAP1A;APH1B;IRAK1;PRKAR2B;PRKAR2A;UBC;PIP5K1B;PSMF1;NFKB1;PTK2;NFKBIA;PTPRA;AGO2;FGF18;MDM2;GRB2;CALM1;YWHAE;PRR5;GSK3B;GSK3A;CDKN1B;YWHAB;RASGRF2;CLTA;ARRB1;AATF;PCSK5;AP2A2;ARHGAP4;PPP2R5E;MCF2L;PEA15;MLST8;RAC1;JAK2;SPTAN1;JAK1;PSENEN;VAV3;DUSP3;ARHGEF12;DUSP1;PDPK1;ARHGEF16;ARHGEF18;PPP2R5A;DUSP8;RHOA;DNM2;PSMA5;PSMA6;PSMA3;PSMA4;CREB1;PSMA1;PSMA2;RASA1;AKT1S1;RAPGEF1;ARHGEF3;ARHGEF1;ARHGEF7;HBEGF;HDAC2;RALA;HDAC3;RALB;HDAC1;ADCY3;FURIN;PSMA7;ADCY5;PSMB6;PPP2CA;PSMB7;PPP2CB;PSMB5;PSMB2;PSMB3;PSMB1;AP2S1;MAPK1;CSK;FYN;RPS27A;CAMK2G;KIDINS220;BAD;STAT3;DAB2IP;PTPN11;AP2B1;PSMB8;MTOR;MAPK13;PSMC5;PSMC6;PSMC3;PSMC4;PSMC1;PSMC2;RPS6KB2;KRAS;LAMTOR3 |
| 119 | EPH-Ephrin signaling Homo sapiens R-HSA-2682334 | 4.44E-08 | 5.34E-07 | 3.18E+00 | 5.38E+01 | ARPC1B;SDC2;ARPC1A;CLTC;CLTB;CLTA;WASL;SHB;AP2A2;EFNA4;MYL12A;ACTG1;MYL12B;APH1A;CDC42;SDCBP;PAK1;APH1B;CFL1;AP2S1;FYN;RAC1;PAK2;MYH10;AP2M1;ACTR3;PSENEN;VAV3;ACTR2;HSP90AA1;EPHA6;YES1;LIMK2;AP2B1;ARPC5;RHOA;PTK2;VEGFA;TIAM1;MYL6;EFNA3;ARPC2;ARPC3;RASA1;ARHGEF7;EPHA2 |

| | | | | | | |
|---|---|---|---|---|---|---|
| 120 | ER to Golgi Anterograde Transport Homo sapiens R-HSA-199977 | 5.01E-08 | 5.97E-07 | 2.69E+00 | 4.52E+01 | ARF4;MCFD2;ARF1;SERPINA1;KDELR1;CTSZ;MIA3;GOLGA2;PPP6C;TFG;TMED3;TMED2;TMED7;SPTAN1;NSF;TMED9;COG6;COG2;COPZ2;ANK2;ARFGAP3;COPZ1;TRAPPC6A;KDELR2;SEC22B;SEC22C;NAPA;DCTN6;COPB2;SPTBN5;TMED10;DCTN2;SAR1B;COPB1;RAB1B;GOSR2;USO1;DCTN3;NAPG;LMAN1;SCFD1;LMAN2;STX5;CD59;CD55;TRAPPC3;DYNC1H1;TRAPPC1;SEC24A;TRAPPC4;TRAPPC10;TRAPPC5;TRAPPC9;ARCN1;CNIH1;TMEM115;GOLGB1;COPE |
| 121 | Beta-catenin independent WNT signaling Homo sapiens R-HSA-3858494 | 6.25E-08 | 7.39E-07 | 2.56E+00 | 4.25E+01 | CLTC;CLTB;CLTA;AP2A2;PSMD8;PSMD9;PSMD6;PPP3CB;PSMD7;PSMD4;PSMD2;PSMD3;PSMD1;RAC1;AP2M1;RHOA;PSMA5;PSMA6;PSMA3;PSMA4;PSMA1;PSMA2;PSME1;PSME2;PFN1;PLCB1;PSMD10;PSMD12;PSMD11;PSMD14;PSMD13;ITPR2;PSMA7;PSMB6;PSMB7;GNG10;PARD6A;WNT11;PSMB5;GNG5;PSMB2;PSMB3;UBC;PSMB1;AP2S1;DVL3;PSMF1;RPS27A;TCF7L1;SMURF2;AP2B1;PSMB8;GNAO1;PSMC5;PSMC6;PSMC3;PSMC4;PSMC1;AGO2;PSMC2;GNB5;CALM1 |
| 122 | Mitotic Anaphase Homo sapiens R-HSA-68882 | 7.54E-08 | 8.85E-07 | 2.36E+00 | 3.87E+01 | ANKLE2;UBE2D1;PPP2R2A;SMC3;PSMD8;PPP1CC;PSMD9;PSMD6;PSMD7;PSMD4;PPP2R5E;CDC26;PSMD2;PSMD3;CDC27;PSMD1;TMPO;NUDC;UBE2E1;PPP2R5A;SMC1A;LEMD2;PSMA5;PSMA6;CLIP1;PSMA3;PSMA4;KIF2A;PSMA1;PSMA2;PSME1;PSME2;ANAPC5;MAPRE1;PAFAH1B1;PSMD10;PSMD12;ANAPC15;PSMD11;ANAPC16;PSMD14;PSMD13;PDS5B;PMF1;PSMA7;ANAPC11;SKA2;CENPC;PPP2CA;PSMB6;PSMB7;PPP2CB;PSMB5;PSMB2;PSMB3;RAD21;UBC;LMNA;PSMB1;PSMF1;BUB3;RPS27A;PSMB8;PSMC5;STAG2;PSMC6;PSMC3;PSMC4;PSMC1;TAOK1;PSMC2 |
| 123 | Synthesis of DNA Homo sapiens R-HSA-69239 | 9.84E-08 | 1.14E-06 | 3.05E+00 | 4.92E+01 | RB1;PSMD10;PSMD12;PSMD11;CDKN1B;PCNA;PSMD14;PSMD13;MCM8;PSMA7;PSMD8;PSMB6;PSMD9;PSMB7;PSMD6;PSMD7;PSMB5;PSMD4;PSMB2;PSMB3;PSMD2;PSMD3;UBC;PSMB1;POLD2;PSMD1;PSMF1;RPS27A;RFC1;PSMB8;PSMA5;PSMA6;PSMA3;PSMC5;PSMA4;PSMC6;PSMA1;PSMC3;PSMA2;PSMC4;PSMC1;RPA3;CDK2;PSMC2;PSME1;PSME2 |

| | | | | | | |
|---|---|---|---|---|---|---|
| 124 | Mitotic Metaphase and Anaphase Homo sapiens R-HSA-2555396 | 9.91E-08 | 1.14E-06 | 2.34E+00 | 3.77E+01 | ANKLE2;UBE2D1;PPP2R2A;SMC3;PSMD8;PPP1CC;PSMD9;PSMD6;PSMD7;PSMD4;PPP2R5E;CDC26;PSMD2;PSMD3;CDC27;PSMD1;TMPO;NUDC;UBE2E1;PPP2R5A;SMC1A;LEMD2;PSMA5;PSMA6;CLIP1;PSMA3;PSMA4;KIF2A;PSMA1;PSMA2;PSME1;PSME2;ANAPC5;MAPRE1;PAFAH1B1;PSMD10;PSMD12;ANAPC15;PSMD11;ANAPC16;PSMD14;PSMD13;PDS5B;PMF1;PSMA7;ANAPC11;SKA2;CENPC;PPP2CA;PSMB6;PSMB7;PPP2CB;PSMB5;PSMB2;PSMB3;RAD21;UBC;LMNA;PSMB1;PSMF1;BUB3;RPS27A;PSMB8;PSMC5;STAG2;PSMC6;PSMC3;PSMC4;PSMC1;TAOK1;PSMC2 |
| 125 | TCR signaling Homo sapiens R-HSA-202403 | 1.05E-07 | 1.20E-06 | 2.76E+00 | 4.44E+01 | UBE2D2;UBE2D1;PSMD8;PSMD9;PSMD6;PSMD7;PSMD4;PSMD2;PSMD3;PSMD1;SKP1;FBXW11;PDPK1;PSMA5;PSMA6;PSMA3;PSMA4;PSMA1;PSMA2;PSME1;PSME2;UBE2V1;HLA-DQB1;PSMD10;PSMD12;PSMD11;PSMD14;PSMD13;CUL1;BCL10;PSMA7;PSMB6;PSMB7;PAK1;PSMB5;PSMB2;PSMB3;UBC;PSMB1;CSK;PSMF1;RPS27A;PAK2;VASP;NFKB1;PSMB8;NFKBIA;PSMC5;PSMC6;PSMC3;PSMC4;PSMC1;PSMC2 |
| 126 | TNFR2 non-canonical NF-kB pathway Homo sapiens R-HSA-5668541 | 1.41E-07 | 1.60E-06 | 3.05E+00 | 4.80E+01 | PSMD10;PSMD12;PSMD11;PSMD14;PSMD13;CUL1;TNF;PSMA7;PSMD8;PSMB6;PSMD9;PSMB7;PSMD6;PSMD7;PSMB5;PSMD4;PSMB2;PSMB3;PSMD2;PSMD3;UBC;PSMB1;PSMD1;PSMF1;RPS27A;SKP1;TNFRSF12A;FBXW11;TNFSF12;PSMB8;PSMA5;PSMA6;PSMA3;PSMC5;PSMA4;PSMC6;PSMA1;PSMC3;PSMA2;PSMC4;PSMC1;PSMC2;PSME1;PSME2;UBE2M |
| 127 | IRS-mediated signalling Homo sapiens R-HSA-112399 | 2.19E-07 | 2.46E-06 | 1.93E+00 | 2.97E+01 | PEBP1;WDR83;FGF2;PSMD8;PSMD9;PSMD6;PSMD7;PSMD4;AKT2;PSMD2;PSMD3;AKT1;PSMD1;MAP2K2;RBX1;BTC;RRAGA;RRAGB;AKAP9;PSME1;PSME2;TLN1;PSMD10;SPTBN5;PSMD12;SHC2;PSMD11;PSMD14;PSMD13;CUL3;PDGFB;RASAL1;PHB;RASGRP3;RAP1B;RAP1A;UBC;PSMF1;PTK2;PTPRA;RHEB;FGF18;GRB2;CALM1;EIF4G1;CAB39L;YWHAB;RASGRF2;PRKAG1;PRKAG2;ARRB1;PPP2R5E;PEA15;MLST8;JAK2;SPTAN1;JAK1;DUSP1;PDPK1;STRADB;PPP2R5A;DUSP8;PSMA5;PSMA6;PSMA3;PSMA4;PSMA1;PSMA2;RASA1;AKT1S1;HBEGF;PRKAA1;PSMA7;PPP2CA;PSMB6;PSMB7;PPP2CB;PSMB5;PSMB2;PSMB3;PSMB1;MAPK1;CSK;FYN;RPS27A;EIF4E;CAMK2G;EIF4B;DAB2IP;PTPN11;PSMB8;MTOR;PSMC5;PSMC6;PSMC3;PSMC4;PSMC1;PSMC2;KRAS;LAMTOR1;LAMTOR4;LAMTOR3;LAMTOR5 |

| | | | | | |
|---|---|---|---|---|---|
| 128 | Signaling by FGFR2 Homo sapiens R-HSA-5654738 | 2.36E-07 | 2.64E-06 | 1.80E+00 | 2.75E+01 | PEBP1;WDR83;FGF2;TIAL1;PSMD8;PSMD9;PSMD6;PSMD7;PSMD4;AKT2;PSMD2;PSMD3;AKT1;PSMD1;PIP4K2C;IER3;MAP2K2;GTF2F2;RBX1;BTC;PRKAR1A;AKAP9;PSME1;PSME2;TLN1;PSMD10;SPTBN5;PSMD12;SHC2;PSMD11;PSMD14;PSMD13;PDE1A;MAPKAP1;CUL3;PDGFB;PIK3R3;ITPR2;RASAL1;PHB;RASGRP3;RAP1B;RAP1A;PRKAR2B;PRKAR2A;UBC;PIP5K1B;PSMF1;PTK2;HNRNPM;PTPRA;HNRNPF;AGO2;FGF18;MDM2;GRB2;CALM1;PRR5;GSK3B;GSK3A;CDKN1B;YWHAB;RASGRF2;ARRB1;PPP2R5E;PEA15;MLST8;JAK2;SPTAN1;JAK1;FGFBP1;NCBP1;DUSP1;PDPK1;PPP2R5A;DUSP8;PSMA5;PSMA6;PSMA3;PSMA4;CREB1;PSMA1;PSMA2;RASA1;ESRP1;AKT1S1;HBEGF;ADCY3;CBL;PSMA7;ADCY5;PPP2CA;PSMB6;PSMB7;PPP2CB;PSMB5;PSMB2;PSMB3;POLR2B;POLR2C;PSMB1;POLR2E;MAPK1;CSK;FYN;POLR2H;RPS27A;POLR2J;CAMK2G;POLR2K;POLR2L;BAD;DAB2IP;PTPN11;PSMB8;MTOR;PSMC5;PSMC6;PSMC3;PSMC4;PSMC1;PSMC2;RPS6KB2;KRAS;LAMTOR3 |
| 129 | Signaling by EGFR Homo sapiens R-HSA-177929 | 2.82E-07 | 3.13E-06 | 1.80E+00 | 2.72E+01 | PEBP1;WDR83;FGF2;PSMD8;PSMD9;PSMD6;PSMD7;PSMD4;AKT2;PSMD2;PSMD3;AKT1;PSMD1;PIP4K2C;AP2M1;IER3;MAP2K2;RBX1;BTC;PRKAR1A;AKAP9;PSME1;PSME2;TLN1;STAM2;PSMD10;SPTBN5;PSMD12;SHC2;PSMD11;PSMD14;PSMD13;PDE1A;MAPKAP1;CUL3;PDGFB;PIK3R3;ITPR2;RASAL1;PHB;RASGRP3;RAP1B;RAP1A;PRKAR2B;PRKAR2A;UBC;PIP5K1B;PSMF1;STAM;PTK2;PTPRA;AGO2;FGF18;MDM2;GRB2;CALM1;PRR5;GSK3B;GSK3A;CDKN1B;SH3KBP1;YWHAB;RASGRF2;CLTA;ARRB1;AP2A2;PPP2R5E;PEA15;MLST8;JAK2;SPTAN1;JAK1;DUSP1;PDPK1;PPP2R5A;DUSP8;EPN1;PSMA5;PSMA6;PSMA3;PSMA4;CREB1;PSMA1;PSMA2;RASA1;AKT1S1;ARHGEF7;HBEGF;ADCY3;CBL;PSMA7;ADCY5;PPP2CA;PSMB6;CDC42;PSMB7;PPP2CB;PSMB5;PSMB2;PSMB3;PSMB1;AP2S1;MAPK1;CSK;FYN;RPS27A;EPS15;CAMK2G;BAD;DAB2IP;PTPN11;AP2B1;PSMB8;MTOR;PSMC5;PSMC6;PSMC3;PSMC4;PSMC1;PSMC2;RPS6KB2;KRAS;LAMTOR3 |
| 130 | Transport to the Golgi and subsequent modification Homo sapiens R-HSA-948021 | 2.95E-07 | 3.22E-06 | 2.33E+00 | 3.50E+01 | B4GALT3;ARF4;MCFD2;ARF1;SERPINA1;KDELR1;CTSZ;MIA3;GOLGA2;PPP6C;TFG;TMED3;TMED2;TMED7;SPTAN1;NSF;TMED9;COG6;COG2;COPZ2;ANK2;ARFGAP3;COPZ1;TRAPPC6A;KDELR2;SEC22B;SEC22C;NAPA;DCTN6;COPB2;SPTBN5;TMED10;DCTN2;SAR1B;COPB1;RAB1B;GOSR2;USO1;DCTN3;NAPG;LMAN1;FUT8;SCFD1;MAN2A2;LMAN2;ST3GAL4;MAN1C1;STX5;CD59;MGAT1;CD55;TRAPPC3;DYNC1H1;TRAPPC1;SEC24A;TRAPPC4;FUCA1;TRAPPC10;TRAPPC5;TRAPPC9;ARCN1;CNIH1;TMEM115;GOLGB1;MGAT4B;COPE |

| | | | | | | |
|---|---|---|---|---|---|---|
| 131 | Separation of Sister Chromatids Homo sapiens R-HSA-2467813 | 2.95E-07 | 3.22E-06 | 2.33E+00 | 3.50E+01 | UBE2D1;SMC3;PSMD8;PPP1CC;PSMD9;PSMD6;PSMD7;PSMD4;PPP2R5E;CDC26;PSMD2;PSMD3;CDC27;PSMD1;NUDC;UBE2E1;PPP2R5A;SMC1A;PSMA5;PSMA6;CLIP1;PSMA3;PSMA4;KIF2A;PSMA1;PSMA2;PSME1;PSME2;ANAPC5;MAPRE1;PAFAH1B1;PSMD10;PSMD12;ANAPC15;PSMD11;ANAPC16;PSMD14;PSMD13;PDS5B;PMF1;PSMA7;ANAPC11;SKA2;CENPC;PPP2CA;PSMB6;PSMB7;PPP2CB;PSMB5;PSMB2;PSMB3;RAD21;UBC;PSMB1;PSMF1;BUB3;RPS27A;PSMB8;PSMC5;STAG2;PSMC6;PSMC3;PSMC4;PSMC1;TAOK1;PSMC2 |
| 132 | Signaling by FGFR Homo sapiens R-HSA-190236 | 3.03E-07 | 3.28E-06 | 1.79E+00 | 2.68E+01 | PEBP1;WDR83;FGF2;TIAL1;PSMD8;PSMD9;PSMD6;PSMD7;PSMD4;AKT2;PSMD2;PSMD3;AKT1;PSMD1;PIP4K2C;IER3;MAP2K2;GTF2F2;RBX1;BTC;PRKAR1A;AKAP9;PSME1;PSME2;TLN1;PSMD10;SPTBN5;PSMD12;SHC2;PSMD11;PSMD14;PSMD13;PDE1A;MAPKAP1;CUL3;PDGFB;PIK3R3;ITPR2;RASAL1;PHB;RASGRP3;RAP1B;RAP1A;PRKAR2B;PRKAR2A;UBC;PIP5K1B;PSMF1;PTK2;HNRNPM;PTPRA;HNRNPF;AGO2;FGF18;MDM2;GRB2;CALM1;PRR5;GSK3B;GSK3A;CDKN1B;YWHAB;RASGRF2;ARRB1;PPP2R5E;PEA15;MLST8;JAK2;SPTAN1;JAK1;FGFBP1;NCBP1;DUSP1;PDPK1;PPP2R5A;DUSP8;PSMA5;PSMA6;PSMA3;PSMA4;CREB1;PSMA1;PSMA2;RASA1;ESRP1;AKT1S1;HBEGF;ADCY3;CBL;PSMA7;ADCY5;PPP2CA;PSMB6;PSMB7;PPP2CB;PSMB5;PSMB2;PSMB3;POLR2B;POLR2C;PSMB1;POLR2E;MAPK1;CSK;FYN;POLR2H;RPS27A;POLR2J;CAMK2G;POLR2K;POLR2L;GALNT3;BAD;DAB2IP;PTPN11;PSMB8;MTOR;PSMC5;PSMC6;PSMC3;PSMC4;PSMC1;PSMC2;RPS6KB2;KRAS;LAMTOR3 |
| 133 | mRNA Splicing - Minor Pathway Homo sapiens R-HSA-72165 | 3.09E-07 | 3.33E-06 | 4.26E+00 | 6.38E+01 | SF3B4;SF3B5;SF3B2;PDCD7;SF3B6;YBX1;SNRPD2;SNRPD1;POLR2B;POLR2C;POLR2E;SNRPD3;POLR2H;TXNL4A;SNRNP35;POLR2J;POLR2K;POLR2L;NCBP1;ZCRB1;GTF2F2;LSM2;SNRNP40;PRPF6;SNRPG;SRSF2;SNRNP48;SNRNP25;SNRPB |

| | | | | | | |
|---|---|---|---|---|---|---|
| 134 | Axon guidance Homo sapiens R-HSA-422475 | 3.71E-07 | 3.96E-06 | 1.64E+00 | 2.42E+01 | PITPNA;PEBP1;WDR83;FGF2;ACTG1;PSMD8;PSMD9;PSMD6;PSMD7;PSMD4;DPYSL2;PSMD2;PSMD3;CFL1;PSMD1;AP2M1;MAP2K2;EPHA6;CSNK2A2;ANK2;RBX1;BTC;TIAM1;MYL6;CACNB4;COL4A4;AKAP9;CSNK2B;COL4A3;PSME1;PSME2;TLN1;PFN1;EZR;EPHA2;PFN2;PSMD10;SHC2;SPTBN5;PSMD12;PSMD11;PSMD14;PSMD13;CUL3;SDC2;PDGFB;RASAL1;CACNA1D;PHB;CACNA1C;EFNA4;MYL12A;MYL12B;RASGRP3;RAP1B;APH1A;SDCBP;RAP1A;APH1B;UBC;PLXNA2;PSMF1;SRGAP3;MYH10;PLXNA4;VASP;PRNP;HSPA8;YES1;ARPC5;L1CAM;PTK2;DLG1;EFNA3;ARPC2;CDK5;ARPC3;PTPRA;FGF18;GRB2;CALM1;CD24;ITGB1;ROBO3;GSK3B;HSP90AB1;YWHAB;ARPC1B;RASGRF2;ARPC1A;CLTC;CLTB;CLTA;ARRB1;WASL;SHB;AP2A2;PPP2R5E;PEA15;RAC1;JAK2;SPTAN1;JAK1;ACTR3;PSENEN;VAV3;CAP1;ACTR2;HSP90AA1;ARHGEF12;DUSP1;RHOG;PPP2R5A;DUSP8;RHOC;RHOA;DNM2;PSMA5;PSMA6;PSMA3;PSMA4;CREB1;PSMA1;PSMA2;RASA1;ARHGEF7;DOCK1;HBEGF;NTN4;CRMP1;NTN1;PSMA7;PSMB6;CDC42;PPP2CA;PSMB7;PPP2CB;PAK1;PSMB5;PSMB2;PSMB3;GPC1;PSMB1;AP2S1;PAK6;MAPK1;CSK;FYN;RPS27A;PAK2;CAMK2G;SEMA4D;LIMK2;DAB2IP;RANBP9;PTPN11;AP2B1;PSMB8;VEGFA;PSMC5;PSMC6;PSMC3;PSMC4;PSMC1;PSMC2;KRAS;LAMTOR3 |
| 135 | Insulin receptor signalling cascade Homo sapiens R-HSA-74751 | 3.92E-07 | 4.15E-06 | 1.90E+00 | 2.81E+01 | PEBP1;WDR83;FGF2;PSMD8;PSMD9;PSMD6;PSMD7;PSMD4;AKT2;PSMD2;PSMD3;AKT1;PSMD1;MAP2K2;RBX1;BTC;RRAGA;RRAGB;AKAP9;PSME1;PSME2;TLN1;PSMD10;SPTBN5;PSMD12;SHC2;PSMD11;PSMD14;PSMD13;CUL3;PDGFB;RASAL1;PHB;RASGRP3;RAP1B;RAP1A;UBC;PSMF1;PTK2;PTPRA;RHEB;FGF18;GRB2;CALM1;EIF4G1;CAB39L;YWHAB;RASGRF2;PRKAG1;PRKAG2;ARRB1;PPP2R5E;PEA15;MLST8;JAK2;SPTAN1;JAK1;DUSP1;PDPK1;STRADB;PPP2R5A;DUSP8;PSMA5;PSMA6;PSMA3;PSMA4;PSMA1;PSMA2;RASA1;AKT1S1;HBEGF;PRKAA1;PSMA7;PPP2CA;PSMB6;PSMB7;PPP2CB;PSMB5;PSMB2;PSMB3;PSMB1;MAPK1;CSK;FYN;RPS27A;EIF4E;CAMK2G;EIF4B;DAB2IP;PTPN11;PSMB8;MTOR;PSMC5;PSMC6;PSMC3;PSMC4;PSMC1;PSMC2;KRAS;LAMTOR1;LAMTOR4;LAMTOR3;LAMTOR5 |

| | | | | | | |
|---|---|---|---|---|---|---|
| 136 | IGF1R signaling cascade Homo sapiens R-HSA-2428924 | 4.74E-07 | 4.91E-06 | 1.89E+00 | 2.75E+01 | PEBP1;WDR83;FGF2;PSMD8;PSMD9;PSMD6;PSMD7;PSMD4;AKT2;PSMD2;PSMD3;AKT1;PSMD1;MAP2K2;RBX1;BTC;RRAGA;RRAGB;AKAP9;PSME1;PSME2;TLN1;PSMD10;SPTBN5;PSMD12;SHC2;PSMD11;PSMD14;PSMD13;CUL3;PDGFB;RASAL1;PHB;RASGRP3;RAP1B;RAP1A;UBC;PSMF1;PTK2;PTPRA;RHEB;FGF18;GRB2;CALM1;EIF4G1;CAB39L;YWHAB;RASGRF2;PRKAG1;PRKAG2;ARRB1;PPP2R5E;PEA15;MLST8;JAK2;SPTAN1;JAK1;DUSP1;PDPK1;STRADB;PPP2R5A;DUSP8;PSMA5;PSMA6;PSMA3;PSMA4;PSMA1;PSMA2;RASA1;AKT1S1;HBEGF;PRKAA1;PSMA7;PPP2CA;PSMB6;PSMB7;PPP2CB;PSMB5;PSMB2;PSMB3;PSMB1;MAPK1;CSK;FYN;RPS27A;EIF4E;CAMK2G;EIF4B;DAB2IP;PTPN11;PSMB8;MTOR;PSMC5;PSMC6;PSMC3;PSMC4;PSMC1;PSMC2;KRAS;LAMTOR1;LAMTOR4;LAMTOR3;LAMTOR5 |
| 137 | Signaling by Type 1 Insulin-like Growth Factor 1 Receptor (IGF1R) Homo sapiens R-HSA-2404192 | 4.74E-07 | 4.91E-06 | 1.89E+00 | 2.75E+01 | PEBP1;WDR83;FGF2;PSMD8;PSMD9;PSMD6;PSMD7;PSMD4;AKT2;PSMD2;PSMD3;AKT1;PSMD1;MAP2K2;RBX1;BTC;RRAGA;RRAGB;AKAP9;PSME1;PSME2;TLN1;PSMD10;SPTBN5;PSMD12;SHC2;PSMD11;PSMD14;PSMD13;CUL3;PDGFB;RASAL1;PHB;RASGRP3;RAP1B;RAP1A;UBC;PSMF1;PTK2;PTPRA;RHEB;FGF18;GRB2;CALM1;EIF4G1;CAB39L;YWHAB;RASGRF2;PRKAG1;PRKAG2;ARRB1;PPP2R5E;PEA15;MLST8;JAK2;SPTAN1;JAK1;DUSP1;PDPK1;STRADB;PPP2R5A;DUSP8;PSMA5;PSMA6;PSMA3;PSMA4;PSMA1;PSMA2;RASA1;AKT1S1;HBEGF;PRKAA1;PSMA7;PPP2CA;PSMB6;PSMB7;PPP2CB;PSMB5;PSMB2;PSMB3;PSMB1;MAPK1;CSK;FYN;RPS27A;EIF4E;CAMK2G;EIF4B;DAB2IP;PTPN11;PSMB8;MTOR;PSMC5;PSMC6;PSMC3;PSMC4;PSMC1;PSMC2;KRAS;LAMTOR1;LAMTOR4;LAMTOR3;LAMTOR5 |
| 138 | IRS-related events triggered by IGF1R Homo sapiens R-HSA-2428928 | 4.74E-07 | 4.91E-06 | 1.89E+00 | 2.75E+01 | PEBP1;WDR83;FGF2;PSMD8;PSMD9;PSMD6;PSMD7;PSMD4;AKT2;PSMD2;PSMD3;AKT1;PSMD1;MAP2K2;RBX1;BTC;RRAGA;RRAGB;AKAP9;PSME1;PSME2;TLN1;PSMD10;SPTBN5;PSMD12;SHC2;PSMD11;PSMD14;PSMD13;CUL3;PDGFB;RASAL1;PHB;RASGRP3;RAP1B;RAP1A;UBC;PSMF1;PTK2;PTPRA;RHEB;FGF18;GRB2;CALM1;EIF4G1;CAB39L;YWHAB;RASGRF2;PRKAG1;PRKAG2;ARRB1;PPP2R5E;PEA15;MLST8;JAK2;SPTAN1;JAK1;DUSP1;PDPK1;STRADB;PPP2R5A;DUSP8;PSMA5;PSMA6;PSMA3;PSMA4;PSMA1;PSMA2;RASA1;AKT1S1;HBEGF;PRKAA1;PSMA7;PPP2CA;PSMB6;PSMB7;PPP2CB;PSMB5;PSMB2;PSMB3;PSMB1;MAPK1;CSK;FYN;RPS27A;EIF4E;CAMK2G;EIF4B;DAB2IP;PTPN11;PSMB8;MTOR;PSMC5;PSMC6;PSMC3;PSMC4;PSMC1;PSMC2;KRAS;LAMTOR1;LAMTOR4;LAMTOR3;LAMTOR5 |

| | | | | | | |
|---|---|---|---|---|---|---|
| 139 | Endosomal Sorting Complex Required For Transport (ESCRT) Homo sapiens R-HSA-917729 | 5.25E-07 | 5.41E-06 | 7.12E+00 | 1.03E+02 | TSG101;VPS4B;VPS4A;VPS37D;STAM;VPS37B;SNF8;UBC;CHMP2B;CHMP2A;CHMP3;CHMP4B;CHMP4A;RPS27A;VPS36;VPS25;STAM2;VPS28;CHMP5 |
| 140 | VEGFA-VEGFR2 Pathway Homo sapiens R-HSA-4420097 | 5.34E-07 | 5.46E-06 | 1.83E+00 | 2.64E+01 | CYFIP2;NCKAP1;PEBP1;WDR83;FGF2;PSMD8;PSMD9;PSMD6;PSMD7;PSMD4;AKT2;PSMD2;PSMD3;AKT1;PSMD1;MAP2K2;CYBA;RBX1;BTC;AKAP9;PSME1;PSME2;ELMO2;TLN1;PSMD10;SPTBN5;PSMD12;SHC2;PSMD11;PSMD14;PSMD13;MAPKAP1;CUL3;PDGFB;ITPR2;RASAL1;PHB;PRKCZ;RASGRP3;RAP1B;RAP1A;UBC;PSMF1;JUP;PTK2;PTPRA;FGF18;GRB2;CALM1;PRR5;YWHAB;RASGRF2;BRK1;HSPB1;ARRB1;SHB;PPP2R5E;PEA15;MLST8;RAC1;JAK2;SPTAN1;JAK1;VAV3;HSP90AA1;DUSP1;PDPK1;PPP2R5A;DUSP8;RHOA;PSMA5;PSMA6;PSMA3;PSMA4;PSMA1;PSMA2;RASA1;DOCK1;HBEGF;PSMA7;PPP2CA;PSMB6;CDC42;PSMB7;PPP2CB;PAK1;PSMB5;PSMB2;PSMB3;PSMB1;MAPK1;CSK;FYN;RPS27A;PAK2;WASF2;CAMK2G;DAB2IP;PSMB8;MTOR;VEGFA;MAPK13;PSMC5;PSMC6;PSMC3;ABI2;PSMC4;PSMC1;PSMC2;KRAS;LAMTOR3;BCAR1 |
| 141 | Nucleotide Excision Repair Homo sapiens R-HSA-5696398 | 6.02E-07 | 6.11E-06 | 2.71E+00 | 3.88E+01 | INO80B;PCNA;CETN2;XPA;COPS7B;YY1;TFPT;SUMO1;POLR2B;SUMO3;POLR2C;UBC;SUMO2;POLD2;POLR2E;POLK;POLR2H;RPS27A;POLR2J;POLR2K;HMGN1;POLR2L;USP7;UBE2I;RFC1;PARP1;MCRS1;ACTL6A;USP45;RAD23A;ACTR8;RAD23B;GTF2H4;RBX1;COPS4;DDB1;COPS3;XAB2;COPS6;COPS5;COPS2;RPA3;ERCC1;UBE2V2;PPIE;TCEA1;ERCC5;MNAT1 |
| 142 | DNA Replication Homo sapiens R-HSA-69306 | 6.10E-07 | 6.14E-06 | 2.74E+00 | 3.92E+01 | RB1;PSMD10;PSMD12;PSMD11;CDKN1B;PCNA;PSMD14;PSMD13;MCM8;GMNN;PSMA7;PSMD8;PSMB6;PSMD9;PSMB7;PSMD6;PSMD7;PSMB5;PSMD4;PSMB2;PSMB3;PSMD2;PSMD3;UBC;PSMB1;POLD2;PSMD1;PSMF1;RPS27A;RFC1;PSMB8;PSMA5;PSMA6;PSMA3;PSMC5;PSMA4;PSMC6;PSMA1;PSMC3;PSMA2;PSMC4;PSMC1;RPA3;CDK2;PSMC2;PSME1;PSME2 |

| | | | | | | |
|---|---|---|---|---|---|---|
| 143 | Metabolism of amino acids and derivatives Homo sapiens R-HSA-71291 | 6.30E-07 | 6.30E-06 | 1.80E+00 | 2.57E+01 | RPL5;RPL30;RPL3;EPRS;RPL8;RPL10A;SAT1;RPL6;GLS;PSMD8;RPS15;PSMD9;PSMD6;PSMD7;PSMD4;RPS19;RPL18A;PSMD2;PSMD3;PSMD1;GLUL;RPS10;RPS13;RPS12;RPS9;RPS8;SECISBP2;RPS5;MCCC1;SLC6A12;SARS;RPSA;KMO;PSME1;PSME2;RPL29;RPL28;ALDH7A1;PSMD10;PSMD12;PSMD11;PSMD14;PSMD13;SHMT1;RPS4Y1;OAZ1;LARS;AFMID;PSMF1;EEFSEC;PDHA1;GOT1;GOT2;PYCR2;NMRAL1;AIMP1;QDPR;GRHPR;QARS;ADI1;RPS21;RPS24;RPS23;HIBADH;RPLP1;AMD1;APIP;RPL7A;ENOPH1;HIBCH;ACAD8;ARG2;BHMT2;AMT;PSMA5;PSMA6;PSMA3;PSMA4;PSMA1;PSMA2;IVD;EEF1E1;ALDH9A1;RPL10;PDHB;PPM1K;HSD17B10;PSMA7;PAPSS1;PSMB6;PSMB7;PSMB5;PSMB2;PSMB3;PSMB1;RPL14;PCBD1;RPL15;RPS2;RPL18;RPS27A;RPL19;TXNRD1;GSR;RPL23A;PSMB8;PSMC5;PSMC6;PSMC3;PSMC4;PSMC1;MARS;PSMC2;NDUFAB1;FAU;RPL26L1 |
| 144 | Intra-Golgi and retrograde Golgi-to-ER traffic Homo sapiens R-HSA-6811442 | 8.22E-07 | 8.17E-06 | 2.18E+00 | 3.05E+01 | ARF4;SCOC;ARF1;SYS1;KDELR1;GCC2;RAB43;KIF5C;KIF5B;TMED3;ARFIP2;TMED2;KIF21A;KIFAP3;TMED7;NSF;TMED9;NAA30;COG6;COG2;PLA2G4A;COPZ2;ARFGAP3;COPZ1;KIF2A;KIFC2;PLIN3;KDELR2;NAA38;SEC22B;PAFAH1B3;PAFAH1B2;VAMP3;PAFAH1B1;NAPA;DCTN6;COPB2;NBAS;TMED10;TMF1;DCTN2;COPB1;RAB1B;GOSR2;DCTN3;ARL1;KLC1;NAPG;STX10;TGOLN2;CYTH3;MAN2A2;VPS51;KIF3A;VPS53;MAN1C1;STX5;RAB6A;CYTH1;DYNC1H1;RIC1;GALNT1;SURF4;M6PR;ARCN1;KIF26B;RAB18;TRIP11;RAB9A;COPE |
| 145 | Golgi Associated Vesicle Biogenesis Homo sapiens R-HSA-432722 | 9.00E-07 | 8.88E-06 | 3.92E+00 | 5.45E+01 | NAPA;ARF1;RAB5C;CLTC;AP4E1;CLTA;ARRB1;CLINT1;TGOLN2;BLOC1S6;SNX2;AP1G1;AP1S2;AP3S1;SNX5;AP1M2;TPD52;HSPA8;SNAPIN;SORT1;DTNBP1;AP3B1;TPD52L1;AP4B1;DNM2;VAMP8;DNAJC6;YIPF6;PICALM |
| 146 | Signaling by the B Cell Receptor (BCR) Homo sapiens R-HSA-983705 | 9.14E-07 | 8.96E-06 | 1.98E+00 | 2.76E+01 | PRR5;GSK3B;GSK3A;CDKN1B;SH3KBP1;FGF2;PSMD8;PSMD9;PSMD6;PSMD7;PSMD4;PPP2R5E;AKT2;PSMD2;PSMD3;MLST8;AKT1;PSMD1;PIP4K2C;IER3;SKP1;FBXW11;PDPK1;PPP2R5A;BTC;PSMA5;PSMA6;PSMA3;PSMA4;CREB1;PSMA1;PSMA2;AKT1S1;PSME1;ORAI1;PSME2;HBEGF;PSMD10;PSMD12;PSMD11;PSMD14;PSMD13;MAPKAP1;PDGFB;CUL1;PIK3R3;ITPR2;BCL10;CBL;PSMA7;RASGRP3;PPP2CA;PSMB6;PSMB7;PPP2CB;PSMB5;PSMB2;PSMB3;PSMB1;UBC;MAPK1;PIP5K1B;FYN;PSMF1;RPS27A;BAD;PTPN11;PSMB8;NFKB1;MTOR;NFKBIA;PSMC5;PSMC6;PSMC3;PSMC4;PSMC1;AGO2;PSMC2;FGF18;RPS6KB2;REL;MDM2;GRB2;PTPN6;KRAS;CALM1 |

| | | | | | | |
|---|---|---|---|---|---|---|
| 147 | Pink/Parkin Mediated Mitophagy Homo sapiens R-HSA-5205685 | 1.00E-06 | 9.68E-06 | 8.99E+00 | 1.24E+02 | TOMM40;TOMM20;TOMM22;PINK1;MAP1LC3B;MAP1LC3A;UBC;MFN1;TOMM7;MFN2;VDAC1;RPS27A;SQSTM1;TOMM5;ATG5;TOMM6 |
| 148 | Mitophagy Homo sapiens R-HSA-5205647 | 1.00E-06 | 9.68E-06 | 8.99E+00 | 1.24E+02 | TOMM40;TOMM20;TOMM22;PINK1;MAP1LC3B;MAP1LC3A;UBC;MFN1;TOMM7;MFN2;VDAC1;RPS27A;SQSTM1;TOMM5;ATG5;TOMM6 |
| 149 | NGF signalling via TRKA from the plasma membrane Homo sapiens R-HSA-187037 | 1.12E-06 | 1.07E-05 | 1.73E+00 | 2.37E+01 | ATF1;PEBP1;WDR83;FGF2;CRKL;PSMD8;PSMD9;PSMD6;PSMD7;PSMD4;AKT2;PSMD2;PSMD3;AKT1;PSMD1;PIP4K2C;AP2M1;IER3;MAP2K2;RBX1;BTC;PRKAR1A;AKAP9;PSME1;PSME2;TLN1;PSMD10;SPTBN5;PSMD12;SHC2;PSMD11;PSMD14;PSMD13;PDE1A;MAPKAP1;CUL3;PDGFB;PIK3R3;ITPR2;RASAL1;PHB;RASGRP3;RAP1B;RAP1A;PRKAR2B;PRKAR2A;UBC;PIP5K1B;PSMF1;PTK2;PTPRA;AGO2;FGF18;MDM2;GRB2;CALM1;PRR5;GSK3B;GSK3A;CDKN1B;YWHAB;RASGRF2;CLTA;ARRB1;AP2A2;PPP2R5E;PEA15;MLST8;JAK2;SPTAN1;JAK1;DUSP3;DUSP1;PDPK1;PPP2R5A;DUSP8;RHOA;DNM2;PSMA5;PSMA6;PSMA3;PSMA4;CREB1;PSMA1;PSMA2;RASA1;AKT1S1;RAPGEF1;HBEGF;RALA;RALB;ADCY3;PSMA7;ADCY5;PPP2CA;PSMB6;PSMB7;PPP2CB;PSMB5;PSMB2;PSMB3;PSMB1;AP2S1;MAPK1;CSK;FYN;RPS27A;CAMK2G;KIDINS220;BAD;STAT3;DAB2IP;PTPN11;AP2B1;PSMB8;MTOR;MAPK13;PSMC5;PSMC6;PSMC3;PSMC4;PSMC1;PSMC2;RPS6KB2;KRAS;LAMTOR3 |
| 150 | Signaling by VEGF Homo sapiens R-HSA-194138 | 1.16E-06 | 1.11E-05 | 1.79E+00 | 2.44E+01 | CYFIP2;NCKAP1;PEBP1;WDR83;FGF2;PSMD8;PSMD9;PSMD6;PSMD7;PSMD4;AKT2;PSMD2;PSMD3;AKT1;PSMD1;MAP2K2;CYBA;RBX1;BTC;AKAP9;PSME1;PSME2;ELMO2;TLN1;PSMD10;SPTBN5;PSMD12;SHC2;PSMD11;PSMD14;PSMD13;MAPKAP1;CUL3;PDGFB;ITPR2;RASAL1;PHB;PRKCZ;RASGRP3;RAP1B;RAP1A;UBC;PSMF1;JUP;PTK2;PTPRA;FGF18;GRB2;CALM1;PRR5;YWHAB;RASGRF2;BRK1;HSPB1;ARRB1;SHB;PPP2R5E;PEA15;MLST8;RAC1;JAK2;SPTAN1;JAK1;VAV3;HSP90AA1;DUSP1;PDPK1;PPP2R5A;DUSP8;RHOA;PSMA5;PSMA6;PSMA3;PSMA4;PSMA1;PSMA2;RASA1;DOCK1;HBEGF;PSMA7;PPP2CA;PSMB6;CDC42;PSMB7;PPP2CB;PAK1;PSMB5;PSMB2;PSMB3;PSMB1;MAPK1;CSK;FYN;RPS27A;PAK2;WASF2;CAMK2G;VEGFB;DAB2IP;PSMB8;MTOR;VEGFA;MAPK13;PSMC5;PSMC6;PSMC3;ABI2;PSMC4;PSMC1;PSMC2;KRAS;LAMTOR3;BCAR1 |

| | | | | | | |
|---|---|---|---|---|---|---|
| 151 | FCERI mediated NF-kB activation Homo sapiens R-HSA-2871837 | 1.59E-06 | 1.51E-05 | 2.58E+00 | 3.44E+01 | PSMD10;PSMD12;PSMD11;PSMD14;PSMD13;UBE2D2;CUL1;UBE2D1;BCL10;PSMA7;PSMD8;PSMB6;PSMD9;PSMB7;PSMD6;PSMD7;PSMB5;PSMD4;PSMB2;PSMB3;PSMD2;PSMD3;UBC;PSMB1;PSMD1;PSMF1;RPS27A;SKP1;FBXW11;PDPK1;NFKB1;PSMB8;NFKBIA;PSMA5;PSMA6;PSMA3;PSMC5;PSMA4;PSMC6;PSMA1;PSMC3;PSMA2;PSMC4;PSMC1;PSMC2;PSME1;PSME2;UBE2V1 |
| 152 | Cell Cycle Checkpoints Homo sapiens R-HSA-69620 | 1.68E-06 | 1.58E-05 | 2.12E+00 | 2.82E+01 | YWHAE;CDKN1B;YWHAB;MCM8;UBE2D1;PSMD8;BABAM1;PSMD9;PSMD6;PSMD7;HERC2;PSMD4;YWHAQ;SUMO1;CDC26;PSMD2;PSMD3;CDC27;UIMC1;PSMD1;YWHAG;YWHAH;UBE2E1;YWHAZ;RNF168;PSMA5;PSMA6;PSMA3;PSMA4;PSMA1;PSMA2;PSME1;UBE2V2;PSME2;ANAPC5;PSMD10;PSMD12;ANAPC15;PSMD11;ANAPC16;PSMD14;PSMD13;BRCC3;PSMA7;ANAPC11;PSMB6;PSMB7;CCNB1;PSMB5;PSMB2;PCBP4;PSMB3;UBC;PSMB1;PSMF1;BUB3;RPS27A;PSMB8;PSMC5;RAD50;PSMC6;PSMC3;PSMC4;PSMC1;RPA3;CDK2;PSMC2;MDM2;RAD17;RAD1 |
| 153 | COPI-mediated anterograde transport Homo sapiens R-HSA-6807878 | 1.70E-06 | 1.59E-05 | 3.05E+00 | 4.05E+01 | NAPA;DCTN6;ARF4;COPB2;ARF1;SPTBN5;TMED10;DCTN2;KDELR1;RAB1B;COPB1;GOSR2;USO1;DCTN3;NAPG;GOLGA2;TMED3;TMED2;STX5;CD59;TMED7;SPTAN1;CD55;NSF;DYNC1H1;TMED9;COG6;COG2;COPZ2;ANK2;ARFGAP3;COPZ1;ARCN1;TMEM115;GOLGB1;KDELR2;COPE |
| 154 | Deadenylation-dependent mRNA decay Homo sapiens R-HSA-429914 | 1.85E-06 | 1.72E-05 | 3.96E+00 | 5.23E+01 | EIF4A2;EIF4A1;EIF4A3;EXOSC7;EXOSC4;EXOSC3;EIF4E;DCPS;EIF4B;EXOSC1;PAIP1;CNOT6L;LSM1;LSM4;PATL1;LSM2;CNOT4;NT5C3B;LSM7;CNOT7;XRN1;CNOT11;CNOT2;CNOT3;PABPC1;DCP2;EIF4G1 |
| 155 | M Phase Homo sapiens R-HSA-68886 | 2.02E-06 | 1.86E-05 | 1.86E+00 | 2.44E+01 | RB1;ANKLE2;UBE2D1;PPP2R2A;SMC3;SMC4;PSMD8;GOLGA2;PSMD9;PPP1CC;PSMD6;PSMD7;PSMD4;PPP2R5E;CDC26;PSMD2;PSMD3;CDC27;ENSA;PSMD1;HIST1H2AC;TMPO;RAB2A;NUDC;H2AFZ;CSNK2A2;UBE2E1;PPP2R5A;SMC1A;LEMD2;NUP93;PSMA5;PSMA6;PSMA3;CLIP1;PSMA4;PSMA1;KIF2A;PSMA2;GORASP2;CNEP1R1;NUP50;CSNK2B;PSME1;PSME2;ANAPC5;MAPRE1;PAFAH1B1;MCPH1;PSMD10;PSMD12;SET;ANAPC15;PSMD11;ANAPC16;PSMD14;PSMD13;RAB1B;USO1;PDS5B;PMF1;PSMA7;ANAPC11;SKA2;CENPC;PPP2CA;PSMB6;PSMB7;PPP2CB;CCNB1;PSMB5;PSMB2;PSMB3;RAD21;PSMB1;LMNA;TPR;UBC;MAPK1;PSMF1;BUB3;RPS27A;H3F3A;NEK7;PSMB8;PSMC5;STAG2;PSMC6;PSMC3;PSMC4;PSMC1;TAOK1;PSMC2;NCAPD3;LPIN2 |

| | | | | | | |
|---|---|---|---|---|---|---|
| 156 | Influenza Infection Homo sapiens R-HSA-168254 | 2.19E-06 | 2.00E-05 | 2.27E+00 | 2.96E+01 | RPL5;RPL30;RPL3;RPLP1;CLTA;RPL8;RPL10A;RPL6;RPS15;RPL7A;RPS19;RPL18A;KPNA4;KPNA2;KPNA3;RPS10;RPS13;KPNA1;RPS12;RPS9;HSP90AA1;RPS8;RPS5;RPSA;GTF2F2;NUP93;DNAJC3;NUP50;RPL29;RPL28;SLC25A6;RPL10;RPS4Y1;POLR2B;POLR2C;TPR;RPL14;POLR2E;RPL15;RPS2;POLR2H;RPL18;RPS27A;POLR2J;POLR2K;RPL19;POLR2L;TGFB1;EIF2AK2;RPL23A;CALR;FAU;GRSF1;RPS21;RPS24;RAN;RPL26L1;RPS23;HSPA1A |
| 157 | Cell Cycle, Mitotic Homo sapiens R-HSA-69278 | 2.19E-06 | 2.00E-05 | 1.62E+00 | 2.11E+01 | RB1;ANKLE2;GMNN;UBE2D1;PPP2R2A;SMC3;SMC4;PSMD8;PPP1CB;GOLGA2;PSMD9;PPP1CC;PSMD6;PSMD7;PSMD4;PSMD2;CDC26;PSMD3;CDC27;PSMD1;PHLDA1;HIST1H2AC;TMPO;SKP1;RAB2A;FKBPL;FBXW11;CSNK2A2;UBE2E1;LEMD2;TUBA4A;CDC25B;NUP93;CLIP1;AKAP9;CSNK2B;PSME1;PSME2;ANAPC5;AJUBA;OPTN;PSMD10;PSMD12;SET;ANAPC15;PSMD11;ANAPC16;PSMD14;TUBGCP2;PSMD13;CUL1;PDS5B;PMF1;ANAPC11;SKA2;CCNB1;PRKAR2B;RAD21;TPR;UBC;PSMF1;H3F3A;NEK7;CDK6;STAG2;CDK4;CDK2;TUBGCP4;MNAT1;CEP57;YWHAE;CDKN1B;HSP90AB1;MCM8;CETN2;CKS1B;ACTR1A;PPP2R5E;ENSA;TK2;RAB8A;YWHAG;OFD1;NUDC;HSP90AA1;RFC1;H2AFZ;TUBB;PPP2R5A;SMC1A;TUBG1;HAUS1;PSMA5;PSMA6;PSMA3;PSMA4;PSMA1;TFDP2;KIF2A;PSMA2;GORASP2;CNEP1R1;NUP50;MAPRE1;PAFAH1B1;MCPH1;PCNA;LIN37;DCTN2;HDAC1;RAB1B;SSNA1;USO1;DCTN3;PSMA7;CENPC;PSMB6;PPP2CA;PSMB7;PPP2CB;PSMB5;PSMB2;RBBP4;HAUS8;PSMB3;HAUS7;PSMB1;LMNA;POLD2;MAPK1;E2F4;BUB3;RPS27A;CDKN2D;DYNC1H1;CDK11A;TUBB4B;CDK11B;PSMB8;PSMC5;PSMC6;PSMC3;PSMC4;PSMC1;TAOK1;RPA3;PSMC2;NCAPD3;LPIN2 |
| 158 | rRNA processing Homo sapiens R-HSA-72312 | 2.28E-06 | 2.05E-05 | 2.11E+00 | 2.74E+01 | LTV1;RPL5;RPL30;RPL3;DDX49;RPLP1;RPL8;RPL10A;WDR46;RPL6;WDR43;RRP9;RPS15;FBL;TRMT112;RPL7A;RPS19;RPL18A;RIOK1;RPS10;RPP38;RPS13;RPS12;RPS9;RPS8;IMP3;DIMT1;RPS5;RPSA;DDX52;UTP18;DKC1;XRN2;C1D;RPP21;NHP2;RPP25;RPL29;RPL28;RPL10;RPS27L;RPS4Y1;HSD17B10;EXOSC7;EMG1;EXOSC4;BMS1;RPL14;RRP36;RPL15;RPS2;EXOSC3;RPL18;RPS27A;EXOSC1;DCAF13;RPL19;NOP14;KRR1;RIOK3;WDR18;RPL23A;FAU;RPS21;NOP10;RPS24;RPL26L1;RPS23;MPHOSPH6 |

| | | | | | | |
|---|---|---|---|---|---|---|
| 159 | Signaling by SCF-KIT Homo sapiens R-HSA-1433557 | 2.30E-06 | 2.05E-05 | 1.76E+00 | 2.29E+01 | PEBP1;WDR83;FGF2;PSMD8;PSMD9;PSMD6;PSMD7;PSMD4;AKT2;PSMD2;PSMD3;AKT1;PSMD1;PIP4K2C;IER3;MAP2K2;RBX1;BTC;AKAP9;PSME1;PSME2;TLN1;PSMD10;GRB7;SPTBN5;PSMD12;SHC2;PSMD11;PSMD14;PSMD13;MAPKAP1;CUL3;PDGFB;PIK3R3;RASAL1;PHB;RASGRP3;RAP1B;RAP1A;SOCS1;UBC;PIP5K1B;PSMF1;YES1;PTK2;PTPRA;AGO2;FGF18;MDM2;GRB2;CALM1;PRR5;GSK3B;GSK3A;CDKN1B;YWHAB;RASGRF2;ARRB1;PPP2R5E;PEA15;MLST8;RAC1;JAK2;SPTAN1;JAK1;DUSP1;PDPK1;PPP2R5A;DUSP8;PSMA5;PSMA6;PSMA3;PSMA4;CREB1;PSMA1;PSMA2;RASA1;AKT1S1;HBEGF;CBL;PSMA7;PPP2CA;PSMB6;PSMB7;PPP2CB;PSMB5;PSMB2;PSMB3;PSMB1;MAPK1;CSK;FYN;RPS27A;CAMK2G;SH2B2;BAD;STAT3;DAB2IP;PTPN11;PSMB8;MTOR;PSMC5;PSMC6;PSMC3;PSMC4;PSMC1;PSMC2;RPS6KB2;PTPN6;KRAS;LAMTOR3 |
| 160 | Nonsense-Mediated Decay (NMD) Homo sapiens R-HSA-927802 | 2.31E-06 | 2.05E-05 | 2.59E+00 | 3.37E+01 | RPL5;RPL30;RPL3;RBM8A;RPL10;RPLP1;EIF4A3;PPP2R2A;RPL8;RPL10A;RPS4Y1;RPL6;SMG5;RPS15;PPP2CA;RPL7A;RPS19;RPL18A;MAGOH;RPL14;RPL15;RPS2;RPL18;RPS27A;RPS10;RPS13;RPS12;RPL19;RPS9;NCBP1;RPS8;RPS5;RPSA;UPF3A;RPL23A;RNPS1;ETF1;PABPC1;FAU;RPL29;RPL28;RPS21;RPS24;RPL26L1;EIF4G1;RPS23 |
| 161 | Nonsense Mediated Decay (NMD) enhanced by the Exon Junction Complex (EJC) Homo sapiens R-HSA-975957 | 2.31E-06 | 2.05E-05 | 2.59E+00 | 3.37E+01 | RPL5;RPL30;RPL3;RBM8A;RPL10;RPLP1;EIF4A3;PPP2R2A;RPL8;RPL10A;RPS4Y1;RPL6;SMG5;RPS15;PPP2CA;RPL7A;RPS19;RPL18A;MAGOH;RPL14;RPL15;RPS2;RPL18;RPS27A;RPS10;RPS13;RPS12;RPL19;RPS9;NCBP1;RPS8;RPS5;RPSA;UPF3A;RPL23A;RNPS1;ETF1;PABPC1;FAU;RPL29;RPL28;RPS21;RPS24;RPL26L1;EIF4G1;RPS23 |

| | | | | | | |
|---|---|---|---|---|---|---|
| 162 | Developmental Biology Homo sapiens R-HSA-1266738 | 2.48E-06 | 2.19E-05 | 1.46E+00 | 1.88E+01 | PITPNA;CCNC;WDR83;RBPJ;PSMD8;PSMD9;PSMD6;PSMD7;PSMD4;DPYSL2;AKT2;PSMD2;PSMD3;AKT1;PSMD1;PKNOX1;EPHA6;CSNK2A2;MED8;ANK2;MED4;DKK1;BTC;TIAM1;MYL6;CSNK2B;AKAP9;PSME1;HOXB4;PSME2;HOXB2;TLN1;PFN1;EZR;EPHA2;PFN2;SPTBN5;PAXIP1;HOXD1;SDC2;PDGFB;MYL12A;MYL12B;APH1A;APH1B;PSMF1;VASP;YES1;H3F3A;L1CAM;PTK2;GRB2;HOXD3;CALM1;CD24;TDGF1;ROBO3;RARG;ARPC1B;ARPC1A;CLTC;CLTB;CLTA;ASH2L;CTCF;WASL;SHB;AP2A2;MED11;MED10;PEA15;RAC1;JAK2;SPTAN1;JAK1;CAP1;PSENEN;VAV3;NCOA2;DUSP1;NCOA3;RHOG;DUSP8;MED27;RHOC;SREBF2;RHOA;PSMA5;NCOR2;PSMA6;PSMA3;PSMA4;MED24;NCOR1;PSMA1;PSMA2;MED21;RARB;ARHGEF7;HBEGF;ADAM22;PSMA7;PSMB6;PPP2CA;PSMB7;PPP2CB;PAK1;PSMB5;PSMB2;PSMB3;GPC1;PSMB1;AP2S1;PAK6;RPS27A;PAK2;SUZ12;PCGF2;STAT3;DAB2IP;RANBP9;AP2B1;PSMB8;GCK;VEGFA;PSMC5;PSMC6;PSMC3;PSMC4;PSMC1;PSMC2;LAMTOR3;FOXA3;PEBP1;TNF;FGF2;ACTG1;YY1;CFL1;AP2M1;HIST1H2AC;MAP2K2;RBX1;CACNB4;COL4A4;COL4A3;AJUBA;PSMD10;SHC2;PSMD12;PSMD11;PSMD14;PSMD13;CUL3;CACNA1D;RASAL1;CACNA1C;PHB;EFNA4;RASGRP3;RAP1B;SDCBP;RAP1A;MED30;UBC;PLXNA2;SRGAP3;MYH10;PLXNA4;PRNP;HSPA8;SMAD4;TGFB1;ARPC5;MAFA;NFKB1;DRAP1;DLG1;EFNA3;ARPC2;CDK5;ARPC3;PTPRA;CDK4;FGF18;TCF4;ITGB1;GSK3B;HSP90AB1;YWHAB;RASGRF2;ARRB1;PAGR1;PPP2R5E;BOC;ACTR3;ACTR2;HSP90AA1;ARHGEF12;H2AFZ;PPP2R5A;D [illegible] |
| 163 | Signaling by ERBB4 Homo sapiens R-HSA-1236394 | 2.91E-06 | 2.55E-05 | 1.74E+00 | 2.22E+01 | PEBP1;WDR83;FGF2;PSMD8;PSMD9;PSMD6;PSMD7;PSMD4;AKT2;PSMD2;PSMD3;AKT1;PSMD1;PIP4K2C;IER3;SKP1;MAP2K2;RBX1;BTC;AKAP9;PSME1;PSME2;TLN1;PSMD10;SPTBN5;PSMD12;SHC2;PSMD11;PSMD14;PSMD13;MAPKAP1;CUL3;PDGFB;CUL1;PIK3R3;RASAL1;PHB;RASGRP3;APH1A;RAP1B;APH1B;RAP1A;UBC;PIP5K1B;PSMF1;PTK2;PTPRA;AGO2;FGF18;MDM2;GRB2;CALM1;PRR5;GSK3B;GSK3A;CDKN1B;YWHAB;RASGRF2;ARRB1;PPP2R5E;PEA15;MLST8;JAK2;SPTAN1;JAK1;PSENEN;DUSP1;PDPK1;PPP2R5A;DUSP8;PSMA5;PSMA6;PSMA3;NCOR1;PSMA4;CREB1;PSMA1;PSMA2;RASA1;AKT1S1;HBEGF;PSMA7;PPP2CA;PSMB6;PSMB7;PPP2CB;PSMB5;PSMB2;PSMB3;PSMB1;MAPK1;CSK;FYN;RPS27A;CAMK2G;WWOX;BAD;DAB2IP;PTPN11;PSMB8;MTOR;PSMC5;ITCH;PSMC6;PSMC3;PSMC4;PSMC1;NEDD4;PSMC2;RPS6KB2;KRAS;LAMTOR3 |

| | | | | | | |
|---|---|---|---|---|---|---|
| 164 | Downstream signal transduction Homo sapiens R-HSA-186763 | 3.04E-06 | 2.66E-05 | 1.73E+00 | 2.20E+01 | PEBP1;WDR83;FGF2;CRKL;PSMD8;PSMD9;PSMD6;PSMD7;PSMD4;AKT2;PSMD2;PSMD3;AKT1;PSMD1;PIP4K2C;IER3;MAP2K2;RBX1;BTC;PRKAR1A;AKAP9;PSME1;PSME2;TLN1;PSMD10;GRB7;SPTBN5;PSMD12;SHC2;PSMD11;PSMD14;PSMD13;PDE1A;MAPKAP1;CUL3;PDGFB;PIK3R3;ITPR2;RASAL1;PHB;RASGRP3;RAP1B;RAP1A;PRKAR2B;PRKAR2A;UBC;PIP5K1B;PSMF1;PTK2;PTPRA;AGO2;FGF18;MDM2;GRB2;CALM1;PRR5;GSK3B;GSK3A;CDKN1B;YWHAB;RASGRF2;ARRB1;PPP2R5E;PEA15;MLST8;JAK2;SPTAN1;JAK1;DUSP1;PDPK1;PPP2R5A;DUSP8;PSMA5;PSMA6;PSMA3;PSMA4;CREB1;PSMA1;PSMA2;RASA1;AKT1S1;RAPGEF1;HBEGF;ADCY3;PSMA7;ADCY5;PPP2CA;PSMB6;PSMB7;PPP2CB;PSMB5;PSMB2;PSMB3;PSMB1;MAPK1;CSK;FYN;RPS27A;CAMK2G;BAD;STAT3;DAB2IP;PTPN11;PSMB8;MTOR;PSMC5;PSMC6;PSMC3;PSMC4;PSMC1;PSMC2;RPS6KB2;KRAS;LAMTOR3;BCAR1 |
| 165 | Signaling by Wnt Homo sapiens R-HSA-195721 | 3.14E-06 | 2.72E-05 | 1.79E+00 | 2.27E+01 | VPS29;PSMD8;PSMD9;FRAT1;PSMD6;PSMD7;PSMD4;AKT2;PSMD2;PSMD3;AKT1;PSMD1;VPS35;HIST1H2AC;AP2M1;SKP1;RNF43;USP8;CSNK2A2;CTNNBIP1;DKK1;RBX1;PYGO1;CSNK2B;PSME1;PSME2;PFN1;CSNK1G2;PSMD10;PSMD12;PSMD11;PSMD14;PSMD13;CUL3;CUL1;ITPR2;VPS26A;SNX3;GNG10;UBC;DVL3;PIP5K1B;PSMF1;SMURF2;H3F3A;GNAO1;AGO2;TCF4;GNB5;CALM1;GSK3B;CLTC;USP34;CLTB;CBY1;CLTA;ASH2L;AP2A2;PPP3CB;PPP2R5E;RAC1;TMED5;H2AFZ;PPP2R5A;YWHAZ;RHOA;PSMA5;PSMA6;PSMA3;PSMA4;PSMA1;PSMA2;PLCB1;HDAC1;PSMA7;PPP2CA;PSMB6;PSMB7;PPP2CB;PSMB5;PARD6A;WNT11;PSMB2;GNG5;TNKS2;PSMB3;PSMB1;AP2S1;LEO1;RPS27A;TCF7L1;CSNK1A1;WNT7B;AP2B1;PSMB8;PSMC5;PSMC6;PSMC3;PSMC4;PSMC1;PSMC2;LGR5 |

| | | | | | | |
|---|---|---|---|---|---|---|
| 166 | Cell Cycle Homo sapiens R-HSA-1640170 | 3.61E-06 | 3.09E-05 | 1.54E+00 | 1.93E+01 | RB1;ANKLE2;GMNN;UBE2D1;PPP2R2A;SMC3;SMC4;PSMD8;PPP1CB;GOLGA2;PSMD9;PPP1CC;PSMD6;PSMD7;HERC2;PSMD4;PSMD2;CDC26;PSMD3;CDC27;PSMD1;PHLDA1;HIST1H2AC;TMPO;SKP1;RAB2A;FKBPL;FBXW11;CSNK2A2;TINF2;UBE2E1;LEMD2;TUBA4A;CDC25B;NUP93;CLIP1;DKC1;AKAP9;CSNK2B;PSME1;NHP2;UBE2V2;PSME2;ANAPC5;AJUBA;OPTN;PSMD10;PSMD12;SET;ANAPC15;PSMD11;ANAPC16;PSMD14;TUBGCP2;PSMD13;CUL1;BRCC3;PDS5B;PMF1;ANAPC11;SKA2;CCNB1;PRKAR2B;PCBP4;RAD21;TPR;UBC;PSMF1;UBE2I;H3F3A;NEK7;SMARCA5;HSPA2;CDK6;STAG2;CDK4;CDK2;MDM2;RAD17;TUBGCP4;MNAT1;CEP57;YWHAE;CDKN1B;HSP90AB1;MCM8;YWHAB;CETN2;RSF1;SYNE2;CKS1B;BABAM1;ACTR1A;SUMO1;YWHAQ;PPP2R5E;ENSA;UIMC1;TK2;RAB8A;YWHAG;YWHAH;OFD1;NUDC;HSP90AA1;RFC1;H2AFZ;TUBB;PPP2R5A;SMC1A;TUBG1;YWHAZ;HAUS1;PSMA5;RNF168;PSMA6;PSMA3;PSMA4;PSMA1;TFDP2;KIF2A;PSMA2;GORASP2;CNEP1R1;NUP50;TERF2IP;MAPRE1;PAFAH1B1;MCPH1;PCNA;LIN37;DCTN2;HDAC1;RAB1B;SSNA1;USO1;DCTN3;PSMA7;CENPC;PSMB6;PPP2CA;PSMB7;PPP2CB;PSMB5;PSMB2;RBBP4;HAUS8;PSMB3;HAUS7;PSMB1;LMNA;POLD2;MAPK1;RBBP7;E2F4;BUB3;RPS27A;CDKN2D;DYNC1H1;NPM1;CDK11A;TUBB4B;CDK11B;PSMB8;PSMC5;RAD50;PSMC6;PSMC3;PSMC4;PSMC1;TAOK1;RPA3;PSMC2;RAD1;NCAPD3;LPIN2 |
| 167 | Eukaryotic Translation Elongation Homo sapiens R-HSA-156842 | 3.61E-06 | 3.09E-05 | 2.76E+00 | 3.46E+01 | RPL5;RPL30;RPL3;RPL10;RPLP1;RPL10A;RPL8;RPS4Y1;RPL6;EEF1B2;RPS15;RPL7A;RPL18A;RPS19;RPL14;RPL15;RPS2;RPL18;RPS27A;RPS10;RPS13;RPS12;RPL19;RPS9;RPS8;RPS5;RPSA;RPL23A;EEF2;EEF1A1;EEF1G;EEF1D;EEF1A2;FAU;RPL29;RPL28;RPS21;RPL26L1;RPS24;RPS23 |
| 168 | Citric acid cycle (TCA cycle) Homo sapiens R-HSA-71403 | 3.84E-06 | 3.27E-05 | 9.44E+00 | 1.18E+02 | FH;MDH2;IDH3G;IDH2;SDHC;SDHD;SDHA;SDHB;SUCLA2;NNT;IDH3B;SUCLG2;SUCLG1;IDH3A |
| 169 | Translocation of GLUT4 to the plasma membrane Homo sapiens R-HSA-1445148 | 4.16E-06 | 3.52E-05 | 3.38E+00 | 4.18E+01 | YWHAE;RALA;STXBP3;YWHAB;SNAP23;PRKAG1;PRKAG2;YWHAQ;KIF3A;AKT2;AKT1;STX4;RAC1;KIFAP3;YWHAG;RAB8A;YWHAH;RAB4A;ASPSCR1;MYO5A;LNPEP;YWHAZ;RAB11A;RAB10;RAB14;RAB13;MYH8;EXOC6;CALM1;RHOQ |

| | | | | | | |
|---|---|---|---|---|---|---|
| 170 | Signaling by FGFR3 Homo sapiens R-HSA-5654741 | 4.69E-06 | 3.94E-05 | 1.72E+00 | 2.11E+01 | PEBP1;WDR83;FGF2;PSMD8;PSMD9;PSMD6;PSMD7;PSMD4;AKT2;PSMD2;PSMD3;AKT1;PSMD1;PIP4K2C;IER3;MAP2K2;RBX1;BTC;PRKAR1A;AKAP9;PSME1;PSME2;TLN1;PSMD10;SPTBN5;PSMD12;SHC2;PSMD11;PSMD14;PSMD13;PDE1A;MAPKAP1;CUL3;PDGFB;PIK3R3;ITPR2;RASAL1;PHB;RASGRP3;RAP1B;RAP1A;PRKAR2B;PRKAR2A;UBC;PIP5K1B;PSMF1;PTK2;PTPRA;AGO2;FGF18;MDM2;GRB2;CALM1;PRR5;GSK3B;GSK3A;CDKN1B;YWHAB;RASGRF2;ARRB1;PPP2R5E;PEA15;MLST8;JAK2;SPTAN1;JAK1;DUSP1;PDPK1;PPP2R5A;DUSP8;PSMA5;PSMA6;PSMA3;PSMA4;CREB1;PSMA1;PSMA2;RASA1;AKT1S1;HBEGF;ADCY3;CBL;PSMA7;ADCY5;PPP2CA;PSMB6;PSMB7;PPP2CB;PSMB5;PSMB2;PSMB3;PSMB1;MAPK1;CSK;FYN;RPS27A;CAMK2G;GALNT3;BAD;DAB2IP;PTPN11;PSMB8;MTOR;PSMC5;PSMC6;PSMC3;PSMC4;PSMC1;PSMC2;RPS6KB2;KRAS;LAMTOR3 |
| 171 | Formation of a pool of free 40S subunits Homo sapiens R-HSA-72689 | 4.87E-06 | 4.07E-05 | 2.63E+00 | 3.22E+01 | RPL5;RPL30;RPL3;RPL10;RPLP1;RPL10A;RPL8;RPS4Y1;RPL6;RPS15;RPL7A;RPL18A;RPS19;RPL14;RPL15;RPS2;RPL18;RPS27A;RPS10;RPS13;RPS12;RPL19;RPS9;RPS8;RPS5;RPSA;RPL23A;EIF3K;EIF3I;EIF3J;EIF3G;EIF3H;FAU;EIF3F;RPL29;RPL28;RPS21;EIF3D;RPL26L1;EIF3A;RPS24;RPS23 |
| 172 | Macroautophagy Homo sapiens R-HSA-1632852 | 6.99E-06 | 5.80E-05 | 3.09E+00 | 3.67E+01 | BECN1;PRKAA1;WDR45B;PRKAG1;PRKAG2;WDR45;GABARAP;MAP1LC3B;MAP1LC3A;ATG101;MLST8;ATG5;ATG3;GABARAPL2;WIPI1;WIPI2;MTOR;RRAGA;RRAGB;RHEB;CHMP2B;CHMP2A;CHMP4B;CHMP3;ULK1;ATG4A;CHMP4A;LAMTOR1;LAMTOR4;LAMTOR3;ATG4D;LAMTOR5 |
| 173 | Signaling by FGFR4 Homo sapiens R-HSA-5654743 | 7.01E-06 | 5.80E-05 | 1.71E+00 | 2.02E+01 | PEBP1;WDR83;FGF2;PSMD8;PSMD9;PSMD6;PSMD7;PSMD4;AKT2;PSMD2;PSMD3;AKT1;PSMD1;PIP4K2C;IER3;MAP2K2;RBX1;BTC;PRKAR1A;AKAP9;PSME1;PSME2;TLN1;PSMD10;SPTBN5;PSMD12;SHC2;PSMD11;PSMD14;PSMD13;PDE1A;MAPKAP1;CUL3;PDGFB;PIK3R3;ITPR2;RASAL1;PHB;RASGRP3;RAP1B;RAP1A;PRKAR2B;PRKAR2A;UBC;PIP5K1B;PSMF1;PTK2;PTPRA;AGO2;FGF18;MDM2;GRB2;CALM1;PRR5;GSK3B;GSK3A;CDKN1B;YWHAB;RASGRF2;ARRB1;PPP2R5E;PEA15;MLST8;JAK2;SPTAN1;JAK1;DUSP1;PDPK1;PPP2R5A;DUSP8;PSMA5;PSMA6;PSMA3;PSMA4;CREB1;PSMA1;PSMA2;RASA1;AKT1S1;HBEGF;ADCY3;CBL;PSMA7;ADCY5;PPP2CA;PSMB6;PSMB7;PPP2CB;PSMB5;PSMB2;PSMB3;PSMB1;MAPK1;CSK;FYN;RPS27A;CAMK2G;BAD;DAB2IP;PTPN11;PSMB8;MTOR;PSMC5;PSMC6;PSMC3;PSMC4;PSMC1;PSMC2;RPS6KB2;KRAS;LAMTOR3 |

| | | | | | | |
|---|---|---|---|---|---|---|
| 174 | Formation of the ternary complex, and subsequently, the 43S complex Homo sapiens R-HSA-72695 | 7.20E-06 | 5.92E-05 | 3.66E+00 | 4.33E+01 | RPS4Y1;RPS15;RPS19;RPS2;RPS27A;RPS10;RPS13;RPS12;RPS9;RPS8;RPS5;RPSA;EIF2S2;EIF2S1;EIF3K;EIF3I;EIF3J;EIF3G;EIF3H;FAU;EIF3F;RPS21;EIF3D;EIF3A;RPS24;RPS23 |
| 175 | Signaling by TGF-beta Receptor Complex Homo sapiens R-HSA-170834 | 7.32E-06 | 5.99E-05 | 2.94E+00 | 3.48E+01 | HDAC1;UBE2D3;UBE2D1;CCNC;NEDD8;FURIN;CBL;PRKCZ;PPP1CB;PPP1CC;PARD6A;UBC;E2F4;RPS27A;SKIL;SMAD4;TGFB1;PARP1;SMURF2;ARHGEF18;CGN;RHOA;UCHL5;PPP1CA;NCOR2;CDK9;FKBP1A;NCOR1;SNW1;TFDP2;BAMBI;STRAP;STUB1;UBE2M |
| 176 | Glucose metabolism Homo sapiens R-HSA-70326 | 7.41E-06 | 6.03E-05 | 2.83E+00 | 3.34E+01 | SLC25A1;GPI;PYGB;PFKFB3;ADPGK;ENO1;ENO2;ENO3;HK1;PPP2CA;PPP2CB;UBC;PHKG2;PGK1;RPS27A;PGM1;TPI1;GOT1;MDH1;MDH2;PGAM1;GAA;PGAM2;GOT2;PHKB;GCK;PFKL;PKM;GNPDA1;PC;ALDOC;ALDOA;CALM1;SLC25A11;GAPDH;PFKP |
| 177 | TCF dependent signaling in response to WNT Homo sapiens R-HSA-201681 | 7.56E-06 | 6.10E-05 | 1.96E+00 | 2.31E+01 | GSK3B;USP34;CBY1;ASH2L;PSMD8;PSMD9;FRAT1;PSMD6;PSMD7;PSMD4;PPP2R5E;AKT2;PSMD2;PSMD3;AKT1;PSMD1;HIST1H2AC;RNF43;USP8;H2AFZ;CSNK2A2;CTNNBIP1;PPP2R5A;DKK1;YWHAZ;RBX1;PSMA5;PYGO1;PSMA6;PSMA3;PSMA4;PSMA1;PSMA2;CSNK2B;PSME1;PSME2;CSNK1G2;PSMD10;PSMD12;PSMD11;PSMD14;PSMD13;HDAC1;CUL3;PSMA7;PPP2CA;PSMB6;PSMB7;PPP2CB;PSMB5;PSMB2;TNKS2;PSMB3;PSMB1;UBC;LEO1;DVL3;PIP5K1B;PSMF1;RPS27A;TCF7L1;SMURF2;CSNK1A1;H3F3A;PSMB8;PSMC5;PSMC6;PSMC3;PSMC4;PSMC1;PSMC2;TCF4;LGR5 |
| 178 | Downstream signaling of activated FGFR2 Homo sapiens R-HSA-5654696 | 7.68E-06 | 6.10E-05 | 1.71E+00 | 2.01E+01 | PEBP1;WDR83;FGF2;PSMD8;PSMD9;PSMD6;PSMD7;PSMD4;AKT2;PSMD2;PSMD3;AKT1;PSMD1;PIP4K2C;IER3;MAP2K2;RBX1;BTC;PRKAR1A;AKAP9;PSME1;PSME2;TLN1;PSMD10;SPTBN5;PSMD12;SHC2;PSMD11;PSMD14;PSMD13;PDE1A;MAPKAP1;CUL3;PDGFB;PIK3R3;ITPR2;RASAL1;PHB;RASGRP3;RAP1B;RAP1A;PRKAR2B;PRKAR2A;UBC;PIP5K1B;PSMF1;PTK2;PTPRA;AGO2;FGF18;MDM2;GRB2;CALM1;PRR5;GSK3B;GSK3A;CDKN1B;YWHAB;RASGRF2;ARRB1;PPP2R5E;PEA15;MLST8;JAK2;SPTAN1;JAK1;DUSP1;PDPK1;PPP2R5A;DUSP8;PSMA5;PSMA6;PSMA3;PSMA4;CREB1;PSMA1;PSMA2;RASA1;AKT1S1;HBEGF;ADCY3;PSMA7;ADCY5;PPP2CA;PSMB6;PSMB7;PPP2CB;PSMB5;PSMB2;PSMB3;PSMB1;MAPK1;CSK;FYN;RPS27A;CAMK2G;BAD;DAB2IP;PTPN11;PSMB8;MTOR;PSMC5;PSMC6;PSMC3;PSMC4;PSMC1;PSMC2;RPS6KB2;KRAS;LAMTOR3 |

| | | | | | | |
|---|---|---|---|---|---|---|
| 179 | Downstream signaling of activated FGFR4 Homo sapiens R-HSA-5654716 | 7.68E-06 | 6.10E-05 | 1.71E+00 | 2.01E+01 | PEBP1;WDR83;FGF2;PSMD8;PSMD9;PSMD6;PSMD7;PSMD4;AKT2;PSMD2;PSMD3;AKT1;PSMD1;PIP4K2C;IER3;MAP2K2;RBX1;BTC;PRKAR1A;AKAP9;PSME1;PSME2;TLN1;PSMD10;SPTBN5;PSMD12;SHC2;PSMD11;PSMD14;PSMD13;PDE1A;MAPKAP1;CUL3;PDGFB;PIK3R3;ITPR2;RASAL1;PHB;RASGRP3;RAP1B;RAP1A;PRKAR2B;PRKAR2A;UBC;PIP5K1B;PSMF1;PTK2;PTPRA;AGO2;FGF18;MDM2;GRB2;CALM1;PRR5;GSK3B;GSK3A;CDKN1B;YWHAB;RASGRF2;ARRB1;PPP2R5E;PEA15;MLST8;JAK2;SPTAN1;JAK1;DUSP1;PDPK1;PPP2R5A;DUSP8;PSMA5;PSMA6;PSMA3;PSMA4;CREB1;PSMA1;PSMA2;RASA1;AKT1S1;HBEGF;ADCY3;PSMA7;ADCY5;PPP2CA;PSMB6;PSMB7;PPP2CB;PSMB5;PSMB2;PSMB3;PSMB1;MAPK1;CSK;FYN;RPS27A;CAMK2G;BAD;DAB2IP;PTPN11;PSMB8;MTOR;PSMC5;PSMC6;PSMC3;PSMC4;PSMC1;PSMC2;RPS6KB2;KRAS;LAMTOR3 |
| 180 | Downstream signaling of activated FGFR3 Homo sapiens R-HSA-5654708 | 7.68E-06 | 6.10E-05 | 1.71E+00 | 2.01E+01 | PEBP1;WDR83;FGF2;PSMD8;PSMD9;PSMD6;PSMD7;PSMD4;AKT2;PSMD2;PSMD3;AKT1;PSMD1;PIP4K2C;IER3;MAP2K2;RBX1;BTC;PRKAR1A;AKAP9;PSME1;PSME2;TLN1;PSMD10;SPTBN5;PSMD12;SHC2;PSMD11;PSMD14;PSMD13;PDE1A;MAPKAP1;CUL3;PDGFB;PIK3R3;ITPR2;RASAL1;PHB;RASGRP3;RAP1B;RAP1A;PRKAR2B;PRKAR2A;UBC;PIP5K1B;PSMF1;PTK2;PTPRA;AGO2;FGF18;MDM2;GRB2;CALM1;PRR5;GSK3B;GSK3A;CDKN1B;YWHAB;RASGRF2;ARRB1;PPP2R5E;PEA15;MLST8;JAK2;SPTAN1;JAK1;DUSP1;PDPK1;PPP2R5A;DUSP8;PSMA5;PSMA6;PSMA3;PSMA4;CREB1;PSMA1;PSMA2;RASA1;AKT1S1;HBEGF;ADCY3;PSMA7;ADCY5;PPP2CA;PSMB6;PSMB7;PPP2CB;PSMB5;PSMB2;PSMB3;PSMB1;MAPK1;CSK;FYN;RPS27A;CAMK2G;BAD;DAB2IP;PTPN11;PSMB8;MTOR;PSMC5;PSMC6;PSMC3;PSMC4;PSMC1;PSMC2;RPS6KB2;KRAS;LAMTOR3 |

| | | | | | | |
|---|---|---|---|---|---|---|
| 181 | Post-translational protein modification Homo sapiens R-HSA-597592 | 8.70E-06 | 6.88E-05 | 1.54E+00 | 1.79E+01 | DPAGT1;SEMA5B;PGAP1;CMAS;TUSC3;KDELR1;GALNT18;SMC3;GOLGA2;C1GALT1C1;HERC2;TFG;NUDT14;PDIA3;NSF;ALG8;COG6;ALG5;COG2;ALG2;ALG3;MOGS;ANK2;NUP93;DPM2;DPM3;KDELR2;COPB2;SPTBN5;TMED10;RPN2;SAR1B;COPB1;SEL1L;SCFD1;FUT8;MAN2A2;RAD21;ARSJ;TPR;DPH3;NEU1;ST3GAL4;MLEC;ST3GAL6;DPH7;MPDU1;EDEM3;CTSA;TRAPPC3;PHC2;ICMT;TRAPPC1;UBE2I;CBX4;FUCA1;TRAPPC4;EDEM2;TRAPPC5;GFPT1;DOLK;TRAPPC9;EEF2;HNRNPK;CNIH1;STAG2;TMEM115;GOLGB1;SP3;MDM2;UBA2;HNRNPC;CALR;SAE1;MUC20;COPE;ARF4;B4GALT3;ARF1;MCFD2;SERPINA1;PIGT;CETN2;PIGP;CTSZ;MIA3;PIGX;GMPPB;PPP6C;SENP5;SUMO1;SUMO3;ADAMTSL2;SUMO2;TMED3;GNPNAT1;TMED2;MUC15;TMED7;SPTAN1;GALNT9;EIF5A;TMED9;PARP1;DHPS;COPZ2;SMC1A;ARFGAP3;POMGNT1;COPZ1;DDOST;RNF168;TRAPPC6A;NSMCE4A;DAD1;NUP50;PIGK;B3GNT3;B3GNT2;TOP1;GAS6;SEC22B;PIGF;PIGH;ST6GALNAC6;SEC22C;NAPA;DCTN6;SP100;PCNA;DCTN2;RAB1B;GOSR2;USO1;DCTN3;DERL2;FURIN;NAPG;LMAN1;MUC1;OS9;LMAN2;TDG;SLC17A5;MAN1C1;STX5;CD59;MGAT1;CD55;SLC35A1;DYNC1H1;GALNT6;SEC24A;GALNT3;PCGF2;TRAPPC10;GALNT1;C1GALT1;DOHH;ARCN1;GLB1;MGAT4B |
| 182 | Major pathway of rRNA processing in the nucleolus Homo sapiens R-HSA-6791226 | 8.96E-06 | 7.04E-05 | 2.07E+00 | 2.41E+01 | LTV1;RPL5;RPL30;RPL3;DDX49;RPLP1;RPL8;RPL10A;WDR46;RPL6;WDR43;RRP9;RPS15;FBL;RPL7A;RPS19;RPL18A;RIOK1;RPS10;RPP38;RPS13;RPS12;RPS9;RPS8;IMP3;RPS5;RPSA;DDX52;UTP18;XRN2;C1D;RPP21;RPP25;RPL29;RPL28;RPL10;RPS27L;RPS4Y1;EXOSC7;EMG1;EXOSC4;BMS1;RPL14;RRP36;RPL15;RPS2;EXOSC3;RPL18;RPS27A;EXOSC1;DCAF13;RPL19;NOP14;KRR1;RIOK3;WDR18;RPL23A;FAU;RPS21;RPS24;RPL26L1;RPS23;MPHOSPH6 |
| 183 | Prefoldin mediated transfer of substrate to CCT/TriC Homo sapiens R-HSA-389957 | 9.80E-06 | 7.66E-05 | 5.73E+00 | 6.61E+01 | CCT3;CCT2;VBP1;TUBB4B;TUBA4A;CCT6A;TUBA1C;TUBB6;TCP1;PFDN1;PFDN2;CCT8;CCT7;PFDN4;PFDN5;CCT5;CCT4 |
| 184 | Glycolysis Homo sapiens R-HSA-70171 | 1.00E-05 | 7.79E-05 | 4.93E+00 | 5.67E+01 | GPI;PFKFB3;TPI1;PGAM1;PGAM2;ENO1;ENO2;ENO3;GCK;HK1;PPP2CA;PPP2CB;PFKL;PKM;PGK1;ALDOC;ALDOA;GAPDH;PFKP |

| | | | | | | |
|---|---|---|---|---|---|---|
| 185 | Prolonged ERK activation events Homo sapiens R-HSA-169893 | 1.02E-05 | 7.86E-05 | 1.84E+00 | 2.11E+01 | YWHAB;RASGRF2;PEBP1;WDR83;ARRB1;FGF2;CRKL;PSMD8;PSMD9;PSMD6;PSMD7;PSMD4;PPP2R5E;PSMD2;PSMD3;PEA15;PSMD1;JAK2;SPTAN1;JAK1;MAP2K2;DUSP1;PPP2R5A;DUSP8;RBX1;BTC;PSMA5;PSMA6;PSMA3;PSMA4;PSMA1;PSMA2;RASA1;AKAP9;PSME1;RAPGEF1;PSME2;TLN1;HBEGF;PSMD10;SPTBN5;PSMD12;SHC2;PSMD11;PSMD14;PSMD13;CUL3;PDGFB;RASAL1;PHB;PSMA7;RASGRP3;PPP2CA;PSMB6;RAP1B;PSMB7;PPP2CB;PSMB5;RAP1A;PSMB2;PSMB3;PSMB1;UBC;MAPK1;CSK;FYN;PSMF1;RPS27A;CAMK2G;KIDINS220;DAB2IP;PTK2;PSMB8;PSMC5;PSMC6;PSMC3;PSMC4;PTPRA;PSMC1;PSMC2;FGF18;GRB2;KRAS;CALM1;LAMTOR3 |
| 186 | Selenoamino acid metabolism Homo sapiens R-HSA-2408522 | 1.03E-05 | 7.93E-05 | 2.39E+00 | 2.75E+01 | RPL5;RPL30;RPL3;RPL10;RPLP1;EPRS;RPL8;RPL10A;RPS4Y1;RPL6;PAPSS1;RPS15;RPL7A;RPS19;RPL18A;LARS;RPL14;RPL15;RPS2;RPL18;RPS27A;RPS10;RPS13;RPS12;RPL19;EEFSEC;RPS9;RPS8;RPS5;SECISBP2;TXNRD1;GSR;SARS;RPSA;RPL23A;AIMP1;QARS;MARS;FAU;RPL29;EEF1E1;RPL28;RPS21;RPS24;RPL26L1;RPS23 |
| 187 | G2/M Checkpoints Homo sapiens R-HSA-69481 | 1.04E-05 | 7.94E-05 | 2.13E+00 | 2.45E+01 | YWHAE;YWHAB;MCM8;PSMD8;BABAM1;PSMD9;PSMD6;PSMD7;HERC2;PSMD4;YWHAQ;SUMO1;PSMD2;PSMD3;UIMC1;PSMD1;YWHAG;YWHAH;YWHAZ;RNF168;PSMA5;PSMA6;PSMA3;PSMA4;PSMA1;PSMA2;PSME1;UBE2V2;PSME2;PSMD10;PSMD12;PSMD11;PSMD14;PSMD13;BRCC3;PSMA7;PSMB6;PSMB7;CCNB1;PSMB5;PSMB2;PSMB3;UBC;PSMB1;PSMF1;RPS27A;PSMB8;PSMC5;RAD50;PSMC6;PSMC3;PSMC4;PSMC1;RPA3;CDK2;PSMC2;RAD17;RAD1 |
| 188 | Signalling to ERKs Homo sapiens R-HSA-187687 | 1.11E-05 | 8.43E-05 | 1.81E+00 | 2.06E+01 | YWHAB;RASGRF2;PEBP1;WDR83;ARRB1;FGF2;CRKL;PSMD8;PSMD9;PSMD6;PSMD7;PSMD4;PPP2R5E;PSMD2;PSMD3;PEA15;PSMD1;JAK2;SPTAN1;JAK1;MAP2K2;DUSP1;PPP2R5A;DUSP8;RBX1;BTC;PSMA5;PSMA6;PSMA3;PSMA4;PSMA1;PSMA2;RASA1;AKAP9;PSME1;RAPGEF1;PSME2;TLN1;HBEGF;PSMD10;SPTBN5;PSMD12;SHC2;RALA;PSMD11;RALB;PSMD14;PSMD13;CUL3;PDGFB;RASAL1;PHB;PSMA7;RASGRP3;PPP2CA;PSMB6;RAP1B;PSMB7;PPP2CB;PSMB5;RAP1A;PSMB2;PSMB3;PSMB1;UBC;MAPK1;CSK;FYN;PSMF1;RPS27A;CAMK2G;KIDINS220;DAB2IP;PTK2;PSMB8;MAPK13;PSMC5;PSMC6;PSMC3;PSMC4;PTPRA;PSMC1;PSMC2;FGF18;GRB2;KRAS;CALM1;LAMTOR3 |

| | | | | | | |
|---|---|---|---|---|---|---|
| 189 | Downstream signaling of activated FGFR1 Homo sapiens R-HSA-5654687 | 1.21E-05 | 8.97E-05 | 1.68E+00 | 1.90E+01 | PEBP1;WDR83;FGF2;PSMD8;PSMD9;PSMD6;PSMD7;PSMD4;AKT2;PSMD2;PSMD3;AKT1;PSMD1;PIP4K2C;IER3;MAP2K2;RBX1;BTC;PRKAR1A;AKAP9;PSME1;PSME2;TLN1;PSMD10;SPTBN5;PSMD12;SHC2;PSMD11;PSMD14;PSMD13;PDE1A;MAPKAP1;CUL3;PDGFB;PIK3R3;ITPR2;RASAL1;PHB;RASGRP3;RAP1B;RAP1A;PRKAR2B;PRKAR2A;UBC;PIP5K1B;PSMF1;PTK2;PTPRA;AGO2;FGF18;MDM2;GRB2;CALM1;PRR5;GSK3B;GSK3A;CDKN1B;YWHAB;RASGRF2;ARRB1;PPP2R5E;PEA15;MLST8;JAK2;SPTAN1;JAK1;DUSP1;PDPK1;PPP2R5A;DUSP8;PSMA5;PSMA6;PSMA3;PSMA4;CREB1;PSMA1;PSMA2;RASA1;AKT1S1;HBEGF;ADCY3;PSMA7;ADCY5;PPP2CA;PSMB6;PSMB7;PPP2CB;PSMB5;PSMB2;PSMB3;PSMB1;MAPK1;CSK;FYN;RPS27A;CAMK2G;BAD;DAB2IP;PTPN11;PSMB8;MTOR;PSMC5;PSMC6;PSMC3;PSMC4;PSMC1;PSMC2;RPS6KB2;KRAS;LAMTOR3 |
| 190 | FRS-mediated FGFR2 signaling Homo sapiens R-HSA-5654700 | 1.22E-05 | 8.97E-05 | 1.84E+00 | 2.08E+01 | YWHAB;RASGRF2;PEBP1;WDR83;ARRB1;FGF2;PSMD8;PSMD9;PSMD6;PSMD7;PSMD4;PPP2R5E;PSMD2;PSMD3;PEA15;PSMD1;JAK2;SPTAN1;JAK1;MAP2K2;DUSP1;PPP2R5A;DUSP8;RBX1;BTC;PSMA5;PSMA6;PSMA3;PSMA4;PSMA1;PSMA2;RASA1;AKAP9;PSME1;PSME2;TLN1;HBEGF;PSMD10;SPTBN5;PSMD12;SHC2;PSMD11;PSMD14;PSMD13;CUL3;PDGFB;RASAL1;PHB;PSMA7;RASGRP3;PPP2CA;PSMB6;RAP1B;PSMB7;PPP2CB;PSMB5;RAP1A;PSMB2;PSMB3;PSMB1;UBC;MAPK1;CSK;FYN;PSMF1;RPS27A;CAMK2G;DAB2IP;PTPN11;PTK2;PSMB8;PSMC5;PSMC6;PSMC3;PSMC4;PTPRA;PSMC1;PSMC2;FGF18;GRB2;KRAS;CALM1;LAMTOR3 |
| 191 | FRS-mediated FGFR4 signaling Homo sapiens R-HSA-5654712 | 1.22E-05 | 8.97E-05 | 1.84E+00 | 2.08E+01 | YWHAB;RASGRF2;PEBP1;WDR83;ARRB1;FGF2;PSMD8;PSMD9;PSMD6;PSMD7;PSMD4;PPP2R5E;PSMD2;PSMD3;PEA15;PSMD1;JAK2;SPTAN1;JAK1;MAP2K2;DUSP1;PPP2R5A;DUSP8;RBX1;BTC;PSMA5;PSMA6;PSMA3;PSMA4;PSMA1;PSMA2;RASA1;AKAP9;PSME1;PSME2;TLN1;HBEGF;PSMD10;SPTBN5;PSMD12;SHC2;PSMD11;PSMD14;PSMD13;CUL3;PDGFB;RASAL1;PHB;PSMA7;RASGRP3;PPP2CA;PSMB6;RAP1B;PSMB7;PPP2CB;PSMB5;RAP1A;PSMB2;PSMB3;PSMB1;UBC;MAPK1;CSK;FYN;PSMF1;RPS27A;CAMK2G;DAB2IP;PTPN11;PTK2;PSMB8;PSMC5;PSMC6;PSMC3;PSMC4;PTPRA;PSMC1;PSMC2;FGF18;GRB2;KRAS;CALM1;LAMTOR3 |

| | | | | | | |
|---|---|---|---|---|---|---|
| 192 | FRS-mediated FGFR3 signaling Homo sapiens R-HSA-5654706 | 1.22E-05 | 8.97E-05 | 1.84E+00 | 2.08E+01 | YWHAB;RASGRF2;PEBP1;WDR83;ARRB1;FGF2;PSMD8;PSMD9;PSMD6;PSMD7;PSMD4;PPP2R5E;PSMD2;PSMD3;PEA15;PSMD1;JAK2;SPTAN1;JAK1;MAP2K2;DUSP1;PPP2R5A;DUSP8;RBX1;BTC;PSMA5;PSMA6;PSMA3;PSMA4;PSMA1;PSMA2;RASA1;AKAP9;PSME1;PSME2;TLN1;HBEGF;PSMD10;SPTBN5;PSMD12;SHC2;PSMD11;PSMD14;PSMD13;CUL3;PDGFB;RASAL1;PHB;PSMA7;RASGRP3;PPP2CA;PSMB6;RAP1B;PSMB7;PPP2CB;PSMB5;RAP1A;PSMB2;PSMB3;PSMB1;UBC;MAPK1;CSK;FYN;PSMF1;RPS27A;CAMK2G;DAB2IP;PTPN11;PTK2;PSMB8;PSMC5;PSMC6;PSMC3;PSMC4;PTPRA;PSMC1;PSMC2;FGF18;GRB2;KRAS;CALM1;LAMTOR3 |
| 193 | FRS-mediated FGFR1 signaling Homo sapiens R-HSA-5654693 | 1.22E-05 | 8.97E-05 | 1.84E+00 | 2.08E+01 | YWHAB;RASGRF2;PEBP1;WDR83;ARRB1;FGF2;PSMD8;PSMD9;PSMD6;PSMD7;PSMD4;PPP2R5E;PSMD2;PSMD3;PEA15;PSMD1;JAK2;SPTAN1;JAK1;MAP2K2;DUSP1;PPP2R5A;DUSP8;RBX1;BTC;PSMA5;PSMA6;PSMA3;PSMA4;PSMA1;PSMA2;RASA1;AKAP9;PSME1;PSME2;TLN1;HBEGF;PSMD10;SPTBN5;PSMD12;SHC2;PSMD11;PSMD14;PSMD13;CUL3;PDGFB;RASAL1;PHB;PSMA7;RASGRP3;PPP2CA;PSMB6;RAP1B;PSMB7;PPP2CB;PSMB5;RAP1A;PSMB2;PSMB3;PSMB1;UBC;MAPK1;CSK;FYN;PSMF1;RPS27A;CAMK2G;DAB2IP;PTPN11;PTK2;PSMB8;PSMC5;PSMC6;PSMC3;PSMC4;PTPRA;PSMC1;PSMC2;FGF18;GRB2;KRAS;CALM1;LAMTOR3 |
| 194 | Signaling by Leptin Homo sapiens R-HSA-2586552 | 1.22E-05 | 8.97E-05 | 1.82E+00 | 2.06E+01 | YWHAB;RASGRF2;PEBP1;WDR83;ARRB1;FGF2;PSMD8;PSMD9;PSMD6;PSMD7;PSMD4;PPP2R5E;PSMD2;PSMD3;PEA15;LEPR;PSMD1;JAK2;SPTAN1;JAK1;MAP2K2;DUSP1;PPP2R5A;DUSP8;RBX1;BTC;PSMA5;PSMA6;PSMA3;PSMA4;PSMA1;PSMA2;RASA1;AKAP9;PSME1;PSME2;TLN1;HBEGF;PSMD10;SPTBN5;PSMD12;SHC2;PSMD11;PSMD14;PSMD13;CUL3;PDGFB;RASAL1;PHB;PSMA7;RASGRP3;PPP2CA;PSMB6;RAP1B;PSMB7;PPP2CB;PSMB5;RAP1A;PSMB2;PSMB3;PSMB1;UBC;MAPK1;CSK;FYN;PSMF1;RPS27A;CAMK2G;STAT3;DAB2IP;PTPN11;PTK2;PSMB8;PSMC5;PSMC6;PSMC3;PSMC4;PTPRA;PSMC1;PSMC2;FGF18;GRB2;KRAS;CALM1;LAMTOR3 |

| | | | | | | |
|---|---|---|---|---|---|---|
| 195 | Signaling by FGFR1 Homo sapiens R-HSA-5654736 | 1.28E-05 | 9.41E-05 | 1.67E+00 | 1.89E+01 | PEBP1;WDR83;FGF2;PSMD8;PSMD9;PSMD6;PSMD7;PSMD4;AKT2;PSMD2;PSMD3;AKT1;PSMD1;PIP4K2C;IER3;MAP2K2;RBX1;BTC;PRKAR1A;AKAP9;PSME1;PSME2;TLN1;PSMD10;SPTBN5;PSMD12;SHC2;PSMD11;PSMD14;PSMD13;PDE1A;MAPKAP1;CUL3;PDGFB;PIK3R3;ITPR2;RASAL1;PHB;RASGRP3;RAP1B;RAP1A;PRKAR2B;PRKAR2A;UBC;PIP5K1B;PSMF1;PTK2;PTPRA;AGO2;FGF18;MDM2;GRB2;CALM1;PRR5;GSK3B;GSK3A;CDKN1B;YWHAB;RASGRF2;ARRB1;PPP2R5E;PEA15;MLST8;JAK2;SPTAN1;JAK1;DUSP1;PDPK1;PPP2R5A;DUSP8;PSMA5;PSMA6;PSMA3;PSMA4;CREB1;PSMA1;PSMA2;RASA1;AKT1S1;HBEGF;ADCY3;CBL;PSMA7;ADCY5;PPP2CA;PSMB6;PSMB7;PPP2CB;PSMB5;PSMB2;PSMB3;PSMB1;MAPK1;CSK;FYN;RPS27A;CAMK2G;BAD;DAB2IP;PTPN11;PSMB8;MTOR;PSMC5;PSMC6;PSMC3;PSMC4;PSMC1;PSMC2;RPS6KB2;KRAS;LAMTOR3 |
| 196 | Lysosome Vesicle Biogenesis Homo sapiens R-HSA-432720 | 1.30E-05 | 9.48E-05 | 4.50E+00 | 5.06E+01 | APP;HSPA8;ARF1;CTSZ;CLTC;CLTB;AP4E1;M6PR;CLTA;ARRB1;AP4B1;DNM2;VAMP8;AP1G2;AP1G1;DNAJC6;AP1S2;CHMP2A;DNASE2;AP1M2 |
| 197 | Interleukin-3, 5 and GM-CSF signaling Homo sapiens R-HSA-512988 | 1.31E-05 | 9.48E-05 | 1.78E+00 | 2.01E+01 | YWHAB;RASGRF2;PEBP1;WDR83;ARRB1;FGF2;CRKL;PSMD8;PSMD9;PSMD6;PSMD7;PSMD4;PPP2R5E;PSMD2;PSMD3;PEA15;PSMD1;JAK2;SPTAN1;JAK1;MAP2K2;DUSP1;PPP2R5A;DUSP8;YWHAZ;RBX1;BTC;PSMA5;PSMA6;PSMA3;PSMA4;PSMA1;PSMA2;RASA1;AKAP9;PSME1;RAPGEF1;PSME2;TLN1;HBEGF;PSMD10;SPTBN5;PSMD12;SHC2;PSMD11;PSMD14;PSMD13;CUL3;PDGFB;PIK3R3;RASAL1;PHB;CBL;PSMA7;RASGRP3;PPP2CA;PSMB6;RAP1B;PSMB7;PPP2CB;PSMB5;RAP1A;PSMB2;PSMB3;PSMB1;UBC;MAPK1;CSK;FYN;PSMF1;RPS27A;CAMK2G;YES1;DAB2IP;PTPN11;PTK2;PSMB8;PSMC5;PSMC6;PSMC3;PSMC4;PTPRA;PSMC1;PSMC2;FGF18;GRB2;PTPN6;KRAS;CALM1;LAMTOR3 |
| 198 | Frs2-mediated activation Homo sapiens R-HSA-170968 | 1.33E-05 | 9.58E-05 | 1.82E+00 | 2.05E+01 | YWHAB;RASGRF2;PEBP1;WDR83;ARRB1;FGF2;CRKL;PSMD8;PSMD9;PSMD6;PSMD7;PSMD4;PPP2R5E;PSMD2;PSMD3;PEA15;PSMD1;JAK2;SPTAN1;JAK1;MAP2K2;DUSP1;PPP2R5A;DUSP8;RBX1;BTC;PSMA5;PSMA6;PSMA3;PSMA4;PSMA1;PSMA2;RASA1;AKAP9;PSME1;RAPGEF1;PSME2;TLN1;HBEGF;PSMD10;SPTBN5;PSMD12;SHC2;PSMD11;PSMD14;PSMD13;CUL3;PDGFB;RASAL1;PHB;PSMA7;RASGRP3;PPP2CA;PSMB6;RAP1B;PSMB7;PPP2CB;PSMB5;RAP1A;PSMB2;PSMB3;PSMB1;UBC;MAPK1;CSK;FYN;PSMF1;RPS27A;CAMK2G;DAB2IP;PTK2;PSMB8;PSMC5;PSMC6;PSMC3;PSMC4;PTPRA;PSMC1;PSMC2;FGF18;GRB2;KRAS;CALM1;LAMTOR3 |

| | | | | | | |
|---|---|---|---|---|---|---|
| 199 | Budding and maturation of HIV virion Homo sapiens R-HSA-162588 | 1.33E-05 | 9.58E-05 | 5.99E+00 | 6.73E+01 | TSG101;PDCD6IP;VPS4B;VPS4A;VPS37D;VPS37B;UBC;CHMP2B;CHMP2A;CHMP3;CHMP4B;CHMP4A;RPS27A;PPIA;VPS28;CHMP5 |
| 200 | DAP12 signaling Homo sapiens R-HSA-2424491 | 1.43E-05 | 1.02E-04 | 1.66E+00 | 1.85E+01 | PEBP1;WDR83;FGF2;PSMD8;PSMD9;PSMD6;PSMD7;PSMD4;AKT2;PSMD2;PSMD3;AKT1;PSMD1;PIP4K2C;IER3;MAP2K2;HLA-E;RBX1;BTC;PRKAR1A;AKAP9;PSME1;PSME2;TLN1;PSMD10;SPTBN5;PSMD12;SHC2;PSMD11;PSMD14;PSMD13;PDE1A;MAPKAP1;CUL3;PDGFB;PIK3R3;ITPR2;RASAL1;PHB;RASGRP3;RAP1B;RAP1A;PRKAR2B;PRKAR2A;UBC;PIP5K1B;PSMF1;PTK2;PTPRA;AGO2;FGF18;MDM2;GRB2;CALM1;PRR5;GSK3B;GSK3A;CDKN1B;YWHAB;RASGRF2;ARRB1;PPP2R5E;PEA15;MLST8;RAC1;JAK2;SPTAN1;JAK1;VAV3;DUSP1;PDPK1;PPP2R5A;DUSP8;PSMA5;PSMA6;PSMA3;PSMA4;CREB1;PSMA1;PSMA2;RASA1;AKT1S1;HBEGF;ADCY3;PSMA7;ADCY5;PPP2CA;PSMB6;PSMB7;PPP2CB;PSMB5;PSMB2;PSMB3;PSMB1;MAPK1;CSK;FYN;RPS27A;CAMK2G;BAD;DAB2IP;PTPN11;PSMB8;MTOR;PSMC5;PSMC6;PSMC3;PSMC4;PSMC1;PSMC2;RPS6KB2;KRAS;LAMTOR3 |
| 201 | VEGFR2 mediated cell proliferation Homo sapiens R-HSA-5218921 | 1.58E-05 | 1.12E-04 | 1.80E+00 | 1.99E+01 | YWHAB;RASGRF2;PEBP1;WDR83;ARRB1;FGF2;PSMD8;PSMD9;PSMD6;PSMD7;PSMD4;PPP2R5E;PSMD2;PSMD3;PEA15;PSMD1;JAK2;SPTAN1;JAK1;MAP2K2;DUSP1;PDPK1;PPP2R5A;DUSP8;RBX1;BTC;PSMA5;PSMA6;PSMA3;PSMA4;PSMA1;PSMA2;RASA1;AKAP9;PSME1;PSME2;TLN1;HBEGF;PSMD10;SPTBN5;PSMD12;SHC2;PSMD11;PSMD14;PSMD13;CUL3;PDGFB;ITPR2;RASAL1;PHB;PRKCZ;PSMA7;RASGRP3;PPP2CA;PSMB6;RAP1B;PSMB7;PPP2CB;PSMB5;RAP1A;PSMB2;PSMB3;PSMB1;UBC;MAPK1;CSK;FYN;PSMF1;RPS27A;CAMK2G;DAB2IP;PTK2;PSMB8;VEGFA;PSMC5;PSMC6;PSMC3;PSMC4;PTPRA;PSMC1;PSMC2;FGF18;GRB2;KRAS;CALM1;LAMTOR3 |

| | | | | | | |
|---|---|---|---|---|---|---|
| 202 | Signaling by PDGF Homo sapiens R-HSA-186797 | 1.85E-05 | 1.31E-04 | 1.63E+00 | 1.77E+01 | PEBP1;WDR83;FGF2;CRKL;PSMD8;PSMD9;PSMD6;PSMD7;PSMD4;AKT2;PSMD2;PSMD3;AKT1;PSMD1;PIP4K2C;IER3;MAP2K2;RBX1;BTC;PRKAR1A;COL4A4;AKAP9;COL4A3;PSME1;PSME2;TLN1;PSMD10;GRB7;SPTBN5;PSMD12;SHC2;PSMD11;PSMD14;PSMD13;PDE1A;MAPKAP1;CUL3;PDGFB;PIK3R3;ITPR2;RASAL1;PHB;RASGRP3;RAP1B;RAP1A;PRKAR2B;PRKAR2A;UBC;PIP5K1B;PSMF1;PTK2;PTPRA;AGO2;FGF18;MDM2;GRB2;CALM1;PRR5;GSK3B;GSK3A;CDKN1B;YWHAB;RASGRF2;ARRB1;PPP2R5E;PEA15;MLST8;JAK2;SPTAN1;JAK1;DUSP1;PDPK1;PPP2R5A;DUSP8;PSMA5;PSMA6;PSMA3;PSMA4;CREB1;PSMA1;PSMA2;RASA1;AKT1S1;RAPGEF1;HBEGF;ADCY3;FURIN;PSMA7;ADCY5;PPP2CA;PSMB6;PSMB7;PPP2CB;PSMB5;PSMB2;PSMB3;PSMB1;MAPK1;CSK;FYN;RPS27A;CAMK2G;BAD;STAT3;DAB2IP;PTPN11;PSMB8;MTOR;PSMC5;PSMC6;PSMC3;PSMC4;PSMC1;PSMC2;RPS6KB2;KRAS;LAMTOR3;BCAR1 |
| 203 | Influenza Life Cycle Homo sapiens R-HSA-168255 | 1.89E-05 | 1.31E-04 | 2.16E+00 | 2.35E+01 | RPL5;RPL30;RPL3;RPLP1;CLTA;RPL8;RPL10A;RPL6;RPS15;RPL7A;RPS19;RPL18A;RPS10;RPS13;KPNA1;RPS12;RPS9;HSP90AA1;RPS8;RPS5;RPSA;GTF2F2;NUP93;DNAJC3;NUP50;RPL29;RPL28;RPL10;RPS4Y1;POLR2B;POLR2C;TPR;RPL14;POLR2E;RPL15;RPS2;POLR2H;RPL18;RPS27A;POLR2J;POLR2K;RPL19;POLR2L;RPL23A;CALR;FAU;GRSF1;RPS21;RPS24;RAN;RPL26L1;RPS23;HSPA1A |
| 204 | GRB2 events in EGFR signaling Homo sapiens R-HSA-179812 | 1.90E-05 | 1.31E-04 | 1.82E+00 | 1.97E+01 | YWHAB;RASGRF2;PEBP1;WDR83;ARRB1;FGF2;PSMD8;PSMD9;PSMD6;PSMD7;PSMD4;PPP2R5E;PSMD2;PSMD3;PEA15;PSMD1;JAK2;SPTAN1;JAK1;MAP2K2;DUSP1;PPP2R5A;DUSP8;RBX1;BTC;PSMA5;PSMA6;PSMA3;PSMA4;PSMA1;PSMA2;RASA1;AKAP9;PSME1;PSME2;TLN1;HBEGF;PSMD10;SPTBN5;PSMD12;SHC2;PSMD11;PSMD14;PSMD13;CUL3;PDGFB;RASAL1;PHB;PSMA7;RASGRP3;PPP2CA;PSMB6;RAP1B;PSMB7;PPP2CB;PSMB5;RAP1A;PSMB2;PSMB3;PSMB1;UBC;MAPK1;CSK;FYN;PSMF1;RPS27A;CAMK2G;DAB2IP;PTK2;PSMB8;PSMC5;PSMC6;PSMC3;PSMC4;PTPRA;PSMC1;PSMC2;FGF18;GRB2;KRAS;CALM1;LAMTOR3 |

| | | | | | | |
|---|---|---|---|---|---|---|
| 205 | SHC1 events in EGFR signaling Homo sapiens R-HSA-180336 | 1.90E-05 | 1.31E-04 | 1.82E+00 | 1.97E+01 | YWHAB;RASGRF2;PEBP1;WDR83;ARRB1;FGF2;PSMD8;PSMD9;PSMD6;PSMD7;PSMD4;PPP2R5E;PSMD2;PSMD3;PEA15;PSMD1;JAK2;SPTAN1;JAK1;MAP2K2;DUSP1;PPP2R5A;DUSP8;RBX1;BTC;PSMA5;PSMA6;PSMA3;PSMA4;PSMA1;PSMA2;RASA1;AKAP9;PSME1;PSME2;TLN1;HBEGF;PSMD10;SPTBN5;PSMD12;SHC2;PSMD11;PSMD14;PSMD13;CUL3;PDGFB;RASAL1;PHB;PSMA7;RASGRP3;PPP2CA;PSMB6;RAP1B;PSMB7;PPP2CB;PSMB5;RAP1A;PSMB2;PSMB3;PSMB1;UBC;MAPK1;CSK;FYN;PSMF1;RPS27A;CAMK2G;DAB2IP;PTK2;PSMB8;PSMC5;PSMC6;PSMC3;PSMC4;PTPRA;PSMC1;PSMC2;FGF18;GRB2;KRAS;CALM1;LAMTOR3 |
| 206 | SOS-mediated signalling Homo sapiens R-HSA-112412 | 1.90E-05 | 1.31E-04 | 1.82E+00 | 1.97E+01 | YWHAB;RASGRF2;PEBP1;WDR83;ARRB1;FGF2;PSMD8;PSMD9;PSMD6;PSMD7;PSMD4;PPP2R5E;PSMD2;PSMD3;PEA15;PSMD1;JAK2;SPTAN1;JAK1;MAP2K2;DUSP1;PPP2R5A;DUSP8;RBX1;BTC;PSMA5;PSMA6;PSMA3;PSMA4;PSMA1;PSMA2;RASA1;AKAP9;PSME1;PSME2;TLN1;HBEGF;PSMD10;SPTBN5;PSMD12;SHC2;PSMD11;PSMD14;PSMD13;CUL3;PDGFB;RASAL1;PHB;PSMA7;RASGRP3;PPP2CA;PSMB6;RAP1B;PSMB7;PPP2CB;PSMB5;RAP1A;PSMB2;PSMB3;PSMB1;UBC;MAPK1;CSK;FYN;PSMF1;RPS27A;CAMK2G;DAB2IP;PTK2;PSMB8;PSMC5;PSMC6;PSMC3;PSMC4;PTPRA;PSMC1;PSMC2;FGF18;GRB2;KRAS;CALM1;LAMTOR3 |
| 207 | SHC1 events in ERBB4 signaling Homo sapiens R-HSA-1250347 | 1.90E-05 | 1.31E-04 | 1.82E+00 | 1.97E+01 | YWHAB;RASGRF2;PEBP1;WDR83;ARRB1;FGF2;PSMD8;PSMD9;PSMD6;PSMD7;PSMD4;PPP2R5E;PSMD2;PSMD3;PEA15;PSMD1;JAK2;SPTAN1;JAK1;MAP2K2;DUSP1;PPP2R5A;DUSP8;RBX1;BTC;PSMA5;PSMA6;PSMA3;PSMA4;PSMA1;PSMA2;RASA1;AKAP9;PSME1;PSME2;TLN1;HBEGF;PSMD10;SPTBN5;PSMD12;SHC2;PSMD11;PSMD14;PSMD13;CUL3;PDGFB;RASAL1;PHB;PSMA7;RASGRP3;PPP2CA;PSMB6;RAP1B;PSMB7;PPP2CB;PSMB5;RAP1A;PSMB2;PSMB3;PSMB1;UBC;MAPK1;CSK;FYN;PSMF1;RPS27A;CAMK2G;DAB2IP;PTK2;PSMB8;PSMC5;PSMC6;PSMC3;PSMC4;PTPRA;PSMC1;PSMC2;FGF18;GRB2;KRAS;CALM1;LAMTOR3 |

| | | | | | | |
|---|---|---|---|---|---|---|
| 208 | RAF/MAP kinase cascade Homo sapiens R-HSA-5673001 | 1.90E-05 | 1.31E-04 | 1.82E+00 | 1.97E+01 | YWHAB;RASGRF2;PEBP1;WDR83;ARRB1;FGF2;PSMD8;PSMD9;PSMD6;PSMD7;PSMD4;PPP2R5E;PSMD2;PSMD3;PEA15;PSMD1;JAK2;SPTAN1;JAK1;MAP2K2;DUSP1;PPP2R5A;DUSP8;RBX1;BTC;PSMA5;PSMA6;PSMA3;PSMA4;PSMA1;PSMA2;RASA1;AKAP9;PSME1;PSME2;TLN1;HBEGF;PSMD10;SPTBN5;PSMD12;SHC2;PSMD11;PSMD14;PSMD13;CUL3;PDGFB;RASAL1;PHB;PSMA7;RASGRP3;PPP2CA;PSMB6;RAP1B;PSMB7;PPP2CB;PSMB5;RAP1A;PSMB2;PSMB3;PSMB1;UBC;MAPK1;CSK;FYN;PSMF1;RPS27A;CAMK2G;DAB2IP;PTK2;PSMB8;PSMC5;PSMC6;PSMC3;PSMC4;PTPRA;PSMC1;PSMC2;FGF18;GRB2;KRAS;CALM1;LAMTOR3 |
| 209 | Signalling to RAS Homo sapiens R-HSA-167044 | 2.05E-05 | 1.40E-04 | 1.79E+00 | 1.93E+01 | YWHAB;RASGRF2;PEBP1;WDR83;ARRB1;FGF2;PSMD8;PSMD9;PSMD6;PSMD7;PSMD4;PPP2R5E;PSMD2;PSMD3;PEA15;PSMD1;JAK2;SPTAN1;JAK1;MAP2K2;DUSP1;PPP2R5A;DUSP8;RBX1;BTC;PSMA5;PSMA6;PSMA3;PSMA4;PSMA1;PSMA2;RASA1;AKAP9;PSME1;PSME2;TLN1;HBEGF;PSMD10;SPTBN5;PSMD12;SHC2;RALA;PSMD11;RALB;PSMD14;PSMD13;CUL3;PDGFB;RASAL1;PHB;PSMA7;RASGRP3;PPP2CA;PSMB6;RAP1B;PSMB7;PPP2CB;PSMB5;RAP1A;PSMB2;PSMB3;PSMB1;UBC;MAPK1;CSK;FYN;PSMF1;RPS27A;CAMK2G;DAB2IP;PTK2;PSMB8;MAPK13;PSMC5;PSMC6;PSMC3;PSMC4;PTPRA;PSMC1;PSMC2;FGF18;GRB2;KRAS;CALM1;LAMTOR3 |
| 210 | Global Genome Nucleotide Excision Repair (GG-NER) Homo sapiens R-HSA-5696399 | 2.06E-05 | 1.40E-04 | 2.64E+00 | 2.85E+01 | INO80B;PCNA;CETN2;XPA;COPS7B;YY1;TFPT;SUMO1;SUMO3;SUMO2;UBC;POLD2;POLK;RPS27A;UBE2I;RFC1;PARP1;MCRS1;ACTL6A;USP45;RAD23A;ACTR8;RAD23B;GTF2H4;RBX1;DDB1;COPS4;COPS3;COPS6;COPS5;COPS2;RPA3;ERCC1;UBE2V2;ERCC5;MNAT1 |
| 211 | ARMS-mediated activation Homo sapiens R-HSA-170984 | 2.07E-05 | 1.40E-04 | 1.80E+00 | 1.94E+01 | YWHAB;RASGRF2;PEBP1;WDR83;ARRB1;FGF2;PSMD8;PSMD9;PSMD6;PSMD7;PSMD4;PPP2R5E;PSMD2;PSMD3;PEA15;PSMD1;JAK2;SPTAN1;JAK1;MAP2K2;DUSP1;PPP2R5A;DUSP8;RBX1;BTC;PSMA5;PSMA6;PSMA3;PSMA4;PSMA1;PSMA2;RASA1;AKAP9;PSME1;PSME2;TLN1;HBEGF;PSMD10;SPTBN5;PSMD12;SHC2;PSMD11;PSMD14;PSMD13;CUL3;PDGFB;RASAL1;PHB;PSMA7;RASGRP3;PPP2CA;PSMB6;RAP1B;PSMB7;PPP2CB;PSMB5;RAP1A;PSMB2;PSMB3;PSMB1;UBC;MAPK1;CSK;FYN;PSMF1;RPS27A;CAMK2G;KIDINS220;DAB2IP;PTK2;PSMB8;PSMC5;PSMC6;PSMC3;PSMC4;PTPRA;PSMC1;PSMC2;FGF18;GRB2;KRAS;CALM1;LAMTOR3 |

| | | | | | | |
|---|---|---|---|---|---|---|
| 212 | Nonsense Mediated Decay (NMD) independent of the Exon Junction Complex (EJC) Homo sapiens R-HSA-975956 | 2.65E-05 | 1.79E-04 | 2.52E+00 | 2.65E+01 | RPL5;RPL30;RPL3;RPL10;RPLP1;RPL10A;RPL8;RPS4Y1;RPL6;RPS15;RPL7A;RPL18A;RPS19;RPL14;RPL15;RPS2;RPL18;RPS27A;RPS10;RPS13;RPS12;RPL19;RPS9;NCBP1;RPS8;RPS5;RPSA;RPL23A;ETF1;PABPC1;FAU;RPL29;RPL28;RPS21;RPL26L1;RPS24;EIF4G1;RPS23 |
| 213 | mTOR signalling Homo sapiens R-HSA-165159 | 2.73E-05 | 1.83E-04 | 3.93E+00 | 4.14E+01 | PRKAA1;YWHAB;CAB39L;STRADB;PRKAG1;PRKAG2;MTOR;RRAGA;RRAGB;RHEB;AKT2;AKT1S1;MLST8;AKT1;LAMTOR1;LAMTOR4;EIF4E;LAMTOR3;EIF4B;EIF4G1;LAMTOR5 |
| 214 | MAPK1/MAPK3 signaling Homo sapiens R-HSA-5684996 | 2.92E-05 | 1.95E-04 | 1.78E+00 | 1.86E+01 | YWHAB;RASGRF2;PEBP1;WDR83;ARRB1;FGF2;PSMD8;PSMD9;PSMD6;PSMD7;PSMD4;PPP2R5E;PSMD2;PSMD3;PEA15;PSMD1;JAK2;SPTAN1;JAK1;MAP2K2;DUSP1;PPP2R5A;DUSP8;RBX1;BTC;PSMA5;PSMA6;PSMA3;PSMA4;PSMA1;PSMA2;RASA1;AKAP9;PSME1;PSME2;TLN1;HBEGF;PSMD10;SPTBN5;PSMD12;SHC2;PSMD11;PSMD14;PSMD13;CUL3;PDGFB;RASAL1;PHB;PSMA7;RASGRP3;PPP2CA;PSMB6;RAP1B;PSMB7;PPP2CB;PSMB5;RAP1A;PSMB2;PSMB3;PSMB1;UBC;MAPK1;CSK;FYN;PSMF1;RPS27A;CAMK2G;DAB2IP;PTPN11;PTK2;PSMB8;PSMC5;PSMC6;PSMC3;PSMC4;PTPRA;PSMC1;PSMC2;FGF18;GRB2;KRAS;CALM1;LAMTOR3 |
| 215 | COPII (Coat Protein 2) Mediated Vesicle Transport Homo sapiens R-HSA-204005 | 3.10E-05 | 2.06E-04 | 2.83E+00 | 2.94E+01 | NAPA;MCFD2;SERPINA1;TMED10;SAR1B;RAB1B;GOSR2;CTSZ;USO1;NAPG;GOLGA2;PPP6C;LMAN1;SCFD1;TFG;LMAN2;TMED2;STX5;CD59;NSF;TRAPPC3;SEC24A;TRAPPC1;TRAPPC10;TRAPPC4;TRAPPC5;TRAPPC9;TRAPPC6A;CNIH1;SEC22B;SEC22C |
| 216 | Interleukin receptor SHC signaling Homo sapiens R-HSA-912526 | 3.16E-05 | 2.09E-04 | 1.77E+00 | 1.83E+01 | YWHAB;RASGRF2;PEBP1;WDR83;ARRB1;FGF2;PSMD8;PSMD9;PSMD6;PSMD7;PSMD4;PPP2R5E;PSMD2;PSMD3;PEA15;PSMD1;JAK2;SPTAN1;JAK1;MAP2K2;DUSP1;PPP2R5A;DUSP8;RBX1;BTC;PSMA5;PSMA6;PSMA3;PSMA4;PSMA1;PSMA2;RASA1;AKAP9;PSME1;PSME2;TLN1;HBEGF;PSMD10;SPTBN5;PSMD12;SHC2;PSMD11;PSMD14;PSMD13;CUL3;PDGFB;PIK3R3;RASAL1;PHB;PSMA7;RASGRP3;PPP2CA;PSMB6;RAP1B;PSMB7;PPP2CB;PSMB5;RAP1A;PSMB2;PSMB3;PSMB1;UBC;MAPK1;CSK;FYN;PSMF1;RPS27A;CAMK2G;DAB2IP;PTK2;PSMB8;PSMC5;PSMC6;PSMC3;PSMC4;PTPRA;PSMC1;PSMC2;FGF18;GRB2;PTPN6;KRAS;CALM1;LAMTOR3 |

| | | | | | | |
|---|---|---|---|---|---|---|
| 217 | Selenocysteine synthesis Homo sapiens R-HSA-2408557 | 3.73E-05 | 2.45E-04 | 2.50E+00 | 2.55E+01 | RPL5;RPL30;RPL3;RPL10;RPLP1;RPL10A;RPL8;RPS4Y1;RPL6;RPS15;RPL7A;RPL18A;RPS19;RPL14;RPL15;RPS2;RPL18;RPS27A;RPS10;RPS13;RPS12;RPL19;EEFSEC;RPS9;RPS8;RPS5;SECISBP2;SARS;RPSA;RPL23A;FAU;RPL29;RPL28;RPS21;RPL26L1;RPS24;RPS23 |
| 218 | Metabolism of nucleotides Homo sapiens R-HSA-15869 | 3.73E-05 | 2.45E-04 | 2.50E+00 | 2.55E+01 | CDA;DTYMK;GMPR2;AK1;ADK;NUDT1;GMPS;AK2;GMPR;NUDT5;AK6;DGUOK;ATIC;GUK1;TK2;TK1;NUDT15;LHPP;ADSL;DUT;TXNRD1;GSR;NME2;AMPD2;NME4;CTPS1;PAICS;APRT;NT5C3A;NUDT9;UCK1;RRM2B;CAT;CMPK1;HPRT1;DCTPP1;ITPA |
| 219 | Pyruvate metabolism and Citric Acid (TCA) cycle Homo sapiens R-HSA-71406 | 3.75E-05 | 2.45E-04 | 3.37E+00 | 3.44E+01 | FH;PDHA1;MDH2;IDH3G;GLO1;IDH2;SDHC;PDHB;SDHD;SDHA;SDHB;LDHB;LDHA;SUCLA2;NNT;MPC2;IDH3B;PDK3;SUCLG2;SUCLG1;HAGH;SLC16A3;PDK2;IDH3A |
| 220 | Signalling to p38 via RIT and RIN Homo sapiens R-HSA-187706 | 3.79E-05 | 2.46E-04 | 1.77E+00 | 1.80E+01 | YWHAB;RASGRF2;PEBP1;WDR83;ARRB1;FGF2;PSMD8;PSMD9;PSMD6;PSMD7;PSMD4;PPP2R5E;PSMD2;PSMD3;PEA15;PSMD1;JAK2;SPTAN1;JAK1;MAP2K2;DUSP1;PPP2R5A;DUSP8;RBX1;BTC;PSMA5;PSMA6;PSMA3;PSMA4;PSMA1;PSMA2;RASA1;AKAP9;PSME1;PSME2;TLN1;HBEGF;PSMD10;SPTBN5;PSMD12;SHC2;PSMD11;PSMD14;PSMD13;CUL3;PDGFB;RASAL1;PHB;PSMA7;RASGRP3;PPP2CA;PSMB6;RAP1B;PSMB7;PPP2CB;PSMB5;RAP1A;PSMB2;PSMB3;PSMB1;UBC;MAPK1;CSK;FYN;PSMF1;RPS27A;CAMK2G;DAB2IP;PTK2;PSMB8;PSMC5;PSMC6;PSMC3;PSMC4;PTPRA;PSMC1;PSMC2;FGF18;GRB2;KRAS;CALM1;LAMTOR3 |
| 221 | Viral mRNA Translation Homo sapiens R-HSA-192823 | 3.88E-05 | 2.50E-04 | 2.53E+00 | 2.57E+01 | RPL5;RPL30;RPL3;RPL10;RPLP1;RPL10A;RPL8;RPS4Y1;RPL6;RPS15;RPL7A;RPL18A;RPS19;RPL14;RPL15;RPS2;RPL18;RPS27A;RPS10;RPS13;RPS12;RPL19;RPS9;RPS8;RPS5;RPSA;RPL23A;DNAJC3;FAU;RPL29;GRSF1;RPL28;RPS21;RPL26L1;RPS24;RPS23 |
| 222 | Peptide chain elongation Homo sapiens R-HSA-156902 | 3.88E-05 | 2.50E-04 | 2.53E+00 | 2.57E+01 | RPL5;RPL30;RPL3;RPL10;RPLP1;RPL10A;RPL8;RPS4Y1;RPL6;RPS15;RPL7A;RPL18A;RPS19;RPL14;RPL15;RPS2;RPL18;RPS27A;RPS10;RPS13;RPS12;RPL19;RPS9;RPS8;RPS5;RPSA;RPL23A;EEF2;EEF1A1;FAU;RPL29;RPL28;RPS21;RPL26L1;RPS24;RPS23 |
| 223 | PKB-mediated events Homo sapiens R-HSA-109703 | 4.51E-05 | 2.89E-04 | 3.73E+00 | 3.73E+01 | PRKAA1;YWHAB;CAB39L;STRADB;PRKAG1;PRKAG2;MTOR;RRAGA;RRAGB;RHEB;AKT2;AKT1S1;MLST8;AKT1;LAMTOR1;LAMTOR4;EIF4E;LAMTOR3;EIF4B;EIF4G1;LAMTOR5 |
| 224 | Gluconeogenesis Homo sapiens R-HSA-70263 | 4.72E-05 | 3.00E-04 | 4.33E+00 | 4.32E+01 | SLC25A1;GPI;TPI1;GOT1;MDH1;MDH2;PGAM1;GOT2;PGAM2;ENO1;ENO2;ENO3;PC;PGK1;ALDOC;ALDOA;SLC25A11;GAPDH |

| | | | | | | |
|---|---|---|---|---|---|---|
| 225 | Cooperation of Prefoldin and TriC/CCT in actin and tubulin folding Homo sapiens R-HSA-389958 | 4.72E-05 | 3.00E-04 | 4.33E+00 | 4.32E+01 | CCT3;CCT2;VBP1;TUBB4B;TUBA4A;CCT6A;TUBA1C;TUBA1B;TUBB6;TCP1;PFDN1;PFDN2;CCT8;CCT7;PFDN4;PFDN5;CCT5;CCT4 |
| 226 | mTORC1-mediated signalling Homo sapiens R-HSA-166208 | 5.17E-05 | 3.27E-04 | 5.90E+00 | 5.82E+01 | YWHAB;MTOR;RRAGA;RRAGB;RHEB;AKT1S1;MLST8;LAMTOR1;LAMTOR4;LAMTOR3;EIF4E;EIF4B;EIF4G1;LAMTOR5 |
| 227 | NCAM signaling for neurite out-growth Homo sapiens R-HSA-375165 | 5.25E-05 | 3.31E-04 | 1.70E+00 | 1.68E+01 | YWHAB;RASGRF2;PEBP1;WDR83;ARRB1;FGF2;PSMD8;PSMD9;PSMD6;PSMD7;PSMD4;PPP2R5E;PSMD2;PSMD3;PEA15;PSMD1;JAK2;SPTAN1;JAK1;MAP2K2;DUSP1;PPP2R5A;DUSP8;RBX1;BTC;PSMA5;PSMA6;PSMA3;PSMA4;CACNB4;CREB1;PSMA1;PSMA2;COL4A4;RASA1;AKAP9;COL4A3;PSME1;PSME2;TLN1;HBEGF;PSMD10;SPTBN5;PSMD12;SHC2;PSMD11;PSMD14;PSMD13;CUL3;PDGFB;RASAL1;CACNA1D;PHB;CACNA1C;PSMA7;RASGRP3;PPP2CA;PSMB6;RAP1B;PSMB7;PPP2CB;PSMB5;RAP1A;PSMB2;PSMB3;PSMB1;UBC;MAPK1;CSK;FYN;PSMF1;RPS27A;CAMK2G;PRNP;DAB2IP;PTK2;PSMB8;PSMC5;PSMC6;PSMC3;PSMC4;PTPRA;PSMC1;PSMC2;FGF18;GRB2;KRAS;CALM1;LAMTOR3 |
| 228 | EGFR downregulation Homo sapiens R-HSA-182971 | 5.27E-05 | 3.31E-04 | 4.90E+00 | 4.83E+01 | SH3KBP1;CLTA;AP2B1;STAM;CBL;AP2A2;EPN1;CDC42;UBC;AP2S1;GRB2;RPS27A;ARHGEF7;EPS15;STAM2;AP2M1 |
| 229 | Retrograde transport at the Trans-Golgi-Network Homo sapiens R-HSA-6811440 | 5.77E-05 | 3.60E-04 | 3.24E+00 | 3.16E+01 | NSF;NAPA;SCOC;NAA30;SYS1;TMF1;RIC1;COG6;COG2;ARL1;M6PR;GCC2;NAPG;STX10;RAB43;TGOLN2;VPS51;VPS53;ARFIP2;PLIN3;RAB9A;NAA38;RAB6A;VAMP3 |
| 230 | HIV Life Cycle Homo sapiens R-HSA-162587 | 6.00E-05 | 3.73E-04 | 2.04E+00 | 1.98E+01 | GTF2B;PSIP1;RCC1;NELFCD;KPNA1;SUPT16H;PDCD6IP;NCBP1;LIG4;VPS37D;VPS37B;GTF2F2;NUP93;NUP50;CHMP4B;CHMP3;CHMP4A;PPIA;VPS28;CHMP5;GTF2A1;GTF2A2;TSG101;VPS4B;VPS4A;SUPT4H1;FURIN;GTF2E2;POLR2B;POLR2C;TPR;UBC;POLR2E;POLR2H;RPS27A;POLR2J;POLR2K;POLR2L;RANBP1;XRCC6;XRCC5;TAF13;TAF10;TAF11;HMGA1;GTF2H4;CDK9;CHMP2B;CHMP2A;NMT1;TCEA1;MNAT1;RAN |

| | | | | | | |
|---|---|---|---|---|---|---|
| 231 | DNA Damage Recognition in GG-NER Homo sapiens R-HSA-5696394 | 6.51E-05 | 4.04E-04 | 3.75E+00 | 3.61E+01 | INO80B;PARP1;CETN2;MCRS1;ACTL6A;COPS7B;RAD23A;ACTR8;RAD23B;RBX1;YY1;COPS4;DDB1;TFPT;COPS3;COPS6;COPS5;COPS2;UBC;RPS27A |
| 232 | DAP12 interactions Homo sapiens R-HSA-2172127 | 6.79E-05 | 4.19E-04 | 1.58E+00 | 1.51E+01 | PEBP1;WDR83;FGF2;PSMD8;PSMD9;PSMD6;PSMD7;PSMD4;AKT2;PSMD2;PSMD3;AKT1;PSMD1;PIP4K2C;IER3;MAP2K2;HLA-C;HLA-E;RBX1;BTC;PRKAR1A;AKAP9;PSME1;PSME2;TLN1;PSMD10;SPTBN5;PSMD12;SHC2;PSMD11;PSMD14;PSMD13;PDE1A;MAPKAP1;CUL3;PDGFB;PIK3R3;ITPR2;RASAL1;PHB;RASGRP3;RAP1B;RAP1A;PRKAR2B;PRKAR2A;UBC;PIP5K1B;PSMF1;PTK2;PTPRA;AGO2;FGF18;MDM2;GRB2;CALM1;PRR5;GSK3B;GSK3A;CDKN1B;YWHAB;RASGRF2;ARRB1;PPP2R5E;PEA15;MLST8;RAC1;JAK2;SPTAN1;JAK1;VAV3;DUSP1;PDPK1;PPP2R5A;DUSP8;PSMA5;PSMA6;PSMA3;PSMA4;CREB1;PSMA1;PSMA2;RASA1;AKT1S1;HBEGF;ADCY3;PSMA7;ADCY5;PPP2CA;PSMB6;PSMB7;PPP2CB;PSMB5;PSMB2;PSMB3;PSMB1;MAPK1;CSK;FYN;RPS27A;CAMK2G;BAD;DAB2IP;PTPN11;PSMB8;MTOR;PSMC5;PSMC6;PSMC3;PSMC4;PSMC1;PSMC2;RPS6KB2;KRAS;LAMTOR3 |
| 233 | Constitutive Signaling by AKT1 E17K in Cancer Homo sapiens R-HSA-5674400 | 7.42E-05 | 4.56E-04 | 5.05E+00 | 4.81E+01 | PRR5;GSK3B;GSK3A;CDKN1B;BAD;PDPK1;MAPKAP1;MTOR;CREB1;AKT2;RPS6KB2;AKT1S1;MLST8;MDM2;AKT1 |
| 234 | Detoxification of Reactive Oxygen Species Homo sapiens R-HSA-3299685 | 8.17E-05 | 4.99E-04 | 4.05E+00 | 3.81E+01 | TXNRD2;GSTP1;TXNRD1;GSR;CYBA;SOD2;SOD3;PRDX6;ATOX1;TXN2;SOD1;PRDX3;PRDX2;PRDX5;PRDX1;CAT;CYCS;P4HB |

| | | | | | | |
|---|---|---|---|---|---|---|
| 235 | Adaptive Immune System Homo sapiens R-HSA-1280218 | 8.61E-05 | 5.24E-04 | 1.37E+00 | 1.29E+01 | KEAP1;UBE2L6;UBE2L3;PSMD8;PSMD9;HERC4;RNF115;PSMD6;HERC3;RNF114;PSMD7;HERC2;PSMD4;AKT2;PSMD2;CDC26;PSMD3;CDC27;AP1S2;AKT1;PSMD1;FBXO3;PIP4K2C;KIFAP3;IER3;SKP1;CXADR;FBXO2;HLA-C;HLA-G;HLA-E;BTC;RNF126;PSME1;ORAI1;PSME2;RBCK1;ANAPC13;SAR1B;MAPKAP1;PDGFB;PIK3R3;BCL10;UBE2J2;ANAPC11;UBE2J1;PIP5K1B;PSMF1;VASP;CTSA;YES1;LNPEP;UBAC1;AGO2;REL;GRB2;CALR;CALM1;ARF1;CDKN1B;SH3KBP1;CLTC;CLTA;AP2A2;ACTR1A;ACTR1B;CTSL;CTSK;SEC61G;ARIH2;CTSH;SEC61B;CTSF;RAC1;CTSD;CTSB;PSMA5;PSMA6;PSMA3;PSMA4;PSMA1;KIF2A;PSMA2;GAN;RAPGEF3;HLA-DQB1;HBEGF;RAB7A;PSMA7;PSMB6;PPP2CA;PSMB7;PPP2CB;HLA-DMA;PAK1;PSMB5;HLA-DMB;PSMB2;PSMB3;PSMB1;AP2S1;RPS27A;PAK2;DYNC1H1;CD74;MIB2;UBE2G1;UBE2G2;AP2B1;PSMB8;PSMC5;ITCH;PSMC6;PSMC3;PSMC4;PSMC1;NEDD4;PSMC2;SIPA1;UBE3C;UBE2D2;FBXO27;UBE2D3;LTN1;UBE2D1;FGF2;NPEPPS;KIF5C;RNF19A;KIF5B;AP2M1;PDIA3;FBXW11;UBE2E3;UBE2E1;TAP1;UBE2E2;CYBA;RBX1;UBE2V2;ANAPC5;UBE2V1;PSMD10;PSMD12;PSMD11;PSMD14;PSMD13;CUL3;CUL2;CUL1;ITPR2;UBR2;KLC1;FBXO44;RAP1GAP;RASGRP3;RAP1B;RAP1A;SOCS1;BTBD1;KIF3A;UBC;AP1M2;UBE2F;UBE2H;UBE2B;HSPA5;SMURF2;UBE2A;NFKB1;NFKBIA;UBE2S;FGF18;MKRN1;MDM2;UBE2K;UBE2M;ITGB1;PRR5;GSK3B;GSK3A;YWHAB;ITGB2;UBE2Q1;AP1G1;PPP2R5E;UBE2Q2;MLS [illegible] |
| 236 | Formation of TC-NER Pre-Incision Complex Homo sapiens R-HSA-6781823 | 8.93E-05 | 5.41E-04 | 3.01E+00 | 2.81E+01 | XPA;COPS7B;POLR2B;POLR2C;UBC;POLR2E;POLR2H;RPS27A;POLR2J;POLR2K;HMGN1;POLR2L;USP7;GTF2H4;RBX1;DDB1;COPS4;COPS3;XAB2;COPS6;COPS5;COPS2;PPIE;TCEA1;MNAT1 |
| 237 | Eukaryotic Translation Termination Homo sapiens R-HSA-72764 | 9.41E-05 | 5.67E-04 | 2.38E+00 | 2.21E+01 | RPL5;RPL30;RPL3;RPL10;RPLP1;RPL10A;RPL8;RPS4Y1;RPL6;RPS15;TRMT112;RPL7A;RPL18A;RPS19;RPL14;RPL15;RPS2;RPL18;RPS27A;RPS10;RPS13;RPS12;RPL19;RPS9;RPS8;RPS5;RPSA;RPL23A;ETF1;FAU;RPL29;RPL28;RPS21;RPL26L1;RPS24;RPS23 |
| 238 | RHO GTPases Activate WASPs and WAVEs Homo sapiens R-HSA-5663213 | 9.43E-05 | 5.67E-04 | 3.77E+00 | 3.49E+01 | CYFIP2;ACTR3;NCKAP1;ACTR2;ARPC1B;ARPC1A;BRK1;WASL;ARPC5;PTK2;ACTG1;CDC42;ABI2;ARPC2;ARPC3;MAPK1;GRB2;RAC1;WASF2 |

| | | | | | | |
|---|---|---|---|---|---|---|
| 239 | Interleukin-2 signaling Homo sapiens R-HSA-451927 | 9.70E-05 | 5.81E-04 | 1.69E+00 | 1.56E+01 | YWHAB;RASGRF2;PEBP1;WDR83;ARRB1;FGF2;PSMD8;PSMD9;PSMD6;PSMD7;PSMD4;PPP2R5E;PSMD2;PSMD3;PEA15;PSMD1;JAK2;SPTAN1;JAK1;MAP2K2;DUSP1;PPP2R5A;DUSP8;RBX1;BTC;PSMA5;PSMA6;PSMA3;PSMA4;PSMA1;PSMA2;RASA1;AKAP9;PSME1;PSME2;TLN1;HBEGF;PSMD10;SPTBN5;PSMD12;SHC2;PSMD11;PSMD14;PSMD13;CUL3;PDGFB;PIK3R3;RASAL1;PHB;PSMA7;RASGRP3;PPP2CA;PSMB6;RAP1B;PSMB7;PPP2CB;PSMB5;RAP1A;PSMB2;PSMB3;PSMB1;UBC;MAPK1;CSK;FYN;PSMF1;RPS27A;CAMK2G;DAB2IP;PTK2;PSMB8;PSMC5;PSMC6;PSMC3;PSMC4;PTPRA;PSMC1;PSMC2;FGF18;GRB2;PTPN6;KRAS;CALM1;LAMTOR3 |
| 240 | Golgi-to-ER retrograde transport Homo sapiens R-HSA-8856688 | 1.02E-04 | 6.09E-04 | 2.17E+00 | 1.99E+01 | NAPA;DCTN6;COPB2;ARF4;ARF1;NBAS;TMED10;DCTN2;KDELR1;COPB1;RAB1B;DCTN3;KLC1;NAPG;KIF5C;KIF3A;KIF5B;TMED3;TMED2;KIF21A;KIFAP3;RAB6A;TMED7;NSF;DYNC1H1;TMED9;GALNT1;SURF4;PLA2G4A;COPZ2;ARFGAP3;COPZ1;ARCN1;KIF2A;KIFC2;KIF26B;RAB18;KDELR2;SEC22B;PAFAH1B3;COPE;PAFAH1B2;PAFAH1B1 |
| 241 | Deadenylation of mRNA Homo sapiens R-HSA-429947 | 1.04E-04 | 6.19E-04 | 5.24E+00 | 4.81E+01 | EIF4A2;EIF4A1;CNOT6L;EIF4A3;CNOT4;CNOT7;CNOT11;CNOT2;CNOT3;PABPC1;EIF4E;EIF4B;EIF4G1;PAIP1 |
| 242 | EPHB-mediated forward signaling Homo sapiens R-HSA-3928662 | 1.14E-04 | 6.73E-04 | 3.37E+00 | 3.06E+01 | ACTR3;ACTR2;YES1;SDC2;ARPC1B;LIMK2;ARPC1A;WASL;ARPC5;RHOA;PTK2;ACTG1;CDC42;TIAM1;PAK1;ARPC2;ARPC3;RASA1;CFL1;FYN;RAC1 |
| 243 | RNA Polymerase II Transcription Homo sapiens R-HSA-73857 | 1.21E-04 | 7.12E-04 | 2.06E+00 | 1.86E+01 | FYTTD1;SLBP;GTF2A1;GTF2A2;RBM8A;POLDIP3;GTF2B;EIF4A3;SUPT4H1;GTF2E2;LSM10;FIP1L1;CLP1;POLR2B;POLR2C;MAGOH;CSTF1;POLR2E;SNRPD3;NELFCD;POLR2H;POLR2J;POLR2K;POLR2L;SUPT16H;NCBP1;TAF13;CPSF3;TAF10;TAF11;THOC3;THOC2;THOC5;GTF2F2;GTF2H4;CDC40;CDK9;DDX39A;SARNP;SNRPG;SRSF2;TCEA1;SRSF4;RNPS1;MNAT1;SNRPB;SRSF9 |
| 244 | Influenza Viral RNA Transcription and Replication Homo sapiens R-HSA-168273 | 1.39E-04 | 8.17E-04 | 2.03E+00 | 1.80E+01 | RPL5;RPL30;RPL3;RPL10;RPLP1;RPL8;RPL10A;RPS4Y1;RPL6;RPS15;RPL7A;RPS19;RPL18A;POLR2B;POLR2C;TPR;RPL14;POLR2E;RPL15;RPS2;POLR2H;RPL18;RPS27A;RPS10;POLR2J;RPS13;POLR2K;RPS12;RPL19;POLR2L;RPS9;HSP90AA1;RPS8;RPS5;RPSA;RPL23A;GTF2F2;NUP93;DNAJC3;NUP50;FAU;RPL29;GRSF1;RPL28;RPS21;RPS24;RPL26L1;RPS23 |

| | | | | | | |
|---|---|---|---|---|---|---|
| 245 | SUMO is transferred from E1 to E2 (UBE2I, UBC9) Homo sapiens R-HSA-3065678 | 1.45E-04 | 8.43E-04 | 9.25E+04 | 8.17E+05 | UBE2I;SUMO1;SUMO3;SUMO2;UBA2;SAE1 |
| 246 | Uptake and function of diphtheria toxin Homo sapiens R-HSA-5336415 | 1.45E-04 | 8.43E-04 | 9.25E+04 | 8.17E+05 | HSP90AA1;HSP90AB1;TXNRD1;CD9;EEF2;HBEGF |
| 247 | MAPK family signaling cascades Homo sapiens R-HSA-5683057 | 1.50E-04 | 8.68E-04 | 1.62E+00 | 1.43E+01 | YWHAB;RASGRF2;PEBP1;HSPB1;WDR83;ARRB1;FGF2;PSMD8;PSMD9;PSMD6;PSMD7;PSMD4;PPP2R5E;PSMD2;PSMD3;PEA15;PSMD1;RAC1;JAK2;SPTAN1;JAK1;MAP2K2;DUSP1;NCOA3;PPP2R5A;DUSP8;RBX1;BTC;PSMA5;PSMA6;PSMA3;PSMA4;PSMA1;PSMA2;RASA1;AKAP9;CDC42EP3;PSME1;PSME2;MAPKAPK5;TLN1;HBEGF;PSMD10;SPTBN5;PSMD12;SHC2;PSMD11;PSMD14;PSMD13;CUL3;PDGFB;RASAL1;PHB;PSMA7;RASGRP3;PPP2CA;PSMB6;RAP1B;CDC42;PSMB7;PPP2CB;PAK1;PSMB5;RAP1A;PSMB2;PSMB3;PSMB1;UBC;MAPK1;CSK;FYN;PSMF1;RPS27A;PAK2;CAMK2G;DAB2IP;PTPN11;PTK2;PSMB8;PSMC5;PSMC6;PSMC3;PSMC4;PTPRA;PSMC1;AGO2;PSMC2;FGF18;GRB2;KRAS;CALM1;LAMTOR3 |
| 248 | Transcription-Coupled Nucleotide Excision Repair (TC-NER) Homo sapiens R-HSA-6781827 | 1.53E-04 | 8.83E-04 | 2.45E+00 | 2.16E+01 | PCNA;XPA;COPS7B;POLR2B;POLR2C;UBC;POLD2;POLR2E;POLK;POLR2H;RPS27A;POLR2J;POLR2K;HMGN1;POLR2L;USP7;RFC1;GTF2H4;RBX1;DDB1;COPS4;COPS3;XAB2;COPS6;COPS5;COPS2;RPA3;ERCC1;PPIE;TCEA1;ERCC5;MNAT1 |
| 249 | Unfolded Protein Response (UPR) Homo sapiens R-HSA-381119 | 1.73E-04 | 9.96E-04 | 2.32E+00 | 2.01E+01 | GSK3A;ACADVL;FKBP14;GOSR2;ADD1;HERPUD1;EXOSC7;SERP1;EXOSC4;SEC61G;LMNA;DNAJB9;SEC61B;SEC62;EXOSC3;EXOSC1;KLHDC3;ZBTB17;XBP1;HSPA5;WFS1;GFPT1;WIPI1;PDIA6;EIF2S1;DNAJC3;DNAJB11;SRPRB;HDGF;CALR;TLN1;ATP6V0D1;ATF6;DCP2;ATF4 |
| 250 | IRE1alpha activates chaperones Homo sapiens R-HSA-381070 | 1.87E-04 | 1.07E-03 | 2.81E+00 | 2.41E+01 | GSK3A;ACADVL;FKBP14;GOSR2;ADD1;SERP1;SEC61G;LMNA;DNAJB9;SEC61B;SEC62;KLHDC3;ZBTB17;XBP1;HSPA5;WFS1;GFPT1;WIPI1;PDIA6;DNAJC3;DNAJB11;SRPRB;HDGF;TLN1;ATP6V0D1 |

| | | | | | | |
|---|---|---|---|---|---|---|
| 251 | TGF-beta receptor signaling activates SMADs Homo sapiens R-HSA-2173789 | 1.98E-04 | 1.13E-03 | 3.82E+00 | 3.26E+01 | SMAD4;TGFB1;SMURF2;FURIN;NEDD8;CBL;UCHL5;PPP1CA;PPP1CB;FKBP1A;PPP1CC;BAMBI;UBC;STRAP;STUB1;RPS27A;UBE2M |
| 252 | Folding of actin by CCT/TriC Homo sapiens R-HSA-390450 | 2.17E-04 | 1.23E-03 | 1.35E+01 | 1.14E+02 | CCT3;CCT6A;CCT2;TCP1;CCT8;CCT7;CCT5;CCT4 |
| 253 | XBP1(S) activates chaperone genes Homo sapiens R-HSA-381038 | 2.68E-04 | 1.52E-03 | 2.79E+00 | 2.29E+01 | KLHDC3;ZBTB17;GSK3A;ACADVL;XBP1;FKBP14;WFS1;GOSR2;GFPT1;WIPI1;PDIA6;ADD1;DNAJC3;SERP1;SEC61G;DNAJB11;LMNA;DNAJB9;SRPRB;HDGF;SEC61B;SEC62;TLN1;ATP6V0D1 |
| 254 | Late Phase of HIV Life Cycle Homo sapiens R-HSA-162599 | 2.87E-04 | 1.62E-03 | 1.96E+00 | 1.60E+01 | GTF2A1;GTF2A2;TSG101;GTF2B;VPS4B;VPS4A;SUPT4H1;FURIN;GTF2E2;POLR2B;POLR2C;TPR;UBC;POLR2E;RCC1;NELFCD;POLR2H;RPS27A;POLR2J;POLR2K;POLR2L;RANBP1;SUPT16H;PDCD6IP;NCBP1;TAF13;TAF10;TAF11;VPS37D;VPS37B;GTF2F2;GTF2H4;NUP93;CDK9;NUP50;CHMP2B;CHMP2A;NMT1;CHMP4B;CHMP3;TCEA1;CHMP4A;MNAT1;PPIA;VPS28;RAN;CHMP5 |
| 255 | Fc epsilon receptor (FCERI) signaling Homo sapiens R-HSA-2454202 | 3.42E-04 | 1.92E-03 | 1.48E+00 | 1.18E+01 | UBE2D2;UBE2D1;PEBP1;WDR83;FGF2;PSMD8;PSMD9;PSMD6;PSMD7;PSMD4;AKT2;PSMD2;PSMD3;AKT1;PSMD1;PIP4K2C;IER3;SKP1;MAP2K2;FBXW11;RBX1;BTC;AKAP9;PSME1;PSME2;UBE2V1;TLN1;PSMD10;SHC2;SPTBN5;PSMD12;PSMD11;PSMD14;PSMD13;MAPKAP1;CUL3;PDGFB;CUL1;ITPR2;PIK3R3;RASAL1;BCL10;PHB;RASGRP3;RAP1B;RAP1A;UBC;PIP5K1B;PSMF1;NFKB1;PTK2;NFKBIA;PTPRA;AGO2;FGF18;MDM2;GRB2;CALM1;PRR5;GSK3B;GSK3A;CDKN1B;YWHAB;RASGRF2;ARRB1;PPP3CB;PPP2R5E;PEA15;MLST8;RAC1;JAK2;SPTAN1;JAK1;VAV3;DUSP1;PDPK1;PPP2R5A;DUSP8;PSMA5;PSMA6;PSMA3;PSMA4;CREB1;PSMA1;PSMA2;RASA1;AKT1S1;HBEGF;PSMA7;PSMB6;PPP2CA;PSMB7;PPP2CB;MAPK9;PAK1;PSMB5;PSMB2;PSMB3;PSMB1;MAPK1;CSK;FYN;RPS27A;PAK2;CAMK2G;MAP3K1;BAD;DAB2IP;PTPN11;PSMB8;MTOR;PSMC5;PSMC6;PSMC3;PSMC4;PSMC1;PSMC2;RPS6KB2;KRAS;LAMTOR3 |

| | | | | | | |
|---|---|---|---|---|---|---|
| 256 | Activation of BAD and translocation to mitochondria Homo sapiens R-HSA-111447 | 3.75E-04 | 2.10E-03 | 6.73E+00 | 5.31E+01 | YWHAE;YWHAQ;YWHAB;BAD;AKT2;AKT1;BID;YWHAZ;YWHAG;YWHAH |
| 257 | Formation of Incision Complex in GG-NER Homo sapiens R-HSA-5696395 | 5.58E-04 | 3.11E-03 | 2.93E+00 | 2.20E+01 | UBE2I;PARP1;CETN2;USP45;XPA;RAD23A;RAD23B;GTF2H4;RBX1;DDB1;SUMO1;SUMO3;RPA3;ERCC1;UBC;SUMO2;UBE2V2;ERCC5;RPS27A;MNAT1 |
| 258 | COPI-dependent Golgi-to-ER retrograde traffic Homo sapiens R-HSA-6811434 | 6.03E-04 | 3.34E-03 | 2.20E+00 | 1.63E+01 | NAPA;ARF4;COPB2;ARF1;NBAS;TMED10;KDELR1;RAB1B;COPB1;KLC1;NAPG;KIF5C;KIF3A;KIF5B;TMED3;TMED2;KIF21A;TMED7;KIFAP3;NSF;TMED9;SURF4;COPZ2;ARFGAP3;COPZ1;ARCN1;KIF2A;KIFC2;KIF26B;KDELR2;SEC22B;COPE |
| 259 | SUMO is conjugated to E1 (UBA2:SAE1) Homo sapiens R-HSA-3065676 | 6.33E-04 | 3.47E-03 | 7.71E+04 | 5.68E+05 | SUMO1;SUMO3;SUMO2;UBA2;SAE1 |
| 260 | Beta oxidation of octanoyl-CoA to hexanoyl-CoA Homo sapiens R-HSA-77348 | 6.33E-04 | 3.47E-03 | 7.71E+04 | 5.68E+05 | HADHB;HADHA;ECHS1;ACADM;HADH |
| 261 | Beta oxidation of decanoyl-CoA to octanoyl-CoA-CoA Homo sapiens R-HSA-77346 | 6.33E-04 | 3.47E-03 | 7.71E+04 | 5.68E+05 | HADHB;HADHA;ECHS1;ACADM;HADH |
| 262 | RHO GTPases activate PAKs Homo sapiens R-HSA-5627123 | 7.38E-04 | 4.03E-03 | 4.49E+00 | 3.24E+01 | CDC42;PPP1CB;MYL6;PAK1;CTTN;NF2;RAC1;CALM1;PAK2;MYH10;MYLK;MYL12B |
| 263 | Processing and activation of SUMO Homo sapiens R-HSA-3215018 | 7.65E-04 | 4.15E-03 | 1.18E+01 | 8.45E+01 | SENP5;UBE2I;SUMO1;SUMO3;SUMO2;UBA2;SAE1 |

| | | | | | | |
|---|---|---|---|---|---|---|
| 264 | Integration of provirus Homo sapiens R-HSA-162592 | 7.65E-04 | 4.15E-03 | 1.18E+01 | 8.45E+01 | XRCC6;XRCC5;HMGA1;PSIP1;LIG4;PPIA;KPNA1 |
| 265 | Cytosolic tRNA aminoacylation Homo sapiens R-HSA-379716 | 8.82E-04 | 4.76E-03 | 3.98E+00 | 2.80E+01 | CARS;SARS;EPRS;AIMP1;NARS;QARS;PPA1;LARS;MARS;GARS;HARS;FARSA;EEF1E1 |
| 266 | Dual incision in TC-NER Homo sapiens R-HSA-6782135 | 8.85E-04 | 4.76E-03 | 2.37E+00 | 1.67E+01 | PCNA;XPA;POLR2B;POLR2C;UBC;POLD2;POLR2E;POLK;POLR2H;RPS27A;POLR2J;POLR2K;HMGN1;POLR2L;USP7;RFC1;GTF2H4;RBX1;DDB1;XAB2;RPA3;ERCC1;PPIE;TCEA1;ERCC5;MNAT1 |
| 267 | Cellular response to heat stress Homo sapiens R-HSA-3371556 | 8.99E-04 | 4.82E-03 | 2.02E+00 | 1.42E+01 | YWHAE;GSK3B;VCP;HSP90AB1;RPS19BP1;MRPL18;HSPA13;BAG4;HSPH1;BAG5;DNAJB6;BAG3;TPR;BAG1;MLST8;MAPK1;CAMK2G;HSPA9;HSPA8;HSP90AA1;HSPA5;HSBP1;HSPA4;ST13;PTGES3;HSPA2;SIRT1;MTOR;NUP93;EEF1A1;DNAJC7;NUP50;RPA3;AKT1S1;FKBP4;HSPA1A |
| 268 | Cleavage of Growing Transcript in the Termination Region Homo sapiens R-HSA-109688 | 9.34E-04 | 4.95E-03 | 2.41E+00 | 1.68E+01 | FYTTD1;SLBP;RBM8A;POLDIP3;EIF4A3;LSM10;FIP1L1;CLP1;MAGOH;CSTF1;SNRPD3;NCBP1;CPSF3;THOC3;THOC2;THOC5;CDC40;DDX39A;SARNP;SNRPG;SRSF2;SRSF4;RNPS1;SNRPB;SRSF9 |
| 269 | RNA Polymerase II Transcription Termination Homo sapiens R-HSA-73856 | 9.34E-04 | 4.95E-03 | 2.41E+00 | 1.68E+01 | FYTTD1;SLBP;RBM8A;POLDIP3;EIF4A3;LSM10;FIP1L1;CLP1;MAGOH;CSTF1;SNRPD3;NCBP1;CPSF3;THOC3;THOC2;THOC5;CDC40;DDX39A;SARNP;SNRPG;SRSF2;SRSF4;RNPS1;SNRPB;SRSF9 |
| 270 | Post-Elongation Processing of the Transcript Homo sapiens R-HSA-76044 | 9.34E-04 | 4.95E-03 | 2.41E+00 | 1.68E+01 | FYTTD1;SLBP;RBM8A;POLDIP3;EIF4A3;LSM10;FIP1L1;CLP1;MAGOH;CSTF1;SNRPD3;NCBP1;CPSF3;THOC3;THOC2;THOC5;CDC40;DDX39A;SARNP;SNRPG;SRSF2;SRSF4;RNPS1;SNRPB;SRSF9 |
| 271 | MHC class II antigen presentation Homo sapiens R-HSA-2132295 | 9.43E-04 | 4.98E-03 | 1.97E+00 | 1.37E+01 | DCTN6;ARF1;DCTN2;SAR1B;CLTC;DCTN3;CLTA;KLC1;AP2A2;ACTR1A;HLA-DMA;ACTR1B;HLA-DMB;KIF5C;KIF3A;CTSL;AP1G1;KIF5B;CTSK;AP1S2;AP2S1;CTSH;CTSF;KIFAP3;CTSD;RILP;AP2M1;AP1M2;CTSB;CTSA;CD74;DYNC1H1;SEC24A;AP2B1;DNM2;KIF2A;HLA-DQB1;RAB7A |

| | | | | | | |
|---|---|---|---|---|---|---|
| 272 | VEGFR2 mediated vascular permeability Homo sapiens R-HSA-5218920 | 9.94E-04 | 5.23E-03 | 3.63E+00 | 2.51E+01 | PRR5;VAV3;HSP90AA1;JUP;PDPK1;MAPKAP1;MTOR;PAK1;AKT2;MLST8;AKT1;RAC1;CALM1;PAK2 |
| 273 | Cooperation of PDCL (PhLP1) and TRiC/CCT in G-protein beta folding Homo sapiens R-HSA-6814122 | 1.16E-03 | 6.07E-03 | 2.78E+00 | 1.88E+01 | CCT3;CCT2;CSNK2A2;GNAI3;GNAI1;GNAI2;CCT6A;GNAO1;GNG10;GNG5;TCP1;CSNK2B;PDCL;CCT8;GNB5;RGS11;CCT7;CCT5;CCT4 |
| 274 | Protein folding Homo sapiens R-HSA-391251 | 1.27E-03 | 6.62E-03 | 1.95E+00 | 1.30E+01 | HDAC3;GNAI3;ARL2;GNAI1;GNAI2;TUBA1C;TUBB6;GNG10;TUBA1B;GNG5;CCT8;CCT7;CCT5;CCT4;CCT3;CCT2;VBP1;CSNK2A2;STAT3;TBCB;TBCA;TUBB4B;TUBA4A;CCT6A;GNAO1;XRN2;CSNK2B;TCP1;PDCL;PFDN1;PFDN2;FKBP9;GNB5;RGS11;FBXL5;PFDN4;PFDN5 |
| 275 | Formation of tubulin folding intermediates by CCT/TriC Homo sapiens R-HSA-389960 | 1.45E-03 | 7.54E-03 | 3.65E+00 | 2.38E+01 | CCT3;CCT2;TUBB4B;TUBA4A;CCT6A;TUBA1C;TUBA1B;TUBB6;TCP1;CCT8;CCT7;CCT5;CCT4 |
| 276 | Synthesis and interconversion of nucleotide di- and triphosphates Homo sapiens R-HSA-499943 | 1.45E-03 | 7.54E-03 | 3.65E+00 | 2.38E+01 | DTYMK;TXNRD1;AK1;GSR;AK2;NME2;NME4;CTPS1;AK6;RRM2B;GUK1;CMPK1;DCTPP1 |
| 277 | Energy dependent regulation of mTOR by LKB1-AMPK Homo sapiens R-HSA-380972 | 1.57E-03 | 8.12E-03 | 3.37E+00 | 2.17E+01 | PRKAA1;CAB39L;STRADB;PRKAG1;PRKAG2;MTOR;RRAGA;RRAGB;RHEB;MLST8;LAMTOR1;LAMTOR4;LAMTOR3;LAMTOR5 |
| 278 | Iron uptake and transport Homo sapiens R-HSA-917937 | 1.63E-03 | 8.41E-03 | 2.67E+00 | 1.71E+01 | ATP6V1A;SLC46A1;ATP6V1G1;ATP6V0B;ATP6V0E1;ATP6AP1;SLC40A1;CYBRD1;MCOLN1;FLVCR1;ATP6V1H;ATP6V0E2;ATP6V1E1;ATP6V0D1;ATP6V1D;ATP6V0C;HMOX2;ATP6V1C1;ATP6V1F |

| | | | | | | |
|---|---|---|---|---|---|---|
| 279 | FCERI mediated MAPK activation Homo sapiens R-HSA-2871796 | 1.81E-03 | 9.31E-03 | 1.48E+00 | 9.35E+00 | YWHAB;RASGRF2;PEBP1;WDR83;ARRB1;FGF2;PSMD8;PSMD9;PSMD6;PSMD7;PSMD4;PPP2R5E;PSMD2;PSMD3;PEA15;PSMD1;RAC1;JAK2;SPTAN1;JAK1;VAV3;MAP2K2;DUSP1;PPP2R5A;DUSP8;RBX1;BTC;PSMA5;PSMA6;PSMA3;PSMA4;PSMA1;PSMA2;RASA1;AKAP9;PSME1;PSME2;TLN1;HBEGF;PSMD10;SPTBN5;PSMD12;SHC2;PSMD11;PSMD14;PSMD13;CUL3;PDGFB;RASAL1;PHB;PSMA7;RASGRP3;PPP2CA;PSMB6;RAP1B;PSMB7;PPP2CB;MAPK9;PAK1;PSMB5;RAP1A;PSMB2;PSMB3;PSMB1;UBC;MAPK1;CSK;FYN;PSMF1;RPS27A;PAK2;CAMK2G;MAP3K1;DAB2IP;PTK2;PSMB8;PSMC5;PSMC6;PSMC3;PSMC4;PTPRA;PSMC1;PSMC2;FGF18;GRB2;KRAS;CALM1;LAMTOR3 |
| 280 | eNOS activation and regulation Homo sapiens R-HSA-203765 | 1.88E-03 | 9.56E-03 | 4.12E+00 | 2.58E+01 | GCHFR;LYPLA1;HSP90AA1;GCH1;SPR;NOSIP;AKT1;WASL;CALM1;PTS;DNM2 |
| 281 | Metabolism of nitric oxide Homo sapiens R-HSA-202131 | 1.88E-03 | 9.56E-03 | 4.12E+00 | 2.58E+01 | GCHFR;LYPLA1;HSP90AA1;GCH1;SPR;NOSIP;AKT1;WASL;CALM1;PTS;DNM2 |
| 282 | Response to elevated platelet cytosolic Ca2+ Homo sapiens R-HSA-76005 | 1.92E-03 | 9.75E-03 | 1.85E+00 | 1.16E+01 | LGALS3BP;APP;CD63;SERPINA1;STXBP3;WDR1;PDGFB;CLU;KNG1;CYB5R1;LAMP2;CFL1;HABP4;TIMP3;QSOX1;STX4;FAM3C;TIMP1;CAP1;TGFB2;TGFB1;HSPA5;APLP2;ACTN1;RARRES2;PCDH7;VEGFB;TUBA4A;VEGFA;SOD1;CALU;CD9;TLN1;PFN1;CALM1;HRG;ALDOA;GAS6;PPIA |
| 283 | Tetrahydrobiopterin (BH4) synthesis, recycling, salvage and regulation Homo sapiens R-HSA-1474151 | 2.04E-03 | 1.03E-02 | 7.85E+00 | 4.86E+01 | GCHFR;HSP90AA1;SPR;GCH1;AKT1;CALM1;PTS |
| 284 | Cholesterol biosynthesis Homo sapiens R-HSA-191273 | 2.13E-03 | 1.07E-02 | 3.67E+00 | 2.26E+01 | FDPS;IDI1;SQLE;EBP;ARV1;MVK;CYP51A1;SC5D;DHCR24;MSMO1;HSD17B7;FDFT1 |
| 285 | FGFR2 alternative splicing Homo sapiens R-HSA-6803529 | 2.30E-03 | 1.15E-02 | 3.37E+00 | 2.05E+01 | NCBP1;GTF2F2;TIAL1;HNRNPM;POLR2B;POLR2C;ESRP1;HNRNPF;POLR2E;POLR2H;POLR2J;POLR2K;POLR2L |

| | | | | | | |
|---|---|---|---|---|---|---|
| 286 | Insulin receptor recycling Homo sapiens R-HSA-77387 | 2.30E-03 | 1.15E-02 | 3.37E+00 | 2.05E+01 | ATP6V1A;ATP6V1G1;ATP6V0B;ATP6V0E1;ATP6AP1;ATP6V1H;ATP6V0E2;ATP6V1E1;ATP6V0D1;ATP6V1D;ATP6V0C;ATP6V1C1;ATP6V1F |
| 287 | Transferrin endocytosis and recycling Homo sapiens R-HSA-917977 | 2.40E-03 | 1.20E-02 | 3.14E+00 | 1.90E+01 | ATP6V1A;ATP6V1G1;ATP6V0B;ATP6V0E1;ATP6AP1;MCOLN1;ATP6V1H;ATP6V0E2;ATP6V1E1;ATP6V0D1;ATP6V1D;ATP6V0C;ATP6V1C1;ATP6V1F |
| 288 | Regulation of PLK1 Activity at G2/M Transition Homo sapiens R-HSA-2565942 | 2.42E-03 | 1.20E-02 | 1.96E+00 | 1.18E+01 | YWHAE;CEP57;DCTN2;CETN2;SSNA1;DCTN3;CUL1;PPP1CB;ACTR1A;CCNB1;HAUS8;HAUS7;PRKAR2B;UBC;RPS27A;YWHAG;RAB8A;SKP1;OFD1;DYNC1H1;HSP90AA1;FBXW11;TUBB;TUBG1;TUBB4B;TUBA4A;HAUS1;AKAP9;MAPRE1;AJUBA;OPTN;PAFAH1B1 |
| 289 | Recycling of eIF2:GDP Homo sapiens R-HSA-72731 | 2.63E-03 | 1.30E-02 | 1.01E+01 | 6.00E+01 | EIF2B5;EIF2B4;EIF2B3;EIF2B2;EIF2S2;EIF2S1 |
| 290 | mitochondrial fatty acid beta-oxidation of saturated fatty acids Homo sapiens R-HSA-77286 | 2.63E-03 | 1.30E-02 | 1.01E+01 | 6.00E+01 | HADHB;HADHA;ACADVL;ECHS1;ACADM;HADH |
| 291 | Oxygen-dependent proline hydroxylation of Hypoxia-inducible Factor Alpha Homo sapiens R-HSA-1234176 | 2.74E-03 | 1.35E-02 | 4.21E+00 | 2.48E+01 | EGLN1;EGLN2;UBE2D2;UBE2D3;CUL2;UBC;UBE2D1;RPS27A;HIF1A;RBX1 |
| 292 | RHO GTPase Effectors Homo sapiens R-HSA-195258 | 2.78E-03 | 1.36E-02 | 1.49E+00 | 8.77E+00 | CYFIP2;YWHAE;ITGB1;NCKAP1;CDKN1B;RTKN;YWHAB;ARPC1B;ARPC1A;BRK1;WASL;MYLK;ACTG1;PPP1CB;PPP1CC;YWHAQ;PPP2R5E;KIF5B;RAC1;HIST1H2AC;YWHAG;YWHAH;ACTR3;NCOA2;ACTR2;NUDC;H2AFZ;PDPK1;RHOG;PPP2R5A;CYBA;RHOC;RHOD;YWHAZ;RHOA;MYL6;CLIP1;CTTN;KIF2A;TAX1BP3;PFN1;MAPRE1;RHOQ;PFN2;PAFAH1B1;KLC1;PMF1;MYL12B;SKA2;CENPC;PPP2CA;CDC42;PPP2CB;PAK1;RHPN1;DVL3;MAPK1;RHPN2;BUB3;PAK2;MYH10;WASF2;LIMK2;H3F3A;ARPC5;PTK2;FMNL3;FMNL1;ABI2;ARPC2;ARPC3;TAOK1;NOXA1;GRB2;GOPC;PKN1;NF2;CALM1 |

| | | | | | | |
|---|---|---|---|---|---|---|
| 293 | Platelet degranulation Homo sapiens R-HSA-114608 | 2.79E-03 | 1.36E-02 | 1.84E+00 | 1.08E+01 | LGALS3BP;APP;CD63;SERPINA1;WDR1;PDGFB;CLU;KNG1;CYB5R1;LAMP2;CFL1;HABP4;TIMP3;QSOX1;FAM3C;TIMP1;CAP1;TGFB2;TGFB1;HSPA5;APLP2;ACTN1;RARRES2;PCDH7;VEGFB;TUBA4A;VEGFA;SOD1;CALU;CD9;TLN1;PFN1;CALM1;HRG;ALDOA;GAS6;PPIA |
| 294 | Chaperonin-mediated protein folding Homo sapiens R-HSA-390466 | 3.04E-03 | 1.48E-02 | 1.88E+00 | 1.09E+01 | HDAC3;GNAI3;GNAI1;GNAI2;TUBA1C;TUBB6;TUBA1B;GNG10;GNG5;CCT8;CCT7;CCT5;CCT4;CCT3;CCT2;VBP1;CSNK2A2;STAT3;TUBB4B;TUBA4A;CCT6A;GNAO1;TCP1;XRN2;CSNK2B;PDCL;PFDN1;PFDN2;FKBP9;GNB5;RGS11;PFDN4;FBXL5;PFDN5 |
| 295 | AKT phosphorylates targets in the cytosol Homo sapiens R-HSA-198323 | 3.16E-03 | 1.53E-02 | 5.39E+00 | 3.10E+01 | GSK3B;GSK3A;CDKN1B;BAD;AKT2;AKT1S1;MDM2;AKT1 |
| 296 | Assembly Of The HIV Virion Homo sapiens R-HSA-175474 | 3.16E-03 | 1.53E-02 | 5.39E+00 | 3.10E+01 | TSG101;UBC;VPS37D;FURIN;VPS37B;RPS27A;PPIA;VPS28 |
| 297 | Cargo concentration in the ER Homo sapiens R-HSA-5694530 | 3.54E-03 | 1.70E-02 | 2.81E+00 | 1.58E+01 | MCFD2;TMED10;SERPINA1;SEC24A;SAR1B;GOSR2;CTSZ;MIA3;LMAN1;CNIH1;LMAN2;TMED2;STX5;CD59;SEC22B |
| 298 | CD28 co-stimulation Homo sapiens R-HSA-389356 | 3.54E-03 | 1.70E-02 | 2.81E+00 | 1.58E+01 | PRR5;YES1;PDPK1;MAPKAP1;PIK3R3;MTOR;CDC42;PAK1;AKT2;MLST8;AKT1;FYN;GRB2;RAC1;PAK2 |
| 299 | Cell-extracellular matrix interactions Homo sapiens R-HSA-446353 | 4.02E-03 | 1.91E-02 | 4.33E+00 | 2.39E+01 | ITGB1;VASP;FBLIM1;ACTN1;TESK1;ILK;RSU1;FERMT2;LIMS1 |
| 300 | Sema3A PAK dependent Axon repulsion Homo sapiens R-HSA-399954 | 4.02E-03 | 1.91E-02 | 4.33E+00 | 2.39E+01 | HSP90AA1;PAK1;HSP90AB1;CFL1;PLXNA2;FYN;RAC1;PAK2;PLXNA4 |

| | | | | | | |
|---|---|---|---|---|---|---|
| 301 | TGF-beta receptor signaling in EMT (epithelial to mesenchymal transition) Homo sapiens R-HSA-2173791 | 4.02E-03 | 1.91E-02 | 4.33E+00 | 2.39E+01 | FKBP1A;TGFB1;PARD6A;UBC;ARHGEF18;CGN;RPS27A;PRKCZ;RHOA |
| 302 | DNA Repair Homo sapiens R-HSA-73894 | 4.04E-03 | 1.91E-02 | 1.44E+00 | 7.92E+00 | MUS81;MPG;CETN2;UBE2L6;BABAM1;YY1;HERC2;PPP4R2;SUMO1;SUMO3;UIMC1;SUMO2;POLK;KPNA2;HMGN1;USP7;PARP1;RFC1;ACTL6A;USP45;LIG4;RAD23A;ACTR8;RAD23B;RBX1;RAD51B;DDB1;RNF168;MSH6;MSH2;MSH3;UBE2V2;PPIE;NEIL1;INO80B;VCP;PAXIP1;PCNA;PRKDC;XPA;COPS7B;BRCC3;UNG;TFPT;PPP4C;NTHL1;POLR2B;POLR2C;TDG;UBC;POLD2;POLR2E;ASCC3;POLR2H;RPS27A;POLR2J;POLR2K;POLR2L;MBD4;XRCC6;UBE2I;UBE2B;XRCC5;MGMT;FANCL;POLM;MCRS1;SMARCA5;GTF2H4;COPS4;XAB2;COPS3;COPS6;RAD50;COPS5;RPA3;UBE2T;COPS2;ERCC1;CDK2;RAD17;TCEA1;RAD1;ERCC5;MNAT1 |
| 303 | Assembly of the primary cilium Homo sapiens R-HSA-5617833 | 4.07E-03 | 1.92E-02 | 1.56E+00 | 8.57E+00 | YWHAE;CEP57;ARF4;CETN2;PKD2;PKD1;HSPB11;ACTR1A;KIFAP3;TNPO1;IFT57;CC2D2A;YWHAG;RAB8A;OFD1;CCT3;CCT2;MKKS;HSP90AA1;TUBB;IFT80;CYS1;TUBG1;WDR34;TUBA4A;HAUS1;AKAP9;TCP1;EXOC6;MAPRE1;PAFAH1B1;UNC119B;DCTN2;ARL3;SSNA1;DCTN3;ASAP1;IFT74;HAUS8;ARL13B;KIF3A;PRKAR2B;HAUS7;NPHP1;TCTEX1D2;INPP5E;PDE6D;CCT8;CCT5;CCT4;DYNC1H1;TUBB4B;RAB11A;IFT20;AHI1;IFT22;WDR60;TRIP11;DYNLRB1 |
| 304 | p75 NTR receptor-mediated signalling Homo sapiens R-HSA-193704 | 4.38E-03 | 2.06E-02 | 1.91E+00 | 1.04E+01 | YWHAE;HDAC2;HDAC3;HDAC1;MAGED1;RASGRF2;AATF;RTN4;ARHGAP4;APH1A;APH1B;IRAK1;MCF2L;UBC;RAC1;RPS27A;PSENEN;VAV3;ARHGEF12;BAD;ARHGEF16;ARHGEF18;RHOA;NFKB1;NFKBIA;TIAM1;ARHGEF3;ARHGEF1;ARHGEF7;SQSTM1 |
| 305 | Signaling by FGFR2 IIIa TM Homo sapiens R-HSA-8851708 | 4.61E-03 | 2.16E-02 | 3.74E+00 | 2.01E+01 | NCBP1;POLR2B;POLR2C;POLR2E;POLR2H;FGF2;GTF2F2;POLR2J;POLR2K;POLR2L |
| 306 | HSF1-dependent transactivation Homo sapiens R-HSA-3371571 | 5.01E-03 | 2.34E-02 | 2.66E+00 | 1.41E+01 | HSPA8;HSP90AA1;HSP90AB1;HSBP1;MRPL18;PTGES3;HSPA2;MTOR;HSPH1;DNAJB6;AKT1S1;MLST8;FKBP4;CAMK2G;HSPA1A |

| | | | | | | |
|---|---|---|---|---|---|---|
| 307 | Negative regulators of RIG-I/MDA5 signaling Homo sapiens R-HSA-936440 | 5.01E-03 | 2.34E-02 | 2.66E+00 | 1.41E+01 | UBE2D2;UBE2D3;UBE2D1;UBE2L6;ITCH;MAVS;TBK1;RNF216;IRF3;UBC;TAX1BP1;PIN1;RPS27A;UBE2K;ATG5 |
| 308 | Semaphorin interactions Homo sapiens R-HSA-373755 | 5.55E-03 | 2.58E-02 | 2.01E+00 | 1.04E+01 | ITGB1;GSK3B;HSP90AB1;CRMP1;MYL12B;CDC42;PAK1;DPYSL2;CFL1;PLXNA2;FYN;RAC1;PAK2;MYH10;PLXNA4;HSP90AA1;ARHGEF12;SEMA4D;LIMK2;RHOG;RHOC;RHOA;MYL6;CDK5;TLN1 |
| 309 | Nef Mediated CD4 Down-regulation Homo sapiens R-HSA-167590 | 6.35E-03 | 2.93E-02 | 6.73E+00 | 3.41E+01 | ARF1;AP2S1;ATP6V1H;AP2B1;AP2A2;AP2M1 |
| 310 | RHO GTPases Activate Rhotekin and Rhophilins Homo sapiens R-HSA-5666185 | 6.35E-03 | 2.93E-02 | 6.73E+00 | 3.41E+01 | RTKN;RHPN1;TAX1BP3;RHPN2;RHOC;RHOA |
| 311 | G1 Phase Homo sapiens R-HSA-69236 | 6.60E-03 | 3.01E-02 | 2.45E+00 | 1.23E+01 | RB1;CDKN2D;CDKN1B;CUL1;PPP2R2A;CKS1B;PPP2CA;PPP2CB;CDK6;TFDP2;CDK4;UBC;E2F4;RPS27A;MNAT1;SKP1 |
| 312 | Cyclin D associated events in G1 Homo sapiens R-HSA-69231 | 6.60E-03 | 3.01E-02 | 2.45E+00 | 1.23E+01 | RB1;CDKN2D;CDKN1B;CUL1;PPP2R2A;CKS1B;PPP2CA;PPP2CB;CDK6;TFDP2;CDK4;UBC;E2F4;RPS27A;MNAT1;SKP1 |
| 313 | mRNA 3'-end processing Homo sapiens R-HSA-72187 | 6.64E-03 | 3.01E-02 | 2.17E+00 | 1.09E+01 | FYTTD1;NCBP1;RBM8A;POLDIP3;CPSF3;EIF4A3;THOC3;THOC2;THOC5;CDC40;DDX39A;FIP1L1;CLP1;SARNP;MAGOH;SRSF2;CSTF1;SRSF4;RNPS1;SRSF9 |
| 314 | Post-Elongation Processing of Intron-Containing pre-mRNA Homo sapiens R-HSA-112296 | 6.64E-03 | 3.01E-02 | 2.17E+00 | 1.09E+01 | FYTTD1;NCBP1;RBM8A;POLDIP3;CPSF3;EIF4A3;THOC3;THOC2;THOC5;CDC40;DDX39A;FIP1L1;CLP1;SARNP;MAGOH;SRSF2;CSTF1;SRSF4;RNPS1;SRSF9 |
| 315 | Cargo trafficking to the periciliary membrane Homo sapiens R-HSA-5620920 | 6.64E-03 | 3.01E-02 | 2.17E+00 | 1.09E+01 | CCT3;ARF4;CCT2;UNC119B;MKKS;ARL3;ASAP1;CYS1;PKD2;PKD1;RAB11A;ARL13B;TCP1;INPP5E;PDE6D;EXOC6;CCT8;RAB8A;CCT5;CCT4 |

| | | | | | | |
|---|---|---|---|---|---|---|
| 316 | N-glycan trimming in the ER and Calnexin/Calreticulin cycle Homo sapiens R-HSA-532668 | 6.82E-03 | 3.09E-02 | 3.79E+00 | 1.89E+01 | PDIA3;EDEM3;OS9;EDEM2;SEL1L;DERL2;MLEC;MOGS;CALR |
| 317 | Regulation of TP53 Degradation Homo sapiens R-HSA-6804757 | 6.94E-03 | 3.13E-02 | 2.53E+00 | 1.26E+01 | PRR5;USP7;PDPK1;MAPKAP1;USP2;MTOR;PPP2CA;PPP2CB;AKT2;UBC;CDK2;MLST8;MDM2;AKT1;RPS27A |
| 318 | EPH-ephrin mediated repulsion of cells Homo sapiens R-HSA-3928665 | 7.16E-03 | 3.22E-02 | 2.21E+00 | 1.09E+01 | PSENEN;VAV3;YES1;EPHA6;CLTC;CLTB;CLTA;AP2B1;EFNA4;AP2A2;APH1A;TIAM1;EFNA3;APH1B;AP2S1;FYN;RAC1;AP2M1;EPHA2 |
| 319 | HSF1 activation Homo sapiens R-HSA-3371511 | 7.47E-03 | 3.35E-02 | 2.74E+00 | 1.34E+01 | YWHAE;VCP;HSP90AA1;HSP90AB1;HSBP1;MRPL18;PTGES3;EEF1A1;HSPH1;DNAJB6;RPA3;FKBP4;HSPA1A |
| 320 | RNA Polymerase III Transcription Termination Homo sapiens R-HSA-73980 | 7.58E-03 | 3.35E-02 | 3.09E+00 | 1.51E+01 | SSB;CRCP;POLR3GL;POLR3D;NFIC;POLR1D;POLR2E;POLR2H;POLR3K;POLR2K;POLR2L |
| 321 | Abortive elongation of HIV-1 transcript in the absence of Tat Homo sapiens R-HSA-167242 | 7.58E-03 | 3.35E-02 | 3.09E+00 | 1.51E+01 | NCBP1;POLR2B;POLR2C;SUPT4H1;POLR2E;NELFCD;POLR2H;GTF2F2;POLR2J;POLR2K;POLR2L |
| 322 | Downregulation of SMAD2/3:SMAD4 transcriptional activity Homo sapiens R-HSA-2173795 | 7.58E-03 | 3.35E-02 | 3.09E+00 | 1.51E+01 | NCOR2;SMAD4;NCOR1;SMURF2;PARP1;HDAC1;UBE2D3;UBC;UBE2D1;RPS27A;SKIL |

| | | | | | | |
|---|---|---|---|---|---|---|
| 323 | APC/C:Cdc20 mediated degradation of Cyclin B Homo sapiens R-HSA-174048 | 7.58E-03 | 3.35E-02 | 3.09E+00 | 1.51E+01 | CCNB1;ANAPC15;ANAPC16;CDC26;UBC;CDC27;UBE2E1;UBE2D1;ANAPC5;RPS27A;ANAPC11 |
| 324 | Regulation of Hypoxia-inducible Factor (HIF) by oxygen Homo sapiens R-HSA-1234174 | 7.60E-03 | 3.35E-02 | 2.89E+00 | 1.41E+01 | EGLN1;EGLN2;UBE2D2;UBE2D3;CUL2;UBC;UBE2D1;ARNT;RPS27A;HIF1A;RBX1;VEGFA |
| 325 | Cellular response to hypoxia Homo sapiens R-HSA-2262749 | 7.60E-03 | 3.35E-02 | 2.89E+00 | 1.41E+01 | EGLN1;EGLN2;UBE2D2;UBE2D3;CUL2;UBC;UBE2D1;ARNT;RPS27A;HIF1A;RBX1;VEGFA |
| 326 | Gap-filling DNA repair synthesis and ligation in TC-NER Homo sapiens R-HSA-6782210 | 8.25E-03 | 3.62E-02 | 1.99E+00 | 9.53E+00 | USP7;PCNA;RFC1;GTF2H4;RBX1;DDB1;XAB2;POLR2B;POLR2C;RPA3;UBC;POLD2;POLR2E;PPIE;TCEA1;POLK;POLR2H;RPS27A;MNAT1;POLR2J;POLR2K;HMGN1;POLR2L |
| 327 | Intrinsic Pathway for Apoptosis Homo sapiens R-HSA-109606 | 8.27E-03 | 3.62E-02 | 2.29E+00 | 1.10E+01 | YWHAE;PPP1R13B;APAF1;YWHAB;BAD;YWHAZ;BBC3;CASP7;YWHAQ;AKT2;NMT1;AKT1;BAX;CYCS;BID;YWHAG;YWHAH |
| 328 | Chk1/Chk2(Cds1) mediated inactivation of Cyclin B:Cdk1 complex Homo sapiens R-HSA-75035 | 8.69E-03 | 3.78E-02 | 4.71E+00 | 2.24E+01 | YWHAE;CCNB1;YWHAQ;YWHAB;YWHAZ;YWHAG;YWHAH |
| 329 | 2-LTR circle formation Homo sapiens R-HSA-164843 | 8.71E-03 | 3.78E-02 | 8.41E+00 | 3.99E+01 | XRCC6;XRCC5;HMGA1;PSIP1;LIG4 |
| 330 | Nef Mediated CD8 Down-regulation Homo sapiens R-HSA-182218 | 8.71E-03 | 3.78E-02 | 8.41E+00 | 3.99E+01 | AP2S1;ATP6V1H;AP2B1;AP2A2;AP2M1 |

| | | | | | | |
|---|---|---|---|---|---|---|
| 331 | Regulation of TP53 Expression and Degradation Homo sapiens R-HSA-6806003 | 9.42E-03 | 4.07E-02 | 2.41E+00 | 1.12E+01 | PRR5;USP7;PDPK1;MAPKAP1;USP2;MTOR;PPP2CA;PPP2CB;AKT2;UBC;CDK2;MLST8;MDM2;AKT1;RPS27A |
| 332 | Innate Immune System Homo sapiens R-HSA-168249 | 1.02E-02 | 4.42E-02 | 1.22E+00 | 5.57E+00 | ATF1;CYFIP2;NCKAP1;APP;UBE2L6;WDR83;PSMD8;PSMD9;LGALS3;PSMD6;PSMD7;TBK1;PSMD4;AKT2;PSMD2;PSMD3;AKT1;PSMD1;PIP4K2C;PRSS3;IER3;SKP1;HLA-C;HLA-E;SIGIRR;BTC;PRKAR1A;AKAP9;PSME1;PSME2;TLN1;SPTBN5;DHX9;PRKDC;MAPKAP1;PDE1A;PDGFB;PIK3R3;BCL10;RNF216;PRKAR2B;PRKAR2A;PIP5K1B;PSMF1;FADD;ATG5;XRCC6;YES1;XRCC5;MYO5A;PTK2;AGO2;GRB2;CALM1;CDKN1B;ARPC1B;ARPC1A;WASL;ECSIT;EEA1;PPP3CB;CTSL;CTSK;PEA15;RAC1;JAK2;SPTAN1;JAK1;CTSB;VAV3;DUSP3;DUSP1;DUSP8;PSMA5;PSMA6;PSMA3;SIKE1;PSMA4;PSMA1;PSMA2;PELI1;TAX1BP1;CD46;TLR5;HBEGF;ADCY3;TANK;PSMA7;ADCY5;PSMB6;PPP2CA;PSMB7;PPP2CB;PAK1;PSMB5;PSMB2;PSMB3;PSMB1;CD59;RPS27A;PAK2;CD55;MAP3K1;DAB2IP;PSMB8;PSMC5;ITCH;PSMC6;PSMC3;ABI2;PSMC4;PSMC1;CAPZA1;PSMC2;CAPZA2;LAMTOR3;CRCP;UBE2D2;UBE2D3;UBE2D1;PEBP1;CLU;FGF2;ACTG1;PYCARD;CFL1;MAP2K3;MAP2K2;FBXW11;CNPY3;RBX1;ELMO2;UBE2V1;PSMD10;SHC2;PSMD12;PSMD11;PSMD14;PSMD13;CUL3;CUL1;ITPR2;RASAL1;PHB;RASGRP3;RAP1B;RAP1A;SOCS1;IRAK1;UBC;POLR3GL;ARPC5;NFKB1;NFKBIA;MAVS;ARPC2;ARPC3;PTPRA;FGF18;MDM2;PIN1;UBE2K;UBE2M;PRR5;GSK3B;GSK3A;HSP90AB1;YWHAB;RASGRF2;ITGB2;BRK1;ARRB1;HMGB1;PPP2R5E;MLST8;LRRFIP1;ACTR3;ACTR2;HSP90AA1;PDPK1;PPP2R5A;IRAK4;DDOST;DNM2;CREB1;IRF3;RASA1;IRF2;AKT1S1;POLR1D;DOCK1;UNC93B1;CDC42;MAPK9;POLR2E;[illegible] |
| 333 | Uptake and actions of bacterial toxins Homo sapiens R-HSA-5339562 | 1.09E-02 | 4.67E-02 | 2.69E+00 | 1.22E+01 | MAP2K3;HSP90AA1;HSP90AB1;MAP2K2;PDCD6IP;SYT1;TXNRD1;CD9;FURIN;EEF2;CALM1;HBEGF |
| 334 | RNA Polymerase III Chain Elongation Homo sapiens R-HSA-73780 | 1.09E-02 | 4.67E-02 | 3.37E+00 | 1.52E+01 | CRCP;POLR3GL;POLR3D;POLR1D;POLR2E;POLR2H;POLR3K;POLR2K;POLR2L |

| | | | | | | |
|---|---|---|---|---|---|---|
| 335 | DARPP-32 events Homo sapiens R-HSA-180024 | 1.11E-02 | 4.74E-02 | 2.85E+00 | 1.28E+01 | PPP2CA;PPP2CB;PPP3CB;CDK5;PRKAR1A;PRKAR2B;PDE4D;PRKAR2A;PDE4A;CALM1;PPP1CA |
| 336 | Sema4D induced cell migration and growth-cone collapse Homo sapiens R-HSA-416572 | 1.11E-02 | 4.74E-02 | 2.85E+00 | 1.28E+01 | CDC42;MYL6;ARHGEF12;SEMA4D;LIMK2;RHOG;RAC1;RHOC;MYH10;RHOA;MYL12B |
| 337 | Beta oxidation of hexanoyl-CoA to butanoyl-CoA Homo sapiens R-HSA-77350 | 1.13E-02 | 4.77E-02 | 1.35E+01 | 6.03E+01 | HADHB;HADHA;ECHS1;HADH |
| 338 | Beta oxidation of lauroyl-CoA to decanoyl-CoA-CoA Homo sapiens R-HSA-77310 | 1.13E-02 | 4.77E-02 | 1.35E+01 | 6.03E+01 | HADHB;HADHA;ECHS1;HADH |
| 339 | Platelet activation, signaling and aggregation Homo sapiens R-HSA-76002 | 1.13E-02 | 4.78E-02 | 1.40E+00 | 6.26E+00 | LGALS3BP;APP;SERPINA1;DAGLA;WDR1;ARRB1;CLU;AKT2;LAMP2;CFL1;HABP4;AKT1;TIMP3;QSOX1;TIMP1;RAC1;FAM3C;JAK2;JAK1;VAV3;CAP1;PDPK1;ACTN1;APLP2;F2R;RHOG;PLA2G4A;YWHAZ;RHOA;TUBA4A;TLN1;PFN1;ALDOA;GAS6;PPIA;RAPGEF3;CD63;STXBP3;PDGFB;GNAI3;PIK3R3;ITPR2;PRKCZ;GNAI1;KNG1;GNAI2;RAP1B;CDC42;CYB5R1;GNG10;RAP1A;GNG5;MAPK1;STX4;CSK;FYN;TGFB2;TGFB1;HSPA5;RARRES2;PCDH7;VEGFB;PTPN11;PTK2;VEGFA;SOD1;CALU;CD9;GRB2;PTPN6;GNB5;CALM1;HRG;BCAR1 |
| 340 | Signaling by PTK6 Homo sapiens R-HSA-8848021 | 1.14E-02 | 4.81E-02 | 1.88E+00 | 8.41E+00 | KHDRBS1;CDKN1B;STAT3;DAB2IP;RASAL1;CBL;HIF1A;RHOA;BTC;DOK1;SFPQ;CDK4;RASA1;UBC;CDK2;AKT1;ELMO2;STAP2;KRAS;RAC1;RPS27A;DOCK1;BCAR1;HBEGF |
| 341 | Constitutive Signaling by NOTCH1 HD+PEST Domain Mutants Homo sapiens R-HSA-2894862 | 1.23E-02 | 5.10E-02 | 1.97E+00 | 8.64E+00 | PSENEN;JAG2;HDAC2;HDAC3;HDAC10;HDAC1;HDAC11;CUL1;MIB2;CCNC;RBPJ;RBX1;NCOR2;APH1A;KAT2B;NCOR1;APH1B;SNW1;UBC;RPS27A;SKP1 |

| | | | | | | |
|---|---|---|---|---|---|---|
| 342 | Signaling by NOTCH1 in Cancer Homo sapiens R-HSA-2644603 | 1.23E-02 | 5.10E-02 | 1.97E+00 | 8.64E+00 | PSENEN;JAG2;HDAC2;HDAC3;HDAC10;HDAC1;HDAC11;CUL1;MIB2;CCNC;RBPJ;RBX1;NCOR2;APH1A;KAT2B;NCOR1;APH1B;SNW1;UBC;RPS27A;SKP1 |
| 343 | Signaling by NOTCH1 PEST Domain Mutants in Cancer Homo sapiens R-HSA-2644602 | 1.23E-02 | 5.10E-02 | 1.97E+00 | 8.64E+00 | PSENEN;JAG2;HDAC2;HDAC3;HDAC10;HDAC1;HDAC11;CUL1;MIB2;CCNC;RBPJ;RBX1;NCOR2;APH1A;KAT2B;NCOR1;APH1B;SNW1;UBC;RPS27A;SKP1 |
| 344 | Constitutive Signaling by NOTCH1 PEST Domain Mutants Homo sapiens R-HSA-2644606 | 1.23E-02 | 5.10E-02 | 1.97E+00 | 8.64E+00 | PSENEN;JAG2;HDAC2;HDAC3;HDAC10;HDAC1;HDAC11;CUL1;MIB2;CCNC;RBPJ;RBX1;NCOR2;APH1A;KAT2B;NCOR1;APH1B;SNW1;UBC;RPS27A;SKP1 |
| 345 | Signaling by NOTCH1 HD+PEST Domain Mutants in Cancer Homo sapiens R-HSA-2894858 | 1.23E-02 | 5.10E-02 | 1.97E+00 | 8.64E+00 | PSENEN;JAG2;HDAC2;HDAC3;HDAC10;HDAC1;HDAC11;CUL1;MIB2;CCNC;RBPJ;RBX1;NCOR2;APH1A;KAT2B;NCOR1;APH1B;SNW1;UBC;RPS27A;SKP1 |
| 346 | RNA Polymerase II Pre-transcription Events Homo sapiens R-HSA-674695 | 1.26E-02 | 5.20E-02 | 1.89E+00 | 8.27E+00 | GTF2A1;SUPT16H;GTF2A2;NCBP1;TAF13;GTF2B;TAF10;TAF11;SUPT4H1;GTF2F2;GTF2H4;GTF2E2;CDK9;POLR2B;POLR2C;POLR2E;TCEA1;NELFCD;POLR2H;MNAT1;POLR2J;POLR2K;POLR2L |
| 347 | Toll-Like Receptors Cascades Homo sapiens R-HSA-168898 | 1.27E-02 | 5.24E-02 | 1.55E+00 | 6.75E+00 | ATF1;APP;DHX9;UNC93B1;UBE2D2;UBE2D3;ITGB2;CUL1;UBE2D1;HMGB1;ECSIT;TANK;EEA1;PPP2CA;PPP2CB;MAPK9;TBK1;SOCS1;IRAK1;CTSL;CTSK;UBC;MAPK1;FADD;RPS27A;SKP1;CTSB;MAP2K3;DUSP3;MAP3K1;FBXW11;PTPN11;CNPY3;IRAK4;NFKB1;DNM2;SIGIRR;NFKBIA;CREB1;IRF3;TNIP2;PELI1;UBE2V1;TLR5 |
| 348 | Base-Excision Repair, AP Site Formation Homo sapiens R-HSA-73929 | 1.28E-02 | 5.25E-02 | 5.05E+00 | 2.20E+01 | MBD4;MPG;NTHL1;TDG;NEIL1;UNG |

| | | | | | | |
|---|---|---|---|---|---|---|
| 349 | Gap junction degradation Homo sapiens R-HSA-190873 | 1.28E-02 | 5.25E-02 | 5.05E+00 | 2.20E+01 | MYO6;CLTC;CLTB;CLTA;AP2M1;DNM2 |
| 350 | Activation of anterior HOX genes in hindbrain development during early embryogenesis Homo sapiens R-HSA-5617472 | 1.32E-02 | 5.38E-02 | 1.71E+00 | 7.42E+00 | RARG;PAXIP1;HDAC3;HOXD1;ASH2L;CTCF;YY1;PAGR1;RBBP4;POLR2B;POLR2C;POLR2E;RBBP7;POLR2H;PKNOX1;POLR2J;HIST1H2AC;POLR2K;POLR2L;SUZ12;H2AFZ;NCOA3;PCGF2;H3F3A;NCOR1;HOXB4;RARB;HOXB2;HOXD3;AJUBA |
| 351 | Activation of HOX genes during differentiation Homo sapiens R-HSA-5619507 | 1.32E-02 | 5.38E-02 | 1.71E+00 | 7.42E+00 | RARG;PAXIP1;HDAC3;HOXD1;ASH2L;CTCF;YY1;PAGR1;RBBP4;POLR2B;POLR2C;POLR2E;RBBP7;POLR2H;PKNOX1;POLR2J;HIST1H2AC;POLR2K;POLR2L;SUZ12;H2AFZ;NCOA3;PCGF2;H3F3A;NCOR1;HOXB4;RARB;HOXB2;HOXD3;AJUBA |
| 352 | EPHA-mediated growth cone collapse Homo sapiens R-HSA-3928663 | 1.34E-02 | 5.46E-02 | 2.36E+00 | 1.02E+01 | HSP90AA1;YES1;EPHA6;SHB;EFNA4;RHOA;PTK2;MYL12B;VEGFA;MYL6;EFNA3;FYN;MYH10;EPHA2 |
| 353 | RNA Polymerase II HIV Promoter Escape Homo sapiens R-HSA-167162 | 1.39E-02 | 5.55E-02 | 2.12E+00 | 9.06E+00 | GTF2A1;GTF2A2;TAF13;GTF2B;TAF10;TAF11;GTF2F2;GTF2H4;GTF2E2;POLR2B;POLR2C;POLR2E;POLR2H;MNAT1;POLR2J;POLR2K;POLR2L |
| 354 | RNA Polymerase II Promoter Escape Homo sapiens R-HSA-73776 | 1.39E-02 | 5.55E-02 | 2.12E+00 | 9.06E+00 | GTF2A1;GTF2A2;TAF13;GTF2B;TAF10;TAF11;GTF2F2;GTF2H4;GTF2E2;POLR2B;POLR2C;POLR2E;POLR2H;MNAT1;POLR2J;POLR2K;POLR2L |
| 355 | RNA Polymerase II Transcription Initiation And Promoter Clearance Homo sapiens R-HSA-76042 | 1.39E-02 | 5.55E-02 | 2.12E+00 | 9.06E+00 | GTF2A1;GTF2A2;TAF13;GTF2B;TAF10;TAF11;GTF2F2;GTF2H4;GTF2E2;POLR2B;POLR2C;POLR2E;POLR2H;MNAT1;POLR2J;POLR2K;POLR2L |

| | | | | | | |
|---|---|---|---|---|---|---|
| 356 | RNA Polymerase II Transcription Initiation Homo sapiens R-HSA-75953 | 1.39E-02 | 5.55E-02 | 2.12E+00 | 9.06E+00 | GTF2A1;GTF2A2;TAF13;GTF2B;TAF10;TAF11;GTF2F2;GTF2H4;GTF2E2;POLR2B;POLR2C;POLR2E;POLR2H;MNAT1;POLR2J;POLR2K;POLR2L |
| 357 | RNA Polymerase II Transcription Pre-Initiation And Promoter Opening Homo sapiens R-HSA-73779 | 1.39E-02 | 5.55E-02 | 2.12E+00 | 9.06E+00 | GTF2A1;GTF2A2;TAF13;GTF2B;TAF10;TAF11;GTF2F2;GTF2H4;GTF2E2;POLR2B;POLR2C;POLR2E;POLR2H;MNAT1;POLR2J;POLR2K;POLR2L |
| 358 | HIV Transcription Initiation Homo sapiens R-HSA-167161 | 1.39E-02 | 5.55E-02 | 2.12E+00 | 9.06E+00 | GTF2A1;GTF2A2;TAF13;GTF2B;TAF10;TAF11;GTF2F2;GTF2H4;GTF2E2;POLR2B;POLR2C;POLR2E;POLR2H;MNAT1;POLR2J;POLR2K;POLR2L |
| 359 | PLC beta mediated events Homo sapiens R-HSA-112043 | 1.39E-02 | 5.55E-02 | 2.12E+00 | 9.06E+00 | PDE1A;GNAI3;ITPR2;ADCY3;PLA2G4A;GNAI1;ADCY5;GNAI2;GNAO1;CREB1;PLCB4;PRKAR2B;PRKAR1A;PRKAR2A;MAPK1;PLCB1;CALM1 |
| 360 | Pausing and recovery of Tat-mediated HIV elongation Homo sapiens R-HSA-167238 | 1.43E-02 | 5.67E-02 | 2.43E+00 | 1.03E+01 | SUPT16H;SUPT4H1;GTF2F2;CDK9;POLR2B;POLR2C;POLR2E;TCEA1;NELFCD;POLR2H;POLR2J;POLR2K;POLR2L |
| 361 | Tat-mediated HIV elongation arrest and recovery Homo sapiens R-HSA-167243 | 1.43E-02 | 5.67E-02 | 2.43E+00 | 1.03E+01 | SUPT16H;SUPT4H1;GTF2F2;CDK9;POLR2B;POLR2C;POLR2E;TCEA1;NELFCD;POLR2H;POLR2J;POLR2K;POLR2L |
| 362 | Centrosome maturation Homo sapiens R-HSA-380287 | 1.50E-02 | 5.86E-02 | 1.75E+00 | 7.35E+00 | YWHAE;CEP57;DCTN2;TUBGCP2;CETN2;SSNA1;DCTN3;ACTR1A;CCNB1;HAUS8;HAUS7;PRKAR2B;YWHAG;OFD1;DYNC1H1;HSP90AA1;CDK11A;TUBB;TUBG1;TUBB4B;CDK11B;TUBA4A;HAUS1;AKAP9;TUBGCP4;MAPRE1;PAFAH1B1 |
| 363 | Recruitment of mitotic centrosome proteins and complexes Homo sapiens R-HSA-380270 | 1.50E-02 | 5.86E-02 | 1.75E+00 | 7.35E+00 | YWHAE;CEP57;DCTN2;TUBGCP2;CETN2;SSNA1;DCTN3;ACTR1A;CCNB1;HAUS8;HAUS7;PRKAR2B;YWHAG;OFD1;DYNC1H1;HSP90AA1;CDK11A;TUBB;TUBG1;TUBB4B;CDK11B;TUBA4A;HAUS1;AKAP9;TUBGCP4;MAPRE1;PAFAH1B1 |

| | | | | | | |
|---|---|---|---|---|---|---|
| 364 | The role of Nef in HIV-1 replication and disease pathogenesis Homo sapiens R-HSA-164952 | 1.51E-02 | 5.86E-02 | 2.52E+00 | 1.06E+01 | ARF1;AP1G1;AP2S1;AP1S2;ATP6V1H;FYN;RAC1;AP2B1;AP2A2;PAK2;AP1M2;AP2M1 |
| 365 | RNA Polymerase III Transcription Initiation From Type 3 Promoter Homo sapiens R-HSA-76071 | 1.51E-02 | 5.86E-02 | 2.52E+00 | 1.06E+01 | ZNF143;CRCP;SNAPC1;POLR3GL;SNAPC3;POLR3D;POLR2E;POLR1D;POLR2H;POLR3K;POLR2K;POLR2L |
| 366 | WNT5A-dependent internalization of FZD2, FZD5 and ROR2 Homo sapiens R-HSA-5140745 | 1.51E-02 | 5.86E-02 | 3.93E+00 | 1.65E+01 | CLTC;CLTB;AP2S1;CLTA;AP2B1;AP2A2;AP2M1 |
| 367 | Advanced glycosylation endproduct receptor signaling Homo sapiens R-HSA-879415 | 1.51E-02 | 5.86E-02 | 3.93E+00 | 1.65E+01 | APP;LGALS3;CAPZA1;CAPZA2;MAPK1;HMGB1;DDOST |
| 368 | Purine salvage Homo sapiens R-HSA-74217 | 1.51E-02 | 5.86E-02 | 3.93E+00 | 1.65E+01 | DGUOK;GMPR2;ADK;GMPR;AMPD2;HPRT1;APRT |
| 369 | RNA Polymerase III Abortive And Retractive Initiation Homo sapiens R-HSA-749476 | 1.51E-02 | 5.86E-02 | 2.16E+00 | 9.03E+00 | ZNF143;CRCP;SSB;GTF3C6;POLR3GL;GTF3A;SNAPC1;SNAPC3;POLR3D;NFIC;POLR2E;POLR1D;POLR2H;POLR3K;POLR2K;POLR2L |
| 370 | RNA Polymerase III Transcription Homo sapiens R-HSA-74158 | 1.51E-02 | 5.86E-02 | 2.16E+00 | 9.03E+00 | ZNF143;CRCP;SSB;GTF3C6;POLR3GL;GTF3A;SNAPC1;SNAPC3;POLR3D;NFIC;POLR2E;POLR1D;POLR2H;POLR3K;POLR2K;POLR2L |

| | | | | | | |
|---|---|---|---|---|---|---|
| 371 | rRNA modification in the nucleus Homo sapiens R-HSA-6790901 | 1.52E-02 | 5.87E-02 | 1.91E+00 | 8.00E+00 | RPS9;NOP14;DDX49;IMP3;DIMT1;KRR1;DDX52;WDR46;UTP18;WDR43;RRP9;FBL;TRMT112;EMG1;DKC1;BMS1;NHP2;RRP36;RPS2;DCAF13;NOP10 |
| 372 | APC-Cdc20 mediated degradation of Nek2A Homo sapiens R-HSA-179409 | 1.58E-02 | 6.09E-02 | 2.64E+00 | 1.10E+01 | ANAPC15;ANAPC16;CDC26;UBC;CDC27;UBE2E1;UBE2D1;ANAPC5;BUB3;RPS27A;ANAPC11 |
| 373 | TP53 Regulates Transcription of Cell Cycle Genes Homo sapiens R-HSA-6791312 | 1.62E-02 | 6.21E-02 | 2.02E+00 | 8.33E+00 | NPM1;PCNA;CDKN1B;CNOT6L;GADD45A;PRMT1;CNOT4;CCNB1;CNOT7;TFDP2;PCBP4;CNOT11;CNOT2;CARM1;CNOT3;CDK2;BAX;E2F4 |
| 374 | Spry regulation of FGF signaling Homo sapiens R-HSA-1295596 | 1.62E-02 | 6.21E-02 | 3.37E+00 | 1.39E+01 | PPP2CA;PPP2CB;UBC;MAPK1;PTPN11;GRB2;CBL;RPS27A |
| 375 | Signaling by Rho GTPases Homo sapiens R-HSA-194315 | 1.63E-02 | 6.21E-02 | 1.30E+00 | 5.36E+00 | CYFIP2;NCKAP1;RTKN;FAM13B;MYLK;ACTG1;PPP1CB;PPP1CC;SYDE1;OPHN1;KIF5B;HIST1H2AC;RALBP1;CYBA;TIAM1;MYL6;CLIP1;PFN1;PFN2;KLC1;PMF1;MYL12B;SKA2;ARHGAP12;RHOT2;RHOT1;RHPN1;DVL3;RHPN2;SRGAP3;MYH10;STARD13;H3F3A;ARHGAP29;ARPC5;PTK2;ARPC2;ARPC3;NOXA1;GRB2;NF2;CALM1;YWHAE;ITGB1;CDKN1B;GDI1;YWHAB;ARPC1B;RASGRF2;ARPC1A;GDI2;BRK1;WASL;ARHGAP4;YWHAQ;PPP2R5E;MCF2L;RAC1;YWHAG;YWHAH;VAV3;ACTR3;NCOA2;ACTR2;NUDC;ARHGEF12;H2AFZ;PDPK1;ARHGEF16;RHOG;ARHGEF18;PPP2R5A;RHOF;RHOC;RHOD;YWHAZ;RHOA;CTTN;KIF2A;ARHGEF3;TAX1BP3;ARHGEF7;MAPRE1;RHOQ;PAFAH1B1;CENPC;PPP2CA;CDC42;PPP2CB;PAK1;MAPK1;BUB3;PAK2;WASF2;LIMK2;FMNL3;FMNL1;ABI2;TAOK1;TRIP10;GOPC;PKN1 |
| 376 | CTLA4 inhibitory signaling Homo sapiens R-HSA-389513 | 1.64E-02 | 6.22E-02 | 2.80E+00 | 1.15E+01 | PPP2CA;PPP2CB;YES1;PDPK1;PPP2R5E;AKT2;PPP2R5A;AKT1;PTPN11;FYN |
| 377 | Phosphorylation of the APC/C Homo sapiens R-HSA-176412 | 1.66E-02 | 6.28E-02 | 3.03E+00 | 1.24E+01 | CCNB1;ANAPC15;ANAPC16;CDC26;CDC27;UBE2E1;UBE2D1;ANAPC5;ANAPC11 |

| | | | | | | |
|---|---|---|---|---|---|---|
| 378 | G-protein mediated events Homo sapiens R-HSA-112040 | 1.77E-02 | 6.71E-02 | 2.04E+00 | 8.25E+00 | PDE1A;GNAI3;ITPR2;ADCY3;PLA2G4A;GNAI1;ADCY5;GNAI2;GNAO1;CREB1;PLCB4;PRKAR2B;PRKAR1A;PRKAR2A;MAPK1;PLCB1;CALM1 |
| 379 | Regulation of HSF1-mediated heat shock response Homo sapiens R-HSA-3371453 | 1.78E-02 | 6.71E-02 | 1.72E+00 | 6.92E+00 | YWHAE;GSK3B;RPS19BP1;MRPL18;HSPA13;BAG4;HSPH1;BAG5;DNAJB6;BAG3;TPR;BAG1;MAPK1;HSPA9;HSPA8;HSPA5;HSBP1;HSPA4;ST13;HSPA2;SIRT1;NUP93;DNAJC7;NUP50;RPA3;FKBP4;HSPA1A |
| 380 | Cleavage of the damaged pyrimidine Homo sapiens R-HSA-110329 | 1.89E-02 | 7.06E-02 | 5.61E+00 | 2.23E+01 | MBD4;NTHL1;TDG;NEIL1;UNG |
| 381 | Recognition and association of DNA glycosylase with site containing an affected pyrimidine Homo sapiens R-HSA-110328 | 1.89E-02 | 7.06E-02 | 5.61E+00 | 2.23E+01 | MBD4;NTHL1;TDG;NEIL1;UNG |
| 382 | Depyrimidination Homo sapiens R-HSA-73928 | 1.89E-02 | 7.06E-02 | 5.61E+00 | 2.23E+01 | MBD4;NTHL1;TDG;NEIL1;UNG |
| 383 | HIV elongation arrest and recovery Homo sapiens R-HSA-167287 | 1.91E-02 | 7.11E-02 | 2.30E+00 | 9.11E+00 | SUPT16H;SUPT4H1;GTF2F2;CDK9;POLR2B;POLR2C;POLR2E;TCEA1;NELFCD;POLR2H;POLR2J;POLR2K;POLR2L |
| 384 | Pausing and recovery of HIV elongation Homo sapiens R-HSA-167290 | 1.91E-02 | 7.11E-02 | 2.30E+00 | 9.11E+00 | SUPT16H;SUPT4H1;GTF2F2;CDK9;POLR2B;POLR2C;POLR2E;TCEA1;NELFCD;POLR2H;POLR2J;POLR2K;POLR2L |
| 385 | Elongation arrest and recovery Homo sapiens R-HSA-112387 | 1.91E-02 | 7.11E-02 | 2.30E+00 | 9.11E+00 | SUPT16H;SUPT4H1;GTF2F2;CDK9;POLR2B;POLR2C;POLR2E;TCEA1;NELFCD;POLR2H;POLR2J;POLR2K;POLR2L |

| | | | | | | |
|---|---|---|---|---|---|---|
| 386 | tRNA Aminoacylation Homo sapiens R-HSA-379724 | 1.94E-02 | 7.18E-02 | 2.07E+00 | 8.17E+00 | CARS;SARS;EPRS;YARS2;NARS;AIMP1;QARS;LARS;PPA1;MARS;IARS2;GARS;HARS;FARSA;EEF1E1;CARS2 |
| 387 | Activation of BH3-only proteins Homo sapiens R-HSA-114452 | 2.05E-02 | 7.59E-02 | 2.38E+00 | 9.23E+00 | YWHAE;PPP1R13B;YWHAQ;YWHAB;BAD;AKT2;AKT1;BID;YWHAZ;YWHAG;BBC3;YWHAH |
| 388 | Opioid Signalling Homo sapiens R-HSA-111885 | 2.10E-02 | 7.73E-02 | 1.68E+00 | 6.51E+00 | PDE1A;GNAI3;ITPR2;ADCY3;GNAI1;ADCY5;GNAI2;PPP2CA;PPP2CB;PPP3CB;GNG10;GNG5;PRKAR2B;PRKAR2A;PDE4A;MAPK1;PDE4D;PLA2G4A;PPP1CA;GNAO1;PLCB4;CREB1;CDK5;PRKAR1A;GNB5;PLCB1;CALM1 |
| 389 | Attenuation phase Homo sapiens R-HSA-3371568 | 2.19E-02 | 8.03E-02 | 2.47E+00 | 9.43E+00 | HSPA8;HSP90AA1;HSP90AB1;HSPH1;DNAJB6;HSBP1;MRPL18;PTGES3;HSPA2;FKBP4;HSPA1A |
| 390 | Downregulation of TGF-beta receptor signaling Homo sapiens R-HSA-2173788 | 2.19E-02 | 8.03E-02 | 2.47E+00 | 9.43E+00 | PPP1CB;PPP1CC;TGFB1;SMURF2;BAMBI;UBC;STRAP;STUB1;RPS27A;UCHL5;PPP1CA |
| 391 | Transcription of the HIV genome Homo sapiens R-HSA-167172 | 2.23E-02 | 8.15E-02 | 1.76E+00 | 6.70E+00 | GTF2A1;SUPT16H;GTF2A2;NCBP1;TAF13;GTF2B;TAF10;TAF11;SUPT4H1;GTF2F2;GTF2H4;GTF2E2;CDK9;POLR2B;POLR2C;POLR2E;TCEA1;NELFCD;POLR2H;MNAT1;POLR2J;POLR2K;POLR2L |
| 392 | SLBP Dependent Processing of Replication-Dependent Histone Pre-mRNAs Homo sapiens R-HSA-77588 | 2.27E-02 | 8.22E-02 | 4.04E+00 | 1.53E+01 | LSM10;SLBP;NCBP1;SNRPG;SNRPD3;SNRPB |
| 393 | Membrane binding and targetting of GAG proteins Homo sapiens R-HSA-174490 | 2.27E-02 | 8.22E-02 | 4.04E+00 | 1.53E+01 | TSG101;UBC;VPS37D;VPS37B;RPS27A;VPS28 |

| | | | | | | |
|---|---|---|---|---|---|---|
| 394 | Synthesis And Processing Of GAG, GAGPOL Polyproteins Homo sapiens R-HSA-174495 | 2.27E-02 | 8.22E-02 | 4.04E+00 | 1.53E+01 | TSG101;UBC;VPS37D;VPS37B;RPS27A;VPS28 |
| 395 | RHO GTPases activate KTN1 Homo sapiens R-HSA-5625970 | 2.27E-02 | 8.22E-02 | 4.04E+00 | 1.53E+01 | CDC42;KIF5B;RHOG;RAC1;KLC1;RHOA |
| 396 | SeMet incorporation into proteins Homo sapiens R-HSA-2408517 | 2.27E-02 | 8.22E-02 | 4.04E+00 | 1.53E+01 | AIMP1;QARS;LARS;MARS;EPRS;EEF1E1 |
| 397 | RNA Polymerase III Transcription Initiation Homo sapiens R-HSA-76046 | 2.31E-02 | 8.31E-02 | 2.14E+00 | 8.08E+00 | ZNF143;CRCP;GTF3C6;POLR3GL;GTF3A;SNAPC1;SNAPC3;POLR3D;POLR2E;POLR1D;POLR2H;POLR3K;POLR2K;POLR2L |
| 398 | MicroRNA (miRNA) biogenesis Homo sapiens R-HSA-203927 | 2.31E-02 | 8.31E-02 | 2.59E+00 | 9.75E+00 | POLR2B;PRKRA;POLR2C;AGO2;POLR2E;POLR2H;POLR2J;POLR2K;RAN;POLR2L |
| 399 | Signaling by Interleukins Homo sapiens R-HSA-449147 | 2.35E-02 | 8.42E-02 | 1.27E+00 | 4.76E+00 | PEBP1;WDR83;FGF2;CRKL;PSMD8;PSMD9;PSMD6;PSMD7;PSMD4;PSMD2;PSMD3;PSMD1;SKP1;MAP2K2;FBXW11;IL1R2;RBX1;BTC;TXLNA;AKAP9;PSME1;PSME2;UBE2V1;TLN1;SQSTM1;PSMD10;SHC2;SPTBN5;PSMD12;PSMD11;PSMD14;PSMD13;CUL3;PDGFB;CUL1;PIK3R3;RASAL1;PHB;RASGRP3;RAP1B;RAP1A;IRAK1;UBC;PSMF1;YES1;NFKB1;PTK2;PTPRA;FGF18;GRB2;CALM1;YWHAB;RASGRF2;ARRB1;PPP2R5E;PEA15;JAK2;SPTAN1;JAK1;DUSP1;PPP2R5A;IRAK4;DUSP8;YWHAZ;IL17RB;PSMA5;PSMA6;PSMA3;PSMA4;PSMA1;PSMA2;RASA1;RAPGEF1;PELI1;HBEGF;CBL;PSMA7;PSMB6;PPP2CA;PSMB7;PPP2CB;PSMB5;PSMB2;PSMB3;PSMB1;MAPK1;STX4;CSK;FYN;RPS27A;CAMK2G;IL10RB;STAT3;DAB2IP;PTPN11;PSMB8;PSMC5;PSMC6;PSMC3;PSMC4;TNIP2;PSMC1;PSMC2;PTPN6;KRAS;LAMTOR3;IL18R1 |

| | | | | | | |
|---|---|---|---|---|---|---|
| 400 | Inhibition of the proteolytic activity of APC/C required for the onset of anaphase by mitotic spindle checkpoint components Homo sapiens R-HSA-141405 | 2.41E-02 | 8.57E-02 | 2.75E+00 | 1.03E+01 | ANAPC15;ANAPC16;CDC26;CDC27;UBE2E1;UBE2D1;ANAPC5;BUB3;ANAPC11 |
| 401 | Inactivation of APC/C via direct inhibition of the APC/C complex Homo sapiens R-HSA-141430 | 2.41E-02 | 8.57E-02 | 2.75E+00 | 1.03E+01 | ANAPC15;ANAPC16;CDC26;CDC27;UBE2E1;UBE2D1;ANAPC5;BUB3;ANAPC11 |
| 402 | Early Phase of HIV Life Cycle Homo sapiens R-HSA-162594 | 2.44E-02 | 8.57E-02 | 3.36E+00 | 1.25E+01 | XRCC6;XRCC5;HMGA1;PSIP1;LIG4;PPIA;KPNA1 |
| 403 | Regulation of IFNG signaling Homo sapiens R-HSA-877312 | 2.44E-02 | 8.57E-02 | 3.36E+00 | 1.25E+01 | SOCS1;SUMO1;IFNGR2;PTPN11;PTPN6;JAK2;JAK1 |
| 404 | Golgi Cisternae Pericentriolar Stack Reorganization Homo sapiens R-HSA-162658 | 2.44E-02 | 8.57E-02 | 3.36E+00 | 1.25E+01 | RAB2A;GOLGA2;CCNB1;GORASP2;RAB1B;USO1;MAPK1 |
| 405 | DCC mediated attractive signaling Homo sapiens R-HSA-418885 | 2.44E-02 | 8.57E-02 | 3.36E+00 | 1.25E+01 | CDC42;FYN;WASL;RAC1;NTN1;DOCK1;PTK2 |
| 406 | HIV Transcription Elongation Homo sapiens R-HSA-167169 | 2.44E-02 | 8.57E-02 | 2.00E+00 | 7.41E+00 | SUPT16H;NCBP1;SUPT4H1;GTF2F2;GTF2H4;CDK9;POLR2B;POLR2C;POLR2E;TCEA1;NELFCD;POLR2H;MNAT1;POLR2J;POLR2K;POLR2L |
| 407 | Tat-mediated elongation of the HIV-1 transcript Homo sapiens R-HSA-167246 | 2.44E-02 | 8.57E-02 | 2.00E+00 | 7.41E+00 | SUPT16H;NCBP1;SUPT4H1;GTF2F2;GTF2H4;CDK9;POLR2B;POLR2C;POLR2E;TCEA1;NELFCD;POLR2H;MNAT1;POLR2J;POLR2K;POLR2L |

| | | | | | | |
|---|---|---|---|---|---|---|
| 408 | Formation of HIV-1 elongation complex containing HIV-1 Tat Homo sapiens R-HSA-167200 | 2.44E-02 | 8.57E-02 | 2.00E+00 | 7.41E+00 | SUPT16H;NCBP1;SUPT4H1;GTF2F2;GTF2H4;CDK9;POLR2B;POLR2C;POLR2E;TCEA1;NELFCD;POLR2H;MNAT1;POLR2J;POLR2K;POLR2L |
| 409 | Tristetraprolin (TTP, ZFP36) binds and destabilizes mRNA Homo sapiens R-HSA-450513 | 2.46E-02 | 8.57E-02 | 2.99E+00 | 1.11E+01 | EXOSC7;EXOSC4;XRN1;YWHAB;TNPO1;EXOSC3;DCP2;EXOSC1 |
| 410 | Butyrate Response Factor 1 (BRF1) binds and destabilizes mRNA Homo sapiens R-HSA-450385 | 2.46E-02 | 8.57E-02 | 2.99E+00 | 1.11E+01 | EXOSC7;EXOSC4;XRN1;YWHAB;AKT1;EXOSC3;DCP2;EXOSC1 |
| 411 | Platelet sensitization by LDL Homo sapiens R-HSA-432142 | 2.46E-02 | 8.57E-02 | 2.99E+00 | 1.11E+01 | PPP2CA;PPP2CB;PPP2R5E;PLA2G4A;PPP2R5A;PTPN11;PTPN6;LRP8 |
| 412 | Formation of the Early Elongation Complex Homo sapiens R-HSA-113418 | 2.51E-02 | 8.70E-02 | 2.19E+00 | 8.06E+00 | NCBP1;SUPT4H1;GTF2F2;GTF2H4;POLR2B;POLR2C;POLR2E;NELFCD;POLR2H;MNAT1;POLR2J;POLR2K;POLR2L |
| 413 | Formation of the HIV-1 Early Elongation Complex Homo sapiens R-HSA-167158 | 2.51E-02 | 8.70E-02 | 2.19E+00 | 8.06E+00 | NCBP1;SUPT4H1;GTF2F2;GTF2H4;POLR2B;POLR2C;POLR2E;NELFCD;POLR2H;MNAT1;POLR2J;POLR2K;POLR2L |
| 414 | Activated TLR4 signalling Homo sapiens R-HSA-166054 | 2.63E-02 | 9.09E-02 | 1.53E+00 | 5.57E+00 | ATF1;APP;DHX9;UBE2D2;UBE2D3;CUL1;UBE2D1;HMGB1;ECSIT;TANK;PPP2CA;PPP2CB;MAPK9;TBK1;SOCS1;IRAK1;UBC;MAPK1;FADD;RPS27A;SKP1;MAP2K3;DUSP3;MAP3K1;FBXW11;PTPN11;IRAK4;NFKB1;SIGIRR;NFKBIA;CREB1;IRF3;TNIP2;PELI1;UBE2V1 |

| | | | | | | |
|---|---|---|---|---|---|---|
| 415 | Transcriptional activation of mitochondrial biogenesis Homo sapiens R-HSA-2151201 | 2.68E-02 | 9.26E-02 | 2.02E+00 | 7.31E+00 | ESRRA;NCOA2;SMARCD3;HDAC3;ALAS1;NR1D1;TFB1M;POLRMT;CREB1;NCOR1;TGS1;CARM1;TFAM;CYCS;SSBP1 |
| 416 | PI3K Cascade Homo sapiens R-HSA-109704 | 2.72E-02 | 9.35E-02 | 1.65E+00 | 5.96E+00 | PRKAA1;YWHAB;CAB39L;PRKAG1;PRKAG2;FGF2;AKT2;MLST8;AKT1;EIF4E;EIF4B;PDPK1;STRADB;PTPN11;MTOR;RRAGA;RRAGB;RHEB;AKT1S1;FGF18;GRB2;LAMTOR1;LAMTOR4;LAMTOR3;EIF4G1;LAMTOR5 |
| 417 | Cell death signalling via NRAGE, NRIF and NADE Homo sapiens R-HSA-204998 | 2.72E-02 | 9.35E-02 | 1.77E+00 | 6.37E+00 | YWHAE;PSENEN;VAV3;ARHGEF12;MAGED1;BAD;RASGRF2;ARHGEF16;ARHGEF18;AATF;ARHGAP4;APH1A;TIAM1;APH1B;MCF2L;UBC;ARHGEF3;RAC1;ARHGEF7;RPS27A;SQSTM1 |
| 418 | NOTCH1 Intracellular Domain Regulates Transcription Homo sapiens R-HSA-2122947 | 2.76E-02 | 9.41E-02 | 1.91E+00 | 6.85E+00 | HDAC2;HDAC3;HDAC10;HDAC1;HDAC11;CUL1;CCNC;RBPJ;HIF1A;RBX1;NCOR2;KAT2B;NCOR1;SNW1;UBC;RPS27A;SKP1 |
| 419 | SUMO is proteolytically processed Homo sapiens R-HSA-3065679 | 2.77E-02 | 9.41E-02 | 6.73E+00 | 2.41E+01 | SENP5;SUMO1;SUMO3;SUMO2 |
| 420 | Acyl chain remodeling of CL Homo sapiens R-HSA-1482798 | 2.77E-02 | 9.41E-02 | 6.73E+00 | 2.41E+01 | HADHB;HADHA;PLA2G4A;TAZ |
| 421 | Methionine salvage pathway Homo sapiens R-HSA-1237112 | 2.77E-02 | 9.41E-02 | 6.73E+00 | 2.41E+01 | GOT1;ADI1;APIP;ENOPH1 |
| 422 | ISG15 antiviral mechanism Homo sapiens R-HSA-1169408 | 2.84E-02 | 9.57E-02 | 1.68E+00 | 6.00E+00 | EIF4A2;EIF4A1;EIF4E3;EIF4A3;UBE2E1;EIF2AK2;UBE2L6;NUP93;IRF3;NUP50;NEDD4;TPR;UBC;PIN1;KPNA4;KPNA2;RPS27A;KPNA3;EIF4E;EIF4G3;EIF4G2;KPNA1;JAK1;EIF4G1 |

| | | | | | | |
|---|---|---|---|---|---|---|
| 423 | Antiviral mechanism by IFN-stimulated genes Homo sapiens R-HSA-1169410 | 2.84E-02 | 9.57E-02 | 1.68E+00 | 6.00E+00 | EIF4A2;EIF4A1;EIF4E3;EIF4A3;UBE2E1;EIF2AK2;UBE2L6;NUP93;IRF3;NUP50;NEDD4;TPR;UBC;PIN1;KPNA4;KPNA2;RPS27A;KPNA3;EIF4E;EIF4G3;EIF4G2;KPNA1;JAK1;EIF4G1 |
| 424 | Signaling by NOTCH1 Homo sapiens R-HSA-1980143 | 2.84E-02 | 9.57E-02 | 1.68E+00 | 6.00E+00 | PSENEN;JAG2;HDAC2;HDAC3;HDAC10;HDAC1;HDAC11;CUL1;MIB2;CCNC;ARRB1;RBPJ;HIF1A;RBX1;NCOR2;APH1A;KAT2B;ITCH;NCOR1;APH1B;SNW1;UBC;RPS27A;SKP1 |
| 425 | Sema4D in semaphorin signaling Homo sapiens R-HSA-400685 | 2.94E-02 | 9.91E-02 | 2.31E+00 | 8.16E+00 | CDC42;MYL6;ARHGEF12;SEMA4D;LIMK2;RHOG;RAC1;RHOC;MYH10;RHOA;MYL12B |
| 426 | Formation of HIV elongation complex in the absence of HIV Tat Homo sapiens R-HSA-167152 | 3.04E-02 | 1.02E-01 | 1.92E+00 | 6.72E+00 | SUPT16H;NCBP1;SUPT4H1;GTF2F2;GTF2H4;CDK9;POLR2B;POLR2C;POLR2E;TCEA1;NELFCD;POLR2H;MNAT1;POLR2J;POLR2K;POLR2L |
| 427 | RNA Polymerase II Transcription Elongation Homo sapiens R-HSA-75955 | 3.04E-02 | 1.02E-01 | 1.92E+00 | 6.72E+00 | SUPT16H;NCBP1;SUPT4H1;GTF2F2;GTF2H4;CDK9;POLR2B;POLR2C;POLR2E;TCEA1;NELFCD;POLR2H;MNAT1;POLR2J;POLR2K;POLR2L |
| 428 | Formation of RNA Pol II elongation complex Homo sapiens R-HSA-112382 | 3.04E-02 | 1.02E-01 | 1.92E+00 | 6.72E+00 | SUPT16H;NCBP1;SUPT4H1;GTF2F2;GTF2H4;CDK9;POLR2B;POLR2C;POLR2E;TCEA1;NELFCD;POLR2H;MNAT1;POLR2J;POLR2K;POLR2L |
| 429 | Transcriptional activity of SMAD2/SMAD3:SMAD4 heterotrimer Homo sapiens R-HSA-2173793 | 3.04E-02 | 1.02E-01 | 1.92E+00 | 6.72E+00 | SMAD4;SMURF2;PARP1;HDAC1;UBE2D3;UBE2D1;CCNC;CDK9;NCOR2;NCOR1;SNW1;TFDP2;UBC;E2F4;RPS27A;SKIL |
| 430 | Post-Elongation Processing of Intronless pre-mRNA Homo sapiens R-HSA-112297 | 3.17E-02 | 1.05E-01 | 2.40E+00 | 8.30E+00 | LSM10;SLBP;NCBP1;FIP1L1;CLP1;CPSF3;CSTF1;SNRPG;SNRPD3;SNRPB |

| | | | | | | |
|---|---|---|---|---|---|---|
| 431 | Processing of Capped Intronless Pre-mRNA Homo sapiens R-HSA-75067 | 3.17E-02 | 1.05E-01 | 2.40E+00 | 8.30E+00 | LSM10;SLBP;NCBP1;FIP1L1;CLP1;CPSF3;CSTF1;SNRPG;SNRPD3;SNRPB |
| 432 | ROS, RNS production in response to bacteria Homo sapiens R-HSA-1222556 | 3.23E-02 | 1.07E-01 | 2.08E+00 | 7.15E+00 | ATP6V1A;ATP6V1G1;ATP6V0B;ATP6V0E1;CYBA;ATP6V1H;ATP6V0E2;ATP6V1E1;ATP6V0D1;ATP6V1D;ATP6V0C;ATP6V1C1;ATP6V1F |
| 433 | Purine metabolism Homo sapiens R-HSA-73847 | 3.23E-02 | 1.07E-01 | 2.08E+00 | 7.15E+00 | ADSL;GMPR2;ADK;GMPS;GMPR;AMPD2;PAICS;APRT;DGUOK;ATIC;CAT;HPRT1;LHPP |
| 434 | Regulation of cholesterol biosynthesis by SREBP (SREBF) Homo sapiens R-HSA-1655829 | 3.34E-02 | 1.10E-01 | 1.78E+00 | 6.04E+00 | NCOA2;IDI1;FDPS;MVK;SMARCD3;SEC24A;SAR1B;INSIG2;INSIG1;CYP51A1;ELOVL6;SREBF2;ACACA;SQLE;TGS1;CARM1;SC5D;RAN;FDFT1 |
| 435 | Nef-mediates down modulation of cell surface receptors by recruiting them to clathrin adapters Homo sapiens R-HSA-164938 | 3.38E-02 | 1.11E-01 | 2.52E+00 | 8.55E+00 | ARF1;AP1G1;AP2S1;AP1S2;ATP6V1H;AP2B1;AP2A2;AP1M2;AP2M1 |
| 436 | Mitotic Spindle Checkpoint Homo sapiens R-HSA-69618 | 3.38E-02 | 1.11E-01 | 2.52E+00 | 8.55E+00 | ANAPC15;ANAPC16;CDC26;CDC27;UBE2E1;UBE2D1;ANAPC5;BUB3;ANAPC11 |
| 437 | Displacement of DNA glycosylase by APEX1 Homo sapiens R-HSA-110357 | 3.45E-02 | 1.12E-01 | 4.20E+00 | 1.42E+01 | MBD4;MPG;NTHL1;TDG;UNG |
| 438 | Cohesin Loading onto Chromatin Homo sapiens R-HSA-2470946 | 3.45E-02 | 1.12E-01 | 4.20E+00 | 1.42E+01 | STAG2;RAD21;PDS5B;SMC1A;SMC3 |

| | | | | | | |
|---|---|---|---|---|---|---|
| 439 | Formation of annular gap junctions Homo sapiens R-HSA-196025 | 3.45E-02 | 1.12E-01 | 4.20E+00 | 1.42E+01 | CLTC;CLTB;CLTA;AP2M1;DNM2 |
| 440 | MAPK1 (ERK2) activation Homo sapiens R-HSA-112411 | 3.45E-02 | 1.12E-01 | 4.20E+00 | 1.42E+01 | MAP2K2;MAPK1;PTPN11;JAK2;JAK1 |
| 441 | PTK6 Regulates RTKs and Their Effectors AKT1 and DOK1 Homo sapiens R-HSA-8849469 | 3.45E-02 | 1.12E-01 | 4.20E+00 | 1.42E+01 | DOK1;UBC;AKT1;CBL;RPS27A |
| 442 | Cytosolic sensors of pathogen-associated DNA Homo sapiens R-HSA-1834949 | 3.48E-02 | 1.13E-01 | 1.68E+00 | 5.66E+00 | APP;XRCC6;CRCP;DHX9;PRKDC;POLR3GL;XRCC5;HMGB1;NFKB1;NFKBIA;TBK1;IRF3;POLR3D;UBC;POLR1D;POLR2E;LRRFIP1;POLR2H;RPS27A;POLR3K;POLR2K;POLR2L |
| 443 | Trafficking of AMPA receptors Homo sapiens R-HSA-399719 | 3.54E-02 | 1.14E-01 | 2.13E+00 | 7.10E+00 | NSF;DLG1;MYO6;TSPAN7;AP2S1;MDM2;PICK1;AKAP5;AP2B1;AP2A2;CAMK2G;AP2M1 |
| 444 | Glutamate Binding, Activation of AMPA Receptors and Synaptic Plasticity Homo sapiens R-HSA-399721 | 3.54E-02 | 1.14E-01 | 2.13E+00 | 7.10E+00 | NSF;DLG1;MYO6;TSPAN7;AP2S1;MDM2;PICK1;AKAP5;AP2B1;AP2A2;CAMK2G;AP2M1 |
| 445 | TP53 Regulates Transcription of Genes Involved in G2 Cell Cycle Arrest Homo sapiens R-HSA-6804114 | 3.56E-02 | 1.14E-01 | 2.69E+00 | 8.98E+00 | CCNB1;PCNA;TFDP2;GADD45A;PRMT1;CARM1;BAX;E2F4 |
| 446 | Pre-NOTCH Processing in Golgi Homo sapiens R-HSA-1912420 | 3.56E-02 | 1.14E-01 | 2.69E+00 | 8.98E+00 | SEL1L;ATP2A3;ST3GAL4;TMED2;FURIN;ST3GAL6;RFNG;RAB6A |

| | | | | | | |
|---|---|---|---|---|---|---|
| 447 | Toll Like Receptor 4 (TLR4) Cascade Homo sapiens R-HSA-166016 | 3.57E-02 | 1.14E-01 | 1.47E+00 | 4.89E+00 | ATF1;APP;DHX9;UBE2D2;UBE2D3;ITGB2;CUL1;UBE2D1;HMGB1;ECSIT;TANK;PPP2CA;PPP2CB;MAPK9;TBK1;SOCS1;IRAK1;UBC;MAPK1;FADD;RPS27A;SKP1;MAP2K3;DUSP3;MAP3K1;FBXW11;PTPN11;IRAK4;NFKB1;DNM2;SIGIRR;NFKBIA;CREB1;IRF3;TNIP2;PELI1;UBE2V1 |
| 448 | WNT5A-dependent internalization of FZD4 Homo sapiens R-HSA-5099900 | 3.68E-02 | 1.17E-01 | 2.94E+00 | 9.72E+00 | CLTC;CLTB;AP2S1;CLTA;AP2B1;AP2A2;AP2M1 |
| 449 | Calnexin/calreticulin cycle Homo sapiens R-HSA-901042 | 3.68E-02 | 1.17E-01 | 2.94E+00 | 9.72E+00 | PDIA3;EDEM3;OS9;EDEM2;SEL1L;DERL2;CALR |
| 450 | mRNA decay by 5' to 3' exoribonuclease Homo sapiens R-HSA-430039 | 3.68E-02 | 1.17E-01 | 2.94E+00 | 9.72E+00 | LSM7;XRN1;LSM1;LSM4;PATL1;DCP2;LSM2 |
| 451 | mRNA decay by 3' to 5' exoribonuclease Homo sapiens R-HSA-429958 | 3.68E-02 | 1.17E-01 | 3.36E+00 | 1.11E+01 | EXOSC7;NT5C3B;EXOSC4;EXOSC3;DCPS;EXOSC1 |
| 452 | CD28 dependent Vav1 pathway Homo sapiens R-HSA-389359 | 3.68E-02 | 1.17E-01 | 3.36E+00 | 1.11E+01 | CDC42;PAK1;FYN;GRB2;RAC1;PAK2 |
| 453 | Signaling by ERBB2 Homo sapiens R-HSA-1227986 | 3.75E-02 | 1.18E-01 | 1.86E+00 | 6.10E+00 | GRB7;USP8;HSP90AA1;YES1;RHOA;BTC;AKT2;CDC37;UBC;AKT1;FYN;KRAS;GRB2;STUB1;RPS27A;HBEGF |
| 454 | Epigenetic regulation of gene expression Homo sapiens R-HSA-212165 | 3.83E-02 | 1.21E-01 | 1.47E+00 | 4.81E+00 | GSK3B;DNMT1;HDAC2;HDAC1;PHF1;DDX21;ARID4B;CHD4;ZNRD1;RBBP4;TDG;SAP30L;TWISTNB;POLR2E;SAP18;SUDS3;RBBP7;POLR2H;JARID2;MTA3;HIST1H2AC;POLR2K;POLR2L;SUZ12;CBX3;H2AFZ;H3F3A;MBD2;SMARCA5;DEK;SIRT1;GTF2H4;KAT2B;POLR1D;MNAT1 |
| 455 | Circadian Clock Homo sapiens R-HSA-400253 | 3.86E-02 | 1.21E-01 | 1.68E+00 | 5.48E+00 | NCOA2;SMARCD3;HDAC3;BHLHE41;CUL1;RORA;NR1D1;SIRT1;HIF1A;ARNTL;PPP1CA;PPP1CB;PPP1CC;NCOR1;CREB1;TGS1;CARM1;NAMPT;UBC;RPS27A;SKP1 |

| | | | | | | |
|---|---|---|---|---|---|---|
| 456 | RNA Polymerase III Transcription Initiation From Type 1 Promoter Homo sapiens R-HSA-76061 | 3.87E-02 | 1.21E-01 | 2.18E+00 | 7.08E+00 | CRCP;GTF3C6;POLR3GL;POLR3D;POLR2E;GTF3A;POLR1D;POLR2H;POLR3K;POLR2K;POLR2L |
| 457 | Transport of Mature Transcript to Cytoplasm Homo sapiens R-HSA-72202 | 3.89E-02 | 1.22E-01 | 1.62E+00 | 5.25E+00 | FYTTD1;SLBP;NXT1;NCBP1;RBM8A;POLDIP3;CPSF3;EIF4A3;THOC3;THOC2;THOC5;CDC40;NUP93;DDX39A;FIP1L1;NUP50;SARNP;MAGOH;TPR;SRSF2;SRSF4;RNPS1;EIF4E;SRSF9 |
| 458 | Mitochondrial biogenesis Homo sapiens R-HSA-1592230 | 4.11E-02 | 1.28E-01 | 1.79E+00 | 5.71E+00 | ESRRA;NCOA2;ALAS1;SMARCD3;HDAC3;PRKAG1;PRKAG2;NR1D1;TFB1M;POLRMT;NCOR1;CREB1;TGS1;CARM1;TFAM;CYCS;SSBP1 |
| 459 | Toll Like Receptor 10 (TLR10) Cascade Homo sapiens R-HSA-168142 | 4.25E-02 | 1.32E-01 | 1.56E+00 | 4.94E+00 | ATF1;APP;DHX9;CUL1;HMGB1;ECSIT;PPP2CA;PPP2CB;MAPK9;IRAK1;UBC;MAPK1;RPS27A;SKP1;MAP2K3;DUSP3;MAP3K1;FBXW11;IRAK4;NFKB1;NFKBIA;CREB1;TNIP2;PELI1;UBE2V1;TLR5 |
| 460 | Toll Like Receptor 5 (TLR5) Cascade Homo sapiens R-HSA-168176 | 4.25E-02 | 1.32E-01 | 1.56E+00 | 4.94E+00 | ATF1;APP;DHX9;CUL1;HMGB1;ECSIT;PPP2CA;PPP2CB;MAPK9;IRAK1;UBC;MAPK1;RPS27A;SKP1;MAP2K3;DUSP3;MAP3K1;FBXW11;IRAK4;NFKB1;NFKBIA;CREB1;TNIP2;PELI1;UBE2V1;TLR5 |
| 461 | MyD88 cascade initiated on plasma membrane Homo sapiens R-HSA-975871 | 4.25E-02 | 1.32E-01 | 1.56E+00 | 4.94E+00 | ATF1;APP;DHX9;CUL1;HMGB1;ECSIT;PPP2CA;PPP2CB;MAPK9;IRAK1;UBC;MAPK1;RPS27A;SKP1;MAP2K3;DUSP3;MAP3K1;FBXW11;IRAK4;NFKB1;NFKBIA;CREB1;TNIP2;PELI1;UBE2V1;TLR5 |
| 462 | HDACs deacetylate histones Homo sapiens R-HSA-3214815 | 4.28E-02 | 1.32E-01 | 1.68E+00 | 5.31E+00 | HDAC2;HDAC3;HDAC10;HDAC1;GPS2;ARID4B;CHD4;NCOR2;REST;NCOR1;BRMS1;RBBP4;SAP30L;HMG20B;SAP18;SUDS3;RBBP7;MTA3;HIST1H2AC;RCOR1 |
| 463 | Regulation of TP53 Activity Homo sapiens R-HSA-5633007 | 4.50E-02 | 1.39E-01 | 1.39E+00 | 4.30E+00 | PRR5;PRKAA1;HDAC2;PPP1R13B;MEAF6;PHF20;SETD9;HDAC1;MAPKAP1;PRKAG1;EHMT1;PRKAG2;CHD4;PPP2CA;PPP2CB;NUAK1;RBBP4;AKT2;UBC;MLST8;AKT1;RBBP7;RPS27A;BRD7;SUPT16H;USP7;PDPK1;TAF13;TAF10;USP2;CSNK2A2;TAF11;HIPK1;MTOR;RAD50;CDK5;RPA3;CSNK2B;CDK2;MDM2;RAD17;PIN1;MAPKAPK5;RAD1 |

| | | | | | | |
|---|---|---|---|---|---|---|
| 464 | Signaling by Robo receptor Homo sapiens R-HSA-376176 | 4.52E-02 | 1.39E-01 | 2.02E+00 | 6.25E+00 | CDC42;CAP1;VASP;ROBO3;PAK1;GPC1;PAK6;RAC1;SRGAP3;PFN1;PAK2;PFN2 |
| 465 | CD28 dependent PI3K/Akt signaling Homo sapiens R-HSA-389357 | 4.58E-02 | 1.41E-01 | 2.33E+00 | 7.18E+00 | PRR5;PDPK1;MAPKAP1;AKT2;MLST8;PIK3R3;AKT1;FYN;MTOR |
| 466 | Dual Incision in GG-NER Homo sapiens R-HSA-5696400 | 4.60E-02 | 1.41E-01 | 1.88E+00 | 5.80E+00 | PCNA;PARP1;RFC1;XPA;GTF2H4;RBX1;DDB1;RPA3;ERCC1;UBC;POLD2;ERCC5;POLK;RPS27A |
| 467 | RIG-I/MDA5 mediated induction of IFN-alpha/beta pathways Homo sapiens R-HSA-168928 | 4.70E-02 | 1.44E-01 | 1.56E+00 | 4.77E+00 | APP;DHX9;UBE2D2;UBE2D3;UBE2D1;UBE2L6;HMGB1;TANK;TBK1;RNF216;UBC;FADD;RPS27A;ATG5;MAP3K1;NFKB1;NFKBIA;SIKE1;ITCH;MAVS;IRF3;IRF2;TAX1BP1;PIN1;UBE2K |
| 468 | Metabolism of water-soluble vitamins and cofactors Homo sapiens R-HSA-196849 | 4.85E-02 | 1.48E-01 | 1.49E+00 | 4.50E+00 | PANK2;SLC46A1;MOCS2;PANK3;SHMT1;QPRT;SLC2A1;NADSYN1;ACACA;PPCS;RFK;NAMPT;PRSS3;NADK;AASDHPPT;CYB5A;GSTO2;GSTO1;MMAB;MCCC1;MTHFS;NMNAT3;COASY;LMBRD1;BTD;PC;MTHFD1;PCCB;MMADHC;CD320 |
| 469 | Conversion from APC/C:Cdc20 to APC/C:Cdh1 in late anaphase Homo sapiens R-HSA-176407 | 4.94E-02 | 1.51E-01 | 2.45E+00 | 7.36E+00 | ANAPC15;ANAPC16;CDC26;CDC27;UBE2E1;UBE2D1;ANAPC5;ANAPC11 |
| 470 | COPI-independent Golgi-to-ER retrograde traffic Homo sapiens R-HSA-6811436 | 4.98E-02 | 1.51E-01 | 2.06E+00 | 6.17E+00 | DCTN6;DYNC1H1;DCTN2;GALNT1;DCTN3;RAB18;PLA2G4A;RAB6A;PAFAH1B3;PAFAH1B2;PAFAH1B1 |
| 471 | mRNA Capping Homo sapiens R-HSA-72086 | 4.98E-02 | 1.51E-01 | 2.06E+00 | 6.17E+00 | NCBP1;POLR2B;POLR2C;POLR2E;POLR2H;MNAT1;POLR2J;GTF2F2;GTF2H4;POLR2K;POLR2L |

| | | | | | | |
|---|---|---|---|---|---|---|
| 472 | Costimulation by the CD28 family Homo sapiens R-HSA-388841 | 5.00E-02 | 1.52E-01 | 1.58E+00 | 4.73E+00 | PRR5;YES1;PDPK1;MAPKAP1;PPP2R5A;PIK3R3;PTPN11;MTOR;PPP2CA;CDC42;PPP2CB;PAK1;PPP2R5E;AKT2;MLST8;AKT1;CSK;FYN;PTPN6;GRB2;RAC1;PAK2;HLA-DQB1 |
| 473 | Hemostasis Homo sapiens R-HSA-109582 | 5.24E-02 | 1.58E-01 | 1.18E+00 | 3.48E+00 | APP;CLU;GRB14;KIF5C;AKT2;KIF5B;LAMP2;CFL1;HABP4;AKT1;KIF21A;FAM3C;KIFAP3;SLC16A3;CXADR;ACTN1;APLP2;F2R;ATP1B3;MIF;ATP1B1;TUBA4A;PRKAR1A;ORAI1;TLN1;PFN1;PPIA;GRB7;STXBP3;PDE1A;PDGFB;AK3;ITPR2;PIK3R3;KLC1;PRKCZ;KNG1;RAP1B;CYB5R1;GNG10;RAP1A;PRKAR2B;KIF3A;PRKAR2A;S100A10;TGFB2;YES1;TGFB1;CBX5;HSPA5;RARRES2;H3F3A;L1CAM;PTK2;CDK5;MAFG;CDK2;CALU;GNAS;ESAM;GRB2;GNB5;HRG;CALM1;RAB5A;RCOR1;ITGB1;LGALS3BP;SERPINA1;DAGLA;WDR1;DOCK9;ITGB2;ATP2A3;ARRB1;SERPINA5;CAPZB;PPP2R5E;C1QBP;TIMP3;HMG20B;QSOX1;TIMP1;RAC1;JAK2;JAK1;VAV3;CAP1;ANXA2;PDPK1;ITPK1;RHOG;PLA2G4A;PPP2R5A;YWHAZ;RHOA;SERPINB6;RAD51B;SLC7A6;SLC7A8;KIFC2;KIF2A;IRF3;IRF2;CD47;GAS6;ALDOA;DOCK1;RAPGEF3;CD63;HDAC2;HDAC1;GNAI3;SRI;LRP8;GNAI1;GNAI2;CDC42;PPP2CA;PPP2CB;GNG5;MFN1;MFN2;MAPK1;STX4;CSK;FYN;CD58;SH2B2;CD74;PCDH7;VEGFB;PTPN11;ATP2B1;VEGFA;SOD1;KIF26B;CAPZA1;CAPZA2;CD9;PTPN6;KRAS;BCAR1 |
| 474 | MAP3K8 (TPL2)-dependent MAPK1/3 activation Homo sapiens R-HSA-5684264 | 5.28E-02 | 1.58E-01 | 2.62E+00 | 7.70E+00 | FBXW11;TNIP2;UBC;CUL1;RPS27A;NFKB1;SKP1 |
| 475 | Regulation of KIT signaling Homo sapiens R-HSA-1433559 | 5.28E-02 | 1.58E-01 | 2.62E+00 | 7.70E+00 | YES1;SOCS1;FYN;GRB2;PTPN6;CBL;SH2B2 |
| 476 | Rap1 signalling Homo sapiens R-HSA-392517 | 5.28E-02 | 1.58E-01 | 2.62E+00 | 7.70E+00 | RAP1B;RAP1A;YWHAB;YWHAZ;RAP1GAP;RAPGEF3;SIPA1 |
| 477 | CRMPs in Sema3A signaling Homo sapiens R-HSA-399956 | 5.28E-02 | 1.58E-01 | 2.62E+00 | 7.70E+00 | GSK3B;CDK5;DPYSL2;PLXNA2;CRMP1;FYN;PLXNA4 |

| | | | | | | |
|---|---|---|---|---|---|---|
| 478 | RHO GTPases activate CIT Homo sapiens R-HSA-5625900 | 5.28E-02 | 1.58E-01 | 2.62E+00 | 7.70E+00 | MYL6;CDKN1B;RAC1;RHOC;MYH10;RHOA;MYL12B |
| 479 | Defective HLCS causes multiple carboxylase deficiency Homo sapiens R-HSA-3371599 | 5.30E-02 | 1.58E-01 | 4.48E+00 | 1.32E+01 | PC;PCCB;MCCC1;ACACA |
| 480 | Defects in biotin (Btn) metabolism Homo sapiens R-HSA-3323169 | 5.30E-02 | 1.58E-01 | 4.48E+00 | 1.32E+01 | PC;PCCB;MCCC1;ACACA |
| 481 | Phosphate bond hydrolysis by NUDT proteins Homo sapiens R-HSA-2393930 | 5.30E-02 | 1.58E-01 | 4.48E+00 | 1.32E+01 | NUDT9;NUDT1;NUDT15;NUDT5 |
| 482 | RNA Polymerase I Transcription Initiation Homo sapiens R-HSA-73762 | 5.49E-02 | 1.61E-01 | 1.74E+00 | 5.04E+00 | HDAC2;HDAC1;CHD4;GTF2H4;KAT2B;ZNRD1;RBBP4;TWISTNB;POLR2E;POLR1D;RBBP7;POLR2H;MNAT1;MTA3;POLR2K;POLR2L |
| 483 | Recycling pathway of L1 Homo sapiens R-HSA-437239 | 5.49E-02 | 1.61E-01 | 2.10E+00 | 6.10E+00 | DPYSL2;AP2S1;CLTA;MAPK1;AP2B1;L1CAM;EZR;AP2A2;AP2M1;DNM2 |
| 484 | Signaling by NOTCH Homo sapiens R-HSA-157118 | 5.50E-02 | 1.61E-01 | 1.41E+00 | 4.09E+00 | HDAC2;POGLUT1;HDAC3;HDAC10;HDAC1;HDAC11;SEL1L;CUL1;ATP2A3;CCNC;ARRB1;FURIN;RFNG;RBPJ;HIF1A;APH1A;APH1B;UBC;ST3GAL4;TMED2;ST3GAL6;RAB6A;RPS27A;SKP1;JAG2;PSENEN;MIB2;RBX1;NCOR2;KAT2B;ITCH;SNW1;CREB1;NCOR1;TFDP2;AGO2 |
| 485 | Mitotic Telophase/Cytokinesis Homo sapiens R-HSA-68884 | 5.54E-02 | 1.61E-01 | 2.88E+00 | 8.34E+00 | NUDC;STAG2;RAD21;PDS5B;SMC1A;SMC3 |

| | | | | | | |
|---|---|---|---|---|---|---|
| 486 | Initiation of Nuclear Envelope Reformation Homo sapiens R-HSA-2995383 | 5.54E-02 | 1.61E-01 | 2.88E+00 | 8.34E+00 | PPP2CA;ANKLE2;LMNA;PPP2R2A;LEMD2;TMPO |
| 487 | Nuclear Envelope Reassembly Homo sapiens R-HSA-2995410 | 5.54E-02 | 1.61E-01 | 2.88E+00 | 8.34E+00 | PPP2CA;ANKLE2;LMNA;PPP2R2A;LEMD2;TMPO |
| 488 | Retrograde neurotrophin signalling Homo sapiens R-HSA-177504 | 5.54E-02 | 1.61E-01 | 2.88E+00 | 8.34E+00 | AP2S1;CLTA;AP2B1;AP2A2;AP2M1;DNM2 |
| 489 | NF-kB is activated and signals survival Homo sapiens R-HSA-209560 | 5.54E-02 | 1.61E-01 | 2.88E+00 | 8.34E+00 | NFKBIA;IRAK1;UBC;RPS27A;SQSTM1;NFKB1 |
| 490 | Signal regulatory protein (SIRP) family interactions Homo sapiens R-HSA-391160 | 5.54E-02 | 1.61E-01 | 2.88E+00 | 8.34E+00 | PTPN11;GRB2;PTPN6;CD47;PTK2;SKAP2 |
| 491 | Synthesis, secretion, and inactivation of Glucose-dependent Insulinotropic Polypeptide (GIP) Homo sapiens R-HSA-400511 | 5.54E-02 | 1.61E-01 | 2.88E+00 | 8.34E+00 | DPP4;SPCS3;SPCS2;SPCS1;SEC11A;SEC11C |
| 492 | Loss of Nlp from mitotic centrosomes Homo sapiens R-HSA-380259 | 5.54E-02 | 1.61E-01 | 1.58E+00 | 4.56E+00 | YWHAE;CEP57;OFD1;DYNC1H1;HSP90AA1;DCTN2;CETN2;TUBB;SSNA1;DCTN3;TUBG1;TUBB4B;TUBA4A;HAUS1;ACTR1A;HAUS8;HAUS7;PRKAR2B;AKAP9;MAPRE1;YWHAG;PAFAH1B1 |

| | | | | | | |
|---|---|---|---|---|---|---|
| 493 | Loss of proteins required for interphase microtubule organization?from the centrosome Homo sapiens R-HSA-380284 | 5.54E-02 | 1.61E-01 | 1.58E+00 | 4.56E+00 | YWHAE;CEP57;OFD1;DYNC1H1;HSP90AA1;DCTN2;CETN2;TUBB;SSNA1;DCTN3;TUBG1;TUBB4B;TUBA4A;HAUS1;ACTR1A;HAUS8;HAUS7;PRKAR2B;AKAP9;MAPRE1;YWHAG;PAFAH1B1 |
| 494 | Establishment of Sister Chromatid Cohesion Homo sapiens R-HSA-2468052 | 5.62E-02 | 1.62E-01 | 3.36E+00 | 9.68E+00 | STAG2;RAD21;PDS5B;SMC1A;SMC3 |
| 495 | ER Quality Control Compartment (ERQC) Homo sapiens R-HSA-901032 | 5.62E-02 | 1.62E-01 | 3.36E+00 | 9.68E+00 | EDEM3;OS9;EDEM2;SEL1L;DERL2 |
| 496 | ATF6-alpha activates chaperone genes Homo sapiens R-HSA-381183 | 5.62E-02 | 1.62E-01 | 3.36E+00 | 9.68E+00 | XBP1;HSPA5;CALR;ATF6;ATF4 |
| 497 | SLBP independent Processing of Histone Pre-mRNAs Homo sapiens R-HSA-111367 | 5.62E-02 | 1.62E-01 | 3.36E+00 | 9.68E+00 | LSM10;NCBP1;SNRPG;SNRPD3;SNRPB |
| 498 | Beta-oxidation of pristanoyl-CoA Homo sapiens R-HSA-389887 | 5.62E-02 | 1.62E-01 | 3.36E+00 | 9.68E+00 | ACOT8;SCP2;HSD17B4;CROT;CRAT |
| 499 | RNA polymerase II transcribes snRNA genes Homo sapiens R-HSA-6807505 | 5.98E-02 | 1.71E-01 | 1.52E+00 | 4.30E+00 | GTF2A1;ZNF143;GTF2A2;ICE1;NCBP1;TAF13;GTF2B;SRRT;TAF11;PHAX;RPRD2;GTF2F2;GTF2E2;CDK9;SNAPC1;POLR2B;SNAPC3;POLR2C;POLR2E;RPRD1A;POLR2H;POLR2J;POLR2K;POLR2L |

| | | | | | | |
|---|---|---|---|---|---|---|
| 500 | Regulation of TP53 Activity through Acetylation Homo sapiens R-HSA-6804758 | 6.04E-02 | 1.73E-01 | 2.16E+00 | 6.07E+00 | HDAC2;MEAF6;RBBP4;HDAC1;AKT2;AKT1;CHD4;RBBP7;BRD7 |
| 501 | Branched-chain amino acid catabolism Homo sapiens R-HSA-70895 | 6.04E-02 | 1.73E-01 | 2.16E+00 | 6.07E+00 | ACAD8;HIBADH;SHMT1;IVD;MCCC1;PPM1K;HSD17B10;HIBCH;ALDH9A1 |
| 502 | Cell-Cell communication Homo sapiens R-HSA-1500931 | 6.26E-02 | 1.78E-01 | 1.38E+00 | 3.81E+00 | ITGB1;CD151;FBLIM1;TESK1;ILK;WASL;CLDN1;NTN1;PARD6B;PAK1;PARD6A;FYN;RSU1;RAC1;SPTAN1;VASP;JUP;LAMB3;CADM2;ACTN1;MAGI2;PTPN11;PTK2;CD2AP;MAPK13;CLDN10;CLDN4;CLDN3;CLDN7;CLDN12;CLDN19;CDH13;GRB2;PTPN6;CD47;FERMT2;SKAP2;LIMS1 |
| 503 | Metabolism of carbohydrates Homo sapiens R-HSA-71387 | 6.30E-02 | 1.79E-01 | 1.24E+00 | 3.44E+00 | SLC35B3;SLC25A1;GPI;PYGB;B4GALT3;CHPF;ADPGK;HEXA;SLC2A1;AKR1B1;ENO1;ENO2;ENO3;HK1;NAGLU;HYAL2;CHP1;PGLS;PGM1;MANBA;TPI1;PGAM1;GAA;ABCC5;DCXR;PGAM2;TALDO1;HSPG2;NUP93;PKM;NUP50;B3GNT3;CHST1;B3GNT2;ALDOC;ALDOA;SLC25A11;B4GALT7;GAPDH;PFKFB3;SDC2;SLC5A3;PAPSS1;PPP2CA;PPP2CB;CHST12;GPC1;TPR;UBC;PHKG2;CHST15;ST3GAL4;PGK1;ST3GAL6;GPC5;CHST14;RPS27A;MDH1;GOT1;MDH2;B3GAT3;GOT2;PHKB;GCK;HS3ST1;DERA;PFKL;PC;GNPDA1;GLB1;SDC1;CRYL1;HPSE;CALM1;PFKP;GALK1 |
| 504 | RNA Polymerase I, RNA Polymerase III, and Mitochondrial Transcription Homo sapiens R-HSA-504046 | 6.60E-02 | 1.87E-01 | 1.40E+00 | 3.80E+00 | HDAC2;CRCP;GTF3C6;HDAC1;CHD4;ZNRD1;SNAPC1;RBBP4;SNAPC3;TWISTNB;POLR2E;RBBP7;POLR2H;MTA3;HIST1H2AC;POLR2K;POLR2L;ZNF143;SSB;POLR3GL;CBX3;H2AFZ;H3F3A;MBD2;GTF3A;POLRMT;GTF2H4;KAT2B;POLR3D;NFIC;POLR1D;TFAM;MNAT1;POLR3K |
| 505 | MyD88-independent TLR3/TLR4 cascade Homo sapiens R-HSA-166166 | 6.79E-02 | 1.92E-01 | 1.44E+00 | 3.86E+00 | ATF1;APP;DHX9;UBE2D2;UBE2D3;CUL1;UBE2D1;HMGB1;TANK;PPP2CA;PPP2CB;MAPK9;TBK1;IRAK1;UBC;MAPK1;FADD;RPS27A;SKP1;MAP2K3;DUSP3;FBXW11;PTPN11;NFKB1;NFKBIA;CREB1;IRF3;TNIP2;UBE2V1 |
| 506 | Toll Like Receptor 3 (TLR3) Cascade Homo sapiens R-HSA-168164 | 6.79E-02 | 1.92E-01 | 1.44E+00 | 3.86E+00 | ATF1;APP;DHX9;UBE2D2;UBE2D3;CUL1;UBE2D1;HMGB1;TANK;PPP2CA;PPP2CB;MAPK9;TBK1;IRAK1;UBC;MAPK1;FADD;RPS27A;SKP1;MAP2K3;DUSP3;FBXW11;PTPN11;NFKB1;NFKBIA;CREB1;IRF3;TNIP2;UBE2V1 |

| | | | | | | |
|---|---|---|---|---|---|---|
| 507 | TRIF-mediated TLR3/TLR4 signaling Homo sapiens R-HSA-937061 | 6.79E-02 | 1.92E-01 | 1.44E+00 | 3.86E+00 | ATF1;APP;DHX9;UBE2D2;UBE2D3;CUL1;UBE2D1;HMGB1;TANK;PPP2CA;PPP2CB;MAPK9;TBK1;IRAK1;UBC;MAPK1;FADD;RPS27A;SKP1;MAP2K3;DUSP3;FBXW11;PTPN11;NFKB1;NFKBIA;CREB1;IRF3;TNIP2;UBE2V1 |
| 508 | Intraflagellar transport Homo sapiens R-HSA-5620924 | 6.81E-02 | 1.92E-01 | 1.75E+00 | 4.69E+00 | IFT80;WDR34;HSPB11;IFT20;IFT74;IFT22;KIF3A;WDR60;TCTEX1D2;TRIP11;DYNLRB1;TNPO1;KIFAP3;IFT57 |
| 509 | SUMOylation Homo sapiens R-HSA-2990846 | 6.92E-02 | 1.94E-01 | 1.42E+00 | 3.80E+00 | SP100;PCNA;CETN2;SMC3;HERC2;SENP5;SUMO1;RAD21;SUMO3;TDG;TPR;SUMO2;PHC2;UBE2I;CBX4;PARP1;PCGF2;SMC1A;NUP93;RNF168;HNRNPK;STAG2;NSMCE4A;NUP50;SP3;MDM2;UBA2;HNRNPC;TOP1;SAE1 |
| 510 | RNA Polymerase III Transcription Initiation From Type 2 Promoter Homo sapiens R-HSA-76066 | 6.99E-02 | 1.94E-01 | 1.98E+00 | 5.27E+00 | CRCP;GTF3C6;POLR3GL;POLR3D;POLR2E;POLR1D;POLR2H;POLR3K;POLR2K;POLR2L |
| 511 | RNA Pol II CTD phosphorylation and interaction with CE Homo sapiens R-HSA-167160 | 6.99E-02 | 1.94E-01 | 1.98E+00 | 5.27E+00 | POLR2B;POLR2C;POLR2E;POLR2H;MNAT1;POLR2J;GTF2F2;GTF2H4;POLR2K;POLR2L |
| 512 | RNA Pol II CTD phosphorylation and interaction with CE Homo sapiens R-HSA-77075 | 6.99E-02 | 1.94E-01 | 1.98E+00 | 5.27E+00 | POLR2B;POLR2C;POLR2E;POLR2H;MNAT1;POLR2J;GTF2F2;GTF2H4;POLR2K;POLR2L |
| 513 | Integrin alphaIIb beta3 signaling Homo sapiens R-HSA-354192 | 6.99E-02 | 1.94E-01 | 1.98E+00 | 5.27E+00 | RAP1B;RAP1A;PDPK1;AKT1;CSK;GRB2;TLN1;PTK2;RAPGEF3;BCAR1 |
| 514 | Peroxisomal lipid metabolism Homo sapiens R-HSA-390918 | 6.99E-02 | 1.94E-01 | 1.98E+00 | 5.27E+00 | ALDH3A2;PECR;ACOT8;SCP2;IDH1;ECI2;HSD17B4;HACL1;CROT;CRAT |

| | | | | | | |
|---|---|---|---|---|---|---|
| 515 | RORA activates gene expression Homo sapiens R-HSA-1368082 | 6.99E-02 | 1.94E-01 | 1.98E+00 | 5.27E+00 | NCOA2;SMARCD3;HDAC3;NCOR1;TGS1;CARM1;RORA;NR1D1;SIRT1;ARNTL |
| 516 | Pyruvate metabolism Homo sapiens R-HSA-70268 | 6.99E-02 | 1.94E-01 | 1.98E+00 | 5.27E+00 | LDHB;LDHA;PDHA1;GLO1;MPC2;PDK3;PDHB;HAGH;SLC16A3;PDK2 |
| 517 | Metabolism of vitamins and cofactors Homo sapiens R-HSA-196854 | 7.13E-02 | 1.97E-01 | 1.33E+00 | 3.53E+00 | VKORC1L1;PANK2;SLC46A1;MOCS2;PANK3;SDC2;SHMT1;QPRT;SLC2A1;NADSYN1;LRP8;ACACA;PPCS;RDH11;RFK;GPC1;NAMPT;GPC5;BCO2;PRSS3;NADK;AASDHPPT;VKORC1;CYB5A;GSTO2;GSTO1;MMAB;AKR1C1;MCCC1;MTHFS;NMNAT3;HSPG2;COASY;LMBRD1;BTD;PC;MTHFD1;PCCB;SDC1;LRP10;MMADHC;CD320 |
| 518 | Trafficking of GluR2-containing AMPA receptors Homo sapiens R-HSA-416993 | 7.25E-02 | 1.99E-01 | 2.35E+00 | 6.18E+00 | NSF;TSPAN7;AP2S1;PICK1;AP2B1;AP2A2;AP2M1 |
| 519 | KSRP (KHSRP) binds and destabilizes mRNA Homo sapiens R-HSA-450604 | 7.25E-02 | 1.99E-01 | 2.35E+00 | 6.18E+00 | EXOSC7;EXOSC4;AKT1;EXOSC3;YWHAZ;DCP2;EXOSC1 |
| 520 | Beta-catenin phosphorylation cascade Homo sapiens R-HSA-196299 | 7.25E-02 | 1.99E-01 | 2.35E+00 | 6.18E+00 | PPP2CA;GSK3B;PPP2CB;FRAT1;CSNK1A1;PPP2R5E;PPP2R5A |
| 521 | RHO GTPases Activate ROCKs Homo sapiens R-HSA-5627117 | 7.25E-02 | 1.99E-01 | 2.35E+00 | 6.18E+00 | PPP1CB;MYL6;LIMK2;RHOC;MYH10;RHOA;MYL12B |
| 522 | Mitochondrial Fatty Acid Beta-Oxidation Homo sapiens R-HSA-77289 | 7.25E-02 | 1.99E-01 | 2.35E+00 | 6.18E+00 | HADHB;HADHA;ACADVL;ECHS1;PCCB;ACADM;HADH |
| 523 | RHO GTPases activate PKNs Homo sapiens R-HSA-5625740 | 7.59E-02 | 2.08E-01 | 1.56E+00 | 4.02E+00 | YWHAE;NCOA2;YWHAB;H2AFZ;PDPK1;H3F3A;RHOC;YWHAZ;RHOA;MYL12B;PPP1CB;MYL6;YWHAQ;RAC1;PKN1;MYH10;YWHAG;HIST1H2AC;YWHAH |

| | | | | | | |
|---|---|---|---|---|---|---|
| 524 | Transcriptional regulation of white adipocyte differentiation Homo sapiens R-HSA-381340 | 7.74E-02 | 2.11E-01 | 1.47E+00 | 3.76E+00 | NCOA2;SMARCD3;HDAC3;TGFB1;CEBPD;NCOA3;MED8;CCNC;MED4;MED27;TNF;SREBF2;NFKB1;NCOR2;MED11;NCOR1;MED24;KLF5;MED30;CDK4;MED10;MED21;TGS1;CARM1 |
| 525 | SUMOylation of transcription factors Homo sapiens R-HSA-3232118 | 7.87E-02 | 2.12E-01 | 2.52E+00 | 6.41E+00 | UBE2I;SUMO1;SUMO3;SP3;SUMO2;MDM2 |
| 526 | TP53 regulates transcription of several additional cell death genes whose specific roles in p53-dependent apoptosis remain uncertain Homo sapiens R-HSA-6803205 | 7.87E-02 | 2.12E-01 | 2.52E+00 | 6.41E+00 | PPP1R13B;TP53I3;BCL6;PERP;CHM;RABGGTB |
| 527 | AMER1 mutants destabilize the destruction complex Homo sapiens R-HSA-4839748 | 7.87E-02 | 2.12E-01 | 2.52E+00 | 6.41E+00 | PPP2CA;GSK3B;PPP2CB;CSNK1A1;PPP2R5E;PPP2R5A |
| 528 | Truncations of AMER1 destabilize the destruction complex Homo sapiens R-HSA-5467348 | 7.87E-02 | 2.12E-01 | 2.52E+00 | 6.41E+00 | PPP2CA;GSK3B;PPP2CB;CSNK1A1;PPP2R5E;PPP2R5A |

| | | | | | | |
|---|---|---|---|---|---|---|
| 529 | truncated APC mutants destabilize the destruction complex Homo sapiens R-HSA-4839744 | 7.87E-02 | 2.12E-01 | 2.52E+00 | 6.41E+00 | PPP2CA;GSK3B;PPP2CB;CSNK1A1;PPP2R5E;PPP2R5A |
| 530 | AXIN missense mutants destabilize the destruction complex Homo sapiens R-HSA-5467340 | 7.87E-02 | 2.12E-01 | 2.52E+00 | 6.41E+00 | PPP2CA;GSK3B;PPP2CB;CSNK1A1;PPP2R5E;PPP2R5A |
| 531 | AXIN mutants destabilize the destruction complex, activating WNT signaling Homo sapiens R-HSA-4839735 | 7.87E-02 | 2.12E-01 | 2.52E+00 | 6.41E+00 | PPP2CA;GSK3B;PPP2CB;CSNK1A1;PPP2R5E;PPP2R5A |
| 532 | APC truncation mutants have impaired AXIN binding Homo sapiens R-HSA-5467337 | 7.87E-02 | 2.12E-01 | 2.52E+00 | 6.41E+00 | PPP2CA;GSK3B;PPP2CB;CSNK1A1;PPP2R5E;PPP2R5A |
| 533 | TRAF6 mediated induction of NFkB and MAP kinases upon TLR7/8 or 9 activation Homo sapiens R-HSA-975138 | 7.92E-02 | 2.13E-01 | 1.45E+00 | 3.68E+00 | ATF1;APP;DHX9;CUL1;HMGB1;ECSIT;PPP2CA;PPP2CB;MAPK9;IRAK1;UBC;MAPK1;RPS27A;SKP1;MAP2K3;DUSP3;MAP3K1;FBXW11;IRAK4;NFKB1;NFKBIA;CREB1;TNIP2;PELI1;UBE2V1 |
| 534 | Activation of gene expression by SREBF (SREBP) Homo sapiens R-HSA-2426168 | 8.14E-02 | 2.18E-01 | 1.68E+00 | 4.22E+00 | IDI1;NCOA2;FDPS;MVK;SMARCD3;CYP51A1;ELOVL6;ACACA;SREBF2;SQLE;TGS1;CARM1;SC5D;FDFT1 |

| | | | | | | |
|---|---|---|---|---|---|---|
| 535 | APOBEC3G mediated resistance to HIV-1 infection Homo sapiens R-HSA-180689 | 8.29E-02 | 2.19E-01 | 5.04E+00 | 1.26E+01 | HMGA1;PSIP1;PPIA |
| 536 | Loss of Function of FBXW7 in Cancer and NOTCH1 Signaling Homo sapiens R-HSA-2644607 | 8.29E-02 | 2.19E-01 | 5.04E+00 | 1.26E+01 | CUL1;RBX1;SKP1 |
| 537 | FBXW7 Mutants and NOTCH1 in Cancer Homo sapiens R-HSA-2644605 | 8.29E-02 | 2.19E-01 | 5.04E+00 | 1.26E+01 | CUL1;RBX1;SKP1 |
| 538 | Cytochrome c-mediated apoptotic response Homo sapiens R-HSA-111461 | 8.29E-02 | 2.19E-01 | 5.04E+00 | 1.26E+01 | CASP7;APAF1;CYCS |
| 539 | Activation of caspases through apoptosome-mediated cleavage Homo sapiens R-HSA-111459 | 8.29E-02 | 2.19E-01 | 5.04E+00 | 1.26E+01 | CASP7;APAF1;CYCS |
| 540 | Trafficking of myristoylated proteins to the cilium Homo sapiens R-HSA-5624138 | 8.29E-02 | 2.19E-01 | 5.04E+00 | 1.26E+01 | UNC119B;ARL3;CYS1 |
| 541 | Signaling by MST1 Homo sapiens R-HSA-8852405 | 8.29E-02 | 2.19E-01 | 5.04E+00 | 1.26E+01 | SPINT1;HPN;SPINT2 |
| 542 | AURKA Activation by TPX2 Homo sapiens R-HSA-8854518 | 8.35E-02 | 2.21E-01 | 1.48E+00 | 3.68E+00 | YWHAE;CEP57;OFD1;DYNC1H1;HSP90AA1;DCTN2;CETN2;TUBB;SSNA1;DCTN3;TUBG1;TUBB4B;TUBA4A;HAUS1;ACTR1A;HAUS8;HAUS7;PRKAR2B;AKAP9;MAPRE1;YWHAG;PAFAH1B1 |

| | | | | | | |
|---|---|---|---|---|---|---|
| 543 | Uptake and function of anthrax toxins Homo sapiens R-HSA-5210891 | 8.41E-02 | 2.21E-01 | 2.80E+00 | 6.94E+00 | MAP2K3;MAP2K2;PDCD6IP;FURIN;CALM1 |
| 544 | Interleukin-6 signaling Homo sapiens R-HSA-1059683 | 8.41E-02 | 2.21E-01 | 2.80E+00 | 6.94E+00 | STAT3;PTPN11;CBL;JAK2;JAK1 |
| 545 | Biotin transport and metabolism Homo sapiens R-HSA-196780 | 8.41E-02 | 2.21E-01 | 2.80E+00 | 6.94E+00 | BTD;PC;PCCB;MCCC1;ACACA |
| 546 | Chromatin organization Homo sapiens R-HSA-4839726 | 8.54E-02 | 2.23E-01 | 1.25E+00 | 3.07E+00 | SUV39H2;SMARCB1;SETD3;HDAC10;PHF20;EHMT1;ASH2L;ARID4B;CHD4;ING4;MECOM;KAT8;HMG20B;JAK2;HIST1H2AC;NCOA2;KDM2A;PRMT1;H2AFZ;ACTL6A;MSL3;JMJD6;NCOR2;TAF6L;KAT2B;MORF4L1;NCOR1;BRMS1;HAT1;SMARCD2;HDAC2;SMARCD3;HDAC3;MEAF6;HDAC1;GPS2;RBBP4;SAP30L;MBIP;SAP18;BRD8;SMYD3;SUDS3;DMAP1;RBBP7;MTA3;SUZ12;DR1;MCRS1;TAF10;SETDB2;ELP5;ELP6;COPRS;REST;MRGBP;CDK4;CARM1;TADA3;OGT;RCOR1 |
| 547 | Chromatin modifying enzymes Homo sapiens R-HSA-3247509 | 8.54E-02 | 2.23E-01 | 1.25E+00 | 3.07E+00 | SUV39H2;SMARCB1;SETD3;HDAC10;PHF20;EHMT1;ASH2L;ARID4B;CHD4;ING4;MECOM;KAT8;HMG20B;JAK2;HIST1H2AC;NCOA2;KDM2A;PRMT1;H2AFZ;ACTL6A;MSL3;JMJD6;NCOR2;TAF6L;KAT2B;MORF4L1;NCOR1;BRMS1;HAT1;SMARCD2;HDAC2;SMARCD3;HDAC3;MEAF6;HDAC1;GPS2;RBBP4;SAP30L;MBIP;SAP18;BRD8;SMYD3;SUDS3;DMAP1;RBBP7;MTA3;SUZ12;DR1;MCRS1;TAF10;SETDB2;ELP5;ELP6;COPRS;REST;MRGBP;CDK4;CARM1;TADA3;OGT;RCOR1 |
| 548 | HS-GAG degradation Homo sapiens R-HSA-2024096 | 8.64E-02 | 2.24E-01 | 2.07E+00 | 5.07E+00 | NAGLU;GLB1;SDC2;GPC1;SDC1;GPC5;HPSE;HSPG2 |
| 549 | Signal transduction by L1 Homo sapiens R-HSA-445144 | 8.64E-02 | 2.24E-01 | 2.07E+00 | 5.07E+00 | ITGB1;PAK1;MAP2K2;CSNK2A2;CSNK2B;MAPK1;RAC1;L1CAM |

| | | | | | | |
|---|---|---|---|---|---|---|
| 550 | Vitamin C (ascorbate) metabolism Homo sapiens R-HSA-196836 | 8.71E-02 | 2.24E-01 | 3.36E+00 | 8.21E+00 | CYB5A;GSTO2;GSTO1;SLC2A1 |
| 551 | WNT mediated activation of DVL Homo sapiens R-HSA-201688 | 8.71E-02 | 2.24E-01 | 3.36E+00 | 8.21E+00 | CSNK2A2;CSNK2B;DVL3;PIP5K1B |
| 552 | Downregulation of ERBB4 signaling Homo sapiens R-HSA-1253288 | 8.71E-02 | 2.24E-01 | 3.36E+00 | 8.21E+00 | ITCH;NEDD4;UBC;RPS27A |
| 553 | Regulation of cytoskeletal remodeling and cell spreading by IPP complex components Homo sapiens R-HSA-446388 | 8.71E-02 | 2.24E-01 | 3.36E+00 | 8.21E+00 | ACTN1;TESK1;RSU1;LIMS1 |
| 554 | Coenzyme A biosynthesis Homo sapiens R-HSA-196783 | 8.71E-02 | 2.24E-01 | 3.36E+00 | 8.21E+00 | PANK2;PANK3;PPCS;COASY |
| 555 | NR1D1 (REV-ERBA) represses gene expression Homo sapiens R-HSA-1368071 | 8.71E-02 | 2.24E-01 | 3.36E+00 | 8.21E+00 | NCOR1;HDAC3;NR1D1;ARNTL |
| 556 | PERK regulates gene expression Homo sapiens R-HSA-381042 | 8.72E-02 | 2.24E-01 | 1.87E+00 | 4.56E+00 | EXOSC7;EXOSC4;HSPA5;EXOSC3;EIF2S1;ATF6;EXOSC1;DCP2;HERPUD1;ATF4 |
| 557 | Ca-dependent events Homo sapiens R-HSA-111996 | 8.72E-02 | 2.24E-01 | 1.87E+00 | 4.56E+00 | CREB1;PRKAR2B;PRKAR1A;PDE1A;PRKAR2A;ADCY3;MAPK1;PLA2G4A;CALM1;ADCY5 |
| 558 | Toll Like Receptor 9 (TLR9) Cascade Homo sapiens R-HSA-168138 | 9.07E-02 | 2.32E-01 | 1.41E+00 | 3.39E+00 | ATF1;APP;DHX9;CUL1;HMGB1;ECSIT;EEA1;PPP2CA;PPP2CB;MAPK9;IRAK1;UBC;MAPK1;RPS27A;SKP1;MAP2K3;DUSP3;MAP3K1;FBXW11;IRAK4;NFKB1;NFKBIA;CREB1;TNIP2;PELI1;UBE2V1 |

| | | | | | | |
|---|---|---|---|---|---|---|
| 559 | Association of TriC/CCT with target proteins during biosynthesis Homo sapiens R-HSA-390471 | 9.10E-02 | 2.33E-01 | 1.68E+00 | 4.03E+00 | CCT3;CCT2;HDAC3;STAT3;CCT6A;TCP1;XRN2;CCT8;FKBP9;CCT7;FBXL5;CCT5;CCT4 |
| 560 | MyD88:Mal cascade initiated on plasma membrane Homo sapiens R-HSA-166058 | 9.20E-02 | 2.34E-01 | 1.40E+00 | 3.34E+00 | ATF1;APP;DHX9;CUL1;HMGB1;ECSIT;PPP2CA;PPP2CB;MAPK9;SOCS1;IRAK1;UBC;MAPK1;RPS27A;SKP1;MAP2K3;DUSP3;MAP3K1;FBXW11;IRAK4;NFKB1;SIGIRR;NFKBIA;CREB1;TNIP2;PELI1;UBE2V1 |
| 561 | Toll Like Receptor TLR1:TLR2 Cascade Homo sapiens R-HSA-168179 | 9.20E-02 | 2.34E-01 | 1.40E+00 | 3.34E+00 | ATF1;APP;DHX9;CUL1;HMGB1;ECSIT;PPP2CA;PPP2CB;MAPK9;SOCS1;IRAK1;UBC;MAPK1;RPS27A;SKP1;MAP2K3;DUSP3;MAP3K1;FBXW11;IRAK4;NFKB1;SIGIRR;NFKBIA;CREB1;TNIP2;PELI1;UBE2V1 |
| 562 | Toll Like Receptor TLR6:TLR2 Cascade Homo sapiens R-HSA-168188 | 9.20E-02 | 2.34E-01 | 1.40E+00 | 3.34E+00 | ATF1;APP;DHX9;CUL1;HMGB1;ECSIT;PPP2CA;PPP2CB;MAPK9;SOCS1;IRAK1;UBC;MAPK1;RPS27A;SKP1;MAP2K3;DUSP3;MAP3K1;FBXW11;IRAK4;NFKB1;SIGIRR;NFKBIA;CREB1;TNIP2;PELI1;UBE2V1 |
| 563 | Toll Like Receptor 2 (TLR2) Cascade Homo sapiens R-HSA-181438 | 9.20E-02 | 2.34E-01 | 1.40E+00 | 3.34E+00 | ATF1;APP;DHX9;CUL1;HMGB1;ECSIT;PPP2CA;PPP2CB;MAPK9;SOCS1;IRAK1;UBC;MAPK1;RPS27A;SKP1;MAP2K3;DUSP3;MAP3K1;FBXW11;IRAK4;NFKB1;SIGIRR;NFKBIA;CREB1;TNIP2;PELI1;UBE2V1 |
| 564 | Positive epigenetic regulation of rRNA expression Homo sapiens R-HSA-5250913 | 9.47E-02 | 2.40E-01 | 1.45E+00 | 3.42E+00 | GSK3B;HDAC2;H2AFZ;HDAC1;CBX3;H3F3A;SMARCA5;DDX21;CHD4;DEK;KAT2B;ZNRD1;RBBP4;TWISTNB;POLR1D;POLR2E;RBBP7;POLR2H;MTA3;HIST1H2AC;POLR2K;POLR2L |
| 565 | Miscellaneous transport and binding events Homo sapiens R-HSA-5223345 | 9.60E-02 | 2.42E-01 | 2.14E+00 | 5.02E+00 | DMTN;MMGT1;TUSC3;NIPAL3;CTNS;NIPA2;ADD1 |
| 566 | Regulation of signaling by CBL Homo sapiens R-HSA-912631 | 9.60E-02 | 2.42E-01 | 2.14E+00 | 5.02E+00 | YES1;RAPGEF1;PIK3R3;FYN;GRB2;CBL;CRKL |

| | | | | | | |
|---|---|---|---|---|---|---|
| 567 | Immune System Homo sapiens R-HSA-168256 | 9.60E-02 | 2.42E-01 | 1.09E+00 | 2.55E+00 | ATF1;EIF4A2;CYFIP2;NCKAP1;EIF4A1;APP;IFITM2;EIF4A3;KEAP1;WDR83;UBE2L6;CRKL;UBE2L3;PSMD8;HERC4;LGALS3;PSMD9;HERC3;PSMD6;RNF115;TBK1;HERC2;PSMD7;RNF114;PSMD4;AKT2;CDC26;PSMD2;CDC27;PSMD3;AP1S2;AKT1;FBXO3;PSMD1;PIP4K2C;ATP6V1E1;PRSS3;KIFAP3;IER3;SKP1;ATP6V1G1;CXADR;ATP6V0E1;FBXO2;IL1R2;HLA-C;HLA-G;HLA-E;SIGIRR;BTC;NUP93;RNF126;PRKAR1A;AKAP9;PSME1;ORAI1;PSME2;RBCK1;TLN1;ATP6V0D1;ANAPC13;SPTBN5;SAR1B;DHX9;PRKDC;MAPKAP1;PDE1A;PDGFB;PIK3R3;BCL10;UBE2J2;ANAPC11;UBE2J1;RNF216;PRKAR2B;PRKAR2A;PIP5K1B;PSMF1;FADD;ATP6V1C1;ATG5;VASP;CTSA;XRCC6;YES1;XRCC5;TNFSF12;EIF2AK2;MYO5A;LNPEP;PTK2;UBAC1;AGO2;REL;GRB2;CALR;CALM1;ARF1;CDKN1B;SH3KBP1;ARPC1B;ARPC1A;CLTC;CLTA;WASL;ECSIT;AP2A2;EEA1;ACTR1A;PPP3CB;ACTR1B;CTSL;TRIM5;SEC61G;CTSK;TRIM2;PEA15;ARIH2;CTSH;RAC1;SEC61B;CTSF;JAK2;SPTAN1;CTSD;JAK1;CTSB;VAV3;ATP6V0B;DUSP3;TNFRSF12A;DUSP1;IFNGR2;DUSP8;IL17RB;PSMA5;PSMA6;SIKE1;PSMA3;PSMA4;PSMA1;KIF2A;GAN;PSMA2;RAPGEF1;TAX1BP1;PELI1;TLR5;CD46;ATP6V0C;RAPGEF3;HLA-DQB1;RAB7A;HBEGF;SP100;ADCY3;TANK;PSMA7;ADCY5;PPP2CA;PSMB6;PPP2CB;PSMB7;HLA-DMA;PAK1;PSMB5;HLA-DMB;PSMB2;PSMB3;PSMB1;AP2S1;STX4;CD59;ATP6V0E2;RPS27A;PAK2;CD55;DYNC1H1;CD74;MAP3K1;IL10RB;STAT3;DAB2IP;MIB2;UBE2G1;UBE2G2;AP2B1;P[illegible] |
| 568 | Senescence-Associated Secretory Phenotype (SASP) Homo sapiens R-HSA-2559582 | 9.66E-02 | 2.43E-01 | 1.43E+00 | 3.35E+00 | CDKN2D;CDKN1B;ANAPC15;ANAPC16;H2AFZ;H3F3A;STAT3;UBE2E1;UBE2D1;EHMT1;NFKB1;ANAPC11;CDK6;CDK4;CDC26;CDC27;UBC;CDK2;MAPK1;ANAPC5;IGFBP7;RPS27A;HIST1H2AC |
| 569 | Post-translational modification: synthesis of GPI-anchored proteins Homo sapiens R-HSA-163125 | 9.76E-02 | 2.45E-01 | 1.89E+00 | 4.40E+00 | DPM2;DPM3;PIGT;PGAP1;PIGP;PIGK;PIGF;PIGX;PIGH |

| | | | | | | |
|---|---|---|---|---|---|---|
| 570 | Antigen Presentation: Folding, assembly and peptide loading of class I MHC Homo sapiens R-HSA-983170 | 9.76E-02 | 2.45E-01 | 1.89E+00 | 4.40E+00 | PDIA3;SEC24A;HSPA5;SAR1B;HLA-C;TAP1;CALR;HLA-G;HLA-E |
| 571 | RAF activation Homo sapiens R-HSA-5673000 | 9.76E-02 | 2.45E-01 | 1.89E+00 | 4.40E+00 | PPP2CA;PPP2CB;MAP2K2;YWHAB;PPP2R5E;PPP2R5A;KRAS;PHB;JAK2 |
| 572 | MyD88 dependent cascade initiated on endosome Homo sapiens R-HSA-975155 | 9.99E-02 | 2.49E-01 | 1.40E+00 | 3.23E+00 | ATF1;APP;DHX9;CUL1;HMGB1;ECSIT;PPP2CA;PPP2CB;MAPK9;IRAK1;UBC;MAPK1;RPS27A;SKP1;MAP2K3;DUSP3;MAP3K1;FBXW11;IRAK4;NFKB1;NFKBIA;CREB1;TNIP2;PELI1;UBE2V1 |
| 573 | Toll Like Receptor 7/8 (TLR7/8) Cascade Homo sapiens R-HSA-168181 | 9.99E-02 | 2.49E-01 | 1.40E+00 | 3.23E+00 | ATF1;APP;DHX9;CUL1;HMGB1;ECSIT;PPP2CA;PPP2CB;MAPK9;IRAK1;UBC;MAPK1;RPS27A;SKP1;MAP2K3;DUSP3;MAP3K1;FBXW11;IRAK4;NFKB1;NFKBIA;CREB1;TNIP2;PELI1;UBE2V1 |
| 574 | Apoptotic execution phase Homo sapiens R-HSA-75153 | 1.05E-01 | 2.61E-01 | 1.54E+00 | 3.47E+00 | DBNL;GSN;HMGB1;ADD1;PTK2;TJP1;CASP7;H1F0;STK24;LMNA;HIST1H1E;ACIN1;DNM1L;SPTAN1;PAK2;KPNA1 |
| 575 | Depolymerisation of the Nuclear Lamina Homo sapiens R-HSA-4419969 | 1.07E-01 | 2.61E-01 | 2.24E+00 | 5.02E+00 | CCNB1;CNEP1R1;LMNA;LEMD2;LPIN2;TMPO |
| 576 | Mismatch Repair Homo sapiens R-HSA-5358508 | 1.07E-01 | 2.61E-01 | 2.24E+00 | 5.02E+00 | MSH6;PCNA;MSH2;MSH3;RPA3;POLD2 |
| 577 | NRIF signals cell death from the nucleus Homo sapiens R-HSA-205043 | 1.07E-01 | 2.61E-01 | 2.24E+00 | 5.02E+00 | PSENEN;APH1A;APH1B;UBC;RPS27A;SQSTM1 |

| | | | | | | |
|---|---|---|---|---|---|---|
| 578 | Glycogen breakdown (glycogenolysis) Homo sapiens R-HSA-70221 | 1.07E-01 | 2.61E-01 | 2.24E+00 | 5.02E+00 | PYGB;GAA;PHKG2;PHKB;CALM1;PGM1 |
| 579 | S33 mutants of beta-catenin aren't phosphorylated Homo sapiens R-HSA-5358747 | 1.07E-01 | 2.61E-01 | 2.24E+00 | 5.02E+00 | PPP2CA;GSK3B;PPP2CB;CSNK1A1;PPP2R5E;PPP2R5A |
| 580 | S45 mutants of beta-catenin aren't phosphorylated Homo sapiens R-HSA-5358751 | 1.07E-01 | 2.61E-01 | 2.24E+00 | 5.02E+00 | PPP2CA;GSK3B;PPP2CB;CSNK1A1;PPP2R5E;PPP2R5A |
| 581 | phosphorylation site mutants of CTNNB1 are not targeted to the proteasome by the destruction complex Homo sapiens R-HSA-4839743 | 1.07E-01 | 2.61E-01 | 2.24E+00 | 5.02E+00 | PPP2CA;GSK3B;PPP2CB;CSNK1A1;PPP2R5E;PPP2R5A |
| 582 | T41 mutants of beta-catenin aren't phosphorylated Homo sapiens R-HSA-5358752 | 1.07E-01 | 2.61E-01 | 2.24E+00 | 5.02E+00 | PPP2CA;GSK3B;PPP2CB;CSNK1A1;PPP2R5E;PPP2R5A |
| 583 | S37 mutants of beta-catenin aren't phosphorylated Homo sapiens R-HSA-5358749 | 1.07E-01 | 2.61E-01 | 2.24E+00 | 5.02E+00 | PPP2CA;GSK3B;PPP2CB;CSNK1A1;PPP2R5E;PPP2R5A |

| | | | | | | |
|---|---|---|---|---|---|---|
| 584 | Misspliced GSK3beta mutants stabilize beta-catenin Homo sapiens R-HSA-5339716 | 1.07E-01 | 2.61E-01 | 2.24E+00 | 5.02E+00 | PPP2CA;GSK3B;PPP2CB;CSNK1A1;PPP2R5E;PPP2R5A |
| 585 | Negative regulation of FGFR3 signaling Homo sapiens R-HSA-5654732 | 1.07E-01 | 2.61E-01 | 1.77E+00 | 3.96E+00 | PPP2CA;PPP2CB;UBC;FGF18;MAPK1;PTPN11;GRB2;CBL;RPS27A;FGF2 |
| 586 | Negative regulation of MAPK pathway Homo sapiens R-HSA-5675221 | 1.08E-01 | 2.63E-01 | 1.62E+00 | 3.61E+00 | MAP2K2;YWHAB;DUSP1;PEBP1;PPP2R5A;DUSP8;PPP2CA;PPP2CB;PPP2R5E;UBC;MAPK1;KRAS;RPS27A |
| 587 | Post-chaperonin tubulin folding pathway Homo sapiens R-HSA-389977 | 1.10E-01 | 2.67E-01 | 1.92E+00 | 4.25E+00 | TUBA1C;TUBA1B;TUBB6;ARL2;TBCB;TBCA;TUBB4B;TUBA4A |
| 588 | Intra-Golgi traffic Homo sapiens R-HSA-6811438 | 1.13E-01 | 2.74E-01 | 1.57E+00 | 3.43E+00 | NAPA;NSF;ARF1;RIC1;COG6;GOSR2;COG2;NAPG;CYTH3;MAN2A2;MAN1C1;STX5;TRIP11;CYTH1 |
| 589 | Signaling by FGFR in disease Homo sapiens R-HSA-1226099 | 1.14E-01 | 2.76E-01 | 1.45E+00 | 3.16E+00 | CPSF6;NCBP1;STAT3;ERLIN2;FGF2;GTF2F2;BAG4;POLR2B;POLR2C;FGFR1OP2;FGF18;POLR2E;KRAS;GRB2;LRRFIP1;POLR2H;POLR2J;POLR2K;POLR2L |
| 590 | FGFR2 mutant receptor activation Homo sapiens R-HSA-1839126 | 1.14E-01 | 2.77E-01 | 1.68E+00 | 3.65E+00 | NCBP1;POLR2B;POLR2C;FGF18;POLR2E;POLR2H;FGF2;POLR2J;GTF2F2;POLR2K;POLR2L |
| 591 | Smooth Muscle Contraction Homo sapiens R-HSA-445355 | 1.14E-01 | 2.77E-01 | 1.68E+00 | 3.65E+00 | MYL6;ANXA2;TPM4;TPM3;TPM1;MYL6B;TLN1;CALM1;MYL12A;MYLK;MYL12B |

| | | | | | | |
|---|---|---|---|---|---|---|
| 592 | Integration of energy metabolism Homo sapiens R-HSA-163685 | 1.16E-01 | 2.82E-01 | 1.32E+00 | 2.84E+00 | SLC2A1;ARL2;ITPR2;PRKAG2;ADCY3;CACNA1D;CACNA1C;AKAP5;ADIPOR1;ACACA;GNAI1;ADCY5;GNAI2;PPP2CA;PPP2CB;GNG10;RAP1A;GNG5;PRKAR2B;PRKAR2A;MLX;TALDO1;ARL2BP;PRKAR1A;GNAS;GNB5;SLC25A5;PLCB1;SLC25A4;RAPGEF3;SLC25A6 |
| 593 | Defective EXT2 causes exostoses 2 Homo sapiens R-HSA-3656237 | 1.18E-01 | 2.82E-01 | 2.40E+00 | 5.14E+00 | GPC1;SDC2;SDC1;GPC5;HSPG2 |
| 594 | Defective EXT1 causes exostoses 1, TRPS2 and CHDS Homo sapiens R-HSA-3656253 | 1.18E-01 | 2.82E-01 | 2.40E+00 | 5.14E+00 | GPC1;SDC2;SDC1;GPC5;HSPG2 |
| 595 | Endosomal/Vacuolar pathway Homo sapiens R-HSA-1236977 | 1.18E-01 | 2.82E-01 | 2.40E+00 | 5.14E+00 | CTSL;HLA-C;LNPEP;HLA-G;HLA-E |
| 596 | ATF6-alpha activates chaperones Homo sapiens R-HSA-381033 | 1.18E-01 | 2.82E-01 | 2.40E+00 | 5.14E+00 | XBP1;HSPA5;CALR;ATF6;ATF4 |
| 597 | Caspase-mediated cleavage of cytoskeletal proteins Homo sapiens R-HSA-264870 | 1.18E-01 | 2.82E-01 | 2.40E+00 | 5.14E+00 | CASP7;GSN;DBNL;SPTAN1;ADD1 |
| 598 | Vitamin B5 (pantothenate) metabolism Homo sapiens R-HSA-199220 | 1.18E-01 | 2.82E-01 | 2.40E+00 | 5.14E+00 | PANK2;PANK3;PPCS;AASDHPPT;COASY |
| 599 | Purine ribonucleoside monophosphate biosynthesis Homo sapiens R-HSA-73817 | 1.18E-01 | 2.82E-01 | 2.40E+00 | 5.14E+00 | ADSL;ATIC;GMPS;PAICS;LHPP |

| | | | | | | |
|---|---|---|---|---|---|---|
| 600 | PTK6 Regulates RHO GTPases, RAS GTPase and MAP kinases Homo sapiens R-HSA-8849471 | 1.20E-01 | 2.87E-01 | 1.78E+00 | 3.77E+00 | RASA1;DAB2IP;ELMO2;RASAL1;KRAS;RAC1;DOCK1;RHOA;BCAR1 |
| 601 | Constitutive Signaling by Ligand-Responsive EGFR Cancer Variants Homo sapiens R-HSA-1236382 | 1.23E-01 | 2.92E-01 | 1.96E+00 | 4.11E+00 | HSP90AA1;CDC37;UBC;KRAS;GRB2;CBL;RPS27A |
| 602 | Signaling by Ligand-Responsive EGFR Variants in Cancer Homo sapiens R-HSA-5637815 | 1.23E-01 | 2.92E-01 | 1.96E+00 | 4.11E+00 | HSP90AA1;CDC37;UBC;KRAS;GRB2;CBL;RPS27A |
| 603 | Signaling by EGFR in Cancer Homo sapiens R-HSA-1643713 | 1.23E-01 | 2.92E-01 | 1.96E+00 | 4.11E+00 | HSP90AA1;CDC37;UBC;KRAS;GRB2;CBL;RPS27A |
| 604 | Ephrin signaling Homo sapiens R-HSA-3928664 | 1.23E-01 | 2.92E-01 | 1.96E+00 | 4.11E+00 | SDCBP;PAK1;FYN;RAC1;ARHGEF7;PAK2;MYL12A |
| 605 | tRNA processing Homo sapiens R-HSA-72306 | 1.27E-01 | 3.00E-01 | 1.32E+00 | 2.73E+00 | TSEN34;POP5;TRMU;DDX1;URM1;RTCB;ZBTB8OS;C2ORF49;EPRS;MTO1;LAGE3;HSD17B10;TSEN54;TRMT112;CLP1;TPR;TYW1;RPP38;TRNT1;PUS7;NUP93;QTRT1;NUP50;TRMT6;RPP21;TRMT11;RPP25;TRMT61A;RAN |
| 606 | AKT phosphorylates targets in the nucleus Homo sapiens R-HSA-198693 | 1.29E-01 | 3.04E-01 | 2.69E+00 | 5.51E+00 | CREB1;AKT2;RPS6KB2;AKT1 |
| 607 | Small interfering RNA (siRNA) biogenesis Homo sapiens R-HSA-426486 | 1.29E-01 | 3.04E-01 | 2.69E+00 | 5.51E+00 | TSNAX;PRKRA;AGO2;TSN |

| | | | | | | |
|---|---|---|---|---|---|---|
| 608 | Inactivation of Cdc42 and Rac Homo sapiens R-HSA-428543 | 1.29E-01 | 3.04E-01 | 2.69E+00 | 5.51E+00 | CDC42;GPC1;RAC1;SRGAP3 |
| 609 | Pyrimidine salvage reactions Homo sapiens R-HSA-73614 | 1.29E-01 | 3.04E-01 | 2.69E+00 | 5.51E+00 | CDA;UCK1;TK2;TK1 |
| 610 | Transport of Mature mRNA derived from an Intron-Containing Transcript Homo sapiens R-HSA-159236 | 1.31E-01 | 3.06E-01 | 1.40E+00 | 2.86E+00 | FYTTD1;NXT1;NCBP1;RBM8A;POLDIP3;EIF4A3;THOC3;THOC2;THOC5;CDC40;NUP93;DDX39A;NUP50;SARNP;MAGOH;TPR;SRSF2;SRSF4;RNPS1;SRSF9 |
| 611 | NRAGE signals death through JNK Homo sapiens R-HSA-193648 | 1.31E-01 | 3.06E-01 | 1.52E+00 | 3.09E+00 | VAV3;ARHGEF12;MAGED1;BAD;RASGRF2;ARHGEF16;ARHGEF18;AATF;ARHGAP4;TIAM1;MCF2L;ARHGEF3;RAC1;ARHGEF7 |
| 612 | Negative epigenetic regulation of rRNA expression Homo sapiens R-HSA-5250941 | 1.34E-01 | 3.13E-01 | 1.37E+00 | 2.76E+00 | DNMT1;HDAC2;H2AFZ;HDAC1;H3F3A;MBD2;SMARCA5;ARID4B;SIRT1;GTF2H4;ZNRD1;SAP30L;TWISTNB;POLR1D;POLR2E;SAP18;SUDS3;POLR2H;MNAT1;HIST1H2AC;POLR2K;POLR2L |
| 613 | Regulation of insulin secretion Homo sapiens R-HSA-422356 | 1.35E-01 | 3.15E-01 | 1.36E+00 | 2.72E+00 | SLC2A1;ITPR2;ARL2;CACNA1D;AKAP5;CACNA1C;GNAI1;ADCY5;GNAI2;GNG10;RAP1A;GNG5;ARL2BP;PRKAR1A;PRKAR2B;PRKAR2A;GNAS;GNB5;SLC25A5;PLCB1;SLC25A4;RAPGEF3;SLC25A6 |
| 614 | RAF-independent MAPK1/3 activation Homo sapiens R-HSA-112409 | 1.36E-01 | 3.16E-01 | 1.79E+00 | 3.58E+00 | MAP2K2;DUSP1;PEA15;MAPK1;PTPN11;DUSP8;JAK2;JAK1 |
| 615 | Signaling by BMP Homo sapiens R-HSA-201451 | 1.36E-01 | 3.16E-01 | 1.79E+00 | 3.58E+00 | SMAD1;SMAD4;ZFYVE16;SMURF2;UBE2D3;UBE2D1;SMAD5;CHRDL1 |
| 616 | GPVI-mediated activation cascade Homo sapiens R-HSA-114604 | 1.38E-01 | 3.17E-01 | 1.46E+00 | 2.89E+00 | VAV3;PDPK1;RHOG;PIK3R3;PTPN11;PRKCZ;RHOA;CDC42;AKT2;AKT1;FYN;PTPN6;GRB2;RAC1;JAK2;JAK1 |

| | | | | | | |
|---|---|---|---|---|---|---|
| 617 | PTK6 Regulates Cell Cycle Homo sapiens R-HSA-8849470 | 1.38E-01 | 3.17E-01 | 3.36E+00 | 6.66E+00 | CDKN1B;CDK4;CDK2 |
| 618 | Scavenging by Class F Receptors Homo sapiens R-HSA-3000484 | 1.38E-01 | 3.17E-01 | 3.36E+00 | 6.66E+00 | HSP90AA1;HSPH1;CALR |
| 619 | p75NTR negatively regulates cell cycle via SC1 Homo sapiens R-HSA-193670 | 1.38E-01 | 3.17E-01 | 3.36E+00 | 6.66E+00 | HDAC2;HDAC3;HDAC1 |
| 620 | Choline catabolism Homo sapiens R-HSA-6798163 | 1.38E-01 | 3.17E-01 | 3.36E+00 | 6.66E+00 | DMGDH;CHDH;ALDH7A1 |
| 621 | mitochondrial fatty acid beta-oxidation of unsaturated fatty acids Homo sapiens R-HSA-77288 | 1.38E-01 | 3.17E-01 | 3.36E+00 | 6.66E+00 | HADHB;HADHA;ACADM |
| 622 | Beta-oxidation of very long chain fatty acids Homo sapiens R-HSA-390247 | 1.38E-01 | 3.17E-01 | 3.36E+00 | 6.66E+00 | ACOT8;HSD17B4;CRAT |
| 623 | Protein repair Homo sapiens R-HSA-5676934 | 1.38E-01 | 3.17E-01 | 3.36E+00 | 6.66E+00 | PCMT1;MSRA;MSRB2 |
| 624 | SUMO E3 ligases SUMOylate target proteins Homo sapiens R-HSA-3108232 | 1.38E-01 | 3.17E-01 | 1.32E+00 | 2.61E+00 | SP100;PCNA;CETN2;SMC3;HERC2;SUMO1;SUMO3;RAD21;TDG;SUMO2;TPR;PHC2;UBE2I;PARP1;CBX4;PCGF2;SMC1A;NUP93;RNF168;NSMCE4A;HNRNPK;STAG2;NUP50;SP3;MDM2;TOP1;HNRNPC |
| 625 | p75NTR signals via NF-kB Homo sapiens R-HSA-193639 | 1.39E-01 | 3.18E-01 | 2.02E+00 | 3.98E+00 | NFKBIA;IRAK1;UBC;RPS27A;SQSTM1;NFKB1 |

| | | | | | | |
|---|---|---|---|---|---|---|
| 626 | MAP2K and MAPK activation Homo sapiens R-HSA-5674135 | 1.42E-01 | 3.23E-01 | 1.55E+00 | 3.04E+00 | RAP1B;RAP1A;MAP2K2;YWHAB;PEBP1;MAPK1;WDR83;ARRB1;CSK;KRAS;TLN1;LAMTOR3 |
| 627 | Glutathione conjugation Homo sapiens R-HSA-156590 | 1.42E-01 | 3.23E-01 | 1.55E+00 | 3.04E+00 | GSTM4;GGCT;GSTM3;GSTK1;GSTO2;GSTO1;GSTP1;MGST3;ESD;MGST2;GSTT2B;GSTM5 |
| 628 | BMAL1:CLOCK, NPAS2 activates circadian gene expression Homo sapiens R-HSA-1368108 | 1.46E-01 | 3.33E-01 | 1.51E+00 | 2.90E+00 | NCOA2;SMARCD3;HDAC3;BHLHE41;RORA;NR1D1;HIF1A;ARNTL;CREB1;NCOR1;TGS1;CARM1;NAMPT |
| 629 | NoRC negatively regulates rRNA expression Homo sapiens R-HSA-427413 | 1.47E-01 | 3.35E-01 | 1.36E+00 | 2.60E+00 | DNMT1;HDAC2;H2AFZ;HDAC1;H3F3A;MBD2;SMARCA5;ARID4B;GTF2H4;ZNRD1;SAP30L;TWISTNB;POLR1D;POLR2E;SAP18;SUDS3;POLR2H;MNAT1;HIST1H2AC;POLR2K;POLR2L |
| 630 | Factors involved in megakaryocyte development and platelet production Homo sapiens R-HSA-983231 | 1.50E-01 | 3.40E-01 | 1.24E+00 | 2.36E+00 | HDAC2;DOCK9;HDAC1;AK3;KLC1;CDC42;CAPZB;KIF5C;KIF3A;PRKAR2B;KIF5B;PRKAR2A;MFN1;HMG20B;MFN2;KIF21A;RAC1;KIFAP3;JAK2;SH2B2;CBX5;H3F3A;ITPK1;RAD51B;CDK5;KIF2A;KIFC2;IRF3;PRKAR1A;MAFG;CAPZA1;KIF26B;CDK2;IRF2;CAPZA2;RAB5A;DOCK1;RCOR1 |
| 631 | GAB1 signalosome Homo sapiens R-HSA-180292 | 1.51E-01 | 3.42E-01 | 1.26E+00 | 2.38E+00 | PRR5;GSK3B;GSK3A;CDKN1B;MAPKAP1;PDGFB;PIK3R3;FGF2;PPP2CA;PPP2CB;PPP2R5E;AKT2;MLST8;AKT1;MAPK1;PIP5K1B;CSK;FYN;PIP4K2C;IER3;PDPK1;BAD;PPP2R5A;PTPN11;MTOR;BTC;CREB1;FGF18;AGO2;AKT1S1;RPS6KB2;MDM2;GRB2;HBEGF |
| 632 | PI3K/AKT activation Homo sapiens R-HSA-198203 | 1.51E-01 | 3.42E-01 | 1.26E+00 | 2.38E+00 | PRR5;GSK3B;GSK3A;CDKN1B;MAPKAP1;PDGFB;PIK3R3;FGF2;PPP2CA;PPP2CB;PPP2R5E;AKT2;MLST8;AKT1;MAPK1;PIP5K1B;FYN;PIP4K2C;IER3;PDPK1;BAD;PPP2R5A;PTPN11;RHOA;MTOR;BTC;CREB1;FGF18;AGO2;AKT1S1;RPS6KB2;MDM2;GRB2;HBEGF |
| 633 | Anchoring of the basal body to the plasma membrane Homo sapiens R-HSA-5620912 | 1.51E-01 | 3.42E-01 | 1.30E+00 | 2.45E+00 | YWHAE;CEP57;DCTN2;CETN2;SSNA1;DCTN3;ACTR1A;HAUS8;PRKAR2B;HAUS7;NPHP1;CC2D2A;YWHAG;RAB8A;OFD1;DYNC1H1;HSP90AA1;TUBB;TUBG1;TUBB4B;TUBA4A;RAB11A;HAUS1;AHI1;AKAP9;MAPRE1;PAFAH1B1 |

| | | | | | | |
|---|---|---|---|---|---|---|
| 634 | Regulation of actin dynamics for phagocytic cup formation Homo sapiens R-HSA-2029482 | 1.51E-01 | 3.42E-01 | 1.30E+00 | 2.45E+00 | CYFIP2;NCKAP1;HSP90AB1;ARPC1B;ARPC1A;BRK1;WASL;ACTG1;CDC42;PAK1;CFL1;MAPK1;RAC1;WASF2;ACTR3;VAV3;ACTR2;HSP90AA1;MYO5A;ARPC5;PTK2;ARPC2;ABI2;ARPC3;ELMO2;GRB2;DOCK1 |
| 635 | Negative regulation of FGFR4 signaling Homo sapiens R-HSA-5654733 | 1.53E-01 | 3.45E-01 | 1.60E+00 | 3.01E+00 | PPP2CA;PPP2CB;UBC;FGF18;MAPK1;PTPN11;GRB2;CBL;RPS27A;FGF2 |
| 636 | Syndecan interactions Homo sapiens R-HSA-3000170 | 1.54E-01 | 3.45E-01 | 1.81E+00 | 3.39E+00 | ITGB1;TGFB1;TRAPPC4;ACTN1;SDC2;SDC1;FGF2 |
| 637 | Signaling by Hippo Homo sapiens R-HSA-2028269 | 1.54E-01 | 3.45E-01 | 1.81E+00 | 3.39E+00 | YWHAE;TJP1;MOB1B;MOB1A;YWHAB;AMOTL2;STK3 |
| 638 | IRF3-mediated induction of type I IFN Homo sapiens R-HSA-3270619 | 1.57E-01 | 3.51E-01 | 2.10E+00 | 3.89E+00 | XRCC6;TBK1;IRF3;XRCC5;PRKDC |
| 639 | Downregulation of ERBB2:ERBB3 signaling Homo sapiens R-HSA-1358803 | 1.57E-01 | 3.51E-01 | 2.10E+00 | 3.89E+00 | USP8;AKT2;UBC;AKT1;RPS27A |
| 640 | Trafficking and processing of endosomal TLR Homo sapiens R-HSA-1679131 | 1.57E-01 | 3.51E-01 | 2.10E+00 | 3.89E+00 | UNC93B1;CTSL;CTSK;CNPY3;CTSB |
| 641 | Base Excision Repair Homo sapiens R-HSA-73884 | 1.59E-01 | 3.55E-01 | 1.54E+00 | 2.83E+00 | MBD4;PCNA;PARP1;RFC1;MPG;NTHL1;TDG;RPA3;POLD2;NEIL1;UNG |

| | | | | | | |
|---|---|---|---|---|---|---|
| 642 | Resolution of Abasic Sites (AP sites) Homo sapiens R-HSA-73933 | 1.59E-01 | 3.55E-01 | 1.54E+00 | 2.83E+00 | MBD4;PCNA;PARP1;RFC1;MPG;NTHL1;TDG;RPA3;POLD2;NEIL1;UNG |
| 643 | PI-3K cascade:FGFR1 Homo sapiens R-HSA-5654689 | 1.64E-01 | 3.61E-01 | 1.25E+00 | 2.26E+00 | PRR5;GSK3B;GSK3A;CDKN1B;MAPKAP1;PDGFB;PIK3R3;FGF2;PPP2CA;PPP2CB;PPP2R5E;AKT2;MLST8;AKT1;MAPK1;PIP5K1B;FYN;PIP4K2C;IER3;PDPK1;BAD;PPP2R5A;PTPN11;MTOR;BTC;CREB1;FGF18;AGO2;AKT1S1;RPS6KB2;MDM2;GRB2;HBEGF |
| 644 | PI-3K cascade:FGFR3 Homo sapiens R-HSA-5654710 | 1.64E-01 | 3.61E-01 | 1.25E+00 | 2.26E+00 | PRR5;GSK3B;GSK3A;CDKN1B;MAPKAP1;PDGFB;PIK3R3;FGF2;PPP2CA;PPP2CB;PPP2R5E;AKT2;MLST8;AKT1;MAPK1;PIP5K1B;FYN;PIP4K2C;IER3;PDPK1;BAD;PPP2R5A;PTPN11;MTOR;BTC;CREB1;FGF18;AGO2;AKT1S1;RPS6KB2;MDM2;GRB2;HBEGF |
| 645 | PI3K events in ERBB4 signaling Homo sapiens R-HSA-1250342 | 1.64E-01 | 3.61E-01 | 1.25E+00 | 2.26E+00 | PRR5;GSK3B;GSK3A;CDKN1B;MAPKAP1;PDGFB;PIK3R3;FGF2;PPP2CA;PPP2CB;PPP2R5E;AKT2;MLST8;AKT1;MAPK1;PIP5K1B;FYN;PIP4K2C;IER3;PDPK1;BAD;PPP2R5A;PTPN11;MTOR;BTC;CREB1;FGF18;AGO2;AKT1S1;RPS6KB2;MDM2;GRB2;HBEGF |
| 646 | PIP3 activates AKT signaling Homo sapiens R-HSA-1257604 | 1.64E-01 | 3.61E-01 | 1.25E+00 | 2.26E+00 | PRR5;GSK3B;GSK3A;CDKN1B;MAPKAP1;PDGFB;PIK3R3;FGF2;PPP2CA;PPP2CB;PPP2R5E;AKT2;MLST8;AKT1;MAPK1;PIP5K1B;FYN;PIP4K2C;IER3;PDPK1;BAD;PPP2R5A;PTPN11;MTOR;BTC;CREB1;FGF18;AGO2;AKT1S1;RPS6KB2;MDM2;GRB2;HBEGF |
| 647 | PI-3K cascade:FGFR4 Homo sapiens R-HSA-5654720 | 1.64E-01 | 3.61E-01 | 1.25E+00 | 2.26E+00 | PRR5;GSK3B;GSK3A;CDKN1B;MAPKAP1;PDGFB;PIK3R3;FGF2;PPP2CA;PPP2CB;PPP2R5E;AKT2;MLST8;AKT1;MAPK1;PIP5K1B;FYN;PIP4K2C;IER3;PDPK1;BAD;PPP2R5A;PTPN11;MTOR;BTC;CREB1;FGF18;AGO2;AKT1S1;RPS6KB2;MDM2;GRB2;HBEGF |
| 648 | PI-3K cascade:FGFR2 Homo sapiens R-HSA-5654695 | 1.64E-01 | 3.61E-01 | 1.25E+00 | 2.26E+00 | PRR5;GSK3B;GSK3A;CDKN1B;MAPKAP1;PDGFB;PIK3R3;FGF2;PPP2CA;PPP2CB;PPP2R5E;AKT2;MLST8;AKT1;MAPK1;PIP5K1B;FYN;PIP4K2C;IER3;PDPK1;BAD;PPP2R5A;PTPN11;MTOR;BTC;CREB1;FGF18;AGO2;AKT1S1;RPS6KB2;MDM2;GRB2;HBEGF |
| 649 | Nuclear signaling by ERBB4 Homo sapiens R-HSA-1251985 | 1.64E-01 | 3.61E-01 | 1.50E+00 | 2.70E+00 | APH1A;PSENEN;BTC;WWOX;APH1B;NCOR1;CUL1;PTPN11;JAK2;SKP1;RBX1;HBEGF |

| | | | | | | |
|---|---|---|---|---|---|---|
| 650 | Biosynthesis of the N-glycan precursor (dolichol lipid-linked oligosaccharide, LLO) and transfer to a nascent protein Homo sapiens R-HSA-446193 | 1.64E-01 | 3.61E-01 | 1.32E+00 | 2.39E+00 | DPAGT1;CTSA;SLC35A1;ALG8;CMAS;ALG5;GFPT1;ALG2;ALG3;DOLK;GMPPB;DPM2;DPM3;GLB1;NEU1;ST3GAL4;SLC17A5;GNPNAT1;ST3GAL6;NUDT14;ST6GALNAC6;MPDU1 |
| 651 | PI3K/AKT Signaling in Cancer Homo sapiens R-HSA-2219528 | 1.65E-01 | 3.63E-01 | 1.30E+00 | 2.35E+00 | PRR5;GSK3B;GSK3A;CDKN1B;PDPK1;BAD;MAPKAP1;PDGFB;PIK3R3;PTPN11;FGF2;MTOR;BTC;CREB1;AKT2;AKT1S1;FGF18;RPS6KB2;MLST8;MDM2;AKT1;FYN;GRB2;HBEGF |
| 652 | TP53 Regulates Transcription of Cell Death Genes Homo sapiens R-HSA-5633008 | 1.68E-01 | 3.68E-01 | 1.46E+00 | 2.60E+00 | BNIP3L;PRELID1;PPP1R13B;APAF1;CHM;RABGGTB;BBC3;TP53I3;BCL6;PERP;TMEM219;BAX;BID |
| 653 | Signaling by FGFR2 in disease Homo sapiens R-HSA-5655253 | 1.68E-01 | 3.68E-01 | 1.46E+00 | 2.60E+00 | NCBP1;FGF2;GTF2F2;POLR2B;POLR2C;FGF18;POLR2E;KRAS;GRB2;POLR2H;POLR2J;POLR2K;POLR2L |
| 654 | Recognition of DNA damage by PCNA-containing replication complex Homo sapiens R-HSA-110314 | 1.73E-01 | 3.78E-01 | 1.59E+00 | 2.79E+00 | DDB1;PCNA;RFC1;UBE2B;RPA3;UBC;POLD2;RPS27A;RBX1 |
| 655 | Ion homeostasis Homo sapiens R-HSA-5578775 | 1.74E-01 | 3.78E-01 | 1.40E+00 | 2.46E+00 | DMPK;ATP2A3;ITPR2;ATP1B3;ATP1A1;SRI;ATP2B1;ATP1B1;ASPH;FXYD3;FXYD2;FKBP1B;ORAI1;CALM1;CAMK2G |
| 656 | Nuclear Receptor transcription pathway Homo sapiens R-HSA-383280 | 1.74E-01 | 3.78E-01 | 1.40E+00 | 2.46E+00 | ESRRA;RARG;THRA;VDR;NR1H2;ESRRB;RORA;NR1D1;NR2C2;NR2C1;NRBP1;ESR2;NR2F6;RARB;NRBF2 |

| | | | | | | |
|---|---|---|---|---|---|---|
| 657 | Translesion synthesis by POLK Homo sapiens R-HSA-5655862 | 1.75E-01 | 3.80E-01 | 1.83E+00 | 3.20E+00 | PCNA;RFC1;RPA3;UBC;POLK;RPS27A |
| 658 | PKA-mediated phosphorylation of CREB Homo sapiens R-HSA-111931 | 1.75E-01 | 3.80E-01 | 1.83E+00 | 3.20E+00 | CREB1;PRKAR1A;PRKAR2B;PRKAR2A;ADCY3;ADCY5 |
| 659 | PKA activation in glucagon signalling Homo sapiens R-HSA-164378 | 1.75E-01 | 3.80E-01 | 1.83E+00 | 3.20E+00 | PRKAR1A;PRKAR2B;PRKAR2A;GNAS;ADCY3;ADCY5 |
| 660 | Regulated proteolysis of p75NTR Homo sapiens R-HSA-193692 | 1.78E-01 | 3.84E-01 | 2.24E+00 | 3.87E+00 | APH1A;PSENEN;APH1B;NFKB1 |
| 661 | MASTL Facilitates Mitotic Progression Homo sapiens R-HSA-2465910 | 1.78E-01 | 3.84E-01 | 2.24E+00 | 3.87E+00 | PPP2CA;PPP2CB;CCNB1;ENSA |
| 662 | Netrin mediated repulsion signals Homo sapiens R-HSA-418886 | 1.78E-01 | 3.84E-01 | 2.24E+00 | 3.87E+00 | PTPN11;FYN;NTN1;PTK2 |
| 663 | eNOS activation Homo sapiens R-HSA-203615 | 1.78E-01 | 3.84E-01 | 2.24E+00 | 3.87E+00 | LYPLA1;HSP90AA1;AKT1;CALM1 |
| 664 | SUMOylation of DNA damage response and repair proteins Homo sapiens R-HSA-3108214 | 1.80E-01 | 3.89E-01 | 1.31E+00 | 2.24E+00 | SP100;PHC2;UBE2I;PARP1;CBX4;CETN2;PCGF2;SMC3;SMC1A;NUP93;RNF168;HERC2;NSMCE4A;STAG2;SUMO1;SUMO3;NUP50;RAD21;TDG;SUMO2;TPR |

| | | | | | | |
|---|---|---|---|---|---|---|
| 665 | TP53 regulates transcription of additional cell cycle genes whose exact role in the p53 pathway remain uncertain Homo sapiens R-HSA-6804115 | 1.87E-01 | 4.02E-01 | 1.68E+00 | 2.82E+00 | CNOT4;NPM1;CNOT6L;CNOT7;CNOT11;CNOT2;CNOT3 |
| 666 | NOTCH2 Activation and Transmission of Signal to the Nucleus Homo sapiens R-HSA-2979096 | 1.87E-01 | 4.02E-01 | 1.68E+00 | 2.82E+00 | PSENEN;JAG2;APH1A;APH1B;UBC;MIB2;RPS27A |
| 667 | VxPx cargo-targeting to cilium Homo sapiens R-HSA-5620916 | 1.87E-01 | 4.02E-01 | 1.68E+00 | 2.82E+00 | ARF4;ASAP1;EXOC6;PKD2;PKD1;RAB11A;RAB8A |
| 668 | Synthesis of very long-chain fatty acyl-CoAs Homo sapiens R-HSA-75876 | 1.87E-01 | 4.02E-01 | 1.68E+00 | 2.82E+00 | ELOVL1;TECR;PPT1;ELOVL6;ACSL4;HSD17B12;PPT2 |
| 669 | Interleukin-1 signaling Homo sapiens R-HSA-446652 | 1.91E-01 | 4.09E-01 | 1.41E+00 | 2.34E+00 | FBXW11;IL1R2;CUL1;IRAK4;NFKB1;IRAK1;TNIP2;UBC;PELI1;UBE2V1;RPS27A;SQSTM1;SKP1 |
| 670 | ATF4 activates genes Homo sapiens R-HSA-380994 | 1.96E-01 | 4.19E-01 | 1.58E+00 | 2.58E+00 | EXOSC7;EXOSC4;EXOSC3;ATF6;EXOSC1;DCP2;HERPUD1;ATF4 |
| 671 | Basigin interactions Homo sapiens R-HSA-210991 | 1.96E-01 | 4.19E-01 | 1.58E+00 | 2.58E+00 | ITGB1;SLC7A6;SLC7A8;ATP1B3;ATP1B1;L1CAM;SLC16A3;PPIA |
| 672 | TRAF6 Mediated Induction of proinflammatory cytokines Homo sapiens R-HSA-168180 | 1.98E-01 | 4.22E-01 | 1.29E+00 | 2.09E+00 | ATF1;MAP2K3;APP;DUSP3;FBXW11;DHX9;CUL1;HMGB1;NFKB1;PPP2CA;NFKBIA;PPP2CB;MAPK9;CREB1;IRAK1;TNIP2;UBC;MAPK1;RPS27A;SKP1 |

| | | | | | | |
|---|---|---|---|---|---|---|
| 673 | Mismatch repair (MMR) directed by MSH2:MSH3 (MutSbeta) Homo sapiens R-HSA-5358606 | 2.00E-01 | 4.22E-01 | 1.87E+00 | 3.00E+00 | PCNA;MSH2;MSH3;RPA3;POLD2 |
| 674 | Mismatch repair (MMR) directed by MSH2:MSH6 (MutSalpha) Homo sapiens R-HSA-5358565 | 2.00E-01 | 4.22E-01 | 1.87E+00 | 3.00E+00 | MSH6;PCNA;MSH2;RPA3;POLD2 |
| 675 | SEMA3A-Plexin repulsion signaling by inhibiting Integrin adhesion Homo sapiens R-HSA-399955 | 2.00E-01 | 4.22E-01 | 1.87E+00 | 3.00E+00 | PLXNA2;FYN;RAC1;TLN1;PLXNA4 |
| 676 | Activation of Rac Homo sapiens R-HSA-428540 | 2.00E-01 | 4.22E-01 | 1.87E+00 | 3.00E+00 | PAK1;GPC1;PAK6;RAC1;PAK2 |
| 677 | Inhibition of adenylate cyclase pathway Homo sapiens R-HSA-997269 | 2.00E-01 | 4.22E-01 | 1.87E+00 | 3.00E+00 | GNAI3;ADCY3;GNAI1;ADCY5;GNAI2 |
| 678 | Adenylate cyclase inhibitory pathway Homo sapiens R-HSA-170670 | 2.00E-01 | 4.22E-01 | 1.87E+00 | 3.00E+00 | GNAI3;ADCY3;GNAI1;ADCY5;GNAI2 |
| 679 | Nicotinate metabolism Homo sapiens R-HSA-196807 | 2.00E-01 | 4.22E-01 | 1.87E+00 | 3.00E+00 | QPRT;NAMPT;NMNAT3;NADSYN1;NADK |
| 680 | Attachment of GPI anchor to uPAR Homo sapiens R-HSA-162791 | 2.02E-01 | 4.24E-01 | 2.52E+00 | 4.04E+00 | PIGT;PGAP1;PIGK |

| | | | | | | |
|---|---|---|---|---|---|---|
| 681 | Apoptotic factor-mediated response Homo sapiens R-HSA-111471 | 2.02E-01 | 4.24E-01 | 2.52E+00 | 4.04E+00 | CASP7;APAF1;CYCS |
| 682 | Sphingolipid de novo biosynthesis Homo sapiens R-HSA-1660661 | 2.08E-01 | 4.36E-01 | 1.46E+00 | 2.30E+00 | ALDH3A2;SGPL1;SPTLC1;VAPA;VAPB;OSBP;DEGS1;KDSR;CSNK1G2;CERS2 |
| 683 | Activation of SMO Homo sapiens R-HSA-5635838 | 2.15E-01 | 4.48E-01 | 1.68E+00 | 2.59E+00 | EVC2;EVC;CSNK1A1;KIF3A;BOC;ARRB1 |
| 684 | Activation of IRF3/IRF7 mediated by TBK1/IKK epsilon Homo sapiens R-HSA-936964 | 2.15E-01 | 4.48E-01 | 1.68E+00 | 2.59E+00 | TBK1;IRF3;UBC;PTPN11;RPS27A;TANK |
| 685 | Defects in vitamin and cofactor metabolism Homo sapiens R-HSA-3296482 | 2.15E-01 | 4.48E-01 | 1.68E+00 | 2.59E+00 | PC;MCCC1;PCCB;ACACA;MMADHC;CD320 |
| 686 | tRNA processing in the nucleus Homo sapiens R-HSA-6784531 | 2.18E-01 | 4.55E-01 | 1.31E+00 | 2.00E+00 | TSEN34;POP5;DDX1;RTCB;ZBTB8OS;C2ORF49;TRNT1;TSEN54;NUP93;CLP1;NUP50;TPR;RPP21;RPP25;RPP38;RAN |
| 687 | Cyclin A/B1 associated events during G2/M transition Homo sapiens R-HSA-69273 | 2.24E-01 | 4.66E-01 | 1.57E+00 | 2.35E+00 | PPP2CA;PPP2CB;CCNB1;CDK2;PPP2R2A;MNAT1;CDC25B |
| 688 | CaM pathway Homo sapiens R-HSA-111997 | 2.30E-01 | 4.77E-01 | 1.49E+00 | 2.20E+00 | CREB1;PRKAR2B;PRKAR1A;PDE1A;PRKAR2A;ADCY3;CALM1;ADCY5 |

| | | | | | | |
|---|---|---|---|---|---|---|
| 689 | Calmodulin induced events Homo sapiens R-HSA-111933 | 2.30E-01 | 4.77E-01 | 1.49E+00 | 2.20E+00 | CREB1;PRKAR2B;PRKAR1A;PDE1A;PRKAR2A;ADCY3;CALM1;ADCY5 |
| 690 | Signaling by NOTCH3 Homo sapiens R-HSA-1980148 | 2.31E-01 | 4.77E-01 | 1.92E+00 | 2.81E+00 | APH1A;PSENEN;JAG2;APH1B |
| 691 | Signaling by NOTCH4 Homo sapiens R-HSA-1980150 | 2.31E-01 | 4.77E-01 | 1.92E+00 | 2.81E+00 | APH1A;PSENEN;JAG2;APH1B |
| 692 | DSCAM interactions Homo sapiens R-HSA-376172 | 2.31E-01 | 4.77E-01 | 1.92E+00 | 2.81E+00 | PAK1;RAC1;NTN1;MAPK13 |
| 693 | Recruitment of NuMA to mitotic centrosomes Homo sapiens R-HSA-380320 | 2.31E-01 | 4.77E-01 | 1.92E+00 | 2.81E+00 | CCNB1;TUBGCP2;TUBGCP4;TUBG1 |
| 694 | Condensation of Prometaphase Chromosomes Homo sapiens R-HSA-2514853 | 2.31E-01 | 4.77E-01 | 1.92E+00 | 2.81E+00 | CCNB1;CSNK2A2;CSNK2B;SMC4 |
| 695 | RHO GTPases activate IQGAPs Homo sapiens R-HSA-5626467 | 2.31E-01 | 4.77E-01 | 1.92E+00 | 2.81E+00 | CDC42;CLIP1;RAC1;CALM1 |
| 696 | Cell junction organization Homo sapiens R-HSA-446728 | 2.34E-01 | 4.80E-01 | 1.23E+00 | 1.78E+00 | ITGB1;VASP;CD151;LAMB3;JUP;FBLIM1;CADM2;ACTN1;TESK1;ILK;CLDN1;CLDN10;CLDN4;PARD6B;CLDN3;PARD6A;CLDN12;CLDN7;CLDN19;CDH13;RSU1;FERMT2;LIMS1 |
| 697 | RNA Polymerase I Promoter Escape Homo sapiens R-HSA-73772 | 2.35E-01 | 4.80E-01 | 1.44E+00 | 2.09E+00 | ZNRD1;TWISTNB;POLR2E;POLR1D;POLR2H;MNAT1;GTF2H4;POLR2K;POLR2L |

| | | | | | | |
|---|---|---|---|---|---|---|
| 698 | Activated NOTCH1 Transmits Signal to the Nucleus Homo sapiens R-HSA-2122948 | 2.35E-01 | 4.80E-01 | 1.44E+00 | 2.09E+00 | APH1A;PSENEN;JAG2;ITCH;APH1B;UBC;MIB2;ARRB1;RPS27A |
| 699 | Tight junction interactions Homo sapiens R-HSA-420029 | 2.35E-01 | 4.80E-01 | 1.44E+00 | 2.09E+00 | CLDN10;PARD6B;CLDN4;PARD6A;CLDN3;CLDN12;CLDN7;CLDN19;CLDN1 |
| 700 | Negative regulation of FGFR2 signaling Homo sapiens R-HSA-5654727 | 2.37E-01 | 4.85E-01 | 1.40E+00 | 2.01E+00 | PPP2CA;PPP2CB;UBC;FGF18;MAPK1;PTPN11;GRB2;CBL;RPS27A;FGF2 |
| 701 | Signaling by Retinoic Acid Homo sapiens R-HSA-5362517 | 2.41E-01 | 4.89E-01 | 1.35E+00 | 1.92E+00 | ALDH1A3;RARG;PDHA1;RDH11;RDH10;PDK3;RARB;RDH14;PDHB;RDH13;PDK2;DHRS4 |
| 702 | Non-integrin membrane-ECM interactions Homo sapiens R-HSA-3000171 | 2.41E-01 | 4.89E-01 | 1.35E+00 | 1.92E+00 | ITGB1;TGFB1;LAMB3;LAMB2;SDC2;TRAPPC4;ACTN1;PDGFB;NTN4;SDC1;FGF2;HSPG2 |
| 703 | Transcriptional regulation of pluripotent stem cells Homo sapiens R-HSA-452723 | 2.41E-01 | 4.89E-01 | 1.35E+00 | 1.92E+00 | SMAD4;POLR2B;POLR2C;STAT3;POLR2E;POLR2H;FGF2;POLR2J;DKK1;TDGF1;POLR2K;POLR2L |
| 704 | Glucagon-like Peptide-1 (GLP1) regulates insulin secretion Homo sapiens R-HSA-381676 | 2.41E-01 | 4.89E-01 | 1.35E+00 | 1.92E+00 | GNG10;RAP1A;GNG5;PRKAR2B;PRKAR1A;PRKAR2A;GNAS;ITPR2;AKAP5;GNB5;RAPGEF3;ADCY5 |
| 705 | Fatty Acyl-CoA Biosynthesis Homo sapiens R-HSA-75105 | 2.41E-01 | 4.89E-01 | 1.35E+00 | 1.92E+00 | SLC25A1;ELOVL1;ACOT9;ACOT8;TECR;ACOT11;PPT1;ELOVL6;ACSL4;HSD17B12;PPT2;ACACA |

| | | | | | | |
|---|---|---|---|---|---|---|
| 706 | Cellular Senescence Homo sapiens R-HSA-2559583 | 2.47E-01 | 4.97E-01 | 1.15E+00 | 1.61E+00 | RB1;ANAPC15;CDKN1B;ANAPC16;UBE2D1;EHMT1;ETS1;ETS2;ANAPC11;MAPK9;RBBP4;CDC26;CDC27;UBC;HIST1H1E;MAPK1;IGFBP7;RBBP7;RPS27A;HIST1H2AC;MAP2K3;SUZ12;CDKN2D;PHC2;CBX4;H2AFZ;TINF2;H3F3A;STAT3;UBE2E1;HMGA1;NFKB1;H1F0;RAD50;CDK6;CDK4;CDK2;MDM2;TERF2IP;MAPKAPK5;ANAPC5 |
| 707 | Processing of Intronless Pre-mRNAs Homo sapiens R-HSA-77595 | 2.47E-01 | 4.97E-01 | 1.68E+00 | 2.35E+00 | NCBP1;FIP1L1;CLP1;CPSF3;CSTF1 |
| 708 | Prolactin receptor signaling Homo sapiens R-HSA-1170546 | 2.47E-01 | 4.97E-01 | 1.68E+00 | 2.35E+00 | CUL1;PTPN11;JAK2;RBX1;SKP1 |
| 709 | Signaling by EGFRvIII in Cancer Homo sapiens R-HSA-5637812 | 2.47E-01 | 4.97E-01 | 1.68E+00 | 2.35E+00 | HSP90AA1;CDC37;KRAS;GRB2;CBL |
| 710 | Constitutive Signaling by EGFRvIII Homo sapiens R-HSA-5637810 | 2.47E-01 | 4.97E-01 | 1.68E+00 | 2.35E+00 | HSP90AA1;CDC37;KRAS;GRB2;CBL |
| 711 | GRB2:SOS provides linkage to MAPK signaling for Integrins Homo sapiens R-HSA-354194 | 2.47E-01 | 4.97E-01 | 1.68E+00 | 2.35E+00 | RAP1B;RAP1A;GRB2;TLN1;PTK2 |
| 712 | p130Cas linkage to MAPK signaling for integrins Homo sapiens R-HSA-372708 | 2.47E-01 | 4.97E-01 | 1.68E+00 | 2.35E+00 | RAP1B;RAP1A;TLN1;PTK2;BCAR1 |

| | | | | | | |
|---|---|---|---|---|---|---|
| 713 | TP53 Regulates Transcription of Genes Involved in Cytochrome C Release Homo sapiens R-HSA-6803204 | 2.57E-01 | 5.14E-01 | 1.55E+00 | 2.11E+00 | BNIP3L;PRELID1;PPP1R13B;BAX;BID;BBC3 |
| 714 | Translesion Synthesis by POLH Homo sapiens R-HSA-110320 | 2.57E-01 | 5.14E-01 | 1.55E+00 | 2.11E+00 | VCP;PCNA;RFC1;RPA3;UBC;RPS27A |
| 715 | RNA Polymerase I Transcription Homo sapiens R-HSA-73864 | 2.57E-01 | 5.14E-01 | 1.22E+00 | 1.66E+00 | HDAC2;H2AFZ;HDAC1;CBX3;H3F3A;MBD2;CHD4;GTF2H4;KAT2B;ZNRD1;RBBP4;TWISTNB;POLR1D;POLR2E;RBBP7;POLR2H;MNAT1;MTA3;HIST1H2AC;POLR2K;POLR2L |
| 716 | G alpha (12/13) signalling events Homo sapiens R-HSA-416482 | 2.58E-01 | 5.17E-01 | 1.22E+00 | 1.66E+00 | VAV3;ARHGEF12;RASGRF2;ARHGEF16;RHOG;ARHGEF18;PIK3R3;RHOC;RHOA;ARHGAP4;CDC42;TIAM1;GNG10;GNG5;MCF2L;ARHGEF3;ARHGEF1;RAC1;GNB5;ARHGEF7 |
| 717 | Gap-filling DNA repair synthesis and ligation in GG-NER Homo sapiens R-HSA-5696397 | 2.62E-01 | 5.23E-01 | 1.47E+00 | 1.97E+00 | PCNA;RFC1;RPA3;UBC;POLD2;POLK;RPS27A |
| 718 | BBSome-mediated cargo-targeting to cilium Homo sapiens R-HSA-5620922 | 2.62E-01 | 5.23E-01 | 1.47E+00 | 1.97E+00 | CCT3;CCT2;MKKS;TCP1;CCT8;CCT5;CCT4 |
| 719 | Synthesis of substrates in N-glycan biosythesis Homo sapiens R-HSA-446219 | 2.63E-01 | 5.24E-01 | 1.24E+00 | 1.66E+00 | CTSA;SLC35A1;CMAS;ALG5;GFPT1;DOLK;GMPPB;DPM2;DPM3;GLB1;NEU1;ST3GAL4;SLC17A5;GNPNAT1;ST3GAL6;NUDT14;ST6GALNAC6 |
| 720 | Synthesis of bile acids and bile salts Homo sapiens R-HSA-192105 | 2.66E-01 | 5.28E-01 | 1.42E+00 | 1.88E+00 | CYP39A1;NCOA2;ACOT8;SCP2;AKR1C1;HSD17B4;AKR1C2;SLC27A5 |

| | | | | | | |
|---|---|---|---|---|---|---|
| 721 | Cytokine Signaling in Immune system Homo sapiens R-HSA-1280215 | 2.68E-01 | 5.31E-01 | 1.07E+00 | 1.40E+00 | EIF4A2;EIF4A1;IFITM2;EIF4A3;PEBP1;WDR83;UBE2L6;TNF;FGF2;CRKL;PSMD8;PSMD9;PSMD6;PSMD7;PSMD4;PSMD2;PSMD3;PSMD1;KPNA4;KPNA2;KPNA3;KPNA1;SKP1;IFNAR2;MAP2K2;FBXW11;IL1R2;UBE2E1;HLA-C;HLA-G;RBX1;HLA-E;NUP93;BTC;TXLNA;AKAP9;PSME1;PSME2;UBE2V1;TLN1;SQSTM1;IFNAR1;PSMD10;SHC2;SPTBN5;PSMD12;PSMD11;PSMD14;PSMD13;CUL3;PDGFB;CUL1;PIK3R3;RASAL1;PHB;RASGRP3;RAP1B;RAP1A;SOCS1;IRAK1;TPR;UBC;PSMF1;IP6K2;YES1;TNFSF12;EIF2AK2;NFKB1;PTK2;ISG20;PTPRA;FGF18;PIN1;GRB2;CALM1;EIF4G3;EIF4G2;EIF4G1;UBE2M;EIF4E3;YWHAB;RASGRF2;ARRB1;SUMO1;TRIM5;PPP2R5E;TRIM2;PEA15;JAK2;SPTAN1;JAK1;TNFRSF12A;DUSP1;IFNGR2;PPP2R5A;IRAK4;DUSP8;YWHAZ;IL17RB;PSMA5;PSMA6;PSMA3;PSMA4;PSMA1;IRF3;PSMA2;RASA1;NUP50;IRF2;RAPGEF1;PELI1;IRF5;HLA-DQB1;HBEGF;SP100;CBL;PSMA7;PSMB6;PPP2CA;PSMB7;PPP2CB;PSMB5;PSMB2;PSMB3;PSMB1;MAPK1;STX4;CSK;FYN;RPS27A;EIF4E;CAMK2G;IL10RB;STAT3;DAB2IP;PTPN11;PSMB8;PSMC5;PSMC6;PSMC3;PSMC4;TNIP2;PSMC1;NEDD4;PSMC2;PTPN6;KRAS;LAMTOR3;IL18R1 |
| 722 | Disassembly of the destruction complex and recruitment of AXIN to the membrane Homo sapiens R-HSA-4641262 | 2.68E-01 | 5.31E-01 | 1.38E+00 | 1.81E+00 | PPP2CA;GSK3B;PPP2CB;FRAT1;PPP2R5E;CSNK1A1;DVL3;PPP2R5A;CSNK1G2 |
| 723 | Synthesis of diphthamide-EEF2 Homo sapiens R-HSA-5358493 | 2.71E-01 | 5.31E-01 | 2.02E+00 | 2.64E+00 | DPH3;DPH7;EEF2 |
| 724 | HuR (ELAVL1) binds and stabilizes mRNA Homo sapiens R-HSA-450520 | 2.71E-01 | 5.31E-01 | 2.02E+00 | 2.64E+00 | SET;ANP32A;ELAVL1 |

| | | | | | | |
|---|---|---|---|---|---|---|
| 725 | BH3-only proteins associate with and inactivate anti-apoptotic BCL-2 members Homo sapiens R-HSA-111453 | 2.71E-01 | 5.31E-01 | 2.02E+00 | 2.64E+00 | BAD;BID;BBC3 |
| 726 | Nef and signal transduction Homo sapiens R-HSA-164944 | 2.71E-01 | 5.31E-01 | 2.02E+00 | 2.64E+00 | FYN;RAC1;PAK2 |
| 727 | Sema4D mediated inhibition of cell attachment and migration Homo sapiens R-HSA-416550 | 2.71E-01 | 5.31E-01 | 2.02E+00 | 2.64E+00 | SEMA4D;RAC1;RHOA |
| 728 | Axonal growth inhibition (RHOA activation) Homo sapiens R-HSA-193634 | 2.71E-01 | 5.31E-01 | 2.02E+00 | 2.64E+00 | ARHGEF1;RHOA;RTN4 |
| 729 | Signaling by TGF-beta Receptor Complex in Cancer Homo sapiens R-HSA-3304351 | 2.71E-01 | 5.31E-01 | 2.02E+00 | 2.64E+00 | FKBP1A;SMAD4;TGFB1 |
| 730 | tRNA modification in the mitochondrion Homo sapiens R-HSA-6787450 | 2.71E-01 | 5.31E-01 | 2.02E+00 | 2.64E+00 | TRMU;MTO1;HSD17B10 |
| 731 | RNA Polymerase I Promoter Clearance Homo sapiens R-HSA-73854 | 2.80E-01 | 5.48E-01 | 1.20E+00 | 1.53E+00 | HDAC2;H2AFZ;HDAC1;H3F3A;MBD2;CHD4;GTF2H4;KAT2B;ZNRD1;RBBP4;TWISTNB;POLR1D;POLR2E;RBBP7;POLR2H;MNAT1;MTA3;HIST1H2AC;POLR2K;POLR2L |

| | | | | | | |
|---|---|---|---|---|---|---|
| 732 | Rho GTPase cycle Homo sapiens R-HSA-194840 | 2.88E-01 | 5.60E-01 | 1.15E+00 | 1.43E+00 | FAM13B;GDI1;RASGRF2;GDI2;ARHGAP12;ARHGAP4;CDC42;SYDE1;RHOT2;RHOT1;OPHN1;MCF2L;SRGAP3;RAC1;VAV3;RALBP1;STARD13;ARHGEF12;ARHGEF16;RHOG;ARHGAP29;ARHGEF18;RHOF;RHOC;RHOD;RHOA;TIAM1;ARHGEF3;TRIP10;ARHGEF7;RHOQ |
| 733 | ERKs are inactivated Homo sapiens R-HSA-202670 | 2.88E-01 | 5.60E-01 | 1.68E+00 | 2.09E+00 | PPP2CA;PPP2CB;DUSP3;MAPK1 |
| 734 | PECAM1 interactions Homo sapiens R-HSA-210990 | 2.88E-01 | 5.60E-01 | 1.68E+00 | 2.09E+00 | YES1;PTPN11;FYN;PTPN6 |
| 735 | Reduction of cytosolic Ca++ levels Homo sapiens R-HSA-418359 | 2.88E-01 | 5.60E-01 | 1.68E+00 | 2.09E+00 | ATP2A3;SRI;ATP2B1;CALM1 |
| 736 | The NLRP3 inflammasome Homo sapiens R-HSA-844456 | 2.88E-01 | 5.60E-01 | 1.68E+00 | 2.09E+00 | PYCARD;APP;HSP90AB1;SUGT1 |
| 737 | MAP kinase activation in TLR cascade Homo sapiens R-HSA-450294 | 2.89E-01 | 5.62E-01 | 1.22E+00 | 1.52E+00 | ATF1;MAP2K3;DUSP3;FBXW11;CUL1;NFKB1;PPP2CA;PPP2CB;MAPK9;CREB1;IRAK1;TNIP2;UBC;MAPK1;RPS27A;SKP1 |
| 738 | Nonhomologous End-Joining (NHEJ) Homo sapiens R-HSA-5693571 | 2.94E-01 | 5.70E-01 | 1.24E+00 | 1.52E+00 | XRCC6;PAXIP1;PRKDC;XRCC5;POLM;LIG4;BRCC3;BABAM1;RNF168;RAD50;HERC2;SUMO1;UIMC1;UBE2V2 |
| 739 | Synthesis of glycosylphosphatidylinositol (GPI) Homo sapiens R-HSA-162710 | 2.97E-01 | 5.72E-01 | 1.53E+00 | 1.86E+00 | DPM2;PIGP;PIGF;PIGX;PIGH |
| 740 | STING mediated induction of host immune responses Homo sapiens R-HSA-1834941 | 2.97E-01 | 5.72E-01 | 1.53E+00 | 1.86E+00 | XRCC6;TBK1;IRF3;XRCC5;PRKDC |

| | | | | | | |
|---|---|---|---|---|---|---|
| 741 | Translesion synthesis by REV1 Homo sapiens R-HSA-110312 | 2.97E-01 | 5.72E-01 | 1.53E+00 | 1.86E+00 | PCNA;RFC1;RPA3;UBC;RPS27A |
| 742 | PKA activation Homo sapiens R-HSA-163615 | 2.97E-01 | 5.72E-01 | 1.53E+00 | 1.86E+00 | PRKAR1A;PRKAR2B;PRKAR2A;ADCY3;ADCY5 |
| 743 | Inositol phosphate metabolism Homo sapiens R-HSA-1483249 | 2.98E-01 | 5.74E-01 | 1.26E+00 | 1.53E+00 | ITPKB;MINPP1;ISYNA1;PLCB4;IMPA1;INPP1;PPIP5K2;IPMK;ITPK1;PLCB1;CALM1;IP6K2 |
| 744 | Synthesis, secretion, and inactivation of Glucagon-like Peptide-1 (GLP-1) Homo sapiens R-HSA-381771 | 3.01E-01 | 5.76E-01 | 1.44E+00 | 1.73E+00 | DPP4;SPCS3;SPCS2;SPCS1;SEC11A;SEC11C |
| 745 | RNA Polymerase I Transcription Termination Homo sapiens R-HSA-73863 | 3.02E-01 | 5.76E-01 | 1.32E+00 | 1.57E+00 | ZNRD1;TWISTNB;POLR2E;POLR1D;POLR2H;MNAT1;GTF2H4;POLR2K;POLR2L |
| 746 | Signaling by NOTCH2 Homo sapiens R-HSA-1980145 | 3.02E-01 | 5.76E-01 | 1.32E+00 | 1.57E+00 | APH1A;PSENEN;JAG2;APH1B;CREB1;UBC;MIB2;RBPJ;RPS27A |
| 747 | Oncogene Induced Senescence Homo sapiens R-HSA-2559585 | 3.02E-01 | 5.76E-01 | 1.32E+00 | 1.57E+00 | CDKN2D;CDK6;CDK4;UBC;MDM2;MAPK1;RPS27A;ETS1;ETS2 |
| 748 | SMAD2/SMAD3:SMAD4 heterotrimer regulates transcription Homo sapiens R-HSA-2173796 | 3.02E-01 | 5.76E-01 | 1.32E+00 | 1.57E+00 | CDK9;SMAD4;SNW1;TFDP2;HDAC1;UBC;CCNC;E2F4;RPS27A |

| | | | | | | |
|---|---|---|---|---|---|---|
| 749 | Negative regulation of FGFR1 signaling Homo sapiens R-HSA-5654726 | 3.02E-01 | 5.76E-01 | 1.32E+00 | 1.57E+00 | PPP2CA;PPP2CB;UBC;MAPK1;PTPN11;GRB2;CBL;RPS27A;FGF2 |
| 750 | DAG and IP3 signaling Homo sapiens R-HSA-1489509 | 3.02E-01 | 5.76E-01 | 1.32E+00 | 1.57E+00 | CREB1;PRKAR2B;PRKAR1A;PDE1A;PRKAR2A;ITPR2;ADCY3;CALM1;ADCY5 |
| 751 | TRAF6 mediated NF-kB activation Homo sapiens R-HSA-933542 | 3.03E-01 | 5.76E-01 | 1.38E+00 | 1.66E+00 | NFKBIA;APP;MAVS;MAP3K1;DHX9;HMGB1;NFKB1 |
| 752 | Synthesis of bile acids and bile salts via 7alpha-hydroxycholesterol Homo sapiens R-HSA-193368 | 3.03E-01 | 5.76E-01 | 1.38E+00 | 1.66E+00 | NCOA2;ACOT8;SCP2;AKR1C1;AKR1C2;HSD17B4;SLC27A5 |
| 753 | Adrenaline,noradrenaline inhibits insulin secretion Homo sapiens R-HSA-400042 | 3.03E-01 | 5.76E-01 | 1.35E+00 | 1.61E+00 | GNG10;GNG5;CACNA1D;CACNA1C;GNB5;GNAI1;ADCY5;GNAI2 |
| 754 | TP53 Regulates Transcription of DNA Repair Genes Homo sapiens R-HSA-6796648 | 3.15E-01 | 5.97E-01 | 1.20E+00 | 1.38E+00 | SUPT16H;SUPT4H1;GTF2F2;GTF2H4;CDK9;MSH2;POLR2B;POLR2C;POLR2E;TCEA1;NELFCD;POLR2H;MNAT1;POLR2J;POLR2K;POLR2L |
| 755 | Mitotic Prophase Homo sapiens R-HSA-68875 | 3.19E-01 | 6.05E-01 | 1.14E+00 | 1.30E+00 | RB1;SET;RAB1B;USO1;SMC4;GOLGA2;PPP2CA;PPP2CB;CCNB1;TPR;ENSA;LMNA;MAPK1;HIST1H2AC;TMPO;RAB2A;H2AFZ;NEK7;H3F3A;LEMD2;NUP93;GORASP2;CNEP1R1;NUP50;NCAPD3;LPIN2;MCPH1 |
| 756 | Catabolism of glucuronate to xylulose-5-phosphate Homo sapiens R-HSA-5661270 | 3.24E-01 | 6.05E-01 | 2.24E+00 | 2.53E+00 | DCXR;CRYL1 |

| | | | | | | |
|---|---|---|---|---|---|---|
| 757 | Disinhibition of SNARE formation Homo sapiens R-HSA-114516 | 3.24E-01 | 6.05E-01 | 2.24E+00 | 2.53E+00 | STXBP3;STX4 |
| 758 | N-glycan trimming and elongation in the cis-Golgi Homo sapiens R-HSA-964739 | 3.24E-01 | 6.05E-01 | 2.24E+00 | 2.53E+00 | MAN1C1;MGAT1 |
| 759 | PTK6 Regulates Proteins Involved in RNA Processing Homo sapiens R-HSA-8849468 | 3.24E-01 | 6.05E-01 | 2.24E+00 | 2.53E+00 | KHDRBS1;SFPQ |
| 760 | Activation of RAS in B cells Homo sapiens R-HSA-1169092 | 3.24E-01 | 6.05E-01 | 2.24E+00 | 2.53E+00 | KRAS;RASGRP3 |
| 761 | LRR FLII-interacting protein 1 (LRRFIP1) activates type I IFN production Homo sapiens R-HSA-3134973 | 3.24E-01 | 6.05E-01 | 2.24E+00 | 2.53E+00 | IRF3;LRRFIP1 |
| 762 | NOSTRIN mediated eNOS trafficking Homo sapiens R-HSA-203641 | 3.24E-01 | 6.05E-01 | 2.24E+00 | 2.53E+00 | WASL;DNM2 |
| 763 | Alpha-oxidation of phytanate Homo sapiens R-HSA-389599 | 3.24E-01 | 6.05E-01 | 2.24E+00 | 2.53E+00 | ALDH3A2;HACL1 |

| | | | | | | |
|---|---|---|---|---|---|---|
| 764 | tRNA processing in the mitochondrion Homo sapiens R-HSA-6785470 | 3.24E-01 | 6.05E-01 | 2.24E+00 | 2.53E+00 | TRNT1;HSD17B10 |
| 765 | Galactose catabolism Homo sapiens R-HSA-70370 | 3.24E-01 | 6.05E-01 | 2.24E+00 | 2.53E+00 | PGM1;GALK1 |
| 766 | Fcgamma receptor (FCGR) dependent phagocytosis Homo sapiens R-HSA-2029480 | 3.27E-01 | 6.11E-01 | 1.12E+00 | 1.25E+00 | CYFIP2;NCKAP1;HSP90AB1;ARPC1B;ARPC1A;ITPR2;BRK1;WASL;ACTG1;CDC42;PAK1;CFL1;MAPK1;FYN;RAC1;WASF2;ACTR3;VAV3;ACTR2;HSP90AA1;YES1;MYO5A;ARPC5;PTK2;ARPC2;ABI2;ARPC3;ELMO2;GRB2;DOCK1 |
| 767 | RMTs methylate histone arginines Homo sapiens R-HSA-3214858 | 3.28E-01 | 6.12E-01 | 1.22E+00 | 1.36E+00 | COPRS;SMARCD2;SMARCD3;SMARCB1;H2AFZ;PRMT1;CDK4;CARM1;ACTL6A;RBBP7;JAK2;HIST1H2AC |
| 768 | Sphingolipid metabolism Homo sapiens R-HSA-428157 | 3.28E-01 | 6.12E-01 | 1.16E+00 | 1.29E+00 | GLTP;CTSA;ASAH1;HEXA;ALDH3A2;SGPL1;SPTLC1;VAPA;GLB1;VAPB;CPTP;ARSJ;OSBP;NEU1;DEGS1;KDSR;GLA;CSNK1G2;CERS2 |
| 769 | Regulation of lipid metabolism by Peroxisome proliferator-activated receptor alpha (PPARalpha) Homo sapiens R-HSA-400206 | 3.31E-01 | 6.16E-01 | 1.12E+00 | 1.24E+00 | ALAS1;SMARCD3;HDAC3;CCNC;RORA;ARNTL;MED11;MED30;MED10;TGS1;ME1;ACADM;TEAD2;FDFT1;ESRRA;NCOA2;NCOA3;TXNRD1;MED8;NR1D1;MED4;MED27;SREBF2;AGT;NCOR2;MED24;NCOR1;MED21;CARM1 |
| 770 | Elastic fibre formation Homo sapiens R-HSA-1566948 | 3.31E-01 | 6.16E-01 | 1.23E+00 | 1.36E+00 | FBN2;ITGB1;TGFB2;TGFB1;MFAP1;ITGB8;FURIN;LTBP3;LTBP1;FBLN5;LOXL2 |

| | | | | | | |
|---|---|---|---|---|---|---|
| 771 | Platelet Aggregation (Plug Formation) Homo sapiens R-HSA-76009 | 3.35E-01 | 6.21E-01 | 1.25E+00 | 1.36E+00 | RAP1B;RAP1A;PDPK1;AKT1;CSK;GRB2;TLN1;PTK2;RAPGEF3;BCAR1 |
| 772 | Apoptotic cleavage of cellular proteins Homo sapiens R-HSA-111465 | 3.35E-01 | 6.21E-01 | 1.25E+00 | 1.36E+00 | TJP1;CASP7;GSN;DBNL;STK24;LMNA;ACIN1;SPTAN1;ADD1;PTK2 |
| 773 | Sialic acid metabolism Homo sapiens R-HSA-4085001 | 3.38E-01 | 6.24E-01 | 1.26E+00 | 1.37E+00 | CTSA;SLC35A1;CMAS;GLB1;NEU1;ST3GAL4;SLC17A5;ST3GAL6;ST6GALNAC6 |
| 774 | PIWI-interacting RNA (piRNA) biogenesis Homo sapiens R-HSA-5601884 | 3.41E-01 | 6.24E-01 | 1.28E+00 | 1.38E+00 | HSP90AA1;POLR2B;POLR2C;POLR2E;POLR2H;POLR2J;POLR2K;POLR2L |
| 775 | Transport of nucleotide sugars Homo sapiens R-HSA-727802 | 3.41E-01 | 6.24E-01 | 1.68E+00 | 1.81E+00 | SLC35B3;SLC35A1;SLC35A3 |
| 776 | A third proteolytic cleavage releases NICD Homo sapiens R-HSA-157212 | 3.41E-01 | 6.24E-01 | 1.68E+00 | 1.81E+00 | APH1A;PSENEN;APH1B |
| 777 | EGFR Transactivation by Gastrin Homo sapiens R-HSA-2179392 | 3.41E-01 | 6.24E-01 | 1.68E+00 | 1.81E+00 | KRAS;GRB2;HBEGF |
| 778 | p75NTR regulates axonogenesis Homo sapiens R-HSA-193697 | 3.41E-01 | 6.24E-01 | 1.68E+00 | 1.81E+00 | ARHGEF1;RHOA;RTN4 |

| | | | | | | |
|---|---|---|---|---|---|---|
| 779 | Mitochondrial iron-sulfur cluster biogenesis Homo sapiens R-HSA-1362409 | 3.41E-01 | 6.24E-01 | 1.68E+00 | 1.81E+00 | SLC25A28;FDX1;ISCU |
| 780 | Pentose phosphate pathway (hexose monophosphate shunt) Homo sapiens R-HSA-71336 | 3.41E-01 | 6.24E-01 | 1.68E+00 | 1.81E+00 | TALDO1;PGLS;DERA |
| 781 | G0 and Early G1 Homo sapiens R-HSA-1538133 | 3.44E-01 | 6.24E-01 | 1.31E+00 | 1.40E+00 | PCNA;LIN37;RBBP4;TFDP2;HDAC1;CDK2;E2F4 |
| 782 | B-WICH complex positively regulates rRNA expression Homo sapiens R-HSA-5250924 | 3.45E-01 | 6.24E-01 | 1.17E+00 | 1.25E+00 | GSK3B;H2AFZ;H3F3A;SMARCA5;DDX21;DEK;KAT2B;ZNRD1;TWISTNB;POLR1D;POLR2E;POLR2H;HIST1H2AC;POLR2K;POLR2L |
| 783 | Facilitative Na+-independent glucose transporters Homo sapiens R-HSA-428790 | 3.47E-01 | 6.24E-01 | 1.49E+00 | 1.58E+00 | SLC2A9;SLC2A11;SLC2A1;SLC2A6 |
| 784 | Cytosolic iron-sulfur cluster assembly Homo sapiens R-HSA-2564830 | 3.47E-01 | 6.24E-01 | 1.49E+00 | 1.58E+00 | ABCB7;MMS19;CIAO1;NUBP2 |
| 785 | Dissolution of Fibrin Clot Homo sapiens R-HSA-75205 | 3.47E-01 | 6.24E-01 | 1.49E+00 | 1.58E+00 | ANXA2;HRG;SERPINB6;S100A10 |
| 786 | Activation of DNA fragmentation factor Homo sapiens R-HSA-211227 | 3.47E-01 | 6.24E-01 | 1.49E+00 | 1.58E+00 | H1F0;HIST1H1E;HMGB1;KPNA1 |

| | | | | | | |
|---|---|---|---|---|---|---|
| 787 | Apoptosis induced DNA fragmentation Homo sapiens R-HSA-140342 | 3.47E-01 | 6.24E-01 | 1.49E+00 | 1.58E+00 | H1F0;HIST1H1E;HMGB1;KPNA1 |
| 788 | p75NTR recruits signalling complexes Homo sapiens R-HSA-209543 | 3.47E-01 | 6.24E-01 | 1.49E+00 | 1.58E+00 | IRAK1;UBC;RPS27A;SQSTM1 |
| 789 | p38MAPK events Homo sapiens R-HSA-171007 | 3.47E-01 | 6.24E-01 | 1.49E+00 | 1.58E+00 | RALA;RALB;KRAS;MAPK13 |
| 790 | DEx/H-box helicases activate type I IFN and inflammatory cytokines production Homo sapiens R-HSA-3134963 | 3.47E-01 | 6.24E-01 | 1.49E+00 | 1.58E+00 | APP;DHX9;HMGB1;NFKB1 |
| 791 | TNFR1-induced proapoptotic signaling Homo sapiens R-HSA-5357786 | 3.47E-01 | 6.24E-01 | 1.49E+00 | 1.58E+00 | TRADD;USP2;FADD;TNF |
| 792 | Synthesis of PE Homo sapiens R-HSA-1483213 | 3.47E-01 | 6.24E-01 | 1.49E+00 | 1.58E+00 | ETNK2;ETNK1;LPIN2;PISD |
| 793 | alpha-linolenic (omega3) and linoleic (omega6) acid metabolism Homo sapiens R-HSA-2046104 | 3.47E-01 | 6.24E-01 | 1.49E+00 | 1.58E+00 | ELOVL1;ACOT8;SCP2;HSD17B4 |
| 794 | alpha-linolenic acid (ALA) metabolism Homo sapiens R-HSA-2046106 | 3.47E-01 | 6.24E-01 | 1.49E+00 | 1.58E+00 | ELOVL1;ACOT8;SCP2;HSD17B4 |

| | | | | | | |
|---|---|---|---|---|---|---|
| 795 | Translesion synthesis by POLI Homo sapiens R-HSA-5656121 | 3.47E-01 | 6.24E-01 | 1.40E+00 | 1.48E+00 | PCNA;RFC1;RPA3;UBC;RPS27A |
| 796 | Synthesis, secretion, and deacylation of Ghrelin Homo sapiens R-HSA-422085 | 3.47E-01 | 6.24E-01 | 1.40E+00 | 1.48E+00 | SPCS3;SPCS2;SPCS1;SEC11A;SEC11C |
| 797 | Chylomicron-mediated lipid transport Homo sapiens R-HSA-174800 | 3.47E-01 | 6.24E-01 | 1.40E+00 | 1.48E+00 | SAR1B;SDC1;P4HB;LDLRAP1;HSPG2 |
| 798 | L1CAM interactions Homo sapiens R-HSA-373760 | 3.52E-01 | 6.31E-01 | 1.12E+00 | 1.17E+00 | ITGB1;HSPA8;SPTBN5;MAP2K2;CSNK2A2;CLTA;RANBP9;ANK2;AP2B1;L1CAM;AP2A2;DNM2;SDCBP;PAK1;DLG1;DPYSL2;CSNK2B;AP2S1;MAPK1;RAC1;EZR;CD24;SPTAN1;AP2M1 |
| 799 | PPARA activates gene expression Homo sapiens R-HSA-1989781 | 3.54E-01 | 6.34E-01 | 1.11E+00 | 1.15E+00 | ALAS1;SMARCD3;HDAC3;CCNC;RORA;ARNTL;MED11;MED30;MED10;TGS1;ME1;ACADM;TEAD2;FDFT1;ESRRA;NCOA2;NCOA3;TXNRD1;MED8;NR1D1;MED4;MED27;SREBF2;AGT;MED24;NCOR1;MED21;CARM1 |
| 800 | DNA Damage Bypass Homo sapiens R-HSA-73893 | 3.59E-01 | 6.42E-01 | 1.19E+00 | 1.22E+00 | DDB1;VCP;PCNA;RFC1;UBE2B;RPA3;UBC;POLD2;UBE2L6;POLK;RPS27A;RBX1 |
| 801 | Viral Messenger RNA Synthesis Homo sapiens R-HSA-168325 | 3.64E-01 | 6.48E-01 | 1.19E+00 | 1.21E+00 | NUP93;POLR2B;POLR2C;NUP50;TPR;POLR2E;POLR2H;POLR2J;GTF2F2;POLR2K;POLR2L |
| 802 | Deactivation of the beta-catenin transactivating complex Homo sapiens R-HSA-3769402 | 3.64E-01 | 6.48E-01 | 1.19E+00 | 1.21E+00 | PYGO1;TCF7L1;HDAC1;AKT2;UBC;CTNNBIP1;CBY1;AKT1;ASH2L;RPS27A;YWHAZ |
| 803 | Netrin-1 signaling Homo sapiens R-HSA-373752 | 3.64E-01 | 6.48E-01 | 1.19E+00 | 1.21E+00 | CDC42;PITPNA;NTN4;FYN;PTPN11;WASL;RAC1;EZR;NTN1;DOCK1;PTK2 |

| | | | | | | |
|---|---|---|---|---|---|---|
| 804 | Signaling by WNT in cancer Homo sapiens R-HSA-4791275 | 3.74E-01 | 6.64E-01 | 1.21E+00 | 1.19E+00 | PPP2CA;GSK3B;PPP2CB;RNF43;TNKS2;PPP2R5E;CSNK1A1;PPP2R5A;DKK1 |
| 805 | EGFR interacts with phospholipase C-gamma Homo sapiens R-HSA-212718 | 3.74E-01 | 6.64E-01 | 1.21E+00 | 1.19E+00 | CREB1;PRKAR2B;PRKAR1A;PDE1A;PRKAR2A;ITPR2;ADCY3;CALM1;ADCY5 |
| 806 | PLC-gamma1 signalling Homo sapiens R-HSA-167021 | 3.74E-01 | 6.64E-01 | 1.21E+00 | 1.19E+00 | CREB1;PRKAR2B;PRKAR1A;PDE1A;PRKAR2A;ITPR2;ADCY3;CALM1;ADCY5 |
| 807 | Molecules associated with elastic fibres Homo sapiens R-HSA-2129379 | 3.80E-01 | 6.73E-01 | 1.22E+00 | 1.18E+00 | ITGB1;TGFB2;TGFB1;MFAP1;ITGB8;LTBP3;LTBP1;FBLN5 |
| 808 | Termination of translesion DNA synthesis Homo sapiens R-HSA-5656169 | 3.80E-01 | 6.73E-01 | 1.22E+00 | 1.18E+00 | PCNA;RFC1;RPA3;UBC;POLD2;UBE2L6;POLK;RPS27A |
| 809 | Regulation of TP53 Activity through Phosphorylation Homo sapiens R-HSA-6804756 | 3.83E-01 | 6.78E-01 | 1.10E+00 | 1.06E+00 | SUPT16H;PRKAA1;TAF13;TAF10;CSNK2A2;TAF11;PRKAG1;PRKAG2;HIPK1;NUAK1;RAD50;CDK5;RPA3;CSNK2B;UBC;CDK2;MDM2;RAD17;PIN1;MAPKAPK5;RAD1;RPS27A |
| 810 | Gastrin-CREB signalling pathway via PKC and MAPK Homo sapiens R-HSA-881907 | 3.85E-01 | 6.79E-01 | 1.04E+00 | 9.92E-01 | APP;PEBP1;WDR83;FGF2;PSMD8;PSMD9;PSMD6;PSMD7;EDNRB;PSMD4;PSMD2;PSMD3;PSMD1;MAP2K2;F2R;RBX1;BTC;AKAP9;PSME1;PSME2;TLN1;PSMD10;SHC2;SPTBN5;PSMD12;PSMD11;PSMD14;ARHGEF25;PSMD13;CUL3;PDGFB;ITPR2;LPAR2;PIK3R3;RASAL1;PHB;KNG1;RASGRP3;RAP1B;GNG10;RAP1A;UBC;PSMF1;PTK2;PTPRA;FGF18;GRB2;GNB5;CALM1;DAGLA;YWHAB;RASGRF2;ARRB1;PPP2R5E;PEA15;BDKRB2;BDKRB1;JAK2;SPTAN1;JAK1;DUSP1;PPP2R5A;DUSP8;PSMA5;PSMA6;PSMA3;PLCB4;PSMA4;CREB1;PSMA1;PSMA2;RASA1;PLCB1;HBEGF;RGS19;PSMA7;PSMB6;PPP2CA;PSMB7;PPP2CB;PSMB5;PSMB2;GNG5;PSMB3;PSMB1;MAPK1;CSK;FYN;RPS27A;CAMK2G;DAB2IP;PSMB8;AGT;PSMC5;PSMC6;PSMC3;PSMC4;PSMC1;PSMC2;F2RL1;KRAS;LAMTOR3 |

| | | | | | | |
|---|---|---|---|---|---|---|
| 811 | A tetrasaccharide linker sequence is required for GAG synthesis Homo sapiens R-HSA-1971475 | 3.86E-01 | 6.79E-01 | 1.24E+00 | 1.18E+00 | B3GAT3;SDC2;GPC1;SDC1;GPC5;HSPG2;B4GALT7 |
| 812 | ZBP1(DAI) mediated induction of type I IFNs Homo sapiens R-HSA-1606322 | 3.86E-01 | 6.79E-01 | 1.24E+00 | 1.18E+00 | NFKBIA;APP;TBK1;IRF3;DHX9;HMGB1;NFKB1 |
| 813 | WNT ligand biogenesis and trafficking Homo sapiens R-HSA-3238698 | 3.86E-01 | 6.79E-01 | 1.24E+00 | 1.18E+00 | SNX3;VPS29;WNT11;WNT7B;VPS26A;VPS35;TMED5 |
| 814 | RA biosynthesis pathway Homo sapiens R-HSA-5365859 | 3.92E-01 | 6.87E-01 | 1.26E+00 | 1.18E+00 | ALDH1A3;RDH11;RDH10;RDH14;RDH13;DHRS4 |
| 815 | Nephrin interactions Homo sapiens R-HSA-373753 | 3.92E-01 | 6.87E-01 | 1.26E+00 | 1.18E+00 | ACTN1;MAGI2;FYN;WASL;SPTAN1;CD2AP |
| 816 | ADP signalling through P2Y purinoceptor 12 Homo sapiens R-HSA-392170 | 3.92E-01 | 6.87E-01 | 1.26E+00 | 1.18E+00 | GNG10;GNG5;GNAI3;GNB5;GNAI1;GNAI2 |
| 817 | Vasopressin regulates renal water homeostasis via Aquaporins Homo sapiens R-HSA-432040 | 3.96E-01 | 6.94E-01 | 1.16E+00 | 1.07E+00 | GNG10;GNG5;PRKAR2B;PRKAR1A;PRKAR2A;GNAS;ADCY3;GNB5;RAB11FIP2;RAB11A;ADCY5 |

| | | | | | | |
|---|---|---|---|---|---|---|
| 818 | Signaling by cytosolic FGFR1 fusion mutants Homo sapiens R-HSA-1839117 | 3.98E-01 | 6.96E-01 | 1.29E+00 | 1.19E+00 | CPSF6;STAT3;FGFR1OP2;GRB2;LRRFIP1 |
| 819 | Tie2 Signaling Homo sapiens R-HSA-210993 | 3.98E-01 | 6.96E-01 | 1.29E+00 | 1.19E+00 | GRB7;GRB14;PTPN11;KRAS;GRB2 |
| 820 | Gamma carboxylation, hypusine formation and arylsulfatase activation Homo sapiens R-HSA-163841 | 4.03E-01 | 7.02E-01 | 1.16E+00 | 1.05E+00 | EIF5A;ICMT;ARSJ;DPH3;DHPS;FURIN;DPH7;DOHH;EEF2;GAS6 |
| 821 | tRNA modification in the nucleus and cytosol Homo sapiens R-HSA-6782315 | 4.03E-01 | 7.02E-01 | 1.16E+00 | 1.05E+00 | QTRT1;TRMT112;URM1;TRMT6;TYW1;TRMT11;EPRS;PUS7;LAGE3;TRMT61A |
| 822 | Regulation of TP53 Activity through Association with Co-factors Homo sapiens R-HSA-6804759 | 4.05E-01 | 7.02E-01 | 1.34E+00 | 1.22E+00 | PPP1R13B;PHF20;AKT2;AKT1 |
| 823 | TRAF3-dependent IRF activation pathway Homo sapiens R-HSA-918233 | 4.05E-01 | 7.02E-01 | 1.34E+00 | 1.22E+00 | MAVS;SIKE1;TBK1;IRF3 |
| 824 | Methylation Homo sapiens R-HSA-156581 | 4.05E-01 | 7.02E-01 | 1.34E+00 | 1.22E+00 | TRMT112;TPMT;GSTO1;COMT |
| 825 | Synthesis of bile acids and bile salts via 24-hydroxycholesterol Homo sapiens R-HSA-193775 | 4.05E-01 | 7.02E-01 | 1.34E+00 | 1.22E+00 | CYP39A1;AKR1C1;AKR1C2;SLC27A5 |

| | | | | | | |
|---|---|---|---|---|---|---|
| 826 | Regulation of pyruvate dehydrogenase (PDH) complex Homo sapiens R-HSA-204174 | 4.05E-01 | 7.02E-01 | 1.34E+00 | 1.22E+00 | PDHA1;PDK3;PDHB;PDK2 |
| 827 | Interactions of Vpr with host cellular proteins Homo sapiens R-HSA-176033 | 4.11E-01 | 7.05E-01 | 1.16E+00 | 1.04E+00 | NUP93;NUP50;TPR;HMGA1;PSIP1;SLC25A5;SLC25A4;KPNA1;SLC25A6 |
| 828 | Bile acid and bile salt metabolism Homo sapiens R-HSA-194068 | 4.11E-01 | 7.05E-01 | 1.16E+00 | 1.04E+00 | CYP39A1;NCOA2;ACOT8;SCP2;AKR1C1;SLCO1A2;HSD17B4;AKR1C2;SLC27A5 |
| 829 | Zinc influx into cells by the SLC39 gene family Homo sapiens R-HSA-442380 | 4.12E-01 | 7.05E-01 | 1.44E+00 | 1.28E+00 | SLC39A7;SLC39A1;SLC39A4 |
| 830 | Nef mediated downregulation of MHC class I complex cell surface expression Homo sapiens R-HSA-164940 | 4.12E-01 | 7.05E-01 | 1.44E+00 | 1.28E+00 | AP1G1;AP1S2;AP1M2 |
| 831 | Reactions specific to the complex N-glycan synthesis pathway Homo sapiens R-HSA-975578 | 4.12E-01 | 7.05E-01 | 1.44E+00 | 1.28E+00 | FUT8;MAN2A2;FUCA1 |
| 832 | Removal of the Flap Intermediate from the C-strand Homo sapiens R-HSA-174437 | 4.12E-01 | 7.05E-01 | 1.44E+00 | 1.28E+00 | PCNA;RPA3;POLD2 |

| | | | | | | |
|---|---|---|---|---|---|---|
| 833 | MAPK3 (ERK1) activation Homo sapiens R-HSA-110056 | 4.12E-01 | 7.05E-01 | 1.44E+00 | 1.28E+00 | PTPN11;JAK2;JAK1 |
| 834 | Regulation of gene expression by Hypoxia-inducible Factor Homo sapiens R-HSA-1234158 | 4.12E-01 | 7.05E-01 | 1.44E+00 | 1.28E+00 | ARNT;HIF1A;VEGFA |
| 835 | IRAK1 recruits IKK complex Homo sapiens R-HSA-937039 | 4.12E-01 | 7.05E-01 | 1.44E+00 | 1.28E+00 | IRAK1;PELI1;UBE2V1 |
| 836 | IRAK1 recruits IKK complex upon TLR7/8 or 9 stimulation Homo sapiens R-HSA-975144 | 4.12E-01 | 7.05E-01 | 1.44E+00 | 1.28E+00 | IRAK1;PELI1;UBE2V1 |
| 837 | DNA Damage Reversal Homo sapiens R-HSA-73942 | 4.16E-01 | 7.05E-01 | 1.68E+00 | 1.47E+00 | MGMT;ASCC3 |
| 838 | IRF3 mediated activation of type 1 IFN Homo sapiens R-HSA-1606341 | 4.16E-01 | 7.05E-01 | 1.68E+00 | 1.47E+00 | TBK1;IRF3 |
| 839 | Telomere Extension By Telomerase Homo sapiens R-HSA-171319 | 4.16E-01 | 7.05E-01 | 1.68E+00 | 1.47E+00 | DKC1;NHP2 |
| 840 | Sensing of DNA Double Strand Breaks Homo sapiens R-HSA-5693548 | 4.16E-01 | 7.05E-01 | 1.68E+00 | 1.47E+00 | RAD50;KPNA2 |

| | | | | | | |
|---|---|---|---|---|---|---|
| 841 | Negative feedback regulation of MAPK pathway Homo sapiens R-HSA-5674499 | 4.16E-01 | 7.05E-01 | 1.68E+00 | 1.47E+00 | MAP2K2;MAPK1 |
| 842 | RSK activation Homo sapiens R-HSA-444257 | 4.16E-01 | 7.05E-01 | 1.68E+00 | 1.47E+00 | PDPK1;MAPK1 |
| 843 | CLEC7A/inflammasome pathway Homo sapiens R-HSA-5660668 | 4.16E-01 | 7.05E-01 | 1.68E+00 | 1.47E+00 | PYCARD;NFKB1 |
| 844 | Transport and synthesis of PAPS Homo sapiens R-HSA-174362 | 4.16E-01 | 7.05E-01 | 1.68E+00 | 1.47E+00 | SLC35B3;PAPSS1 |
| 845 | Synthesis of 5-eicosatetraenoic acids Homo sapiens R-HSA-2142688 | 4.16E-01 | 7.05E-01 | 1.68E+00 | 1.47E+00 | GPX4;ALOX5 |
| 846 | FGFR1 mutant receptor activation Homo sapiens R-HSA-1839124 | 4.19E-01 | 7.08E-01 | 1.17E+00 | 1.02E+00 | BAG4;CPSF6;FGFR1OP2;STAT3;GRB2;LRRFIP1;ERLIN2;FGF2 |
| 847 | Regulation of TNFR1 signaling Homo sapiens R-HSA-5357905 | 4.19E-01 | 7.08E-01 | 1.17E+00 | 1.02E+00 | SHARPIN;TRADD;UBC;USP2;TAX1BP1;RBCK1;RPS27A;TNF |
| 848 | CREB phosphorylation through the activation of Ras Homo sapiens R-HSA-442742 | 4.28E-01 | 7.22E-01 | 1.18E+00 | 9.99E-01 | CREB1;PDPK1;RASGRF2;AKAP9;MAPK1;CALM1;CAMK2G |

| | | | | | | |
|---|---|---|---|---|---|---|
| 849 | Phospholipase C-mediated cascade; FGFR3 Homo sapiens R-HSA-5654227 | 4.29E-01 | 7.23E-01 | 1.12E+00 | 9.49E-01 | CREB1;PRKAR2B;PRKAR1A;PDE1A;PRKAR2A;FGF18;ITPR2;ADCY3;CALM1;FGF2;ADCY5 |
| 850 | Ion channel transport Homo sapiens R-HSA-983712 | 4.31E-01 | 7.25E-01 | 1.04E+00 | 8.77E-01 | ATP6V1A;TTYH3;CLCNKB;ATP2A3;CLCNKA;ATP1A1;SRI;ATP12A;CLCN3;BEST3;UBC;BSND;ANO10;ATP6V1H;TRPM7;ATP6V1E1;RPS27A;ATP6V0E2;ATP6V1D;CAMK2G;ATP6V1C1;ATP6V1F;ATP6V1G1;ATP6V0B;ATP6V0E1;ATP6AP1;ATP8B1;ANO6;ATP1B3;ATP2B1;ATP1B1;MCOLN1;ANO5;TRPC4AP;TPCN1;CLCN7;OSTM1;ASPH;FXYD3;FXYD2;TRPV4;WNK3;FKBP1B;ATP6V0D1;CALM1;SGK2;ATP6V0C;CUTC |
| 851 | Pre-NOTCH Expression and Processing Homo sapiens R-HSA-1912422 | 4.35E-01 | 7.32E-01 | 1.09E+00 | 9.11E-01 | POGLUT1;SEL1L;ATP2A3;FURIN;RFNG;RBPJ;KAT2B;SNW1;TFDP2;AGO2;ST3GAL4;TMED2;ST3GAL6;RAB6A |
| 852 | PKMTs methylate histone lysines Homo sapiens R-HSA-3214841 | 4.38E-01 | 7.33E-01 | 1.12E+00 | 9.26E-01 | SUZ12;SUV39H2;SETD3;MECOM;RBBP4;SETDB2;EHMT1;ASH2L;SMYD3;RBBP7 |
| 853 | The canonical retinoid cycle in rods (twilight vision) Homo sapiens R-HSA-2453902 | 4.38E-01 | 7.33E-01 | 1.19E+00 | 9.80E-01 | RDH11;HSD17B1;RDH10;ABCA4;NAPEPLD;MYO7A |
| 854 | Incretin synthesis, secretion, and inactivation Homo sapiens R-HSA-400508 | 4.38E-01 | 7.33E-01 | 1.19E+00 | 9.80E-01 | DPP4;SPCS3;SPCS2;SPCS1;SEC11A;SEC11C |
| 855 | Stimuli-sensing channels Homo sapiens R-HSA-2672351 | 4.38E-01 | 7.33E-01 | 1.06E+00 | 8.77E-01 | TTYH3;CLCNKB;CLCNKA;ANO6;SRI;CLCN3;MCOLN1;BEST3;TRPC4AP;ANO5;TPCN1;CLCN7;ASPH;OSTM1;TRPV4;UBC;WNK3;FKBP1B;BSND;ANO10;TRPM7;CALM1;RPS27A;SGK2 |

| | | | | | | |
|---|---|---|---|---|---|---|
| 856 | Defective B3GALT6 causes EDSP2 and SEMDJL1 Homo sapiens R-HSA-4420332 | 4.49E-01 | 7.48E-01 | 1.20E+00 | 9.61E-01 | SDC2;GPC1;SDC1;GPC5;HSPG2 |
| 857 | Defective B4GALT7 causes EDS, progeroid type Homo sapiens R-HSA-3560783 | 4.49E-01 | 7.48E-01 | 1.20E+00 | 9.61E-01 | SDC2;GPC1;SDC1;GPC5;HSPG2 |
| 858 | Defective B3GAT3 causes JDSSDHD Homo sapiens R-HSA-3560801 | 4.49E-01 | 7.48E-01 | 1.20E+00 | 9.61E-01 | SDC2;GPC1;SDC1;GPC5;HSPG2 |
| 859 | Regulation of TP53 Activity through Methylation Homo sapiens R-HSA-6804760 | 4.49E-01 | 7.48E-01 | 1.20E+00 | 9.61E-01 | SETD9;UBC;MDM2;EHMT1;RPS27A |
| 860 | Interferon Signaling Homo sapiens R-HSA-913531 | 4.56E-01 | 7.59E-01 | 1.03E+00 | 8.09E-01 | EIF4A2;EIF4A1;SP100;IFITM2;EIF4E3;EIF4A3;UBE2L6;SOCS1;SUMO1;TRIM5;TRIM2;TPR;UBC;KPNA4;JAK2;RPS27A;KPNA2;KPNA3;EIF4E;CAMK2G;JAK1;KPNA1;IP6K2;IFNAR2;IFNGR2;UBE2E1;HLA-C;EIF2AK2;PTPN11;HLA-G;PSMB8;HLA-E;NUP93;ISG20;IRF3;NEDD4;NUP50;IRF2;PIN1;IRF5;PTPN6;EIF4G3;EIF4G2;HLA-DQB1;IFNAR1;EIF4G1 |
| 861 | Keratan sulfate/keratin metabolism Homo sapiens R-HSA-1638074 | 4.58E-01 | 7.61E-01 | 1.12E+00 | 8.76E-01 | B4GALT3;GLB1;HEXA;ST3GAL4;B3GNT3;CHST1;B3GNT2;ST3GAL6 |
| 862 | Cell surface interactions at the vascular wall Homo sapiens R-HSA-202733 | 4.59E-01 | 7.61E-01 | 1.05E+00 | 8.15E-01 | ITGB1;GRB7;CD74;CXADR;YES1;ITGB2;ATP1B3;PTPN11;MIF;ATP1B1;L1CAM;SLC7A6;GRB14;SLC7A8;ESAM;GRB2;FYN;PTPN6;KRAS;CD47;CD58;GAS6;SLC16A3;PPIA |

| | | | | | | |
|---|---|---|---|---|---|---|
| 863 | RHO GTPases Activate Formins Homo sapiens R-HSA-5663220 | 4.60E-01 | 7.61E-01 | 1.04E+00 | 8.10E-01 | ITGB1;PMF1;ACTG1;SKA2;CENPC;PPP2CA;CDC42;PPP1CC;PPP2CB;PPP2R5E;DVL3;RAC1;BUB3;NUDC;PPP2R5A;RHOC;RHOD;RHOA;FMNL3;CLIP1;FMNL1;KIF2A;TAOK1;PFN1;MAPRE1;PFN2;PAFAH1B1 |
| 864 | SUMOylation of RNA binding proteins Homo sapiens R-HSA-4570464 | 4.62E-01 | 7.61E-01 | 1.09E+00 | 8.41E-01 | NUP93;PHC2;UBE2I;HNRNPK;SUMO1;CBX4;PCGF2;NUP50;TPR;SUMO2;HNRNPC |
| 865 | Diseases of metabolism Homo sapiens R-HSA-5668914 | 4.62E-01 | 7.61E-01 | 1.09E+00 | 8.41E-01 | PC;IDH1;UBC;MCCC1;CCDC59;PCCB;FDX1;RPS27A;ACACA;MMADHC;CD320 |
| 866 | Regulation of innate immune responses to cytosolic DNA Homo sapiens R-HSA-3134975 | 4.62E-01 | 7.61E-01 | 1.22E+00 | 9.43E-01 | TBK1;IRF3;UBC;RPS27A |
| 867 | Constitutive Signaling by NOTCH1 HD Domain Mutants Homo sapiens R-HSA-2691232 | 4.62E-01 | 7.61E-01 | 1.22E+00 | 9.43E-01 | JAG2;UBC;MIB2;RPS27A |
| 868 | Signaling by NOTCH1 HD Domain Mutants in Cancer Homo sapiens R-HSA-2691230 | 4.62E-01 | 7.61E-01 | 1.22E+00 | 9.43E-01 | JAG2;UBC;MIB2;RPS27A |
| 869 | CREB phosphorylation through the activation of CaMKII Homo sapiens R-HSA-442729 | 4.62E-01 | 7.61E-01 | 1.22E+00 | 9.43E-01 | CREB1;AKAP9;CALM1;CAMK2G |
| 870 | G-protein activation Homo sapiens R-HSA-202040 | 4.69E-01 | 7.72E-01 | 1.12E+00 | 8.48E-01 | GNAO1;GNG10;GNG5;GNAI3;GNB5;GNAI1;GNAI2 |

| | | | | | | |
|---|---|---|---|---|---|---|
| 871 | Host Interactions with Influenza Factors Homo sapiens R-HSA-168253 | 4.72E-01 | 7.74E-01 | 1.08E+00 | 8.14E-01 | NUP93;TGFB1;NUP50;TPR;EIF2AK2;KPNA4;KPNA2;KPNA3;KPNA1;SLC25A6 |
| 872 | TNF signaling Homo sapiens R-HSA-75893 | 4.72E-01 | 7.74E-01 | 1.08E+00 | 8.14E-01 | BAG4;SHARPIN;TRADD;UBC;USP2;TAX1BP1;FADD;RBCK1;RPS27A;TNF |
| 873 | DNA Double-Strand Break Repair Homo sapiens R-HSA-5693532 | 4.73E-01 | 7.75E-01 | 1.03E+00 | 7.71E-01 | MUS81;PAXIP1;PCNA;PRKDC;BRCC3;BABAM1;HERC2;PPP4C;PPP4R2;SUMO1;UBC;UIMC1;SUMO2;POLD2;POLK;KPNA2;RPS27A;XRCC6;UBE2I;RFC1;PARP1;XRCC5;POLM;SMARCA5;LIG4;RNF168;RAD51B;RAD50;RPA3;ERCC1;CDK2;UBE2V2;RAD17;RAD1 |
| 874 | Transport of organic anions Homo sapiens R-HSA-879518 | 4.79E-01 | 7.78E-01 | 1.26E+00 | 9.28E-01 | SLCO3A1;SLCO4A1;SLCO1A2 |
| 875 | Processive synthesis on the C-strand of the telomere Homo sapiens R-HSA-174414 | 4.79E-01 | 7.78E-01 | 1.26E+00 | 9.28E-01 | PCNA;RPA3;POLD2 |
| 876 | TRAF6 mediated IRF7 activation in TLR7/8 or 9 signaling Homo sapiens R-HSA-975110 | 4.79E-01 | 7.78E-01 | 1.26E+00 | 9.28E-01 | IRAK1;UBE2V1;IRAK4 |
| 877 | CASP8 activity is inhibited Homo sapiens R-HSA-5218900 | 4.79E-01 | 7.78E-01 | 1.26E+00 | 9.28E-01 | TRADD;CFLAR;FADD |
| 878 | Regulation by c-FLIP Homo sapiens R-HSA-3371378 | 4.79E-01 | 7.78E-01 | 1.26E+00 | 9.28E-01 | TRADD;CFLAR;FADD |
| 879 | Dimerization of procaspase-8 Homo sapiens R-HSA-69416 | 4.79E-01 | 7.78E-01 | 1.26E+00 | 9.28E-01 | TRADD;CFLAR;FADD |

| | | | | | | |
|---|---|---|---|---|---|---|
| 880 | CLEC7A (Dectin-1) induces NFAT activation Homo sapiens R-HSA-5607763 | 4.79E-01 | 7.78E-01 | 1.26E+00 | 9.28E-01 | PPP3CB;ITPR2;CALM1 |
| 881 | Synthesis of IP2, IP, and Ins in the cytosol Homo sapiens R-HSA-1855183 | 4.79E-01 | 7.78E-01 | 1.26E+00 | 9.28E-01 | ISYNA1;IMPA1;INPP1 |
| 882 | Nuclear Events (kinase and transcription factor activation) Homo sapiens R-HSA-198725 | 4.83E-01 | 7.78E-01 | 1.12E+00 | 8.16E-01 | ATF1;PPP2CA;PPP2CB;DUSP3;CREB1;MAPK1 |
| 883 | IKK complex recruitment mediated by RIP1 Homo sapiens R-HSA-937041 | 4.83E-01 | 7.78E-01 | 1.12E+00 | 8.16E-01 | UBE2D2;UBE2D3;UBC;UBE2D1;UBE2V1;RPS27A |
| 884 | Insulin processing Homo sapiens R-HSA-264876 | 4.83E-01 | 7.78E-01 | 1.12E+00 | 8.16E-01 | KIF5C;KIF5B;MYO5A;RAB27A;CPE;EXOC6 |
| 885 | Pyrimidine metabolism Homo sapiens R-HSA-73848 | 4.83E-01 | 7.78E-01 | 1.12E+00 | 8.16E-01 | NT5C3A;CDA;DUT;UCK1;TK2;TK1 |
| 886 | Chondroitin sulfate/dermatan sulfate metabolism Homo sapiens R-HSA-1793185 | 4.83E-01 | 7.78E-01 | 1.06E+00 | 7.72E-01 | CHPF;CHST12;B3GAT3;SDC2;GPC1;HEXA;CHST15;SDC1;GPC5;CHST14;HSPG2;B4GALT7 |
| 887 | Metabolism of non-coding RNA Homo sapiens R-HSA-194441 | 4.83E-01 | 7.78E-01 | 1.06E+00 | 7.72E-01 | NUP93;SNRPD2;NCBP1;SNRPD1;NUP50;TGS1;TPR;SNRPG;PHAX;GEMIN6;SNRPD3;SNRPB |
| 888 | snRNP Assembly Homo sapiens R-HSA-191859 | 4.83E-01 | 7.78E-01 | 1.06E+00 | 7.72E-01 | NUP93;SNRPD2;NCBP1;SNRPD1;NUP50;TGS1;TPR;SNRPG;PHAX;GEMIN6;SNRPD3;SNRPB |

| | | | | | | |
|---|---|---|---|---|---|---|
| 889 | Translesion synthesis by Y family DNA polymerases bypasses lesions on DNA template Homo sapiens R-HSA-110313 | 4.83E-01 | 7.78E-01 | 1.08E+00 | 7.86E-01 | VCP;PCNA;RFC1;RPA3;UBC;POLD2;UBE2L6;POLK;RPS27A |
| 890 | Phospholipase C-mediated cascade; FGFR4 Homo sapiens R-HSA-5654228 | 4.94E-01 | 7.91E-01 | 1.06E+00 | 7.45E-01 | CREB1;PRKAR2B;PRKAR1A;PDE1A;PRKAR2A;FGF18;ITPR2;ADCY3;CALM1;FGF2;ADCY5 |
| 891 | Glucagon signaling in metabolic regulation Homo sapiens R-HSA-163359 | 4.96E-01 | 7.91E-01 | 1.08E+00 | 7.55E-01 | GNG10;GNG5;PRKAR2B;PRKAR1A;PRKAR2A;GNAS;ADCY3;ADCY5 |
| 892 | Defective C1GALT1C1 causes Tn polyagglutination syndrome (TNPS) Homo sapiens R-HSA-5083632 | 4.98E-01 | 7.91E-01 | 1.12E+00 | 7.80E-01 | MUC1;C1GALT1C1;C1GALT1;MUC15;MUC20 |
| 893 | Regulation of gene expression in beta cells Homo sapiens R-HSA-210745 | 4.98E-01 | 7.91E-01 | 1.12E+00 | 7.80E-01 | AKT2;AKT1;MAFA;GCK;FOXA3 |
| 894 | Interferon alpha/beta signaling Homo sapiens R-HSA-909733 | 5.00E-01 | 7.91E-01 | 1.03E+00 | 7.16E-01 | IFNAR2;IFITM2;HLA-C;PTPN11;HLA-G;PSMB8;HLA-E;ISG20;SOCS1;IRF3;IRF2;IRF5;PTPN6;JAK1;IFNAR1;IP6K2 |
| 895 | Inositol transporters Homo sapiens R-HSA-429593 | 5.02E-01 | 7.91E-01 | 1.34E+00 | 9.26E-01 | SLC2A13;SLC5A3 |

| | | | | | | |
|---|---|---|---|---|---|---|
| 896 | Clearance of Nuclear Envelope Membranes from Chromatin Homo sapiens R-HSA-2993913 | 5.02E-01 | 7.91E-01 | 1.34E+00 | 9.26E-01 | LEMD2;TMPO |
| 897 | PTK6 promotes HIF1A stabilization Homo sapiens R-HSA-8857538 | 5.02E-01 | 7.91E-01 | 1.34E+00 | 9.26E-01 | HIF1A;HBEGF |
| 898 | PP2A-mediated dephosphorylation of key metabolic factors Homo sapiens R-HSA-163767 | 5.02E-01 | 7.91E-01 | 1.34E+00 | 9.26E-01 | PPP2CA;PPP2CB |
| 899 | IkBA variant leads to EDA-ID Homo sapiens R-HSA-5603029 | 5.02E-01 | 7.91E-01 | 1.34E+00 | 9.26E-01 | NFKBIA;NFKB1 |
| 900 | TRAIL signaling Homo sapiens R-HSA-75158 | 5.02E-01 | 7.91E-01 | 1.34E+00 | 9.26E-01 | CFLAR;FADD |
| 901 | Activation of NIMA Kinases NEK9, NEK6, NEK7 Homo sapiens R-HSA-2980767 | 5.02E-01 | 7.91E-01 | 1.34E+00 | 9.26E-01 | CCNB1;NEK7 |
| 902 | CREB phosphorylation Homo sapiens R-HSA-199920 | 5.02E-01 | 7.91E-01 | 1.34E+00 | 9.26E-01 | ATF1;CREB1 |

| | | | | | | |
|---|---|---|---|---|---|---|
| 903 | CREB phosphorylation through the activation of Adenylate Cyclase Homo sapiens R-HSA-442720 | 5.02E-01 | 7.91E-01 | 1.34E+00 | 9.26E-01 | CREB1;ADCY3 |
| 904 | Phase 3 - rapid repolarisation Homo sapiens R-HSA-5576890 | 5.02E-01 | 7.91E-01 | 1.34E+00 | 9.26E-01 | KCNQ1;AKAP9 |
| 905 | Loss of Function of TGFBR1 in Cancer Homo sapiens R-HSA-3656534 | 5.02E-01 | 7.91E-01 | 1.34E+00 | 9.26E-01 | FKBP1A;TGFB1 |
| 906 | TGFBR1 KD Mutants in Cancer Homo sapiens R-HSA-3656532 | 5.02E-01 | 7.91E-01 | 1.34E+00 | 9.26E-01 | FKBP1A;TGFB1 |
| 907 | Loss of Function of SMAD2/3 in Cancer Homo sapiens R-HSA-3304349 | 5.02E-01 | 7.91E-01 | 1.34E+00 | 9.26E-01 | SMAD4;TGFB1 |
| 908 | SMAD2/3 MH2 Domain Mutants in Cancer Homo sapiens R-HSA-3315487 | 5.02E-01 | 7.91E-01 | 1.34E+00 | 9.26E-01 | SMAD4;TGFB1 |
| 909 | Ion transport by P-type ATPases Homo sapiens R-HSA-936837 | 5.03E-01 | 7.91E-01 | 1.04E+00 | 7.16E-01 | ATP8B1;ATP2A3;ATP1B3;ATP1A1;SRI;ATP2B1;ATP12A;ATP1B1;FXYD3;FXYD2;CALM1;CAMK2G;CUTC |
| 910 | Retinoid metabolism and transport Homo sapiens R-HSA-975634 | 5.06E-01 | 7.95E-01 | 1.05E+00 | 7.16E-01 | RDH11;SDC2;GPC1;AKR1C1;SDC1;GPC5;BCO2;LRP10;LRP8;HSPG2 |

| | | | | | | |
|---|---|---|---|---|---|---|
| 911 | YAP1- and WWTR1 (TAZ)-stimulated gene expression Homo sapiens R-HSA-2032785 | 5.10E-01 | 8.01E-01 | 1.07E+00 | 7.20E-01 | NCOA2;KAT2B;SMARCD3;TGS1;CARM1;HIPK1;TEAD2 |
| 912 | Fatty acid, triacylglycerol, and ketone body metabolism Homo sapiens R-HSA-535734 | 5.11E-01 | 8.02E-01 | 1.01E+00 | 6.76E-01 | SLC25A1;ACADVL;ALAS1;TECR;PRKAG2;CCNC;RORA;MED11;MED10;ME1;ACADM;HADH;TEAD2;ACOT9;NCOA2;ELOVL1;ACOT8;NCOA3;ACOT11;MED8;ACSL4;ELOVL6;MED4;MED27;SREBF2;NCOR2;HADHB;HADHA;NCOR1;MED24;MED21;PCCB;PLIN3;ECHS1;SMARCD3;HDAC3;HSD17B12;ACACA;ARNTL;MED30;TGS1;FDFT1;ESRRA;TXNRD1;NR1D1;AGT;CARM1;PPT1;PPT2;LPIN2 |
| 913 | Metabolism of fat-soluble vitamins Homo sapiens R-HSA-6806667 | 5.14E-01 | 8.04E-01 | 1.03E+00 | 6.89E-01 | VKORC1;VKORC1L1;RDH11;SDC2;GPC1;AKR1C1;SDC1;GPC5;BCO2;LRP10;HSPG2;LRP8 |
| 914 | G-protein beta:gamma signalling Homo sapiens R-HSA-397795 | 5.14E-01 | 8.04E-01 | 1.03E+00 | 6.89E-01 | GNG10;GNG5;PDPK1;AKT2;AKT1;PIK3R3;GRB2;GNB5;JAK2;PLCB1;RHOA;JAK1 |
| 915 | GRB2 events in ERBB2 signaling Homo sapiens R-HSA-1963640 | 5.18E-01 | 8.08E-01 | 1.12E+00 | 7.38E-01 | BTC;KRAS;GRB2;HBEGF |
| 916 | Metabolism of Angiotensinogen to Angiotensins Homo sapiens R-HSA-2022377 | 5.18E-01 | 8.08E-01 | 1.12E+00 | 7.38E-01 | ATP6AP2;CTSZ;CTSD;AGT |
| 917 | Metabolism of folate and pterines Homo sapiens R-HSA-196757 | 5.18E-01 | 8.08E-01 | 1.12E+00 | 7.38E-01 | SLC46A1;MTHFD1;SHMT1;MTHFS |
| 918 | Signaling by FGFR1 in disease Homo sapiens R-HSA-5655302 | 5.19E-01 | 8.09E-01 | 1.04E+00 | 6.85E-01 | BAG4;CPSF6;FGFR1OP2;STAT3;KRAS;GRB2;LRRFIP1;ERLIN2;FGF2 |

| | | | | | | |
|---|---|---|---|---|---|---|
| 919 | Regulation of IFNA signaling Homo sapiens R-HSA-912694 | 5.27E-01 | 8.20E-01 | 1.06E+00 | 6.81E-01 | IFNAR2;SOCS1;PTPN11;PTPN6;JAK1;IFNAR1 |
| 920 | G2/M DNA damage checkpoint Homo sapiens R-HSA-69473 | 5.32E-01 | 8.27E-01 | 1.01E+00 | 6.37E-01 | YWHAE;YWHAB;BRCC3;YWHAZ;BABAM1;RNF168;CCNB1;RAD50;HERC2;YWHAQ;SUMO1;RPA3;UIMC1;UBE2V2;RAD17;RAD1;YWHAG;YWHAH |
| 921 | ERCC6 (CSB) and EHMT2 (G9a) positively regulate rRNA expression Homo sapiens R-HSA-427389 | 5.39E-01 | 8.37E-01 | 1.02E+00 | 6.30E-01 | HDAC2;RBBP4;HDAC1;H2AFZ;CBX3;H3F3A;CHD4;RBBP7;MTA3;HIST1H2AC |
| 922 | Hyaluronan uptake and degradation Homo sapiens R-HSA-2160916 | 5.43E-01 | 8.41E-01 | 1.12E+00 | 6.85E-01 | HYAL2;HEXA;CHP1 |
| 923 | Inhibition of replication initiation of damaged DNA by RB1/E2F1 Homo sapiens R-HSA-113501 | 5.43E-01 | 8.41E-01 | 1.12E+00 | 6.85E-01 | RB1;PPP2CA;PPP2CB |
| 924 | Physiological factors Homo sapiens R-HSA-5578768 | 5.43E-01 | 8.41E-01 | 1.12E+00 | 6.85E-01 | NPPC;KAT2B;HIPK1 |
| 925 | RIP-mediated NFkB activation via ZBP1 Homo sapiens R-HSA-1810476 | 5.46E-01 | 8.41E-01 | 1.05E+00 | 6.36E-01 | NFKBIA;APP;DHX9;HMGB1;NFKB1 |
| 926 | Regulation of FZD by ubiquitination Homo sapiens R-HSA-4641263 | 5.46E-01 | 8.41E-01 | 1.05E+00 | 6.36E-01 | RNF43;USP8;UBC;RPS27A;LGR5 |

| | | | | | | |
|---|---|---|---|---|---|---|
| 927 | CD209 (DC-SIGN) signaling Homo sapiens R-HSA-5621575 | 5.46E-01 | 8.41E-01 | 1.05E+00 | 6.36E-01 | PAK1;KRAS;FYN;PAK2;NFKB1 |
| 928 | Cobalamin (Cbl, vitamin B12) transport and metabolism Homo sapiens R-HSA-196741 | 5.46E-01 | 8.41E-01 | 1.05E+00 | 6.36E-01 | LMBRD1;MMAB;PRSS3;MMADHC;CD320 |
| 929 | Mitochondrial tRNA aminoacylation Homo sapiens R-HSA-379726 | 5.46E-01 | 8.41E-01 | 1.05E+00 | 6.36E-01 | QARS;IARS2;GARS;YARS2;CARS2 |
| 930 | Gap junction trafficking and regulation Homo sapiens R-HSA-157858 | 5.50E-01 | 8.45E-01 | 1.02E+00 | 6.12E-01 | TJP1;CLTC;MYO6;CLTB;CLTA;AP2M1;DNM2 |
| 931 | MAPK targets/ Nuclear events mediated by MAP kinases Homo sapiens R-HSA-450282 | 5.50E-01 | 8.45E-01 | 1.02E+00 | 6.12E-01 | ATF1;PPP2CA;PPP2CB;MAPK9;DUSP3;CREB1;MAPK1 |
| 932 | Collagen degradation Homo sapiens R-HSA-1442490 | 5.53E-01 | 8.50E-01 | 1.01E+00 | 5.97E-01 | COL26A1;COL14A1;CTSL;CTSK;COL12A1;ADAM9;FURIN;CTSD;CTSB |
| 933 | Death Receptor Signalling Homo sapiens R-HSA-73887 | 5.57E-01 | 8.53E-01 | 9.99E-01 | 5.85E-01 | BAG4;SHARPIN;TRADD;UBC;USP2;TAX1BP1;FADD;CFLAR;RBCK1;RPS27A;TNF |
| 934 | G beta:gamma signalling through PI3Kgamma Homo sapiens R-HSA-392451 | 5.57E-01 | 8.53E-01 | 9.99E-01 | 5.85E-01 | GNG10;GNG5;PDPK1;AKT2;PIK3R3;AKT1;GRB2;GNB5;JAK2;RHOA;JAK1 |

| | | | | | | |
|---|---|---|---|---|---|---|
| 935 | RNA Polymerase I Chain Elongation Homo sapiens R-HSA-73777 | 5.60E-01 | 8.58E-01 | 9.93E-01 | 5.75E-01 | H2AFZ;CBX3;H3F3A;GTF2H4;ZNRD1;TWISTNB;POLR1D;POLR2E;POLR2H;MNAT1;HIST1H2AC;POLR2K;POLR2L |
| 936 | N-glycan antennae elongation in the medial/trans-Golgi Homo sapiens R-HSA-975576 | 5.69E-01 | 8.65E-01 | 1.01E+00 | 5.69E-01 | B4GALT3;FUT8;MAN2A2;FUCA1;MGAT4B;ST3GAL4 |
| 937 | TAK1 activates NFkB by phosphorylation and activation of IKKs complex Homo sapiens R-HSA-445989 | 5.69E-01 | 8.65E-01 | 1.01E+00 | 5.69E-01 | NFKBIA;APP;IRAK1;DHX9;HMGB1;NFKB1 |
| 938 | Phase 2 - plateau phase Homo sapiens R-HSA-5576893 | 5.69E-01 | 8.65E-01 | 1.01E+00 | 5.69E-01 | CACNB4;KCNQ1;AKAP9;CACNA2D3;CACNA1D;CACNA1C |
| 939 | Post NMDA receptor activation events Homo sapiens R-HSA-438064 | 5.70E-01 | 8.65E-01 | 9.96E-01 | 5.60E-01 | CREB1;PDPK1;RASGRF2;AKAP9;ADCY3;MAPK1;CALM1;CAMK2G |
| 940 | Hyaluronan metabolism Homo sapiens R-HSA-2142845 | 5.70E-01 | 8.65E-01 | 1.03E+00 | 5.81E-01 | ABCC5;HYAL2;HEXA;CHP1 |
| 941 | Formation of Senescence-Associated Heterochromatin Foci (SAHF) Homo sapiens R-HSA-2559584 | 5.70E-01 | 8.65E-01 | 1.03E+00 | 5.81E-01 | RB1;H1F0;HMGA1;HIST1H1E |

| | | | | | | |
|---|---|---|---|---|---|---|
| 942 | Ras activation uopn Ca2+ infux through NMDA receptor Homo sapiens R-HSA-442982 | 5.70E-01 | 8.65E-01 | 1.03E+00 | 5.81E-01 | RASGRF2;AKAP9;CALM1;CAMK2G |
| 943 | Inflammasomes Homo sapiens R-HSA-622312 | 5.70E-01 | 8.65E-01 | 1.03E+00 | 5.81E-01 | PYCARD;APP;HSP90AB1;SUGT1 |
| 944 | Glycerophospholipid biosynthesis Homo sapiens R-HSA-1483206 | 5.75E-01 | 8.69E-01 | 9.78E-01 | 5.41E-01 | DMGDH;SLC44A3;ABHD3;PITPNB;PLA2G4A;MBOAT2;TAZ;CRLS1;PISD;PGS1;HADHB;PTDSS2;HADHA;ETNK2;PTPMT1;CHDH;ETNK1;CPNE1;CPNE3;CHPT1;ALDH7A1;LPIN2;PNPLA2 |
| 945 | Release of Hh-Np from the secreting cell Homo sapiens R-HSA-5362798 | 5.79E-01 | 8.69E-01 | 1.12E+00 | 6.12E-01 | GPC5;NOTUM |
| 946 | Synthesis of UDP-N-acetyl-glucosamine Homo sapiens R-HSA-446210 | 5.79E-01 | 8.69E-01 | 1.12E+00 | 6.12E-01 | GFPT1;GNPNAT1 |
| 947 | Activation of PUMA and translocation to mitochondria Homo sapiens R-HSA-139915 | 5.79E-01 | 8.69E-01 | 1.12E+00 | 6.12E-01 | PPP1R13B;BBC3 |
| 948 | AMPK inhibits chREBP transcriptional activation activity Homo sapiens R-HSA-163680 | 5.79E-01 | 8.69E-01 | 1.12E+00 | 6.12E-01 | PRKAG2;ADIPOR1 |

| | | | | | | |
|---|---|---|---|---|---|---|
| 949 | IKBKG deficiency causes anhidrotic ectodermal dysplasia with immunodeficiency (EDA-ID) (via TLR) Homo sapiens R-HSA-5603027 | 5.79E-01 | 8.69E-01 | 1.12E+00 | 6.12E-01 | NFKBIA;NFKB1 |
| 950 | Formyl peptide receptors bind formyl peptides and many other ligands Homo sapiens R-HSA-444473 | 5.79E-01 | 8.69E-01 | 1.12E+00 | 6.12E-01 | HEBP1;APP |
| 951 | VEGF ligand-receptor interactions Homo sapiens R-HSA-194313 | 5.79E-01 | 8.69E-01 | 1.12E+00 | 6.12E-01 | VEGFB;VEGFA |
| 952 | VEGF binds to VEGFR leading to receptor dimerization Homo sapiens R-HSA-195399 | 5.79E-01 | 8.69E-01 | 1.12E+00 | 6.12E-01 | VEGFB;VEGFA |
| 953 | Synthesis of PG Homo sapiens R-HSA-1483148 | 5.79E-01 | 8.69E-01 | 1.12E+00 | 6.12E-01 | PGS1;PTPMT1 |
| 954 | ChREBP activates metabolic gene expression Homo sapiens R-HSA-163765 | 5.79E-01 | 8.69E-01 | 1.12E+00 | 6.12E-01 | MLX;ACACA |
| 955 | HATs acetylate histones Homo sapiens R-HSA-3214847 | 5.87E-01 | 8.77E-01 | 9.72E-01 | 5.18E-01 | NCOA2;DR1;MEAF6;PHF20;MCRS1;TAF10;ACTL6A;MSL3;ELP5;ELP6;ING4;MORF4L1;TAF6L;KAT2B;MRGBP;HAT1;TADA3;MBIP;KAT8;DMAP1;RBBP7;BRD8;OGT;HIST1H2AC |

| | | | | | | |
|---|---|---|---|---|---|---|
| 956 | Mitotic Prometaphase Homo sapiens R-HSA-68877 | 5.87E-01 | 8.77E-01 | 9.72E-01 | 5.18E-01 | NUDC;CSNK2A2;PPP2R5A;PDS5B;SMC3;SMC1A;SMC4;PMF1;SKA2;CENPC;PPP2CA;PPP1CC;PPP2CB;CLIP1;CCNB1;STAG2;KIF2A;PPP2R5E;RAD21;TAOK1;CSNK2B;BUB3;MAPRE1;PAFAH1B1 |
| 957 | PRC2 methylates histones and DNA Homo sapiens R-HSA-212300 | 5.87E-01 | 8.77E-01 | 9.76E-01 | 5.20E-01 | SUZ12;DNMT1;RBBP4;H2AFZ;PHF1;H3F3A;RBBP7;JARID2;HIST1H2AC |
| 958 | Phospholipase C-mediated cascade; FGFR2 Homo sapiens R-HSA-5654221 | 5.87E-01 | 8.77E-01 | 9.73E-01 | 5.18E-01 | CREB1;PRKAR1A;PRKAR2B;PDE1A;PRKAR2A;FGF18;ITPR2;ADCY3;CALM1;FGF2;ADCY5 |
| 959 | Resolution of AP sites via the multiple-nucleotide patch replacement pathway Homo sapiens R-HSA-110373 | 5.91E-01 | 8.82E-01 | 9.89E-01 | 5.19E-01 | PCNA;PARP1;RFC1;RPA3;POLD2 |
| 960 | HDR through Homologous Recombination (HR) or Single Strand Annealing (SSA) Homo sapiens R-HSA-5693567 | 5.98E-01 | 8.88E-01 | 9.66E-01 | 4.97E-01 | MUS81;PCNA;BRCC3;BABAM1;HERC2;PPP4C;PPP4R2;SUMO1;UBC;UIMC1;SUMO2;POLD2;POLK;RPS27A;UBE2I;RFC1;RNF168;RAD51B;RAD50;RPA3;ERCC1;CDK2;UBE2V2;RAD17;RAD1 |
| 961 | Glycosaminoglycan metabolism Homo sapiens R-HSA-1630316 | 5.99E-01 | 8.88E-01 | 9.65E-01 | 4.94E-01 | SLC35B3;B4GALT3;CHPF;SDC2;HEXA;PAPSS1;NAGLU;CHST12;HYAL2;GPC1;CHST15;ST3GAL4;CHP1;ST3GAL6;GPC5;CHST14;B3GAT3;ABCC5;HSPG2;HS3ST1;GLB1;B3GNT3;SDC1;CHST1;B3GNT2;HPSE;B4GALT7 |
| 962 | TP53 Regulates Transcription of Genes Involved in G1 Cell Cycle Arrest Homo sapiens R-HSA-6804116 | 6.02E-01 | 8.88E-01 | 1.01E+00 | 5.12E-01 | CDKN1B;PCBP4;CDK2 |

| | | | | | | |
|---|---|---|---|---|---|---|
| 963 | POU5F1 (OCT4), SOX2, NANOG activate genes related to proliferation Homo sapiens R-HSA-2892247 | 6.02E-01 | 8.88E-01 | 1.01E+00 | 5.12E-01 | STAT3;FGF2;TDGF1 |
| 964 | NICD traffics to nucleus Homo sapiens R-HSA-157052 | 6.02E-01 | 8.88E-01 | 1.01E+00 | 5.12E-01 | KAT2B;SNW1;RBPJ |
| 965 | Notch-HLH transcription pathway Homo sapiens R-HSA-350054 | 6.02E-01 | 8.88E-01 | 1.01E+00 | 5.12E-01 | KAT2B;SNW1;RBPJ |
| 966 | FGFR3c ligand binding and activation Homo sapiens R-HSA-190372 | 6.02E-01 | 8.88E-01 | 1.01E+00 | 5.12E-01 | GALNT3;FGF18;FGF2 |
| 967 | FGFR3 ligand binding and activation Homo sapiens R-HSA-190239 | 6.02E-01 | 8.88E-01 | 1.01E+00 | 5.12E-01 | GALNT3;FGF18;FGF2 |
| 968 | Depolarization of the Presynaptic Terminal Triggers the Opening of Calcium Channels Homo sapiens R-HSA-112308 | 6.02E-01 | 8.88E-01 | 1.01E+00 | 5.12E-01 | CACNB4;CACFD1;CACNA2D3 |
| 969 | Signaling by Activin Homo sapiens R-HSA-1502540 | 6.02E-01 | 8.88E-01 | 1.01E+00 | 5.12E-01 | DRAP1;SMAD4;FST |
| 970 | RHO GTPases Activate NADPH Oxidases Homo sapiens R-HSA-5668599 | 6.02E-01 | 8.88E-01 | 1.01E+00 | 5.12E-01 | NOXA1;CYBA;RAC1 |

| | | | | | | |
|---|---|---|---|---|---|---|
| 971 | Processing of DNA double-strand break ends Homo sapiens R-HSA-5693607 | 6.03E-01 | 8.89E-01 | 9.60E-01 | 4.85E-01 | UBE2I;BRCC3;BABAM1;RNF168;RAD50;HERC2;PPP4C;PPP4R2;SUMO1;RPA3;UIMC1;SUMO2;UBC;CDK2;UBE2V2;RAD17;RAD1;RPS27A |
| 972 | Keratan sulfate biosynthesis Homo sapiens R-HSA-2022854 | 6.09E-01 | 8.96E-01 | 9.60E-01 | 4.76E-01 | B4GALT3;ST3GAL4;B3GNT3;CHST1;B3GNT2;ST3GAL6 |
| 973 | Platelet calcium homeostasis Homo sapiens R-HSA-418360 | 6.09E-01 | 8.96E-01 | 9.60E-01 | 4.76E-01 | ATP2A3;ITPR2;ORAI1;SRI;ATP2B1;CALM1 |
| 974 | DNA Double Strand Break Response Homo sapiens R-HSA-5693606 | 6.15E-01 | 9.04E-01 | 9.50E-01 | 4.61E-01 | UBE2I;SMARCA5;BRCC3;BABAM1;RNF168;RAD50;HERC2;SUMO1;UIMC1;UBC;UBE2V2;KPNA2;RPS27A |
| 975 | SHC-mediated cascade:FGFR3 Homo sapiens R-HSA-5654704 | 6.19E-01 | 9.05E-01 | 9.60E-01 | 4.60E-01 | FGF18;KRAS;GRB2;FGF2 |
| 976 | SHC1 events in ERBB2 signaling Homo sapiens R-HSA-1250196 | 6.19E-01 | 9.05E-01 | 9.60E-01 | 4.60E-01 | BTC;KRAS;GRB2;HBEGF |
| 977 | Cytosolic sulfonation of small molecules Homo sapiens R-HSA-156584 | 6.19E-01 | 9.05E-01 | 9.60E-01 | 4.60E-01 | SLC35B3;SULT2B1;ABHD14B;PAPSS1 |
| 978 | Glycosphingolipid metabolism Homo sapiens R-HSA-1660662 | 6.19E-01 | 9.05E-01 | 9.45E-01 | 4.53E-01 | GLTP;CTSA;ASAH1;GLB1;ARSJ;CPTP;HEXA;NEU1;GLA |

| | | | | | | |
|---|---|---|---|---|---|---|
| 979 | Condensation of Prophase Chromosomes Homo sapiens R-HSA-2299718 | 6.19E-01 | 9.05E-01 | 9.45E-01 | 4.53E-01 | RB1;CCNB1;SET;H2AFZ;H3F3A;NCAPD3;SMC4;HIST1H2AC;MCPH1 |
| 980 | Signal amplification Homo sapiens R-HSA-392518 | 6.24E-01 | 9.12E-01 | 9.41E-01 | 4.43E-01 | GNG10;GNG5;GNAI3;PLA2G4A;GNB5;GNAI1;GNAI2 |
| 981 | Homology Directed Repair Homo sapiens R-HSA-5693538 | 6.27E-01 | 9.15E-01 | 9.50E-01 | 4.43E-01 | MUS81;PCNA;BRCC3;BABAM1;HERC2;PPP4C;PPP4R2;SUMO1;UBC;UIMC1;SUMO2;POLD2;POLK;RPS27A;UBE2I;RFC1;PARP1;RNF168;RAD51B;RAD50;RPA3;ERCC1;CDK2;UBE2V2;RAD17;RAD1 |
| 982 | Metabolism of lipids and lipoproteins Homo sapiens R-HSA-556833 | 6.30E-01 | 9.19E-01 | 9.73E-01 | 4.49E-01 | DMGDH;GLTP;PI4K2B;PECR;FAAH;PITPNB;ECI2;GDE1;RORA;CCNC;MSMO1;PPP1CB;PPP1CC;SCP2;LIPH;OSBP;CPNE1;CYP1B1;CPNE3;PIP4K2C;HADH;ACOT9;ACOT8;ACOT11;MED8;ACSL4;MED4;HSPG2;PISD;CYP39A1;ALDH3A2;FDX1;ALDH7A1;SLC27A5;CSNK1G2;PTGES;IDI1;SAR1B;ABHD3;INSIG2;INSIG1;PIK3R3;MTMR6;ACACA;PGS1;PTDSS2;SGPL1;EBP;MED30;INPP5F;PTPMT1;ARSJ;INPP5E;NEU1;PIP5K1B;HACL1;LTA4H;KDSR;FDFT1;ESRRA;CTSA;FDPS;IDH1;DHCR24;NR1D1;CRLS1;COQ5;SQLE;NCEH1;SLCO1A2;PPT1;SDC1;DEGS1;PPT2;SLC25A1;ARF1;SLC44A3;ACADVL;ALAS1;HEXA;TECR;PRKAG2;AKR1B1;CROT;TAZ;MED11;ARV1;SPTLC1;MED10;ME1;ACADM;LDLRAP1;TEAD2;ELOVL1;NCOA2;CBR1;GPX4;NCOA3;PLA2G4A;ELOVL6;MED27;SREBF2;NCOR2;HADHB;HADHA;NCOR1;MED24;MED21;CHDH;CPTP;PCCB;PLIN3;CERS2;ASAH1;ECHS1;SMARCD3;HDAC3;MVK;HSD17B4;HSD17B12;HSD17B7;ARNTL;ALOX5;HSD17B1;TGS1;SC5D;SEC24A;TXNRD1;PTGES3;AKR1C1;CYP51A1;MBOAT2;AKR1C2;AGT;PPP1CA;VAPA;ETNK2;VAPB;GLB1;ETNK1;CARM1;CHPT1;P4HB;CRAT;GLA;LPIN2;RAN;PNPLA2 |
| 983 | G alpha (z) signalling events Homo sapiens R-HSA-418597 | 6.33E-01 | 9.21E-01 | 9.33E-01 | 4.27E-01 | GNG10;RGS19;GNG5;GNAI3;GNAS;ADCY3;GNB5;GNAI1;ADCY5;GNAI2 |
| 984 | SHC-mediated cascade:FGFR4 Homo sapiens R-HSA-5654719 | 6.34E-01 | 9.22E-01 | 9.34E-01 | 4.25E-01 | FGF18;PTPN11;KRAS;GRB2;FGF2 |

| | | | | | | |
|---|---|---|---|---|---|---|
| 985 | Platelet homeostasis Homo sapiens R-HSA-418346 | 6.37E-01 | 9.25E-01 | 9.39E-01 | 4.24E-01 | PDE1A;ATP2A3;PPP2R5A;PLA2G4A;ITPR2;PTPN11;SRI;ATP2B1;LRP8;PPP2CA;PPP2CB;GNG10;GNG5;PPP2R5E;GNAS;ORAI1;PTPN6;GNB5;CALM1 |
| 986 | Aquaporin-mediated transport Homo sapiens R-HSA-445717 | 6.45E-01 | 9.31E-01 | 9.24E-01 | 4.05E-01 | GNG10;GNG5;PRKAR1A;PRKAR2B;PRKAR2A;GNAS;ADCY3;GNB5;RAB11FIP2;RAB11A;ADCY5 |
| 987 | Organic cation transport Homo sapiens R-HSA-549127 | 6.47E-01 | 9.31E-01 | 9.60E-01 | 4.18E-01 | SLC22A15;RUNX1 |
| 988 | Type I hemidesmosome assembly Homo sapiens R-HSA-446107 | 6.47E-01 | 9.31E-01 | 9.60E-01 | 4.18E-01 | CD151;LAMB3 |
| 989 | Crosslinking of collagen fibrils Homo sapiens R-HSA-2243919 | 6.47E-01 | 9.31E-01 | 9.60E-01 | 4.18E-01 | PCOLCE;LOXL2 |
| 990 | SHC-related events triggered by IGF1R Homo sapiens R-HSA-2428933 | 6.47E-01 | 9.31E-01 | 9.60E-01 | 4.18E-01 | KRAS;GRB2 |
| 991 | CHL1 interactions Homo sapiens R-HSA-447041 | 6.47E-01 | 9.31E-01 | 9.60E-01 | 4.18E-01 | ITGB1;HSPA8 |
| 992 | Role of Abl in Robo-Slit signaling Homo sapiens R-HSA-428890 | 6.47E-01 | 9.31E-01 | 9.60E-01 | 4.18E-01 | CAP1;GPC1 |
| 993 | Pregnenolone biosynthesis Homo sapiens R-HSA-196108 | 6.47E-01 | 9.31E-01 | 9.60E-01 | 4.18E-01 | FDX1;AKR1B1 |
| 994 | Gap junction trafficking Homo sapiens R-HSA-190828 | 6.47E-01 | 9.31E-01 | 9.17E-01 | 3.99E-01 | CLTC;MYO6;CLTB;CLTA;AP2M1;DNM2 |
| 995 | Extension of Telomeres Homo sapiens R-HSA-180786 | 6.47E-01 | 9.31E-01 | 9.17E-01 | 3.99E-01 | PCNA;RFC1;DKC1;RPA3;POLD2;NHP2 |

| | | | | | | |
|---|---|---|---|---|---|---|
| 996 | PI5P, PP2A and IER3 Regulate PI3K/AKT Signaling Homo sapiens R-HSA-6811558 | 6.48E-01 | 9.31E-01 | 9.30E-01 | 4.03E-01 | PDGFB;PPP2R5A;PIK3R3;PTPN11;FGF2;BTC;PPP2CA;PPP2CB;PPP2R5E;FGF18;AKT1;MAPK1;PIP5K1B;FYN;GRB2;PIP4K2C;IER3;HBEGF |
| 997 | Polymerase switching on the C-strand of the telomere Homo sapiens R-HSA-174411 | 6.55E-01 | 9.35E-01 | 9.17E-01 | 3.88E-01 | PCNA;RFC1;POLD2 |
| 998 | Polymerase switching Homo sapiens R-HSA-69091 | 6.55E-01 | 9.35E-01 | 9.17E-01 | 3.88E-01 | PCNA;RFC1;POLD2 |
| 999 | Leading Strand Synthesis Homo sapiens R-HSA-69109 | 6.55E-01 | 9.35E-01 | 9.17E-01 | 3.88E-01 | PCNA;RFC1;POLD2 |
| 1000 | Removal of the Flap Intermediate Homo sapiens R-HSA-69166 | 6.55E-01 | 9.35E-01 | 9.17E-01 | 3.88E-01 | PCNA;RPA3;POLD2 |
| 1001 | Receptor-ligand binding initiates the second proteolytic cleavage of Notch receptor Homo sapiens R-HSA-156988 | 6.55E-01 | 9.35E-01 | 9.17E-01 | 3.88E-01 | JAG2;UBC;RPS27A |
| 1002 | Regulation of necroptotic cell death Homo sapiens R-HSA-5675482 | 6.55E-01 | 9.35E-01 | 9.17E-01 | 3.88E-01 | TRADD;CFLAR;FADD |
| 1003 | Synthesis of Prostaglandins (PG) and Thromboxanes (TX) Homo sapiens R-HSA-2162123 | 6.55E-01 | 9.35E-01 | 9.17E-01 | 3.88E-01 | CBR1;PTGES3;PTGES |

| | | | | | | |
|---|---|---|---|---|---|---|
| 1004 | E2F mediated regulation of DNA replication Homo sapiens R-HSA-113510 | 6.59E-01 | 9.39E-01 | 9.05E-01 | 3.77E-01 | RB1;PPP2CA;PPP2CB;CCNB1;PCNA;MCM8;TK2 |
| 1005 | Phospholipase C-mediated cascade: FGFR1 Homo sapiens R-HSA-5654219 | 6.62E-01 | 9.42E-01 | 9.08E-01 | 3.75E-01 | CREB1;PRKAR2B;PRKAR1A;PDE1A;PRKAR2A;ITPR2;ADCY3;CALM1;FGF2;ADCY5 |
| 1006 | PCNA-Dependent Long Patch Base Excision Repair Homo sapiens R-HSA-5651801 | 6.65E-01 | 9.42E-01 | 8.96E-01 | 3.66E-01 | PCNA;RFC1;RPA3;POLD2 |
| 1007 | Other semaphorin interactions Homo sapiens R-HSA-416700 | 6.65E-01 | 9.42E-01 | 8.96E-01 | 3.66E-01 | ITGB1;SEMA4D;PLXNA2;PLXNA4 |
| 1008 | Prostacyclin signalling through prostacyclin receptor Homo sapiens R-HSA-392851 | 6.65E-01 | 9.42E-01 | 8.96E-01 | 3.66E-01 | GNG10;GNG5;GNAS;GNB5 |
| 1009 | Signaling by NODAL Homo sapiens R-HSA-1181150 | 6.65E-01 | 9.42E-01 | 8.96E-01 | 3.66E-01 | DRAP1;SMAD4;FURIN;TDGF1 |
| 1010 | Transport of vitamins, nucleosides, and related molecules Homo sapiens R-HSA-425397 | 6.70E-01 | 9.48E-01 | 8.96E-01 | 3.59E-01 | SLC35B3;SLC35A1;SLC35A3;SLCO3A1;SLCO4A1;LCN12;SLCO1A2;SLC29A2 |
| 1011 | Transport of Mature mRNAs Derived from Intronless Transcripts Homo sapiens R-HSA-159234 | 6.70E-01 | 9.48E-01 | 8.96E-01 | 3.59E-01 | NUP93;SLBP;FIP1L1;NCBP1;CPSF3;NUP50;TPR;EIF4E |

| | | | | | | |
|---|---|---|---|---|---|---|
| 1012 | Glutamate Neurotransmitter Release Cycle Homo sapiens R-HSA-210500 | 6.74E-01 | 9.51E-01 | 8.84E-01 | 3.49E-01 | SYT1;ARL6IP5;PPFIA1;SLC38A2;GLS |
| 1013 | Growth hormone receptor signaling Homo sapiens R-HSA-982772 | 6.74E-01 | 9.51E-01 | 8.84E-01 | 3.49E-01 | SOCS1;STAT3;MAPK1;PTPN6;JAK2 |
| 1014 | Endogenous sterols Homo sapiens R-HSA-211976 | 6.74E-01 | 9.51E-01 | 8.84E-01 | 3.49E-01 | CYP39A1;NCOA2;CYP51A1;FDX1;CYP1B1 |
| 1015 | Meiotic synapsis Homo sapiens R-HSA-1221632 | 6.82E-01 | 9.55E-01 | 8.96E-01 | 3.43E-01 | UBE2I;STAG2;H2AFZ;RAD21;TINF2;LMNA;TERF2IP;HSPA2;SMC3;SMC1A;SYNE2;HIST1H2AC |
| 1016 | Generic Transcription Pathway Homo sapiens R-HSA-212436 | 6.84E-01 | 9.55E-01 | 9.63E-01 | 3.65E-01 | UBE2D3;EHMT1;UBE2D1;RORA;CCNC;RBPJ;COX6A1;GLS;AKT2;AKT1;ZNF568;ZNF567;BNIP3L;SUPT16H;USP7;ZFP1;CSNK2A2;USP2;MED8;MED4;GTF2F2;NRBP1;COX6B1;RRAGA;COX7A2L;ZNF718;ZNF92;MSH2;SCO2;RRAGB;DDIT4;CSNK2B;MAPKAPK5;ZNF675;COX19;MEAF6;COX16;COX18;MAPKAP1;SUPT4H1;PRDX2;CCNB1;PRDX5;NUAK1;MED30;ZNF706;PCBP4;PRDX1;UBC;ZNF667;ZNF700;COX14;NRBF2;SKIL;ESRRA;SURF1;PRELID1;SMAD4;SMURF2;VDR;GADD45A;ESRRB;NR1D1;ESR2;NR2F6;CNOT4;CDK9;SNW1;CNOT7;TP53I3;CDK5;BCL6;CNOT11;RHEB;CNOT2;AGO2;CNOT3;CDK2;MDM2;PIN1;CYCS;RAD17;MNAT1;COX20;YWHAE;PRR5;GPI;ZNF770;COX7B;RARG;PPP1R13B;CDKN1B;ZNF250;THRA;YWHAB;PHF20;SETD9;COX4I1;PRKAG1;PRKAG2;CHM;CHD4;COX7C;BBC3;YWHAQ;MED10;MLST8;NELFCD;YWHAG;TEAD2;YWHAH;COX8A;NCOA2;CNOT6L;APAF1;PARP1;PDPK1;PRMT1;MED27;RABGGTB;YWHAZ;NCOR2;KAT2B;MED24;NCOR1;RRM2B;TFDP2;TMEM219;RARB;ZNF599;PRKAA1;HDAC2;PCNA;SMARCD3;HDAC1;COX5B;NR2C2;COX5A;NR2C1;PPP2CA;PPP2CB;ZFP14;RBBP4;POLR2B;PERP;TGS1;POLR2C;POLR2E;E2F4;RBBP7;POLR2H;RPS27A;BID;BRD7;POLR2J;POLR2K;POLR2L;NPM1;NDUFA4;TAF13;TAF10;NR1H2;TXNRD1;TAF11;GSR;COX6C;HIPK1;GTF2H4;LRPPRC;MTOR;RAD50;RPA3;CARM1;TCEA1;BAX;RAD1;LAMTOR1;LAMTOR4;LAMTOR3;LAMTOR5;ZNF354B |

| | | | | | | |
|---|---|---|---|---|---|---|
| 1017 | Striated Muscle Contraction Homo sapiens R-HSA-390522 | 6.92E-01 | 9.55E-01 | 8.71E-01 | 3.21E-01 | TPM4;TPM3;TNNC1;TMOD3;TPM1;MYL3;MYH8 |
| 1018 | Resolution of Sister Chromatid Cohesion Homo sapiens R-HSA-2500257 | 6.95E-01 | 9.55E-01 | 9.04E-01 | 3.28E-01 | NUDC;PPP2R5A;PDS5B;SMC3;SMC1A;PMF1;SKA2;CENPC;PPP2CA;PPP1CC;PPP2CB;CLIP1;CCNB1;STAG2;KIF2A;PPP2R5E;RAD21;TAOK1;BUB3;MAPRE1;PAFAH1B1 |
| 1019 | Negative regulation of the PI3K/AKT network Homo sapiens R-HSA-199418 | 6.99E-01 | 9.55E-01 | 8.99E-01 | 3.22E-01 | PDGFB;PPP2R5A;PIK3R3;PTPN11;FGF2;BTC;PPP2CA;PPP2CB;PPP2R5E;AKT2;FGF18;AKT1;MAPK1;PIP5K1B;FYN;GRB2;PIP4K2C;IER3;HBEGF |
| 1020 | NS1 Mediated Effects on Host Pathways Homo sapiens R-HSA-168276 | 7.00E-01 | 9.55E-01 | 8.67E-01 | 3.09E-01 | NUP93;NUP50;TPR;EIF2AK2;KPNA4;KPNA2;KPNA3;KPNA1 |
| 1021 | GABA B receptor activation Homo sapiens R-HSA-977444 | 7.00E-01 | 9.55E-01 | 8.67E-01 | 3.09E-01 | GNG10;GNG5;GNAI3;KCNJ15;ADCY3;GNAI1;ADCY5;GNAI2 |
| 1022 | Activation of GABAB receptors Homo sapiens R-HSA-991365 | 7.00E-01 | 9.55E-01 | 8.67E-01 | 3.09E-01 | GNG10;GNG5;GNAI3;KCNJ15;ADCY3;GNAI1;ADCY5;GNAI2 |
| 1023 | Activation of NMDA receptor upon glutamate binding and postsynaptic events Homo sapiens R-HSA-442755 | 7.00E-01 | 9.55E-01 | 8.67E-01 | 3.09E-01 | CREB1;PDPK1;RASGRF2;AKAP9;ADCY3;MAPK1;CALM1;CAMK2G |
| 1024 | N-Glycan antennae elongation Homo sapiens R-HSA-975577 | 7.03E-01 | 9.55E-01 | 8.40E-01 | 2.96E-01 | B4GALT3;MGAT4B;ST3GAL4 |

| | | | | | | |
|---|---|---|---|---|---|---|
| 1025 | Processive synthesis on the lagging strand Homo sapiens R-HSA-69183 | 7.03E-01 | 9.55E-01 | 8.40E-01 | 2.96E-01 | PCNA;RPA3;POLD2 |
| 1026 | Association of licensing factors with the pre-replicative complex Homo sapiens R-HSA-69298 | 7.03E-01 | 9.55E-01 | 8.40E-01 | 2.96E-01 | GMNN;UBC;RPS27A |
| 1027 | Glycogen synthesis Homo sapiens R-HSA-3322077 | 7.03E-01 | 9.55E-01 | 8.40E-01 | 2.96E-01 | UBC;RPS27A;PGM1 |
| 1028 | ERBB2 Regulates Cell Motility Homo sapiens R-HSA-6785631 | 7.03E-01 | 9.55E-01 | 8.40E-01 | 2.96E-01 | BTC;RHOA;HBEGF |
| 1029 | Recycling of bile acids and salts Homo sapiens R-HSA-159418 | 7.03E-01 | 9.55E-01 | 8.40E-01 | 2.96E-01 | NCOA2;SLCO1A2;SLC27A5 |
| 1030 | Synthesis of bile acids and bile salts via 27-hydroxycholesterol Homo sapiens R-HSA-193807 | 7.03E-01 | 9.55E-01 | 8.40E-01 | 2.96E-01 | NCOA2;AKR1C1;AKR1C2 |
| 1031 | Lagging Strand Synthesis Homo sapiens R-HSA-69186 | 7.06E-01 | 9.55E-01 | 8.40E-01 | 2.92E-01 | PCNA;RFC1;RPA3;POLD2 |
| 1032 | G beta:gamma signalling through PLC beta Homo sapiens R-HSA-418217 | 7.06E-01 | 9.55E-01 | 8.40E-01 | 2.92E-01 | GNG10;GNG5;GNB5;PLCB1 |
| 1033 | Synthesis of PC Homo sapiens R-HSA-1483191 | 7.06E-01 | 9.55E-01 | 8.40E-01 | 2.92E-01 | SLC44A3;ABHD3;CHPT1;LPIN2 |

| | | | | | | |
|---|---|---|---|---|---|---|
| 1034 | Removal of aminoterminal propeptides from gamma-carboxylated proteins Homo sapiens R-HSA-159782 | 7.06E-01 | 9.55E-01 | 8.40E-01 | 2.92E-01 | FURIN;GAS6 |
| 1035 | Activation of Ca-permeable Kainate Receptor Homo sapiens R-HSA-451308 | 7.06E-01 | 9.55E-01 | 8.40E-01 | 2.92E-01 | DLG1;CALM1 |
| 1036 | Ionotropic activity of Kainate Receptors Homo sapiens R-HSA-451306 | 7.06E-01 | 9.55E-01 | 8.40E-01 | 2.92E-01 | DLG1;CALM1 |
| 1037 | Activation of PPARGC1A (PGC-1alpha) by phosphorylation Homo sapiens R-HSA-2151209 | 7.06E-01 | 9.55E-01 | 8.40E-01 | 2.92E-01 | PRKAG1;PRKAG2 |
| 1038 | HDR through MMEJ (alt-NHEJ) Homo sapiens R-HSA-5685939 | 7.06E-01 | 9.55E-01 | 8.40E-01 | 2.92E-01 | RAD50;PARP1 |
| 1039 | E2F-enabled inhibition of pre-replication complex formation Homo sapiens R-HSA-113507 | 7.06E-01 | 9.55E-01 | 8.40E-01 | 2.92E-01 | CCNB1;MCM8 |
| 1040 | Signal attenuation Homo sapiens R-HSA-74749 | 7.06E-01 | 9.55E-01 | 8.40E-01 | 2.92E-01 | MAPK1;GRB2 |

| | | | | | | |
|---|---|---|---|---|---|---|
| 1041 | IRAK2 mediated activation of TAK1 complex upon TLR7/8 or 9 stimulation Homo sapiens R-HSA-975163 | 7.06E-01 | 9.55E-01 | 8.40E-01 | 2.92E-01 | UBC;RPS27A |
| 1042 | IRAK2 mediated activation of TAK1 complex Homo sapiens R-HSA-937042 | 7.06E-01 | 9.55E-01 | 8.40E-01 | 2.92E-01 | UBC;RPS27A |
| 1043 | Diseases of carbohydrate metabolism Homo sapiens R-HSA-5663084 | 7.06E-01 | 9.55E-01 | 8.40E-01 | 2.92E-01 | UBC;RPS27A |
| 1044 | Myoclonic epilepsy of Lafora Homo sapiens R-HSA-3785653 | 7.06E-01 | 9.55E-01 | 8.40E-01 | 2.92E-01 | UBC;RPS27A |
| 1045 | Glycogen storage diseases Homo sapiens R-HSA-3229121 | 7.06E-01 | 9.55E-01 | 8.40E-01 | 2.92E-01 | UBC;RPS27A |
| 1046 | FGFR2b ligand binding and activation Homo sapiens R-HSA-190377 | 7.06E-01 | 9.55E-01 | 8.40E-01 | 2.92E-01 | FGFBP1;FGF2 |
| 1047 | Signaling by FGFR3 fusions in cancer Homo sapiens R-HSA-8853334 | 7.06E-01 | 9.55E-01 | 8.40E-01 | 2.92E-01 | KRAS;GRB2 |
| 1048 | Activation of the AP-1 family of transcription factors Homo sapiens R-HSA-450341 | 7.06E-01 | 9.55E-01 | 8.40E-01 | 2.92E-01 | MAPK9;MAPK1 |

| | | | | | | |
|---|---|---|---|---|---|---|
| 1049 | Role of second messengers in netrin-1 signaling Homo sapiens R-HSA-418890 | 7.06E-01 | 9.55E-01 | 8.40E-01 | 2.92E-01 | PITPNA;NTN1 |
| 1050 | Adenylate cyclase activating pathway Homo sapiens R-HSA-170660 | 7.06E-01 | 9.55E-01 | 8.40E-01 | 2.92E-01 | ADCY3;ADCY5 |
| 1051 | Synthesis of pyrophosphates in the cytosol Homo sapiens R-HSA-1855167 | 7.06E-01 | 9.55E-01 | 8.40E-01 | 2.92E-01 | PPIP5K2;ITPK1 |
| 1052 | Ligand-independent caspase activation via DCC Homo sapiens R-HSA-418889 | 7.06E-01 | 9.55E-01 | 8.40E-01 | 2.92E-01 | MAGED1;APPL1 |
| 1053 | Urea cycle Homo sapiens R-HSA-70635 | 7.06E-01 | 9.55E-01 | 8.40E-01 | 2.92E-01 | ARG2;NMRAL1 |
| 1054 | Recruitment and ATM-mediated phosphorylation of repair and signaling proteins at DNA double strand breaks Homo sapiens R-HSA-5693565 | 7.06E-01 | 9.55E-01 | 8.76E-01 | 3.05E-01 | BABAM1;RNF168;RAD50;HERC2;UBE2I;SUMO1;UIMC1;UBC;UBE2V2;SMARCA5;BRCC3;RPS27A |
| 1055 | Transcriptional regulation by small RNAs Homo sapiens R-HSA-5578749 | 7.08E-01 | 9.55E-01 | 8.84E-01 | 3.06E-01 | H2AFZ;H3F3A;NUP93;POLR2B;NUP50;POLR2C;TPR;AGO2;POLR2E;POLR2H;POLR2J;HIST1H2AC;POLR2K;RAN;POLR2L |
| 1056 | Kinesins Homo sapiens R-HSA-983189 | 7.08E-01 | 9.55E-01 | 8.64E-01 | 2.98E-01 | KIF2A;KIFC2;KIF5C;KIF3A;KIF5B;KIF26B;KIF21A;KLC1;KIFAP3 |

| | | | | | | |
|---|---|---|---|---|---|---|
| 1057 | SUMOylation of DNA replication proteins Homo sapiens R-HSA-4615885 | 7.08E-01 | 9.55E-01 | 8.64E-01 | 2.98E-01 | NUP93;PCNA;UBE2I;SUMO1;SUMO3;NUP50;TPR;SUMO2;TOP1 |
| 1058 | Metal ion SLC transporters Homo sapiens R-HSA-425410 | 7.11E-01 | 9.55E-01 | 8.40E-01 | 2.87E-01 | SLC41A2;SLC40A1;SLC39A7;SLC39A1;SLC39A4 |
| 1059 | Amino acid synthesis and interconversion (transamination) Homo sapiens R-HSA-70614 | 7.11E-01 | 9.55E-01 | 8.40E-01 | 2.87E-01 | GOT1;GOT2;PYCR2;GLUL;GLS |
| 1060 | Synthesis of IP3 and IP4 in the cytosol Homo sapiens R-HSA-1855204 | 7.11E-01 | 9.55E-01 | 8.40E-01 | 2.87E-01 | ITPKB;PLCB4;ITPK1;PLCB1;CALM1 |
| 1061 | Sulfur amino acid metabolism Homo sapiens R-HSA-1614635 | 7.11E-01 | 9.55E-01 | 8.40E-01 | 2.87E-01 | GOT1;ADI1;BHMT2;APIP;ENOPH1 |
| 1062 | Glyoxylate metabolism and glycine degradation Homo sapiens R-HSA-389661 | 7.11E-01 | 9.55E-01 | 8.40E-01 | 2.87E-01 | GRHPR;PDHA1;NDUFAB1;AMT;PDHB |
| 1063 | Activation of Kainate Receptors upon glutamate binding Homo sapiens R-HSA-451326 | 7.16E-01 | 9.55E-01 | 8.40E-01 | 2.80E-01 | GNG10;DLG1;GNG5;GNB5;PLCB1;CALM1 |
| 1064 | Glucagon-type ligand receptors Homo sapiens R-HSA-420092 | 7.16E-01 | 9.55E-01 | 8.40E-01 | 2.80E-01 | GNG10;GNG5;GIPR;SCTR;GNAS;GNB5 |

| | | | | | | |
|---|---|---|---|---|---|---|
| 1065 | Interactions of Rev with host cellular proteins Homo sapiens R-HSA-177243 | 7.22E-01 | 9.55E-01 | 8.40E-01 | 2.73E-01 | NUP93;RANBP1;NPM1;NUP50;TPR;RCC1;RAN |
| 1066 | Fanconi Anemia Pathway Homo sapiens R-HSA-6783310 | 7.22E-01 | 9.55E-01 | 8.40E-01 | 2.73E-01 | MUS81;FANCL;RPA3;ERCC1;UBE2T;UBC;RPS27A |
| 1067 | Heparan sulfate/heparin (HS-GAG) metabolism Homo sapiens R-HSA-1638091 | 7.23E-01 | 9.55E-01 | 8.59E-01 | 2.79E-01 | NAGLU;B3GAT3;GLB1;SDC2;GPC1;SDC1;GPC5;HPSE;HSPG2;B4GALT7;HS3ST1 |
| 1068 | Reversal of alkylation damage by DNA dioxygenases Homo sapiens R-HSA-73943 | 7.28E-01 | 9.55E-01 | 8.40E-01 | 2.67E-01 | ASCC3 |
| 1069 | Sperm:Oocyte Membrane Binding Homo sapiens R-HSA-1300652 | 7.28E-01 | 9.55E-01 | 8.40E-01 | 2.67E-01 | CD9 |
| 1070 | Abacavir metabolism Homo sapiens R-HSA-2161541 | 7.28E-01 | 9.55E-01 | 8.40E-01 | 2.67E-01 | GUK1 |
| 1071 | Na+-dependent glucose transporters Homo sapiens R-HSA-428808 | 7.28E-01 | 9.55E-01 | 8.40E-01 | 2.67E-01 | SLC5A3 |
| 1072 | Sodium-coupled phosphate cotransporters Homo sapiens R-HSA-427652 | 7.28E-01 | 9.55E-01 | 8.40E-01 | 2.67E-01 | SLC20A2 |

| | | | | | | |
|---|---|---|---|---|---|---|
| 1073 | Hyaluronan biosynthesis and export Homo sapiens R-HSA-2142850 | 7.28E-01 | 9.55E-01 | 8.40E-01 | 2.67E-01 | ABCC5 |
| 1074 | Negative regulation of TCF-dependent signaling by DVL-interacting proteins Homo sapiens R-HSA-5368598 | 7.28E-01 | 9.55E-01 | 8.40E-01 | 2.67E-01 | DVL3 |
| 1075 | Synthesis of GDP-mannose Homo sapiens R-HSA-446205 | 7.28E-01 | 9.55E-01 | 8.40E-01 | 2.67E-01 | GMPPB |
| 1076 | G2 Phase Homo sapiens R-HSA-68911 | 7.28E-01 | 9.55E-01 | 8.40E-01 | 2.67E-01 | CDK2 |
| 1077 | PTK6 Expression Homo sapiens R-HSA-8849473 | 7.28E-01 | 9.55E-01 | 8.40E-01 | 2.67E-01 | HIF1A |
| 1078 | GRB7 events in ERBB2 signaling Homo sapiens R-HSA-1306955 | 7.28E-01 | 9.55E-01 | 8.40E-01 | 2.67E-01 | GRB7 |
| 1079 | FasL/ CD95L signaling Homo sapiens R-HSA-75157 | 7.28E-01 | 9.55E-01 | 8.40E-01 | 2.67E-01 | FADD |
| 1080 | Cyclin B2 mediated events Homo sapiens R-HSA-157881 | 7.28E-01 | 9.55E-01 | 8.40E-01 | 2.67E-01 | CDC25B |
| 1081 | G2/M DNA replication checkpoint Homo sapiens R-HSA-69478 | 7.28E-01 | 9.55E-01 | 8.40E-01 | 2.67E-01 | CCNB1 |

| | | | | | | |
|---|---|---|---|---|---|---|
| 1082 | SMAC binds to IAPs Homo sapiens R-HSA-111463 | 7.28E-01 | 9.55E-01 | 8.40E-01 | 2.67E-01 | CASP7 |
| 1083 | SMAC-mediated dissociation of IAP:caspase complexes Homo sapiens R-HSA-111464 | 7.28E-01 | 9.55E-01 | 8.40E-01 | 2.67E-01 | CASP7 |
| 1084 | SMAC-mediated apoptotic response Homo sapiens R-HSA-111469 | 7.28E-01 | 9.55E-01 | 8.40E-01 | 2.67E-01 | CASP7 |
| 1085 | Arachidonate production from DAG Homo sapiens R-HSA-426048 | 7.28E-01 | 9.55E-01 | 8.40E-01 | 2.67E-01 | DAGLA |
| 1086 | Vitamin B2 (riboflavin) metabolism Homo sapiens R-HSA-196843 | 7.28E-01 | 9.55E-01 | 8.40E-01 | 2.67E-01 | RFK |
| 1087 | Propionyl-CoA catabolism Homo sapiens R-HSA-71032 | 7.28E-01 | 9.55E-01 | 8.40E-01 | 2.67E-01 | PCCB |
| 1088 | Estrogen biosynthesis Homo sapiens R-HSA-193144 | 7.28E-01 | 9.55E-01 | 8.40E-01 | 2.67E-01 | HSD17B1 |
| 1089 | The fatty acid cycling model Homo sapiens R-HSA-167826 | 7.28E-01 | 9.55E-01 | 8.40E-01 | 2.67E-01 | UCP2 |
| 1090 | The proton buffering model Homo sapiens R-HSA-167827 | 7.28E-01 | 9.55E-01 | 8.40E-01 | 2.67E-01 | UCP2 |
| 1091 | Mitochondrial Uncoupling Proteins Homo sapiens R-HSA-166187 | 7.28E-01 | 9.55E-01 | 8.40E-01 | 2.67E-01 | UCP2 |

| | | | | | | |
|---|---|---|---|---|---|---|
| 1092 | ABC-family proteins mediated transport Homo sapiens R-HSA-382556 | 7.34E-01 | 9.62E-01 | 8.40E-01 | 2.59E-01 | ABCD3;ABCB7;PEX3;ABCC5;ABCA3;ABCA4;EIF2S2;EIF2S1;ABCF1 |
| 1093 | ERK/MAPK targets Homo sapiens R-HSA-198753 | 7.44E-01 | 9.70E-01 | 7.91E-01 | 2.34E-01 | PPP2CA;PPP2CB;DUSP3;MAPK1 |
| 1094 | Presynaptic function of Kainate receptors Homo sapiens R-HSA-500657 | 7.44E-01 | 9.70E-01 | 7.91E-01 | 2.34E-01 | GNG10;GNG5;GNB5;PLCB1 |
| 1095 | Diseases associated with glycosaminoglycan metabolism Homo sapiens R-HSA-3560782 | 7.45E-01 | 9.70E-01 | 8.00E-01 | 2.36E-01 | SDC2;GPC1;SDC1;GPC5;HSPG2 |
| 1096 | SHC-mediated cascade:FGFR2 Homo sapiens R-HSA-5654699 | 7.45E-01 | 9.70E-01 | 8.00E-01 | 2.36E-01 | FGF18;PTPN11;KRAS;GRB2;FGF2 |
| 1097 | TNFR1-induced NFkappaB signaling pathway Homo sapiens R-HSA-5357956 | 7.45E-01 | 9.70E-01 | 8.00E-01 | 2.36E-01 | SHARPIN;TRADD;USP2;RBCK1;TNF |
| 1098 | PI3K events in ERBB2 signaling Homo sapiens R-HSA-1963642 | 7.46E-01 | 9.70E-01 | 7.75E-01 | 2.27E-01 | BTC;GRB2;HBEGF |
| 1099 | Regulated Necrosis Homo sapiens R-HSA-5218859 | 7.46E-01 | 9.70E-01 | 7.75E-01 | 2.27E-01 | TRADD;CFLAR;FADD |

| | | | | | | |
|---|---|---|---|---|---|---|
| 1100 | RIPK1-mediated regulated necrosis Homo sapiens R-HSA-5213460 | 7.46E-01 | 9.70E-01 | 7.75E-01 | 2.27E-01 | TRADD;CFLAR;FADD |
| 1101 | Polo-like kinase mediated events Homo sapiens R-HSA-156711 | 7.46E-01 | 9.70E-01 | 7.75E-01 | 2.27E-01 | CCNB1;LIN37;RBBP4 |
| 1102 | HS-GAG biosynthesis Homo sapiens R-HSA-2022928 | 7.47E-01 | 9.70E-01 | 8.06E-01 | 2.35E-01 | SDC2;GPC1;SDC1;GPC5;HSPG2;HS3ST1 |
| 1103 | Gamma-carboxylation, transport, and amino-terminal cleavage of proteins Homo sapiens R-HSA-159854 | 7.57E-01 | 9.75E-01 | 7.47E-01 | 2.08E-01 | FURIN;GAS6 |
| 1104 | Import of palmitoyl-CoA into the mitochondrial matrix Homo sapiens R-HSA-200425 | 7.57E-01 | 9.75E-01 | 7.47E-01 | 2.08E-01 | PRKAG2;ACACA |
| 1105 | Signaling by FGFR4 in disease Homo sapiens R-HSA-5655291 | 7.57E-01 | 9.75E-01 | 7.47E-01 | 2.08E-01 | KRAS;GRB2 |
| 1106 | Interleukin-7 signaling Homo sapiens R-HSA-1266695 | 7.57E-01 | 9.75E-01 | 7.47E-01 | 2.08E-01 | PIK3R3;JAK1 |
| 1107 | Sodium/Calcium exchangers Homo sapiens R-HSA-425561 | 7.57E-01 | 9.75E-01 | 7.47E-01 | 2.08E-01 | SRI;CALM1 |

| | | | | | | |
|---|---|---|---|---|---|---|
| 1108 | Defects in cobalamin (B12) metabolism Homo sapiens R-HSA-3296469 | 7.57E-01 | 9.75E-01 | 7.47E-01 | 2.08E-01 | MMADHC;CD320 |
| 1109 | Tryptophan catabolism Homo sapiens R-HSA-71240 | 7.57E-01 | 9.75E-01 | 7.47E-01 | 2.08E-01 | AFMID;KMO |
| 1110 | Phenylalanine and tyrosine catabolism Homo sapiens R-HSA-71182 | 7.57E-01 | 9.75E-01 | 7.47E-01 | 2.08E-01 | QDPR;PCBD1 |
| 1111 | Nuclear Envelope Breakdown Homo sapiens R-HSA-2980766 | 7.63E-01 | 9.81E-01 | 8.19E-01 | 2.21E-01 | NUP93;CCNB1;NUP50;CNEP1R1;LMNA;TPR;NEK7;LEMD2;LPIN2;TMPO |
| 1112 | Cell-cell junction organization Homo sapiens R-HSA-421270 | 7.72E-01 | 9.81E-01 | 8.23E-01 | 2.13E-01 | CLDN10;CLDN4;PARD6B;CLDN3;PARD6A;JUP;CADM2;CLDN12;CLDN7;CLDN19;CDH13;CLDN1 |
| 1113 | Muscle contraction Homo sapiens R-HSA-397014 | 7.74E-01 | 9.81E-01 | 8.88E-01 | 2.28E-01 | KCNK5;ATP2A3;ITPR2;MYL6B;CACNA1D;CACNA1C;ATP1A1;SRI;MYL12A;MYLK;MYL12B;NPPC;CAMK2G;DMPK;ANXA2;TPM4;TPM3;TNNC1;TMOD3;TPM1;CACNA2D3;KCNK15;ATP1B3;ATP2B1;HIPK1;ATP1B1;KAT2B;MYL6;CACNB4;ASPH;FXYD3;KCNQ1;FXYD2;MYL3;AKAP9;FKBP1B;MYH8;ORAI1;RANGRF;TLN1;CALM1 |
| 1114 | Interleukin-6 family signaling Homo sapiens R-HSA-6783589 | 7.75E-01 | 9.81E-01 | 7.64E-01 | 1.94E-01 | STAT3;PTPN11;CBL;JAK2;JAK1 |
| 1115 | Caspase activation via extrinsic apoptotic signalig pathway Homo sapiens R-HSA-5357769 | 7.75E-01 | 9.81E-01 | 7.64E-01 | 1.94E-01 | MAGED1;TRADD;FADD;CFLAR;APPL1 |

| | | | | | | |
|---|---|---|---|---|---|---|
| 1116 | Regulation of beta-cell development Homo sapiens R-HSA-186712 | 7.75E-01 | 9.81E-01 | 7.75E-01 | 1.97E-01 | AKT2;AKT1;RBPJ;MAFA;GCK;FOXA3 |
| 1117 | Nuclear import of Rev protein Homo sapiens R-HSA-180746 | 7.75E-01 | 9.81E-01 | 7.75E-01 | 1.97E-01 | NUP93;NPM1;NUP50;TPR;RCC1;RAN |
| 1118 | Vpr-mediated nuclear import of PICs Homo sapiens R-HSA-180910 | 7.75E-01 | 9.81E-01 | 7.75E-01 | 1.97E-01 | NUP93;NUP50;TPR;HMGA1;PSIP1;KPNA1 |
| 1119 | TRAF6 mediated IRF7 activation Homo sapiens R-HSA-933541 | 7.75E-01 | 9.81E-01 | 7.75E-01 | 1.97E-01 | MAVS;SIKE1;TBK1;IRF3;IRF2;TANK |
| 1120 | Thrombin signalling through proteinase activated receptors (PARs) Homo sapiens R-HSA-456926 | 7.75E-01 | 9.81E-01 | 7.75E-01 | 1.97E-01 | GNG10;GNG5;F2R;MAPK1;ARRB1;GNB5 |
| 1121 | O-linked glycosylation of mucins Homo sapiens R-HSA-913709 | 7.77E-01 | 9.81E-01 | 8.24E-01 | 2.08E-01 | GALNT6;GALNT3;GALNT18;GALNT1;C1GALT1;C1GALT1C1;MUC1;B3GNT3;ST3GAL4;B3GNT2;MUC15;MUC20;GALNT9 |
| 1122 | Triglyceride Biosynthesis Homo sapiens R-HSA-75109 | 7.77E-01 | 9.81E-01 | 8.24E-01 | 2.08E-01 | SLC25A1;ACOT9;ELOVL1;ACOT8;TECR;ACOT11;ACSL4;ELOVL6;HSD17B12;ACACA;PPT1;PPT2;LPIN2 |
| 1123 | Transport of Mature mRNA Derived from an Intronless Transcript Homo sapiens R-HSA-159231 | 7.77E-01 | 9.81E-01 | 7.84E-01 | 1.98E-01 | NUP93;FIP1L1;NCBP1;CPSF3;NUP50;TPR;EIF4E |
| 1124 | Collagen formation Homo sapiens R-HSA-1474290 | 7.77E-01 | 9.81E-01 | 8.40E-01 | 2.11E-01 | CRTAP;COL26A1;CD151;LAMB3;COL14A1;COL12A1;PLOD2;PCOLCE;LOXL2;P4HA1;CTSL;P4HA2;COL4A4;COL4A3;P4HB;PPIB;CTSB |

| | | | | | | |
|---|---|---|---|---|---|---|
| 1125 | Meiosis Homo sapiens R-HSA-1500620 | 7.77E-01 | 9.81E-01 | 8.40E-01 | 2.11E-01 | UBE2I;H2AFZ;TINF2;H3F3A;HSPA2;SMC3;SMC1A;SYNE2;RAD50;STAG2;CDK4;RAD21;RPA3;LMNA;CDK2;TERF2IP;HIST1H2AC |
| 1126 | Telomere C-strand (Lagging Strand) Synthesis Homo sapiens R-HSA-174417 | 7.78E-01 | 9.81E-01 | 7.47E-01 | 1.88E-01 | PCNA;RFC1;RPA3;POLD2 |
| 1127 | Signaling by FGFR3 point mutants in cancer Homo sapiens R-HSA-8853338 | 7.78E-01 | 9.81E-01 | 7.47E-01 | 1.88E-01 | FGF18;KRAS;GRB2;FGF2 |
| 1128 | Signaling by FGFR3 in disease Homo sapiens R-HSA-5655332 | 7.78E-01 | 9.81E-01 | 7.47E-01 | 1.88E-01 | FGF18;KRAS;GRB2;FGF2 |
| 1129 | Zinc transporters Homo sapiens R-HSA-435354 | 7.84E-01 | 9.81E-01 | 7.20E-01 | 1.75E-01 | SLC39A7;SLC39A1;SLC39A4 |
| 1130 | ABC transporters in lipid homeostasis Homo sapiens R-HSA-1369062 | 7.84E-01 | 9.81E-01 | 7.20E-01 | 1.75E-01 | ABCD3;PEX3;ABCA3 |
| 1131 | Ligand-dependent caspase activation Homo sapiens R-HSA-140534 | 7.84E-01 | 9.81E-01 | 7.20E-01 | 1.75E-01 | TRADD;CFLAR;FADD |
| 1132 | Peptide hormone metabolism Homo sapiens R-HSA-2980736 | 7.90E-01 | 9.81E-01 | 8.27E-01 | 1.95E-01 | CTSZ;ATP6AP2;MYO5A;RAB27A;AGT;DPP4;SPCS3;SPCS2;SPCS1;KIF5C;KIF5B;CPE;EXOC6;CTSD;SEC11A;SEC11C |

| | | | | | | |
|---|---|---|---|---|---|---|
| 1133 | Neurotransmitter Receptor Binding And Downstream Transmission In The Postsynaptic Cell Homo sapiens R-HSA-112314 | 7.90E-01 | 9.81E-01 | 8.62E-01 | 2.03E-01 | CHRNA5;RASGRF2;GNAI3;ADCY3;AKAP5;AP2A2;GNAI1;ADCY5;GNAI2;GNG10;GNG5;TSPAN7;MYO6;AP2S1;MAPK1;CAMK2G;AP2M1;NSF;PDPK1;KCNJ15;AP2B1;DLG1;CREB1;AKAP9;MDM2;PICK1;GNB5;CALM1;PLCB1 |
| 1134 | Fructose metabolism Homo sapiens R-HSA-5652084 | 7.90E-01 | 9.81E-01 | 6.72E-01 | 1.58E-01 | AKR1B1 |
| 1135 | Synthesis of Dolichyl-phosphate Homo sapiens R-HSA-446199 | 7.90E-01 | 9.81E-01 | 6.72E-01 | 1.58E-01 | DOLK |
| 1136 | Pre-NOTCH Processing in the Endoplasmic Reticulum Homo sapiens R-HSA-1912399 | 7.90E-01 | 9.81E-01 | 6.72E-01 | 1.58E-01 | POGLUT1 |
| 1137 | FGFR1b ligand binding and activation Homo sapiens R-HSA-190370 | 7.90E-01 | 9.81E-01 | 6.72E-01 | 1.58E-01 | FGF2 |
| 1138 | TNFR1-mediated ceramide production Homo sapiens R-HSA-5626978 | 7.90E-01 | 9.81E-01 | 6.72E-01 | 1.58E-01 | TNF |
| 1139 | Phosphorylation of Emi1 Homo sapiens R-HSA-176417 | 7.90E-01 | 9.81E-01 | 6.72E-01 | 1.58E-01 | CCNB1 |

| | | | | | | |
|---|---|---|---|---|---|---|
| 1140 | Misspliced LRP5 mutants have enhanced beta-catenin-dependent signaling Homo sapiens R-HSA-5339717 | 7.90E-01 | 9.81E-01 | 6.72E-01 | 1.58E-01 | DKK1 |
| 1141 | Fibronectin matrix formation Homo sapiens R-HSA-1566977 | 7.90E-01 | 9.81E-01 | 6.72E-01 | 1.58E-01 | ITGB1 |
| 1142 | Proton-coupled monocarboxylate transport Homo sapiens R-HSA-433692 | 7.90E-01 | 9.81E-01 | 6.72E-01 | 1.58E-01 | SLC16A3 |
| 1143 | LDL-mediated lipid transport Homo sapiens R-HSA-171052 | 7.90E-01 | 9.81E-01 | 6.72E-01 | 1.58E-01 | LDLRAP1 |
| 1144 | NADE modulates death signalling Homo sapiens R-HSA-205025 | 7.90E-01 | 9.81E-01 | 6.72E-01 | 1.58E-01 | YWHAE |
| 1145 | SMAD2/3 Phosphorylation Motif Mutants in Cancer Homo sapiens R-HSA-3304356 | 7.90E-01 | 9.81E-01 | 6.72E-01 | 1.58E-01 | TGFB1 |
| 1146 | Molybdenum cofactor biosynthesis Homo sapiens R-HSA-947581 | 7.90E-01 | 9.81E-01 | 6.72E-01 | 1.58E-01 | MOCS2 |

| | | | | | | |
|---|---|---|---|---|---|---|
| 1147 | Metabolic disorders of biological oxidation enzymes Homo sapiens R-HSA-5579029 | 7.90E-01 | 9.81E-01 | 6.72E-01 | 1.58E-01 | FDX1 |
| 1148 | Heme degradation Homo sapiens R-HSA-189483 | 7.90E-01 | 9.81E-01 | 6.72E-01 | 1.58E-01 | HMOX2 |
| 1149 | Synthesis of Lipoxins (LX) Homo sapiens R-HSA-2142700 | 7.90E-01 | 9.81E-01 | 6.72E-01 | 1.58E-01 | ALOX5 |
| 1150 | Synthesis of 15-eicosatetraenoic acid derivatives Homo sapiens R-HSA-2142770 | 7.90E-01 | 9.81E-01 | 6.72E-01 | 1.58E-01 | GPX4 |
| 1151 | Synthesis of 12-eicosatetraenoic acid derivatives Homo sapiens R-HSA-2142712 | 7.90E-01 | 9.81E-01 | 6.72E-01 | 1.58E-01 | GPX4 |
| 1152 | Pyrimidine biosynthesis Homo sapiens R-HSA-500753 | 7.90E-01 | 9.81E-01 | 6.72E-01 | 1.58E-01 | DUT |
| 1153 | Synthesis of wybutosine at G37 of tRNA(Phe) Homo sapiens R-HSA-6782861 | 7.90E-01 | 9.81E-01 | 6.72E-01 | 1.58E-01 | TYW1 |
| 1154 | Phase II conjugation Homo sapiens R-HSA-156580 | 7.91E-01 | 9.81E-01 | 8.40E-01 | 1.97E-01 | SLC35B3;GSTM4;GSTK1;GSTM3;TPMT;GSTO2;GSTO1;MGST3;GSTP1;MGST2;ABHD14B;COMT;PAPSS1;SULT2B1;GGCT;TRMT112;NAT2;ESD;GSTT2B;GSTM5 |

| | | | | | | |
|---|---|---|---|---|---|---|
| 1155 | Oxidative Stress Induced Senescence Homo sapiens R-HSA-2559580 | 7.98E-01 | 9.83E-01 | 8.28E-01 | 1.87E-01 | MAP2K3;SUZ12;CDKN2D;PHC2;CBX4;H2AFZ;H3F3A;MAPK9;CDK6;RBBP4;CDK4;UBC;MDM2;MAPK1;MAPKAPK5;RBBP7;RPS27A;HIST1H2AC |
| 1156 | Erythrocytes take up carbon dioxide and release oxygen Homo sapiens R-HSA-1237044 | 7.99E-01 | 9.83E-01 | 6.72E-01 | 1.50E-01 | CYB5R1;CA2 |
| 1157 | O2/CO2 exchange in erythrocytes Homo sapiens R-HSA-1480926 | 7.99E-01 | 9.83E-01 | 6.72E-01 | 1.50E-01 | CYB5R1;CA2 |
| 1158 | Reversible hydration of carbon dioxide Homo sapiens R-HSA-1475029 | 7.99E-01 | 9.83E-01 | 6.72E-01 | 1.50E-01 | CA5B;CA2 |
| 1159 | Keratan sulfate degradation Homo sapiens R-HSA-2022857 | 7.99E-01 | 9.83E-01 | 6.72E-01 | 1.50E-01 | GLB1;HEXA |
| 1160 | TP53 Regulates Transcription of Death Receptors and Ligands Homo sapiens R-HSA-6803211 | 7.99E-01 | 9.83E-01 | 6.72E-01 | 1.50E-01 | PPP1R13B;TMEM219 |
| 1161 | NOTCH2 intracellular domain regulates transcription Homo sapiens R-HSA-2197563 | 7.99E-01 | 9.83E-01 | 6.72E-01 | 1.50E-01 | CREB1;RBPJ |

| | | | | | | |
|---|---|---|---|---|---|---|
| 1162 | FGFR3 mutant receptor activation Homo sapiens R-HSA-2033514 | 7.99E-01 | 9.83E-01 | 6.72E-01 | 1.50E-01 | FGF18;FGF2 |
| 1163 | Signaling by activated point mutants of FGFR3 Homo sapiens R-HSA-1839130 | 7.99E-01 | 9.83E-01 | 6.72E-01 | 1.50E-01 | FGF18;FGF2 |
| 1164 | NF-kB activation through FADD/RIP-1 pathway mediated by caspase-8 and -10 Homo sapiens R-HSA-933543 | 7.99E-01 | 9.83E-01 | 6.72E-01 | 1.50E-01 | MAVS;FADD |
| 1165 | Rev-mediated nuclear export of HIV RNA Homo sapiens R-HSA-165054 | 8.01E-01 | 9.84E-01 | 7.47E-01 | 1.65E-01 | NUP93;RANBP1;NUP50;TPR;RCC1;RAN |
| 1166 | Role of LAT2/NTAL/LAB on calcium mobilization Homo sapiens R-HSA-2730905 | 8.07E-01 | 9.90E-01 | 8.59E-01 | 1.84E-01 | PRR5;GSK3B;GSK3A;CDKN1B;MAPKAP1;PDGFB;PIK3R3;FGF2;PPP2CA;PPP2CB;PPP2R5E;AKT2;MLST8;AKT1;MAPK1;PIP5K1B;FYN;PIP4K2C;IER3;PDPK1;BAD;PPP2R5A;PTPN11;MTOR;BTC;CREB1;FGF18;AGO2;AKT1S1;RPS6KB2;MDM2;GRB2;HBEGF |
| 1167 | Laminin interactions Homo sapiens R-HSA-3000157 | 8.08E-01 | 9.90E-01 | 7.07E-01 | 1.51E-01 | ITGB1;LAMB3;LAMB2;HSPG2 |
| 1168 | Collagen biosynthesis and modifying enzymes Homo sapiens R-HSA-1650814 | 8.10E-01 | 9.93E-01 | 7.90E-01 | 1.66E-01 | CRTAP;COL26A1;P4HA1;COL14A1;P4HA2;COL4A4;COL12A1;COL4A3;PLOD2;PCOLCE;P4HB;PPIB |
| 1169 | Amyloid fiber formation Homo sapiens R-HSA-977225 | 8.13E-01 | 9.94E-01 | 7.94E-01 | 1.64E-01 | APP;CALCA;GSN;H2AFZ;H3F3A;UBE2L6;FURIN;HSPG2;GGA1;UBC;RPS27A;HIST1H2AC;ITM2B |

| | | | | | | |
|---|---|---|---|---|---|---|
| 1170 | Visual phototransduction Homo sapiens R-HSA-2187338 | 8.17E-01 | 9.94E-01 | 8.18E-01 | 1.66E-01 | SDC2;AKR1C1;ABCA4;MYO7A;HSPG2;LRP8;CAMKMT;RDH11;RDH10;HSD17B1;GPC1;NMT1;NAPEPLD;SDC1;GPC5;GNB5;BCO2;LRP10;CALM1 |
| 1171 | Defective GALNT12 causes colorectal cancer 1 (CRCS1) Homo sapiens R-HSA-5083636 | 8.17E-01 | 9.94E-01 | 6.72E-01 | 1.36E-01 | MUC1;MUC15;MUC20 |
| 1172 | Defective GALNT3 causes familial hyperphosphatemic tumoral calcinosis (HFTC) Homo sapiens R-HSA-5083625 | 8.17E-01 | 9.94E-01 | 6.72E-01 | 1.36E-01 | MUC1;MUC15;MUC20 |
| 1173 | Synthesis of PIPs at the Golgi membrane Homo sapiens R-HSA-1660514 | 8.17E-01 | 9.94E-01 | 6.72E-01 | 1.36E-01 | PI4K2B;ARF1;INPP5E |
| 1174 | Cardiac conduction Homo sapiens R-HSA-5576891 | 8.19E-01 | 9.94E-01 | 8.39E-01 | 1.67E-01 | KCNK5;ATP2A3;ITPR2;CACNA1D;CACNA1C;ATP1A1;SRI;NPPC;CAMK2G;DMPK;CACNA2D3;KCNK15;ATP1B3;ATP2B1;ATP1B1;HIPK1;KAT2B;CACNB4;ASPH;FXYD3;KCNQ1;FXYD2;AKAP9;FKBP1B;ORAI1;RANGRF;CALM1 |
| 1175 | Transport of the SLBP Dependant Mature mRNA Homo sapiens R-HSA-159230 | 8.25E-01 | 9.94E-01 | 7.20E-01 | 1.39E-01 | NUP93;SLBP;NCBP1;NUP50;TPR;EIF4E |
| 1176 | DNA Damage/Telomere Stress Induced Senescence Homo sapiens R-HSA-2559586 | 8.26E-01 | 9.94E-01 | 7.70E-01 | 1.48E-01 | RB1;RAD50;H1F0;CDKN1B;H2AFZ;TINF2;CDK2;HMGA1;HIST1H1E;TERF2IP;HIST1H2AC |

| | | | | | | |
|---|---|---|---|---|---|---|
| 1177 | HDR through Homologous Recombination (HRR) Homo sapiens R-HSA-5685942 | 8.27E-01 | 9.94E-01 | 7.75E-01 | 1.47E-01 | MUS81;RAD51B;RAD50;PCNA;RFC1;RPA3;UBC;POLD2;RAD17;RAD1;POLK;RPS27A |
| 1178 | Interferon gamma signaling Homo sapiens R-HSA-877300 | 8.27E-01 | 9.94E-01 | 8.06E-01 | 1.53E-01 | SP100;IFNGR2;HLA-C;PTPN11;HLA-G;HLA-E;SOCS1;SUMO1;IRF3;TRIM5;TRIM2;IRF2;IRF5;PTPN6;JAK2;CAMK2G;JAK1;HLA-DQB1 |
| 1179 | Surfactant metabolism Homo sapiens R-HSA-5683826 | 8.28E-01 | 9.94E-01 | 7.00E-01 | 1.32E-01 | ABCA3;CCDC59;CTSH;ZDHHC2;LMCD1 |
| 1180 | Myogenesis Homo sapiens R-HSA-525793 | 8.28E-01 | 9.94E-01 | 7.00E-01 | 1.32E-01 | CDC42;SPAG9;BOC;BNIP2;TCF4 |
| 1181 | CDO in myogenesis Homo sapiens R-HSA-375170 | 8.28E-01 | 9.94E-01 | 7.00E-01 | 1.32E-01 | CDC42;SPAG9;BOC;BNIP2;TCF4 |
| 1182 | Transmission across Chemical Synapses Homo sapiens R-HSA-112315 | 8.33E-01 | 9.94E-01 | 8.59E-01 | 1.57E-01 | CHRNA5;RASGRF2;GNAI3;ADCY3;AKAP5;COMT;LIN7C;AP2A2;GNAI1;GLS;ADCY5;GNAI2;GNG10;GNG5;TSPAN7;MYO6;ARL6IP5;AP2S1;PPFIA1;MAPK1;GLUL;SLC38A2;CAMK2G;AP2M1;NSF;HSPA8;SYT1;PDPK1;CACFD1;CACNA2D3;KCNJ15;SLC6A12;AP2B1;DLG1;CACNB4;CREB1;AKAP9;NAAA;MDM2;PICK1;GNB5;CALM1;PLCB1 |
| 1183 | Diseases associated with the TLR signaling cascade Homo sapiens R-HSA-5602358 | 8.35E-01 | 9.94E-01 | 6.72E-01 | 1.22E-01 | NFKBIA;IRAK4;TLR5;NFKB1 |
| 1184 | Diseases of Immune System Homo sapiens R-HSA-5260271 | 8.35E-01 | 9.94E-01 | 6.72E-01 | 1.22E-01 | NFKBIA;IRAK4;TLR5;NFKB1 |
| 1185 | FGFRL1 modulation of FGFR1 signaling Homo sapiens R-HSA-5658623 | 8.35E-01 | 9.94E-01 | 6.11E-01 | 1.10E-01 | FGF18;FGF2 |

| | | | | | | |
|---|---|---|---|---|---|---|
| 1186 | FGFR2c ligand binding and activation Homo sapiens R-HSA-190375 | 8.35E-01 | 9.94E-01 | 6.11E-01 | 1.10E-01 | FGF18;FGF2 |
| 1187 | ERBB2 Activates PTK6 Signaling Homo sapiens R-HSA-8847993 | 8.35E-01 | 9.94E-01 | 6.11E-01 | 1.10E-01 | BTC;HBEGF |
| 1188 | Platelet Adhesion to exposed collagen Homo sapiens R-HSA-75892 | 8.35E-01 | 9.94E-01 | 6.11E-01 | 1.10E-01 | ITGB1;FYN |
| 1189 | Cation-coupled Chloride cotransporters Homo sapiens R-HSA-426117 | 8.39E-01 | 9.94E-01 | 5.60E-01 | 9.86E-02 | SLC12A2 |
| 1190 | Transport of nucleosides and free purine and pyrimidine bases across the plasma membrane Homo sapiens R-HSA-83936 | 8.39E-01 | 9.94E-01 | 5.60E-01 | 9.86E-02 | SLC29A2 |
| 1191 | SLC transporter disorders Homo sapiens R-HSA-5619102 | 8.39E-01 | 9.94E-01 | 5.60E-01 | 9.86E-02 | GCK |
| 1192 | POLB-Dependent Long Patch Base Excision Repair Homo sapiens R-HSA-110362 | 8.39E-01 | 9.94E-01 | 5.60E-01 | 9.86E-02 | PARP1 |

| | | | | | | |
|---|---|---|---|---|---|---|
| 1193 | APEX1-Independent Resolution of AP Sites via the Single Nucleotide Replacement Pathway Homo sapiens R-HSA-5649702 | 8.39E-01 | 9.94E-01 | 5.60E-01 | 9.86E-02 | NEIL1 |
| 1194 | Anchoring fibril formation Homo sapiens R-HSA-2214320 | 8.39E-01 | 9.94E-01 | 5.60E-01 | 9.86E-02 | LAMB3 |
| 1195 | GLI proteins bind promoters of Hh responsive genes to promote transcription Homo sapiens R-HSA-5635851 | 8.39E-01 | 9.94E-01 | 5.60E-01 | 9.86E-02 | BOC |
| 1196 | Neurofascin interactions Homo sapiens R-HSA-447043 | 8.39E-01 | 9.94E-01 | 5.60E-01 | 9.86E-02 | SDCBP |
| 1197 | NrCAM interactions Homo sapiens R-HSA-447038 | 8.39E-01 | 9.94E-01 | 5.60E-01 | 9.86E-02 | DLG1 |
| 1198 | Regulation of gene expression in late stage (branching morphogenesis) pancreatic bud precursor cells Homo sapiens R-HSA-210744 | 8.39E-01 | 9.94E-01 | 5.60E-01 | 9.86E-02 | RBPJ |

| | | | | | | |
|---|---|---|---|---|---|---|
| 1199 | Signaling by NOTCH1 t(7;9)(NOTCH1:M1580 K2555) Translocation Mutant Homo sapiens R-HSA-2660825 | 8.39E-01 | 9.94E-01 | 5.60E-01 | 9.86E-02 | JAG2 |
| 1200 | Constitutive Signaling by NOTCH1 t(7;9)(NOTCH1:M1580 K2555) Translocation Mutant Homo sapiens R-HSA-2660826 | 8.39E-01 | 9.94E-01 | 5.60E-01 | 9.86E-02 | JAG2 |
| 1201 | FGFR3b ligand binding and activation Homo sapiens R-HSA-190371 | 8.39E-01 | 9.94E-01 | 5.60E-01 | 9.86E-02 | FGF18 |
| 1202 | Post-transcriptional silencing by small RNAs Homo sapiens R-HSA-426496 | 8.39E-01 | 9.94E-01 | 5.60E-01 | 9.86E-02 | AGO2 |
| 1203 | Binding of TCF/LEF:CTNNB1 to target gene promoters Homo sapiens R-HSA-4411364 | 8.39E-01 | 9.94E-01 | 5.60E-01 | 9.86E-02 | TCF7L1 |
| 1204 | Glycerophospholipid catabolism Homo sapiens R-HSA-6814848 | 8.39E-01 | 9.94E-01 | 5.60E-01 | 9.86E-02 | GDE1 |
| 1205 | Interleukin-1 processing Homo sapiens R-HSA-448706 | 8.39E-01 | 9.94E-01 | 5.60E-01 | 9.86E-02 | NFKB1 |

| | | | | | | |
|---|---|---|---|---|---|---|
| 1206 | Acyl chain remodeling of DAG and TAG Homo sapiens R-HSA-1482883 | 8.39E-01 | 9.94E-01 | 5.60E-01 | 9.86E-02 | PNPLA2 |
| 1207 | Ubiquinol biosynthesis Homo sapiens R-HSA-2142789 | 8.39E-01 | 9.94E-01 | 5.60E-01 | 9.86E-02 | COQ5 |
| 1208 | GABA synthesis, release, reuptake and degradation Homo sapiens R-HSA-888590 | 8.46E-01 | 1.00E+00 | 6.30E-01 | 1.06E-01 | HSPA8;SYT1;SLC6A12 |
| 1209 | Phospholipid metabolism Homo sapiens R-HSA-1483257 | 8.46E-01 | 1.00E+00 | 8.33E-01 | 1.39E-01 | DMGDH;PI4K2B;ARF1;SLC44A3;ABHD3;PITPNB;GDE1;PIK3R3;TAZ;MTMR6;PGS1;PTDSS2;INPP5F;PTPMT1;INPP5E;CPNE1;CPNE3;PIP5K1B;PIP4K2C;PLA2G4A;MBOAT2;CRLS1;PISD;HADHB;HADHA;ETNK2;CHDH;ETNK1;CHPT1;ALDH7A1;LPIN2;PNPLA2 |
| 1210 | Ca2+ pathway Homo sapiens R-HSA-4086398 | 8.57E-01 | 1.00E+00 | 7.39E-01 | 1.14E-01 | GNAO1;PPP3CB;GNG10;TCF7L1;WNT11;GNG5;AGO2;ITPR2;GNB5;PLCB1;CALM1 |
| 1211 | Neurotransmitter Release Cycle Homo sapiens R-HSA-112310 | 8.58E-01 | 1.00E+00 | 7.20E-01 | 1.10E-01 | HSPA8;SYT1;NAAA;PPFIA1;ARL6IP5;SLC6A12;LIN7C;SLC38A2;GLS |
| 1212 | ADP signalling through P2Y purinoceptor 1 Homo sapiens R-HSA-418592 | 8.58E-01 | 1.00E+00 | 6.40E-01 | 9.79E-02 | GNG10;GNG5;PLA2G4A;GNB5 |
| 1213 | TRP channels Homo sapiens R-HSA-3295583 | 8.58E-01 | 1.00E+00 | 6.40E-01 | 9.79E-02 | TRPV4;TRPM7;MCOLN1;TRPC4AP |
| 1214 | Chromosome Maintenance Homo sapiens R-HSA-73886 | 8.62E-01 | 1.00E+00 | 7.67E-01 | 1.14E-01 | NPM1;PCNA;RFC1;H2AFZ;TINF2;SMARCA5;RSF1;CENPC;RBBP4;DKC1;RPA3;POLD2;NHP2;TERF2IP;RBBP7;HIST1H2AC |

| | | | | | | |
|---|---|---|---|---|---|---|
| 1215 | Organic cation/anion/zwitterion transport Homo sapiens R-HSA-549132 | 8.65E-01 | 1.00E+00 | 5.60E-01 | 8.10E-02 | SLC22A15;RUNX1 |
| 1216 | FGFR4 ligand binding and activation Homo sapiens R-HSA-190322 | 8.65E-01 | 1.00E+00 | 5.60E-01 | 8.10E-02 | FGF18;FGF2 |
| 1217 | Bile salt and organic anion SLC transporters Homo sapiens R-HSA-425471 | 8.65E-01 | 1.00E+00 | 5.60E-01 | 8.10E-02 | SLC47A2;SLC16A3 |
| 1218 | Repression of WNT target genes Homo sapiens R-HSA-4641265 | 8.65E-01 | 1.00E+00 | 5.60E-01 | 8.10E-02 | TCF7L1;HDAC1 |
| 1219 | Lysosphingolipid and LPA receptors Homo sapiens R-HSA-419408 | 8.65E-01 | 1.00E+00 | 5.60E-01 | 8.10E-02 | LPAR2;S1PR3 |
| 1220 | Synthesis of PIPs at the early endosome membrane Homo sapiens R-HSA-1660516 | 8.65E-01 | 1.00E+00 | 5.60E-01 | 8.10E-02 | PI4K2B;INPP5F |
| 1221 | Gene Silencing by RNA Homo sapiens R-HSA-211000 | 8.67E-01 | 1.00E+00 | 7.78E-01 | 1.11E-01 | TSNAX;HSP90AA1;H2AFZ;H3F3A;TSN;NUP93;POLR2B;PRKRA;NUP50;POLR2C;TPR;AGO2;POLR2E;POLR2H;POLR2J;HIST1H2AC;POLR2K;RAN;POLR2L |
| 1222 | Chondroitin sulfate biosynthesis Homo sapiens R-HSA-2022870 | 8.70E-01 | 1.00E+00 | 5.93E-01 | 8.24E-02 | CHPF;CHST12;CHST15 |
| 1223 | FGFR2 ligand binding and activation Homo sapiens R-HSA-190241 | 8.70E-01 | 1.00E+00 | 5.93E-01 | 8.24E-02 | FGFBP1;FGF18;FGF2 |

| | | | | | | |
|---|---|---|---|---|---|---|
| 1224 | Hormone-sensitive lipase (HSL)-mediated triacylglycerol hydrolysis Homo sapiens R-HSA-163560 | 8.70E-01 | 1.00E+00 | 5.93E-01 | 8.24E-02 | PPP1CB;PPP1CC;PPP1CA |
| 1225 | Amino acid transport across the plasma membrane Homo sapiens R-HSA-352230 | 8.71E-01 | 1.00E+00 | 6.46E-01 | 8.96E-02 | SLC7A6;SLC43A2;SLC7A8;SLC6A12;SLC38A2 |
| 1226 | Export of Viral Ribonucleoproteins from Nucleus Homo sapiens R-HSA-168274 | 8.71E-01 | 1.00E+00 | 6.46E-01 | 8.96E-02 | NUP93;NUP50;TPR;RAN;HSPA1A |
| 1227 | Nucleotide-binding domain, leucine rich repeat containing receptor (NLR) signaling pathways Homo sapiens R-HSA-168643 | 8.75E-01 | 1.00E+00 | 6.89E-01 | 9.19E-02 | PYCARD;APP;ITCH;HSP90AB1;IRAK1;SUGT1;UBE2V1;MAPK13 |
| 1228 | Antigen activates B Cell Receptor (BCR) leading to generation of second messengers Homo sapiens R-HSA-983695 | 8.75E-01 | 1.00E+00 | 6.89E-01 | 9.19E-02 | SH3KBP1;ITPR2;ORAI1;FYN;GRB2;PTPN6;CBL;CALM1 |
| 1229 | Erythrocytes take up oxygen and release carbon dioxide Homo sapiens R-HSA-1247673 | 8.76E-01 | 1.00E+00 | 4.80E-01 | 6.38E-02 | CA2 |

| | | | | | | |
|---|---|---|---|---|---|---|
| 1230 | Neurotransmitter Clearance In The Synaptic Cleft Homo sapiens R-HSA-112311 | 8.76E-01 | 1.00E+00 | 4.80E-01 | 6.38E-02 | COMT |
| 1231 | Organic anion transporters Homo sapiens R-HSA-428643 | 8.76E-01 | 1.00E+00 | 4.80E-01 | 6.38E-02 | SLC17A5 |
| 1232 | Transport of fatty acids Homo sapiens R-HSA-804914 | 8.76E-01 | 1.00E+00 | 4.80E-01 | 6.38E-02 | LCN12 |
| 1233 | Ligand-receptor interactions Homo sapiens R-HSA-5632681 | 8.76E-01 | 1.00E+00 | 4.80E-01 | 6.38E-02 | BOC |
| 1234 | RNF mutants show enhanced WNT signaling and proliferation Homo sapiens R-HSA-5340588 | 8.76E-01 | 1.00E+00 | 4.80E-01 | 6.38E-02 | RNF43 |
| 1235 | Terminal pathway of complement Homo sapiens R-HSA-166665 | 8.76E-01 | 1.00E+00 | 4.80E-01 | 6.38E-02 | CLU |
| 1236 | Free fatty acids regulate insulin secretion Homo sapiens R-HSA-400451 | 8.76E-01 | 1.00E+00 | 4.80E-01 | 6.38E-02 | PLCB1 |
| 1237 | Fatty Acids bound to GPR40 (FFAR1) regulate insulin secretion Homo sapiens R-HSA-434316 | 8.76E-01 | 1.00E+00 | 4.80E-01 | 6.38E-02 | PLCB1 |

| | | | | | | |
|---|---|---|---|---|---|---|
| 1238 | Cross-presentation of particulate exogenous antigens (phagosomes) Homo sapiens R-HSA-1236973 | 8.76E-01 | 1.00E+00 | 4.80E-01 | 6.38E-02 | CYBA |
| 1239 | Hydrolysis of LPC Homo sapiens R-HSA-1483115 | 8.76E-01 | 1.00E+00 | 4.80E-01 | 6.38E-02 | PLA2G4A |
| 1240 | Linoleic acid (LA) metabolism Homo sapiens R-HSA-2046105 | 8.76E-01 | 1.00E+00 | 4.80E-01 | 6.38E-02 | ELOVL1 |
| 1241 | Nicotinamide salvaging Homo sapiens R-HSA-197264 | 8.76E-01 | 1.00E+00 | 4.80E-01 | 6.38E-02 | NAMPT |
| 1242 | Synthesis of epoxy (EET) and dihydroxyeicosatrienoic acids (DHET) Homo sapiens R-HSA-2142670 | 8.76E-01 | 1.00E+00 | 4.80E-01 | 6.38E-02 | CYP1B1 |
| 1243 | Interleukin-17 signaling Homo sapiens R-HSA-448424 | 8.76E-01 | 1.00E+00 | 4.80E-01 | 6.38E-02 | IL17RB |
| 1244 | Termination of O-glycan biosynthesis Homo sapiens R-HSA-977068 | 8.79E-01 | 1.00E+00 | 6.11E-01 | 7.89E-02 | MUC1;ST3GAL4;MUC15;MUC20 |
| 1245 | PD-1 signaling Homo sapiens R-HSA-389948 | 8.79E-01 | 1.00E+00 | 6.11E-01 | 7.89E-02 | CSK;PTPN11;PTPN6;HLA-DQB1 |
| 1246 | NCAM1 interactions Homo sapiens R-HSA-419037 | 8.82E-01 | 1.00E+00 | 6.50E-01 | 8.15E-02 | PRNP;CACNB4;COL4A4;COL4A3;CACNA1D;CACNA1C |

| | | | | | | |
|---|---|---|---|---|---|---|
| 1247 | Arachidonic acid metabolism Homo sapiens R-HSA-2142753 | 8.87E-01 | 1.00E+00 | 6.87E-01 | 8.23E-02 | CBR1;FAAH;GPX4;ALOX5;PTGES3;CYP1B1;PLA2G4A;LTA4H;PTGES |
| 1248 | DNA strand elongation Homo sapiens R-HSA-69190 | 8.88E-01 | 1.00E+00 | 6.22E-01 | 7.38E-02 | PCNA;RFC1;MCM8;RPA3;POLD2 |
| 1249 | Elevation of cytosolic Ca2+ levels Homo sapiens R-HSA-139853 | 8.90E-01 | 1.00E+00 | 5.17E-01 | 6.01E-02 | ITPR2;ORAI1 |
| 1250 | SHC-mediated cascade:FGFR1 Homo sapiens R-HSA-5654688 | 8.91E-01 | 1.00E+00 | 5.60E-01 | 6.44E-02 | KRAS;GRB2;FGF2 |
| 1251 | Phase 1 - inactivation of fast Na+ channels Homo sapiens R-HSA-5576894 | 8.97E-01 | 1.00E+00 | 5.84E-01 | 6.37E-02 | CACNB4;CACNA2D3;CACNA1D;CACNA1C |
| 1252 | Telomere Maintenance Homo sapiens R-HSA-157579 | 8.98E-01 | 1.00E+00 | 6.85E-01 | 7.38E-02 | PCNA;RFC1;H2AFZ;DKC1;RPA3;TINF2;POLD2;NHP2;TERF2IP;HIST1H2AC |
| 1253 | Transport of the SLBP independent Mature mRNA Homo sapiens R-HSA-159227 | 9.04E-01 | 1.00E+00 | 6.00E-01 | 6.08E-02 | NUP93;NCBP1;NUP50;TPR;EIF4E |
| 1254 | Bicarbonate transporters Homo sapiens R-HSA-425381 | 9.04E-01 | 1.00E+00 | 4.20E-01 | 4.23E-02 | SLC4A7 |
| 1255 | Retinoid cycle disease events Homo sapiens R-HSA-2453864 | 9.04E-01 | 1.00E+00 | 4.20E-01 | 4.23E-02 | NAPEPLD |

| | | | | | | |
|---|---|---|---|---|---|---|
| 1256 | Diseases associated with visual transduction Homo sapiens R-HSA-2474795 | 9.04E-01 | 1.00E+00 | 4.20E-01 | 4.23E-02 | NAPEPLD |
| 1257 | Sodium/Proton exchangers Homo sapiens R-HSA-425986 | 9.04E-01 | 1.00E+00 | 4.20E-01 | 4.23E-02 | SLC9A3 |
| 1258 | Transport of gamma-carboxylated protein precursors from the endoplasmic reticulum to the Golgi apparatus Homo sapiens R-HSA-159763 | 9.04E-01 | 1.00E+00 | 4.20E-01 | 4.23E-02 | GAS6 |
| 1259 | Alpha-defensins Homo sapiens R-HSA-1462054 | 9.04E-01 | 1.00E+00 | 4.20E-01 | 4.23E-02 | PRSS3 |
| 1260 | Synthesis of (16-20)-hydroxyeicosatetraenoic acids (HETE) Homo sapiens R-HSA-2142816 | 9.04E-01 | 1.00E+00 | 4.20E-01 | 4.23E-02 | CYP1B1 |
| 1261 | Highly calcium permeable nicotinic acetylcholine receptors Homo sapiens R-HSA-629597 | 9.04E-01 | 1.00E+00 | 4.20E-01 | 4.23E-02 | CHRNA5 |
| 1262 | Interleukin-19, 20, 22, 24 Homo sapiens R-HSA-8854691 | 9.04E-01 | 1.00E+00 | 4.20E-01 | 4.23E-02 | IL10RB |

| | | | | | | |
|---|---|---|---|---|---|---|
| 1263 | Phase 0 - rapid depolarisation Homo sapiens R-HSA-5576892 | 9.06E-01 | 1.00E+00 | 6.35E-01 | 6.27E-02 | CACNB4;CACNA2D3;RANGRF;CACNA1D;CACNA1C;CALM1;CAMK2G |
| 1264 | PI Metabolism Homo sapiens R-HSA-1483255 | 9.09E-01 | 1.00E+00 | 6.72E-01 | 6.41E-02 | PI4K2B;ARF1;INPP5F;PITPNB;INPP5E;GDE1;PIK3R3;PIP5K1B;PIP4K2C;MTMR6 |
| 1265 | Intrinsic Pathway of Fibrin Clot Formation Homo sapiens R-HSA-140837 | 9.09E-01 | 1.00E+00 | 5.30E-01 | 5.05E-02 | C1QBP;KNG1;SERPINA5 |
| 1266 | SALM protein interactions at the synapse Homo sapiens R-HSA-8849932 | 9.09E-01 | 1.00E+00 | 5.30E-01 | 5.05E-02 | RTN3;DLG1;FLOT1 |
| 1267 | TRAF6 mediated induction of TAK1 complex Homo sapiens R-HSA-937072 | 9.11E-01 | 1.00E+00 | 4.80E-01 | 4.48E-02 | UBC;RPS27A |
| 1268 | JNK (c-Jun kinases) phosphorylation and activation mediated by activated human TAK1 Homo sapiens R-HSA-450321 | 9.11E-01 | 1.00E+00 | 4.80E-01 | 4.48E-02 | MAPK9;IRAK1 |
| 1269 | Transport of glucose and other sugars, bile salts and organic acids, metal ions and amine compounds Homo sapiens R-HSA-425366 | 9.14E-01 | 1.00E+00 | 7.28E-01 | 6.58E-02 | SLC47A2;SLC14A2;SLC44A3;SLC2A11;SLC41A2;SLC2A13;SLC2A1;SLC40A1;SLC6A12;SLC5A3;SLC2A6;RUNX1;SLC2A9;SLC22A15;SLC39A7;SLC39A1;SLC16A3;SLC39A4 |

| | | | | | | |
|---|---|---|---|---|---|---|
| 1270 | Degradation of the extracellular matrix Homo sapiens R-HSA-1474228 | 9.14E-01 | 1.00E+00 | 7.33E-01 | 6.60E-02 | CAST;COL26A1;LAMB3;COL14A1;COL12A1;FURIN;HSPG2;CAPN15;CAPN14;CAPN6;CAPN7;CTSL;CTSK;CAPN5;ADAM9;CAPN1;TIMP1;CTSD;CTSB |
| 1271 | Nuclear Pore Complex (NPC) Disassembly Homo sapiens R-HSA-3301854 | 9.17E-01 | 1.00E+00 | 5.79E-01 | 5.00E-02 | NUP93;CCNB1;NUP50;TPR;NEK7 |
| 1272 | Activated PKN1 stimulates transcription of AR (androgen receptor) regulated genes KLK2 and KLK3 Homo sapiens R-HSA-5625886 | 9.17E-01 | 1.00E+00 | 5.79E-01 | 5.00E-02 | NCOA2;H2AFZ;H3F3A;PKN1;HIST1H2AC |
| 1273 | Lipoprotein metabolism Homo sapiens R-HSA-174824 | 9.17E-01 | 1.00E+00 | 5.79E-01 | 5.00E-02 | SAR1B;SDC1;P4HB;LDLRAP1;HSPG2 |
| 1274 | Synthesis of PIPs at the plasma membrane Homo sapiens R-HSA-1660499 | 9.17E-01 | 1.00E+00 | 5.79E-01 | 5.00E-02 | PI4K2B;PIK3R3;PIP5K1B;PIP4K2C;MTMR6 |
| 1275 | Dopamine Neurotransmitter Release Cycle Homo sapiens R-HSA-212676 | 9.24E-01 | 1.00E+00 | 5.04E-01 | 3.96E-02 | SYT1;PPFIA1;LIN7C |
| 1276 | Thromboxane signalling through TP receptor Homo sapiens R-HSA-428930 | 9.24E-01 | 1.00E+00 | 5.04E-01 | 3.96E-02 | GNG10;GNG5;GNB5 |

| | | | | | | |
|---|---|---|---|---|---|---|
| 1277 | Interaction between L1 and Ankyrins Homo sapiens R-HSA-445095 | 9.26E-01 | 1.00E+00 | 5.37E-01 | 4.14E-02 | SPTBN5;ANK2;L1CAM;SPTAN1 |
| 1278 | Neurotoxicity of clostridium toxins Homo sapiens R-HSA-168799 | 9.26E-01 | 1.00E+00 | 3.73E-01 | 2.87E-02 | SYT1 |
| 1279 | Creatine metabolism Homo sapiens R-HSA-71288 | 9.26E-01 | 1.00E+00 | 3.73E-01 | 2.87E-02 | SLC6A12 |
| 1280 | Abacavir transport and metabolism Homo sapiens R-HSA-2161522 | 9.26E-01 | 1.00E+00 | 3.73E-01 | 2.87E-02 | GUK1 |
| 1281 | Diseases associated with surfactant metabolism Homo sapiens R-HSA-5687613 | 9.26E-01 | 1.00E+00 | 3.73E-01 | 2.87E-02 | CCDC59 |
| 1282 | Gamma-carboxylation of protein precursors Homo sapiens R-HSA-159740 | 9.26E-01 | 1.00E+00 | 3.73E-01 | 2.87E-02 | GAS6 |
| 1283 | POU5F1 (OCT4), SOX2, NANOG repress genes related to differentiation Homo sapiens R-HSA-2892245 | 9.26E-01 | 1.00E+00 | 3.73E-01 | 2.87E-02 | DKK1 |
| 1284 | TRIF-mediated programmed cell death Homo sapiens R-HSA-2562578 | 9.26E-01 | 1.00E+00 | 3.73E-01 | 2.87E-02 | FADD |

| | | | | | | |
|---|---|---|---|---|---|---|
| 1285 | GP1b-IX-V activation signalling Homo sapiens R-HSA-430116 | 9.26E-01 | 1.00E+00 | 3.73E-01 | 2.87E-02 | YWHAZ |
| 1286 | Dectin-2 family Homo sapiens R-HSA-5621480 | 9.26E-01 | 1.00E+00 | 3.73E-01 | 2.87E-02 | FYN |
| 1287 | Acetylcholine regulates insulin secretion Homo sapiens R-HSA-399997 | 9.26E-01 | 1.00E+00 | 3.73E-01 | 2.87E-02 | PLCB1 |
| 1288 | Regulation of signaling by NODAL Homo sapiens R-HSA-1433617 | 9.26E-01 | 1.00E+00 | 3.73E-01 | 2.87E-02 | TDGF1 |
| 1289 | Purine catabolism Homo sapiens R-HSA-74259 | 9.26E-01 | 1.00E+00 | 3.73E-01 | 2.87E-02 | CAT |
| 1290 | Calcitonin-like ligand receptors Homo sapiens R-HSA-419812 | 9.26E-01 | 1.00E+00 | 3.73E-01 | 2.87E-02 | CALCA |
| 1291 | Acetylcholine Neurotransmitter Release Cycle Homo sapiens R-HSA-264642 | 9.28E-01 | 1.00E+00 | 4.48E-01 | 3.36E-02 | SYT1;PPFIA1 |
| 1292 | TNF receptor superfamily (TNFSF) members mediating non-canonical NF-kB pathway Homo sapiens R-HSA-5676594 | 9.28E-01 | 1.00E+00 | 4.48E-01 | 3.36E-02 | TNFRSF12A;TNFSF12 |
| 1293 | G1/S-Specific Transcription Homo sapiens R-HSA-69205 | 9.28E-01 | 1.00E+00 | 4.48E-01 | 3.36E-02 | PCNA;TK2 |

| | | | | | | |
|---|---|---|---|---|---|---|
| 1294 | Activated point mutants of FGFR2 Homo sapiens R-HSA-2033519 | 9.28E-01 | 1.00E+00 | 4.48E-01 | 3.36E-02 | FGF18;FGF2 |
| 1295 | activated TAK1 mediates p38 MAPK activation Homo sapiens R-HSA-450302 | 9.28E-01 | 1.00E+00 | 4.48E-01 | 3.36E-02 | MAP2K3;IRAK1 |
| 1296 | Unblocking of NMDA receptor, glutamate binding and activation Homo sapiens R-HSA-438066 | 9.28E-01 | 1.00E+00 | 4.48E-01 | 3.36E-02 | AKAP9;CAMK2G |
| 1297 | Acyl chain remodelling of PG Homo sapiens R-HSA-1482925 | 9.28E-01 | 1.00E+00 | 4.48E-01 | 3.36E-02 | PLA2G4A;CRLS1 |
| 1298 | Metabolism of porphyrins Homo sapiens R-HSA-189445 | 9.28E-01 | 1.00E+00 | 4.48E-01 | 3.36E-02 | ALAS1;HMOX2 |
| 1299 | Deposition of new CENPA-containing nucleosomes at the centromere Homo sapiens R-HSA-606279 | 9.34E-01 | 1.00E+00 | 6.10E-01 | 4.18E-02 | NPM1;RBBP4;H2AFZ;SMARCA5;RSF1;RBBP7;HIST1H2AC;CENPC |
| 1300 | Nucleosome assembly Homo sapiens R-HSA-774815 | 9.34E-01 | 1.00E+00 | 6.10E-01 | 4.18E-02 | NPM1;RBBP4;H2AFZ;SMARCA5;RSF1;RBBP7;HIST1H2AC;CENPC |
| 1301 | NEP/NS2 Interacts with the Cellular Export Machinery Homo sapiens R-HSA-168333 | 9.37E-01 | 1.00E+00 | 5.17E-01 | 3.34E-02 | NUP93;NUP50;TPR;RAN |

| | | | | | | |
|---|---|---|---|---|---|---|
| 1302 | Regulation of Glucokinase by Glucokinase Regulatory Protein Homo sapiens R-HSA-170822 | 9.37E-01 | 1.00E+00 | 5.17E-01 | 3.34E-02 | NUP93;NUP50;TPR;GCK |
| 1303 | Transport of Ribonucleoproteins into the Host Nucleus Homo sapiens R-HSA-168271 | 9.37E-01 | 1.00E+00 | 5.17E-01 | 3.34E-02 | NUP93;NUP50;TPR;KPNA1 |
| 1304 | RNA Polymerase I Promoter Opening Homo sapiens R-HSA-73728 | 9.37E-01 | 1.00E+00 | 5.17E-01 | 3.34E-02 | H2AFZ;H3F3A;MBD2;HIST1H2AC |
| 1305 | Metabolism of steroid hormones Homo sapiens R-HSA-196071 | 9.37E-01 | 1.00E+00 | 5.17E-01 | 3.34E-02 | HSD17B1;FDX1;AKR1B1;HSD17B12 |
| 1306 | Formation of the beta-catenin:TCF transactivating complex Homo sapiens R-HSA-201722 | 9.39E-01 | 1.00E+00 | 6.17E-01 | 3.88E-02 | PYGO1;TCF7L1;H2AFZ;HDAC1;H3F3A;LEO1;ASH2L;TCF4;HIST1H2AC |
| 1307 | Serotonin Neurotransmitter Release Cycle Homo sapiens R-HSA-181429 | 9.42E-01 | 1.00E+00 | 4.20E-01 | 2.53E-02 | SYT1;PPFIA1 |
| 1308 | Norepinephrine Neurotransmitter Release Cycle Homo sapiens R-HSA-181430 | 9.42E-01 | 1.00E+00 | 4.20E-01 | 2.53E-02 | SYT1;PPFIA1 |
| 1309 | Glucose transport Homo sapiens R-HSA-70153 | 9.42E-01 | 1.00E+00 | 5.60E-01 | 3.32E-02 | NUP93;NUP50;TPR;SLC2A1;GCK;HK1 |

| | | | | | | |
|---|---|---|---|---|---|---|
| 1310 | Dermatan sulfate biosynthesis Homo sapiens R-HSA-2022923 | 9.43E-01 | 1.00E+00 | 3.36E-01 | 1.97E-02 | CHST14 |
| 1311 | CDC6 association with the ORC:origin complex Homo sapiens R-HSA-68689 | 9.43E-01 | 1.00E+00 | 3.36E-01 | 1.97E-02 | MCM8 |
| 1312 | FGFR1c ligand binding and activation Homo sapiens R-HSA-190373 | 9.43E-01 | 1.00E+00 | 3.36E-01 | 1.97E-02 | FGF2 |
| 1313 | Signaling by activated point mutants of FGFR1 Homo sapiens R-HSA-1839122 | 9.43E-01 | 1.00E+00 | 3.36E-01 | 1.97E-02 | FGF2 |
| 1314 | MyD88 deficiency (TLR2/4) Homo sapiens R-HSA-5602498 | 9.43E-01 | 1.00E+00 | 3.36E-01 | 1.97E-02 | IRAK4 |
| 1315 | IRAK4 deficiency (TLR2/4) Homo sapiens R-HSA-5603041 | 9.43E-01 | 1.00E+00 | 3.36E-01 | 1.97E-02 | IRAK4 |
| 1316 | Apoptotic cleavage of cell adhesion proteins Homo sapiens R-HSA-351906 | 9.43E-01 | 1.00E+00 | 3.36E-01 | 1.97E-02 | TJP1 |
| 1317 | Androgen biosynthesis Homo sapiens R-HSA-193048 | 9.43E-01 | 1.00E+00 | 3.36E-01 | 1.97E-02 | HSD17B12 |
| 1318 | CYP2E1 reactions Homo sapiens R-HSA-211999 | 9.43E-01 | 1.00E+00 | 3.36E-01 | 1.97E-02 | CYP2S1 |

| | | | | | | |
|---|---|---|---|---|---|---|
| 1319 | Heme biosynthesis Homo sapiens R-HSA-189451 | 9.43E-01 | 1.00E+00 | 3.36E-01 | 1.97E-02 | ALAS1 |
| 1320 | Highly calcium permeable postsynaptic nicotinic acetylcholine receptors Homo sapiens R-HSA-629594 | 9.43E-01 | 1.00E+00 | 3.36E-01 | 1.97E-02 | CHRNA5 |
| 1321 | Hexose transport Homo sapiens R-HSA-189200 | 9.46E-01 | 1.00E+00 | 5.73E-01 | 3.19E-02 | NUP93;NUP50;TPR;SLC2A1;SLC5A3;GCK;HK1 |
| 1322 | Packaging Of Telomere Ends Homo sapiens R-HSA-171306 | 9.47E-01 | 1.00E+00 | 4.98E-01 | 2.70E-02 | H2AFZ;TINF2;TERF2IP;HIST1H2AC |
| 1323 | NOD1/2 Signaling Pathway Homo sapiens R-HSA-168638 | 9.47E-01 | 1.00E+00 | 4.98E-01 | 2.70E-02 | ITCH;IRAK1;UBE2V1;MAPK13 |
| 1324 | Activation of G protein gated Potassium channels Homo sapiens R-HSA-1296041 | 9.48E-01 | 1.00E+00 | 4.58E-01 | 2.44E-02 | GNG10;GNG5;KCNJ15 |
| 1325 | Inhibition of voltage gated Ca2+ channels via Gbeta/gamma subunits Homo sapiens R-HSA-997272 | 9.48E-01 | 1.00E+00 | 4.58E-01 | 2.44E-02 | GNG10;GNG5;KCNJ15 |
| 1326 | G protein gated Potassium channels Homo sapiens R-HSA-1296059 | 9.48E-01 | 1.00E+00 | 4.58E-01 | 2.44E-02 | GNG10;GNG5;KCNJ15 |

| | | | | | | |
|---|---|---|---|---|---|---|
| 1327 | HDR through Single Strand Annealing (SSA) Homo sapiens R-HSA-5685938 | 9.48E-01 | 1.00E+00 | 5.25E-01 | 2.78E-02 | RAD50;RPA3;ERCC1;RAD17;RAD1 |
| 1328 | Activation of ATR in response to replication stress Homo sapiens R-HSA-176187 | 9.48E-01 | 1.00E+00 | 5.25E-01 | 2.78E-02 | MCM8;RPA3;CDK2;RAD17;RAD1 |
| 1329 | Assembly of collagen fibrils and other multimeric structures Homo sapiens R-HSA-2022090 | 9.49E-01 | 1.00E+00 | 5.84E-01 | 3.03E-02 | CD151;LAMB3;CTSL;COL4A4;COL4A3;PCOLCE;LOXL2;CTSB |
| 1330 | Transmembrane transport of small molecules Homo sapiens R-HSA-382551 | 9.53E-01 | 1.00E+00 | 8.47E-01 | 4.11E-02 | SLC35B3;SLC46A1;TUSC3;ATP12A;BEST3;NIPAL3;SLC39A7;ATP6V1E1;SLC39A1;SLC16A3;SLC39A4;ATP6V1G1;ATP6V0E1;MMGT1;SLC6A12;ATP1B3;ATP1B1;TRPC4AP;RUNX1;NUP93;SLC9A3;PRKAR1A;ATP6V0D1;ATP6V1A;TTYH3;ABCB7;SLC41A2;CLCNKB;CLCNKA;SLC5A3;GNG10;PRKAR2B;PRKAR2A;LCN12;TPR;UBC;ATP6V1H;TRPM7;RAB11FIP2;SLC38A2;ATP6V1D;HMOX2;ATP6V1C1;ATP6V1F;SLC14A2;ATP8B1;ABCA3;ABCA4;CYBRD1;EIF2S2;EIF2S1;SLC2A9;FXYD3;FXYD2;TRPV4;SLCO1A2;WNK3;GNAS;GNB5;CALM1;CUTC;SLC44A3;ABCD3;SLC2A1;SLC40A1;ATP2A3;SLC2A6;HK1;SLC22A15;FLVCR1;ATP6V0B;SLC2A11;ATP6AP1;SLC2A13;ABCC5;ANO6;NIPA2;ANO5;SLC7A6;SLC7A8;ASPH;SLCO4A1;PEX3;NUP50;SGK2;ATP6V0C;SLC29A2;SLC47A2;SLC20A2;SLC43A2;ADCY3;SRI;ATP1A1;CLCN3;ADD1;ADCY5;SLC25A26;GNG5;SLC17A5;BSND;ANO10;RPS27A;ATP6V0E2;CAMK2G;ABCF1;SLC12A2;SLC35A1;DMTN;SLC35A3;CTNS;ATP2B1;MCOLN1;RAB11A;GCK;TPCN1;CLCN7;SLC4A7;OSTM1;SLCO3A1;FKBP1B |
| 1331 | Phase 4 - resting membrane potential Homo sapiens R-HSA-5576886 | 9.53E-01 | 1.00E+00 | 3.95E-01 | 1.91E-02 | KCNK5;KCNK15 |

| | | | | | | |
|---|---|---|---|---|---|---|
| 1332 | DNA methylation Homo sapiens R-HSA-5334118 | 9.56E-01 | 1.00E+00 | 4.80E-01 | 2.17E-02 | DNMT1;H2AFZ;H3F3A;HIST1H2AC |
| 1333 | GABA receptor activation Homo sapiens R-HSA-977443 | 9.56E-01 | 1.00E+00 | 5.71E-01 | 2.57E-02 | GNG10;GNG5;GNAI3;KCNJ15;ADCY3;GNAI1;ADCY5;GNAI2 |
| 1334 | Ethanol oxidation Homo sapiens R-HSA-71384 | 9.56E-01 | 1.00E+00 | 3.05E-01 | 1.37E-02 | ADH5 |
| 1335 | TP53 Regulates Transcription of Caspase Activators and Caspases Homo sapiens R-HSA-6803207 | 9.56E-01 | 1.00E+00 | 3.05E-01 | 1.37E-02 | APAF1 |
| 1336 | Unwinding of DNA Homo sapiens R-HSA-176974 | 9.56E-01 | 1.00E+00 | 3.05E-01 | 1.37E-02 | MCM8 |
| 1337 | Miscellaneous substrates Homo sapiens R-HSA-211958 | 9.56E-01 | 1.00E+00 | 3.05E-01 | 1.37E-02 | CYP2S1 |
| 1338 | Pyrimidine catabolism Homo sapiens R-HSA-73621 | 9.56E-01 | 1.00E+00 | 3.05E-01 | 1.37E-02 | NT5C3A |
| 1339 | Lysine catabolism Homo sapiens R-HSA-71064 | 9.56E-01 | 1.00E+00 | 3.05E-01 | 1.37E-02 | ALDH7A1 |
| 1340 | Presynaptic nicotinic acetylcholine receptors Homo sapiens R-HSA-622323 | 9.56E-01 | 1.00E+00 | 3.05E-01 | 1.37E-02 | CHRNA5 |
| 1341 | Regulation of Complement cascade Homo sapiens R-HSA-977606 | 9.57E-01 | 1.00E+00 | 4.38E-01 | 1.92E-02 | CD59;CD46;CD55 |

| | | | | | | |
|---|---|---|---|---|---|---|
| 1342 | Constitutive Signaling by Aberrant PI3K in Cancer Homo sapiens R-HSA-2219530 | 9.59E-01 | 1.00E+00 | 5.81E-01 | 2.44E-02 | BTC;FGF18;PDGFB;PIK3R3;FYN;PTPN11;GRB2;FGF2;HBEGF |
| 1343 | IL-6-type cytokine receptor ligand interactions Homo sapiens R-HSA-6788467 | 9.62E-01 | 1.00E+00 | 3.73E-01 | 1.44E-02 | JAK2;JAK1 |
| 1344 | Presynaptic phase of homologous DNA pairing and strand exchange Homo sapiens R-HSA-5693616 | 9.63E-01 | 1.00E+00 | 4.94E-01 | 1.87E-02 | RAD51B;RAD50;RPA3;RAD17;RAD1 |
| 1345 | Inactivation, recovery and regulation of the phototransduction cascade Homo sapiens R-HSA-2514859 | 9.63E-01 | 1.00E+00 | 4.63E-01 | 1.75E-02 | NMT1;GNB5;CALM1;CAMKMT |
| 1346 | The activation of arylsulfatases Homo sapiens R-HSA-1663150 | 9.66E-01 | 1.00E+00 | 2.80E-01 | 9.63E-03 | ARSJ |
| 1347 | The phototransduction cascade Homo sapiens R-HSA-2514856 | 9.69E-01 | 1.00E+00 | 4.48E-01 | 1.41E-02 | NMT1;GNB5;CALM1;CAMKMT |

| | | | | | | |
|---|---|---|---|---|---|---|
| 1348 | Regulation of Insulin-like Growth Factor (IGF) transport and uptake by Insulin-like Growth Factor Binding Proteins (IGFBPs) Homo sapiens R-HSA-381426 | 9.70E-01 | 1.00E+00 | 3.54E-01 | 1.09E-02 | IGFBP5;IGFBP6 |
| 1349 | Synthesis of Leukotrienes (LT) and Eoxins (EX) Homo sapiens R-HSA-2142691 | 9.70E-01 | 1.00E+00 | 3.54E-01 | 1.09E-02 | ALOX5;LTA4H |
| 1350 | Aflatoxin activation and detoxification Homo sapiens R-HSA-5423646 | 9.70E-01 | 1.00E+00 | 3.54E-01 | 1.09E-02 | MGST3;MGST2 |
| 1351 | Pre-NOTCH Transcription and Translation Homo sapiens R-HSA-1912408 | 9.73E-01 | 1.00E+00 | 4.66E-01 | 1.26E-02 | KAT2B;SNW1;TFDP2;AGO2;RBPJ |
| 1352 | Histidine, lysine, phenylalanine, tyrosine, proline and tryptophan catabolism Homo sapiens R-HSA-6788656 | 9.73E-01 | 1.00E+00 | 4.66E-01 | 1.26E-02 | QDPR;AFMID;PCBD1;KMO;ALDH7A1 |
| 1353 | LGI-ADAM interactions Homo sapiens R-HSA-5682910 | 9.74E-01 | 1.00E+00 | 2.58E-01 | 6.82E-03 | ADAM22 |

| | | | | | | |
|---|---|---|---|---|---|---|
| 1354 | CS/DS degradation Homo sapiens R-HSA-2024101 | 9.74E-01 | 1.00E+00 | 2.58E-01 | 6.82E-03 | HEXA |
| 1355 | Glutathione synthesis and recycling Homo sapiens R-HSA-174403 | 9.74E-01 | 1.00E+00 | 2.58E-01 | 6.82E-03 | GGCT |
| 1356 | Activation of Nicotinic Acetylcholine Receptors Homo sapiens R-HSA-629602 | 9.74E-01 | 1.00E+00 | 2.58E-01 | 6.82E-03 | CHRNA5 |
| 1357 | Postsynaptic nicotinic acetylcholine receptors Homo sapiens R-HSA-622327 | 9.74E-01 | 1.00E+00 | 2.58E-01 | 6.82E-03 | CHRNA5 |
| 1358 | Acetylcholine Binding And Downstream Events Homo sapiens R-HSA-181431 | 9.74E-01 | 1.00E+00 | 2.58E-01 | 6.82E-03 | CHRNA5 |
| 1359 | Generation of second messenger molecules Homo sapiens R-HSA-202433 | 9.74E-01 | 1.00E+00 | 4.33E-01 | 1.14E-02 | VASP;PAK1;PAK2;HLA-DQB1 |
| 1360 | SIRT1 negatively regulates rRNA Expression Homo sapiens R-HSA-427359 | 9.74E-01 | 1.00E+00 | 4.33E-01 | 1.14E-02 | H2AFZ;H3F3A;SIRT1;HIST1H2AC |
| 1361 | Common Pathway of Fibrin Clot Formation Homo sapiens R-HSA-140875 | 9.76E-01 | 1.00E+00 | 3.36E-01 | 8.31E-03 | F2R;SERPINA5 |

| | | | | | | |
|---|---|---|---|---|---|---|
| 1362 | Homologous DNA Pairing and Strand Exchange Homo sapiens R-HSA-5693579 | 9.78E-01 | 1.00E+00 | 4.54E-01 | 1.03E-02 | RAD51B;RAD50;RPA3;RAD17;RAD1 |
| 1363 | Lipid digestion, mobilization, and transport Homo sapiens R-HSA-73923 | 9.78E-01 | 1.00E+00 | 5.50E-01 | 1.25E-02 | PPP1CB;PPP1CC;NCEH1;SAR1B;LIPH;SDC1;P4HB;LDLRAP1;HSPG2;PPP1CA |
| 1364 | Meiotic recombination Homo sapiens R-HSA-912446 | 9.78E-01 | 1.00E+00 | 5.00E-01 | 1.12E-02 | RAD50;H2AFZ;CDK4;RPA3;H3F3A;CDK2;HIST1H2AC |
| 1365 | Disorders of transmembrane transporters Homo sapiens R-HSA-5619115 | 9.80E-01 | 1.00E+00 | 2.40E-01 | 4.87E-03 | GCK |
| 1366 | FGFR1 ligand binding and activation Homo sapiens R-HSA-190242 | 9.80E-01 | 1.00E+00 | 2.40E-01 | 4.87E-03 | FGF2 |
| 1367 | Negative regulation of TCF-dependent signaling by WNT ligand antagonists Homo sapiens R-HSA-3772470 | 9.80E-01 | 1.00E+00 | 2.40E-01 | 4.87E-03 | DKK1 |
| 1368 | Acyl chain remodelling of PE Homo sapiens R-HSA-1482839 | 9.80E-01 | 1.00E+00 | 3.20E-01 | 6.32E-03 | PLA2G4A;MBOAT2 |
| 1369 | Amine compound SLC transporters Homo sapiens R-HSA-425428 | 9.80E-01 | 1.00E+00 | 3.73E-01 | 7.35E-03 | SLC44A3;SLC14A2;SLC6A12 |

| | | | | | | |
|---|---|---|---|---|---|---|
| 1370 | Activation of the pre-replicative complex Homo sapiens R-HSA-68962 | 9.80E-01 | 1.00E+00 | 3.73E-01 | 7.35E-03 | MCM8;RPA3;CDK2 |
| 1371 | Amino acid and oligopeptide SLC transporters Homo sapiens R-HSA-425374 | 9.81E-01 | 1.00E+00 | 4.68E-01 | 9.05E-03 | SLC7A6;SLC7A8;SLC43A2;SLC17A5;SLC6A12;SLC38A2 |
| 1372 | Extracellular matrix organization Homo sapiens R-HSA-1474244 | 9.82E-01 | 1.00E+00 | 7.36E-01 | 1.34E-02 | ITGB1;COL14A1;COL12A1;ITGB2;PLOD2;ITGAE;FGF2;CAPN15;LOXL2;CAPN6;CTSL;CAPN7;CTSK;CAPN5;ITGB8;TIMP1;CAPN1;CTSD;CTSB;CAST;ACTN1;PCOLCE;HSPG2;COL4A4;COL4A3;ADAM9;CD47;PPIB;MATN3;FBN2;CD151;SDC2;PDGFB;NTN4;FURIN;LTBP3;LTBP1;FBLN5;TGFB2;CRTAP;COL26A1;TGFB1;LAMB3;LAMB2;TRAPPC4;CAPN14;P4HA1;MFAP1;P4HA2;SDC1;P4HB |
| 1373 | Steroid hormones Homo sapiens R-HSA-209943 | 9.82E-01 | 1.00E+00 | 4.07E-01 | 7.36E-03 | HSD17B1;FDX1;AKR1B1;HSD17B12 |
| 1374 | Adherens junctions interactions Homo sapiens R-HSA-418990 | 9.84E-01 | 1.00E+00 | 3.60E-01 | 5.78E-03 | JUP;CADM2;CDH13 |
| 1375 | Inwardly rectifying K+ channels Homo sapiens R-HSA-1296065 | 9.84E-01 | 1.00E+00 | 3.60E-01 | 5.78E-03 | GNG10;GNG5;KCNJ15 |
| 1376 | Phosphorylation of CD3 and TCR zeta chains Homo sapiens R-HSA-202427 | 9.84E-01 | 1.00E+00 | 3.05E-01 | 4.82E-03 | CSK;HLA-DQB1 |
| 1377 | Acyl chain remodelling of PI Homo sapiens R-HSA-1482922 | 9.85E-01 | 1.00E+00 | 2.24E-01 | 3.49E-03 | PLA2G4A |

| | | | | | | |
|---|---|---|---|---|---|---|
| 1378 | Activation of Matrix Metalloproteinases Homo sapiens R-HSA-1592389 | 9.87E-01 | 1.00E+00 | 3.47E-01 | 4.55E-03 | CTSK;FURIN;TIMP1 |
| 1379 | Resolution of D-loop Structures through Holliday Junction Intermediates Homo sapiens R-HSA-5693568 | 9.87E-01 | 1.00E+00 | 3.47E-01 | 4.55E-03 | MUS81;RAD51B;RAD50 |
| 1380 | HDMs demethylate histones Homo sapiens R-HSA-3214842 | 9.87E-01 | 1.00E+00 | 2.92E-01 | 3.67E-03 | KDM2A;JMJD6 |
| 1381 | Acyl chain remodelling of PC Homo sapiens R-HSA-1482788 | 9.87E-01 | 1.00E+00 | 2.92E-01 | 3.67E-03 | PLA2G4A;MBOAT2 |
| 1382 | Formation of Fibrin Clot (Clotting Cascade) Homo sapiens R-HSA-140877 | 9.88E-01 | 1.00E+00 | 3.84E-01 | 4.75E-03 | C1QBP;F2R;KNG1;SERPINA5 |
| 1383 | Digestion of dietary lipid Homo sapiens R-HSA-192456 | 9.88E-01 | 1.00E+00 | 2.10E-01 | 2.52E-03 | LIPH |
| 1384 | Acyl chain remodelling of PS Homo sapiens R-HSA-1482801 | 9.88E-01 | 1.00E+00 | 2.10E-01 | 2.52E-03 | PLA2G4A |
| 1385 | Resolution of D-Loop Structures Homo sapiens R-HSA-5693537 | 9.89E-01 | 1.00E+00 | 3.36E-01 | 3.58E-03 | MUS81;RAD51B;RAD50 |

| | | | | | | |
|---|---|---|---|---|---|---|
| 1386 | O-linked glycosylation Homo sapiens R-HSA-5173105 | 9.90E-01 | 1.00E+00 | 5.71E-01 | 5.97E-03 | SEMA5B;GALNT6;GALNT3;GALNT18;GALNT1;C1GALT1;POMGNT1;C1GALT1C1;MUC1;ADAMTSL2;ST3GAL4;B3GNT3;B3GNT2;MUC15;MUC20;GALNT9 |
| 1387 | Transport of inorganic cations/anions and amino acids/oligopeptides Homo sapiens R-HSA-425393 | 9.90E-01 | 1.00E+00 | 5.52E-01 | 5.77E-03 | SLC12A2;SLC20A2;SLC43A2;SLC6A12;CTNS;SRI;SLC4A7;SLC9A3;SLC25A26;SLC7A6;SLC7A8;SLC17A5;CALM1;SLC38A2 |
| 1388 | Diseases of glycosylation Homo sapiens R-HSA-3781865 | 9.90E-01 | 1.00E+00 | 5.29E-01 | 5.40E-03 | SEMA5B;C1GALT1C1;MUC1;SDC2;GPC1;ADAMTSL2;C1GALT1;SDC1;GPC5;MUC15;HSPG2;MUC20 |
| 1389 | Resolution of D-loop Structures through Synthesis-Dependent Strand Annealing (SDSA) Homo sapiens R-HSA-5693554 | 9.90E-01 | 1.00E+00 | 2.80E-01 | 2.80E-03 | RAD51B;RAD50 |
| 1390 | CD22 mediated BCR regulation Homo sapiens R-HSA-5690714 | 9.91E-01 | 1.00E+00 | 1.98E-01 | 1.82E-03 | PTPN6 |
| 1391 | cGMP effects Homo sapiens R-HSA-418457 | 9.91E-01 | 1.00E+00 | 1.98E-01 | 1.82E-03 | PDE1A |
| 1392 | Effects of PIP2 hydrolysis Homo sapiens R-HSA-114508 | 9.92E-01 | 1.00E+00 | 2.69E-01 | 2.14E-03 | DAGLA;ITPR2 |
| 1393 | Integrin cell surface interactions Homo sapiens R-HSA-216083 | 9.93E-01 | 1.00E+00 | 4.55E-01 | 3.32E-03 | ITGB1;COL4A4;ITGB2;COL4A3;ITGB8;ITGAE;CD47;HSPG2 |

| | | | | | | |
|---|---|---|---|---|---|---|
| 1394 | Na+/Cl- dependent neurotransmitter transporters Homo sapiens R-HSA-442660 | 9.93E-01 | 1.00E+00 | 1.87E-01 | 1.32E-03 | SLC6A12 |
| 1395 | Scavenging by Class A Receptors Homo sapiens R-HSA-3000480 | 9.93E-01 | 1.00E+00 | 1.87E-01 | 1.32E-03 | CALR |
| 1396 | Diseases associated with O-glycosylation of proteins Homo sapiens R-HSA-3906995 | 9.94E-01 | 1.00E+00 | 4.27E-01 | 2.60E-03 | SEMA5B;C1GALT1C1;MUC1;ADAMTSL2;C1GALT1;MUC15;MUC20 |
| 1397 | Cytochrome P450 - arranged by substrate type Homo sapiens R-HSA-211897 | 9.94E-01 | 1.00E+00 | 4.27E-01 | 2.60E-03 | CYP39A1;NCOA2;POR;CYP2S1;FDX1;CYP51A1;CYP1B1 |
| 1398 | Xenobiotics Homo sapiens R-HSA-211981 | 9.95E-01 | 1.00E+00 | 1.77E-01 | 9.66E-04 | CYP2S1 |
| 1399 | Translocation of ZAP-70 to Immunological synapse Homo sapiens R-HSA-202430 | 9.96E-01 | 1.00E+00 | 1.68E-01 | 7.07E-04 | HLA-DQB1 |
| 1400 | Class B/2 (Secretin family receptors) Homo sapiens R-HSA-373080 | 9.97E-01 | 1.00E+00 | 4.53E-01 | 1.51E-03 | CALCA;GNG10;WNT11;GNG5;GIPR;WNT7B;SCTR;GNAS;GNB5;CD55 |
| 1401 | SLC-mediated transmembrane transport Homo sapiens R-HSA-425407 | 9.97E-01 | 1.00E+00 | 6.57E-01 | 2.18E-03 | SLC35B3;SLC47A2;SLC44A3;SLC20A2;SLC43A2;SLC41A2;SLC2A1;SLC40A1;SRI;SLC5A3;SLC2A6;HK1;SLC25A26;SLC22A15;LCN12;TPR;SLC17A5;SLC39A7;SLC39A1;SLC16A3;SLC38A2;SLC39A4;SLC12A2;SLC35A1;SLC14A2;SLC35A3;SLC2A11;SLC2A13;SLC6A12;CTNS;GCK;RUNX1;NUP93;SLC2A9;SLC4A7;SLC9A3;SLC7A6;SLC7A8;SLCO3A1;SLCO4A1;NUP50;SLCO1A2;CALM1;SLC29A2 |

| | | | | | | |
|---|---|---|---|---|---|---|
| 1402 | Biological oxidations Homo sapiens R-HSA-211859 | 9.98E-01 | 1.00E+00 | 5.94E-01 | 1.29E-03 | SLC35B3;TPMT;MGST3;GSTP1;MGST2;COMT;ADH5;PAPSS1;TRMT112;ESD;CYP1B1;GSTT2B;GSTM4;GSTK1;GSTM3;NCOA2;AOC1;GSTO2;GSTO1;EPHX1;CYP51A1;ABHD14B;CYP39A1;SULT2B1;GGCT;POR;NAT2;CYP2S1;FDX1;GSTM5 |
| 1403 | ECM proteoglycans Homo sapiens R-HSA-3000178 | 9.98E-01 | 1.00E+00 | 3.35E-01 | 7.02E-04 | ITGB1;TGFB2;TGFB1;LAMB2;MATN3 |
| 1404 | Nitric oxide stimulates guanylate cyclase Homo sapiens R-HSA-392154 | 9.99E-01 | 1.00E+00 | 1.40E-01 | 2.07E-04 | PDE1A |
| 1405 | Fertilization Homo sapiens R-HSA-1187000 | 9.99E-01 | 1.00E+00 | 1.34E-01 | 1.53E-04 | CD9 |
| 1406 | Reproduction Homo sapiens R-HSA-1474165 | 9.99E-01 | 1.00E+00 | 1.34E-01 | 1.53E-04 | CD9 |
| 1407 | Phase 1 - Functionalization of compounds Homo sapiens R-HSA-211945 | 9.99E-01 | 1.00E+00 | 4.09E-01 | 3.79E-04 | CYP39A1;NCOA2;POR;AOC1;CYP2S1;FDX1;EPHX1;CYP51A1;CYP1B1;ADH5 |
| 1408 | Defective B3GALTL causes Peters-plus syndrome (PpS) Homo sapiens R-HSA-5083635 | 9.99E-01 | 1.00E+00 | 1.92E-01 | 1.50E-04 | SEMA5B;ADAMTSL2 |
| 1409 | Synthesis of PA Homo sapiens R-HSA-1483166 | 9.99E-01 | 1.00E+00 | 1.24E-01 | 8.44E-05 | PLA2G4A |
| 1410 | O-glycosylation of TSR domain-containing proteins Homo sapiens R-HSA-5173214 | 9.99E-01 | 1.00E+00 | 1.86E-01 | 1.15E-04 | SEMA5B;ADAMTSL2 |

| | | | | | | |
|---|---|---|---|---|---|---|
| 1411 | FCERI mediated Ca+2 mobilization Homo sapiens R-HSA-2871809 | 9.99E-01 | 1.00E+00 | 2.94E-01 | 1.57E-04 | VAV3;PPP3CB;ITPR2;GRB2;CALM1 |
| 1412 | Neuronal System Homo sapiens R-HSA-112316 | 1.00E+00 | 1.00E+00 | 6.02E-01 | 2.14E-04 | RTN3;CHRNA5;RASGRF2;GNAI3;ADCY3;AKAP5;COMT;LIN7C;AP2A2;GNAI1;GLS;ADCY5;GNAI2;GNG10;GNG5;TSPAN7;MYO6;ARL6IP5;AP2S1;PPFIA1;FLOT1;MAPK1;GLUL;SLC38A2;CAMK2G;AP2M1;NSF;HSPA8;SYT1;PDPK1;CACFD1;CACNA2D3;KCNJ15;SLC6A12;AP2B1;DLG1;CACNB4;CREB1;KCNQ1;AKAP9;NAAA;MDM2;PICK1;GNB5;CALM1;PLCB1 |
| 1413 | G alpha (s) signalling events Homo sapiens R-HSA-418555 | 1.00E+00 | 1.00E+00 | 4.55E-01 | 1.32E-04 | CALCA;GIPR;PDE1A;PDE4D;GNAI3;SCTR;GPBAR1;ADCY3;ARRB1;GNAI1;ADCY5;GNAI2;GNG10;GNG5;GNAS;PDE4A;GNB5 |
| 1414 | FCGR activation Homo sapiens R-HSA-2029481 | 1.00E+00 | 1.00E+00 | 1.43E-01 | 6.73E-06 | YES1;FYN |

| | | | | | | |
|---|---|---|---|---|---|---|
| 1415 | Signal Transduction Homo sapiens R-HSA-162582 | 1.00E+00 | 1.00E+00 | 8.05E-01 | 2.17E-05 | ATF1;CYFIP2;VPS29;NCKAP1;WDR83;SPINT2;RBPJ;MYLK;PSMD8;PSMD9;PSMD6;PSMD7;PSMD4;AKT2;PSMD2;PSMD3;AKT1;PSMD1;VPS35;PIP4K2C;IER3;CSNK2A2;HSPG2;DKK1;MYL6;ZFYVE16;SPINT1;PRKAR1A;PSME1;PSME2;RBCK1;MAPKAP1;HPN;PDGFB;VPS26A;PRKCZ;HIF1A;SKA2;RHOT2;RHOT1;RDH11;PRKAR2B;RDH10;PRKAR2A;ST3GAL4;PIP5K1B;RDH14;ST3GAL6;RDH13;PSMF1;JAG2;STARD13;JUP;ABCA4;H3F3A;CGN;HNRNPM;HNRNPF;AGO2;PNOC;NMT1;NAPEPLD;DZIP1;RARG;CDKN1B;ARPC1B;ARPC1A;CLTC;PRKAG1;CLTB;PRKAG2;CLTA;ASH2L;SHB;ARHGAP4;PEA15;JAK2;JAK1;OFD1;FGFBP1;NUDC;DUSP3;DUSP1;DUSP8;NCOR2;PSMA5;KAT2B;PSMA6;PSMA3;NCOR1;PSMA4;PLCB4;PSMA1;PSMA2;CDC42EP3;RAPGEF1;RARB;PLCB1;RAPGEF3;PAFAH1B1;HBEGF;ADCY3;PSMA7;CENPC;ADCY5;PSMB6;PSMB7;PSMB5;PSMB2;PSMB3;PSMB1;SPOP;AP2S1;LEO1;RPS27A;EVC2;WWOX;CSNK1A1;KIDINS220;WNT7B;STAT3;SCTR;DAB2IP;PSMB8;AGT;DHRS4;PSMC5;GPR143;PSMC6;PSMC3;ABI2;PSMC4;TAOK1;PSMC1;PSMC2;GOPC;PKN1;LAMTOR1;NOTUM;LAMTOR4;LGR5;CCL28;LAMTOR3;LAMTOR5;POGLUT1;FAM13B;UBE2D3;UBE2D1;TNF;FGF2;CXCL16;PPP1CB;PPP1CC;LEPR;AP2M1;USP8;RNF43;MAP2K2;FST;CTNNBIP1;USP2;F2R;GTF2F2;RBX1;PYGO1;CLIP1;MAPKAPK5;SQSTM1;CSNK1G2;KHDRBS1;SHC2;CUL3;CUL1;ITPR2;RASAL1;PHB;KLC1;KNG1;RAP1B;BAG4;RAP1A;SOCS1;DVL3;SRGAP3;RAB6A;SKIL;ARPC5;NFKB1;DRAP1;MOB1B;CDK9;MOB1A;ARPC2;CDK5;ARPC3;PTPRA;RHEB;CDK4;NOX[illegible] |
| 1416 | Voltage gated Potassium channels Homo sapiens R-HSA-1296072 | 1.00E+00 | 1.00E+00 | 7.98E-02 | 1.32E-06 | KCNQ1 |
| 1417 | Amine ligand-binding receptors Homo sapiens R-HSA-375280 | 1.00E+00 | 1.00E+00 | 7.98E-02 | 1.32E-06 | GPR143 |
| 1418 | Chemokine receptors bind chemokines Homo sapiens R-HSA-380108 | 1.00E+00 | 1.00E+00 | 1.24E-01 | 1.41E-06 | CCL28;CXCL16 |

| | | | | | | |
|---|---|---|---|---|---|---|
| 1419 | Signaling by GPCR Homo sapiens R-HSA-372790 | 1.00E+00 | 1.00E+00 | 4.39E-01 | 3.62E-06 | APP;PEBP1;WDR83;FGF2;CXCL16;PSMD8;PSMD9;PSMD6;PSMD7;EDNRB;PSMD4;AKT2;PSMD2;PSMD3;AKT1;PSMD1;MAP2K2;F2R;RBX1;BTC;TIAM1;PRKAR1A;AKAP9;PSME1;PSME2;TLN1;PSMD10;SHC2;CALCA;PSMD12;SPTBN5;PSMD11;PSMD14;GIPR;ARHGEF25;PSMD13;CUL3;PDE1A;PDGFB;LPAR2;PIK3R3;ITPR2;RASAL1;PHB;KNG1;RASGRP3;RAP1B;GNG10;RAP1A;PRKAR2B;PRKAR2A;UBC;PSMF1;PTK2;GNAO1;CDK5;PTPRA;FGF18;PNOC;GNAS;GRB2;GNB5;CALM1;DAGLA;YWHAB;RASGRF2;ARRB1;ARHGAP4;HEBP1;PPP3CB;PPP2R5E;GPER1;MCF2L;PEA15;BDKRB2;BDKRB1;PDE4A;RAC1;JAK2;SPTAN1;JAK1;VAV3;ARHGEF12;DUSP1;PDPK1;PDE4D;ARHGEF16;RHOG;PLA2G4A;ARHGEF18;GPBAR1;PPP2R5A;DUSP8;SSTR2;RHOC;RHOA;PSMA5;PSMA6;PSMA3;PSMA4;GAL;CREB1;PLCB4;PSMA1;PSMA2;RASA1;ARHGEF3;ARHGEF1;ARHGEF7;PLCB1;HBEGF;RGS19;GNAI3;ADCY3;PSMA7;GNAI1;GNAI2;ADCY5;PSMB6;CDC42;PPP2CA;PSMB7;PPP2CB;PSMB5;WNT11;PSMB2;GNG5;PSMB3;PSMB1;MAPK1;FYN;CSK;S1PR3;RPS27A;CAMK2G;CD55;WNT7B;SCTR;DAB2IP;PSMB8;AGT;PPP1CA;PSMC5;GPR143;PSMC6;PSMC3;PSMC4;PSMC1;PSMC2;F2RL1;KRAS;RGS12;RGS11;CCL28;LAMTOR3 |
| 1420 | Defensins Homo sapiens R-HSA-1461973 | 1.00E+00 | 1.00E+00 | 7.29E-02 | 5.75E-07 | PRSS3 |
| 1421 | GPCR downstream signaling Homo sapiens R-HSA-388396 | 1.00E+00 | 1.00E+00 | 2.27E-01 | 1.76E-06 | APP;DAGLA;RASGRF2;ARRB1;ARHGAP4;CXCL16;HEBP1;EDNRB;AKT2;GPER1;MCF2L;BDKRB2;AKT1;BDKRB1;PDE4A;RAC1;JAK2;JAK1;VAV3;ARHGEF12;PDPK1;PDE4D;ARHGEF16;F2R;RHOG;ARHGEF18;GPBAR1;SSTR2;RHOC;RHOA;TIAM1;GAL;PLCB4;ARHGEF3;ARHGEF1;ARHGEF7;PLCB1;CALCA;RGS19;GIPR;ARHGEF25;PDE1A;GNAI3;LPAR2;PIK3R3;ADCY3;ITPR2;GNAI1;KNG1;GNAI2;ADCY5;CDC42;GNG10;GNG5;S1PR3;SCTR;AGT;PNOC;GNAS;F2RL1;GRB2;RGS12;GNB5;RGS11;CCL28 |
| 1422 | GPCR ligand binding Homo sapiens R-HSA-500792 | 1.00E+00 | 1.00E+00 | 2.28E-01 | 1.46E-06 | APP;CALCA;GIPR;LPAR2;KNG1;CXCL16;HEBP1;GNG10;EDNRB;WNT11;GNG5;GPER1;BDKRB2;BDKRB1;S1PR3;CD55;WNT7B;F2R;SCTR;GPBAR1;SSTR2;AGT;GPR143;GAL;PNOC;GNAS;F2RL1;GNB5;CCL28 |
| 1423 | Complement cascade Homo sapiens R-HSA-166658 | 1.00E+00 | 1.00E+00 | 1.76E-01 | 1.05E-06 | CD59;CD46;CLU;CD55 |

| | | | | | | |
|---|---|---|---|---|---|---|
| 1424 | G alpha (q) signalling events Homo sapiens R-HSA-416476 | 1.00E+00 | 1.00E+00 | 3.69E-01 | 2.18E-06 | APP;DAGLA;RGS19;ARHGEF25;F2R;LPAR2;ITPR2;PIK3R3;KNG1;AGT;GNG10;EDNRB;PLCB4;GNG5;BDKRB2;BDKRB1;F2RL1;GNB5;PLCB1 |
| 1425 | Class A/1 (Rhodopsin-like receptors) Homo sapiens R-HSA-373076 | 1.00E+00 | 1.00E+00 | 2.07E-01 | 1.21E-06 | APP;F2R;LPAR2;GPBAR1;SSTR2;AGT;KNG1;CXCL16;HEBP1;GPR143;EDNRB;GAL;GPER1;PNOC;BDKRB2;F2RL1;BDKRB1;S1PR3;CCL28 |
| 1426 | G alpha (i) signalling events Homo sapiens R-HSA-418594 | 1.00E+00 | 1.00E+00 | 4.05E-01 | 2.34E-06 | APP;RGS19;GNAI3;LPAR2;ADCY3;GNAI1;KNG1;ADCY5;GNAI2;CXCL16;HEBP1;GNG10;GNG5;GPER1;BDKRB2;BDKRB1;S1PR3;SSTR2;AGT;GAL;PNOC;GNAS;RGS12;GNB5;RGS11;CCL28 |
| 1427 | Peptide ligand-binding receptors Homo sapiens R-HSA-375276 | 1.00E+00 | 1.00E+00 | 2.81E-01 | 1.42E-06 | APP;F2R;SSTR2;AGT;KNG1;CXCL16;HEBP1;EDNRB;GAL;GPER1;PNOC;BDKRB2;F2RL1;BDKRB1;CCL28 |
| 1428 | Binding and Uptake of Ligands by Scavenger Receptors Homo sapiens R-HSA-2173782 | 1.00E+00 | 1.00E+00 | 1.41E-01 | 6.78E-07 | HSP90AA1;HSPH1;CALR |
| 1429 | Immunoregulatory interactions between a Lymphoid and a non-Lymphoid cell Homo sapiens R-HSA-198933 | 1.00E+00 | 1.00E+00 | 1.32E-01 | 6.29E-07 | ITGB1;CXADR;ITGB2;HLA-C;HLA-G;HLA-E |
| 1430 | Potassium Channels Homo sapiens R-HSA-1296071 | 1.00E+00 | 1.00E+00 | 1.41E-01 | 5.81E-07 | GNG10;GNG5;KCNQ1;KCNJ15 |
| 1431 | Role of phospholipids in phagocytosis Homo sapiens R-HSA-2029485 | 1.00E+00 | 1.00E+00 | 5.68E-02 | 2.05E-07 | ITPR2 |

**Pathways enriched by genes enhanced with SGLT2 inhibitor in descending thin limb cells**

| | Term | P value | Adjusted P value | Odds Ratio | Combined Score | Genes |
|---|---|---|---|---|---|---|
| 1 | Diseases of glycosylation Homo sapiens R-HSA-3781865 | 5.78E-04 | 1.14E-01 | 5.32E+00 | 3.97E+01 | ADAMTS15;ADAMTS1;BGN;GPC3;GPC4;THBS1;THSD4 |
| 2 | Integrin cell surface interactions Homo sapiens R-HSA-216083 | 7.63E-04 | 1.14E-01 | 6.05E+00 | 4.34E+01 | COL4A2;COL4A1;ITGB3;SPP1;ITGB6;THBS1 |
| 3 | Extracellular matrix organization Homo sapiens R-HSA-1474244 | 7.81E-04 | 1.14E-01 | 2.99E+00 | 2.14E+01 | ITGB3;BGN;LAMB1;THBS1;BMP4;BMP2;COL4A2;COL5A1;COL4A1;ADAMTS1;COL6A1;SPP1;ITGB6 |
| 4 | Molecules associated with elastic fibres Homo sapiens R-HSA-2129379 | 1.34E-03 | 1.14E-01 | 9.42E+00 | 6.23E+01 | BMP4;BMP2;ITGB3;ITGB6 |
| 5 | NCAM signaling for neurite out-growth Homo sapiens R-HSA-375165 | 1.44E-03 | 1.14E-01 | 2.93E+00 | 1.92E+01 | CACNB2;KL;GRIN2A;FGF9;COL4A2;RASA3;COL4A1;ERBB4;ITGB3;COL6A1;FGFR4;FGFR3 |
| 6 | Axon guidance Homo sapiens R-HSA-422475 | 2.03E-03 | 1.14E-01 | 2.26E+00 | 1.40E+01 | KL;EPHA7;SEMA6D;ITGB3;LAMB1;DNM1;EFNB2;CACNB2;GRIN2A;FGF9;COL4A2;RASA3;COL4A1;ERBB4;COL6A1;SLIT3;FGFR4;FGFR3 |
| 7 | O-linked glycosylation Homo sapiens R-HSA-5173105 | 2.14E-03 | 1.14E-01 | 4.18E+00 | 2.57E+01 | ADAMTS15;GALNT14;ADAMTS1;GCNT4;THBS1;GALNT10;THSD4 |
| 8 | Developmental Biology Homo sapiens R-HSA-1266738 | 2.42E-03 | 1.14E-01 | 1.98E+00 | 1.19E+01 | KL;EPHA7;SMAD3;TSC22D1;SEMA6D;ITGB3;HNF1B;LAMB1;FOXO3;PBX1;DNM1;MED13L;EFNB2;CACNB2;GRIN2A;FGF9;COL4A2;RASA3;COL4A1;ERBB4;COL6A1;SLIT3;FGFR4;FGFR3 |
| 9 | Signaling by PDGF Homo sapiens R-HSA-186797 | 2.67E-03 | 1.14E-01 | 2.49E+00 | 1.47E+01 | KL;ITGB3;FOXO3;THBS1;GRIN2A;FGF9;COL4A2;RASA3;COL4A1;ERBB4;COL6A1;SPP1;FGFR4;FGFR3 |
| 10 | PI3K/AKT Signaling in Cancer Homo sapiens R-HSA-2219528 | 2.78E-03 | 1.14E-01 | 4.61E+00 | 2.71E+01 | KL;FGF9;ERBB4;FOXO3;FGFR4;FGFR3 |
| 11 | Defective B3GALTL causes Peters-plus syndrome (PpS) Homo sapiens R-HSA-5083635 | 2.96E-03 | 1.14E-01 | 7.42E+00 | 4.32E+01 | ADAMTS15;ADAMTS1;THBS1;THSD4 |
| 12 | NCAM1 interactions Homo sapiens R-HSA-419037 | 2.96E-03 | 1.14E-01 | 7.42E+00 | 4.32E+01 | CACNB2;COL4A2;COL4A1;COL6A1 |

| | | | | | | |
|---|---|---|---|---|---|---|
| 13 | Constitutive Signaling by Aberrant PI3K in Cancer Homo sapiens R-HSA-2219530 | 3.11E-03 | 1.14E-01 | 5.47E+00 | 3.16E+01 | KL;FGF9;ERBB4;FGFR4;FGFR3 |
| 14 | O-glycosylation of TSR domain-containing proteins Homo sapiens R-HSA-5173214 | 3.27E-03 | 1.14E-01 | 7.20E+00 | 4.12E+01 | ADAMTS15;ADAMTS1;THBS1;THSD4 |
| 15 | Defective B3GALT6 causes EDSP2 and SEMDJL1 Homo sapiens R-HSA-4420332 | 3.40E-03 | 1.14E-01 | 1.14E+01 | 6.51E+01 | BGN;GPC3;GPC4 |
| 16 | Defective B4GALT7 causes EDS, progeroid type Homo sapiens R-HSA-3560783 | 3.40E-03 | 1.14E-01 | 1.14E+01 | 6.51E+01 | BGN;GPC3;GPC4 |
| 17 | Defective B3GAT3 causes JDSSDHD Homo sapiens R-HSA-3560801 | 3.40E-03 | 1.14E-01 | 1.14E+01 | 6.51E+01 | BGN;GPC3;GPC4 |
| 18 | Elastic fibre formation Homo sapiens R-HSA-1566948 | 4.32E-03 | 1.37E-01 | 6.61E+00 | 3.60E+01 | BMP4;BMP2;ITGB3;ITGB6 |
| 19 | Signaling by PTK6 Homo sapiens R-HSA-8848021 | 4.66E-03 | 1.41E-01 | 4.94E+00 | 2.65E+01 | SOCS3;CCND1;RASA3;GPNMB;ERBB4 |
| 20 | SALM protein interactions at the synapse Homo sapiens R-HSA-8849932 | 5.21E-03 | 1.43E-01 | 9.64E+00 | 5.07E+01 | PTPRD;GRIN2A;PTPRF |
| 21 | FGFR3b ligand binding and activation Homo sapiens R-HSA-190371 | 5.24E-03 | 1.43E-01 | 2.44E+01 | 1.28E+02 | FGF9;FGFR3 |
| 22 | Signaling by BMP Homo sapiens R-HSA-201451 | 5.92E-03 | 1.54E-01 | 9.16E+00 | 4.70E+01 | BMP2;SMAD9;SMAD7 |
| 23 | Diseases of signal transduction Homo sapiens R-HSA-5663202 | 7.74E-03 | 1.72E-01 | 2.45E+00 | 1.19E+01 | KL;JAG1;SMAD3;CTBP2;CUX1;FGF9;ERBB4;RBP1;FOXO3;FGFR4;FGFR3 |
| 24 | Diseases associated with glycosaminoglycan metabolism Homo sapiens R-HSA-3560782 | 8.38E-03 | 1.72E-01 | 7.96E+00 | 3.81E+01 | GPC3;BGN;GPC4 |

| | | | | | | |
|---|---|---|---|---|---|---|
| 25 | A tetrasaccharide linker sequence is required for GAG synthesis Homo sapiens R-HSA-1971475 | 8.38E-03 | 1.72E-01 | 7.96E+00 | 3.81E+01 | GPC3;BGN;GPC4 |
| 26 | Chondroitin sulfate/dermatan sulfate metabolism Homo sapiens R-HSA-1793185 | 8.76E-03 | 1.72E-01 | 5.32E+00 | 2.52E+01 | BGN;GPC3;GPC4;CHST3 |
| 27 | Ca2+ activated K+ channels Homo sapiens R-HSA-1296052 | 8.79E-03 | 1.72E-01 | 1.74E+01 | 8.24E+01 | KCNMB2;KCNMA1 |
| 28 | Formation of annular gap junctions Homo sapiens R-HSA-196025 | 8.79E-03 | 1.72E-01 | 1.74E+01 | 8.24E+01 | DAB2;DNM1 |
| 29 | PI3K Cascade Homo sapiens R-HSA-109704 | 9.30E-03 | 1.72E-01 | 4.14E+00 | 1.94E+01 | KL;FGF9;RRAGD;FGFR4;FGFR3 |
| 30 | Gap junction degradation Homo sapiens R-HSA-190873 | 1.09E-02 | 1.72E-01 | 1.52E+01 | 6.88E+01 | DAB2;DNM1 |
| 31 | Acetylcholine regulates insulin secretion Homo sapiens R-HSA-399997 | 1.09E-02 | 1.72E-01 | 1.52E+01 | 6.88E+01 | MARCKS;GNAQ |
| 32 | PI5P, PP2A and IER3 Regulate PI3K/AKT Signaling Homo sapiens R-HSA-6811558 | 1.14E-02 | 1.72E-01 | 3.93E+00 | 1.76E+01 | KL;FGF9;ERBB4;FGFR4;FGFR3 |
| 33 | Heparan sulfate/heparin (HS-GAG) metabolism Homo sapiens R-HSA-1638091 | 1.14E-02 | 1.72E-01 | 4.89E+00 | 2.19E+01 | BGN;GPC3;GPC4;HS6ST1 |
| 34 | Assembly of collagen fibrils and other multimeric structures Homo sapiens R-HSA-2022090 | 1.14E-02 | 1.72E-01 | 4.89E+00 | 2.19E+01 | COL4A2;COL5A1;COL4A1;COL6A1 |
| 35 | Gastrin-CREB signalling pathway via PKC and MAPK Homo sapiens R-HSA-881907 | 1.15E-02 | 1.72E-01 | 2.07E+00 | 9.26E+00 | KL;EDN1;EDN2;ITGB3;AVPR1A;GNG11;GRIN2A;HRH1;FGF9;RASA3;ERBB4;GNAQ;FGFR4;FGFR3 |
| 36 | ECM proteoglycans Homo sapiens R-HSA-3000178 | 1.22E-02 | 1.72E-01 | 4.79E+00 | 2.11E+01 | ITGB3;BGN;LAMB1;ITGB6 |

| | | | | | | |
|---|---|---|---|---|---|---|
| 37 | Signaling by Interleukins Homo sapiens R-HSA-449147 | 1.23E-02 | 1.72E-01 | 2.12E+00 | 9.34E+00 | KL;IL1R1;ITGB3;PELI2;IL20RA;SOCS3;GRIN2A;FGF9;RASA3;ERBB4;BLNK;FGFR4;FGFR3 |
| 38 | HS-GAG biosynthesis Homo sapiens R-HSA-2022928 | 1.37E-02 | 1.72E-01 | 6.54E+00 | 2.81E+01 | GPC3;GPC4;HS6ST1 |
| 39 | PI-3K cascade:FGFR1 Homo sapiens R-HSA-5654689 | 1.48E-02 | 1.72E-01 | 3.17E+00 | 1.34E+01 | KL;FGF9;ERBB4;FOXO3;FGFR4;FGFR3 |
| 40 | PI-3K cascade:FGFR3 Homo sapiens R-HSA-5654710 | 1.48E-02 | 1.72E-01 | 3.17E+00 | 1.34E+01 | KL;FGF9;ERBB4;FOXO3;FGFR4;FGFR3 |
| 41 | PI3K events in ERBB4 signaling Homo sapiens R-HSA-1250342 | 1.48E-02 | 1.72E-01 | 3.17E+00 | 1.34E+01 | KL;FGF9;ERBB4;FOXO3;FGFR4;FGFR3 |
| 42 | PIP3 activates AKT signaling Homo sapiens R-HSA-1257604 | 1.48E-02 | 1.72E-01 | 3.17E+00 | 1.34E+01 | KL;FGF9;ERBB4;FOXO3;FGFR4;FGFR3 |
| 43 | PI-3K cascade:FGFR4 Homo sapiens R-HSA-5654720 | 1.48E-02 | 1.72E-01 | 3.17E+00 | 1.34E+01 | KL;FGF9;ERBB4;FOXO3;FGFR4;FGFR3 |
| 44 | PI-3K cascade:FGFR2 Homo sapiens R-HSA-5654695 | 1.48E-02 | 1.72E-01 | 3.17E+00 | 1.34E+01 | KL;FGF9;ERBB4;FOXO3;FGFR4;FGFR3 |
| 45 | SMAD2/SMAD3:SMAD4 heterotrimer regulates transcription Homo sapiens R-HSA-2173796 | 1.49E-02 | 1.72E-01 | 6.31E+00 | 2.66E+01 | SMAD3;JUNB;SMAD7 |
| 46 | Defective EXT2 causes exostoses 2 Homo sapiens R-HSA-3656237 | 1.56E-02 | 1.72E-01 | 1.22E+01 | 5.07E+01 | GPC3;GPC4 |
| 47 | Defective EXT1 causes exostoses 1, TRPS2 and CHDS Homo sapiens R-HSA-3656253 | 1.56E-02 | 1.72E-01 | 1.22E+01 | 5.07E+01 | GPC3;GPC4 |
| 48 | Lysine catabolism Homo sapiens R-HSA-71064 | 1.56E-02 | 1.72E-01 | 1.22E+01 | 5.07E+01 | CRYM;AASS |
| 49 | FGFR3 mutant receptor activation Homo sapiens R-HSA-2033514 | 1.56E-02 | 1.72E-01 | 1.22E+01 | 5.07E+01 | FGF9;FGFR3 |

| | | | | | | |
|---|---|---|---|---|---|---|
| 50 | Signaling by activated point mutants of FGFR3 Homo sapiens R-HSA-1839130 | 1.56E-02 | 1.72E-01 | 1.22E+01 | 5.07E+01 | FGF9;FGFR3 |
| 51 | Negative regulation of the PI3K/AKT network Homo sapiens R-HSA-199418 | 1.57E-02 | 1.72E-01 | 3.60E+00 | 1.50E+01 | KL;FGF9;ERBB4;FGFR4;FGFR3 |
| 52 | GAB1 signalosome Homo sapiens R-HSA-180292 | 1.65E-02 | 1.72E-01 | 3.09E+00 | 1.27E+01 | KL;FGF9;ERBB4;FOXO3;FGFR4;FGFR3 |
| 53 | PI3K/AKT activation Homo sapiens R-HSA-198203 | 1.65E-02 | 1.72E-01 | 3.09E+00 | 1.27E+01 | KL;FGF9;ERBB4;FOXO3;FGFR4;FGFR3 |
| 54 | Dissolution of Fibrin Clot Homo sapiens R-HSA-75205 | 1.82E-02 | 1.72E-01 | 1.11E+01 | 4.43E+01 | SERPINE2;SERPINF2 |
| 55 | Signaling by Activin Homo sapiens R-HSA-1502540 | 1.82E-02 | 1.72E-01 | 1.11E+01 | 4.43E+01 | SMAD3;FSTL3 |
| 56 | Activation of DNA fragmentation factor Homo sapiens R-HSA-211227 | 1.82E-02 | 1.72E-01 | 1.11E+01 | 4.43E+01 | HMGB2;HIST1H1D |
| 57 | Apoptosis induced DNA fragmentation Homo sapiens R-HSA-140342 | 1.82E-02 | 1.72E-01 | 1.11E+01 | 4.43E+01 | HMGB2;HIST1H1D |
| 58 | FGFR3c ligand binding and activation Homo sapiens R-HSA-190372 | 1.82E-02 | 1.72E-01 | 1.11E+01 | 4.43E+01 | FGF9;FGFR3 |
| 59 | FGFR3 ligand binding and activation Homo sapiens R-HSA-190239 | 1.82E-02 | 1.72E-01 | 1.11E+01 | 4.43E+01 | FGF9;FGFR3 |
| 60 | Diseases associated with O-glycosylation of proteins Homo sapiens R-HSA-3906995 | 1.83E-02 | 1.72E-01 | 4.21E+00 | 1.69E+01 | ADAMTS15;ADAMTS1;THBS1;THSD4 |
| 61 | Signaling by Leptin Homo sapiens R-HSA-2586552 | 1.83E-02 | 1.72E-01 | 2.37E+00 | 9.47E+00 | SOCS3;KL;GRIN2A;FGF9;RASA3;ERBB4;ITGB3;FGFR4;FGFR3 |
| 62 | Collagen biosynthesis and modifying enzymes Homo sapiens R-HSA-1650814 | 1.93E-02 | 1.75E-01 | 4.14E+00 | 1.64E+01 | COL4A2;COL5A1;COL4A1;COL6A1 |
| 63 | Signaling by FGFR in disease Homo sapiens R-HSA-1226099 | 1.93E-02 | 1.75E-01 | 4.14E+00 | 1.64E+01 | FGF9;CUX1;FGFR4;FGFR3 |

| | | | | | | |
|---|---|---|---|---|---|---|
| 64 | Signal Transduction Homo sapiens R-HSA-162582 | 2.00E-02 | 1.79E-01 | 1.39E+00 | 5.46E+00 | ARHGAP8;ITGB3;LFNG;FGF9;CCND1;TNFSF10;PDE4B;JUNB;SOX4;EDN1;EDN2;TLE1;AVPR1A;PRLR;DNM1;AR;SFRP1;COL4A2;RASA3;COL4A1;RBP1;RRAGD;COL6A1;ALDH1A1;CTBP2;PTGER2;FOXO3;THBS1;FSTL3;SOCS3;GRIN2A;C5;HRH1;GPNMB;FLRT3;ERBB4;GPC3;SPP1;FLNA;GPC4;KL;NTRK2;JAG1;SMAD3;CAV1;SMAD9;GNG11;DHRS3;SMAD7;BMP2;GNAQ;FGFR4;FGFR3 |
| 65 | Repression of WNT target genes Homo sapiens R-HSA-4641265 | 2.11E-02 | 1.80E-01 | 1.01E+01 | 3.92E+01 | TLE1;CTBP2 |
| 66 | Synthesis of bile acids and bile salts via 24-hydroxycholesterol Homo sapiens R-HSA-193775 | 2.11E-02 | 1.80E-01 | 1.01E+01 | 3.92E+01 | CYP27A1;SLC27A2 |
| 67 | FGFR4 ligand binding and activation Homo sapiens R-HSA-190322 | 2.11E-02 | 1.80E-01 | 1.01E+01 | 3.92E+01 | FGF9;FGFR4 |
| 68 | FGFR1 ligand binding and activation Homo sapiens R-HSA-190242 | 2.40E-02 | 2.03E-01 | 9.37E+00 | 3.49E+01 | KL;FGF9 |
| 69 | Cell surface interactions at the vascular wall Homo sapiens R-HSA-202733 | 2.46E-02 | 2.04E-01 | 3.19E+00 | 1.18E+01 | THBD;ANGPT2;CAV1;ITGB3;SIRPA |
| 70 | Neuronal System Homo sapiens R-HSA-112316 | 2.59E-02 | 2.12E-01 | 2.11E+00 | 7.72E+00 | GABRA2;PTPRD;CACNB2;GRIN2A;KCNMB2;KCNMA1;SLC6A13;KCNJ16;GNG11;PTPRF |
| 71 | Interleukin-3, 5 and GM-CSF signaling Homo sapiens R-HSA-512988 | 2.74E-02 | 2.21E-01 | 2.20E+00 | 7.90E+00 | KL;GRIN2A;FGF9;RASA3;ERBB4;ITGB3;BLNK;FGFR4;FGFR3 |
| 72 | Disease Homo sapiens R-HSA-1643685 | 2.85E-02 | 2.26E-01 | 1.67E+00 | 5.94E+00 | KL;JAG1;SMAD3;CTBP2;BGN;AVPR1A;FOXO3;THBS1;THSD4;ADAMTS15;CUX1;FGF9;ADAMTS1;ERBB4;RBP1;GPC3;GPC4;FGFR4;FGFR3 |
| 73 | Retinoid metabolism and transport Homo sapiens R-HSA-975634 | 3.06E-02 | 2.35E-01 | 4.69E+00 | 1.64E+01 | RBP1;GPC3;GPC4 |
| 74 | Non-integrin membrane-ECM interactions Homo sapiens R-HSA-3000171 | 3.06E-02 | 2.35E-01 | 4.69E+00 | 1.64E+01 | ITGB3;LAMB1;THBS1 |
| 75 | cGMP effects Homo sapiens R-HSA-418457 | 3.39E-02 | 2.35E-01 | 7.61E+00 | 2.57E+01 | KCNMB2;KCNMA1 |

| | | | | | | |
|---|---|---|---|---|---|---|
| 76 | SHC-mediated cascade:FGFR3 Homo sapiens R-HSA-5654704 | 3.39E-02 | 2.35E-01 | 7.61E+00 | 2.57E+01 | FGF9;FGFR3 |
| 77 | Transcriptional activity of SMAD2/SMAD3:SMAD4 heterotrimer Homo sapiens R-HSA-2173793 | 3.45E-02 | 2.35E-01 | 4.46E+00 | 1.50E+01 | SMAD3;JUNB;SMAD7 |
| 78 | Scavenging by Class A Receptors Homo sapiens R-HSA-3000480 | 3.75E-02 | 2.35E-01 | 7.16E+00 | 2.35E+01 | COL4A2;COL4A1 |
| 79 | Signaling by NODAL Homo sapiens R-HSA-1181150 | 3.75E-02 | 2.35E-01 | 7.16E+00 | 2.35E+01 | SMAD3;FOXO3 |
| 80 | GRB2 events in EGFR signaling Homo sapiens R-HSA-179812 | 3.88E-02 | 2.35E-01 | 2.16E+00 | 7.03E+00 | KL;GRIN2A;FGF9;RASA3;ERBB4;ITGB3;FGFR4;FGFR3 |
| 81 | SHC1 events in EGFR signaling Homo sapiens R-HSA-180336 | 3.88E-02 | 2.35E-01 | 2.16E+00 | 7.03E+00 | KL;GRIN2A;FGF9;RASA3;ERBB4;ITGB3;FGFR4;FGFR3 |
| 82 | SOS-mediated signalling Homo sapiens R-HSA-112412 | 3.88E-02 | 2.35E-01 | 2.16E+00 | 7.03E+00 | KL;GRIN2A;FGF9;RASA3;ERBB4;ITGB3;FGFR4;FGFR3 |
| 83 | SHC1 events in ERBB4 signaling Homo sapiens R-HSA-1250347 | 3.88E-02 | 2.35E-01 | 2.16E+00 | 7.03E+00 | KL;GRIN2A;FGF9;RASA3;ERBB4;ITGB3;FGFR4;FGFR3 |
| 84 | RAF/MAP kinase cascade Homo sapiens R-HSA-5673001 | 3.88E-02 | 2.35E-01 | 2.16E+00 | 7.03E+00 | KL;GRIN2A;FGF9;RASA3;ERBB4;ITGB3;FGFR4;FGFR3 |
| 85 | FRS-mediated FGFR2 signaling Homo sapiens R-HSA-5654700 | 3.96E-02 | 2.35E-01 | 2.15E+00 | 6.95E+00 | KL;GRIN2A;FGF9;RASA3;ERBB4;ITGB3;FGFR4;FGFR3 |
| 86 | FRS-mediated FGFR4 signaling Homo sapiens R-HSA-5654712 | 3.96E-02 | 2.35E-01 | 2.15E+00 | 6.95E+00 | KL;GRIN2A;FGF9;RASA3;ERBB4;ITGB3;FGFR4;FGFR3 |
| 87 | FRS-mediated FGFR3 signaling Homo sapiens R-HSA-5654706 | 3.96E-02 | 2.35E-01 | 2.15E+00 | 6.95E+00 | KL;GRIN2A;FGF9;RASA3;ERBB4;ITGB3;FGFR4;FGFR3 |
| 88 | FRS-mediated FGFR1 signaling Homo sapiens R-HSA-5654693 | 3.96E-02 | 2.35E-01 | 2.15E+00 | 6.95E+00 | KL;GRIN2A;FGF9;RASA3;ERBB4;ITGB3;FGFR4;FGFR3 |

| | | | | | | |
|---|---|---|---|---|---|---|
| 89 | Chondroitin sulfate biosynthesis Homo sapiens R-HSA-2022870 | 4.13E-02 | 2.35E-01 | 6.76E+00 | 2.16E+01 | BGN;CHST3 |
| 90 | Syndecan interactions Homo sapiens R-HSA-3000170 | 4.13E-02 | 2.35E-01 | 6.76E+00 | 2.16E+01 | ITGB3;THBS1 |
| 91 | Regulation of insulin secretion Homo sapiens R-HSA-422356 | 4.14E-02 | 2.35E-01 | 3.21E+00 | 1.02E+01 | CACNB2;MARCKS;GNAQ;GNG11 |
| 92 | ARMS-mediated activation Homo sapiens R-HSA-170984 | 4.21E-02 | 2.35E-01 | 2.12E+00 | 6.73E+00 | KL;GRIN2A;FGF9;RASA3;ERBB4;ITGB3;FGFR4;FGFR3 |
| 93 | Signalling to p38 via RIT and RIN Homo sapiens R-HSA-187706 | 4.21E-02 | 2.35E-01 | 2.12E+00 | 6.73E+00 | KL;GRIN2A;FGF9;RASA3;ERBB4;ITGB3;FGFR4;FGFR3 |
| 94 | NGF signalling via TRKA from the plasma membrane Homo sapiens R-HSA-187037 | 4.29E-02 | 2.35E-01 | 1.86E+00 | 5.87E+00 | KL;NTRK2;GRIN2A;FGF9;RASA3;ERBB4;ITGB3;FOXO3;FGFR4;FGFR3;DNM1 |
| 95 | EPH-ephrin mediated repulsion of cells Homo sapiens R-HSA-3928665 | 4.30E-02 | 2.35E-01 | 4.06E+00 | 1.28E+01 | EFNB2;EPHA7;DNM1 |
| 96 | Frs2-mediated activation Homo sapiens R-HSA-170968 | 4.30E-02 | 2.35E-01 | 2.11E+00 | 6.66E+00 | KL;GRIN2A;FGF9;RASA3;ERBB4;ITGB3;FGFR4;FGFR3 |
| 97 | MAPK family signaling cascades Homo sapiens R-HSA-5683057 | 4.32E-02 | 2.35E-01 | 2.01E+00 | 6.31E+00 | KL;GRIN2A;FGF9;RASA3;ERBB4;ITGB3;FOXO3;FGFR4;FGFR3 |
| 98 | IRS-mediated signalling Homo sapiens R-HSA-112399 | 4.32E-02 | 2.35E-01 | 2.01E+00 | 6.31E+00 | KL;GRIN2A;FGF9;RASA3;ERBB4;ITGB3;RRAGD;FGFR4;FGFR3 |
| 99 | MAPK1/MAPK3 signaling Homo sapiens R-HSA-5684996 | 4.39E-02 | 2.35E-01 | 2.11E+00 | 6.58E+00 | KL;GRIN2A;FGF9;RASA3;ERBB4;ITGB3;FGFR4;FGFR3 |
| 100 | Signaling by ERBB4 Homo sapiens R-HSA-1236394 | 4.41E-02 | 2.35E-01 | 1.92E+00 | 5.99E+00 | KL;GRIN2A;FGF9;RASA3;ERBB4;ITGB3;FOXO3;FGFR4;PRLR;FGFR3 |
| 101 | Prolonged ERK activation events Homo sapiens R-HSA-169893 | 4.48E-02 | 2.35E-01 | 2.10E+00 | 6.51E+00 | KL;GRIN2A;FGF9;RASA3;ERBB4;ITGB3;FGFR4;FGFR3 |
| 102 | HS-GAG degradation Homo sapiens R-HSA-2024096 | 4.51E-02 | 2.35E-01 | 6.41E+00 | 1.98E+01 | GPC3;GPC4 |

| | | | | | | |
|---|---|---|---|---|---|---|
| 103 | SHC-mediated cascade:FGFR1 Homo sapiens R-HSA-5654688 | 4.51E-02 | 2.35E-01 | 6.41E+00 | 1.98E+01 | KL;FGF9 |
| 104 | Downstream signaling of activated FGFR1 Homo sapiens R-HSA-5654687 | 4.57E-02 | 2.35E-01 | 1.91E+00 | 5.89E+00 | KL;GRIN2A;FGF9;RASA3;FLRT3;ERBB4;ITGB3;FOXO3;FGFR4;FGFR3 |
| 105 | Insulin receptor signalling cascade Homo sapiens R-HSA-74751 | 4.57E-02 | 2.35E-01 | 1.99E+00 | 6.13E+00 | KL;GRIN2A;FGF9;RASA3;ERBB4;ITGB3;RRAGD;FGFR4;FGFR3 |
| 106 | IGF1R signaling cascade Homo sapiens R-HSA-2428924 | 4.65E-02 | 2.35E-01 | 1.98E+00 | 6.08E+00 | KL;GRIN2A;FGF9;RASA3;ERBB4;ITGB3;RRAGD;FGFR4;FGFR3 |
| 107 | Signaling by Type 1 Insulin-like Growth Factor 1 Receptor (IGF1R) Homo sapiens R-HSA-2404192 | 4.65E-02 | 2.35E-01 | 1.98E+00 | 6.08E+00 | KL;GRIN2A;FGF9;RASA3;ERBB4;ITGB3;RRAGD;FGFR4;FGFR3 |
| 108 | IRS-related events triggered by IGF1R Homo sapiens R-HSA-2428928 | 4.65E-02 | 2.35E-01 | 1.98E+00 | 6.08E+00 | KL;GRIN2A;FGF9;RASA3;ERBB4;ITGB3;RRAGD;FGFR4;FGFR3 |
| 109 | Cytokine Signaling in Immune system Homo sapiens R-HSA-1280215 | 4.72E-02 | 2.35E-01 | 1.63E+00 | 4.99E+00 | EGR1;KL;IL1R1;ITGB3;PELI2;IL20RA;PRLR;SOCS3;GRIN2A;FGF9;RASA3;ERBB4;BLNK;FGFR4;FGFR3;TRIM45 |
| 110 | FCERI mediated MAPK activation Homo sapiens R-HSA-2871796 | 4.73E-02 | 2.35E-01 | 1.97E+00 | 6.02E+00 | KL;GRIN2A;FGF9;RASA3;ERBB4;ITGB3;FOS;FGFR4;FGFR3 |
| 111 | Interleukin receptor SHC signaling Homo sapiens R-HSA-912526 | 4.75E-02 | 2.35E-01 | 2.07E+00 | 6.31E+00 | KL;GRIN2A;FGF9;RASA3;ERBB4;ITGB3;FGFR4;FGFR3 |
| 112 | Glycosaminoglycan metabolism Homo sapiens R-HSA-1630316 | 4.77E-02 | 2.35E-01 | 2.63E+00 | 8.02E+00 | BGN;GPC3;GPC4;HS6ST1;CHST3 |
| 113 | Signalling to RAS Homo sapiens R-HSA-167044 | 4.84E-02 | 2.35E-01 | 2.06E+00 | 6.24E+00 | KL;GRIN2A;FGF9;RASA3;ERBB4;ITGB3;FGFR4;FGFR3 |
| 114 | Signaling by FGFR1 Homo sapiens R-HSA-5654736 | 4.88E-02 | 2.35E-01 | 1.88E+00 | 5.69E+00 | KL;GRIN2A;FGF9;RASA3;FLRT3;ERBB4;ITGB3;FOXO3;FGFR4;FGFR3 |
| 115 | RA biosynthesis pathway Homo sapiens R-HSA-5365859 | 4.91E-02 | 2.35E-01 | 6.09E+00 | 1.83E+01 | ALDH1A1;DHRS3 |

| | | | | | | |
|---|---|---|---|---|---|---|
| 116 | Common Pathway of Fibrin Clot Formation Homo sapiens R-HSA-140875 | 4.91E-02 | 2.35E-01 | 6.09E+00 | 1.83E+01 | THBD;SERPINE2 |
| 117 | Signaling by FGFR3 point mutants in cancer Homo sapiens R-HSA-8853338 | 4.91E-02 | 2.35E-01 | 6.09E+00 | 1.83E+01 | FGF9;FGFR3 |
| 118 | Signaling by FGFR3 in disease Homo sapiens R-HSA-5655332 | 4.91E-02 | 2.35E-01 | 6.09E+00 | 1.83E+01 | FGF9;FGFR3 |
| 119 | Role of LAT2/NTAL/LAB on calcium mobilization Homo sapiens R-HSA-2730905 | 4.93E-02 | 2.35E-01 | 2.35E+00 | 7.08E+00 | KL;FGF9;ERBB4;FOXO3;FGFR4;FGFR3 |
| 120 | Collagen formation Homo sapiens R-HSA-1474290 | 4.98E-02 | 2.35E-01 | 3.01E+00 | 9.04E+00 | COL4A2;COL5A1;COL4A1;COL6A1 |
| 121 | Metabolism of fat-soluble vitamins Homo sapiens R-HSA-6806667 | 5.00E-02 | 2.35E-01 | 3.81E+00 | 1.14E+01 | RBP1;GPC3;GPC4 |
| 122 | Apoptotic execution phase Homo sapiens R-HSA-75153 | 5.00E-02 | 2.35E-01 | 3.81E+00 | 1.14E+01 | HMGB2;HIST1H1D;DSG2 |
| 123 | VEGFR2 mediated cell proliferation Homo sapiens R-HSA-5218921 | 5.03E-02 | 2.35E-01 | 2.04E+00 | 6.11E+00 | KL;GRIN2A;FGF9;RASA3;ERBB4;ITGB3;FGFR4;FGFR3 |
| 124 | Thromboxane signalling through TP receptor Homo sapiens R-HSA-428930 | 5.32E-02 | 2.44E-01 | 5.80E+00 | 1.70E+01 | GNAQ;GNG11 |
| 125 | SHC-mediated cascade:FGFR4 Homo sapiens R-HSA-5654719 | 5.32E-02 | 2.44E-01 | 5.80E+00 | 1.70E+01 | FGF9;FGFR4 |
| 126 | Interleukin-2 signaling Homo sapiens R-HSA-451927 | 5.43E-02 | 2.47E-01 | 2.01E+00 | 5.85E+00 | KL;GRIN2A;FGF9;RASA3;ERBB4;ITGB3;FGFR4;FGFR3 |
| 127 | Signalling to ERKs Homo sapiens R-HSA-187687 | 5.53E-02 | 2.50E-01 | 2.00E+00 | 5.79E+00 | KL;GRIN2A;FGF9;RASA3;ERBB4;ITGB3;FGFR4;FGFR3 |
| 128 | Synthesis of bile acids and bile salts via 7alpha-hydroxycholesterol Homo sapiens R-HSA-193368 | 5.75E-02 | 2.55E-01 | 5.53E+00 | 1.58E+01 | CYP27A1;SLC27A2 |

| | | | | | | |
|---|---|---|---|---|---|---|
| 129 | Growth hormone receptor signaling Homo sapiens R-HSA-982772 | 5.75E-02 | 2.55E-01 | 5.53E+00 | 1.58E+01 | SOCS3;PRLR |
| 130 | ADP signalling through P2Y purinoceptor 1 Homo sapiens R-HSA-418592 | 6.18E-02 | 2.70E-01 | 5.29E+00 | 1.47E+01 | GNAQ;GNG11 |
| 131 | Nitric oxide stimulates guanylate cyclase Homo sapiens R-HSA-392154 | 6.18E-02 | 2.70E-01 | 5.29E+00 | 1.47E+01 | KCNMB2;KCNMA1 |
| 132 | Downregulation of TGF-beta receptor signaling Homo sapiens R-HSA-2173788 | 6.62E-02 | 2.88E-01 | 5.07E+00 | 1.38E+01 | SMAD3;SMAD7 |
| 133 | Signaling by Insulin receptor Homo sapiens R-HSA-74752 | 6.85E-02 | 2.95E-01 | 1.83E+00 | 4.90E+00 | KL;GRIN2A;FGF9;RASA3;ERBB4;ITGB3;RRAGD;FGFR4;FGFR3 |
| 134 | Synthesis of bile acids and bile salts Homo sapiens R-HSA-192105 | 7.08E-02 | 3.03E-01 | 4.87E+00 | 1.29E+01 | CYP27A1;SLC27A2 |
| 135 | Visual phototransduction Homo sapiens R-HSA-2187338 | 7.35E-02 | 3.12E-01 | 2.62E+00 | 6.85E+00 | RBP1;GPC3;GPC4;DHRS3 |
| 136 | Adrenaline,noradrenaline inhibits insulin secretion Homo sapiens R-HSA-400042 | 7.55E-02 | 3.15E-01 | 4.68E+00 | 1.21E+01 | CACNB2;GNG11 |
| 137 | Gap junction trafficking Homo sapiens R-HSA-190828 | 7.55E-02 | 3.15E-01 | 4.68E+00 | 1.21E+01 | DAB2;DNM1 |
| 138 | Signaling by FGFR Homo sapiens R-HSA-190236 | 7.68E-02 | 3.15E-01 | 1.72E+00 | 4.42E+00 | KL;GRIN2A;FGF9;RASA3;FLRT3;ERBB4;ITGB3;FOXO3;FGFR4;FGFR3 |
| 139 | VEGFA-VEGFR2 Pathway Homo sapiens R-HSA-4420097 | 7.86E-02 | 3.15E-01 | 1.77E+00 | 4.51E+00 | KL;GRIN2A;FGF9;RASA3;ERBB4;CAV1;ITGB3;FGFR4;FGFR3 |
| 140 | Abacavir transmembrane transport Homo sapiens R-HSA-2161517 | 7.87E-02 | 3.15E-01 | 1.52E+01 | 3.86E+01 | ABCB1 |
| 141 | NOSTRIN mediated eNOS trafficking Homo sapiens R-HSA-203641 | 7.87E-02 | 3.15E-01 | 1.52E+01 | 3.86E+01 | CAV1 |

| | | | | | | |
|---|---|---|---|---|---|---|
| 142 | NGF-independant TRKA activation Homo sapiens R-HSA-187024 | 7.87E-02 | 3.15E-01 | 1.52E+01 | 3.86E+01 | NTRK2 |
| 143 | Alpha-oxidation of phytanate Homo sapiens R-HSA-389599 | 7.87E-02 | 3.15E-01 | 1.52E+01 | 3.86E+01 | SLC27A2 |
| 144 | Negative regulation of FGFR3 signaling Homo sapiens R-HSA-5654732 | 8.02E-02 | 3.15E-01 | 4.51E+00 | 1.14E+01 | FGF9;FGFR3 |
| 145 | Synthesis of substrates in N-glycan biosythesis Homo sapiens R-HSA-446219 | 8.29E-02 | 3.15E-01 | 3.05E+00 | 7.58E+00 | NEU3;GMDS;ST3GAL5 |
| 146 | Signaling by SCF-KIT Homo sapiens R-HSA-1433557 | 8.46E-02 | 3.15E-01 | 1.74E+00 | 4.31E+00 | KL;GRIN2A;FGF9;RASA3;ERBB4;ITGB3;FOXO3;FGFR4;FGFR3 |
| 147 | Gap junction trafficking and regulation Homo sapiens R-HSA-157858 | 8.50E-02 | 3.15E-01 | 4.34E+00 | 1.07E+01 | DAB2;DNM1 |
| 148 | Signaling by the B Cell Receptor (BCR) Homo sapiens R-HSA-983705 | 8.67E-02 | 3.15E-01 | 1.89E+00 | 4.63E+00 | KL;FGF9;ERBB4;BLNK;FOXO3;FGFR4;FGFR3 |
| 149 | MHC class II antigen presentation Homo sapiens R-HSA-2132295 | 8.72E-02 | 3.15E-01 | 2.46E+00 | 6.01E+00 | CTSO;HLA-DOA;CTSC;DNM1 |
| 150 | Signaling by VEGF Homo sapiens R-HSA-194138 | 8.83E-02 | 3.15E-01 | 1.73E+00 | 4.19E+00 | KL;GRIN2A;FGF9;RASA3;ERBB4;CAV1;ITGB3;FGFR4;FGFR3 |
| 151 | Downstream signaling of activated FGFR2 Homo sapiens R-HSA-5654696 | 8.96E-02 | 3.15E-01 | 1.72E+00 | 4.16E+00 | KL;GRIN2A;FGF9;RASA3;ERBB4;ITGB3;FOXO3;FGFR4;FGFR3 |
| 152 | Downstream signaling of activated FGFR4 Homo sapiens R-HSA-5654716 | 8.96E-02 | 3.15E-01 | 1.72E+00 | 4.16E+00 | KL;GRIN2A;FGF9;RASA3;ERBB4;ITGB3;FOXO3;FGFR4;FGFR3 |
| 153 | Downstream signaling of activated FGFR3 Homo sapiens R-HSA-5654708 | 8.96E-02 | 3.15E-01 | 1.72E+00 | 4.16E+00 | KL;GRIN2A;FGF9;RASA3;ERBB4;ITGB3;FOXO3;FGFR4;FGFR3 |
| 154 | Negative regulation of FGFR4 signaling Homo sapiens R-HSA-5654733 | 9.00E-02 | 3.15E-01 | 4.19E+00 | 1.01E+01 | FGF9;FGFR4 |

| | | | | | | |
|---|---|---|---|---|---|---|
| 155 | FGFR1 mutant receptor activation Homo sapiens R-HSA-1839124 | 9.00E-02 | 3.15E-01 | 4.19E+00 | 1.01E+01 | FGF9;CUX1 |
| 156 | Platelet degranulation Homo sapiens R-HSA-114608 | 9.20E-02 | 3.15E-01 | 2.41E+00 | 5.76E+00 | ITGB3;SERPINF2;FLNA;THBS1 |
| 157 | G alpha (q) signalling events Homo sapiens R-HSA-416476 | 9.21E-02 | 3.15E-01 | 1.98E+00 | 4.73E+00 | HRH1;EDN1;EDN2;GNAQ;AVPR1A;GNG11 |
| 158 | O-linked glycosylation of mucins Homo sapiens R-HSA-913709 | 9.23E-02 | 3.15E-01 | 2.90E+00 | 6.91E+00 | GALNT14;GCNT4;GALNT10 |
| 159 | Signaling by FGFR4 Homo sapiens R-HSA-5654743 | 9.35E-02 | 3.15E-01 | 1.71E+00 | 4.04E+00 | KL;GRIN2A;FGF9;RASA3;ERBB4;ITGB3;FOXO3;FGFR4;FGFR3 |
| 160 | Vasopressin-like receptors Homo sapiens R-HSA-388479 | 9.36E-02 | 3.15E-01 | 1.21E+01 | 2.88E+01 | AVPR1A |
| 161 | Fructose metabolism Homo sapiens R-HSA-5652084 | 9.36E-02 | 3.15E-01 | 1.21E+01 | 2.88E+01 | ALDH1A1 |
| 162 | The retinoid cycle in cones (daylight vision) Homo sapiens R-HSA-2187335 | 9.36E-02 | 3.15E-01 | 1.21E+01 | 2.88E+01 | DHRS3 |
| 163 | Activation of C3 and C5 Homo sapiens R-HSA-174577 | 9.36E-02 | 3.15E-01 | 1.21E+01 | 2.88E+01 | C5 |
| 164 | GDP-fucose biosynthesis Homo sapiens R-HSA-6787639 | 9.36E-02 | 3.15E-01 | 1.21E+01 | 2.88E+01 | GMDS |
| 165 | Activation of TRKA receptors Homo sapiens R-HSA-187015 | 9.36E-02 | 3.15E-01 | 1.21E+01 | 2.88E+01 | NTRK2 |
| 166 | SMAD2/3 Phosphorylation Motif Mutants in Cancer Homo sapiens R-HSA-3304356 | 9.36E-02 | 3.15E-01 | 1.21E+01 | 2.88E+01 | SMAD3 |
| 167 | PTK6 Regulates Cell Cycle Homo sapiens R-HSA-8849470 | 9.36E-02 | 3.15E-01 | 1.21E+01 | 2.88E+01 | CCND1 |
| 168 | Downstream signaling events of B Cell Receptor (BCR) Homo sapiens R-HSA-1168372 | 9.38E-02 | 3.15E-01 | 1.97E+00 | 4.66E+00 | KL;FGF9;ERBB4;FOXO3;FGFR4;FGFR3 |

| | | | | | | |
|---|---|---|---|---|---|---|
| 169 | Signaling by FGFR3 Homo sapiens R-HSA-5654741 | 9.48E-02 | 3.15E-01 | 1.70E+00 | 4.01E+00 | KL;GRIN2A;FGF9;RASA3;ERBB4;ITGB3;FOXO3;FGFR4;FGFR3 |
| 170 | Signal amplification Homo sapiens R-HSA-392518 | 9.50E-02 | 3.15E-01 | 4.05E+00 | 9.55E+00 | GNAQ;GNG11 |
| 171 | Thrombin signalling through proteinase activated receptors (PARs) Homo sapiens R-HSA-456926 | 9.50E-02 | 3.15E-01 | 4.05E+00 | 9.55E+00 | GNAQ;GNG11 |
| 172 | TGF-beta receptor signaling activates SMADs Homo sapiens R-HSA-2173789 | 9.50E-02 | 3.15E-01 | 4.05E+00 | 9.55E+00 | SMAD3;SMAD7 |
| 173 | Negative regulation of FGFR1 signaling Homo sapiens R-HSA-5654726 | 9.50E-02 | 3.15E-01 | 4.05E+00 | 9.55E+00 | KL;FGF9 |
| 174 | Sialic acid metabolism Homo sapiens R-HSA-4085001 | 1.00E-01 | 3.22E-01 | 3.92E+00 | 9.03E+00 | NEU3;ST3GAL5 |
| 175 | Cargo concentration in the ER Homo sapiens R-HSA-5694530 | 1.00E-01 | 3.22E-01 | 3.92E+00 | 9.03E+00 | FOLR1;CTSC |
| 176 | Response to elevated platelet cytosolic Ca2+ Homo sapiens R-HSA-76005 | 1.05E-01 | 3.22E-01 | 2.30E+00 | 5.19E+00 | ITGB3;SERPINF2;FLNA;THBS1 |
| 177 | Integration of energy metabolism Homo sapiens R-HSA-163685 | 1.05E-01 | 3.22E-01 | 2.30E+00 | 5.19E+00 | CACNB2;MARCKS;GNAQ;GNG11 |
| 178 | Downstream signal transduction Homo sapiens R-HSA-186763 | 1.06E-01 | 3.22E-01 | 1.66E+00 | 3.73E+00 | KL;GRIN2A;FGF9;RASA3;ERBB4;ITGB3;FOXO3;FGFR4;FGFR3 |
| 179 | Nectin/Necl trans heterodimerization Homo sapiens R-HSA-420597 | 1.08E-01 | 3.22E-01 | 1.01E+01 | 2.25E+01 | CADM1 |
| 180 | Cation-coupled Chloride cotransporters Homo sapiens R-HSA-426117 | 1.08E-01 | 3.22E-01 | 1.01E+01 | 2.25E+01 | SLC12A1 |
| 181 | SLC transporter disorders Homo sapiens R-HSA-5619102 | 1.08E-01 | 3.22E-01 | 1.01E+01 | 2.25E+01 | AVPR1A |

| | | | | | | |
|---|---|---|---|---|---|---|
| 182 | Defective CHST3 causes SEDCJD Homo sapiens R-HSA-3595172 | 1.08E-01 | 3.22E-01 | 1.01E+01 | 2.25E+01 | BGN |
| 183 | Defective CHSY1 causes TPBS Homo sapiens R-HSA-3595177 | 1.08E-01 | 3.22E-01 | 1.01E+01 | 2.25E+01 | BGN |
| 184 | Defective CHST14 causes EDS, musculocontractural type Homo sapiens R-HSA-3595174 | 1.08E-01 | 3.22E-01 | 1.01E+01 | 2.25E+01 | BGN |
| 185 | Loss of Function of TGFBR1 in Cancer Homo sapiens R-HSA-3656534 | 1.08E-01 | 3.22E-01 | 1.01E+01 | 2.25E+01 | SMAD3 |
| 186 | TGFBR1 KD Mutants in Cancer Homo sapiens R-HSA-3656532 | 1.08E-01 | 3.22E-01 | 1.01E+01 | 2.25E+01 | SMAD3 |
| 187 | Loss of Function of SMAD2/3 in Cancer Homo sapiens R-HSA-3304349 | 1.08E-01 | 3.22E-01 | 1.01E+01 | 2.25E+01 | SMAD3 |
| 188 | SMAD2/3 MH2 Domain Mutants in Cancer Homo sapiens R-HSA-3315487 | 1.08E-01 | 3.22E-01 | 1.01E+01 | 2.25E+01 | SMAD3 |
| 189 | Regulation of gene expression in late stage (branching morphogenesis) pancreatic bud precursor cells Homo sapiens R-HSA-210744 | 1.08E-01 | 3.22E-01 | 1.01E+01 | 2.25E+01 | HNF1B |
| 190 | Signaling by NOTCH1 t(7;9)(NOTCH1:M1580 K2555) Translocation Mutant Homo sapiens R-HSA-2660825 | 1.08E-01 | 3.22E-01 | 1.01E+01 | 2.25E+01 | JAG1 |
| 191 | Constitutive Signaling by NOTCH1 t(7;9)(NOTCH1:M1580 K2555) Translocation Mutant Homo sapiens R-HSA-2660826 | 1.08E-01 | 3.22E-01 | 1.01E+01 | 2.25E+01 | JAG1 |
| 192 | PTK6 promotes HIF1A stabilization Homo sapiens R-HSA-8857538 | 1.08E-01 | 3.22E-01 | 1.01E+01 | 2.25E+01 | GPNMB |

| | | | | | | |
|---|---|---|---|---|---|---|
| 193 | TRAIL signaling Homo sapiens R-HSA-75158 | 1.08E-01 | 3.22E-01 | 1.01E+01 | 2.25E+01 | TNFSF10 |
| 194 | DAP12 signaling Homo sapiens R-HSA-2424491 | 1.10E-01 | 3.24E-01 | 1.64E+00 | 3.63E+00 | KL;GRIN2A;FGF9;RASA3;ERBB4;ITGB3;FOXO3;FGFR4;FGFR3 |
| 195 | Bile acid and bile salt metabolism Homo sapiens R-HSA-194068 | 1.10E-01 | 3.25E-01 | 3.69E+00 | 8.12E+00 | CYP27A1;SLC27A2 |
| 196 | Fc epsilon receptor (FCERI) signaling Homo sapiens R-HSA-2454202 | 1.12E-01 | 3.27E-01 | 1.59E+00 | 3.48E+00 | KL;GRIN2A;FGF9;RASA3;ERBB4;ITGB3;FOS;FOXO3;FGFR4;FGFR3 |
| 197 | Ion channel transport Homo sapiens R-HSA-983712 | 1.14E-01 | 3.32E-01 | 1.86E+00 | 4.03E+00 | GABRA2;CLCN5;TSC22D3;ATP13A4;FXYD6;TRPM3 |
| 198 | Signaling by TGF-beta Receptor Complex Homo sapiens R-HSA-170834 | 1.16E-01 | 3.35E-01 | 2.61E+00 | 5.63E+00 | SMAD3;JUNB;SMAD7 |
| 199 | Hemostasis Homo sapiens R-HSA-109582 | 1.17E-01 | 3.36E-01 | 1.48E+00 | 3.17E+00 | ANGPT2;SERPINE2;CAV1;ITGB3;SERPINF2;GNG11;THBS1;THBD;GNAQ;KCNMB2;KCNMA1;FLNA;SIRPA |
| 200 | Adaptive Immune System Homo sapiens R-HSA-1280218 | 1.17E-01 | 3.36E-01 | 1.40E+00 | 3.01E+00 | KL;ZBTB16;KLHL13;FOXO3;DNM1;MRC2;RNF144B;SOCS3;CTSO;FGF9;ERBB4;BLNK;HLA-DOA;FGFR4;FGFR3;CD200;CTSC |
| 201 | Signalling by NGF Homo sapiens R-HSA-166520 | 1.18E-01 | 3.36E-01 | 1.54E+00 | 3.28E+00 | KL;NTRK2;GRIN2A;FGF9;RASA3;ERBB4;ITGB3;FOXO3;FGFR4;FGFR3;DNM1 |
| 202 | Terminal pathway of complement Homo sapiens R-HSA-166665 | 1.23E-01 | 3.38E-01 | 8.67E+00 | 1.82E+01 | C5 |
| 203 | Free fatty acids regulate insulin secretion Homo sapiens R-HSA-400451 | 1.23E-01 | 3.38E-01 | 8.67E+00 | 1.82E+01 | GNAQ |
| 204 | Fatty Acids bound to GPR40 (FFAR1) regulate insulin secretion Homo sapiens R-HSA-434316 | 1.23E-01 | 3.38E-01 | 8.67E+00 | 1.82E+01 | GNAQ |
| 205 | Signaling by TGF-beta Receptor Complex in Cancer Homo sapiens R-HSA-3304351 | 1.23E-01 | 3.38E-01 | 8.67E+00 | 1.82E+01 | SMAD3 |
| 206 | Hydrolysis of LPC Homo sapiens R-HSA-1483115 | 1.23E-01 | 3.38E-01 | 8.67E+00 | 1.82E+01 | GPCPD1 |

| | | | | | | |
|---|---|---|---|---|---|---|
| 207 | Regulation of gene expression in early pancreatic precursor cells Homo sapiens R-HSA-210747 | 1.23E-01 | 3.38E-01 | 8.67E+00 | 1.82E+01 | HNF1B |
| 208 | Downregulation of ERBB4 signaling Homo sapiens R-HSA-1253288 | 1.23E-01 | 3.38E-01 | 8.67E+00 | 1.82E+01 | ERBB4 |
| 209 | Signaling by EGFR Homo sapiens R-HSA-177929 | 1.26E-01 | 3.45E-01 | 1.59E+00 | 3.29E+00 | KL;GRIN2A;FGF9;RASA3;ERBB4;ITGB3;FOXO3;FGFR4;FGFR3 |
| 210 | Signaling by FGFR1 in disease Homo sapiens R-HSA-5655302 | 1.27E-01 | 3.45E-01 | 3.38E+00 | 6.98E+00 | FGF9;CUX1 |
| 211 | Transmission across Chemical Synapses Homo sapiens R-HSA-112315 | 1.30E-01 | 3.54E-01 | 1.79E+00 | 3.64E+00 | GABRA2;CACNB2;GRIN2A;SLC6A13;KCNJ16;GNG11 |
| 212 | Formation of Fibrin Clot (Clotting Cascade) Homo sapiens R-HSA-140877 | 1.32E-01 | 3.55E-01 | 3.29E+00 | 6.66E+00 | THBD;SERPINE2 |
| 213 | Nuclear signaling by ERBB4 Homo sapiens R-HSA-1251985 | 1.32E-01 | 3.55E-01 | 3.29E+00 | 6.66E+00 | ERBB4;PRLR |
| 214 | DAP12 interactions Homo sapiens R-HSA-2172127 | 1.32E-01 | 3.55E-01 | 1.57E+00 | 3.18E+00 | KL;GRIN2A;FGF9;RASA3;ERBB4;ITGB3;FOXO3;FGFR4;FGFR3 |
| 215 | Biosynthesis of the N-glycan precursor (dolichol lipid-linked oligosaccharide, LLO) and transfer to a nascent protein Homo sapiens R-HSA-446193 | 1.34E-01 | 3.55E-01 | 2.43E+00 | 4.90E+00 | NEU3;GMDS;ST3GAL5 |
| 216 | Signaling by FGFR2 Homo sapiens R-HSA-5654738 | 1.36E-01 | 3.55E-01 | 1.56E+00 | 3.12E+00 | KL;GRIN2A;FGF9;RASA3;ERBB4;ITGB3;FOXO3;FGFR4;FGFR3 |
| 217 | Retinoid cycle disease events Homo sapiens R-HSA-2453864 | 1.37E-01 | 3.55E-01 | 7.59E+00 | 1.51E+01 | RBP1 |
| 218 | Diseases associated with visual transduction Homo sapiens R-HSA-2474795 | 1.37E-01 | 3.55E-01 | 7.59E+00 | 1.51E+01 | RBP1 |
| 219 | Prostanoid ligand receptors Homo sapiens R-HSA-391908 | 1.37E-01 | 3.55E-01 | 7.59E+00 | 1.51E+01 | PTGER2 |

| | | | | | | |
|---|---|---|---|---|---|---|
| 220 | AKT phosphorylates targets in the nucleus Homo sapiens R-HSA-198693 | 1.37E-01 | 3.55E-01 | 7.59E+00 | 1.51E+01 | FOXO3 |
| 221 | Interleukin-19, 20, 22, 24 Homo sapiens R-HSA-8854691 | 1.37E-01 | 3.55E-01 | 7.59E+00 | 1.51E+01 | IL20RA |
| 222 | Signaling by NOTCH Homo sapiens R-HSA-157118 | 1.38E-01 | 3.55E-01 | 2.07E+00 | 4.10E+00 | LFNG;TLE1;JAG1;CCND1 |
| 223 | Histidine, lysine, phenylalanine, tyrosine, proline and tryptophan catabolism Homo sapiens R-HSA-6788656 | 1.43E-01 | 3.67E-01 | 3.12E+00 | 6.06E+00 | CRYM;AASS |
| 224 | Post-translational protein modification Homo sapiens R-HSA-597592 | 1.44E-01 | 3.67E-01 | 1.44E+00 | 2.80E+00 | ADAMTS15;NEU3;GALNT14;GMDS;ADAMTS1;ST3GAL5;GCNT4;THBS1;FOLR1;CTSC;GALNT10;THSD4 |
| 225 | Signaling by Retinoic Acid Homo sapiens R-HSA-5362517 | 1.49E-01 | 3.67E-01 | 3.04E+00 | 5.79E+00 | ALDH1A1;DHRS3 |
| 226 | Transcriptional regulation of pluripotent stem cells Homo sapiens R-HSA-452723 | 1.49E-01 | 3.67E-01 | 3.04E+00 | 5.79E+00 | TSC22D1;PBX1 |
| 227 | Deactivation of the beta-catenin transactivating complex Homo sapiens R-HSA-3769402 | 1.49E-01 | 3.67E-01 | 3.04E+00 | 5.79E+00 | TLE1;SOX4 |
| 228 | Creatine metabolism Homo sapiens R-HSA-71288 | 1.51E-01 | 3.67E-01 | 6.74E+00 | 1.27E+01 | SLC6A8 |
| 229 | GP1b-IX-V activation signalling Homo sapiens R-HSA-430116 | 1.51E-01 | 3.67E-01 | 6.74E+00 | 1.27E+01 | FLNA |
| 230 | Abacavir transport and metabolism Homo sapiens R-HSA-2161522 | 1.51E-01 | 3.67E-01 | 6.74E+00 | 1.27E+01 | ABCB1 |
| 231 | eNOS activation Homo sapiens R-HSA-203615 | 1.51E-01 | 3.67E-01 | 6.74E+00 | 1.27E+01 | CAV1 |

| | | | | | | |
|---|---|---|---|---|---|---|
| 232 | POU5F1 (OCT4), SOX2, NANOG repress genes related to differentiation Homo sapiens R-HSA-2892245 | 1.51E-01 | 3.67E-01 | 6.74E+00 | 1.27E+01 | TSC22D1 |
| 233 | Signaling by FGFR3 fusions in cancer Homo sapiens R-HSA-8853334 | 1.51E-01 | 3.67E-01 | 6.74E+00 | 1.27E+01 | FGFR3 |
| 234 | Activation of the AP-1 family of transcription factors Homo sapiens R-HSA-450341 | 1.51E-01 | 3.67E-01 | 6.74E+00 | 1.27E+01 | FOS |
| 235 | IRAK1 recruits IKK complex Homo sapiens R-HSA-937039 | 1.51E-01 | 3.67E-01 | 6.74E+00 | 1.27E+01 | PELI2 |
| 236 | IRAK1 recruits IKK complex upon TLR7/8 or 9 stimulation Homo sapiens R-HSA-975144 | 1.51E-01 | 3.67E-01 | 6.74E+00 | 1.27E+01 | PELI2 |
| 237 | Phospholipase C-mediated cascade; FGFR3 Homo sapiens R-HSA-5654227 | 1.60E-01 | 3.72E-01 | 2.89E+00 | 5.30E+00 | FGF9;FGFR3 |
| 238 | Interleukin-1 signaling Homo sapiens R-HSA-446652 | 1.60E-01 | 3.72E-01 | 2.89E+00 | 5.30E+00 | IL1R1;PELI2 |
| 239 | Cell junction organization Homo sapiens R-HSA-446728 | 1.64E-01 | 3.72E-01 | 2.20E+00 | 3.97E+00 | CADM1;CLDN8;FLNA |
| 240 | Cell-Cell communication Homo sapiens R-HSA-1500931 | 1.65E-01 | 3.72E-01 | 1.92E+00 | 3.46E+00 | CADM1;CLDN8;SIRPA;FLNA |
| 241 | Transport of organic anions Homo sapiens R-HSA-879518 | 1.65E-01 | 3.72E-01 | 6.07E+00 | 1.09E+01 | SLCO2A1 |
| 242 | Dermatan sulfate biosynthesis Homo sapiens R-HSA-2022923 | 1.65E-01 | 3.72E-01 | 6.07E+00 | 1.09E+01 | BGN |
| 243 | Apoptotic cleavage of cell adhesion proteins Homo sapiens R-HSA-351906 | 1.65E-01 | 3.72E-01 | 6.07E+00 | 1.09E+01 | DSG2 |
| 244 | Response to metal ions Homo sapiens R-HSA-5660526 | 1.65E-01 | 3.72E-01 | 6.07E+00 | 1.09E+01 | MT1X |

| | | | | | | |
|---|---|---|---|---|---|---|
| 245 | Metallothioneins bind metals Homo sapiens R-HSA-5661231 | 1.65E-01 | 3.72E-01 | 6.07E+00 | 1.09E+01 | MT1X |
| 246 | Interleukin-6 signaling Homo sapiens R-HSA-1059683 | 1.65E-01 | 3.72E-01 | 6.07E+00 | 1.09E+01 | SOCS3 |
| 247 | Signaling by NOTCH3 Homo sapiens R-HSA-1980148 | 1.65E-01 | 3.72E-01 | 6.07E+00 | 1.09E+01 | JAG1 |
| 248 | Signaling by NOTCH4 Homo sapiens R-HSA-1980150 | 1.65E-01 | 3.72E-01 | 6.07E+00 | 1.09E+01 | JAG1 |
| 249 | FGFR1c ligand binding and activation Homo sapiens R-HSA-190373 | 1.65E-01 | 3.72E-01 | 6.07E+00 | 1.09E+01 | FGF9 |
| 250 | Signaling by activated point mutants of FGFR1 Homo sapiens R-HSA-1839122 | 1.65E-01 | 3.72E-01 | 6.07E+00 | 1.09E+01 | FGF9 |
| 251 | Signaling by FGFR4 in disease Homo sapiens R-HSA-5655291 | 1.65E-01 | 3.72E-01 | 6.07E+00 | 1.09E+01 | FGFR4 |
| 252 | CASP8 activity is inhibited Homo sapiens R-HSA-5218900 | 1.65E-01 | 3.72E-01 | 6.07E+00 | 1.09E+01 | TNFSF10 |
| 253 | Regulation by c-FLIP Homo sapiens R-HSA-3371378 | 1.65E-01 | 3.72E-01 | 6.07E+00 | 1.09E+01 | TNFSF10 |
| 254 | Dimerization of procaspase-8 Homo sapiens R-HSA-69416 | 1.65E-01 | 3.72E-01 | 6.07E+00 | 1.09E+01 | TNFSF10 |
| 255 | RMTs methylate histone arginines Homo sapiens R-HSA-3214858 | 1.66E-01 | 3.73E-01 | 2.83E+00 | 5.08E+00 | HIST3H2A;CCND1 |
| 256 | Platelet homeostasis Homo sapiens R-HSA-418346 | 1.68E-01 | 3.77E-01 | 2.17E+00 | 3.87E+00 | KCNMA1;KCNMB2;GNG11 |
| 257 | Phospholipase C-mediated cascade; FGFR4 Homo sapiens R-HSA-5654228 | 1.72E-01 | 3.82E-01 | 2.76E+00 | 4.87E+00 | FGF9;FGFR4 |
| 258 | Transmembrane transport of small molecules Homo sapiens R-HSA-382551 | 1.72E-01 | 3.82E-01 | 1.37E+00 | 2.41E+00 | GABRA2;ABCB1;TSC22D3;SLC12A1;SLC6A13;SLC3A1;GNG11;CLCN5;SLC22A17;SLCO2A1;ATP13A4;FXYD6;TRPM3 |

| | | | | | | |
|---|---|---|---|---|---|---|
| 259 | Phospholipase C-mediated cascade: FGFR1 Homo sapiens R-HSA-5654219 | 1.77E-01 | 3.90E-01 | 2.70E+00 | 4.67E+00 | KL;FGF9 |
| 260 | PECAM1 interactions Homo sapiens R-HSA-210990 | 1.79E-01 | 3.90E-01 | 5.52E+00 | 9.51E+00 | ITGB3 |
| 261 | Ethanol oxidation Homo sapiens R-HSA-71384 | 1.79E-01 | 3.90E-01 | 5.52E+00 | 9.51E+00 | ALDH1A1 |
| 262 | The NLRP3 inflammasome Homo sapiens R-HSA-844456 | 1.79E-01 | 3.90E-01 | 5.52E+00 | 9.51E+00 | TXNIP |
| 263 | Immune System Homo sapiens R-HSA-168256 | 1.79E-01 | 3.90E-01 | 1.22E+00 | 2.09E+00 | ITGB3;PELI2;IL20RA;DEFB1;KLHL13;FOXO3;MRC2;SOCS3;GRIN2A;C5;CTSO;FGF9;ERBB4;BLNK;HLA-DOA;CTSC;TRIM45;EGR1;KL;IL1R1;ZBTB16;FOS;PRLR;DNM1;RNF144B;RASA3;TXNIP;FGFR4;FGFR3;CD200 |
| 264 | Signal regulatory protein (SIRP) family interactions Homo sapiens R-HSA-391160 | 1.92E-01 | 4.07E-01 | 5.06E+00 | 8.35E+00 | SIRPA |
| 265 | Depolarization of the Presynaptic Terminal Triggers the Opening of Calcium Channels Homo sapiens R-HSA-112308 | 1.92E-01 | 4.07E-01 | 5.06E+00 | 8.35E+00 | CACNB2 |
| 266 | Retrograde neurotrophin signalling Homo sapiens R-HSA-177504 | 1.92E-01 | 4.07E-01 | 5.06E+00 | 8.35E+00 | DNM1 |
| 267 | GABA A receptor activation Homo sapiens R-HSA-977441 | 1.92E-01 | 4.07E-01 | 5.06E+00 | 8.35E+00 | GABRA2 |
| 268 | Synthesis of PE Homo sapiens R-HSA-1483213 | 1.92E-01 | 4.07E-01 | 5.06E+00 | 8.35E+00 | LPIN1 |
| 269 | FGFR2c ligand binding and activation Homo sapiens R-HSA-190375 | 1.92E-01 | 4.07E-01 | 5.06E+00 | 8.35E+00 | FGF9 |
| 270 | ERBB2 Activates PTK6 Signaling Homo sapiens R-HSA-8847993 | 1.92E-01 | 4.07E-01 | 5.06E+00 | 8.35E+00 | ERBB4 |

| | | | | | | |
|---|---|---|---|---|---|---|
| 271 | Neurotransmitter Receptor Binding And Downstream Transmission In The Postsynaptic Cell Homo sapiens R-HSA-112314 | 2.00E-01 | 4.23E-01 | 1.76E+00 | 2.83E+00 | GABRA2;GRIN2A;KCNJ16;GNG11 |
| 272 | EPH-Ephrin signaling Homo sapiens R-HSA-2682334 | 2.01E-01 | 4.23E-01 | 1.98E+00 | 3.18E+00 | EFNB2;EPHA7;DNM1 |
| 273 | CS/DS degradation Homo sapiens R-HSA-2024101 | 2.05E-01 | 4.23E-01 | 4.67E+00 | 7.40E+00 | BGN |
| 274 | TP53 regulates transcription of several additional cell death genes whose specific roles in p53-dependent apoptosis remain uncertain Homo sapiens R-HSA-6803205 | 2.05E-01 | 4.23E-01 | 4.67E+00 | 7.40E+00 | NDRG1 |
| 275 | Regulation of IFNG signaling Homo sapiens R-HSA-877312 | 2.05E-01 | 4.23E-01 | 4.67E+00 | 7.40E+00 | SOCS3 |
| 276 | Receptor-ligand binding initiates the second proteolytic cleavage of Notch receptor Homo sapiens R-HSA-156988 | 2.05E-01 | 4.23E-01 | 4.67E+00 | 7.40E+00 | JAG1 |
| 277 | Regulation of necroptotic cell death Homo sapiens R-HSA-5675482 | 2.05E-01 | 4.23E-01 | 4.67E+00 | 7.40E+00 | TNFSF10 |
| 278 | L1CAM interactions Homo sapiens R-HSA-373760 | 2.05E-01 | 4.23E-01 | 1.96E+00 | 3.11E+00 | ITGB3;LAMB1;DNM1 |
| 279 | Potassium Channels Homo sapiens R-HSA-1296071 | 2.18E-01 | 4.29E-01 | 1.90E+00 | 2.90E+00 | KCNMB2;KCNMA1;KCNJ16 |
| 280 | Negative regulation of TCF-dependent signaling by WNT ligand antagonists Homo sapiens R-HSA-3772470 | 2.18E-01 | 4.29E-01 | 4.33E+00 | 6.60E+00 | SFRP1 |

| | | | | | | |
|---|---|---|---|---|---|---|
| 281 | GRB2:SOS provides linkage to MAPK signaling for Integrins Homo sapiens R-HSA-354194 | 2.18E-01 | 4.29E-01 | 4.33E+00 | 6.60E+00 | ITGB3 |
| 282 | p130Cas linkage to MAPK signaling for integrins Homo sapiens R-HSA-372708 | 2.18E-01 | 4.29E-01 | 4.33E+00 | 6.60E+00 | ITGB3 |
| 283 | Disorders of transmembrane transporters Homo sapiens R-HSA-5619115 | 2.18E-01 | 4.29E-01 | 4.33E+00 | 6.60E+00 | AVPR1A |
| 284 | CREB phosphorylation through the activation of CaMKII Homo sapiens R-HSA-442729 | 2.18E-01 | 4.29E-01 | 4.33E+00 | 6.60E+00 | GRIN2A |
| 285 | Depolymerisation of the Nuclear Lamina Homo sapiens R-HSA-4419969 | 2.18E-01 | 4.29E-01 | 4.33E+00 | 6.60E+00 | LPIN1 |
| 286 | Eicosanoid ligand-binding receptors Homo sapiens R-HSA-391903 | 2.18E-01 | 4.29E-01 | 4.33E+00 | 6.60E+00 | PTGER2 |
| 287 | Synthesis of bile acids and bile salts via 27-hydroxycholesterol Homo sapiens R-HSA-193807 | 2.18E-01 | 4.29E-01 | 4.33E+00 | 6.60E+00 | CYP27A1 |
| 288 | Prolactin receptor signaling Homo sapiens R-HSA-1170546 | 2.18E-01 | 4.29E-01 | 4.33E+00 | 6.60E+00 | PRLR |
| 289 | Constitutive Signaling by NOTCH1 HD Domain Mutants Homo sapiens R-HSA-2691232 | 2.18E-01 | 4.29E-01 | 4.33E+00 | 6.60E+00 | JAG1 |
| 290 | Signaling by NOTCH1 HD Domain Mutants in Cancer Homo sapiens R-HSA-2691230 | 2.18E-01 | 4.29E-01 | 4.33E+00 | 6.60E+00 | JAG1 |
| 291 | ERBB2 Regulates Cell Motility Homo sapiens R-HSA-6785631 | 2.18E-01 | 4.29E-01 | 4.33E+00 | 6.60E+00 | ERBB4 |

| | | | | | | |
|---|---|---|---|---|---|---|
| 292 | Metabolism of vitamins and cofactors Homo sapiens R-HSA-196854 | 2.21E-01 | 4.33E-01 | 1.69E+00 | 2.55E+00 | RBP1;GPC3;AOX1;GPC4 |
| 293 | Stimuli-sensing channels Homo sapiens R-HSA-2672351 | 2.22E-01 | 4.34E-01 | 1.88E+00 | 2.83E+00 | CLCN5;TSC22D3;TRPM3 |
| 294 | GABA receptor activation Homo sapiens R-HSA-977443 | 2.25E-01 | 4.37E-01 | 2.29E+00 | 3.42E+00 | GABRA2;KCNJ16 |
| 295 | Ion transport by P-type ATPases Homo sapiens R-HSA-936837 | 2.25E-01 | 4.37E-01 | 2.29E+00 | 3.42E+00 | ATP13A4;FXYD6 |
| 296 | Platelet activation, signaling and aggregation Homo sapiens R-HSA-76002 | 2.30E-01 | 4.39E-01 | 1.48E+00 | 2.17E+00 | GNAQ;ITGB3;SERPINF2;FLNA;GNG11;THBS1 |
| 297 | Cell-extracellular matrix interactions Homo sapiens R-HSA-446353 | 2.31E-01 | 4.39E-01 | 4.05E+00 | 5.93E+00 | FLNA |
| 298 | PI3K events in ERBB2 signaling Homo sapiens R-HSA-1963642 | 2.31E-01 | 4.39E-01 | 4.05E+00 | 5.93E+00 | ERBB4 |
| 299 | GRB2 events in ERBB2 signaling Homo sapiens R-HSA-1963640 | 2.31E-01 | 4.39E-01 | 4.05E+00 | 5.93E+00 | ERBB4 |
| 300 | Regulated Necrosis Homo sapiens R-HSA-5218859 | 2.31E-01 | 4.39E-01 | 4.05E+00 | 5.93E+00 | TNFSF10 |
| 301 | RIPK1-mediated regulated necrosis Homo sapiens R-HSA-5213460 | 2.31E-01 | 4.39E-01 | 4.05E+00 | 5.93E+00 | TNFSF10 |
| 302 | Pre-NOTCH Expression and Processing Homo sapiens R-HSA-1912422 | 2.37E-01 | 4.49E-01 | 2.21E+00 | 3.18E+00 | LFNG;CCND1 |
| 303 | Ras activation uopn Ca2+ infux through NMDA receptor Homo sapiens R-HSA-442982 | 2.43E-01 | 4.52E-01 | 3.79E+00 | 5.36E+00 | GRIN2A |

| | | | | | | |
|---|---|---|---|---|---|---|
| 304 | Unblocking of NMDA receptor, glutamate binding and activation Homo sapiens R-HSA-438066 | 2.43E-01 | 4.52E-01 | 3.79E+00 | 5.36E+00 | GRIN2A |
| 305 | Inflammasomes Homo sapiens R-HSA-622312 | 2.43E-01 | 4.52E-01 | 3.79E+00 | 5.36E+00 | TXNIP |
| 306 | Formation of Senescence-Associated Heterochromatin Foci (SAHF) Homo sapiens R-HSA-2559584 | 2.43E-01 | 4.52E-01 | 3.79E+00 | 5.36E+00 | HIST1H1D |
| 307 | Activated point mutants of FGFR2 Homo sapiens R-HSA-2033519 | 2.43E-01 | 4.52E-01 | 3.79E+00 | 5.36E+00 | FGF9 |
| 308 | Ligand-dependent caspase activation Homo sapiens R-HSA-140534 | 2.43E-01 | 4.52E-01 | 3.79E+00 | 5.36E+00 | TNFSF10 |
| 309 | Degradation of the extracellular matrix Homo sapiens R-HSA-1474228 | 2.48E-01 | 4.60E-01 | 1.77E+00 | 2.47E+00 | ADAMTS1;SPP1;LAMB1 |
| 310 | Regulation of signaling by CBL Homo sapiens R-HSA-912631 | 2.55E-01 | 4.66E-01 | 3.57E+00 | 4.87E+00 | BLNK |
| 311 | Pre-NOTCH Processing in Golgi Homo sapiens R-HSA-1912420 | 2.55E-01 | 4.66E-01 | 3.57E+00 | 4.87E+00 | LFNG |
| 312 | Signaling by cytosolic FGFR1 fusion mutants Homo sapiens R-HSA-1839117 | 2.55E-01 | 4.66E-01 | 3.57E+00 | 4.87E+00 | CUX1 |
| 313 | SHC1 events in ERBB2 signaling Homo sapiens R-HSA-1250196 | 2.55E-01 | 4.66E-01 | 3.57E+00 | 4.87E+00 | ERBB4 |
| 314 | Tie2 Signaling Homo sapiens R-HSA-210993 | 2.55E-01 | 4.66E-01 | 3.57E+00 | 4.87E+00 | ANGPT2 |
| 315 | Cell-cell junction organization Homo sapiens R-HSA-421270 | 2.61E-01 | 4.75E-01 | 2.06E+00 | 2.77E+00 | CADM1;CLDN8 |
| 316 | GABA synthesis, release, reuptake and degradation Homo sapiens R-HSA-888590 | 2.68E-01 | 4.79E-01 | 3.37E+00 | 4.44E+00 | SLC6A13 |

| | | | | | | |
|---|---|---|---|---|---|---|
| 317 | Na+/Cl- dependent neurotransmitter transporters Homo sapiens R-HSA-442660 | 2.68E-01 | 4.79E-01 | 3.37E+00 | 4.44E+00 | SLC6A13 |
| 318 | Other semaphorin interactions Homo sapiens R-HSA-416700 | 2.68E-01 | 4.79E-01 | 3.37E+00 | 4.44E+00 | SEMA6D |
| 319 | Ephrin signaling Homo sapiens R-HSA-3928664 | 2.68E-01 | 4.79E-01 | 3.37E+00 | 4.44E+00 | EFNB2 |
| 320 | Prostacyclin signalling through prostacyclin receptor Homo sapiens R-HSA-392851 | 2.68E-01 | 4.79E-01 | 3.37E+00 | 4.44E+00 | GNG11 |
| 321 | Apoptosis Homo sapiens R-HSA-109581 | 2.74E-01 | 4.88E-01 | 1.53E+00 | 1.98E+00 | TNFSF10;HMGB2;HIST1H1D;DSG2 |
| 322 | G beta:gamma signalling through PLC beta Homo sapiens R-HSA-418217 | 2.80E-01 | 4.88E-01 | 3.19E+00 | 4.07E+00 | GNG11 |
| 323 | eNOS activation and regulation Homo sapiens R-HSA-203765 | 2.80E-01 | 4.88E-01 | 3.19E+00 | 4.07E+00 | CAV1 |
| 324 | Metabolism of nitric oxide Homo sapiens R-HSA-202131 | 2.80E-01 | 4.88E-01 | 3.19E+00 | 4.07E+00 | CAV1 |
| 325 | Hormone-sensitive lipase (HSL)-mediated triacylglycerol hydrolysis Homo sapiens R-HSA-163560 | 2.80E-01 | 4.88E-01 | 3.19E+00 | 4.07E+00 | CAV1 |
| 326 | Synthesis of PC Homo sapiens R-HSA-1483191 | 2.80E-01 | 4.88E-01 | 3.19E+00 | 4.07E+00 | LPIN1 |
| 327 | Glucuronidation Homo sapiens R-HSA-156588 | 2.80E-01 | 4.88E-01 | 3.19E+00 | 4.07E+00 | UGT2B7 |
| 328 | FGFR2 ligand binding and activation Homo sapiens R-HSA-190241 | 2.80E-01 | 4.88E-01 | 3.19E+00 | 4.07E+00 | FGF9 |
| 329 | Programmed Cell Death Homo sapiens R-HSA-5357801 | 2.85E-01 | 4.96E-01 | 1.50E+00 | 1.89E+00 | TNFSF10;HMGB2;HIST1H1D;DSG2 |
| 330 | Triglyceride Biosynthesis Homo sapiens R-HSA-75109 | 2.91E-01 | 4.98E-01 | 1.90E+00 | 2.34E+00 | SCD5;LPIN1 |
| 331 | RHO GTPases activate PAKs Homo sapiens R-HSA-5627123 | 2.91E-01 | 4.98E-01 | 3.03E+00 | 3.74E+00 | FLNA |

| | | | | | | |
|---|---|---|---|---|---|---|
| 332 | Regulation of Insulin-like Growth Factor (IGF) transport and uptake by Insulin-like Growth Factor Binding Proteins (IGFBPs) Homo sapiens R-HSA-381426 | 2.91E-01 | 4.98E-01 | 3.03E+00 | 3.74E+00 | IGFBP4 |
| 333 | Presynaptic function of Kainate receptors Homo sapiens R-HSA-500657 | 2.91E-01 | 4.98E-01 | 3.03E+00 | 3.74E+00 | GNG11 |
| 334 | Signal transduction by L1 Homo sapiens R-HSA-445144 | 2.91E-01 | 4.98E-01 | 3.03E+00 | 3.74E+00 | ITGB3 |
| 335 | NOTCH2 Activation and Transmission of Signal to the Nucleus Homo sapiens R-HSA-2979096 | 2.91E-01 | 4.98E-01 | 3.03E+00 | 3.74E+00 | JAG1 |
| 336 | Post-chaperonin tubulin folding pathway Homo sapiens R-HSA-389977 | 3.03E-01 | 5.09E-01 | 2.89E+00 | 3.45E+00 | TUBB2A |
| 337 | ADP signalling through P2Y purinoceptor 12 Homo sapiens R-HSA-392170 | 3.03E-01 | 5.09E-01 | 2.89E+00 | 3.45E+00 | GNG11 |
| 338 | Intrinsic Pathway of Fibrin Clot Formation Homo sapiens R-HSA-140837 | 3.03E-01 | 5.09E-01 | 2.89E+00 | 3.45E+00 | SERPINE2 |
| 339 | mTORC1-mediated signalling Homo sapiens R-HSA-166208 | 3.03E-01 | 5.09E-01 | 2.89E+00 | 3.45E+00 | RRAGD |
| 340 | COPII (Coat Protein 2) Mediated Vesicle Transport Homo sapiens R-HSA-204005 | 3.03E-01 | 5.09E-01 | 1.84E+00 | 2.20E+00 | FOLR1;CTSC |
| 341 | Interferon alpha/beta signaling Homo sapiens R-HSA-909733 | 3.03E-01 | 5.09E-01 | 1.84E+00 | 2.20E+00 | SOCS3;EGR1 |
| 342 | Metabolism of carbohydrates Homo sapiens R-HSA-71387 | 3.10E-01 | 5.20E-01 | 1.32E+00 | 1.55E+00 | ALDH1A1;BGN;GPC3;GPC4;HS6ST1;CHST3 |
| 343 | The canonical retinoid cycle in rods (twilight vision) Homo sapiens R-HSA-2453902 | 3.14E-01 | 5.22E-01 | 2.76E+00 | 3.19E+00 | RBP1 |

| | | | | | | |
|---|---|---|---|---|---|---|
| 344 | Laminin interactions Homo sapiens R-HSA-3000157 | 3.14E-01 | 5.22E-01 | 2.76E+00 | 3.19E+00 | LAMB1 |
| 345 | Downregulation of SMAD2/3:SMAD4 transcriptional activity Homo sapiens R-HSA-2173795 | 3.14E-01 | 5.22E-01 | 2.76E+00 | 3.19E+00 | SMAD3 |
| 346 | Toll Like Receptor 4 (TLR4) Cascade Homo sapiens R-HSA-166016 | 3.19E-01 | 5.27E-01 | 1.53E+00 | 1.75E+00 | PELI2;FOS;DNM1 |
| 347 | Endogenous sterols Homo sapiens R-HSA-211976 | 3.25E-01 | 5.35E-01 | 2.64E+00 | 2.96E+00 | CYP27A1 |
| 348 | DARPP-32 events Homo sapiens R-HSA-180024 | 3.25E-01 | 5.35E-01 | 2.64E+00 | 2.96E+00 | PDE4B |
| 349 | Signaling by NOTCH1 Homo sapiens R-HSA-1980143 | 3.27E-01 | 5.35E-01 | 1.73E+00 | 1.94E+00 | TLE1;JAG1 |
| 350 | Innate Immune System Homo sapiens R-HSA-168249 | 3.34E-01 | 5.35E-01 | 1.15E+00 | 1.27E+00 | KL;ITGB3;PELI2;DEFB1;FOS;FOXO3;DNM1;GRIN2A;C5;FGF9;RASA3;ERBB4;TXNIP;FGFR4;FGFR3 |
| 351 | Basigin interactions Homo sapiens R-HSA-210991 | 3.36E-01 | 5.35E-01 | 2.53E+00 | 2.75E+00 | CAV1 |
| 352 | Amino acid synthesis and interconversion (transamination) Homo sapiens R-HSA-70614 | 3.36E-01 | 5.35E-01 | 2.53E+00 | 2.75E+00 | PHGDH |
| 353 | Formation of tubulin folding intermediates by CCT/TriC Homo sapiens R-HSA-389960 | 3.36E-01 | 5.35E-01 | 2.53E+00 | 2.75E+00 | TUBB2A |
| 354 | Activation of G protein gated Potassium channels Homo sapiens R-HSA-1296041 | 3.36E-01 | 5.35E-01 | 2.53E+00 | 2.75E+00 | KCNJ16 |
| 355 | Inhibition of voltage gated Ca2+ channels via Gbeta/gamma subunits Homo sapiens R-HSA-997272 | 3.36E-01 | 5.35E-01 | 2.53E+00 | 2.75E+00 | KCNJ16 |
| 356 | G protein gated Potassium channels Homo sapiens R-HSA-1296059 | 3.36E-01 | 5.35E-01 | 2.53E+00 | 2.75E+00 | KCNJ16 |

| | | | | | | |
|---|---|---|---|---|---|---|
| 357 | TRP channels Homo sapiens R-HSA-3295583 | 3.36E-01 | 5.35E-01 | 2.53E+00 | 2.75E+00 | TRPM3 |
| 358 | Constitutive Signaling by AKT1 E17K in Cancer Homo sapiens R-HSA-5674400 | 3.36E-01 | 5.35E-01 | 2.53E+00 | 2.75E+00 | FOXO3 |
| 359 | HDMs demethylate histones Homo sapiens R-HSA-3214842 | 3.36E-01 | 5.35E-01 | 2.53E+00 | 2.75E+00 | ARID5B |
| 360 | Regulation of IFNA signaling Homo sapiens R-HSA-912694 | 3.36E-01 | 5.35E-01 | 2.53E+00 | 2.75E+00 | SOCS3 |
| 361 | Binding and Uptake of Ligands by Scavenger Receptors Homo sapiens R-HSA-2173782 | 3.39E-01 | 5.38E-01 | 1.69E+00 | 1.82E+00 | COL4A2;COL4A1 |
| 362 | Regulation of Complement cascade Homo sapiens R-HSA-977606 | 3.47E-01 | 5.41E-01 | 2.43E+00 | 2.57E+00 | C5 |
| 363 | Phase 2 - plateau phase Homo sapiens R-HSA-5576893 | 3.47E-01 | 5.41E-01 | 2.43E+00 | 2.57E+00 | CACNB2 |
| 364 | Recycling pathway of L1 Homo sapiens R-HSA-437239 | 3.47E-01 | 5.41E-01 | 2.43E+00 | 2.57E+00 | DNM1 |
| 365 | Ligand-gated ion channel transport Homo sapiens R-HSA-975298 | 3.47E-01 | 5.41E-01 | 2.43E+00 | 2.57E+00 | GABRA2 |
| 366 | SHC-mediated cascade:FGFR2 Homo sapiens R-HSA-5654699 | 3.47E-01 | 5.41E-01 | 2.43E+00 | 2.57E+00 | FGF9 |
| 367 | PTK6 Regulates RHO GTPases, RAS GTPase and MAP kinases Homo sapiens R-HSA-8849471 | 3.47E-01 | 5.41E-01 | 2.43E+00 | 2.57E+00 | RASA3 |
| 368 | Signaling by Wnt Homo sapiens R-HSA-195721 | 3.47E-01 | 5.41E-01 | 1.26E+00 | 1.33E+00 | SFRP1;TLE1;CTBP2;CAV1;GNG11;SOX4 |
| 369 | Integrin alphaIIb beta3 signaling Homo sapiens R-HSA-354192 | 3.58E-01 | 5.44E-01 | 2.33E+00 | 2.40E+00 | ITGB3 |

| | | | | | | |
|---|---|---|---|---|---|---|
| 370 | Prefoldin mediated transfer of substrate to CCT/TriC Homo sapiens R-HSA-389957 | 3.58E-01 | 5.44E-01 | 2.33E+00 | 2.40E+00 | TUBB2A |
| 371 | CREB phosphorylation through the activation of Ras Homo sapiens R-HSA-442742 | 3.58E-01 | 5.44E-01 | 2.33E+00 | 2.40E+00 | GRIN2A |
| 372 | Phase 1 - inactivation of fast Na+ channels Homo sapiens R-HSA-5576894 | 3.58E-01 | 5.44E-01 | 2.33E+00 | 2.40E+00 | CACNB2 |
| 373 | VEGFR2 mediated vascular permeability Homo sapiens R-HSA-5218920 | 3.58E-01 | 5.44E-01 | 2.33E+00 | 2.40E+00 | CAV1 |
| 374 | Peroxisomal lipid metabolism Homo sapiens R-HSA-390918 | 3.58E-01 | 5.44E-01 | 2.33E+00 | 2.40E+00 | SLC27A2 |
| 375 | RORA activates gene expression Homo sapiens R-HSA-1368082 | 3.58E-01 | 5.44E-01 | 2.33E+00 | 2.40E+00 | NRIP1 |
| 376 | Interleukin-6 family signaling Homo sapiens R-HSA-6783589 | 3.58E-01 | 5.44E-01 | 2.33E+00 | 2.40E+00 | SOCS3 |
| 377 | Caspase activation via extrinsic apoptotic signalig pathway Homo sapiens R-HSA-5357769 | 3.58E-01 | 5.44E-01 | 2.33E+00 | 2.40E+00 | TNFSF10 |
| 378 | G-protein activation Homo sapiens R-HSA-202040 | 3.68E-01 | 5.57E-01 | 2.25E+00 | 2.24E+00 | GNG11 |
| 379 | Energy dependent regulation of mTOR by LKB1-AMPK Homo sapiens R-HSA-380972 | 3.68E-01 | 5.57E-01 | 2.25E+00 | 2.24E+00 | RRAGD |
| 380 | Surfactant metabolism Homo sapiens R-HSA-5683826 | 3.78E-01 | 5.68E-01 | 2.17E+00 | 2.10E+00 | NAPSA |
| 381 | Opioid Signalling Homo sapiens R-HSA-111885 | 3.80E-01 | 5.68E-01 | 1.54E+00 | 1.49E+00 | PDE4B;GNG11 |
| 382 | Peptide ligand-binding receptors Homo sapiens R-HSA-375276 | 3.84E-01 | 5.68E-01 | 1.28E+00 | 1.23E+00 | C5;EDN1;EDN2;AVPR1A |

| | | | | | | |
|---|---|---|---|---|---|---|
| 383 | Degradation of beta-catenin by the destruction complex Homo sapiens R-HSA-195253 | 3.86E-01 | 5.68E-01 | 1.52E+00 | 1.44E+00 | TLE1;CTBP2 |
| 384 | Toll Like Receptor 10 (TLR10) Cascade Homo sapiens R-HSA-168142 | 3.86E-01 | 5.68E-01 | 1.52E+00 | 1.44E+00 | PELI2;FOS |
| 385 | Toll Like Receptor 5 (TLR5) Cascade Homo sapiens R-HSA-168176 | 3.86E-01 | 5.68E-01 | 1.52E+00 | 1.44E+00 | PELI2;FOS |
| 386 | MyD88 cascade initiated on plasma membrane Homo sapiens R-HSA-975871 | 3.86E-01 | 5.68E-01 | 1.52E+00 | 1.44E+00 | PELI2;FOS |
| 387 | Tight junction interactions Homo sapiens R-HSA-420029 | 3.89E-01 | 5.68E-01 | 2.09E+00 | 1.98E+00 | CLDN8 |
| 388 | Amine compound SLC transporters Homo sapiens R-HSA-425428 | 3.89E-01 | 5.68E-01 | 2.09E+00 | 1.98E+00 | SLC6A13 |
| 389 | Activation of Kainate Receptors upon glutamate binding Homo sapiens R-HSA-451326 | 3.89E-01 | 5.68E-01 | 2.09E+00 | 1.98E+00 | GNG11 |
| 390 | Glucagon-type ligand receptors Homo sapiens R-HSA-420092 | 3.89E-01 | 5.68E-01 | 2.09E+00 | 1.98E+00 | GNG11 |
| 391 | Activated NOTCH1 Transmits Signal to the Nucleus Homo sapiens R-HSA-2122948 | 3.89E-01 | 5.68E-01 | 2.09E+00 | 1.98E+00 | JAG1 |
| 392 | MAPK targets/ Nuclear events mediated by MAP kinases Homo sapiens R-HSA-450282 | 3.89E-01 | 5.68E-01 | 2.09E+00 | 1.98E+00 | FOS |
| 393 | TRAF6 mediated induction of NFkB and MAP kinases upon TLR7/8 or 9 activation Homo sapiens R-HSA-975138 | 3.92E-01 | 5.71E-01 | 1.50E+00 | 1.40E+00 | PELI2;FOS |
| 394 | TP53 Regulates Metabolic Genes Homo sapiens R-HSA-5628897 | 3.97E-01 | 5.71E-01 | 1.48E+00 | 1.37E+00 | SESN3;RRAGD |

| | | | | | | |
|---|---|---|---|---|---|---|
| 395 | Metabolism of polyamines Homo sapiens R-HSA-351202 | 3.97E-01 | 5.71E-01 | 1.48E+00 | 1.37E+00 | SLC6A8;ODC1 |
| 396 | Toll-Like Receptors Cascades Homo sapiens R-HSA-168898 | 3.98E-01 | 5.71E-01 | 1.33E+00 | 1.22E+00 | PELI2;FOS;DNM1 |
| 397 | Adherens junctions interactions Homo sapiens R-HSA-418990 | 3.98E-01 | 5.71E-01 | 2.02E+00 | 1.86E+00 | CADM1 |
| 398 | Amino acid transport across the plasma membrane Homo sapiens R-HSA-352230 | 3.98E-01 | 5.71E-01 | 2.02E+00 | 1.86E+00 | SLC3A1 |
| 399 | Inwardly rectifying K+ channels Homo sapiens R-HSA-1296065 | 3.98E-01 | 5.71E-01 | 2.02E+00 | 1.86E+00 | KCNJ16 |
| 400 | Disassembly of the destruction complex and recruitment of AXIN to the membrane Homo sapiens R-HSA-4641262 | 3.98E-01 | 5.71E-01 | 2.02E+00 | 1.86E+00 | CAV1 |
| 401 | MyD88 dependent cascade initiated on endosome Homo sapiens R-HSA-975155 | 4.03E-01 | 5.73E-01 | 1.46E+00 | 1.33E+00 | PELI2;FOS |
| 402 | Toll Like Receptor 7/8 (TLR7/8) Cascade Homo sapiens R-HSA-168181 | 4.03E-01 | 5.73E-01 | 1.46E+00 | 1.33E+00 | PELI2;FOS |
| 403 | TCF dependent signaling in response to WNT Homo sapiens R-HSA-201681 | 4.06E-01 | 5.73E-01 | 1.24E+00 | 1.12E+00 | SFRP1;TLE1;CAV1;SOX4 |
| 404 | G alpha (s) signalling events Homo sapiens R-HSA-418555 | 4.07E-01 | 5.73E-01 | 1.31E+00 | 1.18E+00 | PTGER2;PDE4B;GNG11 |
| 405 | Cooperation of Prefoldin and TriC/CCT in actin and tubulin folding Homo sapiens R-HSA-389958 | 4.08E-01 | 5.73E-01 | 1.96E+00 | 1.75E+00 | TUBB2A |
| 406 | Regulation of beta-cell development Homo sapiens R-HSA-186712 | 4.08E-01 | 5.73E-01 | 1.96E+00 | 1.75E+00 | HNF1B |
| 407 | Signaling by NOTCH2 Homo sapiens R-HSA-1980145 | 4.08E-01 | 5.73E-01 | 1.96E+00 | 1.75E+00 | JAG1 |

| | | | | | | |
|---|---|---|---|---|---|---|
| 408 | Oncogene Induced Senescence Homo sapiens R-HSA-2559585 | 4.08E-01 | 5.73E-01 | 1.96E+00 | 1.75E+00 | ID1 |
| 409 | Asparagine N-linked glycosylation Homo sapiens R-HSA-446203 | 4.12E-01 | 5.78E-01 | 1.19E+00 | 1.06E+00 | NEU3;GMDS;ST3GAL5;FOLR1;CTSC |
| 410 | Glucagon signaling in metabolic regulation Homo sapiens R-HSA-163359 | 4.18E-01 | 5.83E-01 | 1.89E+00 | 1.65E+00 | GNG11 |
| 411 | FGFR2 mutant receptor activation Homo sapiens R-HSA-1839126 | 4.18E-01 | 5.83E-01 | 1.89E+00 | 1.65E+00 | FGF9 |
| 412 | Toll Like Receptor 9 (TLR9) Cascade Homo sapiens R-HSA-168138 | 4.20E-01 | 5.84E-01 | 1.41E+00 | 1.22E+00 | PELI2;FOS |
| 413 | EPHA-mediated growth cone collapse Homo sapiens R-HSA-3928663 | 4.27E-01 | 5.89E-01 | 1.84E+00 | 1.56E+00 | EPHA7 |
| 414 | Activated PKN1 stimulates transcription of AR (androgen receptor) regulated genes KLK2 and KLK3 Homo sapiens R-HSA-5625886 | 4.27E-01 | 5.89E-01 | 1.84E+00 | 1.56E+00 | AR |
| 415 | Signaling by WNT in cancer Homo sapiens R-HSA-4791275 | 4.27E-01 | 5.89E-01 | 1.84E+00 | 1.56E+00 | CTBP2 |
| 416 | Negative regulation of FGFR2 signaling Homo sapiens R-HSA-5654727 | 4.27E-01 | 5.89E-01 | 1.84E+00 | 1.56E+00 | FGF9 |
| 417 | Class A/1 (Rhodopsin-like receptors) Homo sapiens R-HSA-373076 | 4.28E-01 | 5.89E-01 | 1.15E+00 | 9.74E-01 | C5;EDN1;HRH1;EDN2;PTGER2;AVPR1A |
| 418 | Post NMDA receptor activation events Homo sapiens R-HSA-438064 | 4.37E-01 | 5.99E-01 | 1.78E+00 | 1.48E+00 | GRIN2A |
| 419 | Phase 1 - Functionalization of compounds Homo sapiens R-HSA-211945 | 4.42E-01 | 5.99E-01 | 1.35E+00 | 1.10E+00 | CYP27A1;ALDH1A1 |

| | | | | | | |
|---|---|---|---|---|---|---|
| 420 | MyD88:Mal cascade initiated on plasma membrane Homo sapiens R-HSA-166058 | 4.42E-01 | 5.99E-01 | 1.35E+00 | 1.10E+00 | PELI2;FOS |
| 421 | Toll Like Receptor TLR1:TLR2 Cascade Homo sapiens R-HSA-168179 | 4.42E-01 | 5.99E-01 | 1.35E+00 | 1.10E+00 | PELI2;FOS |
| 422 | Toll Like Receptor TLR6:TLR2 Cascade Homo sapiens R-HSA-168188 | 4.42E-01 | 5.99E-01 | 1.35E+00 | 1.10E+00 | PELI2;FOS |
| 423 | Toll Like Receptor 2 (TLR2) Cascade Homo sapiens R-HSA-181438 | 4.42E-01 | 5.99E-01 | 1.35E+00 | 1.10E+00 | PELI2;FOS |
| 424 | Interferon gamma signaling Homo sapiens R-HSA-877300 | 4.48E-01 | 6.05E-01 | 1.33E+00 | 1.07E+00 | SOCS3;TRIM45 |
| 425 | Platelet Aggregation (Plug Formation) Homo sapiens R-HSA-76009 | 4.55E-01 | 6.12E-01 | 1.68E+00 | 1.33E+00 | ITGB3 |
| 426 | Apoptotic cleavage of cellular proteins Homo sapiens R-HSA-111465 | 4.55E-01 | 6.12E-01 | 1.68E+00 | 1.33E+00 | DSG2 |
| 427 | Chaperonin-mediated protein folding Homo sapiens R-HSA-390466 | 4.59E-01 | 6.15E-01 | 1.30E+00 | 1.02E+00 | TUBB2A;GNG11 |
| 428 | MAP2K and MAPK activation Homo sapiens R-HSA-5674135 | 4.64E-01 | 6.15E-01 | 1.64E+00 | 1.26E+00 | ITGB3 |
| 429 | Transport of vitamins, nucleosides, and related molecules Homo sapiens R-HSA-425397 | 4.64E-01 | 6.15E-01 | 1.64E+00 | 1.26E+00 | SLCO2A1 |
| 430 | Beta defensins Homo sapiens R-HSA-1461957 | 4.64E-01 | 6.15E-01 | 1.64E+00 | 1.26E+00 | DEFB1 |
| 431 | G1 Phase Homo sapiens R-HSA-69236 | 4.64E-01 | 6.15E-01 | 1.64E+00 | 1.26E+00 | CCND1 |
| 432 | Cyclin D associated events in G1 Homo sapiens R-HSA-69231 | 4.64E-01 | 6.15E-01 | 1.64E+00 | 1.26E+00 | CCND1 |
| 433 | GABA B receptor activation Homo sapiens R-HSA-977444 | 4.72E-01 | 6.21E-01 | 1.59E+00 | 1.20E+00 | KCNJ16 |

| | | | | | | |
|---|---|---|---|---|---|---|
| 434 | Activation of GABAB receptors Homo sapiens R-HSA-991365 | 4.72E-01 | 6.21E-01 | 1.59E+00 | 1.20E+00 | KCNJ16 |
| 435 | Activation of NMDA receptor upon glutamate binding and postsynaptic events Homo sapiens R-HSA-442755 | 4.72E-01 | 6.21E-01 | 1.59E+00 | 1.20E+00 | GRIN2A |
| 436 | mTOR signalling Homo sapiens R-HSA-165159 | 4.72E-01 | 6.21E-01 | 1.59E+00 | 1.20E+00 | RRAGD |
| 437 | Transport of inorganic cations/anions and amino acids/oligopeptides Homo sapiens R-HSA-425393 | 4.80E-01 | 6.29E-01 | 1.25E+00 | 9.17E-01 | SLC12A1;SLC3A1 |
| 438 | PKB-mediated events Homo sapiens R-HSA-109703 | 4.81E-01 | 6.29E-01 | 1.55E+00 | 1.14E+00 | RRAGD |
| 439 | Cellular Senescence Homo sapiens R-HSA-2559583 | 4.88E-01 | 6.35E-01 | 1.15E+00 | 8.27E-01 | ID1;HIST1H1D;FOS |
| 440 | Glycosphingolipid metabolism Homo sapiens R-HSA-1660662 | 4.90E-01 | 6.35E-01 | 1.52E+00 | 1.08E+00 | NEU3 |
| 441 | Pre-NOTCH Transcription and Translation Homo sapiens R-HSA-1912408 | 4.90E-01 | 6.35E-01 | 1.52E+00 | 1.08E+00 | CCND1 |
| 442 | Protein folding Homo sapiens R-HSA-391251 | 4.90E-01 | 6.35E-01 | 1.22E+00 | 8.72E-01 | TUBB2A;GNG11 |
| 443 | Glycerophospholipid biosynthesis Homo sapiens R-HSA-1483206 | 4.96E-01 | 6.35E-01 | 1.21E+00 | 8.51E-01 | GPCPD1;LPIN1 |
| 444 | Netrin-1 signaling Homo sapiens R-HSA-373752 | 4.98E-01 | 6.35E-01 | 1.48E+00 | 1.03E+00 | SLIT3 |
| 445 | EPHB-mediated forward signaling Homo sapiens R-HSA-3928662 | 4.98E-01 | 6.35E-01 | 1.48E+00 | 1.03E+00 | EFNB2 |
| 446 | Cooperation of PDCL (PhLP1) and TRiC/CCT in G-protein beta folding Homo sapiens R-HSA-6814122 | 4.98E-01 | 6.35E-01 | 1.48E+00 | 1.03E+00 | GNG11 |

| | | | | | | |
|---|---|---|---|---|---|---|
| 447 | Glucagon-like Peptide-1 (GLP1) regulates insulin secretion Homo sapiens R-HSA-381676 | 4.98E-01 | 6.35E-01 | 1.48E+00 | 1.03E+00 | GNG11 |
| 448 | Fatty Acyl-CoA Biosynthesis Homo sapiens R-HSA-75105 | 4.98E-01 | 6.35E-01 | 1.48E+00 | 1.03E+00 | SCD5 |
| 449 | BMAL1:CLOCK,NPAS2 activates circadian gene expression Homo sapiens R-HSA-1368108 | 4.98E-01 | 6.35E-01 | 1.48E+00 | 1.03E+00 | NRIP1 |
| 450 | Chromatin organization Homo sapiens R-HSA-4839726 | 5.02E-01 | 6.36E-01 | 1.09E+00 | 7.52E-01 | HIST3H2A;CCND1;JADE1;ARID5B |
| 451 | Chromatin modifying enzymes Homo sapiens R-HSA-3247509 | 5.02E-01 | 6.36E-01 | 1.09E+00 | 7.52E-01 | HIST3H2A;CCND1;JADE1;ARID5B |
| 452 | Vasopressin regulates renal water homeostasis via Aquaporins Homo sapiens R-HSA-432040 | 5.06E-01 | 6.36E-01 | 1.44E+00 | 9.83E-01 | GNG11 |
| 453 | Amine ligand-binding receptors Homo sapiens R-HSA-375280 | 5.06E-01 | 6.36E-01 | 1.44E+00 | 9.83E-01 | HRH1 |
| 454 | TP53 Regulates Transcription of Cell Death Genes Homo sapiens R-HSA-5633008 | 5.06E-01 | 6.36E-01 | 1.44E+00 | 9.83E-01 | NDRG1 |
| 455 | Iron uptake and transport Homo sapiens R-HSA-917937 | 5.06E-01 | 6.36E-01 | 1.44E+00 | 9.83E-01 | SLC22A17 |
| 456 | Signaling by FGFR2 in disease Homo sapiens R-HSA-5655253 | 5.06E-01 | 6.36E-01 | 1.44E+00 | 9.83E-01 | FGF9 |
| 457 | Phase 0 - rapid depolarisation Homo sapiens R-HSA-5576892 | 5.14E-01 | 6.43E-01 | 1.41E+00 | 9.38E-01 | CACNB2 |
| 458 | Intra-Golgi traffic Homo sapiens R-HSA-6811438 | 5.14E-01 | 6.43E-01 | 1.41E+00 | 9.38E-01 | CUX1 |

| | | | | | | |
|---|---|---|---|---|---|---|
| 459 | HATs acetylate histones Homo sapiens R-HSA-3214847 | 5.21E-01 | 6.49E-01 | 1.15E+00 | 7.52E-01 | HIST3H2A;JADE1 |
| 460 | ABC-family proteins mediated transport Homo sapiens R-HSA-382556 | 5.22E-01 | 6.49E-01 | 1.38E+00 | 8.95E-01 | ABCB1 |
| 461 | Signaling by ERBB2 Homo sapiens R-HSA-1227986 | 5.22E-01 | 6.49E-01 | 1.38E+00 | 8.95E-01 | ERBB4 |
| 462 | G alpha (z) signalling events Homo sapiens R-HSA-418597 | 5.30E-01 | 6.57E-01 | 1.35E+00 | 8.55E-01 | GNG11 |
| 463 | Antigen activates B Cell Receptor (BCR) leading to generation of second messengers Homo sapiens R-HSA-983695 | 5.37E-01 | 6.61E-01 | 1.32E+00 | 8.18E-01 | BLNK |
| 464 | Defensins Homo sapiens R-HSA-1461973 | 5.37E-01 | 6.61E-01 | 1.32E+00 | 8.18E-01 | DEFB1 |
| 465 | NOTCH1 Intracellular Domain Regulates Transcription Homo sapiens R-HSA-2122947 | 5.37E-01 | 6.61E-01 | 1.32E+00 | 8.18E-01 | TLE1 |
| 466 | Nucleotide-binding domain, leucine rich repeat containing receptor (NLR) signaling pathways Homo sapiens R-HSA-168643 | 5.37E-01 | 6.61E-01 | 1.32E+00 | 8.18E-01 | TXNIP |
| 467 | G beta:gamma signalling through PI3Kgamma Homo sapiens R-HSA-392451 | 5.45E-01 | 6.67E-01 | 1.29E+00 | 7.82E-01 | GNG11 |
| 468 | Death Receptor Signalling Homo sapiens R-HSA-73887 | 5.45E-01 | 6.67E-01 | 1.29E+00 | 7.82E-01 | TNFSF10 |
| 469 | Activated TLR4 signalling Homo sapiens R-HSA-166054 | 5.46E-01 | 6.67E-01 | 1.10E+00 | 6.67E-01 | PELI2;FOS |
| 470 | Amino acid and oligopeptide SLC transporters Homo sapiens R-HSA-425374 | 5.52E-01 | 6.71E-01 | 1.26E+00 | 7.49E-01 | SLC3A1 |

| | | | | | | |
|---|---|---|---|---|---|---|
| 471 | Phospholipase C-mediated cascade; FGFR2 Homo sapiens R-HSA-5654221 | 5.52E-01 | 6.71E-01 | 1.26E+00 | 7.49E-01 | FGF9 |
| 472 | Cross-presentation of soluble exogenous antigens (endosomes) Homo sapiens R-HSA-1236978 | 5.52E-01 | 6.71E-01 | 1.26E+00 | 7.49E-01 | MRC2 |
| 473 | Class I MHC mediated antigen processing & presentation Homo sapiens R-HSA-983169 | 5.54E-01 | 6.71E-01 | 1.01E+00 | 5.95E-01 | MRC2;RNF144B;SOCS3;ZBTB16;KLHL13 |
| 474 | Ubiquitin-dependent degradation of Cyclin D1 Homo sapiens R-HSA-69229 | 5.60E-01 | 6.71E-01 | 1.24E+00 | 7.18E-01 | CCND1 |
| 475 | Ubiquitin-dependent degradation of Cyclin D Homo sapiens R-HSA-75815 | 5.60E-01 | 6.71E-01 | 1.24E+00 | 7.18E-01 | CCND1 |
| 476 | Regulation of ornithine decarboxylase (ODC) Homo sapiens R-HSA-350562 | 5.60E-01 | 6.71E-01 | 1.24E+00 | 7.18E-01 | ODC1 |
| 477 | Neurotransmitter Release Cycle Homo sapiens R-HSA-112310 | 5.67E-01 | 6.71E-01 | 1.21E+00 | 6.88E-01 | SLC6A13 |
| 478 | G-protein beta:gamma signalling Homo sapiens R-HSA-397795 | 5.67E-01 | 6.71E-01 | 1.21E+00 | 6.88E-01 | GNG11 |
| 479 | Aquaporin-mediated transport Homo sapiens R-HSA-445717 | 5.67E-01 | 6.71E-01 | 1.21E+00 | 6.88E-01 | GNG11 |
| 480 | Ion homeostasis Homo sapiens R-HSA-5578775 | 5.67E-01 | 6.71E-01 | 1.21E+00 | 6.88E-01 | FXYD6 |
| 481 | Nuclear Envelope Breakdown Homo sapiens R-HSA-2980766 | 5.67E-01 | 6.71E-01 | 1.21E+00 | 6.88E-01 | LPIN1 |
| 482 | mRNA 3'-end processing Homo sapiens R-HSA-72187 | 5.67E-01 | 6.71E-01 | 1.21E+00 | 6.88E-01 | SRSF3 |

| | | | | | | |
|---|---|---|---|---|---|---|
| 483 | Post-Elongation Processing of Intron-Containing pre-mRNA Homo sapiens R-HSA-112296 | 5.67E-01 | 6.71E-01 | 1.21E+00 | 6.88E-01 | SRSF3 |
| 484 | Nuclear Receptor transcription pathway Homo sapiens R-HSA-383280 | 5.67E-01 | 6.71E-01 | 1.21E+00 | 6.88E-01 | AR |
| 485 | GPCR ligand binding Homo sapiens R-HSA-500792 | 5.92E-01 | 7.00E-01 | 9.62E-01 | 5.04E-01 | C5;EDN1;HRH1;EDN2;PTGER2;AVPR1A;GNG11 |
| 486 | Constitutive Signaling by NOTCH1 HD+PEST Domain Mutants Homo sapiens R-HSA-2894862 | 6.07E-01 | 7.10E-01 | 1.08E+00 | 5.39E-01 | JAG1 |
| 487 | Signaling by NOTCH1 in Cancer Homo sapiens R-HSA-2644603 | 6.07E-01 | 7.10E-01 | 1.08E+00 | 5.39E-01 | JAG1 |
| 488 | Signaling by NOTCH1 PEST Domain Mutants in Cancer Homo sapiens R-HSA-2644602 | 6.07E-01 | 7.10E-01 | 1.08E+00 | 5.39E-01 | JAG1 |
| 489 | Constitutive Signaling by NOTCH1 PEST Domain Mutants Homo sapiens R-HSA-2644606 | 6.07E-01 | 7.10E-01 | 1.08E+00 | 5.39E-01 | JAG1 |
| 490 | Signaling by NOTCH1 HD+PEST Domain Mutants in Cancer Homo sapiens R-HSA-2894858 | 6.07E-01 | 7.10E-01 | 1.08E+00 | 5.39E-01 | JAG1 |
| 491 | Antigen processing: Ubiquitination & Proteasome degradation Homo sapiens R-HSA-983168 | 6.13E-01 | 7.15E-01 | 9.45E-01 | 4.63E-01 | RNF144B;SOCS3;ZBTB16;KLHL13 |
| 492 | Formation of the beta-catenin:TCF transactivating complex Homo sapiens R-HSA-201722 | 6.14E-01 | 7.15E-01 | 1.06E+00 | 5.18E-01 | TLE1 |

| | | | | | | |
|---|---|---|---|---|---|---|
| 493 | DNA Damage/Telomere Stress Induced Senescence Homo sapiens R-HSA-2559586 | 6.20E-01 | 7.16E-01 | 1.04E+00 | 4.99E-01 | HIST1H1D |
| 494 | SCF(Skp2)-mediated degradation of p27/p21 Homo sapiens R-HSA-187577 | 6.20E-01 | 7.16E-01 | 1.04E+00 | 4.99E-01 | CCND1 |
| 495 | Interferon Signaling Homo sapiens R-HSA-913531 | 6.20E-01 | 7.16E-01 | 9.40E-01 | 4.49E-01 | EGR1;SOCS3;TRIM45 |
| 496 | Cleavage of Growing Transcript in the Termination Region Homo sapiens R-HSA-109688 | 6.26E-01 | 7.16E-01 | 1.03E+00 | 4.80E-01 | SRSF3 |
| 497 | RNA Polymerase II Transcription Termination Homo sapiens R-HSA-73856 | 6.26E-01 | 7.16E-01 | 1.03E+00 | 4.80E-01 | SRSF3 |
| 498 | Post-Elongation Processing of the Transcript Homo sapiens R-HSA-76044 | 6.26E-01 | 7.16E-01 | 1.03E+00 | 4.80E-01 | SRSF3 |
| 499 | RHO GTPases activate PKNs Homo sapiens R-HSA-5625740 | 6.26E-01 | 7.16E-01 | 1.03E+00 | 4.80E-01 | AR |
| 500 | HDACs deacetylate histones Homo sapiens R-HSA-3214815 | 6.26E-01 | 7.16E-01 | 1.03E+00 | 4.80E-01 | HIST3H2A |
| 501 | MAP kinase activation in TLR cascade Homo sapiens R-HSA-450294 | 6.26E-01 | 7.16E-01 | 1.03E+00 | 4.80E-01 | FOS |
| 502 | Biological oxidations Homo sapiens R-HSA-211859 | 6.30E-01 | 7.18E-01 | 9.26E-01 | 4.27E-01 | CYP27A1;ALDH1A1;UGT2B7 |
| 503 | ER to Golgi Anterograde Transport Homo sapiens R-HSA-199977 | 6.31E-01 | 7.18E-01 | 9.38E-01 | 4.32E-01 | FOLR1;CTSC |
| 504 | Ca2+ pathway Homo sapiens R-HSA-4086398 | 6.32E-01 | 7.18E-01 | 1.01E+00 | 4.62E-01 | GNG11 |

| | | | | | | |
|---|---|---|---|---|---|---|
| 505 | TP53 Regulates Transcription of DNA Repair Genes Homo sapiens R-HSA-6796648 | 6.32E-01 | 7.18E-01 | 1.01E+00 | 4.62E-01 | FOS |
| 506 | SLC-mediated transmembrane transport Homo sapiens R-HSA-425407 | 6.36E-01 | 7.20E-01 | 9.16E-01 | 4.14E-01 | SLCO2A1;SLC12A1;SLC6A13;SLC3A1 |
| 507 | Metabolism of amino acids and derivatives Homo sapiens R-HSA-71291 | 6.37E-01 | 7.20E-01 | 9.16E-01 | 4.12E-01 | SLC6A8;ODC1;PHGDH;CRYM;AASS |
| 508 | Cytochrome P450 - arranged by substrate type Homo sapiens R-HSA-211897 | 6.38E-01 | 7.20E-01 | 9.92E-01 | 4.45E-01 | CYP27A1 |
| 509 | Circadian Clock Homo sapiens R-HSA-400253 | 6.44E-01 | 7.25E-01 | 9.76E-01 | 4.29E-01 | NRIP1 |
| 510 | Cardiac conduction Homo sapiens R-HSA-5576891 | 6.47E-01 | 7.27E-01 | 9.10E-01 | 3.96E-01 | CACNB2;FXYD6 |
| 511 | Semaphorin interactions Homo sapiens R-HSA-373755 | 6.67E-01 | 7.45E-01 | 9.17E-01 | 3.71E-01 | SEMA6D |
| 512 | Macroautophagy Homo sapiens R-HSA-1632852 | 6.67E-01 | 7.45E-01 | 9.17E-01 | 3.71E-01 | RRAGD |
| 513 | Regulation of RAS by GAPs Homo sapiens R-HSA-5658442 | 6.67E-01 | 7.45E-01 | 9.17E-01 | 3.71E-01 | RASA3 |
| 514 | Transport of Mature mRNA derived from an Intron-Containing Transcript Homo sapiens R-HSA-159236 | 6.72E-01 | 7.48E-01 | 9.03E-01 | 3.59E-01 | SRSF3 |
| 515 | Cyclin E associated events during G1/S transition Homo sapiens R-HSA-69202 | 6.72E-01 | 7.48E-01 | 9.03E-01 | 3.59E-01 | CCND1 |
| 516 | Metabolism of proteins Homo sapiens R-HSA-392499 | 6.76E-01 | 7.51E-01 | 9.11E-01 | 3.57E-01 | NAPSA;GALNT14;IGFBP4;GNG11;THBS1;GALNT10;THSD4;ADAMTS15;NEU3;TUBB2A;GMDS;ADAMTS1;ST3GAL5;GCNT4;FOLR1;CTSC |
| 517 | Cyclin A:Cdk2-associated events at S phase entry Homo sapiens R-HSA-69656 | 6.78E-01 | 7.51E-01 | 8.90E-01 | 3.46E-01 | CCND1 |

| | | | | | | |
|---|---|---|---|---|---|---|
| 518 | Fatty acid, triacylglycerol, and ketone body metabolism Homo sapiens R-HSA-535734 | 6.88E-01 | 7.60E-01 | 8.47E-01 | 3.17E-01 | SCD5;LPIN1;MED13L |
| 519 | Lipid digestion, mobilization, and transport Homo sapiens R-HSA-73923 | 6.88E-01 | 7.60E-01 | 8.64E-01 | 3.23E-01 | CAV1 |
| 520 | Vesicle-mediated transport Homo sapiens R-HSA-5653656 | 6.91E-01 | 7.61E-01 | 8.71E-01 | 3.22E-01 | DAB2;CUX1;COL4A2;COL4A1;FOLR1;CTSC;DNM1 |
| 521 | TRAF6 Mediated Induction of proinflammatory cytokines Homo sapiens R-HSA-168180 | 6.93E-01 | 7.62E-01 | 8.52E-01 | 3.12E-01 | FOS |
| 522 | Sphingolipid metabolism Homo sapiens R-HSA-428157 | 7.03E-01 | 7.70E-01 | 8.29E-01 | 2.92E-01 | NEU3 |
| 523 | Transport of Mature Transcript to Cytoplasm Homo sapiens R-HSA-72202 | 7.03E-01 | 7.70E-01 | 8.29E-01 | 2.92E-01 | SRSF3 |
| 524 | G alpha (12/13) signalling events Homo sapiens R-HSA-416482 | 7.08E-01 | 7.74E-01 | 8.18E-01 | 2.82E-01 | GNG11 |
| 525 | Senescence-Associated Secretory Phenotype (SASP) Homo sapiens R-HSA-2559582 | 7.17E-01 | 7.82E-01 | 7.96E-01 | 2.64E-01 | FOS |
| 526 | RNA polymerase II transcribes snRNA genes Homo sapiens R-HSA-6807505 | 7.17E-01 | 7.82E-01 | 7.96E-01 | 2.64E-01 | NABP1 |
| 527 | COPI-mediated anterograde transport Homo sapiens R-HSA-6807878 | 7.22E-01 | 7.85E-01 | 7.86E-01 | 2.56E-01 | FOLR1 |
| 528 | Transcriptional regulation of white adipocyte differentiation Homo sapiens R-HSA-381340 | 7.27E-01 | 7.89E-01 | 7.75E-01 | 2.48E-01 | MED13L |
| 529 | Complement cascade Homo sapiens R-HSA-166658 | 7.31E-01 | 7.92E-01 | 7.66E-01 | 2.40E-01 | C5 |

| | | | | | | |
|---|---|---|---|---|---|---|
| 530 | Phospholipid metabolism Homo sapiens R-HSA-1483257 | 7.40E-01 | 7.98E-01 | 7.60E-01 | 2.29E-01 | GPCPD1;LPIN1 |
| 531 | Antigen processing-Cross presentation Homo sapiens R-HSA-1236975 | 7.40E-01 | 7.98E-01 | 7.47E-01 | 2.25E-01 | MRC2 |
| 532 | Transport to the Golgi and subsequent modification Homo sapiens R-HSA-948021 | 7.43E-01 | 7.99E-01 | 7.55E-01 | 2.25E-01 | FOLR1;CTSC |
| 533 | Metabolism Homo sapiens R-HSA-1430728 | 7.43E-01 | 7.99E-01 | 8.92E-01 | 2.65E-01 | ABCB1;ODC1;MT1X;HS6ST1;NEU3;GPC3;AOX1;PHGDH;GPC4;CRYM;AASS;CAV1;SCD5;BGN;GPCPD1;GNG11;MED13L;CYP27A1;CACNB2;MARCKS;SLC6A8;GNAQ;RBP1;ALDH1A1;LPIN1;SLC27A2;UGT2B7;CHST3 |
| 534 | Class B/2 (Secretin family receptors) Homo sapiens R-HSA-373080 | 7.48E-01 | 8.03E-01 | 7.29E-01 | 2.11E-01 | GNG11 |
| 535 | Activation of anterior HOX genes in hindbrain development during early embryogenesis Homo sapiens R-HSA-5617472 | 7.68E-01 | 8.21E-01 | 6.87E-01 | 1.81E-01 | PBX1 |
| 536 | Activation of HOX genes during differentiation Homo sapiens R-HSA-5619507 | 7.68E-01 | 8.21E-01 | 6.87E-01 | 1.81E-01 | PBX1 |
| 537 | Oxidative Stress Induced Senescence Homo sapiens R-HSA-2559580 | 7.76E-01 | 8.28E-01 | 6.72E-01 | 1.71E-01 | FOS |
| 538 | MAPK6/MAPK4 signaling Homo sapiens R-HSA-5687128 | 7.79E-01 | 8.30E-01 | 6.64E-01 | 1.66E-01 | FOXO3 |
| 539 | MyD88-independent TLR3/TLR4 cascade Homo sapiens R-HSA-166166 | 7.97E-01 | 8.44E-01 | 6.29E-01 | 1.43E-01 | FOS |
| 540 | Toll Like Receptor 3 (TLR3) Cascade Homo sapiens R-HSA-168164 | 7.97E-01 | 8.44E-01 | 6.29E-01 | 1.43E-01 | FOS |

| | | | | | | |
|---|---|---|---|---|---|---|
| 541 | TRIF-mediated TLR3/TLR4 signaling Homo sapiens R-HSA-937061 | 7.97E-01 | 8.44E-01 | 6.29E-01 | 1.43E-01 | FOS |
| 542 | Metabolism of water-soluble vitamins and cofactors Homo sapiens R-HSA-196849 | 8.00E-01 | 8.46E-01 | 6.23E-01 | 1.39E-01 | AOX1 |
| 543 | Phase II conjugation Homo sapiens R-HSA-156580 | 8.06E-01 | 8.51E-01 | 6.10E-01 | 1.31E-01 | UGT2B7 |
| 544 | Transport of glucose and other sugars, bile salts and organic acids, metal ions and amine compounds Homo sapiens R-HSA-425366 | 8.10E-01 | 8.53E-01 | 6.04E-01 | 1.28E-01 | SLC6A13 |
| 545 | Membrane Trafficking Homo sapiens R-HSA-199991 | 8.16E-01 | 8.58E-01 | 7.25E-01 | 1.48E-01 | DAB2;CUX1;FOLR1;CTSC;DNM1 |
| 546 | Transcriptional Regulation by TP53 Homo sapiens R-HSA-3700989 | 8.20E-01 | 8.61E-01 | 7.00E-01 | 1.39E-01 | SESN3;RRAGD;FOS;NDRG1 |
| 547 | Mitotic Prophase Homo sapiens R-HSA-68875 | 8.28E-01 | 8.67E-01 | 5.70E-01 | 1.08E-01 | LPIN1 |
| 548 | Muscle contraction Homo sapiens R-HSA-397014 | 8.31E-01 | 8.68E-01 | 6.22E-01 | 1.15E-01 | CACNB2;FXYD6 |
| 549 | G1/S Transition Homo sapiens R-HSA-69206 | 8.41E-01 | 8.77E-01 | 5.44E-01 | 9.41E-02 | CCND1 |
| 550 | Metabolism of lipids and lipoproteins Homo sapiens R-HSA-556833 | 8.44E-01 | 8.77E-01 | 7.37E-01 | 1.25E-01 | CYP27A1;NEU3;CAV1;SCD5;GPCPD1;LPIN1;SLC27A2;MED13L |
| 551 | PPARA activates gene expression Homo sapiens R-HSA-1989781 | 8.44E-01 | 8.77E-01 | 5.39E-01 | 9.16E-02 | MED13L |
| 552 | Signaling by GPCR Homo sapiens R-HSA-372790 | 8.48E-01 | 8.81E-01 | 7.96E-01 | 1.31E-01 | KL;EDN1;EDN2;PTGER2;ITGB3;AVPR1A;GNG11;GRIN2A;C5;HRH1;FGF9;RASA3;ERBB4;GNAQ;PDE4B;FGFR4;FGFR3 |
| 553 | Cellular responses to stress Homo sapiens R-HSA-2262752 | 8.50E-01 | 8.81E-01 | 6.63E-01 | 1.08E-01 | RRAGD;ID1;HIST1H1D;FOS |

| | | | | | | |
|---|---|---|---|---|---|---|
| 554 | Regulation of lipid metabolism by Peroxisome proliferator-activated receptor alpha (PPARalpha) Homo sapiens R-HSA-400206 | 8.51E-01 | 8.81E-01 | 5.25E-01 | 8.45E-02 | MED13L |
| 555 | Rho GTPase cycle Homo sapiens R-HSA-194840 | 8.65E-01 | 8.93E-01 | 4.99E-01 | 7.21E-02 | ARHGAP8 |
| 556 | RNA Polymerase II Transcription Homo sapiens R-HSA-73857 | 8.70E-01 | 8.95E-01 | 4.91E-01 | 6.85E-02 | SRSF3 |
| 557 | S Phase Homo sapiens R-HSA-69242 | 8.70E-01 | 8.95E-01 | 4.91E-01 | 6.85E-02 | CCND1 |
| 558 | mRNA Splicing - Major Pathway Homo sapiens R-HSA-72163 | 8.90E-01 | 9.13E-01 | 4.53E-01 | 5.31E-02 | SRSF3 |
| 559 | Mitotic G1-G1/S phases Homo sapiens R-HSA-453279 | 8.93E-01 | 9.15E-01 | 4.47E-01 | 5.05E-02 | CCND1 |
| 560 | G alpha (i) signalling events Homo sapiens R-HSA-418594 | 9.04E-01 | 9.24E-01 | 5.06E-01 | 5.10E-02 | C5;GNG11 |
| 561 | mRNA Splicing Homo sapiens R-HSA-72172 | 9.06E-01 | 9.24E-01 | 4.22E-01 | 4.15E-02 | SRSF3 |
| 562 | Beta-catenin independent WNT signaling Homo sapiens R-HSA-3858494 | 9.06E-01 | 9.24E-01 | 4.22E-01 | 4.15E-02 | GNG11 |
| 563 | Generic Transcription Pathway Homo sapiens R-HSA-212436 | 9.15E-01 | 9.31E-01 | 6.69E-01 | 5.94E-02 | AR;SMAD3;SESN3;RRAGD;FOS;JUNB;NDRG1;ZNF124;SMAD7 |
| 564 | RHO GTPase Effectors Homo sapiens R-HSA-195258 | 9.21E-01 | 9.36E-01 | 4.75E-01 | 3.89E-02 | AR;FLNA |
| 565 | Immunoregulatory interactions between a Lymphoid and a non-Lymphoid cell Homo sapiens R-HSA-198933 | 9.24E-01 | 9.37E-01 | 3.86E-01 | 3.04E-02 | CD200 |
| 566 | Signaling by Rho GTPases Homo sapiens R-HSA-194315 | 9.40E-01 | 9.51E-01 | 4.94E-01 | 3.08E-02 | AR;ARHGAP8;FLNA |
| 567 | Intra-Golgi and retrograde Golgi-to-ER traffic Homo sapiens R-HSA-6811442 | 9.47E-01 | 9.57E-01 | 3.38E-01 | 1.83E-02 | CUX1 |

| | | | | | | |
|---|---|---|---|---|---|---|
| 568 | Processing of Capped Intron-Containing Pre-mRNA Homo sapiens R-HSA-72203 | 9.58E-01 | 9.67E-01 | 3.13E-01 | 1.33E-02 | SRSF3 |
| 569 | GPCR downstream signaling Homo sapiens R-HSA-388396 | 9.81E-01 | 9.88E-01 | 5.47E-01 | 1.06E-02 | C5;EDN1;HRH1;EDN2;PTGER2;GNAQ;PDE4B;AVPR1A;GNG11 |
| 570 | M Phase Homo sapiens R-HSA-68886 | 9.88E-01 | 9.93E-01 | 2.24E-01 | 2.72E-03 | LPIN1 |
| 571 | Cell Cycle, Mitotic Homo sapiens R-HSA-69278 | 9.96E-01 | 9.99E-01 | 2.59E-01 | 1.08E-03 | CCND1;LPIN1 |
| 572 | Cell Cycle Homo sapiens R-HSA-1640170 | 9.99E-01 | 1.00E+00 | 2.10E-01 | 1.84E-04 | CCND1;LPIN1 |
| 573 | Gene Expression Homo sapiens R-HSA-74160 | 1.00E+00 | 1.00E+00 | 3.90E-01 | 5.25E-05 | AR;SMAD3;SESN3;RRAGD;SRSF3;NABP1;FOS;JUNB;NDRG1;ZNF124;SMAD7 |

**Pathways enriched by genes suppressed with SGLT2 inhibitor in thick ascending limb cells**

| | Term | P value | Adjusted P value | Odds Ratio | Combined Score | Genes |
|---|---|---|---|---|---|---|
| 1 | Platelet degranulation Homo sapiens R-HSA-114608 | 9.55E-06 | 4.65E-03 | 3.90E+00 | 4.51E+01 | SRGN;TGFB2;SPARC;SERPINA1;VWF;ACTN1;RARRES2;PCDH7;PDGFB;SELP;TMSB4X;PECAM1;TIMP3;TIMP1;A2M;CALM1;GAS6 |
| 2 | Smooth Muscle Contraction Homo sapiens R-HSA-445355 | 1.71E-05 | 4.65E-03 | 7.53E+00 | 8.27E+01 | ACTA2;ANXA1;ANXA2;TPM4;CALD1;TPM2;TPM1;CALM1;MYL12A |
| 3 | Elastic fibre formation Homo sapiens R-HSA-1566948 | 1.72E-05 | 4.65E-03 | 6.48E+00 | 7.11E+01 | FBN2;ITGB1;TGFB2;MFAP2;LOXL4;FURIN;ITGAV;FBLN2;LTBP1;LOXL2 |
| 4 | Response to elevated platelet cytosolic Ca2+ Homo sapiens R-HSA-76005 | 1.80E-05 | 4.65E-03 | 3.69E+00 | 4.03E+01 | SRGN;TGFB2;SPARC;SERPINA1;VWF;ACTN1;RARRES2;PCDH7;PDGFB;SELP;TMSB4X;PECAM1;TIMP3;TIMP1;A2M;CALM1;GAS6 |
| 5 | Muscle contraction Homo sapiens R-HSA-397014 | 2.29E-05 | 4.75E-03 | 2.83E+00 | 3.02E+01 | ANXA1;ANXA2;TPM4;KCNE4;NPR1;TMOD3;TPM2;TPM1;ATP2A3;KCNK15;ATP1B3;ITPR3;CACNA1C;MYL12A;ACTA2;FXYD4;CALD1;MYH8;TNNI1;ORAI1;VIM;CALM1;FGF12;KCNK3 |
| 6 | Hemostasis Homo sapiens R-HSA-109582 | 4.87E-05 | 8.40E-03 | 1.94E+00 | 1.93E+01 | ITGB1;SERPINA1;SPARC;PDGFB;ATP2A3;PIK3R3;ITPR3;ARRB1;IL2RG;SERPINA5;RAP1B;GRB14;PRKAR2B;TMSB4X;KCNMB4;TIMP3;KIF1C;ITGAV;TIMP1;A2M;JAM2;SRGN;TGFB2;CXADR;ANGPT2;ANXA2;VWF;PCDH7;ACTN1;RARRES2;CAV1;F2R;PDE2A;ATP1B3;SERPINB6;SELP;EHD2;SLC7A6;MAFG;KIF26B;IL3RA;CAPZA2;PECAM1;ORAI1;GAS6;CALM1;DOCK1;RAPGEF3 |
| 7 | Axon guidance Homo sapiens R-HSA-422475 | 7.42E-05 | 1.10E-02 | 1.95E+00 | 1.86E+01 | ITGB1;PLXND1;ARPC1B;RASGRF2;ARPC1A;CLTC;PDGFB;NTN4;CRMP1;ARRB1;CACNA1C;SHB;IL2RG;FGF2;EGFR;MYL12A;RASGRP3;RAP1B;EFNB2;DPYSL2;UBC;KDR;ITGAV;SLIT3;SRGAP2;PAK2;PLXNA4;SPTBN1;KSR1;VWF;SEMA4D;DUSP1;RANBP9;ANK2;DUSP6;PSMB9;DLG4;COL4A4;IL3RA;FGF18;MYH9;COL9A2;CALM1;DOCK1;EPHA2 |
| 8 | Extracellular matrix organization Homo sapiens R-HSA-1474244 | 9.69E-05 | 1.25E-02 | 2.31E+00 | 2.14E+01 | FBN2;ITGB1;SPARC;COL12A1;PDGFB;HTRA1;NTN4;LOXL4;PLOD2;FURIN;FGF2;FBLN2;LTBP1;LOXL2;KDR;ITGAV;TIMP1;A2M;JAM2;TGFB2;LAMB2;ACTN1;HSPG2;CAPN14;P4HA1;COL4A4;MFAP2;PECAM1;COL9A2 |

| | | | | | | |
|---|---|---|---|---|---|---|
| 9 | SLC-mediated transmembrane transport Homo sapiens R-HSA-425407 | 2.20E-04 | 2.53E-02 | 2.27E+00 | 1.91E+01 | SLC47A2;SLC2A1;SLC5A3;SLC2A6;HK2;SLC38A2;SLC14A1;SLC15A1;SLC38A1;SLC14A2;SLC35A3;NUP210;SLC6A12;RUNX1;SLC2A9;SLC6A6;SLC5A8;SLC7A6;SLC6A9;SLCO4A1;SLCO4C1;SLCO1A2;SLC29A4;SLC26A7;CALM1;SLC29A1;SLC26A6 |
| 10 | Transmembrane transport of small molecules Homo sapiens R-HSA-382551 | 2.79E-04 | 2.89E-02 | 1.79E+00 | 1.47E+01 | SLC47A2;TTYH3;ABCB1;SLC2A1;ATP2A3;CLCNKA;SLC5A3;SLC2A6;HK2;AQP1;PRKAR2B;NIPAL3;UBC;BSND;ASIC2;SLC38A2;ATP9A;SLC14A1;SLC15A1;SLC38A1;SLC14A2;SLC35A3;NUP210;ABCA4;SLC6A12;ATP1B3;MCOLN1;RUNX1;SLC2A9;CLCN7;SLC6A6;SLC5A8;SLC7A6;CLCN4;SLC6A9;FXYD4;OSTM1;SLCO4A1;TRPV4;SLCO4C1;SLCO1A2;LCN2;SLC29A4;SLC26A7;SLC29A1;CALM1;SGK1;SLC26A6 |
| 11 | Developmental Biology Homo sapiens R-HSA-1266738 | 4.12E-04 | 3.88E-02 | 1.66E+00 | 1.29E+01 | ITGB1;PLXND1;RASGRF2;ARPC1B;ARPC1A;CLTC;ARRB1;SHB;FGF2;CDH4;DPYSL2;BOC;KDR;ITGAV;HIST1H2AC;KSR1;VWF;DUSP1;ANK2;DUSP6;COL4A4;IL3RA;MYH9;PPARG;ANGPTL4;DOCK1;EPHA2;CEBPD;PDGFB;NTN4;CRMP1;FURIN;CACNA1C;IL2RG;EGFR;MYL12A;RASGRP3;RAP1B;EFNB2;UBC;SLIT3;PCK1;SRGAP2;PAK2;PLXNA4;SPTBN1;SEMA4D;STAT3;LGI2;RANBP9;MAFA;PSMB9;DLG4;FGF18;COL9A2;CALM1;FOXA3;EZH2;FOXA2 |
| 12 | Platelet activation, signaling and aggregation Homo sapiens R-HSA-76002 | 4.94E-04 | 4.26E-02 | 2.22E+00 | 1.69E+01 | SPARC;SERPINA1;PDGFB;PIK3R3;ITPR3;ARRB1;IL2RG;RAP1B;TMSB4X;TIMP3;TIMP1;A2M;SRGN;TGFB2;VWF;ACTN1;RARRES2;PCDH7;F2R;SELP;IL3RA;PECAM1;CALM1;GAS6;RAPGEF3 |
| 13 | Non-integrin membrane-ECM interactions Homo sapiens R-HSA-3000171 | 7.33E-04 | 5.84E-02 | 4.72E+00 | 3.41E+01 | ITGB1;LAMB2;ACTN1;PDGFB;NTN4;ITGAV;FGF2;HSPG2 |
| 14 | Calcitonin-like ligand receptors Homo sapiens R-HSA-419812 | 8.64E-04 | 6.39E-02 | 1.33E+01 | 9.40E+01 | CALCA;RAMP2;CALCRL;CALCB |
| 15 | Striated Muscle Contraction Homo sapiens R-HSA-390522 | 9.66E-04 | 6.67E-02 | 5.20E+00 | 3.61E+01 | TPM4;TPM2;TMOD3;TPM1;TNNI1;MYH8;VIM |
| 16 | eNOS activation and regulation Homo sapiens R-HSA-203765 | 2.10E-03 | 1.28E-01 | 6.67E+00 | 4.11E+01 | DHFR;SPR;CAV1;NOSTRIN;CALM1 |

| | | | | | | |
|---|---|---|---|---|---|---|
| 17 | Metabolism of nitric oxide Homo sapiens R-HSA-202131 | 2.10E-03 | 1.28E-01 | 6.67E+00 | 4.11E+01 | DHFR;SPR;CAV1;NOSTRIN;CALM1 |
| 18 | Signaling by VEGF Homo sapiens R-HSA-194138 | 2.26E-03 | 1.30E-01 | 1.89E+00 | 1.15E+01 | FLT1;RASGRF2;PDGFB;ITPR3;ARRB1;SHB;IL2RG;FGF2;EGFR;RASGRP3;RAP1B;UBC;KDR;ITGAV;PAK2;SPTBN1;KSR1;VWF;DUSP1;CAV1;DUSP6;MTOR;PSMB9;DLG4;FGF18;IL3RA;CALM1;DOCK1 |
| 19 | Molecules associated with elastic fibres Homo sapiens R-HSA-2129379 | 2.60E-03 | 1.42E-01 | 5.01E+00 | 2.98E+01 | ITGB1;TGFB2;MFAP2;ITGAV;FBLN2;LTBP1 |
| 20 | Stimuli-sensing channels Homo sapiens R-HSA-2672351 | 2.91E-03 | 1.48E-01 | 2.74E+00 | 1.60E+01 | CLCN7;TTYH3;CLCN4;OSTM1;TRPV4;UBC;BSND;CLCNKA;ASIC2;CALM1;SGK1;MCOLN1 |
| 21 | VEGFA-VEGFR2 Pathway Homo sapiens R-HSA-4420097 | 3.15E-03 | 1.48E-01 | 1.86E+00 | 1.07E+01 | RASGRF2;PDGFB;ITPR3;ARRB1;SHB;IL2RG;FGF2;EGFR;RASGRP3;RAP1B;UBC;KDR;ITGAV;PAK2;SPTBN1;KSR1;VWF;DUSP1;CAV1;DUSP6;MTOR;PSMB9;DLG4;FGF18;IL3RA;CALM1;DOCK1 |
| 22 | Cell surface interactions at the vascular wall Homo sapiens R-HSA-202733 | 3.16E-03 | 1.48E-01 | 2.71E+00 | 1.56E+01 | SELP;ITGB1;SLC7A6;CXADR;ANGPT2;GRB14;CAV1;PECAM1;ATP1B3;ITGAV;GAS6;JAM2 |
| 23 | PTK6 promotes HIF1A stabilization Homo sapiens R-HSA-8857538 | 3.30E-03 | 1.48E-01 | 1.50E+01 | 8.56E+01 | LRRK2;HIF1A;EGFR |
| 24 | Synthesis of bile acids and bile salts via 27-hydroxycholesterol Homo sapiens R-HSA-193807 | 4.64E-03 | 2.00E-01 | 7.27E+00 | 3.91E+01 | AKR1C1;NR1H4;AKR1C3;AKR1C2 |
| 25 | Bile acid and bile salt metabolism Homo sapiens R-HSA-194068 | 5.80E-03 | 2.37E-01 | 4.14E+00 | 2.13E+01 | CH25H;AKR1C1;SLCO1A2;NR1H4;AKR1C3;AKR1C2 |
| 26 | Cell-extracellular matrix interactions Homo sapiens R-HSA-446353 | 5.95E-03 | 2.37E-01 | 6.66E+00 | 3.41E+01 | ITGB1;FBLIM1;ACTN1;FERMT2 |
| 27 | Class B/2 (Secretin family receptors) Homo sapiens R-HSA-373080 | 6.66E-03 | 2.56E-01 | 2.71E+00 | 1.36E+01 | RAMP2;CALCA;CALCRL;CALCB;WNT11;FZD2;GIPR;WNT7B;GLP2R;FZD9 |

| | | | | | | |
|---|---|---|---|---|---|---|
| 28 | PTK6 Regulates RTKs and Their Effectors AKT1 and DOK1 Homo sapiens R-HSA-8849469 | 7.36E-03 | 2.72E-01 | 9.99E+00 | 4.91E+01 | DOK1;UBC;ARAP1 |
| 29 | VEGFR2 mediated cell proliferation Homo sapiens R-HSA-5218921 | 8.21E-03 | 2.75E-01 | 1.86E+00 | 8.94E+00 | KSR1;VWF;DUSP1;RASGRF2;PDGFB;ITPR3;ARRB1;IL2RG;FGF2;DUSP6;EGFR;PSMB9;RASGRP3;RAP1B;DLG4;UBC;FGF18;IL3RA;KDR;CALM1;SPTBN1 |
| 30 | Synthesis of bile acids and bile salts Homo sapiens R-HSA-192105 | 8.29E-03 | 2.75E-01 | 4.55E+00 | 2.18E+01 | CH25H;AKR1C1;NR1H4;AKR1C3;AKR1C2 |
| 31 | Amino acid and oligopeptide SLC transporters Homo sapiens R-HSA-425374 | 8.29E-03 | 2.75E-01 | 3.34E+00 | 1.60E+01 | SLC6A6;SLC15A1;SLC38A1;SLC7A6;SLC5A8;SLC6A12;SLC38A2 |
| 32 | Transport of vitamins, nucleosides, and related molecules Homo sapiens R-HSA-425397 | 8.74E-03 | 2.75E-01 | 3.75E+00 | 1.78E+01 | SLC35A3;SLCO4A1;SLCO4C1;SLCO1A2;SLC29A4;SLC29A1 |
| 33 | Transport of glucose and other sugars, bile salts and organic acids, metal ions and amine compounds Homo sapiens R-HSA-425366 | 8.83E-03 | 2.75E-01 | 2.45E+00 | 1.16E+01 | SLC2A9;SLC47A2;SLC14A1;SLC6A6;SLC14A2;SLC6A9;SLC2A1;SLC6A12;SLC5A3;SLC2A6;RUNX1 |
| 34 | NCAM signaling for neurite out-growth Homo sapiens R-HSA-375165 | 9.03E-03 | 2.75E-01 | 1.81E+00 | 8.54E+00 | KSR1;VWF;DUSP1;RASGRF2;PDGFB;ARRB1;CACNA1C;IL2RG;FGF2;DUSP6;EGFR;PSMB9;RASGRP3;RAP1B;DLG4;COL4A4;UBC;FGF18;IL3RA;COL9A2;CALM1;SPTBN1 |
| 35 | Tetrahydrobiopterin (BH4) synthesis, recycling, salvage and regulation Homo sapiens R-HSA-1474151 | 1.01E-02 | 3.00E-01 | 8.56E+00 | 3.93E+01 | DHFR;SPR;CALM1 |
| 36 | Other semaphorin interactions Homo sapiens R-HSA-416700 | 1.13E-02 | 3.11E-01 | 5.33E+00 | 2.39E+01 | ITGB1;PLXND1;SEMA4D;PLXNA4 |
| 37 | Amine compound SLC transporters Homo sapiens R-HSA-425428 | 1.30E-02 | 3.11E-01 | 4.00E+00 | 1.74E+01 | SLC14A1;SLC6A6;SLC14A2;SLC6A9;SLC6A12 |

| | | | | | | |
|---|---|---|---|---|---|---|
| 38 | Activated NOTCH1 Transmits Signal to the Nucleus Homo sapiens R-HSA-2122948 | 1.30E-02 | 3.11E-01 | 4.00E+00 | 1.74E+01 | JAG1;UBC;DNER;DTX1;ARRB1 |
| 39 | Signaling by Leptin Homo sapiens R-HSA-2586552 | 1.30E-02 | 3.11E-01 | 1.80E+00 | 7.83E+00 | KSR1;VWF;DUSP1;RASGRF2;STAT3;PDGFB;ARRB1;IL2RG;FGF2;DUSP6;EGFR;PSMB9;RASGRP3;RAP1B;DLG4;UBC;FGF18;IL3RA;CALM1;SPTBN1 |
| 40 | Transport of organic anions Homo sapiens R-HSA-879518 | 1.35E-02 | 3.11E-01 | 7.49E+00 | 3.23E+01 | SLCO4A1;SLCO4C1;SLCO1A2 |
| 41 | Syndecan interactions Homo sapiens R-HSA-3000170 | 1.36E-02 | 3.11E-01 | 5.00E+00 | 2.15E+01 | ITGB1;ACTN1;ITGAV;FGF2 |
| 42 | Interleukin receptor SHC signaling Homo sapiens R-HSA-912526 | 1.41E-02 | 3.11E-01 | 1.79E+00 | 7.61E+00 | KSR1;VWF;DUSP1;RASGRF2;PDGFB;PIK3R3;ARRB1;IL2RG;FGF2;DUSP6;EGFR;PSMB9;RASGRP3;RAP1B;DLG4;UBC;FGF18;IL3RA;CALM1;SPTBN1 |
| 43 | Signaling by Retinoic Acid Homo sapiens R-HSA-5362517 | 1.41E-02 | 3.11E-01 | 3.33E+00 | 1.42E+01 | ALDH1A3;CPT1A;RDH11;RDH10;AKR1C3;PDK3 |
| 44 | Integrin cell surface interactions Homo sapiens R-HSA-216083 | 1.42E-02 | 3.11E-01 | 2.72E+00 | 1.15E+01 | ITGB1;COL4A4;KDR;PECAM1;ITGAV;COL9A2;HSPG2;JAM2 |
| 45 | Signaling by PTK6 Homo sapiens R-HSA-8848021 | 1.42E-02 | 3.11E-01 | 2.72E+00 | 1.15E+01 | DOK1;LRRK2;UBC;STAT3;ARAP1;DOCK1;HIF1A;EGFR |
| 46 | Semaphorin interactions Homo sapiens R-HSA-373755 | 1.42E-02 | 3.11E-01 | 2.72E+00 | 1.15E+01 | ITGB1;PLXND1;SEMA4D;DPYSL2;MYH9;CRMP1;PAK2;PLXNA4 |
| 47 | Amino acid transport across the plasma membrane Homo sapiens R-HSA-352230 | 1.49E-02 | 3.11E-01 | 3.85E+00 | 1.62E+01 | SLC6A6;SLC38A1;SLC7A6;SLC6A12;SLC38A2 |
| 48 | EPH-Ephrin signaling Homo sapiens R-HSA-2682334 | 1.53E-02 | 3.11E-01 | 2.36E+00 | 9.86E+00 | EFNB2;ARPC1B;ARPC1A;CLTC;KDR;MYH9;SHB;PAK2;MYL12A;EPHA2 |
| 49 | Amyloid fiber formation Homo sapiens R-HSA-977225 | 1.55E-02 | 3.11E-01 | 2.67E+00 | 1.11E+01 | CALCA;GSN;UBC;FURIN;SNCAIP;HSPG2;HIST1H2AC;LTF |

| | | | | | | |
|---|---|---|---|---|---|---|
| 50 | Signaling by PDGF Homo sapiens R-HSA-186797 | 1.63E-02 | 3.11E-01 | 1.61E+00 | 6.65E+00 | RASGRF2;PDGFB;PIK3R3;ITPR3;ARRB1;FURIN;IL2RG;FGF2;EGFR;RASGRP3;RAP1B;PRKAR2B;UBC;SPTBN1;KSR1;VWF;DUSP1;STAT3;DUSP6;MTOR;PSMB9;DLG4;COL4A4;FGF18;IL3RA;COL9A2;CALM1 |
| 51 | Keratan sulfate/keratin metabolism Homo sapiens R-HSA-1638074 | 1.70E-02 | 3.11E-01 | 3.70E+00 | 1.51E+01 | CHST6;B3GNT3;CHST1;B4GALT6;GNS |
| 52 | GRB2 events in EGFR signaling Homo sapiens R-HSA-179812 | 1.80E-02 | 3.11E-01 | 1.77E+00 | 7.10E+00 | KSR1;VWF;DUSP1;RASGRF2;PDGFB;ARRB1;IL2RG;FGF2;DUSP6;EGFR;PSMB9;RASGRP3;RAP1B;DLG4;UBC;FGF18;IL3RA;CALM1;SPTBN1 |
| 53 | SHC1 events in EGFR signaling Homo sapiens R-HSA-180336 | 1.80E-02 | 3.11E-01 | 1.77E+00 | 7.10E+00 | KSR1;VWF;DUSP1;RASGRF2;PDGFB;ARRB1;IL2RG;FGF2;DUSP6;EGFR;PSMB9;RASGRP3;RAP1B;DLG4;UBC;FGF18;IL3RA;CALM1;SPTBN1 |
| 54 | SOS-mediated signalling Homo sapiens R-HSA-112412 | 1.80E-02 | 3.11E-01 | 1.77E+00 | 7.10E+00 | KSR1;VWF;DUSP1;RASGRF2;PDGFB;ARRB1;IL2RG;FGF2;DUSP6;EGFR;PSMB9;RASGRP3;RAP1B;DLG4;UBC;FGF18;IL3RA;CALM1;SPTBN1 |
| 55 | SHC1 events in ERBB4 signaling Homo sapiens R-HSA-1250347 | 1.80E-02 | 3.11E-01 | 1.77E+00 | 7.10E+00 | KSR1;VWF;DUSP1;RASGRF2;PDGFB;ARRB1;IL2RG;FGF2;DUSP6;EGFR;PSMB9;RASGRP3;RAP1B;DLG4;UBC;FGF18;IL3RA;CALM1;SPTBN1 |
| 56 | RAF/MAP kinase cascade Homo sapiens R-HSA-5673001 | 1.80E-02 | 3.11E-01 | 1.77E+00 | 7.10E+00 | KSR1;VWF;DUSP1;RASGRF2;PDGFB;ARRB1;IL2RG;FGF2;DUSP6;EGFR;PSMB9;RASGRP3;RAP1B;DLG4;UBC;FGF18;IL3RA;CALM1;SPTBN1 |
| 57 | MAPK family signaling cascades Homo sapiens R-HSA-5683057 | 1.82E-02 | 3.11E-01 | 1.69E+00 | 6.77E+00 | KSR1;VWF;DUSP1;RASGRF2;PDGFB;ARRB1;IL2RG;FGF2;DUSP6;EGFR;PSMB9;RASGRP3;RAP1B;DLG4;CDC42EP3;UBC;FGF18;IL3RA;CDC42EP2;CALM1;PAK2;SPTBN1 |
| 58 | Interleukin-2 signaling Homo sapiens R-HSA-451927 | 1.86E-02 | 3.11E-01 | 1.73E+00 | 6.90E+00 | KSR1;VWF;DUSP1;RASGRF2;PDGFB;PIK3R3;ARRB1;IL2RG;FGF2;DUSP6;EGFR;PSMB9;RASGRP3;RAP1B;DLG4;UBC;FGF18;IL3RA;CALM1;SPTBN1 |
| 59 | FRS-mediated FGFR2 signaling Homo sapiens R-HSA-5654700 | 1.88E-02 | 3.11E-01 | 1.76E+00 | 6.99E+00 | KSR1;VWF;DUSP1;RASGRF2;PDGFB;ARRB1;IL2RG;FGF2;DUSP6;EGFR;PSMB9;RASGRP3;RAP1B;DLG4;UBC;FGF18;IL3RA;CALM1;SPTBN1 |
| 60 | FRS-mediated FGFR4 signaling Homo sapiens R-HSA-5654712 | 1.88E-02 | 3.11E-01 | 1.76E+00 | 6.99E+00 | KSR1;VWF;DUSP1;RASGRF2;PDGFB;ARRB1;IL2RG;FGF2;DUSP6;EGFR;PSMB9;RASGRP3;RAP1B;DLG4;UBC;FGF18;IL3RA;CALM1;SPTBN1 |
| 61 | FRS-mediated FGFR3 signaling Homo sapiens R-HSA-5654706 | 1.88E-02 | 3.11E-01 | 1.76E+00 | 6.99E+00 | KSR1;VWF;DUSP1;RASGRF2;PDGFB;ARRB1;IL2RG;FGF2;DUSP6;EGFR;PSMB9;RASGRP3;RAP1B;DLG4;UBC;FGF18;IL3RA;CALM1;SPTBN1 |

| | | | | | | |
|---|---|---|---|---|---|---|
| 62 | FRS-mediated FGFR1 signaling Homo sapiens R-HSA-5654693 | 1.88E-02 | 3.11E-01 | 1.76E+00 | 6.99E+00 | KSR1;VWF;DUSP1;RASGRF2;PDGFB;ARRB1;IL2RG;FGF2;DUSP6;EGFR;PSMB9;RASGRP3;RAP1B;DLG4;UBC;FGF18;IL3RA;CALM1;SPTBN1 |
| 63 | RA biosynthesis pathway Homo sapiens R-HSA-5365859 | 1.91E-02 | 3.11E-01 | 4.44E+00 | 1.76E+01 | ALDH1A3;RDH11;RDH10;AKR1C3 |
| 64 | Transport of inorganic cations/anions and amino acids/oligopeptides Homo sapiens R-HSA-425393 | 1.99E-02 | 3.11E-01 | 2.25E+00 | 8.82E+00 | SLC6A6;SLC15A1;SLC38A1;SLC5A8;SLC7A6;SLC6A12;SLC26A7;CALM1;SLC38A2;SLC26A6 |
| 65 | NOSTRIN mediated eNOS trafficking Homo sapiens R-HSA-203641 | 2.07E-02 | 3.11E-01 | 1.33E+01 | 5.16E+01 | CAV1;NOSTRIN |
| 66 | ARMS-mediated activation Homo sapiens R-HSA-170984 | 2.11E-02 | 3.11E-01 | 1.74E+00 | 6.69E+00 | KSR1;VWF;DUSP1;RASGRF2;PDGFB;ARRB1;IL2RG;FGF2;DUSP6;EGFR;PSMB9;RASGRP3;RAP1B;DLG4;UBC;FGF18;IL3RA;CALM1;SPTBN1 |
| 67 | Signalling to p38 via RIT and RIN Homo sapiens R-HSA-187706 | 2.11E-02 | 3.11E-01 | 1.74E+00 | 6.69E+00 | KSR1;VWF;DUSP1;RASGRF2;PDGFB;ARRB1;IL2RG;FGF2;DUSP6;EGFR;PSMB9;RASGRP3;RAP1B;DLG4;UBC;FGF18;IL3RA;CALM1;SPTBN1 |
| 68 | Cell junction organization Homo sapiens R-HSA-446728 | 2.15E-02 | 3.11E-01 | 2.34E+00 | 8.99E+00 | ITGB1;CLDN5;CDH4;CADM1;FBLIM1;ACTN1;PVR;CLDN1;FERMT2 |
| 69 | Facilitative Na+-independent glucose transporters Homo sapiens R-HSA-428790 | 2.17E-02 | 3.11E-01 | 5.99E+00 | 2.29E+01 | SLC2A9;SLC2A1;SLC2A6 |
| 70 | FGFR3c ligand binding and activation Homo sapiens R-HSA-190372 | 2.17E-02 | 3.11E-01 | 5.99E+00 | 2.29E+01 | GALNT3;FGF18;FGF2 |
| 71 | FGFR3 ligand binding and activation Homo sapiens R-HSA-190239 | 2.17E-02 | 3.11E-01 | 5.99E+00 | 2.29E+01 | GALNT3;FGF18;FGF2 |
| 72 | Frs2-mediated activation Homo sapiens R-HSA-170968 | 2.19E-02 | 3.11E-01 | 1.73E+00 | 6.60E+00 | KSR1;VWF;DUSP1;RASGRF2;PDGFB;ARRB1;IL2RG;FGF2;DUSP6;EGFR;PSMB9;RASGRP3;RAP1B;DLG4;UBC;FGF18;IL3RA;CALM1;SPTBN1 |

| | | | | | | |
|---|---|---|---|---|---|---|
| 73 | The canonical retinoid cycle in rods (twilight vision) Homo sapiens R-HSA-2453902 | 2.22E-02 | 3.11E-01 | 4.21E+00 | 1.60E+01 | RDH11;RDH10;ABCA4;NAPEPLD |
| 74 | Laminin interactions Homo sapiens R-HSA-3000157 | 2.22E-02 | 3.11E-01 | 4.21E+00 | 1.60E+01 | ITGB1;LAMB2;ITGAV;HSPG2 |
| 75 | Cytokine Signaling in Immune system Homo sapiens R-HSA-1280215 | 2.27E-02 | 3.11E-01 | 1.43E+00 | 5.41E+00 | IFITM3;IFITM2;RASGRF2;PDGFB;PIK3R3;ARRB1;IL2RG;FGF2;EGFR;RASGRP3;SOCS2;RAP1B;IRAK2;UBC;STX3;GBP2;SPTBN1;HLA-DPA1;IP6K2;EDARADD;NUP210;KSR1;VWF;TNFRSF12A;DUSP1;DDX58;STAT3;ISG15;HLA-A;TNFRSF1B;DUSP6;IL17RB;PSMB9;IFI27;DLG4;NEDD4;IL3RA;FGF18;HLA-DPB1;CALM1;EIF4G3 |
| 76 | MAPK1/MAPK3 signaling Homo sapiens R-HSA-5684996 | 2.28E-02 | 3.11E-01 | 1.72E+00 | 6.50E+00 | KSR1;VWF;DUSP1;RASGRF2;PDGFB;ARRB1;IL2RG;FGF2;DUSP6;EGFR;PSMB9;RASGRP3;RAP1B;DLG4;UBC;FGF18;IL3RA;CALM1;SPTBN1 |
| 77 | Prolonged ERK activation events Homo sapiens R-HSA-169893 | 2.37E-02 | 3.18E-01 | 1.71E+00 | 6.41E+00 | KSR1;VWF;DUSP1;RASGRF2;PDGFB;ARRB1;IL2RG;FGF2;DUSP6;EGFR;PSMB9;RASGRP3;RAP1B;DLG4;UBC;FGF18;IL3RA;CALM1;SPTBN1 |
| 78 | Synthesis of bile acids and bile salts via 7alpha-hydroxycholesterol Homo sapiens R-HSA-193368 | 2.57E-02 | 3.37E-01 | 4.00E+00 | 1.46E+01 | AKR1C1;NR1H4;AKR1C3;AKR1C2 |
| 79 | Interleukin-3, 5 and GM-CSF signaling Homo sapiens R-HSA-512988 | 2.60E-02 | 3.37E-01 | 1.67E+00 | 6.09E+00 | KSR1;VWF;DUSP1;RASGRF2;PDGFB;PIK3R3;ARRB1;IL2RG;FGF2;DUSP6;EGFR;PSMB9;RASGRP3;RAP1B;DLG4;UBC;FGF18;IL3RA;CALM1;SPTBN1 |
| 80 | Receptor-ligand binding initiates the second proteolytic cleavage of Notch receptor Homo sapiens R-HSA-156988 | 2.67E-02 | 3.37E-01 | 5.45E+00 | 1.97E+01 | JAG1;NOTCH4;UBC |
| 81 | Synthesis of bile acids and bile salts via 24-hydroxycholesterol Homo sapiens R-HSA-193775 | 2.67E-02 | 3.37E-01 | 5.45E+00 | 1.97E+01 | AKR1C1;AKR1C3;AKR1C2 |

| | | | | | | |
|---|---|---|---|---|---|---|
| 82 | Synthesis of Prostaglandins (PG) and Thromboxanes (TX) Homo sapiens R-HSA-2162123 | 2.67E-02 | 3.37E-01 | 5.45E+00 | 1.97E+01 | AKR1C3;PTGS1;PTGES |
| 83 | Signalling to RAS Homo sapiens R-HSA-167044 | 2.74E-02 | 3.43E-01 | 1.68E+00 | 6.04E+00 | KSR1;VWF;DUSP1;RASGRF2;PDGFB;ARRB1;IL2RG;FGF2;DUSP6;EGFR;PSMB9;RASGRP3;RAP1B;DLG4;UBC;FGF18;IL3RA;CALM1;SPTBN1 |
| 84 | Cardiac conduction Homo sapiens R-HSA-5576891 | 2.84E-02 | 3.51E-01 | 1.96E+00 | 6.96E+00 | FXYD4;KCNE4;NPR1;ATP2A3;KCNK15;ORAI1;ATP1B3;ITPR3;CACNA1C;CALM1;FGF12;KCNK3 |
| 85 | Basigin interactions Homo sapiens R-HSA-210991 | 2.95E-02 | 3.59E-01 | 3.81E+00 | 1.34E+01 | ITGB1;SLC7A6;CAV1;ATP1B3 |
| 86 | Choline catabolism Homo sapiens R-HSA-6798163 | 3.01E-02 | 3.60E-01 | 9.98E+00 | 3.50E+01 | DMGDH;CHDH |
| 87 | Signaling by FGFR3 Homo sapiens R-HSA-5654741 | 3.03E-02 | 3.60E-01 | 1.56E+00 | 5.46E+00 | KSR1;VWF;GALNT3;DUSP1;RASGRF2;PDGFB;PIK3R3;ITPR3;ARRB1;IL2RG;FGF2;DUSP6;EGFR;MTOR;PSMB9;RASGRP3;RAP1B;PRKAR2B;DLG4;UBC;FGF18;IL3RA;CALM1;SPTBN1 |
| 88 | IRS-mediated signalling Homo sapiens R-HSA-112399 | 3.22E-02 | 3.79E-01 | 1.60E+00 | 5.51E+00 | KSR1;VWF;CAB39L;DUSP1;RASGRF2;PDGFB;ARRB1;IL2RG;FGF2;DUSP6;EGFR;MTOR;PSMB9;RASGRP3;RAP1B;DLG4;UBC;FGF18;IL3RA;CALM1;SPTBN1 |
| 89 | Ion homeostasis Homo sapiens R-HSA-5578775 | 3.39E-02 | 3.93E-01 | 2.67E+00 | 9.02E+00 | FXYD4;ATP2A3;ORAI1;ATP1B3;ITPR3;CALM1 |
| 90 | Ion channel transport Homo sapiens R-HSA-983712 | 3.46E-02 | 3.93E-01 | 1.72E+00 | 5.77E+00 | TTYH3;ATP2A3;CLCNKA;ATP1B3;MCOLN1;CLCN7;CLCN4;FXYD4;OSTM1;TRPV4;UBC;BSND;CALM1;SGK1;ASIC2;ATP9A |
| 91 | Signalling to ERKs Homo sapiens R-HSA-187687 | 3.51E-02 | 3.93E-01 | 1.63E+00 | 5.46E+00 | KSR1;VWF;DUSP1;RASGRF2;PDGFB;ARRB1;IL2RG;FGF2;DUSP6;EGFR;PSMB9;RASGRP3;RAP1B;DLG4;UBC;FGF18;IL3RA;CALM1;SPTBN1 |
| 92 | Insulin receptor signalling cascade Homo sapiens R-HSA-74751 | 3.54E-02 | 3.93E-01 | 1.59E+00 | 5.30E+00 | KSR1;VWF;CAB39L;DUSP1;RASGRF2;PDGFB;ARRB1;IL2RG;FGF2;DUSP6;EGFR;MTOR;PSMB9;RASGRP3;RAP1B;DLG4;UBC;FGF18;IL3RA;CALM1;SPTBN1 |
| 93 | IGF1R signaling cascade Homo sapiens R-HSA-2428924 | 3.66E-02 | 3.93E-01 | 1.58E+00 | 5.22E+00 | KSR1;VWF;CAB39L;DUSP1;RASGRF2;PDGFB;ARRB1;IL2RG;FGF2;DUSP6;EGFR;MTOR;PSMB9;RASGRP3;RAP1B;DLG4;UBC;FGF18;IL3RA;CALM1;SPTBN1 |

| | | | | | | |
|---|---|---|---|---|---|---|
| 94 | Signaling by Type 1 Insulin-like Growth Factor 1 Receptor (IGF1R) Homo sapiens R-HSA-2404192 | 3.66E-02 | 3.93E-01 | 1.58E+00 | 5.22E+00 | KSR1;VWF;CAB39L;DUSP1;RASGRF2;PDGFB;ARRB1;IL2RG;FGF2;DUSP6;EGFR;MTOR;PSMB9;RASGRP3;RAP1B;DLG4;UBC;FGF18;IL3RA;CALM1;SPTBN1 |
| 95 | IRS-related events triggered by IGF1R Homo sapiens R-HSA-2428928 | 3.66E-02 | 3.93E-01 | 1.58E+00 | 5.22E+00 | KSR1;VWF;CAB39L;DUSP1;RASGRF2;PDGFB;ARRB1;IL2RG;FGF2;DUSP6;EGFR;MTOR;PSMB9;RASGRP3;RAP1B;DLG4;UBC;FGF18;IL3RA;CALM1;SPTBN1 |
| 96 | Keratan sulfate biosynthesis Homo sapiens R-HSA-2022854 | 3.79E-02 | 3.93E-01 | 3.47E+00 | 1.14E+01 | CHST6;B3GNT3;CHST1;B4GALT6 |
| 97 | VEGFR2 mediated vascular permeability Homo sapiens R-HSA-5218920 | 3.79E-02 | 3.93E-01 | 3.47E+00 | 1.14E+01 | CAV1;CALM1;PAK2;MTOR |
| 98 | Platelet calcium homeostasis Homo sapiens R-HSA-418360 | 3.79E-02 | 3.93E-01 | 3.47E+00 | 1.14E+01 | ATP2A3;ORAI1;ITPR3;CALM1 |
| 99 | Rap1 signalling Homo sapiens R-HSA-392517 | 3.83E-02 | 3.93E-01 | 4.61E+00 | 1.50E+01 | RAP1B;RAPGEF3;SIPA1 |
| 100 | CRMPs in Sema3A signaling Homo sapiens R-HSA-399956 | 3.83E-02 | 3.93E-01 | 4.61E+00 | 1.50E+01 | DPYSL2;CRMP1;PLXNA4 |
| 101 | Downstream signal transduction Homo sapiens R-HSA-186763 | 3.83E-02 | 3.93E-01 | 1.52E+00 | 4.96E+00 | KSR1;VWF;DUSP1;RASGRF2;STAT3;PDGFB;PIK3R3;ITPR3;ARRB1;IL2RG;FGF2;DUSP6;EGFR;MTOR;PSMB9;RASGRP3;RAP1B;PRKAR2B;DLG4;UBC;FGF18;IL3RA;CALM1;SPTBN1 |
| 102 | Signaling by SCF-KIT Homo sapiens R-HSA-1433557 | 3.98E-02 | 4.03E-01 | 1.53E+00 | 4.93E+00 | KSR1;VWF;DUSP1;RASGRF2;STAT3;PDGFB;PIK3R3;ARRB1;IL2RG;FGF2;DUSP6;EGFR;MTOR;PSMB9;RASGRP3;RAP1B;DLG4;UBC;FGF18;IL3RA;GRAP;CALM1;SPTBN1 |
| 103 | Transport of nucleosides and free purine and pyrimidine bases across the plasma membrane Homo sapiens R-HSA-83936 | 4.08E-02 | 4.03E-01 | 7.98E+00 | 2.55E+01 | SLC29A4;SLC29A1 |
| 104 | Nectin/Necl trans heterodimerization Homo sapiens R-HSA-420597 | 4.08E-02 | 4.03E-01 | 7.98E+00 | 2.55E+01 | CADM1;PVR |

| | | | | | | |
|---|---|---|---|---|---|---|
| 105 | Multifunctional anion exchangers Homo sapiens R-HSA-427601 | 4.08E-02 | 4.03E-01 | 7.98E+00 | 2.55E+01 | SLC26A7;SLC26A6 |
| 106 | Gastrin-CREB signalling pathway via PKC and MAPK Homo sapiens R-HSA-881907 | 4.19E-02 | 4.09E-01 | 1.45E+00 | 4.59E+00 | RGS19;GPR68;RASGRF2;PDGFB;PIK3R3;ITPR3;ARRB1;IL2RG;NTS;FGF2;EGFR;RASGRP3;RAP1B;RGS3;UBC;BDKRB2;SPTBN1;ANXA1;KSR1;VWF;DUSP1;F2R;GPR4;DUSP6;PSMB9;DLG4;IL3RA;FGF18;CALM1 |
| 107 | Interferon alpha/beta signaling Homo sapiens R-HSA-909733 | 4.32E-02 | 4.19E-01 | 2.30E+00 | 7.21E+00 | IFITM3;IFITM2;IFI27;ISG15;HLA-A;GBP2;IP6K2 |
| 108 | Downstream signaling of activated FGFR2 Homo sapiens R-HSA-5654696 | 4.46E-02 | 4.19E-01 | 1.51E+00 | 4.70E+00 | KSR1;VWF;DUSP1;RASGRF2;PDGFB;PIK3R3;ITPR3;ARRB1;IL2RG;FGF2;DUSP6;EGFR;MTOR;PSMB9;RASGRP3;RAP1B;PRKAR2B;DLG4;UBC;FGF18;IL3RA;CALM1;SPTBN1 |
| 109 | Downstream signaling of activated FGFR4 Homo sapiens R-HSA-5654716 | 4.46E-02 | 4.19E-01 | 1.51E+00 | 4.70E+00 | KSR1;VWF;DUSP1;RASGRF2;PDGFB;PIK3R3;ITPR3;ARRB1;IL2RG;FGF2;DUSP6;EGFR;MTOR;PSMB9;RASGRP3;RAP1B;PRKAR2B;DLG4;UBC;FGF18;IL3RA;CALM1;SPTBN1 |
| 110 | Downstream signaling of activated FGFR3 Homo sapiens R-HSA-5654708 | 4.46E-02 | 4.19E-01 | 1.51E+00 | 4.70E+00 | KSR1;VWF;DUSP1;RASGRF2;PDGFB;PIK3R3;ITPR3;ARRB1;IL2RG;FGF2;DUSP6;EGFR;MTOR;PSMB9;RASGRP3;RAP1B;PRKAR2B;DLG4;UBC;FGF18;IL3RA;CALM1;SPTBN1 |
| 111 | Ras activation uopn Ca2+ infux through NMDA receptor Homo sapiens R-HSA-442982 | 4.49E-02 | 4.19E-01 | 4.28E+00 | 1.33E+01 | DLG4;RASGRF2;CALM1 |
| 112 | ECM proteoglycans Homo sapiens R-HSA-3000178 | 4.66E-02 | 4.31E-01 | 2.45E+00 | 7.51E+00 | ITGB1;TGFB2;SPARC;LAMB2;ITGAV;COL9A2 |
| 113 | Signaling by FGFR4 Homo sapiens R-HSA-5654743 | 4.85E-02 | 4.41E-01 | 1.49E+00 | 4.52E+00 | KSR1;VWF;DUSP1;RASGRF2;PDGFB;PIK3R3;ITPR3;ARRB1;IL2RG;FGF2;DUSP6;EGFR;MTOR;PSMB9;RASGRP3;RAP1B;PRKAR2B;DLG4;UBC;FGF18;IL3RA;CALM1;SPTBN1 |
| 114 | Downstream signaling of activated FGFR1 Homo sapiens R-HSA-5654687 | 4.85E-02 | 4.41E-01 | 1.49E+00 | 4.52E+00 | KSR1;VWF;DUSP1;RASGRF2;PDGFB;PIK3R3;ITPR3;ARRB1;IL2RG;FGF2;DUSP6;EGFR;MTOR;PSMB9;RASGRP3;RAP1B;PRKAR2B;DLG4;UBC;FGF18;IL3RA;CALM1;SPTBN1 |
| 115 | Pre-NOTCH Processing in Golgi Homo sapiens R-HSA-1912420 | 5.20E-02 | 4.55E-01 | 3.99E+00 | 1.18E+01 | NOTCH4;ATP2A3;FURIN |

| | | | | | | |
|---|---|---|---|---|---|---|
| 116 | Erythrocytes take up oxygen and release carbon dioxide Homo sapiens R-HSA-1247673 | 5.28E-02 | 4.55E-01 | 6.65E+00 | 1.96E+01 | CA2;AQP1 |
| 117 | Downregulation of ERBB4 signaling Homo sapiens R-HSA-1253288 | 5.28E-02 | 4.55E-01 | 6.65E+00 | 1.96E+01 | NEDD4;UBC |
| 118 | VEGF ligand-receptor interactions Homo sapiens R-HSA-194313 | 5.28E-02 | 4.55E-01 | 6.65E+00 | 1.96E+01 | FLT1;KDR |
| 119 | VEGF binds to VEGFR leading to receptor dimerization Homo sapiens R-HSA-195399 | 5.28E-02 | 4.55E-01 | 6.65E+00 | 1.96E+01 | FLT1;KDR |
| 120 | Coenzyme A biosynthesis Homo sapiens R-HSA-196783 | 5.28E-02 | 4.55E-01 | 6.65E+00 | 1.96E+01 | PANK2;COASY |
| 121 | Signaling by FGFR1 Homo sapiens R-HSA-5654736 | 5.41E-02 | 4.63E-01 | 1.48E+00 | 4.30E+00 | KSR1;VWF;DUSP1;RASGRF2;PDGFB;PIK3R3;ITPR3;ARRB1;IL2RG;FGF2;DUSP6;EGFR;MTOR;PSMB9;RASGRP3;RAP1B;PRKAR2B;DLG4;UBC;FGF18;IL3RA;CALM1;SPTBN1 |
| 122 | Phospholipase C-mediated cascade; FGFR3 Homo sapiens R-HSA-5654227 | 5.76E-02 | 4.84E-01 | 2.56E+00 | 7.31E+00 | PRKAR2B;FGF18;ITPR3;CALM1;FGF2 |
| 123 | Inositol phosphate metabolism Homo sapiens R-HSA-1483249 | 5.76E-02 | 4.84E-01 | 2.56E+00 | 7.31E+00 | ITPKB;INPP4B;INPP1;CALM1;IP6K2 |
| 124 | NGF signalling via TRKA from the plasma membrane Homo sapiens R-HSA-187037 | 5.79E-02 | 4.84E-01 | 1.44E+00 | 4.10E+00 | RASGRF2;PDGFB;PIK3R3;ITPR3;ARRB1;IL2RG;FGF2;EGFR;RASGRP3;RAP1B;PRKAR2B;UBC;SPTBN1;DUSP3;KSR1;VWF;DUSP1;STAT3;DUSP6;MTOR;PSMB9;DLG4;FGF18;IL3RA;CALM1 |
| 125 | Na+/Cl- dependent neurotransmitter transporters Homo sapiens R-HSA-442660 | 5.96E-02 | 4.90E-01 | 3.74E+00 | 1.06E+01 | SLC6A6;SLC6A9;SLC6A12 |
| 126 | Ephrin signaling Homo sapiens R-HSA-3928664 | 5.96E-02 | 4.90E-01 | 3.74E+00 | 1.06E+01 | EFNB2;PAK2;MYL12A |

| | | | | | | |
|---|---|---|---|---|---|---|
| 127 | FCERI mediated MAPK activation Homo sapiens R-HSA-2871796 | 6.30E-02 | 5.14E-01 | 1.49E+00 | 4.12E+00 | KSR1;VWF;DUSP1;RASGRF2;PDGFB;ARRB1;IL2RG;FGF2;DUSP6;EGFR;PSMB9;RASGRP3;RAP1B;DLG4;UBC;FGF18;IL3RA;CALM1;PAK2;SPTBN1 |
| 128 | Oncogene Induced Senescence Homo sapiens R-HSA-2559585 | 6.46E-02 | 5.23E-01 | 2.85E+00 | 7.82E+00 | UBC;E2F3;ETS1;ETS2 |
| 129 | Crosslinking of collagen fibrils Homo sapiens R-HSA-2243919 | 6.57E-02 | 5.24E-01 | 5.70E+00 | 1.55E+01 | LOXL4;LOXL2 |
| 130 | Ca2+ activated K+ channels Homo sapiens R-HSA-1296052 | 6.57E-02 | 5.24E-01 | 5.70E+00 | 1.55E+01 | KCNMB4;KCNN3 |
| 131 | DAP12 signaling Homo sapiens R-HSA-2424491 | 6.65E-02 | 5.24E-01 | 1.44E+00 | 3.90E+00 | KSR1;VWF;DUSP1;RASGRF2;PDGFB;PIK3R3;ITPR3;ARRB1;IL2RG;FGF2;DUSP6;EGFR;MTOR;PSMB9;RASGRP3;RAP1B;PRKAR2B;DLG4;UBC;FGF18;IL3RA;CALM1;SPTBN1 |
| 132 | Signaling by NOTCH Homo sapiens R-HSA-157118 | 6.73E-02 | 5.24E-01 | 1.79E+00 | 4.82E+00 | JAG1;NOTCH4;UBC;DNER;ATP2A3;DTX1;ARRB1;FURIN;E2F3;HIF1A |
| 133 | Phospholipase C-mediated cascade; FGFR4 Homo sapiens R-HSA-5654228 | 6.74E-02 | 5.24E-01 | 2.44E+00 | 6.57E+00 | PRKAR2B;FGF18;ITPR3;CALM1;FGF2 |
| 134 | Regulation of gene expression in beta cells Homo sapiens R-HSA-210745 | 6.77E-02 | 5.24E-01 | 3.52E+00 | 9.49E+00 | MAFA;FOXA3;FOXA2 |
| 135 | Signaling by Insulin receptor Homo sapiens R-HSA-74752 | 7.09E-02 | 5.43E-01 | 1.45E+00 | 3.84E+00 | KSR1;VWF;CAB39L;DUSP1;RASGRF2;PDGFB;ARRB1;IL2RG;FGF2;DUSP6;EGFR;MTOR;PSMB9;RASGRP3;RAP1B;DLG4;UBC;FGF18;IL3RA;CALM1;SPTBN1 |
| 136 | Metabolism of lipids and lipoproteins Homo sapiens R-HSA-556833 | 7.24E-02 | 5.43E-01 | 1.30E+00 | 3.41E+00 | DMGDH;GLTP;AGPAT5;ACHE;MVK;FHL2;PIK3R3;AKR1B1;ABHD5;NPAS2;ACACA;PTGS1;PIK3C2B;SMPD3;LIPG;HMGCS2;A2M;ARSA;CPT1A;PLA2G4F;AKR1C1;CAV1;NR1H4;ACSL6;AKR1C3;AKR1C2;DHCR24;HSPG2;PISD;CH25H;INPP4B;CHDH;LPCAT2;SLCO1A2;CYP1A1;PPARG;FAR2;ANGPTL4;LPIN2;PTGES |
| 137 | Signaling by ERBB4 Homo sapiens R-HSA-1236394 | 7.32E-02 | 5.43E-01 | 1.43E+00 | 3.75E+00 | KSR1;VWF;DUSP1;RASGRF2;PDGFB;PIK3R3;ARRB1;IL2RG;FGF2;DUSP6;EGFR;MTOR;PSMB9;RASGRP3;RAP1B;DLG4;NEDD4;UBC;FGF18;IL3RA;CALM1;SPTBN1 |

| | | | | | | |
|---|---|---|---|---|---|---|
| 138 | Signaling by FGFR Homo sapiens R-HSA-190236 | 7.36E-02 | 5.43E-01 | 1.41E+00 | 3.67E+00 | KSR1;VWF;GALNT3;DUSP1;RASGRF2;PDGFB;PIK3R3;ITPR3;ARRB1;IL2RG;FGF2;DUSP6;EGFR;MTOR;PSMB9;RASGRP3;RAP1B;PRKAR2B;DLG4;UBC;FGF18;IL3RA;CALM1;SPTBN1 |
| 139 | Signal transduction by L1 Homo sapiens R-HSA-445144 | 7.63E-02 | 5.43E-01 | 3.33E+00 | 8.56E+00 | ITGB1;ITGAV;EGFR |
| 140 | Regulation of FZD by ubiquitination Homo sapiens R-HSA-4641263 | 7.63E-02 | 5.43E-01 | 3.33E+00 | 8.56E+00 | UBC;RSPO3;LGR5 |
| 141 | RHO GTPases activate PAKs Homo sapiens R-HSA-5627123 | 7.63E-02 | 5.43E-01 | 3.33E+00 | 8.56E+00 | MYH9;CALM1;PAK2 |
| 142 | EPHA-mediated growth cone collapse Homo sapiens R-HSA-3928663 | 7.74E-02 | 5.43E-01 | 2.66E+00 | 6.82E+00 | KDR;MYH9;SHB;EPHA2 |
| 143 | EGFR interacts with phospholipase C-gamma Homo sapiens R-HSA-212718 | 7.74E-02 | 5.43E-01 | 2.66E+00 | 6.82E+00 | PRKAR2B;ITPR3;CALM1;EGFR |
| 144 | Abacavir transport and metabolism Homo sapiens R-HSA-2161522 | 7.96E-02 | 5.43E-01 | 4.99E+00 | 1.26E+01 | ABCB1;PCK1 |
| 145 | Removal of aminoterminal propeptides from gamma-carboxylated proteins Homo sapiens R-HSA-159782 | 7.96E-02 | 5.43E-01 | 4.99E+00 | 1.26E+01 | FURIN;GAS6 |
| 146 | Nef mediated downregulation of MHC class I complex cell surface expression Homo sapiens R-HSA-164940 | 7.96E-02 | 5.43E-01 | 4.99E+00 | 1.26E+01 | AP1S2;HLA-A |
| 147 | Activation of Ca-permeable Kainate Receptor Homo sapiens R-HSA-451308 | 7.96E-02 | 5.43E-01 | 4.99E+00 | 1.26E+01 | DLG4;CALM1 |

| | | | | | | |
|---|---|---|---|---|---|---|
| 148 | Ionotropic activity of Kainate Receptors Homo sapiens R-HSA-451306 | 7.96E-02 | 5.43E-01 | 4.99E+00 | 1.26E+01 | DLG4;CALM1 |
| 149 | Reactions specific to the complex N-glycan synthesis pathway Homo sapiens R-HSA-975578 | 7.96E-02 | 5.43E-01 | 4.99E+00 | 1.26E+01 | MAN2A2;FUCA1 |
| 150 | IRAK2 mediated activation of TAK1 complex upon TLR7/8 or 9 stimulation Homo sapiens R-HSA-975163 | 7.96E-02 | 5.43E-01 | 4.99E+00 | 1.26E+01 | IRAK2;UBC |
| 151 | IRAK2 mediated activation of TAK1 complex Homo sapiens R-HSA-937042 | 7.96E-02 | 5.43E-01 | 4.99E+00 | 1.26E+01 | IRAK2;UBC |
| 152 | eNOS activation Homo sapiens R-HSA-203615 | 7.96E-02 | 5.43E-01 | 4.99E+00 | 1.26E+01 | CAV1;CALM1 |
| 153 | Phospholipase C-mediated cascade; FGFR2 Homo sapiens R-HSA-5654221 | 8.36E-02 | 5.63E-01 | 2.27E+00 | 5.63E+00 | PRKAR2B;FGF18;ITPR3;CALM1;FGF2 |
| 154 | Metabolism of carbohydrates Homo sapiens R-HSA-71387 | 8.40E-02 | 5.63E-01 | 1.45E+00 | 3.59E+00 | CHST6;PFKFB3;NUP210;CHPF2;SLC2A1;AKR1B1;HS6ST1;HSPG2;SLC5A3;GNS;HK2;UBC;CHST15;B3GNT3;CHST1;PCK1;CALM1;B4GALT6;PFKP |
| 155 | Lysosome Vesicle Biogenesis Homo sapiens R-HSA-432720 | 8.42E-02 | 5.63E-01 | 2.58E+00 | 6.38E+00 | CLTC;AP1S2;ARRB1;GNS |
| 156 | Intrinsic Pathway of Fibrin Clot Formation Homo sapiens R-HSA-140837 | 8.53E-02 | 5.67E-01 | 3.15E+00 | 7.76E+00 | VWF;A2M;SERPINA5 |
| 157 | Signaling by EGFR Homo sapiens R-HSA-177929 | 8.68E-02 | 5.73E-01 | 1.39E+00 | 3.40E+00 | KSR1;VWF;DUSP1;RASGRF2;PDGFB;PIK3R3;ITPR3;ARRB1;IL2RG;FGF2;DUSP6;EGFR;MTOR;PSMB9;RASGRP3;RAP1B;PRKAR2B;DLG4;UBC;FGF18;IL3RA;CALM1;SPTBN1 |
| 158 | Interferon Signaling Homo sapiens R-HSA-913531 | 8.77E-02 | 5.75E-01 | 1.54E+00 | 3.75E+00 | IFITM3;IFITM2;NUP210;DDX58;ISG15;HLA-A;IFI27;NEDD4;UBC;HLA-DPB1;GBP2;EIF4G3;HLA-DPA1;IP6K2 |

| | | | | | | |
|---|---|---|---|---|---|---|
| 159 | L1CAM interactions Homo sapiens R-HSA-373760 | 8.85E-02 | 5.76E-01 | 1.82E+00 | 4.41E+00 | ITGB1;DLG4;DPYSL2;RANBP9;ITGAV;ANK2;EGFR;SPTBN1 |
| 160 | Visual phototransduction Homo sapiens R-HSA-2187338 | 9.26E-02 | 5.78E-01 | 1.80E+00 | 4.28E+00 | RDH11;RDH10;AKR1C1;ABCA4;NAPEPLD;AKR1C3;CALM1;HSPG2 |
| 161 | Fc epsilon receptor (FCERI) signaling Homo sapiens R-HSA-2454202 | 9.40E-02 | 5.78E-01 | 1.36E+00 | 3.20E+00 | RASGRF2;PDGFB;PIK3R3;ITPR3;ARRB1;IL2RG;FGF2;EGFR;MALT1;RASGRP3;RAP1B;UBC;PAK2;SPTBN1;KSR1;VWF;DUSP1;DUSP6;MTOR;PSMB9;NFKBIA;DLG4;IL3RA;FGF18;CALM1 |
| 162 | Uptake and function of anthrax toxins Homo sapiens R-HSA-5210891 | 9.43E-02 | 5.78E-01 | 4.43E+00 | 1.05E+01 | FURIN;CALM1 |
| 163 | Gamma-carboxylation, transport, and amino-terminal cleavage of proteins Homo sapiens R-HSA-159854 | 9.43E-02 | 5.78E-01 | 4.43E+00 | 1.05E+01 | FURIN;GAS6 |
| 164 | Import of palmitoyl-CoA into the mitochondrial matrix Homo sapiens R-HSA-200425 | 9.43E-02 | 5.78E-01 | 4.43E+00 | 1.05E+01 | CPT1A;ACACA |
| 165 | CDC6 association with the ORC:origin complex Homo sapiens R-HSA-68689 | 9.43E-02 | 5.78E-01 | 4.43E+00 | 1.05E+01 | MCM8;E2F3 |
| 166 | Signaling by NOTCH4 Homo sapiens R-HSA-1980150 | 9.43E-02 | 5.78E-01 | 4.43E+00 | 1.05E+01 | JAG1;NOTCH4 |
| 167 | Interleukin-7 signaling Homo sapiens R-HSA-1266695 | 9.43E-02 | 5.78E-01 | 4.43E+00 | 1.05E+01 | PIK3R3;IL2RG |
| 168 | CLEC7A (Dectin-1) induces NFAT activation Homo sapiens R-HSA-5607763 | 9.43E-02 | 5.78E-01 | 4.43E+00 | 1.05E+01 | ITPR3;CALM1 |
| 169 | Synthesis of IP2, IP, and Ins in the cytosol Homo sapiens R-HSA-1855183 | 9.43E-02 | 5.78E-01 | 4.43E+00 | 1.05E+01 | INPP4B;INPP1 |

| | | | | | | |
|---|---|---|---|---|---|---|
| 170 | DAP12 interactions Homo sapiens R-HSA-2172127 | 9.50E-02 | 5.79E-01 | 1.37E+00 | 3.23E+00 | KSR1;VWF;DUSP1;RASGRF2;PDGFB;PIK3R3;ITPR3;ARRB1;IL2RG;FGF2;DUSP6;EGFR;MTOR;PSMB9;RASGRP3;RAP1B;PRKAR2B;DLG4;UBC;FGF18;IL3RA;CALM1;SPTBN1 |
| 171 | Cell-Cell communication Homo sapiens R-HSA-1500931 | 9.72E-02 | 5.89E-01 | 1.65E+00 | 3.85E+00 | ITGB1;CLDN5;CDH4;CADM1;FBLIM1;ACTN1;PVR;CLDN1;SPTBN1;FERMT2 |
| 172 | Signaling by FGFR2 Homo sapiens R-HSA-5654738 | 9.94E-02 | 5.98E-01 | 1.36E+00 | 3.15E+00 | KSR1;VWF;DUSP1;RASGRF2;PDGFB;PIK3R3;ITPR3;ARRB1;IL2RG;FGF2;DUSP6;EGFR;MTOR;PSMB9;RASGRP3;RAP1B;PRKAR2B;DLG4;UBC;FGF18;IL3RA;CALM1;SPTBN1 |
| 173 | DARPP-32 events Homo sapiens R-HSA-180024 | 1.05E-01 | 6.14E-01 | 2.85E+00 | 6.44E+00 | PRKAR2B;PDE4D;CALM1 |
| 174 | MAP2K and MAPK activation Homo sapiens R-HSA-5674135 | 1.06E-01 | 6.14E-01 | 2.35E+00 | 5.26E+00 | RAP1B;VWF;KSR1;ARRB1 |
| 175 | Erythrocytes take up carbon dioxide and release oxygen Homo sapiens R-HSA-1237044 | 1.10E-01 | 6.14E-01 | 3.99E+00 | 8.82E+00 | CA2;AQP1 |
| 176 | O2/CO2 exchange in erythrocytes Homo sapiens R-HSA-1480926 | 1.10E-01 | 6.14E-01 | 3.99E+00 | 8.82E+00 | CA2;AQP1 |
| 177 | Reversible hydration of carbon dioxide Homo sapiens R-HSA-1475029 | 1.10E-01 | 6.14E-01 | 3.99E+00 | 8.82E+00 | CA5B;CA2 |
| 178 | Endosomal/Vacuolar pathway Homo sapiens R-HSA-1236977 | 1.10E-01 | 6.14E-01 | 3.99E+00 | 8.82E+00 | LNPEP;HLA-A |
| 179 | FGFR3 mutant receptor activation Homo sapiens R-HSA-2033514 | 1.10E-01 | 6.14E-01 | 3.99E+00 | 8.82E+00 | FGF18;FGF2 |
| 180 | Signaling by activated point mutants of FGFR3 Homo sapiens R-HSA-1839130 | 1.10E-01 | 6.14E-01 | 3.99E+00 | 8.82E+00 | FGF18;FGF2 |
| 181 | ERKs are inactivated Homo sapiens R-HSA-202670 | 1.10E-01 | 6.14E-01 | 3.99E+00 | 8.82E+00 | DUSP3;DUSP6 |

| | | | | | | |
|---|---|---|---|---|---|---|
| 182 | PECAM1 interactions Homo sapiens R-HSA-210990 | 1.10E-01 | 6.14E-01 | 3.99E+00 | 8.82E+00 | PECAM1;ITGAV |
| 183 | Reduction of cytosolic Ca++ levels Homo sapiens R-HSA-418359 | 1.10E-01 | 6.14E-01 | 3.99E+00 | 8.82E+00 | ATP2A3;CALM1 |
| 184 | Caspase-mediated cleavage of cytoskeletal proteins Homo sapiens R-HSA-264870 | 1.10E-01 | 6.14E-01 | 3.99E+00 | 8.82E+00 | GSN;VIM |
| 185 | Vitamin B5 (pantothenate) metabolism Homo sapiens R-HSA-199220 | 1.10E-01 | 6.14E-01 | 3.99E+00 | 8.82E+00 | PANK2;COASY |
| 186 | Collagen formation Homo sapiens R-HSA-1474290 | 1.12E-01 | 6.22E-01 | 1.79E+00 | 3.93E+00 | P4HA1;COL4A4;COL12A1;LOXL4;PLOD2;COL9A2;LOXL2 |
| 187 | Formation of Fibrin Clot (Clotting Cascade) Homo sapiens R-HSA-140877 | 1.14E-01 | 6.33E-01 | 2.28E+00 | 4.95E+00 | VWF;F2R;A2M;SERPINA5 |
| 188 | Glycerophospholipid biosynthesis Homo sapiens R-HSA-1483206 | 1.15E-01 | 6.34E-01 | 1.70E+00 | 3.68E+00 | DMGDH;AGPAT5;ACHE;PLA2G4F;CHDH;LPCAT2;LPIN2;PISD |
| 189 | Ion transport by P-type ATPases Homo sapiens R-HSA-936837 | 1.22E-01 | 6.42E-01 | 2.00E+00 | 4.21E+00 | FXYD4;ATP2A3;ATP1B3;CALM1;ATP9A |
| 190 | Platelet homeostasis Homo sapiens R-HSA-418346 | 1.22E-01 | 6.42E-01 | 1.75E+00 | 3.67E+00 | PDE2A;ATP2A3;PECAM1;ORAI1;KCNMB4;ITPR3;CALM1 |
| 191 | Negative regulation of MAPK pathway Homo sapiens R-HSA-5675221 | 1.22E-01 | 6.42E-01 | 2.22E+00 | 4.66E+00 | KSR1;DUSP1;UBC;DUSP6 |
| 192 | Glycosaminoglycan metabolism Homo sapiens R-HSA-1630316 | 1.25E-01 | 6.42E-01 | 1.61E+00 | 3.34E+00 | CHST6;CHPF2;CHST15;B3GNT3;CHST1;B4GALT6;HS6ST1;HSPG2;GNS |
| 193 | N-glycan antennae elongation in the medial/trans-Golgi Homo sapiens R-HSA-975576 | 1.26E-01 | 6.42E-01 | 2.60E+00 | 5.40E+00 | MAN2A2;FUCA1;B4GALT6 |

| | | | | | | |
|---|---|---|---|---|---|---|
| 194 | WNT ligand biogenesis and trafficking Homo sapiens R-HSA-3238698 | 1.26E-01 | 6.42E-01 | 2.60E+00 | 5.40E+00 | WNT11;WNT7B;TMED5 |
| 195 | Dissolution of Fibrin Clot Homo sapiens R-HSA-75205 | 1.26E-01 | 6.42E-01 | 3.63E+00 | 7.52E+00 | ANXA2;SERPINB6 |
| 196 | WNT5A-dependent internalization of FZD2, FZD5 and ROR2 Homo sapiens R-HSA-5140745 | 1.26E-01 | 6.42E-01 | 3.63E+00 | 7.52E+00 | FZD2;CLTC |
| 197 | POU5F1 (OCT4), SOX2, NANOG activate genes related to proliferation Homo sapiens R-HSA-2892247 | 1.26E-01 | 6.42E-01 | 3.63E+00 | 7.52E+00 | STAT3;FGF2 |
| 198 | NF-kB is activated and signals survival Homo sapiens R-HSA-209560 | 1.26E-01 | 6.42E-01 | 3.63E+00 | 7.52E+00 | NFKBIA;UBC |
| 199 | Assembly Of The HIV Virion Homo sapiens R-HSA-175474 | 1.26E-01 | 6.42E-01 | 3.63E+00 | 7.52E+00 | UBC;FURIN |
| 200 | FGFRL1 modulation of FGFR1 signaling Homo sapiens R-HSA-5658623 | 1.26E-01 | 6.42E-01 | 3.63E+00 | 7.52E+00 | FGF18;FGF2 |
| 201 | FGFR2c ligand binding and activation Homo sapiens R-HSA-190375 | 1.26E-01 | 6.42E-01 | 3.63E+00 | 7.52E+00 | FGF18;FGF2 |
| 202 | Platelet Adhesion to exposed collagen Homo sapiens R-HSA-75892 | 1.26E-01 | 6.42E-01 | 3.63E+00 | 7.52E+00 | ITGB1;VWF |
| 203 | Synthesis of PE Homo sapiens R-HSA-1483213 | 1.26E-01 | 6.42E-01 | 3.63E+00 | 7.52E+00 | LPIN2;PISD |
| 204 | Signaling by Interleukins Homo sapiens R-HSA-449147 | 1.29E-01 | 6.48E-01 | 1.31E+00 | 2.68E+00 | KSR1;VWF;DUSP1;RASGRF2;STAT3;PDGFB;PIK3R3;ARRB1;IL2RG;FGF2;DUSP6;EGFR;IL17RB;PSMB9;RASGRP3;RAP1B;IRAK2;DLG4;IL3RA;UBC;FGF18;STX3;CALM1;SPTBN1 |

| | | | | | | |
|---|---|---|---|---|---|---|
| 205 | ISG15 antiviral mechanism Homo sapiens R-HSA-1169408 | 1.29E-01 | 6.48E-01 | 1.82E+00 | 3.71E+00 | NUP210;DDX58;NEDD4;UBC;ISG15;EIF4G3 |
| 206 | Antiviral mechanism by IFN-stimulated genes Homo sapiens R-HSA-1169410 | 1.29E-01 | 6.48E-01 | 1.82E+00 | 3.71E+00 | NUP210;DDX58;NEDD4;UBC;ISG15;EIF4G3 |
| 207 | Signaling by NOTCH1 Homo sapiens R-HSA-1980143 | 1.29E-01 | 6.48E-01 | 1.82E+00 | 3.71E+00 | JAG1;DNER;UBC;DTX1;ARRB1;HIF1A |
| 208 | Signalling by NGF Homo sapiens R-HSA-166520 | 1.34E-01 | 6.65E-01 | 1.28E+00 | 2.58E+00 | RASGRF2;PDGFB;PIK3R3;ITPR3;ARRB1;FURIN;IL2RG;FGF2;EGFR;RASGRP3;RAP1B;PRKAR2B;UBC;SPTBN1;DUSP3;KSR1;VWF;DUSP1;STAT3;DUSP6;MTOR;PSMB9;NFKBIA;DLG4;IL3RA;FGF18;CALM1 |
| 209 | Degradation of the extracellular matrix Homo sapiens R-HSA-1474228 | 1.35E-01 | 6.67E-01 | 1.63E+00 | 3.27E+00 | CAPN14;COL12A1;HTRA1;FURIN;COL9A2;TIMP1;A2M;HSPG2 |
| 210 | Integrin alphaIIb beta3 signaling Homo sapiens R-HSA-354192 | 1.37E-01 | 6.67E-01 | 2.49E+00 | 4.97E+00 | RAP1B;VWF;RAPGEF3 |
| 211 | CREB phosphorylation through the activation of Ras Homo sapiens R-HSA-442742 | 1.37E-01 | 6.67E-01 | 2.49E+00 | 4.97E+00 | DLG4;RASGRF2;CALM1 |
| 212 | Pyruvate metabolism Homo sapiens R-HSA-70268 | 1.37E-01 | 6.67E-01 | 2.49E+00 | 4.97E+00 | LDHA;PDK3;HAGH |
| 213 | Retinoid metabolism and transport Homo sapiens R-HSA-975634 | 1.40E-01 | 6.79E-01 | 2.10E+00 | 4.14E+00 | RDH11;AKR1C1;AKR1C3;HSPG2 |
| 214 | LGI-ADAM interactions Homo sapiens R-HSA-5682910 | 1.42E-01 | 6.84E-01 | 3.32E+00 | 6.49E+00 | DLG4;LGI2 |
| 215 | FGFR4 ligand binding and activation Homo sapiens R-HSA-190322 | 1.42E-01 | 6.84E-01 | 3.32E+00 | 6.49E+00 | FGF18;FGF2 |
| 216 | G alpha (s) signalling events Homo sapiens R-HSA-418555 | 1.43E-01 | 6.84E-01 | 1.51E+00 | 2.95E+00 | CALCA;RAMP2;CALCRL;CALCB;GIPR;PDE4D;PDE2A;GLP2R;GPR83;ARRB1 |

| | | | | | | |
|---|---|---|---|---|---|---|
| 217 | The role of Nef in HIV-1 replication and disease pathogenesis Homo sapiens R-HSA-164952 | 1.48E-01 | 7.06E-01 | 2.39E+00 | 4.58E+00 | AP1S2;HLA-A;PAK2 |
| 218 | Phospholipid metabolism Homo sapiens R-HSA-1483257 | 1.49E-01 | 7.07E-01 | 1.47E+00 | 2.79E+00 | DMGDH;AGPAT5;INPP4B;ACHE;PLA2G4F;CHDH;LPCAT2;PIK3R3;LPIN2;PISD;PIK3C2B |
| 219 | Signaling by the B Cell Receptor (BCR) Homo sapiens R-HSA-983705 | 1.50E-01 | 7.07E-01 | 1.38E+00 | 2.62E+00 | PDGFB;PIK3R3;ITPR3;FGF2;EGFR;MALT1;MTOR;PSMB9;RASGRP3;NFKBIA;UBC;FGF18;REL;ORAI1;CALM1 |
| 220 | Association of licensing factors with the pre-replicative complex Homo sapiens R-HSA-69298 | 1.59E-01 | 7.24E-01 | 3.07E+00 | 5.64E+00 | UBC;E2F3 |
| 221 | Constitutive Signaling by NOTCH1 HD Domain Mutants Homo sapiens R-HSA-2691232 | 1.59E-01 | 7.24E-01 | 3.07E+00 | 5.64E+00 | JAG1;UBC |
| 222 | Signaling by NOTCH1 HD Domain Mutants in Cancer Homo sapiens R-HSA-2691230 | 1.59E-01 | 7.24E-01 | 3.07E+00 | 5.64E+00 | JAG1;UBC |
| 223 | GRB2:SOS provides linkage to MAPK signaling for Integrins Homo sapiens R-HSA-354194 | 1.59E-01 | 7.24E-01 | 3.07E+00 | 5.64E+00 | RAP1B;VWF |
| 224 | p130Cas linkage to MAPK signaling for integrins Homo sapiens R-HSA-372708 | 1.59E-01 | 7.24E-01 | 3.07E+00 | 5.64E+00 | RAP1B;VWF |
| 225 | CREB phosphorylation through the activation of CaMKII Homo sapiens R-HSA-442729 | 1.59E-01 | 7.24E-01 | 3.07E+00 | 5.64E+00 | DLG4;CALM1 |
| 226 | Elevation of cytosolic Ca2+ levels Homo sapiens R-HSA-139853 | 1.59E-01 | 7.24E-01 | 3.07E+00 | 5.64E+00 | ORAI1;ITPR3 |

| | | | | | | |
|---|---|---|---|---|---|---|
| 227 | Recycling of bile acids and salts Homo sapiens R-HSA-159418 | 1.59E-01 | 7.24E-01 | 3.07E+00 | 5.64E+00 | SLCO1A2;NR1H4 |
| 228 | Negative regulation of FGFR3 signaling Homo sapiens R-HSA-5654732 | 1.59E-01 | 7.24E-01 | 2.30E+00 | 4.23E+00 | UBC;FGF18;FGF2 |
| 229 | Constitutive Signaling by Aberrant PI3K in Cancer Homo sapiens R-HSA-2219530 | 1.66E-01 | 7.45E-01 | 1.78E+00 | 3.20E+00 | FGF18;PDGFB;PIK3R3;FGF2;EGFR |
| 230 | Cell-cell junction organization Homo sapiens R-HSA-421270 | 1.66E-01 | 7.45E-01 | 1.78E+00 | 3.20E+00 | CLDN5;CDH4;CADM1;PVR;CLDN1 |
| 231 | Ca2+ pathway Homo sapiens R-HSA-4086398 | 1.66E-01 | 7.45E-01 | 1.78E+00 | 3.20E+00 | TCF7L1;WNT11;FZD2;ITPR3;CALM1 |
| 232 | Metabolism of vitamins and cofactors Homo sapiens R-HSA-196854 | 1.71E-01 | 7.65E-01 | 1.45E+00 | 2.55E+00 | DHFR;PANK2;RDH11;AKR1C1;QPRT;SLC2A1;AKR1C3;HSPG2;ACACA;COASY |
| 233 | PPARA activates gene expression Homo sapiens R-HSA-1989781 | 1.73E-01 | 7.70E-01 | 1.52E+00 | 2.67E+00 | CPT1A;CYP1A1;FHL2;NR1H4;PPARG;HMGCS2;ANGPTL4;NPAS2 |
| 234 | Glucose metabolism Homo sapiens R-HSA-70326 | 1.76E-01 | 7.71E-01 | 1.64E+00 | 2.85E+00 | PFKFB3;UBC;PCK1;CALM1;HK2;PFKP |
| 235 | p75NTR signals via NF-kB Homo sapiens R-HSA-193639 | 1.76E-01 | 7.71E-01 | 2.85E+00 | 4.94E+00 | NFKBIA;UBC |
| 236 | Sema3A PAK dependent Axon repulsion Homo sapiens R-HSA-399954 | 1.76E-01 | 7.71E-01 | 2.85E+00 | 4.94E+00 | PAK2;PLXNA4 |
| 237 | RHO GTPases activate CIT Homo sapiens R-HSA-5625900 | 1.76E-01 | 7.71E-01 | 2.85E+00 | 4.94E+00 | DLG4;MYH9 |
| 238 | Collagen biosynthesis and modifying enzymes Homo sapiens R-HSA-1650814 | 1.82E-01 | 7.81E-01 | 1.72E+00 | 2.93E+00 | P4HA1;COL4A4;COL12A1;PLOD2;COL9A2 |

| | | | | | | |
|---|---|---|---|---|---|---|
| 239 | Circadian Clock Homo sapiens R-HSA-400253 | 1.82E-01 | 7.81E-01 | 1.72E+00 | 2.93E+00 | CPT1A;UBC;HIF1A;NPAS2;ARNTL2 |
| 240 | Disassembly of the destruction complex and recruitment of AXIN to the membrane Homo sapiens R-HSA-4641262 | 1.83E-01 | 7.81E-01 | 2.14E+00 | 3.63E+00 | FZD2;CAV1;DVL3 |
| 241 | Negative regulation of FGFR4 signaling Homo sapiens R-HSA-5654733 | 1.83E-01 | 7.81E-01 | 2.14E+00 | 3.63E+00 | UBC;FGF18;FGF2 |
| 242 | FGFR1 mutant receptor activation Homo sapiens R-HSA-1839124 | 1.83E-01 | 7.81E-01 | 2.14E+00 | 3.63E+00 | STAT3;LRRFIP1;FGF2 |
| 243 | Adherens junctions interactions Homo sapiens R-HSA-418990 | 1.83E-01 | 7.81E-01 | 2.14E+00 | 3.63E+00 | CDH4;CADM1;PVR |
| 244 | Phospholipase C-mediated cascade: FGFR1 Homo sapiens R-HSA-5654219 | 1.86E-01 | 7.89E-01 | 1.86E+00 | 3.12E+00 | PRKAR2B;ITPR3;CALM1;FGF2 |
| 245 | Regulation of lipid metabolism by Peroxisome proliferator-activated receptor alpha (PPARalpha) Homo sapiens R-HSA-400206 | 1.91E-01 | 8.07E-01 | 1.48E+00 | 2.45E+00 | CPT1A;CYP1A1;FHL2;NR1H4;PPARG;HMGCS2;ANGPTL4;NPAS2 |
| 246 | Activated point mutants of FGFR2 Homo sapiens R-HSA-2033519 | 1.94E-01 | 8.07E-01 | 2.66E+00 | 4.36E+00 | FGF18;FGF2 |
| 247 | Regulation of beta-cell development Homo sapiens R-HSA-186712 | 1.95E-01 | 8.07E-01 | 2.06E+00 | 3.37E+00 | MAFA;FOXA3;FOXA2 |
| 248 | Glycolysis Homo sapiens R-HSA-70171 | 1.95E-01 | 8.07E-01 | 2.06E+00 | 3.37E+00 | PFKFB3;HK2;PFKP |
| 249 | DAG and IP3 signaling Homo sapiens R-HSA-1489509 | 1.95E-01 | 8.07E-01 | 2.06E+00 | 3.37E+00 | PRKAR2B;ITPR3;CALM1 |

| | | | | | | |
|---|---|---|---|---|---|---|
| 250 | TGF-beta receptor signaling activates SMADs Homo sapiens R-HSA-2173789 | 1.95E-01 | 8.07E-01 | 2.06E+00 | 3.37E+00 | UBC;PMEPA1;FURIN |
| 251 | Hexose transport Homo sapiens R-HSA-189200 | 1.96E-01 | 8.07E-01 | 1.81E+00 | 2.96E+00 | NUP210;SLC2A1;SLC5A3;HK2 |
| 252 | Toll Like Receptor 10 (TLR10) Cascade Homo sapiens R-HSA-168142 | 1.98E-01 | 8.07E-01 | 1.58E+00 | 2.55E+00 | NFKBIA;DUSP3;IRAK2;UBC;TLR5;DUSP6 |
| 253 | Toll Like Receptor 5 (TLR5) Cascade Homo sapiens R-HSA-168176 | 1.98E-01 | 8.07E-01 | 1.58E+00 | 2.55E+00 | NFKBIA;DUSP3;IRAK2;UBC;TLR5;DUSP6 |
| 254 | MyD88 cascade initiated on plasma membrane Homo sapiens R-HSA-975871 | 1.98E-01 | 8.07E-01 | 1.58E+00 | 2.55E+00 | NFKBIA;DUSP3;IRAK2;UBC;TLR5;DUSP6 |
| 255 | Metabolism Homo sapiens R-HSA-1430728 | 2.04E-01 | 8.10E-01 | 1.10E+00 | 1.75E+00 | DMGDH;GLTP;PANK2;SAT1;HS6ST1;GLS;AQP1;LIPG;NUDT18;PDK3;RPS10;ARSA;CPT1A;NUP210;ACSL6;SLC6A12;HSPG2;PISD;SULT2B1;CH25H;INPP4B;FAR2;PTGES;ABCB1;PFKFB3;NNMT;PIK3R3;ITPR3;CACNA1C;ABHD5;RPS4Y1;SLC5A3;GNS;NPAS2;ACACA;LDHA;PRKAR2B;RDH11;AFMID;UBC;CHST15;PCK1;A2M;IP6K2;GSTM3;CAV1;EPHX1;DHCR24;DHFR;CYP2S1;SLCO1A2;NOSTRIN;CYP1A1;CALM1;ACHE;QPRT;FHL2;SLC2A1;AKR1B1;HK2;PIK3C2B;SMPD3;SPR;CA5B;CA2;TK1;HMGCS2;SULT4A1;ARG2;CHST6;PLA2G4F;CHPF2;AMPD2;COASY;NT5C3A;ITPKB;CHDH;CAT;B3GNT3;CHST1;PPARG;ANGPTL4;HAGH;B4GALT6;RAPGEF3;AGPAT5;MVK;INPP1;PTGS1;UCP2;AKR1C1;AKR1C3;NR1H4;AKR1C2;PSMB9;LPCAT2;CDO1;LPIN2;PFKP |
| 256 | O-linked glycosylation of mucins Homo sapiens R-HSA-913709 | 2.07E-01 | 8.10E-01 | 1.64E+00 | 2.57E+00 | GALNT6;ST6GALNAC2;GALNT3;B3GNT3;B4GALT6 |
| 257 | Activation of NF-kappaB in B cells Homo sapiens R-HSA-1169091 | 2.07E-01 | 8.10E-01 | 1.64E+00 | 2.57E+00 | NFKBIA;UBC;REL;MALT1;PSMB9 |

| | | | | | | |
|---|---|---|---|---|---|---|
| 258 | CD28 co-stimulation Homo sapiens R-HSA-389356 | 2.08E-01 | 8.10E-01 | 2.00E+00 | 3.13E+00 | PIK3R3;PAK2;MTOR |
| 259 | Downstream signaling events of B Cell Receptor (BCR) Homo sapiens R-HSA-1168372 | 2.09E-01 | 8.10E-01 | 1.33E+00 | 2.08E+00 | NFKBIA;UBC;FGF18;PDGFB;REL;PIK3R3;FGF2;EGFR;MALT1;MTOR;RASGRP3;PSMB9 |
| 260 | Activation of SMO Homo sapiens R-HSA-5635838 | 2.12E-01 | 8.10E-01 | 2.49E+00 | 3.87E+00 | BOC;ARRB1 |
| 261 | Oxygen-dependent proline hydroxylation of Hypoxia-inducible Factor Alpha Homo sapiens R-HSA-1234176 | 2.12E-01 | 8.10E-01 | 2.49E+00 | 3.87E+00 | UBC;HIF1A |
| 262 | SHC-mediated cascade:FGFR3 Homo sapiens R-HSA-5654704 | 2.12E-01 | 8.10E-01 | 2.49E+00 | 3.87E+00 | FGF18;FGF2 |
| 263 | Signaling by cytosolic FGFR1 fusion mutants Homo sapiens R-HSA-1839117 | 2.12E-01 | 8.10E-01 | 2.49E+00 | 3.87E+00 | STAT3;LRRFIP1 |
| 264 | Tie2 Signaling Homo sapiens R-HSA-210993 | 2.12E-01 | 8.10E-01 | 2.49E+00 | 3.87E+00 | ANGPT2;GRB14 |
| 265 | cGMP effects Homo sapiens R-HSA-418457 | 2.12E-01 | 8.10E-01 | 2.49E+00 | 3.87E+00 | PDE2A;KCNMB4 |
| 266 | Cytosolic sulfonation of small molecules Homo sapiens R-HSA-156584 | 2.12E-01 | 8.10E-01 | 2.49E+00 | 3.87E+00 | SULT2B1;SULT4A1 |
| 267 | Abacavir transmembrane transport Homo sapiens R-HSA-2161517 | 2.17E-01 | 8.10E-01 | 4.98E+00 | 7.61E+00 | ABCB1 |
| 268 | Abacavir metabolism Homo sapiens R-HSA-2161541 | 2.17E-01 | 8.10E-01 | 4.98E+00 | 7.61E+00 | PCK1 |
| 269 | Na+-dependent glucose transporters Homo sapiens R-HSA-428808 | 2.17E-01 | 8.10E-01 | 4.98E+00 | 7.61E+00 | SLC5A3 |

| | | | | | | |
|---|---|---|---|---|---|---|
| 270 | Negative regulation of TCF-dependent signaling by DVL-interacting proteins Homo sapiens R-HSA-5368598 | 2.17E-01 | 8.10E-01 | 4.98E+00 | 7.61E+00 | DVL3 |
| 271 | G2 Phase Homo sapiens R-HSA-68911 | 2.17E-01 | 8.10E-01 | 4.98E+00 | 7.61E+00 | E2F3 |
| 272 | PTK6 Expression Homo sapiens R-HSA-8849473 | 2.17E-01 | 8.10E-01 | 4.98E+00 | 7.61E+00 | HIF1A |
| 273 | Activation of RAS in B cells Homo sapiens R-HSA-1169092 | 2.17E-01 | 8.10E-01 | 4.98E+00 | 7.61E+00 | RASGRP3 |
| 274 | LRR FLII-interacting protein 1 (LRRFIP1) activates type I IFN production Homo sapiens R-HSA-3134973 | 2.17E-01 | 8.10E-01 | 4.98E+00 | 7.61E+00 | LRRFIP1 |
| 275 | Cyclin B2 mediated events Homo sapiens R-HSA-157881 | 2.17E-01 | 8.10E-01 | 4.98E+00 | 7.61E+00 | CDC25B |
| 276 | The fatty acid cycling model Homo sapiens R-HSA-167826 | 2.17E-01 | 8.10E-01 | 4.98E+00 | 7.61E+00 | UCP2 |
| 277 | The proton buffering model Homo sapiens R-HSA-167827 | 2.17E-01 | 8.10E-01 | 4.98E+00 | 7.61E+00 | UCP2 |
| 278 | Mitochondrial Uncoupling Proteins Homo sapiens R-HSA-166187 | 2.17E-01 | 8.10E-01 | 4.98E+00 | 7.61E+00 | UCP2 |
| 279 | Negative regulation of FGFR2 signaling Homo sapiens R-HSA-5654727 | 2.21E-01 | 8.10E-01 | 1.93E+00 | 2.92E+00 | UBC;FGF18;FGF2 |
| 280 | Negative regulators of RIG-I/MDA5 signaling Homo sapiens R-HSA-936440 | 2.21E-01 | 8.10E-01 | 1.93E+00 | 2.92E+00 | DDX58;UBC;ISG15 |
| 281 | PLC-gamma1 signalling Homo sapiens R-HSA-167021 | 2.21E-01 | 8.10E-01 | 1.93E+00 | 2.92E+00 | PRKAR2B;ITPR3;CALM1 |

| | | | | | | |
|---|---|---|---|---|---|---|
| 282 | Synthesis of PIPs at the plasma membrane Homo sapiens R-HSA-1660499 | 2.21E-01 | 8.10E-01 | 1.93E+00 | 2.92E+00 | INPP4B;PIK3R3;PIK3C2B |
| 283 | Metabolism of fat-soluble vitamins Homo sapiens R-HSA-6806667 | 2.26E-01 | 8.20E-01 | 1.70E+00 | 2.53E+00 | RDH11;AKR1C1;AKR1C3;HSPG2 |
| 284 | Apoptotic execution phase Homo sapiens R-HSA-75153 | 2.26E-01 | 8.20E-01 | 1.70E+00 | 2.53E+00 | GSN;VIM;PAK2;HIST1H1C |
| 285 | PI3K/AKT Signaling in Cancer Homo sapiens R-HSA-2219528 | 2.29E-01 | 8.20E-01 | 1.50E+00 | 2.21E+00 | FGF18;PDGFB;PIK3R3;FGF2;EGFR;MTOR |
| 286 | Rho GTPase cycle Homo sapiens R-HSA-194840 | 2.29E-01 | 8.20E-01 | 1.40E+00 | 2.07E+00 | STARD13;ARHGAP31;RASGRF2;DLC1;RHOJ;ARAP1;A2M;SRGAP2 |
| 287 | Constitutive Signaling by Ligand-Responsive EGFR Cancer Variants Homo sapiens R-HSA-1236382 | 2.30E-01 | 8.20E-01 | 2.35E+00 | 3.45E+00 | UBC;EGFR |
| 288 | Signaling by Ligand-Responsive EGFR Variants in Cancer Homo sapiens R-HSA-5637815 | 2.30E-01 | 8.20E-01 | 2.35E+00 | 3.45E+00 | UBC;EGFR |
| 289 | Signaling by EGFR in Cancer Homo sapiens R-HSA-1643713 | 2.30E-01 | 8.20E-01 | 2.35E+00 | 3.45E+00 | UBC;EGFR |
| 290 | Phase 4 - resting membrane potential Homo sapiens R-HSA-5576886 | 2.30E-01 | 8.20E-01 | 2.35E+00 | 3.45E+00 | KCNK15;KCNK3 |
| 291 | Generation of second messenger molecules Homo sapiens R-HSA-202433 | 2.33E-01 | 8.25E-01 | 1.87E+00 | 2.72E+00 | HLA-DPB1;PAK2;HLA-DPA1 |
| 292 | Regulation of TP53 Degradation Homo sapiens R-HSA-6804757 | 2.33E-01 | 8.25E-01 | 1.87E+00 | 2.72E+00 | UBC;SGK1;MTOR |

| | | | | | | |
|---|---|---|---|---|---|---|
| 293 | Post NMDA receptor activation events Homo sapiens R-HSA-438064 | 2.33E-01 | 8.25E-01 | 1.87E+00 | 2.72E+00 | DLG4;RASGRF2;CALM1 |
| 294 | trans-Golgi Network Vesicle Budding Homo sapiens R-HSA-199992 | 2.43E-01 | 8.49E-01 | 1.54E+00 | 2.17E+00 | CLTC;AP1S2;ARRB1;CLINT1;GNS |
| 295 | Clathrin derived vesicle budding Homo sapiens R-HSA-421837 | 2.43E-01 | 8.49E-01 | 1.54E+00 | 2.17E+00 | CLTC;AP1S2;ARRB1;CLINT1;GNS |
| 296 | Regulation of TP53 Expression and Degradation Homo sapiens R-HSA-6806003 | 2.46E-01 | 8.49E-01 | 1.81E+00 | 2.54E+00 | UBC;SGK1;MTOR |
| 297 | Arachidonic acid metabolism Homo sapiens R-HSA-2142753 | 2.47E-01 | 8.49E-01 | 1.63E+00 | 2.28E+00 | AKR1C3;CYP1A1;PTGES;PTGS1 |
| 298 | Chondroitin sulfate biosynthesis Homo sapiens R-HSA-2022870 | 2.48E-01 | 8.49E-01 | 2.22E+00 | 3.09E+00 | CHPF2;CHST15 |
| 299 | FGFR2 ligand binding and activation Homo sapiens R-HSA-190241 | 2.48E-01 | 8.49E-01 | 2.22E+00 | 3.09E+00 | FGF18;FGF2 |
| 300 | Hormone-sensitive lipase (HSL)-mediated triacylglycerol hydrolysis Homo sapiens R-HSA-163560 | 2.48E-01 | 8.49E-01 | 2.22E+00 | 3.09E+00 | CAV1;ABHD5 |
| 301 | Synthesis of PC Homo sapiens R-HSA-1483191 | 2.48E-01 | 8.49E-01 | 2.22E+00 | 3.09E+00 | ACHE;LPIN2 |
| 302 | Xenobiotics Homo sapiens R-HSA-211981 | 2.48E-01 | 8.49E-01 | 2.22E+00 | 3.09E+00 | CYP2S1;CYP1A1 |
| 303 | Lipid digestion, mobilization, and transport Homo sapiens R-HSA-73923 | 2.52E-01 | 8.51E-01 | 1.51E+00 | 2.09E+00 | CAV1;LIPG;ABHD5;A2M;HSPG2 |

| | | | | | | |
|---|---|---|---|---|---|---|
| 304 | Beta-catenin independent WNT signaling Homo sapiens R-HSA-3858494 | 2.52E-01 | 8.51E-01 | 1.33E+00 | 1.84E+00 | TCF7L1;FZD2;WNT11;UBC;CLTC;DVL3;ITPR3;CALM1;PSMB9 |
| 305 | Fructose metabolism Homo sapiens R-HSA-5652084 | 2.55E-01 | 8.51E-01 | 3.99E+00 | 5.45E+00 | AKR1B1 |
| 306 | Pre-NOTCH Processing in the Endoplasmic Reticulum Homo sapiens R-HSA-1912399 | 2.55E-01 | 8.51E-01 | 3.99E+00 | 5.45E+00 | NOTCH4 |
| 307 | FGFR1b ligand binding and activation Homo sapiens R-HSA-190370 | 2.55E-01 | 8.51E-01 | 3.99E+00 | 5.45E+00 | FGF2 |
| 308 | TNFR1-mediated ceramide production Homo sapiens R-HSA-5626978 | 2.55E-01 | 8.51E-01 | 3.99E+00 | 5.45E+00 | SMPD3 |
| 309 | Fibronectin matrix formation Homo sapiens R-HSA-1566977 | 2.55E-01 | 8.51E-01 | 3.99E+00 | 5.45E+00 | ITGB1 |
| 310 | CLEC7A/inflammasome pathway Homo sapiens R-HSA-5660668 | 2.55E-01 | 8.51E-01 | 3.99E+00 | 5.45E+00 | MALT1 |
| 311 | Golgi Associated Vesicle Biogenesis Homo sapiens R-HSA-432722 | 2.57E-01 | 8.51E-01 | 1.60E+00 | 2.17E+00 | CLTC;AP1S2;ARRB1;CLINT1 |
| 312 | Platelet Aggregation (Plug Formation) Homo sapiens R-HSA-76009 | 2.59E-01 | 8.51E-01 | 1.76E+00 | 2.38E+00 | RAP1B;VWF;RAPGEF3 |
| 313 | NCAM1 interactions Homo sapiens R-HSA-419037 | 2.59E-01 | 8.51E-01 | 1.76E+00 | 2.38E+00 | COL4A4;CACNA1C;COL9A2 |
| 314 | Costimulation by the CD28 family Homo sapiens R-HSA-388841 | 2.61E-01 | 8.51E-01 | 1.49E+00 | 2.00E+00 | HLA-DPB1;PIK3R3;PAK2;MTOR;HLA-DPA1 |

| | | | | | | |
|---|---|---|---|---|---|---|
| 315 | TRAF6 Mediated Induction of proinflammatory cytokines Homo sapiens R-HSA-168180 | 2.61E-01 | 8.51E-01 | 1.49E+00 | 2.00E+00 | NFKBIA;DUSP3;IRAK2;UBC;DUSP6 |
| 316 | Nef-mediates down modulation of cell surface receptors by recruiting them to clathrin adapters Homo sapiens R-HSA-164938 | 2.66E-01 | 8.51E-01 | 2.10E+00 | 2.78E+00 | AP1S2;HLA-A |
| 317 | Translocation of ZAP-70 to Immunological synapse Homo sapiens R-HSA-202430 | 2.66E-01 | 8.51E-01 | 2.10E+00 | 2.78E+00 | HLA-DPB1;HLA-DPA1 |
| 318 | NOTCH2 Activation and Transmission of Signal to the Nucleus Homo sapiens R-HSA-2979096 | 2.66E-01 | 8.51E-01 | 2.10E+00 | 2.78E+00 | JAG1;UBC |
| 319 | ERK/MAPK targets Homo sapiens R-HSA-198753 | 2.66E-01 | 8.51E-01 | 2.10E+00 | 2.78E+00 | DUSP3;DUSP6 |
| 320 | Regulation of Insulin-like Growth Factor (IGF) transport and uptake by Insulin-like Growth Factor Binding Proteins (IGFBPs) Homo sapiens R-HSA-381426 | 2.66E-01 | 8.51E-01 | 2.10E+00 | 2.78E+00 | IGFBP5;IGFBP2 |
| 321 | PCP/CE pathway Homo sapiens R-HSA-4086400 | 2.69E-01 | 8.51E-01 | 1.41E+00 | 1.85E+00 | WNT11;FZD2;CLTC;UBC;DVL3;PSMB9 |
| 322 | Signaling by FGFR1 in disease Homo sapiens R-HSA-5655302 | 2.72E-01 | 8.51E-01 | 1.71E+00 | 2.22E+00 | STAT3;LRRFIP1;FGF2 |
| 323 | Common Pathway of Fibrin Clot Formation Homo sapiens R-HSA-140875 | 2.84E-01 | 8.51E-01 | 1.99E+00 | 2.51E+00 | F2R;SERPINA5 |
| 324 | CD28 dependent PI3K/Akt signaling Homo sapiens R-HSA-389357 | 2.84E-01 | 8.51E-01 | 1.99E+00 | 2.51E+00 | PIK3R3;MTOR |

| | | | | | | |
|---|---|---|---|---|---|---|
| 325 | Signaling by FGFR3 point mutants in cancer Homo sapiens R-HSA-8853338 | 2.84E-01 | 8.51E-01 | 1.99E+00 | 2.51E+00 | FGF18;FGF2 |
| 326 | Signaling by FGFR3 in disease Homo sapiens R-HSA-5655332 | 2.84E-01 | 8.51E-01 | 1.99E+00 | 2.51E+00 | FGF18;FGF2 |
| 327 | Nephrin interactions Homo sapiens R-HSA-373753 | 2.84E-01 | 8.51E-01 | 1.99E+00 | 2.51E+00 | ACTN1;SPTBN1 |
| 328 | SALM protein interactions at the synapse Homo sapiens R-HSA-8849932 | 2.84E-01 | 8.51E-01 | 1.99E+00 | 2.51E+00 | RTN3;DLG4 |
| 329 | Collagen degradation Homo sapiens R-HSA-1442490 | 2.86E-01 | 8.51E-01 | 1.66E+00 | 2.08E+00 | COL12A1;FURIN;COL9A2 |
| 330 | Gamma carboxylation, hypusine formation and arylsulfatase activation Homo sapiens R-HSA-163841 | 2.86E-01 | 8.51E-01 | 1.66E+00 | 2.08E+00 | ARSA;FURIN;GAS6 |
| 331 | Activation of NMDA receptor upon glutamate binding and postsynaptic events Homo sapiens R-HSA-442755 | 2.86E-01 | 8.51E-01 | 1.66E+00 | 2.08E+00 | DLG4;RASGRF2;CALM1 |
| 332 | Pre-NOTCH Expression and Processing Homo sapiens R-HSA-1912422 | 2.90E-01 | 8.51E-01 | 1.51E+00 | 1.87E+00 | NOTCH4;ATP2A3;FURIN;E2F3 |
| 333 | Inositol transporters Homo sapiens R-HSA-429593 | 2.90E-01 | 8.51E-01 | 3.32E+00 | 4.11E+00 | SLC5A3 |
| 334 | GLI proteins bind promoters of Hh responsive genes to promote transcription Homo sapiens R-HSA-5635851 | 2.90E-01 | 8.51E-01 | 3.32E+00 | 4.11E+00 | BOC |

| | | | | | | |
|---|---|---|---|---|---|---|
| 335 | Attachment of GPI anchor to uPAR Homo sapiens R-HSA-162791 | 2.90E-01 | 8.51E-01 | 3.32E+00 | 4.11E+00 | PGAP1 |
| 336 | NrCAM interactions Homo sapiens R-HSA-447038 | 2.90E-01 | 8.51E-01 | 3.32E+00 | 4.11E+00 | DLG4 |
| 337 | Signaling by NOTCH1 t(7;9)(NOTCH1:M1580 K2555) Translocation Mutant Homo sapiens R-HSA-2660825 | 2.90E-01 | 8.51E-01 | 3.32E+00 | 4.11E+00 | JAG1 |
| 338 | Constitutive Signaling by NOTCH1 t(7;9)(NOTCH1:M1580 K2555) Translocation Mutant Homo sapiens R-HSA-2660826 | 2.90E-01 | 8.51E-01 | 3.32E+00 | 4.11E+00 | JAG1 |
| 339 | FGFR3b ligand binding and activation Homo sapiens R-HSA-190371 | 2.90E-01 | 8.51E-01 | 3.32E+00 | 4.11E+00 | FGF18 |
| 340 | IkBA variant leads to EDA-ID Homo sapiens R-HSA-5603029 | 2.90E-01 | 8.51E-01 | 3.32E+00 | 4.11E+00 | NFKBIA |
| 341 | Binding of TCF/LEF:CTNNB1 to target gene promoters Homo sapiens R-HSA-4411364 | 2.90E-01 | 8.51E-01 | 3.32E+00 | 4.11E+00 | TCF7L1 |
| 342 | Phase 3 - rapid repolarisation Homo sapiens R-HSA-5576890 | 2.90E-01 | 8.51E-01 | 3.32E+00 | 4.11E+00 | KCNE4 |
| 343 | Defective HLCS causes multiple carboxylase deficiency Homo sapiens R-HSA-3371599 | 2.90E-01 | 8.51E-01 | 3.32E+00 | 4.11E+00 | ACACA |

| | | | | | | |
|---|---|---|---|---|---|---|
| 344 | Defects in biotin (Btn) metabolism Homo sapiens R-HSA-3323169 | 2.90E-01 | 8.51E-01 | 3.32E+00 | 4.11E+00 | ACACA |
| 345 | Phosphate bond hydrolysis by NUDT proteins Homo sapiens R-HSA-2393930 | 2.90E-01 | 8.51E-01 | 3.32E+00 | 4.11E+00 | NUDT18 |
| 346 | Synthesis of Ketone Bodies Homo sapiens R-HSA-77111 | 2.90E-01 | 8.51E-01 | 3.32E+00 | 4.11E+00 | HMGCS2 |
| 347 | RAF-independent MAPK1/3 activation Homo sapiens R-HSA-112409 | 3.02E-01 | 8.51E-01 | 1.90E+00 | 2.28E+00 | DUSP1;DUSP6 |
| 348 | Downregulation of SMAD2/3:SMAD4 transcriptional activity Homo sapiens R-HSA-2173795 | 3.02E-01 | 8.51E-01 | 1.90E+00 | 2.28E+00 | UBC;SKIL |
| 349 | SHC-mediated cascade:FGFR4 Homo sapiens R-HSA-5654719 | 3.02E-01 | 8.51E-01 | 1.90E+00 | 2.28E+00 | FGF18;FGF2 |
| 350 | Cholesterol biosynthesis Homo sapiens R-HSA-191273 | 3.02E-01 | 8.51E-01 | 1.90E+00 | 2.28E+00 | MVK;DHCR24 |
| 351 | Glycosphingolipid metabolism Homo sapiens R-HSA-1660662 | 3.12E-01 | 8.51E-01 | 1.57E+00 | 1.83E+00 | GLTP;SMPD3;ARSA |
| 352 | Glutamate Neurotransmitter Release Cycle Homo sapiens R-HSA-210500 | 3.19E-01 | 8.51E-01 | 1.81E+00 | 2.07E+00 | SLC38A2;GLS |
| 353 | TNFs bind their physiological receptors Homo sapiens R-HSA-5669034 | 3.19E-01 | 8.51E-01 | 1.81E+00 | 2.07E+00 | EDARADD;TNFRSF1B |
| 354 | Phosphorylation of CD3 and TCR zeta chains Homo sapiens R-HSA-202427 | 3.19E-01 | 8.51E-01 | 1.81E+00 | 2.07E+00 | HLA-DPB1;HLA-DPA1 |

| | | | | | | |
|---|---|---|---|---|---|---|
| 355 | Growth hormone receptor signaling Homo sapiens R-HSA-982772 | 3.19E-01 | 8.51E-01 | 1.81E+00 | 2.07E+00 | SOCS2;STAT3 |
| 356 | Diseases associated with the TLR signaling cascade Homo sapiens R-HSA-5602358 | 3.19E-01 | 8.51E-01 | 1.81E+00 | 2.07E+00 | NFKBIA;TLR5 |
| 357 | Diseases of Immune System Homo sapiens R-HSA-5260271 | 3.19E-01 | 8.51E-01 | 1.81E+00 | 2.07E+00 | NFKBIA;TLR5 |
| 358 | TRAF6 mediated NF-kB activation Homo sapiens R-HSA-933542 | 3.19E-01 | 8.51E-01 | 1.81E+00 | 2.07E+00 | NFKBIA;DDX58 |
| 359 | Nuclear Events (kinase and transcription factor activation) Homo sapiens R-HSA-198725 | 3.19E-01 | 8.51E-01 | 1.81E+00 | 2.07E+00 | DUSP3;DUSP6 |
| 360 | Sema4D induced cell migration and growth-cone collapse Homo sapiens R-HSA-416572 | 3.19E-01 | 8.51E-01 | 1.81E+00 | 2.07E+00 | SEMA4D;MYH9 |
| 361 | Pyrimidine metabolism Homo sapiens R-HSA-73848 | 3.19E-01 | 8.51E-01 | 1.81E+00 | 2.07E+00 | NT5C3A;TK1 |
| 362 | MAP kinase activation in TLR cascade Homo sapiens R-HSA-450294 | 3.22E-01 | 8.51E-01 | 1.42E+00 | 1.61E+00 | DUSP3;IRAK2;UBC;DUSP6 |
| 363 | Vitamin C (ascorbate) metabolism Homo sapiens R-HSA-196836 | 3.24E-01 | 8.51E-01 | 2.85E+00 | 3.21E+00 | SLC2A1 |
| 364 | Neurotransmitter Clearance In The Synaptic Cleft Homo sapiens R-HSA-112311 | 3.24E-01 | 8.51E-01 | 2.85E+00 | 3.21E+00 | ACHE |

| | | | | | | |
|---|---|---|---|---|---|---|
| 365 | Organic anion transporters Homo sapiens R-HSA-428643 | 3.24E-01 | 8.51E-01 | 2.85E+00 | 3.21E+00 | SLC5A8 |
| 366 | Insulin-like Growth Factor-2 mRNA Binding Proteins (IGF2BPs/IMPs/VICKZs) bind RNA Homo sapiens R-HSA-428359 | 3.24E-01 | 8.51E-01 | 2.85E+00 | 3.21E+00 | H19 |
| 367 | Release of Hh-Np from the secreting cell Homo sapiens R-HSA-5362798 | 3.24E-01 | 8.51E-01 | 2.85E+00 | 3.21E+00 | NOTUM |
| 368 | Ligand-receptor interactions Homo sapiens R-HSA-5632681 | 3.24E-01 | 8.51E-01 | 2.85E+00 | 3.21E+00 | BOC |
| 369 | WNT mediated activation of DVL Homo sapiens R-HSA-201688 | 3.24E-01 | 8.51E-01 | 2.85E+00 | 3.21E+00 | DVL3 |
| 370 | Synthesis of UDP-N-acetyl-glucosamine Homo sapiens R-HSA-446210 | 3.24E-01 | 8.51E-01 | 2.85E+00 | 3.21E+00 | GNPNAT1 |
| 371 | Nef and signal transduction Homo sapiens R-HSA-164944 | 3.24E-01 | 8.51E-01 | 2.85E+00 | 3.21E+00 | PAK2 |
| 372 | IKBKG deficiency causes anhidrotic ectodermal dysplasia with immunodeficiency (EDA-ID) (via TLR) Homo sapiens R-HSA-5603027 | 3.24E-01 | 8.51E-01 | 2.85E+00 | 3.21E+00 | NFKBIA |
| 373 | Formyl peptide receptors bind formyl peptides and many other ligands Homo sapiens R-HSA-444473 | 3.24E-01 | 8.51E-01 | 2.85E+00 | 3.21E+00 | ANXA1 |

| | | | | | | |
|---|---|---|---|---|---|---|
| 374 | Regulation of cytoskeletal remodeling and cell spreading by IPP complex components Homo sapiens R-HSA-446388 | 3.24E-01 | 8.51E-01 | 2.85E+00 | 3.21E+00 | ACTN1 |
| 375 | Sema4D mediated inhibition of cell attachment and migration Homo sapiens R-HSA-416550 | 3.24E-01 | 8.51E-01 | 2.85E+00 | 3.21E+00 | SEMA4D |
| 376 | Cross-presentation of particulate exogenous antigens (phagosomes) Homo sapiens R-HSA-1236973 | 3.24E-01 | 8.51E-01 | 2.85E+00 | 3.21E+00 | ITGAV |
| 377 | Hydrolysis of LPC Homo sapiens R-HSA-1483115 | 3.24E-01 | 8.51E-01 | 2.85E+00 | 3.21E+00 | PLA2G4F |
| 378 | ChREBP activates metabolic gene expression Homo sapiens R-HSA-163765 | 3.24E-01 | 8.51E-01 | 2.85E+00 | 3.21E+00 | ACACA |
| 379 | Metabolism of ingested SeMet, Sec, MeSec into H2Se Homo sapiens R-HSA-2408508 | 3.24E-01 | 8.51E-01 | 2.85E+00 | 3.21E+00 | NNMT |
| 380 | NR1D1 (REV-ERBA) represses gene expression Homo sapiens R-HSA-1368071 | 3.24E-01 | 8.51E-01 | 2.85E+00 | 3.21E+00 | NPAS2 |
| 381 | Synthesis of epoxy (EET) and dihydroxyeicosatrienoic acids (DHET) Homo sapiens R-HSA-2142670 | 3.24E-01 | 8.51E-01 | 2.85E+00 | 3.21E+00 | CYP1A1 |
| 382 | Ketone body metabolism Homo sapiens R-HSA-74182 | 3.24E-01 | 8.51E-01 | 2.85E+00 | 3.21E+00 | HMGCS2 |

| | | | | | | |
|---|---|---|---|---|---|---|
| 383 | Interleukin-17 signaling Homo sapiens R-HSA-448424 | 3.24E-01 | 8.51E-01 | 2.85E+00 | 3.21E+00 | IL17RB |
| 384 | Glucose transport Homo sapiens R-HSA-70153 | 3.25E-01 | 8.51E-01 | 1.53E+00 | 1.72E+00 | NUP210;SLC2A1;HK2 |
| 385 | Deactivation of the beta-catenin transactivating complex Homo sapiens R-HSA-3769402 | 3.25E-01 | 8.51E-01 | 1.53E+00 | 1.72E+00 | TCF7L1;SOX17;UBC |
| 386 | Netrin-1 signaling Homo sapiens R-HSA-373752 | 3.25E-01 | 8.51E-01 | 1.53E+00 | 1.72E+00 | NTN4;SLIT3;DOCK1 |
| 387 | EPHB-mediated forward signaling Homo sapiens R-HSA-3928662 | 3.25E-01 | 8.51E-01 | 1.53E+00 | 1.72E+00 | EFNB2;ARPC1B;ARPC1A |
| 388 | Glucagon-like Peptide-1 (GLP1) regulates insulin secretion Homo sapiens R-HSA-381676 | 3.25E-01 | 8.51E-01 | 1.53E+00 | 1.72E+00 | PRKAR2B;ITPR3;RAPGEF3 |
| 389 | BMAL1:CLOCK,NPAS2 activates circadian gene expression Homo sapiens R-HSA-1368108 | 3.25E-01 | 8.51E-01 | 1.53E+00 | 1.72E+00 | HIF1A;NPAS2;ARNTL2 |
| 390 | Metabolism of water-soluble vitamins and cofactors Homo sapiens R-HSA-196849 | 3.27E-01 | 8.51E-01 | 1.30E+00 | 1.45E+00 | DHFR;PANK2;QPRT;SLC2A1;ACACA;COASY |
| 391 | TCR signaling Homo sapiens R-HSA-202403 | 3.35E-01 | 8.51E-01 | 1.26E+00 | 1.38E+00 | NFKBIA;UBC;HLA-DPB1;PAK2;MALT1;HLA-DPA1;PSMB9 |
| 392 | CLEC7A (Dectin-1) signaling Homo sapiens R-HSA-5607764 | 3.36E-01 | 8.51E-01 | 1.29E+00 | 1.40E+00 | NFKBIA;UBC;ITPR3;CALM1;MALT1;PSMB9 |
| 393 | Downstream TCR signaling Homo sapiens R-HSA-202424 | 3.36E-01 | 8.51E-01 | 1.29E+00 | 1.40E+00 | NFKBIA;UBC;HLA-DPB1;MALT1;HLA-DPA1;PSMB9 |
| 394 | Potassium Channels Homo sapiens R-HSA-1296071 | 3.36E-01 | 8.51E-01 | 1.29E+00 | 1.40E+00 | HCN4;KCNJ5;KCNJ15;KCNMB4;KCNN3;KCNK3 |

| | | | | | | |
|---|---|---|---|---|---|---|
| 395 | Regulation of insulin secretion Homo sapiens R-HSA-422356 | 3.36E-01 | 8.51E-01 | 1.33E+00 | 1.45E+00 | PRKAR2B;SLC2A1;ITPR3;CACNA1C;RAPGEF3 |
| 396 | ATF4 activates genes Homo sapiens R-HSA-380994 | 3.37E-01 | 8.51E-01 | 1.73E+00 | 1.88E+00 | CCL2;DCP2 |
| 397 | Activation of G protein gated Potassium channels Homo sapiens R-HSA-1296041 | 3.37E-01 | 8.51E-01 | 1.73E+00 | 1.88E+00 | KCNJ5;KCNJ15 |
| 398 | Inhibition of voltage gated Ca2+ channels via Gbeta/gamma subunits Homo sapiens R-HSA-997272 | 3.37E-01 | 8.51E-01 | 1.73E+00 | 1.88E+00 | KCNJ5;KCNJ15 |
| 399 | G protein gated Potassium channels Homo sapiens R-HSA-1296059 | 3.37E-01 | 8.51E-01 | 1.73E+00 | 1.88E+00 | KCNJ5;KCNJ15 |
| 400 | Nitric oxide stimulates guanylate cyclase Homo sapiens R-HSA-392154 | 3.37E-01 | 8.51E-01 | 1.73E+00 | 1.88E+00 | PDE2A;KCNMB4 |
| 401 | Synthesis of IP3 and IP4 in the cytosol Homo sapiens R-HSA-1855204 | 3.37E-01 | 8.51E-01 | 1.73E+00 | 1.88E+00 | ITPKB;CALM1 |
| 402 | TRP channels Homo sapiens R-HSA-3295583 | 3.37E-01 | 8.51E-01 | 1.73E+00 | 1.88E+00 | TRPV4;MCOLN1 |
| 403 | Acyl chain remodelling of PC Homo sapiens R-HSA-1482788 | 3.37E-01 | 8.51E-01 | 1.73E+00 | 1.88E+00 | PLA2G4F;LPCAT2 |
| 404 | Signaling by Wnt Homo sapiens R-HSA-195721 | 3.38E-01 | 8.51E-01 | 1.14E+00 | 1.24E+00 | TCF7L1;FZD2;WNT7B;CAV1;CLTC;ITPR3;PSMB9;WNT11;SOX17;UBC;RSPO3;DVL3;CALM1;LGR5;TMED5;HIST1H2AC |
| 405 | Fatty acid, triacylglycerol, and ketone body metabolism Homo sapiens R-HSA-535734 | 3.44E-01 | 8.51E-01 | 1.17E+00 | 1.25E+00 | AGPAT5;CPT1A;ACSL6;CYP1A1;NR1H4;FHL2;PPARG;ANGPTL4;HMGCS2;LPIN2;ACACA;NPAS2 |

| | | | | | | |
|---|---|---|---|---|---|---|
| 406 | PLC beta mediated events Homo sapiens R-HSA-112043 | 3.52E-01 | 8.51E-01 | 1.46E+00 | 1.52E+00 | PRKAR2B;ITPR3;CALM1 |
| 407 | Phase 0 - rapid depolarisation Homo sapiens R-HSA-5576892 | 3.52E-01 | 8.51E-01 | 1.46E+00 | 1.52E+00 | CACNA1C;CALM1;FGF12 |
| 408 | PD-1 signaling Homo sapiens R-HSA-389948 | 3.55E-01 | 8.51E-01 | 1.66E+00 | 1.72E+00 | HLA-DPB1;HLA-DPA1 |
| 409 | Regulation of Hypoxia-inducible Factor (HIF) by oxygen Homo sapiens R-HSA-1234174 | 3.55E-01 | 8.51E-01 | 1.66E+00 | 1.72E+00 | UBC;HIF1A |
| 410 | Cellular response to hypoxia Homo sapiens R-HSA-2262749 | 3.55E-01 | 8.51E-01 | 1.66E+00 | 1.72E+00 | UBC;HIF1A |
| 411 | SHC-mediated cascade:FGFR2 Homo sapiens R-HSA-5654699 | 3.55E-01 | 8.51E-01 | 1.66E+00 | 1.72E+00 | FGF18;FGF2 |
| 412 | TAK1 activates NFkB by phosphorylation and activation of IKKs complex Homo sapiens R-HSA-445989 | 3.55E-01 | 8.51E-01 | 1.66E+00 | 1.72E+00 | NFKBIA;IRAK2 |
| 413 | CaM pathway Homo sapiens R-HSA-111997 | 3.55E-01 | 8.51E-01 | 1.66E+00 | 1.72E+00 | PRKAR2B;CALM1 |
| 414 | Calmodulin induced events Homo sapiens R-HSA-111933 | 3.55E-01 | 8.51E-01 | 1.66E+00 | 1.72E+00 | PRKAR2B;CALM1 |
| 415 | Phase 2 - plateau phase Homo sapiens R-HSA-5576893 | 3.55E-01 | 8.51E-01 | 1.66E+00 | 1.72E+00 | KCNE4;CACNA1C |
| 416 | Downregulation of TGF-beta receptor signaling Homo sapiens R-HSA-2173788 | 3.55E-01 | 8.51E-01 | 1.66E+00 | 1.72E+00 | UBC;PMEPA1 |
| 417 | Antigen processing-Cross presentation Homo sapiens R-HSA-1236975 | 3.55E-01 | 8.51E-01 | 1.30E+00 | 1.34E+00 | UBC;ITGAV;LNPEP;HLA-A;PSMB9 |

| | | | | | | |
|---|---|---|---|---|---|---|
| 418 | DNA Replication Pre-Initiation Homo sapiens R-HSA-69002 | 3.55E-01 | 8.51E-01 | 1.30E+00 | 1.34E+00 | DBF4;MCM8;UBC;E2F3;PSMB9 |
| 419 | M/G1 Transition Homo sapiens R-HSA-68874 | 3.55E-01 | 8.51E-01 | 1.30E+00 | 1.34E+00 | DBF4;MCM8;UBC;E2F3;PSMB9 |
| 420 | Signaling by FGFR in disease Homo sapiens R-HSA-1226099 | 3.55E-01 | 8.51E-01 | 1.35E+00 | 1.40E+00 | STAT3;FGF18;LRRFIP1;FGF2 |
| 421 | Organic cation transport Homo sapiens R-HSA-549127 | 3.57E-01 | 8.51E-01 | 2.49E+00 | 2.57E+00 | RUNX1 |
| 422 | Transport of nucleotide sugars Homo sapiens R-HSA-727802 | 3.57E-01 | 8.51E-01 | 2.49E+00 | 2.57E+00 | SLC35A3 |
| 423 | Retinoid cycle disease events Homo sapiens R-HSA-2453864 | 3.57E-01 | 8.51E-01 | 2.49E+00 | 2.57E+00 | NAPEPLD |
| 424 | Diseases associated with visual transduction Homo sapiens R-HSA-2474795 | 3.57E-01 | 8.51E-01 | 2.49E+00 | 2.57E+00 | NAPEPLD |
| 425 | Transport of gamma-carboxylated protein precursors from the endoplasmic reticulum to the Golgi apparatus Homo sapiens R-HSA-159763 | 3.57E-01 | 8.51E-01 | 2.49E+00 | 2.57E+00 | GAS6 |
| 426 | Formation of annular gap junctions Homo sapiens R-HSA-196025 | 3.57E-01 | 8.51E-01 | 2.49E+00 | 2.57E+00 | CLTC |
| 427 | A third proteolytic cleavage releases NICD Homo sapiens R-HSA-157212 | 3.57E-01 | 8.51E-01 | 2.49E+00 | 2.57E+00 | NOTCH4 |
| 428 | EGFR Transactivation by Gastrin Homo sapiens R-HSA-2179392 | 3.57E-01 | 8.51E-01 | 2.49E+00 | 2.57E+00 | EGFR |

| | | | | | | |
|---|---|---|---|---|---|---|
| 429 | CHL1 interactions Homo sapiens R-HSA-447041 | 3.57E-01 | 8.51E-01 | 2.49E+00 | 2.57E+00 | ITGB1 |
| 430 | Inactivation of Cdc42 and Rac Homo sapiens R-HSA-428543 | 3.57E-01 | 8.51E-01 | 2.49E+00 | 2.57E+00 | SRGAP2 |
| 431 | Degradation of cysteine and homocysteine Homo sapiens R-HSA-1614558 | 3.57E-01 | 8.51E-01 | 2.49E+00 | 2.57E+00 | CDO1 |
| 432 | Pregnenolone biosynthesis Homo sapiens R-HSA-196108 | 3.57E-01 | 8.51E-01 | 2.49E+00 | 2.57E+00 | AKR1B1 |
| 433 | Synthesis of (16-20)-hydroxyeicosatetraenoic acids (HETE) Homo sapiens R-HSA-2142816 | 3.57E-01 | 8.51E-01 | 2.49E+00 | 2.57E+00 | CYP1A1 |
| 434 | Pyrimidine salvage reactions Homo sapiens R-HSA-73614 | 3.57E-01 | 8.51E-01 | 2.49E+00 | 2.57E+00 | TK1 |
| 435 | Cellular Senescence Homo sapiens R-HSA-2559583 | 3.64E-01 | 8.64E-01 | 1.18E+00 | 1.19E+00 | PHC2;UBC;STAT3;E2F3;ETS1;ETS2;HIST1H2AC;EZH2;HIST1H1C |
| 436 | PI5P, PP2A and IER3 Regulate PI3K/AKT Signaling Homo sapiens R-HSA-6811558 | 3.65E-01 | 8.64E-01 | 1.28E+00 | 1.29E+00 | FGF18;PDGFB;PIK3R3;FGF2;EGFR |
| 437 | TRAF6 mediated induction of NFkB and MAP kinases upon TLR7/8 or 9 activation Homo sapiens R-HSA-975138 | 3.65E-01 | 8.64E-01 | 1.28E+00 | 1.29E+00 | NFKBIA;DUSP3;IRAK2;UBC;DUSP6 |
| 438 | G-protein mediated events Homo sapiens R-HSA-112040 | 3.65E-01 | 8.64E-01 | 1.42E+00 | 1.43E+00 | PRKAR2B;ITPR3;CALM1 |
| 439 | Neuronal System Homo sapiens R-HSA-112316 | 3.67E-01 | 8.67E-01 | 1.12E+00 | 1.12E+00 | HCN4;ACHE;RTN3;KCNJ5;SLC38A1;RASGRF2;KCNJ15;SLC6A12;GLS;GRIP1;DLG4;KCNMB4;KCNN3;CALM1;SLC38A2;KCNK3 |
| 440 | Uptake and actions of bacterial toxins Homo sapiens R-HSA-5339562 | 3.72E-01 | 8.70E-01 | 1.59E+00 | 1.58E+00 | FURIN;CALM1 |

| | | | | | | |
|---|---|---|---|---|---|---|
| 441 | EGFR downregulation Homo sapiens R-HSA-182971 | 3.72E-01 | 8.70E-01 | 1.59E+00 | 1.58E+00 | UBC;EGFR |
| 442 | Sema4D in semaphorin signaling Homo sapiens R-HSA-400685 | 3.72E-01 | 8.70E-01 | 1.59E+00 | 1.58E+00 | SEMA4D;MYH9 |
| 443 | RORA activates gene expression Homo sapiens R-HSA-1368082 | 3.72E-01 | 8.70E-01 | 1.59E+00 | 1.58E+00 | CPT1A;NPAS2 |
| 444 | C-type lectin receptors (CLRs) Homo sapiens R-HSA-5621481 | 3.74E-01 | 8.70E-01 | 1.20E+00 | 1.18E+00 | NFKBIA;UBC;ITPR3;CALM1;PAK2;MALT1;PSMB9 |
| 445 | MyD88 dependent cascade initiated on endosome Homo sapiens R-HSA-975155 | 3.84E-01 | 8.70E-01 | 1.25E+00 | 1.19E+00 | NFKBIA;DUSP3;IRAK2;UBC;DUSP6 |
| 446 | Toll Like Receptor 7/8 (TLR7/8) Cascade Homo sapiens R-HSA-168181 | 3.84E-01 | 8.70E-01 | 1.25E+00 | 1.19E+00 | NFKBIA;DUSP3;IRAK2;UBC;DUSP6 |
| 447 | Creatine metabolism Homo sapiens R-HSA-71288 | 3.87E-01 | 8.70E-01 | 2.21E+00 | 2.10E+00 | SLC6A12 |
| 448 | Gamma-carboxylation of protein precursors Homo sapiens R-HSA-159740 | 3.87E-01 | 8.70E-01 | 2.21E+00 | 2.10E+00 | GAS6 |
| 449 | Gap junction degradation Homo sapiens R-HSA-190873 | 3.87E-01 | 8.70E-01 | 2.21E+00 | 2.10E+00 | CLTC |
| 450 | E2F-enabled inhibition of pre-replication complex formation Homo sapiens R-HSA-113507 | 3.87E-01 | 8.70E-01 | 2.21E+00 | 2.10E+00 | MCM8 |
| 451 | Regulation of gene expression by Hypoxia-inducible Factor Homo sapiens R-HSA-1234158 | 3.87E-01 | 8.70E-01 | 2.21E+00 | 2.10E+00 | HIF1A |

| | | | | | | |
|---|---|---|---|---|---|---|
| 452 | Diseases of carbohydrate metabolism Homo sapiens R-HSA-5663084 | 3.87E-01 | 8.70E-01 | 2.21E+00 | 2.10E+00 | UBC |
| 453 | Myoclonic epilepsy of Lafora Homo sapiens R-HSA-3785653 | 3.87E-01 | 8.70E-01 | 2.21E+00 | 2.10E+00 | UBC |
| 454 | Glycogen storage diseases Homo sapiens R-HSA-3229121 | 3.87E-01 | 8.70E-01 | 2.21E+00 | 2.10E+00 | UBC |
| 455 | FGFR2b ligand binding and activation Homo sapiens R-HSA-190377 | 3.87E-01 | 8.70E-01 | 2.21E+00 | 2.10E+00 | FGF2 |
| 456 | GP1b-IX-V activation signalling Homo sapiens R-HSA-430116 | 3.87E-01 | 8.70E-01 | 2.21E+00 | 2.10E+00 | VWF |
| 457 | Purine catabolism Homo sapiens R-HSA-74259 | 3.87E-01 | 8.70E-01 | 2.21E+00 | 2.10E+00 | CAT |
| 458 | Urea cycle Homo sapiens R-HSA-70635 | 3.87E-01 | 8.70E-01 | 2.21E+00 | 2.10E+00 | ARG2 |
| 459 | Cytosolic sensors of pathogen-associated DNA Homo sapiens R-HSA-1834949 | 3.88E-01 | 8.70E-01 | 1.29E+00 | 1.22E+00 | NFKBIA;IFI16;UBC;LRRFIP1 |
| 460 | Triglyceride Biosynthesis Homo sapiens R-HSA-75109 | 3.88E-01 | 8.70E-01 | 1.29E+00 | 1.22E+00 | AGPAT5;ACSL6;LPIN2;ACACA |
| 461 | Gap junction trafficking Homo sapiens R-HSA-190828 | 3.90E-01 | 8.70E-01 | 1.53E+00 | 1.45E+00 | CLTC;GJA4 |
| 462 | PERK regulates gene expression Homo sapiens R-HSA-381042 | 3.90E-01 | 8.70E-01 | 1.53E+00 | 1.45E+00 | CCL2;DCP2 |
| 463 | Energy dependent regulation of mTOR by LKB1-AMPK Homo sapiens R-HSA-380972 | 3.90E-01 | 8.70E-01 | 1.53E+00 | 1.45E+00 | CAB39L;MTOR |

| | | | | | | |
|---|---|---|---|---|---|---|
| 464 | Ca-dependent events Homo sapiens R-HSA-111996 | 3.90E-01 | 8.70E-01 | 1.53E+00 | 1.45E+00 | PRKAR2B;CALM1 |
| 465 | Antigen activates B Cell Receptor (BCR) leading to generation of second messengers Homo sapiens R-HSA-983695 | 3.91E-01 | 8.72E-01 | 1.36E+00 | 1.28E+00 | ORAI1;ITPR3;CALM1 |
| 466 | Signaling by Rho GTPases Homo sapiens R-HSA-194315 | 3.94E-01 | 8.75E-01 | 1.09E+00 | 1.02E+00 | ITGB1;STARD13;ARPC1B;RASGRF2;ARPC1A;ARAP1;PMF1;ARHGAP31;DLG4;DLC1;RHOJ;DVL3;MYH9;PKN1;A2M;CALM1;SRGAP2;PAK2;HIST1H2AC |
| 467 | Assembly of the pre-replicative complex Homo sapiens R-HSA-68867 | 3.99E-01 | 8.81E-01 | 1.27E+00 | 1.16E+00 | MCM8;UBC;E2F3;PSMB9 |
| 468 | Metabolism of nucleotides Homo sapiens R-HSA-15869 | 4.03E-01 | 8.81E-01 | 1.22E+00 | 1.11E+00 | NT5C3A;CAT;NUDT18;AMPD2;TK1 |
| 469 | EPH-ephrin mediated repulsion of cells Homo sapiens R-HSA-3928665 | 4.04E-01 | 8.81E-01 | 1.33E+00 | 1.20E+00 | EFNB2;CLTC;EPHA2 |
| 470 | G beta:gamma signalling through PI3Kgamma Homo sapiens R-HSA-392451 | 4.04E-01 | 8.81E-01 | 1.33E+00 | 1.20E+00 | IL3RA;PIK3R3;IL2RG |
| 471 | Pyruvate metabolism and Citric Acid (TCA) cycle Homo sapiens R-HSA-71406 | 4.04E-01 | 8.81E-01 | 1.33E+00 | 1.20E+00 | LDHA;PDK3;HAGH |
| 472 | Surfactant metabolism Homo sapiens R-HSA-5683826 | 4.07E-01 | 8.81E-01 | 1.48E+00 | 1.33E+00 | CTSH;ZDHHC2 |
| 473 | Interaction between L1 and Ankyrins Homo sapiens R-HSA-445095 | 4.07E-01 | 8.81E-01 | 1.48E+00 | 1.33E+00 | ANK2;SPTBN1 |
| 474 | PIWI-interacting RNA (piRNA) biogenesis Homo sapiens R-HSA-5601884 | 4.07E-01 | 8.81E-01 | 1.48E+00 | 1.33E+00 | TDRD12;TDRD6 |

| | | | | | | |
|---|---|---|---|---|---|---|
| 475 | Myogenesis Homo sapiens R-HSA-525793 | 4.07E-01 | 8.81E-01 | 1.48E+00 | 1.33E+00 | CDH4;BOC |
| 476 | CDO in myogenesis Homo sapiens R-HSA-375170 | 4.07E-01 | 8.81E-01 | 1.48E+00 | 1.33E+00 | CDH4;BOC |
| 477 | Toll Like Receptor 9 (TLR9) Cascade Homo sapiens R-HSA-168138 | 4.12E-01 | 8.81E-01 | 1.20E+00 | 1.06E+00 | NFKBIA;DUSP3;IRAK2;UBC;DUSP6 |
| 478 | Interleukin-6 signaling Homo sapiens R-HSA-1059683 | 4.17E-01 | 8.81E-01 | 1.99E+00 | 1.75E+00 | STAT3 |
| 479 | Signaling by NOTCH3 Homo sapiens R-HSA-1980148 | 4.17E-01 | 8.81E-01 | 1.99E+00 | 1.75E+00 | JAG1 |
| 480 | Membrane binding and targetting of GAG proteins Homo sapiens R-HSA-174490 | 4.17E-01 | 8.81E-01 | 1.99E+00 | 1.75E+00 | UBC |
| 481 | Synthesis And Processing Of GAG, GAGPOL Polyproteins Homo sapiens R-HSA-174495 | 4.17E-01 | 8.81E-01 | 1.99E+00 | 1.75E+00 | UBC |
| 482 | FGFR1c ligand binding and activation Homo sapiens R-HSA-190373 | 4.17E-01 | 8.81E-01 | 1.99E+00 | 1.75E+00 | FGF2 |
| 483 | Signaling by activated point mutants of FGFR1 Homo sapiens R-HSA-1839122 | 4.17E-01 | 8.81E-01 | 1.99E+00 | 1.75E+00 | FGF2 |
| 484 | Condensation of Prometaphase Chromosomes Homo sapiens R-HSA-2514853 | 4.17E-01 | 8.81E-01 | 1.99E+00 | 1.75E+00 | SMC4 |
| 485 | RHO GTPases activate IQGAPs Homo sapiens R-HSA-5626467 | 4.17E-01 | 8.81E-01 | 1.99E+00 | 1.75E+00 | CALM1 |

| | | | | | | |
|---|---|---|---|---|---|---|
| 486 | Sodium/Calcium exchangers Homo sapiens R-HSA-425561 | 4.17E-01 | 8.81E-01 | 1.99E+00 | 1.75E+00 | CALM1 |
| 487 | Biotin transport and metabolism Homo sapiens R-HSA-196780 | 4.17E-01 | 8.81E-01 | 1.99E+00 | 1.75E+00 | ACACA |
| 488 | CYP2E1 reactions Homo sapiens R-HSA-211999 | 4.17E-01 | 8.81E-01 | 1.99E+00 | 1.75E+00 | CYP2S1 |
| 489 | Tryptophan catabolism Homo sapiens R-HSA-71240 | 4.17E-01 | 8.81E-01 | 1.99E+00 | 1.75E+00 | AFMID |
| 490 | Regulation of activated PAK-2p34 by proteasome mediated degradation Homo sapiens R-HSA-211733 | 4.17E-01 | 8.81E-01 | 1.30E+00 | 1.14E+00 | UBC;PAK2;PSMB9 |
| 491 | Host Interactions of HIV factors Homo sapiens R-HSA-162909 | 4.21E-01 | 8.81E-01 | 1.14E+00 | 9.90E-01 | NUP210;UBC;AP1S2;RCC1;HLA-A;PAK2;PSMB9 |
| 492 | Gap junction trafficking and regulation Homo sapiens R-HSA-157858 | 4.24E-01 | 8.81E-01 | 1.42E+00 | 1.22E+00 | CLTC;GJA4 |
| 493 | Termination of translesion DNA synthesis Homo sapiens R-HSA-5656169 | 4.24E-01 | 8.81E-01 | 1.42E+00 | 1.22E+00 | UBC;ISG15 |
| 494 | Activation of the pre-replicative complex Homo sapiens R-HSA-68962 | 4.24E-01 | 8.81E-01 | 1.42E+00 | 1.22E+00 | DBF4;MCM8 |
| 495 | MAPK targets/ Nuclear events mediated by MAP kinases Homo sapiens R-HSA-450282 | 4.24E-01 | 8.81E-01 | 1.42E+00 | 1.22E+00 | DUSP3;DUSP6 |
| 496 | Tight junction interactions Homo sapiens R-HSA-420029 | 4.24E-01 | 8.81E-01 | 1.42E+00 | 1.22E+00 | CLDN5;CLDN1 |

| | | | | | | |
|---|---|---|---|---|---|---|
| 497 | Activation of Kainate Receptors upon glutamate binding Homo sapiens R-HSA-451326 | 4.24E-01 | 8.81E-01 | 1.42E+00 | 1.22E+00 | DLG4;CALM1 |
| 498 | Glucagon-type ligand receptors Homo sapiens R-HSA-420092 | 4.24E-01 | 8.81E-01 | 1.42E+00 | 1.22E+00 | GIPR;GLP2R |
| 499 | Transmission across Chemical Synapses Homo sapiens R-HSA-112315 | 4.28E-01 | 8.81E-01 | 1.10E+00 | 9.31E-01 | ACHE;GRIP1;KCNJ5;SLC38A1;DLG4;RASGRF2;KCNJ15;SLC6A12;CALM1;SLC38A2;GLS |
| 500 | G alpha (q) signalling events Homo sapiens R-HSA-416476 | 4.30E-01 | 8.81E-01 | 1.10E+00 | 9.29E-01 | RGS3;ANXA1;RGS19;GPR68;F2R;BDKRB2;PIK3R3;ITPR3;GPR4;NTS |
| 501 | Chondroitin sulfate/dermatan sulfate metabolism Homo sapiens R-HSA-1793185 | 4.30E-01 | 8.81E-01 | 1.27E+00 | 1.07E+00 | CHPF2;CHST15;HSPG2 |
| 502 | Regulation of Apoptosis Homo sapiens R-HSA-169911 | 4.30E-01 | 8.81E-01 | 1.27E+00 | 1.07E+00 | UBC;PAK2;PSMB9 |
| 503 | O-linked glycosylation Homo sapiens R-HSA-5173105 | 4.30E-01 | 8.81E-01 | 1.15E+00 | 9.70E-01 | SEMA5B;GALNT6;ST6GALNAC2;GALNT3;B3GNT3;B4GALT6 |
| 504 | Integration of energy metabolism Homo sapiens R-HSA-163685 | 4.30E-01 | 8.81E-01 | 1.15E+00 | 9.70E-01 | PRKAR2B;SLC2A1;ITPR3;CACNA1C;ACACA;RAPGEF3 |
| 505 | Negative regulation of the PI3K/AKT network Homo sapiens R-HSA-199418 | 4.31E-01 | 8.81E-01 | 1.17E+00 | 9.86E-01 | FGF18;PDGFB;PIK3R3;FGF2;EGFR |
| 506 | HS-GAG biosynthesis Homo sapiens R-HSA-2022928 | 4.40E-01 | 8.81E-01 | 1.37E+00 | 1.13E+00 | HS6ST1;HSPG2 |
| 507 | Trafficking of AMPA receptors Homo sapiens R-HSA-399719 | 4.40E-01 | 8.81E-01 | 1.37E+00 | 1.13E+00 | GRIP1;DLG4 |

| | | | | | | |
|---|---|---|---|---|---|---|
| 508 | Glutamate Binding, Activation of AMPA Receptors and Synaptic Plasticity Homo sapiens R-HSA-399721 | 4.40E-01 | 8.81E-01 | 1.37E+00 | 1.13E+00 | GRIP1;DLG4 |
| 509 | Inwardly rectifying K+ channels Homo sapiens R-HSA-1296065 | 4.40E-01 | 8.81E-01 | 1.37E+00 | 1.13E+00 | KCNJ5;KCNJ15 |
| 510 | Oxidative Stress Induced Senescence Homo sapiens R-HSA-2559580 | 4.41E-01 | 8.81E-01 | 1.16E+00 | 9.49E-01 | PHC2;UBC;E2F3;HIST1H2AC;EZH2 |
| 511 | Neurotransmitter Release Cycle Homo sapiens R-HSA-112310 | 4.43E-01 | 8.81E-01 | 1.25E+00 | 1.01E+00 | SLC6A12;SLC38A2;GLS |
| 512 | G-protein beta:gamma signalling Homo sapiens R-HSA-397795 | 4.43E-01 | 8.81E-01 | 1.25E+00 | 1.01E+00 | IL3RA;PIK3R3;IL2RG |
| 513 | Passive transport by Aquaporins Homo sapiens R-HSA-432047 | 4.45E-01 | 8.81E-01 | 1.81E+00 | 1.47E+00 | AQP1 |
| 514 | Defective EXT2 causes exostoses 2 Homo sapiens R-HSA-3656237 | 4.45E-01 | 8.81E-01 | 1.81E+00 | 1.47E+00 | HSPG2 |
| 515 | Defective EXT1 causes exostoses 1, TRPS2 and CHDS Homo sapiens R-HSA-3656253 | 4.45E-01 | 8.81E-01 | 1.81E+00 | 1.47E+00 | HSPG2 |
| 516 | Keratan sulfate degradation Homo sapiens R-HSA-2022857 | 4.45E-01 | 8.81E-01 | 1.81E+00 | 1.47E+00 | GNS |
| 517 | CD28 dependent Vav1 pathway Homo sapiens R-HSA-389359 | 4.45E-01 | 8.81E-01 | 1.81E+00 | 1.47E+00 | PAK2 |
| 518 | Unwinding of DNA Homo sapiens R-HSA-176974 | 4.45E-01 | 8.81E-01 | 1.81E+00 | 1.47E+00 | MCM8 |

| | | | | | | |
|---|---|---|---|---|---|---|
| 519 | NF-kB activation through FADD/RIP-1 pathway mediated by caspase-8 and -10 Homo sapiens R-HSA-933543 | 4.45E-01 | 8.81E-01 | 1.81E+00 | 1.47E+00 | DDX58 |
| 520 | Physiological factors Homo sapiens R-HSA-5578768 | 4.45E-01 | 8.81E-01 | 1.81E+00 | 1.47E+00 | NPR1 |
| 521 | Miscellaneous substrates Homo sapiens R-HSA-211958 | 4.45E-01 | 8.81E-01 | 1.81E+00 | 1.47E+00 | CYP2S1 |
| 522 | Pyrimidine catabolism Homo sapiens R-HSA-73621 | 4.45E-01 | 8.81E-01 | 1.81E+00 | 1.47E+00 | NT5C3A |
| 523 | Tandem pore domain potassium channels Homo sapiens R-HSA-1296346 | 4.45E-01 | 8.81E-01 | 1.81E+00 | 1.47E+00 | KCNK3 |
| 524 | MAPK6/MAPK4 signaling Homo sapiens R-HSA-5687128 | 4.50E-01 | 8.82E-01 | 1.15E+00 | 9.14E-01 | CDC42EP3;UBC;CDC42EP2;PAK2;PSMB9 |
| 525 | MyD88:Mal cascade initiated on plasma membrane Homo sapiens R-HSA-166058 | 4.50E-01 | 8.82E-01 | 1.15E+00 | 9.14E-01 | NFKBIA;DUSP3;IRAK2;UBC;DUSP6 |
| 526 | Toll Like Receptor TLR1:TLR2 Cascade Homo sapiens R-HSA-168179 | 4.50E-01 | 8.82E-01 | 1.15E+00 | 9.14E-01 | NFKBIA;DUSP3;IRAK2;UBC;DUSP6 |
| 527 | Toll Like Receptor TLR6:TLR2 Cascade Homo sapiens R-HSA-168188 | 4.50E-01 | 8.82E-01 | 1.15E+00 | 9.14E-01 | NFKBIA;DUSP3;IRAK2;UBC;DUSP6 |
| 528 | Toll Like Receptor 2 (TLR2) Cascade Homo sapiens R-HSA-181438 | 4.50E-01 | 8.82E-01 | 1.15E+00 | 9.14E-01 | NFKBIA;DUSP3;IRAK2;UBC;DUSP6 |
| 529 | Phase 1 - Functionalization of compounds Homo sapiens R-HSA-211945 | 4.50E-01 | 8.82E-01 | 1.15E+00 | 9.14E-01 | CYP2S1;EPHX1;NR1H4;CYP1A1;PTGS1 |

| | | | | | | |
|---|---|---|---|---|---|---|
| 530 | Activation of Matrix Metalloproteinases Homo sapiens R-HSA-1592389 | 4.56E-01 | 8.82E-01 | 1.33E+00 | 1.04E+00 | FURIN;TIMP1 |
| 531 | Nuclear import of Rev protein Homo sapiens R-HSA-180746 | 4.56E-01 | 8.82E-01 | 1.33E+00 | 1.04E+00 | NUP210;RCC1 |
| 532 | Signaling by NOTCH2 Homo sapiens R-HSA-1980145 | 4.56E-01 | 8.82E-01 | 1.33E+00 | 1.04E+00 | JAG1;UBC |
| 533 | Negative regulation of FGFR1 signaling Homo sapiens R-HSA-5654726 | 4.56E-01 | 8.82E-01 | 1.33E+00 | 1.04E+00 | UBC;FGF2 |
| 534 | Signaling by Robo receptor Homo sapiens R-HSA-376176 | 4.56E-01 | 8.82E-01 | 1.33E+00 | 1.04E+00 | SRGAP2;PAK2 |
| 535 | Thrombin signalling through proteinase activated receptors (PARs) Homo sapiens R-HSA-456926 | 4.56E-01 | 8.82E-01 | 1.33E+00 | 1.04E+00 | F2R;ARRB1 |
| 536 | Signaling by TGF-beta Receptor Complex Homo sapiens R-HSA-170834 | 4.64E-01 | 8.82E-01 | 1.16E+00 | 8.88E-01 | UBC;PMEPA1;FURIN;SKIL |
| 537 | Immune System Homo sapiens R-HSA-168256 | 4.66E-01 | 8.82E-01 | 1.02E+00 | 7.75E-01 | ITGB1;IFITM3;IFITM2;RASGRF2;ARPC1B;CLTC;ARPC1A;ARRB1;FGF2;PVR;AP1S2;CTSH;ITGAV;LRRFIP1;CD34;HLA-DPA1;EDARADD;CXADR;DUSP3;KSR1;TNFRSF12A;NUP210;VWF;DUSP1;DDX58;FBXO2;HLA-A;TNFRSF1B;DUSP6;IL17RB;IFI27;IL3RA;ORAI1;MYH9;TLR5;DOCK1;RAPGEF3;LTF;PDGFB;UBR4;PIK3R3;RNF6;ITPR3;IL2RG;EGFR;MALT1;RASGRP3;SOCS2;RAP1B;ZNRF1;IFI16;PRKAR2B;IRAK2;UBC;STX3;GBP2;PAK2;SPTBN1;IP6K2;STAT3;LNPEP;ISG15;MTOR;PSMB9;NFKBIA;DLG4;NEDD4;FGF18;CAPZA2;HLA-DPB1;REL;RNF182;CALM1;EIF4G3;SIPA1 |
| 538 | GPVI-mediated activation cascade Homo sapiens R-HSA-114604 | 4.68E-01 | 8.82E-01 | 1.20E+00 | 9.07E-01 | IL3RA;PIK3R3;IL2RG |

| | | | | | | |
|---|---|---|---|---|---|---|
| 539 | Signal Transduction Homo sapiens R-HSA-162582 | 4.68E-01 | 8.82E-01 | 1.01E+00 | 7.67E-01 | GPR68;FGF2;RGS5;RGS3;SOX17;KDR;PDK3;HIST1H2AC;CPT1A;FST;F2R;HSPG2;DOK1;ZFYVE16;COL4A4;CALCA;CALCB;CALCRL;GIPR;NOTCH4;PDGFB;PIK3R3;DTX1;GPR83;ITPR3;NTS;HIF1A;PMF1;RASGRP3;RAP1B;PRKAR2B;RDH11;RDH10;UBC;DVL3;A2M;SRGAP2;SKIL;SPTBN1;STARD13;JAG1;FZD2;CAV1;ABCA4;PDE2A;FZD9;NFKBIA;ARHGAP31;DLG4;FGF18;NAPEPLD;COL9A2;GRAP;CALM1;ITGB1;FLT1;CAB39L;RASGRF2;LRRK2;ARPC1B;CLTC;ARPC1A;ATP2A3;ARRB1;SHB;CX3CL1;SMPD3;GPER1;BOC;DNER;BDKRB2;PMEPA1;RSPO3;ITGAV;TMED5;DUSP3;ANXA1;KSR1;VWF;DUSP1;PDE4D;GLP2R;ARAP1;SSTR2;DUSP6;ALDH1A3;GAL;CDC42EP3;IL3RA;RHOJ;CDC42EP2;MYH9;DOCK1;RAPGEF3;RAMP2;RGS19;FURIN;IL2RG;EGFR;WNT11;CCL2;E2F3;S1PR3;PAK2;TCF7L1;GALNT3;WNT7B;AKR1C1;STAT3;AKR1C3;GPR4;MTOR;PSMB9;NEDD4;DLC1;RGS12;PKN1;NOTUM;LGR5 |
| 540 | The activation of arylsulfatases Homo sapiens R-HSA-1663150 | 4.71E-01 | 8.82E-01 | 1.66E+00 | 1.25E+00 | ARSA |
| 541 | IRF3-mediated induction of type I IFN Homo sapiens R-HSA-3270619 | 4.71E-01 | 8.82E-01 | 1.66E+00 | 1.25E+00 | IFI16 |
| 542 | Activation of DNA fragmentation factor Homo sapiens R-HSA-211227 | 4.71E-01 | 8.82E-01 | 1.66E+00 | 1.25E+00 | HIST1H1C |
| 543 | Apoptosis induced DNA fragmentation Homo sapiens R-HSA-140342 | 4.71E-01 | 8.82E-01 | 1.66E+00 | 1.25E+00 | HIST1H1C |
| 544 | NICD traffics to nucleus Homo sapiens R-HSA-157052 | 4.71E-01 | 8.82E-01 | 1.66E+00 | 1.25E+00 | NOTCH4 |
| 545 | Notch-HLH transcription pathway Homo sapiens R-HSA-350054 | 4.71E-01 | 8.82E-01 | 1.66E+00 | 1.25E+00 | NOTCH4 |

| | | | | | | |
|---|---|---|---|---|---|---|
| 546 | p75NTR recruits signalling complexes Homo sapiens R-HSA-209543 | 4.71E-01 | 8.82E-01 | 1.66E+00 | 1.25E+00 | UBC |
| 547 | ERBB2 Activates PTK6 Signaling Homo sapiens R-HSA-8847993 | 4.71E-01 | 8.82E-01 | 1.66E+00 | 1.25E+00 | EGFR |
| 548 | Downregulation of ERBB2:ERBB3 signaling Homo sapiens R-HSA-1358803 | 4.71E-01 | 8.82E-01 | 1.66E+00 | 1.25E+00 | UBC |
| 549 | Advanced glycosylation endproduct receptor signaling Homo sapiens R-HSA-879415 | 4.71E-01 | 8.82E-01 | 1.66E+00 | 1.25E+00 | CAPZA2 |
| 550 | Signaling by Activin Homo sapiens R-HSA-1502540 | 4.71E-01 | 8.82E-01 | 1.66E+00 | 1.25E+00 | FST |
| 551 | Purine salvage Homo sapiens R-HSA-74217 | 4.71E-01 | 8.82E-01 | 1.66E+00 | 1.25E+00 | AMPD2 |
| 552 | Rev-mediated nuclear export of HIV RNA Homo sapiens R-HSA-165054 | 4.72E-01 | 8.82E-01 | 1.29E+00 | 9.64E-01 | NUP210;RCC1 |
| 553 | E2F mediated regulation of DNA replication Homo sapiens R-HSA-113510 | 4.72E-01 | 8.82E-01 | 1.29E+00 | 9.64E-01 | DHFR;MCM8 |
| 554 | FGFR2 mutant receptor activation Homo sapiens R-HSA-1839126 | 4.72E-01 | 8.82E-01 | 1.29E+00 | 9.64E-01 | FGF18;FGF2 |
| 555 | Detoxification of Reactive Oxygen Species Homo sapiens R-HSA-3299685 | 4.72E-01 | 8.82E-01 | 1.29E+00 | 9.64E-01 | CAT;SOD3 |
| 556 | GPCR ligand binding Homo sapiens R-HSA-500792 | 4.75E-01 | 8.85E-01 | 1.03E+00 | 7.68E-01 | RAMP2;CALCA;FZD2;ANXA1;CALCRL;CALCB;GPR68;GIPR;WNT7B;F2R;GLP2R;FZD9;GPR4;NTS;SSTR2;CX3CL1;GAL;WNT11;GPER1;BDKRB2;CCL2;S1PR3 |
| 557 | TNFR2 non-canonical NF-kB pathway Homo sapiens R-HSA-5668541 | 4.78E-01 | 8.87E-01 | 1.11E+00 | 8.16E-01 | EDARADD;TNFRSF12A;UBC;TNFRSF1B;PSMB9 |

| | | | | | | |
|---|---|---|---|---|---|---|
| 558 | Assembly of collagen fibrils and other multimeric structures Homo sapiens R-HSA-2022090 | 4.81E-01 | 8.87E-01 | 1.17E+00 | 8.59E-01 | COL4A4;LOXL4;LOXL2 |
| 559 | TCF dependent signaling in response to WNT Homo sapiens R-HSA-201681 | 4.82E-01 | 8.87E-01 | 1.05E+00 | 7.70E-01 | TCF7L1;FZD2;SOX17;CAV1;UBC;RSPO3;DVL3;LGR5;HIST1H2AC;PSMB9 |
| 560 | Regulation of DNA replication Homo sapiens R-HSA-69304 | 4.85E-01 | 8.87E-01 | 1.12E+00 | 8.13E-01 | MCM8;UBC;E2F3;PSMB9 |
| 561 | Adaptive Immune System Homo sapiens R-HSA-1280218 | 4.85E-01 | 8.87E-01 | 1.02E+00 | 7.36E-01 | ITGB1;CLTC;PDGFB;UBR4;PIK3R3;ITPR3;RNF6;FGF2;PVR;EGFR;MALT1;RASGRP3;RAP1B;ZNRF1;UBC;AP1S2;CTSH;ITGAV;CD34;PAK2;HLA-DPA1;CXADR;FBXO2;LNPEP;HLA-A;MTOR;PSMB9;NFKBIA;NEDD4;FGF18;REL;HLA-DPB1;RNF182;ORAI1;CALM1;RAPGEF3;SIPA1 |
| 562 | Activated PKN1 stimulates transcription of AR (androgen receptor) regulated genes KLK2 and KLK3 Homo sapiens R-HSA-5625886 | 4.88E-01 | 8.87E-01 | 1.25E+00 | 8.93E-01 | PKN1;HIST1H2AC |
| 563 | Lipoprotein metabolism Homo sapiens R-HSA-174824 | 4.88E-01 | 8.87E-01 | 1.25E+00 | 8.93E-01 | A2M;HSPG2 |
| 564 | Purine metabolism Homo sapiens R-HSA-73847 | 4.88E-01 | 8.87E-01 | 1.25E+00 | 8.93E-01 | CAT;AMPD2 |
| 565 | Organic cation/anion/zwitterion transport Homo sapiens R-HSA-549132 | 4.96E-01 | 8.87E-01 | 1.53E+00 | 1.07E+00 | RUNX1 |

| | | | | | | |
|---|---|---|---|---|---|---|
| 566 | TP53 regulates transcription of several additional cell death genes whose specific roles in p53-dependent apoptosis remain uncertain Homo sapiens R-HSA-6803205 | 4.96E-01 | 8.87E-01 | 1.53E+00 | 1.07E+00 | BCL6 |
| 567 | TRAF3-dependent IRF activation pathway Homo sapiens R-HSA-918233 | 4.96E-01 | 8.87E-01 | 1.53E+00 | 1.07E+00 | DDX58 |
| 568 | Bile salt and organic anion SLC transporters Homo sapiens R-HSA-425471 | 4.96E-01 | 8.87E-01 | 1.53E+00 | 1.07E+00 | SLC47A2 |
| 569 | DCC mediated attractive signaling Homo sapiens R-HSA-418885 | 4.96E-01 | 8.87E-01 | 1.53E+00 | 1.07E+00 | DOCK1 |
| 570 | SEMA3A-Plexin repulsion signaling by inhibiting Integrin adhesion Homo sapiens R-HSA-399955 | 4.96E-01 | 8.87E-01 | 1.53E+00 | 1.07E+00 | PLXNA4 |
| 571 | Activation of Rac Homo sapiens R-HSA-428540 | 4.96E-01 | 8.87E-01 | 1.53E+00 | 1.07E+00 | PAK2 |
| 572 | Repression of WNT target genes Homo sapiens R-HSA-4641265 | 4.96E-01 | 8.87E-01 | 1.53E+00 | 1.07E+00 | TCF7L1 |
| 573 | Lysosphingolipid and LPA receptors Homo sapiens R-HSA-419408 | 4.96E-01 | 8.87E-01 | 1.53E+00 | 1.07E+00 | S1PR3 |
| 574 | Synthesis of PIPs at the early endosome membrane Homo sapiens R-HSA-1660516 | 4.96E-01 | 8.87E-01 | 1.53E+00 | 1.07E+00 | INPP4B |
| 575 | Methylation Homo sapiens R-HSA-156581 | 4.96E-01 | 8.87E-01 | 1.53E+00 | 1.07E+00 | NNMT |

| | | | | | | |
|---|---|---|---|---|---|---|
| 576 | Nicotinate metabolism Homo sapiens R-HSA-196807 | 4.96E-01 | 8.87E-01 | 1.53E+00 | 1.07E+00 | QPRT |
| 577 | Regulation of pyruvate dehydrogenase (PDH) complex Homo sapiens R-HSA-204174 | 4.96E-01 | 8.87E-01 | 1.53E+00 | 1.07E+00 | PDK3 |
| 578 | MyD88-independent TLR3/TLR4 cascade Homo sapiens R-HSA-166166 | 4.97E-01 | 8.87E-01 | 1.08E+00 | 7.58E-01 | NFKBIA;DUSP3;IRAK2;UBC;DUSP6 |
| 579 | Toll Like Receptor 3 (TLR3) Cascade Homo sapiens R-HSA-168164 | 4.97E-01 | 8.87E-01 | 1.08E+00 | 7.58E-01 | NFKBIA;DUSP3;IRAK2;UBC;DUSP6 |
| 580 | TRIF-mediated TLR3/TLR4 signaling Homo sapiens R-HSA-937061 | 4.97E-01 | 8.87E-01 | 1.08E+00 | 7.58E-01 | NFKBIA;DUSP3;IRAK2;UBC;DUSP6 |
| 581 | Interactions of Rev with host cellular proteins Homo sapiens R-HSA-177243 | 5.04E-01 | 8.90E-01 | 1.21E+00 | 8.28E-01 | NUP210;RCC1 |
| 582 | Fanconi Anemia Pathway Homo sapiens R-HSA-6783310 | 5.04E-01 | 8.90E-01 | 1.21E+00 | 8.28E-01 | UBE2T;UBC |
| 583 | Degradation of DVL Homo sapiens R-HSA-4641258 | 5.05E-01 | 8.90E-01 | 1.13E+00 | 7.71E-01 | UBC;DVL3;PSMB9 |
| 584 | Factors involved in megakaryocyte development and platelet production Homo sapiens R-HSA-983231 | 5.14E-01 | 8.90E-01 | 1.04E+00 | 6.93E-01 | EHD2;PRKAR2B;MAFG;KIF26B;CAPZA2;KIF1C;DOCK1 |
| 585 | Ribosomal scanning and start codon recognition Homo sapiens R-HSA-72702 | 5.17E-01 | 8.90E-01 | 1.11E+00 | 7.30E-01 | EIF5;RPS4Y1;RPS10 |
| 586 | RHO GTPases Activate WASPs and WAVEs Homo sapiens R-HSA-5663213 | 5.19E-01 | 8.90E-01 | 1.17E+00 | 7.69E-01 | ARPC1B;ARPC1A |

| | | | | | | |
|---|---|---|---|---|---|---|
| 587 | Depolymerisation of the Nuclear Lamina Homo sapiens R-HSA-4419969 | 5.20E-01 | 8.90E-01 | 1.42E+00 | 9.29E-01 | LPIN2 |
| 588 | WNT5A-dependent internalization of FZD4 Homo sapiens R-HSA-5099900 | 5.20E-01 | 8.90E-01 | 1.42E+00 | 9.29E-01 | CLTC |
| 589 | N-Glycan antennae elongation Homo sapiens R-HSA-975577 | 5.20E-01 | 8.90E-01 | 1.42E+00 | 9.29E-01 | B4GALT6 |
| 590 | mRNA decay by 5' to 3' exoribonuclease Homo sapiens R-HSA-430039 | 5.20E-01 | 8.90E-01 | 1.42E+00 | 9.29E-01 | DCP2 |
| 591 | Regulation of innate immune responses to cytosolic DNA Homo sapiens R-HSA-3134975 | 5.20E-01 | 8.90E-01 | 1.42E+00 | 9.29E-01 | UBC |
| 592 | NRIF signals cell death from the nucleus Homo sapiens R-HSA-205043 | 5.20E-01 | 8.90E-01 | 1.42E+00 | 9.29E-01 | UBC |
| 593 | Glycogen synthesis Homo sapiens R-HSA-3322077 | 5.20E-01 | 8.90E-01 | 1.42E+00 | 9.29E-01 | UBC |
| 594 | Glycogen breakdown (glycogenolysis) Homo sapiens R-HSA-70221 | 5.20E-01 | 8.90E-01 | 1.42E+00 | 9.29E-01 | CALM1 |
| 595 | FGFR1 ligand binding and activation Homo sapiens R-HSA-190242 | 5.20E-01 | 8.90E-01 | 1.42E+00 | 9.29E-01 | FGF2 |
| 596 | ERBB2 Regulates Cell Motility Homo sapiens R-HSA-6785631 | 5.20E-01 | 8.90E-01 | 1.42E+00 | 9.29E-01 | EGFR |
| 597 | Signaling by EGFRvIII in Cancer Homo sapiens R-HSA-5637812 | 5.20E-01 | 8.90E-01 | 1.42E+00 | 9.29E-01 | EGFR |
| 598 | Constitutive Signaling by EGFRvIII Homo sapiens R-HSA-5637810 | 5.20E-01 | 8.90E-01 | 1.42E+00 | 9.29E-01 | EGFR |

| | | | | | | |
|---|---|---|---|---|---|---|
| 599 | PI3K Cascade Homo sapiens R-HSA-109704 | 5.26E-01 | 8.90E-01 | 1.06E+00 | 6.83E-01 | CAB39L;FGF18;FGF2;MTOR |
| 600 | RIG-I/MDA5 mediated induction of IFN-alpha/beta pathways Homo sapiens R-HSA-168928 | 5.26E-01 | 8.90E-01 | 1.06E+00 | 6.83E-01 | NFKBIA;DDX58;UBC;ISG15 |
| 601 | Transcriptional regulation of white adipocyte differentiation Homo sapiens R-HSA-381340 | 5.26E-01 | 8.90E-01 | 1.06E+00 | 6.83E-01 | CEBPD;PPARG;ANGPTL4;PCK1 |
| 602 | CDT1 association with the CDC6:ORC:origin complex Homo sapiens R-HSA-68827 | 5.29E-01 | 8.90E-01 | 1.09E+00 | 6.93E-01 | MCM8;UBC;PSMB9 |
| 603 | PI-3K cascade:FGFR1 Homo sapiens R-HSA-5654689 | 5.31E-01 | 8.90E-01 | 1.03E+00 | 6.53E-01 | FGF18;PDGFB;PIK3R3;FGF2;EGFR;MTOR |
| 604 | PI-3K cascade:FGFR3 Homo sapiens R-HSA-5654710 | 5.31E-01 | 8.90E-01 | 1.03E+00 | 6.53E-01 | FGF18;PDGFB;PIK3R3;FGF2;EGFR;MTOR |
| 605 | PI3K events in ERBB4 signaling Homo sapiens R-HSA-1250342 | 5.31E-01 | 8.90E-01 | 1.03E+00 | 6.53E-01 | FGF18;PDGFB;PIK3R3;FGF2;EGFR;MTOR |
| 606 | PIP3 activates AKT signaling Homo sapiens R-HSA-1257604 | 5.31E-01 | 8.90E-01 | 1.03E+00 | 6.53E-01 | FGF18;PDGFB;PIK3R3;FGF2;EGFR;MTOR |
| 607 | PI-3K cascade:FGFR4 Homo sapiens R-HSA-5654720 | 5.31E-01 | 8.90E-01 | 1.03E+00 | 6.53E-01 | FGF18;PDGFB;PIK3R3;FGF2;EGFR;MTOR |
| 608 | PI-3K cascade:FGFR2 Homo sapiens R-HSA-5654695 | 5.31E-01 | 8.90E-01 | 1.03E+00 | 6.53E-01 | FGF18;PDGFB;PIK3R3;FGF2;EGFR;MTOR |
| 609 | Translesion synthesis by Y family DNA polymerases bypasses lesions on DNA template Homo sapiens R-HSA-110313 | 5.34E-01 | 8.90E-01 | 1.14E+00 | 7.15E-01 | UBC;ISG15 |

| | | | | | | |
|---|---|---|---|---|---|---|
| 610 | Activation of ATR in response to replication stress Homo sapiens R-HSA-176187 | 5.34E-01 | 8.90E-01 | 1.14E+00 | 7.15E-01 | DBF4;MCM8 |
| 611 | Apoptotic cleavage of cellular proteins Homo sapiens R-HSA-111465 | 5.34E-01 | 8.90E-01 | 1.14E+00 | 7.15E-01 | GSN;VIM |
| 612 | STING mediated induction of host immune responses Homo sapiens R-HSA-1834941 | 5.43E-01 | 8.90E-01 | 1.33E+00 | 8.10E-01 | IFI16 |
| 613 | Translesion synthesis by REV1 Homo sapiens R-HSA-110312 | 5.43E-01 | 8.90E-01 | 1.33E+00 | 8.10E-01 | UBC |
| 614 | TRAF6 mediated induction of TAK1 complex Homo sapiens R-HSA-937072 | 5.43E-01 | 8.90E-01 | 1.33E+00 | 8.10E-01 | UBC |
| 615 | Spry regulation of FGF signaling Homo sapiens R-HSA-1295596 | 5.43E-01 | 8.90E-01 | 1.33E+00 | 8.10E-01 | UBC |
| 616 | PI3K events in ERBB2 signaling Homo sapiens R-HSA-1963642 | 5.43E-01 | 8.90E-01 | 1.33E+00 | 8.10E-01 | EGFR |
| 617 | GRB2 events in ERBB2 signaling Homo sapiens R-HSA-1963640 | 5.43E-01 | 8.90E-01 | 1.33E+00 | 8.10E-01 | EGFR |
| 618 | JNK (c-Jun kinases) phosphorylation and activation mediated by activated human TAK1 Homo sapiens R-HSA-450321 | 5.43E-01 | 8.90E-01 | 1.33E+00 | 8.10E-01 | IRAK2 |
| 619 | MAP3K8 (TPL2)-dependent MAPK1/3 activation Homo sapiens R-HSA-5684264 | 5.43E-01 | 8.90E-01 | 1.33E+00 | 8.10E-01 | UBC |
| 620 | PKA activation Homo sapiens R-HSA-163615 | 5.43E-01 | 8.90E-01 | 1.33E+00 | 8.10E-01 | PRKAR2B |

| | | | | | | |
|---|---|---|---|---|---|---|
| 621 | TGF-beta receptor signaling in EMT (epithelial to mesenchymal transition) Homo sapiens R-HSA-2173791 | 5.43E-01 | 8.90E-01 | 1.33E+00 | 8.10E-01 | UBC |
| 622 | Acyl chain remodelling of PI Homo sapiens R-HSA-1482922 | 5.43E-01 | 8.90E-01 | 1.33E+00 | 8.10E-01 | PLA2G4F |
| 623 | Metabolism of folate and pterines Homo sapiens R-HSA-196757 | 5.43E-01 | 8.90E-01 | 1.33E+00 | 8.10E-01 | DHFR |
| 624 | Opioid Signalling Homo sapiens R-HSA-111885 | 5.46E-01 | 8.90E-01 | 1.03E+00 | 6.27E-01 | PRKAR2B;PDE4D;ITPR3;CALM1 |
| 625 | G1 Phase Homo sapiens R-HSA-69236 | 5.48E-01 | 8.90E-01 | 1.11E+00 | 6.65E-01 | UBC;E2F3 |
| 626 | Cyclin D associated events in G1 Homo sapiens R-HSA-69231 | 5.48E-01 | 8.90E-01 | 1.11E+00 | 6.65E-01 | UBC;E2F3 |
| 627 | MHC class II antigen presentation Homo sapiens R-HSA-2132295 | 5.50E-01 | 8.90E-01 | 1.02E+00 | 6.07E-01 | CLTC;AP1S2;HLA-DPB1;CTSH;HLA-DPA1 |
| 628 | Translocation of GLUT4 to the plasma membrane Homo sapiens R-HSA-1445148 | 5.52E-01 | 8.90E-01 | 1.05E+00 | 6.23E-01 | MYH8;LNPEP;CALM1 |
| 629 | RHO GTPases activate PKNs Homo sapiens R-HSA-5625740 | 5.52E-01 | 8.90E-01 | 1.05E+00 | 6.23E-01 | MYH9;PKN1;HIST1H2AC |
| 630 | PI Metabolism Homo sapiens R-HSA-1483255 | 5.52E-01 | 8.90E-01 | 1.05E+00 | 6.23E-01 | INPP4B;PIK3R3;PIK3C2B |
| 631 | GAB1 signalosome Homo sapiens R-HSA-180292 | 5.55E-01 | 8.90E-01 | 1.00E+00 | 5.92E-01 | FGF18;PDGFB;PIK3R3;FGF2;EGFR;MTOR |
| 632 | PI3K/AKT activation Homo sapiens R-HSA-198203 | 5.55E-01 | 8.90E-01 | 1.00E+00 | 5.92E-01 | FGF18;PDGFB;PIK3R3;FGF2;EGFR;MTOR |
| 633 | mTOR signalling Homo sapiens R-HSA-165159 | 5.62E-01 | 8.90E-01 | 1.08E+00 | 6.20E-01 | CAB39L;MTOR |

| | | | | | | |
|---|---|---|---|---|---|---|
| 634 | NS1 Mediated Effects on Host Pathways Homo sapiens R-HSA-168276 | 5.62E-01 | 8.90E-01 | 1.08E+00 | 6.20E-01 | NUP210;ISG15 |
| 635 | GABA B receptor activation Homo sapiens R-HSA-977444 | 5.62E-01 | 8.90E-01 | 1.08E+00 | 6.20E-01 | KCNJ5;KCNJ15 |
| 636 | Activation of GABAB receptors Homo sapiens R-HSA-991365 | 5.62E-01 | 8.90E-01 | 1.08E+00 | 6.20E-01 | KCNJ5;KCNJ15 |
| 637 | Innate Immune System Homo sapiens R-HSA-168249 | 5.63E-01 | 8.90E-01 | 9.84E-01 | 5.65E-01 | RASGRF2;ARPC1B;ARPC1A;PDGFB;PIK3R3;ARRB1;ITPR3;IL2RG;FGF2;EGFR;MALT1;RASGRP3;RAP1B;IFI16;PRKAR2B;IRAK2;UBC;LRRFIP1;PAK2;SPTBN1;DUSP3;KSR1;VWF;DUSP1;DDX58;ISG15;DUSP6;MTOR;PSMB9;NFKBIA;DLG4;FGF18;IL3RA;CAPZA2;MYH9;CALM1;TLR5;DOCK1 |
| 638 | RHO GTPase Effectors Homo sapiens R-HSA-195258 | 5.64E-01 | 8.90E-01 | 9.84E-01 | 5.63E-01 | ITGB1;DLG4;ARPC1B;ARPC1A;DVL3;MYH9;PKN1;CALM1;SRGAP2;PAK2;PMF1;HIST1H2AC |
| 639 | TNF receptor superfamily (TNFSF) members mediating non-canonical NF-kB pathway Homo sapiens R-HSA-5676594 | 5.65E-01 | 8.90E-01 | 1.25E+00 | 7.10E-01 | TNFRSF12A |
| 640 | Trafficking of GluR2-containing AMPA receptors Homo sapiens R-HSA-416993 | 5.65E-01 | 8.90E-01 | 1.25E+00 | 7.10E-01 | GRIP1 |
| 641 | KSRP (KHSRP) binds and destabilizes mRNA Homo sapiens R-HSA-450604 | 5.65E-01 | 8.90E-01 | 1.25E+00 | 7.10E-01 | DCP2 |
| 642 | Tristetraprolin (TTP, ZFP36) binds and destabilizes mRNA Homo sapiens R-HSA-450513 | 5.65E-01 | 8.90E-01 | 1.25E+00 | 7.10E-01 | DCP2 |
| 643 | Butyrate Response Factor 1 (BRF1) binds and destabilizes mRNA Homo sapiens R-HSA-450385 | 5.65E-01 | 8.90E-01 | 1.25E+00 | 7.10E-01 | DCP2 |

| | | | | | | |
|---|---|---|---|---|---|---|
| 644 | Formation of Senescence-Associated Heterochromatin Foci (SAHF) Homo sapiens R-HSA-2559584 | 5.65E-01 | 8.90E-01 | 1.25E+00 | 7.10E-01 | HIST1H1C |
| 645 | Translesion synthesis by POLK Homo sapiens R-HSA-5655862 | 5.65E-01 | 8.90E-01 | 1.25E+00 | 7.10E-01 | UBC |
| 646 | Translesion synthesis by POLI Homo sapiens R-HSA-5656121 | 5.65E-01 | 8.90E-01 | 1.25E+00 | 7.10E-01 | UBC |
| 647 | G1/S-Specific Transcription Homo sapiens R-HSA-69205 | 5.65E-01 | 8.90E-01 | 1.25E+00 | 7.10E-01 | DHFR |
| 648 | Platelet sensitization by LDL Homo sapiens R-HSA-432142 | 5.65E-01 | 8.90E-01 | 1.25E+00 | 7.10E-01 | PECAM1 |
| 649 | activated TAK1 mediates p38 MAPK activation Homo sapiens R-HSA-450302 | 5.65E-01 | 8.90E-01 | 1.25E+00 | 7.10E-01 | IRAK2 |
| 650 | RHO GTPases Activate ROCKs Homo sapiens R-HSA-5627117 | 5.65E-01 | 8.90E-01 | 1.25E+00 | 7.10E-01 | MYH9 |
| 651 | Synthesis, secretion, and deacylation of Ghrelin Homo sapiens R-HSA-422085 | 5.65E-01 | 8.90E-01 | 1.25E+00 | 7.10E-01 | ACHE |
| 652 | Unblocking of NMDA receptor, glutamate binding and activation Homo sapiens R-HSA-438066 | 5.65E-01 | 8.90E-01 | 1.25E+00 | 7.10E-01 | DLG4 |
| 653 | PKA-mediated phosphorylation of CREB Homo sapiens R-HSA-111931 | 5.65E-01 | 8.90E-01 | 1.25E+00 | 7.10E-01 | PRKAR2B |
| 654 | PKA activation in glucagon signalling Homo sapiens R-HSA-164378 | 5.65E-01 | 8.90E-01 | 1.25E+00 | 7.10E-01 | PRKAR2B |

| | | | | | | |
|---|---|---|---|---|---|---|
| 655 | Chylomicron-mediated lipid transport Homo sapiens R-HSA-174800 | 5.65E-01 | 8.90E-01 | 1.25E+00 | 7.10E-01 | HSPG2 |
| 656 | Digestion of dietary lipid Homo sapiens R-HSA-192456 | 5.65E-01 | 8.90E-01 | 1.25E+00 | 7.10E-01 | LIPG |
| 657 | Acyl chain remodelling of PG Homo sapiens R-HSA-1482925 | 5.65E-01 | 8.90E-01 | 1.25E+00 | 7.10E-01 | PLA2G4F |
| 658 | Acyl chain remodelling of PS Homo sapiens R-HSA-1482801 | 5.65E-01 | 8.90E-01 | 1.25E+00 | 7.10E-01 | PLA2G4F |
| 659 | DNA Replication Homo sapiens R-HSA-69306 | 5.68E-01 | 8.92E-01 | 9.96E-01 | 5.64E-01 | DBF4;MCM8;UBC;E2F3;PSMB9 |
| 660 | Cytochrome P450 - arranged by substrate type Homo sapiens R-HSA-211897 | 5.74E-01 | 9.00E-01 | 1.01E+00 | 5.62E-01 | CYP2S1;NR1H4;CYP1A1 |
| 661 | Metabolism of polyamines Homo sapiens R-HSA-351202 | 5.75E-01 | 9.00E-01 | 9.96E-01 | 5.52E-01 | ARG2;SLC6A12;SAT1;PSMB9 |
| 662 | PKB-mediated events Homo sapiens R-HSA-109703 | 5.76E-01 | 9.00E-01 | 1.05E+00 | 5.78E-01 | CAB39L;MTOR |
| 663 | PRC2 methylates histones and DNA Homo sapiens R-HSA-212300 | 5.76E-01 | 9.00E-01 | 1.05E+00 | 5.78E-01 | HIST1H2AC;EZH2 |
| 664 | Peptide ligand-binding receptors Homo sapiens R-HSA-375276 | 5.78E-01 | 9.02E-01 | 9.74E-01 | 5.34E-01 | ANXA1;GAL;GPER1;F2R;BDKRB2;CCL2;NTS;SSTR2;CX3CL1 |
| 665 | Hedgehog 'on' state Homo sapiens R-HSA-5632684 | 5.84E-01 | 9.05E-01 | 9.84E-01 | 5.29E-01 | BOC;UBC;ARRB1;PSMB9 |
| 666 | Asymmetric localization of PCP proteins Homo sapiens R-HSA-4608870 | 5.85E-01 | 9.05E-01 | 9.96E-01 | 5.34E-01 | FZD2;UBC;PSMB9 |

| | | | | | | |
|---|---|---|---|---|---|---|
| 667 | Miscellaneous transport and binding events Homo sapiens R-HSA-5223345 | 5.86E-01 | 9.05E-01 | 1.17E+00 | 6.26E-01 | NIPAL3 |
| 668 | SHC1 events in ERBB2 signaling Homo sapiens R-HSA-1250196 | 5.86E-01 | 9.05E-01 | 1.17E+00 | 6.26E-01 | EGFR |
| 669 | Activation of IRF3/IRF7 mediated by TBK1/IKK epsilon Homo sapiens R-HSA-936964 | 5.86E-01 | 9.05E-01 | 1.17E+00 | 6.26E-01 | UBC |
| 670 | Regulation of signaling by CBL Homo sapiens R-HSA-912631 | 5.86E-01 | 9.05E-01 | 1.17E+00 | 6.26E-01 | PIK3R3 |
| 671 | Defects in vitamin and cofactor metabolism Homo sapiens R-HSA-3296482 | 5.86E-01 | 9.05E-01 | 1.17E+00 | 6.26E-01 | ACACA |
| 672 | Host Interactions with Influenza Factors Homo sapiens R-HSA-168253 | 5.90E-01 | 9.05E-01 | 1.02E+00 | 5.39E-01 | NUP210;ISG15 |
| 673 | Condensation of Prophase Chromosomes Homo sapiens R-HSA-2299718 | 5.90E-01 | 9.05E-01 | 1.02E+00 | 5.39E-01 | SMC4;HIST1H2AC |
| 674 | Pre-NOTCH Transcription and Translation Homo sapiens R-HSA-1912408 | 5.90E-01 | 9.05E-01 | 1.02E+00 | 5.39E-01 | NOTCH4;E2F3 |
| 675 | TNF signaling Homo sapiens R-HSA-75893 | 5.90E-01 | 9.05E-01 | 1.02E+00 | 5.39E-01 | SMPD3;UBC |
| 676 | Hedgehog ligand biogenesis Homo sapiens R-HSA-5358346 | 5.96E-01 | 9.07E-01 | 9.80E-01 | 5.07E-01 | UBC;NOTUM;PSMB9 |
| 677 | Mitotic G2-G2/M phases Homo sapiens R-HSA-453274 | 6.02E-01 | 9.07E-01 | 9.54E-01 | 4.84E-01 | PRKAR2B;UBC;E2F3;PHLDA1;OPTN;CDK5RAP2;CDC25B;PSMB9 |

| | | | | | | |
|---|---|---|---|---|---|---|
| 678 | Regulation of PLK1 Activity at G2/M Transition Homo sapiens R-HSA-2565942 | 6.03E-01 | 9.07E-01 | 9.60E-01 | 4.86E-01 | PRKAR2B;UBC;OPTN;CDK5RAP2 |
| 679 | Transcriptional regulation of pluripotent stem cells Homo sapiens R-HSA-452723 | 6.03E-01 | 9.07E-01 | 9.96E-01 | 5.04E-01 | STAT3;FGF2 |
| 680 | Fatty Acyl-CoA Biosynthesis Homo sapiens R-HSA-75105 | 6.03E-01 | 9.07E-01 | 9.96E-01 | 5.04E-01 | ACSL6;ACACA |
| 681 | Activation of gene expression by SREBF (SREBP) Homo sapiens R-HSA-2426168 | 6.03E-01 | 9.07E-01 | 9.96E-01 | 5.04E-01 | MVK;ACACA |
| 682 | GABA synthesis, release, reuptake and degradation Homo sapiens R-HSA-888590 | 6.06E-01 | 9.07E-01 | 1.11E+00 | 5.55E-01 | SLC6A12 |
| 683 | Defective B3GALT6 causes EDSP2 and SEMDJL1 Homo sapiens R-HSA-4420332 | 6.06E-01 | 9.07E-01 | 1.11E+00 | 5.55E-01 | HSPG2 |
| 684 | Defective B4GALT7 causes EDS, progeroid type Homo sapiens R-HSA-3560783 | 6.06E-01 | 9.07E-01 | 1.11E+00 | 5.55E-01 | HSPG2 |
| 685 | Defective B3GAT3 causes JDSSDHD Homo sapiens R-HSA-3560801 | 6.06E-01 | 9.07E-01 | 1.11E+00 | 5.55E-01 | HSPG2 |
| 686 | Gap junction assembly Homo sapiens R-HSA-190861 | 6.06E-01 | 9.07E-01 | 1.11E+00 | 5.55E-01 | GJA4 |
| 687 | TP53 Regulates Transcription of Genes Involved in Cytochrome C Release Homo sapiens R-HSA-6803204 | 6.06E-01 | 9.07E-01 | 1.11E+00 | 5.55E-01 | BNIP3L |

| | | | | | | |
|---|---|---|---|---|---|---|
| 688 | Signaling by FGFR2 IIIa TM Homo sapiens R-HSA-8851708 | 6.06E-01 | 9.07E-01 | 1.11E+00 | 5.55E-01 | FGF2 |
| 689 | Regulation of TP53 Activity through Methylation Homo sapiens R-HSA-6804760 | 6.06E-01 | 9.07E-01 | 1.11E+00 | 5.55E-01 | UBC |
| 690 | Translesion Synthesis by POLH Homo sapiens R-HSA-110320 | 6.06E-01 | 9.07E-01 | 1.11E+00 | 5.55E-01 | UBC |
| 691 | HDL-mediated lipid transport Homo sapiens R-HSA-194223 | 6.06E-01 | 9.07E-01 | 1.11E+00 | 5.55E-01 | A2M |
| 692 | Signaling by NODAL Homo sapiens R-HSA-1181150 | 6.06E-01 | 9.07E-01 | 1.11E+00 | 5.55E-01 | FURIN |
| 693 | ER-Phagosome pathway Homo sapiens R-HSA-1236974 | 6.07E-01 | 9.07E-01 | 9.64E-01 | 4.82E-01 | UBC;HLA-A;PSMB9 |
| 694 | Biological oxidations Homo sapiens R-HSA-211859 | 6.15E-01 | 9.14E-01 | 9.43E-01 | 4.58E-01 | SULT2B1;GSTM3;NNMT;CYP2S1;EPHX1;CYP1A1;NR1H4;SULT4A1;PTGS1 |
| 695 | TP53 Regulates Transcription of Cell Death Genes Homo sapiens R-HSA-5633008 | 6.16E-01 | 9.14E-01 | 9.72E-01 | 4.71E-01 | BNIP3L;BCL6 |
| 696 | Signaling by FGFR2 in disease Homo sapiens R-HSA-5655253 | 6.16E-01 | 9.14E-01 | 9.72E-01 | 4.71E-01 | FGF18;FGF2 |
| 697 | Vasopressin regulates renal water homeostasis via Aquaporins Homo sapiens R-HSA-432040 | 6.16E-01 | 9.14E-01 | 9.72E-01 | 4.71E-01 | PRKAR2B;AQP1 |
| 698 | Iron uptake and transport Homo sapiens R-HSA-917937 | 6.16E-01 | 9.14E-01 | 9.72E-01 | 4.71E-01 | LCN2;MCOLN1 |
| 699 | G1/S Transition Homo sapiens R-HSA-69206 | 6.25E-01 | 9.25E-01 | 9.31E-01 | 4.37E-01 | DHFR;DBF4;MCM8;UBC;PSMB9 |

| | | | | | |
|---|---|---|---|---|---|
| 700 | Activated TLR4 signalling Homo sapiens R-HSA-166054 | 6.25E-01 | 9.25E-01 | 9.31E-01 | 4.37E-01 | NFKBIA;DUSP3;IRAK2;UBC;DUSP6 |
| 701 | Kinesins Homo sapiens R-HSA-983189 | 6.28E-01 | 9.26E-01 | 9.48E-01 | 4.41E-01 | KIF26B;KIF1C |
| 702 | Transcriptional activity of SMAD2/SMAD3:SMAD4 heterotrimer Homo sapiens R-HSA-2173793 | 6.28E-01 | 9.26E-01 | 9.48E-01 | 4.41E-01 | UBC;SKIL |
| 703 | Interleukin-1 signaling Homo sapiens R-HSA-446652 | 6.28E-01 | 9.26E-01 | 9.48E-01 | 4.41E-01 | IRAK2;UBC |
| 704 | Membrane Trafficking Homo sapiens R-HSA-199991 | 6.31E-01 | 9.29E-01 | 9.43E-01 | 4.34E-01 | SERPINA1;CLTC;LNPEP;ARRB1;ANK2;GNS;CLINT1;MAN2A2;KIF26B;GJA4;UBC;AP1S2;RAB18;TMED3;MYH8;KIF1C;RAB9A;CALM1;SPTBN1 |
| 705 | Signaling by Hedgehog Homo sapiens R-HSA-5358351 | 6.37E-01 | 9.30E-01 | 9.19E-01 | 4.14E-01 | PRKAR2B;UBC;BOC;ARRB1;NOTUM;PSMB9 |
| 706 | Mitotic G1-G1/S phases Homo sapiens R-HSA-453279 | 6.37E-01 | 9.30E-01 | 9.19E-01 | 4.14E-01 | DHFR;DBF4;MCM8;UBC;E2F3;PSMB9 |
| 707 | ABC-family proteins mediated transport Homo sapiens R-HSA-382556 | 6.41E-01 | 9.30E-01 | 9.26E-01 | 4.12E-01 | ABCB1;ABCA4 |
| 708 | SUMOylation of RNA binding proteins Homo sapiens R-HSA-4570464 | 6.41E-01 | 9.30E-01 | 9.26E-01 | 4.12E-01 | PHC2;NUP210 |
| 709 | Signaling by ERBB2 Homo sapiens R-HSA-1227986 | 6.41E-01 | 9.30E-01 | 9.26E-01 | 4.12E-01 | UBC;EGFR |
| 710 | Diseases of metabolism Homo sapiens R-HSA-5668914 | 6.41E-01 | 9.30E-01 | 9.26E-01 | 4.12E-01 | UBC;ACACA |
| 711 | HS-GAG degradation Homo sapiens R-HSA-2024096 | 6.43E-01 | 9.30E-01 | 9.96E-01 | 4.40E-01 | HSPG2 |

| | | | | | | |
|---|---|---|---|---|---|---|
| 712 | TP53 regulates transcription of additional cell cycle genes whose exact role in the p53 pathway remain uncertain Homo sapiens R-HSA-6804115 | 6.43E-01 | 9.30E-01 | 9.96E-01 | 4.40E-01 | RGCC |
| 713 | SHC-mediated cascade:FGFR1 Homo sapiens R-HSA-5654688 | 6.43E-01 | 9.30E-01 | 9.96E-01 | 4.40E-01 | FGF2 |
| 714 | RIP-mediated NFkB activation via ZBP1 Homo sapiens R-HSA-1810476 | 6.43E-01 | 9.30E-01 | 9.96E-01 | 4.40E-01 | NFKBIA |
| 715 | CD209 (DC-SIGN) signaling Homo sapiens R-HSA-5621575 | 6.43E-01 | 9.30E-01 | 9.96E-01 | 4.40E-01 | PAK2 |
| 716 | Synthesis of very long-chain fatty acyl-CoAs Homo sapiens R-HSA-75876 | 6.43E-01 | 9.30E-01 | 9.96E-01 | 4.40E-01 | ACSL6 |
| 717 | Cyclin A:Cdk2-associated events at S phase entry Homo sapiens R-HSA-69656 | 6.47E-01 | 9.35E-01 | 9.05E-01 | 3.94E-01 | UBC;CDC25B;PSMB9 |
| 718 | DNA Damage Bypass Homo sapiens R-HSA-73893 | 6.53E-01 | 9.42E-01 | 9.05E-01 | 3.86E-01 | UBC;ISG15 |
| 719 | Interferon gamma signaling Homo sapiens R-HSA-877300 | 6.55E-01 | 9.43E-01 | 8.95E-01 | 3.79E-01 | HLA-DPB1;HLA-A;GBP2;HLA-DPA1 |
| 720 | Switching of origins to a post-replicative state Homo sapiens R-HSA-69052 | 6.57E-01 | 9.43E-01 | 8.92E-01 | 3.75E-01 | MCM8;UBC;PSMB9 |
| 721 | Orc1 removal from chromatin Homo sapiens R-HSA-68949 | 6.57E-01 | 9.43E-01 | 8.92E-01 | 3.75E-01 | MCM8;UBC;PSMB9 |
| 722 | mTORC1-mediated signalling Homo sapiens R-HSA-166208 | 6.60E-01 | 9.43E-01 | 9.49E-01 | 3.94E-01 | MTOR |

| | | | | | | |
|---|---|---|---|---|---|---|
| 723 | Pink/Parkin Mediated Mitophagy Homo sapiens R-HSA-5205685 | 6.60E-01 | 9.43E-01 | 9.49E-01 | 3.94E-01 | UBC |
| 724 | Mitophagy Homo sapiens R-HSA-5205647 | 6.60E-01 | 9.43E-01 | 9.49E-01 | 3.94E-01 | UBC |
| 725 | Cyclin A/B1 associated events during G2/M transition Homo sapiens R-HSA-69273 | 6.60E-01 | 9.43E-01 | 9.49E-01 | 3.94E-01 | CDC25B |
| 726 | Transport to the Golgi and subsequent modification Homo sapiens R-HSA-948021 | 6.61E-01 | 9.43E-01 | 8.99E-01 | 3.72E-01 | SERPINA1;MAN2A2;FUCA1;TMED3;ANK2;B4GALT6;SPTBN1 |
| 727 | NOTCH1 Intracellular Domain Regulates Transcription Homo sapiens R-HSA-2122947 | 6.64E-01 | 9.46E-01 | 8.85E-01 | 3.62E-01 | UBC;HIF1A |
| 728 | Toll-Like Receptors Cascades Homo sapiens R-HSA-168898 | 6.65E-01 | 9.46E-01 | 8.91E-01 | 3.64E-01 | NFKBIA;DUSP3;IRAK2;UBC;TLR5;DUSP6 |
| 729 | Removal of licensing factors from origins Homo sapiens R-HSA-69300 | 6.75E-01 | 9.54E-01 | 8.66E-01 | 3.40E-01 | MCM8;UBC;PSMB9 |
| 730 | Death Receptor Signalling Homo sapiens R-HSA-73887 | 6.76E-01 | 9.54E-01 | 8.66E-01 | 3.40E-01 | SMPD3;UBC |
| 731 | Gap-filling DNA repair synthesis and ligation in GG-NER Homo sapiens R-HSA-5696397 | 6.76E-01 | 9.54E-01 | 9.05E-01 | 3.54E-01 | UBC |
| 732 | APC/C:Cdc20 mediated degradation of Cyclin B Homo sapiens R-HSA-174048 | 6.76E-01 | 9.54E-01 | 9.05E-01 | 3.54E-01 | UBC |
| 733 | Signaling by BMP Homo sapiens R-HSA-201451 | 6.76E-01 | 9.54E-01 | 9.05E-01 | 3.54E-01 | ZFYVE16 |

| | | | | | | |
|---|---|---|---|---|---|---|
| 734 | Acyl chain remodelling of PE Homo sapiens R-HSA-1482839 | 6.76E-01 | 9.54E-01 | 9.05E-01 | 3.54E-01 | PLA2G4F |
| 735 | Neurotransmitter Receptor Binding And Downstream Transmission In The Postsynaptic Cell Homo sapiens R-HSA-112314 | 6.78E-01 | 9.56E-01 | 8.78E-01 | 3.41E-01 | GRIP1;KCNJ5;DLG4;RASGRF2;KCNJ15;CALM1 |
| 736 | Fcgamma receptor (FCGR) dependent phagocytosis Homo sapiens R-HSA-2029480 | 6.85E-01 | 9.62E-01 | 8.65E-01 | 3.27E-01 | ARPC1B;ARPC1A;MYH9;ITPR3;DOCK1 |
| 737 | Regulation of actin dynamics for phagocytic cup formation Homo sapiens R-HSA-2029482 | 6.87E-01 | 9.62E-01 | 8.56E-01 | 3.21E-01 | ARPC1B;ARPC1A;MYH9;DOCK1 |
| 738 | IKK complex recruitment mediated by RIP1 Homo sapiens R-HSA-937041 | 6.92E-01 | 9.62E-01 | 8.66E-01 | 3.19E-01 | UBC |
| 739 | Insulin processing Homo sapiens R-HSA-264876 | 6.92E-01 | 9.62E-01 | 8.66E-01 | 3.19E-01 | CPE |
| 740 | Endogenous sterols Homo sapiens R-HSA-211976 | 6.92E-01 | 9.62E-01 | 8.66E-01 | 3.19E-01 | NR1H4 |
| 741 | Sphingolipid metabolism Homo sapiens R-HSA-428157 | 6.93E-01 | 9.62E-01 | 8.41E-01 | 3.08E-01 | GLTP;SMPD3;ARSA |
| 742 | Metabolism of non-coding RNA Homo sapiens R-HSA-194441 | 6.97E-01 | 9.62E-01 | 8.30E-01 | 2.99E-01 | NUP210;PHAX |
| 743 | snRNP Assembly Homo sapiens R-HSA-191859 | 6.97E-01 | 9.62E-01 | 8.30E-01 | 2.99E-01 | NUP210;PHAX |
| 744 | Formation of the ternary complex, and subsequently, the 43S complex Homo sapiens R-HSA-72695 | 6.97E-01 | 9.62E-01 | 8.30E-01 | 2.99E-01 | RPS4Y1;RPS10 |

| | | | | | | |
|---|---|---|---|---|---|---|
| 745 | CDK-mediated phosphorylation and removal of Cdc6 Homo sapiens R-HSA-69017 | 6.97E-01 | 9.62E-01 | 8.30E-01 | 2.99E-01 | UBC;PSMB9 |
| 746 | Ubiquitin-dependent degradation of Cyclin D1 Homo sapiens R-HSA-69229 | 6.97E-01 | 9.62E-01 | 8.30E-01 | 2.99E-01 | UBC;PSMB9 |
| 747 | Ubiquitin-dependent degradation of Cyclin D Homo sapiens R-HSA-75815 | 6.97E-01 | 9.62E-01 | 8.30E-01 | 2.99E-01 | UBC;PSMB9 |
| 748 | Toll Like Receptor 4 (TLR4) Cascade Homo sapiens R-HSA-166016 | 6.99E-01 | 9.62E-01 | 8.51E-01 | 3.05E-01 | NFKBIA;DUSP3;IRAK2;UBC;DUSP6 |
| 749 | Amino acid synthesis and interconversion (transamination) Homo sapiens R-HSA-70614 | 7.06E-01 | 9.62E-01 | 8.30E-01 | 2.89E-01 | GLS |
| 750 | Post-translational modification: synthesis of GPI-anchored proteins Homo sapiens R-HSA-163125 | 7.06E-01 | 9.62E-01 | 8.30E-01 | 2.89E-01 | PGAP1 |
| 751 | Antigen Presentation: Folding, assembly and peptide loading of class I MHC Homo sapiens R-HSA-983170 | 7.06E-01 | 9.62E-01 | 8.30E-01 | 2.89E-01 | HLA-A |
| 752 | Constitutive Signaling by AKT1 E17K in Cancer Homo sapiens R-HSA-5674400 | 7.06E-01 | 9.62E-01 | 8.30E-01 | 2.89E-01 | MTOR |
| 753 | HDMs demethylate histones Homo sapiens R-HSA-3214842 | 7.06E-01 | 9.62E-01 | 8.30E-01 | 2.89E-01 | UTY |
| 754 | Budding and maturation of HIV virion Homo sapiens R-HSA-162588 | 7.06E-01 | 9.62E-01 | 8.30E-01 | 2.89E-01 | UBC |

| | | | | | | |
|---|---|---|---|---|---|---|
| 755 | RAF activation Homo sapiens R-HSA-5673000 | 7.06E-01 | 9.62E-01 | 8.30E-01 | 2.89E-01 | KSR1 |
| 756 | APC-Cdc20 mediated degradation of Nek2A Homo sapiens R-HSA-179409 | 7.06E-01 | 9.62E-01 | 8.30E-01 | 2.89E-01 | UBC |
| 757 | Sulfur amino acid metabolism Homo sapiens R-HSA-1614635 | 7.06E-01 | 9.62E-01 | 8.30E-01 | 2.89E-01 | CDO1 |
| 758 | Nuclear Envelope Breakdown Homo sapiens R-HSA-2980766 | 7.08E-01 | 9.62E-01 | 8.13E-01 | 2.81E-01 | NUP210;LPIN2 |
| 759 | Vpu mediated degradation of CD4 Homo sapiens R-HSA-180534 | 7.08E-01 | 9.62E-01 | 8.13E-01 | 2.81E-01 | UBC;PSMB9 |
| 760 | Autodegradation of the E3 ubiquitin ligase COP1 Homo sapiens R-HSA-349425 | 7.08E-01 | 9.62E-01 | 8.13E-01 | 2.81E-01 | UBC;PSMB9 |
| 761 | Aquaporin-mediated transport Homo sapiens R-HSA-445717 | 7.08E-01 | 9.62E-01 | 8.13E-01 | 2.81E-01 | PRKAR2B;AQP1 |
| 762 | Nuclear Receptor transcription pathway Homo sapiens R-HSA-383280 | 7.08E-01 | 9.62E-01 | 8.13E-01 | 2.81E-01 | NR1H4;PPARG |
| 763 | Phase II conjugation Homo sapiens R-HSA-156580 | 7.10E-01 | 9.62E-01 | 8.29E-01 | 2.84E-01 | SULT2B1;GSTM3;NNMT;SULT4A1 |
| 764 | Gene Silencing by RNA Homo sapiens R-HSA-211000 | 7.17E-01 | 9.62E-01 | 8.21E-01 | 2.73E-01 | NUP210;TDRD12;TDRD6;HIST1H2AC |
| 765 | Deposition of new CENPA-containing nucleosomes at the centromere Homo sapiens R-HSA-606279 | 7.18E-01 | 9.62E-01 | 7.96E-01 | 2.64E-01 | CENPW;HIST1H2AC |
| 766 | Nucleosome assembly Homo sapiens R-HSA-774815 | 7.18E-01 | 9.62E-01 | 7.96E-01 | 2.64E-01 | CENPW;HIST1H2AC |

| | | | | | | |
|---|---|---|---|---|---|---|
| 767 | p53-Independent DNA Damage Response Homo sapiens R-HSA-69610 | 7.18E-01 | 9.62E-01 | 7.96E-01 | 2.64E-01 | UBC;PSMB9 |
| 768 | p53-Independent G1/S DNA damage checkpoint Homo sapiens R-HSA-69613 | 7.18E-01 | 9.62E-01 | 7.96E-01 | 2.64E-01 | UBC;PSMB9 |
| 769 | Ubiquitin Mediated Degradation of Phosphorylated Cdc25A Homo sapiens R-HSA-69601 | 7.18E-01 | 9.62E-01 | 7.96E-01 | 2.64E-01 | UBC;PSMB9 |
| 770 | Senescence-Associated Secretory Phenotype (SASP) Homo sapiens R-HSA-2559582 | 7.19E-01 | 9.62E-01 | 8.07E-01 | 2.66E-01 | UBC;STAT3;HIST1H2AC |
| 771 | Diseases associated with glycosaminoglycan metabolism Homo sapiens R-HSA-3560782 | 7.20E-01 | 9.62E-01 | 7.97E-01 | 2.61E-01 | HSPG2 |
| 772 | A tetrasaccharide linker sequence is required for GAG synthesis Homo sapiens R-HSA-1971475 | 7.20E-01 | 9.62E-01 | 7.97E-01 | 2.61E-01 | HSPG2 |
| 773 | Termination of O-glycan biosynthesis Homo sapiens R-HSA-977068 | 7.20E-01 | 9.62E-01 | 7.97E-01 | 2.61E-01 | ST6GALNAC2 |
| 774 | Recycling pathway of L1 Homo sapiens R-HSA-437239 | 7.20E-01 | 9.62E-01 | 7.97E-01 | 2.61E-01 | DPYSL2 |
| 775 | PTK6 Regulates RHO GTPases, RAS GTPase and MAP kinases Homo sapiens R-HSA-8849471 | 7.20E-01 | 9.62E-01 | 7.97E-01 | 2.61E-01 | DOCK1 |
| 776 | ZBP1(DAI) mediated induction of type I IFNs Homo sapiens R-HSA-1606322 | 7.20E-01 | 9.62E-01 | 7.97E-01 | 2.61E-01 | NFKBIA |
| 777 | G2/M Transition Homo sapiens R-HSA-69275 | 7.26E-01 | 9.66E-01 | 8.39E-01 | 2.69E-01 | PRKAR2B;UBC;PHLDA1;OPTN;CDK5RAP2;CDC25B;PSMB9 |

| | | | | | | |
|---|---|---|---|---|---|---|
| 778 | COPI-mediated anterograde transport Homo sapiens R-HSA-6807878 | 7.27E-01 | 9.66E-01 | 7.96E-01 | 2.54E-01 | TMED3;ANK2;SPTBN1 |
| 779 | Vif-mediated degradation of APOBEC3G Homo sapiens R-HSA-180585 | 7.27E-01 | 9.66E-01 | 7.81E-01 | 2.48E-01 | UBC;PSMB9 |
| 780 | Diseases of signal transduction Homo sapiens R-HSA-5663202 | 7.28E-01 | 9.66E-01 | 8.64E-01 | 2.75E-01 | JAG1;STAT3;UBC;FGF18;PDGFB;NAPEPLD;PIK3R3;LRRFIP1;FGF2;EGFR;MTOR;PSMB9 |
| 781 | Interleukin-6 family signaling Homo sapiens R-HSA-6783589 | 7.34E-01 | 9.69E-01 | 7.66E-01 | 2.37E-01 | STAT3 |
| 782 | Phase 1 - inactivation of fast Na+ channels Homo sapiens R-HSA-5576894 | 7.34E-01 | 9.69E-01 | 7.66E-01 | 2.37E-01 | CACNA1C |
| 783 | Effects of PIP2 hydrolysis Homo sapiens R-HSA-114508 | 7.34E-01 | 9.69E-01 | 7.66E-01 | 2.37E-01 | ITPR3 |
| 784 | Heparan sulfate/heparin (HS-GAG) metabolism Homo sapiens R-HSA-1638091 | 7.37E-01 | 9.69E-01 | 7.66E-01 | 2.34E-01 | HS6ST1;HSPG2 |
| 785 | SCF-beta-TrCP mediated degradation of Emi1 Homo sapiens R-HSA-174113 | 7.37E-01 | 9.69E-01 | 7.66E-01 | 2.34E-01 | UBC;PSMB9 |
| 786 | Stabilization of p53 Homo sapiens R-HSA-69541 | 7.37E-01 | 9.69E-01 | 7.66E-01 | 2.34E-01 | UBC;PSMB9 |
| 787 | AUF1 (hnRNP D0) binds and destabilizes mRNA Homo sapiens R-HSA-450408 | 7.37E-01 | 9.69E-01 | 7.66E-01 | 2.34E-01 | UBC;PSMB9 |
| 788 | Degradation of AXIN Homo sapiens R-HSA-4641257 | 7.37E-01 | 9.69E-01 | 7.66E-01 | 2.34E-01 | UBC;PSMB9 |

| | | | | | | |
|---|---|---|---|---|---|---|
| 789 | Hh mutants that don't undergo autocatalytic processing are degraded by ERAD Homo sapiens R-HSA-5362768 | 7.46E-01 | 9.69E-01 | 7.51E-01 | 2.20E-01 | UBC;PSMB9 |
| 790 | GABA receptor activation Homo sapiens R-HSA-977443 | 7.46E-01 | 9.69E-01 | 7.51E-01 | 2.20E-01 | KCNJ5;KCNJ15 |
| 791 | Regulation of cholesterol biosynthesis by SREBP (SREBF) Homo sapiens R-HSA-1655829 | 7.46E-01 | 9.69E-01 | 7.51E-01 | 2.20E-01 | MVK;ACACA |
| 792 | RNA Polymerase III Transcription Initiation From Type 3 Promoter Homo sapiens R-HSA-76071 | 7.46E-01 | 9.69E-01 | 7.38E-01 | 2.16E-01 | SNAPC1 |
| 793 | Recognition of DNA damage by PCNA-containing replication complex Homo sapiens R-HSA-110314 | 7.46E-01 | 9.69E-01 | 7.38E-01 | 2.16E-01 | UBC |
| 794 | Endosomal Sorting Complex Required For Transport (ESCRT) Homo sapiens R-HSA-917729 | 7.46E-01 | 9.69E-01 | 7.38E-01 | 2.16E-01 | UBC |
| 795 | Adrenaline,noradrenaline inhibits insulin secretion Homo sapiens R-HSA-400042 | 7.46E-01 | 9.69E-01 | 7.38E-01 | 2.16E-01 | CACNA1C |
| 796 | Synthesis of PA Homo sapiens R-HSA-1483166 | 7.46E-01 | 9.69E-01 | 7.38E-01 | 2.16E-01 | AGPAT5 |
| 797 | COPI-dependent Golgi-to-ER retrograde traffic Homo sapiens R-HSA-6811434 | 7.50E-01 | 9.69E-01 | 7.65E-01 | 2.20E-01 | KIF26B;TMED3;KIF1C |

| | | | | | | |
|---|---|---|---|---|---|---|
| 798 | Chemokine receptors bind chemokines Homo sapiens R-HSA-380108 | 7.55E-01 | 9.69E-01 | 7.37E-01 | 2.07E-01 | CCL2;CX3CL1 |
| 799 | Degradation of beta-catenin by the destruction complex Homo sapiens R-HSA-195253 | 7.58E-01 | 9.69E-01 | 7.56E-01 | 2.10E-01 | TCF7L1;UBC;PSMB9 |
| 800 | Mitotic Prophase Homo sapiens R-HSA-68875 | 7.58E-01 | 9.69E-01 | 7.73E-01 | 2.14E-01 | NUP210;SMC4;LPIN2;HIST1H2AC |
| 801 | COPI-independent Golgi-to-ER retrograde traffic Homo sapiens R-HSA-6811436 | 7.59E-01 | 9.69E-01 | 7.11E-01 | 1.96E-01 | RAB18 |
| 802 | Transferrin endocytosis and recycling Homo sapiens R-HSA-917977 | 7.59E-01 | 9.69E-01 | 7.11E-01 | 1.96E-01 | MCOLN1 |
| 803 | Translation initiation complex formation Homo sapiens R-HSA-72649 | 7.64E-01 | 9.69E-01 | 7.24E-01 | 1.95E-01 | RPS4Y1;RPS10 |
| 804 | Meiotic synapsis Homo sapiens R-HSA-1221632 | 7.64E-01 | 9.69E-01 | 7.24E-01 | 1.95E-01 | SYNE2;HIST1H2AC |
| 805 | Constitutive Signaling by NOTCH1 HD+PEST Domain Mutants Homo sapiens R-HSA-2894862 | 7.64E-01 | 9.69E-01 | 7.24E-01 | 1.95E-01 | JAG1;UBC |
| 806 | Signaling by NOTCH1 in Cancer Homo sapiens R-HSA-2644603 | 7.64E-01 | 9.69E-01 | 7.24E-01 | 1.95E-01 | JAG1;UBC |
| 807 | Signaling by NOTCH1 PEST Domain Mutants in Cancer Homo sapiens R-HSA-2644602 | 7.64E-01 | 9.69E-01 | 7.24E-01 | 1.95E-01 | JAG1;UBC |
| 808 | Constitutive Signaling by NOTCH1 PEST Domain Mutants Homo sapiens R-HSA-2644606 | 7.64E-01 | 9.69E-01 | 7.24E-01 | 1.95E-01 | JAG1;UBC |

| | | | | | | |
|---|---|---|---|---|---|---|
| 809 | Signaling by NOTCH1 HD+PEST Domain Mutants in Cancer Homo sapiens R-HSA-2894858 | 7.64E-01 | 9.69E-01 | 7.24E-01 | 1.95E-01 | JAG1;UBC |
| 810 | Hh mutants abrogate ligand secretion Homo sapiens R-HSA-5387390 | 7.64E-01 | 9.69E-01 | 7.24E-01 | 1.95E-01 | UBC;PSMB9 |
| 811 | p75 NTR receptor-mediated signalling Homo sapiens R-HSA-193704 | 7.65E-01 | 9.69E-01 | 7.46E-01 | 2.00E-01 | NFKBIA;RASGRF2;UBC |
| 812 | NEP/NS2 Interacts with the Cellular Export Machinery Homo sapiens R-HSA-168333 | 7.70E-01 | 9.69E-01 | 6.87E-01 | 1.79E-01 | NUP210 |
| 813 | Regulation of Glucokinase by Glucokinase Regulatory Protein Homo sapiens R-HSA-170822 | 7.70E-01 | 9.69E-01 | 6.87E-01 | 1.79E-01 | NUP210 |
| 814 | Transport of Ribonucleoproteins into the Host Nucleus Homo sapiens R-HSA-168271 | 7.70E-01 | 9.69E-01 | 6.87E-01 | 1.79E-01 | NUP210 |
| 815 | RNA Polymerase I Promoter Opening Homo sapiens R-HSA-73728 | 7.70E-01 | 9.69E-01 | 6.87E-01 | 1.79E-01 | HIST1H2AC |
| 816 | Metabolism of steroid hormones Homo sapiens R-HSA-196071 | 7.70E-01 | 9.69E-01 | 6.87E-01 | 1.79E-01 | AKR1B1 |
| 817 | TP53 Regulates Metabolic Genes Homo sapiens R-HSA-5628897 | 7.72E-01 | 9.69E-01 | 7.37E-01 | 1.91E-01 | SESN3;MTOR;GLS |
| 818 | Activation of the mRNA upon binding of the cap-binding complex and eIFs, and subsequent binding to 43S Homo sapiens R-HSA-72662 | 7.72E-01 | 9.69E-01 | 7.11E-01 | 1.84E-01 | RPS4Y1;RPS10 |

| | | | | | | |
|---|---|---|---|---|---|---|
| 819 | Formation of the beta-catenin:TCF transactivating complex Homo sapiens R-HSA-201722 | 7.72E-01 | 9.69E-01 | 7.11E-01 | 1.84E-01 | TCF7L1;HIST1H2AC |
| 820 | NIK-->noncanonical NF-kB signaling Homo sapiens R-HSA-5676590 | 7.72E-01 | 9.69E-01 | 7.11E-01 | 1.84E-01 | UBC;PSMB9 |
| 821 | Golgi-to-ER retrograde transport Homo sapiens R-HSA-8856688 | 7.77E-01 | 9.69E-01 | 7.51E-01 | 1.89E-01 | KIF26B;TMED3;RAB18;KIF1C |
| 822 | DNA Damage/Telomere Stress Induced Senescence Homo sapiens R-HSA-2559586 | 7.80E-01 | 9.69E-01 | 6.98E-01 | 1.73E-01 | HIST1H2AC;HIST1H1C |
| 823 | Degradation of GLI1 by the proteasome Homo sapiens R-HSA-5610780 | 7.80E-01 | 9.69E-01 | 6.98E-01 | 1.73E-01 | UBC;PSMB9 |
| 824 | GLI3 is processed to GLI3R by the proteasome Homo sapiens R-HSA-5610785 | 7.80E-01 | 9.69E-01 | 6.98E-01 | 1.73E-01 | UBC;PSMB9 |
| 825 | Degradation of GLI2 by the proteasome Homo sapiens R-HSA-5610783 | 7.80E-01 | 9.69E-01 | 6.98E-01 | 1.73E-01 | UBC;PSMB9 |
| 826 | The role of GTSE1 in G2/M progression after G2 checkpoint Homo sapiens R-HSA-8852276 | 7.80E-01 | 9.69E-01 | 6.98E-01 | 1.73E-01 | UBC;PSMB9 |
| 827 | SCF(Skp2)-mediated degradation of p27/p21 Homo sapiens R-HSA-187577 | 7.80E-01 | 9.69E-01 | 6.98E-01 | 1.73E-01 | UBC;PSMB9 |
| 828 | Export of Viral Ribonucleoproteins from Nucleus Homo sapiens R-HSA-168274 | 7.81E-01 | 9.69E-01 | 6.64E-01 | 1.64E-01 | NUP210 |

| | | | | | | |
|---|---|---|---|---|---|---|
| 829 | Packaging Of Telomere Ends Homo sapiens R-HSA-171306 | 7.81E-01 | 9.69E-01 | 6.64E-01 | 1.64E-01 | HIST1H2AC |
| 830 | NOD1/2 Signaling Pathway Homo sapiens R-HSA-168638 | 7.81E-01 | 9.69E-01 | 6.64E-01 | 1.64E-01 | IRAK2 |
| 831 | Regulation of TNFR1 signaling Homo sapiens R-HSA-5357905 | 7.81E-01 | 9.69E-01 | 6.64E-01 | 1.64E-01 | UBC |
| 832 | FCERI mediated NF-kB activation Homo sapiens R-HSA-2871837 | 7.83E-01 | 9.69E-01 | 7.44E-01 | 1.82E-01 | NFKBIA;UBC;MALT1;PSMB9 |
| 833 | Unfolded Protein Response (UPR) Homo sapiens R-HSA-381119 | 7.85E-01 | 9.69E-01 | 7.19E-01 | 1.74E-01 | WFS1;CCL2;DCP2 |
| 834 | Regulation of mRNA stability by proteins that bind AU-rich elements Homo sapiens R-HSA-450531 | 7.85E-01 | 9.69E-01 | 7.19E-01 | 1.74E-01 | UBC;DCP2;PSMB9 |
| 835 | Vpr-mediated nuclear import of PICs Homo sapiens R-HSA-180910 | 7.92E-01 | 9.69E-01 | 6.42E-01 | 1.50E-01 | NUP210 |
| 836 | DNA methylation Homo sapiens R-HSA-5334118 | 7.92E-01 | 9.69E-01 | 6.42E-01 | 1.50E-01 | HIST1H2AC |
| 837 | DNA strand elongation Homo sapiens R-HSA-69190 | 7.92E-01 | 9.69E-01 | 6.42E-01 | 1.50E-01 | MCM8 |
| 838 | SMAD2/SMAD3:SMAD4 heterotrimer regulates transcription Homo sapiens R-HSA-2173796 | 7.92E-01 | 9.69E-01 | 6.42E-01 | 1.50E-01 | UBC |
| 839 | Gluconeogenesis Homo sapiens R-HSA-70263 | 7.92E-01 | 9.69E-01 | 6.42E-01 | 1.50E-01 | PCK1 |
| 840 | TRAF6 mediated IRF7 activation Homo sapiens R-HSA-933541 | 7.92E-01 | 9.69E-01 | 6.42E-01 | 1.50E-01 | DDX58 |

| | | | | | | |
|---|---|---|---|---|---|---|
| 841 | Eukaryotic Translation Termination Homo sapiens R-HSA-72764 | 7.92E-01 | 9.69E-01 | 7.11E-01 | 1.66E-01 | ETF1;RPS4Y1;RPS10 |
| 842 | Role of LAT2/NTAL/LAB on calcium mobilization Homo sapiens R-HSA-2730905 | 7.92E-01 | 9.69E-01 | 7.65E-01 | 1.78E-01 | FGF18;PDGFB;PIK3R3;FGF2;EGFR;MTOR |
| 843 | Dectin-1 mediated noncanonical NF-kB signaling Homo sapiens R-HSA-5607761 | 7.96E-01 | 9.69E-01 | 6.75E-01 | 1.54E-01 | UBC;PSMB9 |
| 844 | Cell death signalling via NRAGE, NRIF and NADE Homo sapiens R-HSA-204998 | 7.96E-01 | 9.69E-01 | 6.75E-01 | 1.54E-01 | RASGRF2;UBC |
| 845 | Apoptosis Homo sapiens R-HSA-109581 | 7.97E-01 | 9.69E-01 | 7.60E-01 | 1.73E-01 | GSN;UBC;VIM;PAK2;HIST1H1C;PSMB9 |
| 846 | Asparagine N-linked glycosylation Homo sapiens R-HSA-446203 | 7.98E-01 | 9.69E-01 | 7.98E-01 | 1.81E-01 | SERPINA1;ST6GALNAC2;MAN2A2;FUCA1;RPN1;TMED3;GNPNAT1;ANK2;B4GALT6;SPTBN1 |
| 847 | Diseases of glycosylation Homo sapiens R-HSA-3781865 | 7.98E-01 | 9.69E-01 | 7.02E-01 | 1.58E-01 | SEMA5B;NOTCH4;HSPG2 |
| 848 | RHO GTPases Activate Formins Homo sapiens R-HSA-5663220 | 8.00E-01 | 9.69E-01 | 7.23E-01 | 1.61E-01 | ITGB1;DVL3;SRGAP2;PMF1 |
| 849 | Sialic acid metabolism Homo sapiens R-HSA-4085001 | 8.02E-01 | 9.69E-01 | 6.22E-01 | 1.38E-01 | ST6GALNAC2 |
| 850 | Cargo concentration in the ER Homo sapiens R-HSA-5694530 | 8.02E-01 | 9.69E-01 | 6.22E-01 | 1.38E-01 | SERPINA1 |
| 851 | Transport of the SLBP independent Mature mRNA Homo sapiens R-HSA-159227 | 8.02E-01 | 9.69E-01 | 6.22E-01 | 1.38E-01 | NUP210 |
| 852 | Glucagon signaling in metabolic regulation Homo sapiens R-HSA-163359 | 8.02E-01 | 9.69E-01 | 6.22E-01 | 1.38E-01 | PRKAR2B |

| | | | | | | |
|---|---|---|---|---|---|---|
| 853 | Inactivation, recovery and regulation of the phototransduction cascade Homo sapiens R-HSA-2514859 | 8.02E-01 | 9.69E-01 | 6.22E-01 | 1.38E-01 | CALM1 |
| 854 | Vesicle-mediated transport Homo sapiens R-HSA-5653656 | 8.03E-01 | 9.69E-01 | 8.41E-01 | 1.85E-01 | SERPINA1;SPARC;CLTC;LNPEP;ARRB1;ANK2;GNS;CLINT1;MAN2A2;KIF26B;GJA4;UBC;AP1S2;RAB18;TMED3;MYH8;KIF1C;RAB9A;CALM1;SPTBN1 |
| 855 | Diseases associated with O-glycosylation of proteins Homo sapiens R-HSA-3906995 | 8.03E-01 | 9.69E-01 | 6.63E-01 | 1.45E-01 | SEMA5B;NOTCH4 |
| 856 | Autodegradation of Cdh1 by Cdh1:APC/C Homo sapiens R-HSA-174084 | 8.03E-01 | 9.69E-01 | 6.63E-01 | 1.45E-01 | UBC;PSMB9 |
| 857 | FCERI mediated Ca+2 mobilization Homo sapiens R-HSA-2871809 | 8.03E-01 | 9.69E-01 | 6.63E-01 | 1.45E-01 | ITPR3;CALM1 |
| 858 | Nonsense Mediated Decay (NMD) independent of the Exon Junction Complex (EJC) Homo sapiens R-HSA-975956 | 8.05E-01 | 9.69E-01 | 6.94E-01 | 1.51E-01 | ETF1;RPS4Y1;RPS10 |
| 859 | Synthesis of substrates in N-glycan biosythesis Homo sapiens R-HSA-446219 | 8.10E-01 | 9.69E-01 | 6.52E-01 | 1.37E-01 | ST6GALNAC2;GNPNAT1 |
| 860 | p53-Dependent G1/S DNA damage checkpoint Homo sapiens R-HSA-69580 | 8.10E-01 | 9.69E-01 | 6.52E-01 | 1.37E-01 | UBC;PSMB9 |
| 861 | p53-Dependent G1 DNA Damage Response Homo sapiens R-HSA-69563 | 8.10E-01 | 9.69E-01 | 6.52E-01 | 1.37E-01 | UBC;PSMB9 |
| 862 | Programmed Cell Death Homo sapiens R-HSA-5357801 | 8.11E-01 | 9.69E-01 | 7.45E-01 | 1.57E-01 | GSN;UBC;VIM;PAK2;HIST1H1C;PSMB9 |

| | | | | | | |
|---|---|---|---|---|---|---|
| 863 | HSF1-dependent transactivation Homo sapiens R-HSA-3371571 | 8.11E-01 | 9.69E-01 | 6.03E-01 | 1.26E-01 | MTOR |
| 864 | Transport of the SLBP Dependant Mature mRNA Homo sapiens R-HSA-159230 | 8.11E-01 | 9.69E-01 | 6.03E-01 | 1.26E-01 | NUP210 |
| 865 | Nuclear Pore Complex (NPC) Disassembly Homo sapiens R-HSA-3301854 | 8.11E-01 | 9.69E-01 | 6.03E-01 | 1.26E-01 | NUP210 |
| 866 | The phototransduction cascade Homo sapiens R-HSA-2514856 | 8.11E-01 | 9.69E-01 | 6.03E-01 | 1.26E-01 | CALM1 |
| 867 | ROS, RNS production in response to bacteria Homo sapiens R-HSA-1222556 | 8.11E-01 | 9.69E-01 | 6.03E-01 | 1.26E-01 | LTF |
| 868 | G alpha (i) signalling events Homo sapiens R-HSA-418594 | 8.15E-01 | 9.73E-01 | 7.74E-01 | 1.58E-01 | RGS5;ANXA1;GAL;RGS19;GPER1;BDKRB2;S1PR3;RGS12;SSTR2 |
| 869 | Interactions of Vpr with host cellular proteins Homo sapiens R-HSA-176033 | 8.20E-01 | 9.77E-01 | 5.85E-01 | 1.16E-01 | NUP210 |
| 870 | SIRT1 negatively regulates rRNA Expression Homo sapiens R-HSA-427359 | 8.20E-01 | 9.77E-01 | 5.85E-01 | 1.16E-01 | HIST1H2AC |
| 871 | G1/S DNA Damage Checkpoints Homo sapiens R-HSA-69615 | 8.24E-01 | 9.80E-01 | 6.32E-01 | 1.22E-01 | UBC;PSMB9 |
| 872 | RNA Polymerase III Transcription Initiation Homo sapiens R-HSA-76046 | 8.29E-01 | 9.85E-01 | 5.69E-01 | 1.07E-01 | SNAPC1 |

| | | | | | | |
|---|---|---|---|---|---|---|
| 873 | APC/C:Cdc20 mediated degradation of Securin Homo sapiens R-HSA-174154 | 8.30E-01 | 9.85E-01 | 6.22E-01 | 1.16E-01 | UBC;PSMB9 |
| 874 | Regulation of RAS by GAPs Homo sapiens R-HSA-5658442 | 8.37E-01 | 9.88E-01 | 6.12E-01 | 1.09E-01 | UBC;PSMB9 |
| 875 | Defective B3GALTL causes Peters-plus syndrome (PpS) Homo sapiens R-HSA-5083635 | 8.37E-01 | 9.88E-01 | 5.53E-01 | 9.84E-02 | SEMA5B |
| 876 | Transport of Mature mRNA Derived from an Intronless Transcript Homo sapiens R-HSA-159231 | 8.37E-01 | 9.88E-01 | 5.53E-01 | 9.84E-02 | NUP210 |
| 877 | Steroid hormones Homo sapiens R-HSA-209943 | 8.37E-01 | 9.88E-01 | 5.53E-01 | 9.84E-02 | AKR1B1 |
| 878 | HIV Infection Homo sapiens R-HSA-162906 | 8.38E-01 | 9.88E-01 | 7.43E-01 | 1.31E-01 | NUP210;UBC;AP1S2;RCC1;FURIN;HLA-A;PAK2;PSMB9 |
| 879 | Cellular responses to stress Homo sapiens R-HSA-2262752 | 8.41E-01 | 9.88E-01 | 7.87E-01 | 1.36E-01 | PHC2;NUP210;STAT3;HIF1A;ETS1;SOD3;ETS2;MTOR;UBC;CAT;E2F3;HIST1H2AC;EZH2;HIST1H1C |
| 880 | Cyclin E associated events during G1/S transition Homo sapiens R-HSA-69202 | 8.43E-01 | 9.88E-01 | 6.03E-01 | 1.03E-01 | UBC;PSMB9 |
| 881 | O-glycosylation of TSR domain-containing proteins Homo sapiens R-HSA-5173214 | 8.45E-01 | 9.88E-01 | 5.38E-01 | 9.07E-02 | SEMA5B |
| 882 | Transport of Mature mRNAs Derived from Intronless Transcripts Homo sapiens R-HSA-159234 | 8.45E-01 | 9.88E-01 | 5.38E-01 | 9.07E-02 | NUP210 |
| 883 | DNA Damage Recognition in GG-NER Homo sapiens R-HSA-5696394 | 8.45E-01 | 9.88E-01 | 5.38E-01 | 9.07E-02 | UBC |

| | | | | | | |
|---|---|---|---|---|---|---|
| 884 | Glutathione conjugation Homo sapiens R-HSA-156590 | 8.45E-01 | 9.88E-01 | 5.38E-01 | 9.07E-02 | GSTM3 |
| 885 | Loss of Nlp from mitotic centrosomes Homo sapiens R-HSA-380259 | 8.48E-01 | 9.88E-01 | 5.94E-01 | 9.76E-02 | PRKAR2B;CDK5RAP2 |
| 886 | Loss of proteins required for interphase microtubule organization?from the centrosome Homo sapiens R-HSA-380284 | 8.48E-01 | 9.88E-01 | 5.94E-01 | 9.76E-02 | PRKAR2B;CDK5RAP2 |
| 887 | Synthesis of DNA Homo sapiens R-HSA-69239 | 8.49E-01 | 9.88E-01 | 6.35E-01 | 1.04E-01 | MCM8;UBC;PSMB9 |
| 888 | Anchoring of the basal body to the plasma membrane Homo sapiens R-HSA-5620912 | 8.49E-01 | 9.88E-01 | 6.35E-01 | 1.04E-01 | AHI1;PRKAR2B;CDK5RAP2 |
| 889 | S Phase Homo sapiens R-HSA-69242 | 8.50E-01 | 9.88E-01 | 6.63E-01 | 1.08E-01 | MCM8;UBC;CDC25B;PSMB9 |
| 890 | Class A/1 (Rhodopsin-like receptors) Homo sapiens R-HSA-373076 | 8.51E-01 | 9.88E-01 | 7.66E-01 | 1.23E-01 | ANXA1;GAL;GPR68;GPER1;F2R;BDKRB2;CCL2;GPR4;S1PR3;NTS;SSTR2;CX3CL1 |
| 891 | Dual Incision in GG-NER Homo sapiens R-HSA-5696400 | 8.52E-01 | 9.88E-01 | 5.24E-01 | 8.37E-02 | UBC |
| 892 | Association of TriC/CCT with target proteins during biosynthesis Homo sapiens R-HSA-390471 | 8.52E-01 | 9.88E-01 | 5.24E-01 | 8.37E-02 | STAT3 |
| 893 | Translation Homo sapiens R-HSA-72766 | 8.53E-01 | 9.88E-01 | 6.81E-01 | 1.08E-01 | EIF5;RPN1;ETF1;RPS4Y1;RPS10 |
| 894 | Infectious disease Homo sapiens R-HSA-5663205 | 8.54E-01 | 9.88E-01 | 7.70E-01 | 1.21E-01 | NUP210;ISG15;FURIN;HLA-A;RPS4Y1;PSMB9;UBC;AP1S2;RCC1;CALM1;RPS10;PAK2;LTF |
| 895 | Hedgehog 'off' state Homo sapiens R-HSA-5610787 | 8.58E-01 | 9.88E-01 | 6.21E-01 | 9.51E-02 | PRKAR2B;UBC;PSMB9 |

| | | | | | | |
|---|---|---|---|---|---|---|
| 896 | PKMTs methylate histone lysines Homo sapiens R-HSA-3214841 | 8.59E-01 | 9.88E-01 | 5.10E-01 | 7.74E-02 | EZH2 |
| 897 | APC/C:Cdh1 mediated degradation of Cdc20 and other APC/C:Cdh1 targeted proteins in late mitosis/early G1 Homo sapiens R-HSA-174178 | 8.60E-01 | 9.88E-01 | 5.77E-01 | 8.73E-02 | UBC;PSMB9 |
| 898 | Cdc20:Phospho-APC/C mediated degradation of Cyclin A Homo sapiens R-HSA-174184 | 8.60E-01 | 9.88E-01 | 5.77E-01 | 8.73E-02 | UBC;PSMB9 |
| 899 | Intra-Golgi and retrograde Golgi-to-ER traffic Homo sapiens R-HSA-6811442 | 8.62E-01 | 9.88E-01 | 6.89E-01 | 1.02E-01 | MAN2A2;KIF26B;TMED3;RAB18;KIF1C;RAB9A |
| 900 | Disease Homo sapiens R-HSA-1643685 | 8.64E-01 | 9.88E-01 | 8.25E-01 | 1.21E-01 | SEMA5B;NOTCH4;PDGFB;PIK3R3;FURIN;RPS4Y1;FGF2;EGFR;ACACA;UBC;AP1S2;RCC1;LRRFIP1;RPS10;PAK2;JAG1;NUP210;STAT3;ISG15;HLA-A;HSPG2;MTOR;PSMB9;NFKBIA;FGF18;NAPEPLD;CALM1;TLR5;LTF |
| 901 | Transcriptional regulation by small RNAs Homo sapiens R-HSA-5578749 | 8.65E-01 | 9.88E-01 | 5.68E-01 | 8.26E-02 | NUP210;HIST1H2AC |
| 902 | APC:Cdc20 mediated degradation of cell cycle proteins prior to satisfation of the cell cycle checkpoint Homo sapiens R-HSA-179419 | 8.65E-01 | 9.88E-01 | 5.68E-01 | 8.26E-02 | UBC;PSMB9 |
| 903 | AURKA Activation by TPX2 Homo sapiens R-HSA-8854518 | 8.65E-01 | 9.88E-01 | 5.68E-01 | 8.26E-02 | PRKAR2B;CDK5RAP2 |
| 904 | Intraflagellar transport Homo sapiens R-HSA-5620924 | 8.66E-01 | 9.88E-01 | 4.97E-01 | 7.15E-02 | TRAF3IP1 |

| | | | | | | |
|---|---|---|---|---|---|---|
| 905 | RNA Polymerase III Abortive And Retractive Initiation Homo sapiens R-HSA-749476 | 8.66E-01 | 9.88E-01 | 4.97E-01 | 7.15E-02 | SNAPC1 |
| 906 | RNA Polymerase III Transcription Homo sapiens R-HSA-74158 | 8.66E-01 | 9.88E-01 | 4.97E-01 | 7.15E-02 | SNAPC1 |
| 907 | Histidine, lysine, phenylalanine, tyrosine, proline and tryptophan catabolism Homo sapiens R-HSA-6788656 | 8.66E-01 | 9.88E-01 | 4.97E-01 | 7.15E-02 | AFMID |
| 908 | Late Phase of HIV Life Cycle Homo sapiens R-HSA-162599 | 8.66E-01 | 9.88E-01 | 6.41E-01 | 9.21E-02 | NUP210;UBC;RCC1;FURIN |
| 909 | Class I MHC mediated antigen processing & presentation Homo sapiens R-HSA-983169 | 8.68E-01 | 9.89E-01 | 7.42E-01 | 1.05E-01 | ZNRF1;FBXO2;NEDD4;UBC;UBR4;RNF182;LNPEP;ITGAV;RNF6;HLA-A;PSMB9 |
| 910 | Post-translational protein modification Homo sapiens R-HSA-597592 | 8.72E-01 | 9.90E-01 | 7.91E-01 | 1.09E-01 | SEMA5B;ARSA;GALNT6;PHC2;PGAP1;SERPINA1;ST6GALNAC2;NUP210;GALNT3;FUCA1;RPN1;FURIN;ANK2;MAN2A2;TMED3;B3GNT3;GNPNAT1;GAS6;B4GALT6;SPTBN1 |
| 911 | Viral Messenger RNA Synthesis Homo sapiens R-HSA-168325 | 8.72E-01 | 9.90E-01 | 4.85E-01 | 6.62E-02 | NUP210 |
| 912 | APC/C:Cdc20 mediated degradation of mitotic proteins Homo sapiens R-HSA-176409 | 8.75E-01 | 9.90E-01 | 5.52E-01 | 7.39E-02 | UBC;PSMB9 |
| 913 | Immunoregulatory interactions between a Lymphoid and a non-Lymphoid cell Homo sapiens R-HSA-198933 | 8.75E-01 | 9.90E-01 | 6.53E-01 | 8.69E-02 | ITGB1;CXADR;HLA-A;PVR;CD34 |

| | | | | | | |
|---|---|---|---|---|---|---|
| 914 | ER to Golgi Anterograde Transport Homo sapiens R-HSA-199977 | 8.78E-01 | 9.90E-01 | 6.26E-01 | 8.17E-02 | SERPINA1;TMED3;ANK2;SPTBN1 |
| 915 | ERCC6 (CSB) and EHMT2 (G9a) positively regulate rRNA expression Homo sapiens R-HSA-427389 | 8.79E-01 | 9.90E-01 | 4.74E-01 | 6.13E-02 | HIST1H2AC |
| 916 | Formation of Incision Complex in GG-NER Homo sapiens R-HSA-5696395 | 8.79E-01 | 9.90E-01 | 4.74E-01 | 6.13E-02 | UBC |
| 917 | Antigen processing: Ubiquitination & Proteasome degradation Homo sapiens R-HSA-983168 | 8.79E-01 | 9.90E-01 | 7.12E-01 | 9.20E-02 | ZNRF1;FBXO2;NEDD4;UBC;UBR4;RNF182;LNPEP;RNF6;PSMB9 |
| 918 | SUMOylation of DNA damage response and repair proteins Homo sapiens R-HSA-3108214 | 8.80E-01 | 9.90E-01 | 5.45E-01 | 7.00E-02 | PHC2;NUP210 |
| 919 | Activation of APC/C and APC/C:Cdc20 mediated degradation of mitotic proteins Homo sapiens R-HSA-176814 | 8.80E-01 | 9.90E-01 | 5.45E-01 | 7.00E-02 | UBC;PSMB9 |
| 920 | G alpha (12/13) signalling events Homo sapiens R-HSA-416482 | 8.80E-01 | 9.90E-01 | 5.45E-01 | 7.00E-02 | RASGRF2;PIK3R3 |
| 921 | Intra-Golgi traffic Homo sapiens R-HSA-6811438 | 8.84E-01 | 9.93E-01 | 4.63E-01 | 5.68E-02 | MAN2A2 |
| 922 | SUMOylation of DNA replication proteins Homo sapiens R-HSA-4615885 | 8.84E-01 | 9.93E-01 | 4.63E-01 | 5.68E-02 | NUP210 |
| 923 | Nonsense-Mediated Decay (NMD) Homo sapiens R-HSA-927802 | 8.87E-01 | 9.93E-01 | 5.79E-01 | 6.91E-02 | ETF1;RPS4Y1;RPS10 |

| | | | | | | |
|---|---|---|---|---|---|---|
| 924 | Nonsense Mediated Decay (NMD) enhanced by the Exon Junction Complex (EJC) Homo sapiens R-HSA-975957 | 8.87E-01 | 9.93E-01 | 5.79E-01 | 6.91E-02 | ETF1;RPS4Y1;RPS10 |
| 925 | RNA polymerase II transcribes snRNA genes Homo sapiens R-HSA-6807505 | 8.88E-01 | 9.93E-01 | 5.30E-01 | 6.27E-02 | SNAPC1;PHAX |
| 926 | RMTs methylate histone arginines Homo sapiens R-HSA-3214858 | 8.90E-01 | 9.93E-01 | 4.52E-01 | 5.27E-02 | HIST1H2AC |
| 927 | NRAGE signals death through JNK Homo sapiens R-HSA-193648 | 8.90E-01 | 9.93E-01 | 4.52E-01 | 5.27E-02 | RASGRF2 |
| 928 | SRP-dependent cotranslational protein targeting to membrane Homo sapiens R-HSA-1799339 | 8.91E-01 | 9.93E-01 | 5.73E-01 | 6.60E-02 | RPN1;RPS4Y1;RPS10 |
| 929 | GTP hydrolysis and joining of the 60S ribosomal subunit Homo sapiens R-HSA-72706 | 8.91E-01 | 9.93E-01 | 5.73E-01 | 6.60E-02 | EIF5;RPS4Y1;RPS10 |
| 930 | HATs acetylate histones Homo sapiens R-HSA-3214847 | 8.91E-01 | 9.93E-01 | 5.73E-01 | 6.60E-02 | JADE2;ELP5;HIST1H2AC |
| 931 | Biosynthesis of the N-glycan precursor (dolichol lipid-linked oligosaccharide, LLO) and transfer to a nascent protein Homo sapiens R-HSA-446193 | 8.93E-01 | 9.93E-01 | 5.23E-01 | 5.94E-02 | ST6GALNAC2;GNPNAT1 |
| 932 | G alpha (z) signalling events Homo sapiens R-HSA-418597 | 8.95E-01 | 9.94E-01 | 4.42E-01 | 4.89E-02 | RGS19 |

| | | | | | | |
|---|---|---|---|---|---|---|
| 933 | Regulation of APC/C activators between G1/S and early anaphase Homo sapiens R-HSA-176408 | 8.97E-01 | 9.94E-01 | 5.16E-01 | 5.62E-02 | UBC;PSMB9 |
| 934 | Centrosome maturation Homo sapiens R-HSA-380287 | 8.97E-01 | 9.94E-01 | 5.16E-01 | 5.62E-02 | PRKAR2B;CDK5RAP2 |
| 935 | Recruitment of mitotic centrosome proteins and complexes Homo sapiens R-HSA-380270 | 8.97E-01 | 9.94E-01 | 5.16E-01 | 5.62E-02 | PRKAR2B;CDK5RAP2 |
| 936 | Nucleotide-binding domain, leucine rich repeat containing receptor (NLR) signaling pathways Homo sapiens R-HSA-168643 | 9.00E-01 | 9.96E-01 | 4.32E-01 | 4.55E-02 | IRAK2 |
| 937 | Peptide hormone metabolism Homo sapiens R-HSA-2980736 | 9.05E-01 | 9.99E-01 | 5.03E-01 | 5.05E-02 | ACHE;CPE |
| 938 | TP53 Regulates Transcription of Cell Cycle Genes Homo sapiens R-HSA-6791312 | 9.05E-01 | 9.99E-01 | 4.23E-01 | 4.22E-02 | RGCC |
| 939 | Selenoamino acid metabolism Homo sapiens R-HSA-2408522 | 9.05E-01 | 9.99E-01 | 5.52E-01 | 5.51E-02 | NNMT;RPS4Y1;RPS10 |
| 940 | Retrograde transport at the Trans-Golgi-Network Homo sapiens R-HSA-6811440 | 9.10E-01 | 1.00E+00 | 4.14E-01 | 3.93E-02 | RAB9A |
| 941 | Cross-presentation of soluble exogenous antigens (endosomes) Homo sapiens R-HSA-1236978 | 9.10E-01 | 1.00E+00 | 4.14E-01 | 3.93E-02 | PSMB9 |
| 942 | HIV Life Cycle Homo sapiens R-HSA-162587 | 9.10E-01 | 1.00E+00 | 5.80E-01 | 5.48E-02 | NUP210;UBC;RCC1;FURIN |

| | | | | | | |
|---|---|---|---|---|---|---|
| 943 | Deadenylation-dependent mRNA decay Homo sapiens R-HSA-429914 | 9.14E-01 | 1.00E+00 | 4.06E-01 | 3.66E-02 | DCP2 |
| 944 | Regulation of ornithine decarboxylase (ODC) Homo sapiens R-HSA-350562 | 9.14E-01 | 1.00E+00 | 4.06E-01 | 3.66E-02 | PSMB9 |
| 945 | Cap-dependent Translation Initiation Homo sapiens R-HSA-72737 | 9.14E-01 | 1.00E+00 | 5.37E-01 | 4.81E-02 | EIF5;RPS4Y1;RPS10 |
| 946 | Eukaryotic Translation Initiation Homo sapiens R-HSA-72613 | 9.14E-01 | 1.00E+00 | 5.37E-01 | 4.81E-02 | EIF5;RPS4Y1;RPS10 |
| 947 | Viral mRNA Translation Homo sapiens R-HSA-192823 | 9.15E-01 | 1.00E+00 | 4.85E-01 | 4.29E-02 | RPS4Y1;RPS10 |
| 948 | Peptide chain elongation Homo sapiens R-HSA-156902 | 9.15E-01 | 1.00E+00 | 4.85E-01 | 4.29E-02 | RPS4Y1;RPS10 |
| 949 | Meiosis Homo sapiens R-HSA-1500620 | 9.19E-01 | 1.00E+00 | 4.79E-01 | 4.07E-02 | SYNE2;HIST1H2AC |
| 950 | APC/C-mediated degradation of cell cycle proteins Homo sapiens R-HSA-174143 | 9.19E-01 | 1.00E+00 | 4.79E-01 | 4.07E-02 | UBC;PSMB9 |
| 951 | Regulation of mitotic cell cycle Homo sapiens R-HSA-453276 | 9.19E-01 | 1.00E+00 | 4.79E-01 | 4.07E-02 | UBC;PSMB9 |
| 952 | Chromosome Maintenance Homo sapiens R-HSA-73886 | 9.22E-01 | 1.00E+00 | 4.73E-01 | 3.86E-02 | CENPW;HIST1H2AC |
| 953 | Selenocysteine synthesis Homo sapiens R-HSA-2408557 | 9.25E-01 | 1.00E+00 | 4.68E-01 | 3.66E-02 | RPS4Y1;RPS10 |
| 954 | Influenza Infection Homo sapiens R-HSA-168254 | 9.25E-01 | 1.00E+00 | 5.55E-01 | 4.31E-02 | NUP210;ISG15;RPS4Y1;RPS10 |

| | | | | | | |
|---|---|---|---|---|---|---|
| 955 | XBP1(S) activates chaperone genes Homo sapiens R-HSA-381038 | 9.26E-01 | 1.00E+00 | 3.82E-01 | 2.95E-02 | WFS1 |
| 956 | Formation of TC-NER Pre-Incision Complex Homo sapiens R-HSA-6781823 | 9.26E-01 | 1.00E+00 | 3.82E-01 | 2.95E-02 | UBC |
| 957 | Meiotic recombination Homo sapiens R-HSA-912446 | 9.29E-01 | 1.00E+00 | 3.75E-01 | 2.75E-02 | HIST1H2AC |
| 958 | Eukaryotic Translation Elongation Homo sapiens R-HSA-156842 | 9.30E-01 | 1.00E+00 | 4.57E-01 | 3.29E-02 | RPS4Y1;RPS10 |
| 959 | Activation of anterior HOX genes in hindbrain development during early embryogenesis Homo sapiens R-HSA-5617472 | 9.30E-01 | 1.00E+00 | 4.57E-01 | 3.29E-02 | HIST1H2AC;EZH2 |
| 960 | Activation of HOX genes during differentiation Homo sapiens R-HSA-5619507 | 9.30E-01 | 1.00E+00 | 4.57E-01 | 3.29E-02 | HIST1H2AC;EZH2 |
| 961 | Regulation of TP53 Activity through Phosphorylation Homo sapiens R-HSA-6804756 | 9.30E-01 | 1.00E+00 | 4.57E-01 | 3.29E-02 | NUAK1;UBC |
| 962 | G2/M Checkpoints Homo sapiens R-HSA-69481 | 9.32E-01 | 1.00E+00 | 5.44E-01 | 3.82E-02 | DBF4;MCM8;UBC;PSMB9 |
| 963 | Cell Cycle, Mitotic Homo sapiens R-HSA-69278 | 9.33E-01 | 1.00E+00 | 7.10E-01 | 4.95E-02 | NUP210;MCM8;SMC4;PMF1;PSMB9;CDC25B;DHFR;DBF4;PRKAR2B;UBC;E2F3;PHLDA1;LPIN2;OPTN;HIST1H2AC;CDK5RAP2 |
| 964 | IRE1alpha activates chaperones Homo sapiens R-HSA-381070 | 9.33E-01 | 1.00E+00 | 3.68E-01 | 2.57E-02 | WFS1 |
| 965 | Regulation of TP53 Activity Homo sapiens R-HSA-5633007 | 9.34E-01 | 1.00E+00 | 5.40E-01 | 3.67E-02 | NUAK1;UBC;SGK1;MTOR |

| | | | | | | |
|---|---|---|---|---|---|---|
| 966 | The citric acid (TCA) cycle and respiratory electron transport Homo sapiens R-HSA-1428517 | 9.38E-01 | 1.00E+00 | 5.33E-01 | 3.39E-02 | LDHA;UCP2;PDK3;HAGH |
| 967 | tRNA processing in the nucleus Homo sapiens R-HSA-6784531 | 9.39E-01 | 1.00E+00 | 3.55E-01 | 2.24E-02 | NUP210 |
| 968 | RNA Polymerase I Chain Elongation Homo sapiens R-HSA-73777 | 9.39E-01 | 1.00E+00 | 3.55E-01 | 2.24E-02 | HIST1H2AC |
| 969 | Signaling by GPCR Homo sapiens R-HSA-372790 | 9.39E-01 | 1.00E+00 | 8.08E-01 | 5.05E-02 | GPR68;RASGRF2;ARRB1;FGF2;CX3CL1;RGS5;RGS3;GPER1;BDKRB2;ANXA1;KSR1;VWF;DUSP1;PDE4D;F2R;GLP2R;SSTR2;DUSP6;GAL;IL3RA;RAMP2;CALCA;CALCB;CALCRL;RGS19;GIPR;PDGFB;PIK3R3;GPR83;ITPR3;IL2RG;NTS;EGFR;RASGRP3;RAP1B;WNT11;PRKAR2B;UBC;CCL2;S1PR3;SPTBN1;FZD2;WNT7B;PDE2A;FZD9;GPR4;PSMB9;DLG4;FGF18;RGS12;CALM1 |
| 970 | B-WICH complex positively regulates rRNA expression Homo sapiens R-HSA-5250924 | 9.42E-01 | 1.00E+00 | 3.49E-01 | 2.09E-02 | HIST1H2AC |
| 971 | Recruitment and ATM-mediated phosphorylation of repair and signaling proteins at DNA double strand breaks Homo sapiens R-HSA-5693565 | 9.42E-01 | 1.00E+00 | 3.49E-01 | 2.09E-02 | UBC |
| 972 | Telomere Maintenance Homo sapiens R-HSA-157579 | 9.45E-01 | 1.00E+00 | 3.43E-01 | 1.95E-02 | HIST1H2AC |
| 973 | DNA Double Strand Break Response Homo sapiens R-HSA-5693606 | 9.45E-01 | 1.00E+00 | 3.43E-01 | 1.95E-02 | UBC |
| 974 | HDACs deacetylate histones Homo sapiens R-HSA-3214815 | 9.47E-01 | 1.00E+00 | 3.37E-01 | 1.82E-02 | HIST1H2AC |

| | | | | | | |
|---|---|---|---|---|---|---|
| 975 | Role of phospholipids in phagocytosis Homo sapiens R-HSA-2029485 | 9.47E-01 | 1.00E+00 | 3.37E-01 | 1.82E-02 | ITPR3 |
| 976 | Cellular response to heat stress Homo sapiens R-HSA-3371556 | 9.48E-01 | 1.00E+00 | 4.23E-01 | 2.28E-02 | NUP210;MTOR |
| 977 | SUMO E3 ligases SUMOylate target proteins Homo sapiens R-HSA-3108232 | 9.48E-01 | 1.00E+00 | 4.23E-01 | 2.28E-02 | PHC2;NUP210 |
| 978 | Formation of a pool of free 40S subunits Homo sapiens R-HSA-72689 | 9.48E-01 | 1.00E+00 | 4.23E-01 | 2.28E-02 | RPS4Y1;RPS10 |
| 979 | Influenza Viral RNA Transcription and Replication Homo sapiens R-HSA-168273 | 9.48E-01 | 1.00E+00 | 4.76E-01 | 2.56E-02 | NUP210;RPS4Y1;RPS10 |
| 980 | Metabolism of proteins Homo sapiens R-HSA-392499 | 9.48E-01 | 1.00E+00 | 7.81E-01 | 4.15E-02 | ACHE;SEMA5B;CALCA;PGAP1;ST6GALNAC2;SERPINA1;RPN1;FURIN;RPS4Y1;ZDHHC2;MAN2A2;UBC;TMED3;CCL2;CTSH;GNPNAT1;RPS10;SPTBN1;HIST1H2AC;ARSA;GALNT6;PHC2;GSN;GALNT3;NUP210;IGFBP5;WFS1;FUCA1;STAT3;IGFBP2;ANK2;SNCAIP;HSPG2;EIF5;B3GNT3;CPE;ETF1;GAS6;B4GALT6;DCP2;LTF |
| 981 | Gap-filling DNA repair synthesis and ligation in TC-NER Homo sapiens R-HSA-6782210 | 9.52E-01 | 1.00E+00 | 3.26E-01 | 1.59E-02 | UBC |
| 982 | Dual incision in TC-NER Homo sapiens R-HSA-6782135 | 9.55E-01 | 1.00E+00 | 3.21E-01 | 1.49E-02 | UBC |
| 983 | HDR through Homologous Recombination (HRR) Homo sapiens R-HSA-5685942 | 9.57E-01 | 1.00E+00 | 3.15E-01 | 1.40E-02 | UBC |
| 984 | SUMOylation Homo sapiens R-HSA-2990846 | 9.57E-01 | 1.00E+00 | 4.01E-01 | 1.75E-02 | PHC2;NUP210 |
| 985 | Influenza Life Cycle Homo sapiens R-HSA-168255 | 9.61E-01 | 1.00E+00 | 4.48E-01 | 1.79E-02 | NUP210;RPS4Y1;RPS10 |

| | | | | | | |
|---|---|---|---|---|---|---|
| 986 | Metabolism of amino acids and derivatives Homo sapiens R-HSA-71291 | 9.62E-01 | 1.00E+00 | 6.09E-01 | 2.38E-02 | ARG2;NNMT;AFMID;SLC6A12;RPS4Y1;CDO1;RPS10;SAT1;GLS;PSMB9 |
| 987 | Macroautophagy Homo sapiens R-HSA-1632852 | 9.63E-01 | 1.00E+00 | 3.01E-01 | 1.15E-02 | MTOR |
| 988 | COPII (Coat Protein 2) Mediated Vesicle Transport Homo sapiens R-HSA-204005 | 9.64E-01 | 1.00E+00 | 2.97E-01 | 1.07E-02 | SERPINA1 |
| 989 | Transport of Mature mRNA derived from an Intron-Containing Transcript Homo sapiens R-HSA-159236 | 9.64E-01 | 1.00E+00 | 2.97E-01 | 1.07E-02 | NUP210 |
| 990 | L13a-mediated translational silencing of Ceruloplasmin expression Homo sapiens R-HSA-156827 | 9.65E-01 | 1.00E+00 | 3.82E-01 | 1.35E-02 | RPS4Y1;RPS10 |
| 991 | 3' -UTR-mediated translational regulation Homo sapiens R-HSA-157279 | 9.65E-01 | 1.00E+00 | 3.82E-01 | 1.35E-02 | RPS4Y1;RPS10 |
| 992 | Mitotic Prometaphase Homo sapiens R-HSA-68877 | 9.67E-01 | 1.00E+00 | 3.78E-01 | 1.29E-02 | SMC4;PMF1 |
| 993 | Positive epigenetic regulation of rRNA expression Homo sapiens R-HSA-5250913 | 9.72E-01 | 1.00E+00 | 2.76E-01 | 7.78E-03 | HIST1H2AC |
| 994 | NoRC negatively regulates rRNA expression Homo sapiens R-HSA-427413 | 9.72E-01 | 1.00E+00 | 2.76E-01 | 7.78E-03 | HIST1H2AC |
| 995 | Transport of Mature Transcript to Cytoplasm Homo sapiens R-HSA-72202 | 9.74E-01 | 1.00E+00 | 2.72E-01 | 7.30E-03 | NUP210 |

| | | | | | | |
|---|---|---|---|---|---|---|
| 996 | Binding and Uptake of Ligands by Scavenger Receptors Homo sapiens R-HSA-2173782 | 9.74E-01 | 1.00E+00 | 2.72E-01 | 7.30E-03 | SPARC |
| 997 | M Phase Homo sapiens R-HSA-68886 | 9.75E-01 | 1.00E+00 | 5.31E-01 | 1.37E-02 | NUP210;UBC;SMC4;PMF1;LPIN2;HIST1H2AC;PSMB9 |
| 998 | Negative epigenetic regulation of rRNA expression Homo sapiens R-HSA-5250941 | 9.76E-01 | 1.00E+00 | 2.65E-01 | 6.43E-03 | HIST1H2AC |
| 999 | RNA Polymerase I Promoter Clearance Homo sapiens R-HSA-73854 | 9.76E-01 | 1.00E+00 | 2.65E-01 | 6.43E-03 | HIST1H2AC |
| 1000 | Transcription-Coupled Nucleotide Excision Repair (TC-NER) Homo sapiens R-HSA-6781827 | 9.76E-01 | 1.00E+00 | 2.65E-01 | 6.43E-03 | UBC |
| 1001 | Epigenetic regulation of gene expression Homo sapiens R-HSA-212165 | 9.76E-01 | 1.00E+00 | 3.51E-01 | 8.52E-03 | HIST1H2AC;EZH2 |
| 1002 | Cell Cycle Checkpoints Homo sapiens R-HSA-69620 | 9.77E-01 | 1.00E+00 | 4.45E-01 | 1.05E-02 | DBF4;MCM8;UBC;PSMB9 |
| 1003 | RNA Polymerase I, RNA Polymerase III, and Mitochondrial Transcription Homo sapiens R-HSA-504046 | 9.77E-01 | 1.00E+00 | 3.48E-01 | 8.09E-03 | SNAPC1;HIST1H2AC |
| 1004 | Cell Cycle Homo sapiens R-HSA-1640170 | 9.77E-01 | 1.00E+00 | 6.48E-01 | 1.48E-02 | CENPW;NUP210;MCM8;SMC4;PMF1;SYNE2;PSMB9;CDC25B;DHFR;DBF4;PRKAR2B;UBC;E2F3;PHLDA1;LPIN2;OPTN;HIST1H2AC;CDK5RAP2 |
| 1005 | RNA Polymerase I Transcription Homo sapiens R-HSA-73864 | 9.79E-01 | 1.00E+00 | 2.55E-01 | 5.33E-03 | HIST1H2AC |
| 1006 | Regulation of HSF1-mediated heat shock response Homo sapiens R-HSA-3371453 | 9.80E-01 | 1.00E+00 | 2.51E-01 | 5.01E-03 | NUP210 |
| 1007 | Assembly of the primary cilium Homo sapiens R-HSA-5617833 | 9.80E-01 | 1.00E+00 | 4.33E-01 | 8.56E-03 | AHI1;TRAF3IP1;PRKAR2B;CDK5RAP2 |

| | | | | | | |
|---|---|---|---|---|---|---|
| 1008 | Processing of DNA double-strand break ends Homo sapiens R-HSA-5693607 | 9.81E-01 | 1.00E+00 | 2.48E-01 | 4.70E-03 | UBC |
| 1009 | Global Genome Nucleotide Excision Repair (GG-NER) Homo sapiens R-HSA-5696399 | 9.82E-01 | 1.00E+00 | 2.45E-01 | 4.42E-03 | UBC |
| 1010 | Mitochondrial translation initiation Homo sapiens R-HSA-5368286 | 9.84E-01 | 1.00E+00 | 2.39E-01 | 3.91E-03 | MRPS6 |
| 1011 | Mitochondrial translation termination Homo sapiens R-HSA-5419276 | 9.84E-01 | 1.00E+00 | 2.39E-01 | 3.91E-03 | MRPS6 |
| 1012 | Mitochondrial translation elongation Homo sapiens R-HSA-5389840 | 9.84E-01 | 1.00E+00 | 2.39E-01 | 3.91E-03 | MRPS6 |
| 1013 | Chromatin organization Homo sapiens R-HSA-4839726 | 9.85E-01 | 1.00E+00 | 4.48E-01 | 6.77E-03 | JADE2;UTY;ELP5;HIST1H2AC;EZH2 |
| 1014 | Chromatin modifying enzymes Homo sapiens R-HSA-3247509 | 9.85E-01 | 1.00E+00 | 4.48E-01 | 6.77E-03 | JADE2;UTY;ELP5;HIST1H2AC;EZH2 |
| 1015 | Separation of Sister Chromatids Homo sapiens R-HSA-2467813 | 9.85E-01 | 1.00E+00 | 3.74E-01 | 5.56E-03 | UBC;PMF1;PSMB9 |
| 1016 | Transcriptional Regulation by TP53 Homo sapiens R-HSA-3700989 | 9.87E-01 | 1.00E+00 | 5.24E-01 | 6.93E-03 | BNIP3L;NUAK1;RGCC;SESN3;BCL6;UBC;SGK1;MTOR;GLS |
| 1017 | Mitochondrial translation Homo sapiens R-HSA-5368287 | 9.88E-01 | 1.00E+00 | 2.23E-01 | 2.71E-03 | MRPS6 |
| 1018 | Mitotic Anaphase Homo sapiens R-HSA-68882 | 9.90E-01 | 1.00E+00 | 3.49E-01 | 3.39E-03 | UBC;PMF1;PSMB9 |

| | | | | | | |
|---|---|---|---|---|---|---|
| 1019 | Chaperonin-mediated protein folding Homo sapiens R-HSA-390466 | 9.91E-01 | 1.00E+00 | 2.11E-01 | 2.00E-03 | STAT3 |
| 1020 | Mitotic Metaphase and Anaphase Homo sapiens R-HSA-2555396 | 9.91E-01 | 1.00E+00 | 3.47E-01 | 3.24E-03 | UBC;PMF1;PSMB9 |
| 1021 | Resolution of Sister Chromatid Cohesion Homo sapiens R-HSA-2500257 | 9.92E-01 | 1.00E+00 | 2.02E-01 | 1.57E-03 | PMF1 |
| 1022 | Protein folding Homo sapiens R-HSA-391251 | 9.93E-01 | 1.00E+00 | 1.98E-01 | 1.40E-03 | STAT3 |
| 1023 | tRNA processing Homo sapiens R-HSA-72306 | 9.94E-01 | 1.00E+00 | 1.94E-01 | 1.24E-03 | NUP210 |
| 1024 | Nucleotide Excision Repair Homo sapiens R-HSA-5696398 | 9.95E-01 | 1.00E+00 | 1.85E-01 | 9.24E-04 | UBC |
| 1025 | Respiratory electron transport, ATP synthesis by chemiosmotic coupling, and heat production by uncoupling proteins. Homo sapiens R-HSA-163200 | 9.95E-01 | 1.00E+00 | 1.84E-01 | 8.71E-04 | UCP2 |
| 1026 | HDR through Homologous Recombination (HR) or Single Strand Annealing (SSA) Homo sapiens R-HSA-5693567 | 9.96E-01 | 1.00E+00 | 1.79E-01 | 7.31E-04 | UBC |
| 1027 | Homology Directed Repair Homo sapiens R-HSA-5693538 | 9.97E-01 | 1.00E+00 | 1.69E-01 | 5.16E-04 | UBC |
| 1028 | Major pathway of rRNA processing in the nucleolus Homo sapiens R-HSA-6791226 | 9.97E-01 | 1.00E+00 | 2.41E-01 | 6.47E-04 | RPS4Y1;RPS10 |
| 1029 | rRNA processing Homo sapiens R-HSA-72312 | 9.99E-01 | 1.00E+00 | 2.22E-01 | 3.21E-04 | RPS4Y1;RPS10 |

| | | | | | | |
|---|---|---|---|---|---|---|
| 1030 | GPCR downstream signaling Homo sapiens R-HSA-388396 | 9.99E-01 | 1.00E+00 | 5.93E-01 | 7.65E-04 | RAMP2;CALCA;CALCB;CALCRL;RGS19;GPR68;GIPR;RASGRF2;PIK3R3;GPR83;ARRB1;ITPR3;IL2RG;NTS;RGS5;RGS3;GPER1;BDKRB2;S1PR3;ANXA1;PDE4D;F2R;PDE2A;GLP2R;GPR4;SSTR2;GAL;IL3RA;RGS12 |
| 1031 | DNA Double-Strand Break Repair Homo sapiens R-HSA-5693532 | 9.99E-01 | 1.00E+00 | 1.37E-01 | 1.11E-04 | UBC |
| 1032 | Organelle biogenesis and maintenance Homo sapiens R-HSA-1852241 | 1.00E+00 | 1.00E+00 | 3.07E-01 | 1.27E-04 | AHI1;TRAF3IP1;PRKAR2B;MRPS6;CDK5RAP2 |
| 1033 | DNA Repair Homo sapiens R-HSA-73894 | 1.00E+00 | 1.00E+00 | 2.09E-01 | 2.05E-05 | UBE2T;UBC;ISG15 |
| 1034 | Processing of Capped Intron-Containing Pre-mRNA Homo sapiens R-HSA-72203 | 1.00E+00 | 1.00E+00 | 1.03E-01 | 8.02E-06 | NUP210 |
| 1035 | Gene Expression Homo sapiens R-HSA-74160 | 1.00E+00 | 1.00E+00 | 3.41E-01 | 2.10E-06 | NOTCH4;RPN1;PHAX;RPS4Y1;GLS;NUAK1;SESN3;SNAPC1;ZNF703;UBC;SKIL;RPS10;HIST1H2AC;BNIP3L;NUP210;NR1H4;TDRD6;MTOR;PSMB9;H19;EIF5;RGCC;BCL6;TDRD12;PPARG;ETF1;SGK1;DCP2;EZH2 |
| 1036 | Generic Transcription Pathway Homo sapiens R-HSA-212436 | 1.00E+00 | 1.00E+00 | 3.40E-01 | 1.97E-06 | BNIP3L;NOTCH4;NR1H4;MTOR;GLS;NUAK1;RGCC;SESN3;BCL6;ZNF703;UBC;PPARG;SGK1;SKIL |

**Pathways enriched by genes enhanced with SGLT2 inhibitor in thick ascending limb cells**

| | Term | P value | Adjusted P value | Odds Ratio | Combined Score | Genes |
|---|---|---|---|---|---|---|
| 1 | Metabolism Homo sapiens R-HSA-1430728 | 5.12E-50 | 4.55E-47 | 4.31E+00 | 4.89E+02 | RPL3;RPL32;RPL31;RPL34;ECI1;ECI2;MT1X;RPL8;NUDT4;RPS14;MT2A;RPS16;RPS18;BSG;RPL36;RPL35;RPL38;RPL37;PTGDS;ACAD11;RPL39;GSTK1;RPL21;RPL23;MCCC1;CMBL;MT1F;RPL24;MT1G;SUCLG1;RPL26;UQCRC2;HPRT1;MT1E;SDC4;MAOA;SLC22A2;MGST3;MGST1;PIK3R1;MTMR7;SULT1A1;LDHB;UGT8;APOE;PPARGC1A;DECR1;RPL41;GLYATL1;ADHFE1;ACADSB;RPS26;RPS25;RPS28;ALDH6A1;SLC6A8;RPS27;RPS29;RPL27A;SDC1;RPS21;RPS23;COX7B;TECR;GPT;PYGL;COX7C;KHK;GYS1;MCEE;PHGDH;COX8A;CBR1;SREBF1;TPI1;NME2;SUMF2;BDH1;ALDOB;RPL10;GSTP1;RPL11;UQCR11;UQCR10;COX5B;COX5A;RPS15A;RPL14;GPC3;RPL13;CKB;RPL15;RPS2;RPS27A;RPL19;CA12;MDH1;CKM;GCGR;SCD5;COX6C;FABP3;GATM;NDUFAB1;FAU;ALDH1L1;NDUFA13;NDUFA12;COMT;COX6A1;MPC1;NUDT16;PCYT2;RPS8;RPS6;ACOT11;SDHB;COX6B1;FOLH1;UQCRB;FECH;NDUFB10;NDUFB11;GMPR;LPL;UQCRH;ACAT1;SLC9A1;EBP;PSAP;NDUFV3;NDUFV2;NDUFV1;GOT1;EPHX2;MT3;ACSF2;UQCRQ;DHCR7;GALK1;COX4I1;GADL1;ETFB;OPLAH;RPS4X;ALDH2;NAPRT;DBT;AOX1;PGM1;GPX4;DCXR;AMT;NDUFC2;NDUFC1;LSS;NDUFS8;NDUFS7;NDUFS6;NDUFS5;RPL37A;ALDH1A1;NDUFS4;SLC25A5;BC [illegible] |
| 2 | Respiratory electron transport Homo sapiens R-HSA-611105 | 1.82E-41 | 8.10E-39 | 3.09E+01 | 2.90E+03 | NDUFB9;COX7B;NDUFA13;NDUFB8;NDUFB7;NDUFB6;NDUFB10;UQCRB;NDUFA12;NDUFB11;COX4I1;NDUFB4;NDUFB3;NDUFB1;UQCR11;ETFB;UQCR10;COX6A1;COX5B;COX7C;COX5A;UQCRH;NDUFV3;NDUFV2;NDUFV1;COX8A;NDUFA8;NDUFA7;NDUFA4;NDUFA3;NDUFA1;NDUFC2;NDUFC1;COX6C;SDHB;COX6B1;NDUFS8;NDUFS7;NDUFS6;UQCRQ;NDUFS5;NDUFS4;NDUFAB1;UQCRC2 |
| 3 | Viral mRNA Translation Homo sapiens R-HSA-192823 | 3.98E-41 | 8.86E-39 | 3.24E+01 | 3.01E+03 | RPL3;RPL32;RPL10;RPL31;RPL34;RPL11;RPL36A;RPL8;RPS4X;RPS14;RPS16;RPS15A;RPS18;RPL36;RPL14;RPL35;RPL13;RPL38;RPL37;RPL15;RPS2;RPS27A;RPL39;RPL19;RPL41;RPL21;RPS8;RPL23;RPS6;RPL35A;RPL23A;RPS26;RPS25;RPS28;RPS27;RPS29;RPL27A;RPL37A;RPL24;RPL26;FAU;RPS21;RPS23 |

| | | | | | | |
|---|---|---|---|---|---|---|
| 4 | Peptide chain elongation Homo sapiens R-HSA-156902 | 3.98E-41 | 8.86E-39 | 3.24E+01 | 3.01E+03 | RPL3;RPL32;RPL10;RPL31;RPL34;RPL11;RPL36A;RPL8;RPS4X;RPS14;RPS16;RPS15A;RPS18;RPL36;RPL14;RPL35;RPL13;RPL38;RPL37;RPL15;RPS2;RPS27A;RPL39;RPL19;RPL41;RPL21;RPS8;RPL23;RPS6;RPL35A;RPL23A;RPS26;RPS25;RPS28;RPS27;RPS29;RPL27A;RPL37A;RPL24;RPL26;FAU;RPS21;RPS23 |
| 5 | Selenocysteine synthesis Homo sapiens R-HSA-2408557 | 2.90E-40 | 4.31E-38 | 3.02E+01 | 2.75E+03 | RPL3;RPL32;RPL10;RPL31;RPL34;RPL11;RPL36A;RPL8;RPS4X;RPS14;RPS16;RPS15A;RPS18;RPL36;RPL14;RPL35;RPL13;RPL38;RPL37;RPL15;RPS2;RPS27A;RPL39;RPL19;RPL41;RPL21;RPS8;RPL23;RPS6;RPL35A;RPL23A;RPS26;RPS25;RPS28;RPS27;RPS29;RPL27A;RPL37A;RPL24;RPL26;FAU;RPS21;RPS23 |
| 6 | Eukaryotic Translation Termination Homo sapiens R-HSA-72764 | 2.90E-40 | 4.31E-38 | 3.02E+01 | 2.75E+03 | RPL3;RPL32;RPL10;RPL31;RPL34;RPL11;RPL36A;RPL8;RPS4X;RPS14;RPS16;RPS15A;RPS18;RPL36;RPL14;RPL35;RPL13;RPL38;RPL37;RPL15;RPS2;RPS27A;RPL39;RPL19;RPL41;RPL21;RPS8;RPL23;RPS6;RPL35A;RPL23A;RPS26;RPS25;RPS28;RPS27;RPS29;RPL27A;RPL37A;RPL24;RPL26;FAU;RPS21;RPS23 |
| 7 | Eukaryotic Translation Elongation Homo sapiens R-HSA-156842 | 1.03E-39 | 1.15E-37 | 2.88E+01 | 2.59E+03 | RPL3;RPL32;RPL10;RPL31;RPL34;RPL11;RPL36A;RPL8;RPS4X;RPS14;RPS16;RPS15A;RPS18;RPL36;RPL14;RPL35;RPL13;RPL38;RPL37;RPL15;RPS2;RPS27A;RPL39;RPL19;RPL41;RPL21;RPS8;RPL23;RPS6;RPL35A;RPL23A;RPS26;RPS25;RPS28;RPS27;RPS29;RPL27A;RPL37A;RPL24;RPL26;FAU;RPS21;RPS23 |
| 8 | Nonsense Mediated Decay (NMD) independent of the Exon Junction Complex (EJC) Homo sapiens R-HSA-975956 | 1.03E-39 | 1.15E-37 | 2.88E+01 | 2.59E+03 | RPL3;RPL32;RPL10;RPL31;RPL34;RPL11;RPL36A;RPL8;RPS4X;RPS14;RPS16;RPS15A;RPS18;RPL36;RPL14;RPL35;RPL13;RPL38;RPL37;RPL15;RPS2;RPS27A;RPL39;RPL19;RPL41;RPL21;RPS8;RPL23;RPS6;RPL35A;RPL23A;RPS26;RPS25;RPS28;RPS27;RPS29;RPL27A;RPL37A;RPL24;RPL26;FAU;RPS21;RPS23 |
| 9 | Formation of a pool of free 40S subunits Homo sapiens R-HSA-72689 | 2.50E-39 | 2.47E-37 | 2.61E+01 | 2.32E+03 | RPL3;RPL32;RPL10;RPL31;RPL34;RPL11;RPL36A;RPL8;RPS4X;RPS14;RPS16;RPS15A;RPS18;RPL36;RPL14;RPL35;RPL13;RPL38;RPL37;RPL15;RPS2;RPS27A;RPL39;RPL19;RPL41;RPL21;RPS8;RPL23;RPS6;RPL35A;RPL23A;RPS26;RPS25;RPS28;RPS27;EIF3K;RPS29;RPL27A;RPL37A;RPL24;RPL26;FAU;RPS21;RPS23 |

| | | | | | | |
|---|---|---|---|---|---|---|
| 10 | L13a-mediated translational silencing of Ceruloplasmin expression Homo sapiens R-HSA-156827 | 5.43E-37 | 4.40E-35 | 2.19E+01 | 1.83E+03 | RPL3;RPL32;RPL10;RPL31;RPL34;RPL11;RPL36A;RPL8;RPS4X;RPS14;RPS16;RPS15A;RPS18;RPL36;RPL14;RPL35;RPL13;RPL38;RPL37;RPL15;RPS2;RPS27A;RPL39;RPL19;RPL41;RPL21;RPS8;RPL23;RPS6;RPL35A;RPL23A;RPS26;RPS25;RPS28;RPS27;EIF3K;RPS29;RPL27A;RPL37A;RPL24;RPL26;FAU;RPS21;RPS23 |
| 11 | 3' -UTR-mediated translational regulation Homo sapiens R-HSA-157279 | 5.43E-37 | 4.40E-35 | 2.19E+01 | 1.83E+03 | RPL3;RPL32;RPL10;RPL31;RPL34;RPL11;RPL36A;RPL8;RPS4X;RPS14;RPS16;RPS15A;RPS18;RPL36;RPL14;RPL35;RPL13;RPL38;RPL37;RPL15;RPS2;RPS27A;RPL39;RPL19;RPL41;RPL21;RPS8;RPL23;RPS6;RPL35A;RPL23A;RPS26;RPS25;RPS28;RPS27;EIF3K;RPS29;RPL27A;RPL37A;RPL24;RPL26;FAU;RPS21;RPS23 |
| 12 | SRP-dependent cotranslational protein targeting to membrane Homo sapiens R-HSA-1799339 | 8.94E-37 | 6.12E-35 | 2.16E+01 | 1.79E+03 | RPL3;RPL32;RPL10;RPL31;RPL34;RPL11;RPL36A;RPL8;RPS4X;RPS14;RPS16;RPS15A;RPS18;RPL36;RPL14;RPL35;RPL13;RPL38;RPL37;RPL15;RPS2;RPS27A;RPL39;RPL19;RPL41;RPL21;RPS8;RPL23;RPS6;SSR3;RPL35A;RPL23A;RPS26;RPS25;RPS28;RPS27;RPS29;RPL27A;RPL37A;RPL24;RPL26;FAU;RPS21;RPS23 |
| 13 | GTP hydrolysis and joining of the 60S ribosomal subunit Homo sapiens R-HSA-72706 | 8.94E-37 | 6.12E-35 | 2.16E+01 | 1.79E+03 | RPL3;RPL32;RPL10;RPL31;RPL34;RPL11;RPL36A;RPL8;RPS4X;RPS14;RPS16;RPS15A;RPS18;RPL36;RPL14;RPL35;RPL13;RPL38;RPL37;RPL15;RPS2;RPS27A;RPL39;RPL19;RPL41;RPL21;RPS8;RPL23;RPS6;RPL35A;RPL23A;RPS26;RPS25;RPS28;RPS27;EIF3K;RPS29;RPL27A;RPL37A;RPL24;RPL26;FAU;RPS21;RPS23 |
| 14 | Respiratory electron transport, ATP synthesis by chemiosmotic coupling, and heat production by uncoupling proteins. Homo sapiens R-HSA-163200 | 2.38E-36 | 1.51E-34 | 2.09E+01 | 1.71E+03 | NDUFB9;NDUFA13;COX7B;NDUFB8;NDUFB7;NDUFB10;UQCRB;NDUFB6;NDUFA12;NDUFB11;COX4I1;NDUFB4;NDUFB3;NDUFB1;UQCR11;ETFB;UQCR10;COX6A1;COX5B;COX7C;COX5A;UQCRH;NDUFV3;NDUFV2;NDUFV1;COX8A;NDUFA8;NDUFA7;NDUFA4;NDUFA3;NDUFA1;NDUFC2;NDUFC1;COX6C;SDHB;COX6B1;NDUFS8;NDUFS7;UQCRQ;NDUFS6;NDUFS5;NDUFS4;NDUFAB1;UQCRC2 |
| 15 | Nonsense-Mediated Decay (NMD) Homo sapiens R-HSA-927802 | 1.17E-35 | 6.52E-34 | 2.10E+01 | 1.69E+03 | RPL3;RPL32;RPL10;RPL31;RPL34;RPL11;RPL36A;RPL8;RPS4X;RPS14;RPS16;RPS15A;RPS18;RPL36;RPL14;RPL35;RPL13;RPL38;RPL37;RPL15;RPS2;RPS27A;RPL39;RPL19;RPL41;RPL21;RPS8;RPL23;RPS6;RPL35A;RPL23A;RPS26;RPS25;RPS28;RPS27;RPS29;RPL27A;RPL37A;RPL24;RPL26;FAU;RPS21;RPS23 |

| | | | | | | |
|---|---|---|---|---|---|---|
| 16 | Nonsense Mediated Decay (NMD) enhanced by the Exon Junction Complex (EJC) Homo sapiens R-HSA-975957 | 1.17E-35 | 6.52E-34 | 2.10E+01 | 1.69E+03 | RPL3;RPL32;RPL10;RPL31;RPL34;RPL11;RPL36A;RPL8;RPS4X;RPS14;RPS16;RPS15A;RPS18;RPL36;RPL14;RPL35;RPL13;RPL38;RPL37;RPL15;RPS2;RPS27A;RPL39;RPL19;RPL41;RPL21;RPS8;RPL23;RPS6;RPL35A;RPL23A;RPS26;RPS25;RPS28;RPS27;RPS29;RPL27A;RPL37A;RPL24;RPL26;FAU;RPS21;RPS23 |
| 17 | Cap-dependent Translation Initiation Homo sapiens R-HSA-72737 | 2.47E-35 | 1.22E-33 | 1.94E+01 | 1.55E+03 | RPL3;RPL32;RPL10;RPL31;RPL34;RPL11;RPL36A;RPL8;RPS4X;RPS14;RPS16;RPS15A;RPS18;RPL36;RPL14;RPL35;RPL13;RPL38;RPL37;RPL15;RPS2;RPS27A;RPL39;RPL19;RPL41;RPL21;RPS8;RPL23;RPS6;RPL35A;RPL23A;RPS26;RPS25;RPS28;RPS27;EIF3K;RPS29;RPL27A;RPL37A;RPL24;RPL26;FAU;RPS21;RPS23 |
| 18 | Eukaryotic Translation Initiation Homo sapiens R-HSA-72613 | 2.47E-35 | 1.22E-33 | 1.94E+01 | 1.55E+03 | RPL3;RPL32;RPL10;RPL31;RPL34;RPL11;RPL36A;RPL8;RPS4X;RPS14;RPS16;RPS15A;RPS18;RPL36;RPL14;RPL35;RPL13;RPL38;RPL37;RPL15;RPS2;RPS27A;RPL39;RPL19;RPL41;RPL21;RPS8;RPL23;RPS6;RPL35A;RPL23A;RPS26;RPS25;RPS28;RPS27;EIF3K;RPS29;RPL27A;RPL37A;RPL24;RPL26;FAU;RPS21;RPS23 |
| 19 | The citric acid (TCA) cycle and respiratory electron transport Homo sapiens R-HSA-1428517 | 1.03E-34 | 4.82E-33 | 1.46E+01 | 1.15E+03 | NDUFA13;COX7B;NDUFA12;COX4I1;ETFB;COX6A1;COX7C;MPC1;BSG;COX8A;NDUFC2;NDUFC1;SDHB;COX6B1;NDUFS8;NDUFS7;NDUFS6;NDUFS5;NDUFS4;SUCLG1;UQCRC2;NDUFB9;NDUFB8;NDUFB7;NDUFB10;UQCRB;NDUFB6;NDUFB11;NDUFB4;NDUFB3;NDUFB1;UQCR11;UQCR10;COX5B;COX5A;UQCRH;LDHB;NDUFV3;NDUFV2;NDUFV1;NDUFA8;NDUFA7;NDUFA4;ADHFE1;NDUFA3;NDUFA1;COX6C;UQCRQ;NDUFAB1 |
| 20 | Selenoamino acid metabolism Homo sapiens R-HSA-2408522 | 1.23E-34 | 5.49E-33 | 1.95E+01 | 1.52E+03 | RPL3;RPL32;RPL10;RPL31;RPL34;RPL11;RPL36A;RPL8;RPS4X;RPS14;RPS16;RPS15A;RPS18;RPL36;RPL14;RPL35;RPL13;RPL38;RPL37;RPL15;RPS2;RPS27A;RPL39;RPL19;RPL41;RPL21;RPS8;RPL23;RPS6;RPL35A;RPL23A;RPS26;RPS25;RPS28;RPS27;RPS29;RPL27A;RPL37A;RPL24;RPL26;FAU;RPS21;RPS23 |

| | | | | | | |
|---|---|---|---|---|---|---|
| 21 | Influenza Viral RNA Transcription and Replication Homo sapiens R-HSA-168273 | 8.76E-33 | 3.71E-31 | 1.62E+01 | 1.19E+03 | RPL3;RPL32;RPL10;RPL31;RPL34;RPL11;RPL36A;RPL8;RPS4X;RPS14;RPS16;RPS15A;RPS18;RPL36;RPL14;RPL35;RPL13;RPL38;RPL37;RPL15;RPS2;RPS27A;RPL39;RPL19;POLR2L;RPL41;RPL21;RPS8;RPL23;RPS6;RPL35A;RPL23A;RPS26;RPS25;RPS28;RPS27;RPS29;RPL27A;RPL37A;RPL24;RPL26;FAU;RPS21;RPS23 |
| 22 | Influenza Life Cycle Homo sapiens R-HSA-168255 | 1.14E-32 | 4.62E-31 | 1.53E+01 | 1.12E+03 | RPL3;RPL32;RPL10;RPL31;RPL34;RPL11;RPL36A;RPL8;RPS4X;RPS14;RPS16;RPS15A;RPS18;RPL36;RPL14;RPL35;RPL13;RPL38;RPL37;RPL15;RPS2;RPS27A;RPL39;RPL19;POLR2L;RPL41;RPL21;RPS8;RPL23;RPS6;RPL35A;RPL23A;RPS26;RPS25;RPS28;RPS27;RPS29;RPL27A;RPL37A;RPL24;RPL26;CALR;FAU;RPS21;RPS23 |
| 23 | Metabolism of amino acids and derivatives Homo sapiens R-HSA-71291 | 3.60E-32 | 1.39E-30 | 7.77E+00 | 5.63E+02 | RPL3;RPL32;RPL31;RPL34;GADL1;GPT;RPL8;RPS4X;RPS14;RPS16;RPS18;DBT;RPL36;RPL35;RPL38;PHGDH;RPL37;RPL39;RPL21;RPS8;RPL23;MCCC1;RPS6;AMT;FOLH1;RPL37A;RPL24;RPL26;BCAT1;BCAT2;RPL10;RPL11;RPL36A;PPM1K;HSD17B10;ACAT1;CKMT2;RPS15A;RPL14;RPL13;PCBD1;RPL15;CKB;RPS2;RPS27A;RPL19;RPL41;CKM;GOT1;RPL35A;RPL23A;ACADSB;ASS1;RPS26;RPS25;RPS28;ALDH6A1;GATM;RPS27;SLC6A8;RPS29;RPL27A;NDUFAB1;FAU;RPS21;RPS23 |
| 24 | Influenza Infection Homo sapiens R-HSA-168254 | 5.38E-31 | 2.00E-29 | 1.36E+01 | 9.49E+02 | RPL3;RPL32;RPL10;RPL31;RPL34;RPL11;RPL36A;RPL8;RPS4X;RPS14;RPS16;RPS15A;RPS18;RPL36;RPL14;RPL35;RPL13;RPL38;RPL37;RPL15;RPS2;RPS27A;RPL39;RPL19;POLR2L;RPL41;RPL21;RPS8;RPL23;RPS6;RPL35A;RPL23A;RPS26;RPS25;RPS28;RPS27;RPS29;RPL27A;RPL37A;RPL24;RPL26;CALR;FAU;RPS21;RPS23 |
| 25 | Translation Homo sapiens R-HSA-72766 | 1.99E-30 | 7.09E-29 | 1.31E+01 | 8.96E+02 | RPL3;RPL32;RPL10;RPL31;RPL34;RPL11;RPL36A;RPL8;RPS4X;RPS14;RPS16;RPS15A;RPS18;RPL36;RPL14;RPL35;RPL13;RPL38;RPL37;RPL15;RPS2;RPS27A;RPL39;RPL19;RPL41;RPL21;RPS8;RPL23;RPS6;SSR3;RPL35A;RPL23A;RPS26;RPS25;RPS28;RPS27;EIF3K;RPS29;RPL27A;RPL37A;RPL24;RPL26;FAU;RPS21;RPS23 |

| | | | | | | |
|---|---|---|---|---|---|---|
| 26 | Major pathway of rRNA processing in the nucleolus Homo sapiens R-HSA-6791226 | 2.32E-26 | 7.94E-25 | 1.07E+01 | 6.34E+02 | RPL3;RPL32;RPL10;RPL31;RPL34;RPL11;RPL36A;RPL8;RPS4X;RPS14;RPS16;RPS15A;RPS18;RPL36;RPL14;RPL35;RPL13;RPL38;RPL37;RPL15;RPS2;RPS27A;RPL39;RPL19;RPL41;RPL21;RPS8;RPL23;RPS6;RPL35A;RPL23A;RPS26;RPS25;RPS28;RPS27;RPS29;RPL27A;RPL37A;RPL24;RPL26;FAU;RPS21;RPS23 |
| 27 | Complex I biogenesis Homo sapiens R-HSA-6799198 | 7.27E-26 | 2.40E-24 | 3.40E+01 | 1.97E+03 | NDUFB9;NDUFA13;NDUFB8;NDUFB7;NDUFB6;NDUFB10;NDUFA12;NDUFB11;NDUFB4;NDUFB3;NDUFB1;NDUFV3;NDUFV2;NDUFV1;NDUFA8;NDUFA7;NDUFA3;NDUFA1;NDUFC2;NDUFC1;NDUFS8;NDUFS7;NDUFS6;NDUFS5;NDUFS4;NDUFAB1 |
| 28 | rRNA processing Homo sapiens R-HSA-72312 | 8.18E-26 | 2.60E-24 | 9.95E+00 | 5.75E+02 | RPL3;RPL32;RPL10;RPL31;RPL34;RPL11;RPL36A;RPL8;HSD17B10;RPS4X;RPS14;RPS16;RPS15A;RPS18;RPL36;RPL14;RPL35;RPL13;RPL38;RPL37;RPL15;RPS2;RPS27A;RPL39;RPL19;RPL41;RPL21;RPS8;RPL23;RPS6;RPL35A;RPL23A;RPS26;RPS25;RPS28;RPS27;RPS29;RPL27A;RPL37A;RPL24;RPL26;FAU;RPS21;RPS23 |
| 29 | Infectious disease Homo sapiens R-HSA-5663205 | 1.22E-17 | 3.75E-16 | 5.04E+00 | 1.96E+02 | RPL3;RPL32;RPL31;RPL34;RPL8;RPS4X;RPS14;RPS16;RPS18;RPL36;RPL35;RPL38;RPL37;B2M;RPL39;RPL21;RPS8;RPL23;RPS6;RPL37A;RPL24;RPL26;SLC25A5;SLC25A4;RPL10;RPL11;RPL36A;RPS15A;RPL14;RPL13;RPL15;RPS2;RPS27A;RPL19;POLR2L;RPL41;RPL35A;RPL23A;GTF2H2;RPS26;RPS25;RPS28;RPS27;RPS29;RPL27A;CALR;FAU;RPS21;RPS23 |
| 30 | Formation of the ternary complex, and subsequently, the 43S complex Homo sapiens R-HSA-72695 | 1.42E-14 | 4.21E-13 | 1.67E+01 | 5.33E+02 | RPS8;RPS6;RPS4X;RPS26;RPS14;RPS25;RPS28;RPS16;RPS15A;RPS27;EIF3K;RPS18;RPS29;RPS2;FAU;RPS27A;RPS21;RPS23 |
| 31 | Ribosomal scanning and start codon recognition Homo sapiens R-HSA-72702 | 1.96E-13 | 5.45E-12 | 1.37E+01 | 4.01E+02 | RPS8;RPS6;RPS4X;RPS26;RPS14;RPS25;RPS28;RPS16;RPS15A;RPS27;EIF3K;RPS18;RPS29;RPS2;FAU;RPS27A;RPS21;RPS23 |
| 32 | Translation initiation complex formation Homo sapiens R-HSA-72649 | 1.96E-13 | 5.45E-12 | 1.37E+01 | 4.01E+02 | RPS8;RPS6;RPS4X;RPS26;RPS14;RPS25;RPS28;RPS16;RPS15A;RPS27;EIF3K;RPS18;RPS29;RPS2;FAU;RPS27A;RPS21;RPS23 |

| | | | | | | |
|---|---|---|---|---|---|---|
| 33 | Activation of the mRNA upon binding of the cap-binding complex and eIFs, and subsequent binding to 43S Homo sapiens R-HSA-72662 | 2.76E-13 | 7.43E-12 | 1.34E+01 | 3.86E+02 | RPS8;RPS6;RPS4X;RPS26;RPS14;RPS25;RPS28;RPS16;RPS15A;RPS27;EIF3K;RPS18;RPS29;RPS2;FAU;RPS27A;RPS21;RPS23 |
| 34 | Disease Homo sapiens R-HSA-1643685 | 4.21E-13 | 1.10E-11 | 3.05E+00 | 8.69E+01 | RPL3;RPL32;RPL31;RPL34;RPL8;RPS4X;RPS14;GYS1;RPS16;FGF9;RPS18;RPL36;RPL35;RPL38;RPL37;THSD7A;MUC15;RPL39;B2M;HRAS;RPL21;RPS8;RPL23;RPS6;MCCC1;RPL37A;RPL24;RPL26;SLC25A5;SLC25A4;NOTCH3;RPL10;SDC4;RPL11;RPL36A;PIK3R1;FOXO3;MUC1;RPS15A;ERBB4;RPL14;GPC3;RPL13;HES1;RPL15;RPS2;RPS27A;RPL19;POLR2L;RPL41;EGF;RPL35A;RPL23A;GTF2H2;RPS26;RPS25;RPS28;RPS27;RPS29;RPL27A;SDC1;CALR;FAU;RPS21;RPS23 |
| 35 | Metabolism of proteins Homo sapiens R-HSA-392499 | 1.07E-09 | 2.73E-08 | 2.32E+00 | 4.79E+01 | GALNT14;RPL3;RPL32;GALNT16;RPL31;RPL34;RPL8;HERPUD1;RPS4X;RPS14;RPS16;RPS18;RPL36;RPL35;RPL38;RPL37;THSD7A;MUC15;RPL39;B2M;CTSC;RGS6;DYNC2H1;TRAPPC2L;RPL21;RPS8;RPL23;SSR3;RPS6;PAM16;SUMF2;TUBB2B;TUBB2A;RPL37A;RPL24;RPL26;SLC25A4;RPL10;RPL11;RPL36A;HSP90B1;HSPD1;CST3;MUC1;RPS15A;GMDS;LDHD;RPL14;RPL13;RPL15;RPS2;RPS27A;RPL19;DNAJC19;RPL41;XBP1;PMM1;HSPA5;TRAPPC5;RPL35A;RPL23A;RPS26;RPS25;RPS28;RPS27;EIF3K;RPS29;RPL27A;MSRB2;TOMM7;CALR;FAU;FOLR1;RPS21;RPS23 |
| 36 | Branched-chain amino acid catabolism Homo sapiens R-HSA-70895 | 2.63E-08 | 6.51E-07 | 1.88E+01 | 3.29E+02 | ALDH6A1;MCCC1;DBT;PPM1K;BCAT1;ACADSB;HSD17B10;ACAT1;BCAT2 |
| 37 | Response to metal ions Homo sapiens R-HSA-5660526 | 5.43E-07 | 1.27E-05 | 3.50E+01 | 5.05E+02 | MT2A;MT1F;MT1G;MT1X;MT3;MT1E |
| 38 | Metallothioneins bind metals Homo sapiens R-HSA-5661231 | 5.43E-07 | 1.27E-05 | 3.50E+01 | 5.05E+02 | MT2A;MT1F;MT1G;MT1X;MT3;MT1E |

| | | | | | | |
|---|---|---|---|---|---|---|
| 39 | Creatine metabolism Homo sapiens R-HSA-71288 | 8.98E-06 | 2.05E-04 | 2.92E+01 | 3.39E+02 | CKMT2;GATM;SLC6A8;CKM;CKB |
| 40 | TP53 Regulates Metabolic Genes Homo sapiens R-HSA-5628897 | 2.19E-05 | 4.53E-04 | 4.89E+00 | 5.25E+01 | COX8A;PRDX2;COX7B;PRDX5;NDUFA4;COX4I1;COX6A1;COX5B;COX6C;COX7C;COX5A;COX6B1 |
| 41 | Mitochondrial translation initiation Homo sapiens R-HSA-5368286 | 2.19E-05 | 4.53E-04 | 4.89E+00 | 5.25E+01 | MRPL41;MRPS28;MRPS24;MRPS18B;MRPL16;MRPL36;MRPL15;MRPL12;MRPL23;MRPL13;MRPL24;MRPL33 |
| 42 | Mitochondrial translation termination Homo sapiens R-HSA-5419276 | 2.19E-05 | 4.53E-04 | 4.89E+00 | 5.25E+01 | MRPL41;MRPS28;MRPS24;MRPS18B;MRPL16;MRPL36;MRPL15;MRPL12;MRPL23;MRPL13;MRPL24;MRPL33 |
| 43 | Mitochondrial translation elongation Homo sapiens R-HSA-5389840 | 2.19E-05 | 4.53E-04 | 4.89E+00 | 5.25E+01 | MRPL41;MRPS28;MRPS24;MRPS18B;MRPL16;MRPL36;MRPL15;MRPL12;MRPL23;MRPL13;MRPL24;MRPL33 |
| 44 | Mitochondrial translation Homo sapiens R-HSA-5368287 | 4.42E-05 | 8.94E-04 | 4.52E+00 | 4.53E+01 | MRPL41;MRPS28;MRPS24;MRPS18B;MRPL16;MRPL36;MRPL15;MRPL12;MRPL23;MRPL13;MRPL24;MRPL33 |
| 45 | Intrinsic Pathway of Fibrin Clot Formation Homo sapiens R-HSA-140837 | 6.41E-05 | 1.27E-03 | 1.09E+01 | 1.06E+02 | SERPINE2;C1QBP;F11;SERPING1;PRCP;KNG1 |
| 46 | Neurotransmitter Clearance In The Synaptic Cleft Homo sapiens R-HSA-112311 | 7.76E-05 | 1.50E-03 | 2.91E+01 | 2.75E+02 | ALDH2;MAOA;SLC22A2;COMT |
| 47 | Detoxification of Reactive Oxygen Species Homo sapiens R-HSA-3299685 | 9.00E-05 | 1.70E-03 | 7.86E+00 | 7.33E+01 | PRDX3;PRDX2;PRDX5;GSTP1;NOX4;PRDX6;TXN2 |
| 48 | ATF6-alpha activates chaperone genes Homo sapiens R-HSA-381183 | 2.21E-04 | 4.09E-03 | 1.94E+01 | 1.63E+02 | XBP1;HSPA5;CALR;HSP90B1 |

| | | | | | | |
|---|---|---|---|---|---|---|
| 49 | Formation of Fibrin Clot (Clotting Cascade) Homo sapiens R-HSA-140877 | 2.72E-04 | 4.94E-03 | 6.39E+00 | 5.24E+01 | SERPINE2;C1QBP;F11;SERPING1;PRCP;KNG1;PF4V1 |
| 50 | Glucose metabolism Homo sapiens R-HSA-70326 | 2.92E-04 | 5.19E-03 | 4.24E+00 | 3.45E+01 | GYS1;TPI1;GOT1;MDH1;PYGL;ALDOB;RPS27A;GAPDH;PFKM;PGM1 |
| 51 | Scavenging by Class A Receptors Homo sapiens R-HSA-3000480 | 3.22E-04 | 5.63E-03 | 1.04E+01 | 8.37E+01 | FTH1;APOE;CALR;HSP90B1;FTL |
| 52 | ATF6-alpha activates chaperones Homo sapiens R-HSA-381033 | 4.93E-04 | 8.43E-03 | 1.46E+01 | 1.11E+02 | XBP1;HSPA5;CALR;HSP90B1 |
| 53 | Fructose metabolism Homo sapiens R-HSA-5652084 | 6.88E-04 | 1.16E-02 | 2.91E+01 | 2.12E+02 | ALDH1A1;ALDOB;KHK |
| 54 | Amino acid synthesis and interconversion (transamination) Homo sapiens R-HSA-70614 | 1.25E-03 | 2.06E-02 | 7.28E+00 | 4.87E+01 | FOLH1;GOT1;GADL1;GPT;PHGDH |
| 55 | Mitochondrial Fatty Acid Beta-Oxidation Homo sapiens R-HSA-77289 | 2.07E-03 | 3.36E-02 | 8.95E+00 | 5.53E+01 | MCEE;ECI1;ACAD11;DECR1 |
| 56 | Gene Expression Homo sapiens R-HSA-74160 | 2.87E-03 | 4.51E-02 | 1.44E+00 | 8.46E+00 | COX7B;RPL3;RPL32;RPL31;RPL34;COX4I1;HSPB1;RPL8;COX6A1;COX7C;RPS4X;RPS14;RPS16;RPS18;ZNF208;RPL36;RPL35;RPL38;RPL37;RPL39;COX8A;RPL21;RPS8;RPL23;SSR3;RPS6;USP2;COX6B1;ZNF439;RPL37A;SRSF2;RPL24;SRSF3;RPL26;SRSF5;SRSF6;NOTCH3;RPL10;RPL11;SRSF1;RPL36A;RNPC3;COX5B;HSD17B10;COX5A;PRDX2;PRDX5;RPS15A;PCBP2;RPL14;RPL13;RPL15;RPS2;RPS27A;RPL19;POLR2L;HSPA8;RPL41;FUS;NDUFA4;RPL35A;RPL23A;GTF2H2;COX6C;RPS26;RPS25;RPS28;RPS27;TP53I3;EIF3K;RPS29;RPL27A;FAU;RPS21;RPS23 |

| | | | | | | |
|---|---|---|---|---|---|---|
| 57 | Biological oxidations Homo sapiens R-HSA-211859 | 2.89E-03 | 4.51E-02 | 2.39E+00 | 1.40E+01 | GSTK1;GLYATL1;MAOA;MGST3;GSTP1;MGST1;CMBL;COMT;OPLAH;BPHL;SULT1A1;ALDH2;MAT2A;ALDH1A1;ACSS1 |
| 58 | Constitutive Signaling by Ligand-Responsive EGFR Cancer Variants Homo sapiens R-HSA-1236382 | 3.20E-03 | 4.75E-02 | 7.76E+00 | 4.46E+01 | EGF;PIK3R1;RPS27A;HRAS |
| 59 | Signaling by Ligand-Responsive EGFR Variants in Cancer Homo sapiens R-HSA-5637815 | 3.20E-03 | 4.75E-02 | 7.76E+00 | 4.46E+01 | EGF;PIK3R1;RPS27A;HRAS |
| 60 | Signaling by EGFR in Cancer Homo sapiens R-HSA-1643713 | 3.20E-03 | 4.75E-02 | 7.76E+00 | 4.46E+01 | EGF;PIK3R1;RPS27A;HRAS |
| 61 | Gluconeogenesis Homo sapiens R-HSA-70263 | 3.90E-03 | 5.69E-02 | 5.39E+00 | 2.99E+01 | TPI1;GOT1;MDH1;ALDOB;GAPDH |
| 62 | Pyruvate metabolism and Citric Acid (TCA) cycle Homo sapiens R-HSA-71406 | 5.05E-03 | 7.25E-02 | 4.16E+00 | 2.20E+01 | LDHB;MPC1;ADHFE1;BSG;SUCLG1;SDHB |
| 63 | Phase II conjugation Homo sapiens R-HSA-156580 | 6.27E-03 | 8.78E-02 | 2.89E+00 | 1.46E+01 | SULT1A1;GSTK1;GLYATL1;MAT2A;MGST3;GSTP1;MGST1;COMT;OPLAH |
| 64 | Ethanol oxidation Homo sapiens R-HSA-71384 | 6.51E-03 | 8.78E-02 | 9.69E+00 | 4.88E+01 | ALDH2;ALDH1A1;ACSS1 |
| 65 | Defective EXT2 causes exostoses 2 Homo sapiens R-HSA-3656237 | 6.51E-03 | 8.78E-02 | 9.69E+00 | 4.88E+01 | SDC4;GPC3;SDC1 |
| 66 | Defective EXT1 causes exostoses 1, TRPS2 and CHDS Homo sapiens R-HSA-3656253 | 6.51E-03 | 8.78E-02 | 9.69E+00 | 4.88E+01 | SDC4;GPC3;SDC1 |
| 67 | Metabolism of carbohydrates Homo sapiens R-HSA-71387 | 6.82E-03 | 9.06E-02 | 2.00E+00 | 9.98E+00 | TPI1;SDC4;MDH1;GOT1;DCXR;PYGL;KHK;SLC9A1;GYS1;ALDH1A1;GPC3;SDC1;ALDOB;RPS27A;GAPDH;PFKM;PGM1;GALK1 |

| | | | | | | |
|---|---|---|---|---|---|---|
| 68 | Glutathione conjugation Homo sapiens R-HSA-156590 | 8.26E-03 | 1.07E-01 | 4.41E+00 | 2.12E+01 | GSTK1;GSTP1;MGST3;MGST1;OPLAH |
| 69 | Dissolution of Fibrin Clot Homo sapiens R-HSA-75205 | 8.26E-03 | 1.07E-01 | 8.72E+00 | 4.18E+01 | SERPINE2;PLAU;HRG |
| 70 | Mitochondrial protein import Homo sapiens R-HSA-1268020 | 8.99E-03 | 1.14E-01 | 3.64E+00 | 1.72E+01 | DNAJC19;PAM16;LDHD;TOMM7;SLC25A4;HSPD1 |
| 71 | Bile salt and organic anion SLC transporters Homo sapiens R-HSA-425471 | 1.03E-02 | 1.29E-01 | 7.92E+00 | 3.63E+01 | SLC47A1;BSG;SLC16A7 |
| 72 | Galactose catabolism Homo sapiens R-HSA-70370 | 1.04E-02 | 1.29E-01 | 1.93E+01 | 8.83E+01 | PGM1;GALK1 |
| 73 | Metabolism of vitamins and cofactors Homo sapiens R-HSA-196854 | 1.10E-02 | 1.34E-01 | 2.35E+00 | 1.06E+01 | CYB5A;ALDH1L1;PDXK;SDC4;NAPRT;MCCC1;GPC3;LPL;SDC1;AOX1;APOE |
| 74 | Glycogen synthesis Homo sapiens R-HSA-3322077 | 1.25E-02 | 1.45E-01 | 7.26E+00 | 3.18E+01 | GYS1;RPS27A;PGM1 |
| 75 | Signaling by EGFRvIII in Cancer Homo sapiens R-HSA-5637812 | 1.25E-02 | 1.45E-01 | 7.26E+00 | 3.18E+01 | EGF;PIK3R1;HRAS |
| 76 | Constitutive Signaling by EGFRvIII Homo sapiens R-HSA-5637810 | 1.25E-02 | 1.45E-01 | 7.26E+00 | 3.18E+01 | EGF;PIK3R1;HRAS |
| 77 | Retinoid metabolism and transport Homo sapiens R-HSA-975634 | 1.26E-02 | 1.45E-01 | 3.93E+00 | 1.72E+01 | SDC4;GPC3;LPL;SDC1;APOE |
| 78 | PI3K events in ERBB2 signaling Homo sapiens R-HSA-1963642 | 1.50E-02 | 1.68E-01 | 6.70E+00 | 2.82E+01 | EGF;ERBB4;PIK3R1 |

| | | | | | | |
|---|---|---|---|---|---|---|
| 79 | GRB2 events in ERBB2 signaling Homo sapiens R-HSA-1963640 | 1.50E-02 | 1.68E-01 | 6.70E+00 | 2.82E+01 | EGF;ERBB4;HRAS |
| 80 | Proton-coupled monocarboxylate transport Homo sapiens R-HSA-433692 | 1.53E-02 | 1.68E-01 | 1.45E+01 | 6.07E+01 | BSG;SLC16A7 |
| 81 | mitochondrial fatty acid beta-oxidation of unsaturated fatty acids Homo sapiens R-HSA-77288 | 1.53E-02 | 1.68E-01 | 1.45E+01 | 6.07E+01 | ECI1;DECR1 |
| 82 | Signaling by ERBB2 Homo sapiens R-HSA-1227986 | 1.66E-02 | 1.80E-01 | 3.64E+00 | 1.49E+01 | ERBB4;EGF;PIK3R1;RPS27A;HRAS |
| 83 | Chylomicron-mediated lipid transport Homo sapiens R-HSA-174800 | 1.78E-02 | 1.91E-01 | 6.22E+00 | 2.51E+01 | LPL;SDC1;APOE |
| 84 | SHC1 events in ERBB2 signaling Homo sapiens R-HSA-1250196 | 2.08E-02 | 2.19E-01 | 5.81E+00 | 2.25E+01 | EGF;ERBB4;HRAS |
| 85 | Synthesis of Ketone Bodies Homo sapiens R-HSA-77111 | 2.09E-02 | 2.19E-01 | 1.16E+01 | 4.49E+01 | BDH1;ACAT1 |
| 86 | Glycolysis Homo sapiens R-HSA-70171 | 2.11E-02 | 2.19E-01 | 4.15E+00 | 1.60E+01 | TPI1;ALDOB;PFKM;GAPDH |
| 87 | Metabolism of polyamines Homo sapiens R-HSA-351202 | 2.22E-02 | 2.27E-01 | 2.65E+00 | 1.01E+01 | CKMT2;GATM;SLC6A8;CKM;GOT1;CKB;ASS1 |
| 88 | Metabolism of lipids and lipoproteins Homo sapiens R-HSA-556833 | 2.26E-02 | 2.28E-01 | 1.50E+00 | 5.69E+00 | ECI1;ECI2;TECR;LPL;PIK3R1;AGPAT2;MTMR7;ACAT1;UGT8;MCEE;EBP;PSAP;APOE;PTGDS;ACAD11;PPARGC1A;DECR1;CBR1;SREBF1;PCYT2;GPX4;EPHX2;SCD5;ACOT11;LSS;SUMF2;ACSF2;FABP3;BDH1;SDC1;CHPT1;DHCR7 |
| 89 | mRNA Splicing Homo sapiens R-HSA-72172 | 2.28E-02 | 2.28E-01 | 2.18E+00 | 8.23E+00 | HSPA8;FUS;PCBP2;SRSF1;SRSF2;SRSF3;SRSF5;SRSF6;RNPC3;POLR2L |

| | | | | | | |
|---|---|---|---|---|---|---|
| 90 | Defective B3GALT6 causes EDSP2 and SEMDJL1 Homo sapiens R-HSA-4420332 | 2.41E-02 | 2.31E-01 | 5.45E+00 | 2.03E+01 | SDC4;GPC3;SDC1 |
| 91 | Defective B4GALT7 causes EDS, progeroid type Homo sapiens R-HSA-3560783 | 2.41E-02 | 2.31E-01 | 5.45E+00 | 2.03E+01 | SDC4;GPC3;SDC1 |
| 92 | Defective B3GAT3 causes JDSSDHD Homo sapiens R-HSA-3560801 | 2.41E-02 | 2.31E-01 | 5.45E+00 | 2.03E+01 | SDC4;GPC3;SDC1 |
| 93 | Signaling by PTK6 Homo sapiens R-HSA-8848021 | 2.42E-02 | 2.31E-01 | 2.86E+00 | 1.07E+01 | KHDRBS3;CCND1;ERBB4;EGF;RPS27A;HRAS |
| 94 | Platelet degranulation Homo sapiens R-HSA-114608 | 2.44E-02 | 2.31E-01 | 2.40E+00 | 8.92E+00 | LGALS3BP;HSPA5;EGF;APLP2;PSAP;SERPING1;HRG;KNG1 |
| 95 | Metabolism of fat-soluble vitamins Homo sapiens R-HSA-6806667 | 2.72E-02 | 2.45E-01 | 3.16E+00 | 1.14E+01 | SDC4;GPC3;LPL;SDC1;APOE |
| 96 | mRNA 3'-end processing Homo sapiens R-HSA-72187 | 2.72E-02 | 2.45E-01 | 3.16E+00 | 1.14E+01 | SRSF1;SRSF2;SRSF3;SRSF5;SRSF6 |
| 97 | Post-Elongation Processing of Intron-Containing pre-mRNA Homo sapiens R-HSA-112296 | 2.72E-02 | 2.45E-01 | 3.16E+00 | 1.14E+01 | SRSF1;SRSF2;SRSF3;SRSF5;SRSF6 |
| 98 | Downregulation of ERBB4 signaling Homo sapiens R-HSA-1253288 | 2.73E-02 | 2.45E-01 | 9.67E+00 | 3.48E+01 | ERBB4;RPS27A |
| 99 | Ketone body metabolism Homo sapiens R-HSA-74182 | 2.73E-02 | 2.45E-01 | 9.67E+00 | 3.48E+01 | BDH1;ACAT1 |
| 100 | Diseases of glycosylation Homo sapiens R-HSA-3781865 | 2.78E-02 | 2.47E-01 | 2.52E+00 | 9.02E+00 | NOTCH3;MUC1;SDC4;GPC3;SDC1;MUC15;THSD7A |

| | | | | | | |
|---|---|---|---|---|---|---|
| 101 | mRNA Splicing - Minor Pathway Homo sapiens R-HSA-72165 | 2.93E-02 | 2.58E-01 | 3.09E+00 | 1.09E+01 | SRSF1;SRSF2;SRSF6;RNPC3;POLR2L |
| 102 | Fatty acid, triacylglycerol, and ketone body metabolism Homo sapiens R-HSA-535734 | 3.09E-02 | 2.69E-01 | 1.86E+00 | 6.47E+00 | SREBF1;ECI1;TECR;SCD5;ACOT11;AGPAT2;ACSF2;ACAT1;MCEE;BDH1;ACAD11;PPARGC1A;DECR1 |
| 103 | Response to elevated platelet cytosolic Ca2+ Homo sapiens R-HSA-76005 | 3.11E-02 | 2.69E-01 | 2.29E+00 | 7.93E+00 | LGALS3BP;HSPA5;EGF;APLP2;PSAP;SERPING1;HRG;KNG1 |
| 104 | HS-GAG degradation Homo sapiens R-HSA-2024096 | 3.15E-02 | 2.70E-01 | 4.84E+00 | 1.67E+01 | SDC4;GPC3;SDC1 |
| 105 | Signaling by FGFR3 point mutants in cancer Homo sapiens R-HSA-8853338 | 3.56E-02 | 2.98E-01 | 4.59E+00 | 1.53E+01 | FGF9;PIK3R1;HRAS |
| 106 | Signaling by FGFR3 in disease Homo sapiens R-HSA-5655332 | 3.56E-02 | 2.98E-01 | 4.59E+00 | 1.53E+01 | FGF9;PIK3R1;HRAS |
| 107 | mRNA Splicing - Major Pathway Homo sapiens R-HSA-72163 | 3.59E-02 | 2.98E-01 | 2.10E+00 | 6.98E+00 | HSPA8;FUS;PCBP2;SRSF1;SRSF2;SRSF3;SRSF5;SRSF6;POLR2L |
| 108 | Ion transport by P-type ATPases Homo sapiens R-HSA-936837 | 3.62E-02 | 2.98E-01 | 2.91E+00 | 9.65E+00 | ATP8B2;FXYD2;ATP2B3;ATP1A1;ATP1B1 |
| 109 | Cholesterol biosynthesis Homo sapiens R-HSA-191273 | 4.00E-02 | 3.26E-01 | 4.36E+00 | 1.40E+01 | EBP;DHCR7;LSS |
| 110 | Diseases of carbohydrate metabolism Homo sapiens R-HSA-5663084 | 4.20E-02 | 3.30E-01 | 7.25E+00 | 2.30E+01 | GYS1;RPS27A |
| 111 | Myoclonic epilepsy of Lafora Homo sapiens R-HSA-3785653 | 4.20E-02 | 3.30E-01 | 7.25E+00 | 2.30E+01 | GYS1;RPS27A |

| | | | | | | |
|---|---|---|---|---|---|---|
| 112 | Glycogen storage diseases Homo sapiens R-HSA-3229121 | 4.20E-02 | 3.30E-01 | 7.25E+00 | 2.30E+01 | GYS1;RPS27A |
| 113 | Signaling by FGFR3 fusions in cancer Homo sapiens R-HSA-8853334 | 4.20E-02 | 3.30E-01 | 7.25E+00 | 2.30E+01 | PIK3R1;HRAS |
| 114 | Organelle biogenesis and maintenance Homo sapiens R-HSA-1852241 | 4.75E-02 | 3.71E-01 | 1.61E+00 | 4.90E+00 | DYNC2H1;MRPS28;MEF2C;MRPS24;TUBB;MRPS18B;MRPL16;MRPL36;MRPL15;CYS1;MRPL12;MRPL23;MRPL13;MRPL24;MRPL33;MRPL41;PPARGC1A |
| 115 | Antigen Presentation: Folding, assembly and peptide loading of class I MHC Homo sapiens R-HSA-983170 | 4.95E-02 | 3.72E-01 | 3.96E+00 | 1.19E+01 | HSPA5;CALR;B2M |
| 116 | Glyoxylate metabolism and glycine degradation Homo sapiens R-HSA-389661 | 4.95E-02 | 3.72E-01 | 3.96E+00 | 1.19E+01 | DBT;NDUFAB1;AMT |
| 117 | Cleavage of Growing Transcript in the Termination Region Homo sapiens R-HSA-109688 | 4.97E-02 | 3.72E-01 | 2.64E+00 | 7.94E+00 | SRSF1;SRSF2;SRSF3;SRSF5;SRSF6 |
| 118 | RNA Polymerase II Transcription Termination Homo sapiens R-HSA-73856 | 4.97E-02 | 3.72E-01 | 2.64E+00 | 7.94E+00 | SRSF1;SRSF2;SRSF3;SRSF5;SRSF6 |
| 119 | Post-Elongation Processing of the Transcript Homo sapiens R-HSA-76044 | 4.97E-02 | 3.72E-01 | 2.64E+00 | 7.94E+00 | SRSF1;SRSF2;SRSF3;SRSF5;SRSF6 |
| 120 | Signaling by FGFR4 in disease Homo sapiens R-HSA-5655291 | 5.02E-02 | 3.72E-01 | 6.45E+00 | 1.93E+01 | PIK3R1;HRAS |

| | | | | | | |
|---|---|---|---|---|---|---|
| 121 | Fatty Acyl-CoA Biosynthesis Homo sapiens R-HSA-75105 | 5.08E-02 | 3.73E-01 | 3.06E+00 | 9.12E+00 | TECR;ACOT11;SCD5;ACSF2 |
| 122 | Signaling by FGFR2 in disease Homo sapiens R-HSA-5655253 | 5.46E-02 | 3.92E-01 | 2.98E+00 | 8.67E+00 | FGF9;PIK3R1;HRAS;POLR2L |
| 123 | Diseases associated with glycosaminoglycan metabolism Homo sapiens R-HSA-3560782 | 5.46E-02 | 3.92E-01 | 3.79E+00 | 1.10E+01 | SDC4;GPC3;SDC1 |
| 124 | A tetrasaccharide linker sequence is required for GAG synthesis Homo sapiens R-HSA-1971475 | 5.46E-02 | 3.92E-01 | 3.79E+00 | 1.10E+01 | SDC4;GPC3;SDC1 |
| 125 | Circadian Clock Homo sapiens R-HSA-400253 | 5.90E-02 | 4.20E-01 | 2.51E+00 | 7.09E+00 | SREBF1;MEF2C;BHLHE40;RPS27A;PPARGC1A |
| 126 | Pyruvate metabolism Homo sapiens R-HSA-70268 | 5.99E-02 | 4.23E-01 | 3.63E+00 | 1.02E+01 | LDHB;MPC1;BSG |
| 127 | ERBB2 Activates PTK6 Signaling Homo sapiens R-HSA-8847993 | 6.81E-02 | 4.74E-01 | 5.27E+00 | 1.42E+01 | EGF;ERBB4 |
| 128 | Purine salvage Homo sapiens R-HSA-74217 | 6.81E-02 | 4.74E-01 | 5.27E+00 | 1.42E+01 | GMPR;HPRT1 |
| 129 | Triglyceride Biosynthesis Homo sapiens R-HSA-75109 | 6.93E-02 | 4.78E-01 | 2.38E+00 | 6.36E+00 | TECR;ACOT11;SCD5;AGPAT2;ACSF2 |
| 130 | Transport of Mature mRNA derived from an Intron-Containing Transcript Homo sapiens R-HSA-159236 | 7.67E-02 | 5.13E-01 | 2.31E+00 | 5.92E+00 | SRSF1;SRSF2;SRSF3;SRSF5;SRSF6 |
| 131 | Transcriptional Regulation by TP53 Homo sapiens R-HSA-3700989 | 7.68E-02 | 5.13E-01 | 1.50E+00 | 3.85E+00 | COX8A;COX7B;NDUFA4;COX4I1;USP2;GTF2H2;COX5B;COX6C;COX6A1;COX7C;COX5A;COX6B1;PRDX2;PRDX5;TP53I3;RPS27A;POLR2L |

| | | | | | | |
|---|---|---|---|---|---|---|
| 132 | Hemostasis Homo sapiens R-HSA-109582 | 7.72E-02 | 5.13E-01 | 1.39E+00 | 3.55E+00 | LGALS3BP;SERPINE2;PDE1A;PRCP;PIK3R1;KNG1;AKAP1;PLAU;C1QBP;KCNMB2;PSAP;BSG;HRAS;PF4V1;HSPA5;EGF;APLP2;F11;ATP2B3;MIF;ATP1B1;WEE1;KIF26A;SERPING1;HRG |
| 133 | Receptor-ligand binding initiates the second proteolytic cleavage of Notch receptor Homo sapiens R-HSA-156988 | 7.78E-02 | 5.13E-01 | 4.83E+00 | 1.23E+01 | NOTCH3;RPS27A |
| 134 | Methylation Homo sapiens R-HSA-156581 | 7.78E-02 | 5.13E-01 | 4.83E+00 | 1.23E+01 | MAT2A;COMT |
| 135 | Synthesis of Prostaglandins (PG) and Thromboxanes (TX) Homo sapiens R-HSA-2162123 | 7.78E-02 | 5.13E-01 | 4.83E+00 | 1.23E+01 | CBR1;PTGDS |
| 136 | Neuronal System Homo sapiens R-HSA-112316 | 8.11E-02 | 5.31E-01 | 1.53E+00 | 3.84E+00 | HSPA8;MAOA;KCNJ12;SLC22A2;KCNC4;KCNJ16;COMT;PTPRF;ALDH2;KCNMB2;TSPAN7;NAAA;NEFL;HRAS;KCNJ1 |
| 137 | HS-GAG biosynthesis Homo sapiens R-HSA-2022928 | 8.36E-02 | 5.39E-01 | 3.11E+00 | 7.72E+00 | SDC4;GPC3;SDC1 |
| 138 | Inwardly rectifying K+ channels Homo sapiens R-HSA-1296065 | 8.36E-02 | 5.39E-01 | 3.11E+00 | 7.72E+00 | KCNJ12;KCNJ16;KCNJ1 |
| 139 | Phase 1 - Functionalization of compounds Homo sapiens R-HSA-211945 | 8.75E-02 | 5.54E-01 | 2.03E+00 | 4.94E+00 | ALDH2;MAOA;ALDH1A1;CMBL;ACSS1;BPHL |
| 140 | Glycogen breakdown (glycogenolysis) Homo sapiens R-HSA-70221 | 8.78E-02 | 5.54E-01 | 4.46E+00 | 1.09E+01 | PYGL;PGM1 |
| 141 | ERBB2 Regulates Cell Motility Homo sapiens R-HSA-6785631 | 8.78E-02 | 5.54E-01 | 4.46E+00 | 1.09E+01 | EGF;ERBB4 |

| | | | | | | |
|---|---|---|---|---|---|---|
| 142 | Signaling by NOTCH2 Homo sapiens R-HSA-1980145 | 9.01E-02 | 5.57E-01 | 3.00E+00 | 7.23E+00 | CNTN1;HES1;RPS27A |
| 143 | Neurotransmitter Release Cycle Homo sapiens R-HSA-112310 | 9.01E-02 | 5.57E-01 | 2.47E+00 | 5.95E+00 | HSPA8;MAOA;SLC22A2;NAAA |
| 144 | Ion homeostasis Homo sapiens R-HSA-5578775 | 9.01E-02 | 5.57E-01 | 2.47E+00 | 5.95E+00 | FXYD2;ATP2B3;ATP1A1;ATP1B1 |
| 145 | Transmission across Chemical Synapses Homo sapiens R-HSA-112315 | 9.71E-02 | 5.96E-01 | 1.60E+00 | 3.73E+00 | HSPA8;ALDH2;MAOA;KCNJ12;SLC22A2;TSPAN7;NAAA;NEFL;KCNJ16;COMT;HRAS |
| 146 | Arachidonic acid metabolism Homo sapiens R-HSA-2142753 | 1.00E-01 | 6.09E-01 | 2.37E+00 | 5.45E+00 | CBR1;GPX4;EPHX2;PTGDS |
| 147 | Transport of Mature Transcript to Cytoplasm Homo sapiens R-HSA-72202 | 1.01E-01 | 6.09E-01 | 2.11E+00 | 4.82E+00 | SRSF1;SRSF2;SRSF3;SRSF5;SRSF6 |
| 148 | Binding and Uptake of Ligands by Scavenger Receptors Homo sapiens R-HSA-2173782 | 1.01E-01 | 6.09E-01 | 2.11E+00 | 4.82E+00 | FTH1;APOE;CALR;FTL;HSP90B1 |
| 149 | Lipoprotein metabolism Homo sapiens R-HSA-174824 | 1.04E-01 | 6.19E-01 | 2.81E+00 | 6.37E+00 | LPL;SDC1;APOE |
| 150 | Ras activation uopn Ca2+ infux through NMDA receptor Homo sapiens R-HSA-442982 | 1.09E-01 | 6.42E-01 | 3.87E+00 | 8.57E+00 | NEFL;HRAS |
| 151 | Inflammasomes Homo sapiens R-HSA-622312 | 1.09E-01 | 6.42E-01 | 3.87E+00 | 8.57E+00 | BCL2;TXNIP |
| 152 | Metabolism of water-soluble vitamins and cofactors Homo sapiens R-HSA-196849 | 1.10E-01 | 6.44E-01 | 1.90E+00 | 4.19E+00 | CYB5A;ALDH1L1;PDXK;NAPRT;MCCC1;AOX1 |

| | | | | | | |
|---|---|---|---|---|---|---|
| 153 | Processing of Capped Intron-Containing Pre-mRNA Homo sapiens R-HSA-72203 | 1.13E-01 | 6.59E-01 | 1.59E+00 | 3.46E+00 | HSPA8;FUS;PCBP2;SRSF1;SRSF2;SRSF3;SRSF5;RNPC3;SRSF6;POLR2L |
| 154 | Norepinephrine Neurotransmitter Release Cycle Homo sapiens R-HSA-181430 | 1.20E-01 | 6.71E-01 | 3.63E+00 | 7.69E+00 | MAOA;SLC22A2 |
| 155 | Defective GALNT12 causes colorectal cancer 1 (CRCS1) Homo sapiens R-HSA-5083636 | 1.20E-01 | 6.71E-01 | 3.63E+00 | 7.69E+00 | MUC1;MUC15 |
| 156 | Defective GALNT3 causes familial hyperphosphatemic tumoral calcinosis (HFTC) Homo sapiens R-HSA-5083625 | 1.20E-01 | 6.71E-01 | 3.63E+00 | 7.69E+00 | MUC1;MUC15 |
| 157 | SHC-mediated cascade:FGFR3 Homo sapiens R-HSA-5654704 | 1.20E-01 | 6.71E-01 | 3.63E+00 | 7.69E+00 | FGF9;HRAS |
| 158 | Tie2 Signaling Homo sapiens R-HSA-210993 | 1.20E-01 | 6.71E-01 | 3.63E+00 | 7.69E+00 | PIK3R1;HRAS |
| 159 | cGMP effects Homo sapiens R-HSA-418457 | 1.20E-01 | 6.71E-01 | 3.63E+00 | 7.69E+00 | PDE1A;KCNMB2 |
| 160 | RNA Polymerase II Transcription Homo sapiens R-HSA-73857 | 1.22E-01 | 6.75E-01 | 1.74E+00 | 3.66E+00 | SRSF1;SRSF2;SRSF3;SRSF5;GTF2H2;SRSF6;POLR2L |
| 161 | Cell surface interactions at the vascular wall Homo sapiens R-HSA-202733 | 1.22E-01 | 6.75E-01 | 1.84E+00 | 3.86E+00 | BSG;MIF;PIK3R1;ATP1B1;HRAS;PF4V1 |
| 162 | Signaling by NODAL Homo sapiens R-HSA-1181150 | 1.31E-01 | 7.12E-01 | 3.41E+00 | 6.93E+00 | FOXO3;TDGF1 |
| 163 | Citric acid cycle (TCA cycle) Homo sapiens R-HSA-71403 | 1.31E-01 | 7.12E-01 | 3.41E+00 | 6.93E+00 | SUCLG1;SDHB |

| | | | | | | |
|---|---|---|---|---|---|---|
| 164 | G1 Phase Homo sapiens R-HSA-69236 | 1.33E-01 | 7.12E-01 | 2.49E+00 | 5.02E+00 | CCND3;CCND1;RPS27A |
| 165 | Cyclin D associated events in G1 Homo sapiens R-HSA-69231 | 1.33E-01 | 7.12E-01 | 2.49E+00 | 5.02E+00 | CCND3;CCND1;RPS27A |
| 166 | Signaling by FGFR1 in disease Homo sapiens R-HSA-5655302 | 1.33E-01 | 7.12E-01 | 2.49E+00 | 5.02E+00 | FGF9;PIK3R1;HRAS |
| 167 | Nuclear signaling by ERBB4 Homo sapiens R-HSA-1251985 | 1.41E-01 | 7.36E-01 | 2.42E+00 | 4.74E+00 | CXCL12;ERBB4;PRLR |
| 168 | Defective C1GALT1C1 causes Tn polyagglutination syndrome (TNPS) Homo sapiens R-HSA-5083632 | 1.43E-01 | 7.36E-01 | 3.22E+00 | 6.27E+00 | MUC1;MUC15 |
| 169 | Syndecan interactions Homo sapiens R-HSA-3000170 | 1.43E-01 | 7.36E-01 | 3.22E+00 | 6.27E+00 | SDC4;SDC1 |
| 170 | Ion channel transport Homo sapiens R-HSA-983712 | 1.43E-01 | 7.36E-01 | 1.51E+00 | 2.93E+00 | CLCN5;ATP8B2;FXYD2;TSC22D3;WNK4;CLCNKB;ATP2B3;ATP1A1;RPS27A;ATP1B1 |
| 171 | Constitutive Signaling by Aberrant PI3K in Cancer Homo sapiens R-HSA-2219530 | 1.46E-01 | 7.36E-01 | 2.04E+00 | 3.91E+00 | FGF9;ERBB4;EGF;PIK3R1 |
| 172 | Negative regulation of MAPK pathway Homo sapiens R-HSA-5675221 | 1.48E-01 | 7.36E-01 | 2.35E+00 | 4.49E+00 | RPS27A;HRAS;DUSP9 |
| 173 | Diseases associated with O-glycosylation of proteins Homo sapiens R-HSA-3906995 | 1.53E-01 | 7.36E-01 | 2.00E+00 | 3.76E+00 | NOTCH3;MUC1;MUC15;THSD7A |

| | | | | | | |
|---|---|---|---|---|---|---|
| 174 | NOTCH2 Activation and Transmission of Signal to the Nucleus Homo sapiens R-HSA-2979096 | 1.54E-01 | 7.36E-01 | 3.05E+00 | 5.70E+00 | CNTN1;RPS27A |
| 175 | SHC-mediated cascade:FGFR1 Homo sapiens R-HSA-5654688 | 1.54E-01 | 7.36E-01 | 3.05E+00 | 5.70E+00 | FGF9;HRAS |
| 176 | Aflatoxin activation and detoxification Homo sapiens R-HSA-5423646 | 1.54E-01 | 7.36E-01 | 3.05E+00 | 5.70E+00 | MGST3;MGST1 |
| 177 | Abacavir transmembrane transport Homo sapiens R-HSA-2161517 | 1.56E-01 | 7.36E-01 | 7.24E+00 | 1.34E+01 | SLC22A2 |
| 178 | Sodium-coupled phosphate cotransporters Homo sapiens R-HSA-427652 | 1.56E-01 | 7.36E-01 | 7.24E+00 | 1.34E+01 | SLC20A2 |
| 179 | Catabolism of glucuronate to xylulose-5-phosphate Homo sapiens R-HSA-5661270 | 1.56E-01 | 7.36E-01 | 7.24E+00 | 1.34E+01 | DCXR |
| 180 | Synthesis of GDP-mannose Homo sapiens R-HSA-446205 | 1.56E-01 | 7.36E-01 | 7.24E+00 | 1.34E+01 | PMM1 |
| 181 | PTK6 Regulates Proteins Involved in RNA Processing Homo sapiens R-HSA-8849468 | 1.56E-01 | 7.36E-01 | 7.24E+00 | 1.34E+01 | KHDRBS3 |
| 182 | Activation of RAS in B cells Homo sapiens R-HSA-1169092 | 1.56E-01 | 7.36E-01 | 7.24E+00 | 1.34E+01 | HRAS |
| 183 | G2/M DNA replication checkpoint Homo sapiens R-HSA-69478 | 1.56E-01 | 7.36E-01 | 7.24E+00 | 1.34E+01 | WEE1 |

| | | | | | | |
|---|---|---|---|---|---|---|
| 184 | NGF-independant TRKA activation Homo sapiens R-HSA-187024 | 1.56E-01 | 7.36E-01 | 7.24E+00 | 1.34E+01 | NTRK2 |
| 185 | Propionyl-CoA catabolism Homo sapiens R-HSA-71032 | 1.56E-01 | 7.36E-01 | 7.24E+00 | 1.34E+01 | MCEE |
| 186 | tRNA processing in the mitochondrion Homo sapiens R-HSA-6785470 | 1.56E-01 | 7.36E-01 | 7.24E+00 | 1.34E+01 | HSD17B10 |
| 187 | Trafficking of myristoylated proteins to the cilium Homo sapiens R-HSA-5624138 | 1.56E-01 | 7.36E-01 | 7.24E+00 | 1.34E+01 | CYS1 |
| 188 | Signaling by MST1 Homo sapiens R-HSA-8852405 | 1.56E-01 | 7.36E-01 | 7.24E+00 | 1.34E+01 | HPN |
| 189 | Glycosphingolipid metabolism Homo sapiens R-HSA-1660662 | 1.56E-01 | 7.36E-01 | 2.29E+00 | 4.25E+00 | UGT8;PSAP;SUMF2 |
| 190 | Signaling by FGFR in disease Homo sapiens R-HSA-1226099 | 1.59E-01 | 7.43E-01 | 1.97E+00 | 3.62E+00 | FGF9;PIK3R1;HRAS;POLR2L |
| 191 | Unfolded Protein Response (UPR) Homo sapiens R-HSA-381119 | 1.60E-01 | 7.43E-01 | 1.79E+00 | 3.28E+00 | XBP1;HSPA5;CALR;HERPUD1;HSP90B1 |
| 192 | PI3K/AKT Signaling in Cancer Homo sapiens R-HSA-2219528 | 1.60E-01 | 7.43E-01 | 1.79E+00 | 3.28E+00 | FGF9;ERBB4;EGF;PIK3R1;FOXO3 |
| 193 | Activation of gene expression by SREBF (SREBP) Homo sapiens R-HSA-2426168 | 1.65E-01 | 7.44E-01 | 2.23E+00 | 4.03E+00 | SREBF1;DHCR7;LSS |
| 194 | Metabolism of nucleotides Homo sapiens R-HSA-15869 | 1.66E-01 | 7.44E-01 | 1.77E+00 | 3.18E+00 | NME2;GMPR;GLRX;NUDT16;HPRT1 |
| 195 | Cardiac conduction Homo sapiens R-HSA-5576891 | 1.66E-01 | 7.44E-01 | 1.59E+00 | 2.85E+00 | KCNJ12;FXYD2;KCNIP3;ATP2B3;ATP1A1;ATP1B1;SCN1B |

| | | | | | | |
|---|---|---|---|---|---|---|
| 196 | Common Pathway of Fibrin Clot Formation Homo sapiens R-HSA-140875 | 1.66E-01 | 7.44E-01 | 2.90E+00 | 5.20E+00 | SERPINE2;PF4V1 |
| 197 | Pink/Parkin Mediated Mitophagy Homo sapiens R-HSA-5205685 | 1.66E-01 | 7.44E-01 | 2.90E+00 | 5.20E+00 | TOMM7;RPS27A |
| 198 | Mitophagy Homo sapiens R-HSA-5205647 | 1.66E-01 | 7.44E-01 | 2.90E+00 | 5.20E+00 | TOMM7;RPS27A |
| 199 | Post-chaperonin tubulin folding pathway Homo sapiens R-HSA-389977 | 1.66E-01 | 7.44E-01 | 2.90E+00 | 5.20E+00 | TUBB2B;TUBB2A |
| 200 | SHC-mediated cascade:FGFR4 Homo sapiens R-HSA-5654719 | 1.78E-01 | 7.71E-01 | 2.76E+00 | 4.76E+00 | FGF9;HRAS |
| 201 | APC/C:Cdc20 mediated degradation of Cyclin B Homo sapiens R-HSA-174048 | 1.78E-01 | 7.71E-01 | 2.76E+00 | 4.76E+00 | RPS27A;ANAPC11 |
| 202 | O-linked glycosylation of mucins Homo sapiens R-HSA-913709 | 1.79E-01 | 7.71E-01 | 1.87E+00 | 3.23E+00 | GALNT14;MUC1;GALNT16;MUC15 |
| 203 | GDP-fucose biosynthesis Homo sapiens R-HSA-6787639 | 1.84E-01 | 7.71E-01 | 5.80E+00 | 9.80E+00 | GMDS |
| 204 | PTK6 Regulates Cell Cycle Homo sapiens R-HSA-8849470 | 1.84E-01 | 7.71E-01 | 5.80E+00 | 9.80E+00 | CCND1 |
| 205 | Pre-NOTCH Processing in the Endoplasmic Reticulum Homo sapiens R-HSA-1912399 | 1.84E-01 | 7.71E-01 | 5.80E+00 | 9.80E+00 | NOTCH3 |

| | | | | | | |
|---|---|---|---|---|---|---|
| 206 | Scavenging by Class F Receptors Homo sapiens R-HSA-3000484 | 1.84E-01 | 7.71E-01 | 5.80E+00 | 9.80E+00 | CALR |
| 207 | Activation of TRKA receptors Homo sapiens R-HSA-187015 | 1.84E-01 | 7.71E-01 | 5.80E+00 | 9.80E+00 | NTRK2 |
| 208 | Methionine salvage pathway Homo sapiens R-HSA-1237112 | 1.84E-01 | 7.71E-01 | 5.80E+00 | 9.80E+00 | GOT1 |
| 209 | Conjugation of benzoate with glycine Homo sapiens R-HSA-177135 | 1.84E-01 | 7.71E-01 | 5.80E+00 | 9.80E+00 | GLYATL1 |
| 210 | Synthesis of 5-eicosatetraenoic acids Homo sapiens R-HSA-2142688 | 1.84E-01 | 7.71E-01 | 5.80E+00 | 9.80E+00 | GPX4 |
| 211 | Synthesis of 15-eicosatetraenoic acid derivatives Homo sapiens R-HSA-2142770 | 1.84E-01 | 7.71E-01 | 5.80E+00 | 9.80E+00 | GPX4 |
| 212 | Synthesis of 12-eicosatetraenoic acid derivatives Homo sapiens R-HSA-2142712 | 1.84E-01 | 7.71E-01 | 5.80E+00 | 9.80E+00 | GPX4 |
| 213 | Protein repair Homo sapiens R-HSA-5676934 | 1.84E-01 | 7.71E-01 | 5.80E+00 | 9.80E+00 | MSRB2 |
| 214 | Diseases of metabolism Homo sapiens R-HSA-5668914 | 1.90E-01 | 7.88E-01 | 2.07E+00 | 3.45E+00 | GYS1;MCCC1;RPS27A |
| 215 | COPII (Coat Protein 2) Mediated Vesicle Transport Homo sapiens R-HSA-204005 | 1.92E-01 | 7.95E-01 | 1.81E+00 | 2.99E+00 | TRAPPC2L;TRAPPC5;FOLR1;CTSC |
| 216 | RAF activation Homo sapiens R-HSA-5673000 | 2.03E-01 | 8.04E-01 | 2.52E+00 | 4.02E+00 | PHB;HRAS |

| | | | | | | |
|---|---|---|---|---|---|---|
| 217 | APC-Cdc20 mediated degradation of Nek2A Homo sapiens R-HSA-179409 | 2.03E-01 | 8.04E-01 | 2.52E+00 | 4.02E+00 | RPS27A;ANAPC11 |
| 218 | Basigin interactions Homo sapiens R-HSA-210991 | 2.03E-01 | 8.04E-01 | 2.52E+00 | 4.02E+00 | BSG;ATP1B1 |
| 219 | Formation of tubulin folding intermediates by CCT/TriC Homo sapiens R-HSA-389960 | 2.03E-01 | 8.04E-01 | 2.52E+00 | 4.02E+00 | TUBB2B;TUBB2A |
| 220 | Activation of G protein gated Potassium channels Homo sapiens R-HSA-1296041 | 2.03E-01 | 8.04E-01 | 2.52E+00 | 4.02E+00 | KCNJ12;KCNJ16 |
| 221 | Inhibition of voltage gated Ca2+ channels via Gbeta/gamma subunits Homo sapiens R-HSA-997272 | 2.03E-01 | 8.04E-01 | 2.52E+00 | 4.02E+00 | KCNJ12;KCNJ16 |
| 222 | G protein gated Potassium channels Homo sapiens R-HSA-1296059 | 2.03E-01 | 8.04E-01 | 2.52E+00 | 4.02E+00 | KCNJ12;KCNJ16 |
| 223 | Nitric oxide stimulates guanylate cyclase Homo sapiens R-HSA-392154 | 2.03E-01 | 8.04E-01 | 2.52E+00 | 4.02E+00 | PDE1A;KCNMB2 |
| 224 | Synthesis and interconversion of nucleotide di- and triphosphates Homo sapiens R-HSA-499943 | 2.03E-01 | 8.04E-01 | 2.52E+00 | 4.02E+00 | NME2;GLRX |

| | | | | | | |
|---|---|---|---|---|---|---|
| 225 | NOTCH1 Intracellular Domain Regulates Transcription Homo sapiens R-HSA-2122947 | 2.07E-01 | 8.04E-01 | 1.98E+00 | 3.12E+00 | TLE1;HES1;RPS27A |
| 226 | Zinc efflux and compartmentalization by the SLC30 family Homo sapiens R-HSA-435368 | 2.12E-01 | 8.04E-01 | 4.83E+00 | 7.50E+00 | SLC30A1 |
| 227 | Cation-coupled Chloride cotransporters Homo sapiens R-HSA-426117 | 2.12E-01 | 8.04E-01 | 4.83E+00 | 7.50E+00 | SLC12A1 |
| 228 | Neurofascin interactions Homo sapiens R-HSA-447043 | 2.12E-01 | 8.04E-01 | 4.83E+00 | 7.50E+00 | CNTN1 |
| 229 | Regulation of gene expression in late stage (branching morphogenesis) pancreatic bud precursor cells Homo sapiens R-HSA-210744 | 2.12E-01 | 8.04E-01 | 4.83E+00 | 7.50E+00 | HES1 |
| 230 | FGFR3b ligand binding and activation Homo sapiens R-HSA-190371 | 2.12E-01 | 8.04E-01 | 4.83E+00 | 7.50E+00 | FGF9 |
| 231 | Defective HLCS causes multiple carboxylase deficiency Homo sapiens R-HSA-3371599 | 2.12E-01 | 8.04E-01 | 4.83E+00 | 7.50E+00 | MCCC1 |
| 232 | Defects in biotin (Btn) metabolism Homo sapiens R-HSA-3323169 | 2.12E-01 | 8.04E-01 | 4.83E+00 | 7.50E+00 | MCCC1 |
| 233 | Phosphate bond hydrolysis by NUDT proteins Homo sapiens R-HSA-2393930 | 2.12E-01 | 8.04E-01 | 4.83E+00 | 7.50E+00 | NUDT16 |

| | | | | | | |
|---|---|---|---|---|---|---|
| 234 | Lipid digestion, mobilization, and transport Homo sapiens R-HSA-73923 | 2.13E-01 | 8.04E-01 | 1.73E+00 | 2.68E+00 | FABP3;LPL;SDC1;APOE |
| 235 | Termination of O-glycan biosynthesis Homo sapiens R-HSA-977068 | 2.15E-01 | 8.04E-01 | 2.42E+00 | 3.71E+00 | MUC1;MUC15 |
| 236 | Regulation of Hypoxia-inducible Factor (HIF) by oxygen Homo sapiens R-HSA-1234174 | 2.15E-01 | 8.04E-01 | 2.42E+00 | 3.71E+00 | CITED2;RPS27A |
| 237 | Cellular response to hypoxia Homo sapiens R-HSA-2262749 | 2.15E-01 | 8.04E-01 | 2.42E+00 | 3.71E+00 | CITED2;RPS27A |
| 238 | SHC-mediated cascade:FGFR2 Homo sapiens R-HSA-5654699 | 2.15E-01 | 8.04E-01 | 2.42E+00 | 3.71E+00 | FGF9;HRAS |
| 239 | Signaling by NOTCH1 Homo sapiens R-HSA-1980143 | 2.20E-01 | 8.16E-01 | 1.71E+00 | 2.58E+00 | TLE1;CNTN1;HES1;RPS27A |
| 240 | Signaling by NOTCH Homo sapiens R-HSA-157118 | 2.24E-01 | 8.16E-01 | 1.50E+00 | 2.25E+00 | NOTCH3;TLE1;CCND1;CNTN1;HES1;RPS27A |
| 241 | Visual phototransduction Homo sapiens R-HSA-2187338 | 2.24E-01 | 8.16E-01 | 1.58E+00 | 2.36E+00 | SDC4;GPC3;LPL;SDC1;APOE |
| 242 | Retrograde transport at the Trans-Golgi-Network Homo sapiens R-HSA-6811440 | 2.24E-01 | 8.16E-01 | 1.89E+00 | 2.83E+00 | RHOBTB3;NAA38;RAB6B |
| 243 | EGFR downregulation Homo sapiens R-HSA-182971 | 2.27E-01 | 8.16E-01 | 2.32E+00 | 3.43E+00 | EGF;RPS27A |

| | | | | | | |
|---|---|---|---|---|---|---|
| 244 | RNA Pol II CTD phosphorylation and interaction with CE Homo sapiens R-HSA-167160 | 2.27E-01 | 8.16E-01 | 2.32E+00 | 3.43E+00 | GTF2H2;POLR2L |
| 245 | RNA Pol II CTD phosphorylation and interaction with CE Homo sapiens R-HSA-77075 | 2.27E-01 | 8.16E-01 | 2.32E+00 | 3.43E+00 | GTF2H2;POLR2L |
| 246 | Prefoldin mediated transfer of substrate to CCT/TriC Homo sapiens R-HSA-389957 | 2.27E-01 | 8.16E-01 | 2.32E+00 | 3.43E+00 | TUBB2B;TUBB2A |
| 247 | CREB phosphorylation through the activation of Ras Homo sapiens R-HSA-442742 | 2.27E-01 | 8.16E-01 | 2.32E+00 | 3.43E+00 | NEFL;HRAS |
| 248 | RORA activates gene expression Homo sapiens R-HSA-1368082 | 2.27E-01 | 8.16E-01 | 2.32E+00 | 3.43E+00 | SREBF1;PPARGC1A |
| 249 | Chondroitin sulfate/dermatan sulfate metabolism Homo sapiens R-HSA-1793185 | 2.33E-01 | 8.21E-01 | 1.85E+00 | 2.69E+00 | SDC4;GPC3;SDC1 |
| 250 | Transport of inorganic cations/anions and amino acids/oligopeptides Homo sapiens R-HSA-425393 | 2.36E-01 | 8.21E-01 | 1.54E+00 | 2.23E+00 | SLC20A2;SLC43A1;SLC12A1;SLC3A1;SLC9A1 |
| 251 | Potassium Channels Homo sapiens R-HSA-1296071 | 2.36E-01 | 8.21E-01 | 1.54E+00 | 2.23E+00 | KCNJ12;KCNMB2;KCNC4;KCNJ16;KCNJ1 |
| 252 | Vitamin C (ascorbate) metabolism Homo sapiens R-HSA-196836 | 2.38E-01 | 8.21E-01 | 4.14E+00 | 5.94E+00 | CYB5A |

| | | | | | | |
|---|---|---|---|---|---|---|
| 253 | BH3-only proteins associate with and inactivate anti-apoptotic BCL-2 members Homo sapiens R-HSA-111453 | 2.38E-01 | 8.21E-01 | 4.14E+00 | 5.94E+00 | BCL2 |
| 254 | NR1D1 (REV-ERBA) represses gene expression Homo sapiens R-HSA-1368071 | 2.38E-01 | 8.21E-01 | 4.14E+00 | 5.94E+00 | PPARGC1A |
| 255 | Nicotinamide salvaging Homo sapiens R-HSA-197264 | 2.38E-01 | 8.21E-01 | 4.14E+00 | 5.94E+00 | NAPRT |
| 256 | Synthesis of epoxy (EET) and dihydroxyeicosatrienoic acids (DHET) Homo sapiens R-HSA-2142670 | 2.38E-01 | 8.21E-01 | 4.14E+00 | 5.94E+00 | EPHX2 |
| 257 | Conjugation of salicylate with glycine Homo sapiens R-HSA-177128 | 2.38E-01 | 8.21E-01 | 4.14E+00 | 5.94E+00 | GLYATL1 |
| 258 | tRNA modification in the mitochondrion Homo sapiens R-HSA-6787450 | 2.38E-01 | 8.21E-01 | 4.14E+00 | 5.94E+00 | HSD17B10 |
| 259 | PERK regulates gene expression Homo sapiens R-HSA-381042 | 2.40E-01 | 8.25E-01 | 2.23E+00 | 3.18E+00 | HSPA5;HERPUD1 |
| 260 | Stimuli-sensing channels Homo sapiens R-HSA-2672351 | 2.42E-01 | 8.29E-01 | 1.53E+00 | 2.16E+00 | CLCN5;TSC22D3;WNK4;CLCNKB;RPS27A |
| 261 | Transport of glucose and other sugars, bile salts and organic acids, metal ions and amine compounds Homo sapiens R-HSA-425366 | 2.49E-01 | 8.39E-01 | 1.51E+00 | 2.10E+00 | SLC47A1;SLC22A2;BSG;SLC16A7;SLC30A1 |

| | | | | | | |
|---|---|---|---|---|---|---|
| 262 | COPI-independent Golgi-to-ER retrograde traffic Homo sapiens R-HSA-6811436 | 2.52E-01 | 8.39E-01 | 2.15E+00 | 2.96E+00 | DYNC2H1;RAB6B |
| 263 | mRNA Capping Homo sapiens R-HSA-72086 | 2.52E-01 | 8.39E-01 | 2.15E+00 | 2.96E+00 | GTF2H2;POLR2L |
| 264 | Negative regulation of FGFR3 signaling Homo sapiens R-HSA-5654732 | 2.52E-01 | 8.39E-01 | 2.15E+00 | 2.96E+00 | FGF9;RPS27A |
| 265 | Formation of TC-NER Pre-Incision Complex Homo sapiens R-HSA-6781823 | 2.60E-01 | 8.39E-01 | 1.74E+00 | 2.34E+00 | GTF2H2;RPS27A;POLR2L |
| 266 | Organic cation transport Homo sapiens R-HSA-549127 | 2.63E-01 | 8.39E-01 | 3.62E+00 | 4.83E+00 | SLC22A2 |
| 267 | Type I hemidesmosome assembly Homo sapiens R-HSA-446107 | 2.63E-01 | 8.39E-01 | 3.62E+00 | 4.83E+00 | DST |
| 268 | Sodium/Proton exchangers Homo sapiens R-HSA-425986 | 2.63E-01 | 8.39E-01 | 3.62E+00 | 4.83E+00 | SLC9A1 |
| 269 | AKT phosphorylates targets in the nucleus Homo sapiens R-HSA-198693 | 2.63E-01 | 8.39E-01 | 3.62E+00 | 4.83E+00 | FOXO3 |
| 270 | A third proteolytic cleavage releases NICD Homo sapiens R-HSA-157212 | 2.63E-01 | 8.39E-01 | 3.62E+00 | 4.83E+00 | NOTCH3 |
| 271 | PTK6 Regulates RTKs and Their Effectors AKT1 and DOK1 Homo sapiens R-HSA-8849469 | 2.63E-01 | 8.39E-01 | 3.62E+00 | 4.83E+00 | RPS27A |

| | | | | | | |
|---|---|---|---|---|---|---|
| 272 | EGFR Transactivation by Gastrin Homo sapiens R-HSA-2179392 | 2.63E-01 | 8.39E-01 | 3.62E+00 | 4.83E+00 | HRAS |
| 273 | SHC-related events triggered by IGF1R Homo sapiens R-HSA-2428933 | 2.63E-01 | 8.39E-01 | 3.62E+00 | 4.83E+00 | HRAS |
| 274 | CHL1 interactions Homo sapiens R-HSA-447041 | 2.63E-01 | 8.39E-01 | 3.62E+00 | 4.83E+00 | HSPA8 |
| 275 | Ca2+ activated K+ channels Homo sapiens R-HSA-1296052 | 2.63E-01 | 8.39E-01 | 3.62E+00 | 4.83E+00 | KCNMB2 |
| 276 | Prostanoid ligand receptors Homo sapiens R-HSA-391908 | 2.63E-01 | 8.39E-01 | 3.62E+00 | 4.83E+00 | PTGER3 |
| 277 | Conjugation of carboxylic acids Homo sapiens R-HSA-159424 | 2.63E-01 | 8.39E-01 | 3.62E+00 | 4.83E+00 | GLYATL1 |
| 278 | Amino Acid conjugation Homo sapiens R-HSA-156587 | 2.63E-01 | 8.39E-01 | 3.62E+00 | 4.83E+00 | GLYATL1 |
| 279 | RNA Polymerase I Promoter Escape Homo sapiens R-HSA-73772 | 2.65E-01 | 8.39E-01 | 2.07E+00 | 2.75E+00 | GTF2H2;POLR2L |
| 280 | Activated NOTCH1 Transmits Signal to the Nucleus Homo sapiens R-HSA-2122948 | 2.65E-01 | 8.39E-01 | 2.07E+00 | 2.75E+00 | CNTN1;RPS27A |
| 281 | Tight junction interactions Homo sapiens R-HSA-420029 | 2.65E-01 | 8.39E-01 | 2.07E+00 | 2.75E+00 | CLDN10;CLDN14 |
| 282 | Heparan sulfate/heparin (HS-GAG) metabolism Homo sapiens R-HSA-1638091 | 2.69E-01 | 8.44E-01 | 1.71E+00 | 2.24E+00 | SDC4;GPC3;SDC1 |

| | | | | | | |
|---|---|---|---|---|---|---|
| 283 | Golgi Associated Vesicle Biogenesis Homo sapiens R-HSA-432722 | 2.69E-01 | 8.44E-01 | 1.71E+00 | 2.24E+00 | HSPA8;FTH1;FTL |
| 284 | AUF1 (hnRNP D0) binds and destabilizes mRNA Homo sapiens R-HSA-450408 | 2.69E-01 | 8.44E-01 | 1.71E+00 | 2.24E+00 | HSPA8;HSPB1;RPS27A |
| 285 | Transcriptional regulation of white adipocyte differentiation Homo sapiens R-HSA-381340 | 2.71E-01 | 8.46E-01 | 1.55E+00 | 2.02E+00 | SREBF1;CCND3;LPL;PPARGC1A |
| 286 | Amino acid transport across the plasma membrane Homo sapiens R-HSA-352230 | 2.77E-01 | 8.46E-01 | 2.00E+00 | 2.56E+00 | SLC43A1;SLC3A1 |
| 287 | Negative regulation of FGFR4 signaling Homo sapiens R-HSA-5654733 | 2.77E-01 | 8.46E-01 | 2.00E+00 | 2.56E+00 | FGF9;RPS27A |
| 288 | FGFR1 mutant receptor activation Homo sapiens R-HSA-1839124 | 2.77E-01 | 8.46E-01 | 2.00E+00 | 2.56E+00 | FGF9;PIK3R1 |
| 289 | Regulation of TNFR1 signaling Homo sapiens R-HSA-5357905 | 2.77E-01 | 8.46E-01 | 2.00E+00 | 2.56E+00 | USP2;RPS27A |
| 290 | Regulation of cholesterol biosynthesis by SREBP (SREBF) Homo sapiens R-HSA-1655829 | 2.78E-01 | 8.46E-01 | 1.67E+00 | 2.14E+00 | SREBF1;DHCR7;LSS |
| 291 | Abacavir transport and metabolism Homo sapiens R-HSA-2161522 | 2.88E-01 | 8.46E-01 | 3.22E+00 | 4.01E+00 | SLC22A2 |

| | | | | | | |
|---|---|---|---|---|---|---|
| 292 | Nef mediated downregulation of MHC class I complex cell surface expression Homo sapiens R-HSA-164940 | 2.88E-01 | 8.46E-01 | 3.22E+00 | 4.01E+00 | B2M |
| 293 | Activation of PPARGC1A (PGC-1alpha) by phosphorylation Homo sapiens R-HSA-2151209 | 2.88E-01 | 8.46E-01 | 3.22E+00 | 4.01E+00 | PPARGC1A |
| 294 | IRAK2 mediated activation of TAK1 complex upon TLR7/8 or 9 stimulation Homo sapiens R-HSA-975163 | 2.88E-01 | 8.46E-01 | 3.22E+00 | 4.01E+00 | RPS27A |
| 295 | IRAK2 mediated activation of TAK1 complex Homo sapiens R-HSA-937042 | 2.88E-01 | 8.46E-01 | 3.22E+00 | 4.01E+00 | RPS27A |
| 296 | Regulation of gene expression by Hypoxia-inducible Factor Homo sapiens R-HSA-1234158 | 2.88E-01 | 8.46E-01 | 3.22E+00 | 4.01E+00 | CITED2 |
| 297 | GP1b-IX-V activation signalling Homo sapiens R-HSA-430116 | 2.88E-01 | 8.46E-01 | 3.22E+00 | 4.01E+00 | PIK3R1 |
| 298 | Synthesis of pyrophosphates in the cytosol Homo sapiens R-HSA-1855167 | 2.88E-01 | 8.46E-01 | 3.22E+00 | 4.01E+00 | NUDT4 |
| 299 | Synthesis of PIPs at the late endosome membrane Homo sapiens R-HSA-1660517 | 2.88E-01 | 8.46E-01 | 3.22E+00 | 4.01E+00 | MTMR7 |
| 300 | Regulation of signaling by NODAL Homo sapiens R-HSA-1433617 | 2.88E-01 | 8.46E-01 | 3.22E+00 | 4.01E+00 | TDGF1 |

| | | | | | | |
|---|---|---|---|---|---|---|
| 301 | Urea cycle Homo sapiens R-HSA-70635 | 2.88E-01 | 8.46E-01 | 3.22E+00 | 4.01E+00 | ASS1 |
| 302 | RNA Polymerase I Transcription Termination Homo sapiens R-HSA-73863 | 2.90E-01 | 8.46E-01 | 1.93E+00 | 2.39E+00 | GTF2H2;POLR2L |
| 303 | Oncogene Induced Senescence Homo sapiens R-HSA-2559585 | 2.90E-01 | 8.46E-01 | 1.93E+00 | 2.39E+00 | ID1;RPS27A |
| 304 | Negative regulation of FGFR1 signaling Homo sapiens R-HSA-5654726 | 2.90E-01 | 8.46E-01 | 1.93E+00 | 2.39E+00 | FGF9;RPS27A |
| 305 | Cooperation of Prefoldin and TriC/CCT in actin and tubulin folding Homo sapiens R-HSA-389958 | 2.90E-01 | 8.46E-01 | 1.93E+00 | 2.39E+00 | TUBB2B;TUBB2A |
| 306 | PI5P, PP2A and IER3 Regulate PI3K/AKT Signaling Homo sapiens R-HSA-6811558 | 3.01E-01 | 8.60E-01 | 1.47E+00 | 1.76E+00 | FGF9;ERBB4;EGF;PIK3R1 |
| 307 | Cargo concentration in the ER Homo sapiens R-HSA-5694530 | 3.02E-01 | 8.60E-01 | 1.87E+00 | 2.24E+00 | FOLR1;CTSC |
| 308 | Formation of the Early Elongation Complex Homo sapiens R-HSA-113418 | 3.02E-01 | 8.60E-01 | 1.87E+00 | 2.24E+00 | GTF2H2;POLR2L |
| 309 | Formation of the HIV-1 Early Elongation Complex Homo sapiens R-HSA-167158 | 3.02E-01 | 8.60E-01 | 1.87E+00 | 2.24E+00 | GTF2H2;POLR2L |
| 310 | FGFR2 mutant receptor activation Homo sapiens R-HSA-1839126 | 3.02E-01 | 8.60E-01 | 1.87E+00 | 2.24E+00 | FGF9;POLR2L |

| | | | | | | |
|---|---|---|---|---|---|---|
| 311 | rRNA modification in the nucleus Homo sapiens R-HSA-6790901 | 3.06E-01 | 8.60E-01 | 1.58E+00 | 1.87E+00 | RPS14;RPS6;RPS2 |
| 312 | O-linked glycosylation Homo sapiens R-HSA-5173105 | 3.06E-01 | 8.60E-01 | 1.38E+00 | 1.63E+00 | GALNT14;MUC1;GALNT16;THSD7A;MUC15 |
| 313 | Signaling by NOTCH3 Homo sapiens R-HSA-1980148 | 3.12E-01 | 8.60E-01 | 2.90E+00 | 3.38E+00 | NOTCH3 |
| 314 | Membrane binding and targetting of GAG proteins Homo sapiens R-HSA-174490 | 3.12E-01 | 8.60E-01 | 2.90E+00 | 3.38E+00 | RPS27A |
| 315 | Synthesis And Processing Of GAG, GAGPOL Polyproteins Homo sapiens R-HSA-174495 | 3.12E-01 | 8.60E-01 | 2.90E+00 | 3.38E+00 | RPS27A |
| 316 | FGFR1c ligand binding and activation Homo sapiens R-HSA-190373 | 3.12E-01 | 8.60E-01 | 2.90E+00 | 3.38E+00 | FGF9 |
| 317 | Signaling by activated point mutants of FGFR1 Homo sapiens R-HSA-1839122 | 3.12E-01 | 8.60E-01 | 2.90E+00 | 3.38E+00 | FGF9 |
| 318 | Interleukin-7 signaling Homo sapiens R-HSA-1266695 | 3.12E-01 | 8.60E-01 | 2.90E+00 | 3.38E+00 | PIK3R1 |
| 319 | Apoptotic cleavage of cell adhesion proteins Homo sapiens R-HSA-351906 | 3.12E-01 | 8.60E-01 | 2.90E+00 | 3.38E+00 | TJP2 |
| 320 | Biotin transport and metabolism Homo sapiens R-HSA-196780 | 3.12E-01 | 8.60E-01 | 2.90E+00 | 3.38E+00 | MCCC1 |
| 321 | Heme biosynthesis Homo sapiens R-HSA-189451 | 3.12E-01 | 8.60E-01 | 2.90E+00 | 3.38E+00 | FECH |

| | | | | | | |
|---|---|---|---|---|---|---|
| 322 | Phenylalanine and tyrosine catabolism Homo sapiens R-HSA-71182 | 3.12E-01 | 8.60E-01 | 2.90E+00 | 3.38E+00 | PCBD1 |
| 323 | Negative regulation of FGFR2 signaling Homo sapiens R-HSA-5654727 | 3.15E-01 | 8.60E-01 | 1.81E+00 | 2.09E+00 | FGF9;RPS27A |
| 324 | Negative regulators of RIG-I/MDA5 signaling Homo sapiens R-HSA-936440 | 3.15E-01 | 8.60E-01 | 1.81E+00 | 2.09E+00 | PCBP2;RPS27A |
| 325 | EGFR interacts with phospholipase C-gamma Homo sapiens R-HSA-212718 | 3.15E-01 | 8.60E-01 | 1.81E+00 | 2.09E+00 | EGF;PDE1A |
| 326 | Purine metabolism Homo sapiens R-HSA-73847 | 3.15E-01 | 8.60E-01 | 1.81E+00 | 2.09E+00 | GMPR;HPRT1 |
| 327 | Interactions of Vpr with host cellular proteins Homo sapiens R-HSA-176033 | 3.27E-01 | 8.77E-01 | 1.76E+00 | 1.96E+00 | SLC25A5;SLC25A4 |
| 328 | Regulation of TP53 Degradation Homo sapiens R-HSA-6804757 | 3.27E-01 | 8.77E-01 | 1.76E+00 | 1.96E+00 | USP2;RPS27A |
| 329 | Fanconi Anemia Pathway Homo sapiens R-HSA-6783310 | 3.27E-01 | 8.77E-01 | 1.76E+00 | 1.96E+00 | POLN;RPS27A |
| 330 | Post NMDA receptor activation events Homo sapiens R-HSA-438064 | 3.27E-01 | 8.77E-01 | 1.76E+00 | 1.96E+00 | NEFL;HRAS |
| 331 | Muscle contraction Homo sapiens R-HSA-397014 | 3.33E-01 | 8.77E-01 | 1.23E+00 | 1.36E+00 | KCNJ12;FXYD2;KCNIP3;ATP2B3;NEB;ATP1A1;ATP1B1;SCN1B |
| 332 | Reversible hydration of carbon dioxide Homo sapiens R-HSA-1475029 | 3.35E-01 | 8.77E-01 | 2.63E+00 | 2.88E+00 | CA12 |

| | | | | | | |
|---|---|---|---|---|---|---|
| 333 | Hyaluronan uptake and degradation Homo sapiens R-HSA-2160916 | 3.35E-01 | 8.77E-01 | 2.63E+00 | 2.88E+00 | SLC9A1 |
| 334 | Endosomal/Vacuolar pathway Homo sapiens R-HSA-1236977 | 3.35E-01 | 8.77E-01 | 2.63E+00 | 2.88E+00 | B2M |
| 335 | Chk1/Chk2(Cds1) mediated inactivation of Cyclin B:Cdk1 complex Homo sapiens R-HSA-75035 | 3.35E-01 | 8.77E-01 | 2.63E+00 | 2.88E+00 | WEE1 |
| 336 | NOTCH2 intracellular domain regulates transcription Homo sapiens R-HSA-2197563 | 3.35E-01 | 8.77E-01 | 2.63E+00 | 2.88E+00 | HES1 |
| 337 | FGFR3 mutant receptor activation Homo sapiens R-HSA-2033514 | 3.35E-01 | 8.77E-01 | 2.63E+00 | 2.88E+00 | FGF9 |
| 338 | Signaling by activated point mutants of FGFR3 Homo sapiens R-HSA-1839130 | 3.35E-01 | 8.77E-01 | 2.63E+00 | 2.88E+00 | FGF9 |
| 339 | Reduction of cytosolic Ca++ levels Homo sapiens R-HSA-418359 | 3.35E-01 | 8.77E-01 | 2.63E+00 | 2.88E+00 | ATP2B3 |
| 340 | The NLRP3 inflammasome Homo sapiens R-HSA-844456 | 3.35E-01 | 8.77E-01 | 2.63E+00 | 2.88E+00 | TXNIP |
| 341 | Regulation of TP53 Expression and Degradation Homo sapiens R-HSA-6806003 | 3.40E-01 | 8.78E-01 | 1.70E+00 | 1.84E+00 | USP2;RPS27A |
| 342 | Transmembrane transport of small molecules Homo sapiens R-HSA-382551 | 3.40E-01 | 8.78E-01 | 1.12E+00 | 1.21E+00 | SLC47A1;ATP8B2;SLC20A2;SLC22A2;TSC22D3;SLC12A1;SLC43A1;CLCNKB;WNK4;SLC3A1;ATP2B3;SLC30A1;ATP1A1;ATP1B1;SLC9A1;CLCN5;FXYD2;FTH1;BSG;SLC16A7;RPS27A;FTL |

| | | | | | | |
|---|---|---|---|---|---|---|
| 343 | Gap-filling DNA repair synthesis and ligation in TC-NER Homo sapiens R-HSA-6782210 | 3.43E-01 | 8.78E-01 | 1.47E+00 | 1.58E+00 | GTF2H2;RPS27A;POLR2L |
| 344 | Cellular responses to stress Homo sapiens R-HSA-2262752 | 3.44E-01 | 8.78E-01 | 1.15E+00 | 1.23E+00 | HSPA8;HSPA5;CITED2;GSTP1;PRDX6;ANAPC11;TXN2;PRDX3;PRDX2;PRDX5;ID1;NOX4;IGFBP7;RPS27A |
| 345 | MAPK family signaling cascades Homo sapiens R-HSA-5683057 | 3.51E-01 | 8.78E-01 | 1.17E+00 | 1.22E+00 | CCND3;FGF9;ERBB4;EGF;NEFL;HSPB1;PHB;RPS27A;FOXO3;HRAS;DUSP9 |
| 346 | Dual incision in TC-NER Homo sapiens R-HSA-6782135 | 3.52E-01 | 8.78E-01 | 1.45E+00 | 1.51E+00 | GTF2H2;RPS27A;POLR2L |
| 347 | Negative regulation of the PI3K/AKT network Homo sapiens R-HSA-199418 | 3.54E-01 | 8.78E-01 | 1.35E+00 | 1.40E+00 | FGF9;ERBB4;EGF;PIK3R1 |
| 348 | The activation of arylsulfatases Homo sapiens R-HSA-1663150 | 3.57E-01 | 8.78E-01 | 2.41E+00 | 2.49E+00 | SUMF2 |
| 349 | POU5F1 (OCT4), SOX2, NANOG activate genes related to proliferation Homo sapiens R-HSA-2892247 | 3.57E-01 | 8.78E-01 | 2.41E+00 | 2.49E+00 | TDGF1 |
| 350 | NICD traffics to nucleus Homo sapiens R-HSA-157052 | 3.57E-01 | 8.78E-01 | 2.41E+00 | 2.49E+00 | NOTCH3 |
| 351 | Notch-HLH transcription pathway Homo sapiens R-HSA-350054 | 3.57E-01 | 8.78E-01 | 2.41E+00 | 2.49E+00 | NOTCH3 |
| 352 | p75NTR recruits signalling complexes Homo sapiens R-HSA-209543 | 3.57E-01 | 8.78E-01 | 2.41E+00 | 2.49E+00 | RPS27A |
| 353 | NF-kB is activated and signals survival Homo sapiens R-HSA-209560 | 3.57E-01 | 8.78E-01 | 2.41E+00 | 2.49E+00 | RPS27A |

| | | | | | | |
|---|---|---|---|---|---|---|
| 354 | Assembly Of The HIV Virion Homo sapiens R-HSA-175474 | 3.57E-01 | 8.78E-01 | 2.41E+00 | 2.49E+00 | RPS27A |
| 355 | FGFR2c ligand binding and activation Homo sapiens R-HSA-190375 | 3.57E-01 | 8.78E-01 | 2.41E+00 | 2.49E+00 | FGF9 |
| 356 | FGFR3c ligand binding and activation Homo sapiens R-HSA-190372 | 3.57E-01 | 8.78E-01 | 2.41E+00 | 2.49E+00 | FGF9 |
| 357 | FGFR3 ligand binding and activation Homo sapiens R-HSA-190239 | 3.57E-01 | 8.78E-01 | 2.41E+00 | 2.49E+00 | FGF9 |
| 358 | Downregulation of ERBB2:ERBB3 signaling Homo sapiens R-HSA-1358803 | 3.57E-01 | 8.78E-01 | 2.41E+00 | 2.49E+00 | RPS27A |
| 359 | p38MAPK events Homo sapiens R-HSA-171007 | 3.57E-01 | 8.78E-01 | 2.41E+00 | 2.49E+00 | HRAS |
| 360 | Trafficking and processing of endosomal TLR Homo sapiens R-HSA-1679131 | 3.57E-01 | 8.78E-01 | 2.41E+00 | 2.49E+00 | HSP90B1 |
| 361 | TNFR1-induced proapoptotic signaling Homo sapiens R-HSA-5357786 | 3.57E-01 | 8.78E-01 | 2.41E+00 | 2.49E+00 | USP2 |
| 362 | Synthesis of PE Homo sapiens R-HSA-1483213 | 3.57E-01 | 8.78E-01 | 2.41E+00 | 2.49E+00 | PCYT2 |
| 363 | MAPK6/MAPK4 signaling Homo sapiens R-HSA-5687128 | 3.69E-01 | 8.84E-01 | 1.32E+00 | 1.31E+00 | CCND3;HSPB1;FOXO3;RPS27A |
| 364 | ER-Phagosome pathway Homo sapiens R-HSA-1236974 | 3.70E-01 | 8.84E-01 | 1.40E+00 | 1.39E+00 | CALR;RPS27A;B2M |

| | | | | | | |
|---|---|---|---|---|---|---|
| 365 | Dual Incision in GG-NER Homo sapiens R-HSA-5696400 | 3.76E-01 | 8.84E-01 | 1.57E+00 | 1.53E+00 | GTF2H2;RPS27A |
| 366 | GABA B receptor activation Homo sapiens R-HSA-977444 | 3.76E-01 | 8.84E-01 | 1.57E+00 | 1.53E+00 | KCNJ12;KCNJ16 |
| 367 | Activation of GABAB receptors Homo sapiens R-HSA-991365 | 3.76E-01 | 8.84E-01 | 1.57E+00 | 1.53E+00 | KCNJ12;KCNJ16 |
| 368 | Activation of NMDA receptor upon glutamate binding and postsynaptic events Homo sapiens R-HSA-442755 | 3.76E-01 | 8.84E-01 | 1.57E+00 | 1.53E+00 | NEFL;HRAS |
| 369 | Organic cation/anion/zwitterion transport Homo sapiens R-HSA-549132 | 3.79E-01 | 8.84E-01 | 2.23E+00 | 2.16E+00 | SLC22A2 |
| 370 | TP53 regulates transcription of several additional cell death genes whose specific roles in p53-dependent apoptosis remain uncertain Homo sapiens R-HSA-6803205 | 3.79E-01 | 8.84E-01 | 2.23E+00 | 2.16E+00 | TP53I3 |
| 371 | FGFR4 ligand binding and activation Homo sapiens R-HSA-190322 | 3.79E-01 | 8.84E-01 | 2.23E+00 | 2.16E+00 | FGF9 |
| 372 | Repression of WNT target genes Homo sapiens R-HSA-4641265 | 3.79E-01 | 8.84E-01 | 2.23E+00 | 2.16E+00 | TLE1 |
| 373 | Nicotinate metabolism Homo sapiens R-HSA-196807 | 3.79E-01 | 8.84E-01 | 2.23E+00 | 2.16E+00 | NAPRT |

| | | | | | | |
|---|---|---|---|---|---|---|
| 374 | Glutathione synthesis and recycling Homo sapiens R-HSA-174403 | 3.79E-01 | 8.84E-01 | 2.23E+00 | 2.16E+00 | OPLAH |
| 375 | PI-3K cascade:FGFR1 Homo sapiens R-HSA-5654689 | 3.86E-01 | 8.84E-01 | 1.24E+00 | 1.18E+00 | FGF9;ERBB4;EGF;PIK3R1;FOXO3 |
| 376 | PI-3K cascade:FGFR3 Homo sapiens R-HSA-5654710 | 3.86E-01 | 8.84E-01 | 1.24E+00 | 1.18E+00 | FGF9;ERBB4;EGF;PIK3R1;FOXO3 |
| 377 | PI3K events in ERBB4 signaling Homo sapiens R-HSA-1250342 | 3.86E-01 | 8.84E-01 | 1.24E+00 | 1.18E+00 | FGF9;ERBB4;EGF;PIK3R1;FOXO3 |
| 378 | PIP3 activates AKT signaling Homo sapiens R-HSA-1257604 | 3.86E-01 | 8.84E-01 | 1.24E+00 | 1.18E+00 | FGF9;ERBB4;EGF;PIK3R1;FOXO3 |
| 379 | PI-3K cascade:FGFR4 Homo sapiens R-HSA-5654720 | 3.86E-01 | 8.84E-01 | 1.24E+00 | 1.18E+00 | FGF9;ERBB4;EGF;PIK3R1;FOXO3 |
| 380 | PI-3K cascade:FGFR2 Homo sapiens R-HSA-5654695 | 3.86E-01 | 8.84E-01 | 1.24E+00 | 1.18E+00 | FGF9;ERBB4;EGF;PIK3R1;FOXO3 |
| 381 | Transcriptional activation of mitochondrial biogenesis Homo sapiens R-HSA-2151201 | 3.88E-01 | 8.84E-01 | 1.52E+00 | 1.44E+00 | MEF2C;PPARGC1A |
| 382 | Amyloid fiber formation Homo sapiens R-HSA-977225 | 3.98E-01 | 8.84E-01 | 1.34E+00 | 1.23E+00 | CST3;RPS27A;B2M |
| 383 | Cyclin E associated events during G1/S transition Homo sapiens R-HSA-69202 | 3.98E-01 | 8.84E-01 | 1.34E+00 | 1.23E+00 | WEE1;CCND1;RPS27A |
| 384 | Calnexin/calreticulin cycle Homo sapiens R-HSA-901042 | 3.99E-01 | 8.84E-01 | 2.07E+00 | 1.90E+00 | CALR |

| | | | | | | |
|---|---|---|---|---|---|---|
| 385 | Activation of BAD and translocation to mitochondria Homo sapiens R-HSA-111447 | 3.99E-01 | 8.84E-01 | 2.07E+00 | 1.90E+00 | BCL2 |
| 386 | Regulation of innate immune responses to cytosolic DNA Homo sapiens R-HSA-3134975 | 3.99E-01 | 8.84E-01 | 2.07E+00 | 1.90E+00 | RPS27A |
| 387 | Association of licensing factors with the pre-replicative complex Homo sapiens R-HSA-69298 | 3.99E-01 | 8.84E-01 | 2.07E+00 | 1.90E+00 | RPS27A |
| 388 | Prolactin receptor signaling Homo sapiens R-HSA-1170546 | 3.99E-01 | 8.84E-01 | 2.07E+00 | 1.90E+00 | PRLR |
| 389 | Constitutive Signaling by NOTCH1 HD Domain Mutants Homo sapiens R-HSA-2691232 | 3.99E-01 | 8.84E-01 | 2.07E+00 | 1.90E+00 | RPS27A |
| 390 | Signaling by NOTCH1 HD Domain Mutants in Cancer Homo sapiens R-HSA-2691230 | 3.99E-01 | 8.84E-01 | 2.07E+00 | 1.90E+00 | RPS27A |
| 391 | NRIF signals cell death from the nucleus Homo sapiens R-HSA-205043 | 3.99E-01 | 8.84E-01 | 2.07E+00 | 1.90E+00 | RPS27A |
| 392 | FGFR1 ligand binding and activation Homo sapiens R-HSA-190242 | 3.99E-01 | 8.84E-01 | 2.07E+00 | 1.90E+00 | FGF9 |
| 393 | CREB phosphorylation through the activation of CaMKII Homo sapiens R-HSA-442729 | 3.99E-01 | 8.84E-01 | 2.07E+00 | 1.90E+00 | NEFL |

| | | | | | | |
|---|---|---|---|---|---|---|
| 394 | Eicosanoid ligand-binding receptors Homo sapiens R-HSA-391903 | 3.99E-01 | 8.84E-01 | 2.07E+00 | 1.90E+00 | PTGER3 |
| 395 | L1CAM interactions Homo sapiens R-HSA-373760 | 4.00E-01 | 8.84E-01 | 1.26E+00 | 1.16E+00 | HSPA8;ALCAM;CNTN1;SCN1B |
| 396 | Pre-NOTCH Transcription and Translation Homo sapiens R-HSA-1912408 | 4.00E-01 | 8.84E-01 | 1.49E+00 | 1.36E+00 | NOTCH3;CCND1 |
| 397 | TNF signaling Homo sapiens R-HSA-75893 | 4.00E-01 | 8.84E-01 | 1.49E+00 | 1.36E+00 | USP2;RPS27A |
| 398 | GAB1 signalosome Homo sapiens R-HSA-180292 | 4.06E-01 | 8.84E-01 | 1.21E+00 | 1.09E+00 | FGF9;ERBB4;EGF;PIK3R1;FOXO3 |
| 399 | PI3K/AKT activation Homo sapiens R-HSA-198203 | 4.06E-01 | 8.84E-01 | 1.21E+00 | 1.09E+00 | FGF9;ERBB4;EGF;PIK3R1;FOXO3 |
| 400 | SLC-mediated transmembrane transport Homo sapiens R-HSA-425407 | 4.06E-01 | 8.84E-01 | 1.12E+00 | 1.01E+00 | SLC47A1;SLC20A2;SLC22A2;BSG;SLC43A1;SLC12A1;SLC16A7;SLC3A1;SLC30A1;SLC9A1 |
| 401 | Cyclin A:Cdk2-associated events at S phase entry Homo sapiens R-HSA-69656 | 4.07E-01 | 8.84E-01 | 1.32E+00 | 1.18E+00 | WEE1;CCND1;RPS27A |
| 402 | Non-integrin membrane-ECM interactions Homo sapiens R-HSA-3000171 | 4.12E-01 | 8.84E-01 | 1.45E+00 | 1.29E+00 | SDC4;SDC1 |
| 403 | Transcriptional regulation of pluripotent stem cells Homo sapiens R-HSA-452723 | 4.12E-01 | 8.84E-01 | 1.45E+00 | 1.29E+00 | TDGF1;POLR2L |
| 404 | Deactivation of the beta-catenin transactivating complex Homo sapiens R-HSA-3769402 | 4.12E-01 | 8.84E-01 | 1.45E+00 | 1.29E+00 | TLE1;RPS27A |

| | | | | | | |
|---|---|---|---|---|---|---|
| 405 | BMAL1:CLOCK,NPAS 2 activates circadian gene expression Homo sapiens R-HSA-1368108 | 4.12E-01 | 8.84E-01 | 1.45E+00 | 1.29E+00 | BHLHE40;PPARGC1A |
| 406 | trans-Golgi Network Vesicle Budding Homo sapiens R-HSA-199992 | 4.16E-01 | 8.84E-01 | 1.30E+00 | 1.14E+00 | HSPA8;FTH1;FTL |
| 407 | Clathrin derived vesicle budding Homo sapiens R-HSA-421837 | 4.16E-01 | 8.84E-01 | 1.30E+00 | 1.14E+00 | HSPA8;FTH1;FTL |
| 408 | Translesion synthesis by REV1 Homo sapiens R-HSA-110312 | 4.19E-01 | 8.84E-01 | 1.93E+00 | 1.68E+00 | RPS27A |
| 409 | p75NTR signals via NF-kB Homo sapiens R-HSA-193639 | 4.19E-01 | 8.84E-01 | 1.93E+00 | 1.68E+00 | RPS27A |
| 410 | TRAF6 mediated induction of TAK1 complex Homo sapiens R-HSA-937072 | 4.19E-01 | 8.84E-01 | 1.93E+00 | 1.68E+00 | RPS27A |
| 411 | Spry regulation of FGF signaling Homo sapiens R-HSA-1295596 | 4.19E-01 | 8.84E-01 | 1.93E+00 | 1.68E+00 | RPS27A |
| 412 | MAP3K8 (TPL2)-dependent MAPK1/3 activation Homo sapiens R-HSA-5684264 | 4.19E-01 | 8.84E-01 | 1.93E+00 | 1.68E+00 | RPS27A |
| 413 | Polo-like kinase mediated events Homo sapiens R-HSA-156711 | 4.19E-01 | 8.84E-01 | 1.93E+00 | 1.68E+00 | WEE1 |
| 414 | Rap1 signalling Homo sapiens R-HSA-392517 | 4.19E-01 | 8.84E-01 | 1.93E+00 | 1.68E+00 | RAP1GAP |

| | | | | | | |
|---|---|---|---|---|---|---|
| 415 | TGF-beta receptor signaling in EMT (epithelial to mesenchymal transition) Homo sapiens R-HSA-2173791 | 4.19E-01 | 8.84E-01 | 1.93E+00 | 1.68E+00 | RPS27A |
| 416 | Metabolism of folate and pterines Homo sapiens R-HSA-196757 | 4.19E-01 | 8.84E-01 | 1.93E+00 | 1.68E+00 | ALDH1L1 |
| 417 | HIV Transcription Elongation Homo sapiens R-HSA-167169 | 4.23E-01 | 8.84E-01 | 1.41E+00 | 1.21E+00 | GTF2H2;POLR2L |
| 418 | Tat-mediated elongation of the HIV-1 transcript Homo sapiens R-HSA-167246 | 4.23E-01 | 8.84E-01 | 1.41E+00 | 1.21E+00 | GTF2H2;POLR2L |
| 419 | Formation of HIV-1 elongation complex containing HIV-1 Tat Homo sapiens R-HSA-167200 | 4.23E-01 | 8.84E-01 | 1.41E+00 | 1.21E+00 | GTF2H2;POLR2L |
| 420 | Formation of Incision Complex in GG-NER Homo sapiens R-HSA-5696395 | 4.23E-01 | 8.84E-01 | 1.41E+00 | 1.21E+00 | GTF2H2;RPS27A |
| 421 | Iron uptake and transport Homo sapiens R-HSA-917937 | 4.23E-01 | 8.84E-01 | 1.41E+00 | 1.21E+00 | FTH1;FTL |
| 422 | Signaling by ERBB4 Homo sapiens R-HSA-1236394 | 4.23E-01 | 8.84E-01 | 1.09E+00 | 9.40E-01 | CXCL12;FGF9;ERBB4;EGF;NEFL;PIK3R1;PHB;RPS27A;FOXO3;HRAS;DUSP9;PRLR |
| 423 | Interleukin receptor SHC signaling Homo sapiens R-HSA-912526 | 4.34E-01 | 8.84E-01 | 1.11E+00 | 9.24E-01 | FGF9;ERBB4;EGF;NEFL;PIK3R1;PHB;RPS27A;HRAS;DUSP9 |
| 424 | Formation of HIV elongation complex in the absence of HIV Tat Homo sapiens R-HSA-167152 | 4.35E-01 | 8.84E-01 | 1.38E+00 | 1.15E+00 | GTF2H2;POLR2L |

| | | | | | | |
|---|---|---|---|---|---|---|
| 425 | RNA Polymerase II Transcription Elongation Homo sapiens R-HSA-75955 | 4.35E-01 | 8.84E-01 | 1.38E+00 | 1.15E+00 | GTF2H2;POLR2L |
| 426 | Formation of RNA Pol II elongation complex Homo sapiens R-HSA-112382 | 4.35E-01 | 8.84E-01 | 1.38E+00 | 1.15E+00 | GTF2H2;POLR2L |
| 427 | RNA Polymerase II HIV Promoter Escape Homo sapiens R-HSA-167162 | 4.35E-01 | 8.84E-01 | 1.38E+00 | 1.15E+00 | GTF2H2;POLR2L |
| 428 | RNA Polymerase II Promoter Escape Homo sapiens R-HSA-73776 | 4.35E-01 | 8.84E-01 | 1.38E+00 | 1.15E+00 | GTF2H2;POLR2L |
| 429 | RNA Polymerase II Transcription Initiation And Promoter Clearance Homo sapiens R-HSA-76042 | 4.35E-01 | 8.84E-01 | 1.38E+00 | 1.15E+00 | GTF2H2;POLR2L |
| 430 | RNA Polymerase II Transcription Initiation Homo sapiens R-HSA-75953 | 4.35E-01 | 8.84E-01 | 1.38E+00 | 1.15E+00 | GTF2H2;POLR2L |
| 431 | RNA Polymerase II Transcription Pre-Initiation And Promoter Opening Homo sapiens R-HSA-73779 | 4.35E-01 | 8.84E-01 | 1.38E+00 | 1.15E+00 | GTF2H2;POLR2L |
| 432 | HIV Transcription Initiation Homo sapiens R-HSA-167161 | 4.35E-01 | 8.84E-01 | 1.38E+00 | 1.15E+00 | GTF2H2;POLR2L |
| 433 | Phospholipase C-mediated cascade; FGFR3 Homo sapiens R-HSA-5654227 | 4.35E-01 | 8.84E-01 | 1.38E+00 | 1.15E+00 | FGF9;PDE1A |
| 434 | Zinc transporters Homo sapiens R-HSA-435354 | 4.39E-01 | 8.84E-01 | 1.81E+00 | 1.49E+00 | SLC30A1 |

| | | | | | | |
|---|---|---|---|---|---|---|
| 435 | Hyaluronan metabolism Homo sapiens R-HSA-2142845 | 4.39E-01 | 8.84E-01 | 1.81E+00 | 1.49E+00 | SLC9A1 |
| 436 | Trafficking of GluR2-containing AMPA receptors Homo sapiens R-HSA-416993 | 4.39E-01 | 8.84E-01 | 1.81E+00 | 1.49E+00 | TSPAN7 |
| 437 | N-glycan trimming in the ER and Calnexin/Calreticulin cycle Homo sapiens R-HSA-532668 | 4.39E-01 | 8.84E-01 | 1.81E+00 | 1.49E+00 | CALR |
| 438 | Translesion synthesis by POLK Homo sapiens R-HSA-5655862 | 4.39E-01 | 8.84E-01 | 1.81E+00 | 1.49E+00 | RPS27A |
| 439 | Translesion synthesis by POLI Homo sapiens R-HSA-5656121 | 4.39E-01 | 8.84E-01 | 1.81E+00 | 1.49E+00 | RPS27A |
| 440 | Activated point mutants of FGFR2 Homo sapiens R-HSA-2033519 | 4.39E-01 | 8.84E-01 | 1.81E+00 | 1.49E+00 | FGF9 |
| 441 | Unblocking of NMDA receptor, glutamate binding and activation Homo sapiens R-HSA-438066 | 4.39E-01 | 8.84E-01 | 1.81E+00 | 1.49E+00 | NEFL |
| 442 | Metabolism of porphyrins Homo sapiens R-HSA-189445 | 4.39E-01 | 8.84E-01 | 1.81E+00 | 1.49E+00 | FECH |
| 443 | ER to Golgi Anterograde Transport Homo sapiens R-HSA-199977 | 4.46E-01 | 8.95E-01 | 1.15E+00 | 9.29E-01 | DYNC2H1;TRAPPC2L;TRAPPC5;FOLR1;CTSC |
| 444 | Sphingolipid metabolism Homo sapiens R-HSA-428157 | 4.51E-01 | 8.96E-01 | 1.22E+00 | 9.74E-01 | UGT8;PSAP;SUMF2 |

| | | | | | | |
|---|---|---|---|---|---|---|
| 445 | MHC class II antigen presentation Homo sapiens R-HSA-2132295 | 4.52E-01 | 8.96E-01 | 1.17E+00 | 9.29E-01 | DYNC2H1;CTSO;KIF26A;CTSC |
| 446 | Phospholipase C-mediated cascade; FGFR4 Homo sapiens R-HSA-5654228 | 4.57E-01 | 8.96E-01 | 1.32E+00 | 1.03E+00 | FGF9;PDE1A |
| 447 | RNA Polymerase III Chain Elongation Homo sapiens R-HSA-73780 | 4.58E-01 | 8.96E-01 | 1.70E+00 | 1.33E+00 | POLR2L |
| 448 | Pre-NOTCH Processing in Golgi Homo sapiens R-HSA-1912420 | 4.58E-01 | 8.96E-01 | 1.70E+00 | 1.33E+00 | NOTCH3 |
| 449 | Oxygen-dependent proline hydroxylation of Hypoxia-inducible Factor Alpha Homo sapiens R-HSA-1234176 | 4.58E-01 | 8.96E-01 | 1.70E+00 | 1.33E+00 | RPS27A |
| 450 | Signaling by cytosolic FGFR1 fusion mutants Homo sapiens R-HSA-1839117 | 4.58E-01 | 8.96E-01 | 1.70E+00 | 1.33E+00 | PIK3R1 |
| 451 | Activation of IRF3/IRF7 mediated by TBK1/IKK epsilon Homo sapiens R-HSA-936964 | 4.58E-01 | 8.96E-01 | 1.70E+00 | 1.33E+00 | RPS27A |
| 452 | Regulation of signaling by CBL Homo sapiens R-HSA-912631 | 4.58E-01 | 8.96E-01 | 1.70E+00 | 1.33E+00 | PIK3R1 |
| 453 | Defects in vitamin and cofactor metabolism Homo sapiens R-HSA-3296482 | 4.58E-01 | 8.96E-01 | 1.70E+00 | 1.33E+00 | MCCC1 |
| 454 | Cytosolic sulfonation of small molecules Homo sapiens R-HSA-156584 | 4.58E-01 | 8.96E-01 | 1.70E+00 | 1.33E+00 | SULT1A1 |

| | | | | | | |
|---|---|---|---|---|---|---|
| 455 | Downstream signaling events of B Cell Receptor (BCR) Homo sapiens R-HSA-1168372 | 4.61E-01 | 8.96E-01 | 1.10E+00 | 8.48E-01 | FGF9;ERBB4;EGF;PIK3R1;FOXO3;RPS27A;HRAS |
| 456 | Interleukin-2 signaling Homo sapiens R-HSA-451927 | 4.67E-01 | 8.96E-01 | 1.07E+00 | 8.17E-01 | FGF9;ERBB4;EGF;NEFL;PIK3R1;PHB;RPS27A;HRAS;DUSP9 |
| 457 | Transcription-Coupled Nucleotide Excision Repair (TC-NER) Homo sapiens R-HSA-6781827 | 4.69E-01 | 8.96E-01 | 1.19E+00 | 9.02E-01 | GTF2H2;RPS27A;POLR2L |
| 458 | RNA Polymerase I Transcription Initiation Homo sapiens R-HSA-73762 | 4.69E-01 | 8.96E-01 | 1.29E+00 | 9.76E-01 | GTF2H2;POLR2L |
| 459 | Nucleotide-binding domain, leucine rich repeat containing receptor (NLR) signaling pathways Homo sapiens R-HSA-168643 | 4.69E-01 | 8.96E-01 | 1.29E+00 | 9.76E-01 | BCL2;TXNIP |
| 460 | Antigen activates B Cell Receptor (BCR) leading to generation of second messengers Homo sapiens R-HSA-983695 | 4.69E-01 | 8.96E-01 | 1.29E+00 | 9.76E-01 | PIK3R1;PIK3AP1 |
| 461 | Phospholipase C-mediated cascade: FGFR1 Homo sapiens R-HSA-5654219 | 4.69E-01 | 8.96E-01 | 1.29E+00 | 9.76E-01 | FGF9;PDE1A |
| 462 | Platelet activation, signaling and aggregation Homo sapiens R-HSA-76002 | 4.71E-01 | 8.96E-01 | 1.07E+00 | 8.03E-01 | LGALS3BP;HSPA5;EGF;APLP2;PSAP;SERPING1;PIK3R1;HRG;KNG1 |

| | | | | | | |
|---|---|---|---|---|---|---|
| 463 | GABA synthesis, release, reuptake and degradation Homo sapiens R-HSA-888590 | 4.76E-01 | 8.96E-01 | 1.61E+00 | 1.20E+00 | HSPA8 |
| 464 | Na+/Cl- dependent neurotransmitter transporters Homo sapiens R-HSA-442660 | 4.76E-01 | 8.96E-01 | 1.61E+00 | 1.20E+00 | SLC22A2 |
| 465 | Signaling by FGFR2 IIIa TM Homo sapiens R-HSA-8851708 | 4.76E-01 | 8.96E-01 | 1.61E+00 | 1.20E+00 | POLR2L |
| 466 | Regulation of TP53 Activity through Methylation Homo sapiens R-HSA-6804760 | 4.76E-01 | 8.96E-01 | 1.61E+00 | 1.20E+00 | RPS27A |
| 467 | Translesion Synthesis by POLH Homo sapiens R-HSA-110320 | 4.76E-01 | 8.96E-01 | 1.61E+00 | 1.20E+00 | RPS27A |
| 468 | Phosphorylation of the APC/C Homo sapiens R-HSA-176412 | 4.76E-01 | 8.96E-01 | 1.61E+00 | 1.20E+00 | ANAPC11 |
| 469 | Conversion from APC/C:Cdc20 to APC/C:Cdh1 in late anaphase Homo sapiens R-HSA-176407 | 4.76E-01 | 8.96E-01 | 1.61E+00 | 1.20E+00 | ANAPC11 |
| 470 | Other semaphorin interactions Homo sapiens R-HSA-416700 | 4.76E-01 | 8.96E-01 | 1.61E+00 | 1.20E+00 | SEMA6D |
| 471 | HDL-mediated lipid transport Homo sapiens R-HSA-194223 | 4.76E-01 | 8.96E-01 | 1.61E+00 | 1.20E+00 | APOE |
| 472 | Phase 4 - resting membrane potential Homo sapiens R-HSA-5576886 | 4.76E-01 | 8.96E-01 | 1.61E+00 | 1.20E+00 | KCNJ12 |

| | | | | | | |
|---|---|---|---|---|---|---|
| 473 | Senescence-Associated Secretory Phenotype (SASP) Homo sapiens R-HSA-2559582 | 4.77E-01 | 8.96E-01 | 1.17E+00 | 8.69E-01 | IGFBP7;RPS27A;ANAPC11 |
| 474 | IRS-mediated signalling Homo sapiens R-HSA-112399 | 4.78E-01 | 8.96E-01 | 1.06E+00 | 7.81E-01 | FGF9;ERBB4;EGF;RPS6;NEFL;PIK3R1;PHB;RPS27A;HRAS;DUSP9 |
| 475 | Vesicle-mediated transport Homo sapiens R-HSA-5653656 | 4.78E-01 | 8.96E-01 | 1.04E+00 | 7.65E-01 | DYNC2H1;HSPA8;TRAPPC2L;TRAPPC5;RHOBTB3;HSP90B1;TBC1D1;KIF26A;FTH1;NAA38;APOE;CALR;RPS27A;RAB6B;FOLR1;CTSC;FTL |
| 476 | Death Receptor Signalling Homo sapiens R-HSA-73887 | 4.80E-01 | 8.96E-01 | 1.26E+00 | 9.25E-01 | USP2;RPS27A |
| 477 | DAP12 signaling Homo sapiens R-HSA-2424491 | 4.80E-01 | 8.96E-01 | 1.05E+00 | 7.67E-01 | FGF9;ERBB4;EGF;PDE1A;NEFL;PIK3R1;PHB;RPS27A;FOXO3;B2M;HRAS;DUSP9 |
| 478 | NGF signalling via TRKA from the plasma membrane Homo sapiens R-HSA-187037 | 4.82E-01 | 8.96E-01 | 1.04E+00 | 7.62E-01 | NTRK2;MEF2C;EGF;PDE1A;PIK3R1;PHB;FOXO3;DUSP9;FGF9;ERBB4;NEFL;RPS27A;HRAS |
| 479 | Amino acid and oligopeptide SLC transporters Homo sapiens R-HSA-425374 | 4.90E-01 | 8.96E-01 | 1.23E+00 | 8.78E-01 | SLC43A1;SLC3A1 |
| 480 | Phospholipase C-mediated cascade; FGFR2 Homo sapiens R-HSA-5654221 | 4.90E-01 | 8.96E-01 | 1.23E+00 | 8.78E-01 | FGF9;PDE1A |
| 481 | Mitochondrial biogenesis Homo sapiens R-HSA-1592230 | 4.90E-01 | 8.96E-01 | 1.23E+00 | 8.78E-01 | MEF2C;PPARGC1A |
| 482 | Insulin receptor signalling cascade Homo sapiens R-HSA-74751 | 4.91E-01 | 8.96E-01 | 1.05E+00 | 7.43E-01 | FGF9;ERBB4;EGF;RPS6;NEFL;PIK3R1;PHB;RPS27A;HRAS;DUSP9 |
| 483 | FGFR2 ligand binding and activation Homo sapiens R-HSA-190241 | 4.93E-01 | 8.96E-01 | 1.52E+00 | 1.08E+00 | FGF9 |

| | | | | | | |
|---|---|---|---|---|---|---|
| 484 | Inhibition of the proteolytic activity of APC/C required for the onset of anaphase by mitotic spindle checkpoint components Homo sapiens R-HSA-141405 | 4.93E-01 | 8.96E-01 | 1.52E+00 | 1.08E+00 | ANAPC11 |
| 485 | Inactivation of APC/C via direct inhibition of the APC/C complex Homo sapiens R-HSA-141430 | 4.93E-01 | 8.96E-01 | 1.52E+00 | 1.08E+00 | ANAPC11 |
| 486 | Hormone-sensitive lipase (HSL)-mediated triacylglycerol hydrolysis Homo sapiens R-HSA-163560 | 4.93E-01 | 8.96E-01 | 1.52E+00 | 1.08E+00 | FABP3 |
| 487 | Signaling by Hippo Homo sapiens R-HSA-2028269 | 4.93E-01 | 8.96E-01 | 1.52E+00 | 1.08E+00 | TJP2 |
| 488 | Synthesis of PC Homo sapiens R-HSA-1483191 | 4.93E-01 | 8.96E-01 | 1.52E+00 | 1.08E+00 | CHPT1 |
| 489 | PI3K Cascade Homo sapiens R-HSA-109704 | 4.94E-01 | 8.96E-01 | 1.14E+00 | 8.06E-01 | FGF9;RPS6;PIK3R1 |
| 490 | IGF1R signaling cascade Homo sapiens R-HSA-2428924 | 4.96E-01 | 8.96E-01 | 1.04E+00 | 7.31E-01 | FGF9;ERBB4;EGF;RPS6;NEFL;PIK3R1;PHB;RPS27A;HRAS;DUSP9 |
| 491 | Signaling by Type 1 Insulin-like Growth Factor 1 Receptor (IGF1R) Homo sapiens R-HSA-2404192 | 4.96E-01 | 8.96E-01 | 1.04E+00 | 7.31E-01 | FGF9;ERBB4;EGF;RPS6;NEFL;PIK3R1;PHB;RPS27A;HRAS;DUSP9 |
| 492 | IRS-related events triggered by IGF1R Homo sapiens R-HSA-2428928 | 4.96E-01 | 8.96E-01 | 1.04E+00 | 7.31E-01 | FGF9;ERBB4;EGF;RPS6;NEFL;PIK3R1;PHB;RPS27A;HRAS;DUSP9 |

| | | | | | | |
|---|---|---|---|---|---|---|
| 493 | Ubiquitin-dependent degradation of Cyclin D1 Homo sapiens R-HSA-69229 | 5.01E-01 | 8.99E-01 | 1.21E+00 | 8.33E-01 | CCND1;RPS27A |
| 494 | Ubiquitin-dependent degradation of Cyclin D Homo sapiens R-HSA-75815 | 5.01E-01 | 8.99E-01 | 1.21E+00 | 8.33E-01 | CCND1;RPS27A |
| 495 | VEGFA-VEGFR2 Pathway Homo sapiens R-HSA-4420097 | 5.04E-01 | 8.99E-01 | 1.03E+00 | 7.05E-01 | CYFIP2;FGF9;ERBB4;EGF;NEFL;HSPB1;PIK3R1;PHB;RPS27A;HRAS;DUSP9 |
| 496 | Interleukin-3, 5 and GM-CSF signaling Homo sapiens R-HSA-512988 | 5.09E-01 | 8.99E-01 | 1.03E+00 | 6.99E-01 | FGF9;ERBB4;EGF;NEFL;PIK3R1;PHB;RPS27A;HRAS;DUSP9 |
| 497 | Nef-mediates down modulation of cell surface receptors by recruiting them to clathrin adapters Homo sapiens R-HSA-164938 | 5.10E-01 | 8.99E-01 | 1.45E+00 | 9.74E-01 | B2M |
| 498 | Mitotic Spindle Checkpoint Homo sapiens R-HSA-69618 | 5.10E-01 | 8.99E-01 | 1.45E+00 | 9.74E-01 | ANAPC11 |
| 499 | ERK/MAPK targets Homo sapiens R-HSA-198753 | 5.10E-01 | 8.99E-01 | 1.45E+00 | 9.74E-01 | MEF2C |
| 500 | Regulation of FZD by ubiquitination Homo sapiens R-HSA-4641263 | 5.10E-01 | 8.99E-01 | 1.45E+00 | 9.74E-01 | RPS27A |
| 501 | CD209 (DC-SIGN) signaling Homo sapiens R-HSA-5621575 | 5.10E-01 | 8.99E-01 | 1.45E+00 | 9.74E-01 | HRAS |
| 502 | Synthesis of very long-chain fatty acyl-CoAs Homo sapiens R-HSA-75876 | 5.10E-01 | 8.99E-01 | 1.45E+00 | 9.74E-01 | TECR |

| | | | | | | |
|---|---|---|---|---|---|---|
| 503 | Neurotransmitter Receptor Binding And Downstream Transmission In The Postsynaptic Cell Homo sapiens R-HSA-112314 | 5.16E-01 | 8.99E-01 | 1.06E+00 | 6.99E-01 | KCNJ12;TSPAN7;NEFL;KCNJ16;HRAS |
| 504 | Signaling by the B Cell Receptor (BCR) Homo sapiens R-HSA-983705 | 5.19E-01 | 8.99E-01 | 1.03E+00 | 6.76E-01 | FGF9;ERBB4;EGF;PIK3R1;RPS27A;FOXO3;PIK3AP1;HRAS |
| 505 | Antigen processing-Cross presentation Homo sapiens R-HSA-1236975 | 5.19E-01 | 8.99E-01 | 1.10E+00 | 7.21E-01 | CALR;RPS27A;B2M |
| 506 | RA biosynthesis pathway Homo sapiens R-HSA-5365859 | 5.27E-01 | 8.99E-01 | 1.38E+00 | 8.84E-01 | ALDH1A1 |
| 507 | mTORC1-mediated signalling Homo sapiens R-HSA-166208 | 5.27E-01 | 8.99E-01 | 1.38E+00 | 8.84E-01 | RPS6 |
| 508 | CD28 dependent PI3K/Akt signaling Homo sapiens R-HSA-389357 | 5.27E-01 | 8.99E-01 | 1.38E+00 | 8.84E-01 | PIK3R1 |
| 509 | Cyclin A/B1 associated events during G2/M transition Homo sapiens R-HSA-69273 | 5.27E-01 | 8.99E-01 | 1.38E+00 | 8.84E-01 | WEE1 |
| 510 | Nephrin interactions Homo sapiens R-HSA-373753 | 5.27E-01 | 8.99E-01 | 1.38E+00 | 8.84E-01 | PIK3R1 |
| 511 | SALM protein interactions at the synapse Homo sapiens R-HSA-8849932 | 5.27E-01 | 8.99E-01 | 1.38E+00 | 8.84E-01 | PTPRF |
| 512 | GRB2 events in EGFR signaling Homo sapiens R-HSA-179812 | 5.29E-01 | 8.99E-01 | 1.02E+00 | 6.51E-01 | FGF9;ERBB4;EGF;NEFL;PHB;RPS27A;HRAS;DUSP9 |
| 513 | SHC1 events in EGFR signaling Homo sapiens R-HSA-180336 | 5.29E-01 | 8.99E-01 | 1.02E+00 | 6.51E-01 | FGF9;ERBB4;EGF;NEFL;PHB;RPS27A;HRAS;DUSP9 |

| | | | | | | |
|---|---|---|---|---|---|---|
| 514 | SOS-mediated signalling Homo sapiens R-HSA-112412 | 5.29E-01 | 8.99E-01 | 1.02E+00 | 6.51E-01 | FGF9;ERBB4;EGF;NEFL;PHB;RPS27A;HRAS;DUSP9 |
| 515 | SHC1 events in ERBB4 signaling Homo sapiens R-HSA-1250347 | 5.29E-01 | 8.99E-01 | 1.02E+00 | 6.51E-01 | FGF9;ERBB4;EGF;NEFL;PHB;RPS27A;HRAS;DUSP9 |
| 516 | RAF/MAP kinase cascade Homo sapiens R-HSA-5673001 | 5.29E-01 | 8.99E-01 | 1.02E+00 | 6.51E-01 | FGF9;ERBB4;EGF;NEFL;PHB;RPS27A;HRAS;DUSP9 |
| 517 | FRS-mediated FGFR2 signaling Homo sapiens R-HSA-5654700 | 5.33E-01 | 8.99E-01 | 1.02E+00 | 6.38E-01 | FGF9;ERBB4;EGF;NEFL;PHB;RPS27A;HRAS;DUSP9 |
| 518 | FRS-mediated FGFR4 signaling Homo sapiens R-HSA-5654712 | 5.33E-01 | 8.99E-01 | 1.02E+00 | 6.38E-01 | FGF9;ERBB4;EGF;NEFL;PHB;RPS27A;HRAS;DUSP9 |
| 519 | FRS-mediated FGFR3 signaling Homo sapiens R-HSA-5654706 | 5.33E-01 | 8.99E-01 | 1.02E+00 | 6.38E-01 | FGF9;ERBB4;EGF;NEFL;PHB;RPS27A;HRAS;DUSP9 |
| 520 | FRS-mediated FGFR1 signaling Homo sapiens R-HSA-5654693 | 5.33E-01 | 8.99E-01 | 1.02E+00 | 6.38E-01 | FGF9;ERBB4;EGF;NEFL;PHB;RPS27A;HRAS;DUSP9 |
| 521 | Signaling by VEGF Homo sapiens R-HSA-194138 | 5.38E-01 | 8.99E-01 | 1.00E+00 | 6.23E-01 | CYFIP2;FGF9;ERBB4;EGF;NEFL;HSPB1;PIK3R1;PHB;RPS27A;HRAS;DUSP9 |
| 522 | DAP12 interactions Homo sapiens R-HSA-2172127 | 5.40E-01 | 8.99E-01 | 1.00E+00 | 6.16E-01 | FGF9;ERBB4;EGF;PDE1A;NEFL;PIK3R1;PHB;RPS27A;FOXO3;B2M;HRAS;DUSP9 |
| 523 | Membrane Trafficking Homo sapiens R-HSA-199991 | 5.42E-01 | 8.99E-01 | 9.98E-01 | 6.12E-01 | DYNC2H1;HSPA8;TRAPPC2L;TRAPPC5;RHOBTB3;TBC1D1;KIF26A;FTH1;NAA38;RPS27A;RAB6B;FOLR1;CTSC;FTL |
| 524 | Downstream signaling of activated FGFR2 Homo sapiens R-HSA-5654696 | 5.42E-01 | 8.99E-01 | 1.00E+00 | 6.14E-01 | FGF9;ERBB4;EGF;PDE1A;NEFL;PIK3R1;PHB;RPS27A;FOXO3;HRAS;DUSP9 |
| 525 | Downstream signaling of activated FGFR4 Homo sapiens R-HSA-5654716 | 5.42E-01 | 8.99E-01 | 1.00E+00 | 6.14E-01 | FGF9;ERBB4;EGF;PDE1A;NEFL;PIK3R1;PHB;RPS27A;FOXO3;HRAS;DUSP9 |

| | | | | | | |
|---|---|---|---|---|---|---|
| 526 | Downstream signaling of activated FGFR3 Homo sapiens R-HSA-5654708 | 5.42E-01 | 8.99E-01 | 1.00E+00 | 6.14E-01 | FGF9;ERBB4;EGF;PDE1A;NEFL;PIK3R1;PHB;RPS27A;FOXO3;HRAS;DUSP9 |
| 527 | Laminin interactions Homo sapiens R-HSA-3000157 | 5.42E-01 | 8.99E-01 | 1.32E+00 | 8.05E-01 | ITGA7 |
| 528 | RNA Polymerase III Transcription Termination Homo sapiens R-HSA-73980 | 5.42E-01 | 8.99E-01 | 1.32E+00 | 8.05E-01 | POLR2L |
| 529 | MicroRNA (miRNA) biogenesis Homo sapiens R-HSA-203927 | 5.42E-01 | 8.99E-01 | 1.32E+00 | 8.05E-01 | POLR2L |
| 530 | Abortive elongation of HIV-1 transcript in the absence of Tat Homo sapiens R-HSA-167242 | 5.42E-01 | 8.99E-01 | 1.32E+00 | 8.05E-01 | POLR2L |
| 531 | Gap-filling DNA repair synthesis and ligation in GG-NER Homo sapiens R-HSA-5696397 | 5.42E-01 | 8.99E-01 | 1.32E+00 | 8.05E-01 | RPS27A |
| 532 | RAF-independent MAPK1/3 activation Homo sapiens R-HSA-112409 | 5.42E-01 | 8.99E-01 | 1.32E+00 | 8.05E-01 | DUSP9 |
| 533 | Downregulation of SMAD2/3:SMAD4 transcriptional activity Homo sapiens R-HSA-2173795 | 5.42E-01 | 8.99E-01 | 1.32E+00 | 8.05E-01 | RPS27A |
| 534 | ARMS-mediated activation Homo sapiens R-HSA-170984 | 5.48E-01 | 8.99E-01 | 1.00E+00 | 6.03E-01 | FGF9;ERBB4;EGF;NEFL;PHB;RPS27A;HRAS;DUSP9 |
| 535 | Signalling to p38 via RIT and RIN Homo sapiens R-HSA-187706 | 5.48E-01 | 8.99E-01 | 1.00E+00 | 6.03E-01 | FGF9;ERBB4;EGF;NEFL;PHB;RPS27A;HRAS;DUSP9 |

| | | | | | | |
|---|---|---|---|---|---|---|
| 536 | Signaling by FGFR2 Homo sapiens R-HSA-5654738 | 5.48E-01 | 8.99E-01 | 9.95E-01 | 5.98E-01 | FGF9;ERBB4;EGF;PDE1A;NEFL;PIK3R1;PHB;RPS27A;FOXO3;HRAS;DUSP9;POLR2L |
| 537 | Regulation of mRNA stability by proteins that bind AU-rich elements Homo sapiens R-HSA-450531 | 5.52E-01 | 8.99E-01 | 1.05E+00 | 6.23E-01 | HSPA8;HSPB1;RPS27A |
| 538 | Cell junction organization Homo sapiens R-HSA-446728 | 5.52E-01 | 8.99E-01 | 1.05E+00 | 6.23E-01 | CLDN10;DST;CLDN14 |
| 539 | Frs2-mediated activation Homo sapiens R-HSA-170968 | 5.53E-01 | 8.99E-01 | 9.98E-01 | 5.92E-01 | FGF9;ERBB4;EGF;NEFL;PHB;RPS27A;HRAS;DUSP9 |
| 540 | IRE1alpha activates chaperones Homo sapiens R-HSA-381070 | 5.53E-01 | 8.99E-01 | 1.09E+00 | 6.48E-01 | XBP1;HSPA5 |
| 541 | GABA receptor activation Homo sapiens R-HSA-977443 | 5.53E-01 | 8.99E-01 | 1.09E+00 | 6.48E-01 | KCNJ12;KCNJ16 |
| 542 | Signaling by FGFR4 Homo sapiens R-HSA-5654743 | 5.54E-01 | 8.99E-01 | 9.92E-01 | 5.86E-01 | FGF9;ERBB4;EGF;PDE1A;NEFL;PIK3R1;PHB;RPS27A;FOXO3;HRAS;DUSP9 |
| 543 | Downstream signaling of activated FGFR1 Homo sapiens R-HSA-5654687 | 5.54E-01 | 8.99E-01 | 9.92E-01 | 5.86E-01 | FGF9;ERBB4;EGF;PDE1A;NEFL;PIK3R1;PHB;RPS27A;FOXO3;HRAS;DUSP9 |
| 544 | MAPK1/MAPK3 signaling Homo sapiens R-HSA-5684996 | 5.57E-01 | 8.99E-01 | 9.94E-01 | 5.81E-01 | FGF9;ERBB4;EGF;NEFL;PHB;RPS27A;HRAS;DUSP9 |
| 545 | Growth hormone receptor signaling Homo sapiens R-HSA-982772 | 5.58E-01 | 8.99E-01 | 1.26E+00 | 7.35E-01 | PRLR |
| 546 | Nuclear Events (kinase and transcription factor activation) Homo sapiens R-HSA-198725 | 5.58E-01 | 8.99E-01 | 1.26E+00 | 7.35E-01 | MEF2C |

| | | | | | | |
|---|---|---|---|---|---|---|
| 547 | IKK complex recruitment mediated by RIP1 Homo sapiens R-HSA-937041 | 5.58E-01 | 8.99E-01 | 1.26E+00 | 7.35E-01 | RPS27A |
| 548 | Signaling by FGFR3 Homo sapiens R-HSA-5654741 | 5.58E-01 | 8.99E-01 | 9.88E-01 | 5.77E-01 | FGF9;ERBB4;EGF;PDE1A;NEFL;PIK3R1;PHB;RPS27A;FOXO3;HRAS;DUSP9 |
| 549 | Platelet homeostasis Homo sapiens R-HSA-418346 | 5.59E-01 | 8.99E-01 | 1.03E+00 | 6.01E-01 | PDE1A;KCNMB2;ATP2B3 |
| 550 | Signaling by PDGF Homo sapiens R-HSA-186797 | 5.60E-01 | 8.99E-01 | 9.86E-01 | 5.72E-01 | FGF9;ERBB4;EGF;PDE1A;PDGFD;NEFL;PIK3R1;PHB;RPS27A;FOXO3;HRAS;DUSP9 |
| 551 | Prolonged ERK activation events Homo sapiens R-HSA-169893 | 5.62E-01 | 8.99E-01 | 9.89E-01 | 5.70E-01 | FGF9;ERBB4;EGF;NEFL;PHB;RPS27A;HRAS;DUSP9 |
| 552 | Signaling by Leptin Homo sapiens R-HSA-2586552 | 5.67E-01 | 8.99E-01 | 9.85E-01 | 5.59E-01 | FGF9;ERBB4;EGF;NEFL;PHB;RPS27A;HRAS;DUSP9 |
| 553 | Signaling by FGFR Homo sapiens R-HSA-190236 | 5.67E-01 | 8.99E-01 | 9.81E-01 | 5.56E-01 | FGF9;ERBB4;EGF;PDE1A;NEFL;PIK3R1;PHB;RPS27A;FOXO3;HRAS;DUSP9;POLR2L |
| 554 | Signaling by FGFR1 Homo sapiens R-HSA-5654736 | 5.70E-01 | 8.99E-01 | 9.79E-01 | 5.50E-01 | FGF9;ERBB4;EGF;PDE1A;NEFL;PIK3R1;PHB;RPS27A;FOXO3;HRAS;DUSP9 |
| 555 | RNA Polymerase I Chain Elongation Homo sapiens R-HSA-73777 | 5.72E-01 | 8.99E-01 | 1.05E+00 | 5.88E-01 | GTF2H2;POLR2L |
| 556 | Constitutive Signaling by NOTCH1 HD+PEST Domain Mutants Homo sapiens R-HSA-2894862 | 5.72E-01 | 8.99E-01 | 1.05E+00 | 5.88E-01 | HES1;RPS27A |
| 557 | Signaling by NOTCH1 in Cancer Homo sapiens R-HSA-2644603 | 5.72E-01 | 8.99E-01 | 1.05E+00 | 5.88E-01 | HES1;RPS27A |
| 558 | Signaling by NOTCH1 PEST Domain Mutants in Cancer Homo sapiens R-HSA-2644602 | 5.72E-01 | 8.99E-01 | 1.05E+00 | 5.88E-01 | HES1;RPS27A |

| | | | | | | |
|---|---|---|---|---|---|---|
| 559 | Constitutive Signaling by NOTCH1 PEST Domain Mutants Homo sapiens R-HSA-2644606 | 5.72E-01 | 8.99E-01 | 1.05E+00 | 5.88E-01 | HES1;RPS27A |
| 560 | Signaling by NOTCH1 HD+PEST Domain Mutants in Cancer Homo sapiens R-HSA-2894858 | 5.72E-01 | 8.99E-01 | 1.05E+00 | 5.88E-01 | HES1;RPS27A |
| 561 | Pre-NOTCH Expression and Processing Homo sapiens R-HSA-1912422 | 5.72E-01 | 8.99E-01 | 1.05E+00 | 5.88E-01 | NOTCH3;CCND1 |
| 562 | Metal ion SLC transporters Homo sapiens R-HSA-425410 | 5.72E-01 | 8.99E-01 | 1.21E+00 | 6.73E-01 | SLC30A1 |
| 563 | ATF4 activates genes Homo sapiens R-HSA-380994 | 5.72E-01 | 8.99E-01 | 1.21E+00 | 6.73E-01 | HERPUD1 |
| 564 | Constitutive Signaling by AKT1 E17K in Cancer Homo sapiens R-HSA-5674400 | 5.72E-01 | 8.99E-01 | 1.21E+00 | 6.73E-01 | FOXO3 |
| 565 | HDMs demethylate histones Homo sapiens R-HSA-3214842 | 5.72E-01 | 8.99E-01 | 1.21E+00 | 6.73E-01 | ARID5B |
| 566 | Budding and maturation of HIV virion Homo sapiens R-HSA-162588 | 5.72E-01 | 8.99E-01 | 1.21E+00 | 6.73E-01 | RPS27A |
| 567 | Sulfur amino acid metabolism Homo sapiens R-HSA-1614635 | 5.72E-01 | 8.99E-01 | 1.21E+00 | 6.73E-01 | GOT1 |
| 568 | Glycosaminoglycan metabolism Homo sapiens R-HSA-1630316 | 5.79E-01 | 9.05E-01 | 9.89E-01 | 5.41E-01 | SDC4;GPC3;SDC1;SLC9A1 |
| 569 | Signalling to RAS Homo sapiens R-HSA-167044 | 5.81E-01 | 9.05E-01 | 9.72E-01 | 5.29E-01 | FGF9;ERBB4;EGF;NEFL;PHB;RPS27A;HRAS;DUSP9 |

| | | | | | | |
|---|---|---|---|---|---|---|
| 570 | Attenuation phase Homo sapiens R-HSA-3371568 | 5.87E-01 | 9.05E-01 | 1.16E+00 | 6.17E-01 | HSPA8 |
| 571 | FGFR2 alternative splicing Homo sapiens R-HSA-6803529 | 5.87E-01 | 9.05E-01 | 1.16E+00 | 6.17E-01 | POLR2L |
| 572 | PTK6 Regulates RHO GTPases, RAS GTPase and MAP kinases Homo sapiens R-HSA-8849471 | 5.87E-01 | 9.05E-01 | 1.16E+00 | 6.17E-01 | HRAS |
| 573 | TNFR1-induced NFkappaB signaling pathway Homo sapiens R-HSA-5357956 | 5.87E-01 | 9.05E-01 | 1.16E+00 | 6.17E-01 | USP2 |
| 574 | WNT ligand biogenesis and trafficking Homo sapiens R-HSA-3238698 | 5.87E-01 | 9.05E-01 | 1.16E+00 | 6.17E-01 | WLS |
| 575 | CaM pathway Homo sapiens R-HSA-111997 | 5.87E-01 | 9.05E-01 | 1.16E+00 | 6.17E-01 | PDE1A |
| 576 | Calmodulin induced events Homo sapiens R-HSA-111933 | 5.87E-01 | 9.05E-01 | 1.16E+00 | 6.17E-01 | PDE1A |
| 577 | Downregulation of TGF-beta receptor signaling Homo sapiens R-HSA-2173788 | 5.87E-01 | 9.05E-01 | 1.16E+00 | 6.17E-01 | RPS27A |
| 578 | Downstream signal transduction Homo sapiens R-HSA-186763 | 5.90E-01 | 9.07E-01 | 9.64E-01 | 5.09E-01 | FGF9;ERBB4;EGF;PDE1A;NEFL;PIK3R1;PHB;RPS27A;FOXO3;HRAS;DUSP9 |
| 579 | VEGFR2 mediated cell proliferation Homo sapiens R-HSA-5218921 | 5.90E-01 | 9.07E-01 | 9.64E-01 | 5.09E-01 | FGF9;ERBB4;EGF;NEFL;PHB;RPS27A;HRAS;DUSP9 |
| 580 | SCF(Skp2)-mediated degradation of p27/p21 Homo sapiens R-HSA-187577 | 5.91E-01 | 9.07E-01 | 1.02E+00 | 5.34E-01 | CCND1;RPS27A |

| | | | | | | |
|---|---|---|---|---|---|---|
| 581 | Signaling by Insulin receptor Homo sapiens R-HSA-74752 | 5.94E-01 | 9.09E-01 | 9.61E-01 | 5.01E-01 | FGF9;ERBB4;EGF;RPS6;NEFL;PIK3R1;PHB;RPS27A;HRAS;DUSP9 |
| 582 | MAP kinase activation in TLR cascade Homo sapiens R-HSA-450294 | 6.00E-01 | 9.11E-01 | 9.98E-01 | 5.10E-01 | MEF2C;RPS27A |
| 583 | PI Metabolism Homo sapiens R-HSA-1483255 | 6.00E-01 | 9.11E-01 | 9.98E-01 | 5.10E-01 | PIK3R1;MTMR7 |
| 584 | RNA Polymerase III Transcription Initiation From Type 2 Promoter Homo sapiens R-HSA-76066 | 6.01E-01 | 9.11E-01 | 1.11E+00 | 5.68E-01 | POLR2L |
| 585 | Platelet calcium homeostasis Homo sapiens R-HSA-418360 | 6.01E-01 | 9.11E-01 | 1.11E+00 | 5.68E-01 | ATP2B3 |
| 586 | Phase 1 - inactivation of fast Na+ channels Homo sapiens R-HSA-5576894 | 6.01E-01 | 9.11E-01 | 1.11E+00 | 5.68E-01 | KCNIP3 |
| 587 | Peroxisomal lipid metabolism Homo sapiens R-HSA-390918 | 6.01E-01 | 9.11E-01 | 1.11E+00 | 5.68E-01 | ECI2 |
| 588 | Interferon gamma signaling Homo sapiens R-HSA-877300 | 6.05E-01 | 9.14E-01 | 9.65E-01 | 4.85E-01 | MT2A;B2M;TRIM22 |
| 589 | TP53 Regulates Transcription of DNA Repair Genes Homo sapiens R-HSA-6796648 | 6.09E-01 | 9.14E-01 | 9.81E-01 | 4.86E-01 | GTF2H2;POLR2L |
| 590 | Cell-cell junction organization Homo sapiens R-HSA-421270 | 6.09E-01 | 9.14E-01 | 9.81E-01 | 4.86E-01 | CLDN10;CLDN14 |
| 591 | Signalling to ERKs Homo sapiens R-HSA-187687 | 6.12E-01 | 9.14E-01 | 9.44E-01 | 4.63E-01 | FGF9;ERBB4;EGF;NEFL;PHB;RPS27A;HRAS;DUSP9 |

| | | | | | | |
|---|---|---|---|---|---|---|
| 592 | The role of Nef in HIV-1 replication and disease pathogenesis Homo sapiens R-HSA-164952 | 6.14E-01 | 9.14E-01 | 1.07E+00 | 5.23E-01 | B2M |
| 593 | RNA Polymerase III Transcription Initiation From Type 1 Promoter Homo sapiens R-HSA-76061 | 6.14E-01 | 9.14E-01 | 1.07E+00 | 5.23E-01 | POLR2L |
| 594 | RNA Polymerase III Transcription Initiation From Type 3 Promoter Homo sapiens R-HSA-76071 | 6.14E-01 | 9.14E-01 | 1.07E+00 | 5.23E-01 | POLR2L |
| 595 | Recognition of DNA damage by PCNA-containing replication complex Homo sapiens R-HSA-110314 | 6.14E-01 | 9.14E-01 | 1.07E+00 | 5.23E-01 | RPS27A |
| 596 | Endosomal Sorting Complex Required For Transport (ESCRT) Homo sapiens R-HSA-917729 | 6.14E-01 | 9.14E-01 | 1.07E+00 | 5.23E-01 | RPS27A |
| 597 | Ca-dependent events Homo sapiens R-HSA-111996 | 6.14E-01 | 9.14E-01 | 1.07E+00 | 5.23E-01 | PDE1A |
| 598 | Synthesis of PA Homo sapiens R-HSA-1483166 | 6.14E-01 | 9.14E-01 | 1.07E+00 | 5.23E-01 | AGPAT2 |
| 599 | Autodegradation of Cdh1 by Cdh1:APC/C Homo sapiens R-HSA-174084 | 6.18E-01 | 9.18E-01 | 9.65E-01 | 4.64E-01 | RPS27A;ANAPC11 |
| 600 | Chaperonin-mediated protein folding Homo sapiens R-HSA-390466 | 6.19E-01 | 9.19E-01 | 9.43E-01 | 4.52E-01 | TUBB2B;TUBB2A;RGS6 |

| | | | | | | |
|---|---|---|---|---|---|---|
| 601 | Synthesis of substrates in N-glycan biosythesis Homo sapiens R-HSA-446219 | 6.27E-01 | 9.19E-01 | 9.49E-01 | 4.43E-01 | GMDS;PMM1 |
| 602 | Interaction between L1 and Ankyrins Homo sapiens R-HSA-445095 | 6.27E-01 | 9.19E-01 | 1.03E+00 | 4.83E-01 | SCN1B |
| 603 | Activation of BH3-only proteins Homo sapiens R-HSA-114452 | 6.27E-01 | 9.19E-01 | 1.03E+00 | 4.83E-01 | BCL2 |
| 604 | PIWI-interacting RNA (piRNA) biogenesis Homo sapiens R-HSA-5601884 | 6.27E-01 | 9.19E-01 | 1.03E+00 | 4.83E-01 | POLR2L |
| 605 | Myogenesis Homo sapiens R-HSA-525793 | 6.27E-01 | 9.19E-01 | 1.03E+00 | 4.83E-01 | MEF2C |
| 606 | CDO in myogenesis Homo sapiens R-HSA-375170 | 6.27E-01 | 9.19E-01 | 1.03E+00 | 4.83E-01 | MEF2C |
| 607 | Diseases of signal transduction Homo sapiens R-HSA-5663202 | 6.27E-01 | 9.19E-01 | 9.33E-01 | 4.35E-01 | FGF9;ERBB4;EGF;HES1;PIK3R1;RPS27A;FOXO3;HRAS;POLR2L |
| 608 | Phospholipid metabolism Homo sapiens R-HSA-1483257 | 6.28E-01 | 9.19E-01 | 9.27E-01 | 4.32E-01 | PCYT2;CHPT1;PIK3R1;AGPAT2;MTMR7 |
| 609 | Host Interactions of HIV factors Homo sapiens R-HSA-162909 | 6.29E-01 | 9.19E-01 | 9.26E-01 | 4.29E-01 | SLC25A5;RPS27A;B2M;SLC25A4 |
| 610 | Transport to the Golgi and subsequent modification Homo sapiens R-HSA-948021 | 6.33E-01 | 9.21E-01 | 9.21E-01 | 4.21E-01 | DYNC2H1;TRAPPC2L;TRAPPC5;FOLR1;CTSC |
| 611 | Role of LAT2/NTAL/LAB on calcium mobilization Homo sapiens R-HSA-2730905 | 6.33E-01 | 9.21E-01 | 9.21E-01 | 4.21E-01 | FGF9;ERBB4;EGF;PIK3R1;FOXO3 |

| | | | | | | |
|---|---|---|---|---|---|---|
| 612 | RNA Polymerase II Pre-transcription Events Homo sapiens R-HSA-674695 | 6.35E-01 | 9.21E-01 | 9.33E-01 | 4.23E-01 | GTF2H2;POLR2L |
| 613 | Asparagine N-linked glycosylation Homo sapiens R-HSA-446203 | 6.39E-01 | 9.21E-01 | 9.21E-01 | 4.13E-01 | DYNC2H1;TRAPPC2L;PMM1;GMDS;TRAPPC5;CALR;FOLR1;CTSC |
| 614 | Amine compound SLC transporters Homo sapiens R-HSA-425428 | 6.39E-01 | 9.21E-01 | 9.98E-01 | 4.46E-01 | SLC22A2 |
| 615 | Molecules associated with elastic fibres Homo sapiens R-HSA-2129379 | 6.39E-01 | 9.21E-01 | 9.98E-01 | 4.46E-01 | BMP4 |
| 616 | Termination of translesion DNA synthesis Homo sapiens R-HSA-5656169 | 6.39E-01 | 9.21E-01 | 9.98E-01 | 4.46E-01 | RPS27A |
| 617 | MAPK targets/ Nuclear events mediated by MAP kinases Homo sapiens R-HSA-450282 | 6.39E-01 | 9.21E-01 | 9.98E-01 | 4.46E-01 | MEF2C |
| 618 | Glucagon-type ligand receptors Homo sapiens R-HSA-420092 | 6.39E-01 | 9.21E-01 | 9.98E-01 | 4.46E-01 | GCGR |
| 619 | Cell-Cell communication Homo sapiens R-HSA-1500931 | 6.41E-01 | 9.21E-01 | 9.11E-01 | 4.05E-01 | CLDN10;DST;CLDN14;PIK3R1 |
| 620 | Interferon Signaling Homo sapiens R-HSA-913531 | 6.43E-01 | 9.21E-01 | 9.13E-01 | 4.04E-01 | EGR1;MT2A;IFIT1;RPS27A;B2M;TRIM22 |
| 621 | Signaling by EGFR Homo sapiens R-HSA-177929 | 6.43E-01 | 9.21E-01 | 9.24E-01 | 4.08E-01 | FGF9;ERBB4;EGF;PDE1A;NEFL;PIK3R1;PHB;RPS27A;FOXO3;HRAS;DUSP9 |
| 622 | Signaling by SCF-KIT Homo sapiens R-HSA-1433557 | 6.49E-01 | 9.24E-01 | 9.17E-01 | 3.97E-01 | FGF9;ERBB4;EGF;NEFL;PIK3R1;PHB;RPS27A;FOXO3;HRAS;DUSP9 |

| | | | | | | |
|---|---|---|---|---|---|---|
| 623 | Trafficking of AMPA receptors Homo sapiens R-HSA-399719 | 6.51E-01 | 9.24E-01 | 9.65E-01 | 4.13E-01 | TSPAN7 |
| 624 | Glutamate Binding, Activation of AMPA Receptors and Synaptic Plasticity Homo sapiens R-HSA-399721 | 6.51E-01 | 9.24E-01 | 9.65E-01 | 4.13E-01 | TSPAN7 |
| 625 | Pausing and recovery of Tat-mediated HIV elongation Homo sapiens R-HSA-167238 | 6.51E-01 | 9.24E-01 | 9.65E-01 | 4.13E-01 | POLR2L |
| 626 | Tat-mediated HIV elongation arrest and recovery Homo sapiens R-HSA-167243 | 6.51E-01 | 9.24E-01 | 9.65E-01 | 4.13E-01 | POLR2L |
| 627 | APC/C:Cdc20 mediated degradation of Securin Homo sapiens R-HSA-174154 | 6.52E-01 | 9.24E-01 | 9.04E-01 | 3.87E-01 | RPS27A;ANAPC11 |
| 628 | Cytosolic sensors of pathogen-associated DNA Homo sapiens R-HSA-1834949 | 6.52E-01 | 9.24E-01 | 9.04E-01 | 3.87E-01 | RPS27A;POLR2L |
| 629 | Integrin cell surface interactions Homo sapiens R-HSA-216083 | 6.60E-01 | 9.24E-01 | 8.90E-01 | 3.70E-01 | BSG;ITGA7 |
| 630 | Transcription of the HIV genome Homo sapiens R-HSA-167172 | 6.60E-01 | 9.24E-01 | 8.90E-01 | 3.70E-01 | GTF2H2;POLR2L |
| 631 | Regulation of RAS by GAPs Homo sapiens R-HSA-5658442 | 6.60E-01 | 9.24E-01 | 8.90E-01 | 3.70E-01 | RPS27A;HRAS |
| 632 | Protein folding Homo sapiens R-HSA-391251 | 6.60E-01 | 9.24E-01 | 8.85E-01 | 3.67E-01 | TUBB2B;TUBB2A;RGS6 |

| | | | | | | |
|---|---|---|---|---|---|---|
| 633 | Regulation of beta-cell development Homo sapiens R-HSA-186712 | 6.63E-01 | 9.24E-01 | 9.33E-01 | 3.84E-01 | HES1 |
| 634 | HIV elongation arrest and recovery Homo sapiens R-HSA-167287 | 6.63E-01 | 9.24E-01 | 9.33E-01 | 3.84E-01 | POLR2L |
| 635 | Pausing and recovery of HIV elongation Homo sapiens R-HSA-167290 | 6.63E-01 | 9.24E-01 | 9.33E-01 | 3.84E-01 | POLR2L |
| 636 | Elongation arrest and recovery Homo sapiens R-HSA-112387 | 6.63E-01 | 9.24E-01 | 9.33E-01 | 3.84E-01 | POLR2L |
| 637 | SMAD2/SMAD3:SMAD4 heterotrimer regulates transcription Homo sapiens R-HSA-2173796 | 6.63E-01 | 9.24E-01 | 9.33E-01 | 3.84E-01 | RPS27A |
| 638 | DAG and IP3 signaling Homo sapiens R-HSA-1489509 | 6.63E-01 | 9.24E-01 | 9.33E-01 | 3.84E-01 | PDE1A |
| 639 | TGF-beta receptor signaling activates SMADs Homo sapiens R-HSA-2173789 | 6.63E-01 | 9.24E-01 | 9.33E-01 | 3.84E-01 | RPS27A |
| 640 | Glycerophospholipid biosynthesis Homo sapiens R-HSA-1483206 | 6.67E-01 | 9.26E-01 | 8.76E-01 | 3.55E-01 | PCYT2;CHPT1;AGPAT2 |
| 641 | NCAM signaling for neurite out-growth Homo sapiens R-HSA-375165 | 6.68E-01 | 9.26E-01 | 8.96E-01 | 3.62E-01 | FGF9;ERBB4;EGF;NEFL;PHB;RPS27A;HRAS;DUSP9 |
| 642 | Interferon alpha/beta signaling Homo sapiens R-HSA-909733 | 6.68E-01 | 9.26E-01 | 8.77E-01 | 3.54E-01 | EGR1;IFIT1 |

| | | | | | | |
|---|---|---|---|---|---|---|
| 643 | Mitotic G1-G1/S phases Homo sapiens R-HSA-453279 | 6.70E-01 | 9.27E-01 | 8.76E-01 | 3.51E-01 | CCND3;WEE1;CCND1;RPS27A |
| 644 | CD28 co-stimulation Homo sapiens R-HSA-389356 | 6.74E-01 | 9.31E-01 | 9.04E-01 | 3.56E-01 | PIK3R1 |
| 645 | Glucagon signaling in metabolic regulation Homo sapiens R-HSA-163359 | 6.74E-01 | 9.31E-01 | 9.04E-01 | 3.56E-01 | GCGR |
| 646 | HSF1-dependent transactivation Homo sapiens R-HSA-3371571 | 6.85E-01 | 9.40E-01 | 8.77E-01 | 3.31E-01 | HSPA8 |
| 647 | PLC-gamma1 signalling Homo sapiens R-HSA-167021 | 6.85E-01 | 9.40E-01 | 8.77E-01 | 3.31E-01 | PDE1A |
| 648 | Striated Muscle Contraction Homo sapiens R-HSA-390522 | 6.85E-01 | 9.40E-01 | 8.77E-01 | 3.31E-01 | NEB |
| 649 | Synthesis of PIPs at the plasma membrane Homo sapiens R-HSA-1660499 | 6.85E-01 | 9.40E-01 | 8.77E-01 | 3.31E-01 | PIK3R1 |
| 650 | APC/C:Cdh1 mediated degradation of Cdc20 and other APC/C:Cdh1 targeted proteins in late mitosis/early G1 Homo sapiens R-HSA-174178 | 6.91E-01 | 9.45E-01 | 8.38E-01 | 3.10E-01 | RPS27A;ANAPC11 |
| 651 | Cdc20:Phospho-APC/C mediated degradation of Cyclin A Homo sapiens R-HSA-174184 | 6.91E-01 | 9.45E-01 | 8.38E-01 | 3.10E-01 | RPS27A;ANAPC11 |
| 652 | Lysosome Vesicle Biogenesis Homo sapiens R-HSA-432720 | 6.96E-01 | 9.47E-01 | 8.51E-01 | 3.09E-01 | HSPA8 |

| | | | | | | |
|---|---|---|---|---|---|---|
| 653 | ISG15 antiviral mechanism Homo sapiens R-HSA-1169408 | 6.98E-01 | 9.47E-01 | 8.26E-01 | 2.97E-01 | IFIT1;RPS27A |
| 654 | Antiviral mechanism by IFN-stimulated genes Homo sapiens R-HSA-1169410 | 6.98E-01 | 9.47E-01 | 8.26E-01 | 2.97E-01 | IFIT1;RPS27A |
| 655 | APC:Cdc20 mediated degradation of cell cycle proteins prior to satisfation of the cell cycle checkpoint Homo sapiens R-HSA-179419 | 6.98E-01 | 9.47E-01 | 8.26E-01 | 2.97E-01 | RPS27A;ANAPC11 |
| 656 | TRAF6 Mediated Induction of proinflammatory cytokines Homo sapiens R-HSA-168180 | 6.98E-01 | 9.47E-01 | 8.26E-01 | 2.97E-01 | MEF2C;RPS27A |
| 657 | Nucleotide Excision Repair Homo sapiens R-HSA-5696398 | 7.04E-01 | 9.52E-01 | 8.26E-01 | 2.90E-01 | GTF2H2;RPS27A;POLR2L |
| 658 | NoRC negatively regulates rRNA expression Homo sapiens R-HSA-427413 | 7.06E-01 | 9.52E-01 | 8.15E-01 | 2.84E-01 | GTF2H2;POLR2L |
| 659 | RNA Polymerase III Transcription Initiation Homo sapiens R-HSA-76046 | 7.06E-01 | 9.52E-01 | 8.27E-01 | 2.88E-01 | POLR2L |
| 660 | RHO GTPases Activate WASPs and WAVEs Homo sapiens R-HSA-5663213 | 7.06E-01 | 9.52E-01 | 8.27E-01 | 2.88E-01 | CYFIP2 |
| 661 | APC/C:Cdc20 mediated degradation of mitotic proteins Homo sapiens R-HSA-176409 | 7.13E-01 | 9.57E-01 | 8.03E-01 | 2.72E-01 | RPS27A;ANAPC11 |

| | | | | | | |
|---|---|---|---|---|---|---|
| 662 | Golgi-to-ER retrograde transport Homo sapiens R-HSA-8856688 | 7.16E-01 | 9.57E-01 | 8.11E-01 | 2.71E-01 | DYNC2H1;KIF26A;RAB6B |
| 663 | Integration of energy metabolism Homo sapiens R-HSA-163685 | 7.16E-01 | 9.57E-01 | 8.11E-01 | 2.71E-01 | GCGR;SLC25A5;SLC25A4 |
| 664 | Defective B3GALTL causes Peters-plus syndrome (PpS) Homo sapiens R-HSA-5083635 | 7.16E-01 | 9.57E-01 | 8.04E-01 | 2.69E-01 | THSD7A |
| 665 | Translesion synthesis by Y family DNA polymerases bypasses lesions on DNA template Homo sapiens R-HSA-110313 | 7.16E-01 | 9.57E-01 | 8.04E-01 | 2.69E-01 | RPS27A |
| 666 | Apoptotic cleavage of cellular proteins Homo sapiens R-HSA-111465 | 7.16E-01 | 9.57E-01 | 8.04E-01 | 2.69E-01 | TJP2 |
| 667 | Intra-Golgi and retrograde Golgi-to-ER traffic Homo sapiens R-HSA-6811442 | 7.18E-01 | 9.58E-01 | 8.30E-01 | 2.75E-01 | DYNC2H1;KIF26A;RHOBTB3;NAA38;RAB6B |
| 668 | Activation of APC/C and APC/C:Cdc20 mediated degradation of mitotic proteins Homo sapiens R-HSA-176814 | 7.20E-01 | 9.59E-01 | 7.92E-01 | 2.61E-01 | RPS27A;ANAPC11 |
| 669 | O-glycosylation of TSR domain-containing proteins Homo sapiens R-HSA-5173214 | 7.25E-01 | 9.59E-01 | 7.82E-01 | 2.51E-01 | THSD7A |
| 670 | Beta defensins Homo sapiens R-HSA-1461957 | 7.25E-01 | 9.59E-01 | 7.82E-01 | 2.51E-01 | DEFB1 |

| | | | | | | |
|---|---|---|---|---|---|---|
| 671 | DNA Damage Recognition in GG-NER Homo sapiens R-HSA-5696394 | 7.25E-01 | 9.59E-01 | 7.82E-01 | 2.51E-01 | RPS27A |
| 672 | MAP2K and MAPK activation Homo sapiens R-HSA-5674135 | 7.25E-01 | 9.59E-01 | 7.82E-01 | 2.51E-01 | HRAS |
| 673 | Negative epigenetic regulation of rRNA expression Homo sapiens R-HSA-5250941 | 7.26E-01 | 9.59E-01 | 7.82E-01 | 2.50E-01 | GTF2H2;POLR2L |
| 674 | RNA Polymerase I Promoter Clearance Homo sapiens R-HSA-73854 | 7.26E-01 | 9.59E-01 | 7.82E-01 | 2.50E-01 | GTF2H2;POLR2L |
| 675 | G1/S Transition Homo sapiens R-HSA-69206 | 7.27E-01 | 9.59E-01 | 7.96E-01 | 2.54E-01 | WEE1;CCND1;RPS27A |
| 676 | Gamma carboxylation, hypusine formation and arylsulfatase activation Homo sapiens R-HSA-163841 | 7.34E-01 | 9.66E-01 | 7.61E-01 | 2.35E-01 | SUMF2 |
| 677 | mTOR signalling Homo sapiens R-HSA-165159 | 7.34E-01 | 9.66E-01 | 7.61E-01 | 2.35E-01 | RPS6 |
| 678 | Biosynthesis of the N-glycan precursor (dolichol lipid-linked oligosaccharide, LLO) and transfer to a nascent protein Homo sapiens R-HSA-446193 | 7.39E-01 | 9.66E-01 | 7.61E-01 | 2.30E-01 | GMDS;PMM1 |
| 679 | COPI-mediated anterograde transport Homo sapiens R-HSA-6807878 | 7.39E-01 | 9.66E-01 | 7.61E-01 | 2.30E-01 | DYNC2H1;FOLR1 |

| | | | | | | |
|---|---|---|---|---|---|---|
| 680 | Signalling by NGF Homo sapiens R-HSA-166520 | 7.42E-01 | 9.66E-01 | 8.58E-01 | 2.56E-01 | NTRK2;MEF2C;EGF;PDE1A;PHB;PIK3R1;FOXO3;DUSP9;FGF9;ERBB4;NEFL;RPS27A;HRAS |
| 681 | PKB-mediated events Homo sapiens R-HSA-109703 | 7.43E-01 | 9.66E-01 | 7.42E-01 | 2.20E-01 | RPS6 |
| 682 | Class C/3 (Metabotropic glutamate/pheromone receptors) Homo sapiens R-HSA-420499 | 7.43E-01 | 9.66E-01 | 7.42E-01 | 2.20E-01 | GRM1 |
| 683 | RNA Polymerase I Transcription Homo sapiens R-HSA-73864 | 7.46E-01 | 9.66E-01 | 7.51E-01 | 2.20E-01 | GTF2H2;POLR2L |
| 684 | Regulation of APC/C activators between G1/S and early anaphase Homo sapiens R-HSA-176408 | 7.46E-01 | 9.66E-01 | 7.51E-01 | 2.20E-01 | RPS27A;ANAPC11 |
| 685 | RIG-I/MDA5 mediated induction of IFN-alpha/beta pathways Homo sapiens R-HSA-168928 | 7.46E-01 | 9.66E-01 | 7.51E-01 | 2.20E-01 | PCBP2;RPS27A |
| 686 | Elastic fibre formation Homo sapiens R-HSA-1566948 | 7.52E-01 | 9.66E-01 | 7.23E-01 | 2.06E-01 | BMP4 |
| 687 | Intraflagellar transport Homo sapiens R-HSA-5620924 | 7.52E-01 | 9.66E-01 | 7.23E-01 | 2.06E-01 | DYNC2H1 |
| 688 | RNA Polymerase III Abortive And Retractive Initiation Homo sapiens R-HSA-749476 | 7.52E-01 | 9.66E-01 | 7.23E-01 | 2.06E-01 | POLR2L |
| 689 | RNA Polymerase III Transcription Homo sapiens R-HSA-74158 | 7.52E-01 | 9.66E-01 | 7.23E-01 | 2.06E-01 | POLR2L |

| | | | | | | |
|---|---|---|---|---|---|---|
| 690 | Histidine, lysine, phenylalanine, tyrosine, proline and tryptophan catabolism Homo sapiens R-HSA-6788656 | 7.52E-01 | 9.66E-01 | 7.23E-01 | 2.06E-01 | PCBD1 |
| 691 | Regulation of HSF1-mediated heat shock response Homo sapiens R-HSA-3371453 | 7.52E-01 | 9.66E-01 | 7.41E-01 | 2.11E-01 | HSPA8;HSPA5 |
| 692 | Regulation of insulin secretion Homo sapiens R-HSA-422356 | 7.52E-01 | 9.66E-01 | 7.41E-01 | 2.11E-01 | SLC25A5;SLC25A4 |
| 693 | FCERI mediated MAPK activation Homo sapiens R-HSA-2871796 | 7.53E-01 | 9.66E-01 | 8.22E-01 | 2.33E-01 | FGF9;ERBB4;EGF;NEFL;PHB;RPS27A;HRAS;DUSP9 |
| 694 | HIV Infection Homo sapiens R-HSA-162906 | 7.55E-01 | 9.66E-01 | 8.02E-01 | 2.25E-01 | GTF2H2;RPS27A;SLC25A5;B2M;SLC25A4;POLR2L |
| 695 | Signaling by Retinoic Acid Homo sapiens R-HSA-5362517 | 7.60E-01 | 9.66E-01 | 7.05E-01 | 1.93E-01 | ALDH1A1 |
| 696 | Intrinsic Pathway for Apoptosis Homo sapiens R-HSA-109606 | 7.60E-01 | 9.66E-01 | 7.05E-01 | 1.93E-01 | BCL2 |
| 697 | Viral Messenger RNA Synthesis Homo sapiens R-HSA-168325 | 7.60E-01 | 9.66E-01 | 7.05E-01 | 1.93E-01 | POLR2L |
| 698 | EPHB-mediated forward signaling Homo sapiens R-HSA-3928662 | 7.60E-01 | 9.66E-01 | 7.05E-01 | 1.93E-01 | HRAS |
| 699 | Cooperation of PDCL (PhLP1) and TRiC/CCT in G-protein beta folding Homo sapiens R-HSA-6814122 | 7.60E-01 | 9.66E-01 | 7.05E-01 | 1.93E-01 | RGS6 |

| | | | | | | |
|---|---|---|---|---|---|---|
| 700 | Global Genome Nucleotide Excision Repair (GG-NER) Homo sapiens R-HSA-5696399 | 7.64E-01 | 9.66E-01 | 7.23E-01 | 1.94E-01 | GTF2H2;RPS27A |
| 701 | Degradation of beta-catenin by the destruction complex Homo sapiens R-HSA-195253 | 7.64E-01 | 9.66E-01 | 7.23E-01 | 1.94E-01 | TLE1;RPS27A |
| 702 | Toll Like Receptor 10 (TLR10) Cascade Homo sapiens R-HSA-168142 | 7.64E-01 | 9.66E-01 | 7.23E-01 | 1.94E-01 | MEF2C;RPS27A |
| 703 | Toll Like Receptor 5 (TLR5) Cascade Homo sapiens R-HSA-168176 | 7.64E-01 | 9.66E-01 | 7.23E-01 | 1.94E-01 | MEF2C;RPS27A |
| 704 | MyD88 cascade initiated on plasma membrane Homo sapiens R-HSA-975871 | 7.64E-01 | 9.66E-01 | 7.23E-01 | 1.94E-01 | MEF2C;RPS27A |
| 705 | TP53 Regulates Transcription of Cell Death Genes Homo sapiens R-HSA-5633008 | 7.68E-01 | 9.66E-01 | 6.89E-01 | 1.81E-01 | TP53I3 |
| 706 | Voltage gated Potassium channels Homo sapiens R-HSA-1296072 | 7.68E-01 | 9.66E-01 | 6.89E-01 | 1.81E-01 | KCNC4 |
| 707 | TRAF6 mediated induction of NFkB and MAP kinases upon TLR7/8 or 9 activation Homo sapiens R-HSA-975138 | 7.70E-01 | 9.66E-01 | 7.14E-01 | 1.87E-01 | MEF2C;RPS27A |
| 708 | Class B/2 (Secretin family receptors) Homo sapiens R-HSA-373080 | 7.76E-01 | 9.66E-01 | 7.05E-01 | 1.79E-01 | GCGR;PTH2R |
| 709 | Kinesins Homo sapiens R-HSA-983189 | 7.76E-01 | 9.66E-01 | 6.73E-01 | 1.71E-01 | KIF26A |

| | | | | | | |
|---|---|---|---|---|---|---|
| 710 | Transcriptional activity of SMAD2/SMAD3:SMAD4 heterotrimer Homo sapiens R-HSA-2173793 | 7.76E-01 | 9.66E-01 | 6.73E-01 | 1.71E-01 | RPS27A |
| 711 | Interleukin-1 signaling Homo sapiens R-HSA-446652 | 7.76E-01 | 9.66E-01 | 6.73E-01 | 1.71E-01 | RPS27A |
| 712 | PLC beta mediated events Homo sapiens R-HSA-112043 | 7.76E-01 | 9.66E-01 | 6.73E-01 | 1.71E-01 | PDE1A |
| 713 | Phase 0 - rapid depolarisation Homo sapiens R-HSA-5576892 | 7.76E-01 | 9.66E-01 | 6.73E-01 | 1.71E-01 | SCN1B |
| 714 | Inositol phosphate metabolism Homo sapiens R-HSA-1483249 | 7.76E-01 | 9.66E-01 | 6.73E-01 | 1.71E-01 | NUDT4 |
| 715 | Rho GTPase cycle Homo sapiens R-HSA-194840 | 7.78E-01 | 9.66E-01 | 7.28E-01 | 1.82E-01 | ARHGAP18;MYO9A;ARHGAP24 |
| 716 | APC/C-mediated degradation of cell cycle proteins Homo sapiens R-HSA-174143 | 7.81E-01 | 9.66E-01 | 6.96E-01 | 1.72E-01 | RPS27A;ANAPC11 |
| 717 | Regulation of mitotic cell cycle Homo sapiens R-HSA-453276 | 7.81E-01 | 9.66E-01 | 6.96E-01 | 1.72E-01 | RPS27A;ANAPC11 |
| 718 | MyD88 dependent cascade initiated on endosome Homo sapiens R-HSA-975155 | 7.81E-01 | 9.66E-01 | 6.96E-01 | 1.72E-01 | MEF2C;RPS27A |
| 719 | Toll Like Receptor 7/8 (TLR7/8) Cascade Homo sapiens R-HSA-168181 | 7.81E-01 | 9.66E-01 | 6.96E-01 | 1.72E-01 | MEF2C;RPS27A |
| 720 | Gastrin-CREB signalling pathway via PKC and MAPK Homo sapiens R-HSA-881907 | 7.82E-01 | 9.66E-01 | 8.24E-01 | 2.03E-01 | FGF9;ERBB4;EGF;GCGR;NEFL;PHB;PIK3R1;RPS27A;DUSP9;HRAS;KNG1;GRM1 |

| | | | | | | |
|---|---|---|---|---|---|---|
| 721 | RMTs methylate histone arginines Homo sapiens R-HSA-3214858 | 7.84E-01 | 9.66E-01 | 6.57E-01 | 1.60E-01 | CCND1 |
| 722 | G-protein mediated events Homo sapiens R-HSA-112040 | 7.84E-01 | 9.66E-01 | 6.57E-01 | 1.60E-01 | PDE1A |
| 723 | S Phase Homo sapiens R-HSA-69242 | 7.88E-01 | 9.70E-01 | 7.16E-01 | 1.71E-01 | WEE1;CCND1;RPS27A |
| 724 | Cellular Senescence Homo sapiens R-HSA-2559583 | 7.90E-01 | 9.71E-01 | 7.36E-01 | 1.73E-01 | ID1;IGFBP7;RPS27A;ANAPC11 |
| 725 | DNA Damage Bypass Homo sapiens R-HSA-73893 | 7.91E-01 | 9.71E-01 | 6.43E-01 | 1.51E-01 | RPS27A |
| 726 | Regulation of PLK1 Activity at G2/M Transition Homo sapiens R-HSA-2565942 | 7.92E-01 | 9.71E-01 | 6.80E-01 | 1.58E-01 | TUBB;RPS27A |
| 727 | Toll Like Receptor 9 (TLR9) Cascade Homo sapiens R-HSA-168138 | 7.97E-01 | 9.75E-01 | 6.72E-01 | 1.52E-01 | MEF2C;RPS27A |
| 728 | Defensins Homo sapiens R-HSA-1461973 | 7.98E-01 | 9.75E-01 | 6.29E-01 | 1.42E-01 | DEFB1 |
| 729 | G beta:gamma signalling through PI3Kgamma Homo sapiens R-HSA-392451 | 8.05E-01 | 9.82E-01 | 6.15E-01 | 1.34E-01 | PIK3R1 |
| 730 | Late Phase of HIV Life Cycle Homo sapiens R-HSA-162599 | 8.05E-01 | 9.82E-01 | 6.93E-01 | 1.50E-01 | GTF2H2;RPS27A;POLR2L |
| 731 | Regulation of activated PAK-2p34 by proteasome mediated degradation Homo sapiens R-HSA-211733 | 8.11E-01 | 9.87E-01 | 6.02E-01 | 1.26E-01 | RPS27A |

| | | | | | | |
|---|---|---|---|---|---|---|
| 732 | MyD88:Mal cascade initiated on plasma membrane Homo sapiens R-HSA-166058 | 8.17E-01 | 9.87E-01 | 6.42E-01 | 1.30E-01 | MEF2C;RPS27A |
| 733 | Toll Like Receptor TLR1:TLR2 Cascade Homo sapiens R-HSA-168179 | 8.17E-01 | 9.87E-01 | 6.42E-01 | 1.30E-01 | MEF2C;RPS27A |
| 734 | Toll Like Receptor TLR6:TLR2 Cascade Homo sapiens R-HSA-168188 | 8.17E-01 | 9.87E-01 | 6.42E-01 | 1.30E-01 | MEF2C;RPS27A |
| 735 | Toll Like Receptor 2 (TLR2) Cascade Homo sapiens R-HSA-181438 | 8.17E-01 | 9.87E-01 | 6.42E-01 | 1.30E-01 | MEF2C;RPS27A |
| 736 | CDK-mediated phosphorylation and removal of Cdc6 Homo sapiens R-HSA-69017 | 8.17E-01 | 9.87E-01 | 5.90E-01 | 1.19E-01 | RPS27A |
| 737 | Regulation of Apoptosis Homo sapiens R-HSA-169911 | 8.17E-01 | 9.87E-01 | 5.90E-01 | 1.19E-01 | RPS27A |
| 738 | Vpu mediated degradation of CD4 Homo sapiens R-HSA-180534 | 8.24E-01 | 9.87E-01 | 5.78E-01 | 1.12E-01 | RPS27A |
| 739 | Autodegradation of the E3 ubiquitin ligase COP1 Homo sapiens R-HSA-349425 | 8.24E-01 | 9.87E-01 | 5.78E-01 | 1.12E-01 | RPS27A |
| 740 | Cargo trafficking to the periciliary membrane Homo sapiens R-HSA-5620920 | 8.24E-01 | 9.87E-01 | 5.78E-01 | 1.12E-01 | CYS1 |
| 741 | G-protein beta:gamma signalling Homo sapiens R-HSA-397795 | 8.24E-01 | 9.87E-01 | 5.78E-01 | 1.12E-01 | PIK3R1 |

| | | | | | | |
|---|---|---|---|---|---|---|
| 742 | Apoptotic execution phase Homo sapiens R-HSA-75153 | 8.24E-01 | 9.87E-01 | 5.78E-01 | 1.12E-01 | TJP2 |
| 743 | p53-Independent DNA Damage Response Homo sapiens R-HSA-69610 | 8.29E-01 | 9.87E-01 | 5.67E-01 | 1.06E-01 | RPS27A |
| 744 | p53-Independent G1/S DNA damage checkpoint Homo sapiens R-HSA-69613 | 8.29E-01 | 9.87E-01 | 5.67E-01 | 1.06E-01 | RPS27A |
| 745 | Ubiquitin Mediated Degradation of Phosphorylated Cdc25A Homo sapiens R-HSA-69601 | 8.29E-01 | 9.87E-01 | 5.67E-01 | 1.06E-01 | RPS27A |
| 746 | Developmental Biology Homo sapiens R-HSA-1266738 | 8.32E-01 | 9.87E-01 | 8.28E-01 | 1.52E-01 | SREBF1;HSPA8;MEF2C;EGF;SEMA6D;LPL;PHB;FOXO3;DUSP9;CCND3;ALCAM;FGF9;ERBB4;CNTN1;NEFL;HES1;RPS27A;PPARGC1A;TDGF1;HRAS;SCN1B;POLR2L |
| 747 | Post-translational protein modification Homo sapiens R-HSA-597592 | 8.32E-01 | 9.87E-01 | 7.95E-01 | 1.46E-01 | DYNC2H1;GALNT14;TRAPPC2L;PMM1;GALNT16;TRAPPC5;SUMF2;MUC1;GMDS;THSD7A;CALR;MUC15;FOLR1;CTSC |
| 748 | Cellular response to heat stress Homo sapiens R-HSA-3371556 | 8.35E-01 | 9.87E-01 | 6.15E-01 | 1.11E-01 | HSPA8;HSPA5 |
| 749 | XBP1(S) activates chaperone genes Homo sapiens R-HSA-381038 | 8.35E-01 | 9.87E-01 | 5.56E-01 | 1.00E-01 | XBP1 |
| 750 | Vif-mediated degradation of APOBEC3G Homo sapiens R-HSA-180585 | 8.35E-01 | 9.87E-01 | 5.56E-01 | 1.00E-01 | RPS27A |
| 751 | GPVI-mediated activation cascade Homo sapiens R-HSA-114604 | 8.35E-01 | 9.87E-01 | 5.56E-01 | 1.00E-01 | PIK3R1 |

| | | | | | | |
|---|---|---|---|---|---|---|
| 752 | MyD88-independent TLR3/TLR4 cascade Homo sapiens R-HSA-166166 | 8.39E-01 | 9.87E-01 | 6.08E-01 | 1.06E-01 | MEF2C;RPS27A |
| 753 | Toll Like Receptor 3 (TLR3) Cascade Homo sapiens R-HSA-168164 | 8.39E-01 | 9.87E-01 | 6.08E-01 | 1.06E-01 | MEF2C;RPS27A |
| 754 | TRIF-mediated TLR3/TLR4 signaling Homo sapiens R-HSA-937061 | 8.39E-01 | 9.87E-01 | 6.08E-01 | 1.06E-01 | MEF2C;RPS27A |
| 755 | Assembly of collagen fibrils and other multimeric structures Homo sapiens R-HSA-2022090 | 8.41E-01 | 9.87E-01 | 5.45E-01 | 9.46E-02 | DST |
| 756 | SCF-beta-TrCP mediated degradation of Emi1 Homo sapiens R-HSA-174113 | 8.41E-01 | 9.87E-01 | 5.45E-01 | 9.46E-02 | RPS27A |
| 757 | Stabilization of p53 Homo sapiens R-HSA-69541 | 8.41E-01 | 9.87E-01 | 5.45E-01 | 9.46E-02 | RPS27A |
| 758 | Degradation of AXIN Homo sapiens R-HSA-4641257 | 8.41E-01 | 9.87E-01 | 5.45E-01 | 9.46E-02 | RPS27A |
| 759 | ECM proteoglycans Homo sapiens R-HSA-3000178 | 8.46E-01 | 9.88E-01 | 5.35E-01 | 8.95E-02 | ITGA7 |
| 760 | Hh mutants that don't undergo autocatalytic processing are degraded by ERAD Homo sapiens R-HSA-5362768 | 8.46E-01 | 9.88E-01 | 5.35E-01 | 8.95E-02 | RPS27A |
| 761 | Downstream TCR signaling Homo sapiens R-HSA-202424 | 8.48E-01 | 9.88E-01 | 5.95E-01 | 9.85E-02 | PIK3R1;RPS27A |

| | | | | | | |
|---|---|---|---|---|---|---|
| 762 | Hedgehog 'off' state Homo sapiens R-HSA-5610787 | 8.48E-01 | 9.88E-01 | 5.95E-01 | 9.85E-02 | DYNC2H1;RPS27A |
| 763 | Toll-Like Receptors Cascades Homo sapiens R-HSA-168898 | 8.51E-01 | 9.88E-01 | 6.32E-01 | 1.02E-01 | MEF2C;RPS27A;HSP90B1 |
| 764 | Degradation of DVL Homo sapiens R-HSA-4641258 | 8.51E-01 | 9.88E-01 | 5.25E-01 | 8.47E-02 | RPS27A |
| 765 | Chemokine receptors bind chemokines Homo sapiens R-HSA-380108 | 8.51E-01 | 9.88E-01 | 5.25E-01 | 8.47E-02 | CXCL12 |
| 766 | Fc epsilon receptor (FCERI) signaling Homo sapiens R-HSA-2454202 | 8.54E-01 | 9.88E-01 | 7.48E-01 | 1.18E-01 | FGF9;ERBB4;EGF;NEFL;PHB;PIK3R1;RPS27A;FOXO3;DUSP9;HRAS |
| 767 | Factors involved in megakaryocyte development and platelet production Homo sapiens R-HSA-983231 | 8.54E-01 | 9.88E-01 | 6.27E-01 | 9.89E-02 | WEE1;KIF26A;AKAP1 |
| 768 | HIV Life Cycle Homo sapiens R-HSA-162587 | 8.54E-01 | 9.88E-01 | 6.27E-01 | 9.89E-02 | GTF2H2;RPS27A;POLR2L |
| 769 | Hh mutants abrogate ligand secretion Homo sapiens R-HSA-5387390 | 8.56E-01 | 9.88E-01 | 5.16E-01 | 8.01E-02 | RPS27A |
| 770 | G alpha (s) signalling events Homo sapiens R-HSA-418555 | 8.57E-01 | 9.88E-01 | 6.23E-01 | 9.58E-02 | PDE1A;GCGR;PTH2R |
| 771 | Formation of the beta-catenin:TCF transactivating complex Homo sapiens R-HSA-201722 | 8.61E-01 | 9.88E-01 | 5.07E-01 | 7.59E-02 | TLE1 |
| 772 | B-WICH complex positively regulates rRNA expression Homo sapiens R-HSA-5250924 | 8.61E-01 | 9.88E-01 | 5.07E-01 | 7.59E-02 | POLR2L |

| | | | | | | |
|---|---|---|---|---|---|---|
| 773 | Recruitment and ATM-mediated phosphorylation of repair and signaling proteins at DNA double strand breaks Homo sapiens R-HSA-5693565 | 8.61E-01 | 9.88E-01 | 5.07E-01 | 7.59E-02 | RPS27A |
| 774 | NIK-->noncanonical NF-kB signaling Homo sapiens R-HSA-5676590 | 8.61E-01 | 9.88E-01 | 5.07E-01 | 7.59E-02 | RPS27A |
| 775 | CDT1 association with the CDC6:ORC:origin complex Homo sapiens R-HSA-68827 | 8.61E-01 | 9.88E-01 | 5.07E-01 | 7.59E-02 | RPS27A |
| 776 | DNA Double Strand Break Response Homo sapiens R-HSA-5693606 | 8.66E-01 | 9.88E-01 | 4.98E-01 | 7.19E-02 | RPS27A |
| 777 | Degradation of GLI1 by the proteasome Homo sapiens R-HSA-5610780 | 8.66E-01 | 9.88E-01 | 4.98E-01 | 7.19E-02 | RPS27A |
| 778 | GLI3 is processed to GLI3R by the proteasome Homo sapiens R-HSA-5610785 | 8.66E-01 | 9.88E-01 | 4.98E-01 | 7.19E-02 | RPS27A |
| 779 | Degradation of GLI2 by the proteasome Homo sapiens R-HSA-5610783 | 8.66E-01 | 9.88E-01 | 4.98E-01 | 7.19E-02 | RPS27A |
| 780 | The role of GTSE1 in G2/M progression after G2 checkpoint Homo sapiens R-HSA-8852276 | 8.66E-01 | 9.88E-01 | 4.98E-01 | 7.19E-02 | RPS27A |

| | | | | | | |
|---|---|---|---|---|---|---|
| 781 | Translocation of GLUT4 to the plasma membrane Homo sapiens R-HSA-1445148 | 8.70E-01 | 9.90E-01 | 4.90E-01 | 6.81E-02 | TBC1D1 |
| 782 | Role of phospholipids in phagocytosis Homo sapiens R-HSA-2029485 | 8.70E-01 | 9.90E-01 | 4.90E-01 | 6.81E-02 | PIK3R1 |
| 783 | Dectin-1 mediated noncanonical NF-kB signaling Homo sapiens R-HSA-5607761 | 8.74E-01 | 9.93E-01 | 4.82E-01 | 6.46E-02 | RPS27A |
| 784 | Cell death signalling via NRAGE, NRIF and NADE Homo sapiens R-HSA-204998 | 8.74E-01 | 9.93E-01 | 4.82E-01 | 6.46E-02 | RPS27A |
| 785 | Axon guidance Homo sapiens R-HSA-422475 | 8.82E-01 | 9.97E-01 | 7.44E-01 | 9.34E-02 | HSPA8;EGF;SEMA6D;PHB;DUSP9;ALCAM;FGF9;ERBB4;CNTN1;NEFL;RPS27A;HRAS;SCN1B |
| 786 | p53-Dependent G1/S DNA damage checkpoint Homo sapiens R-HSA-69580 | 8.83E-01 | 9.97E-01 | 4.66E-01 | 5.81E-02 | RPS27A |
| 787 | p53-Dependent G1 DNA Damage Response Homo sapiens R-HSA-69563 | 8.83E-01 | 9.97E-01 | 4.66E-01 | 5.81E-02 | RPS27A |
| 788 | Asymmetric localization of PCP proteins Homo sapiens R-HSA-4608870 | 8.83E-01 | 9.97E-01 | 4.66E-01 | 5.81E-02 | RPS27A |
| 789 | G alpha (q) signalling events Homo sapiens R-HSA-416476 | 8.85E-01 | 9.97E-01 | 6.17E-01 | 7.54E-02 | GCGR;PIK3R1;KNG1;GRM1 |
| 790 | HDR through Homologous Recombination (HRR) Homo sapiens R-HSA-5685942 | 8.87E-01 | 9.97E-01 | 4.59E-01 | 5.51E-02 | RPS27A |

| | | | | | | |
|---|---|---|---|---|---|---|
| 791 | Hedgehog ligand biogenesis Homo sapiens R-HSA-5358346 | 8.87E-01 | 9.97E-01 | 4.59E-01 | 5.51E-02 | RPS27A |
| 792 | G1/S DNA Damage Checkpoints Homo sapiens R-HSA-69615 | 8.90E-01 | 9.97E-01 | 4.51E-01 | 5.24E-02 | RPS27A |
| 793 | Activated TLR4 signalling Homo sapiens R-HSA-166054 | 8.92E-01 | 9.97E-01 | 5.25E-01 | 5.98E-02 | MEF2C;RPS27A |
| 794 | Activation of NF-kappaB in B cells Homo sapiens R-HSA-1169091 | 8.94E-01 | 9.97E-01 | 4.44E-01 | 4.97E-02 | RPS27A |
| 795 | PPARA activates gene expression Homo sapiens R-HSA-1989781 | 8.95E-01 | 9.97E-01 | 5.20E-01 | 5.76E-02 | SREBF1;PPARGC1A |
| 796 | Assembly of the pre-replicative complex Homo sapiens R-HSA-68867 | 8.98E-01 | 9.97E-01 | 4.38E-01 | 4.72E-02 | RPS27A |
| 797 | Semaphorin interactions Homo sapiens R-HSA-373755 | 8.98E-01 | 9.97E-01 | 4.38E-01 | 4.72E-02 | SEMA6D |
| 798 | Epigenetic regulation of gene expression Homo sapiens R-HSA-212165 | 9.01E-01 | 9.97E-01 | 5.11E-01 | 5.34E-02 | GTF2H2;POLR2L |
| 799 | RNA Polymerase I, RNA Polymerase III, and Mitochondrial Transcription Homo sapiens R-HSA-504046 | 9.03E-01 | 9.97E-01 | 5.06E-01 | 5.15E-02 | GTF2H2;POLR2L |
| 800 | Regulation of lipid metabolism by Peroxisome proliferator-activated receptor alpha (PPARalpha) Homo sapiens R-HSA-400206 | 9.03E-01 | 9.97E-01 | 5.06E-01 | 5.15E-02 | SREBF1;PPARGC1A |

| | | | | | | |
|---|---|---|---|---|---|---|
| 801 | Loss of Nlp from mitotic centrosomes Homo sapiens R-HSA-380259 | 9.04E-01 | 9.97E-01 | 4.25E-01 | 4.27E-02 | TUBB |
| 802 | Loss of proteins required for interphase microtubule organization?from the centrosome Homo sapiens R-HSA-380284 | 9.04E-01 | 9.97E-01 | 4.25E-01 | 4.27E-02 | TUBB |
| 803 | Switching of origins to a post-replicative state Homo sapiens R-HSA-69052 | 9.08E-01 | 9.97E-01 | 4.19E-01 | 4.06E-02 | RPS27A |
| 804 | Orc1 removal from chromatin Homo sapiens R-HSA-68949 | 9.08E-01 | 9.97E-01 | 4.19E-01 | 4.06E-02 | RPS27A |
| 805 | TCR signaling Homo sapiens R-HSA-202403 | 9.08E-01 | 9.97E-01 | 4.97E-01 | 4.78E-02 | PIK3R1;RPS27A |
| 806 | Signaling by Interleukins Homo sapiens R-HSA-449147 | 9.10E-01 | 9.97E-01 | 6.76E-01 | 6.35E-02 | FGF9;ERBB4;EGF;NEFL;PHB;PIK3R1;RPS27A;DUSP9;HRAS |
| 807 | Separation of Sister Chromatids Homo sapiens R-HSA-2467813 | 9.11E-01 | 9.97E-01 | 5.44E-01 | 5.10E-02 | RPS27;RPS27A;ANAPC11 |
| 808 | Apoptosis Homo sapiens R-HSA-109581 | 9.13E-01 | 9.97E-01 | 5.41E-01 | 4.94E-02 | BCL2;RPS27A;TJP2 |
| 809 | Fcgamma receptor (FCGR) dependent phagocytosis Homo sapiens R-HSA-2029480 | 9.13E-01 | 9.97E-01 | 4.89E-01 | 4.43E-02 | CYFIP2;PIK3R1 |
| 810 | Costimulation by the CD28 family Homo sapiens R-HSA-388841 | 9.14E-01 | 9.97E-01 | 4.07E-01 | 3.67E-02 | PIK3R1 |
| 811 | Transcriptional regulation by small RNAs Homo sapiens R-HSA-5578749 | 9.14E-01 | 9.97E-01 | 4.07E-01 | 3.67E-02 | POLR2L |

| | | | | | | |
|---|---|---|---|---|---|---|
| 812 | Removal of licensing factors from origins Homo sapiens R-HSA-69300 | 9.14E-01 | 9.97E-01 | 4.07E-01 | 3.67E-02 | RPS27A |
| 813 | AURKA Activation by TPX2 Homo sapiens R-HSA-8854518 | 9.14E-01 | 9.97E-01 | 4.07E-01 | 3.67E-02 | TUBB |
| 814 | Extracellular matrix organization Homo sapiens R-HSA-1474244 | 9.14E-01 | 9.97E-01 | 6.23E-01 | 5.58E-02 | BMP4;SDC4;DST;BSG;SDC1;ITGA7 |
| 815 | Positive epigenetic regulation of rRNA expression Homo sapiens R-HSA-5250913 | 9.17E-01 | 9.97E-01 | 4.01E-01 | 3.49E-02 | POLR2L |
| 816 | Signaling by TGF-beta Receptor Complex Homo sapiens R-HSA-170834 | 9.17E-01 | 9.97E-01 | 4.01E-01 | 3.49E-02 | RPS27A |
| 817 | Toll Like Receptor 4 (TLR4) Cascade Homo sapiens R-HSA-166016 | 9.18E-01 | 9.97E-01 | 4.81E-01 | 4.12E-02 | MEF2C;RPS27A |
| 818 | Programmed Cell Death Homo sapiens R-HSA-5357801 | 9.19E-01 | 9.97E-01 | 5.31E-01 | 4.49E-02 | BCL2;RPS27A;TJP2 |
| 819 | C-type lectin receptors (CLRs) Homo sapiens R-HSA-5621481 | 9.20E-01 | 9.97E-01 | 4.77E-01 | 3.97E-02 | RPS27A;HRAS |
| 820 | Regulation of DNA replication Homo sapiens R-HSA-69304 | 9.22E-01 | 9.97E-01 | 3.90E-01 | 3.16E-02 | RPS27A |
| 821 | G alpha (12/13) signalling events Homo sapiens R-HSA-416482 | 9.22E-01 | 9.97E-01 | 3.90E-01 | 3.16E-02 | PIK3R1 |
| 822 | Cytokine Signaling in Immune system Homo sapiens R-HSA-1280215 | 9.26E-01 | 9.97E-01 | 7.11E-01 | 5.43E-02 | EGR1;EGF;PHB;PIK3R1;IFIT1;DUSP9;PRLR;MT2A;FGF9;ERBB4;NEFL;RPS27A;B2M;HRAS;TRIM22 |

| | | | | | | |
|---|---|---|---|---|---|---|
| 823 | RNA polymerase II transcribes snRNA genes Homo sapiens R-HSA-6807505 | 9.27E-01 | 9.97E-01 | 3.80E-01 | 2.87E-02 | POLR2L |
| 824 | Adaptive Immune System Homo sapiens R-HSA-1280218 | 9.29E-01 | 9.97E-01 | 7.32E-01 | 5.42E-02 | DYNC2H1;HSPA5;CD81;EGF;ZBTB16;PIK3R1;FOXO3;RAP1GAP;ANAPC11;CTSO;FGF9;KIF26A;ERBB4;CALR;RPS27A;PIK3AP1;B2M;HRAS;CTSC |
| 825 | G2/M DNA damage checkpoint Homo sapiens R-HSA-69473 | 9.30E-01 | 9.97E-01 | 3.75E-01 | 2.73E-02 | WEE1 |
| 826 | Mitotic Anaphase Homo sapiens R-HSA-68882 | 9.31E-01 | 9.97E-01 | 5.09E-01 | 3.61E-02 | RPS27;RPS27A;ANAPC11 |
| 827 | G2/M Transition Homo sapiens R-HSA-69275 | 9.31E-01 | 9.97E-01 | 5.09E-01 | 3.61E-02 | WEE1;TUBB;RPS27A |
| 828 | Centrosome maturation Homo sapiens R-HSA-380287 | 9.32E-01 | 9.97E-01 | 3.70E-01 | 2.60E-02 | TUBB |
| 829 | Recruitment of mitotic centrosome proteins and complexes Homo sapiens R-HSA-380270 | 9.32E-01 | 9.97E-01 | 3.70E-01 | 2.60E-02 | TUBB |
| 830 | Mitotic Metaphase and Anaphase Homo sapiens R-HSA-2555396 | 9.33E-01 | 9.97E-01 | 5.06E-01 | 3.50E-02 | RPS27;RPS27A;ANAPC11 |
| 831 | Mitotic G2-G2/M phases Homo sapiens R-HSA-453274 | 9.35E-01 | 9.97E-01 | 5.03E-01 | 3.39E-02 | WEE1;TUBB;RPS27A |
| 832 | COPI-dependent Golgi-to-ER retrograde traffic Homo sapiens R-HSA-6811434 | 9.37E-01 | 9.97E-01 | 3.61E-01 | 2.37E-02 | KIF26A |
| 833 | Processing of DNA double-strand break ends Homo sapiens R-HSA-5693607 | 9.37E-01 | 9.97E-01 | 3.61E-01 | 2.37E-02 | RPS27A |

| | | | | | | |
|---|---|---|---|---|---|---|
| 834 | Opioid Signalling Homo sapiens R-HSA-111885 | 9.37E-01 | 9.97E-01 | 3.61E-01 | 2.37E-02 | PDE1A |
| 835 | Innate Immune System Homo sapiens R-HSA-168249 | 9.37E-01 | 9.97E-01 | 7.27E-01 | 4.71E-02 | CYFIP2;MEF2C;EGF;PDE1A;DEFB1;PHB;PIK3R1;FOXO3;DUSP9;HSP90B1;FGF9;ERBB4;PCBP2;BCL2;TXNIP;NEFL;RPS27A;B2M;HRAS;POLR2L |
| 836 | DNA Replication Pre-Initiation Homo sapiens R-HSA-69002 | 9.39E-01 | 9.97E-01 | 3.56E-01 | 2.26E-02 | RPS27A |
| 837 | M/G1 Transition Homo sapiens R-HSA-68874 | 9.39E-01 | 9.97E-01 | 3.56E-01 | 2.26E-02 | RPS27A |
| 838 | p75 NTR receptor-mediated signalling Homo sapiens R-HSA-193704 | 9.41E-01 | 9.97E-01 | 3.52E-01 | 2.15E-02 | RPS27A |
| 839 | Generic Transcription Pathway Homo sapiens R-HSA-212436 | 9.41E-01 | 9.97E-01 | 7.23E-01 | 4.38E-02 | COX8A;NOTCH3;COX7B;NDUFA4;COX4I1;USP2;GTF2H2;COX5B;COX6C;COX6A1;COX7C;COX5A;COX6B1;PRDX2;PRDX5;TP53I3;ZNF208;ZNF439;RPS27A;POLR2L |
| 840 | Class I MHC mediated antigen processing & presentation Homo sapiens R-HSA-983169 | 9.44E-01 | 9.97E-01 | 5.77E-01 | 3.33E-02 | HSPA5;ZBTB16;CALR;RPS27A;B2M;ANAPC11 |
| 841 | Signaling by Hedgehog Homo sapiens R-HSA-5358351 | 9.44E-01 | 9.97E-01 | 4.30E-01 | 2.46E-02 | DYNC2H1;RPS27A |
| 842 | Collagen formation Homo sapiens R-HSA-1474290 | 9.45E-01 | 9.97E-01 | 3.44E-01 | 1.96E-02 | DST |
| 843 | Hedgehog 'on' state Homo sapiens R-HSA-5632684 | 9.45E-01 | 9.97E-01 | 3.44E-01 | 1.96E-02 | RPS27A |
| 844 | Cell Cycle Checkpoints Homo sapiens R-HSA-69620 | 9.45E-01 | 9.97E-01 | 4.83E-01 | 2.73E-02 | WEE1;RPS27A;ANAPC11 |
| 845 | Assembly of the primary cilium Homo sapiens R-HSA-5617833 | 9.52E-01 | 9.98E-01 | 4.69E-01 | 2.33E-02 | DYNC2H1;TUBB;CYS1 |

| | | | | | | |
|---|---|---|---|---|---|---|
| 846 | Activation of anterior HOX genes in hindbrain development during early embryogenesis Homo sapiens R-HSA-5617472 | 9.52E-01 | 9.98E-01 | 3.28E-01 | 1.62E-02 | POLR2L |
| 847 | Activation of HOX genes during differentiation Homo sapiens R-HSA-5619507 | 9.52E-01 | 9.98E-01 | 3.28E-01 | 1.62E-02 | POLR2L |
| 848 | Regulation of TP53 Activity through Phosphorylation Homo sapiens R-HSA-6804756 | 9.52E-01 | 9.98E-01 | 3.28E-01 | 1.62E-02 | RPS27A |
| 849 | Oxidative Stress Induced Senescence Homo sapiens R-HSA-2559580 | 9.55E-01 | 9.98E-01 | 3.21E-01 | 1.48E-02 | RPS27A |
| 850 | PCP/CE pathway Homo sapiens R-HSA-4086400 | 9.55E-01 | 9.98E-01 | 3.21E-01 | 1.48E-02 | RPS27A |
| 851 | Peptide ligand-binding receptors Homo sapiens R-HSA-375276 | 9.58E-01 | 9.98E-01 | 4.54E-01 | 1.93E-02 | CXCL12;PSAP;KNG1 |
| 852 | TNFR2 non-canonical NF-kB pathway Homo sapiens R-HSA-5668541 | 9.61E-01 | 9.98E-01 | 3.07E-01 | 1.23E-02 | RPS27A |
| 853 | EPH-Ephrin signaling Homo sapiens R-HSA-2682334 | 9.61E-01 | 9.98E-01 | 3.07E-01 | 1.23E-02 | HRAS |
| 854 | G alpha (i) signalling events Homo sapiens R-HSA-418594 | 9.61E-01 | 9.98E-01 | 4.87E-01 | 1.93E-02 | CXCL12;PTGER3;RGS6;KNG1 |
| 855 | G2/M Checkpoints Homo sapiens R-HSA-69481 | 9.63E-01 | 9.98E-01 | 3.89E-01 | 1.48E-02 | WEE1;RPS27A |
| 856 | Synthesis of DNA Homo sapiens R-HSA-69239 | 9.63E-01 | 9.98E-01 | 3.00E-01 | 1.13E-02 | RPS27A |

| | | | | | | |
|---|---|---|---|---|---|---|
| 857 | Anchoring of the basal body to the plasma membrane Homo sapiens R-HSA-5620912 | 9.63E-01 | 9.98E-01 | 3.00E-01 | 1.13E-02 | TUBB |
| 858 | Regulation of actin dynamics for phagocytic cup formation Homo sapiens R-HSA-2029482 | 9.63E-01 | 9.98E-01 | 3.00E-01 | 1.13E-02 | CYFIP2 |
| 859 | Regulation of TP53 Activity Homo sapiens R-HSA-5633007 | 9.64E-01 | 9.98E-01 | 3.87E-01 | 1.43E-02 | USP2;RPS27A |
| 860 | CLEC7A (Dectin-1) signaling Homo sapiens R-HSA-5607764 | 9.66E-01 | 9.98E-01 | 2.94E-01 | 1.03E-02 | RPS27A |
| 861 | Resolution of Sister Chromatid Cohesion Homo sapiens R-HSA-2500257 | 9.66E-01 | 9.98E-01 | 2.94E-01 | 1.03E-02 | RPS27 |
| 862 | Gene Silencing by RNA Homo sapiens R-HSA-211000 | 9.68E-01 | 9.99E-01 | 2.88E-01 | 9.40E-03 | POLR2L |
| 863 | Immunoregulatory interactions between a Lymphoid and a non-Lymphoid cell Homo sapiens R-HSA-198933 | 9.69E-01 | 9.99E-01 | 3.72E-01 | 1.15E-02 | CD81;B2M |
| 864 | tRNA processing Homo sapiens R-HSA-72306 | 9.70E-01 | 9.99E-01 | 2.83E-01 | 8.60E-03 | HSD17B10 |
| 865 | DNA Replication Homo sapiens R-HSA-69306 | 9.72E-01 | 1.00E+00 | 2.77E-01 | 7.87E-03 | RPS27A |
| 866 | Degradation of the extracellular matrix Homo sapiens R-HSA-1474228 | 9.73E-01 | 1.00E+00 | 2.75E-01 | 7.53E-03 | BSG |
| 867 | Mitotic Prometaphase Homo sapiens R-HSA-68877 | 9.74E-01 | 1.00E+00 | 2.72E-01 | 7.20E-03 | RPS27 |

| | | | | | | |
|---|---|---|---|---|---|---|
| 868 | FCERI mediated NF-kB activation Homo sapiens R-HSA-2871837 | 9.77E-01 | 1.00E+00 | 2.62E-01 | 6.04E-03 | RPS27A |
| 869 | HDR through Homologous Recombination (HR) or Single Strand Annealing (SSA) Homo sapiens R-HSA-5693567 | 9.78E-01 | 1.00E+00 | 2.60E-01 | 5.78E-03 | RPS27A |
| 870 | RHO GTPases Activate Formins Homo sapiens R-HSA-5663220 | 9.79E-01 | 1.00E+00 | 2.55E-01 | 5.30E-03 | RPS27 |
| 871 | Homology Directed Repair Homo sapiens R-HSA-5693538 | 9.82E-01 | 1.00E+00 | 2.46E-01 | 4.46E-03 | RPS27A |
| 872 | DNA Repair Homo sapiens R-HSA-73894 | 9.87E-01 | 1.00E+00 | 4.08E-01 | 5.37E-03 | POLN;GTF2H2;RPS27A;POLR2L |
| 873 | TCF dependent signaling in response to WNT Homo sapiens R-HSA-201681 | 9.91E-01 | 1.00E+00 | 2.92E-01 | 2.61E-03 | TLE1;RPS27A |
| 874 | Beta-catenin independent WNT signaling Homo sapiens R-HSA-3858494 | 9.93E-01 | 1.00E+00 | 2.01E-01 | 1.49E-03 | RPS27A |
| 875 | DNA Double-Strand Break Repair Homo sapiens R-HSA-5693532 | 9.93E-01 | 1.00E+00 | 2.00E-01 | 1.43E-03 | RPS27A |
| 876 | Antigen processing: Ubiquitination & Proteasome degradation Homo sapiens R-HSA-983168 | 9.93E-01 | 1.00E+00 | 3.35E-01 | 2.38E-03 | ZBTB16;RPS27A;ANAPC11 |
| 877 | GPCR ligand binding Homo sapiens R-HSA-500792 | 9.93E-01 | 1.00E+00 | 4.55E-01 | 3.09E-03 | CXCL12;GCGR;PSAP;PTGER3;PTH2R;KNG1;GRM1 |

| | | | | | | |
|---|---|---|---|---|---|---|
| 878 | Immune System Homo sapiens R-HSA-168256 | 9.94E-01 | 1.00E+00 | 6.72E-01 | 4.32E-03 | CYFIP2;CD81;PDE1A;DEFB1;PHB;PIK3R1;FOXO3;IFIT1;RAP1GAP;ANAPC11;HSP90B1;MT2A;CTSO;FGF9;ERBB4;PCBP2;NEFL;RPS27A;B2M;HRAS;TRIM22;CTSC;POLR2L;DYNC2H1;EGR1;MEF2C;HSPA5;EGF;ZBTB16;DUSP9;PRLR;KIF26A;BCL2;TXNIP;CALR;PIK3AP1 |
| 879 | M Phase Homo sapiens R-HSA-68886 | 9.94E-01 | 1.00E+00 | 3.25E-01 | 1.85E-03 | RPS27;RPS27A;ANAPC11 |
| 880 | Signaling by Rho GTPases Homo sapiens R-HSA-194315 | 9.95E-01 | 1.00E+00 | 3.95E-01 | 2.15E-03 | CYFIP2;RPS27;ARHGAP18;MYO9A;ARHGAP24 |
| 881 | Class A/1 (Rhodopsin-like receptors) Homo sapiens R-HSA-373076 | 9.95E-01 | 1.00E+00 | 3.59E-01 | 1.79E-03 | CXCL12;PTGER3;PSAP;KNG1 |
| 882 | Cell Cycle, Mitotic Homo sapiens R-HSA-69278 | 9.95E-01 | 1.00E+00 | 4.39E-01 | 2.14E-03 | CCND3;WEE1;RPS27;CCND1;TUBB;RPS27A;ANAPC11 |
| 883 | Chromatin organization Homo sapiens R-HSA-4839726 | 9.96E-01 | 1.00E+00 | 2.56E-01 | 1.01E-03 | CCND1;ARID5B |
| 884 | Chromatin modifying enzymes Homo sapiens R-HSA-3247509 | 9.96E-01 | 1.00E+00 | 2.56E-01 | 1.01E-03 | CCND1;ARID5B |
| 885 | Signaling by Wnt Homo sapiens R-HSA-195721 | 9.97E-01 | 1.00E+00 | 2.94E-01 | 7.86E-04 | TLE1;RPS27A;WLS |
| 886 | RHO GTPase Effectors Homo sapiens R-HSA-195258 | 9.98E-01 | 1.00E+00 | 2.26E-01 | 3.69E-04 | CYFIP2;RPS27 |
| 887 | Cell Cycle Homo sapiens R-HSA-1640170 | 1.00E+00 | 1.00E+00 | 3.56E-01 | 1.49E-04 | CCND3;WEE1;RPS27;CCND1;TUBB;RPS27A;ANAPC11 |
| 888 | Signal Transduction Homo sapiens R-HSA-162582 | 1.00E+00 | 1.00E+00 | 5.54E-01 | 7.78E-06 | CYFIP2;HSPB1;GRM1;CCND3;FGF9;CCND1;NEFL;HRAS;RGS6;WLS;DYNC2H1;MEF2C;TLE1;RPS6;USP2;PTH2R;DUSP9;PRLR;ALDH1A1;NOTCH3;KHDRBS3;SDC4;PDE1A;HPN;PTGER3;LPL;ARHGAP18;PHB;PIK3R1;FOXO3;KNG1;ERBB4;PDGFD;PSAP;GPC3;HES1;APOE;RPS27A;POLR2L;NTRK2;EGF;GCGR;MYO9A;ARHGAP24;CXCL12;RPS27;CNTN1;SDC1;TJP2 |

| | | | | | | |
|---|---|---|---|---|---|---|
| 889 | Signaling by GPCR Homo sapiens R-HSA-372790 | 1.00E+00 | 1.00E+00 | 3.92E-01 | 2.91E-06 | EGF;PDE1A;GCGR;PTGER3;PTH2R;PHB;PIK3R1;DUSP9;KNG1;GRM1;CXCL12;FGF9;ERBB4;PSAP;NEFL;RPS27A;HRAS;RGS6 |
| 890 | GPCR downstream signaling Homo sapiens R-HSA-388396 | 1.00E+00 | 1.00E+00 | 2.57E-01 | 1.28E-06 | CXCL12;PDE1A;GCGR;PTGER3;PTH2R;PIK3R1;KNG1;RGS6;GRM1 |

**Pathways enriched by genes suppressed with SGLT2 inhibitor in principal cells**

| | Term | P value | Adjusted P value | Odds Ratio | Combined Score | Genes |
|---|---|---|---|---|---|---|
| 1 | Transmembrane transport of small molecules Homo sapiens R-HSA-382551 | 5.76E-05 | 3.91E-02 | 1.75E+00 | 1.71E+01 | SLC35B3;SLC44A3;SLC44A4;SLC44A1;AQP6;SLC2A1;ATP2A3;SLC4A4;AQP3;SLC2A6;HK1;SLC22A15;NIPAL3;FLVCR1;SLC39A7;ATP7A;SLC39A1;SLC16A3;SLC12A7;ABCC3;ABCC4;SLC15A1;TSC22D3;SLC6A13;SLC39A10;ATP11C;SLC6A12;ATP1B1;NUP93;SLC7A6;ADCY9;SLC9A6;SLCO4A1;NUP50;RAF1;ABCG1;TTYH3;SLC41A2;ATP1A1;SLC5A3;SLC7A2;SLC17A5;ATP6V0A2;ANO10;ATP6V0A4;ASIC2;SLC38A2;SLC14A2;SLC35A3;ABCA6;ABCA3;SLC33A1;EIF2S2;TPCN1;CLCN7;SLC4A7;CLCN4;FXYD4;OSTM1;SLCO3A1;TRPV4;SLCO4C1;SLCO1A2;ATP13A2;CALM1 |
| 2 | SLC-mediated transmembrane transport Homo sapiens R-HSA-425407 | 1.09E-04 | 3.91E-02 | 2.13E+00 | 1.94E+01 | SLC35B3;SLC44A3;SLC44A4;SLC44A1;SLC41A2;SLC2A1;SLC4A4;SLC5A3;SLC2A6;SLC7A2;HK1;SLC22A15;SLC17A5;SLC39A7;SLC12A7;SLC39A1;SLC16A3;SLC38A2;SLC15A1;SLC14A2;SLC35A3;SLC33A1;SLC6A13;SLC6A12;SLC39A10;NUP93;SLC4A7;SLC7A6;SLCO3A1;SLC9A6;SLCO4A1;NUP50;SLCO4C1;SLCO1A2;CALM1 |
| 3 | Interferon alpha/beta signaling Homo sapiens R-HSA-909733 | 1.33E-04 | 3.91E-02 | 3.65E+00 | 3.26E+01 | IFITM3;IFNAR2;IFITM1;IFITM2;ISG15;HLA-E;BST2;OAS1;IRF3;OAS2;IRF1;GBP2;IFNAR1;IP6K2 |
| 4 | Collagen formation Homo sapiens R-HSA-1474290 | 1.39E-04 | 3.91E-02 | 3.27E+00 | 2.90E+01 | COL16A1;MMP7;CD151;DST;COL14A1;COL11A1;COL12A1;LOXL4;PLOD2;CTSS;LOXL2;P4HA1;P4HA2;COL4A4;COL4A3;PLEC |
| 5 | Extracellular matrix organization Homo sapiens R-HSA-1474244 | 3.16E-04 | 6.52E-02 | 2.00E+00 | 1.61E+01 | ITGB1;COL16A1;CD151;ITGB5;COL14A1;COL11A1;COL12A1;NTN4;LOXL4;PLOD2;FBLN1;FGF2;NID2;CAPN15;CTSS;LOXL2;CAPN8;CAPN6;ITGAV;TGFB1;MMP7;DST;LAMB2;ACTN1;DCN;P4HA1;P4HA2;COL4A4;ITGA10;COL4A3;PECAM1;FMOD;AGRN;DDR2;PLEC |
| 6 | Non-integrin membrane-ECM interactions Homo sapiens R-HSA-3000171 | 3.49E-04 | 6.52E-02 | 4.39E+00 | 3.50E+01 | ITGB1;TGFB1;ITGB5;LAMB2;ACTN1;NTN4;ITGAV;AGRN;FGF2;DDR2 |

| | | | | | | |
|---|---|---|---|---|---|---|
| 7 | Intra-Golgi and retrograde Golgi-to-ER traffic Homo sapiens R-HSA-6811442 | 8.52E-04 | 1.37E-01 | 2.19E+00 | 1.55E+01 | DYNC1H1;NBAS;SYS1;TMF1;DCTN2;KDELR1;GALNT2;M6PR;PLA2G4A;COPZ2;GCC2;KIF23;DYNC1LI2;MAN2A2;KIFC2;KIF5B;VPS53;KIF26B;KIF13B;MAN1C1;RAB9A;KIF21A;MAN1A1;KIFAP3 |
| 8 | Syndecan interactions Homo sapiens R-HSA-3000170 | 1.50E-03 | 2.01E-01 | 6.01E+00 | 3.91E+01 | ITGB1;TGFB1;ITGB5;ACTN1;ITGAV;FGF2 |
| 9 | Nuclear Receptor transcription pathway Homo sapiens R-HSA-383280 | 1.75E-03 | 2.01E-01 | 3.43E+00 | 2.18E+01 | NR4A2;NR4A3;THRA;VDR;NR1H4;RORA;PPARG;NRBF2;NR1D1;ESR2 |
| 10 | Bile acid and bile salt metabolism Homo sapiens R-HSA-194068 | 1.79E-03 | 2.01E-01 | 4.16E+00 | 2.63E+01 | ABCC3;CYP39A1;ACOX2;AKR1C1;SLCO1A2;NR1H4;AKR1C3;AKR1C2 |
| 11 | RA biosynthesis pathway Homo sapiens R-HSA-5365859 | 2.58E-03 | 2.32E-01 | 5.26E+00 | 3.14E+01 | ALDH1A3;RDH11;RDH10;AKR1C3;RDH13;ALDH8A1 |
| 12 | Assembly of collagen fibrils and other multimeric structures Homo sapiens R-HSA-2022090 | 2.74E-03 | 2.32E-01 | 3.19E+00 | 1.88E+01 | MMP7;CD151;DST;COL4A4;COL11A1;COL4A3;LOXL4;CTSS;LOXL2;PLEC |
| 13 | Transport of glucose and other sugars, bile salts and organic acids, metal ions and amine compounds Homo sapiens R-HSA-425366 | 2.82E-03 | 2.32E-01 | 2.45E+00 | 1.44E+01 | SLC14A2;SLC44A3;SLC44A4;SLC44A1;SLC41A2;SLC6A13;SLC2A1;SLC6A12;SLC39A10;SLC5A3;SLC2A6;SLC22A15;SLC39A7;SLC39A1;SLC16A3 |
| 14 | Interferon Signaling Homo sapiens R-HSA-913531 | 2.95E-03 | 2.32E-01 | 1.97E+00 | 1.15E+01 | IFITM3;IFNAR2;IFITM1;SP100;IFITM2;ISG15;HLA-E;NUP93;BST2;OAS1;IRF3;TRIM5;OAS2;IRF1;NUP50;HLA-DRA;TRIM38;GBP2;EIF4G3;GBP4;HLA-DPA1;HLA-DQB1;IFNAR1;IP6K2 |

| | | | | | | |
|---|---|---|---|---|---|---|
| 15 | Hemostasis Homo sapiens R-HSA-109582 | 3.10E-03 | 2.32E-01 | 1.54E+00 | 8.88E+00 | ITGB1;KDM1A;DOCK8;ATP2A3;CLU;GRB14;KIF5B;KIF13B;TIMP3;ITGAV;KIF21A;FAM3C;KIFAP3;SLC16A3;SRGN;ABCC4;ANXA2;SYK;ACTN1;F2R;PLA2G4A;KIF23;ADRA2C;ATP1B1;SLC7A6;KIFC2;IRF3;LCK;IRF1;PECAM1;RAF1;PDE9A;GRB7;HDAC1;PDE1B;ITPR2;PIK3R3;TFPI;LRP8;RAP1B;KCNMB4;CD58;S100A10;CD74;ANGPT2;YES1;TGFB1;ITGA10;MAFG;KIF26B;CD9;ESAM;MAFK;CALM1 |
| 16 | Membrane Trafficking Homo sapiens R-HSA-199991 | 3.48E-03 | 2.44E-01 | 1.61E+00 | 9.14E+00 | NBAS;TMF1;SYS1;DCTN2;KDELR1;CTSZ;GCC2;MIA3;CLINT1;MAN2A2;VPS53;KIF5B;GJA4;KIF13B;MAN1C1;KIF21A;MAN1A1;GJA8;KIFAP3;SPTAN1;SPTBN1;DYNC1H1;SORT1;GALNT2;TRAPPC10;M6PR;COPZ2;PLA2G4A;ANK2;KIF23;ANK3;TPD52L1;DYNC1LI2;GJB1;KIFC2;GJB3;GOLGB1;KIF26B;RAB13;EXOC4;RAB9A;DNASE2;CALM1 |
| 17 | Transport of organic anions Homo sapiens R-HSA-879518 | 4.46E-03 | 2.75E-01 | 8.01E+00 | 4.34E+01 | SLCO3A1;SLCO4A1;SLCO4C1;SLCO1A2 |
| 18 | SMAD2/3 Phosphorylation Motif Mutants in Cancer Homo sapiens R-HSA-3304356 | 5.09E-03 | 2.75E-01 | 1.40E+01 | 7.40E+01 | TGFB1;TGFBR1;TGFBR2 |
| 19 | Choline catabolism Homo sapiens R-HSA-6798163 | 5.09E-03 | 2.75E-01 | 1.40E+01 | 7.40E+01 | DMGDH;SLC44A1;CHDH |
| 20 | Transport to the Golgi and subsequent modification Homo sapiens R-HSA-948021 | 5.73E-03 | 2.75E-01 | 1.98E+00 | 1.02E+01 | DYNC1H1;ST6GAL1;DCTN2;KDELR1;FUCA1;TRAPPC10;CTSZ;COPZ2;ANK2;MIA3;ANK3;DYNC1LI2;FUT8;MAN2A2;GOLGB1;MGAT4B;MAN1C1;MAN1A1;SPTAN1;SPTBN1 |
| 21 | Signaling by Retinoic Acid Homo sapiens R-HSA-5362517 | 5.94E-03 | 2.75E-01 | 3.30E+00 | 1.69E+01 | ALDH1A3;RDH11;RDH10;PDK4;AKR1C3;RDH13;ALDH8A1;PDK1 |
| 22 | BMAL1:CLOCK,NPAS2 activates circadian gene expression Homo sapiens R-HSA-1368108 | 5.94E-03 | 2.75E-01 | 3.30E+00 | 1.69E+01 | PER1;CHD9;TGS1;RORA;TBL1X;NR1D1;HIF1A;NPAS2 |

| | | | | | | |
|---|---|---|---|---|---|---|
| 23 | Apoptotic execution phase Homo sapiens R-HSA-75153 | 6.11E-03 | 2.75E-01 | 3.01E+00 | 1.53E+01 | DSP;H1F0;GSN;STK24;VIM;SPTAN1;PAK2;HIST1H1C;PLEC |
| 24 | PECAM1 interactions Homo sapiens R-HSA-210990 | 6.34E-03 | 2.75E-01 | 7.01E+00 | 3.55E+01 | YES1;LCK;PECAM1;ITGAV |
| 25 | Caspase-mediated cleavage of cytoskeletal proteins Homo sapiens R-HSA-264870 | 6.34E-03 | 2.75E-01 | 7.01E+00 | 3.55E+01 | GSN;VIM;SPTAN1;PLEC |
| 26 | Golgi-to-ER retrograde transport Homo sapiens R-HSA-8856688 | 6.37E-03 | 2.75E-01 | 2.22E+00 | 1.12E+01 | DYNC1H1;NBAS;DCTN2;KDELR1;GALNT2;PLA2G4A;COPZ2;KIF23;DYNC1LI2;KIFC2;KIF5B;KIF26B;KIF13B;KIF21A;KIFAP3 |
| 27 | Cell surface interactions at the vascular wall Homo sapiens R-HSA-202733 | 7.13E-03 | 2.79E-01 | 2.26E+00 | 1.12E+01 | ITGB1;GRB7;CD74;YES1;ANGPT2;ATP1B1;SLC7A6;GRB14;LCK;PECAM1;ESAM;ITGAV;CD58;SLC16A3 |
| 28 | RORA activates gene expression Homo sapiens R-HSA-1368082 | 7.69E-03 | 2.79E-01 | 4.01E+00 | 1.95E+01 | CHD9;TGS1;RORA;TBL1X;NR1D1;NPAS2 |
| 29 | Synthesis of bile acids and bile salts Homo sapiens R-HSA-192105 | 7.69E-03 | 2.79E-01 | 4.01E+00 | 1.95E+01 | CYP39A1;ACOX2;AKR1C1;NR1H4;AKR1C3;AKR1C2 |
| 30 | MHC class II antigen presentation Homo sapiens R-HSA-2132295 | 8.46E-03 | 2.79E-01 | 2.21E+00 | 1.06E+01 | CD74;DYNC1H1;DCTN2;KIF23;CTSS;DYNC1LI2;HLA-DMA;HLA-DMB;KIF5B;HLA-DRA;CTSH;KIFAP3;HLA-DPA1;HLA-DQB1 |
| 31 | Loss of Function of TGFBR1 in Cancer Homo sapiens R-HSA-3656534 | 8.47E-03 | 2.79E-01 | 1.05E+01 | 5.01E+01 | TGFB1;TGFBR1;TGFBR2 |
| 32 | TGFBR1 KD Mutants in Cancer Homo sapiens R-HSA-3656532 | 8.47E-03 | 2.79E-01 | 1.05E+01 | 5.01E+01 | TGFB1;TGFBR1;TGFBR2 |
| 33 | Loss of Function of SMAD2/3 in Cancer Homo sapiens R-HSA-3304349 | 8.47E-03 | 2.79E-01 | 1.05E+01 | 5.01E+01 | TGFB1;TGFBR1;TGFBR2 |

| | | | | | | |
|---|---|---|---|---|---|---|
| 34 | SMAD2/3 MH2 Domain Mutants in Cancer Homo sapiens R-HSA-3315487 | 8.47E-03 | 2.79E-01 | 1.05E+01 | 5.01E+01 | TGFB1;TGFBR1;TGFBR2 |
| 35 | Interferon gamma signaling Homo sapiens R-HSA-877300 | 8.70E-03 | 2.79E-01 | 2.28E+00 | 1.08E+01 | SP100;HLA-E;OAS1;IRF3;TRIM5;OAS2;IRF1;HLA-DRA;TRIM38;GBP2;GBP4;HLA-DPA1;HLA-DQB1 |
| 36 | Signal transduction by L1 Homo sapiens R-HSA-445144 | 1.09E-02 | 3.32E-01 | 4.38E+00 | 1.98E+01 | ITGB1;CSNK2A2;ITGAV;EGFR;FGFR1 |
| 37 | L1CAM interactions Homo sapiens R-HSA-373760 | 1.13E-02 | 3.32E-01 | 2.20E+00 | 9.88E+00 | ITGB1;CSNK2A2;RANBP9;ANK2;ANK3;EGFR;DPYSL2;ITGA10;ITGAV;CD24;SPTAN1;SPTBN1;FGFR1 |
| 38 | Synthesis of bile acids and bile salts via 24-hydroxycholesterol Homo sapiens R-HSA-193775 | 1.15E-02 | 3.32E-01 | 5.61E+00 | 2.50E+01 | CYP39A1;AKR1C1;AKR1C3;AKR1C2 |
| 39 | Synthesis of Prostaglandins (PG) and Thromboxanes (TX) Homo sapiens R-HSA-2162123 | 1.15E-02 | 3.32E-01 | 5.61E+00 | 2.50E+01 | AKR1C3;PTGR1;PTGS1;PTGES |
| 40 | Transport of vitamins, nucleosides, and related molecules Homo sapiens R-HSA-425397 | 1.18E-02 | 3.32E-01 | 3.17E+00 | 1.41E+01 | SLC35B3;SLC35A3;SLCO3A1;SLCO4A1;SLC33A1;SLCO4C1;SLCO1A2 |
| 41 | Visual phototransduction Homo sapiens R-HSA-2187338 | 1.22E-02 | 3.32E-01 | 2.18E+00 | 9.58E+00 | AKR1C1;AKR1C3;LRP8;RDH11;RDH10;CYP4V2;NMT1;NAPEPLD;CNGA1;GPC3;LRP10;CALM1;AGRN |
| 42 | Sema4D mediated inhibition of cell attachment and migration Homo sapiens R-HSA-416550 | 1.29E-02 | 3.32E-01 | 8.41E+00 | 3.66E+01 | SEMA4D;RRAS;MET |
| 43 | Signaling by TGF-beta Receptor Complex in Cancer Homo sapiens R-HSA-3304351 | 1.29E-02 | 3.32E-01 | 8.41E+00 | 3.66E+01 | TGFB1;TGFBR1;TGFBR2 |

| | | | | | | |
|---|---|---|---|---|---|---|
| 44 | Amine compound SLC transporters Homo sapiens R-HSA-425428 | 1.30E-02 | 3.32E-01 | 3.51E+00 | 1.52E+01 | SLC44A3;SLC14A2;SLC44A4;SLC44A1;SLC6A13;SLC6A12 |
| 45 | Synthesis of bile acids and bile salts via 27-hydroxycholesterol Homo sapiens R-HSA-193807 | 1.49E-02 | 3.72E-01 | 5.10E+00 | 2.14E+01 | AKR1C1;NR1H4;AKR1C3;AKR1C2 |
| 46 | Elastic fibre formation Homo sapiens R-HSA-1566948 | 1.77E-02 | 4.24E-01 | 2.89E+00 | 1.17E+01 | ITGB1;TGFB1;ITGB5;LOXL4;FBLN1;ITGAV;LOXL2 |
| 47 | Glycerophospholipid biosynthesis Homo sapiens R-HSA-1483206 | 1.81E-02 | 4.24E-01 | 2.05E+00 | 8.23E+00 | DMGDH;SLC44A3;SLC44A4;SLC44A1;CPNE7;PLA2G4A;MBOAT2;AGPAT2;PISD;CHDH;LPCAT3;CDIPT;LPIN2 |
| 48 | Type I hemidesmosome assembly Homo sapiens R-HSA-446107 | 1.84E-02 | 4.24E-01 | 7.00E+00 | 2.80E+01 | CD151;DST;PLEC |
| 49 | TGF-beta receptor signaling in EMT (epithelial to mesenchymal transition) Homo sapiens R-HSA-2173791 | 1.88E-02 | 4.24E-01 | 4.67E+00 | 1.86E+01 | TGFB1;CGN;TGFBR1;TGFBR2 |
| 50 | Metabolism of lipids and lipoproteins Homo sapiens R-HSA-556833 | 1.89E-02 | 4.24E-01 | 1.37E+00 | 5.42E+00 | DMGDH;SLC44A3;SLC44A4;SLC44A1;CHD9;CPNE7;FHL2;AKR1B1;RORA;PIK3C2B;LIPC;SPTLC1;LIPH;ME1;ABCC3;ELOVL1;GPX2;PLA2G4A;PTGR1;PISD;CYP39A1;ACOX2;CHDH;PPARG;ANGPTL4;ABCG1;PTGES;INSIG2;PIK3R3;HSD17B11;AGPAT2;NPAS2;PTGS1;SGPL1;AGMO;TGS1;INPP5J;THEM4;HACL1;TBL1X;CDIPT;FDFT1;WWTR1;AKR1C1;SRD5A3;NR1H4;AKR1C3;MBOAT2;AKR1C2;NR1D1;SYNJ2;CYP4F11;GALC;NCEH1;LPCAT3;SLCO1A2;PNPLA6;LPIN2 |
| 51 | Synthesis of bile acids and bile salts via 7alpha-hydroxycholesterol Homo sapiens R-HSA-193368 | 1.93E-02 | 4.24E-01 | 3.69E+00 | 1.46E+01 | ACOX2;AKR1C1;NR1H4;AKR1C3;AKR1C2 |

| | | | | | | |
|---|---|---|---|---|---|---|
| 52 | Retinoid metabolism and transport Homo sapiens R-HSA-975634 | 2.01E-02 | 4.25E-01 | 2.81E+00 | 1.10E+01 | RDH11;AKR1C1;GPC3;AKR1C3;LRP10;AGRN;LRP8 |
| 53 | CD28 co-stimulation Homo sapiens R-HSA-389356 | 2.05E-02 | 4.25E-01 | 3.12E+00 | 1.21E+01 | PRR5;YES1;LCK;PIK3R3;THEM4;PAK2 |
| 54 | Smooth Muscle Contraction Homo sapiens R-HSA-445355 | 2.05E-02 | 4.25E-01 | 3.12E+00 | 1.21E+01 | ITGB5;ANXA2;TPM4;CALD1;TPM1;CALM1 |
| 55 | Costimulation by the CD28 family Homo sapiens R-HSA-388841 | 2.08E-02 | 4.25E-01 | 2.26E+00 | 8.77E+00 | PRR5;YES1;LCK;PIK3R3;THEM4;HLA-DRA;PAK2;ICOSLG;HLA-DPA1;HLA-DQB1 |
| 56 | Muscle contraction Homo sapiens R-HSA-397014 | 2.17E-02 | 4.31E-01 | 1.69E+00 | 6.47E+00 | KCNK5;WWTR1;ANXA2;TPM4;ITGB5;NPR1;TMOD3;TPM1;ATP2A3;ITPR2;ATP1A1;ATP1B1;HIPK2;DES;FXYD4;CALD1;AKAP9;VIM;CALM1;KCNJ2;KCNK3 |
| 57 | Phospholipid metabolism Homo sapiens R-HSA-1483257 | 2.19E-02 | 4.31E-01 | 1.77E+00 | 6.77E+00 | DMGDH;SLC44A3;SLC44A4;SLC44A1;CPNE7;PIK3R3;PLA2G4A;MBOAT2;SYNJ2;AGPAT2;PISD;PIK3C2B;CHDH;LPCAT3;INPP5J;PNPLA6;CDIPT;LPIN2 |
| 58 | Amino acid synthesis and interconversion (transamination) Homo sapiens R-HSA-70614 | 2.28E-02 | 4.37E-01 | 3.50E+00 | 1.33E+01 | FOLH1;GPT2;ALDH18A1;GLUL;ASPA |
| 59 | Metabolism of carbohydrates Homo sapiens R-HSA-71387 | 2.31E-02 | 4.37E-01 | 1.55E+00 | 5.85E+00 | SLC35B3;PFKFB3;GXYLT1;SLC2A1;AKR1B1;ENO1;HS6ST1;ENO3;SLC5A3;HK1;CHST11;CHST12;CHST15;GPC3;HS3ST3B1;CHST6;DCN;HS3ST1;NUP93;PC;MAN2B2;NUP50;B3GNT3;CALM1;FMOD;AGRN;FBP1;PFKP |
| 60 | Collagen biosynthesis and modifying enzymes Homo sapiens R-HSA-1650814 | 2.33E-02 | 4.37E-01 | 2.34E+00 | 8.79E+00 | COL16A1;P4HA1;COL14A1;P4HA2;COL4A4;COL11A1;COL12A1;COL4A3;PLOD2 |
| 61 | Zinc influx into cells by the SLC39 gene family Homo sapiens R-HSA-442380 | 2.50E-02 | 4.52E-01 | 6.00E+00 | 2.22E+01 | SLC39A10;SLC39A7;SLC39A1 |

| | | | | | | |
|---|---|---|---|---|---|---|
| 62 | Reactions specific to the complex N-glycan synthesis pathway Homo sapiens R-HSA-975578 | 2.50E-02 | 4.52E-01 | 6.00E+00 | 2.22E+01 | FUT8;MAN2A2;FUCA1 |
| 63 | Kinesins Homo sapiens R-HSA-983189 | 2.54E-02 | 4.53E-01 | 2.65E+00 | 9.75E+00 | KIFC2;KIF5B;KIF26B;KIF13B;KIF21A;KIF23;KIFAP3 |
| 64 | N-glycan antennae elongation in the medial/trans-Golgi Homo sapiens R-HSA-975576 | 2.67E-02 | 4.63E-01 | 3.34E+00 | 1.21E+01 | FUT8;ST6GAL1;MAN2A2;FUCA1;MGAT4B |
| 65 | Generation of second messenger molecules Homo sapiens R-HSA-202433 | 2.68E-02 | 4.63E-01 | 2.90E+00 | 1.05E+01 | VASP;LCK;HLA-DRA;PAK2;HLA-DPA1;HLA-DQB1 |
| 66 | ECM proteoglycans Homo sapiens R-HSA-3000178 | 2.86E-02 | 4.80E-01 | 2.39E+00 | 8.48E+00 | ITGB1;TGFB1;ITGB5;LAMB2;ITGAV;AGRN;FMOD;DCN |
| 67 | Ion transport by P-type ATPases Homo sapiens R-HSA-936837 | 2.86E-02 | 4.80E-01 | 2.39E+00 | 8.48E+00 | FXYD4;ATP2A3;ATP11C;ATP13A2;ATP1A1;ATP7A;ATP1B1;CALM1 |
| 68 | Vesicle-mediated transport Homo sapiens R-HSA-5653656 | 2.94E-02 | 4.86E-01 | 1.39E+00 | 4.89E+00 | NBAS;TMF1;SYS1;DCTN2;KDELR1;CTSZ;GCC2;MIA3;CLINT1;MAN2A2;VPS53;KIF5B;GJA4;KIF13B;MAN1C1;KIF21A;MAN1A1;GJA8;APOL1;KIFAP3;SPTAN1;SPTBN1;DYNC1H1;SORT1;GALNT2;TRAPPC10;M6PR;COPZ2;PLA2G4A;ANK2;KIF23;ANK3;TPD52L1;DYNC1LI2;GJB1;KIFC2;GJB3;GOLGB1;KIF26B;RAB13;EXOC4;RAB9A;DNASE2;CALM1 |
| 69 | O-linked glycosylation of mucins Homo sapiens R-HSA-913709 | 3.06E-02 | 4.91E-01 | 2.22E+00 | 7.73E+00 | ST6GAL1;MUC1;GALNT3;GALNT2;GALNT18;C1GALT1;MUC19;B3GNT3;GCNT3 |
| 70 | Glycosaminoglycan metabolism Homo sapiens R-HSA-1630316 | 3.10E-02 | 4.91E-01 | 1.84E+00 | 6.39E+00 | SLC35B3;HS3ST3B1;CHST6;GXYLT1;HS6ST1;DCN;HS3ST1;CHST11;CHST12;CHST15;B3GNT3;GPC3;AGRN;FMOD |
| 71 | Pyruvate metabolism Homo sapiens R-HSA-70268 | 3.10E-02 | 4.91E-01 | 3.19E+00 | 1.11E+01 | LDHA;PDK4;HAGH;SLC16A3;PDK1 |

| | | | | | | |
|---|---|---|---|---|---|---|
| 72 | Integrin cell surface interactions Homo sapiens R-HSA-216083 | 3.33E-02 | 5.00E-01 | 2.18E+00 | 7.41E+00 | ITGB1;COL16A1;ITGB5;COL4A4;ITGA10;COL4A3;PECAM1;ITGAV;AGRN |
| 73 | Semaphorin interactions Homo sapiens R-HSA-373755 | 3.33E-02 | 5.00E-01 | 2.18E+00 | 7.41E+00 | ITGB1;PLXND1;RRAS;SEMA4D;DPYSL2;MET;PAK2;MYH10;PLXNA4 |
| 74 | Gap junction assembly Homo sapiens R-HSA-190861 | 3.42E-02 | 5.00E-01 | 3.74E+00 | 1.26E+01 | GJB1;GJB3;GJA4;GJA8 |
| 75 | Other semaphorin interactions Homo sapiens R-HSA-416700 | 3.42E-02 | 5.00E-01 | 3.74E+00 | 1.26E+01 | ITGB1;PLXND1;SEMA4D;PLXNA4 |
| 76 | COPI-mediated anterograde transport Homo sapiens R-HSA-6807878 | 3.42E-02 | 5.00E-01 | 2.06E+00 | 6.97E+00 | DYNC1H1;DYNC1LI2;DCTN2;KDELR1;GOLGB1;COPZ2;ANK2;ANK3;SPTAN1;SPTBN1 |
| 77 | Apoptotic cleavage of cellular proteins Homo sapiens R-HSA-111465 | 3.43E-02 | 5.00E-01 | 2.71E+00 | 9.15E+00 | DSP;GSN;STK24;VIM;SPTAN1;PLEC |
| 78 | PPARA activates gene expression Homo sapiens R-HSA-1989781 | 3.82E-02 | 5.47E-01 | 1.83E+00 | 5.96E+00 | WWTR1;CHD9;FHL2;NR1H4;RORA;NR1D1;NPAS2;TGS1;ME1;PPARG;ANGPTL4;TBL1X;FDFT1 |
| 79 | N-glycan trimming and elongation in the cis-Golgi Homo sapiens R-HSA-964739 | 3.89E-02 | 5.47E-01 | 9.33E+00 | 3.03E+01 | MAN1C1;MAN1A1 |
| 80 | Axon guidance Homo sapiens R-HSA-422475 | 3.90E-02 | 5.47E-01 | 1.35E+00 | 4.38E+00 | ITGB1;KLB;PLXND1;ARPC1B;NTN4;PEBP1;SHB;FGF2;EGFR;PSMB10;RAP1B;RRAS;DPYSL2;ITGAV;SRGAP3;SPTAN1;PAK2;MYH10;MARK3;PLXNA4;SPTBN1;VASP;PRNP;EPHA6;YES1;SEMA4D;DUSP1;CSNK2A2;RANBP9;ANK2;ANK3;PSMB9;ARPC2;ITGA10;COL4A4;AKAP9;COL4A3;PSME1;AGRN;CALM1;CD24;RAF1;MET;EPHA2;FGFR1 |
| 81 | Chondroitin sulfate biosynthesis Homo sapiens R-HSA-2022870 | 4.06E-02 | 5.47E-01 | 3.50E+00 | 1.12E+01 | CHST11;CHST12;CHST15;DCN |

| | | | | | | |
|---|---|---|---|---|---|---|
| 82 | Synthesis of PC Homo sapiens R-HSA-1483191 | 4.06E-02 | 5.47E-01 | 3.50E+00 | 1.12E+01 | SLC44A3;SLC44A4;SLC44A1;LPIN2 |
| 83 | COPI-independent Golgi-to-ER retrograde traffic Homo sapiens R-HSA-6811436 | 4.09E-02 | 5.47E-01 | 2.92E+00 | 9.33E+00 | DYNC1H1;DYNC1LI2;DCTN2;GALNT2;PLA2G4A |
| 84 | YAP1- and WWTR1 (TAZ)-stimulated gene expression Homo sapiens R-HSA-2032785 | 4.09E-02 | 5.47E-01 | 2.92E+00 | 9.33E+00 | WWTR1;CHD9;TGS1;TBL1X;HIPK2 |
| 85 | Physiological factors Homo sapiens R-HSA-5578768 | 4.14E-02 | 5.48E-01 | 4.67E+00 | 1.49E+01 | WWTR1;NPR1;HIPK2 |
| 86 | COPI-dependent Golgi-to-ER retrograde traffic Homo sapiens R-HSA-6811434 | 4.28E-02 | 5.59E-01 | 1.98E+00 | 6.23E+00 | NBAS;KIFC2;KDELR1;KIF5B;KIF26B;KIF13B;COPZ2;KIF21A;KIF23;KIFAP3 |
| 87 | Regulation of lipid metabolism by Peroxisome proliferator-activated receptor alpha (PPARalpha) Homo sapiens R-HSA-400206 | 4.56E-02 | 5.89E-01 | 1.77E+00 | 5.47E+00 | WWTR1;CHD9;FHL2;NR1H4;RORA;NR1D1;NPAS2;TGS1;ME1;PPARG;ANGPTL4;TBL1X;FDFT1 |
| 88 | Molecules associated with elastic fibres Homo sapiens R-HSA-2129379 | 4.65E-02 | 5.93E-01 | 2.80E+00 | 8.60E+00 | ITGB1;TGFB1;ITGB5;FBLN1;ITGAV |
| 89 | Chondroitin sulfate/dermatan sulfate metabolism Homo sapiens R-HSA-1793185 | 4.70E-02 | 5.93E-01 | 2.28E+00 | 6.98E+00 | CHST11;CHST12;GXYLT1;GPC3;CHST15;AGRN;DCN |
| 90 | Translocation of ZAP-70 to Immunological synapse Homo sapiens R-HSA-202430 | 4.76E-02 | 5.93E-01 | 3.30E+00 | 1.00E+01 | LCK;HLA-DRA;HLA-DPA1;HLA-DQB1 |

| | | | | | | |
|---|---|---|---|---|---|---|
| 91 | Initiation of Nuclear Envelope Reformation Homo sapiens R-HSA-2995383 | 5.13E-02 | 6.15E-01 | 4.20E+00 | 1.25E+01 | ANKLE2;VRK2;LEMD2 |
| 92 | Nuclear Envelope Reassembly Homo sapiens R-HSA-2995410 | 5.13E-02 | 6.15E-01 | 4.20E+00 | 1.25E+01 | ANKLE2;VRK2;LEMD2 |
| 93 | IRF3-mediated induction of type I IFN Homo sapiens R-HSA-3270619 | 5.13E-02 | 6.15E-01 | 4.20E+00 | 1.25E+01 | IRF3;PRKDC;DTX4 |
| 94 | Metabolism of fat-soluble vitamins Homo sapiens R-HSA-6806667 | 5.15E-02 | 6.15E-01 | 2.23E+00 | 6.62E+00 | RDH11;AKR1C1;GPC3;AKR1C3;LRP10;AGRN;LRP8 |
| 95 | Ion channel transport Homo sapiens R-HSA-983712 | 5.23E-02 | 6.15E-01 | 1.54E+00 | 4.53E+00 | TTYH3;TSC22D3;ATP2A3;ATP11C;ATP1A1;ATP1B1;TPCN1;CLCN7;CLCN4;FXYD4;OSTM1;TRPV4;ATP6V0A2;ANO10;ATP6V0A4;ATP13A2;CALM1;RAF1;ASIC2;ATP7A |
| 96 | HS-GAG biosynthesis Homo sapiens R-HSA-2022928 | 5.25E-02 | 6.15E-01 | 2.70E+00 | 7.94E+00 | HS3ST3B1;GPC3;AGRN;HS6ST1;HS3ST1 |
| 97 | Condensation of Prophase Chromosomes Homo sapiens R-HSA-2299718 | 5.31E-02 | 6.15E-01 | 2.40E+00 | 7.06E+00 | RB1;H2AFZ;NCAPD3;SMC4;HIST1H2AC;MCPH1 |
| 98 | CD28 dependent PI3K/Akt signaling Homo sapiens R-HSA-389357 | 5.52E-02 | 6.20E-01 | 3.11E+00 | 9.02E+00 | PRR5;LCK;PIK3R3;THEM4 |
| 99 | Synthesis of Dolichyl-phosphate Homo sapiens R-HSA-446199 | 5.58E-02 | 6.20E-01 | 7.00E+00 | 2.02E+01 | SRD5A3;DOLK |
| 100 | IRF3 mediated activation of type 1 IFN Homo sapiens R-HSA-1606341 | 5.58E-02 | 6.20E-01 | 7.00E+00 | 2.02E+01 | IRF3;DTX4 |
| 101 | FGFR1b ligand binding and activation Homo sapiens R-HSA-190370 | 5.58E-02 | 6.20E-01 | 7.00E+00 | 2.02E+01 | FGF2;FGFR1 |

| | | | | | | |
|---|---|---|---|---|---|---|
| 102 | Circadian Clock Homo sapiens R-HSA-400253 | 5.74E-02 | 6.32E-01 | 2.04E+00 | 5.83E+00 | PER1;CHD9;TGS1;RORA;TBL1X;NR1D1;HIF1A;NPAS2 |
| 103 | Glycolysis Homo sapiens R-HSA-70171 | 5.90E-02 | 6.43E-01 | 2.60E+00 | 7.35E+00 | PFKFB3;ENO1;ENO3;PFKP;HK1 |
| 104 | Arachidonic acid metabolism Homo sapiens R-HSA-2142753 | 6.13E-02 | 6.61E-01 | 2.13E+00 | 5.96E+00 | GPX2;AKR1C3;PLA2G4A;PTGR1;CYP4F11;PTGES;PTGS1 |
| 105 | O-linked glycosylation Homo sapiens R-HSA-5173105 | 6.30E-02 | 6.65E-01 | 1.72E+00 | 4.75E+00 | SBSPON;ST6GAL1;MUC1;GALNT3;GALNT2;GALNT18;MUC19;C1GALT1;B3GNT3;GCNT3;THSD7A;ADAMTS6 |
| 106 | The canonical retinoid cycle in rods (twilight vision) Homo sapiens R-HSA-2453902 | 6.34E-02 | 6.65E-01 | 2.95E+00 | 8.14E+00 | RDH11;CYP4V2;RDH10;NAPEPLD |
| 107 | Laminin interactions Homo sapiens R-HSA-3000157 | 6.34E-02 | 6.65E-01 | 2.95E+00 | 8.14E+00 | ITGB1;LAMB2;ITGAV;NID2 |
| 108 | Asparagine N-linked glycosylation Homo sapiens R-HSA-446203 | 6.48E-02 | 6.72E-01 | 1.44E+00 | 3.93E+00 | DYNC1H1;ST6GAL1;DCTN2;KDELR1;FUCA1;TRAPPC10;CTSZ;SRD5A3;COPZ2;ANK2;MIA3;DOLK;ANK3;DYNC1LI2;FUT8;MAN2A2;GOLGB1;MGAT4B;SLC17A5;MAN1C1;MAN1A1;SPTAN1;SPTBN1;ST6GALNAC6 |
| 109 | Transport of inorganic cations/anions and amino acids/oligopeptides Homo sapiens R-HSA-425393 | 6.57E-02 | 6.72E-01 | 1.75E+00 | 4.78E+00 | SLC4A7;SLC15A1;SLC7A6;SLC9A6;SLC17A5;SLC6A12;CALM1;SLC4A4;SLC12A7;SLC38A2;SLC7A2 |
| 110 | Detoxification of Reactive Oxygen Species Homo sapiens R-HSA-3299685 | 6.59E-02 | 6.72E-01 | 2.50E+00 | 6.81E+00 | GPX2;PRDX1;CAT;GPX8;ATP7A |
| 111 | Heparan sulfate/heparin (HS-GAG) metabolism Homo sapiens R-HSA-1638091 | 6.65E-02 | 6.73E-01 | 2.09E+00 | 5.66E+00 | HS3ST3B1;GXYLT1;GPC3;AGRN;HS6ST1;DCN;HS3ST1 |

| | | | | | | |
|---|---|---|---|---|---|---|
| 112 | Diseases of glycosylation Homo sapiens R-HSA-3781865 | 6.81E-02 | 6.82E-01 | 1.80E+00 | 4.83E+00 | SBSPON;MUC1;C1GALT1;MUC19;GPC3;THSD7A;AGRN;FMOD;ADAMTS6;DCN |
| 113 | Stimuli-sensing channels Homo sapiens R-HSA-2672351 | 6.96E-02 | 6.82E-01 | 1.73E+00 | 4.62E+00 | CLCN7;TTYH3;CLCN4;OSTM1;TRPV4;TSC22D3;ANO10;RAF1;ASIC2;CALM1;TPCN1 |
| 114 | PLC beta mediated events Homo sapiens R-HSA-112043 | 7.06E-02 | 6.82E-01 | 2.21E+00 | 5.87E+00 | ADCY9;PDE1B;ITPR2;PLA2G4A;CALM1;PLCB2 |
| 115 | Cytokine Signaling in Immune system Homo sapiens R-HSA-1280215 | 7.13E-02 | 6.82E-01 | 1.26E+00 | 3.34E+00 | IFITM3;KLB;IFITM1;IFITM2;PEBP1;FGF2;TRIM5;SPTAN1;HLA-DPA1;IFNAR2;SYK;DUSP1;OSMR;TNFRSF1A;HLA-E;NUP93;OAS1;IRF3;LCK;OAS2;IRF1;NUP50;AKAP9;PSME1;RAF1;HLA-DQB1;IFNAR1;SP100;PIK3R3;TNFRSF11B;EGFR;PSMB10;RAP1B;GBP2;MARK3;SPTBN1;GBP4;IP6K2;MAP3K3;YES1;IL10RB;ISG15;PSMB9;BST2;TNIP2;HLA-DRA;TRIM38;CALM1;EIF4G3;IL18R1;FGFR1 |
| 116 | Phosphorylation of CD3 and TCR zeta chains Homo sapiens R-HSA-202427 | 7.22E-02 | 6.82E-01 | 2.80E+00 | 7.36E+00 | LCK;HLA-DRA;HLA-DPA1;HLA-DQB1 |
| 117 | Insulin processing Homo sapiens R-HSA-264876 | 7.22E-02 | 6.82E-01 | 2.80E+00 | 7.36E+00 | KIF5B;EXOC4;RAB27A;CPE |
| 118 | EPHA-mediated growth cone collapse Homo sapiens R-HSA-3928663 | 7.32E-02 | 6.82E-01 | 2.42E+00 | 6.32E+00 | YES1;EPHA6;SHB;MYH10;EPHA2 |
| 119 | EGFR interacts with phospholipase C-gamma Homo sapiens R-HSA-212718 | 7.32E-02 | 6.82E-01 | 2.42E+00 | 6.32E+00 | ADCY9;PDE1B;ITPR2;CALM1;EGFR |
| 120 | Striated Muscle Contraction Homo sapiens R-HSA-390522 | 7.32E-02 | 6.82E-01 | 2.42E+00 | 6.32E+00 | DES;TPM4;TMOD3;TPM1;VIM |

| | | | | | | |
|---|---|---|---|---|---|---|
| 121 | Recycling of bile acids and salts Homo sapiens R-HSA-159418 | 7.39E-02 | 6.82E-01 | 3.50E+00 | 9.12E+00 | ABCC3;SLCO1A2;NR1H4 |
| 122 | Clearance of Nuclear Envelope Membranes from Chromatin Homo sapiens R-HSA-2993913 | 7.47E-02 | 6.82E-01 | 5.60E+00 | 1.45E+01 | VRK2;LEMD2 |
| 123 | PTK6 promotes HIF1A stabilization Homo sapiens R-HSA-8857538 | 7.47E-02 | 6.82E-01 | 5.60E+00 | 1.45E+01 | HIF1A;EGFR |
| 124 | ABC-family proteins mediated transport Homo sapiens R-HSA-382556 | 7.71E-02 | 6.92E-01 | 2.16E+00 | 5.53E+00 | ABCC3;ABCC4;ABCA6;ABCA3;EIF2S2;ABCG1 |
| 125 | G-protein mediated events Homo sapiens R-HSA-112040 | 7.71E-02 | 6.92E-01 | 2.16E+00 | 5.53E+00 | ADCY9;PDE1B;ITPR2;PLA2G4A;CALM1;PLCB2 |
| 126 | Apoptosis Homo sapiens R-HSA-109581 | 7.83E-02 | 6.98E-01 | 1.53E+00 | 3.89E+00 | DSP;GSN;DAPK1;DAPK2;PSMB10;PSMB9;H1F0;STK24;TNFSF10;PSME1;NMT1;VIM;SPTAN1;PAK2;HIST1H1C;PLEC |
| 127 | Metal ion SLC transporters Homo sapiens R-HSA-425410 | 8.16E-02 | 7.00E-01 | 2.67E+00 | 6.69E+00 | SLC41A2;SLC39A10;SLC39A7;SLC39A1 |
| 128 | Basigin interactions Homo sapiens R-HSA-210991 | 8.16E-02 | 7.00E-01 | 2.67E+00 | 6.69E+00 | ITGB1;SLC7A6;ATP1B1;SLC16A3 |
| 129 | Activation of G protein gated Potassium channels Homo sapiens R-HSA-1296041 | 8.16E-02 | 7.00E-01 | 2.67E+00 | 6.69E+00 | GABBR1;KCNJ16;KCNJ2;KCNJ3 |
| 130 | Inhibition of voltage gated Ca2+ channels via Gbeta/gamma subunits Homo sapiens R-HSA-997272 | 8.16E-02 | 7.00E-01 | 2.67E+00 | 6.69E+00 | GABBR1;KCNJ16;KCNJ2;KCNJ3 |

| | | | | | | |
|---|---|---|---|---|---|---|
| 131 | G protein gated Potassium channels Homo sapiens R-HSA-1296059 | 8.16E-02 | 7.00E-01 | 2.67E+00 | 6.69E+00 | GABBR1;KCNJ16;KCNJ2;KCNJ3 |
| 132 | Phospholipase C-mediated cascade; FGFR4 Homo sapiens R-HSA-5654228 | 8.39E-02 | 7.13E-01 | 2.10E+00 | 5.21E+00 | KLB;ADCY9;PDE1B;ITPR2;CALM1;FGF2 |
| 133 | Synthesis of glycosylphosphatidylinositol (GPI) Homo sapiens R-HSA-162710 | 8.66E-02 | 7.13E-01 | 3.23E+00 | 7.90E+00 | PIGB;PIGF;PIGH |
| 134 | STING mediated induction of host immune responses Homo sapiens R-HSA-1834941 | 8.66E-02 | 7.13E-01 | 3.23E+00 | 7.90E+00 | IRF3;PRKDC;DTX4 |
| 135 | Cell-extracellular matrix interactions Homo sapiens R-HSA-446353 | 8.66E-02 | 7.13E-01 | 3.23E+00 | 7.90E+00 | ITGB1;VASP;ACTN1 |
| 136 | Rap1 signalling Homo sapiens R-HSA-392517 | 8.66E-02 | 7.13E-01 | 3.23E+00 | 7.90E+00 | RAP1B;RAF1;SIPA1 |
| 137 | Factors involved in megakaryocyte development and platelet production Homo sapiens R-HSA-983231 | 8.84E-02 | 7.13E-01 | 1.55E+00 | 3.75E+00 | KDM1A;HDAC1;DOCK8;KIF23;KIFC2;IRF3;KIF5B;MAFG;IRF1;KIF26B;KIF13B;KIF21A;MAFK;KIFAP3 |
| 138 | Programmed Cell Death Homo sapiens R-HSA-5357801 | 8.87E-02 | 7.13E-01 | 1.50E+00 | 3.63E+00 | DSP;GSN;DAPK1;DAPK2;PSMB10;PSMB9;H1F0;STK24;TNFSF10;PSME1;NMT1;VIM;SPTAN1;PAK2;HIST1H1C;PLEC |
| 139 | Phospholipase C-mediated cascade: FGFR1 Homo sapiens R-HSA-5654219 | 9.10E-02 | 7.13E-01 | 2.05E+00 | 4.92E+00 | ADCY9;PDE1B;ITPR2;CALM1;FGF2;FGFR1 |
| 140 | Diseases associated with glycosaminoglycan metabolism Homo sapiens R-HSA-3560782 | 9.16E-02 | 7.13E-01 | 2.55E+00 | 6.09E+00 | GPC3;FMOD;AGRN;DCN |

| | | | | | | |
|---|---|---|---|---|---|---|
| 141 | A tetrasaccharide linker sequence is required for GAG synthesis Homo sapiens R-HSA-1971475 | 9.16E-02 | 7.13E-01 | 2.55E+00 | 6.09E+00 | GXYLT1;GPC3;AGRN;DCN |
| 142 | PD-1 signaling Homo sapiens R-HSA-389948 | 9.16E-02 | 7.13E-01 | 2.55E+00 | 6.09E+00 | LCK;HLA-DRA;HLA-DPA1;HLA-DQB1 |
| 143 | Downregulation of TGF-beta receptor signaling Homo sapiens R-HSA-2173788 | 9.16E-02 | 7.13E-01 | 2.55E+00 | 6.09E+00 | USP15;TGFB1;TGFBR1;TGFBR2 |
| 144 | Cleavage of the damaged pyrimidine Homo sapiens R-HSA-110329 | 9.53E-02 | 7.13E-01 | 4.67E+00 | 1.10E+01 | MBD4;NEIL1 |
| 145 | Recognition and association of DNA glycosylase with site containing an affected pyrimidine Homo sapiens R-HSA-110328 | 9.53E-02 | 7.13E-01 | 4.67E+00 | 1.10E+01 | MBD4;NEIL1 |
| 146 | Depyrimidination Homo sapiens R-HSA-73928 | 9.53E-02 | 7.13E-01 | 4.67E+00 | 1.10E+01 | MBD4;NEIL1 |
| 147 | Nef and signal transduction Homo sapiens R-HSA-164944 | 9.53E-02 | 7.13E-01 | 4.67E+00 | 1.10E+01 | LCK;PAK2 |
| 148 | Cross-presentation of particulate exogenous antigens (phagosomes) Homo sapiens R-HSA-1236973 | 9.53E-02 | 7.13E-01 | 4.67E+00 | 1.10E+01 | ITGB5;ITGAV |
| 149 | Coenzyme A biosynthesis Homo sapiens R-HSA-196783 | 9.53E-02 | 7.13E-01 | 4.67E+00 | 1.10E+01 | PANK2;COASY |
| 150 | NR1D1 (REV-ERBA) represses gene expression Homo sapiens R-HSA-1368071 | 9.53E-02 | 7.13E-01 | 4.67E+00 | 1.10E+01 | NR1D1;NPAS2 |

| | | | | | | |
|---|---|---|---|---|---|---|
| 151 | ER to Golgi Anterograde Transport Homo sapiens R-HSA-199977 | 9.82E-02 | 7.21E-01 | 1.55E+00 | 3.59E+00 | DYNC1H1;DCTN2;KDELR1;TRAPPC10;CTSZ;COPZ2;ANK2;MIA3;ANK3;DYNC1LI2;GOLGB1;SPTAN1;SPTBN1 |
| 152 | Zinc transporters Homo sapiens R-HSA-435354 | 1.00E-01 | 7.21E-01 | 3.00E+00 | 6.90E+00 | SLC39A10;SLC39A7;SLC39A1 |
| 153 | ABC transporters in lipid homeostasis Homo sapiens R-HSA-1369062 | 1.00E-01 | 7.21E-01 | 3.00E+00 | 6.90E+00 | ABCA6;ABCA3;ABCG1 |
| 154 | Formation of Senescence-Associated Heterochromatin Foci (SAHF) Homo sapiens R-HSA-2559584 | 1.00E-01 | 7.21E-01 | 3.00E+00 | 6.90E+00 | RB1;H1F0;HIST1H1C |
| 155 | Platelet sensitization by LDL Homo sapiens R-HSA-432142 | 1.00E-01 | 7.21E-01 | 3.00E+00 | 6.90E+00 | PECAM1;PLA2G4A;LRP8 |
| 156 | Mitotic Prophase Homo sapiens R-HSA-68875 | 1.01E-01 | 7.21E-01 | 1.61E+00 | 3.69E+00 | RB1;NUP93;H2AFZ;NUP50;VRK2;NCAPD3;SMC4;LEMD2;LPIN2;HIST1H2AC;MCPH1 |
| 157 | VEGFR2 mediated vascular permeability Homo sapiens R-HSA-5218920 | 1.02E-01 | 7.21E-01 | 2.44E+00 | 5.56E+00 | PRR5;THEM4;CALM1;PAK2 |
| 158 | Sema4D in semaphorin signaling Homo sapiens R-HSA-400685 | 1.02E-01 | 7.21E-01 | 2.44E+00 | 5.56E+00 | RRAS;SEMA4D;MYH10;MET |
| 159 | CREB phosphorylation through the activation of Ras Homo sapiens R-HSA-442742 | 1.02E-01 | 7.21E-01 | 2.44E+00 | 5.56E+00 | RRAS;AKAP9;RAF1;CALM1 |
| 160 | Amino acid and oligopeptide SLC transporters Homo sapiens R-HSA-425374 | 1.06E-01 | 7.41E-01 | 1.96E+00 | 4.38E+00 | SLC15A1;SLC7A6;SLC17A5;SLC6A12;SLC38A2;SLC7A2 |

| | | | | | | |
|---|---|---|---|---|---|---|
| 161 | Retrograde transport at the Trans-Golgi-Network Homo sapiens R-HSA-6811440 | 1.06E-01 | 7.41E-01 | 1.96E+00 | 4.38E+00 | SYS1;TMF1;VPS53;M6PR;RAB9A;GCC2 |
| 162 | Constitutive Signaling by Aberrant PI3K in Cancer Homo sapiens R-HSA-2219530 | 1.11E-01 | 7.46E-01 | 1.82E+00 | 4.00E+00 | KLB;LCK;GAB1;PIK3R3;FGF2;EGFR;FGFR1 |
| 163 | Signaling by TGF-beta Receptor Complex Homo sapiens R-HSA-170834 | 1.13E-01 | 7.46E-01 | 1.73E+00 | 3.77E+00 | WWTR1;USP15;TGFB1;HDAC1;CGN;SKIL;TGFBR1;TGFBR2 |
| 164 | Gap junction trafficking Homo sapiens R-HSA-190828 | 1.13E-01 | 7.46E-01 | 2.33E+00 | 5.09E+00 | GJB1;GJB3;GJA4;GJA8 |
| 165 | Ca-dependent events Homo sapiens R-HSA-111996 | 1.13E-01 | 7.46E-01 | 2.33E+00 | 5.09E+00 | ADCY9;PDE1B;PLA2G4A;CALM1 |
| 166 | Activation of SMO Homo sapiens R-HSA-5635838 | 1.15E-01 | 7.46E-01 | 2.80E+00 | 6.07E+00 | EVC2;EVC;BOC |
| 167 | Tie2 Signaling Homo sapiens R-HSA-210993 | 1.15E-01 | 7.46E-01 | 2.80E+00 | 6.07E+00 | GRB7;ANGPT2;GRB14 |
| 168 | Regulation of signaling by CBL Homo sapiens R-HSA-912631 | 1.15E-01 | 7.46E-01 | 2.80E+00 | 6.07E+00 | YES1;SYK;PIK3R3 |
| 169 | cGMP effects Homo sapiens R-HSA-418457 | 1.15E-01 | 7.46E-01 | 2.80E+00 | 6.07E+00 | PDE1B;KCNMB4;PDE9A |
| 170 | GABA B receptor activation Homo sapiens R-HSA-977444 | 1.16E-01 | 7.46E-01 | 2.06E+00 | 4.44E+00 | GABBR1;ADCY9;KCNJ16;KCNJ2;KCNJ3 |
| 171 | Activation of GABAB receptors Homo sapiens R-HSA-991365 | 1.16E-01 | 7.46E-01 | 2.06E+00 | 4.44E+00 | GABBR1;ADCY9;KCNJ16;KCNJ2;KCNJ3 |
| 172 | Cardiac conduction Homo sapiens R-HSA-5576891 | 1.17E-01 | 7.46E-01 | 1.49E+00 | 3.21E+00 | KCNK5;WWTR1;NPR1;ATP2A3;ITPR2;ATP1A1;ATP1B1;HIPK2;FXYD4;AKAP9;CALM1;KCNJ2;KCNK3 |

| | | | | | | |
|---|---|---|---|---|---|---|
| 173 | Bicarbonate transporters Homo sapiens R-HSA-425381 | 1.17E-01 | 7.46E-01 | 4.00E+00 | 8.57E+00 | SLC4A7;SLC4A4 |
| 174 | Transport of nucleotide sugars Homo sapiens R-HSA-727802 | 1.17E-01 | 7.46E-01 | 4.00E+00 | 8.57E+00 | SLC35B3;SLC35A3 |
| 175 | Crosslinking of collagen fibrils Homo sapiens R-HSA-2243919 | 1.17E-01 | 7.46E-01 | 4.00E+00 | 8.57E+00 | LOXL4;LOXL2 |
| 176 | CHL1 interactions Homo sapiens R-HSA-447041 | 1.17E-01 | 7.46E-01 | 4.00E+00 | 8.57E+00 | ITGB1;ITGA10 |
| 177 | Post-translational protein modification Homo sapiens R-HSA-597592 | 1.18E-01 | 7.46E-01 | 1.23E+00 | 2.64E+00 | SP100;DCTN2;KDELR1;GALNT18;CTSZ;MIA3;FUT8;MUC1;HERC2;MAN2A2;SLC17A5;MAN1C1;GCNT3;MAN1A1;THSD7A;SPTAN1;ADAMTS6;SPTBN1;SBSPON;DYNC1H1;ST6GAL1;GALNT3;GALNT2;FUCA1;TRAPPC10;SRD5A3;C1GALT1;MUC19;COPZ2;ANK2;DOLK;ANK3;NUP93;PIGB;DYNC1LI2;GOLGB1;NUP50;MGAT4B;B3GNT3;PIGF;PIGH;ST6GALNAC6 |
| 178 | Cell junction organization Homo sapiens R-HSA-446728 | 1.19E-01 | 7.53E-01 | 1.64E+00 | 3.48E+00 | ITGB1;VASP;CLDN4;CD151;DST;CADM2;ACTN1;CLDN1;PLEC |
| 179 | Developmental Biology Homo sapiens R-HSA-1266738 | 1.22E-01 | 7.65E-01 | 1.18E+00 | 2.49E+00 | ITGB1;KLB;PLXND1;CHD9;ARPC1B;PEBP1;SHB;FGF2;DPYSL2;BOC;ITGAV;SPTAN1;HIST1H2AC;EPHA6;DUSP1;H2AFZ;CSNK2A2;ANK2;ANK3;COL4A4;AKAP9;COL4A3;PSME1;PPARG;HOXB1;ANGPTL4;RAF1;MET;EPHA2;CEBPD;KMT2C;ADAM22;NTN4;EGFR;PSMB10;RAP1B;RRAS;TGS1;SRGAP3;TBL1X;PAK2;MYH10;MARK3;PLXNA4;SPTBN1;VASP;PRNP;YES1;TGFB1;SEMA4D;LGI2;RANBP9;PSMB9;KLF5;ARPC2;ITGA10;AGRN;CALM1;CD24;FOXA3;FGFR1 |
| 180 | Ion homeostasis Homo sapiens R-HSA-5578775 | 1.23E-01 | 7.65E-01 | 1.87E+00 | 3.92E+00 | FXYD4;ATP2A3;ITPR2;ATP1A1;ATP1B1;CALM1 |
| 181 | Interaction between L1 and Ankyrins Homo sapiens R-HSA-445095 | 1.25E-01 | 7.74E-01 | 2.24E+00 | 4.66E+00 | ANK2;ANK3;SPTAN1;SPTBN1 |

| | | | | | | |
|---|---|---|---|---|---|---|
| 182 | Platelet homeostasis Homo sapiens R-HSA-418346 | 1.26E-01 | 7.75E-01 | 1.62E+00 | 3.35E+00 | PDE1B;ATP2A3;PECAM1;PLA2G4A;ITPR2;KCNMB4;CALM1;LRP8;PDE9A |
| 183 | Defective B3GALT6 causes EDSP2 and SEMDJL1 Homo sapiens R-HSA-4420332 | 1.30E-01 | 7.83E-01 | 2.63E+00 | 5.36E+00 | GPC3;AGRN;DCN |
| 184 | Defective B4GALT7 causes EDS, progeroid type Homo sapiens R-HSA-3560783 | 1.30E-01 | 7.83E-01 | 2.63E+00 | 5.36E+00 | GPC3;AGRN;DCN |
| 185 | Defective B3GAT3 causes JDSSDHD Homo sapiens R-HSA-3560801 | 1.30E-01 | 7.83E-01 | 2.63E+00 | 5.36E+00 | GPC3;AGRN;DCN |
| 186 | Phase 4 - resting membrane potential Homo sapiens R-HSA-5576886 | 1.30E-01 | 7.83E-01 | 2.63E+00 | 5.36E+00 | KCNK5;KCNK3;KCNJ2 |
| 187 | Gap junction trafficking and regulation Homo sapiens R-HSA-157858 | 1.37E-01 | 8.21E-01 | 2.15E+00 | 4.29E+00 | GJB1;GJB3;GJA4;GJA8 |
| 188 | Base-Excision Repair, AP Site Formation Homo sapiens R-HSA-73929 | 1.40E-01 | 8.30E-01 | 3.50E+00 | 6.87E+00 | MBD4;NEIL1 |
| 189 | Tetrahydrobiopterin (BH4) synthesis, recycling, salvage and regulation Homo sapiens R-HSA-1474151 | 1.40E-01 | 8.30E-01 | 3.50E+00 | 6.87E+00 | GCHFR;CALM1 |
| 190 | Ligand-independent caspase activation via DCC Homo sapiens R-HSA-418889 | 1.40E-01 | 8.30E-01 | 3.50E+00 | 6.87E+00 | DAPK1;DAPK2 |

| | | | | | | |
|---|---|---|---|---|---|---|
| 191 | Defective C1GALT1C1 causes Tn polyagglutination syndrome (TNPS) Homo sapiens R-HSA-5083632 | 1.45E-01 | 8.54E-01 | 2.47E+00 | 4.77E+00 | MUC1;C1GALT1;MUC19 |
| 192 | Amino acid transport across the plasma membrane Homo sapiens R-HSA-352230 | 1.49E-01 | 8.68E-01 | 2.07E+00 | 3.95E+00 | SLC7A6;SLC6A12;SLC38A2;SLC7A2 |
| 193 | Inwardly rectifying K+ channels Homo sapiens R-HSA-1296065 | 1.49E-01 | 8.68E-01 | 2.07E+00 | 3.95E+00 | GABBR1;KCNJ16;KCNJ2;KCNJ3 |
| 194 | Glucose metabolism Homo sapiens R-HSA-70326 | 1.56E-01 | 8.87E-01 | 1.58E+00 | 2.93E+00 | PFKFB3;PC;ENO1;CALM1;ENO3;FBP1;PFKP;HK1 |
| 195 | Transcriptional regulation of white adipocyte differentiation Homo sapiens R-HSA-381340 | 1.56E-01 | 8.87E-01 | 1.58E+00 | 2.93E+00 | TGFB1;KLF5;CEBPD;CHD9;TGS1;PPARG;ANGPTL4;TBL1X |
| 196 | RHO GTPases activate PAKs Homo sapiens R-HSA-5627123 | 1.62E-01 | 8.87E-01 | 2.33E+00 | 4.25E+00 | CALM1;PAK2;MYH10 |
| 197 | Gluconeogenesis Homo sapiens R-HSA-70263 | 1.62E-01 | 8.87E-01 | 2.00E+00 | 3.64E+00 | PC;ENO1;ENO3;FBP1 |
| 198 | DAG and IP3 signaling Homo sapiens R-HSA-1489509 | 1.62E-01 | 8.87E-01 | 2.00E+00 | 3.64E+00 | ADCY9;PDE1B;ITPR2;CALM1 |
| 199 | TGF-beta receptor signaling activates SMADs Homo sapiens R-HSA-2173789 | 1.62E-01 | 8.87E-01 | 2.00E+00 | 3.64E+00 | USP15;TGFB1;TGFBR1;TGFBR2 |
| 200 | Platelet degranulation Homo sapiens R-HSA-114608 | 1.63E-01 | 8.87E-01 | 1.48E+00 | 2.68E+00 | SRGN;ABCC4;TGFB1;ACTN1;PECAM1;TIMP3;CD9;FAM3C;CALM1;CLU |
| 201 | TCR signaling Homo sapiens R-HSA-202403 | 1.64E-01 | 8.87E-01 | 1.44E+00 | 2.60E+00 | VASP;NFKBIA;LCK;PSME1;HLA-DRA;PAK2;PSMB10;MALT1;HLA-DPA1;HLA-DQB1;PSMB9 |

| | | | | | | |
|---|---|---|---|---|---|---|
| 202 | FGFR1c ligand binding and activation Homo sapiens R-HSA-190373 | 1.64E-01 | 8.87E-01 | 3.11E+00 | 5.62E+00 | FGF2;FGFR1 |
| 203 | Signaling by activated point mutants of FGFR1 Homo sapiens R-HSA-1839122 | 1.64E-01 | 8.87E-01 | 3.11E+00 | 5.62E+00 | FGF2;FGFR1 |
| 204 | Condensation of Prometaphase Chromosomes Homo sapiens R-HSA-2514853 | 1.64E-01 | 8.87E-01 | 3.11E+00 | 5.62E+00 | CSNK2A2;SMC4 |
| 205 | RHO GTPases activate KTN1 Homo sapiens R-HSA-5625970 | 1.64E-01 | 8.87E-01 | 3.11E+00 | 5.62E+00 | KIF5B;KTN1 |
| 206 | RHO GTPases activate IQGAPs Homo sapiens R-HSA-5626467 | 1.64E-01 | 8.87E-01 | 3.11E+00 | 5.62E+00 | CLIP1;CALM1 |
| 207 | CLEC7A (Dectin-1) induces NFAT activation Homo sapiens R-HSA-5607763 | 1.64E-01 | 8.87E-01 | 3.11E+00 | 5.62E+00 | ITPR2;CALM1 |
| 208 | Synthesis of IP2, IP, and Ins in the cytosol Homo sapiens R-HSA-1855183 | 1.64E-01 | 8.87E-01 | 3.11E+00 | 5.62E+00 | ISYNA1;INPP5J |
| 209 | Phospholipase C-mediated cascade; FGFR3 Homo sapiens R-HSA-5654227 | 1.68E-01 | 8.98E-01 | 1.80E+00 | 3.20E+00 | ADCY9;PDE1B;ITPR2;CALM1;FGF2 |
| 210 | Inositol phosphate metabolism Homo sapiens R-HSA-1483249 | 1.68E-01 | 8.98E-01 | 1.80E+00 | 3.20E+00 | ISYNA1;INPP5J;CALM1;PLCB2;IP6K2 |
| 211 | Nephrin interactions Homo sapiens R-HSA-373753 | 1.78E-01 | 9.49E-01 | 2.21E+00 | 3.81E+00 | ACTN1;SPTAN1;SPTBN1 |
| 212 | Signaling by ERBB2 Homo sapiens R-HSA-1227986 | 1.79E-01 | 9.49E-01 | 1.75E+00 | 3.01E+00 | GRB7;DIAPH1;YES1;GAB1;EGFR |

| | | | | | | |
|---|---|---|---|---|---|---|
| 213 | Antigen processing-Cross presentation Homo sapiens R-HSA-1236975 | 1.80E-01 | 9.50E-01 | 1.51E+00 | 2.60E+00 | ITGB5;PSME1;TAP1;ITGAV;CTSS;PSMB10;HLA-E;PSMB9 |
| 214 | Passive transport by Aquaporins Homo sapiens R-HSA-432047 | 1.89E-01 | 9.52E-01 | 2.80E+00 | 4.67E+00 | AQP6;AQP3 |
| 215 | Defective EXT2 causes exostoses 2 Homo sapiens R-HSA-3656237 | 1.89E-01 | 9.52E-01 | 2.80E+00 | 4.67E+00 | GPC3;AGRN |
| 216 | Defective EXT1 causes exostoses 1, TRPS2 and CHDS Homo sapiens R-HSA-3656253 | 1.89E-01 | 9.52E-01 | 2.80E+00 | 4.67E+00 | GPC3;AGRN |
| 217 | Endosomal/Vacuolar pathway Homo sapiens R-HSA-1236977 | 1.89E-01 | 9.52E-01 | 2.80E+00 | 4.67E+00 | CTSS;HLA-E |
| 218 | CD28 dependent Vav1 pathway Homo sapiens R-HSA-389359 | 1.89E-01 | 9.52E-01 | 2.80E+00 | 4.67E+00 | LCK;PAK2 |
| 219 | Inhibition of replication initiation of damaged DNA by RB1/E2F1 Homo sapiens R-HSA-113501 | 1.89E-01 | 9.52E-01 | 2.80E+00 | 4.67E+00 | RB1;POLA2 |
| 220 | Reduction of cytosolic Ca++ levels Homo sapiens R-HSA-418359 | 1.89E-01 | 9.52E-01 | 2.80E+00 | 4.67E+00 | ATP2A3;CALM1 |
| 221 | Vitamin B5 (pantothenate) metabolism Homo sapiens R-HSA-199220 | 1.89E-01 | 9.52E-01 | 2.80E+00 | 4.67E+00 | PANK2;COASY |
| 222 | PLC-gamma1 signalling Homo sapiens R-HSA-167021 | 1.89E-01 | 9.52E-01 | 1.87E+00 | 3.11E+00 | ADCY9;PDE1B;ITPR2;CALM1 |

| | | | | | | |
|---|---|---|---|---|---|---|
| 223 | Synthesis of PIPs at the plasma membrane Homo sapiens R-HSA-1660499 | 1.89E-01 | 9.52E-01 | 1.87E+00 | 3.11E+00 | INPP5J;PIK3R3;SYNJ2;PIK3C2B |
| 224 | Acyl chain remodelling of PE Homo sapiens R-HSA-1482839 | 1.96E-01 | 9.78E-01 | 2.10E+00 | 3.42E+00 | LPCAT3;PLA2G4A;MBOAT2 |
| 225 | C-type lectin receptors (CLRs) Homo sapiens R-HSA-5621481 | 1.98E-01 | 9.78E-01 | 1.38E+00 | 2.23E+00 | NFKBIA;SYK;CARD9;PSME1;ITPR2;RAF1;CALM1;PAK2;PSMB10;MALT1;PSMB9 |
| 226 | Response to elevated platelet cytosolic Ca2+ Homo sapiens R-HSA-76005 | 1.99E-01 | 9.78E-01 | 1.40E+00 | 2.26E+00 | SRGN;ABCC4;TGFB1;ACTN1;PECAM1;TIMP3;CD9;FAM3C;CALM1;CLU |
| 227 | DNA Damage/Telomere Stress Induced Senescence Homo sapiens R-HSA-2559586 | 1.99E-01 | 9.78E-01 | 1.59E+00 | 2.56E+00 | RB1;RAD50;H1F0;H2AFZ;HIST1H2AC;HIST1H1C |
| 228 | Fatty acid, triacylglycerol, and ketone body metabolism Homo sapiens R-HSA-535734 | 2.01E-01 | 9.78E-01 | 1.27E+00 | 2.03E+00 | WWTR1;ELOVL1;CHD9;NR1H4;FHL2;RORA;NR1D1;AGPAT2;NPAS2;AGMO;TGS1;ME1;THEM4;PPARG;ANGPTL4;TBL1X;LPIN2;FDFT1 |
| 229 | Post NMDA receptor activation events Homo sapiens R-HSA-438064 | 2.03E-01 | 9.78E-01 | 1.81E+00 | 2.88E+00 | RRAS;AKAP9;RAF1;CALM1 |
| 230 | Downstream signaling events of B Cell Receptor (BCR) Homo sapiens R-HSA-1168372 | 2.13E-01 | 9.78E-01 | 1.27E+00 | 1.97E+00 | KLB;PRR5;GAB1;PIK3R3;FGF2;EGFR;PSMB10;MALT1;PSMB9;NFKBIA;AGO3;LCK;PSME1;REL;THEM4;FGFR1 |
| 231 | Endogenous sterols Homo sapiens R-HSA-211976 | 2.13E-01 | 9.78E-01 | 2.00E+00 | 3.09E+00 | CYP39A1;CYP4V2;NR1H4 |
| 232 | Signaling by the B Cell Receptor (BCR) Homo sapiens R-HSA-983705 | 2.14E-01 | 9.78E-01 | 1.24E+00 | 1.92E+00 | KLB;PRR5;SYK;GAB1;PIK3R3;ITPR2;FGF2;EGFR;PSMB10;MALT1;PSMB9;NFKBIA;AGO3;LCK;PSME1;REL;THEM4;CALM1;FGFR1 |

| | | | | | | |
|---|---|---|---|---|---|---|
| 233 | Facilitative Na+-independent glucose transporters Homo sapiens R-HSA-428790 | 2.14E-01 | 9.78E-01 | 2.54E+00 | 3.92E+00 | SLC2A1;SLC2A6 |
| 234 | Dissolution of Fibrin Clot Homo sapiens R-HSA-75205 | 2.14E-01 | 9.78E-01 | 2.54E+00 | 3.92E+00 | ANXA2;S100A10 |
| 235 | Activation of DNA fragmentation factor Homo sapiens R-HSA-211227 | 2.14E-01 | 9.78E-01 | 2.54E+00 | 3.92E+00 | H1F0;HIST1H1C |
| 236 | Apoptosis induced DNA fragmentation Homo sapiens R-HSA-140342 | 2.14E-01 | 9.78E-01 | 2.54E+00 | 3.92E+00 | H1F0;HIST1H1C |
| 237 | FGFR3c ligand binding and activation Homo sapiens R-HSA-190372 | 2.14E-01 | 9.78E-01 | 2.54E+00 | 3.92E+00 | GALNT3;FGF2 |
| 238 | FGFR3 ligand binding and activation Homo sapiens R-HSA-190239 | 2.14E-01 | 9.78E-01 | 2.54E+00 | 3.92E+00 | GALNT3;FGF2 |
| 239 | Trafficking and processing of endosomal TLR Homo sapiens R-HSA-1679131 | 2.14E-01 | 9.78E-01 | 2.54E+00 | 3.92E+00 | CNPY3;CTSS |
| 240 | Platelet Adhesion to exposed collagen Homo sapiens R-HSA-75892 | 2.14E-01 | 9.78E-01 | 2.54E+00 | 3.92E+00 | ITGB1;ITGA10 |
| 241 | Synthesis of PE Homo sapiens R-HSA-1483213 | 2.14E-01 | 9.78E-01 | 2.54E+00 | 3.92E+00 | LPIN2;PISD |
| 242 | Purine salvage Homo sapiens R-HSA-74217 | 2.14E-01 | 9.78E-01 | 2.54E+00 | 3.92E+00 | GMPR;AMPD2 |
| 243 | Hexose transport Homo sapiens R-HSA-189200 | 2.15E-01 | 9.78E-01 | 1.63E+00 | 2.50E+00 | NUP93;NUP50;SLC2A1;SLC5A3;HK1 |

| | | | | | | |
|---|---|---|---|---|---|---|
| 244 | Pyruvate metabolism and Citric Acid (TCA) cycle Homo sapiens R-HSA-71406 | 2.15E-01 | 9.78E-01 | 1.63E+00 | 2.50E+00 | LDHA;PDK4;HAGH;SLC16A3;PDK1 |
| 245 | PI3K/AKT Signaling in Cancer Homo sapiens R-HSA-2219528 | 2.15E-01 | 9.78E-01 | 1.44E+00 | 2.21E+00 | KLB;PRR5;LCK;GAB1;PIK3R3;FGF2;EGFR;FGFR1 |
| 246 | CLEC7A (Dectin-1) signaling Homo sapiens R-HSA-5607764 | 2.15E-01 | 9.78E-01 | 1.40E+00 | 2.15E+00 | NFKBIA;SYK;CARD9;PSME1;ITPR2;CALM1;PSMB10;MALT1;PSMB9 |
| 247 | Downstream TCR signaling Homo sapiens R-HSA-202424 | 2.15E-01 | 9.78E-01 | 1.40E+00 | 2.15E+00 | NFKBIA;LCK;PSME1;HLA-DRA;PSMB10;MALT1;HLA-DPA1;HLA-DQB1;PSMB9 |
| 248 | DAP12 signaling Homo sapiens R-HSA-2424491 | 2.17E-01 | 9.78E-01 | 1.19E+00 | 1.83E+00 | KLB;PRR5;PDE1B;PIK3R3;PEBP1;ITPR2;FGF2;EGFR;PSMB10;RAP1B;THEM4;SPTAN1;MARK3;SPTBN1;SYK;DUSP1;GAB1;HLA-E;PSMB9;ADCY9;AGO3;LCK;AKAP9;PSME1;CALM1;RAF1;FGFR1 |
| 249 | Ca2+ pathway Homo sapiens R-HSA-4086398 | 2.21E-01 | 9.78E-01 | 1.53E+00 | 2.31E+00 | WNT11;AGO3;WNT5A;ITPR2;CALM1;PLCB2 |
| 250 | Downstream signaling of activated FGFR1 Homo sapiens R-HSA-5654687 | 2.25E-01 | 9.78E-01 | 1.19E+00 | 1.78E+00 | KLB;PRR5;PDE1B;PIK3R3;PEBP1;ITPR2;FGF2;EGFR;PSMB10;RAP1B;FLRT3;THEM4;SPTAN1;MARK3;SPTBN1;DUSP1;GAB1;PSMB9;ADCY9;AGO3;LCK;AKAP9;PSME1;CALM1;RAF1;FGFR1 |
| 251 | Phospholipase C-mediated cascade; FGFR2 Homo sapiens R-HSA-5654221 | 2.27E-01 | 9.78E-01 | 1.59E+00 | 2.36E+00 | ADCY9;PDE1B;ITPR2;CALM1;FGF2 |
| 252 | Signaling by FGFR3 Homo sapiens R-HSA-5654741 | 2.30E-01 | 9.78E-01 | 1.19E+00 | 1.75E+00 | KLB;PRR5;PDE1B;PIK3R3;PEBP1;ITPR2;FGF2;EGFR;PSMB10;RAP1B;THEM4;SPTAN1;MARK3;SPTBN1;GALNT3;DUSP1;GAB1;PSMB9;ADCY9;AGO3;LCK;AKAP9;PSME1;CALM1;RAF1;FGFR1 |
| 253 | Post-translational modification: synthesis of GPI-anchored proteins Homo sapiens R-HSA-163125 | 2.31E-01 | 9.78E-01 | 1.91E+00 | 2.79E+00 | PIGB;PIGF;PIGH |

| | | | | | | |
|---|---|---|---|---|---|---|
| 254 | Antigen Presentation: Folding, assembly and peptide loading of class I MHC Homo sapiens R-HSA-983170 | 2.31E-01 | 9.78E-01 | 1.91E+00 | 2.79E+00 | ERAP2;TAP1;HLA-E |
| 255 | ATF4 activates genes Homo sapiens R-HSA-380994 | 2.31E-01 | 9.78E-01 | 1.91E+00 | 2.79E+00 | CCL2;DCP2;ATF4 |
| 256 | Nitric oxide stimulates guanylate cyclase Homo sapiens R-HSA-392154 | 2.31E-01 | 9.78E-01 | 1.91E+00 | 2.79E+00 | PDE1B;KCNMB4;PDE9A |
| 257 | Synthesis of IP3 and IP4 in the cytosol Homo sapiens R-HSA-1855204 | 2.31E-01 | 9.78E-01 | 1.91E+00 | 2.79E+00 | INPP5J;CALM1;PLCB2 |
| 258 | Acyl chain remodelling of PC Homo sapiens R-HSA-1482788 | 2.31E-01 | 9.78E-01 | 1.91E+00 | 2.79E+00 | LPCAT3;PLA2G4A;MBOAT2 |
| 259 | Diseases associated with O-glycosylation of proteins Homo sapiens R-HSA-3906995 | 2.32E-01 | 9.78E-01 | 1.50E+00 | 2.19E+00 | SBSPON;MUC1;C1GALT1;MUC19;THSD7A;ADAMTS6 |
| 260 | NCAM1 interactions Homo sapiens R-HSA-419037 | 2.32E-01 | 9.78E-01 | 1.70E+00 | 2.48E+00 | PRNP;COL4A4;COL4A3;AGRN |
| 261 | LGI-ADAM interactions Homo sapiens R-HSA-5682910 | 2.39E-01 | 9.78E-01 | 2.33E+00 | 3.34E+00 | LGI2;ADAM22 |
| 262 | Polymerase switching on the C-strand of the telomere Homo sapiens R-HSA-174411 | 2.39E-01 | 9.78E-01 | 2.33E+00 | 3.34E+00 | POLA2;RFC1 |
| 263 | Polymerase switching Homo sapiens R-HSA-69091 | 2.39E-01 | 9.78E-01 | 2.33E+00 | 3.34E+00 | POLA2;RFC1 |

| | | | | | | |
|---|---|---|---|---|---|---|
| 264 | Leading Strand Synthesis Homo sapiens R-HSA-69109 | 2.39E-01 | 9.78E-01 | 2.33E+00 | 3.34E+00 | POLA2;RFC1 |
| 265 | FGFR4 ligand binding and activation Homo sapiens R-HSA-190322 | 2.39E-01 | 9.78E-01 | 2.33E+00 | 3.34E+00 | KLB;FGF2 |
| 266 | SEMA3A-Plexin repulsion signaling by inhibiting Integrin adhesion Homo sapiens R-HSA-399955 | 2.39E-01 | 9.78E-01 | 2.33E+00 | 3.34E+00 | RRAS;PLXNA4 |
| 267 | Regulation of pyruvate dehydrogenase (PDH) complex Homo sapiens R-HSA-204174 | 2.39E-01 | 9.78E-01 | 2.33E+00 | 3.34E+00 | PDK4;PDK1 |
| 268 | Signaling by PDGF Homo sapiens R-HSA-186797 | 2.44E-01 | 9.78E-01 | 1.17E+00 | 1.65E+00 | KLB;PRR5;GRB7;PDE1B;PIK3R3;PEBP1;ITPR2;FGF2;EGFR;PSMB10;RAP1B;THEM4;SPTAN1;MARK3;SPTBN1;DUSP1;GAB1;PSMB9;ADCY9;AGO3;LCK;COL4A4;AKAP9;COL4A3;PSME1;CALM1;RAF1;FGFR1 |

| | | | | | | |
|---|---|---|---|---|---|---|
| 269 | Metabolism Homo sapiens R-HSA-1430728 | 2.44E-01 | 9.78E-01 | 1.07E+00 | 1.51E+00 | DMGDH;SLC35B3;PANK2;CPNE7;GXYLT1;RORA;ENO1;ENO3;HS6ST1;LIPC;LIPH;PDK4;SLC16A3;GLUL;PDK1;UGT2B11;HGD;GPT2;SLC6A12;MAT1A;PISD;CYP39A1;NUP93;POR;ADCY9;FOLH1;PSME1;ABCG1;PTGES;ISYNA1;PFKFB3;INSIG2;GMPR;PIK3R3;ITPR2;RPS4Y1;ADIPOR1;SLC5A3;NPAS2;LDHA;SGPL1;CHST11;RDH11;CHST12;AGMO;AFMID;CHST15;INPP5J;THEM4;HACL1;IP6K2;FDFT1;WWTR1;SRD5A3;SLC52A3;EPHX1;NR1D1;SYNJ2;CYP4F11;NCEH1;MAN2B2;SLCO1A2;ALDH18A1;CALM1;LRP10;SLC44A3;SLC44A4;SLC44A1;CHD9;FHL2;APIP;SLC2A1;AKR1B1;PIK3C2B;HK1;SPTLC1;CA5B;CYP4V2;ME1;MLX;TK1;HS3ST3B1;ABCC3;ELOVL1;ARG2;CHST6;GPX2;PLA2G4A;AMPD2;PTGR1;ADRA2C;COASY;DCN;NT5C3A;GCHFR;ACOX2;NUP50;CHDH;CAT;B3GNT3;PPARG;ANGPTL4;HAGH;FBP1;PLCB2;ASPA;HSD17B11;LRP8;AGPAT2;PSMB10;PTGS1;SRM;TGS1;UCP2;GPC3;TBL1X;CDIPT;AOC1;AKR1C1;AKR1C3;NR1H4;MBOAT2;AKR1C2;HS3ST1;PSMB9;GALC;SLC25A37;PC;KCNS3;LPCAT3;PNPLA6;AGRN;FMOD;LPIN2;PFKP |
| 270 | Signaling by FGFR1 Homo sapiens R-HSA-5654736 | 2.44E-01 | 9.78E-01 | 1.18E+00 | 1.66E+00 | KLB;PRR5;PDE1B;PIK3R3;PEBP1;ITPR2;FGF2;EGFR;PSMB10;RAP1B;FLRT3;THEM4;SPTAN1;MARK3;SPTBN1;DUSP1;GAB1;PSMB9;ADCY9;AGO3;LCK;AKAP9;PSME1;CALM1;RAF1;FGFR1 |
| 271 | MAP2K and MAPK activation Homo sapiens R-HSA-5674135 | 2.47E-01 | 9.78E-01 | 1.65E+00 | 2.31E+00 | RAP1B;PEBP1;RAF1;MARK3 |
| 272 | Termination of O-glycan biosynthesis Homo sapiens R-HSA-977068 | 2.50E-01 | 9.78E-01 | 1.83E+00 | 2.53E+00 | MUC1;ST6GAL1;MUC19 |
| 273 | ZBP1(DAI) mediated induction of type I IFNs Homo sapiens R-HSA-1606322 | 2.50E-01 | 9.78E-01 | 1.83E+00 | 2.53E+00 | NFKBIA;IRF3;DTX4 |
| 274 | WNT ligand biogenesis and trafficking Homo sapiens R-HSA-3238698 | 2.50E-01 | 9.78E-01 | 1.83E+00 | 2.53E+00 | WNT11;WNT5A;TMED5 |

| | | | | | | |
|---|---|---|---|---|---|---|
| 275 | CaM pathway Homo sapiens R-HSA-111997 | 2.50E-01 | 9.78E-01 | 1.83E+00 | 2.53E+00 | ADCY9;PDE1B;CALM1 |
| 276 | Calmodulin induced events Homo sapiens R-HSA-111933 | 2.50E-01 | 9.78E-01 | 1.83E+00 | 2.53E+00 | ADCY9;PDE1B;CALM1 |
| 277 | Negative regulation of the PI3K/AKT network Homo sapiens R-HSA-199418 | 2.52E-01 | 9.78E-01 | 1.37E+00 | 1.88E+00 | KLB;LCK;GAB1;PIK3R3;THEM4;FGF2;EGFR;FGFR1 |
| 278 | Nuclear Envelope Breakdown Homo sapiens R-HSA-2980766 | 2.52E-01 | 9.78E-01 | 1.52E+00 | 2.09E+00 | NUP93;NUP50;VRK2;LEMD2;LPIN2 |
| 279 | Signaling by Rho GTPases Homo sapiens R-HSA-194315 | 2.57E-01 | 9.78E-01 | 1.16E+00 | 1.57E+00 | ITGB1;KDM1A;ARPC1B;ARHGAP6;CENPC;OPHN1;KIF5B;MCF2L;ARHGDIB;RHPN2;SRGAP3;ECT2;PAK2;MYH10;HIST1H2AC;H2AFZ;ARHGEF16;ARAP1;BAIAP2;KTN1;FMNL3;DIAPH1;CLIP1;FMNL2;ARPC2;RHOJ;TRIP10;CALM1 |
| 280 | Cell-Cell communication Homo sapiens R-HSA-1500931 | 2.59E-01 | 9.78E-01 | 1.28E+00 | 1.74E+00 | VASP;ITGB1;CLDN4;CD151;DST;CADM2;ACTN1;CLDN1;SPTAN1;SPTBN1;PLEC |
| 281 | Signaling by SCF-KIT Homo sapiens R-HSA-1433557 | 2.59E-01 | 9.78E-01 | 1.17E+00 | 1.58E+00 | KLB;PRR5;GRB7;PIK3R3;PEBP1;FGF2;EGFR;PSMB10;RAP1B;THEM4;SPTAN1;MARK3;SPTBN1;YES1;DUSP1;GAB1;PSMB9;FER;AGO3;LCK;AKAP9;PSME1;CALM1;RAF1;FGFR1 |
| 282 | Collagen degradation Homo sapiens R-HSA-1442490 | 2.61E-01 | 9.78E-01 | 1.60E+00 | 2.15E+00 | COL16A1;MMP7;COL14A1;COL12A1 |
| 283 | Activation of NMDA receptor upon glutamate binding and postsynaptic events Homo sapiens R-HSA-442755 | 2.61E-01 | 9.78E-01 | 1.60E+00 | 2.15E+00 | RRAS;AKAP9;RAF1;CALM1 |
| 284 | Depolymerisation of the Nuclear Lamina Homo sapiens R-HSA-4419969 | 2.65E-01 | 9.78E-01 | 2.15E+00 | 2.86E+00 | LEMD2;LPIN2 |

| | | | | | | |
|---|---|---|---|---|---|---|
| 285 | N-Glycan antennae elongation Homo sapiens R-HSA-975577 | 2.65E-01 | 9.78E-01 | 2.15E+00 | 2.86E+00 | ST6GAL1;MGAT4B |
| 286 | Regulation of innate immune responses to cytosolic DNA Homo sapiens R-HSA-3134975 | 2.65E-01 | 9.78E-01 | 2.15E+00 | 2.86E+00 | IRF3;DTX4 |
| 287 | FGFR1 ligand binding and activation Homo sapiens R-HSA-190242 | 2.65E-01 | 9.78E-01 | 2.15E+00 | 2.86E+00 | FGF2;FGFR1 |
| 288 | ERBB2 Regulates Cell Motility Homo sapiens R-HSA-6785631 | 2.65E-01 | 9.78E-01 | 2.15E+00 | 2.86E+00 | DIAPH1;EGFR |
| 289 | Signaling by EGFRvIII in Cancer Homo sapiens R-HSA-5637812 | 2.65E-01 | 9.78E-01 | 2.15E+00 | 2.86E+00 | GAB1;EGFR |
| 290 | Constitutive Signaling by EGFRvIII Homo sapiens R-HSA-5637810 | 2.65E-01 | 9.78E-01 | 2.15E+00 | 2.86E+00 | GAB1;EGFR |
| 291 | CREB phosphorylation through the activation of CaMKII Homo sapiens R-HSA-442729 | 2.65E-01 | 9.78E-01 | 2.15E+00 | 2.86E+00 | AKAP9;CALM1 |
| 292 | Keratan sulfate biosynthesis Homo sapiens R-HSA-2022854 | 2.68E-01 | 9.78E-01 | 1.75E+00 | 2.30E+00 | CHST6;B3GNT3;FMOD |
| 293 | Caspase activation via extrinsic apoptotic signalig pathway Homo sapiens R-HSA-5357769 | 2.68E-01 | 9.78E-01 | 1.75E+00 | 2.30E+00 | DAPK1;DAPK2;TNFSF10 |
| 294 | Platelet calcium homeostasis Homo sapiens R-HSA-418360 | 2.68E-01 | 9.78E-01 | 1.75E+00 | 2.30E+00 | ATP2A3;ITPR2;CALM1 |

| | | | | | | |
|---|---|---|---|---|---|---|
| 295 | Downstream signal transduction Homo sapiens R-HSA-186763 | 2.68E-01 | 9.78E-01 | 1.16E+00 | 1.52E+00 | KLB;PRR5;GRB7;PDE1B;PIK3R3;PEBP1;ITPR2;FGF2;EGFR;PSMB10;RAP1B;THEM4;SPTAN1;MARK3;SPTBN1;DUSP1;GAB1;PSMB9;ADCY9;AGO3;LCK;AKAP9;PSME1;CALM1;RAF1;FGFR1 |
| 296 | Phase 1 - Functionalization of compounds Homo sapiens R-HSA-211945 | 2.71E-01 | 9.78E-01 | 1.33E+00 | 1.74E+00 | CYP39A1;POR;AOC1;CYP4V2;EPHX1;NR1H4;CYP4F11;PTGS1 |
| 297 | Degradation of the extracellular matrix Homo sapiens R-HSA-1474228 | 2.76E-01 | 9.78E-01 | 1.30E+00 | 1.67E+00 | CAPN8;COL16A1;MMP7;CAPN6;COL14A1;COL12A1;CAPN15;CTSS;DCN |
| 298 | Negative regulation of MAPK pathway Homo sapiens R-HSA-5675221 | 2.77E-01 | 9.78E-01 | 1.56E+00 | 2.00E+00 | DUSP1;PEBP1;RAF1;MARK3 |
| 299 | Transcriptional activation of mitochondrial biogenesis Homo sapiens R-HSA-2151201 | 2.77E-01 | 9.78E-01 | 1.56E+00 | 2.00E+00 | CHD9;TGS1;TBL1X;NR1D1 |
| 300 | Activation of NF-kappaB in B cells Homo sapiens R-HSA-1169091 | 2.77E-01 | 9.78E-01 | 1.40E+00 | 1.80E+00 | NFKBIA;PSME1;REL;PSMB10;MALT1;PSMB9 |
| 301 | Downstream signaling of activated FGFR2 Homo sapiens R-HSA-5654696 | 2.79E-01 | 9.78E-01 | 1.15E+00 | 1.47E+00 | KLB;PRR5;PDE1B;PIK3R3;PEBP1;ITPR2;FGF2;EGFR;PSMB10;RAP1B;THEM4;SPTAN1;MARK3;SPTBN1;DUSP1;GAB1;PSMB9;ADCY9;AGO3;LCK;AKAP9;PSME1;CALM1;RAF1;FGFR1 |
| 302 | Downstream signaling of activated FGFR4 Homo sapiens R-HSA-5654716 | 2.79E-01 | 9.78E-01 | 1.15E+00 | 1.47E+00 | KLB;PRR5;PDE1B;PIK3R3;PEBP1;ITPR2;FGF2;EGFR;PSMB10;RAP1B;THEM4;SPTAN1;MARK3;SPTBN1;DUSP1;GAB1;PSMB9;ADCY9;AGO3;LCK;AKAP9;PSME1;CALM1;RAF1;FGFR1 |
| 303 | Downstream signaling of activated FGFR3 Homo sapiens R-HSA-5654708 | 2.79E-01 | 9.78E-01 | 1.15E+00 | 1.47E+00 | KLB;PRR5;PDE1B;PIK3R3;PEBP1;ITPR2;FGF2;EGFR;PSMB10;RAP1B;THEM4;SPTAN1;MARK3;SPTBN1;DUSP1;GAB1;PSMB9;ADCY9;AGO3;LCK;AKAP9;PSME1;CALM1;RAF1;FGFR1 |
| 304 | Metabolism of vitamins and cofactors Homo sapiens R-HSA-196854 | 2.85E-01 | 9.78E-01 | 1.24E+00 | 1.55E+00 | PANK2;PC;RDH11;AKR1C1;SLC52A3;SLC2A1;GPC3;AKR1C3;LRP10;AGRN;LRP8;COASY |

| | | | | | | |
|---|---|---|---|---|---|---|
| 305 | PERK regulates gene expression Homo sapiens R-HSA-381042 | 2.87E-01 | 9.78E-01 | 1.68E+00 | 2.10E+00 | CCL2;DCP2;ATF4 |
| 306 | RNA Polymerase III Transcription Initiation From Type 3 Promoter Homo sapiens R-HSA-76071 | 2.87E-01 | 9.78E-01 | 1.68E+00 | 2.10E+00 | CRCP;SNAPC1;BDP1 |
| 307 | DAP12 interactions Homo sapiens R-HSA-2172127 | 2.87E-01 | 9.78E-01 | 1.14E+00 | 1.42E+00 | KLB;PRR5;PDE1B;PIK3R3;PEBP1;ITPR2;FGF2;EGFR;PSMB10;RAP1B;THEM4;SPTAN1;MARK3;SPTBN1;SYK;DUSP1;GAB1;HLA-E;PSMB9;ADCY9;AGO3;LCK;AKAP9;PSME1;CALM1;RAF1;FGFR1 |
| 308 | PI3K events in ERBB2 signaling Homo sapiens R-HSA-1963642 | 2.90E-01 | 9.78E-01 | 2.00E+00 | 2.47E+00 | GAB1;EGFR |
| 309 | Regulation of KIT signaling Homo sapiens R-HSA-1433559 | 2.90E-01 | 9.78E-01 | 2.00E+00 | 2.47E+00 | YES1;LCK |
| 310 | CRMPs in Sema3A signaling Homo sapiens R-HSA-399956 | 2.90E-01 | 9.78E-01 | 2.00E+00 | 2.47E+00 | DPYSL2;PLXNA4 |
| 311 | Sema3A PAK dependent Axon repulsion Homo sapiens R-HSA-399954 | 2.90E-01 | 9.78E-01 | 2.00E+00 | 2.47E+00 | PAK2;PLXNA4 |
| 312 | Host Interactions with Influenza Factors Homo sapiens R-HSA-168253 | 2.92E-01 | 9.78E-01 | 1.51E+00 | 1.86E+00 | NUP93;TGFB1;NUP50;ISG15 |
| 313 | Sperm:Oocyte Membrane Binding Homo sapiens R-HSA-1300652 | 2.92E-01 | 9.78E-01 | 3.50E+00 | 4.30E+00 | CD9 |
| 314 | Na+-dependent glucose transporters Homo sapiens R-HSA-428808 | 2.92E-01 | 9.78E-01 | 3.50E+00 | 4.30E+00 | SLC5A3 |

| | | | | | | |
|---|---|---|---|---|---|---|
| 315 | Extrinsic Pathway of Fibrin Clot Formation Homo sapiens R-HSA-140834 | 2.92E-01 | 9.78E-01 | 3.50E+00 | 4.30E+00 | TFPI |
| 316 | PTK6 Expression Homo sapiens R-HSA-8849473 | 2.92E-01 | 9.78E-01 | 3.50E+00 | 4.30E+00 | HIF1A |
| 317 | GRB7 events in ERBB2 signaling Homo sapiens R-HSA-1306955 | 2.92E-01 | 9.78E-01 | 3.50E+00 | 4.30E+00 | GRB7 |
| 318 | LRR FLII-interacting protein 1 (LRRFIP1) activates type I IFN production Homo sapiens R-HSA-3134973 | 2.92E-01 | 9.78E-01 | 3.50E+00 | 4.30E+00 | IRF3 |
| 319 | Cyclin B2 mediated events Homo sapiens R-HSA-157881 | 2.92E-01 | 9.78E-01 | 3.50E+00 | 4.30E+00 | CDC25B |
| 320 | Leukotriene receptors Homo sapiens R-HSA-391906 | 2.92E-01 | 9.78E-01 | 3.50E+00 | 4.30E+00 | CYSLTR2 |
| 321 | Vitamin B2 (riboflavin) metabolism Homo sapiens R-HSA-196843 | 2.92E-01 | 9.78E-01 | 3.50E+00 | 4.30E+00 | SLC52A3 |
| 322 | Estrogen biosynthesis Homo sapiens R-HSA-193144 | 2.92E-01 | 9.78E-01 | 3.50E+00 | 4.30E+00 | HSD17B11 |
| 323 | Alpha-oxidation of phytanate Homo sapiens R-HSA-389599 | 2.92E-01 | 9.78E-01 | 3.50E+00 | 4.30E+00 | HACL1 |
| 324 | The fatty acid cycling model Homo sapiens R-HSA-167826 | 2.92E-01 | 9.78E-01 | 3.50E+00 | 4.30E+00 | UCP2 |
| 325 | The proton buffering model Homo sapiens R-HSA-167827 | 2.92E-01 | 9.78E-01 | 3.50E+00 | 4.30E+00 | UCP2 |

| | | | | | | |
|---|---|---|---|---|---|---|
| 326 | Mitochondrial Uncoupling Proteins Homo sapiens R-HSA-166187 | 2.92E-01 | 9.78E-01 | 3.50E+00 | 4.30E+00 | UCP2 |
| 327 | Signaling by FGFR4 Homo sapiens R-HSA-5654743 | 2.94E-01 | 9.78E-01 | 1.14E+00 | 1.40E+00 | KLB;PRR5;PDE1B;PIK3R3;PEBP1;ITPR2;FGF2;EGFR;PSMB10;RAP1B;THEM4;SPTAN1;MARK3;SPTBN1;DUSP1;GAB1;PSMB9;ADCY9;AGO3;LCK;AKAP9;PSME1;CALM1;RAF1;FGFR1 |
| 328 | Peptide hormone metabolism Homo sapiens R-HSA-2980736 | 2.96E-01 | 9.78E-01 | 1.32E+00 | 1.61E+00 | GRP;KIF5B;CTSZ;EXOC4;RAB27A;CPE;SEC11C |
| 329 | PI-3K cascade:FGFR1 Homo sapiens R-HSA-5654689 | 2.97E-01 | 9.78E-01 | 1.25E+00 | 1.52E+00 | KLB;PRR5;AGO3;LCK;GAB1;PIK3R3;THEM4;FGF2;EGFR;FGFR1 |
| 330 | PI-3K cascade:FGFR3 Homo sapiens R-HSA-5654710 | 2.97E-01 | 9.78E-01 | 1.25E+00 | 1.52E+00 | KLB;PRR5;AGO3;LCK;GAB1;PIK3R3;THEM4;FGF2;EGFR;FGFR1 |
| 331 | PI3K events in ERBB4 signaling Homo sapiens R-HSA-1250342 | 2.97E-01 | 9.78E-01 | 1.25E+00 | 1.52E+00 | KLB;PRR5;AGO3;LCK;GAB1;PIK3R3;THEM4;FGF2;EGFR;FGFR1 |
| 332 | PIP3 activates AKT signaling Homo sapiens R-HSA-1257604 | 2.97E-01 | 9.78E-01 | 1.25E+00 | 1.52E+00 | KLB;PRR5;AGO3;LCK;GAB1;PIK3R3;THEM4;FGF2;EGFR;FGFR1 |
| 333 | PI-3K cascade:FGFR4 Homo sapiens R-HSA-5654720 | 2.97E-01 | 9.78E-01 | 1.25E+00 | 1.52E+00 | KLB;PRR5;AGO3;LCK;GAB1;PIK3R3;THEM4;FGF2;EGFR;FGFR1 |
| 334 | PI-3K cascade:FGFR2 Homo sapiens R-HSA-5654695 | 2.97E-01 | 9.78E-01 | 1.25E+00 | 1.52E+00 | KLB;PRR5;AGO3;LCK;GAB1;PIK3R3;THEM4;FGF2;EGFR;FGFR1 |
| 335 | Rho GTPase cycle Homo sapiens R-HSA-194840 | 2.97E-01 | 9.78E-01 | 1.25E+00 | 1.52E+00 | OPHN1;MCF2L;ARHGDIB;ARHGEF16;RHOJ;TRIP10;SRGAP3;ARAP1;ECT2;ARHGAP6 |
| 336 | EPH-Ephrin signaling Homo sapiens R-HSA-2682334 | 3.01E-01 | 9.78E-01 | 1.29E+00 | 1.55E+00 | EPHA6;YES1;ARPC2;ARPC1B;SHB;PAK2;MYH10;EPHA2 |
| 337 | Amyloid fiber formation Homo sapiens R-HSA-977225 | 3.01E-01 | 9.78E-01 | 1.35E+00 | 1.63E+00 | CALCA;GSN;H2AFZ;ODAM;HIST1H2AC;ITM2B |

| | | | | | | |
|---|---|---|---|---|---|---|
| 338 | GABA receptor activation Homo sapiens R-HSA-977443 | 3.05E-01 | 9.78E-01 | 1.40E+00 | 1.66E+00 | GABBR1;ADCY9;KCNJ16;KCNJ2;KCNJ3 |
| 339 | Regulation of cholesterol biosynthesis by SREBP (SREBF) Homo sapiens R-HSA-1655829 | 3.05E-01 | 9.78E-01 | 1.40E+00 | 1.66E+00 | CHD9;INSIG2;TGS1;TBL1X;FDFT1 |
| 340 | Surfactant metabolism Homo sapiens R-HSA-5683826 | 3.05E-01 | 9.78E-01 | 1.61E+00 | 1.92E+00 | ABCA3;CTSH;ADRA2C |
| 341 | Glucose transport Homo sapiens R-HSA-70153 | 3.07E-01 | 9.78E-01 | 1.47E+00 | 1.74E+00 | NUP93;NUP50;SLC2A1;HK1 |
| 342 | Activation of gene expression by SREBF (SREBP) Homo sapiens R-HSA-2426168 | 3.07E-01 | 9.78E-01 | 1.47E+00 | 1.74E+00 | CHD9;TGS1;TBL1X;FDFT1 |
| 343 | Ras activation uopn Ca2+ infux through NMDA receptor Homo sapiens R-HSA-442982 | 3.15E-01 | 9.78E-01 | 1.87E+00 | 2.15E+00 | AKAP9;CALM1 |
| 344 | Digestion of dietary lipid Homo sapiens R-HSA-192456 | 3.15E-01 | 9.78E-01 | 1.87E+00 | 2.15E+00 | LIPC;LIPH |
| 345 | Acyl chain remodelling of PS Homo sapiens R-HSA-1482801 | 3.15E-01 | 9.78E-01 | 1.87E+00 | 2.15E+00 | LPCAT3;PLA2G4A |
| 346 | PI5P, PP2A and IER3 Regulate PI3K/AKT Signaling Homo sapiens R-HSA-6811558 | 3.18E-01 | 9.78E-01 | 1.29E+00 | 1.48E+00 | KLB;LCK;GAB1;PIK3R3;FGF2;EGFR;FGFR1 |
| 347 | Signaling by FGFR Homo sapiens R-HSA-190236 | 3.22E-01 | 9.78E-01 | 1.12E+00 | 1.26E+00 | KLB;PRR5;PDE1B;PIK3R3;PEBP1;ITPR2;FGF2;EGFR;PSMB10;RAP1B;FLRT3;THEM4;SPTAN1;MARK3;SPTBN1;GALNT3;DUSP1;GAB1;PSMB9;ADCY9;AGO3;LCK;AKAP9;PSME1;CALM1;RAF1;FGFR1 |
| 348 | NCAM signaling for neurite out-growth Homo sapiens R-HSA-375165 | 3.23E-01 | 9.78E-01 | 1.14E+00 | 1.29E+00 | KLB;PRNP;DUSP1;PEBP1;FGF2;EGFR;PSMB10;PSMB9;RAP1B;COL4A4;AKAP9;COL4A3;PSME1;CALM1;AGRN;RAF1;SPTAN1;MARK3;SPTBN1;FGFR1 |

| | | | | | | |
|---|---|---|---|---|---|---|
| 349 | GAB1 signalosome Homo sapiens R-HSA-180292 | 3.24E-01 | 9.78E-01 | 1.22E+00 | 1.37E+00 | KLB;PRR5;AGO3;LCK;GAB1;PIK3R3;THEM4;FGF2;EGFR;FGFR1 |
| 350 | PI3K/AKT activation Homo sapiens R-HSA-198203 | 3.24E-01 | 9.78E-01 | 1.22E+00 | 1.37E+00 | KLB;PRR5;AGO3;LCK;GAB1;PIK3R3;THEM4;FGF2;EGFR;FGFR1 |
| 351 | RNA Polymerase I Promoter Opening Homo sapiens R-HSA-73728 | 3.24E-01 | 9.78E-01 | 1.55E+00 | 1.75E+00 | H2AFZ;MBD2;HIST1H2AC |
| 352 | Termination of translesion DNA synthesis Homo sapiens R-HSA-5656169 | 3.24E-01 | 9.78E-01 | 1.55E+00 | 1.75E+00 | RFC1;USP43;ISG15 |
| 353 | Metabolism of steroid hormones Homo sapiens R-HSA-196071 | 3.24E-01 | 9.78E-01 | 1.55E+00 | 1.75E+00 | SRD5A3;AKR1B1;HSD17B11 |
| 354 | Interleukin-2 signaling Homo sapiens R-HSA-451927 | 3.24E-01 | 9.78E-01 | 1.14E+00 | 1.29E+00 | KLB;SYK;DUSP1;PIK3R3;PEBP1;FGF2;EGFR;PSMB10;PSMB9;RAP1B;LCK;AKAP9;PSME1;CALM1;RAF1;SPTAN1;MARK3;SPTBN1;FGFR1 |
| 355 | trans-Golgi Network Vesicle Budding Homo sapiens R-HSA-199992 | 3.25E-01 | 9.78E-01 | 1.31E+00 | 1.48E+00 | SORT1;CTSZ;M6PR;DNASE2;TPD52L1;CLINT1 |
| 356 | Clathrin derived vesicle budding Homo sapiens R-HSA-421837 | 3.25E-01 | 9.78E-01 | 1.31E+00 | 1.48E+00 | SORT1;CTSZ;M6PR;DNASE2;TPD52L1;CLINT1 |
| 357 | Metabolism of polyamines Homo sapiens R-HSA-351202 | 3.28E-01 | 9.78E-01 | 1.27E+00 | 1.42E+00 | ARG2;APIP;PSME1;SLC6A12;PSMB10;SRM;PSMB9 |
| 358 | Platelet activation, signaling and aggregation Homo sapiens R-HSA-76002 | 3.31E-01 | 9.78E-01 | 1.14E+00 | 1.26E+00 | SRGN;ABCC4;TGFB1;SYK;ACTN1;F2R;PIK3R3;ITPR2;PLA2G4A;ADRA2C;CLU;RAP1B;LCK;PECAM1;TIMP3;CD9;FAM3C;CALM1;RAF1 |
| 359 | Meiotic synapsis Homo sapiens R-HSA-1221632 | 3.32E-01 | 9.78E-01 | 1.35E+00 | 1.49E+00 | SUN2;H2AFZ;HSPA2;SYNE2;HIST1H2AC |
| 360 | Fructose metabolism Homo sapiens R-HSA-5652084 | 3.39E-01 | 9.78E-01 | 2.80E+00 | 3.02E+00 | AKR1B1 |

| | | | | | | |
|---|---|---|---|---|---|---|
| 361 | Sensing of DNA Double Strand Breaks Homo sapiens R-HSA-5693548 | 3.39E-01 | 9.78E-01 | 2.80E+00 | 3.02E+00 | RAD50 |
| 362 | DNA replication initiation Homo sapiens R-HSA-68952 | 3.39E-01 | 9.78E-01 | 2.80E+00 | 3.02E+00 | POLA2 |
| 363 | Telomere C-strand synthesis initiation Homo sapiens R-HSA-174430 | 3.39E-01 | 9.78E-01 | 2.80E+00 | 3.02E+00 | POLA2 |
| 364 | Negative feedback regulation of MAPK pathway Homo sapiens R-HSA-5674499 | 3.39E-01 | 9.78E-01 | 2.80E+00 | 3.02E+00 | RAF1 |
| 365 | TNFR1-mediated ceramide production Homo sapiens R-HSA-5626978 | 3.39E-01 | 9.78E-01 | 2.80E+00 | 3.02E+00 | TNFRSF1A |
| 366 | Fibronectin matrix formation Homo sapiens R-HSA-1566977 | 3.39E-01 | 9.78E-01 | 2.80E+00 | 3.02E+00 | ITGB1 |
| 367 | Proton-coupled monocarboxylate transport Homo sapiens R-HSA-433692 | 3.39E-01 | 9.78E-01 | 2.80E+00 | 3.02E+00 | SLC16A3 |
| 368 | Activation of C3 and C5 Homo sapiens R-HSA-174577 | 3.39E-01 | 9.78E-01 | 2.80E+00 | 3.02E+00 | CFB |
| 369 | p75NTR negatively regulates cell cycle via SC1 Homo sapiens R-HSA-193670 | 3.39E-01 | 9.78E-01 | 2.80E+00 | 3.02E+00 | HDAC1 |
| 370 | CLEC7A/inflammasome pathway Homo sapiens R-HSA-5660668 | 3.39E-01 | 9.78E-01 | 2.80E+00 | 3.02E+00 | MALT1 |
| 371 | Acyl chain remodeling of CL Homo sapiens R-HSA-1482798 | 3.39E-01 | 9.78E-01 | 2.80E+00 | 3.02E+00 | PLA2G4A |

| | | | | | | |
|---|---|---|---|---|---|---|
| 372 | Methionine salvage pathway Homo sapiens R-HSA-1237112 | 3.39E-01 | 9.78E-01 | 2.80E+00 | 3.02E+00 | APIP |
| 373 | Transport and synthesis of PAPS Homo sapiens R-HSA-174362 | 3.39E-01 | 9.78E-01 | 2.80E+00 | 3.02E+00 | SLC35B3 |
| 374 | Uptake and function of diphtheria toxin Homo sapiens R-HSA-5336415 | 3.39E-01 | 9.78E-01 | 2.80E+00 | 3.02E+00 | CD9 |
| 375 | Synthesis of Lipoxins (LX) Homo sapiens R-HSA-2142700 | 3.39E-01 | 9.78E-01 | 2.80E+00 | 3.02E+00 | PTGR1 |
| 376 | Synthesis of 5-eicosatetraenoic acids Homo sapiens R-HSA-2142688 | 3.39E-01 | 9.78E-01 | 2.80E+00 | 3.02E+00 | GPX2 |
| 377 | Synthesis of 15-eicosatetraenoic acid derivatives Homo sapiens R-HSA-2142770 | 3.39E-01 | 9.78E-01 | 2.80E+00 | 3.02E+00 | GPX2 |
| 378 | Synthesis of 12-eicosatetraenoic acid derivatives Homo sapiens R-HSA-2142712 | 3.39E-01 | 9.78E-01 | 2.80E+00 | 3.02E+00 | GPX2 |
| 379 | Synthesis of wybutosine at G37 of tRNA(Phe) Homo sapiens R-HSA-6782861 | 3.39E-01 | 9.78E-01 | 2.80E+00 | 3.02E+00 | TYW1 |
| 380 | Defective GALNT12 causes colorectal cancer 1 (CRCS1) Homo sapiens R-HSA-5083636 | 3.40E-01 | 9.78E-01 | 1.75E+00 | 1.88E+00 | MUC1;MUC19 |
| 381 | Defective GALNT3 causes familial hyperphosphatemic tumoral calcinosis (HFTC) Homo sapiens R-HSA-5083625 | 3.40E-01 | 9.78E-01 | 1.75E+00 | 1.88E+00 | MUC1;MUC19 |

| | | | | | | |
|---|---|---|---|---|---|---|
| 382 | Oxygen-dependent proline hydroxylation of Hypoxia-inducible Factor Alpha Homo sapiens R-HSA-1234176 | 3.40E-01 | 9.78E-01 | 1.75E+00 | 1.88E+00 | EGLN3;HIF1A |
| 383 | Potassium Channels Homo sapiens R-HSA-1296071 | 3.41E-01 | 9.78E-01 | 1.23E+00 | 1.32E+00 | KCNG1;GABBR1;KCNS3;KCNMB4;KCNJ16;KCNJ2;KCNK3;KCNJ3 |
| 384 | NRAGE signals death through JNK Homo sapiens R-HSA-193648 | 3.54E-01 | 9.78E-01 | 1.37E+00 | 1.42E+00 | MCF2L;ARHGEF16;AATF;ECT2 |
| 385 | Keratan sulfate/keratin metabolism Homo sapiens R-HSA-1638074 | 3.61E-01 | 9.78E-01 | 1.45E+00 | 1.47E+00 | CHST6;B3GNT3;FMOD |
| 386 | DNA methylation Homo sapiens R-HSA-5334118 | 3.61E-01 | 9.78E-01 | 1.45E+00 | 1.47E+00 | DNMT1;H2AFZ;HIST1H2AC |
| 387 | Oncogene Induced Senescence Homo sapiens R-HSA-2559585 | 3.61E-01 | 9.78E-01 | 1.45E+00 | 1.47E+00 | CDKN2D;AGO3;ETS2 |
| 388 | Signaling by Robo receptor Homo sapiens R-HSA-376176 | 3.61E-01 | 9.78E-01 | 1.45E+00 | 1.47E+00 | VASP;SRGAP3;PAK2 |
| 389 | GABA synthesis, release, reuptake and degradation Homo sapiens R-HSA-888590 | 3.65E-01 | 9.78E-01 | 1.65E+00 | 1.66E+00 | SLC6A13;SLC6A12 |
| 390 | Na+/Cl- dependent neurotransmitter transporters Homo sapiens R-HSA-442660 | 3.65E-01 | 9.78E-01 | 1.65E+00 | 1.66E+00 | SLC6A13;SLC6A12 |
| 391 | Constitutive Signaling by Ligand-Responsive EGFR Cancer Variants Homo sapiens R-HSA-1236382 | 3.65E-01 | 9.78E-01 | 1.65E+00 | 1.66E+00 | GAB1;EGFR |

| | | | | | | |
|---|---|---|---|---|---|---|
| 392 | Signaling by Ligand-Responsive EGFR Variants in Cancer Homo sapiens R-HSA-5637815 | 3.65E-01 | 9.78E-01 | 1.65E+00 | 1.66E+00 | GAB1;EGFR |
| 393 | Signaling by EGFR in Cancer Homo sapiens R-HSA-1643713 | 3.65E-01 | 9.78E-01 | 1.65E+00 | 1.66E+00 | GAB1;EGFR |
| 394 | PI Metabolism Homo sapiens R-HSA-1483255 | 3.72E-01 | 9.78E-01 | 1.27E+00 | 1.26E+00 | INPP5J;PIK3R3;SYNJ2;PNPLA6;PIK3C2B |
| 395 | Sialic acid metabolism Homo sapiens R-HSA-4085001 | 3.80E-01 | 9.78E-01 | 1.40E+00 | 1.35E+00 | ST6GAL1;SLC17A5;ST6GALNAC6 |
| 396 | Inactivation, recovery and regulation of the phototransduction cascade Homo sapiens R-HSA-2514859 | 3.80E-01 | 9.78E-01 | 1.40E+00 | 1.35E+00 | CNGA1;NMT1;CALM1 |
| 397 | Signalling by NGF Homo sapiens R-HSA-166520 | 3.81E-01 | 9.78E-01 | 1.07E+00 | 1.04E+00 | KLB;PRR5;HDAC1;PDE1B;ITPR2;PIK3R3;PEBP1;AATF;FGF2;EGFR;PSMB10;RAP1B;MCF2L;THEM4;ECT2;SPTAN1;MARK3;SPTBN1;DUSP1;KIDINS220;ARHGEF16;GAB1;PSMB9;NFKBIA;ADCY9;AGO3;LCK;AKAP9;PSME1;CALM1;RAF1;FGFR1 |
| 398 | Interleukin-3, 5 and GM-CSF signaling Homo sapiens R-HSA-512988 | 3.81E-01 | 9.78E-01 | 1.10E+00 | 1.06E+00 | KLB;YES1;SYK;DUSP1;PIK3R3;PEBP1;FGF2;EGFR;PSMB10;PSMB9;RAP1B;AKAP9;PSME1;CALM1;RAF1;SPTAN1;MARK3;SPTBN1;FGFR1 |
| 399 | Cation-coupled Chloride cotransporters Homo sapiens R-HSA-426117 | 3.83E-01 | 9.78E-01 | 2.33E+00 | 2.23E+00 | SLC12A7 |
| 400 | Inositol transporters Homo sapiens R-HSA-429593 | 3.83E-01 | 9.78E-01 | 2.33E+00 | 2.23E+00 | SLC5A3 |

| | | | | | | |
|---|---|---|---|---|---|---|
| 401 | APEX1-Independent Resolution of AP Sites via the Single Nucleotide Replacement Pathway Homo sapiens R-HSA-5649702 | 3.83E-01 | 9.78E-01 | 2.33E+00 | 2.23E+00 | NEIL1 |
| 402 | Defective CHST3 causes SEDCJD Homo sapiens R-HSA-3595172 | 3.83E-01 | 9.78E-01 | 2.33E+00 | 2.23E+00 | DCN |
| 403 | Defective CHSY1 causes TPBS Homo sapiens R-HSA-3595177 | 3.83E-01 | 9.78E-01 | 2.33E+00 | 2.23E+00 | DCN |
| 404 | Defective CHST14 causes EDS, musculocontractural type Homo sapiens R-HSA-3595174 | 3.83E-01 | 9.78E-01 | 2.33E+00 | 2.23E+00 | DCN |
| 405 | Defective CHST6 causes MCDC1 Homo sapiens R-HSA-3656225 | 3.83E-01 | 9.78E-01 | 2.33E+00 | 2.23E+00 | FMOD |
| 406 | Defective ST3GAL3 causes MCT12 and EIEE15 Homo sapiens R-HSA-3656243 | 3.83E-01 | 9.78E-01 | 2.33E+00 | 2.23E+00 | FMOD |
| 407 | Defective B4GALT1 causes B4GALT1-CDG (CDG-2d) Homo sapiens R-HSA-3656244 | 3.83E-01 | 9.78E-01 | 2.33E+00 | 2.23E+00 | FMOD |
| 408 | GLI proteins bind promoters of Hh responsive genes to promote transcription Homo sapiens R-HSA-5635851 | 3.83E-01 | 9.78E-01 | 2.33E+00 | 2.23E+00 | BOC |
| 409 | IkBA variant leads to EDA-ID Homo sapiens R-HSA-5603029 | 3.83E-01 | 9.78E-01 | 2.33E+00 | 2.23E+00 | NFKBIA |

| | | | | | | |
|---|---|---|---|---|---|---|
| 410 | TRAIL signaling Homo sapiens R-HSA-75158 | 3.83E-01 | 9.78E-01 | 2.33E+00 | 2.23E+00 | TNFSF10 |
| 411 | Post-transcriptional silencing by small RNAs Homo sapiens R-HSA-426496 | 3.83E-01 | 9.78E-01 | 2.33E+00 | 2.23E+00 | AGO3 |
| 412 | Phase 3 - rapid repolarisation Homo sapiens R-HSA-5576890 | 3.83E-01 | 9.78E-01 | 2.33E+00 | 2.23E+00 | AKAP9 |
| 413 | Glycerophospholipid catabolism Homo sapiens R-HSA-6814848 | 3.83E-01 | 9.78E-01 | 2.33E+00 | 2.23E+00 | PNPLA6 |
| 414 | Defective HLCS causes multiple carboxylase deficiency Homo sapiens R-HSA-3371599 | 3.83E-01 | 9.78E-01 | 2.33E+00 | 2.23E+00 | PC |
| 415 | Defects in biotin (Btn) metabolism Homo sapiens R-HSA-3323169 | 3.83E-01 | 9.78E-01 | 2.33E+00 | 2.23E+00 | PC |
| 416 | VEGFA-VEGFR2 Pathway Homo sapiens R-HSA-4420097 | 3.87E-01 | 9.78E-01 | 1.08E+00 | 1.03E+00 | KLB;PRR5;DUSP1;PEBP1;ITPR2;SHB;BAIAP2;FGF2;EGFR;PSMB10;PSMB9;RAP1B;AKAP9;PSME1;THEM4;ITGAV;CALM1;RAF1;SPTAN1;PAK2;MARK3;SPTBN1;FGFR1 |
| 417 | Lagging Strand Synthesis Homo sapiens R-HSA-69186 | 3.90E-01 | 9.78E-01 | 1.55E+00 | 1.47E+00 | POLA2;RFC1 |
| 418 | Synthesis, secretion, and inactivation of Glucagon-like Peptide-1 (GLP-1) Homo sapiens R-HSA-381771 | 3.90E-01 | 9.78E-01 | 1.55E+00 | 1.47E+00 | GRP;SEC11C |
| 419 | eNOS activation and regulation Homo sapiens R-HSA-203765 | 3.90E-01 | 9.78E-01 | 1.55E+00 | 1.47E+00 | GCHFR;CALM1 |
| 420 | Metabolism of nitric oxide Homo sapiens R-HSA-202131 | 3.90E-01 | 9.78E-01 | 1.55E+00 | 1.47E+00 | GCHFR;CALM1 |

| | | | | | | |
|---|---|---|---|---|---|---|
| 421 | Adaptive Immune System Homo sapiens R-HSA-1280218 | 3.97E-01 | 9.78E-01 | 1.05E+00 | 9.67E-01 | ITGB1;KLB;PRR5;IFITM1;ITGB5;RNF14;FGF2;CTSS;UFL1;HERC2;KIF5B;CTSH;ITGAV;KIFAP3;ICOSLG;HLA-DPA1;SYK;GAB1;TAP1;KIF23;HLA-E;DYNC1LI2;LCK;PSME1;RAF1;HLA-DQB1;DCTN2;UBR4;ITPR2;PIK3R3;EGFR;MALT1;PSMB10;RAP1B;ZNRF1;HLA-DMA;RNF213;HLA-DMB;THEM4;PAK2;VASP;DYNC1H1;CD74;YES1;ERAP2;PSMB9;NFKBIA;AGO3;REL;HLA-DRA;CALM1;SIPA1;FGFR1 |
| 422 | Activated PKN1 stimulates transcription of AR (androgen receptor) regulated genes KLK2 and KLK3 Homo sapiens R-HSA-5625886 | 3.98E-01 | 9.78E-01 | 1.35E+00 | 1.25E+00 | KDM1A;H2AFZ;HIST1H2AC |
| 423 | The phototransduction cascade Homo sapiens R-HSA-2514856 | 3.98E-01 | 9.78E-01 | 1.35E+00 | 1.25E+00 | CNGA1;NMT1;CALM1 |
| 424 | Purine metabolism Homo sapiens R-HSA-73847 | 3.98E-01 | 9.78E-01 | 1.35E+00 | 1.25E+00 | CAT;GMPR;AMPD2 |
| 425 | Fc epsilon receptor (FCERI) signaling Homo sapiens R-HSA-2454202 | 3.98E-01 | 9.78E-01 | 1.07E+00 | 9.83E-01 | KLB;PRR5;ITPR2;PIK3R3;PEBP1;FGF2;EGFR;MALT1;PSMB10;RAP1B;THEM4;SPTAN1;PAK2;MARK3;SPTBN1;SYK;DUSP1;GAB1;PSMB9;NFKBIA;MAPK10;AGO3;LCK;AKAP9;PSME1;CALM1;RAF1;FGFR1 |
| 426 | Cytochrome P450 - arranged by substrate type Homo sapiens R-HSA-211897 | 3.99E-01 | 9.78E-01 | 1.23E+00 | 1.13E+00 | CYP39A1;POR;CYP4V2;NR1H4;CYP4F11 |
| 427 | Synthesis of substrates in N-glycan biosythesis Homo sapiens R-HSA-446219 | 4.13E-01 | 9.78E-01 | 1.21E+00 | 1.07E+00 | ST6GAL1;SRD5A3;SLC17A5;DOLK;ST6GALNAC6 |
| 428 | HS-GAG degradation Homo sapiens R-HSA-2024096 | 4.14E-01 | 9.78E-01 | 1.47E+00 | 1.30E+00 | GPC3;AGRN |

| | | | | | | |
|---|---|---|---|---|---|---|
| 429 | TP53 regulates transcription of additional cell cycle genes whose exact role in the p53 pathway remain uncertain Homo sapiens R-HSA-6804115 | 4.14E-01 | 9.78E-01 | 1.47E+00 | 1.30E+00 | RGCC;TNKS1BP1 |
| 430 | SHC-mediated cascade:FGFR1 Homo sapiens R-HSA-5654688 | 4.14E-01 | 9.78E-01 | 1.47E+00 | 1.30E+00 | FGF2;FGFR1 |
| 431 | CD209 (DC-SIGN) signaling Homo sapiens R-HSA-5621575 | 4.14E-01 | 9.78E-01 | 1.47E+00 | 1.30E+00 | RAF1;PAK2 |
| 432 | Regulation of Insulin-like Growth Factor (IGF) transport and uptake by Insulin-like Growth Factor Binding Proteins (IGFBPs) Homo sapiens R-HSA-381426 | 4.14E-01 | 9.78E-01 | 1.47E+00 | 1.30E+00 | PAPPA;IGFBP2 |
| 433 | VxPx cargo-targeting to cilium Homo sapiens R-HSA-5620916 | 4.14E-01 | 9.78E-01 | 1.47E+00 | 1.30E+00 | EXOC4;PKD2 |
| 434 | Synthesis of Leukotrienes (LT) and Eoxins (EX) Homo sapiens R-HSA-2142691 | 4.14E-01 | 9.78E-01 | 1.47E+00 | 1.30E+00 | CYP4F11;PTGR1 |
| 435 | Mitochondrial tRNA aminoacylation Homo sapiens R-HSA-379726 | 4.14E-01 | 9.78E-01 | 1.47E+00 | 1.30E+00 | IARS2;GARS |
| 436 | Regulation of activated PAK-2p34 by proteasome mediated degradation Homo sapiens R-HSA-211733 | 4.15E-01 | 9.78E-01 | 1.24E+00 | 1.09E+00 | PSME1;PAK2;PSMB10;PSMB9 |

| | | | | | | |
|---|---|---|---|---|---|---|
| 437 | Mitochondrial biogenesis Homo sapiens R-HSA-1592230 | 4.15E-01 | 9.78E-01 | 1.24E+00 | 1.09E+00 | CHD9;TGS1;TBL1X;NR1D1 |
| 438 | Lysosome Vesicle Biogenesis Homo sapiens R-HSA-432720 | 4.17E-01 | 9.78E-01 | 1.31E+00 | 1.15E+00 | CTSZ;M6PR;DNASE2 |
| 439 | Base Excision Repair Homo sapiens R-HSA-73884 | 4.17E-01 | 9.78E-01 | 1.31E+00 | 1.15E+00 | MBD4;RFC1;NEIL1 |
| 440 | Resolution of Abasic Sites (AP sites) Homo sapiens R-HSA-73933 | 4.17E-01 | 9.78E-01 | 1.31E+00 | 1.15E+00 | MBD4;RFC1;NEIL1 |
| 441 | Signaling by EGFR Homo sapiens R-HSA-177929 | 4.20E-01 | 9.78E-01 | 1.06E+00 | 9.19E-01 | KLB;PRR5;PDE1B;PIK3R3;PEBP1;ITPR2;FGF2;EGFR;PSMB10;RAP1B;THEM4;SPTAN1;MARK3;SPTBN1;DUSP1;GAB1;PSMB9;ADCY9;AGO3;LCK;AKAP9;PSME1;CALM1;RAF1;FGFR1 |
| 442 | Vitamin C (ascorbate) metabolism Homo sapiens R-HSA-196836 | 4.25E-01 | 9.78E-01 | 2.00E+00 | 1.71E+00 | SLC2A1 |
| 443 | Organic anion transporters Homo sapiens R-HSA-428643 | 4.25E-01 | 9.78E-01 | 2.00E+00 | 1.71E+00 | SLC17A5 |
| 444 | Ligand-receptor interactions Homo sapiens R-HSA-5632681 | 4.25E-01 | 9.78E-01 | 2.00E+00 | 1.71E+00 | BOC |
| 445 | WNT mediated activation of DVL Homo sapiens R-HSA-201688 | 4.25E-01 | 9.78E-01 | 2.00E+00 | 1.71E+00 | CSNK2A2 |
| 446 | AMPK inhibits chREBP transcriptional activation activity Homo sapiens R-HSA-163680 | 4.25E-01 | 9.78E-01 | 2.00E+00 | 1.71E+00 | ADIPOR1 |
| 447 | Recycling of eIF2:GDP Homo sapiens R-HSA-72731 | 4.25E-01 | 9.78E-01 | 2.00E+00 | 1.71E+00 | EIF2S2 |

| | | | | | | |
|---|---|---|---|---|---|---|
| 448 | IKBKG deficiency causes anhidrotic ectodermal dysplasia with immunodeficiency (EDA-ID) (via TLR) Homo sapiens R-HSA-5603027 | 4.25E-01 | 9.78E-01 | 2.00E+00 | 1.71E+00 | NFKBIA |
| 449 | Regulation of cytoskeletal remodeling and cell spreading by IPP complex components Homo sapiens R-HSA-446388 | 4.25E-01 | 9.78E-01 | 2.00E+00 | 1.71E+00 | ACTN1 |
| 450 | Terminal pathway of complement Homo sapiens R-HSA-166665 | 4.25E-01 | 9.78E-01 | 2.00E+00 | 1.71E+00 | CLU |
| 451 | Orexin and neuropeptides FF and QRFP bind to their respective receptors Homo sapiens R-HSA-389397 | 4.25E-01 | 9.78E-01 | 2.00E+00 | 1.71E+00 | NPFF |
| 452 | Free fatty acids regulate insulin secretion Homo sapiens R-HSA-400451 | 4.25E-01 | 9.78E-01 | 2.00E+00 | 1.71E+00 | PLCB2 |
| 453 | Fatty Acids bound to GPR40 (FFAR1) regulate insulin secretion Homo sapiens R-HSA-434316 | 4.25E-01 | 9.78E-01 | 2.00E+00 | 1.71E+00 | PLCB2 |
| 454 | Hydrolysis of LPC Homo sapiens R-HSA-1483115 | 4.25E-01 | 9.78E-01 | 2.00E+00 | 1.71E+00 | PLA2G4A |
| 455 | ChREBP activates metabolic gene expression Homo sapiens R-HSA-163765 | 4.25E-01 | 9.78E-01 | 2.00E+00 | 1.71E+00 | MLX |

| | | | | | | |
|---|---|---|---|---|---|---|
| 456 | Metabolism of ingested SeMet, Sec, MeSec into H2Se Homo sapiens R-HSA-2408508 | 4.25E-01 | 9.78E-01 | 2.00E+00 | 1.71E+00 | MAT1A |
| 457 | Linoleic acid (LA) metabolism Homo sapiens R-HSA-2046105 | 4.25E-01 | 9.78E-01 | 2.00E+00 | 1.71E+00 | ELOVL1 |
| 458 | M Phase Homo sapiens R-HSA-68886 | 4.27E-01 | 9.78E-01 | 1.07E+00 | 9.09E-01 | RB1;ANKLE2;H2AFZ;CSNK2A2;KIF23;VRK2;SMC4;LEMD2;PSMB10;CENPC;PSMB9;NUP93;CLIP1;NUP50;PSME1;NCAPD3;LPIN2;HIST1H2AC;MCPH1 |
| 459 | ARMS-mediated activation Homo sapiens R-HSA-170984 | 4.29E-01 | 9.78E-01 | 1.07E+00 | 9.07E-01 | KLB;DUSP1;KIDINS220;PEBP1;FGF2;EGFR;PSMB10;PSMB9;RAP1B;AKAP9;PSME1;CALM1;RAF1;SPTAN1;MARK3;SPTBN1;FGFR1 |
| 460 | Metabolism of non-coding RNA Homo sapiens R-HSA-194441 | 4.31E-01 | 9.78E-01 | 1.22E+00 | 1.03E+00 | NUP93;NUP50;TGS1;PHAX |
| 461 | snRNP Assembly Homo sapiens R-HSA-191859 | 4.31E-01 | 9.78E-01 | 1.22E+00 | 1.03E+00 | NUP93;NUP50;TGS1;PHAX |
| 462 | Regulation of Apoptosis Homo sapiens R-HSA-169911 | 4.31E-01 | 9.78E-01 | 1.22E+00 | 1.03E+00 | PSME1;PAK2;PSMB10;PSMB9 |
| 463 | PI3K Cascade Homo sapiens R-HSA-109704 | 4.34E-01 | 9.78E-01 | 1.15E+00 | 9.61E-01 | KLB;CAB39L;GAB1;THEM4;FGF2;FGFR1 |
| 464 | Signaling by VEGF Homo sapiens R-HSA-194138 | 4.34E-01 | 9.78E-01 | 1.06E+00 | 8.80E-01 | KLB;PRR5;DUSP1;PEBP1;ITPR2;SHB;BAIAP2;FGF2;EGFR;PSMB10;PSMB9;RAP1B;AKAP9;PSME1;THEM4;ITGAV;CALM1;RAF1;SPTAN1;PAK2;MARK3;SPTBN1;FGFR1 |
| 465 | RNA Polymerase III Transcription Initiation Homo sapiens R-HSA-76046 | 4.35E-01 | 9.78E-01 | 1.27E+00 | 1.06E+00 | CRCP;SNAPC1;BDP1 |
| 466 | RHO GTPases Activate WASPs and WAVEs Homo sapiens R-HSA-5663213 | 4.35E-01 | 9.78E-01 | 1.27E+00 | 1.06E+00 | ARPC2;ARPC1B;BAIAP2 |

| | | | | | | |
|---|---|---|---|---|---|---|
| 467 | Telomere C-strand (Lagging Strand) Synthesis Homo sapiens R-HSA-174417 | 4.37E-01 | 9.78E-01 | 1.40E+00 | 1.16E+00 | POLA2;RFC1 |
| 468 | CTLA4 inhibitory signaling Homo sapiens R-HSA-389513 | 4.37E-01 | 9.78E-01 | 1.40E+00 | 1.16E+00 | YES1;LCK |
| 469 | Signaling by FGFR3 point mutants in cancer Homo sapiens R-HSA-8853338 | 4.37E-01 | 9.78E-01 | 1.40E+00 | 1.16E+00 | GAB1;FGF2 |
| 470 | Signaling by FGFR3 in disease Homo sapiens R-HSA-5655332 | 4.37E-01 | 9.78E-01 | 1.40E+00 | 1.16E+00 | GAB1;FGF2 |
| 471 | SALM protein interactions at the synapse Homo sapiens R-HSA-8849932 | 4.37E-01 | 9.78E-01 | 1.40E+00 | 1.16E+00 | RTN3;LRFN4 |
| 472 | RHO GTPase Effectors Homo sapiens R-HSA-195258 | 4.38E-01 | 9.78E-01 | 1.06E+00 | 8.77E-01 | ITGB1;KDM1A;H2AFZ;ARPC1B;BAIAP2;CENPC;KTN1;FMNL3;DIAPH1;CLIP1;FMNL2;ARPC2;KIF5B;RHPN2;CALM1;PAK2;MYH10;HIST1H2AC |
| 473 | Transmission across Chemical Synapses Homo sapiens R-HSA-112315 | 4.39E-01 | 9.78E-01 | 1.07E+00 | 8.82E-01 | GABBR1;SLC6A13;SLC6A12;KCNJ16;ADCY9;RRAS;AKAP9;ARL6IP5;CALM1;RAF1;GLUL;PLCB2;SLC38A2;KCNJ2;KCNJ3 |
| 474 | ER-Phagosome pathway Homo sapiens R-HSA-1236974 | 4.39E-01 | 9.78E-01 | 1.17E+00 | 9.59E-01 | PSME1;TAP1;PSMB10;HLA-E;PSMB9 |
| 475 | NGF signalling via TRKA from the plasma membrane Homo sapiens R-HSA-187037 | 4.44E-01 | 9.78E-01 | 1.05E+00 | 8.49E-01 | KLB;PRR5;PDE1B;PIK3R3;PEBP1;ITPR2;FGF2;EGFR;PSMB10;RAP1B;THEM4;SPTAN1;MARK3;SPTBN1;DUSP1;KIDINS220;GAB1;PSMB9;ADCY9;AGO3;LCK;AKAP9;PSME1;CALM1;RAF1;FGFR1 |
| 476 | Neurotransmitter Release Cycle Homo sapiens R-HSA-112310 | 4.46E-01 | 9.78E-01 | 1.19E+00 | 9.62E-01 | ARL6IP5;SLC6A13;SLC6A12;SLC38A2 |
| 477 | Neuronal System Homo sapiens R-HSA-112316 | 4.49E-01 | 9.78E-01 | 1.05E+00 | 8.40E-01 | KCNG1;RTN3;GABBR1;SLC6A13;SLC6A12;KCNJ16;ADCY9;LRFN4;RRAS;KCNS3;AKAP9;ARL6IP5;KCNMB4;CALM1;RAF1;GLUL;PLCB2;SLC38A2;KCNJ2;KCNK3;KCNJ3 |

| | | | | | | |
|---|---|---|---|---|---|---|
| 478 | Prolonged ERK activation events Homo sapiens R-HSA-169893 | 4.50E-01 | 9.78E-01 | 1.06E+00 | 8.45E-01 | KLB;DUSP1;KIDINS220;PEBP1;FGF2;EGFR;PSMB10;PSMB9;RAP1B;AKAP9;PSME1;CALM1;RAF1;SPTAN1;MARK3;SPTBN1;FGFR1 |
| 479 | Cytosolic sensors of pathogen-associated DNA Homo sapiens R-HSA-1834949 | 4.53E-01 | 9.78E-01 | 1.15E+00 | 9.09E-01 | NFKBIA;CRCP;IRF3;PRKDC;DTX4 |
| 480 | Triglyceride Biosynthesis Homo sapiens R-HSA-75109 | 4.53E-01 | 9.78E-01 | 1.15E+00 | 9.09E-01 | ELOVL1;AGMO;THEM4;AGPAT2;LPIN2 |
| 481 | Defective B3GALTL causes Peters-plus syndrome (PpS) Homo sapiens R-HSA-5083635 | 4.53E-01 | 9.78E-01 | 1.23E+00 | 9.78E-01 | SBSPON;THSD7A;ADAMTS6 |
| 482 | Translesion synthesis by Y family DNA polymerases bypasses lesions on DNA template Homo sapiens R-HSA-110313 | 4.53E-01 | 9.78E-01 | 1.23E+00 | 9.78E-01 | RFC1;USP43;ISG15 |
| 483 | Platelet Aggregation (Plug Formation) Homo sapiens R-HSA-76009 | 4.53E-01 | 9.78E-01 | 1.23E+00 | 9.78E-01 | RAP1B;SYK;ADRA2C |
| 484 | Steroid hormones Homo sapiens R-HSA-209943 | 4.53E-01 | 9.78E-01 | 1.23E+00 | 9.78E-01 | SRD5A3;AKR1B1;HSD17B11 |
| 485 | Metabolism of proteins Homo sapiens R-HSA-392499 | 4.53E-01 | 9.78E-01 | 1.02E+00 | 8.08E-01 | KDELR1;GALNT18;CTSZ;MIA3;TUBA1C;HERC2;KIF5B;CTSH;MAN1A1;THSD7A;SEC62;SPTAN1;ADAMTS6;HIST1H2AC;SBSPON;ST6GAL1;H2AFZ;CSNK2A2;IGFBP2;COPZ2;ANK2;ANK3;ADRA2C;NUP93;DNAJC3;PIGB;DYNC1LI2;NUP50;PAPPA;EXOC4;B3GNT3;ETF1;PIGF;DCP2;ATF4;PIGH;ST6GALNAC6;CALCA;SP100;DCTN2;GRP;COX17;RPS4Y1;FUT8;MUC1;MAN2A2;MAN1C1;SLC17A5;CCL2;GCNT3;CCT5;SPTBN1;SEC11C;DYNC1H1;GSN;GALNT3;ODAM;GALNT2;WFS1;ABCA3;FUCA1;TRAPPC10;SRD5A3;MUC19;C1GALT1;RAB27A;DOLK;EIF2S2;GOLGB1;MGAT4B;CPE;EIF3C;ITM2B |

| | | | | | | |
|---|---|---|---|---|---|---|
| 486 | Integration of energy metabolism Homo sapiens R-HSA-163685 | 4.54E-01 | 9.78E-01 | 1.10E+00 | 8.66E-01 | ADCY9;KCNS3;SLC2A1;MLX;ITPR2;ADIPOR1;ADRA2C;PLCB2 |
| 487 | Signaling by FGFR2 Homo sapiens R-HSA-5654738 | 4.54E-01 | 9.78E-01 | 1.04E+00 | 8.22E-01 | KLB;PRR5;PDE1B;PIK3R3;PEBP1;ITPR2;FGF2;EGFR;PSMB10;RAP1B;THEM4;SPTAN1;MARK3;SPTBN1;DUSP1;GAB1;PSMB9;ADCY9;AGO3;LCK;AKAP9;PSME1;CALM1;RAF1;FGFR1 |
| 488 | Opioid Signalling Homo sapiens R-HSA-111885 | 4.58E-01 | 9.78E-01 | 1.12E+00 | 8.75E-01 | ADCY9;PDE1B;PLA2G4A;ITPR2;CALM1;PLCB2 |
| 489 | Regulation of TP53 Activity through Acetylation Homo sapiens R-HSA-6804758 | 4.60E-01 | 9.78E-01 | 1.33E+00 | 1.03E+00 | HDAC1;BRD7 |
| 490 | Downregulation of SMAD2/3:SMAD4 transcriptional activity Homo sapiens R-HSA-2173795 | 4.60E-01 | 9.78E-01 | 1.33E+00 | 1.03E+00 | HDAC1;SKIL |
| 491 | SHC-mediated cascade:FGFR4 Homo sapiens R-HSA-5654719 | 4.60E-01 | 9.78E-01 | 1.33E+00 | 1.03E+00 | KLB;FGF2 |
| 492 | Incretin synthesis, secretion, and inactivation Homo sapiens R-HSA-400508 | 4.60E-01 | 9.78E-01 | 1.33E+00 | 1.03E+00 | GRP;SEC11C |
| 493 | Signaling by BMP Homo sapiens R-HSA-201451 | 4.60E-01 | 9.78E-01 | 1.33E+00 | 1.03E+00 | ZFYVE16;BMPR1A |
| 494 | Deposition of new CENPA-containing nucleosomes at the centromere Homo sapiens R-HSA-606279 | 4.61E-01 | 9.78E-01 | 1.17E+00 | 9.03E-01 | MIS18BP1;H2AFZ;HIST1H2AC;CENPC |
| 495 | Nucleosome assembly Homo sapiens R-HSA-774815 | 4.61E-01 | 9.78E-01 | 1.17E+00 | 9.03E-01 | MIS18BP1;H2AFZ;HIST1H2AC;CENPC |
| 496 | Nonhomologous End-Joining (NHEJ) Homo sapiens R-HSA-5693571 | 4.61E-01 | 9.78E-01 | 1.17E+00 | 9.03E-01 | RAD50;HERC2;PRKDC;POLM |

| | | | | | | |
|---|---|---|---|---|---|---|
| 497 | Sperm Motility And Taxes Homo sapiens R-HSA-1300642 | 4.63E-01 | 9.78E-01 | 1.75E+00 | 1.35E+00 | HVCN1 |
| 498 | Organic cation transport Homo sapiens R-HSA-549127 | 4.63E-01 | 9.78E-01 | 1.75E+00 | 1.35E+00 | SLC22A15 |
| 499 | Displacement of DNA glycosylase by APEX1 Homo sapiens R-HSA-110357 | 4.63E-01 | 9.78E-01 | 1.75E+00 | 1.35E+00 | MBD4 |
| 500 | Retinoid cycle disease events Homo sapiens R-HSA-2453864 | 4.63E-01 | 9.78E-01 | 1.75E+00 | 1.35E+00 | NAPEPLD |
| 501 | Diseases associated with visual transduction Homo sapiens R-HSA-2474795 | 4.63E-01 | 9.78E-01 | 1.75E+00 | 1.35E+00 | NAPEPLD |
| 502 | Sodium/Proton exchangers Homo sapiens R-HSA-425986 | 4.63E-01 | 9.78E-01 | 1.75E+00 | 1.35E+00 | SLC9A6 |
| 503 | Nef Mediated CD4 Down-regulation Homo sapiens R-HSA-167590 | 4.63E-01 | 9.78E-01 | 1.75E+00 | 1.35E+00 | LCK |
| 504 | PTK6 Regulates RTKs and Their Effectors AKT1 and DOK1 Homo sapiens R-HSA-8849469 | 4.63E-01 | 9.78E-01 | 1.75E+00 | 1.35E+00 | ARAP1 |
| 505 | EGFR Transactivation by Gastrin Homo sapiens R-HSA-2179392 | 4.63E-01 | 9.78E-01 | 1.75E+00 | 1.35E+00 | EGFR |
| 506 | Small interfering RNA (siRNA) biogenesis Homo sapiens R-HSA-426486 | 4.63E-01 | 9.78E-01 | 1.75E+00 | 1.35E+00 | AGO3 |

| | | | | | | |
|---|---|---|---|---|---|---|
| 507 | RHO GTPases Activate Rhotekin and Rhophilins Homo sapiens R-HSA-5666185 | 4.63E-01 | 9.78E-01 | 1.75E+00 | 1.35E+00 | RHPN2 |
| 508 | Inactivation of Cdc42 and Rac Homo sapiens R-HSA-428543 | 4.63E-01 | 9.78E-01 | 1.75E+00 | 1.35E+00 | SRGAP3 |
| 509 | Ca2+ activated K+ channels Homo sapiens R-HSA-1296052 | 4.63E-01 | 9.78E-01 | 1.75E+00 | 1.35E+00 | KCNMB4 |
| 510 | Adrenoceptors Homo sapiens R-HSA-390696 | 4.63E-01 | 9.78E-01 | 1.75E+00 | 1.35E+00 | ADRA2C |
| 511 | Mitochondrial iron-sulfur cluster biogenesis Homo sapiens R-HSA-1362409 | 4.63E-01 | 9.78E-01 | 1.75E+00 | 1.35E+00 | SLC25A37 |
| 512 | Pregnenolone biosynthesis Homo sapiens R-HSA-196108 | 4.63E-01 | 9.78E-01 | 1.75E+00 | 1.35E+00 | AKR1B1 |
| 513 | Pyrimidine salvage reactions Homo sapiens R-HSA-73614 | 4.63E-01 | 9.78E-01 | 1.75E+00 | 1.35E+00 | TK1 |
| 514 | Interleukin-19, 20, 22, 24 Homo sapiens R-HSA-8854691 | 4.63E-01 | 9.78E-01 | 1.75E+00 | 1.35E+00 | IL10RB |
| 515 | O-glycosylation of TSR domain-containing proteins Homo sapiens R-HSA-5173214 | 4.70E-01 | 9.78E-01 | 1.20E+00 | 9.04E-01 | SBSPON;THSD7A;ADAMTS6 |
| 516 | Signaling by FGFR1 in disease Homo sapiens R-HSA-5655302 | 4.70E-01 | 9.78E-01 | 1.20E+00 | 9.04E-01 | GAB1;FGF2;FGFR1 |
| 517 | Interleukin receptor SHC signaling Homo sapiens R-HSA-912526 | 4.70E-01 | 9.78E-01 | 1.04E+00 | 7.86E-01 | KLB;DUSP1;PIK3R3;PEBP1;FGF2;EGFR;PSMB10;PSMB9;RAP1B;AKAP9;PSME1;CALM1;RAF1;SPTAN1;MARK3;SPTBN1;FGFR1 |

| | | | | | | |
|---|---|---|---|---|---|---|
| 518 | Neurotransmitter Receptor Binding And Downstream Transmission In The Postsynaptic Cell Homo sapiens R-HSA-112314 | 4.78E-01 | 9.78E-01 | 1.06E+00 | 7.83E-01 | GABBR1;ADCY9;RRAS;AKAP9;KCNJ16;RAF1;CALM1;PLCB2;KCNJ2;KCNJ3 |
| 519 | p75 NTR receptor-mediated signalling Homo sapiens R-HSA-193704 | 4.81E-01 | 9.78E-01 | 1.09E+00 | 7.97E-01 | NFKBIA;HDAC1;MCF2L;ARHGEF16;AATF;ECT2 |
| 520 | Glutamate Neurotransmitter Release Cycle Homo sapiens R-HSA-210500 | 4.83E-01 | 9.78E-01 | 1.27E+00 | 9.27E-01 | ARL6IP5;SLC38A2 |
| 521 | TNFs bind their physiological receptors Homo sapiens R-HSA-5669034 | 4.83E-01 | 9.78E-01 | 1.27E+00 | 9.27E-01 | TNFRSF11B;TNFRSF1A |
| 522 | Diseases associated with the TLR signaling cascade Homo sapiens R-HSA-5602358 | 4.83E-01 | 9.78E-01 | 1.27E+00 | 9.27E-01 | NFKBIA;TLR5 |
| 523 | Diseases of Immune System Homo sapiens R-HSA-5260271 | 4.83E-01 | 9.78E-01 | 1.27E+00 | 9.27E-01 | NFKBIA;TLR5 |
| 524 | Sema4D induced cell migration and growth-cone collapse Homo sapiens R-HSA-416572 | 4.83E-01 | 9.78E-01 | 1.27E+00 | 9.27E-01 | SEMA4D;MYH10 |
| 525 | Pyrimidine metabolism Homo sapiens R-HSA-73848 | 4.83E-01 | 9.78E-01 | 1.27E+00 | 9.27E-01 | NT5C3A;TK1 |
| 526 | NS1 Mediated Effects on Host Pathways Homo sapiens R-HSA-168276 | 4.88E-01 | 9.78E-01 | 1.17E+00 | 8.37E-01 | NUP93;NUP50;ISG15 |

| | | | | | | |
|---|---|---|---|---|---|---|
| 527 | VEGFR2 mediated cell proliferation Homo sapiens R-HSA-5218921 | 4.91E-01 | 9.78E-01 | 1.03E+00 | 7.32E-01 | KLB;DUSP1;PEBP1;ITPR2;FGF2;EGFR;PSMB10;PSMB9;RAP1B;AKAP9;PSME1;CALM1;RAF1;SPTAN1;MARK3;SPTBN1;FGFR1 |
| 528 | Cyclin A:Cdk2-associated events at S phase entry Homo sapiens R-HSA-69656 | 4.92E-01 | 9.78E-01 | 1.09E+00 | 7.75E-01 | RB1;PSME1;PSMB10;CDC25B;PSMB9 |
| 529 | Loss of Nlp from mitotic centrosomes Homo sapiens R-HSA-380259 | 4.92E-01 | 9.78E-01 | 1.09E+00 | 7.75E-01 | DYNC1H1;DCTN2;AKAP9;CDK5RAP2;HAUS1 |
| 530 | Loss of proteins required for interphase microtubule organization?from the centrosome Homo sapiens R-HSA-380284 | 4.92E-01 | 9.78E-01 | 1.09E+00 | 7.75E-01 | DYNC1H1;DCTN2;AKAP9;CDK5RAP2;HAUS1 |
| 531 | Creatine metabolism Homo sapiens R-HSA-71288 | 4.99E-01 | 9.78E-01 | 1.55E+00 | 1.08E+00 | SLC6A12 |
| 532 | Activation of Ca-permeable Kainate Receptor Homo sapiens R-HSA-451308 | 4.99E-01 | 9.78E-01 | 1.55E+00 | 1.08E+00 | CALM1 |
| 533 | Ionotropic activity of Kainate Receptors Homo sapiens R-HSA-451306 | 4.99E-01 | 9.78E-01 | 1.55E+00 | 1.08E+00 | CALM1 |
| 534 | ATF6-alpha activates chaperone genes Homo sapiens R-HSA-381183 | 4.99E-01 | 9.78E-01 | 1.55E+00 | 1.08E+00 | ATF4 |
| 535 | HDR through MMEJ (alt-NHEJ) Homo sapiens R-HSA-5685939 | 4.99E-01 | 9.78E-01 | 1.55E+00 | 1.08E+00 | RAD50 |

| | | | | | | |
|---|---|---|---|---|---|---|
| 536 | Regulation of gene expression by Hypoxia-inducible Factor Homo sapiens R-HSA-1234158 | 4.99E-01 | 9.78E-01 | 1.55E+00 | 1.08E+00 | HIF1A |
| 537 | FGFR2b ligand binding and activation Homo sapiens R-HSA-190377 | 4.99E-01 | 9.78E-01 | 1.55E+00 | 1.08E+00 | FGF2 |
| 538 | Signaling by FGFR3 fusions in cancer Homo sapiens R-HSA-8853334 | 4.99E-01 | 9.78E-01 | 1.55E+00 | 1.08E+00 | GAB1 |
| 539 | Activation of the AP-1 family of transcription factors Homo sapiens R-HSA-450341 | 4.99E-01 | 9.78E-01 | 1.55E+00 | 1.08E+00 | MAPK10 |
| 540 | GP1b-IX-V activation signalling Homo sapiens R-HSA-430116 | 4.99E-01 | 9.78E-01 | 1.55E+00 | 1.08E+00 | RAF1 |
| 541 | Dectin-2 family Homo sapiens R-HSA-5621480 | 4.99E-01 | 9.78E-01 | 1.55E+00 | 1.08E+00 | SYK |
| 542 | Folding of actin by CCT/TriC Homo sapiens R-HSA-390450 | 4.99E-01 | 9.78E-01 | 1.55E+00 | 1.08E+00 | CCT5 |
| 543 | Adenylate cyclase activating pathway Homo sapiens R-HSA-170660 | 4.99E-01 | 9.78E-01 | 1.55E+00 | 1.08E+00 | ADCY9 |
| 544 | eNOS activation Homo sapiens R-HSA-203615 | 4.99E-01 | 9.78E-01 | 1.55E+00 | 1.08E+00 | CALM1 |
| 545 | Acetylcholine regulates insulin secretion Homo sapiens R-HSA-399997 | 4.99E-01 | 9.78E-01 | 1.55E+00 | 1.08E+00 | PLCB2 |
| 546 | Beta-oxidation of pristanoyl-CoA Homo sapiens R-HSA-389887 | 4.99E-01 | 9.78E-01 | 1.55E+00 | 1.08E+00 | ACOX2 |

| | | | | | | |
|---|---|---|---|---|---|---|
| 547 | Purine catabolism Homo sapiens R-HSA-74259 | 4.99E-01 | 9.78E-01 | 1.55E+00 | 1.08E+00 | CAT |
| 548 | Calcitonin-like ligand receptors Homo sapiens R-HSA-419812 | 4.99E-01 | 9.78E-01 | 1.55E+00 | 1.08E+00 | CALCA |
| 549 | Urea cycle Homo sapiens R-HSA-70635 | 4.99E-01 | 9.78E-01 | 1.55E+00 | 1.08E+00 | ARG2 |
| 550 | HDMs demethylate histones Homo sapiens R-HSA-3214842 | 5.04E-01 | 9.78E-01 | 1.22E+00 | 8.32E-01 | KDM5B;KDM1A |
| 551 | Regulation of IFNA signaling Homo sapiens R-HSA-912694 | 5.04E-01 | 9.78E-01 | 1.22E+00 | 8.32E-01 | IFNAR2;IFNAR1 |
| 552 | RAF activation Homo sapiens R-HSA-5673000 | 5.04E-01 | 9.78E-01 | 1.22E+00 | 8.32E-01 | RAF1;MARK3 |
| 553 | Formation of tubulin folding intermediates by CCT/TriC Homo sapiens R-HSA-389960 | 5.04E-01 | 9.78E-01 | 1.22E+00 | 8.32E-01 | TUBA1C;CCT5 |
| 554 | Sulfur amino acid metabolism Homo sapiens R-HSA-1614635 | 5.04E-01 | 9.78E-01 | 1.22E+00 | 8.32E-01 | APIP;MAT1A |
| 555 | GRB2 events in EGFR signaling Homo sapiens R-HSA-179812 | 5.05E-01 | 9.78E-01 | 1.02E+00 | 6.99E-01 | KLB;DUSP1;PEBP1;FGF2;EGFR;PSMB10;PSMB9;RAP1B;AKAP9;PSME1;CALM1;RAF1;SPTAN1;MARK3;SPTBN1;FGFR1 |
| 556 | SHC1 events in EGFR signaling Homo sapiens R-HSA-180336 | 5.05E-01 | 9.78E-01 | 1.02E+00 | 6.99E-01 | KLB;DUSP1;PEBP1;FGF2;EGFR;PSMB10;PSMB9;RAP1B;AKAP9;PSME1;CALM1;RAF1;SPTAN1;MARK3;SPTBN1;FGFR1 |
| 557 | SOS-mediated signalling Homo sapiens R-HSA-112412 | 5.05E-01 | 9.78E-01 | 1.02E+00 | 6.99E-01 | KLB;DUSP1;PEBP1;FGF2;EGFR;PSMB10;PSMB9;RAP1B;AKAP9;PSME1;CALM1;RAF1;SPTAN1;MARK3;SPTBN1;FGFR1 |
| 558 | SHC1 events in ERBB4 signaling Homo sapiens R-HSA-1250347 | 5.05E-01 | 9.78E-01 | 1.02E+00 | 6.99E-01 | KLB;DUSP1;PEBP1;FGF2;EGFR;PSMB10;PSMB9;RAP1B;AKAP9;PSME1;CALM1;RAF1;SPTAN1;MARK3;SPTBN1;FGFR1 |

| | | | | | | |
|---|---|---|---|---|---|---|
| 559 | RAF/MAP kinase cascade Homo sapiens R-HSA-5673001 | 5.05E-01 | 9.78E-01 | 1.02E+00 | 6.99E-01 | KLB;DUSP1;PEBP1;FGF2;EGFR;PSMB10;PSMB9;RAP1B;AKAP9;PSME1;CALM1;RAF1;SPTAN1;MARK3;SPTBN1;FGFR1 |
| 560 | Meiosis Homo sapiens R-HSA-1500620 | 5.05E-01 | 9.78E-01 | 1.06E+00 | 7.26E-01 | SUN2;RAD50;H2AFZ;HSPA2;SYNE2;HIST1H2AC |
| 561 | Hedgehog 'on' state Homo sapiens R-HSA-5632684 | 5.05E-01 | 9.78E-01 | 1.06E+00 | 7.26E-01 | EVC2;EVC;BOC;PSME1;PSMB10;PSMB9 |
| 562 | PRC2 methylates histones and DNA Homo sapiens R-HSA-212300 | 5.05E-01 | 9.78E-01 | 1.13E+00 | 7.75E-01 | DNMT1;H2AFZ;HIST1H2AC |
| 563 | PKMTs methylate histone lysines Homo sapiens R-HSA-3214841 | 5.05E-01 | 9.78E-01 | 1.13E+00 | 7.75E-01 | SETD3;KMT2C;EHMT1 |
| 564 | Hh mutants that don't undergo autocatalytic processing are degraded by ERAD Homo sapiens R-HSA-5362768 | 5.05E-01 | 9.78E-01 | 1.10E+00 | 7.50E-01 | ERLEC1;PSME1;PSMB10;PSMB9 |
| 565 | Cell Cycle Homo sapiens R-HSA-1640170 | 5.10E-01 | 9.78E-01 | 1.01E+00 | 6.78E-01 | RB1;ANKLE2;DCTN2;HDAC1;GMNN;SMC4;SYNE2;PSMB10;CENPC;HERC2;MIS18BP1;HIST1H2AC;CDK5RAP2;CDKN2D;DYNC1H1;SUN2;RFC1;H2AFZ;CSNK2A2;KIF23;VRK2;HSPA2;LEMD2;PSMB9;CDC25B;HAUS1;NUP93;CLIP1;RAD50;POLA2;NUP50;AKAP9;PSME1;RAD17;NCAPD3;LPIN2;OPTN;MCPH1 |
| 566 | Signaling by Insulin receptor Homo sapiens R-HSA-74752 | 5.11E-01 | 9.78E-01 | 1.01E+00 | 6.80E-01 | KLB;CAB39L;DUSP1;GAB1;PEBP1;FGF2;EGFR;PSMB10;PSMB9;RAP1B;AKAP9;PSME1;ATP6V0A2;THEM4;ATP6V0A4;CALM1;RAF1;SPTAN1;MARK3;SPTBN1;FGFR1 |
| 567 | FRS-mediated FGFR2 signaling Homo sapiens R-HSA-5654700 | 5.12E-01 | 9.78E-01 | 1.02E+00 | 6.82E-01 | KLB;DUSP1;PEBP1;FGF2;EGFR;PSMB10;PSMB9;RAP1B;AKAP9;PSME1;CALM1;RAF1;SPTAN1;MARK3;SPTBN1;FGFR1 |
| 568 | FRS-mediated FGFR4 signaling Homo sapiens R-HSA-5654712 | 5.12E-01 | 9.78E-01 | 1.02E+00 | 6.82E-01 | KLB;DUSP1;PEBP1;FGF2;EGFR;PSMB10;PSMB9;RAP1B;AKAP9;PSME1;CALM1;RAF1;SPTAN1;MARK3;SPTBN1;FGFR1 |
| 569 | FRS-mediated FGFR3 signaling Homo sapiens R-HSA-5654706 | 5.12E-01 | 9.78E-01 | 1.02E+00 | 6.82E-01 | KLB;DUSP1;PEBP1;FGF2;EGFR;PSMB10;PSMB9;RAP1B;AKAP9;PSME1;CALM1;RAF1;SPTAN1;MARK3;SPTBN1;FGFR1 |

| | | | | | | |
|---|---|---|---|---|---|---|
| 570 | FRS-mediated FGFR1 signaling Homo sapiens R-HSA-5654693 | 5.12E-01 | 9.78E-01 | 1.02E+00 | 6.82E-01 | KLB;DUSP1;PEBP1;FGF2;EGFR;PSMB10;PSMB9;RAP1B;AKAP9;PSME1;CALM1;RAF1;SPTAN1;MARK3;SPTBN1;FGFR1 |
| 571 | Cellular Senescence Homo sapiens R-HSA-2559583 | 5.13E-01 | 9.78E-01 | 1.03E+00 | 6.84E-01 | RB1;CDKN2D;MAPK10;H1F0;RAD50;AGO3;H2AFZ;EHMT1;ETS2;HIST1H2AC;HIST1H1C |
| 572 | Cell Cycle, Mitotic Homo sapiens R-HSA-69278 | 5.15E-01 | 9.78E-01 | 1.01E+00 | 6.67E-01 | RB1;ANKLE2;DCTN2;HDAC1;GMNN;SMC4;PSMB10;CENPC;HIST1H2AC;CDK5RAP2;CDKN2D;DYNC1H1;RFC1;H2AFZ;CSNK2A2;KIF23;VRK2;LEMD2;PSMB9;CDC25B;HAUS1;NUP93;CLIP1;POLA2;NUP50;AKAP9;PSME1;NCAPD3;LPIN2;OPTN;MCPH1 |
| 573 | Gene Silencing by RNA Homo sapiens R-HSA-211000 | 5.15E-01 | 9.78E-01 | 1.04E+00 | 6.90E-01 | NUP93;AGO3;H2AFZ;NUP50;TDRD12;TDRD6;HIST1H2AC |
| 574 | Unfolded Protein Response (UPR) Homo sapiens R-HSA-381119 | 5.16E-01 | 9.78E-01 | 1.05E+00 | 6.93E-01 | DNAJC3;WFS1;CCL2;SEC62;DCP2;ATF4 |
| 575 | Chromosome Maintenance Homo sapiens R-HSA-73886 | 5.16E-01 | 9.78E-01 | 1.05E+00 | 6.93E-01 | POLA2;RFC1;MIS18BP1;H2AFZ;HIST1H2AC;CENPC |
| 576 | Role of LAT2/NTAL/LAB on calcium mobilization Homo sapiens R-HSA-2730905 | 5.21E-01 | 9.78E-01 | 1.02E+00 | 6.63E-01 | KLB;PRR5;AGO3;SYK;LCK;GAB1;PIK3R3;THEM4;FGF2;EGFR;FGFR1 |
| 577 | RNA Polymerase III Abortive And Retractive Initiation Homo sapiens R-HSA-749476 | 5.22E-01 | 9.78E-01 | 1.10E+00 | 7.18E-01 | CRCP;SNAPC1;BDP1 |
| 578 | RNA Polymerase III Transcription Homo sapiens R-HSA-74158 | 5.22E-01 | 9.78E-01 | 1.10E+00 | 7.18E-01 | CRCP;SNAPC1;BDP1 |
| 579 | Signalling to ERKs Homo sapiens R-HSA-187687 | 5.25E-01 | 9.78E-01 | 1.01E+00 | 6.49E-01 | KLB;DUSP1;KIDINS220;PEBP1;FGF2;EGFR;PSMB10;PSMB9;RAP1B;AKAP9;PSME1;CALM1;RAF1;SPTAN1;MARK3;SPTBN1;FGFR1 |
| 580 | Fertilization Homo sapiens R-HSA-1187000 | 5.26E-01 | 9.78E-01 | 1.17E+00 | 7.50E-01 | HVCN1;CD9 |

| | | | | | | |
|---|---|---|---|---|---|---|
| 581 | Reproduction Homo sapiens R-HSA-1474165 | 5.26E-01 | 9.78E-01 | 1.17E+00 | 7.50E-01 | HVCN1;CD9 |
| 582 | Regulation of Hypoxia-inducible Factor (HIF) by oxygen Homo sapiens R-HSA-1234174 | 5.26E-01 | 9.78E-01 | 1.17E+00 | 7.50E-01 | EGLN3;HIF1A |
| 583 | Cellular response to hypoxia Homo sapiens R-HSA-2262749 | 5.26E-01 | 9.78E-01 | 1.17E+00 | 7.50E-01 | EGLN3;HIF1A |
| 584 | Insulin receptor recycling Homo sapiens R-HSA-77387 | 5.26E-01 | 9.78E-01 | 1.17E+00 | 7.50E-01 | ATP6V0A2;ATP6V0A4 |
| 585 | Immune System Homo sapiens R-HSA-168256 | 5.27E-01 | 9.78E-01 | 9.97E-01 | 6.39E-01 | KLB;IFITM3;IFITM1;CRCP;IFITM2;PEBP1;FGF2;CLU;HERC2;KIF5B;KIFAP3;ICOSLG;IFNAR2;TAP1;CNPY3;HLA-E;NUP93;DYNC1LI2;ADCY9;AKAP9;PSME1;RAF1;CFB;IFNAR1;PRKDC;PDE1B;UBR4;PIK3R3;ITPR2;TNFRSF11B;DTX4;RAP1B;RNF213;ATP6V0A2;THEM4;ATP6V0A4;MARK3;SPTBN1;IP6K2;VASP;YES1;ISG15;BST2;NFKBIA;AGO3;ARPC2;REL;HLA-DRA;CALM1;EIF4G3;FGFR1;ITGB1;PRR5;ITGB5;RNF14;ARPC1B;CTSS;EEA1;UFL1;TRIM5;CTSH;ITGAV;SPTAN1;HLA-DPA1;SYK;DUSP1;GAB1;KIF23;OSMR;TNFRSF1A;IRF3;OAS1;LCK;OAS2;NUP50;IRF1;TLR5;HLA-DQB1;SP100;DCTN2;EGFR;MALT1;PSMB10;ZNRF1;HLA-DMA;HLA-DMB;GBP2;PAK2;GBP4;CD74;DYNC1H1;MAP3K3;ERAP2;IL10RB;CARD9;BAIAP2;PSMB9;MAPK10;TNIP2;TRIM38;SUGT1;IL18R1;SIPA1 |
| 586 | Regulation of PLK1 Activity at G2/M Transition Homo sapiens R-HSA-2565942 | 5.28E-01 | 9.78E-01 | 1.04E+00 | 6.62E-01 | DYNC1H1;DCTN2;AKAP9;OPTN;CDK5RAP2;HAUS1 |
| 587 | Peptide ligand-binding receptors Homo sapiens R-HSA-375276 | 5.28E-01 | 9.78E-01 | 1.01E+00 | 6.45E-01 | NMB;GRP;NPFF;F2R;NTS;SSTR2;EDNRB;SST;GPER1;ACKR4;F2RL1;CCL2;ACKR3 |

| | | | | | | |
|---|---|---|---|---|---|---|
| 588 | ISG15 antiviral mechanism Homo sapiens R-HSA-1169408 | 5.30E-01 | 9.78E-01 | 1.04E+00 | 6.62E-01 | NUP93;IRF3;NUP50;ISG15;EIF4G3 |
| 589 | Antiviral mechanism by IFN-stimulated genes Homo sapiens R-HSA-1169410 | 5.30E-01 | 9.78E-01 | 1.04E+00 | 6.62E-01 | NUP93;IRF3;NUP50;ISG15;EIF4G3 |
| 590 | Removal of licensing factors from origins Homo sapiens R-HSA-69300 | 5.30E-01 | 9.78E-01 | 1.04E+00 | 6.62E-01 | RB1;GMNN;PSME1;PSMB10;PSMB9 |
| 591 | AURKA Activation by TPX2 Homo sapiens R-HSA-8854518 | 5.30E-01 | 9.78E-01 | 1.04E+00 | 6.62E-01 | DYNC1H1;DCTN2;AKAP9;CDK5RAP2;HAUS1 |
| 592 | MAPK family signaling cascades Homo sapiens R-HSA-5683057 | 5.30E-01 | 9.78E-01 | 1.00E+00 | 6.36E-01 | KLB;DUSP1;PEBP1;FGF2;EGFR;PSMB10;PSMB9;RAP1B;AGO3;CDC42EP5;AKAP9;PSME1;CALM1;RAF1;SPTAN1;PAK2;MARK3;SPTBN1;FGFR1 |
| 593 | IRS-mediated signalling Homo sapiens R-HSA-112399 | 5.30E-01 | 9.78E-01 | 1.00E+00 | 6.36E-01 | KLB;CAB39L;DUSP1;GAB1;PEBP1;FGF2;EGFR;PSMB10;PSMB9;RAP1B;AKAP9;PSME1;THEM4;CALM1;RAF1;SPTAN1;MARK3;SPTBN1;FGFR1 |
| 594 | Uptake and function of anthrax toxins Homo sapiens R-HSA-5210891 | 5.32E-01 | 9.78E-01 | 1.40E+00 | 8.82E-01 | CALM1 |
| 595 | Dermatan sulfate biosynthesis Homo sapiens R-HSA-2022923 | 5.32E-01 | 9.78E-01 | 1.40E+00 | 8.82E-01 | DCN |
| 596 | Signaling by FGFR4 in disease Homo sapiens R-HSA-5655291 | 5.32E-01 | 9.78E-01 | 1.40E+00 | 8.82E-01 | GAB1 |
| 597 | CASP8 activity is inhibited Homo sapiens R-HSA-5218900 | 5.32E-01 | 9.78E-01 | 1.40E+00 | 8.82E-01 | TNFSF10 |
| 598 | Regulation by c-FLIP Homo sapiens R-HSA-3371378 | 5.32E-01 | 9.78E-01 | 1.40E+00 | 8.82E-01 | TNFSF10 |
| 599 | Dimerization of procaspase-8 Homo sapiens R-HSA-69416 | 5.32E-01 | 9.78E-01 | 1.40E+00 | 8.82E-01 | TNFSF10 |

| | | | | | | |
|---|---|---|---|---|---|---|
| 600 | Interleukin-7 signaling Homo sapiens R-HSA-1266695 | 5.32E-01 | 9.78E-01 | 1.40E+00 | 8.82E-01 | PIK3R3 |
| 601 | Activation of the phototransduction cascade Homo sapiens R-HSA-2485179 | 5.32E-01 | 9.78E-01 | 1.40E+00 | 8.82E-01 | CNGA1 |
| 602 | Sodium/Calcium exchangers Homo sapiens R-HSA-425561 | 5.32E-01 | 9.78E-01 | 1.40E+00 | 8.82E-01 | CALM1 |
| 603 | Apoptotic cleavage of cell adhesion proteins Homo sapiens R-HSA-351906 | 5.32E-01 | 9.78E-01 | 1.40E+00 | 8.82E-01 | DSP |
| 604 | Biotin transport and metabolism Homo sapiens R-HSA-196780 | 5.32E-01 | 9.78E-01 | 1.40E+00 | 8.82E-01 | PC |
| 605 | Androgen biosynthesis Homo sapiens R-HSA-193048 | 5.32E-01 | 9.78E-01 | 1.40E+00 | 8.82E-01 | SRD5A3 |
| 606 | Tryptophan catabolism Homo sapiens R-HSA-71240 | 5.32E-01 | 9.78E-01 | 1.40E+00 | 8.82E-01 | AFMID |
| 607 | Phenylalanine and tyrosine catabolism Homo sapiens R-HSA-71182 | 5.32E-01 | 9.78E-01 | 1.40E+00 | 8.82E-01 | HGD |
| 608 | Signalling to p38 via RIT and RIN Homo sapiens R-HSA-187706 | 5.32E-01 | 9.78E-01 | 1.00E+00 | 6.32E-01 | KLB;DUSP1;PEBP1;FGF2;EGFR;PSMB10;PSMB9;RAP1B;AKAP9;PSME1;CALM1;RAF1;SPTAN1;MARK3;SPTBN1;FGFR1 |
| 609 | Hh mutants abrogate ligand secretion Homo sapiens R-HSA-5387390 | 5.33E-01 | 9.78E-01 | 1.06E+00 | 6.63E-01 | ERLEC1;PSME1;PSMB10;PSMB9 |
| 610 | Signaling by ERBB4 Homo sapiens R-HSA-1236394 | 5.34E-01 | 9.78E-01 | 9.99E-01 | 6.26E-01 | KLB;PRR5;DUSP1;GAB1;PIK3R3;PEBP1;FGF2;EGFR;PSMB10;PSMB9;RAP1B;AGO3;LCK;AKAP9;PSME1;THEM4;CALM1;RAF1;SPTAN1;MARK3;SPTBN1;FGFR1 |

| | | | | | | |
|---|---|---|---|---|---|---|
| 611 | EPHB-mediated forward signaling Homo sapiens R-HSA-3928662 | 5.38E-01 | 9.78E-01 | 1.08E+00 | 6.66E-01 | YES1;ARPC2;ARPC1B |
| 612 | Frs2-mediated activation Homo sapiens R-HSA-170968 | 5.39E-01 | 9.78E-01 | 9.99E-01 | 6.17E-01 | KLB;DUSP1;PEBP1;FGF2;EGFR;PSMB10;PSMB9;RAP1B;AKAP9;PSME1;CALM1;RAF1;SPTAN1;MARK3;SPTBN1;FGFR1 |
| 613 | NoRC negatively regulates rRNA expression Homo sapiens R-HSA-427413 | 5.43E-01 | 9.78E-01 | 1.03E+00 | 6.29E-01 | DNMT1;H2AFZ;HDAC1;MBD2;HIST1H2AC |
| 614 | MAPK1/MAPK3 signaling Homo sapiens R-HSA-5684996 | 5.46E-01 | 9.78E-01 | 9.94E-01 | 6.01E-01 | KLB;DUSP1;PEBP1;FGF2;EGFR;PSMB10;PSMB9;RAP1B;AKAP9;PSME1;CALM1;RAF1;SPTAN1;MARK3;SPTBN1;FGFR1 |
| 615 | Uptake and actions of bacterial toxins Homo sapiens R-HSA-5339562 | 5.46E-01 | 9.78E-01 | 1.12E+00 | 6.76E-01 | CD9;CALM1 |
| 616 | RNA Polymerase III Transcription Initiation From Type 2 Promoter Homo sapiens R-HSA-76066 | 5.46E-01 | 9.78E-01 | 1.12E+00 | 6.76E-01 | CRCP;BDP1 |
| 617 | Integrin alphaIIb beta3 signaling Homo sapiens R-HSA-354192 | 5.46E-01 | 9.78E-01 | 1.12E+00 | 6.76E-01 | RAP1B;SYK |
| 618 | Prefoldin mediated transfer of substrate to CCT/TriC Homo sapiens R-HSA-389957 | 5.46E-01 | 9.78E-01 | 1.12E+00 | 6.76E-01 | TUBA1C;CCT5 |
| 619 | Peroxisomal lipid metabolism Homo sapiens R-HSA-390918 | 5.46E-01 | 9.78E-01 | 1.12E+00 | 6.76E-01 | ACOX2;HACL1 |
| 620 | rRNA modification in the nucleus Homo sapiens R-HSA-6790901 | 5.47E-01 | 9.78E-01 | 1.04E+00 | 6.24E-01 | NOP14;WDR75;DDX52;UTP18 |

| | | | | | | |
|---|---|---|---|---|---|---|
| 621 | Recruitment and ATM-mediated phosphorylation of repair and signaling proteins at DNA double strand breaks Homo sapiens R-HSA-5693565 | 5.47E-01 | 9.78E-01 | 1.04E+00 | 6.24E-01 | RAD50;HERC2;EYA4;BAZ1B |
| 622 | CDT1 association with the CDC6:ORC:origin complex Homo sapiens R-HSA-68827 | 5.47E-01 | 9.78E-01 | 1.04E+00 | 6.24E-01 | GMNN;PSME1;PSMB10;PSMB9 |
| 623 | Insulin receptor signalling cascade Homo sapiens R-HSA-74751 | 5.49E-01 | 9.78E-01 | 9.91E-01 | 5.94E-01 | KLB;CAB39L;DUSP1;GAB1;PEBP1;FGF2;EGFR;PSMB10;PSMB9;RAP1B;AKAP9;PSME1;THEM4;CALM1;RAF1;SPTAN1;MARK3;SPTBN1;FGFR1 |
| 624 | ERCC6 (CSB) and EHMT2 (G9a) positively regulate rRNA expression Homo sapiens R-HSA-427389 | 5.54E-01 | 9.78E-01 | 1.05E+00 | 6.19E-01 | HDAC1;H2AFZ;HIST1H2AC |
| 625 | Iron uptake and transport Homo sapiens R-HSA-917937 | 5.54E-01 | 9.78E-01 | 1.05E+00 | 6.19E-01 | FLVCR1;ATP6V0A2;ATP6V0A4 |
| 626 | IGF1R signaling cascade Homo sapiens R-HSA-2428924 | 5.55E-01 | 9.78E-01 | 9.87E-01 | 5.81E-01 | KLB;CAB39L;DUSP1;GAB1;PEBP1;FGF2;EGFR;PSMB10;PSMB9;RAP1B;AKAP9;PSME1;THEM4;CALM1;RAF1;SPTAN1;MARK3;SPTBN1;FGFR1 |
| 627 | Signaling by Type 1 Insulin-like Growth Factor 1 Receptor (IGF1R) Homo sapiens R-HSA-2404192 | 5.55E-01 | 9.78E-01 | 9.87E-01 | 5.81E-01 | KLB;CAB39L;DUSP1;GAB1;PEBP1;FGF2;EGFR;PSMB10;PSMB9;RAP1B;AKAP9;PSME1;THEM4;CALM1;RAF1;SPTAN1;MARK3;SPTBN1;FGFR1 |
| 628 | IRS-related events triggered by IGF1R Homo sapiens R-HSA-2428928 | 5.55E-01 | 9.78E-01 | 9.87E-01 | 5.81E-01 | KLB;CAB39L;DUSP1;GAB1;PEBP1;FGF2;EGFR;PSMB10;PSMB9;RAP1B;AKAP9;PSME1;THEM4;CALM1;RAF1;SPTAN1;MARK3;SPTBN1;FGFR1 |
| 629 | DNA Replication Homo sapiens R-HSA-69306 | 5.57E-01 | 9.78E-01 | 9.99E-01 | 5.84E-01 | RB1;POLA2;RFC1;GMNN;PSME1;PSMB10;PSMB9 |
| 630 | Signaling by Leptin Homo sapiens R-HSA-2586552 | 5.60E-01 | 9.78E-01 | 9.85E-01 | 5.72E-01 | KLB;DUSP1;PEBP1;FGF2;EGFR;PSMB10;PSMB9;RAP1B;AKAP9;PSME1;CALM1;RAF1;SPTAN1;MARK3;SPTBN1;FGFR1 |

| | | | | | | |
|---|---|---|---|---|---|---|
| 631 | Telomere Maintenance Homo sapiens R-HSA-157579 | 5.61E-01 | 9.78E-01 | 1.02E+00 | 5.88E-01 | POLA2;RFC1;H2AFZ;HIST1H2AC |
| 632 | DNA Double Strand Break Response Homo sapiens R-HSA-5693606 | 5.61E-01 | 9.78E-01 | 1.02E+00 | 5.88E-01 | RAD50;HERC2;EYA4;BAZ1B |
| 633 | FCERI mediated MAPK activation Homo sapiens R-HSA-2871796 | 5.62E-01 | 9.78E-01 | 9.84E-01 | 5.67E-01 | KLB;SYK;DUSP1;PEBP1;FGF2;EGFR;PSMB10;PSMB9;RAP1B;MAPK10;AKAP9;PSME1;CALM1;RAF1;SPTAN1;PAK2;MARK3;SPTBN1;FGFR1 |
| 634 | Reversible hydration of carbon dioxide Homo sapiens R-HSA-1475029 | 5.64E-01 | 9.78E-01 | 1.27E+00 | 7.29E-01 | CA5B |
| 635 | Keratan sulfate degradation Homo sapiens R-HSA-2022857 | 5.64E-01 | 9.78E-01 | 1.27E+00 | 7.29E-01 | FMOD |
| 636 | ATF6-alpha activates chaperones Homo sapiens R-HSA-381033 | 5.64E-01 | 9.78E-01 | 1.27E+00 | 7.29E-01 | ATF4 |
| 637 | FGFR3 mutant receptor activation Homo sapiens R-HSA-2033514 | 5.64E-01 | 9.78E-01 | 1.27E+00 | 7.29E-01 | FGF2 |
| 638 | Signaling by activated point mutants of FGFR3 Homo sapiens R-HSA-1839130 | 5.64E-01 | 9.78E-01 | 1.27E+00 | 7.29E-01 | FGF2 |
| 639 | The NLRP3 inflammasome Homo sapiens R-HSA-844456 | 5.64E-01 | 9.78E-01 | 1.27E+00 | 7.29E-01 | SUGT1 |
| 640 | Miscellaneous substrates Homo sapiens R-HSA-211958 | 5.64E-01 | 9.78E-01 | 1.27E+00 | 7.29E-01 | CYP4F11 |
| 641 | Eicosanoids Homo sapiens R-HSA-211979 | 5.64E-01 | 9.78E-01 | 1.27E+00 | 7.29E-01 | CYP4F11 |

| | | | | | | |
|---|---|---|---|---|---|---|
| 642 | Pyrimidine catabolism Homo sapiens R-HSA-73621 | 5.64E-01 | 9.78E-01 | 1.27E+00 | 7.29E-01 | NT5C3A |
| 643 | Tandem pore domain potassium channels Homo sapiens R-HSA-1296346 | 5.64E-01 | 9.78E-01 | 1.27E+00 | 7.29E-01 | KCNK3 |
| 644 | The role of Nef in HIV-1 replication and disease pathogenesis Homo sapiens R-HSA-164952 | 5.66E-01 | 9.78E-01 | 1.08E+00 | 6.12E-01 | LCK;PAK2 |
| 645 | RNA Polymerase III Transcription Initiation From Type 1 Promoter Homo sapiens R-HSA-76061 | 5.66E-01 | 9.78E-01 | 1.08E+00 | 6.12E-01 | CRCP;BDP1 |
| 646 | Extension of Telomeres Homo sapiens R-HSA-180786 | 5.66E-01 | 9.78E-01 | 1.08E+00 | 6.12E-01 | POLA2;RFC1 |
| 647 | Synthesis of PA Homo sapiens R-HSA-1483166 | 5.66E-01 | 9.78E-01 | 1.08E+00 | 6.12E-01 | PLA2G4A;AGPAT2 |
| 648 | Regulation of DNA replication Homo sapiens R-HSA-69304 | 5.67E-01 | 9.78E-01 | 9.99E-01 | 5.67E-01 | RB1;GMNN;PSME1;PSMB10;PSMB9 |
| 649 | Intra-Golgi traffic Homo sapiens R-HSA-6811438 | 5.70E-01 | 9.78E-01 | 1.02E+00 | 5.75E-01 | MAN2A2;MAN1C1;MAN1A1 |
| 650 | Transcriptional activity of SMAD2/SMAD3:SMAD4 heterotrimer Homo sapiens R-HSA-2173793 | 5.70E-01 | 9.78E-01 | 1.02E+00 | 5.75E-01 | WWTR1;HDAC1;SKIL |
| 651 | Translocation of GLUT4 to the plasma membrane Homo sapiens R-HSA-1445148 | 5.75E-01 | 9.78E-01 | 9.99E-01 | 5.53E-01 | RAB13;EXOC4;KIFAP3;CALM1 |

| | | | | | | |
|---|---|---|---|---|---|---|
| 652 | RHO GTPases activate PKNs Homo sapiens R-HSA-5625740 | 5.75E-01 | 9.78E-01 | 9.99E-01 | 5.53E-01 | KDM1A;H2AFZ;MYH10;HIST1H2AC |
| 653 | HDACs deacetylate histones Homo sapiens R-HSA-3214815 | 5.75E-01 | 9.78E-01 | 9.99E-01 | 5.53E-01 | KDM1A;HDAC1;TBL1X;HIST1H2AC |
| 654 | Negative epigenetic regulation of rRNA expression Homo sapiens R-HSA-5250941 | 5.79E-01 | 9.78E-01 | 9.85E-01 | 5.38E-01 | DNMT1;H2AFZ;HDAC1;MBD2;HIST1H2AC |
| 655 | Signalling to RAS Homo sapiens R-HSA-167044 | 5.80E-01 | 9.78E-01 | 9.72E-01 | 5.30E-01 | KLB;DUSP1;PEBP1;FGF2;EGFR;PSMB10;PSMB9;RAP1B;AKAP9;PSME1;CALM1;RAF1;SPTAN1;MARK3;SPTBN1;FGFR1 |
| 656 | MAPK6/MAPK4 signaling Homo sapiens R-HSA-5687128 | 5.83E-01 | 9.78E-01 | 9.75E-01 | 5.26E-01 | AGO3;CDC42EP5;PSME1;PAK2;PSMB10;PSMB9 |
| 657 | PIWI-interacting RNA (piRNA) biogenesis Homo sapiens R-HSA-5601884 | 5.86E-01 | 9.78E-01 | 1.04E+00 | 5.54E-01 | TDRD12;TDRD6 |
| 658 | Transferrin endocytosis and recycling Homo sapiens R-HSA-917977 | 5.86E-01 | 9.78E-01 | 1.04E+00 | 5.54E-01 | ATP6V0A2;ATP6V0A4 |
| 659 | Cell death signalling via NRAGE, NRIF and NADE Homo sapiens R-HSA-204998 | 5.88E-01 | 9.78E-01 | 9.81E-01 | 5.21E-01 | MCF2L;ARHGEF16;AATF;ECT2 |
| 660 | Mitotic Telophase/Cytokinesis Homo sapiens R-HSA-68884 | 5.93E-01 | 9.78E-01 | 1.17E+00 | 6.09E-01 | KIF23 |
| 661 | WNT5A-dependent internalization of FZD2, FZD5 and ROR2 Homo sapiens R-HSA-5140745 | 5.93E-01 | 9.78E-01 | 1.17E+00 | 6.09E-01 | WNT5A |

| | | | | | | |
|---|---|---|---|---|---|---|
| 662 | POU5F1 (OCT4), SOX2, NANOG activate genes related to proliferation Homo sapiens R-HSA-2892247 | 5.93E-01 | 9.78E-01 | 1.17E+00 | 6.09E-01 | FGF2 |
| 663 | NF-kB is activated and signals survival Homo sapiens R-HSA-209560 | 5.93E-01 | 9.78E-01 | 1.17E+00 | 6.09E-01 | NFKBIA |
| 664 | FGFRL1 modulation of FGFR1 signaling Homo sapiens R-HSA-5658623 | 5.93E-01 | 9.78E-01 | 1.17E+00 | 6.09E-01 | FGF2 |
| 665 | FGFR2c ligand binding and activation Homo sapiens R-HSA-190375 | 5.93E-01 | 9.78E-01 | 1.17E+00 | 6.09E-01 | FGF2 |
| 666 | ERBB2 Activates PTK6 Signaling Homo sapiens R-HSA-8847993 | 5.93E-01 | 9.78E-01 | 1.17E+00 | 6.09E-01 | EGFR |
| 667 | TNFR1-induced proapoptotic signaling Homo sapiens R-HSA-5357786 | 5.93E-01 | 9.78E-01 | 1.17E+00 | 6.09E-01 | TNFRSF1A |
| 668 | Synthesis, secretion, and inactivation of Glucose-dependent Insulinotropic Polypeptide (GIP) Homo sapiens R-HSA-400511 | 5.93E-01 | 9.78E-01 | 1.17E+00 | 6.09E-01 | SEC11C |
| 669 | alpha-linolenic (omega3) and linoleic (omega6) acid metabolism Homo sapiens R-HSA-2046104 | 5.93E-01 | 9.78E-01 | 1.17E+00 | 6.09E-01 | ELOVL1 |

| | | | | | | |
|---|---|---|---|---|---|---|
| 670 | alpha-linolenic acid (ALA) metabolism Homo sapiens R-HSA-2046106 | 5.93E-01 | 9.78E-01 | 1.17E+00 | 6.09E-01 | ELOVL1 |
| 671 | Gastrin-CREB signalling pathway via PKC and MAPK Homo sapiens R-HSA-881907 | 5.93E-01 | 9.78E-01 | 9.68E-01 | 5.06E-01 | KLB;RGS19;GRP;ITPR2;PIK3R3;PEBP1;NTS;FGF2;EGFR;PSMB10;RAP1B;CYSLTR2;EDNRB;SPTAN1;MARK3;SPTBN1;NMB;DUSP1;NPFF;F2R;PSMB9;AKAP9;PSME1;F2RL1;CALM1;RAF1;PLCB2;FGFR1 |
| 672 | DNA Damage Bypass Homo sapiens R-HSA-73893 | 6.01E-01 | 9.78E-01 | 9.75E-01 | 4.97E-01 | RFC1;USP43;ISG15 |
| 673 | G alpha (z) signalling events Homo sapiens R-HSA-418597 | 6.01E-01 | 9.78E-01 | 9.75E-01 | 4.97E-01 | RGS19;ADCY9;ADRA2C |
| 674 | Biosynthesis of the N-glycan precursor (dolichol lipid-linked oligosaccharide, LLO) and transfer to a nascent protein Homo sapiens R-HSA-446193 | 6.02E-01 | 9.78E-01 | 9.57E-01 | 4.85E-01 | ST6GAL1;SRD5A3;SLC17A5;DOLK;ST6GALNAC6 |
| 675 | NEP/NS2 Interacts with the Cellular Export Machinery Homo sapiens R-HSA-168333 | 6.04E-01 | 9.78E-01 | 9.99E-01 | 5.03E-01 | NUP93;NUP50 |
| 676 | Regulation of Glucokinase by Glucokinase Regulatory Protein Homo sapiens R-HSA-170822 | 6.04E-01 | 9.78E-01 | 9.99E-01 | 5.03E-01 | NUP93;NUP50 |
| 677 | Transport of Ribonucleoproteins into the Host Nucleus Homo sapiens R-HSA-168271 | 6.04E-01 | 9.78E-01 | 9.99E-01 | 5.03E-01 | NUP93;NUP50 |
| 678 | Tight junction interactions Homo sapiens R-HSA-420029 | 6.04E-01 | 9.78E-01 | 9.99E-01 | 5.03E-01 | CLDN4;CLDN1 |

| | | | | | | |
|---|---|---|---|---|---|---|
| 679 | Activation of Kainate Receptors upon glutamate binding Homo sapiens R-HSA-451326 | 6.04E-01 | 9.78E-01 | 9.99E-01 | 5.03E-01 | CALM1;PLCB2 |
| 680 | Asymmetric localization of PCP proteins Homo sapiens R-HSA-4608870 | 6.14E-01 | 9.78E-01 | 9.48E-01 | 4.63E-01 | WNT5A;PSME1;PSMB10;PSMB9 |
| 681 | Centrosome maturation Homo sapiens R-HSA-380287 | 6.14E-01 | 9.78E-01 | 9.44E-01 | 4.61E-01 | DYNC1H1;DCTN2;AKAP9;CDK5RAP2;HAUS1 |
| 682 | Recruitment of mitotic centrosome proteins and complexes Homo sapiens R-HSA-380270 | 6.14E-01 | 9.78E-01 | 9.44E-01 | 4.61E-01 | DYNC1H1;DCTN2;AKAP9;CDK5RAP2;HAUS1 |
| 683 | NOTCH1 Intracellular Domain Regulates Transcription Homo sapiens R-HSA-2122947 | 6.16E-01 | 9.78E-01 | 9.53E-01 | 4.63E-01 | HDAC1;TBL1X;HIF1A |
| 684 | Antigen activates B Cell Receptor (BCR) leading to generation of second messengers Homo sapiens R-HSA-983695 | 6.16E-01 | 9.78E-01 | 9.53E-01 | 4.63E-01 | SYK;ITPR2;CALM1 |
| 685 | Organic cation/anion/zwitterion transport Homo sapiens R-HSA-549132 | 6.20E-01 | 9.78E-01 | 1.08E+00 | 5.14E-01 | SLC22A15 |
| 686 | CS/DS degradation Homo sapiens R-HSA-2024101 | 6.20E-01 | 9.78E-01 | 1.08E+00 | 5.14E-01 | DCN |

| | | | | | | |
|---|---|---|---|---|---|---|
| 687 | TP53 regulates transcription of several additional cell death genes whose specific roles in p53-dependent apoptosis remain uncertain Homo sapiens R-HSA-6803205 | 6.20E-01 | 9.78E-01 | 1.08E+00 | 5.14E-01 | BCL6 |
| 688 | Mismatch repair (MMR) directed by MSH2:MSH6 (MutSalpha) Homo sapiens R-HSA-5358565 | 6.20E-01 | 9.78E-01 | 1.08E+00 | 5.14E-01 | MSH6 |
| 689 | Removal of the Flap Intermediate Homo sapiens R-HSA-69166 | 6.20E-01 | 9.78E-01 | 1.08E+00 | 5.14E-01 | POLA2 |
| 690 | TRAF3-dependent IRF activation pathway Homo sapiens R-HSA-918233 | 6.20E-01 | 9.78E-01 | 1.08E+00 | 5.14E-01 | IRF3 |
| 691 | Regulation of necroptotic cell death Homo sapiens R-HSA-5675482 | 6.20E-01 | 9.78E-01 | 1.08E+00 | 5.14E-01 | TNFSF10 |
| 692 | Bile salt and organic anion SLC transporters Homo sapiens R-HSA-425471 | 6.20E-01 | 9.78E-01 | 1.08E+00 | 5.14E-01 | SLC16A3 |
| 693 | Activation of Rac Homo sapiens R-HSA-428540 | 6.20E-01 | 9.78E-01 | 1.08E+00 | 5.14E-01 | PAK2 |
| 694 | Repression of WNT target genes Homo sapiens R-HSA-4641265 | 6.20E-01 | 9.78E-01 | 1.08E+00 | 5.14E-01 | HDAC1 |
| 695 | Inhibition of adenylate cyclase pathway Homo sapiens R-HSA-997269 | 6.20E-01 | 9.78E-01 | 1.08E+00 | 5.14E-01 | ADCY9 |

| | | | | | | |
|---|---|---|---|---|---|---|
| 696 | Adenylate cyclase inhibitory pathway Homo sapiens R-HSA-170670 | 6.20E-01 | 9.78E-01 | 1.08E+00 | 5.14E-01 | ADCY9 |
| 697 | Methylation Homo sapiens R-HSA-156581 | 6.20E-01 | 9.78E-01 | 1.08E+00 | 5.14E-01 | MAT1A |
| 698 | Export of Viral Ribonucleoproteins from Nucleus Homo sapiens R-HSA-168274 | 6.22E-01 | 9.78E-01 | 9.64E-01 | 4.57E-01 | NUP93;NUP50 |
| 699 | Packaging Of Telomere Ends Homo sapiens R-HSA-171306 | 6.22E-01 | 9.78E-01 | 9.64E-01 | 4.57E-01 | H2AFZ;HIST1H2AC |
| 700 | Negative regulation of FGFR4 signaling Homo sapiens R-HSA-5654733 | 6.22E-01 | 9.78E-01 | 9.64E-01 | 4.57E-01 | KLB;FGF2 |
| 701 | FGFR1 mutant receptor activation Homo sapiens R-HSA-1839124 | 6.22E-01 | 9.78E-01 | 9.64E-01 | 4.57E-01 | FGF2;FGFR1 |
| 702 | Signaling by Interleukins Homo sapiens R-HSA-449147 | 6.24E-01 | 9.78E-01 | 9.51E-01 | 4.49E-01 | KLB;PIK3R3;PEBP1;FGF2;EGFR;PSMB10;RAP1B;SPTAN1;MARK3;SPTBN1;MAP3K3;YES1;SYK;DUSP1;IL10RB;OSMR;PSMB9;LCK;TNIP2;AKAP9;PSME1;CALM1;RAF1;IL18R1;FGFR1 |
| 703 | Regulation of insulin secretion Homo sapiens R-HSA-422356 | 6.25E-01 | 9.78E-01 | 9.32E-01 | 4.38E-01 | KCNS3;SLC2A1;ITPR2;ADRA2C;PLCB2 |
| 704 | HDR through Homologous Recombination (HRR) Homo sapiens R-HSA-5685942 | 6.26E-01 | 9.78E-01 | 9.32E-01 | 4.36E-01 | RAD50;RFC1;RAD17;SPIDR |
| 705 | Hedgehog ligand biogenesis Homo sapiens R-HSA-5358346 | 6.26E-01 | 9.78E-01 | 9.32E-01 | 4.36E-01 | ERLEC1;PSME1;PSMB10;PSMB9 |
| 706 | G alpha (q) signalling events Homo sapiens R-HSA-416476 | 6.28E-01 | 9.78E-01 | 9.37E-01 | 4.35E-01 | NMB;CYSLTR2;EDNRB;RGS19;GRP;NPFF;F2R;ITPR2;PIK3R3;F2RL1;NTS;PLCB2 |

| | | | | | | |
|---|---|---|---|---|---|---|
| 707 | Beta-catenin independent WNT signaling Homo sapiens R-HSA-3858494 | 6.29E-01 | 9.78E-01 | 9.32E-01 | 4.31E-01 | WNT11;AGO3;WNT5A;PSME1;ITPR2;CALM1;PLCB2;PSMB10;PSMB9 |
| 708 | EPH-ephrin mediated repulsion of cells Homo sapiens R-HSA-3928665 | 6.30E-01 | 9.78E-01 | 9.32E-01 | 4.31E-01 | YES1;EPHA6;EPHA2 |
| 709 | Synthesis of DNA Homo sapiens R-HSA-69239 | 6.35E-01 | 9.78E-01 | 9.21E-01 | 4.18E-01 | RB1;POLA2;RFC1;PSME1;PSMB10;PSMB9 |
| 710 | Anchoring of the basal body to the plasma membrane Homo sapiens R-HSA-5620912 | 6.35E-01 | 9.78E-01 | 9.21E-01 | 4.18E-01 | DYNC1H1;DCTN2;AKAP9;CC2D2A;CDK5RAP2;HAUS1 |
| 711 | DNA Double-Strand Break Repair Homo sapiens R-HSA-5693532 | 6.38E-01 | 9.78E-01 | 9.25E-01 | 4.16E-01 | RAD50;HERC2;RFC1;PRKDC;POLM;EYA4;RAD17;BAZ1B;SPIDR |
| 712 | Nuclear import of Rev protein Homo sapiens R-HSA-180746 | 6.40E-01 | 9.78E-01 | 9.32E-01 | 4.16E-01 | NUP93;NUP50 |
| 713 | Vpr-mediated nuclear import of PICs Homo sapiens R-HSA-180910 | 6.40E-01 | 9.78E-01 | 9.32E-01 | 4.16E-01 | NUP93;NUP50 |
| 714 | Resolution of D-loop Structures through Holliday Junction Intermediates Homo sapiens R-HSA-5693568 | 6.40E-01 | 9.78E-01 | 9.32E-01 | 4.16E-01 | RAD50;SPIDR |
| 715 | DNA strand elongation Homo sapiens R-HSA-69190 | 6.40E-01 | 9.78E-01 | 9.32E-01 | 4.16E-01 | POLA2;RFC1 |
| 716 | SMAD2/SMAD3:SMAD4 heterotrimer regulates transcription Homo sapiens R-HSA-2173796 | 6.40E-01 | 9.78E-01 | 9.32E-01 | 4.16E-01 | WWTR1;HDAC1 |

| | | | | | | |
|---|---|---|---|---|---|---|
| 717 | Negative regulation of FGFR1 signaling Homo sapiens R-HSA-5654726 | 6.40E-01 | 9.78E-01 | 9.32E-01 | 4.16E-01 | FGF2;FGFR1 |
| 718 | TRAF6 mediated IRF7 activation Homo sapiens R-HSA-933541 | 6.40E-01 | 9.78E-01 | 9.32E-01 | 4.16E-01 | IRF3;IRF1 |
| 719 | Cooperation of Prefoldin and TriC/CCT in actin and tubulin folding Homo sapiens R-HSA-389958 | 6.40E-01 | 9.78E-01 | 9.32E-01 | 4.16E-01 | TUBA1C;CCT5 |
| 720 | Cross-presentation of soluble exogenous antigens (endosomes) Homo sapiens R-HSA-1236978 | 6.44E-01 | 9.78E-01 | 9.12E-01 | 4.01E-01 | PSME1;PSMB10;PSMB9 |
| 721 | WNT5A-dependent internalization of FZD4 Homo sapiens R-HSA-5099900 | 6.45E-01 | 9.78E-01 | 9.99E-01 | 4.37E-01 | WNT5A |
| 722 | mRNA decay by 5' to 3' exoribonuclease Homo sapiens R-HSA-430039 | 6.45E-01 | 9.78E-01 | 9.99E-01 | 4.37E-01 | DCP2 |
| 723 | Mismatch Repair Homo sapiens R-HSA-5358508 | 6.45E-01 | 9.78E-01 | 9.99E-01 | 4.37E-01 | MSH6 |
| 724 | Processive synthesis on the lagging strand Homo sapiens R-HSA-69183 | 6.45E-01 | 9.78E-01 | 9.99E-01 | 4.37E-01 | POLA2 |
| 725 | Association of licensing factors with the pre-replicative complex Homo sapiens R-HSA-69298 | 6.45E-01 | 9.78E-01 | 9.99E-01 | 4.37E-01 | GMNN |

| | | | | | | |
|---|---|---|---|---|---|---|
| 726 | Glycogen breakdown (glycogenolysis) Homo sapiens R-HSA-70221 | 6.45E-01 | 9.78E-01 | 9.99E-01 | 4.37E-01 | CALM1 |
| 727 | Negative regulation of TCF-dependent signaling by WNT ligand antagonists Homo sapiens R-HSA-3772470 | 6.45E-01 | 9.78E-01 | 9.99E-01 | 4.37E-01 | WNT5A |
| 728 | GRB2:SOS provides linkage to MAPK signaling for Integrins Homo sapiens R-HSA-354194 | 6.45E-01 | 9.78E-01 | 9.99E-01 | 4.37E-01 | RAP1B |
| 729 | p130Cas linkage to MAPK signaling for integrins Homo sapiens R-HSA-372708 | 6.45E-01 | 9.78E-01 | 9.99E-01 | 4.37E-01 | RAP1B |
| 730 | Elevation of cytosolic Ca2+ levels Homo sapiens R-HSA-139853 | 6.45E-01 | 9.78E-01 | 9.99E-01 | 4.37E-01 | ITPR2 |
| 731 | Eicosanoid ligand-binding receptors Homo sapiens R-HSA-391903 | 6.45E-01 | 9.78E-01 | 9.99E-01 | 4.37E-01 | CYSLTR2 |
| 732 | Fatty acids Homo sapiens R-HSA-211935 | 6.45E-01 | 9.78E-01 | 9.99E-01 | 4.37E-01 | CYP4F11 |
| 733 | DNA Replication Pre-Initiation Homo sapiens R-HSA-69002 | 6.47E-01 | 9.78E-01 | 9.08E-01 | 3.95E-01 | POLA2;GMNN;PSME1;PSMB10;PSMB9 |
| 734 | M/G1 Transition Homo sapiens R-HSA-68874 | 6.47E-01 | 9.78E-01 | 9.08E-01 | 3.95E-01 | POLA2;GMNN;PSME1;PSMB10;PSMB9 |
| 735 | Diseases of signal transduction Homo sapiens R-HSA-5663202 | 6.49E-01 | 9.78E-01 | 9.31E-01 | 4.03E-01 | KLB;PRR5;TGFB1;HDAC1;GAB1;PIK3R3;FGF2;EGFR;TGFBR1;PSMB10;PSMB9;TGFBR2;ERLEC1;LCK;PSME1;NAPEPLD;TBL1X;FGFR1 |

| | | | | | | |
|---|---|---|---|---|---|---|
| 736 | Cellular responses to stress Homo sapiens R-HSA-2262752 | 6.54E-01 | 9.78E-01 | 9.34E-01 | 3.96E-01 | RB1;CDKN2D;EGLN3;GPX2;H2AFZ;GPX8;EHMT1;HSPA2;HIF1A;ETS2;NUP93;MAPK10;H1F0;RAD50;AGO3;ATG101;PRDX1;NUP50;BAG1;CAT;ATP7A;HIST1H2AC;HIST1H1C |
| 737 | Sphingolipid de novo biosynthesis Homo sapiens R-HSA-1660661 | 6.56E-01 | 9.78E-01 | 9.02E-01 | 3.80E-01 | SGPL1;SPTLC1 |
| 738 | Cargo concentration in the ER Homo sapiens R-HSA-5694530 | 6.56E-01 | 9.78E-01 | 9.02E-01 | 3.80E-01 | CTSZ;MIA3 |
| 739 | Transport of the SLBP independent Mature mRNA Homo sapiens R-HSA-159227 | 6.56E-01 | 9.78E-01 | 9.02E-01 | 3.80E-01 | NUP93;NUP50 |
| 740 | Rev-mediated nuclear export of HIV RNA Homo sapiens R-HSA-165054 | 6.56E-01 | 9.78E-01 | 9.02E-01 | 3.80E-01 | NUP93;NUP50 |
| 741 | Resolution of D-Loop Structures Homo sapiens R-HSA-5693537 | 6.56E-01 | 9.78E-01 | 9.02E-01 | 3.80E-01 | RAD50;SPIDR |
| 742 | E2F mediated regulation of DNA replication Homo sapiens R-HSA-113510 | 6.56E-01 | 9.78E-01 | 9.02E-01 | 3.80E-01 | RB1;POLA2 |
| 743 | Formation of the ternary complex, and subsequently, the 43S complex Homo sapiens R-HSA-72695 | 6.58E-01 | 9.78E-01 | 8.92E-01 | 3.74E-01 | RPS4Y1;EIF2S2;EIF3C |
| 744 | CDK-mediated phosphorylation and removal of Cdc6 Homo sapiens R-HSA-69017 | 6.58E-01 | 9.78E-01 | 8.92E-01 | 3.74E-01 | PSME1;PSMB10;PSMB9 |

| | | | | | | |
|---|---|---|---|---|---|---|
| 745 | Ubiquitin-dependent degradation of Cyclin D1 Homo sapiens R-HSA-69229 | 6.58E-01 | 9.78E-01 | 8.92E-01 | 3.74E-01 | PSME1;PSMB10;PSMB9 |
| 746 | Ubiquitin-dependent degradation of Cyclin D Homo sapiens R-HSA-75815 | 6.58E-01 | 9.78E-01 | 8.92E-01 | 3.74E-01 | PSME1;PSMB10;PSMB9 |
| 747 | Regulation of ornithine decarboxylase (ODC) Homo sapiens R-HSA-350562 | 6.58E-01 | 9.78E-01 | 8.92E-01 | 3.74E-01 | PSME1;PSMB10;PSMB9 |
| 748 | Assembly of the pre-replicative complex Homo sapiens R-HSA-68867 | 6.62E-01 | 9.78E-01 | 8.87E-01 | 3.66E-01 | GMNN;PSME1;PSMB10;PSMB9 |
| 749 | RNA Polymerase I, RNA Polymerase III, and Mitochondrial Transcription Homo sapiens R-HSA-504046 | 6.63E-01 | 9.78E-01 | 8.97E-01 | 3.69E-01 | CRCP;SNAPC1;H2AFZ;HDAC1;MBD2;BDP1;HIST1H2AC |
| 750 | TP53 Regulates Metabolic Genes Homo sapiens R-HSA-5628897 | 6.68E-01 | 9.78E-01 | 8.84E-01 | 3.57E-01 | GPX2;SESN3;AGO3;PRDX1;DDIT4 |
| 751 | Translesion synthesis by REV1 Homo sapiens R-HSA-110312 | 6.69E-01 | 9.78E-01 | 9.32E-01 | 3.75E-01 | RFC1 |
| 752 | p75NTR signals via NF-kB Homo sapiens R-HSA-193639 | 6.69E-01 | 9.78E-01 | 9.32E-01 | 3.75E-01 | NFKBIA |
| 753 | GRB2 events in ERBB2 signaling Homo sapiens R-HSA-1963640 | 6.69E-01 | 9.78E-01 | 9.32E-01 | 3.75E-01 | EGFR |

| | | | | | | |
|---|---|---|---|---|---|---|
| 754 | JNK (c-Jun kinases) phosphorylation and activation mediated by activated human TAK1 Homo sapiens R-HSA-450321 | 6.69E-01 | 9.78E-01 | 9.32E-01 | 3.75E-01 | MAPK10 |
| 755 | MAP3K8 (TPL2)-dependent MAPK1/3 activation Homo sapiens R-HSA-5684264 | 6.69E-01 | 9.78E-01 | 9.32E-01 | 3.75E-01 | TNIP2 |
| 756 | Regulated Necrosis Homo sapiens R-HSA-5218859 | 6.69E-01 | 9.78E-01 | 9.32E-01 | 3.75E-01 | TNFSF10 |
| 757 | RIPK1-mediated regulated necrosis Homo sapiens R-HSA-5213460 | 6.69E-01 | 9.78E-01 | 9.32E-01 | 3.75E-01 | TNFSF10 |
| 758 | RHO GTPases activate CIT Homo sapiens R-HSA-5625900 | 6.69E-01 | 9.78E-01 | 9.32E-01 | 3.75E-01 | MYH10 |
| 759 | Metabolism of Angiotensinogen to Angiotensins Homo sapiens R-HSA-2022377 | 6.69E-01 | 9.78E-01 | 9.32E-01 | 3.75E-01 | CTSZ |
| 760 | PKA activation Homo sapiens R-HSA-163615 | 6.69E-01 | 9.78E-01 | 9.32E-01 | 3.75E-01 | ADCY9 |
| 761 | Acyl chain remodelling of PI Homo sapiens R-HSA-1482922 | 6.69E-01 | 9.78E-01 | 9.32E-01 | 3.75E-01 | PLA2G4A |
| 762 | Vpu mediated degradation of CD4 Homo sapiens R-HSA-180534 | 6.71E-01 | 9.78E-01 | 8.74E-01 | 3.49E-01 | PSME1;PSMB10;PSMB9 |
| 763 | Autodegradation of the E3 ubiquitin ligase COP1 Homo sapiens R-HSA-349425 | 6.71E-01 | 9.78E-01 | 8.74E-01 | 3.49E-01 | PSME1;PSMB10;PSMB9 |

| | | | | | | |
|---|---|---|---|---|---|---|
| 764 | Cargo trafficking to the periciliary membrane Homo sapiens R-HSA-5620920 | 6.71E-01 | 9.78E-01 | 8.74E-01 | 3.49E-01 | EXOC4;PKD2;CCT5 |
| 765 | Aquaporin-mediated transport Homo sapiens R-HSA-445717 | 6.71E-01 | 9.78E-01 | 8.74E-01 | 3.49E-01 | ADCY9;AQP6;AQP3 |
| 766 | Transport of the SLBP Dependant Mature mRNA Homo sapiens R-HSA-159230 | 6.73E-01 | 9.78E-01 | 8.74E-01 | 3.47E-01 | NUP93;NUP50 |
| 767 | Nuclear Pore Complex (NPC) Disassembly Homo sapiens R-HSA-3301854 | 6.73E-01 | 9.78E-01 | 8.74E-01 | 3.47E-01 | NUP93;NUP50 |
| 768 | Negative regulators of RIG-I/MDA5 signaling Homo sapiens R-HSA-936440 | 6.73E-01 | 9.78E-01 | 8.74E-01 | 3.47E-01 | IRF3;ISG15 |
| 769 | Lipoprotein metabolism Homo sapiens R-HSA-174824 | 6.73E-01 | 9.78E-01 | 8.74E-01 | 3.47E-01 | LIPC;ABCG1 |
| 770 | ROS, RNS production in response to bacteria Homo sapiens R-HSA-1222556 | 6.73E-01 | 9.78E-01 | 8.74E-01 | 3.47E-01 | ATP6V0A2;ATP6V0A4 |
| 771 | Cyclin E associated events during G1/S transition Homo sapiens R-HSA-69202 | 6.74E-01 | 9.78E-01 | 8.73E-01 | 3.45E-01 | RB1;PSME1;PSMB10;PSMB9 |
| 772 | p53-Independent DNA Damage Response Homo sapiens R-HSA-69610 | 6.84E-01 | 9.78E-01 | 8.56E-01 | 3.25E-01 | PSME1;PSMB10;PSMB9 |
| 773 | p53-Independent G1/S DNA damage checkpoint Homo sapiens R-HSA-69613 | 6.84E-01 | 9.78E-01 | 8.56E-01 | 3.25E-01 | PSME1;PSMB10;PSMB9 |

| | | | | | | |
|---|---|---|---|---|---|---|
| 774 | Ubiquitin Mediated Degradation of Phosphorylated Cdc25A Homo sapiens R-HSA-69601 | 6.84E-01 | 9.78E-01 | 8.56E-01 | 3.25E-01 | PSME1;PSMB10;PSMB9 |
| 775 | Interactions of Rev with host cellular proteins Homo sapiens R-HSA-177243 | 6.88E-01 | 9.78E-01 | 8.47E-01 | 3.17E-01 | NUP93;NUP50 |
| 776 | Interactions of Vpr with host cellular proteins Homo sapiens R-HSA-176033 | 6.88E-01 | 9.78E-01 | 8.47E-01 | 3.17E-01 | NUP93;NUP50 |
| 777 | SIRT1 negatively regulates rRNA Expression Homo sapiens R-HSA-427359 | 6.88E-01 | 9.78E-01 | 8.47E-01 | 3.17E-01 | H2AFZ;HIST1H2AC |
| 778 | Fanconi Anemia Pathway Homo sapiens R-HSA-6783310 | 6.88E-01 | 9.78E-01 | 8.47E-01 | 3.17E-01 | FANCC;FAN1 |
| 779 | KSRP (KHSRP) binds and destabilizes mRNA Homo sapiens R-HSA-450604 | 6.91E-01 | 9.78E-01 | 8.74E-01 | 3.23E-01 | DCP2 |
| 780 | Tristetraprolin (TTP, ZFP36) binds and destabilizes mRNA Homo sapiens R-HSA-450513 | 6.91E-01 | 9.78E-01 | 8.74E-01 | 3.23E-01 | DCP2 |
| 781 | Butyrate Response Factor 1 (BRF1) binds and destabilizes mRNA Homo sapiens R-HSA-450385 | 6.91E-01 | 9.78E-01 | 8.74E-01 | 3.23E-01 | DCP2 |
| 782 | Translesion synthesis by POLK Homo sapiens R-HSA-5655862 | 6.91E-01 | 9.78E-01 | 8.74E-01 | 3.23E-01 | RFC1 |

| | | | | | | |
|---|---|---|---|---|---|---|
| 783 | Translesion synthesis by POLI Homo sapiens R-HSA-5656121 | 6.91E-01 | 9.78E-01 | 8.74E-01 | 3.23E-01 | RFC1 |
| 784 | Activated point mutants of FGFR2 Homo sapiens R-HSA-2033519 | 6.91E-01 | 9.78E-01 | 8.74E-01 | 3.23E-01 | FGF2 |
| 785 | Ligand-dependent caspase activation Homo sapiens R-HSA-140534 | 6.91E-01 | 9.78E-01 | 8.74E-01 | 3.23E-01 | TNFSF10 |
| 786 | RHO GTPases Activate ROCKs Homo sapiens R-HSA-5627117 | 6.91E-01 | 9.78E-01 | 8.74E-01 | 3.23E-01 | MYH10 |
| 787 | Synthesis, secretion, and deacylation of Ghrelin Homo sapiens R-HSA-422085 | 6.91E-01 | 9.78E-01 | 8.74E-01 | 3.23E-01 | SEC11C |
| 788 | Unblocking of NMDA receptor, glutamate binding and activation Homo sapiens R-HSA-438066 | 6.91E-01 | 9.78E-01 | 8.74E-01 | 3.23E-01 | AKAP9 |
| 789 | PKA-mediated phosphorylation of CREB Homo sapiens R-HSA-111931 | 6.91E-01 | 9.78E-01 | 8.74E-01 | 3.23E-01 | ADCY9 |
| 790 | PKA activation in glucagon signalling Homo sapiens R-HSA-164378 | 6.91E-01 | 9.78E-01 | 8.74E-01 | 3.23E-01 | ADCY9 |
| 791 | Chylomicron-mediated lipid transport Homo sapiens R-HSA-174800 | 6.91E-01 | 9.78E-01 | 8.74E-01 | 3.23E-01 | LIPC |
| 792 | Inflammasomes Homo sapiens R-HSA-622312 | 6.91E-01 | 9.78E-01 | 8.74E-01 | 3.23E-01 | SUGT1 |
| 793 | Acyl chain remodelling of PG Homo sapiens R-HSA-1482925 | 6.91E-01 | 9.78E-01 | 8.74E-01 | 3.23E-01 | PLA2G4A |

| | | | | | | |
|---|---|---|---|---|---|---|
| 794 | Signaling by Hedgehog Homo sapiens R-HSA-5358351 | 6.94E-01 | 9.78E-01 | 8.73E-01 | 3.19E-01 | EVC2;ERLEC1;ADCY9;EVC;BOC;PSME1;PSMB10;PSMB9 |
| 795 | Switching of origins to a post-replicative state Homo sapiens R-HSA-69052 | 6.96E-01 | 9.78E-01 | 8.47E-01 | 3.07E-01 | RB1;PSME1;PSMB10;PSMB9 |
| 796 | Orc1 removal from chromatin Homo sapiens R-HSA-68949 | 6.96E-01 | 9.78E-01 | 8.47E-01 | 3.07E-01 | RB1;PSME1;PSMB10;PSMB9 |
| 797 | XBP1(S) activates chaperone genes Homo sapiens R-HSA-381038 | 6.96E-01 | 9.78E-01 | 8.39E-01 | 3.04E-01 | DNAJC3;WFS1;SEC62 |
| 798 | Vif-mediated degradation of APOBEC3G Homo sapiens R-HSA-180585 | 6.96E-01 | 9.78E-01 | 8.39E-01 | 3.04E-01 | PSME1;PSMB10;PSMB9 |
| 799 | GPVI-mediated activation cascade Homo sapiens R-HSA-114604 | 6.96E-01 | 9.78E-01 | 8.39E-01 | 3.04E-01 | SYK;LCK;PIK3R3 |
| 800 | Metabolism of nucleotides Homo sapiens R-HSA-15869 | 6.98E-01 | 9.79E-01 | 8.52E-01 | 3.07E-01 | NT5C3A;CAT;GMPR;AMPD2;TK1 |
| 801 | Lipid digestion, mobilization, and transport Homo sapiens R-HSA-73923 | 7.06E-01 | 9.79E-01 | 8.34E-01 | 2.90E-01 | LIPC;NCEH1;LIPH;ABCG1 |
| 802 | Golgi Associated Vesicle Biogenesis Homo sapiens R-HSA-432722 | 7.08E-01 | 9.79E-01 | 8.22E-01 | 2.84E-01 | SORT1;TPD52L1;CLINT1 |
| 803 | Meiotic recombination Homo sapiens R-HSA-912446 | 7.08E-01 | 9.79E-01 | 8.22E-01 | 2.84E-01 | RAD50;H2AFZ;HIST1H2AC |
| 804 | SCF-beta-TrCP mediated degradation of Emi1 Homo sapiens R-HSA-174113 | 7.08E-01 | 9.79E-01 | 8.22E-01 | 2.84E-01 | PSME1;PSMB10;PSMB9 |

| | | | | | | |
|---|---|---|---|---|---|---|
| 805 | Stabilization of p53 Homo sapiens R-HSA-69541 | 7.08E-01 | 9.79E-01 | 8.22E-01 | 2.84E-01 | PSME1;PSMB10;PSMB9 |
| 806 | AUF1 (hnRNP D0) binds and destabilizes mRNA Homo sapiens R-HSA-450408 | 7.08E-01 | 9.79E-01 | 8.22E-01 | 2.84E-01 | PSME1;PSMB10;PSMB9 |
| 807 | Degradation of AXIN Homo sapiens R-HSA-4641257 | 7.08E-01 | 9.79E-01 | 8.22E-01 | 2.84E-01 | PSME1;PSMB10;PSMB9 |
| 808 | Assembly of the primary cilium Homo sapiens R-HSA-5617833 | 7.11E-01 | 9.79E-01 | 8.73E-01 | 2.98E-01 | DYNC1H1;DCTN2;WDR60;AKAP9;EXOC4;KIFAP3;PKD2;CC2D2A;CCT5;CDK5RAP2;HAUS1 |
| 809 | Miscellaneous transport and binding events Homo sapiens R-HSA-5223345 | 7.12E-01 | 9.79E-01 | 8.22E-01 | 2.80E-01 | NIPAL3 |
| 810 | RNA Polymerase III Chain Elongation Homo sapiens R-HSA-73780 | 7.12E-01 | 9.79E-01 | 8.22E-01 | 2.80E-01 | CRCP |
| 811 | Pre-NOTCH Processing in Golgi Homo sapiens R-HSA-1912420 | 7.12E-01 | 9.79E-01 | 8.22E-01 | 2.80E-01 | ATP2A3 |
| 812 | SHC-mediated cascade:FGFR3 Homo sapiens R-HSA-5654704 | 7.12E-01 | 9.79E-01 | 8.22E-01 | 2.80E-01 | FGF2 |
| 813 | SHC1 events in ERBB2 signaling Homo sapiens R-HSA-1250196 | 7.12E-01 | 9.79E-01 | 8.22E-01 | 2.80E-01 | EGFR |
| 814 | Activation of IRF3/IRF7 mediated by TBK1/IKK epsilon Homo sapiens R-HSA-936964 | 7.12E-01 | 9.79E-01 | 8.22E-01 | 2.80E-01 | IRF3 |
| 815 | Defects in vitamin and cofactor metabolism Homo sapiens R-HSA-3296482 | 7.12E-01 | 9.79E-01 | 8.22E-01 | 2.80E-01 | PC |

| | | | | | | |
|---|---|---|---|---|---|---|
| 816 | Cytosolic sulfonation of small molecules Homo sapiens R-HSA-156584 | 7.12E-01 | 9.79E-01 | 8.22E-01 | 2.80E-01 | SLC35B3 |
| 817 | Transcriptional regulation by small RNAs Homo sapiens R-HSA-5578749 | 7.17E-01 | 9.79E-01 | 8.22E-01 | 2.74E-01 | NUP93;H2AFZ;NUP50;HIST1H2AC |
| 818 | Signaling by NOTCH1 Homo sapiens R-HSA-1980143 | 7.17E-01 | 9.79E-01 | 8.22E-01 | 2.74E-01 | HDAC1;TBL1X;DTX4;HIF1A |
| 819 | Transport of Mature mRNA Derived from an Intronless Transcript Homo sapiens R-HSA-159231 | 7.17E-01 | 9.79E-01 | 7.99E-01 | 2.65E-01 | NUP93;NUP50 |
| 820 | HDR through Single Strand Annealing (SSA) Homo sapiens R-HSA-5685938 | 7.17E-01 | 9.79E-01 | 7.99E-01 | 2.65E-01 | RAD50;RAD17 |
| 821 | DNA Repair Homo sapiens R-HSA-73894 | 7.20E-01 | 9.79E-01 | 8.85E-01 | 2.91E-01 | MBD4;RFC1;PRKDC;USP43;POLM;EYA4;ISG15;FANCC;BAZ1B;SPIDR;MSH6;RAD50;HERC2;RAD17;ERCC5;NEIL1;FAN1 |
| 822 | IRE1alpha activates chaperones Homo sapiens R-HSA-381070 | 7.20E-01 | 9.79E-01 | 8.06E-01 | 2.65E-01 | DNAJC3;WFS1;SEC62 |
| 823 | Toll-Like Receptors Cascades Homo sapiens R-HSA-168898 | 7.25E-01 | 9.79E-01 | 8.46E-01 | 2.73E-01 | NFKBIA;EEA1;MAPK10;IRF3;TNIP2;CNPY3;TLR5;CTSS |
| 824 | G2/M Transition Homo sapiens R-HSA-69275 | 7.26E-01 | 9.79E-01 | 8.57E-01 | 2.75E-01 | DYNC1H1;DCTN2;AKAP9;PSME1;OPTN;PSMB10;CDK5RAP2;CDC25B;HAUS1;PSMB9 |
| 825 | Positive epigenetic regulation of rRNA expression Homo sapiens R-HSA-5250913 | 7.27E-01 | 9.79E-01 | 8.10E-01 | 2.59E-01 | H2AFZ;HDAC1;BAZ1B;HIST1H2AC |
| 826 | S Phase Homo sapiens R-HSA-69242 | 7.29E-01 | 9.79E-01 | 8.36E-01 | 2.64E-01 | RB1;POLA2;RFC1;PSME1;PSMB10;CDC25B;PSMB9 |

| | | | | | | |
|---|---|---|---|---|---|---|
| 827 | Transport of Mature mRNAs Derived from Intronless Transcripts Homo sapiens R-HSA-159234 | 7.31E-01 | 9.79E-01 | 7.76E-01 | 2.43E-01 | NUP93;NUP50 |
| 828 | G1 Phase Homo sapiens R-HSA-69236 | 7.31E-01 | 9.79E-01 | 7.76E-01 | 2.43E-01 | RB1;CDKN2D |
| 829 | Cyclin D associated events in G1 Homo sapiens R-HSA-69231 | 7.31E-01 | 9.79E-01 | 7.76E-01 | 2.43E-01 | RB1;CDKN2D |
| 830 | TP53 Regulates Transcription of Genes Involved in Cytochrome C Release Homo sapiens R-HSA-6803204 | 7.31E-01 | 9.79E-01 | 7.77E-01 | 2.43E-01 | BNIP3L |
| 831 | Signaling by FGFR2 IIIa TM Homo sapiens R-HSA-8851708 | 7.31E-01 | 9.79E-01 | 7.77E-01 | 2.43E-01 | FGF2 |
| 832 | Regulation of TP53 Activity through Methylation Homo sapiens R-HSA-6804760 | 7.31E-01 | 9.79E-01 | 7.77E-01 | 2.43E-01 | EHMT1 |
| 833 | Translesion Synthesis by POLH Homo sapiens R-HSA-110320 | 7.31E-01 | 9.79E-01 | 7.77E-01 | 2.43E-01 | RFC1 |
| 834 | PCNA-Dependent Long Patch Base Excision Repair Homo sapiens R-HSA-5651801 | 7.31E-01 | 9.79E-01 | 7.77E-01 | 2.43E-01 | RFC1 |
| 835 | Ephrin signaling Homo sapiens R-HSA-3928664 | 7.31E-01 | 9.79E-01 | 7.77E-01 | 2.43E-01 | PAK2 |
| 836 | HDL-mediated lipid transport Homo sapiens R-HSA-194223 | 7.31E-01 | 9.79E-01 | 7.77E-01 | 2.43E-01 | ABCG1 |
| 837 | Degradation of DVL Homo sapiens R-HSA-4641258 | 7.31E-01 | 9.79E-01 | 7.91E-01 | 2.48E-01 | PSME1;PSMB10;PSMB9 |

| | | | | | | |
|---|---|---|---|---|---|---|
| 838 | Chemokine receptors bind chemokines Homo sapiens R-HSA-380108 | 7.31E-01 | 9.79E-01 | 7.91E-01 | 2.48E-01 | ACKR4;CCL2;ACKR3 |
| 839 | Innate Immune System Homo sapiens R-HSA-168249 | 7.31E-01 | 9.79E-01 | 9.20E-01 | 2.88E-01 | KLB;PRR5;CRCP;ARPC1B;PEBP1;CLU;FGF2;CTSS;EEA1;SPTAN1;SYK;DUSP1;GAB1;CNPY3;HLA-E;ADCY9;IRF3;LCK;IRF1;AKAP9;PSME1;RAF1;TLR5;CFB;PRKDC;PDE1B;PIK3R3;ITPR2;DTX4;EGFR;MALT1;PSMB10;RAP1B;THEM4;PAK2;MARK3;SPTBN1;YES1;CARD9;ISG15;BAIAP2;PSMB9;MAPK10;NFKBIA;AGO3;ARPC2;TNIP2;SUGT1;CALM1;FGFR1 |
| 840 | Oxidative Stress Induced Senescence Homo sapiens R-HSA-2559580 | 7.35E-01 | 9.81E-01 | 8.12E-01 | 2.50E-01 | CDKN2D;MAPK10;AGO3;H2AFZ;HIST1H2AC |
| 841 | PCP/CE pathway Homo sapiens R-HSA-4086400 | 7.35E-01 | 9.81E-01 | 8.12E-01 | 2.50E-01 | WNT11;WNT5A;PSME1;PSMB10;PSMB9 |
| 842 | Mitotic G2-G2/M phases Homo sapiens R-HSA-453274 | 7.39E-01 | 9.81E-01 | 8.46E-01 | 2.57E-01 | DYNC1H1;DCTN2;AKAP9;PSME1;OPTN;PSMB10;CDK5RAP2;CDC25B;HAUS1;PSMB9 |
| 843 | tRNA processing in the nucleus Homo sapiens R-HSA-6784531 | 7.42E-01 | 9.81E-01 | 7.76E-01 | 2.31E-01 | NUP93;NUP50;C2ORF49 |
| 844 | Ribosomal scanning and start codon recognition Homo sapiens R-HSA-72702 | 7.42E-01 | 9.81E-01 | 7.76E-01 | 2.31E-01 | RPS4Y1;EIF2S2;EIF3C |
| 845 | Translation initiation complex formation Homo sapiens R-HSA-72649 | 7.42E-01 | 9.81E-01 | 7.76E-01 | 2.31E-01 | RPS4Y1;EIF2S2;EIF3C |
| 846 | Formation of Fibrin Clot (Clotting Cascade) Homo sapiens R-HSA-140877 | 7.44E-01 | 9.81E-01 | 7.55E-01 | 2.23E-01 | F2R;TFPI |

| | | | | | | |
|---|---|---|---|---|---|---|
| 847 | Presynaptic phase of homologous DNA pairing and strand exchange Homo sapiens R-HSA-5693616 | 7.44E-01 | 9.81E-01 | 7.55E-01 | 2.23E-01 | RAD50;RAD17 |
| 848 | Dual Incision in GG-NER Homo sapiens R-HSA-5696400 | 7.44E-01 | 9.81E-01 | 7.55E-01 | 2.23E-01 | RFC1;ERCC5 |
| 849 | tRNA modification in the nucleus and cytosol Homo sapiens R-HSA-6782315 | 7.44E-01 | 9.81E-01 | 7.55E-01 | 2.23E-01 | TYW1;TRMT44 |
| 850 | SUMOylation of DNA damage response and repair proteins Homo sapiens R-HSA-3108214 | 7.46E-01 | 9.81E-01 | 7.87E-01 | 2.31E-01 | NUP93;SP100;HERC2;NUP50 |
| 851 | G alpha (12/13) signalling events Homo sapiens R-HSA-416482 | 7.46E-01 | 9.81E-01 | 7.87E-01 | 2.31E-01 | MCF2L;ARHGEF16;PIK3R3;ECT2 |
| 852 | Regulation of gene expression in beta cells Homo sapiens R-HSA-210745 | 7.49E-01 | 9.81E-01 | 7.36E-01 | 2.13E-01 | FOXA3 |
| 853 | IL-6-type cytokine receptor ligand interactions Homo sapiens R-HSA-6788467 | 7.49E-01 | 9.81E-01 | 7.36E-01 | 2.13E-01 | OSMR |
| 854 | FGFR2 ligand binding and activation Homo sapiens R-HSA-190241 | 7.49E-01 | 9.81E-01 | 7.36E-01 | 2.13E-01 | FGF2 |
| 855 | G beta:gamma signalling through PLC beta Homo sapiens R-HSA-418217 | 7.49E-01 | 9.81E-01 | 7.36E-01 | 2.13E-01 | PLCB2 |
| 856 | Signaling by Hippo Homo sapiens R-HSA-2028269 | 7.49E-01 | 9.81E-01 | 7.36E-01 | 2.13E-01 | WWTR1 |
| 857 | Glucuronidation Homo sapiens R-HSA-156588 | 7.49E-01 | 9.81E-01 | 7.36E-01 | 2.13E-01 | UGT2B11 |

| | | | | | | |
|---|---|---|---|---|---|---|
| 858 | Activation of the mRNA upon binding of the cap-binding complex and eIFs, and subsequent binding to 43S Homo sapiens R-HSA-72662 | 7.53E-01 | 9.82E-01 | 7.62E-01 | 2.16E-01 | RPS4Y1;EIF2S2;EIF3C |
| 859 | Formation of the beta-catenin:TCF transactivating complex Homo sapiens R-HSA-201722 | 7.53E-01 | 9.82E-01 | 7.62E-01 | 2.16E-01 | H2AFZ;HDAC1;HIST1H2AC |
| 860 | B-WICH complex positively regulates rRNA expression Homo sapiens R-HSA-5250924 | 7.53E-01 | 9.82E-01 | 7.62E-01 | 2.16E-01 | H2AFZ;BAZ1B;HIST1H2AC |
| 861 | NIK-->noncanonical NF-kB signaling Homo sapiens R-HSA-5676590 | 7.53E-01 | 9.82E-01 | 7.62E-01 | 2.16E-01 | PSME1;PSMB10;PSMB9 |
| 862 | RNA Polymerase I Promoter Clearance Homo sapiens R-HSA-73854 | 7.55E-01 | 9.84E-01 | 7.76E-01 | 2.18E-01 | H2AFZ;HDAC1;MBD2;HIST1H2AC |
| 863 | Degradation of GLI1 by the proteasome Homo sapiens R-HSA-5610780 | 7.63E-01 | 9.85E-01 | 7.48E-01 | 2.02E-01 | PSME1;PSMB10;PSMB9 |
| 864 | GLI3 is processed to GLI3R by the proteasome Homo sapiens R-HSA-5610785 | 7.63E-01 | 9.85E-01 | 7.48E-01 | 2.02E-01 | PSME1;PSMB10;PSMB9 |
| 865 | Degradation of GLI2 by the proteasome Homo sapiens R-HSA-5610783 | 7.63E-01 | 9.85E-01 | 7.48E-01 | 2.02E-01 | PSME1;PSMB10;PSMB9 |

| | | | | | | |
|---|---|---|---|---|---|---|
| 866 | The role of GTSE1 in G2/M progression after G2 checkpoint Homo sapiens R-HSA-8852276 | 7.63E-01 | 9.85E-01 | 7.48E-01 | 2.02E-01 | PSME1;PSMB10;PSMB9 |
| 867 | SCF(Skp2)-mediated degradation of p27/p21 Homo sapiens R-HSA-187577 | 7.63E-01 | 9.85E-01 | 7.48E-01 | 2.02E-01 | PSME1;PSMB10;PSMB9 |
| 868 | Senescence-Associated Secretory Phenotype (SASP) Homo sapiens R-HSA-2559582 | 7.64E-01 | 9.85E-01 | 7.65E-01 | 2.06E-01 | CDKN2D;H2AFZ;EHMT1;HIST1H2AC |
| 869 | Host Interactions of HIV factors Homo sapiens R-HSA-162909 | 7.66E-01 | 9.85E-01 | 8.01E-01 | 2.14E-01 | NUP93;LCK;NUP50;PSME1;PAK2;PSMB10;PSMB9 |
| 870 | Nef-mediates down modulation of cell surface receptors by recruiting them to clathrin adapters Homo sapiens R-HSA-164938 | 7.66E-01 | 9.85E-01 | 6.99E-01 | 1.86E-01 | LCK |
| 871 | RIP-mediated NFkB activation via ZBP1 Homo sapiens R-HSA-1810476 | 7.66E-01 | 9.85E-01 | 6.99E-01 | 1.86E-01 | NFKBIA |
| 872 | Presynaptic function of Kainate receptors Homo sapiens R-HSA-500657 | 7.66E-01 | 9.85E-01 | 6.99E-01 | 1.86E-01 | PLCB2 |
| 873 | Synthesis of very long-chain fatty acyl-CoAs Homo sapiens R-HSA-75876 | 7.66E-01 | 9.85E-01 | 6.99E-01 | 1.86E-01 | ELOVL1 |
| 874 | TNFR2 non-canonical NF-kB pathway Homo sapiens R-HSA-5668541 | 7.68E-01 | 9.85E-01 | 7.76E-01 | 2.05E-01 | PSME1;TNFRSF11B;PSMB10;TNFRSF1A;PSMB9 |

| | | | | | | |
|---|---|---|---|---|---|---|
| 875 | Intraflagellar transport Homo sapiens R-HSA-5620924 | 7.69E-01 | 9.85E-01 | 7.17E-01 | 1.88E-01 | WDR60;KIFAP3 |
| 876 | Histidine, lysine, phenylalanine, tyrosine, proline and tryptophan catabolism Homo sapiens R-HSA-6788656 | 7.69E-01 | 9.85E-01 | 7.17E-01 | 1.88E-01 | HGD;AFMID |
| 877 | Viral Messenger RNA Synthesis Homo sapiens R-HSA-168325 | 7.80E-01 | 9.85E-01 | 6.99E-01 | 1.73E-01 | NUP93;NUP50 |
| 878 | Homologous DNA Pairing and Strand Exchange Homo sapiens R-HSA-5693579 | 7.80E-01 | 9.85E-01 | 6.99E-01 | 1.73E-01 | RAD50;RAD17 |
| 879 | Cooperation of PDCL (PhLP1) and TRiC/CCT in G-protein beta folding Homo sapiens R-HSA-6814122 | 7.80E-01 | 9.85E-01 | 6.99E-01 | 1.73E-01 | CSNK2A2;CCT5 |
| 880 | Glucagon-like Peptide-1 (GLP1) regulates insulin secretion Homo sapiens R-HSA-381676 | 7.80E-01 | 9.85E-01 | 6.99E-01 | 1.73E-01 | KCNS3;ITPR2 |
| 881 | Fatty Acyl-CoA Biosynthesis Homo sapiens R-HSA-75105 | 7.80E-01 | 9.85E-01 | 6.99E-01 | 1.73E-01 | ELOVL1;THEM4 |
| 882 | tRNA Aminoacylation Homo sapiens R-HSA-379724 | 7.80E-01 | 9.85E-01 | 6.99E-01 | 1.73E-01 | IARS2;GARS |
| 883 | RHO GTPases Activate Formins Homo sapiens R-HSA-5663220 | 7.81E-01 | 9.85E-01 | 7.76E-01 | 1.92E-01 | ITGB1;FMNL3;DIAPH1;CLIP1;FMNL2;CENPC |
| 884 | Common Pathway of Fibrin Clot Formation Homo sapiens R-HSA-140875 | 7.81E-01 | 9.85E-01 | 6.66E-01 | 1.64E-01 | F2R |

| | | | | | | |
|---|---|---|---|---|---|---|
| 885 | Resolution of AP sites via the multiple-nucleotide patch replacement pathway Homo sapiens R-HSA-110373 | 7.81E-01 | 9.85E-01 | 6.66E-01 | 1.64E-01 | RFC1 |
| 886 | Cyclin A/B1 associated events during G2/M transition Homo sapiens R-HSA-69273 | 7.81E-01 | 9.85E-01 | 6.66E-01 | 1.64E-01 | CDC25B |
| 887 | Post-chaperonin tubulin folding pathway Homo sapiens R-HSA-389977 | 7.81E-01 | 9.85E-01 | 6.66E-01 | 1.64E-01 | TUBA1C |
| 888 | RNA Polymerase I Transcription Homo sapiens R-HSA-73864 | 7.82E-01 | 9.85E-01 | 7.45E-01 | 1.84E-01 | H2AFZ;HDAC1;MBD2;HIST1H2AC |
| 889 | RIG-I/MDA5 mediated induction of IFN-alpha/beta pathways Homo sapiens R-HSA-168928 | 7.82E-01 | 9.85E-01 | 7.45E-01 | 1.84E-01 | NFKBIA;IRF3;IRF1;ISG15 |
| 890 | Biological oxidations Homo sapiens R-HSA-211859 | 7.82E-01 | 9.85E-01 | 8.17E-01 | 2.01E-01 | SLC35B3;CYP39A1;POR;AOC1;UGT2B11;CYP4V2;EPHX1;NR1H4;MAT1A;CYP4F11;PTGS1 |
| 891 | Dectin-1 mediated noncanonical NF-kB signaling Homo sapiens R-HSA-5607761 | 7.83E-01 | 9.85E-01 | 7.23E-01 | 1.77E-01 | PSME1;PSMB10;PSMB9 |
| 892 | Cell-cell junction organization Homo sapiens R-HSA-421270 | 7.83E-01 | 9.85E-01 | 7.23E-01 | 1.77E-01 | CLDN4;CADM2;CLDN1 |
| 893 | Epigenetic regulation of gene expression Homo sapiens R-HSA-212165 | 7.88E-01 | 9.85E-01 | 7.69E-01 | 1.84E-01 | DNMT1;H2AFZ;HDAC1;MBD2;BAZ1B;HIST1H2AC |

| | | | | | | |
|---|---|---|---|---|---|---|
| 894 | Regulation of HSF1-mediated heat shock response Homo sapiens R-HSA-3371453 | 7.90E-01 | 9.85E-01 | 7.35E-01 | 1.73E-01 | NUP93;NUP50;BAG1;HSPA2 |
| 895 | Metabolism of water-soluble vitamins and cofactors Homo sapiens R-HSA-196849 | 7.91E-01 | 9.85E-01 | 7.51E-01 | 1.76E-01 | PANK2;PC;SLC52A3;SLC2A1;COASY |
| 896 | TP53 Regulates Transcription of Cell Death Genes Homo sapiens R-HSA-5633008 | 7.91E-01 | 9.85E-01 | 6.82E-01 | 1.59E-01 | BNIP3L;BCL6 |
| 897 | Signaling by FGFR2 in disease Homo sapiens R-HSA-5655253 | 7.91E-01 | 9.85E-01 | 6.82E-01 | 1.59E-01 | GAB1;FGF2 |
| 898 | Vasopressin regulates renal water homeostasis via Aquaporins Homo sapiens R-HSA-432040 | 7.91E-01 | 9.85E-01 | 6.82E-01 | 1.59E-01 | ADCY9;AQP3 |
| 899 | Voltage gated Potassium channels Homo sapiens R-HSA-1296072 | 7.91E-01 | 9.85E-01 | 6.82E-01 | 1.59E-01 | KCNG1;KCNS3 |
| 900 | Autodegradation of Cdh1 by Cdh1:APC/C Homo sapiens R-HSA-174084 | 7.92E-01 | 9.85E-01 | 7.10E-01 | 1.66E-01 | PSME1;PSMB10;PSMB9 |
| 901 | FCERI mediated Ca+2 mobilization Homo sapiens R-HSA-2871809 | 7.92E-01 | 9.85E-01 | 7.10E-01 | 1.66E-01 | SYK;ITPR2;CALM1 |
| 902 | Deadenylation of mRNA Homo sapiens R-HSA-429947 | 7.96E-01 | 9.85E-01 | 6.35E-01 | 1.45E-01 | TNKS1BP1 |
| 903 | RNA Polymerase III Transcription Termination Homo sapiens R-HSA-73980 | 7.96E-01 | 9.85E-01 | 6.35E-01 | 1.45E-01 | CRCP |

| | | | | | | |
|---|---|---|---|---|---|---|
| 904 | MicroRNA (miRNA) biogenesis Homo sapiens R-HSA-203927 | 7.96E-01 | 9.85E-01 | 6.35E-01 | 1.45E-01 | AGO3 |
| 905 | Gap-filling DNA repair synthesis and ligation in GG-NER Homo sapiens R-HSA-5696397 | 7.96E-01 | 9.85E-01 | 6.35E-01 | 1.45E-01 | RFC1 |
| 906 | RAF-independent MAPK1/3 activation Homo sapiens R-HSA-112409 | 7.96E-01 | 9.85E-01 | 6.35E-01 | 1.45E-01 | DUSP1 |
| 907 | BBSome-mediated cargo-targeting to cilium Homo sapiens R-HSA-5620922 | 7.96E-01 | 9.85E-01 | 6.35E-01 | 1.45E-01 | CCT5 |
| 908 | Cholesterol biosynthesis Homo sapiens R-HSA-191273 | 7.96E-01 | 9.85E-01 | 6.35E-01 | 1.45E-01 | FDFT1 |
| 909 | Regulation of TP53 Activity Homo sapiens R-HSA-5633007 | 7.97E-01 | 9.85E-01 | 7.81E-01 | 1.77E-01 | PRR5;RAD50;HDAC1;CSNK2A2;EHMT1;RAD17;BRD7;HIPK2 |
| 910 | Signaling by FGFR in disease Homo sapiens R-HSA-1226099 | 8.01E-01 | 9.85E-01 | 6.98E-01 | 1.55E-01 | GAB1;FGF2;FGFR1 |
| 911 | p53-Dependent G1/S DNA damage checkpoint Homo sapiens R-HSA-69580 | 8.01E-01 | 9.85E-01 | 6.98E-01 | 1.55E-01 | PSME1;PSMB10;PSMB9 |
| 912 | p53-Dependent G1 DNA Damage Response Homo sapiens R-HSA-69563 | 8.01E-01 | 9.85E-01 | 6.98E-01 | 1.55E-01 | PSME1;PSMB10;PSMB9 |
| 913 | SUMOylation of DNA replication proteins Homo sapiens R-HSA-4615885 | 8.02E-01 | 9.85E-01 | 6.65E-01 | 1.47E-01 | NUP93;NUP50 |
| 914 | Interleukin-1 signaling Homo sapiens R-HSA-446652 | 8.02E-01 | 9.85E-01 | 6.65E-01 | 1.47E-01 | MAP3K3;TNIP2 |

| | | | | | | |
|---|---|---|---|---|---|---|
| 915 | Degradation of beta-catenin by the destruction complex Homo sapiens R-HSA-195253 | 8.06E-01 | 9.85E-01 | 7.16E-01 | 1.55E-01 | HDAC1;PSME1;PSMB10;PSMB9 |
| 916 | Toll Like Receptor 10 (TLR10) Cascade Homo sapiens R-HSA-168142 | 8.06E-01 | 9.85E-01 | 7.16E-01 | 1.55E-01 | NFKBIA;MAPK10;TNIP2;TLR5 |
| 917 | Toll Like Receptor 5 (TLR5) Cascade Homo sapiens R-HSA-168176 | 8.06E-01 | 9.85E-01 | 7.16E-01 | 1.55E-01 | NFKBIA;MAPK10;TNIP2;TLR5 |
| 918 | MyD88 cascade initiated on plasma membrane Homo sapiens R-HSA-975871 | 8.06E-01 | 9.85E-01 | 7.16E-01 | 1.55E-01 | NFKBIA;MAPK10;TNIP2;TLR5 |
| 919 | DARPP-32 events Homo sapiens R-HSA-180024 | 8.10E-01 | 9.87E-01 | 6.08E-01 | 1.28E-01 | CALM1 |
| 920 | TRAF6 mediated NF-kB activation Homo sapiens R-HSA-933542 | 8.10E-01 | 9.87E-01 | 6.08E-01 | 1.28E-01 | NFKBIA |
| 921 | Cytosolic tRNA aminoacylation Homo sapiens R-HSA-379716 | 8.10E-01 | 9.87E-01 | 6.08E-01 | 1.28E-01 | GARS |
| 922 | Mitotic G1-G1/S phases Homo sapiens R-HSA-453279 | 8.11E-01 | 9.87E-01 | 7.57E-01 | 1.59E-01 | RB1;CDKN2D;POLA2;HDAC1;PSME1;PSMB10;PSMB9 |
| 923 | SUMOylation of RNA binding proteins Homo sapiens R-HSA-4570464 | 8.12E-01 | 9.87E-01 | 6.50E-01 | 1.35E-01 | NUP93;NUP50 |
| 924 | RMTs methylate histone arginines Homo sapiens R-HSA-3214858 | 8.12E-01 | 9.87E-01 | 6.50E-01 | 1.35E-01 | H2AFZ;HIST1H2AC |
| 925 | G1/S DNA Damage Checkpoints Homo sapiens R-HSA-69615 | 8.18E-01 | 9.89E-01 | 6.76E-01 | 1.36E-01 | PSME1;PSMB10;PSMB9 |

| | | | | | | |
|---|---|---|---|---|---|---|
| 926 | Fcgamma receptor (FCGR) dependent phagocytosis Homo sapiens R-HSA-2029480 | 8.20E-01 | 9.89E-01 | 7.35E-01 | 1.46E-01 | YES1;ARPC2;SYK;ARPC1B;ITPR2;BAIAP2 |
| 927 | Class B/2 (Secretin family receptors) Homo sapiens R-HSA-373080 | 8.20E-01 | 9.89E-01 | 6.98E-01 | 1.38E-01 | CALCA;WNT11;GIPR;WNT5A |
| 928 | Constitutive Signaling by AKT1 E17K in Cancer Homo sapiens R-HSA-5674400 | 8.22E-01 | 9.89E-01 | 5.82E-01 | 1.14E-01 | PRR5 |
| 929 | G0 and Early G1 Homo sapiens R-HSA-1538133 | 8.22E-01 | 9.89E-01 | 5.82E-01 | 1.14E-01 | HDAC1 |
| 930 | ADP signalling through P2Y purinoceptor 1 Homo sapiens R-HSA-418592 | 8.22E-01 | 9.89E-01 | 5.82E-01 | 1.14E-01 | PLA2G4A |
| 931 | TRP channels Homo sapiens R-HSA-3295583 | 8.22E-01 | 9.89E-01 | 5.82E-01 | 1.14E-01 | TRPV4 |
| 932 | tRNA processing Homo sapiens R-HSA-72306 | 8.25E-01 | 9.89E-01 | 7.12E-01 | 1.37E-01 | NUP93;NUP50;TYW1;C2ORF49;TRMT44 |
| 933 | APC/C:Cdc20 mediated degradation of Securin Homo sapiens R-HSA-174154 | 8.26E-01 | 9.89E-01 | 6.65E-01 | 1.27E-01 | PSME1;PSMB10;PSMB9 |
| 934 | Organelle biogenesis and maintenance Homo sapiens R-HSA-1852241 | 8.29E-01 | 9.89E-01 | 8.15E-01 | 1.52E-01 | DYNC1H1;GFM1;DCTN2;CHD9;MTIF2;NR1D1;PKD2;MRPS6;HAUS1;WDR60;TGS1;AKAP9;EXOC4;TBL1X;KIFAP3;CC2D2A;CCT5;CDK5RAP2 |
| 935 | Nucleotide-binding domain, leucine rich repeat containing receptor (NLR) signaling pathways Homo sapiens R-HSA-168643 | 8.31E-01 | 9.89E-01 | 6.21E-01 | 1.15E-01 | CARD9;SUGT1 |

| | | | | | | |
|---|---|---|---|---|---|---|
| 936 | Signaling by NOTCH Homo sapiens R-HSA-157118 | 8.32E-01 | 9.89E-01 | 7.22E-01 | 1.33E-01 | AGO3;HDAC1;ATP2A3;TBL1X;DTX4;HIF1A |
| 937 | Regulation of RAS by GAPs Homo sapiens R-HSA-5658442 | 8.33E-01 | 9.89E-01 | 6.55E-01 | 1.19E-01 | PSME1;PSMB10;PSMB9 |
| 938 | Signaling by PTK6 Homo sapiens R-HSA-8848021 | 8.33E-01 | 9.89E-01 | 6.55E-01 | 1.19E-01 | ARAP1;HIF1A;EGFR |
| 939 | Regulation of mRNA stability by proteins that bind AU-rich elements Homo sapiens R-HSA-450531 | 8.34E-01 | 9.89E-01 | 6.81E-01 | 1.23E-01 | PSME1;DCP2;PSMB10;PSMB9 |
| 940 | Recycling pathway of L1 Homo sapiens R-HSA-437239 | 8.34E-01 | 9.89E-01 | 5.59E-01 | 1.01E-01 | DPYSL2 |
| 941 | Attenuation phase Homo sapiens R-HSA-3371568 | 8.34E-01 | 9.89E-01 | 5.59E-01 | 1.01E-01 | HSPA2 |
| 942 | Resolution of D-loop Structures through Synthesis-Dependent Strand Annealing (SDSA) Homo sapiens R-HSA-5693554 | 8.34E-01 | 9.89E-01 | 5.59E-01 | 1.01E-01 | RAD50 |
| 943 | SHC-mediated cascade:FGFR2 Homo sapiens R-HSA-5654699 | 8.34E-01 | 9.89E-01 | 5.59E-01 | 1.01E-01 | FGF2 |
| 944 | TAK1 activates NFkB by phosphorylation and activation of IKKs complex Homo sapiens R-HSA-445989 | 8.34E-01 | 9.89E-01 | 5.59E-01 | 1.01E-01 | NFKBIA |
| 945 | TNFR1-induced NFkappaB signaling pathway Homo sapiens R-HSA-5357956 | 8.34E-01 | 9.89E-01 | 5.59E-01 | 1.01E-01 | TNFRSF1A |

| | | | | | | |
|---|---|---|---|---|---|---|
| 946 | Regulation of Complement cascade Homo sapiens R-HSA-977606 | 8.34E-01 | 9.89E-01 | 5.59E-01 | 1.01E-01 | CFB |
| 947 | Phase 2 - plateau phase Homo sapiens R-HSA-5576893 | 8.34E-01 | 9.89E-01 | 5.59E-01 | 1.01E-01 | AKAP9 |
| 948 | TP53 Regulates Transcription of Cell Cycle Genes Homo sapiens R-HSA-6791312 | 8.39E-01 | 9.93E-01 | 6.07E-01 | 1.06E-01 | RGCC;TNKS1BP1 |
| 949 | Death Receptor Signalling Homo sapiens R-HSA-73887 | 8.39E-01 | 9.93E-01 | 6.07E-01 | 1.06E-01 | TNFSF10;TNFRSF1A |
| 950 | EGFR downregulation Homo sapiens R-HSA-182971 | 8.45E-01 | 9.97E-01 | 5.37E-01 | 9.03E-02 | EGFR |
| 951 | Interleukin-6 family signaling Homo sapiens R-HSA-6783589 | 8.45E-01 | 9.97E-01 | 5.37E-01 | 9.03E-02 | OSMR |
| 952 | Effects of PIP2 hydrolysis Homo sapiens R-HSA-114508 | 8.45E-01 | 9.97E-01 | 5.37E-01 | 9.03E-02 | ITPR2 |
| 953 | Toll Like Receptor 9 (TLR9) Cascade Homo sapiens R-HSA-168138 | 8.47E-01 | 9.98E-01 | 6.65E-01 | 1.10E-01 | EEA1;NFKBIA;MAPK10;TNIP2 |
| 954 | FCGR activation Homo sapiens R-HSA-2029481 | 8.48E-01 | 9.98E-01 | 5.94E-01 | 9.82E-02 | YES1;SYK |
| 955 | Activation of anterior HOX genes in hindbrain development during early embryogenesis Homo sapiens R-HSA-5617472 | 8.53E-01 | 1.00E+00 | 6.57E-01 | 1.04E-01 | H2AFZ;KMT2C;HOXB1;HIST1H2AC |
| 956 | Activation of HOX genes during differentiation Homo sapiens R-HSA-5619507 | 8.53E-01 | 1.00E+00 | 6.57E-01 | 1.04E-01 | H2AFZ;KMT2C;HOXB1;HIST1H2AC |

| | | | | | | |
|---|---|---|---|---|---|---|
| 957 | Regulation of TP53 Activity through Phosphorylation Homo sapiens R-HSA-6804756 | 8.53E-01 | 1.00E+00 | 6.57E-01 | 1.04E-01 | RAD50;CSNK2A2;RAD17;HIPK2 |
| 958 | Deadenylation-dependent mRNA decay Homo sapiens R-HSA-429914 | 8.56E-01 | 1.00E+00 | 5.82E-01 | 9.07E-02 | TNKS1BP1;DCP2 |
| 959 | Energy dependent regulation of mTOR by LKB1-AMPK Homo sapiens R-HSA-380972 | 8.56E-01 | 1.00E+00 | 5.17E-01 | 8.07E-02 | CAB39L |
| 960 | Recognition of DNA damage by PCNA-containing replication complex Homo sapiens R-HSA-110314 | 8.56E-01 | 1.00E+00 | 5.17E-01 | 8.07E-02 | RFC1 |
| 961 | Adrenaline,noradrenaline inhibits insulin secretion Homo sapiens R-HSA-400042 | 8.56E-01 | 1.00E+00 | 5.17E-01 | 8.07E-02 | ADRA2C |
| 962 | APC/C:Cdh1 mediated degradation of Cdc20 and other APC/C:Cdh1 targeted proteins in late mitosis/early G1 Homo sapiens R-HSA-174178 | 8.61E-01 | 1.00E+00 | 6.16E-01 | 9.20E-02 | PSME1;PSMB10;PSMB9 |
| 963 | Cdc20:Phospho-APC/C mediated degradation of Cyclin A Homo sapiens R-HSA-174184 | 8.61E-01 | 1.00E+00 | 6.16E-01 | 9.20E-02 | PSME1;PSMB10;PSMB9 |
| 964 | G-protein beta:gamma signalling Homo sapiens R-HSA-397795 | 8.63E-01 | 1.00E+00 | 5.70E-01 | 8.39E-02 | PIK3R3;PLCB2 |

| | | | | | | |
|---|---|---|---|---|---|---|
| 965 | Negative regulation of FGFR3 signaling Homo sapiens R-HSA-5654732 | 8.65E-01 | 1.00E+00 | 4.99E-01 | 7.22E-02 | FGF2 |
| 966 | Myogenesis Homo sapiens R-HSA-525793 | 8.65E-01 | 1.00E+00 | 4.99E-01 | 7.22E-02 | BOC |
| 967 | CDO in myogenesis Homo sapiens R-HSA-375170 | 8.65E-01 | 1.00E+00 | 4.99E-01 | 7.22E-02 | BOC |
| 968 | APC:Cdc20 mediated degradation of cell cycle proteins prior to satisfation of the cell cycle checkpoint Homo sapiens R-HSA-179419 | 8.67E-01 | 1.00E+00 | 6.07E-01 | 8.63E-02 | PSME1;PSMB10;PSMB9 |
| 969 | TRAF6 Mediated Induction of proinflammatory cytokines Homo sapiens R-HSA-168180 | 8.67E-01 | 1.00E+00 | 6.07E-01 | 8.63E-02 | NFKBIA;MAPK10;TNIP2 |
| 970 | FCERI mediated NF-kB activation Homo sapiens R-HSA-2871837 | 8.71E-01 | 1.00E+00 | 6.58E-01 | 9.11E-02 | NFKBIA;PSME1;PSMB10;MALT1;PSMB9 |
| 971 | Activation of the pre-replicative complex Homo sapiens R-HSA-68962 | 8.74E-01 | 1.00E+00 | 4.82E-01 | 6.47E-02 | POLA2 |
| 972 | Activated NOTCH1 Transmits Signal to the Nucleus Homo sapiens R-HSA-2122948 | 8.74E-01 | 1.00E+00 | 4.82E-01 | 6.47E-02 | DTX4 |
| 973 | MAPK targets/ Nuclear events mediated by MAP kinases Homo sapiens R-HSA-450282 | 8.74E-01 | 1.00E+00 | 4.82E-01 | 6.47E-02 | MAPK10 |

| | | | | | | |
|---|---|---|---|---|---|---|
| 974 | Glucagon-type ligand receptors Homo sapiens R-HSA-420092 | 8.74E-01 | 1.00E+00 | 4.82E-01 | 6.47E-02 | GIPR |
| 975 | HDR through Homologous Recombination (HR) or Single Strand Annealing (SSA) Homo sapiens R-HSA-5693567 | 8.76E-01 | 1.00E+00 | 6.52E-01 | 8.66E-02 | RAD50;HERC2;RFC1;RAD17;SPIDR |
| 976 | G1/S Transition Homo sapiens R-HSA-69206 | 8.76E-01 | 1.00E+00 | 6.52E-01 | 8.66E-02 | RB1;POLA2;PSME1;PSMB10;PSMB9 |
| 977 | Sphingolipid metabolism Homo sapiens R-HSA-428157 | 8.79E-01 | 1.00E+00 | 5.90E-01 | 7.59E-02 | GALC;SGPL1;SPTLC1 |
| 978 | APC/C:Cdc20 mediated degradation of mitotic proteins Homo sapiens R-HSA-176409 | 8.79E-01 | 1.00E+00 | 5.90E-01 | 7.59E-02 | PSME1;PSMB10;PSMB9 |
| 979 | NOD1/2 Signaling Pathway Homo sapiens R-HSA-168638 | 8.83E-01 | 1.00E+00 | 4.66E-01 | 5.81E-02 | CARD9 |
| 980 | Regulation of TNFR1 signaling Homo sapiens R-HSA-5357905 | 8.83E-01 | 1.00E+00 | 4.66E-01 | 5.81E-02 | TNFRSF1A |
| 981 | Adherens junctions interactions Homo sapiens R-HSA-418990 | 8.83E-01 | 1.00E+00 | 4.66E-01 | 5.81E-02 | CADM2 |
| 982 | Activation of APC/C and APC/C:Cdc20 mediated degradation of mitotic proteins Homo sapiens R-HSA-176814 | 8.85E-01 | 1.00E+00 | 5.82E-01 | 7.11E-02 | PSME1;PSMB10;PSMB9 |

| | | | | | | |
|---|---|---|---|---|---|---|
| 983 | Activation of Matrix Metalloproteinases Homo sapiens R-HSA-1592389 | 8.91E-01 | 1.00E+00 | 4.51E-01 | 5.22E-02 | MMP7 |
| 984 | Regulation of beta-cell development Homo sapiens R-HSA-186712 | 8.91E-01 | 1.00E+00 | 4.51E-01 | 5.22E-02 | FOXA3 |
| 985 | Signal amplification Homo sapiens R-HSA-392518 | 8.91E-01 | 1.00E+00 | 4.51E-01 | 5.22E-02 | PLA2G4A |
| 986 | Thrombin signalling through proteinase activated receptors (PARs) Homo sapiens R-HSA-456926 | 8.91E-01 | 1.00E+00 | 4.51E-01 | 5.22E-02 | F2R |
| 987 | Cellular response to heat stress Homo sapiens R-HSA-3371556 | 8.91E-01 | 1.00E+00 | 6.07E-01 | 7.03E-02 | NUP93;NUP50;BAG1;HSPA2 |
| 988 | SUMO E3 ligases SUMOylate target proteins Homo sapiens R-HSA-3108232 | 8.91E-01 | 1.00E+00 | 6.07E-01 | 7.03E-02 | NUP93;SP100;HERC2;NUP50 |
| 989 | Signaling by Wnt Homo sapiens R-HSA-195721 | 8.93E-01 | 1.00E+00 | 7.46E-01 | 8.48E-02 | H2AFZ;HDAC1;CSNK2A2;WNT5A;USP34;ITPR2;PSMB10;PSMB9;WNT11;AGO3;PSME1;CALM1;PLCB2;TMED5;HIST1H2AC |
| 990 | MyD88-independent TLR3/TLR4 cascade Homo sapiens R-HSA-166166 | 8.95E-01 | 1.00E+00 | 6.00E-01 | 6.65E-02 | NFKBIA;MAPK10;IRF3;TNIP2 |
| 991 | Toll Like Receptor 3 (TLR3) Cascade Homo sapiens R-HSA-168164 | 8.95E-01 | 1.00E+00 | 6.00E-01 | 6.65E-02 | NFKBIA;MAPK10;IRF3;TNIP2 |
| 992 | TRIF-mediated TLR3/TLR4 signaling Homo sapiens R-HSA-937061 | 8.95E-01 | 1.00E+00 | 6.00E-01 | 6.65E-02 | NFKBIA;MAPK10;IRF3;TNIP2 |

| | | | | | | |
|---|---|---|---|---|---|---|
| 993 | Regulation of actin dynamics for phagocytic cup formation Homo sapiens R-HSA-2029482 | 8.95E-01 | 1.00E+00 | 6.00E-01 | 6.65E-02 | ARPC2;SYK;ARPC1B;BAIAP2 |
| 994 | Transcriptional Regulation by TP53 Homo sapiens R-HSA-3700989 | 8.96E-01 | 1.00E+00 | 7.59E-01 | 8.30E-02 | PRR5;BNIP3L;GPX2;HDAC1;CSNK2A2;EHMT1;FANCC;HIPK2;RAD50;RGCC;AGO3;SESN3;BCL6;PRDX1;DDIT4;RAD17;TNKS1BP1;BRD7 |
| 995 | FGFR2 mutant receptor activation Homo sapiens R-HSA-1839126 | 8.98E-01 | 1.00E+00 | 4.36E-01 | 4.70E-02 | FGF2 |
| 996 | Glucagon signaling in metabolic regulation Homo sapiens R-HSA-163359 | 8.98E-01 | 1.00E+00 | 4.36E-01 | 4.70E-02 | ADCY9 |
| 997 | Mitotic Anaphase Homo sapiens R-HSA-68882 | 8.98E-01 | 1.00E+00 | 6.76E-01 | 7.28E-02 | ANKLE2;CLIP1;PSME1;VRK2;LEMD2;PSMB10;CENPC;PSMB9 |
| 998 | G2/M DNA damage checkpoint Homo sapiens R-HSA-69473 | 9.00E-01 | 1.00E+00 | 5.58E-01 | 5.87E-02 | RAD50;HERC2;RAD17 |
| 999 | Mitotic Metaphase and Anaphase Homo sapiens R-HSA-2555396 | 9.01E-01 | 1.00E+00 | 6.72E-01 | 6.99E-02 | ANKLE2;CLIP1;PSME1;VRK2;LEMD2;PSMB10;CENPC;PSMB9 |
| 1000 | RNA Polymerase I Chain Elongation Homo sapiens R-HSA-73777 | 9.01E-01 | 1.00E+00 | 5.08E-01 | 5.27E-02 | H2AFZ;HIST1H2AC |
| 1001 | Constitutive Signaling by NOTCH1 HD+PEST Domain Mutants Homo sapiens R-HSA-2894862 | 9.01E-01 | 1.00E+00 | 5.08E-01 | 5.27E-02 | HDAC1;TBL1X |
| 1002 | Signaling by NOTCH1 in Cancer Homo sapiens R-HSA-2644603 | 9.01E-01 | 1.00E+00 | 5.08E-01 | 5.27E-02 | HDAC1;TBL1X |

| | | | | | | |
|---|---|---|---|---|---|---|
| 1003 | Signaling by NOTCH1 PEST Domain Mutants in Cancer Homo sapiens R-HSA-2644602 | 9.01E-01 | 1.00E+00 | 5.08E-01 | 5.27E-02 | HDAC1;TBL1X |
| 1004 | Constitutive Signaling by NOTCH1 PEST Domain Mutants Homo sapiens R-HSA-2644606 | 9.01E-01 | 1.00E+00 | 5.08E-01 | 5.27E-02 | HDAC1;TBL1X |
| 1005 | Signaling by NOTCH1 HD+PEST Domain Mutants in Cancer Homo sapiens R-HSA-2894858 | 9.01E-01 | 1.00E+00 | 5.08E-01 | 5.27E-02 | HDAC1;TBL1X |
| 1006 | Pre-NOTCH Expression and Processing Homo sapiens R-HSA-1912422 | 9.01E-01 | 1.00E+00 | 5.08E-01 | 5.27E-02 | AGO3;ATP2A3 |
| 1007 | Homology Directed Repair Homo sapiens R-HSA-5693538 | 9.02E-01 | 1.00E+00 | 6.17E-01 | 6.38E-02 | RAD50;HERC2;RFC1;RAD17;SPIDR |
| 1008 | Hedgehog 'off' state Homo sapiens R-HSA-5610787 | 9.04E-01 | 1.00E+00 | 5.87E-01 | 5.94E-02 | ADCY9;PSME1;PSMB10;PSMB9 |
| 1009 | HSF1-dependent transactivation Homo sapiens R-HSA-3371571 | 9.05E-01 | 1.00E+00 | 4.23E-01 | 4.24E-02 | HSPA2 |
| 1010 | Negative regulation of FGFR2 signaling Homo sapiens R-HSA-5654727 | 9.05E-01 | 1.00E+00 | 4.23E-01 | 4.24E-02 | FGF2 |
| 1011 | Regulation of APC/C activators between G1/S and early anaphase Homo sapiens R-HSA-176408 | 9.05E-01 | 1.00E+00 | 5.51E-01 | 5.51E-02 | PSME1;PSMB10;PSMB9 |
| 1012 | Regulation of TP53 Degradation Homo sapiens R-HSA-6804757 | 9.11E-01 | 1.00E+00 | 4.11E-01 | 3.83E-02 | PRR5 |

| | | | | | | |
|---|---|---|---|---|---|---|
| 1013 | SUMOylation Homo sapiens R-HSA-2990846 | 9.12E-01 | 1.00E+00 | 5.75E-01 | 5.31E-02 | NUP93;SP100;HERC2;NUP50 |
| 1014 | Processing of DNA double-strand break ends Homo sapiens R-HSA-5693607 | 9.14E-01 | 1.00E+00 | 5.37E-01 | 4.85E-02 | RAD50;HERC2;RAD17 |
| 1015 | Disease Homo sapiens R-HSA-1643685 | 9.14E-01 | 1.00E+00 | 8.11E-01 | 7.27E-02 | KLB;PRR5;HDAC1;PIK3R3;RPS4Y1;FGF2;EGFR;PSMB10;ERLEC1;MUC1;GPC3;THSD7A;TBL1X;PAK2;ADAMTS6;SBSPON;TGFB1;C1GALT1;MUC19;GAB1;ISG15;TGFBR1;DCN;PSMB9;TGFBR2;NUP93;DNAJC3;NFKBIA;PC;LCK;NUP50;PSME1;NMT1;NAPEPLD;CD9;FMOD;AGRN;CALM1;TLR5;FGFR1 |
| 1016 | MAP kinase activation in TLR cascade Homo sapiens R-HSA-450294 | 9.16E-01 | 1.00E+00 | 4.81E-01 | 4.20E-02 | MAPK10;TNIP2 |
| 1017 | Role of phospholipids in phagocytosis Homo sapiens R-HSA-2029485 | 9.16E-01 | 1.00E+00 | 4.81E-01 | 4.20E-02 | SYK;ITPR2 |
| 1018 | Regulation of TP53 Expression and Degradation Homo sapiens R-HSA-6806003 | 9.17E-01 | 1.00E+00 | 3.99E-01 | 3.46E-02 | PRR5 |
| 1019 | Class I MHC mediated antigen processing & presentation Homo sapiens R-HSA-983169 | 9.17E-01 | 1.00E+00 | 7.20E-01 | 6.21E-02 | ITGB5;ERAP2;RNF14;UBR4;TAP1;CTSS;PSMB10;HLA-E;PSMB9;UFL1;ZNRF1;HERC2;RNF213;PSME1;ITGAV |
| 1020 | TCF dependent signaling in response to WNT Homo sapiens R-HSA-201681 | 9.21E-01 | 1.00E+00 | 6.60E-01 | 5.44E-02 | H2AFZ;HDAC1;CSNK2A2;WNT5A;USP34;PSME1;HIST1H2AC;PSMB10;PSMB9 |
| 1021 | TRAF6 mediated induction of NFkB and MAP kinases upon TLR7/8 or 9 activation Homo sapiens R-HSA-975138 | 9.22E-01 | 1.00E+00 | 5.23E-01 | 4.27E-02 | NFKBIA;MAPK10;TNIP2 |

| | | | | | | |
|---|---|---|---|---|---|---|
| 1022 | Activation of ATR in response to replication stress Homo sapiens R-HSA-176187 | 9.23E-01 | 1.00E+00 | 3.88E-01 | 3.13E-02 | RAD17 |
| 1023 | APC/C-mediated degradation of cell cycle proteins Homo sapiens R-HSA-174143 | 9.29E-01 | 1.00E+00 | 5.10E-01 | 3.76E-02 | PSME1;PSMB10;PSMB9 |
| 1024 | Regulation of mitotic cell cycle Homo sapiens R-HSA-453276 | 9.29E-01 | 1.00E+00 | 5.10E-01 | 3.76E-02 | PSME1;PSMB10;PSMB9 |
| 1025 | MyD88 dependent cascade initiated on endosome Homo sapiens R-HSA-975155 | 9.29E-01 | 1.00E+00 | 5.10E-01 | 3.76E-02 | NFKBIA;MAPK10;TNIP2 |
| 1026 | Toll Like Receptor 7/8 (TLR7/8) Cascade Homo sapiens R-HSA-168181 | 9.29E-01 | 1.00E+00 | 5.10E-01 | 3.76E-02 | NFKBIA;MAPK10;TNIP2 |
| 1027 | Dual incision in TC-NER Homo sapiens R-HSA-6782135 | 9.29E-01 | 1.00E+00 | 4.58E-01 | 3.35E-02 | RFC1;ERCC5 |
| 1028 | Mitotic Prometaphase Homo sapiens R-HSA-68877 | 9.33E-01 | 1.00E+00 | 5.42E-01 | 3.78E-02 | CLIP1;CSNK2A2;SMC4;CENPC |
| 1029 | mTOR signalling Homo sapiens R-HSA-165159 | 9.33E-01 | 1.00E+00 | 3.67E-01 | 2.56E-02 | CAB39L |
| 1030 | Association of TriC/CCT with target proteins during biosynthesis Homo sapiens R-HSA-390471 | 9.33E-01 | 1.00E+00 | 3.67E-01 | 2.56E-02 | CCT5 |
| 1031 | Influenza Infection Homo sapiens R-HSA-168254 | 9.33E-01 | 1.00E+00 | 5.93E-01 | 4.12E-02 | NUP93;DNAJC3;TGFB1;NUP50;ISG15;RPS4Y1 |
| 1032 | PKB-mediated events Homo sapiens R-HSA-109703 | 9.37E-01 | 1.00E+00 | 3.58E-01 | 2.33E-02 | CAB39L |

| | | | | | | |
|---|---|---|---|---|---|---|
| 1033 | Class C/3 (Metabotropic glutamate/pheromone receptors) Homo sapiens R-HSA-420499 | 9.37E-01 | 1.00E+00 | 3.58E-01 | 2.33E-02 | GABBR1 |
| 1034 | Chromatin organization Homo sapiens R-HSA-4839726 | 9.40E-01 | 1.00E+00 | 6.45E-01 | 3.97E-02 | KDM5B;SETD3;KDM1A;H2AFZ;HDAC1;KMT2C;EHMT1;TBL1X;ELP5;HIST1H2AC |
| 1035 | Chromatin modifying enzymes Homo sapiens R-HSA-3247509 | 9.40E-01 | 1.00E+00 | 6.45E-01 | 3.97E-02 | KDM5B;SETD3;KDM1A;H2AFZ;HDAC1;KMT2C;EHMT1;TBL1X;ELP5;HIST1H2AC |
| 1036 | G2/M Checkpoints Homo sapiens R-HSA-69481 | 9.40E-01 | 1.00E+00 | 5.81E-01 | 3.57E-02 | RAD50;HERC2;PSME1;RAD17;PSMB10;PSMB9 |
| 1037 | Metabolism of amino acids and derivatives Homo sapiens R-HSA-71291 | 9.41E-01 | 1.00E+00 | 6.98E-01 | 4.24E-02 | ARG2;HGD;GPT2;APIP;SLC6A12;MAT1A;RPS4Y1;PSMB10;PSMB9;SRM;FOLH1;AFMID;PSME1;ALDH18A1;GLUL;ASPA |
| 1038 | Glycosphingolipid metabolism Homo sapiens R-HSA-1660662 | 9.41E-01 | 1.00E+00 | 3.49E-01 | 2.11E-02 | GALC |
| 1039 | Pre-NOTCH Transcription and Translation Homo sapiens R-HSA-1912408 | 9.41E-01 | 1.00E+00 | 3.49E-01 | 2.11E-02 | AGO3 |
| 1040 | TNF signaling Homo sapiens R-HSA-75893 | 9.41E-01 | 1.00E+00 | 3.49E-01 | 2.11E-02 | TNFRSF1A |
| 1041 | Mitochondrial translation Homo sapiens R-HSA-5368287 | 9.45E-01 | 1.00E+00 | 4.81E-01 | 2.74E-02 | GFM1;MTIF2;MRPS6 |
| 1042 | Intrinsic Pathway for Apoptosis Homo sapiens R-HSA-109606 | 9.45E-01 | 1.00E+00 | 3.41E-01 | 1.92E-02 | NMT1 |
| 1043 | Transcriptional regulation of pluripotent stem cells Homo sapiens R-HSA-452723 | 9.45E-01 | 1.00E+00 | 3.41E-01 | 1.92E-02 | FGF2 |

| | | | | | | |
|---|---|---|---|---|---|---|
| 1044 | Deactivation of the beta-catenin transactivating complex Homo sapiens R-HSA-3769402 | 9.45E-01 | 1.00E+00 | 3.41E-01 | 1.92E-02 | HDAC1 |
| 1045 | Netrin-1 signaling Homo sapiens R-HSA-373752 | 9.45E-01 | 1.00E+00 | 3.41E-01 | 1.92E-02 | NTN4 |
| 1046 | Activated TLR4 signalling Homo sapiens R-HSA-166054 | 9.46E-01 | 1.00E+00 | 5.16E-01 | 2.85E-02 | NFKBIA;MAPK10;IRF3;TNIP2 |
| 1047 | COPII (Coat Protein 2) Mediated Vesicle Transport Homo sapiens R-HSA-204005 | 9.47E-01 | 1.00E+00 | 4.23E-01 | 2.31E-02 | TRAPPC10;CTSZ |
| 1048 | Transport of Mature mRNA derived from an Intron-Containing Transcript Homo sapiens R-HSA-159236 | 9.47E-01 | 1.00E+00 | 4.23E-01 | 2.31E-02 | NUP93;NUP50 |
| 1049 | The citric acid (TCA) cycle and respiratory electron transport Homo sapiens R-HSA-1428517 | 9.47E-01 | 1.00E+00 | 5.69E-01 | 3.09E-02 | LDHA;UCP2;PDK4;HAGH;SLC16A3;PDK1 |
| 1050 | Formation of Incision Complex in GG-NER Homo sapiens R-HSA-5696395 | 9.49E-01 | 1.00E+00 | 3.32E-01 | 1.74E-02 | ERCC5 |
| 1051 | Amine ligand-binding receptors Homo sapiens R-HSA-375280 | 9.49E-01 | 1.00E+00 | 3.32E-01 | 1.74E-02 | ADRA2C |
| 1052 | MyD88:Mal cascade initiated on plasma membrane Homo sapiens R-HSA-166058 | 9.50E-01 | 1.00E+00 | 4.70E-01 | 2.41E-02 | NFKBIA;MAPK10;TNIP2 |

| | | | | | | |
|---|---|---|---|---|---|---|
| 1053 | Toll Like Receptor TLR1:TLR2 Cascade Homo sapiens R-HSA-168179 | 9.50E-01 | 1.00E+00 | 4.70E-01 | 2.41E-02 | NFKBIA;MAPK10;TNIP2 |
| 1054 | Toll Like Receptor TLR6:TLR2 Cascade Homo sapiens R-HSA-168188 | 9.50E-01 | 1.00E+00 | 4.70E-01 | 2.41E-02 | NFKBIA;MAPK10;TNIP2 |
| 1055 | Toll Like Receptor 2 (TLR2) Cascade Homo sapiens R-HSA-181438 | 9.50E-01 | 1.00E+00 | 4.70E-01 | 2.41E-02 | NFKBIA;MAPK10;TNIP2 |
| 1056 | Class A/1 (Rhodopsin-like receptors) Homo sapiens R-HSA-373076 | 9.50E-01 | 1.00E+00 | 6.77E-01 | 3.47E-02 | NMB;GRP;NPFF;F2R;NTS;ADRA2C;SSTR2;EDNRB;CYSLTR2;SST;GPER1;ACKR4;F2RL1;CCL2;ACKR3 |
| 1057 | Phase 0 - rapid depolarisation Homo sapiens R-HSA-5576892 | 9.52E-01 | 1.00E+00 | 3.25E-01 | 1.59E-02 | CALM1 |
| 1058 | Diseases of metabolism Homo sapiens R-HSA-5668914 | 9.55E-01 | 1.00E+00 | 3.17E-01 | 1.44E-02 | PC |
| 1059 | Chaperonin-mediated protein folding Homo sapiens R-HSA-390466 | 9.57E-01 | 1.00E+00 | 4.55E-01 | 1.99E-02 | TUBA1C;CSNK2A2;CCT5 |
| 1060 | Scavenging of heme from plasma Homo sapiens R-HSA-2168880 | 9.58E-01 | 1.00E+00 | 3.10E-01 | 1.32E-02 | APOL1 |
| 1061 | RNA Polymerase I Transcription Initiation Homo sapiens R-HSA-73762 | 9.61E-01 | 1.00E+00 | 3.03E-01 | 1.20E-02 | HDAC1 |
| 1062 | Transport of Mature Transcript to Cytoplasm Homo sapiens R-HSA-72202 | 9.62E-01 | 1.00E+00 | 3.87E-01 | 1.49E-02 | NUP93;NUP50 |
| 1063 | G beta:gamma signalling through PI3Kgamma Homo sapiens R-HSA-392451 | 9.64E-01 | 1.00E+00 | 2.97E-01 | 1.09E-02 | PIK3R3 |

| | | | | | | |
|---|---|---|---|---|---|---|
| 1064 | Toll Like Receptor 4 (TLR4) Cascade Homo sapiens R-HSA-166016 | 9.66E-01 | 1.00E+00 | 4.72E-01 | 1.62E-02 | NFKBIA;MAPK10;IRF3;TNIP2 |
| 1065 | Transcription-Coupled Nucleotide Excision Repair (TC-NER) Homo sapiens R-HSA-6781827 | 9.66E-01 | 1.00E+00 | 3.77E-01 | 1.29E-02 | RFC1;ERCC5 |
| 1066 | Phase II conjugation Homo sapiens R-HSA-156580 | 9.67E-01 | 1.00E+00 | 4.31E-01 | 1.45E-02 | SLC35B3;UGT2B11;MAT1A |
| 1067 | RNA polymerase II transcribes snRNA genes Homo sapiens R-HSA-6807505 | 9.68E-01 | 1.00E+00 | 3.72E-01 | 1.20E-02 | SNAPC1;PHAX |
| 1068 | Protein folding Homo sapiens R-HSA-391251 | 9.69E-01 | 1.00E+00 | 4.27E-01 | 1.36E-02 | TUBA1C;CSNK2A2;CCT5 |
| 1069 | Complement cascade Homo sapiens R-HSA-166658 | 9.73E-01 | 1.00E+00 | 3.58E-01 | 9.62E-03 | CLU;CFB |
| 1070 | Influenza Viral RNA Transcription and Replication Homo sapiens R-HSA-168273 | 9.75E-01 | 1.00E+00 | 4.49E-01 | 1.15E-02 | NUP93;DNAJC3;NUP50;RPS4Y1 |
| 1071 | Nonsense-Mediated Decay (NMD) Homo sapiens R-HSA-927802 | 9.76E-01 | 1.00E+00 | 4.06E-01 | 9.95E-03 | SMG1;ETF1;RPS4Y1 |
| 1072 | Nonsense Mediated Decay (NMD) enhanced by the Exon Junction Complex (EJC) Homo sapiens R-HSA-975957 | 9.76E-01 | 1.00E+00 | 4.06E-01 | 9.95E-03 | SMG1;ETF1;RPS4Y1 |

| | | | | | | |
|---|---|---|---|---|---|---|
| 1073 | L13a-mediated translational silencing of Ceruloplasmin expression Homo sapiens R-HSA-156827 | 9.76E-01 | 1.00E+00 | 4.06E-01 | 9.95E-03 | RPS4Y1;EIF2S2;EIF3C |
| 1074 | 3' -UTR-mediated translational regulation Homo sapiens R-HSA-157279 | 9.76E-01 | 1.00E+00 | 4.06E-01 | 9.95E-03 | RPS4Y1;EIF2S2;EIF3C |
| 1075 | Mitochondrial protein import Homo sapiens R-HSA-1268020 | 9.76E-01 | 1.00E+00 | 2.63E-01 | 6.36E-03 | COX17 |
| 1076 | Translation Homo sapiens R-HSA-72766 | 9.76E-01 | 1.00E+00 | 4.77E-01 | 1.15E-02 | ETF1;RPS4Y1;EIF2S2;EIF3C;SEC11C |
| 1077 | Global Genome Nucleotide Excision Repair (GG-NER) Homo sapiens R-HSA-5696399 | 9.76E-01 | 1.00E+00 | 3.49E-01 | 8.33E-03 | RFC1;ERCC5 |
| 1078 | GTP hydrolysis and joining of the 60S ribosomal subunit Homo sapiens R-HSA-72706 | 9.77E-01 | 1.00E+00 | 4.02E-01 | 9.34E-03 | RPS4Y1;EIF2S2;EIF3C |
| 1079 | Viral mRNA Translation Homo sapiens R-HSA-192823 | 9.79E-01 | 1.00E+00 | 3.40E-01 | 7.21E-03 | DNAJC3;RPS4Y1 |
| 1080 | Mitochondrial translation initiation Homo sapiens R-HSA-5368286 | 9.79E-01 | 1.00E+00 | 3.40E-01 | 7.21E-03 | MTIF2;MRPS6 |
| 1081 | Mitochondrial translation elongation Homo sapiens R-HSA-5389840 | 9.79E-01 | 1.00E+00 | 3.40E-01 | 7.21E-03 | GFM1;MRPS6 |
| 1082 | GPCR ligand binding Homo sapiens R-HSA-500792 | 9.81E-01 | 1.00E+00 | 6.50E-01 | 1.24E-02 | NMB;GABBR1;CALCA;GRP;GIPR;NPFF;F2R;WNT5A;ADRA2C;NTS;SSTR2;CYSLTR2;EDNRB;WNT11;SST;GPER1;ACKR4;CCL2;F2RL1;ACKR3 |

| | | | | | | |
|---|---|---|---|---|---|---|
| 1083 | G alpha (i) signalling events Homo sapiens R-HSA-418594 | 9.82E-01 | 1.00E+00 | 5.42E-01 | 9.91E-03 | RGS5;GABBR1;ADCY9;RGS19;SST;GPER1;ACKR3;ADRA2C;SSTR2 |
| 1084 | Initial triggering of complement Homo sapiens R-HSA-166663 | 9.82E-01 | 1.00E+00 | 2.45E-01 | 4.47E-03 | CFB |
| 1085 | Eukaryotic Translation Termination Homo sapiens R-HSA-72764 | 9.82E-01 | 1.00E+00 | 3.28E-01 | 5.81E-03 | ETF1;RPS4Y1 |
| 1086 | Influenza Life Cycle Homo sapiens R-HSA-168255 | 9.83E-01 | 1.00E+00 | 4.22E-01 | 7.24E-03 | NUP93;DNAJC3;NUP50;RPS4Y1 |
| 1087 | HIV Infection Homo sapiens R-HSA-162906 | 9.83E-01 | 1.00E+00 | 5.20E-01 | 8.71E-03 | NUP93;LCK;NUP50;PSME1;NMT1;PAK2;PSMB10;PSMB9 |
| 1088 | Cap-dependent Translation Initiation Homo sapiens R-HSA-72737 | 9.84E-01 | 1.00E+00 | 3.76E-01 | 5.99E-03 | RPS4Y1;EIF2S2;EIF3C |
| 1089 | Eukaryotic Translation Initiation Homo sapiens R-HSA-72613 | 9.84E-01 | 1.00E+00 | 3.76E-01 | 5.99E-03 | RPS4Y1;EIF2S2;EIF3C |
| 1090 | Nonsense Mediated Decay (NMD) independent of the Exon Junction Complex (EJC) Homo sapiens R-HSA-975956 | 9.84E-01 | 1.00E+00 | 3.20E-01 | 5.03E-03 | ETF1;RPS4Y1 |
| 1091 | Cell Cycle Checkpoints Homo sapiens R-HSA-69620 | 9.85E-01 | 1.00E+00 | 4.74E-01 | 7.40E-03 | RAD50;HERC2;PSME1;RAD17;PSMB10;PSMB9 |
| 1092 | TP53 Regulates Transcription of DNA Repair Genes Homo sapiens R-HSA-6796648 | 9.85E-01 | 1.00E+00 | 2.32E-01 | 3.44E-03 | FANCC |

| | | | | | | |
|---|---|---|---|---|---|---|
| 1093 | Generic Transcription Pathway Homo sapiens R-HSA-212436 | 9.86E-01 | 1.00E+00 | 7.16E-01 | 1.04E-02 | PRR5;ZNF331;ZNF670;THRA;CHD9;HDAC1;EHMT1;RORA;SESN3;TGS1;PRDX1;TNKS1BP1;NRBF2;TBL1X;BRD7;SKIL;BNIP3L;WWTR1;GPX2;ZNF680;ZNF141;VDR;CSNK2A2;NR1H4;FANCC;NR1D1;ESR2;HIPK2;NR4A2;RAD50;RGCC;NR4A3;AGO3;BCL6;ZNF519;DDIT4;RAD17;PPARG;ZNF138;ZNF433 |
| 1094 | Separation of Sister Chromatids Homo sapiens R-HSA-2467813 | 9.86E-01 | 1.00E+00 | 4.43E-01 | 6.38E-03 | CLIP1;PSME1;PSMB10;CENPC;PSMB9 |
| 1095 | Gap-filling DNA repair synthesis and ligation in TC-NER Homo sapiens R-HSA-6782210 | 9.86E-01 | 1.00E+00 | 2.29E-01 | 3.16E-03 | RFC1 |
| 1096 | G alpha (s) signalling events Homo sapiens R-HSA-418555 | 9.87E-01 | 1.00E+00 | 4.03E-01 | 5.11E-03 | CALCA;ADCY9;PDE1B;GIPR |
| 1097 | Infectious disease Homo sapiens R-HSA-5663205 | 9.87E-01 | 1.00E+00 | 5.82E-01 | 7.34E-03 | TGFB1;ISG15;RPS4Y1;PSMB10;PSMB9;NUP93;DNAJC3;LCK;NUP50;PSME1;NMT1;CD9;CALM1;PAK2 |
| 1098 | Major pathway of rRNA processing in the nucleolus Homo sapiens R-HSA-6791226 | 9.88E-01 | 1.00E+00 | 4.32E-01 | 5.15E-03 | NOP14;WDR75;DDX52;RPS4Y1;UTP18 |
| 1099 | Formation of a pool of free 40S subunits Homo sapiens R-HSA-72689 | 9.90E-01 | 1.00E+00 | 2.96E-01 | 3.05E-03 | RPS4Y1;EIF3C |
| 1100 | Macroautophagy Homo sapiens R-HSA-1632852 | 9.90E-01 | 1.00E+00 | 2.11E-01 | 2.06E-03 | ATG101 |
| 1101 | Resolution of Sister Chromatid Cohesion Homo sapiens R-HSA-2500257 | 9.91E-01 | 1.00E+00 | 2.87E-01 | 2.47E-03 | CLIP1;CENPC |
| 1102 | Antigen processing: Ubiquitination & Proteasome degradation Homo sapiens R-HSA-983168 | 9.92E-01 | 1.00E+00 | 4.98E-01 | 4.10E-03 | UFL1;ZNRF1;HERC2;RNF213;RNF14;PSME1;UBR4;PSMB10;PSMB9 |

| | | | | | | |
|---|---|---|---|---|---|---|
| 1103 | Late Phase of HIV Life Cycle Homo sapiens R-HSA-162599 | 9.93E-01 | 1.00E+00 | 3.34E-01 | 2.46E-03 | NUP93;NUP50;NMT1 |
| 1104 | rRNA processing Homo sapiens R-HSA-72312 | 9.94E-01 | 1.00E+00 | 3.97E-01 | 2.40E-03 | NOP14;WDR75;DDX52;RPS4Y1;UTP18 |
| 1105 | Binding and Uptake of Ligands by Scavenger Receptors Homo sapiens R-HSA-2173782 | 9.94E-01 | 1.00E+00 | 1.91E-01 | 1.14E-03 | APOL1 |
| 1106 | SRP-dependent cotranslational protein targeting to membrane Homo sapiens R-HSA-1799339 | 9.95E-01 | 1.00E+00 | 2.65E-01 | 1.40E-03 | RPS4Y1;SEC11C |
| 1107 | HATs acetylate histones Homo sapiens R-HSA-3214847 | 9.95E-01 | 1.00E+00 | 2.65E-01 | 1.40E-03 | ELP5;HIST1H2AC |
| 1108 | Nucleotide Excision Repair Homo sapiens R-HSA-5696398 | 9.95E-01 | 1.00E+00 | 2.63E-01 | 1.30E-03 | RFC1;ERCC5 |
| 1109 | Selenoamino acid metabolism Homo sapiens R-HSA-2408522 | 9.96E-01 | 1.00E+00 | 2.55E-01 | 1.05E-03 | MAT1A;RPS4Y1 |
| 1110 | HIV Life Cycle Homo sapiens R-HSA-162587 | 9.96E-01 | 1.00E+00 | 3.02E-01 | 1.07E-03 | NUP93;NUP50;NMT1 |
| 1111 | Peptide chain elongation Homo sapiens R-HSA-156902 | 9.97E-01 | 1.00E+00 | 1.68E-01 | 5.02E-04 | RPS4Y1 |
| 1112 | Mitochondrial translation termination Homo sapiens R-HSA-5419276 | 9.97E-01 | 1.00E+00 | 1.68E-01 | 5.02E-04 | MRPS6 |
| 1113 | Selenocysteine synthesis Homo sapiens R-HSA-2408557 | 9.98E-01 | 1.00E+00 | 1.62E-01 | 3.93E-04 | RPS4Y1 |

| | | | | | | |
|---|---|---|---|---|---|---|
| 1114 | Signal Transduction Homo sapiens R-HSA-162582 | 9.98E-01 | 1.00E+00 | 7.75E-01 | 1.81E-03 | KLB;KDM1A;PEBP1;FGF2;RGS5;CYSLTR2;EDNRB;OPHN1;KIF5B;ARHGDIB;TNFSF10;PDK4;HIST1H2AC;PDK1;NPFF;CSNK2A2;F2R;WNT5A;CLIP1;ZFYVE16;ADCY9;COL4A4;AKAP9;COL4A3;PSME1;RAF1;GRB7;CALCA;GIPR;PDE1B;PIK3R3;ITPR2;NTS;DTX4;HIF1A;RAP1B;RDH11;FLRT3;RDH10;ATP6V0A2;THEM4;ATP6V0A4;RDH13;RHPN2;SRGAP3;ECT2;SKIL;MYH10;MARK3;SPTBN1;WWTR1;NMB;YES1;TGFB1;CGN;NFKBIA;DIAPH1;FER;AGO3;ARPC2;NAPEPLD;NMT1;CALM1;LRP10;FGFR1;BMPR1A;ITGB1;PRR5;CAB39L;ARPC1B;USP34;ATP2A3;SHB;AATF;ARHGAP6;GPER1;MCF2L;CYP4V2;BOC;CNGA1;ITGAV;SPTAN1;TMED5;SYK;DUSP1;H2AFZ;ARHGEF16;GAB1;PLA2G4A;ARAP1;SSTR2;ADRA2C;TGFBR1;TNFRSF1A;TGFBR2;ALDH1A3;EVC;CDC42EP5;LCK;RHOJ;PLCB2;USP15;RGS19;GRP;HDAC1;LRP8;EGFR;PSMB10;CENPC;ERLEC1;WNT11;GPC3;CCL2;TBL1X;PAK2;EVC2;GABBR1;GALNT3;KIDINS220;AKR1C1;AKR1C3;BAIAP2;PSMB9;KTN1;FMNL3;FMNL2;SST;ACKR4;TRIP10;ACKR3;F2RL1;AGRN;ALDH8A1 |
| 1115 | Eukaryotic Translation Elongation Homo sapiens R-HSA-156842 | 9.98E-01 | 1.00E+00 | 1.58E-01 | 3.34E-04 | RPS4Y1 |
| 1116 | Immunoregulatory interactions between a Lymphoid and a non-Lymphoid cell Homo sapiens R-HSA-198933 | 9.99E-01 | 1.00E+00 | 2.71E-01 | 3.85E-04 | ITGB1;IFITM1;HLA-E |
| 1117 | mRNA Splicing - Major Pathway Homo sapiens R-HSA-72163 | 9.99E-01 | 1.00E+00 | 2.11E-01 | 2.08E-04 | DHX16;HNRNPR |
| 1118 | Processing of Capped Intron-Containing Pre-mRNA Homo sapiens R-HSA-72203 | 9.99E-01 | 1.00E+00 | 2.94E-01 | 2.49E-04 | NUP93;NUP50;DHX16;HNRNPR |

| | | | | | | |
|---|---|---|---|---|---|---|
| 1119 | Respiratory electron transport, ATP synthesis by chemiosmotic coupling, and heat production by uncoupling proteins. Homo sapiens R-HSA-163200 | 9.99E-01 | 1.00E+00 | 1.29E-01 | 6.81E-05 | UCP2 |
| 1120 | mRNA Splicing Homo sapiens R-HSA-72172 | 9.99E-01 | 1.00E+00 | 1.96E-01 | 1.03E-04 | DHX16;HNRNPR |
| 1121 | Gene Expression Homo sapiens R-HSA-74160 | 1.00E+00 | 1.00E+00 | 6.45E-01 | 4.32E-05 | PRR5;ZNF331;SMG1;CRCP;THRA;CHD9;EHMT1;RORA;HNRNPR;PHAX;C2ORF49;SESN3;SNAPC1;DHX16;TNKS1BP1;TRMT44;HIST1H2AC;BNIP3L;GPX2;ZNF680;H2AFZ;CSNK2A2;WDR75;DDX52;BAZ1B;UTP18;NUP93;ZNF519;NUP50;DDIT4;PSME1;PPARG;ETF1;GARS;BDP1;ZNF433;DCP2;ZNF670;DNMT1;HDAC1;RPS4Y1;PSMB10;TGS1;PRDX1;TYW1;IARS2;NRBF2;TBL1X;BRD7;SKIL;SEC11C;WWTR1;NOP14;ZNF141;VDR;MBD2;NR1H4;FANCC;NR1D1;TDRD6;EIF2S2;ESR2;PSMB9;HIPK2;NR4A2;RAD50;RGCC;NR4A3;AGO3;BCL6;TDRD12;RAD17;ZNF138;EIF3C |
| 1122 | Signaling by GPCR Homo sapiens R-HSA-372790 | 1.00E+00 | 1.00E+00 | 5.10E-01 | 3.42E-06 | KLB;CALCA;RGS19;GRP;GIPR;PDE1B;PIK3R3;PEBP1;ITPR2;NTS;FGF2;EGFR;PSMB10;RAP1B;RGS5;CYSLTR2;EDNRB;WNT11;GPER1;MCF2L;CCL2;ECT2;SPTAN1;MARK3;SPTBN1;GABBR1;NMB;DUSP1;NPFF;ARHGEF16;F2R;WNT5A;PLA2G4A;SSTR2;ADRA2C;PSMB9;ADCY9;SST;AKAP9;PSME1;ACKR4;ACKR3;F2RL1;CALM1;RAF1;PLCB2;FGFR1 |
| 1123 | GPCR downstream signaling Homo sapiens R-HSA-388396 | 1.00E+00 | 1.00E+00 | 3.68E-01 | 2.17E-06 | CALCA;RGS19;GRP;GIPR;PDE1B;PIK3R3;ITPR2;NTS;RGS5;CYSLTR2;EDNRB;GPER1;MCF2L;ECT2;GABBR1;NMB;NPFF;ARHGEF16;F2R;SSTR2;ADRA2C;ADCY9;SST;ACKR3;F2RL1;PLCB2 |

**Pathways enriched by genes enhanced with SGLT2 inhibitor in principal cells**

| | Term | P value | Adjusted P value | Odds Ratio | Combined Score | Genes |
|---|---|---|---|---|---|---|
| 1 | Eukaryotic Translation Elongation Homo sapiens R-HSA-156842 | 1.04E-26 | 3.81E-24 | 1.64E+01 | 9.80E+02 | RPL4;RPL32;RPL10;RPL34;RPL12;RPL11;RPL36A;RPL9;RPL6;RPL7;RPS4X;RPS14;RPS17;RPS3;RPL38;RPL15;RPS11;RPL39;RPL41;RPL21;RPL23;RPL22;RPL13A;RPL35A;RPS3A;RPS26;RPS28;EEF1D;RPS29;RPL27A;RPL37A;RPL24;RPL27;RPS20;RPS24;RPS23 |
| 2 | Nonsense Mediated Decay (NMD) independent of the Exon Junction Complex (EJC) Homo sapiens R-HSA-975956 | 1.04E-26 | 3.81E-24 | 1.64E+01 | 9.80E+02 | RPL4;RPL32;RPL10;RPL34;RPL12;RPL11;RPL36A;RPL9;RPL6;RPL7;RPS4X;RPS14;RPS17;RPS3;RPL38;RPL15;RPS11;RPL39;RPL41;RPL21;RPL23;RPL22;RPL13A;RPL35A;RPS3A;RPS26;RPS28;RPS29;RPL27A;RPL37A;RPL24;RPL27;RPS20;PABPC1;RPS24;RPS23 |
| 3 | Viral mRNA Translation Homo sapiens R-HSA-192823 | 1.56E-26 | 3.81E-24 | 1.72E+01 | 1.02E+03 | RPL4;RPL32;RPL10;RPL34;RPL12;RPL11;RPL36A;RPL9;RPL6;RPL7;RPS4X;RPS14;RPS17;RPS3;RPL38;RPL15;RPS11;RPL39;RPL41;RPL21;RPL23;RPL22;RPL13A;RPL35A;RPS3A;RPS26;RPS28;RPS29;RPL27A;RPL37A;RPL24;RPL27;RPS20;RPS24;RPS23 |
| 4 | Peptide chain elongation Homo sapiens R-HSA-156902 | 1.56E-26 | 3.81E-24 | 1.72E+01 | 1.02E+03 | RPL4;RPL32;RPL10;RPL34;RPL12;RPL11;RPL36A;RPL9;RPL6;RPL7;RPS4X;RPS14;RPS17;RPS3;RPL38;RPL15;RPS11;RPL39;RPL41;RPL21;RPL23;RPL22;RPL13A;RPL35A;RPS3A;RPS26;RPS28;RPS29;RPL27A;RPL37A;RPL24;RPL27;RPS20;RPS24;RPS23 |
| 5 | Selenocysteine synthesis Homo sapiens R-HSA-2408557 | 6.63E-26 | 9.48E-24 | 1.62E+01 | 9.40E+02 | RPL4;RPL32;RPL10;RPL34;RPL12;RPL11;RPL36A;RPL9;RPL6;RPL7;RPS4X;RPS14;RPS17;RPS3;RPL38;RPL15;RPS11;RPL39;RPL41;RPL21;RPL23;RPL22;RPL13A;RPL35A;RPS3A;RPS26;RPS28;RPS29;RPL27A;RPL37A;RPL24;RPL27;RPS20;RPS24;RPS23 |
| 6 | Eukaryotic Translation Termination Homo sapiens R-HSA-72764 | 6.63E-26 | 9.48E-24 | 1.62E+01 | 9.40E+02 | RPL4;RPL32;RPL10;RPL34;RPL12;RPL11;RPL36A;RPL9;RPL6;RPL7;RPS4X;RPS14;RPS17;RPS3;RPL38;RPL15;RPS11;RPL39;RPL41;RPL21;RPL23;RPL22;RPL13A;RPL35A;RPS3A;RPS26;RPS28;RPS29;RPL27A;RPL37A;RPL24;RPL27;RPS20;RPS24;RPS23 |
| 7 | L13a-mediated translational silencing of Ceruloplasmin expression Homo sapiens R-HSA-156827 | 7.75E-26 | 9.48E-24 | 1.35E+01 | 7.80E+02 | RPL4;RPL32;RPL10;RPL34;RPL12;RPL11;RPL36A;RPL9;RPL6;RPL7;RPS4X;RPS14;RPS17;RPS3;RPL38;RPL15;RPS11;RPL39;RPL41;RPL21;EIF1AX;RPL23;RPL22;RPL13A;RPL35A;RPS3A;RPS26;RPS28;EIF2S3;RPS29;RPL27A;RPL37A;RPL24;RPL27;RPS20;PABPC1;RPS24;RPS23 |

| | | | | | | |
|---|---|---|---|---|---|---|
| 8 | 3' -UTR-mediated translational regulation Homo sapiens R-HSA-157279 | 7.75E-26 | 9.48E-24 | 1.35E+01 | 7.80E+02 | RPL4;RPL32;RPL10;RPL34;RPL12;RPL11;RPL36A;RPL9;RPL6;RPL7;RPS4X;RPS14;RPS17;RPS3;RPL38;RPL15;RPS11;RPL39;RPL41;RPL21;EIF1AX;RPL23;RPL22;RPL13A;RPL35A;RPS3A;RPS26;RPS28;EIF2S3;RPS29;RPL27A;RPL37A;RPL24;RPL27;RPS20;PABPC1;RPS24;RPS23 |
| 9 | Formation of a pool of free 40S subunits Homo sapiens R-HSA-72689 | 2.44E-25 | 2.65E-23 | 1.45E+01 | 8.19E+02 | RPL4;RPL32;RPL10;RPL34;RPL12;RPL11;RPL36A;RPL9;RPL6;RPL7;RPS4X;RPS14;RPS17;RPS3;RPL38;RPL15;RPS11;RPL39;RPL41;RPL21;EIF1AX;RPL23;RPL22;RPL13A;RPL35A;RPS3A;RPS26;RPS28;RPS29;RPL27A;RPL37A;RPL24;RPL27;RPS20;RPS24;RPS23 |
| 10 | GTP hydrolysis and joining of the 60S ribosomal subunit Homo sapiens R-HSA-72706 | 1.52E-24 | 1.35E-22 | 1.27E+01 | 6.99E+02 | RPL4;RPL32;RPL10;RPL34;RPL12;RPL11;RPL36A;RPL9;RPL6;RPL7;RPS4X;RPS14;RPS17;RPS3;RPL38;RPL15;RPS11;RPL39;RPL41;RPL21;EIF1AX;RPL23;RPL22;RPL13A;RPL35A;RPS3A;RPS26;RPS28;EIF2S3;RPS29;RPL27A;RPL37A;RPL24;RPL27;RPS20;RPS24;RPS23 |
| 11 | Cap-dependent Translation Initiation Homo sapiens R-HSA-72737 | 1.65E-24 | 1.35E-22 | 1.21E+01 | 6.61E+02 | RPL4;RPL32;RPL10;RPL34;RPL12;RPL11;RPL36A;RPL9;RPL6;RPL7;RPS4X;RPS14;RPS17;RPS3;RPL38;RPL15;RPS11;RPL39;RPL41;RPL21;EIF1AX;RPL23;RPL22;RPL13A;RPL35A;RPS3A;RPS26;RPS28;EIF2S3;RPS29;RPL27A;RPL37A;RPL24;RPL27;RPS20;PABPC1;RPS24;RPS23 |
| 12 | Eukaryotic Translation Initiation Homo sapiens R-HSA-72613 | 1.65E-24 | 1.35E-22 | 1.21E+01 | 6.61E+02 | RPL4;RPL32;RPL10;RPL34;RPL12;RPL11;RPL36A;RPL9;RPL6;RPL7;RPS4X;RPS14;RPS17;RPS3;RPL38;RPL15;RPS11;RPL39;RPL41;RPL21;EIF1AX;RPL23;RPL22;RPL13A;RPL35A;RPS3A;RPS26;RPS28;EIF2S3;RPS29;RPL27A;RPL37A;RPL24;RPL27;RPS20;PABPC1;RPS24;RPS23 |
| 13 | Nonsense-Mediated Decay (NMD) Homo sapiens R-HSA-927802 | 1.32E-23 | 9.23E-22 | 1.24E+01 | 6.53E+02 | RPL4;RPL32;RPL10;RPL34;RPL12;RPL11;RPL36A;RPL9;RPL6;RPL7;RPS4X;RPS14;RPS17;RPS3;RPL38;RPL15;RPS11;RPL39;RPL41;RPL21;RPL23;RPL22;RPL13A;RPL35A;RPS3A;RPS26;RPS28;RPS29;RPL27A;RPL37A;RPL24;RPL27;RPS20;PABPC1;RPS24;RPS23 |
| 14 | Nonsense Mediated Decay (NMD) enhanced by the Exon Junction Complex (EJC) Homo sapiens R-HSA-975957 | 1.32E-23 | 9.23E-22 | 1.24E+01 | 6.53E+02 | RPL4;RPL32;RPL10;RPL34;RPL12;RPL11;RPL36A;RPL9;RPL6;RPL7;RPS4X;RPS14;RPS17;RPS3;RPL38;RPL15;RPS11;RPL39;RPL41;RPL21;RPL23;RPL22;RPL13A;RPL35A;RPS3A;RPS26;RPS28;RPS29;RPL27A;RPL37A;RPL24;RPL27;RPS20;PABPC1;RPS24;RPS23 |

| | | | | | | |
|---|---|---|---|---|---|---|
| 15 | SRP-dependent cotranslational protein targeting to membrane Homo sapiens R-HSA-1799339 | 1.91E-23 | 1.25E-21 | 1.22E+01 | 6.39E+02 | RPL4;RPL32;RPL10;RPL34;RPL12;RPL11;RPL36A;RPL9;RPL6;RPL7;RPS4X;RPS14;RPS17;SEC61G;RPS3;RPL38;RPL15;RPS11;RPL39;RPL41;RPL21;RPL23;RPL22;RPL13A;RPL35A;RPS3A;RPS26;RPS28;RPS29;RPL27A;RPL37A;RPL24;RPL27;RPS20;RPS24;RPS23 |
| 16 | Selenoamino acid metabolism Homo sapiens R-HSA-2408522 | 8.05E-23 | 4.92E-21 | 1.16E+01 | 5.88E+02 | RPL4;RPL32;RPL10;RPL34;RPL12;RPL11;RPL36A;RPL9;RPL6;PAPSS2;RPL7;RPS4X;RPS14;RPS17;RPS3;RPL38;RPL15;RPS11;RPL39;RPL41;RPL21;RPL23;RPL22;RPL13A;RPL35A;RPS3A;RPS26;RPS28;RPS29;RPL27A;RPL37A;RPL24;RPL27;RPS20;RPS24;RPS23 |
| 17 | Translation Homo sapiens R-HSA-72766 | 1.52E-21 | 8.75E-20 | 8.71E+00 | 4.17E+02 | RPL4;RPL32;RPL10;RPL34;RPL12;RPL11;RPL36A;RPL9;RPL6;RPL7;RPS4X;RPS14;RPS17;SEC61G;RPS3;RPL38;RPL15;RPS11;RPL39;RPL41;RPL21;EIF1AX;RPL23;RPL22;RPL13A;RPL35A;RPS3A;RPS26;RPS28;EIF2S3;RPS29;EEF1D;RPL27A;RPL37A;RPL24;RPL27;RPS20;PABPC1;RPS24;RPS23 |
| 18 | Influenza Viral RNA Transcription and Replication Homo sapiens R-HSA-168273 | 1.79E-20 | 9.76E-19 | 9.41E+00 | 4.28E+02 | RPL4;RPL32;RPL10;RPL34;RPL12;RPL11;RPL36A;RPL9;RPL6;RPL7;RPS4X;RPS14;RPS17;POLR2A;RPS3;RPL38;RPL15;RPS11;RPL39;RPL41;RPL21;RPL23;RPL22;RPL13A;RPL35A;RPS3A;RPS26;RPS28;RPS29;RPL27A;RPL37A;RPL24;RPL27;RPS20;RPS24;RPS23 |
| 19 | Influenza Life Cycle Homo sapiens R-HSA-168255 | 1.66E-19 | 8.53E-18 | 8.66E+00 | 3.74E+02 | RPL4;RPL32;RPL10;RPL34;RPL12;RPL11;RPL36A;RPL9;RPL6;RPL7;RPS4X;RPS14;RPS17;POLR2A;RPS3;RPL38;RPL15;RPS11;RPL39;RPL41;RPL21;RPL23;RPL22;RPL13A;RPL35A;RPS3A;RPS26;RPS28;RPS29;RPL27A;RPL37A;RPL24;RPL27;RPS20;RPS24;RPS23 |
| 20 | Influenza Infection Homo sapiens R-HSA-168254 | 2.68E-18 | 1.31E-16 | 7.80E+00 | 3.15E+02 | RPL4;RPL32;RPL10;RPL34;RPL12;RPL11;RPL36A;RPL9;RPL6;RPL7;RPS4X;RPS14;RPS17;POLR2A;RPS3;RPL38;RPL15;RPS11;RPL39;RPL41;RPL21;RPL23;RPL22;RPL13A;RPL35A;RPS3A;RPS26;RPS28;RPS29;RPL27A;RPL37A;RPL24;RPL27;RPS20;RPS24;RPS23 |
| 21 | Major pathway of rRNA processing in the nucleolus Homo sapiens R-HSA-6791226 | 1.19E-15 | 5.56E-14 | 6.41E+00 | 2.20E+02 | RPL4;RPL32;RPL10;RPL34;RPL12;RPL11;RPL36A;RPL9;RPL6;RPL7;RPS4X;RPS14;RPS17;RPS3;RPL38;RPL15;RPS11;RPL39;RPL41;RPL21;RPL23;RPL22;RPL13A;RPL35A;RPS3A;RPS26;RPS28;RPS29;RPL27A;RPL37A;RPL24;RPL27;RPS20;RPS24;RPS23 |

| | | | | | | |
|---|---|---|---|---|---|---|
| 22 | Metabolism of amino acids and derivatives Homo sapiens R-HSA-71291 | 3.10E-15 | 1.38E-13 | 4.25E+00 | 1.42E+02 | RPL4;RPL32;GLDC;RPL34;RPL9;SAT1;RPL6;RPL7;RPS4X;RPS14;RPS17;RPL38;RPS11;RPL39;RPL21;RPL23;RPL22;RPL13A;RPS3A;RPL37A;RPL24;RPL27;DDC;OAT;RPL10;RPL12;RPL11;RPL36A;PAPSS2;RPS3;RPL15;CKB;RPL41;RPL35A;FAH;ASS1;AGMAT;RPS26;ALDH4A1;RPS28;ALDH6A1;GATM;SLC6A8;RPS29;PSAT1;RPL27A;RPS20;CDO1;RPS24;RPS23 |
| 23 | rRNA processing Homo sapiens R-HSA-72312 | 1.61E-14 | 6.86E-13 | 5.78E+00 | 1.84E+02 | RPL4;RPL32;RPL10;RPL34;RPL12;RPL11;RPL36A;RPL9;RPL6;RPL7;RPS4X;RPS14;RPS17;RPS3;RPL38;RPL15;RPS11;RPL39;RPL41;RPL21;RPL23;RPL22;RPL13A;RPL35A;RPS3A;RPS26;RPS28;RPS29;RPL27A;RPL37A;RPL24;RPL27;RPS20;RPS24;RPS23 |
| 24 | Metabolism Homo sapiens R-HSA-1430728 | 5.71E-13 | 2.33E-11 | 2.07E+00 | 5.85E+01 | RPL4;CDA;ALDH1L1;SLC46A1;AHCYL1;RPL32;GLDC;RPL34;MT1X;SAT1;RPL9;NUDT3;RPL6;RPL7;RPS14;MT2A;RPS17;MPC1;MPC2;RPL38;NUDT16;RPS11;RPL39;FADS1;RPL21;RPL23;ACOT11;RPL22;CMBL;CYP27A1;PRKAR1B;TBL1XR1;MT1F;MT1G;RPL24;MT1H;RPL27;MT1E;ABCB1;UQCRB;SDC4;COX16;FECH;SLC22A2;MGST1;PIK3R2;PIK3R1;GLYAT;UQCRH;SULT1A1;PLCG2;ST3GAL4;APOE;PPARGC1A;DECR1;SLC19A2;RPL41;GLYATL1;EPHX2;IDH2;DHCR24;MT3;RPS26;ALDH6A1;RPS28;SLC6A8;RPS29;RPL27A;ALB;CYCS;RPS20;RPS24;RPS23;GPI;COX7B;ACSM2A;PYGL;OPLAH;ADAL;RPS4X;SPTLC2;HYAL1;IMPA2;NAPRT;COX8A;CHST9;CERS4;SREBF1;SORD;RPL13A;RPS3A;BPGM;GPCPD1;EHHADH;RPL37A;L2HGDH;ALDOB;CHST2;VAMP2;UGT2B7;MGLL;PRPS2;DDC;OAT;LRP1;SGMS1;RPL10;ADH1C;HPGD;RPL12;RPL11;RPL36A;NDUFB1;GLRX;PAPSS2;RXRA;MAT2A;ALDH1B1;GPC1;RPS3;CKB;RPL15;ACSS1;CA12;NDUFA5;NDUFA4;MMAB;GCGR;SCD5;MBOAT1;NDUFA1;RPL35A;FAH;ASS1;AGMAT;FABP1;ALDH4A1;FABP3;GATM;MARCKS;PSAT1;CDO1;LPIN1;PLCD1 |

| | | | | | | |
|---|---|---|---|---|---|---|
| 25 | Disease Homo sapiens R-HSA-1643685 | 1.48E-11 | 5.80E-10 | 2.65E+00 | 6.61E+01 | RPL4;CDKN1B;MAML2;RPL32;RPL34;RPL9;RPL6;RPL7;RPS4X;ADAMTS5;RPS14;RPS17;FGF9;ADAMTSL3;CNTRL;RPL38;BANF1;RPS11;RPL39;MAP2K1;RPL21;RPL23;RPL22;RPL13A;RPS3A;TBL1XR1;RPL37A;RPL24;RPL27;VAMP2;NOTCH1;RPL10;SDC4;RPL12;RPL11;RPL36A;LRP5;NEDD4L;PDGFA;PIK3R2;PIK3R1;FOXO3;FOXO1;ADAMTS16;ADAMTS15;POLR2A;ERBB4;GPC1;RPS3;RPL15;TCF7L2;RPL41;JAG1;EGF;HMGA1;RPL35A;GTF2H2;GTF2H5;MIB1;RPS26;RPS28;RPS29;RPL27A;ALB;CTNNB1;RPS20;MUC20;FGFR3;RPS24;FGFR2;RPS23 |
| 26 | Infectious disease Homo sapiens R-HSA-5663205 | 1.60E-10 | 6.03E-09 | 3.38E+00 | 7.63E+01 | RPL4;RPL32;RPL10;RPL34;RPL12;RPL11;RPL36A;NEDD4L;RPL9;RPL6;RPL7;RPS4X;RPS14;RPS17;POLR2A;RPS3;RPL38;BANF1;RPL15;RPS11;RPL39;RPL41;MAP2K1;RPL21;RPL23;RPL22;HMGA1;RPL13A;RPL35A;RPS3A;GTF2H2;GTF2H5;RPS26;RPS28;RPS29;RPL27A;RPL37A;RPL24;RPL27;RPS20;RPS24;VAMP2;RPS23 |
| 27 | Translation initiation complex formation Homo sapiens R-HSA-72649 | 7.00E-09 | 2.54E-07 | 8.39E+00 | 1.58E+02 | EIF1AX;RPS3A;RPS4X;RPS26;RPS14;RPS17;RPS28;EIF2S3;RPS29;RPS3;RPS20;PABPC1;RPS11;RPS24;RPS23 |
| 28 | Activation of the mRNA upon binding of the cap-binding complex and eIFs, and subsequent binding to 43S Homo sapiens R-HSA-72662 | 9.08E-09 | 3.12E-07 | 8.20E+00 | 1.52E+02 | EIF1AX;RPS3A;RPS4X;RPS26;RPS14;RPS17;RPS28;EIF2S3;RPS29;RPS3;RPS20;PABPC1;RPS11;RPS24;RPS23 |
| 29 | Formation of the ternary complex, and subsequently, the 43S complex Homo sapiens R-HSA-72695 | 9.25E-09 | 3.12E-07 | 9.13E+00 | 1.69E+02 | EIF1AX;RPS3A;RPS4X;RPS26;RPS14;RPS17;RPS28;EIF2S3;RPS29;RPS3;RPS20;RPS11;RPS24;RPS23 |
| 30 | Ribosomal scanning and start codon recognition Homo sapiens R-HSA-72702 | 5.80E-08 | 1.79E-06 | 7.64E+00 | 1.27E+02 | EIF1AX;RPS3A;RPS4X;RPS26;RPS14;RPS17;RPS28;EIF2S3;RPS29;RPS3;RPS20;RPS11;RPS24;RPS23 |
| 31 | Response to metal ions Homo sapiens R-HSA-5660526 | 5.85E-08 | 1.79E-06 | 4.08E+01 | 6.80E+02 | MT2A;MT1F;MT1G;MT1X;MT1H;MT3;MT1E |
| 32 | Metallothioneins bind metals Homo sapiens R-HSA-5661231 | 5.85E-08 | 1.79E-06 | 4.08E+01 | 6.80E+02 | MT2A;MT1F;MT1G;MT1X;MT1H;MT3;MT1E |

| | | | | | | |
|---|---|---|---|---|---|---|
| 33 | Metabolism of proteins Homo sapiens R-HSA-392499 | 3.84E-05 | 1.14E-03 | 1.72E+00 | 1.75E+01 | RPL4;RPL32;PIGQ;RPL34;RPL9;ACTB;RPL6;DCAF7;RPL7;RPS4X;ADAMTS5;RPS14;RPS17;ANPEP;ADAMTSL3;SEC61G;KIF5A;RPL38;RPS11;LMCD1;CTSD;RPL39;CTSC;RPL21;EIF1AX;RPL23;RPL22;RPL13A;RPS3A;DYNLL1;DYNLL2;B3GNT9;EEF1D;CTDSP2;CSNK2B;RPL37A;RPL24;RPL27;PABPC1;VAMP2;RPL10;RPL12;RAB1B;USP11;RPL11;RPL36A;ADAMTS16;ADAMTS15;CHST10;ST3GAL4;RPS3;ST3GAL5;GCNT4;RPL15;NUS1;TCF7L2;RPL41;PMM1;RPA1;RPL35A;TBCA;RPS26;RPS28;EIF2S3;RPS29;RPL27A;CTNNB1;FBXL3;RPS20;MUC20;RPS24;RPS23 |
| 34 | Gene Expression Homo sapiens R-HSA-74160 | 6.62E-04 | 1.91E-02 | 1.47E+00 | 1.07E+01 | RPL4;GPI;COX7B;CDKN1B;MAML2;RPL32;RPL34;RORC;YBX1;RPL9;ACTB;RPL6;NR3C2;RPL7;RPS4X;RPS14;RPS17;HNF4A;SEC61G;RPL38;RPS11;JUNB;RPL39;ZNF124;COX8A;RPL21;EIF1AX;RPL23;RPL22;RPL13A;RPS3A;FOS;PRKAB1;EEF1D;HNRNPH1;TBL1XR1;CSNK2B;RPL37A;RRAGD;SRSF3;RPL24;RPL27;SRSF5;PABPC1;ZNF556;SRSF6;NOTCH1;COX16;RPL10;RPL12;RPL11;RPL36A;NEDD4L;NDRG1;LAGE3;ZFP36L1;RXRA;POLR2A;PCBP4;PLAGL1;PCBP1;PCBP2;ZNF549;RPS3;ZNF426;ZNF304;SNRPB2;RPL15;HSPA8;JUN;RPL41;BRPF1;NDUFA4;GADD45A;RPA1;RPL35A;ARID3A;GTF2H2;GTF2H5;SMAD7;TNRC6C;RPS26;NUDT21;RPS28;EIF2S3;RPS29;RPL27A;NFIC;ZNF615;CYCS;RPS20;RPS24;ZNF772;RPS23 |
| 35 | PI3K/AKT Signaling in Cancer Homo sapiens R-HSA-2219528 | 8.33E-04 | 2.33E-02 | 3.42E+00 | 2.43E+01 | CDKN1B;FGF9;ERBB4;EGF;PDGFA;PIK3R2;PIK3R1;FOXO3;FGFR3;FOXO1;FGFR2 |
| 36 | Scavenging by Class A Receptors Homo sapiens R-HSA-3000480 | 8.65E-04 | 2.35E-02 | 8.30E+00 | 5.86E+01 | COL4A2;COL4A1;FTH1;APOE;FTL |
| 37 | Golgi Associated Vesicle Biogenesis Homo sapiens R-HSA-432722 | 1.62E-03 | 4.28E-02 | 4.05E+00 | 2.60E+01 | TPD52;HSPA8;BLOC1S6;FTH1;HIP1R;VAMP2;FTL;PICALM |
| 38 | The citric acid (TCA) cycle and respiratory electron transport Homo sapiens R-HSA-1428517 | 1.76E-03 | 4.54E-02 | 2.54E+00 | 1.61E+01 | COX8A;COX7B;COX16;UQCRB;NDUFA5;NDUFA4;IDH2;NDUFA1;NDUFB1;UQCRH;RXRA;MPC1;MPC2;CYCS;L2HGDH |

| | | | | | | |
|---|---|---|---|---|---|---|
| 39 | GAB1 signalosome Homo sapiens R-HSA-180292 | 2.06E-03 | 5.18E-02 | 2.71E+00 | 1.68E+01 | CDKN1B;EGF;PDGFA;PIK3R2;PIK3R1;FOXO3;FOXO1;TNRC6C;FGF9;ERBB4;FGFR3;PAG1;FGFR2 |
| 40 | Conjugation of salicylate with glycine Homo sapiens R-HSA-177128 | 3.40E-03 | 8.24E-02 | 1.39E+01 | 7.91E+01 | GLYATL1;ACSM2A;GLYAT |
| 41 | Respiratory electron transport Homo sapiens R-HSA-611105 | 3.45E-03 | 8.24E-02 | 2.99E+00 | 1.69E+01 | COX8A;COX7B;COX16;UQCRB;NDUFA5;NDUFA4;NDUFA1;NDUFB1;CYCS;UQCRH |
| 42 | Constitutive Signaling by Aberrant PI3K in Cancer Homo sapiens R-HSA-2219530 | 3.55E-03 | 8.27E-02 | 3.51E+00 | 1.98E+01 | FGF9;ERBB4;EGF;PDGFA;PIK3R2;PIK3R1;FGFR3;FGFR2 |
| 43 | PI-3K cascade:FGFR1 Homo sapiens R-HSA-5654689 | 4.75E-03 | 9.67E-02 | 2.55E+00 | 1.36E+01 | TNRC6C;CDKN1B;FGF9;ERBB4;EGF;PDGFA;PIK3R2;PIK3R1;FOXO3;FGFR3;FOXO1;FGFR2 |
| 44 | PI-3K cascade:FGFR3 Homo sapiens R-HSA-5654710 | 4.75E-03 | 9.67E-02 | 2.55E+00 | 1.36E+01 | TNRC6C;CDKN1B;FGF9;ERBB4;EGF;PDGFA;PIK3R2;PIK3R1;FOXO3;FGFR3;FOXO1;FGFR2 |
| 45 | PI3K events in ERBB4 signaling Homo sapiens R-HSA-1250342 | 4.75E-03 | 9.67E-02 | 2.55E+00 | 1.36E+01 | TNRC6C;CDKN1B;FGF9;ERBB4;EGF;PDGFA;PIK3R2;PIK3R1;FOXO3;FGFR3;FOXO1;FGFR2 |
| 46 | PIP3 activates AKT signaling Homo sapiens R-HSA-1257604 | 4.75E-03 | 9.67E-02 | 2.55E+00 | 1.36E+01 | TNRC6C;CDKN1B;FGF9;ERBB4;EGF;PDGFA;PIK3R2;PIK3R1;FOXO3;FGFR3;FOXO1;FGFR2 |
| 47 | PI-3K cascade:FGFR4 Homo sapiens R-HSA-5654720 | 4.75E-03 | 9.67E-02 | 2.55E+00 | 1.36E+01 | TNRC6C;CDKN1B;FGF9;ERBB4;EGF;PDGFA;PIK3R2;PIK3R1;FOXO3;FGFR3;FOXO1;FGFR2 |
| 48 | PI-3K cascade:FGFR2 Homo sapiens R-HSA-5654695 | 4.75E-03 | 9.67E-02 | 2.55E+00 | 1.36E+01 | TNRC6C;CDKN1B;FGF9;ERBB4;EGF;PDGFA;PIK3R2;PIK3R1;FOXO3;FGFR3;FOXO1;FGFR2 |
| 49 | Conjugation of carboxylic acids Homo sapiens R-HSA-159424 | 4.94E-03 | 9.67E-02 | 1.16E+01 | 6.16E+01 | GLYATL1;ACSM2A;GLYAT |
| 50 | Amino Acid conjugation Homo sapiens R-HSA-156587 | 4.94E-03 | 9.67E-02 | 1.16E+01 | 6.16E+01 | GLYATL1;ACSM2A;GLYAT |

| | | | | | | |
|---|---|---|---|---|---|---|
| 51 | Norepinephrine Neurotransmitter Release Cycle Homo sapiens R-HSA-181430 | 5.63E-03 | 1.08E-01 | 6.63E+00 | 3.44E+01 | SLC22A2;CPLX1;VAMP2;PPFIA3 |
| 52 | PI3K/AKT activation Homo sapiens R-HSA-198203 | 5.77E-03 | 1.09E-01 | 2.48E+00 | 1.28E+01 | TNRC6C;CDKN1B;FGF9;ERBB4;EGF;PDGFA;PIK3R2;PIK3R1;FOXO3;FGFR3;FOXO1;FGFR2 |
| 53 | Creatine metabolism Homo sapiens R-HSA-71288 | 6.84E-03 | 1.24E-01 | 9.94E+00 | 4.96E+01 | GATM;SLC6A8;CKB |
| 54 | Abacavir transport and metabolism Homo sapiens R-HSA-2161522 | 6.84E-03 | 1.24E-01 | 9.94E+00 | 4.96E+01 | ADAL;ABCB1;SLC22A2 |
| 55 | Deactivation of the beta-catenin transactivating complex Homo sapiens R-HSA-3769402 | 7.31E-03 | 1.30E-01 | 3.88E+00 | 1.91E+01 | TLE3;TCF7L2;BCL9;SOX13;CTNNB1;SOX4 |
| 56 | TP53 Regulates Metabolic Genes Homo sapiens R-HSA-5628897 | 7.93E-03 | 1.39E-01 | 2.79E+00 | 1.35E+01 | COX8A;TNRC6C;GPI;COX7B;COX16;NDUFA4;RRAGD;CYCS;PRKAB1 |
| 57 | trans-Golgi Network Vesicle Budding Homo sapiens R-HSA-199992 | 8.23E-03 | 1.39E-01 | 3.00E+00 | 1.44E+01 | TPD52;HSPA8;BLOC1S6;FTH1;HIP1R;VAMP2;FTL;PICALM |
| 58 | Clathrin derived vesicle budding Homo sapiens R-HSA-421837 | 8.23E-03 | 1.39E-01 | 3.00E+00 | 1.44E+01 | TPD52;HSPA8;BLOC1S6;FTH1;HIP1R;VAMP2;FTL;PICALM |
| 59 | Diseases of signal transduction Homo sapiens R-HSA-5663202 | 9.01E-03 | 1.50E-01 | 1.84E+00 | 8.67E+00 | TCF7L2;JAG1;CDKN1B;NOTCH1;MAML2;EGF;LRP5;PDGFA;PIK3R2;PIK3R1;FOXO3;FOXO1;MIB1;FGF9;POLR2A;TBL1XR1;ERBB4;CNTRL;CTNNB1;FGFR3;FGFR2 |
| 60 | Signaling by PDGF Homo sapiens R-HSA-186797 | 9.70E-03 | 1.58E-01 | 1.73E+00 | 8.01E+00 | AHCYL1;CDKN1B;PDGFA;PIK3R2;PIK3R1;FOXO3;FOXO1;FGF9;ERBB4;SPP1;NEFL;DUSP2;MAP2K1;EGF;IL17RD;GRIN2B;DUSP9;TNRC6C;PRKAR1B;COL4A2;COL4A1;COL6A1;COL9A2;FGFR3;FGFR2 |
| 61 | Ethanol oxidation Homo sapiens R-HSA-71384 | 1.18E-02 | 1.89E-01 | 7.73E+00 | 3.43E+01 | ALDH1B1;ADH1C;ACSS1 |

| | | | | | | |
|---|---|---|---|---|---|---|
| 62 | TP53 Regulates Transcription of Genes Involved in G1 Cell Cycle Arrest Homo sapiens R-HSA-6804116 | 1.49E-02 | 2.34E-01 | 6.96E+00 | 2.93E+01 | CDKN1B;PCBP4;ARID3A |
| 63 | Respiratory electron transport, ATP synthesis by chemiosmotic coupling, and heat production by uncoupling proteins. Homo sapiens R-HSA-163200 | 1.51E-02 | 2.34E-01 | 2.35E+00 | 9.87E+00 | COX8A;COX7B;COX16;UQCRB;NDUFA5;NDUFA4;NDUFA1;CYCS;NDUFB1;UQCRH |
| 64 | Abacavir transmembrane transport Homo sapiens R-HSA-2161517 | 1.58E-02 | 2.38E-01 | 1.55E+01 | 6.41E+01 | ABCB1;SLC22A2 |
| 65 | APOBEC3G mediated resistance to HIV-1 infection Homo sapiens R-HSA-180689 | 1.58E-02 | 2.38E-01 | 1.55E+01 | 6.41E+01 | HMGA1;BANF1 |
| 66 | Growth hormone receptor signaling Homo sapiens R-HSA-982772 | 1.61E-02 | 2.38E-01 | 4.64E+00 | 1.92E+01 | GHR;SOCS2;SOCS1;CISH |
| 67 | PI3K Cascade Homo sapiens R-HSA-109704 | 1.65E-02 | 2.41E-01 | 2.62E+00 | 1.08E+01 | FGF9;CAB39;RRAGD;PIK3R2;PIK3R1;PRKAB1;FGFR3;FGFR2 |
| 68 | Repression of WNT target genes Homo sapiens R-HSA-4641265 | 1.83E-02 | 2.60E-01 | 6.33E+00 | 2.53E+01 | TLE3;TCF7L2;CTNNB1 |
| 69 | Neurotransmitter Release Cycle Homo sapiens R-HSA-112310 | 1.84E-02 | 2.60E-01 | 3.10E+00 | 1.24E+01 | HSPA8;SLC22A2;NAAA;CPLX1;PPFIA3;VAMP2 |
| 70 | Biological oxidations Homo sapiens R-HSA-211859 | 1.89E-02 | 2.65E-01 | 1.90E+00 | 7.54E+00 | GLYATL1;ADH1C;MGST1;ACSM2A;CMBL;GLYAT;OPLAH;PAPSS2;CYP27A1;SULT1A1;RXRA;ALDH1B1;MAT2A;ACSS1;UGT2B7 |
| 71 | Transcriptional Regulation by TP53 Homo sapiens R-HSA-3700989 | 1.92E-02 | 2.65E-01 | 1.65E+00 | 6.54E+00 | COX8A;GPI;JUN;COX7B;CDKN1B;BRPF1;COX16;GADD45A;NDUFA4;RPA1;ARID3A;FOS;GTF2H2;PRKAB1;NDRG1;GTF2H5;TNRC6C;POLR2A;PCBP4;PLAGL1;RRAGD;CSNK2B;CYCS |

| | | | | | | |
|---|---|---|---|---|---|---|
| 72 | Downregulation of TGF-beta receptor signaling Homo sapiens R-HSA-2173788 | 2.12E-02 | 2.85E-01 | 4.22E+00 | 1.63E+01 | PPP1R15A;BAMBI;NEDD4L;SMAD7 |
| 73 | PI5P, PP2A and IER3 Regulate PI3K/AKT Signaling Homo sapiens R-HSA-6811558 | 2.16E-02 | 2.85E-01 | 2.48E+00 | 9.52E+00 | FGF9;ERBB4;EGF;PDGFA;PIK3R2;PIK3R1;FGFR3;FGFR2 |
| 74 | Constitutive Signaling by NOTCH1 HD Domain Mutants Homo sapiens R-HSA-2691232 | 2.22E-02 | 2.85E-01 | 5.80E+00 | 2.21E+01 | NOTCH1;JAG1;MIB1 |
| 75 | Signaling by NOTCH1 HD Domain Mutants in Cancer Homo sapiens R-HSA-2691230 | 2.22E-02 | 2.85E-01 | 5.80E+00 | 2.21E+01 | NOTCH1;JAG1;MIB1 |
| 76 | Phase II conjugation Homo sapiens R-HSA-156580 | 2.30E-02 | 2.85E-01 | 2.30E+00 | 8.69E+00 | SULT1A1;GLYATL1;MAT2A;MGST1;ACSM2A;GLYAT;OPLAH;UGT2B7;PAPSS2 |
| 77 | Fructose metabolism Homo sapiens R-HSA-5652084 | 2.30E-02 | 2.85E-01 | 1.16E+01 | 4.37E+01 | SORD;ALDOB |
| 78 | PTK6 Regulates Cell Cycle Homo sapiens R-HSA-8849470 | 2.30E-02 | 2.85E-01 | 1.16E+01 | 4.37E+01 | CDKN1B;CCND1 |
| 79 | Conjugation of benzoate with glycine Homo sapiens R-HSA-177135 | 2.30E-02 | 2.85E-01 | 1.16E+01 | 4.37E+01 | GLYATL1;GLYAT |
| 80 | mRNA Splicing - Major Pathway Homo sapiens R-HSA-72163 | 2.37E-02 | 2.90E-01 | 2.08E+00 | 7.80E+00 | HSPA8;NUDT21;POLR2A;HNRNPH1;PCBP1;PCBP2;SRSF3;SNRPB2;SRSF5;YBX1;SRSF6 |
| 81 | RORA activates gene expression Homo sapiens R-HSA-1368082 | 2.41E-02 | 2.91E-01 | 4.04E+00 | 1.50E+01 | SREBF1;RXRA;TBL1XR1;PPARGC1A |
| 82 | Pre-NOTCH Transcription and Translation Homo sapiens R-HSA-1912408 | 2.64E-02 | 3.13E-01 | 3.23E+00 | 1.17E+01 | TNRC6C;JUN;NOTCH1;MAML2;CCND1 |

| | | | | | | |
|---|---|---|---|---|---|---|
| 83 | PI3K events in ERBB2 signaling Homo sapiens R-HSA-1963642 | 2.65E-02 | 3.13E-01 | 5.35E+00 | 1.94E+01 | EGF;ERBB4;PIK3R1 |
| 84 | Diseases of glycosylation Homo sapiens R-HSA-3781865 | 2.94E-02 | 3.26E-01 | 2.33E+00 | 8.20E+00 | ADAMTS16;ADAMTS5;ADAMTS15;NOTCH1;SDC4;ADAMTSL3;GPC1;MUC20 |
| 85 | Pre-NOTCH Expression and Processing Homo sapiens R-HSA-1912422 | 3.01E-02 | 3.26E-01 | 2.73E+00 | 9.58E+00 | TNRC6C;JUN;NOTCH1;MAML2;CCND1;ST3GAL4 |
| 86 | Signaling by NOTCH Homo sapiens R-HSA-157118 | 3.04E-02 | 3.26E-01 | 2.08E+00 | 7.26E+00 | TNRC6C;TLE3;JUN;NOTCH1;JAG1;MAML2;CCND1;TBL1XR1;ST3GAL4;MIB1 |
| 87 | Acetylcholine Neurotransmitter Release Cycle Homo sapiens R-HSA-264642 | 3.13E-02 | 3.26E-01 | 4.97E+00 | 1.72E+01 | CPLX1;VAMP2;PPFIA3 |
| 88 | 2-LTR circle formation Homo sapiens R-HSA-164843 | 3.14E-02 | 3.26E-01 | 9.27E+00 | 3.21E+01 | HMGA1;BANF1 |
| 89 | Clearance of Nuclear Envelope Membranes from Chromatin Homo sapiens R-HSA-2993913 | 3.14E-02 | 3.26E-01 | 9.27E+00 | 3.21E+01 | BANF1;EMD |
| 90 | Signaling by NOTCH1 t(7;9)(NOTCH1:M1580K2555) Translocation Mutant Homo sapiens R-HSA-2660825 | 3.14E-02 | 3.26E-01 | 9.27E+00 | 3.21E+01 | NOTCH1;JAG1 |
| 91 | Constitutive Signaling by NOTCH1 t(7;9)(NOTCH1:M1580K2555) Translocation Mutant Homo sapiens R-HSA-2660826 | 3.14E-02 | 3.26E-01 | 9.27E+00 | 3.21E+01 | NOTCH1;JAG1 |

| | | | | | | |
|---|---|---|---|---|---|---|
| 92 | FGFR3b ligand binding and activation Homo sapiens R-HSA-190371 | 3.14E-02 | 3.26E-01 | 9.27E+00 | 3.21E+01 | FGF9;FGFR3 |
| 93 | Binding of TCF/LEF:CTNNB1 to target gene promoters Homo sapiens R-HSA-4411364 | 3.14E-02 | 3.26E-01 | 9.27E+00 | 3.21E+01 | TCF7L2;CTNNB1 |
| 94 | Iron uptake and transport Homo sapiens R-HSA-917937 | 3.17E-02 | 3.26E-01 | 3.06E+00 | 1.05E+01 | SLC46A1;SLC22A17;FTH1;CYBRD1;FTL |
| 95 | Signaling by FGFR Homo sapiens R-HSA-190236 | 3.19E-02 | 3.26E-01 | 1.57E+00 | 5.40E+00 | DUSP2;MAP2K1;AHCYL1;CDKN1B;EGF;PDGFA;PIK3R2;PIK3R1;IL17RD;FOXO3;GRIN2B;DUSP9;FOXO1;TNRC6C;PRKAR1B;FGF9;POLR2A;HNRNPH1;ERBB4;FLRT1;NEFL;FGFR3;FGFR2 |
| 96 | Degradation of the extracellular matrix Homo sapiens R-HSA-1474228 | 3.19E-02 | 3.26E-01 | 2.16E+00 | 7.43E+00 | ADAMTS16;COL18A1;ADAMTS5;COL26A1;CAPN2;TIMP2;SPP1;COL9A2;CTSD |
| 97 | Negative regulation of the PI3K/AKT network Homo sapiens R-HSA-199418 | 3.31E-02 | 3.34E-01 | 2.27E+00 | 7.73E+00 | FGF9;ERBB4;EGF;PDGFA;PIK3R2;PIK3R1;FGFR3;FGFR2 |
| 98 | Binding and Uptake of Ligands by Scavenger Receptors Homo sapiens R-HSA-2173782 | 3.34E-02 | 3.34E-01 | 2.43E+00 | 8.25E+00 | COL4A2;LRP1;COL4A1;FTH1;ALB;APOE;FTL |
| 99 | Serotonin Neurotransmitter Release Cycle Homo sapiens R-HSA-181429 | 3.64E-02 | 3.43E-01 | 4.64E+00 | 1.54E+01 | CPLX1;VAMP2;PPFIA3 |
| 100 | Downstream signaling of activated FGFR1 Homo sapiens R-HSA-5654687 | 3.68E-02 | 3.43E-01 | 1.58E+00 | 5.20E+00 | DUSP2;MAP2K1;AHCYL1;CDKN1B;EGF;PDGFA;PIK3R2;PIK3R1;IL17RD;FOXO3;GRIN2B;DUSP9;FOXO1;TNRC6C;PRKAR1B;FGF9;ERBB4;FLRT1;NEFL;FGFR3;FGFR2 |
| 101 | MyD88:Mal cascade initiated on plasma membrane Homo sapiens R-HSA-166058 | 3.70E-02 | 3.43E-01 | 2.21E+00 | 7.30E+00 | MEF2A;MAP2K1;JUN;SOCS1;MAPKAPK3;PELI2;TAB2;FOS |

| | | | | | | |
|---|---|---|---|---|---|---|
| 102 | Toll Like Receptor TLR1:TLR2 Cascade Homo sapiens R-HSA-168179 | 3.70E-02 | 3.43E-01 | 2.21E+00 | 7.30E+00 | MEF2A;MAP2K1;JUN;SOCS1;MAPKAPK3;PELI2;TAB2;FOS |
| 103 | Toll Like Receptor TLR6:TLR2 Cascade Homo sapiens R-HSA-168188 | 3.70E-02 | 3.43E-01 | 2.21E+00 | 7.30E+00 | MEF2A;MAP2K1;JUN;SOCS1;MAPKAPK3;PELI2;TAB2;FOS |
| 104 | Toll Like Receptor 2 (TLR2) Cascade Homo sapiens R-HSA-181438 | 3.70E-02 | 3.43E-01 | 2.21E+00 | 7.30E+00 | MEF2A;MAP2K1;JUN;SOCS1;MAPKAPK3;PELI2;TAB2;FOS |
| 105 | mRNA Splicing Homo sapiens R-HSA-72172 | 3.74E-02 | 3.43E-01 | 1.92E+00 | 6.33E+00 | HSPA8;NUDT21;POLR2A;HNRNPH1;PCBP1;PCBP2;SRSF3;SNRPB2;SRSF5;YBX1;SRSF6 |
| 106 | MAP kinase activation in TLR cascade Homo sapiens R-HSA-450294 | 3.74E-02 | 3.43E-01 | 2.58E+00 | 8.48E+00 | MEF2A;MAP2K1;JUN;MAPKAPK3;TAB2;FOS |
| 107 | Role of LAT2/NTAL/LAB on calcium mobilization Homo sapiens R-HSA-2730905 | 3.75E-02 | 3.43E-01 | 1.86E+00 | 6.12E+00 | TNRC6C;CDKN1B;FGF9;ERBB4;EGF;PDGFA;PIK3R2;PIK3R1;FOXO3;FGFR3;FOXO1;FGFR2 |
| 108 | Adherens junctions interactions Homo sapiens R-HSA-418990 | 3.79E-02 | 3.44E-01 | 3.44E+00 | 1.12E+01 | CDH4;CDH3;CADM1;CTNNB1 |
| 109 | Developmental Biology Homo sapiens R-HSA-1266738 | 3.92E-02 | 3.52E-01 | 1.36E+00 | 4.40E+00 | NOTCH1;PDGFA;FOXO3;EFNA5;FOXO1;ACTB;ACTG1;CDH4;ABLIM1;RXRA;SALL1;FGF9;POLR2A;CHL1;ERBB4;HNF4A;GPC1;NEFL;HOXC4;SCN3B;PPARGC1A;PAK4;MEF2A;SREBF1;HSPA8;JUN;DUSP2;MAP2K1;EGF;IL17RD;GRIN2B;DUSP9;PBX1;RHOB;COL4A2;COL4A1;TBL1XR1;CSNK2B;COL6A1;CTNNB1;COL9A2;FGFR3;FGFR2 |
| 110 | Cell-cell junction organization Homo sapiens R-HSA-421270 | 4.01E-02 | 3.54E-01 | 2.53E+00 | 8.15E+00 | CLDN10;CDH4;CDH3;CADM1;CLDN19;CTNNB1 |
| 111 | Signaling by EGFR Homo sapiens R-HSA-177929 | 4.02E-02 | 3.54E-01 | 1.54E+00 | 4.96E+00 | DUSP2;MAP2K1;AHCYL1;CDKN1B;EGF;PDGFA;PIK3R2;PIK3R1;IL17RD;FOXO3;GRIN2B;DUSP9;FOXO1;TNRC6C;PRKAR1B;FGF9;ERBB4;NEFL;SPRY1;FGFR3;PAG1;FGFR2 |
| 112 | Signaling by FGFR1 Homo sapiens R-HSA-5654736 | 4.10E-02 | 3.58E-01 | 1.56E+00 | 4.97E+00 | DUSP2;MAP2K1;AHCYL1;CDKN1B;EGF;PDGFA;PIK3R2;PIK3R1;IL17RD;FOXO3;GRIN2B;DUSP9;FOXO1;TNRC6C;PRKAR1B;FGF9;ERBB4;FLRT1;NEFL;FGFR3;FGFR2 |

| | | | | | | |
|---|---|---|---|---|---|---|
| 113 | GABA synthesis, release, reuptake and degradation Homo sapiens R-HSA-888590 | 4.19E-02 | 3.60E-01 | 4.35E+00 | 1.38E+01 | HSPA8;CPLX1;VAMP2 |
| 114 | TGF-beta receptor signaling activates SMADs Homo sapiens R-HSA-2173789 | 4.20E-02 | 3.60E-01 | 3.31E+00 | 1.05E+01 | PPP1R15A;BAMBI;NEDD4L;SMAD7 |
| 115 | Diseases associated with O-glycosylation of proteins Homo sapiens R-HSA-3906995 | 4.29E-02 | 3.66E-01 | 2.49E+00 | 7.83E+00 | ADAMTS16;ADAMTS5;ADAMTS15;NOTCH1;ADAMTSL3;MUC20 |
| 116 | DAP12 interactions Homo sapiens R-HSA-2172127 | 4.45E-02 | 3.75E-01 | 1.52E+00 | 4.74E+00 | DUSP2;MAP2K1;AHCYL1;CDKN1B;EGF;HLA-B;PDGFA;PIK3R2;PIK3R1;IL17RD;FOXO3;GRIN2B;DUSP9;FOXO1;TNRC6C;PRKAR1B;FGF9;ERBB4;PLCG2;NEFL;FGFR3;FGFR2 |
| 117 | Collagen biosynthesis and modifying enzymes Homo sapiens R-HSA-1650814 | 4.59E-02 | 3.75E-01 | 2.44E+00 | 7.53E+00 | COL18A1;COL26A1;COL4A2;COL4A1;COL6A1;COL9A2 |
| 118 | Signaling by FGFR in disease Homo sapiens R-HSA-1226099 | 4.59E-02 | 3.75E-01 | 2.44E+00 | 7.53E+00 | FGF9;POLR2A;CNTRL;PIK3R1;FGFR3;FGFR2 |
| 119 | Cell-Cell communication Homo sapiens R-HSA-1500931 | 4.60E-02 | 3.75E-01 | 1.92E+00 | 5.92E+00 | CLDN10;CDH4;CDH3;CADM1;CLDN19;FLNA;CTNNB1;PIK3R2;PIK3R1;SKAP2 |
| 120 | Detoxification of Reactive Oxygen Species Homo sapiens R-HSA-3299685 | 4.63E-02 | 3.75E-01 | 3.20E+00 | 9.84E+00 | PRDX3;NOX4;CYCS;SOD1 |
| 121 | Signaling by FGFR2 Homo sapiens R-HSA-5654738 | 4.68E-02 | 3.75E-01 | 1.51E+00 | 4.64E+00 | DUSP2;MAP2K1;AHCYL1;CDKN1B;EGF;PDGFA;PIK3R2;PIK3R1;IL17RD;FOXO3;GRIN2B;DUSP9;FOXO1;TNRC6C;PRKAR1B;FGF9;POLR2A;HNRNPH1;ERBB4;NEFL;FGFR3;FGFR2 |
| 122 | TP53 Regulates Transcription of Cell Cycle Genes Homo sapiens R-HSA-6791312 | 4.77E-02 | 3.75E-01 | 2.70E+00 | 8.21E+00 | CDKN1B;PCBP4;GADD45A;PLAGL1;ARID3A |

| | | | | | | |
|---|---|---|---|---|---|---|
| 123 | Pyruvate metabolism and Citric Acid (TCA) cycle Homo sapiens R-HSA-71406 | 4.77E-02 | 3.75E-01 | 2.70E+00 | 8.21E+00 | RXRA;MPC1;MPC2;IDH2;L2HGDH |
| 124 | Hormone-sensitive lipase (HSL)-mediated triacylglycerol hydrolysis Homo sapiens R-HSA-163560 | 4.78E-02 | 3.75E-01 | 4.09E+00 | 1.24E+01 | FABP1;FABP3;MGLL |
| 125 | Signaling by Hippo Homo sapiens R-HSA-2028269 | 4.78E-02 | 3.75E-01 | 4.09E+00 | 1.24E+01 | WWC1;AMOTL2;AMOTL1 |
| 126 | Extracellular matrix organization Homo sapiens R-HSA-1474244 | 4.82E-02 | 3.75E-01 | 1.58E+00 | 4.80E+00 | COL18A1;COL26A1;SDC4;LTBP4;PDGFA;ADAMTS16;BMP4;ADAMTS5;EFEMP2;COL4A2;COL4A1;COL6A1;TIMP2;SPP1;CAPN2;COL9A2;CTSD;JAM3 |
| 127 | DAP12 signaling Homo sapiens R-HSA-2424491 | 5.04E-02 | 3.80E-01 | 1.52E+00 | 4.53E+00 | DUSP2;MAP2K1;AHCYL1;CDKN1B;EGF;PDGFA;PIK3R2;PIK3R1;IL17RD;FOXO3;GRIN2B;DUSP9;FOXO1;TNRC6C;PRKAR1B;FGF9;ERBB4;PLCG2;NEFL;FGFR3;FGFR2 |
| 128 | Integration of provirus Homo sapiens R-HSA-162592 | 5.09E-02 | 3.80E-01 | 6.62E+00 | 1.97E+01 | HMGA1;BANF1 |
| 129 | AKT phosphorylates targets in the nucleus Homo sapiens R-HSA-198693 | 5.09E-02 | 3.80E-01 | 6.62E+00 | 1.97E+01 | FOXO3;FOXO1 |
| 130 | CHL1 interactions Homo sapiens R-HSA-447041 | 5.09E-02 | 3.80E-01 | 6.62E+00 | 1.97E+01 | HSPA8;CHL1 |
| 131 | Prostanoid ligand receptors Homo sapiens R-HSA-391908 | 5.09E-02 | 3.80E-01 | 6.62E+00 | 1.97E+01 | PTGFR;PTGER3 |
| 132 | Mitochondrial biogenesis Homo sapiens R-HSA-1592230 | 5.14E-02 | 3.81E-01 | 2.64E+00 | 7.83E+00 | RXRA;TBL1XR1;CYCS;PRKAB1;PPARGC1A |
| 133 | Toll Like Receptor 10 (TLR10) Cascade Homo sapiens R-HSA-168142 | 5.35E-02 | 3.88E-01 | 2.17E+00 | 6.35E+00 | MEF2A;MAP2K1;JUN;MAPKAPK3;PELI2;TAB2;FOS |

| | | | | | | |
|---|---|---|---|---|---|---|
| 134 | Toll Like Receptor 5 (TLR5) Cascade Homo sapiens R-HSA-168176 | 5.35E-02 | 3.88E-01 | 2.17E+00 | 6.35E+00 | MEF2A;MAP2K1;JUN;MAPKAPK3;PELI2;TAB2;FOS |
| 135 | MyD88 cascade initiated on plasma membrane Homo sapiens R-HSA-975871 | 5.35E-02 | 3.88E-01 | 2.17E+00 | 6.35E+00 | MEF2A;MAP2K1;JUN;MAPKAPK3;PELI2;TAB2;FOS |
| 136 | Cellular responses to stress Homo sapiens R-HSA-2262752 | 5.41E-02 | 3.89E-01 | 1.49E+00 | 4.34E+00 | HSPA8;JUN;CBX6;CDKN1B;ST13;CITED2;HSBP1;RPA1;HMGA1;FOS;DYNLL1;PRKAB1;DYNLL2;SOD1;PRDX3;TNRC6C;MAPKAPK3;RRAGD;ID1;NOX4;CYCS;HSPA1B |
| 137 | TRAF6 mediated induction of NFkB and MAP kinases upon TLR7/8 or 9 activation Homo sapiens R-HSA-975138 | 5.64E-02 | 3.99E-01 | 2.14E+00 | 6.15E+00 | MEF2A;MAP2K1;JUN;MAPKAPK3;PELI2;TAB2;FOS |
| 138 | Downstream signaling of activated FGFR2 Homo sapiens R-HSA-5654696 | 5.71E-02 | 3.99E-01 | 1.51E+00 | 4.32E+00 | DUSP2;MAP2K1;AHCYL1;CDKN1B;EGF;PDGFA;PIK3R2;PIK3R1;IL17RD;FOXO3;GRIN2B;DUSP9;FOXO1;TNRC6C;PRKAR1B;FGF9;ERBB4;NEFL;FGFR3;FGFR2 |
| 139 | Downstream signaling of activated FGFR4 Homo sapiens R-HSA-5654716 | 5.71E-02 | 3.99E-01 | 1.51E+00 | 4.32E+00 | DUSP2;MAP2K1;AHCYL1;CDKN1B;EGF;PDGFA;PIK3R2;PIK3R1;IL17RD;FOXO3;GRIN2B;DUSP9;FOXO1;TNRC6C;PRKAR1B;FGF9;ERBB4;NEFL;FGFR3;FGFR2 |
| 140 | Downstream signaling of activated FGFR3 Homo sapiens R-HSA-5654708 | 5.71E-02 | 3.99E-01 | 1.51E+00 | 4.32E+00 | DUSP2;MAP2K1;AHCYL1;CDKN1B;EGF;PDGFA;PIK3R2;PIK3R1;IL17RD;FOXO3;GRIN2B;DUSP9;FOXO1;TNRC6C;PRKAR1B;FGF9;ERBB4;NEFL;FGFR3;FGFR2 |
| 141 | Signaling by ERBB4 Homo sapiens R-HSA-1236394 | 5.85E-02 | 4.05E-01 | 1.50E+00 | 4.27E+00 | DUSP2;MAP2K1;CDKN1B;EGF;PDGFA;PIK3R2;PIK3R1;IL17RD;FOXO3;GRIN2B;DUSP9;FOXO1;GHR;TNRC6C;FGF9;ERBB4;NEFL;TAB2;FGFR3;FGFR2 |
| 142 | Integrin cell surface interactions Homo sapiens R-HSA-216083 | 5.88E-02 | 4.05E-01 | 2.28E+00 | 6.47E+00 | COL18A1;COL4A2;COL4A1;SPP1;COL9A2;JAM3 |
| 143 | Signaling by FGFR3 point mutants in cancer Homo sapiens R-HSA-8853338 | 6.08E-02 | 4.06E-01 | 3.66E+00 | 1.02E+01 | FGF9;PIK3R1;FGFR3 |
| 144 | Signaling by FGFR3 in disease Homo sapiens R-HSA-5655332 | 6.08E-02 | 4.06E-01 | 3.66E+00 | 1.02E+01 | FGF9;PIK3R1;FGFR3 |

| | | | | | | |
|---|---|---|---|---|---|---|
| 145 | Signaling by FGFR4 Homo sapiens R-HSA-5654743 | 6.15E-02 | 4.06E-01 | 1.49E+00 | 4.17E+00 | DUSP2;MAP2K1;AHCYL1;CDKN1B;EGF;PDGFA;PIK3R2;PIK3R1;IL17RD;FOXO3;GRIN2B;DUSP9;FOXO1;TNRC6C;PRKAR1B;FGF9;ERBB4;NEFL;FGFR3;FGFR2 |
| 146 | Activation of PPARGC1A (PGC-1alpha) by phosphorylation Homo sapiens R-HSA-2151209 | 6.19E-02 | 4.06E-01 | 5.79E+00 | 1.61E+01 | PPARGC1A;PRKAB1 |
| 147 | Signaling by FGFR3 fusions in cancer Homo sapiens R-HSA-8853334 | 6.19E-02 | 4.06E-01 | 5.79E+00 | 1.61E+01 | PIK3R1;FGFR3 |
| 148 | Activation of the AP-1 family of transcription factors Homo sapiens R-HSA-450341 | 6.19E-02 | 4.06E-01 | 5.79E+00 | 1.61E+01 | JUN;FOS |
| 149 | GP1b-IX-V activation signalling Homo sapiens R-HSA-430116 | 6.19E-02 | 4.06E-01 | 5.79E+00 | 1.61E+01 | FLNA;PIK3R1 |
| 150 | MyD88 dependent cascade initiated on endosome Homo sapiens R-HSA-975155 | 6.26E-02 | 4.06E-01 | 2.08E+00 | 5.78E+00 | MEF2A;MAP2K1;JUN;MAPKAPK3;PELI2;TAB2;FOS |
| 151 | Toll Like Receptor 7/8 (TLR7/8) Cascade Homo sapiens R-HSA-168181 | 6.26E-02 | 4.06E-01 | 2.08E+00 | 5.78E+00 | MEF2A;MAP2K1;JUN;MAPKAPK3;PELI2;TAB2;FOS |
| 152 | Signaling by FGFR3 Homo sapiens R-HSA-5654741 | 6.30E-02 | 4.06E-01 | 1.49E+00 | 4.12E+00 | DUSP2;MAP2K1;AHCYL1;CDKN1B;EGF;PDGFA;PIK3R2;PIK3R1;IL17RD;FOXO3;GRIN2B;DUSP9;FOXO1;TNRC6C;PRKAR1B;FGF9;ERBB4;NEFL;FGFR3;FGFR2 |
| 153 | NGF signalling via TRKA from the plasma membrane Homo sapiens R-HSA-187037 | 6.37E-02 | 4.07E-01 | 1.46E+00 | 4.01E+00 | MEF2A;DUSP2;MAP2K1;AHCYL1;CDKN1B;EGF;PDGFA;PIK3R2;PIK3R1;IL17RD;FOXO3;GRIN2B;DUSP9;FOXO1;TNRC6C;PRKAR1B;FGF9;MAPKAPK3;ERBB4;NEFL;FGFR3;FGFR2 |
| 154 | Fc epsilon receptor (FCERI) signaling Homo sapiens R-HSA-2454202 | 6.45E-02 | 4.10E-01 | 1.44E+00 | 3.95E+00 | JUN;DUSP2;MAP2K1;CDKN1B;AHCYL1;EGF;PDGFA;PIK3R2;FOS;PIK3R1;FOXO3;IL17RD;GRIN2B;DUSP9;FOXO1;TNRC6C;FGF9;ERBB4;PLCG2;NEFL;TAB2;FGFR3;FGFR2 |

| | | | | | | |
|---|---|---|---|---|---|---|
| 155 | Defective B3GALTL causes Peters-plus syndrome (PpS) Homo sapiens R-HSA-5083635 | 6.57E-02 | 4.10E-01 | 2.81E+00 | 7.66E+00 | ADAMTS16;ADAMTS5;ADAMTS15;ADAMTSL3 |
| 156 | NCAM1 interactions Homo sapiens R-HSA-419037 | 6.57E-02 | 4.10E-01 | 2.81E+00 | 7.66E+00 | COL4A2;COL4A1;COL6A1;COL9A2 |
| 157 | Cell junction organization Homo sapiens R-HSA-446728 | 6.58E-02 | 4.10E-01 | 2.06E+00 | 5.60E+00 | CLDN10;CDH4;CDH3;CADM1;CLDN19;FLNA;CTNNB1 |
| 158 | Toll Like Receptor 4 (TLR4) Cascade Homo sapiens R-HSA-166016 | 6.71E-02 | 4.15E-01 | 1.85E+00 | 5.00E+00 | MEF2A;MAP2K1;JUN;SOCS1;MAPKAPK3;PELI2;PLCG2;TAB2;FOS |
| 159 | Dopamine Neurotransmitter Release Cycle Homo sapiens R-HSA-212676 | 6.79E-02 | 4.15E-01 | 3.48E+00 | 9.36E+00 | CPLX1;VAMP2;PPFIA3 |
| 160 | RAF-independent MAPK1/3 activation Homo sapiens R-HSA-112409 | 6.79E-02 | 4.15E-01 | 3.48E+00 | 9.36E+00 | DUSP2;MAP2K1;DUSP9 |
| 161 | Platelet degranulation Homo sapiens R-HSA-114608 | 7.02E-02 | 4.27E-01 | 1.92E+00 | 5.09E+00 | CFD;EGF;ALB;PDGFA;FLNA;QSOX1;KNG1;SOD1 |
| 162 | O-glycosylation of TSR domain-containing proteins Homo sapiens R-HSA-5173214 | 7.11E-02 | 4.30E-01 | 2.73E+00 | 7.21E+00 | ADAMTS16;ADAMTS5;ADAMTS15;ADAMTSL3 |
| 163 | Toll Like Receptor 9 (TLR9) Cascade Homo sapiens R-HSA-168138 | 7.26E-02 | 4.34E-01 | 2.01E+00 | 5.26E+00 | MEF2A;MAP2K1;JUN;MAPKAPK3;PELI2;TAB2;FOS |
| 164 | Transport of organic anions Homo sapiens R-HSA-879518 | 7.37E-02 | 4.34E-01 | 5.15E+00 | 1.34E+01 | ALB;SLCO2A1 |
| 165 | Interleukin-7 signaling Homo sapiens R-HSA-1266695 | 7.37E-02 | 4.34E-01 | 5.15E+00 | 1.34E+01 | PIK3R2;PIK3R1 |

| | | | | | | |
|---|---|---|---|---|---|---|
| 166 | Apoptotic cleavage of cell adhesion proteins Homo sapiens R-HSA-351906 | 7.37E-02 | 4.34E-01 | 5.15E+00 | 1.34E+01 | CTNNB1;DSG2 |
| 167 | Glutamate Neurotransmitter Release Cycle Homo sapiens R-HSA-210500 | 7.53E-02 | 4.41E-01 | 3.31E+00 | 8.57E+00 | CPLX1;VAMP2;PPFIA3 |
| 168 | Downstream signal transduction Homo sapiens R-HSA-186763 | 7.60E-02 | 4.42E-01 | 1.45E+00 | 3.74E+00 | DUSP2;MAP2K1;AHCYL1;CDKN1B;EGF;PDGFA;PIK3R2;PIK3R1;IL17RD;FOXO3;GRIN2B;DUSP9;FOXO1;TNRC6C;PRKAR1B;FGF9;ERBB4;NEFL;FGFR3;FGFR2 |
| 169 | Collagen degradation Homo sapiens R-HSA-1442490 | 7.68E-02 | 4.42E-01 | 2.65E+00 | 6.80E+00 | COL18A1;COL26A1;COL9A2;CTSD |
| 170 | Dual Incision in GG-NER Homo sapiens R-HSA-5696400 | 7.68E-02 | 4.42E-01 | 2.65E+00 | 6.80E+00 | XPA;RPA1;GTF2H2;GTF2H5 |
| 171 | Signaling by NOTCH1 Homo sapiens R-HSA-1980143 | 7.77E-02 | 4.42E-01 | 2.11E+00 | 5.39E+00 | TLE3;NOTCH1;JAG1;MAML2;TBL1XR1;MIB1 |
| 172 | TRAF6 Mediated Induction of proinflammatory cytokines Homo sapiens R-HSA-168180 | 7.77E-02 | 4.42E-01 | 2.11E+00 | 5.39E+00 | MEF2A;MAP2K1;JUN;MAPKAPK3;TAB2;FOS |
| 173 | Transcriptional activation of mitochondrial biogenesis Homo sapiens R-HSA-2151201 | 8.27E-02 | 4.54E-01 | 2.58E+00 | 6.42E+00 | RXRA;TBL1XR1;CYCS;PPARGC1A |
| 174 | Constitutive Signaling by AKT1 E17K in Cancer Homo sapiens R-HSA-5674400 | 8.30E-02 | 4.54E-01 | 3.16E+00 | 7.87E+00 | CDKN1B;FOXO3;FOXO1 |
| 175 | IRS-mediated signalling Homo sapiens R-HSA-112399 | 8.34E-02 | 4.54E-01 | 1.48E+00 | 3.68E+00 | DUSP2;MAP2K1;CAB39;EGF;PDGFA;PIK3R2;PIK3R1;IL17RD;PRKAB1;GRIN2B;DUSP9;FGF9;ERBB4;RRAGD;NEFL;FGFR3;FGFR2 |
| 176 | Signaling by SCF-KIT Homo sapiens R-HSA-1433557 | 8.40E-02 | 4.54E-01 | 1.45E+00 | 3.58E+00 | DUSP2;MAP2K1;CDKN1B;EGF;PDGFA;PIK3R2;PIK3R1;IL17RD;FOXO3;GRIN2B;DUSP9;FOXO1;TNRC6C;SOCS1;FGF9;ERBB4;NEFL;FGFR3;FGFR2 |

| | | | | | | |
|---|---|---|---|---|---|---|
| 177 | Defective EXT2 causes exostoses 2 Homo sapiens R-HSA-3656237 | 8.60E-02 | 4.54E-01 | 4.63E+00 | 1.14E+01 | SDC4;GPC1 |
| 178 | Defective EXT1 causes exostoses 1, TRPS2 and CHDS Homo sapiens R-HSA-3656253 | 8.60E-02 | 4.54E-01 | 4.63E+00 | 1.14E+01 | SDC4;GPC1 |
| 179 | FGFR3 mutant receptor activation Homo sapiens R-HSA-2033514 | 8.60E-02 | 4.54E-01 | 4.63E+00 | 1.14E+01 | FGF9;FGFR3 |
| 180 | Signaling by activated point mutants of FGFR3 Homo sapiens R-HSA-1839130 | 8.60E-02 | 4.54E-01 | 4.63E+00 | 1.14E+01 | FGF9;FGFR3 |
| 181 | O-linked glycosylation Homo sapiens R-HSA-5173105 | 8.66E-02 | 4.54E-01 | 1.82E+00 | 4.46E+00 | ADAMTS16;ADAMTS15;ADAMTS5;B3GNT9;ADAMTSL3;ST3GAL4;GCNT4;MUC20 |
| 182 | Response to elevated platelet cytosolic Ca2+ Homo sapiens R-HSA-76005 | 8.66E-02 | 4.54E-01 | 1.82E+00 | 4.46E+00 | CFD;EGF;ALB;PDGFA;FLNA;QSOX1;KNG1;SOD1 |
| 183 | Constitutive Signaling by NOTCH1 HD+PEST Domain Mutants Homo sapiens R-HSA-2894862 | 8.67E-02 | 4.54E-01 | 2.23E+00 | 5.46E+00 | NOTCH1;JAG1;MAML2;TBL1XR1;MIB1 |
| 184 | Signaling by NOTCH1 in Cancer Homo sapiens R-HSA-2644603 | 8.67E-02 | 4.54E-01 | 2.23E+00 | 5.46E+00 | NOTCH1;JAG1;MAML2;TBL1XR1;MIB1 |
| 185 | Signaling by NOTCH1 PEST Domain Mutants in Cancer Homo sapiens R-HSA-2644602 | 8.67E-02 | 4.54E-01 | 2.23E+00 | 5.46E+00 | NOTCH1;JAG1;MAML2;TBL1XR1;MIB1 |
| 186 | Constitutive Signaling by NOTCH1 PEST Domain Mutants Homo sapiens R-HSA-2644606 | 8.67E-02 | 4.54E-01 | 2.23E+00 | 5.46E+00 | NOTCH1;JAG1;MAML2;TBL1XR1;MIB1 |

| | | | | | | |
|---|---|---|---|---|---|---|
| 187 | Signaling by NOTCH1 HD+PEST Domain Mutants in Cancer Homo sapiens R-HSA-2894858 | 8.67E-02 | 4.54E-01 | 2.23E+00 | 5.46E+00 | NOTCH1;JAG1;MAML2;TBL1XR1;MIB1 |
| 188 | NCAM signaling for neurite out-growth Homo sapiens R-HSA-375165 | 8.80E-02 | 4.58E-01 | 1.49E+00 | 3.62E+00 | DUSP2;MAP2K1;EGF;PDGFA;IL17RD;GRIN2B;DUSP9;FGF9;COL4A2;COL4A1;ERBB4;COL6A1;NEFL;COL9A2;FGFR3;FGFR2 |
| 189 | Insulin receptor signalling cascade Homo sapiens R-HSA-74751 | 8.97E-02 | 4.64E-01 | 1.47E+00 | 3.53E+00 | DUSP2;MAP2K1;CAB39;EGF;PDGFA;PIK3R2;PIK3R1;IL17RD;PRKAB1;GRIN2B;DUSP9;FGF9;ERBB4;RRAGD;NEFL;FGFR3;FGFR2 |
| 190 | Attenuation phase Homo sapiens R-HSA-3371568 | 9.11E-02 | 4.66E-01 | 3.02E+00 | 7.24E+00 | HSPA8;HSBP1;HSPA1B |
| 191 | IGF1R signaling cascade Homo sapiens R-HSA-2428924 | 9.18E-02 | 4.66E-01 | 1.46E+00 | 3.49E+00 | DUSP2;MAP2K1;CAB39;EGF;PDGFA;PIK3R2;PIK3R1;IL17RD;PRKAB1;GRIN2B;DUSP9;FGF9;ERBB4;RRAGD;NEFL;FGFR3;FGFR2 |
| 192 | Signaling by Type 1 Insulin-like Growth Factor 1 Receptor (IGF1R) Homo sapiens R-HSA-2404192 | 9.18E-02 | 4.66E-01 | 1.46E+00 | 3.49E+00 | DUSP2;MAP2K1;CAB39;EGF;PDGFA;PIK3R2;PIK3R1;IL17RD;PRKAB1;GRIN2B;DUSP9;FGF9;ERBB4;RRAGD;NEFL;FGFR3;FGFR2 |
| 193 | IRS-related events triggered by IGF1R Homo sapiens R-HSA-2428928 | 9.18E-02 | 4.66E-01 | 1.46E+00 | 3.49E+00 | DUSP2;MAP2K1;CAB39;EGF;PDGFA;PIK3R2;PIK3R1;IL17RD;PRKAB1;GRIN2B;DUSP9;FGF9;ERBB4;RRAGD;NEFL;FGFR3;FGFR2 |
| 194 | Activated TLR4 signalling Homo sapiens R-HSA-166054 | 9.37E-02 | 4.70E-01 | 1.79E+00 | 4.23E+00 | MEF2A;MAP2K1;JUN;SOCS1;MAPKAPK3;PELI2;TAB2;FOS |
| 195 | Retinoid metabolism and transport Homo sapiens R-HSA-975634 | 9.50E-02 | 4.70E-01 | 2.44E+00 | 5.75E+00 | LRP1;SDC4;GPC1;APOE |
| 196 | Intrinsic Pathway for Apoptosis Homo sapiens R-HSA-109606 | 9.50E-02 | 4.70E-01 | 2.44E+00 | 5.75E+00 | CYCS;DYNLL1;DYNLL2;BCL2L1 |
| 197 | Initiation of Nuclear Envelope Reformation Homo sapiens R-HSA-2995383 | 9.90E-02 | 4.70E-01 | 4.21E+00 | 9.74E+00 | BANF1;EMD |

| | | | | | | |
|---|---|---|---|---|---|---|
| 198 | Nuclear Envelope Reassembly Homo sapiens R-HSA-2995410 | 9.90E-02 | 4.70E-01 | 4.21E+00 | 9.74E+00 | BANF1;EMD |
| 199 | NICD traffics to nucleus Homo sapiens R-HSA-157052 | 9.90E-02 | 4.70E-01 | 4.21E+00 | 9.74E+00 | NOTCH1;MAML2 |
| 200 | Notch-HLH transcription pathway Homo sapiens R-HSA-350054 | 9.90E-02 | 4.70E-01 | 4.21E+00 | 9.74E+00 | NOTCH1;MAML2 |
| 201 | FGFR2c ligand binding and activation Homo sapiens R-HSA-190375 | 9.90E-02 | 4.70E-01 | 4.21E+00 | 9.74E+00 | FGF9;FGFR2 |
| 202 | FGFR3c ligand binding and activation Homo sapiens R-HSA-190372 | 9.90E-02 | 4.70E-01 | 4.21E+00 | 9.74E+00 | FGF9;FGFR3 |
| 203 | FGFR3 ligand binding and activation Homo sapiens R-HSA-190239 | 9.90E-02 | 4.70E-01 | 4.21E+00 | 9.74E+00 | FGF9;FGFR3 |
| 204 | ERBB2 Activates PTK6 Signaling Homo sapiens R-HSA-8847993 | 9.90E-02 | 4.70E-01 | 4.21E+00 | 9.74E+00 | EGF;ERBB4 |
| 205 | RNA Pol II CTD phosphorylation and interaction with CE Homo sapiens R-HSA-167160 | 9.95E-02 | 4.70E-01 | 2.90E+00 | 6.69E+00 | POLR2A;GTF2H2;GTF2H5 |
| 206 | RNA Pol II CTD phosphorylation and interaction with CE Homo sapiens R-HSA-77075 | 9.95E-02 | 4.70E-01 | 2.90E+00 | 6.69E+00 | POLR2A;GTF2H2;GTF2H5 |
| 207 | Pyruvate metabolism Homo sapiens R-HSA-70268 | 9.95E-02 | 4.70E-01 | 2.90E+00 | 6.69E+00 | RXRA;MPC1;MPC2 |
| 208 | Formation of Incision Complex in GG-NER Homo sapiens R-HSA-5696395 | 1.02E-01 | 4.76E-01 | 2.38E+00 | 5.44E+00 | XPA;RPA1;GTF2H2;GTF2H5 |

| | | | | | | |
|---|---|---|---|---|---|---|
| 209 | Signaling by FGFR2 in disease Homo sapiens R-HSA-5655253 | 1.02E-01 | 4.76E-01 | 2.38E+00 | 5.44E+00 | POLR2A;FGF9;PIK3R1;FGFR2 |
| 210 | Downstream signaling events of B Cell Receptor (BCR) Homo sapiens R-HSA-1168372 | 1.03E-01 | 4.82E-01 | 1.55E+00 | 3.52E+00 | TNRC6C;CDKN1B;FGF9;ERBB4;EGF;PDGFA;PIK3R2;PIK3R1;FOXO3;FGFR3;FOXO1;FGFR2 |
| 211 | Signaling by the B Cell Receptor (BCR) Homo sapiens R-HSA-983705 | 1.06E-01 | 4.92E-01 | 1.49E+00 | 3.34E+00 | AHCYL1;CDKN1B;EGF;PDGFA;PIK3R2;PIK3R1;FOXO3;FOXO1;TNRC6C;FGF9;ERBB4;PLCG2;FGFR3;FGFR2 |
| 212 | TP53 Regulates Transcription of DNA Repair Genes Homo sapiens R-HSA-6796648 | 1.08E-01 | 4.93E-01 | 2.07E+00 | 4.61E+00 | JUN;POLR2A;FOS;GTF2H2;GTF2H5 |
| 213 | Energy dependent regulation of mTOR by LKB1-AMPK Homo sapiens R-HSA-380972 | 1.08E-01 | 4.93E-01 | 2.78E+00 | 6.19E+00 | CAB39;RRAGD;PRKAB1 |
| 214 | Phospholipase C-mediated cascade; FGFR3 Homo sapiens R-HSA-5654227 | 1.08E-01 | 4.93E-01 | 2.32E+00 | 5.16E+00 | AHCYL1;FGF9;PRKAR1B;FGFR3 |
| 215 | Inositol phosphate metabolism Homo sapiens R-HSA-1483249 | 1.08E-01 | 4.93E-01 | 2.32E+00 | 5.16E+00 | IMPA2;PLCG2;NUDT3;PLCD1 |
| 216 | Early Phase of HIV Life Cycle Homo sapiens R-HSA-162594 | 1.12E-01 | 5.05E-01 | 3.86E+00 | 8.44E+00 | HMGA1;BANF1 |
| 217 | Receptor-ligand binding initiates the second proteolytic cleavage of Notch receptor Homo sapiens R-HSA-156988 | 1.12E-01 | 5.05E-01 | 3.86E+00 | 8.44E+00 | NOTCH1;JAG1 |
| 218 | Activation of Rac Homo sapiens R-HSA-428540 | 1.12E-01 | 5.05E-01 | 3.86E+00 | 8.44E+00 | GPC1;PAK4 |
| 219 | COPI-independent Golgi-to-ER retrograde traffic Homo sapiens R-HSA-6811436 | 1.17E-01 | 5.14E-01 | 2.67E+00 | 5.74E+00 | DYNLL1;BICD1;DYNLL2 |

| | | | | | | |
|---|---|---|---|---|---|---|
| 220 | HSF1 activation Homo sapiens R-HSA-3371511 | 1.17E-01 | 5.14E-01 | 2.67E+00 | 5.74E+00 | HSBP1;RPA1;HSPA1B |
| 221 | mRNA Capping Homo sapiens R-HSA-72086 | 1.17E-01 | 5.14E-01 | 2.67E+00 | 5.74E+00 | POLR2A;GTF2H2;GTF2H5 |
| 222 | Myogenesis Homo sapiens R-HSA-525793 | 1.17E-01 | 5.14E-01 | 2.67E+00 | 5.74E+00 | MEF2A;CDH4;CTNNB1 |
| 223 | CDO in myogenesis Homo sapiens R-HSA-375170 | 1.17E-01 | 5.14E-01 | 2.67E+00 | 5.74E+00 | MEF2A;CDH4;CTNNB1 |
| 224 | VEGFA-VEGFR2 Pathway Homo sapiens R-HSA-4420097 | 1.18E-01 | 5.15E-01 | 1.39E+00 | 2.96E+00 | CYFIP2;DUSP2;MAP2K1;AHCYL1;EGF;PDGFA;PIK3R2;PIK3R1;IL17RD;GRIN2B;DUSP9;FGF9;MAPKAPK3;ERBB4;NEFL;CTNNB1;FGFR3;FGFR2 |
| 225 | Peptide hormone metabolism Homo sapiens R-HSA-2980736 | 1.19E-01 | 5.15E-01 | 1.86E+00 | 3.95E+00 | TCF7L2;ANPEP;KIF5A;CTNNB1;CTSD;VAMP2 |
| 226 | Dual incision in TC-NER Homo sapiens R-HSA-6782135 | 1.20E-01 | 5.15E-01 | 2.00E+00 | 4.25E+00 | POLR2A;XPA;RPA1;GTF2H2;GTF2H5 |
| 227 | Circadian Clock Homo sapiens R-HSA-400253 | 1.20E-01 | 5.15E-01 | 2.00E+00 | 4.25E+00 | SREBF1;RXRA;TBL1XR1;FBXL3;PPARGC1A |
| 228 | Signalling by NGF Homo sapiens R-HSA-166520 | 1.25E-01 | 5.15E-01 | 1.31E+00 | 2.73E+00 | MEF2A;DUSP2;MAP2K1;CDKN1B;AHCYL1;EGF;PDGFA;PIK3R2;PIK3R1;FOXO3;IL17RD;GRIN2B;DUSP9;FOXO1;FGD3;TNRC6C;OBSCN;PRKAR1B;FGF9;MAPKAPK3;ERBB4;NEFL;FGFR3;FGFR2 |
| 229 | Axon guidance Homo sapiens R-HSA-422475 | 1.26E-01 | 5.15E-01 | 1.29E+00 | 2.67E+00 | PDGFA;EFNA5;ACTB;ACTG1;ABLIM1;FGF9;CHL1;ERBB4;GPC1;NEFL;SCN3B;PAK4;HSPA8;DUSP2;MAP2K1;EGF;IL17RD;GRIN2B;DUSP9;RHOB;COL4A2;COL4A1;CSNK2B;COL6A1;COL9A2;FGFR3;FGFR2 |
| 230 | Depolymerisation of the Nuclear Lamina Homo sapiens R-HSA-4419969 | 1.26E-01 | 5.15E-01 | 3.56E+00 | 7.37E+00 | LPIN1;EMD |
| 231 | ERBB2 Regulates Cell Motility Homo sapiens R-HSA-6785631 | 1.26E-01 | 5.15E-01 | 3.56E+00 | 7.37E+00 | EGF;ERBB4 |
| 232 | Signaling by EGFRvIII in Cancer Homo sapiens R-HSA-5637812 | 1.26E-01 | 5.15E-01 | 3.56E+00 | 7.37E+00 | EGF;PIK3R1 |

| | | | | | | |
|---|---|---|---|---|---|---|
| 233 | Constitutive Signaling by EGFRvIII Homo sapiens R-HSA-5637810 | 1.26E-01 | 5.15E-01 | 3.56E+00 | 7.37E+00 | EGF;PIK3R1 |
| 234 | CREB phosphorylation through the activation of CaMKII Homo sapiens R-HSA-442729 | 1.26E-01 | 5.15E-01 | 3.56E+00 | 7.37E+00 | NEFL;GRIN2B |
| 235 | Eicosanoid ligand-binding receptors Homo sapiens R-HSA-391903 | 1.26E-01 | 5.15E-01 | 3.56E+00 | 7.37E+00 | PTGFR;PTGER3 |
| 236 | Recycling of bile acids and salts Homo sapiens R-HSA-159418 | 1.26E-01 | 5.15E-01 | 3.56E+00 | 7.37E+00 | RXRA;ALB |
| 237 | Synthesis of bile acids and bile salts via 27-hydroxycholesterol Homo sapiens R-HSA-193807 | 1.26E-01 | 5.15E-01 | 3.56E+00 | 7.37E+00 | CYP27A1;RXRA |
| 238 | Molecules associated with elastic fibres Homo sapiens R-HSA-2129379 | 1.26E-01 | 5.15E-01 | 2.58E+00 | 5.33E+00 | BMP4;EFEMP2;LTBP4 |
| 239 | Activated NOTCH1 Transmits Signal to the Nucleus Homo sapiens R-HSA-2122948 | 1.26E-01 | 5.15E-01 | 2.58E+00 | 5.33E+00 | NOTCH1;JAG1;MIB1 |
| 240 | MAPK targets/ Nuclear events mediated by MAP kinases Homo sapiens R-HSA-450282 | 1.26E-01 | 5.15E-01 | 2.58E+00 | 5.33E+00 | MEF2A;JUN;FOS |
| 241 | Toll-Like Receptors Cascades Homo sapiens R-HSA-168898 | 1.28E-01 | 5.19E-01 | 1.60E+00 | 3.28E+00 | MEF2A;MAP2K1;JUN;SOCS1;MAPKAPK3;PELI2;PLCG2;TAB2;FOS |
| 242 | NOTCH1 Intracellular Domain Regulates Transcription Homo sapiens R-HSA-2122947 | 1.29E-01 | 5.24E-01 | 2.16E+00 | 4.41E+00 | TLE3;NOTCH1;MAML2;TBL1XR1 |

| | | | | | | |
|---|---|---|---|---|---|---|
| 243 | MAPK family signaling cascades Homo sapiens R-HSA-5683057 | 1.33E-01 | 5.36E-01 | 1.39E+00 | 2.80E+00 | JUN;DUSP2;MAP2K1;EGF;PDGFA;IL17RD;FOXO3;GRIN2B;DUSP9;FOXO1;TNRC6C;FGF9;ERBB4;NEFL;FGFR3;FGFR2 |
| 244 | Metabolism of polyamines Homo sapiens R-HSA-351202 | 1.35E-01 | 5.36E-01 | 1.78E+00 | 3.57E+00 | GATM;SLC6A8;CKB;SAT1;ASS1;AGMAT |
| 245 | MHC class II antigen presentation Homo sapiens R-HSA-2132295 | 1.36E-01 | 5.36E-01 | 1.69E+00 | 3.38E+00 | KIF26A;KIF5A;CTSF;DYNLL1;CTSD;DYNLL2;CTSC |
| 246 | Disassembly of the destruction complex and recruitment of AXIN to the membrane Homo sapiens R-HSA-4641262 | 1.36E-01 | 5.36E-01 | 2.48E+00 | 4.96E+00 | FZD1;LRP5;CTNNB1 |
| 247 | FGFR1 mutant receptor activation Homo sapiens R-HSA-1839124 | 1.36E-01 | 5.36E-01 | 2.48E+00 | 4.96E+00 | FGF9;CNTRL;PIK3R1 |
| 248 | Inwardly rectifying K+ channels Homo sapiens R-HSA-1296065 | 1.36E-01 | 5.36E-01 | 2.48E+00 | 4.96E+00 | KCNJ10;KCNJ15;KCNJ1 |
| 249 | Signaling by VEGF Homo sapiens R-HSA-194138 | 1.39E-01 | 5.42E-01 | 1.35E+00 | 2.67E+00 | CYFIP2;DUSP2;MAP2K1;AHCYL1;EGF;PDGFA;PIK3R2;PIK3R1;IL17RD;GRIN2B;DUSP9;FGF9;MAPKAPK3;ERBB4;NEFL;CTNNB1;FGFR3;FGFR2 |
| 250 | GRB2 events in ERBB2 signaling Homo sapiens R-HSA-1963640 | 1.41E-01 | 5.42E-01 | 3.31E+00 | 6.49E+00 | EGF;ERBB4 |
| 251 | RHO GTPases activate CIT Homo sapiens R-HSA-5625900 | 1.41E-01 | 5.42E-01 | 3.31E+00 | 6.49E+00 | CDKN1B;RHOB |
| 252 | Metabolism of Angiotensinogen to Angiotensins Homo sapiens R-HSA-2022377 | 1.41E-01 | 5.42E-01 | 3.31E+00 | 6.49E+00 | ANPEP;CTSD |
| 253 | Metabolism of folate and pterines Homo sapiens R-HSA-196757 | 1.41E-01 | 5.42E-01 | 3.31E+00 | 6.49E+00 | ALDH1L1;SLC46A1 |

| | | | | | | |
|---|---|---|---|---|---|---|
| 254 | Collagen formation Homo sapiens R-HSA-1474290 | 1.41E-01 | 5.42E-01 | 1.76E+00 | 3.45E+00 | COL18A1;COL26A1;COL4A2;COL4A1;COL6A1;COL9A2 |
| 255 | Interleukin receptor SHC signaling Homo sapiens R-HSA-912526 | 1.41E-01 | 5.42E-01 | 1.41E+00 | 2.76E+00 | DUSP2;MAP2K1;EGF;PDGFA;PIK3R2;PIK3R1;IL17RD;GRIN2B;DUSP9;FGF9;ERBB4;NEFL;FGFR3;FGFR2 |
| 256 | Phospholipase C-mediated cascade; FGFR2 Homo sapiens R-HSA-5654221 | 1.44E-01 | 5.52E-01 | 2.06E+00 | 3.99E+00 | AHCYL1;FGF9;PRKAR1B;FGFR2 |
| 257 | Regulation of beta-cell development Homo sapiens R-HSA-186712 | 1.45E-01 | 5.52E-01 | 2.40E+00 | 4.62E+00 | NOTCH1;HNF4A;FOXO1 |
| 258 | Signaling by NOTCH2 Homo sapiens R-HSA-1980145 | 1.45E-01 | 5.52E-01 | 2.40E+00 | 4.62E+00 | JAG1;MAML2;MIB1 |
| 259 | Generic Transcription Pathway Homo sapiens R-HSA-212436 | 1.47E-01 | 5.56E-01 | 1.21E+00 | 2.32E+00 | GPI;COX7B;CDKN1B;NOTCH1;COX16;MAML2;RORC;NEDD4L;NDRG1;NR3C2;RXRA;POLR2A;PCBP4;PLAGL1;HNF4A;ZNF549;ZNF426;ZNF304;JUNB;ZNF124;COX8A;JUN;BRPF1;NDUFA4;GADD45A;RPA1;ARID3A;FOS;GTF2H2;PRKAB1;GTF2H5;SMAD7;TNRC6C;TBL1XR1;ZNF615;CSNK2B;RRAGD;CYCS;ZNF556;ZNF772 |
| 260 | Signaling by Insulin receptor Homo sapiens R-HSA-74752 | 1.50E-01 | 5.63E-01 | 1.34E+00 | 2.55E+00 | DUSP2;MAP2K1;CAB39;EGF;PDGFA;PIK3R2;PIK3R1;IL17RD;PRKAB1;GRIN2B;DUSP9;FGF9;ERBB4;RRAGD;NEFL;FGFR3;FGFR2 |
| 261 | Chondroitin sulfate/dermatan sulfate metabolism Homo sapiens R-HSA-1793185 | 1.52E-01 | 5.63E-01 | 2.02E+00 | 3.79E+00 | CHST9;SDC4;HYAL1;GPC1 |
| 262 | Activated point mutants of FGFR2 Homo sapiens R-HSA-2033519 | 1.55E-01 | 5.63E-01 | 3.09E+00 | 5.76E+00 | FGF9;FGFR2 |
| 263 | activated TAK1 mediates p38 MAPK activation Homo sapiens R-HSA-450302 | 1.55E-01 | 5.63E-01 | 3.09E+00 | 5.76E+00 | MAPKAPK3;TAB2 |
| 264 | Ras activation uopn Ca2+ infux through NMDA receptor Homo sapiens R-HSA-442982 | 1.55E-01 | 5.63E-01 | 3.09E+00 | 5.76E+00 | NEFL;GRIN2B |

| | | | | | | |
|---|---|---|---|---|---|---|
| 265 | Unblocking of NMDA receptor, glutamate binding and activation Homo sapiens R-HSA-438066 | 1.55E-01 | 5.63E-01 | 3.09E+00 | 5.76E+00 | NEFL;GRIN2B |
| 266 | Inflammasomes Homo sapiens R-HSA-622312 | 1.55E-01 | 5.63E-01 | 3.09E+00 | 5.76E+00 | TXNIP;BCL2L1 |
| 267 | Sphingolipid de novo biosynthesis Homo sapiens R-HSA-1660661 | 1.55E-01 | 5.63E-01 | 2.32E+00 | 4.31E+00 | CERS4;SGMS1;SPTLC2 |
| 268 | Formation of the Early Elongation Complex Homo sapiens R-HSA-113418 | 1.55E-01 | 5.63E-01 | 2.32E+00 | 4.31E+00 | POLR2A;GTF2H2;GTF2H5 |
| 269 | Formation of the HIV-1 Early Elongation Complex Homo sapiens R-HSA-167158 | 1.55E-01 | 5.63E-01 | 2.32E+00 | 4.31E+00 | POLR2A;GTF2H2;GTF2H5 |
| 270 | FGFR2 mutant receptor activation Homo sapiens R-HSA-1839126 | 1.55E-01 | 5.63E-01 | 2.32E+00 | 4.31E+00 | POLR2A;FGF9;FGFR2 |
| 271 | Metabolism of fat-soluble vitamins Homo sapiens R-HSA-6806667 | 1.60E-01 | 5.72E-01 | 1.97E+00 | 3.61E+00 | LRP1;SDC4;GPC1;APOE |
| 272 | mRNA 3'-end processing Homo sapiens R-HSA-72187 | 1.60E-01 | 5.72E-01 | 1.97E+00 | 3.61E+00 | NUDT21;SRSF3;SRSF5;SRSF6 |
| 273 | Post-Elongation Processing of Intron-Containing pre-mRNA Homo sapiens R-HSA-112296 | 1.60E-01 | 5.72E-01 | 1.97E+00 | 3.61E+00 | NUDT21;SRSF3;SRSF5;SRSF6 |
| 274 | Nuclear Receptor transcription pathway Homo sapiens R-HSA-383280 | 1.60E-01 | 5.72E-01 | 1.97E+00 | 3.61E+00 | RXRA;HNF4A;RORC;NR3C2 |
| 275 | Metabolism of vitamins and cofactors Homo sapiens R-HSA-196854 | 1.62E-01 | 5.76E-01 | 1.50E+00 | 2.74E+00 | ALDH1L1;SLC46A1;SDC4;LRP1;MMAB;NAPRT;GPC1;APOE;SLC19A2 |

| | | | | | | |
|---|---|---|---|---|---|---|
| 276 | Interleukin-2 signaling Homo sapiens R-HSA-451927 | 1.64E-01 | 5.76E-01 | 1.37E+00 | 2.47E+00 | DUSP2;MAP2K1;EGF;PDGFA;PIK3R2;PIK3R1;IL17RD;GRIN2B;DUSP9;FGF9;ERBB4;NEFL;FGFR3;FGFR2 |
| 277 | HSF1-dependent transactivation Homo sapiens R-HSA-3371571 | 1.66E-01 | 5.76E-01 | 2.24E+00 | 4.03E+00 | HSPA8;HSBP1;HSPA1B |
| 278 | Signaling by WNT in cancer Homo sapiens R-HSA-4791275 | 1.66E-01 | 5.76E-01 | 2.24E+00 | 4.03E+00 | TCF7L2;LRP5;CTNNB1 |
| 279 | EGFR interacts with phospholipase C-gamma Homo sapiens R-HSA-212718 | 1.66E-01 | 5.76E-01 | 2.24E+00 | 4.03E+00 | AHCYL1;PRKAR1B;EGF |
| 280 | Transmission across Chemical Synapses Homo sapiens R-HSA-112315 | 1.68E-01 | 5.76E-01 | 1.40E+00 | 2.50E+00 | HSPA8;KCNJ10;EPB41L1;CACNA2D1;SLC22A2;NAAA;KCNJ15;NEFL;GRIN2B;CPLX1;VAMP2;PPFIA3 |
| 281 | Miscellaneous transport and binding events Homo sapiens R-HSA-5223345 | 1.70E-01 | 5.76E-01 | 2.90E+00 | 5.13E+00 | NIPAL1;ADD3 |
| 282 | Pre-NOTCH Processing in Golgi Homo sapiens R-HSA-1912420 | 1.70E-01 | 5.76E-01 | 2.90E+00 | 5.13E+00 | NOTCH1;ST3GAL4 |
| 283 | SHC-mediated cascade:FGFR3 Homo sapiens R-HSA-5654704 | 1.70E-01 | 5.76E-01 | 2.90E+00 | 5.13E+00 | FGF9;FGFR3 |
| 284 | Signaling by cytosolic FGFR1 fusion mutants Homo sapiens R-HSA-1839117 | 1.70E-01 | 5.76E-01 | 2.90E+00 | 5.13E+00 | CNTRL;PIK3R1 |
| 285 | SHC1 events in ERBB2 signaling Homo sapiens R-HSA-1250196 | 1.70E-01 | 5.76E-01 | 2.90E+00 | 5.13E+00 | EGF;ERBB4 |
| 286 | Tie2 Signaling Homo sapiens R-HSA-210993 | 1.70E-01 | 5.76E-01 | 2.90E+00 | 5.13E+00 | PIK3R2;PIK3R1 |
| 287 | Regulation of signaling by CBL Homo sapiens R-HSA-912631 | 1.70E-01 | 5.76E-01 | 2.90E+00 | 5.13E+00 | PIK3R2;PIK3R1 |

| | | | | | | |
|---|---|---|---|---|---|---|
| 288 | cGMP effects Homo sapiens R-HSA-418457 | 1.70E-01 | 5.76E-01 | 2.90E+00 | 5.13E+00 | KCNMA1;PDE3A |
| 289 | Cytosolic sulfonation of small molecules Homo sapiens R-HSA-156584 | 1.70E-01 | 5.76E-01 | 2.90E+00 | 5.13E+00 | SULT1A1;PAPSS2 |
| 290 | Lipid digestion, mobilization, and transport Homo sapiens R-HSA-73923 | 1.71E-01 | 5.76E-01 | 1.76E+00 | 3.10E+00 | FABP1;FABP3;ALB;APOE;MGLL |
| 291 | Bile acid and bile salt metabolism Homo sapiens R-HSA-194068 | 1.76E-01 | 5.76E-01 | 2.17E+00 | 3.78E+00 | CYP27A1;RXRA;ALB |
| 292 | Formation of TC-NER Pre-Incision Complex Homo sapiens R-HSA-6781823 | 1.76E-01 | 5.76E-01 | 1.89E+00 | 3.28E+00 | POLR2A;XPA;GTF2H2;GTF2H5 |
| 293 | GPVI-mediated activation cascade Homo sapiens R-HSA-114604 | 1.76E-01 | 5.76E-01 | 1.89E+00 | 3.28E+00 | PLCG2;PIK3R2;PIK3R1;RHOB |
| 294 | Processing of Capped Intron-Containing Pre-mRNA Homo sapiens R-HSA-72203 | 1.79E-01 | 5.76E-01 | 1.40E+00 | 2.41E+00 | NUDT21;HSPA8;POLR2A;HNRNPH1;PCBP1;PCBP2;SRSF3;SNRPB2;SRSF5;YBX1;SRSF6 |
| 295 | Innate Immune System Homo sapiens R-HSA-168249 | 1.80E-01 | 5.76E-01 | 1.18E+00 | 2.03E+00 | CYFIP2;CFD;AHCYL1;CDKN1B;PELI2;PDGFA;DEFB1;PIK3R2;PIK3R1;FOXO3;FOXO1;ACTB;ACTG1;LGALS3;SOCS1;FGF9;ERBB4;PCBP2;PLCG2;NEFL;MEF2A;JUN;DUSP2;MAP2K1;EGF;HLA-B;FOS;IL17RD;GRIN2B;DUSP9;TNRC6C;PRKAR1B;MAPKAPK3;TXNIP;CTNNB1;TAB2;FGFR3;FGFR2;BCL2L1 |
| 296 | Phase 1 - Functionalization of compounds Homo sapiens R-HSA-211945 | 1.82E-01 | 5.76E-01 | 1.62E+00 | 2.75E+00 | CYP27A1;RXRA;ADH1C;ALDH1B1;CMBL;ACSS1 |
| 297 | Assembly of collagen fibrils and other multimeric structures Homo sapiens R-HSA-2022090 | 1.85E-01 | 5.76E-01 | 1.85E+00 | 3.13E+00 | COL18A1;COL4A2;COL4A1;COL6A1 |

| | | | | | | |
|---|---|---|---|---|---|---|
| 298 | Defective B3GALT6 causes EDSP2 and SEMDJL1 Homo sapiens R-HSA-4420332 | 1.85E-01 | 5.76E-01 | 2.72E+00 | 4.60E+00 | SDC4;GPC1 |
| 299 | Defective B4GALT7 causes EDS, progeroid type Homo sapiens R-HSA-3560783 | 1.85E-01 | 5.76E-01 | 2.72E+00 | 4.60E+00 | SDC4;GPC1 |
| 300 | Defective B3GAT3 causes JDSSDHD Homo sapiens R-HSA-3560801 | 1.85E-01 | 5.76E-01 | 2.72E+00 | 4.60E+00 | SDC4;GPC1 |
| 301 | Signaling by FGFR2 IIIa TM Homo sapiens R-HSA-8851708 | 1.85E-01 | 5.76E-01 | 2.72E+00 | 4.60E+00 | POLR2A;FGFR2 |
| 302 | Constitutive Signaling by Ligand-Responsive EGFR Cancer Variants Homo sapiens R-HSA-1236382 | 1.85E-01 | 5.76E-01 | 2.72E+00 | 4.60E+00 | EGF;PIK3R1 |
| 303 | Signaling by Ligand-Responsive EGFR Variants in Cancer Homo sapiens R-HSA-5637815 | 1.85E-01 | 5.76E-01 | 2.72E+00 | 4.60E+00 | EGF;PIK3R1 |
| 304 | Signaling by EGFR in Cancer Homo sapiens R-HSA-1643713 | 1.85E-01 | 5.76E-01 | 2.72E+00 | 4.60E+00 | EGF;PIK3R1 |
| 305 | HDL-mediated lipid transport Homo sapiens R-HSA-194223 | 1.85E-01 | 5.76E-01 | 2.72E+00 | 4.60E+00 | ALB;APOE |
| 306 | Signaling by TGF-beta Receptor Complex Homo sapiens R-HSA-170834 | 1.85E-01 | 5.76E-01 | 1.70E+00 | 2.87E+00 | PPP1R15A;BAMBI;NEDD4L;JUNB;SMAD7 |
| 307 | RHO GTPases Activate WASPs and WAVEs Homo sapiens R-HSA-5663213 | 1.86E-01 | 5.76E-01 | 2.11E+00 | 3.54E+00 | CYFIP2;ACTB;ACTG1 |

| | | | | | | |
|---|---|---|---|---|---|---|
| 308 | Toxicity of botulinum toxin type D (BoNT/D) Homo sapiens R-HSA-5250955 | 1.91E-01 | 5.76E-01 | 5.79E+00 | 9.59E+00 | VAMP2 |
| 309 | Toxicity of botulinum toxin type F (BoNT/F) Homo sapiens R-HSA-5250981 | 1.91E-01 | 5.76E-01 | 5.79E+00 | 9.59E+00 | VAMP2 |
| 310 | Abacavir metabolism Homo sapiens R-HSA-2161541 | 1.91E-01 | 5.76E-01 | 5.79E+00 | 9.59E+00 | ADAL |
| 311 | Sodium-coupled phosphate cotransporters Homo sapiens R-HSA-427652 | 1.91E-01 | 5.76E-01 | 5.79E+00 | 9.59E+00 | SLC20A1 |
| 312 | Catabolism of glucuronate to xylulose-5-phosphate Homo sapiens R-HSA-5661270 | 1.91E-01 | 5.76E-01 | 5.79E+00 | 9.59E+00 | SORD |
| 313 | Negative regulation of TCF-dependent signaling by DVL-interacting proteins Homo sapiens R-HSA-5368598 | 1.91E-01 | 5.76E-01 | 5.79E+00 | 9.59E+00 | CCDC88C |
| 314 | Synthesis of GDP-mannose Homo sapiens R-HSA-446205 | 1.91E-01 | 5.76E-01 | 5.79E+00 | 9.59E+00 | PMM1 |
| 315 | Loss of Function of FBXW7 in Cancer and NOTCH1 Signaling Homo sapiens R-HSA-2644607 | 1.91E-01 | 5.76E-01 | 5.79E+00 | 9.59E+00 | NOTCH1 |
| 316 | FBXW7 Mutants and NOTCH1 in Cancer Homo sapiens R-HSA-2644605 | 1.91E-01 | 5.76E-01 | 5.79E+00 | 9.59E+00 | NOTCH1 |
| 317 | LRR FLII-interacting protein 1 (LRRFIP1) activates type I IFN production Homo sapiens R-HSA-3134973 | 1.91E-01 | 5.76E-01 | 5.79E+00 | 9.59E+00 | CTNNB1 |

| | | | | | | |
|---|---|---|---|---|---|---|
| 318 | G2/M DNA replication checkpoint Homo sapiens R-HSA-69478 | 1.91E-01 | 5.76E-01 | 5.79E+00 | 9.59E+00 | WEE1 |
| 319 | Cytochrome c-mediated apoptotic response Homo sapiens R-HSA-111461 | 1.91E-01 | 5.76E-01 | 5.79E+00 | 9.59E+00 | CYCS |
| 320 | Activation of caspases through apoptosome-mediated cleavage Homo sapiens R-HSA-111459 | 1.91E-01 | 5.76E-01 | 5.79E+00 | 9.59E+00 | CYCS |
| 321 | Arachidonate production from DAG Homo sapiens R-HSA-426048 | 1.91E-01 | 5.76E-01 | 5.79E+00 | 9.59E+00 | MGLL |
| 322 | Serotonin and melatonin biosynthesis Homo sapiens R-HSA-209931 | 1.91E-01 | 5.76E-01 | 5.79E+00 | 9.59E+00 | DDC |
| 323 | Trafficking of myristoylated proteins to the cilium Homo sapiens R-HSA-5624138 | 1.91E-01 | 5.76E-01 | 5.79E+00 | 9.59E+00 | CYS1 |
| 324 | Signaling by MST1 Homo sapiens R-HSA-8852405 | 1.91E-01 | 5.76E-01 | 5.79E+00 | 9.59E+00 | HPN |
| 325 | Interleukin-3, 5 and GM-CSF signaling Homo sapiens R-HSA-512988 | 1.97E-01 | 5.92E-01 | 1.32E+00 | 2.14E+00 | DUSP2;MAP2K1;EGF;PDGFA;PIK3R2;PIK3R1;IL17RD;GRIN2B;DUSP9;FGF9;ERBB4;NEFL;FGFR3;FGFR2 |
| 326 | Cardiac conduction Homo sapiens R-HSA-5576891 | 1.98E-01 | 5.94E-01 | 1.46E+00 | 2.37E+00 | AHCYL1;KCNE3;CACNA2D1;FXYD2;KCNK13;ATP2B4;SCN3B;FGF12 |
| 327 | G alpha (12/13) signalling events Homo sapiens R-HSA-416482 | 2.00E-01 | 5.94E-01 | 1.66E+00 | 2.67E+00 | FGD3;OBSCN;PIK3R2;PIK3R1;RHOB |
| 328 | Regulation of gene expression in beta cells Homo sapiens R-HSA-210745 | 2.00E-01 | 5.94E-01 | 2.57E+00 | 4.14E+00 | HNF4A;FOXO1 |

| | | | | | | |
|---|---|---|---|---|---|---|
| 329 | FGFR2 ligand binding and activation Homo sapiens R-HSA-190241 | 2.00E-01 | 5.94E-01 | 2.57E+00 | 4.14E+00 | FGF9;FGFR2 |
| 330 | Synthesis, secretion, and inactivation of Glucagon-like Peptide-1 (GLP-1) Homo sapiens R-HSA-381771 | 2.00E-01 | 5.94E-01 | 2.57E+00 | 4.14E+00 | TCF7L2;CTNNB1 |
| 331 | TCF dependent signaling in response to WNT Homo sapiens R-HSA-201681 | 2.05E-01 | 6.05E-01 | 1.36E+00 | 2.15E+00 | FZD1;TLE3;BCL9;TCF7L2;ZNRF3;CSNK2B;CCDC88C;SOX13;LRP5;CTNNB1;SOX4 |
| 332 | Transcription-Coupled Nucleotide Excision Repair (TC-NER) Homo sapiens R-HSA-6781827 | 2.07E-01 | 6.09E-01 | 1.63E+00 | 2.57E+00 | POLR2A;XPA;RPA1;GTF2H2;GTF2H5 |
| 333 | Transport of vitamins, nucleosides, and related molecules Homo sapiens R-HSA-425397 | 2.08E-01 | 6.09E-01 | 1.99E+00 | 3.12E+00 | SLC35B4;ALB;SLCO2A1 |
| 334 | Signaling by FGFR1 in disease Homo sapiens R-HSA-5655302 | 2.08E-01 | 6.09E-01 | 1.99E+00 | 3.12E+00 | FGF9;CNTRL;PIK3R1 |
| 335 | Visual phototransduction Homo sapiens R-HSA-2187338 | 2.14E-01 | 6.17E-01 | 1.53E+00 | 2.35E+00 | SDC4;LRP1;PDE6G;GPC1;APOE;METAP2 |
| 336 | MyD88-independent TLR3/TLR4 cascade Homo sapiens R-HSA-166166 | 2.14E-01 | 6.17E-01 | 1.53E+00 | 2.35E+00 | MEF2A;MAP2K1;JUN;MAPKAPK3;TAB2;FOS |
| 337 | Toll Like Receptor 3 (TLR3) Cascade Homo sapiens R-HSA-168164 | 2.14E-01 | 6.17E-01 | 1.53E+00 | 2.35E+00 | MEF2A;MAP2K1;JUN;MAPKAPK3;TAB2;FOS |
| 338 | TRIF-mediated TLR3/TLR4 signaling Homo sapiens R-HSA-937061 | 2.14E-01 | 6.17E-01 | 1.53E+00 | 2.35E+00 | MEF2A;MAP2K1;JUN;MAPKAPK3;TAB2;FOS |
| 339 | HS-GAG degradation Homo sapiens R-HSA-2024096 | 2.16E-01 | 6.17E-01 | 2.44E+00 | 3.74E+00 | SDC4;GPC1 |

| | | | | | | |
|---|---|---|---|---|---|---|
| 340 | Signal transduction by L1 Homo sapiens R-HSA-445144 | 2.16E-01 | 6.17E-01 | 2.44E+00 | 3.74E+00 | MAP2K1;CSNK2B |
| 341 | NOTCH2 Activation and Transmission of Signal to the Nucleus Homo sapiens R-HSA-2979096 | 2.16E-01 | 6.17E-01 | 2.44E+00 | 3.74E+00 | JAG1;MIB1 |
| 342 | Regulation of FZD by ubiquitination Homo sapiens R-HSA-4641263 | 2.16E-01 | 6.17E-01 | 2.44E+00 | 3.74E+00 | ZNRF3;LRP5 |
| 343 | mTOR signalling Homo sapiens R-HSA-165159 | 2.19E-01 | 6.19E-01 | 1.93E+00 | 2.93E+00 | CAB39;RRAGD;PRKAB1 |
| 344 | Nuclear signaling by ERBB4 Homo sapiens R-HSA-1251985 | 2.19E-01 | 6.19E-01 | 1.93E+00 | 2.93E+00 | GHR;ERBB4;TAB2 |
| 345 | Association of TriC/CCT with target proteins during biosynthesis Homo sapiens R-HSA-390471 | 2.19E-01 | 6.19E-01 | 1.93E+00 | 2.93E+00 | USP11;FBXL3;DCAF7 |
| 346 | Formation of the beta-catenin:TCF transactivating complex Homo sapiens R-HSA-201722 | 2.19E-01 | 6.19E-01 | 1.72E+00 | 2.60E+00 | TLE3;TCF7L2;BCL9;CTNNB1 |
| 347 | FCERI mediated MAPK activation Homo sapiens R-HSA-2871796 | 2.21E-01 | 6.19E-01 | 1.27E+00 | 1.92E+00 | JUN;DUSP2;MAP2K1;EGF;PDGFA;FOS;IL17RD;GRIN2B;DUSP9;FGF9;ERBB4;PLCG2;NEFL;FGFR3;FGFR2 |
| 348 | Signalling to RAS Homo sapiens R-HSA-167044 | 2.23E-01 | 6.19E-01 | 1.29E+00 | 1.95E+00 | DUSP2;MAP2K1;EGF;PDGFA;IL17RD;GRIN2B;DUSP9;FGF9;MAPKAPK3;ERBB4;NEFL;FGFR3;FGFR2 |
| 349 | Synthesis of Dolichyl-phosphate Homo sapiens R-HSA-446199 | 2.24E-01 | 6.19E-01 | 4.63E+00 | 6.92E+00 | NUS1 |
| 350 | Negative feedback regulation of MAPK pathway Homo sapiens R-HSA-5674499 | 2.24E-01 | 6.19E-01 | 4.63E+00 | 6.92E+00 | MAP2K1 |

| | | | | | | |
|---|---|---|---|---|---|---|
| 351 | Pre-NOTCH Processing in the Endoplasmic Reticulum Homo sapiens R-HSA-1912399 | 2.24E-01 | 6.19E-01 | 4.63E+00 | 6.92E+00 | NOTCH1 |
| 352 | Misspliced LRP5 mutants have enhanced beta-catenin-dependent signaling Homo sapiens R-HSA-5339717 | 2.24E-01 | 6.19E-01 | 4.63E+00 | 6.92E+00 | LRP5 |
| 353 | mitochondrial fatty acid beta-oxidation of unsaturated fatty acids Homo sapiens R-HSA-77288 | 2.24E-01 | 6.19E-01 | 4.63E+00 | 6.92E+00 | DECR1 |
| 354 | Transport and synthesis of PAPS Homo sapiens R-HSA-174362 | 2.24E-01 | 6.19E-01 | 4.63E+00 | 6.92E+00 | PAPSS2 |
| 355 | Synthesis of Lipoxins (LX) Homo sapiens R-HSA-2142700 | 2.24E-01 | 6.19E-01 | 4.63E+00 | 6.92E+00 | HPGD |
| 356 | PKB-mediated events Homo sapiens R-HSA-109703 | 2.30E-01 | 6.27E-01 | 1.88E+00 | 2.76E+00 | CAB39;RRAGD;PRKAB1 |
| 357 | Negative regulation of MAPK pathway Homo sapiens R-HSA-5675221 | 2.30E-01 | 6.27E-01 | 1.88E+00 | 2.76E+00 | DUSP2;MAP2K1;DUSP9 |
| 358 | Fcgamma receptor (FCGR) dependent phagocytosis Homo sapiens R-HSA-2029480 | 2.30E-01 | 6.27E-01 | 1.44E+00 | 2.11E+00 | CYFIP2;AHCYL1;PLCG2;PIK3R2;PIK3R1;ACTB;ACTG1 |
| 359 | VEGFR2 mediated cell proliferation Homo sapiens R-HSA-5218921 | 2.31E-01 | 6.27E-01 | 1.28E+00 | 1.88E+00 | DUSP2;MAP2K1;AHCYL1;EGF;PDGFA;IL17RD;GRIN2B;DUSP9;FGF9;ERBB4;NEFL;FGFR3;FGFR2 |
| 360 | CD28 dependent PI3K/Akt signaling Homo sapiens R-HSA-389357 | 2.31E-01 | 6.27E-01 | 2.32E+00 | 3.39E+00 | PIK3R2;PIK3R1 |
| 361 | Nephrin interactions Homo sapiens R-HSA-373753 | 2.31E-01 | 6.27E-01 | 2.32E+00 | 3.39E+00 | PIK3R2;PIK3R1 |

| | | | | | | |
|---|---|---|---|---|---|---|
| 362 | Glycosaminoglycan metabolism Homo sapiens R-HSA-1630316 | 2.37E-01 | 6.33E-01 | 1.42E+00 | 2.05E+00 | CHST9;SDC4;HYAL1;GPC1;ST3GAL4;CHST2;PAPSS2 |
| 363 | Cleavage of Growing Transcript in the Termination Region Homo sapiens R-HSA-109688 | 2.37E-01 | 6.33E-01 | 1.65E+00 | 2.38E+00 | NUDT21;SRSF3;SRSF5;SRSF6 |
| 364 | RNA Polymerase II Transcription Termination Homo sapiens R-HSA-73856 | 2.37E-01 | 6.33E-01 | 1.65E+00 | 2.38E+00 | NUDT21;SRSF3;SRSF5;SRSF6 |
| 365 | Post-Elongation Processing of the Transcript Homo sapiens R-HSA-76044 | 2.37E-01 | 6.33E-01 | 1.65E+00 | 2.38E+00 | NUDT21;SRSF3;SRSF5;SRSF6 |
| 366 | Role of phospholipids in phagocytosis Homo sapiens R-HSA-2029485 | 2.37E-01 | 6.33E-01 | 1.65E+00 | 2.38E+00 | AHCYL1;PLCG2;PIK3R2;PIK3R1 |
| 367 | Regulation of HSF1-mediated heat shock response Homo sapiens R-HSA-3371453 | 2.37E-01 | 6.33E-01 | 1.54E+00 | 2.22E+00 | HSPA8;HSBP1;ST13;RPA1;HSPA1B |
| 368 | Elastic fibre formation Homo sapiens R-HSA-1566948 | 2.41E-01 | 6.34E-01 | 1.83E+00 | 2.60E+00 | BMP4;EFEMP2;LTBP4 |
| 369 | Protein folding Homo sapiens R-HSA-391251 | 2.41E-01 | 6.34E-01 | 1.46E+00 | 2.08E+00 | USP11;CSNK2B;FBXL3;TBCA;DCAF7;ACTB |
| 370 | Rho GTPase cycle Homo sapiens R-HSA-194840 | 2.43E-01 | 6.34E-01 | 1.41E+00 | 2.00E+00 | ARHGAP44;FGD3;OBSCN;ARHGAP29;PIK3R2;ARHGAP24;RHOB |
| 371 | Signaling by Wnt Homo sapiens R-HSA-195721 | 2.44E-01 | 6.34E-01 | 1.24E+00 | 1.75E+00 | FZD1;TLE3;TCF7L2;PDE6G;SOX13;WNT9B;LRP5;TNRC6C;BCL9;VANGL2;ZNRF3;CSNK2B;CCDC88C;CTNNB1;SOX4 |
| 372 | Ca2+ pathway Homo sapiens R-HSA-4086398 | 2.46E-01 | 6.34E-01 | 1.63E+00 | 2.28E+00 | TNRC6C;TCF7L2;PDE6G;CTNNB1 |
| 373 | Incretin synthesis, secretion, and inactivation Homo sapiens R-HSA-400508 | 2.47E-01 | 6.34E-01 | 2.21E+00 | 3.09E+00 | TCF7L2;CTNNB1 |

| | | | | | | |
|---|---|---|---|---|---|---|
| 374 | Signaling by BMP Homo sapiens R-HSA-201451 | 2.47E-01 | 6.34E-01 | 2.21E+00 | 3.09E+00 | SMAD1;SMAD7 |
| 375 | Signaling by Retinoic Acid Homo sapiens R-HSA-5362517 | 2.52E-01 | 6.34E-01 | 1.78E+00 | 2.46E+00 | RXRA;ADH1C;CRABP2 |
| 376 | Transcriptional regulation of pluripotent stem cells Homo sapiens R-HSA-452723 | 2.52E-01 | 6.34E-01 | 1.78E+00 | 2.46E+00 | SALL1;POLR2A;PBX1 |
| 377 | EPHB-mediated forward signaling Homo sapiens R-HSA-3928662 | 2.52E-01 | 6.34E-01 | 1.78E+00 | 2.46E+00 | GRIN2B;ACTB;ACTG1 |
| 378 | BMAL1:CLOCK,NPAS2 activates circadian gene expression Homo sapiens R-HSA-1368108 | 2.52E-01 | 6.34E-01 | 1.78E+00 | 2.46E+00 | RXRA;TBL1XR1;PPARGC1A |
| 379 | Activation of gene expression by SREBF (SREBP) Homo sapiens R-HSA-2426168 | 2.52E-01 | 6.34E-01 | 1.78E+00 | 2.46E+00 | SREBF1;RXRA;TBL1XR1 |
| 380 | Signalling to ERKs Homo sapiens R-HSA-187687 | 2.52E-01 | 6.34E-01 | 1.26E+00 | 1.73E+00 | DUSP2;MAP2K1;EGF;PDGFA;IL17RD;GRIN2B;DUSP9;FGF9;MAPKAPK3;ERBB4;NEFL;FGFR3;FGFR2 |
| 381 | Platelet activation, signaling and aggregation Homo sapiens R-HSA-76002 | 2.52E-01 | 6.34E-01 | 1.26E+00 | 1.73E+00 | CFD;EGF;PDGFA;PIK3R2;PIK3R1;KNG1;RHOB;SOD1;ALB;PLCG2;FLNA;QSOX1;MGLL |
| 382 | Global Genome Nucleotide Excision Repair (GG-NER) Homo sapiens R-HSA-5696399 | 2.53E-01 | 6.34E-01 | 1.50E+00 | 2.07E+00 | XPA;RPA1;GTF2H2;ACTB;GTF2H5 |
| 383 | Gap-filling DNA repair synthesis and ligation in TC-NER Homo sapiens R-HSA-6782210 | 2.55E-01 | 6.34E-01 | 1.60E+00 | 2.18E+00 | POLR2A;RPA1;GTF2H2;GTF2H5 |
| 384 | RNA Polymerase II Transcription Homo sapiens R-HSA-73857 | 2.55E-01 | 6.34E-01 | 1.39E+00 | 1.89E+00 | NUDT21;POLR2A;SRSF3;SRSF5;GTF2H2;SRSF6;GTF2H5 |
| 385 | Cation-coupled Chloride cotransporters Homo sapiens R-HSA-426117 | 2.56E-01 | 6.34E-01 | 3.86E+00 | 5.25E+00 | SLC12A1 |

| | | | | | | |
|---|---|---|---|---|---|---|
| 386 | SLC transporter disorders Homo sapiens R-HSA-5619102 | 2.56E-01 | 6.34E-01 | 3.86E+00 | 5.25E+00 | ALB |
| 387 | Regulation of gene expression in late stage (branching morphogenesis) pancreatic bud precursor cells Homo sapiens R-HSA-210744 | 2.56E-01 | 6.34E-01 | 3.86E+00 | 5.25E+00 | NOTCH1 |
| 388 | Post-transcriptional silencing by small RNAs Homo sapiens R-HSA-426496 | 2.56E-01 | 6.34E-01 | 3.86E+00 | 5.25E+00 | TNRC6C |
| 389 | Nectin/Necl trans heterodimerization Homo sapiens R-HSA-420597 | 2.56E-01 | 6.34E-01 | 3.86E+00 | 5.25E+00 | CADM1 |
| 390 | Phase 3 - rapid repolarisation Homo sapiens R-HSA-5576890 | 2.56E-01 | 6.34E-01 | 3.86E+00 | 5.25E+00 | KCNE3 |
| 391 | Apoptotic factor-mediated response Homo sapiens R-HSA-111471 | 2.56E-01 | 6.34E-01 | 3.86E+00 | 5.25E+00 | CYCS |
| 392 | Acyl chain remodeling of DAG and TAG Homo sapiens R-HSA-1482883 | 2.56E-01 | 6.34E-01 | 3.86E+00 | 5.25E+00 | MGLL |
| 393 | Phosphate bond hydrolysis by NUDT proteins Homo sapiens R-HSA-2393930 | 2.56E-01 | 6.34E-01 | 3.86E+00 | 5.25E+00 | NUDT16 |
| 394 | Insulin processing Homo sapiens R-HSA-264876 | 2.62E-01 | 6.34E-01 | 2.10E+00 | 2.82E+00 | KIF5A;VAMP2 |
| 395 | Synthesis of bile acids and bile salts via 7alpha-hydroxycholesterol Homo sapiens R-HSA-193368 | 2.62E-01 | 6.34E-01 | 2.10E+00 | 2.82E+00 | CYP27A1;RXRA |
| 396 | Endogenous sterols Homo sapiens R-HSA-211976 | 2.62E-01 | 6.34E-01 | 2.10E+00 | 2.82E+00 | CYP27A1;RXRA |

| | | | | | | |
|---|---|---|---|---|---|---|
| 397 | HIV Transcription Elongation Homo sapiens R-HSA-167169 | 2.63E-01 | 6.34E-01 | 1.74E+00 | 2.32E+00 | POLR2A;GTF2H2;GTF2H5 |
| 398 | Tat-mediated elongation of the HIV-1 transcript Homo sapiens R-HSA-167246 | 2.63E-01 | 6.34E-01 | 1.74E+00 | 2.32E+00 | POLR2A;GTF2H2;GTF2H5 |
| 399 | Formation of HIV-1 elongation complex containing HIV-1 Tat Homo sapiens R-HSA-167200 | 2.63E-01 | 6.34E-01 | 1.74E+00 | 2.32E+00 | POLR2A;GTF2H2;GTF2H5 |
| 400 | Synthesis of substrates in N-glycan biosythesis Homo sapiens R-HSA-446219 | 2.64E-01 | 6.34E-01 | 1.57E+00 | 2.09E+00 | NUS1;PMM1;ST3GAL4;ST3GAL5 |
| 401 | Neuronal System Homo sapiens R-HSA-112316 | 2.68E-01 | 6.34E-01 | 1.22E+00 | 1.60E+00 | HSPA8;KCNJ10;CACNA2D1;SLC22A2;KCNK13;KCNJ15;GRIN2B;CPLX1;EPB41L1;KCNMA1;NAAA;NEFL;KCNJ1;VAMP2;PPFIA3 |
| 402 | GRB2 events in EGFR signaling Homo sapiens R-HSA-179812 | 2.70E-01 | 6.34E-01 | 1.25E+00 | 1.63E+00 | DUSP2;MAP2K1;FGF9;ERBB4;EGF;PDGFA;NEFL;IL17RD;GRIN2B;DUSP9;FGFR3;FGFR2 |
| 403 | SHC1 events in EGFR signaling Homo sapiens R-HSA-180336 | 2.70E-01 | 6.34E-01 | 1.25E+00 | 1.63E+00 | DUSP2;MAP2K1;FGF9;ERBB4;EGF;PDGFA;NEFL;IL17RD;GRIN2B;DUSP9;FGFR3;FGFR2 |
| 404 | SOS-mediated signalling Homo sapiens R-HSA-112412 | 2.70E-01 | 6.34E-01 | 1.25E+00 | 1.63E+00 | DUSP2;MAP2K1;FGF9;ERBB4;EGF;PDGFA;NEFL;IL17RD;GRIN2B;DUSP9;FGFR3;FGFR2 |
| 405 | SHC1 events in ERBB4 signaling Homo sapiens R-HSA-1250347 | 2.70E-01 | 6.34E-01 | 1.25E+00 | 1.63E+00 | DUSP2;MAP2K1;FGF9;ERBB4;EGF;PDGFA;NEFL;IL17RD;GRIN2B;DUSP9;FGFR3;FGFR2 |
| 406 | RAF/MAP kinase cascade Homo sapiens R-HSA-5673001 | 2.70E-01 | 6.34E-01 | 1.25E+00 | 1.63E+00 | DUSP2;MAP2K1;FGF9;ERBB4;EGF;PDGFA;NEFL;IL17RD;GRIN2B;DUSP9;FGFR3;FGFR2 |
| 407 | Formation of HIV elongation complex in the absence of HIV Tat Homo sapiens R-HSA-167152 | 2.75E-01 | 6.34E-01 | 1.69E+00 | 2.19E+00 | POLR2A;GTF2H2;GTF2H5 |

| | | | | | | |
|---|---|---|---|---|---|---|
| 408 | RNA Polymerase II Transcription Elongation Homo sapiens R-HSA-75955 | 2.75E-01 | 6.34E-01 | 1.69E+00 | 2.19E+00 | POLR2A;GTF2H2;GTF2H5 |
| 409 | Formation of RNA Pol II elongation complex Homo sapiens R-HSA-112382 | 2.75E-01 | 6.34E-01 | 1.69E+00 | 2.19E+00 | POLR2A;GTF2H2;GTF2H5 |
| 410 | RNA Polymerase II HIV Promoter Escape Homo sapiens R-HSA-167162 | 2.75E-01 | 6.34E-01 | 1.69E+00 | 2.19E+00 | POLR2A;GTF2H2;GTF2H5 |
| 411 | RNA Polymerase II Promoter Escape Homo sapiens R-HSA-73776 | 2.75E-01 | 6.34E-01 | 1.69E+00 | 2.19E+00 | POLR2A;GTF2H2;GTF2H5 |
| 412 | RNA Polymerase II Transcription Initiation And Promoter Clearance Homo sapiens R-HSA-76042 | 2.75E-01 | 6.34E-01 | 1.69E+00 | 2.19E+00 | POLR2A;GTF2H2;GTF2H5 |
| 413 | RNA Polymerase II Transcription Initiation Homo sapiens R-HSA-75953 | 2.75E-01 | 6.34E-01 | 1.69E+00 | 2.19E+00 | POLR2A;GTF2H2;GTF2H5 |
| 414 | RNA Polymerase II Transcription Pre-Initiation And Promoter Opening Homo sapiens R-HSA-73779 | 2.75E-01 | 6.34E-01 | 1.69E+00 | 2.19E+00 | POLR2A;GTF2H2;GTF2H5 |
| 415 | HIV Transcription Initiation Homo sapiens R-HSA-167161 | 2.75E-01 | 6.34E-01 | 1.69E+00 | 2.19E+00 | POLR2A;GTF2H2;GTF2H5 |
| 416 | Transcriptional activity of SMAD2/SMAD3:SMAD4 heterotrimer Homo sapiens R-HSA-2173793 | 2.75E-01 | 6.34E-01 | 1.69E+00 | 2.19E+00 | NEDD4L;JUNB;SMAD7 |
| 417 | Interleukin-1 signaling Homo sapiens R-HSA-446652 | 2.75E-01 | 6.34E-01 | 1.69E+00 | 2.19E+00 | MAP2K1;PELI2;TAB2 |

| | | | | | | |
|---|---|---|---|---|---|---|
| 418 | Phase 0 - rapid depolarisation Homo sapiens R-HSA-5576892 | 2.75E-01 | 6.34E-01 | 1.69E+00 | 2.19E+00 | CACNA2D1;SCN3B;FGF12 |
| 419 | FRS-mediated FGFR2 signaling Homo sapiens R-HSA-5654700 | 2.75E-01 | 6.34E-01 | 1.24E+00 | 1.60E+00 | DUSP2;MAP2K1;FGF9;ERBB4;EGF;PDGFA;NEFL;IL17RD;GRIN2B;DUSP9;FGFR3;FGFR2 |
| 420 | FRS-mediated FGFR4 signaling Homo sapiens R-HSA-5654712 | 2.75E-01 | 6.34E-01 | 1.24E+00 | 1.60E+00 | DUSP2;MAP2K1;FGF9;ERBB4;EGF;PDGFA;NEFL;IL17RD;GRIN2B;DUSP9;FGFR3;FGFR2 |
| 421 | FRS-mediated FGFR3 signaling Homo sapiens R-HSA-5654706 | 2.75E-01 | 6.34E-01 | 1.24E+00 | 1.60E+00 | DUSP2;MAP2K1;FGF9;ERBB4;EGF;PDGFA;NEFL;IL17RD;GRIN2B;DUSP9;FGFR3;FGFR2 |
| 422 | FRS-mediated FGFR1 signaling Homo sapiens R-HSA-5654693 | 2.75E-01 | 6.34E-01 | 1.24E+00 | 1.60E+00 | DUSP2;MAP2K1;FGF9;ERBB4;EGF;PDGFA;NEFL;IL17RD;GRIN2B;DUSP9;FGFR3;FGFR2 |
| 423 | Amino acid synthesis and interconversion (transamination) Homo sapiens R-HSA-70614 | 2.78E-01 | 6.34E-01 | 2.01E+00 | 2.58E+00 | OAT;PSAT1 |
| 424 | Activation of G protein gated Potassium channels Homo sapiens R-HSA-1296041 | 2.78E-01 | 6.34E-01 | 2.01E+00 | 2.58E+00 | KCNJ10;KCNJ15 |
| 425 | Inhibition of voltage gated Ca2+ channels via Gbeta/gamma subunits Homo sapiens R-HSA-997272 | 2.78E-01 | 6.34E-01 | 2.01E+00 | 2.58E+00 | KCNJ10;KCNJ15 |
| 426 | G protein gated Potassium channels Homo sapiens R-HSA-1296059 | 2.78E-01 | 6.34E-01 | 2.01E+00 | 2.58E+00 | KCNJ10;KCNJ15 |
| 427 | Nitric oxide stimulates guanylate cyclase Homo sapiens R-HSA-392154 | 2.78E-01 | 6.34E-01 | 2.01E+00 | 2.58E+00 | KCNMA1;PDE3A |
| 428 | Synthesis of IP3 and IP4 in the cytosol Homo sapiens R-HSA-1855204 | 2.78E-01 | 6.34E-01 | 2.01E+00 | 2.58E+00 | PLCG2;PLCD1 |

| | | | | | | |
|---|---|---|---|---|---|---|
| 429 | Signaling by ERBB2 Homo sapiens R-HSA-1227986 | 2.86E-01 | 6.34E-01 | 1.65E+00 | 2.07E+00 | ERBB4;EGF;PIK3R1 |
| 430 | Neurotransmitter Clearance In The Synaptic Cleft Homo sapiens R-HSA-112311 | 2.87E-01 | 6.34E-01 | 3.31E+00 | 4.12E+00 | SLC22A2 |
| 431 | Insulin-like Growth Factor-2 mRNA Binding Proteins (IGF2BPs/IMPs/VICKZs) bind RNA Homo sapiens R-HSA-428359 | 2.87E-01 | 6.34E-01 | 3.31E+00 | 4.12E+00 | ACTB |
| 432 | Ligand-receptor interactions Homo sapiens R-HSA-5632681 | 2.87E-01 | 6.34E-01 | 3.31E+00 | 4.12E+00 | GAS1 |
| 433 | WNT mediated activation of DVL Homo sapiens R-HSA-201688 | 2.87E-01 | 6.34E-01 | 3.31E+00 | 4.12E+00 | CSNK2B |
| 434 | BH3-only proteins associate with and inactivate anti-apoptotic BCL-2 members Homo sapiens R-HSA-111453 | 2.87E-01 | 6.34E-01 | 3.31E+00 | 4.12E+00 | BCL2L1 |
| 435 | Recycling of eIF2:GDP Homo sapiens R-HSA-72731 | 2.87E-01 | 6.34E-01 | 3.31E+00 | 4.12E+00 | EIF2S3 |
| 436 | Downregulation of ERBB4 signaling Homo sapiens R-HSA-1253288 | 2.87E-01 | 6.34E-01 | 3.31E+00 | 4.12E+00 | ERBB4 |
| 437 | RNF mutants show enhanced WNT signaling and proliferation Homo sapiens R-HSA-5340588 | 2.87E-01 | 6.34E-01 | 3.31E+00 | 4.12E+00 | LRP5 |
| 438 | Hydrolysis of LPC Homo sapiens R-HSA-1483115 | 2.87E-01 | 6.34E-01 | 3.31E+00 | 4.12E+00 | GPCPD1 |
| 439 | Linoleic acid (LA) metabolism Homo sapiens R-HSA-2046105 | 2.87E-01 | 6.34E-01 | 3.31E+00 | 4.12E+00 | FADS1 |

| | | | | | | |
|---|---|---|---|---|---|---|
| 440 | NR1D1 (REV-ERBA) represses gene expression Homo sapiens R-HSA-1368071 | 2.87E-01 | 6.34E-01 | 3.31E+00 | 4.12E+00 | PPARGC1A |
| 441 | Nicotinamide salvaging Homo sapiens R-HSA-197264 | 2.87E-01 | 6.34E-01 | 3.31E+00 | 4.12E+00 | NAPRT |
| 442 | Synthesis of epoxy (EET) and dihydroxyeicosatrienoic acids (DHET) Homo sapiens R-HSA-2142670 | 2.87E-01 | 6.34E-01 | 3.31E+00 | 4.12E+00 | EPHX2 |
| 443 | ARMS-mediated activation Homo sapiens R-HSA-170984 | 2.89E-01 | 6.34E-01 | 1.23E+00 | 1.52E+00 | DUSP2;MAP2K1;FGF9;ERBB4;EGF;PDGFA;NEFL;IL17RD;GRIN2B;DUSP9;FGFR3;FGFR2 |
| 444 | Signalling to p38 via RIT and RIN Homo sapiens R-HSA-187706 | 2.89E-01 | 6.34E-01 | 1.23E+00 | 1.52E+00 | DUSP2;MAP2K1;FGF9;ERBB4;EGF;PDGFA;NEFL;IL17RD;GRIN2B;DUSP9;FGFR3;FGFR2 |
| 445 | Nucleotide Excision Repair Homo sapiens R-HSA-5696398 | 2.91E-01 | 6.34E-01 | 1.36E+00 | 1.68E+00 | POLR2A;XPA;RPA1;GTF2H2;ACTB;GTF2H5 |
| 446 | O-linked glycosylation of mucins Homo sapiens R-HSA-913709 | 2.92E-01 | 6.34E-01 | 1.49E+00 | 1.84E+00 | B3GNT9;ST3GAL4;GCNT4;MUC20 |
| 447 | Diseases associated with glycosaminoglycan metabolism Homo sapiens R-HSA-3560782 | 2.93E-01 | 6.34E-01 | 1.93E+00 | 2.37E+00 | SDC4;GPC1 |
| 448 | A tetrasaccharide linker sequence is required for GAG synthesis Homo sapiens R-HSA-1971475 | 2.93E-01 | 6.34E-01 | 1.93E+00 | 2.37E+00 | SDC4;GPC1 |
| 449 | Termination of O-glycan biosynthesis Homo sapiens R-HSA-977068 | 2.93E-01 | 6.34E-01 | 1.93E+00 | 2.37E+00 | ST3GAL4;MUC20 |

| | | | | | | |
|---|---|---|---|---|---|---|
| 450 | N-glycan antennae elongation in the medial/trans-Golgi Homo sapiens R-HSA-975576 | 2.93E-01 | 6.34E-01 | 1.93E+00 | 2.37E+00 | CHST10;ST3GAL4 |
| 451 | FGFR2 alternative splicing Homo sapiens R-HSA-6803529 | 2.93E-01 | 6.34E-01 | 1.93E+00 | 2.37E+00 | POLR2A;HNRNPH1 |
| 452 | SHC-mediated cascade:FGFR2 Homo sapiens R-HSA-5654699 | 2.93E-01 | 6.34E-01 | 1.93E+00 | 2.37E+00 | FGF9;FGFR2 |
| 453 | Phase 2 - plateau phase Homo sapiens R-HSA-5576893 | 2.93E-01 | 6.34E-01 | 1.93E+00 | 2.37E+00 | KCNE3;CACNA2D1 |
| 454 | Frs2-mediated activation Homo sapiens R-HSA-170968 | 2.94E-01 | 6.34E-01 | 1.22E+00 | 1.49E+00 | DUSP2;MAP2K1;FGF9;ERBB4;EGF;PDGFA;NEFL;IL17RD;GRIN2B;DUSP9;FGFR3;FGFR2 |
| 455 | Muscle contraction Homo sapiens R-HSA-397014 | 2.96E-01 | 6.37E-01 | 1.25E+00 | 1.52E+00 | AHCYL1;KCNE3;CACNA2D1;FXYD2;TNNC1;KCNK13;ATP2B4;SCN3B;SORBS3;FGF12 |
| 456 | Phospholipase C-mediated cascade; FGFR4 Homo sapiens R-HSA-5654228 | 2.97E-01 | 6.39E-01 | 1.62E+00 | 1.96E+00 | AHCYL1;FGF9;PRKAR1B |
| 457 | MAPK1/MAPK3 signaling Homo sapiens R-HSA-5684996 | 2.99E-01 | 6.40E-01 | 1.21E+00 | 1.47E+00 | DUSP2;MAP2K1;FGF9;ERBB4;EGF;PDGFA;NEFL;IL17RD;GRIN2B;DUSP9;FGFR3;FGFR2 |
| 458 | Macroautophagy Homo sapiens R-HSA-1632852 | 3.02E-01 | 6.40E-01 | 1.47E+00 | 1.76E+00 | RRAGD;DYNLL1;PRKAB1;DYNLL2 |
| 459 | Signaling by PTK6 Homo sapiens R-HSA-8848021 | 3.02E-01 | 6.40E-01 | 1.47E+00 | 1.76E+00 | CDKN1B;CCND1;ERBB4;EGF |
| 460 | Prolonged ERK activation events Homo sapiens R-HSA-169893 | 3.04E-01 | 6.40E-01 | 1.21E+00 | 1.44E+00 | DUSP2;MAP2K1;FGF9;ERBB4;EGF;PDGFA;NEFL;IL17RD;GRIN2B;DUSP9;FGFR3;FGFR2 |
| 461 | Golgi-to-ER retrograde transport Homo sapiens R-HSA-8856688 | 3.05E-01 | 6.40E-01 | 1.34E+00 | 1.59E+00 | KIF26A;RAB1B;KIF5A;DYNLL1;BICD1;DYNLL2 |
| 462 | Vesicle-mediated transport Homo sapiens R-HSA-5653656 | 3.06E-01 | 6.40E-01 | 1.14E+00 | 1.35E+00 | TPD52;HSPA8;LRP1;RAB1B;GOLIM4;HIP1R;DYNLL1;PRKAB1;DYNLL2;BICD1;TBC1D1;BLOC1S6;COL4A2;KIF26A;COL4A1;FTH1;ALB;KIF5A;APOE;VAMP2;CTSC;FTL;PICALM |

| | | | | | | |
|---|---|---|---|---|---|---|
| 463 | Signaling by Leptin Homo sapiens R-HSA-2586552 | 3.08E-01 | 6.40E-01 | 1.20E+00 | 1.42E+00 | DUSP2;MAP2K1;FGF9;ERBB4;EGF;PDGFA;NEFL;IL17RD;GRIN2B;DUSP9;FGFR3;FGFR2 |
| 464 | Uptake and actions of bacterial toxins Homo sapiens R-HSA-5339562 | 3.09E-01 | 6.40E-01 | 1.85E+00 | 2.18E+00 | MAP2K1;VAMP2 |
| 465 | Keratan sulfate biosynthesis Homo sapiens R-HSA-2022854 | 3.09E-01 | 6.40E-01 | 1.85E+00 | 2.18E+00 | ST3GAL4;CHST2 |
| 466 | EGFR downregulation Homo sapiens R-HSA-182971 | 3.09E-01 | 6.40E-01 | 1.85E+00 | 2.18E+00 | EGF;SPRY1 |
| 467 | CREB phosphorylation through the activation of Ras Homo sapiens R-HSA-442742 | 3.09E-01 | 6.40E-01 | 1.85E+00 | 2.18E+00 | NEFL;GRIN2B |
| 468 | Synthesis of bile acids and bile salts Homo sapiens R-HSA-192105 | 3.09E-01 | 6.40E-01 | 1.85E+00 | 2.18E+00 | CYP27A1;RXRA |
| 469 | Nucleotide-binding domain, leucine rich repeat containing receptor (NLR) signaling pathways Homo sapiens R-HSA-168643 | 3.09E-01 | 6.40E-01 | 1.58E+00 | 1.85E+00 | TXNIP;TAB2;BCL2L1 |
| 470 | Antigen activates B Cell Receptor (BCR) leading to generation of second messengers Homo sapiens R-HSA-983695 | 3.09E-01 | 6.40E-01 | 1.58E+00 | 1.85E+00 | AHCYL1;PLCG2;PIK3R1 |
| 471 | Phospholipase C-mediated cascade: FGFR1 Homo sapiens R-HSA-5654219 | 3.09E-01 | 6.40E-01 | 1.58E+00 | 1.85E+00 | AHCYL1;FGF9;PRKAR1B |
| 472 | Activation of anterior HOX genes in hindbrain development during early embryogenesis Homo sapiens R-HSA-5617472 | 3.09E-01 | 6.40E-01 | 1.38E+00 | 1.62E+00 | JUN;RXRA;POLR2A;HOXC4;PBX1 |

| | | | | | | |
|---|---|---|---|---|---|---|
| 473 | Activation of HOX genes during differentiation Homo sapiens R-HSA-5619507 | 3.09E-01 | 6.40E-01 | 1.38E+00 | 1.62E+00 | JUN;RXRA;POLR2A;HOXC4;PBX1 |
| 474 | Organic cation transport Homo sapiens R-HSA-549127 | 3.17E-01 | 6.44E-01 | 2.89E+00 | 3.32E+00 | SLC22A2 |
| 475 | Transport of nucleotide sugars Homo sapiens R-HSA-727802 | 3.17E-01 | 6.44E-01 | 2.89E+00 | 3.32E+00 | SLC35B4 |
| 476 | A third proteolytic cleavage releases NICD Homo sapiens R-HSA-157212 | 3.17E-01 | 6.44E-01 | 2.89E+00 | 3.32E+00 | NOTCH1 |
| 477 | RHO GTPases Activate Rhotekin and Rhophilins Homo sapiens R-HSA-5666185 | 3.17E-01 | 6.44E-01 | 2.89E+00 | 3.32E+00 | RHOB |
| 478 | Role of Abl in Robo-Slit signaling Homo sapiens R-HSA-428890 | 3.17E-01 | 6.44E-01 | 2.89E+00 | 3.32E+00 | GPC1 |
| 479 | Inactivation of Cdc42 and Rac Homo sapiens R-HSA-428543 | 3.17E-01 | 6.44E-01 | 2.89E+00 | 3.32E+00 | GPC1 |
| 480 | Ca2+ activated K+ channels Homo sapiens R-HSA-1296052 | 3.17E-01 | 6.44E-01 | 2.89E+00 | 3.32E+00 | KCNMA1 |
| 481 | Degradation of cysteine and homocysteine Homo sapiens R-HSA-1614558 | 3.17E-01 | 6.44E-01 | 2.89E+00 | 3.32E+00 | CDO1 |
| 482 | Pyrimidine salvage reactions Homo sapiens R-HSA-73614 | 3.17E-01 | 6.44E-01 | 2.89E+00 | 3.32E+00 | CDA |
| 483 | Loss of Nlp from mitotic centrosomes Homo sapiens R-HSA-380259 | 3.21E-01 | 6.49E-01 | 1.43E+00 | 1.62E+00 | CNTRL;CEP131;CEP41;DYNLL1 |

| | | | | | | |
|---|---|---|---|---|---|---|
| 484 | Loss of proteins required for interphase microtubule organization?from the centrosome Homo sapiens R-HSA-380284 | 3.21E-01 | 6.49E-01 | 1.43E+00 | 1.62E+00 | CNTRL;CEP131;CEP41;DYNLL1 |
| 485 | Oxidative Stress Induced Senescence Homo sapiens R-HSA-2559580 | 3.26E-01 | 6.57E-01 | 1.35E+00 | 1.51E+00 | TNRC6C;JUN;CBX6;MAPKAPK3;FOS |
| 486 | PPARA activates gene expression Homo sapiens R-HSA-1989781 | 3.27E-01 | 6.59E-01 | 1.30E+00 | 1.45E+00 | FABP1;SREBF1;RXRA;TBL1XR1;PPARGC1A;FADS1 |
| 487 | Complex I biogenesis Homo sapiens R-HSA-6799198 | 3.32E-01 | 6.67E-01 | 1.51E+00 | 1.67E+00 | NDUFA5;NDUFA1;NDUFB1 |
| 488 | Activation of BH3-only proteins Homo sapiens R-HSA-114452 | 3.40E-01 | 6.69E-01 | 1.71E+00 | 1.85E+00 | DYNLL1;DYNLL2 |
| 489 | Negative regulation of FGFR3 signaling Homo sapiens R-HSA-5654732 | 3.40E-01 | 6.69E-01 | 1.71E+00 | 1.85E+00 | FGF9;FGFR3 |
| 490 | YAP1- and WWTR1 (TAZ)-stimulated gene expression Homo sapiens R-HSA-2032785 | 3.40E-01 | 6.69E-01 | 1.71E+00 | 1.85E+00 | RXRA;TBL1XR1 |
| 491 | Neurotoxicity of clostridium toxins Homo sapiens R-HSA-168799 | 3.45E-01 | 6.69E-01 | 2.57E+00 | 2.73E+00 | VAMP2 |
| 492 | Reactions specific to the complex N-glycan synthesis pathway Homo sapiens R-HSA-975578 | 3.45E-01 | 6.69E-01 | 2.57E+00 | 2.73E+00 | CHST10 |
| 493 | Removal of the Flap Intermediate from the C-strand Homo sapiens R-HSA-174437 | 3.45E-01 | 6.69E-01 | 2.57E+00 | 2.73E+00 | RPA1 |

| | | | | | | |
|---|---|---|---|---|---|---|
| 494 | MAPK3 (ERK1) activation Homo sapiens R-HSA-110056 | 3.45E-01 | 6.69E-01 | 2.57E+00 | 2.73E+00 | MAP2K1 |
| 495 | IRAK2 mediated activation of TAK1 complex upon TLR7/8 or 9 stimulation Homo sapiens R-HSA-975163 | 3.45E-01 | 6.69E-01 | 2.57E+00 | 2.73E+00 | TAB2 |
| 496 | IRAK2 mediated activation of TAK1 complex Homo sapiens R-HSA-937042 | 3.45E-01 | 6.69E-01 | 2.57E+00 | 2.73E+00 | TAB2 |
| 497 | Regulation of gene expression by Hypoxia-inducible Factor Homo sapiens R-HSA-1234158 | 3.45E-01 | 6.69E-01 | 2.57E+00 | 2.73E+00 | CITED2 |
| 498 | FGFR2b ligand binding and activation Homo sapiens R-HSA-190377 | 3.45E-01 | 6.69E-01 | 2.57E+00 | 2.73E+00 | FGFR2 |
| 499 | IRAK1 recruits IKK complex Homo sapiens R-HSA-937039 | 3.45E-01 | 6.69E-01 | 2.57E+00 | 2.73E+00 | PELI2 |
| 500 | IRAK1 recruits IKK complex upon TLR7/8 or 9 stimulation Homo sapiens R-HSA-975144 | 3.45E-01 | 6.69E-01 | 2.57E+00 | 2.73E+00 | PELI2 |
| 501 | Dectin-2 family Homo sapiens R-HSA-5621480 | 3.45E-01 | 6.69E-01 | 2.57E+00 | 2.73E+00 | PLCG2 |
| 502 | Folding of actin by CCT/TriC Homo sapiens R-HSA-390450 | 3.45E-01 | 6.69E-01 | 2.57E+00 | 2.73E+00 | ACTB |
| 503 | Acetylcholine regulates insulin secretion Homo sapiens R-HSA-399997 | 3.45E-01 | 6.69E-01 | 2.57E+00 | 2.73E+00 | MARCKS |

| | | | | | | |
|---|---|---|---|---|---|---|
| 504 | Synthesis of pyrophosphates in the cytosol Homo sapiens R-HSA-1855167 | 3.45E-01 | 6.69E-01 | 2.57E+00 | 2.73E+00 | NUDT3 |
| 505 | Urea cycle Homo sapiens R-HSA-70635 | 3.45E-01 | 6.69E-01 | 2.57E+00 | 2.73E+00 | ASS1 |
| 506 | AURKA Activation by TPX2 Homo sapiens R-HSA-8854518 | 3.49E-01 | 6.75E-01 | 1.36E+00 | 1.43E+00 | CNTRL;CEP131;CEP41;DYNLL1 |
| 507 | Regulation of lipid metabolism by Peroxisome proliferator-activated receptor alpha (PPARalpha) Homo sapiens R-HSA-400206 | 3.50E-01 | 6.75E-01 | 1.26E+00 | 1.33E+00 | FABP1;SREBF1;RXRA;TBL1XR1;PPARGC1A;FADS1 |
| 508 | Cellular Senescence Homo sapiens R-HSA-2559583 | 3.51E-01 | 6.77E-01 | 1.21E+00 | 1.27E+00 | TNRC6C;JUN;CBX6;CDKN1B;MAPKAPK3;ID1;HMGA1;FOS |
| 509 | Nuclear Envelope Breakdown Homo sapiens R-HSA-2980766 | 3.55E-01 | 6.77E-01 | 1.45E+00 | 1.50E+00 | BANF1;LPIN1;EMD |
| 510 | Ion homeostasis Homo sapiens R-HSA-5578775 | 3.55E-01 | 6.77E-01 | 1.45E+00 | 1.50E+00 | AHCYL1;FXYD2;ATP2B4 |
| 511 | Apoptotic execution phase Homo sapiens R-HSA-75153 | 3.55E-01 | 6.77E-01 | 1.45E+00 | 1.50E+00 | HMGB2;CTNNB1;DSG2 |
| 512 | RNA Polymerase I Promoter Escape Homo sapiens R-HSA-73772 | 3.55E-01 | 6.77E-01 | 1.65E+00 | 1.71E+00 | GTF2H2;GTF2H5 |
| 513 | Tight junction interactions Homo sapiens R-HSA-420029 | 3.55E-01 | 6.77E-01 | 1.65E+00 | 1.71E+00 | CLDN10;CLDN19 |
| 514 | Chaperonin-mediated protein folding Homo sapiens R-HSA-390466 | 3.59E-01 | 6.83E-01 | 1.29E+00 | 1.32E+00 | USP11;CSNK2B;FBXL3;DCAF7;ACTB |
| 515 | mRNA Splicing - Minor Pathway Homo sapiens R-HSA-72165 | 3.66E-01 | 6.83E-01 | 1.42E+00 | 1.42E+00 | POLR2A;YBX1;SRSF6 |

| | | | | | | |
|---|---|---|---|---|---|---|
| 516 | L1CAM interactions Homo sapiens R-HSA-373760 | 3.67E-01 | 6.83E-01 | 1.27E+00 | 1.28E+00 | HSPA8;MAP2K1;CHL1;CSNK2B;SCN3B |
| 517 | Cellular response to heat stress Homo sapiens R-HSA-3371556 | 3.67E-01 | 6.83E-01 | 1.27E+00 | 1.28E+00 | HSPA8;HSBP1;ST13;RPA1;HSPA1B |
| 518 | HS-GAG biosynthesis Homo sapiens R-HSA-2022928 | 3.70E-01 | 6.83E-01 | 1.60E+00 | 1.59E+00 | SDC4;GPC1 |
| 519 | Uptake and function of anthrax toxins Homo sapiens R-HSA-5210891 | 3.72E-01 | 6.83E-01 | 2.31E+00 | 2.29E+00 | MAP2K1 |
| 520 | Import of palmitoyl-CoA into the mitochondrial matrix Homo sapiens R-HSA-200425 | 3.72E-01 | 6.83E-01 | 2.31E+00 | 2.29E+00 | RXRA |
| 521 | Processive synthesis on the C-strand of the telomere Homo sapiens R-HSA-174414 | 3.72E-01 | 6.83E-01 | 2.31E+00 | 2.29E+00 | RPA1 |
| 522 | Signaling by NOTCH3 Homo sapiens R-HSA-1980148 | 3.72E-01 | 6.83E-01 | 2.31E+00 | 2.29E+00 | JAG1 |
| 523 | Signaling by NOTCH4 Homo sapiens R-HSA-1980150 | 3.72E-01 | 6.83E-01 | 2.31E+00 | 2.29E+00 | JAG1 |
| 524 | FGFR1c ligand binding and activation Homo sapiens R-HSA-190373 | 3.72E-01 | 6.83E-01 | 2.31E+00 | 2.29E+00 | FGF9 |
| 525 | Signaling by activated point mutants of FGFR1 Homo sapiens R-HSA-1839122 | 3.72E-01 | 6.83E-01 | 2.31E+00 | 2.29E+00 | FGF9 |
| 526 | Signaling by FGFR4 in disease Homo sapiens R-HSA-5655291 | 3.72E-01 | 6.83E-01 | 2.31E+00 | 2.29E+00 | PIK3R1 |
| 527 | Condensation of Prometaphase Chromosomes Homo sapiens R-HSA-2514853 | 3.72E-01 | 6.83E-01 | 2.31E+00 | 2.29E+00 | CSNK2B |

| | | | | | | |
|---|---|---|---|---|---|---|
| 528 | RHO GTPases activate KTN1 Homo sapiens R-HSA-5625970 | 3.72E-01 | 6.83E-01 | 2.31E+00 | 2.29E+00 | KIF5A |
| 529 | RHO GTPases activate IQGAPs Homo sapiens R-HSA-5626467 | 3.72E-01 | 6.83E-01 | 2.31E+00 | 2.29E+00 | CTNNB1 |
| 530 | Activation of the phototransduction cascade Homo sapiens R-HSA-2485179 | 3.72E-01 | 6.83E-01 | 2.31E+00 | 2.29E+00 | PDE6G |
| 531 | CLEC7A (Dectin-1) induces NFAT activation Homo sapiens R-HSA-5607763 | 3.72E-01 | 6.83E-01 | 2.31E+00 | 2.29E+00 | AHCYL1 |
| 532 | Synthesis of IP2, IP, and Ins in the cytosol Homo sapiens R-HSA-1855183 | 3.72E-01 | 6.83E-01 | 2.31E+00 | 2.29E+00 | IMPA2 |
| 533 | Heme biosynthesis Homo sapiens R-HSA-189451 | 3.72E-01 | 6.83E-01 | 2.31E+00 | 2.29E+00 | FECH |
| 534 | Phenylalanine and tyrosine catabolism Homo sapiens R-HSA-71182 | 3.72E-01 | 6.83E-01 | 2.31E+00 | 2.29E+00 | FAH |
| 535 | Metabolism of water-soluble vitamins and cofactors Homo sapiens R-HSA-196849 | 3.84E-01 | 6.92E-01 | 1.24E+00 | 1.19E+00 | ALDH1L1;SLC46A1;MMAB;NAPRT;SLC19A2 |
| 536 | Keratan sulfate/keratin metabolism Homo sapiens R-HSA-1638074 | 3.85E-01 | 6.92E-01 | 1.54E+00 | 1.47E+00 | ST3GAL4;CHST2 |
| 537 | Activation of Matrix Metalloproteinases Homo sapiens R-HSA-1592389 | 3.85E-01 | 6.92E-01 | 1.54E+00 | 1.47E+00 | COL18A1;TIMP2 |
| 538 | Vpr-mediated nuclear import of PICs Homo sapiens R-HSA-180910 | 3.85E-01 | 6.92E-01 | 1.54E+00 | 1.47E+00 | HMGA1;BANF1 |

| | | | | | | |
|---|---|---|---|---|---|---|
| 539 | RNA Polymerase I Transcription Termination Homo sapiens R-HSA-73863 | 3.85E-01 | 6.92E-01 | 1.54E+00 | 1.47E+00 | GTF2H2;GTF2H5 |
| 540 | Oncogene Induced Senescence Homo sapiens R-HSA-2559585 | 3.85E-01 | 6.92E-01 | 1.54E+00 | 1.47E+00 | TNRC6C;ID1 |
| 541 | SMAD2/SMAD3:SMAD4 heterotrimer regulates transcription Homo sapiens R-HSA-2173796 | 3.85E-01 | 6.92E-01 | 1.54E+00 | 1.47E+00 | JUNB;SMAD7 |
| 542 | Gluconeogenesis Homo sapiens R-HSA-70263 | 3.85E-01 | 6.92E-01 | 1.54E+00 | 1.47E+00 | GPI;ALDOB |
| 543 | Glycolysis Homo sapiens R-HSA-70171 | 3.85E-01 | 6.92E-01 | 1.54E+00 | 1.47E+00 | GPI;ALDOB |
| 544 | Signaling by Robo receptor Homo sapiens R-HSA-376176 | 3.85E-01 | 6.92E-01 | 1.54E+00 | 1.47E+00 | GPC1;PAK4 |
| 545 | DAG and IP3 signaling Homo sapiens R-HSA-1489509 | 3.85E-01 | 6.92E-01 | 1.54E+00 | 1.47E+00 | AHCYL1;PRKAR1B |
| 546 | Metabolism of carbohydrates Homo sapiens R-HSA-71387 | 3.87E-01 | 6.95E-01 | 1.12E+00 | 1.06E+00 | PRPS2;GPI;CHST9;SDC4;SORD;PYGL;BPGM;PAPSS2;HYAL1;GPC1;ST3GAL4;ALDOB;CHST2 |
| 547 | Potassium Channels Homo sapiens R-HSA-1296071 | 3.92E-01 | 6.97E-01 | 1.23E+00 | 1.15E+00 | KCNJ10;KCNK13;KCNMA1;KCNJ15;KCNJ1 |
| 548 | Reversible hydration of carbon dioxide Homo sapiens R-HSA-1475029 | 3.98E-01 | 6.97E-01 | 2.10E+00 | 1.94E+00 | CA12 |
| 549 | Hyaluronan uptake and degradation Homo sapiens R-HSA-2160916 | 3.98E-01 | 6.97E-01 | 2.10E+00 | 1.94E+00 | HYAL1 |
| 550 | Endosomal/Vacuolar pathway Homo sapiens R-HSA-1236977 | 3.98E-01 | 6.97E-01 | 2.10E+00 | 1.94E+00 | HLA-B |

| | | | | | | |
|---|---|---|---|---|---|---|
| 551 | Chk1/Chk2(Cds1) mediated inactivation of Cyclin B:Cdk1 complex Homo sapiens R-HSA-75035 | 3.98E-01 | 6.97E-01 | 2.10E+00 | 1.94E+00 | WEE1 |
| 552 | NOTCH2 intracellular domain regulates transcription Homo sapiens R-HSA-2197563 | 3.98E-01 | 6.97E-01 | 2.10E+00 | 1.94E+00 | MAML2 |
| 553 | Reduction of cytosolic Ca++ levels Homo sapiens R-HSA-418359 | 3.98E-01 | 6.97E-01 | 2.10E+00 | 1.94E+00 | ATP2B4 |
| 554 | The NLRP3 inflammasome Homo sapiens R-HSA-844456 | 3.98E-01 | 6.97E-01 | 2.10E+00 | 1.94E+00 | TXNIP |
| 555 | Tandem pore domain potassium channels Homo sapiens R-HSA-1296346 | 3.98E-01 | 6.97E-01 | 2.10E+00 | 1.94E+00 | KCNK13 |
| 556 | Sialic acid metabolism Homo sapiens R-HSA-4085001 | 4.00E-01 | 6.97E-01 | 1.49E+00 | 1.37E+00 | ST3GAL4;ST3GAL5 |
| 557 | CD28 co-stimulation Homo sapiens R-HSA-389356 | 4.00E-01 | 6.97E-01 | 1.49E+00 | 1.37E+00 | PIK3R2;PIK3R1 |
| 558 | Glucagon signaling in metabolic regulation Homo sapiens R-HSA-163359 | 4.00E-01 | 6.97E-01 | 1.49E+00 | 1.37E+00 | PRKAR1B;GCGR |
| 559 | Inactivation, recovery and regulation of the phototransduction cascade Homo sapiens R-HSA-2514859 | 4.00E-01 | 6.97E-01 | 1.49E+00 | 1.37E+00 | PDE6G;METAP2 |
| 560 | Stimuli-sensing channels Homo sapiens R-HSA-2672351 | 4.00E-01 | 6.97E-01 | 1.22E+00 | 1.12E+00 | CLCN5;WNK4;CLCNKB;NEDD4L;TRPM3 |
| 561 | Ion transport by P-type ATPases Homo sapiens R-HSA-936837 | 4.00E-01 | 6.97E-01 | 1.34E+00 | 1.22E+00 | ATP8B2;FXYD2;ATP2B4 |

| | | | | | | |
|---|---|---|---|---|---|---|
| 562 | Regulation of cholesterol biosynthesis by SREBP (SREBF) Homo sapiens R-HSA-1655829 | 4.00E-01 | 6.97E-01 | 1.34E+00 | 1.22E+00 | SREBF1;RXRA;TBL1XR1 |
| 563 | Biosynthesis of the N-glycan precursor (dolichol lipid-linked oligosaccharide, LLO) and transfer to a nascent protein Homo sapiens R-HSA-446193 | 4.06E-01 | 7.06E-01 | 1.25E+00 | 1.13E+00 | NUS1;PMM1;ST3GAL4;ST3GAL5 |
| 564 | Fatty acid, triacylglycerol, and ketone body metabolism Homo sapiens R-HSA-535734 | 4.12E-01 | 7.06E-01 | 1.12E+00 | 9.92E-01 | FABP1;SREBF1;RXRA;TBL1XR1;SCD5;ACOT11;LPIN1;PPARGC1A;FADS1;DECR1 |
| 565 | Negative regulation of FGFR2 signaling Homo sapiens R-HSA-5654727 | 4.14E-01 | 7.06E-01 | 1.45E+00 | 1.27E+00 | FGF9;FGFR2 |
| 566 | PLC-gamma1 signalling Homo sapiens R-HSA-167021 | 4.14E-01 | 7.06E-01 | 1.45E+00 | 1.27E+00 | AHCYL1;PRKAR1B |
| 567 | The phototransduction cascade Homo sapiens R-HSA-2514856 | 4.14E-01 | 7.06E-01 | 1.45E+00 | 1.27E+00 | PDE6G;METAP2 |
| 568 | Lipoprotein metabolism Homo sapiens R-HSA-174824 | 4.14E-01 | 7.06E-01 | 1.45E+00 | 1.27E+00 | ALB;APOE |
| 569 | Synthesis of PIPs at the plasma membrane Homo sapiens R-HSA-1660499 | 4.14E-01 | 7.06E-01 | 1.45E+00 | 1.27E+00 | PIK3R2;PIK3R1 |
| 570 | Glucose metabolism Homo sapiens R-HSA-70326 | 4.15E-01 | 7.06E-01 | 1.23E+00 | 1.08E+00 | GPI;PYGL;BPGM;ALDOB |
| 571 | Centrosome maturation Homo sapiens R-HSA-380287 | 4.15E-01 | 7.06E-01 | 1.23E+00 | 1.08E+00 | CNTRL;CEP131;CEP41;DYNLL1 |

| | | | | | | |
|---|---|---|---|---|---|---|
| 572 | Recruitment of mitotic centrosome proteins and complexes Homo sapiens R-HSA-380270 | 4.15E-01 | 7.06E-01 | 1.23E+00 | 1.08E+00 | CNTRL;CEP131;CEP41;DYNLL1 |
| 573 | Transcriptional regulation of white adipocyte differentiation Homo sapiens R-HSA-381340 | 4.15E-01 | 7.06E-01 | 1.23E+00 | 1.08E+00 | SREBF1;RXRA;TBL1XR1;PPARGC1A |
| 574 | Transmembrane transport of small molecules Homo sapiens R-HSA-382551 | 4.16E-01 | 7.06E-01 | 1.06E+00 | 9.31E-01 | SLC46A1;ABCB1;SLC47A1;SLC35B4;SLC20A1;SLC22A2;CLCNKB;WNK4;NEDD4L;SLC3A1;ADD3;NIPAL1;SLC22A17;FTH1;TRPM3;ATP8B2;SLC12A1;ATP2B4;CYBRD1;CLCN5;EIF2S3;PRKAR1B;FXYD2;SLCO2A1;ALB;FTL |
| 575 | Dissolution of Fibrin Clot Homo sapiens R-HSA-75205 | 4.23E-01 | 7.06E-01 | 1.93E+00 | 1.66E+00 | PLAU |
| 576 | AKT phosphorylates targets in the cytosol Homo sapiens R-HSA-198323 | 4.23E-01 | 7.06E-01 | 1.93E+00 | 1.66E+00 | CDKN1B |
| 577 | Activation of DNA fragmentation factor Homo sapiens R-HSA-211227 | 4.23E-01 | 7.06E-01 | 1.93E+00 | 1.66E+00 | HMGB2 |
| 578 | Apoptosis induced DNA fragmentation Homo sapiens R-HSA-140342 | 4.23E-01 | 7.06E-01 | 1.93E+00 | 1.66E+00 | HMGB2 |
| 579 | POU5F1 (OCT4), SOX2, NANOG activate genes related to proliferation Homo sapiens R-HSA-2892247 | 4.23E-01 | 7.06E-01 | 1.93E+00 | 1.66E+00 | SALL1 |
| 580 | p38MAPK events Homo sapiens R-HSA-171007 | 4.23E-01 | 7.06E-01 | 1.93E+00 | 1.66E+00 | MAPKAPK3 |
| 581 | Advanced glycosylation endproduct receptor signaling Homo sapiens R-HSA-879415 | 4.23E-01 | 7.06E-01 | 1.93E+00 | 1.66E+00 | LGALS3 |

| | | | | | | |
|---|---|---|---|---|---|---|
| 582 | Signal regulatory protein (SIRP) family interactions Homo sapiens R-HSA-391160 | 4.23E-01 | 7.06E-01 | 1.93E+00 | 1.66E+00 | SKAP2 |
| 583 | Depolarization of the Presynaptic Terminal Triggers the Opening of Calcium Channels Homo sapiens R-HSA-112308 | 4.23E-01 | 7.06E-01 | 1.93E+00 | 1.66E+00 | CACNA2D1 |
| 584 | Synthesis of PE Homo sapiens R-HSA-1483213 | 4.23E-01 | 7.06E-01 | 1.93E+00 | 1.66E+00 | LPIN1 |
| 585 | alpha-linolenic (omega3) and linoleic (omega6) acid metabolism Homo sapiens R-HSA-2046104 | 4.23E-01 | 7.06E-01 | 1.93E+00 | 1.66E+00 | FADS1 |
| 586 | alpha-linolenic acid (ALA) metabolism Homo sapiens R-HSA-2046106 | 4.23E-01 | 7.06E-01 | 1.93E+00 | 1.66E+00 | FADS1 |
| 587 | Purine salvage Homo sapiens R-HSA-74217 | 4.23E-01 | 7.06E-01 | 1.93E+00 | 1.66E+00 | ADAL |
| 588 | Regulation of insulin secretion Homo sapiens R-HSA-422356 | 4.25E-01 | 7.07E-01 | 1.22E+00 | 1.04E+00 | MARCKS;AHCYL1;PRKAR1B;VAMP2 |
| 589 | Lysosome Vesicle Biogenesis Homo sapiens R-HSA-432720 | 4.29E-01 | 7.11E-01 | 1.40E+00 | 1.19E+00 | HSPA8;VAMP2 |
| 590 | Interactions of Vpr with host cellular proteins Homo sapiens R-HSA-176033 | 4.29E-01 | 7.11E-01 | 1.40E+00 | 1.19E+00 | HMGA1;BANF1 |
| 591 | Post NMDA receptor activation events Homo sapiens R-HSA-438064 | 4.29E-01 | 7.11E-01 | 1.40E+00 | 1.19E+00 | NEFL;GRIN2B |
| 592 | Hemostasis Homo sapiens R-HSA-109582 | 4.35E-01 | 7.19E-01 | 1.05E+00 | 8.77E-01 | CFD;EGF;ATP2B4;PDGFA;AK3;PIK3R2;PIK3R1;GATA2;KNG1;RHOB;SOD1;WEE1;PRKAR1B;KIF26A;PLAU;KCNMA1;ALB;KIF5A;PLCG2;PDE3A;FLNA;QSOX1;MGLL;JAM3 |

| | | | | | | |
|---|---|---|---|---|---|---|
| 593 | Organic cation/anion/zwitterion transport Homo sapiens R-HSA-549132 | 4.47E-01 | 7.19E-01 | 1.78E+00 | 1.43E+00 | SLC22A2 |
| 594 | CS/DS degradation Homo sapiens R-HSA-2024101 | 4.47E-01 | 7.19E-01 | 1.78E+00 | 1.43E+00 | HYAL1 |
| 595 | TP53 regulates transcription of several additional cell death genes whose specific roles in p53-dependent apoptosis remain uncertain Homo sapiens R-HSA-6803205 | 4.47E-01 | 7.19E-01 | 1.78E+00 | 1.43E+00 | NDRG1 |
| 596 | Mismatch repair (MMR) directed by MSH2:MSH3 (MutSbeta) Homo sapiens R-HSA-5358606 | 4.47E-01 | 7.19E-01 | 1.78E+00 | 1.43E+00 | RPA1 |
| 597 | Mismatch repair (MMR) directed by MSH2:MSH6 (MutSalpha) Homo sapiens R-HSA-5358565 | 4.47E-01 | 7.19E-01 | 1.78E+00 | 1.43E+00 | RPA1 |
| 598 | Removal of the Flap Intermediate Homo sapiens R-HSA-69166 | 4.47E-01 | 7.19E-01 | 1.78E+00 | 1.43E+00 | RPA1 |
| 599 | Regulation of IFNG signaling Homo sapiens R-HSA-877312 | 4.47E-01 | 7.19E-01 | 1.78E+00 | 1.43E+00 | SOCS1 |
| 600 | FGFR4 ligand binding and activation Homo sapiens R-HSA-190322 | 4.47E-01 | 7.19E-01 | 1.78E+00 | 1.43E+00 | FGF9 |
| 601 | Golgi Cisternae Pericentriolar Stack Reorganization Homo sapiens R-HSA-162658 | 4.47E-01 | 7.19E-01 | 1.78E+00 | 1.43E+00 | RAB1B |

| | | | | | | |
|---|---|---|---|---|---|---|
| 602 | Bile salt and organic anion SLC transporters Homo sapiens R-HSA-425471 | 4.47E-01 | 7.19E-01 | 1.78E+00 | 1.43E+00 | SLC47A1 |
| 603 | DCC mediated attractive signaling Homo sapiens R-HSA-418885 | 4.47E-01 | 7.19E-01 | 1.78E+00 | 1.43E+00 | ABLIM1 |
| 604 | Methylation Homo sapiens R-HSA-156581 | 4.47E-01 | 7.19E-01 | 1.78E+00 | 1.43E+00 | MAT2A |
| 605 | Synthesis of bile acids and bile salts via 24-hydroxycholesterol Homo sapiens R-HSA-193775 | 4.47E-01 | 7.19E-01 | 1.78E+00 | 1.43E+00 | CYP27A1 |
| 606 | Nicotinate metabolism Homo sapiens R-HSA-196807 | 4.47E-01 | 7.19E-01 | 1.78E+00 | 1.43E+00 | NAPRT |
| 607 | Synthesis of Prostaglandins (PG) and Thromboxanes (TX) Homo sapiens R-HSA-2162123 | 4.47E-01 | 7.19E-01 | 1.78E+00 | 1.43E+00 | HPGD |
| 608 | Glutathione synthesis and recycling Homo sapiens R-HSA-174403 | 4.47E-01 | 7.19E-01 | 1.78E+00 | 1.43E+00 | OPLAH |
| 609 | Regulation of pyruvate dehydrogenase (PDH) complex Homo sapiens R-HSA-204174 | 4.47E-01 | 7.19E-01 | 1.78E+00 | 1.43E+00 | RXRA |
| 610 | Translocation of GLUT4 to the plasma membrane Homo sapiens R-HSA-1445148 | 4.56E-01 | 7.31E-01 | 1.22E+00 | 9.57E-01 | TBC1D1;PRKAB1;VAMP2 |
| 611 | Apoptotic cleavage of cellular proteins Homo sapiens R-HSA-111465 | 4.57E-01 | 7.31E-01 | 1.32E+00 | 1.03E+00 | CTNNB1;DSG2 |

| | | | | | | |
|---|---|---|---|---|---|---|
| 612 | Class B/2 (Secretin family receptors) Homo sapiens R-HSA-373080 | 4.62E-01 | 7.31E-01 | 1.16E+00 | 8.94E-01 | FZD1;GCGR;WNT9B;PTH1R |
| 613 | Disorders of transmembrane transporters Homo sapiens R-HSA-5619115 | 4.70E-01 | 7.31E-01 | 1.65E+00 | 1.25E+00 | ALB |
| 614 | N-Glycan antennae elongation Homo sapiens R-HSA-975577 | 4.70E-01 | 7.31E-01 | 1.65E+00 | 1.25E+00 | ST3GAL4 |
| 615 | Processing of Intronless Pre-mRNAs Homo sapiens R-HSA-77595 | 4.70E-01 | 7.31E-01 | 1.65E+00 | 1.25E+00 | NUDT21 |
| 616 | Mismatch Repair Homo sapiens R-HSA-5358508 | 4.70E-01 | 7.31E-01 | 1.65E+00 | 1.25E+00 | RPA1 |
| 617 | Processive synthesis on the lagging strand Homo sapiens R-HSA-69183 | 4.70E-01 | 7.31E-01 | 1.65E+00 | 1.25E+00 | RPA1 |
| 618 | Prolactin receptor signaling Homo sapiens R-HSA-1170546 | 4.70E-01 | 7.31E-01 | 1.65E+00 | 1.25E+00 | GHR |
| 619 | Glycogen breakdown (glycogenolysis) Homo sapiens R-HSA-70221 | 4.70E-01 | 7.31E-01 | 1.65E+00 | 1.25E+00 | PYGL |
| 620 | S33 mutants of beta-catenin aren't phosphorylated Homo sapiens R-HSA-5358747 | 4.70E-01 | 7.31E-01 | 1.65E+00 | 1.25E+00 | CTNNB1 |
| 621 | S45 mutants of beta-catenin aren't phosphorylated Homo sapiens R-HSA-5358751 | 4.70E-01 | 7.31E-01 | 1.65E+00 | 1.25E+00 | CTNNB1 |
| 622 | phosphorylation site mutants of CTNNB1 are not targeted to the proteasome by the destruction complex Homo sapiens R-HSA-4839743 | 4.70E-01 | 7.31E-01 | 1.65E+00 | 1.25E+00 | CTNNB1 |

| | | | | | | |
|---|---|---|---|---|---|---|
| 623 | T41 mutants of beta-catenin aren't phosphorylated Homo sapiens R-HSA-5358752 | 4.70E-01 | 7.31E-01 | 1.65E+00 | 1.25E+00 | CTNNB1 |
| 624 | S37 mutants of beta-catenin aren't phosphorylated Homo sapiens R-HSA-5358749 | 4.70E-01 | 7.31E-01 | 1.65E+00 | 1.25E+00 | CTNNB1 |
| 625 | Misspliced GSK3beta mutants stabilize beta-catenin Homo sapiens R-HSA-5339716 | 4.70E-01 | 7.31E-01 | 1.65E+00 | 1.25E+00 | CTNNB1 |
| 626 | FGFR1 ligand binding and activation Homo sapiens R-HSA-190242 | 4.70E-01 | 7.31E-01 | 1.65E+00 | 1.25E+00 | FGF9 |
| 627 | Negative regulation of TCF-dependent signaling by WNT ligand antagonists Homo sapiens R-HSA-3772470 | 4.70E-01 | 7.31E-01 | 1.65E+00 | 1.25E+00 | LRP5 |
| 628 | G1 Phase Homo sapiens R-HSA-69236 | 4.71E-01 | 7.31E-01 | 1.29E+00 | 9.67E-01 | CDKN1B;CCND1 |
| 629 | Cyclin D associated events in G1 Homo sapiens R-HSA-69231 | 4.71E-01 | 7.31E-01 | 1.29E+00 | 9.67E-01 | CDKN1B;CCND1 |
| 630 | MAP2K and MAPK activation Homo sapiens R-HSA-5674135 | 4.71E-01 | 7.31E-01 | 1.29E+00 | 9.67E-01 | MAP2K1;IL17RD |
| 631 | Glutathione conjugation Homo sapiens R-HSA-156590 | 4.71E-01 | 7.31E-01 | 1.29E+00 | 9.67E-01 | MGST1;OPLAH |
| 632 | Membrane Trafficking Homo sapiens R-HSA-199991 | 4.76E-01 | 7.38E-01 | 1.04E+00 | 7.68E-01 | TPD52;HSPA8;RAB1B;GOLIM4;HIP1R;DYNLL1;PRKAB1;DYNLL2;BICD1;TBC1D1;BLOC1S6;KIF26A;FTH1;KIF5A;VAMP2;CTSC;FTL;PICALM |
| 633 | Integration of energy metabolism Homo sapiens R-HSA-163685 | 4.82E-01 | 7.44E-01 | 1.10E+00 | 8.05E-01 | MARCKS;AHCYL1;PRKAR1B;GCGR;VAMP2 |
| 634 | GABA B receptor activation Homo sapiens R-HSA-977444 | 4.85E-01 | 7.44E-01 | 1.25E+00 | 9.06E-01 | KCNJ10;KCNJ15 |

| | | | | | | |
|---|---|---|---|---|---|---|
| 635 | Activation of GABAB receptors Homo sapiens R-HSA-991365 | 4.85E-01 | 7.44E-01 | 1.25E+00 | 9.06E-01 | KCNJ10;KCNJ15 |
| 636 | Activation of NMDA receptor upon glutamate binding and postsynaptic events Homo sapiens R-HSA-442755 | 4.85E-01 | 7.44E-01 | 1.25E+00 | 9.06E-01 | NEFL;GRIN2B |
| 637 | Regulation of PLK1 Activity at G2/M Transition Homo sapiens R-HSA-2565942 | 4.89E-01 | 7.44E-01 | 1.12E+00 | 7.98E-01 | CNTRL;CEP131;CEP41;DYNLL1 |
| 638 | Metabolism of nucleotides Homo sapiens R-HSA-15869 | 4.89E-01 | 7.44E-01 | 1.12E+00 | 7.98E-01 | ADAL;CDA;GLRX;NUDT16 |
| 639 | Synthesis of glycosylphosphatidylinositol (GPI) Homo sapiens R-HSA-162710 | 4.92E-01 | 7.44E-01 | 1.54E+00 | 1.09E+00 | PIGQ |
| 640 | Translesion synthesis by REV1 Homo sapiens R-HSA-110312 | 4.92E-01 | 7.44E-01 | 1.54E+00 | 1.09E+00 | RPA1 |
| 641 | TRAF6 mediated induction of TAK1 complex Homo sapiens R-HSA-937072 | 4.92E-01 | 7.44E-01 | 1.54E+00 | 1.09E+00 | TAB2 |
| 642 | JNK (c-Jun kinases) phosphorylation and activation mediated by activated human TAK1 Homo sapiens R-HSA-450321 | 4.92E-01 | 7.44E-01 | 1.54E+00 | 1.09E+00 | TAB2 |
| 643 | MAP3K8 (TPL2)-dependent MAPK1/3 activation Homo sapiens R-HSA-5684264 | 4.92E-01 | 7.44E-01 | 1.54E+00 | 1.09E+00 | MAP2K1 |
| 644 | Polo-like kinase mediated events Homo sapiens R-HSA-156711 | 4.92E-01 | 7.44E-01 | 1.54E+00 | 1.09E+00 | WEE1 |

| | | | | | | |
|---|---|---|---|---|---|---|
| 645 | Cell-extracellular matrix interactions Homo sapiens R-HSA-446353 | 4.92E-01 | 7.44E-01 | 1.54E+00 | 1.09E+00 | FLNA |
| 646 | Regulation of KIT signaling Homo sapiens R-HSA-1433559 | 4.92E-01 | 7.44E-01 | 1.54E+00 | 1.09E+00 | SOCS1 |
| 647 | Rap1 signalling Homo sapiens R-HSA-392517 | 4.92E-01 | 7.44E-01 | 1.54E+00 | 1.09E+00 | RAP1GAP2 |
| 648 | PKA activation Homo sapiens R-HSA-163615 | 4.92E-01 | 7.44E-01 | 1.54E+00 | 1.09E+00 | PRKAR1B |
| 649 | Adaptive Immune System Homo sapiens R-HSA-1280218 | 4.96E-01 | 7.48E-01 | 1.01E+00 | 7.12E-01 | CDKN1B;AHCYL1;UBE3D;NEDD4L;PDGFA;PIK3R2;PIK3R1;FOXO3;FOXO1;SOCS1;FGF9;ERBB4;SEC61G;KIF5A;PLCG2;CTSF;CTSD;PAG1;CTSC;RAP1GAP2;WSB1;EGF;HLA-B;DYNLL1;DYNLL2;PJA1;TNRC6C;KIF26A;TAB2;FBXL4;FGFR3;FGFR2 |
| 650 | PKMTs methylate histone lysines Homo sapiens R-HSA-3214841 | 4.98E-01 | 7.50E-01 | 1.22E+00 | 8.48E-01 | ATF7IP;SETD7 |
| 651 | RNA Polymerase II Pre-transcription Events Homo sapiens R-HSA-674695 | 4.98E-01 | 7.50E-01 | 1.14E+00 | 7.92E-01 | POLR2A;GTF2H2;GTF2H5 |
| 652 | Intraflagellar transport Homo sapiens R-HSA-5620924 | 5.11E-01 | 7.54E-01 | 1.19E+00 | 7.96E-01 | DYNLL1;DYNLL2 |
| 653 | Histidine, lysine, phenylalanine, tyrosine, proline and tryptophan catabolism Homo sapiens R-HSA-6788656 | 5.11E-01 | 7.54E-01 | 1.19E+00 | 7.96E-01 | ALDH4A1;FAH |
| 654 | Hyaluronan metabolism Homo sapiens R-HSA-2142845 | 5.13E-01 | 7.54E-01 | 1.45E+00 | 9.64E-01 | HYAL1 |
| 655 | Butyrate Response Factor 1 (BRF1) binds and destabilizes mRNA Homo sapiens R-HSA-450385 | 5.13E-01 | 7.54E-01 | 1.45E+00 | 9.64E-01 | ZFP36L1 |

| | | | | | | |
|---|---|---|---|---|---|---|
| 656 | Formation of Senescence-Associated Heterochromatin Foci (SAHF) Homo sapiens R-HSA-2559584 | 5.13E-01 | 7.54E-01 | 1.45E+00 | 9.64E-01 | HMGA1 |
| 657 | Translesion synthesis by POLK Homo sapiens R-HSA-5655862 | 5.13E-01 | 7.54E-01 | 1.45E+00 | 9.64E-01 | RPA1 |
| 658 | Translesion synthesis by POLI Homo sapiens R-HSA-5656121 | 5.13E-01 | 7.54E-01 | 1.45E+00 | 9.64E-01 | RPA1 |
| 659 | Beta-catenin phosphorylation cascade Homo sapiens R-HSA-196299 | 5.13E-01 | 7.54E-01 | 1.45E+00 | 9.64E-01 | CTNNB1 |
| 660 | RHO GTPases Activate ROCKs Homo sapiens R-HSA-5627117 | 5.13E-01 | 7.54E-01 | 1.45E+00 | 9.64E-01 | RHOB |
| 661 | PKA-mediated phosphorylation of CREB Homo sapiens R-HSA-111931 | 5.13E-01 | 7.54E-01 | 1.45E+00 | 9.64E-01 | PRKAR1B |
| 662 | PKA activation in glucagon signalling Homo sapiens R-HSA-164378 | 5.13E-01 | 7.54E-01 | 1.45E+00 | 9.64E-01 | PRKAR1B |
| 663 | Chylomicron-mediated lipid transport Homo sapiens R-HSA-174800 | 5.13E-01 | 7.54E-01 | 1.45E+00 | 9.64E-01 | APOE |
| 664 | Acyl chain remodelling of PS Homo sapiens R-HSA-1482801 | 5.13E-01 | 7.54E-01 | 1.45E+00 | 9.64E-01 | MBOAT1 |
| 665 | Mitochondrial Fatty Acid Beta-Oxidation Homo sapiens R-HSA-77289 | 5.13E-01 | 7.54E-01 | 1.45E+00 | 9.64E-01 | DECR1 |
| 666 | Metabolism of porphyrins Homo sapiens R-HSA-189445 | 5.13E-01 | 7.54E-01 | 1.45E+00 | 9.64E-01 | FECH |

| | | | | | | |
|---|---|---|---|---|---|---|
| 667 | Apoptosis Homo sapiens R-HSA-109581 | 5.16E-01 | 7.58E-01 | 1.04E+00 | 6.86E-01 | HMGB2;CYCS;CTNNB1;DSG2;DYNLL1;DYNLL2;BCL2L1 |
| 668 | Triglyceride Biosynthesis Homo sapiens R-HSA-75109 | 5.19E-01 | 7.61E-01 | 1.10E+00 | 7.22E-01 | ACOT11;SCD5;LPIN1 |
| 669 | Non-integrin membrane-ECM interactions Homo sapiens R-HSA-3000171 | 5.24E-01 | 7.66E-01 | 1.16E+00 | 7.47E-01 | SDC4;PDGFA |
| 670 | Fatty Acyl-CoA Biosynthesis Homo sapiens R-HSA-75105 | 5.24E-01 | 7.66E-01 | 1.16E+00 | 7.47E-01 | ACOT11;SCD5 |
| 671 | Gastrin-CREB signalling pathway via PKC and MAPK Homo sapiens R-HSA-881907 | 5.25E-01 | 7.66E-01 | 1.01E+00 | 6.47E-01 | PTGFR;DUSP2;MAP2K1;EGF;GCGR;PDGFA;PIK3R2;PIK3R1;IL17RD;GRIN2B;DUSP9;KNG1;FGF9;ERBB4;NEFL;FGFR3;MGLL;FGFR2 |
| 672 | Transcription of the HIV genome Homo sapiens R-HSA-167172 | 5.29E-01 | 7.68E-01 | 1.08E+00 | 6.90E-01 | POLR2A;GTF2H2;GTF2H5 |
| 673 | Factors involved in megakaryocyte development and platelet production Homo sapiens R-HSA-983231 | 5.32E-01 | 7.68E-01 | 1.03E+00 | 6.48E-01 | WEE1;PRKAR1B;KIF26A;KIF5A;AK3;GATA2 |
| 674 | HIV Life Cycle Homo sapiens R-HSA-162587 | 5.32E-01 | 7.68E-01 | 1.03E+00 | 6.48E-01 | POLR2A;HMGA1;NEDD4L;BANF1;GTF2H2;GTF2H5 |
| 675 | MAPK6/MAPK4 signaling Homo sapiens R-HSA-5687128 | 5.33E-01 | 7.68E-01 | 1.05E+00 | 6.61E-01 | TNRC6C;JUN;FOXO3;FOXO1 |
| 676 | Defective GALNT12 causes colorectal cancer 1 (CRCS1) Homo sapiens R-HSA-5083636 | 5.33E-01 | 7.68E-01 | 1.36E+00 | 8.55E-01 | MUC20 |
| 677 | Defective GALNT3 causes familial hyperphosphatemic tumoral calcinosis (HFTC) Homo sapiens R-HSA-5083625 | 5.33E-01 | 7.68E-01 | 1.36E+00 | 8.55E-01 | MUC20 |

| | | | | | | |
|---|---|---|---|---|---|---|
| 678 | Activation of SMO Homo sapiens R-HSA-5635838 | 5.33E-01 | 7.68E-01 | 1.36E+00 | 8.55E-01 | GAS1 |
| 679 | TP53 Regulates Transcription of Genes Involved in G2 Cell Cycle Arrest Homo sapiens R-HSA-6804114 | 5.33E-01 | 7.68E-01 | 1.36E+00 | 8.55E-01 | GADD45A |
| 680 | Amine-derived hormones Homo sapiens R-HSA-209776 | 5.33E-01 | 7.68E-01 | 1.36E+00 | 8.55E-01 | DDC |
| 681 | Programmed Cell Death Homo sapiens R-HSA-5357801 | 5.36E-01 | 7.70E-01 | 1.02E+00 | 6.36E-01 | HMGB2;CYCS;CTNNB1;DSG2;DYNLL1;DYNLL2;BCL2L1 |
| 682 | Transport of Mature mRNA derived from an Intron-Containing Transcript Homo sapiens R-HSA-159236 | 5.39E-01 | 7.72E-01 | 1.07E+00 | 6.59E-01 | SRSF3;SRSF5;SRSF6 |
| 683 | Interferon alpha/beta signaling Homo sapiens R-HSA-909733 | 5.39E-01 | 7.72E-01 | 1.07E+00 | 6.59E-01 | EGR1;SOCS1;HLA-B |
| 684 | Cyclin E associated events during G1/S transition Homo sapiens R-HSA-69202 | 5.39E-01 | 7.72E-01 | 1.07E+00 | 6.59E-01 | WEE1;CDKN1B;CCND1 |
| 685 | Interferon gamma signaling Homo sapiens R-HSA-877300 | 5.42E-01 | 7.74E-01 | 1.04E+00 | 6.37E-01 | MT2A;SOCS1;TRIM2;HLA-B |
| 686 | Cyclin A:Cdk2-associated events at S phase entry Homo sapiens R-HSA-69656 | 5.49E-01 | 7.79E-01 | 1.05E+00 | 6.30E-01 | WEE1;CDKN1B;CCND1 |
| 687 | Kinesins Homo sapiens R-HSA-983189 | 5.50E-01 | 7.79E-01 | 1.10E+00 | 6.59E-01 | KIF26A;KIF5A |
| 688 | PLC beta mediated events Homo sapiens R-HSA-112043 | 5.50E-01 | 7.79E-01 | 1.10E+00 | 6.59E-01 | AHCYL1;PRKAR1B |
| 689 | Na+/Cl- dependent neurotransmitter transporters Homo sapiens R-HSA-442660 | 5.53E-01 | 7.79E-01 | 1.29E+00 | 7.62E-01 | SLC22A2 |

| | | | | | | |
|---|---|---|---|---|---|---|
| 690 | Translesion Synthesis by POLH Homo sapiens R-HSA-110320 | 5.53E-01 | 7.79E-01 | 1.29E+00 | 7.62E-01 | RPA1 |
| 691 | PCNA-Dependent Long Patch Base Excision Repair Homo sapiens R-HSA-5651801 | 5.53E-01 | 7.79E-01 | 1.29E+00 | 7.62E-01 | RPA1 |
| 692 | Signaling by NODAL Homo sapiens R-HSA-1181150 | 5.53E-01 | 7.79E-01 | 1.29E+00 | 7.62E-01 | FOXO3 |
| 693 | Citric acid cycle (TCA cycle) Homo sapiens R-HSA-71403 | 5.53E-01 | 7.79E-01 | 1.29E+00 | 7.62E-01 | IDH2 |
| 694 | Phase 4 - resting membrane potential Homo sapiens R-HSA-5576886 | 5.53E-01 | 7.79E-01 | 1.29E+00 | 7.62E-01 | KCNK13 |
| 695 | Beta-catenin independent WNT signaling Homo sapiens R-HSA-3858494 | 5.53E-01 | 7.79E-01 | 1.01E+00 | 5.95E-01 | FZD1;TNRC6C;TCF7L2;VANGL2;PDE6G;CTNNB1 |
| 696 | EPH-Ephrin signaling Homo sapiens R-HSA-2682334 | 5.59E-01 | 7.85E-01 | 1.02E+00 | 5.92E-01 | EFNA5;GRIN2B;ACTB;ACTG1 |
| 697 | Signaling by Interleukins Homo sapiens R-HSA-449147 | 5.61E-01 | 7.85E-01 | 9.84E-01 | 5.68E-01 | DUSP2;MAP2K1;EGF;PELI2;PDGFA;PIK3R2;PIK3R1;IL17RD;GRIN2B;DUSP9;FGF9;ERBB4;NEFL;TAB2;FGFR3;FGFR2 |
| 698 | ABC-family proteins mediated transport Homo sapiens R-HSA-382556 | 5.62E-01 | 7.85E-01 | 1.08E+00 | 6.20E-01 | ABCB1;EIF2S3 |
| 699 | RMTs methylate histone arginines Homo sapiens R-HSA-3214858 | 5.62E-01 | 7.85E-01 | 1.08E+00 | 6.20E-01 | CCND1;ARID1A |
| 700 | G-protein mediated events Homo sapiens R-HSA-112040 | 5.62E-01 | 7.85E-01 | 1.08E+00 | 6.20E-01 | AHCYL1;PRKAR1B |
| 701 | NRAGE signals death through JNK Homo sapiens R-HSA-193648 | 5.62E-01 | 7.85E-01 | 1.08E+00 | 6.20E-01 | FGD3;OBSCN |

| | | | | | | |
|---|---|---|---|---|---|---|
| 702 | Chondroitin sulfate biosynthesis Homo sapiens R-HSA-2022870 | 5.71E-01 | 7.91E-01 | 1.22E+00 | 6.81E-01 | CHST9 |
| 703 | Defective C1GALT1C1 causes Tn polyagglutination syndrome (TNPS) Homo sapiens R-HSA-5083632 | 5.71E-01 | 7.91E-01 | 1.22E+00 | 6.81E-01 | MUC20 |
| 704 | Syndecan interactions Homo sapiens R-HSA-3000170 | 5.71E-01 | 7.91E-01 | 1.22E+00 | 6.81E-01 | SDC4 |
| 705 | Lagging Strand Synthesis Homo sapiens R-HSA-69186 | 5.71E-01 | 7.91E-01 | 1.22E+00 | 6.81E-01 | RPA1 |
| 706 | Synthesis of PC Homo sapiens R-HSA-1483191 | 5.71E-01 | 7.91E-01 | 1.22E+00 | 6.81E-01 | LPIN1 |
| 707 | Glucuronidation Homo sapiens R-HSA-156588 | 5.71E-01 | 7.91E-01 | 1.22E+00 | 6.81E-01 | UGT2B7 |
| 708 | Scavenging of heme from plasma Homo sapiens R-HSA-2168880 | 5.74E-01 | 7.94E-01 | 1.05E+00 | 5.84E-01 | LRP1;ALB |
| 709 | Anchoring of the basal body to the plasma membrane Homo sapiens R-HSA-5620912 | 5.75E-01 | 7.94E-01 | 9.95E-01 | 5.50E-01 | CEP131;CNTRL;CEP41;DYNLL1 |
| 710 | RNA Polymerase I Transcription Initiation Homo sapiens R-HSA-73762 | 5.86E-01 | 8.06E-01 | 1.03E+00 | 5.50E-01 | GTF2H2;GTF2H5 |
| 711 | TP53 regulates transcription of additional cell cycle genes whose exact role in the p53 pathway remain uncertain Homo sapiens R-HSA-6804115 | 5.89E-01 | 8.06E-01 | 1.16E+00 | 6.12E-01 | PLAGL1 |
| 712 | SHC-mediated cascade:FGFR1 Homo sapiens R-HSA-5654688 | 5.89E-01 | 8.06E-01 | 1.16E+00 | 6.12E-01 | FGF9 |

| | | | | | | |
|---|---|---|---|---|---|---|
| 713 | ERK/MAPK targets Homo sapiens R-HSA-198753 | 5.89E-01 | 8.06E-01 | 1.16E+00 | 6.12E-01 | MEF2A |
| 714 | RHO GTPases activate PAKs Homo sapiens R-HSA-5627123 | 5.89E-01 | 8.06E-01 | 1.16E+00 | 6.12E-01 | FLNA |
| 715 | Cobalamin (Cbl, vitamin B12) transport and metabolism Homo sapiens R-HSA-196741 | 5.89E-01 | 8.06E-01 | 1.16E+00 | 6.12E-01 | MMAB |
| 716 | Aflatoxin activation and detoxification Homo sapiens R-HSA-5423646 | 5.89E-01 | 8.06E-01 | 1.16E+00 | 6.12E-01 | MGST1 |
| 717 | Chromatin organization Homo sapiens R-HSA-4839726 | 5.96E-01 | 8.11E-01 | 9.59E-01 | 4.96E-01 | ATF7IP;BRPF1;CCND1;TBL1XR1;SETD7;JADE1;ARID5B;ARID1A;ACTB |
| 718 | Chromatin modifying enzymes Homo sapiens R-HSA-3247509 | 5.96E-01 | 8.11E-01 | 9.59E-01 | 4.96E-01 | ATF7IP;BRPF1;CCND1;TBL1XR1;SETD7;JADE1;ARID5B;ARID1A;ACTB |
| 719 | Sphingolipid metabolism Homo sapiens R-HSA-428157 | 5.97E-01 | 8.11E-01 | 9.77E-01 | 5.04E-01 | CERS4;SPTLC2;SGMS1 |
| 720 | Transport of Mature Transcript to Cytoplasm Homo sapiens R-HSA-72202 | 5.97E-01 | 8.11E-01 | 9.77E-01 | 5.04E-01 | SRSF3;SRSF5;SRSF6 |
| 721 | G beta:gamma signalling through PI3Kgamma Homo sapiens R-HSA-392451 | 5.97E-01 | 8.11E-01 | 1.01E+00 | 5.18E-01 | PIK3R2;PIK3R1 |
| 722 | RA biosynthesis pathway Homo sapiens R-HSA-5365859 | 6.06E-01 | 8.14E-01 | 1.10E+00 | 5.51E-01 | ADH1C |
| 723 | Intrinsic Pathway of Fibrin Clot Formation Homo sapiens R-HSA-140837 | 6.06E-01 | 8.14E-01 | 1.10E+00 | 5.51E-01 | KNG1 |

| | | | | | | |
|---|---|---|---|---|---|---|
| 724 | mTORC1-mediated signalling Homo sapiens R-HSA-166208 | 6.06E-01 | 8.14E-01 | 1.10E+00 | 5.51E-01 | RRAGD |
| 725 | Resolution of AP sites via the multiple-nucleotide patch replacement pathway Homo sapiens R-HSA-110373 | 6.06E-01 | 8.14E-01 | 1.10E+00 | 5.51E-01 | RPA1 |
| 726 | Telomere C-strand (Lagging Strand) Synthesis Homo sapiens R-HSA-174417 | 6.06E-01 | 8.14E-01 | 1.10E+00 | 5.51E-01 | RPA1 |
| 727 | Cyclin A/B1 associated events during G2/M transition Homo sapiens R-HSA-69273 | 6.06E-01 | 8.14E-01 | 1.10E+00 | 5.51E-01 | WEE1 |
| 728 | Post-chaperonin tubulin folding pathway Homo sapiens R-HSA-389977 | 6.06E-01 | 8.14E-01 | 1.10E+00 | 5.51E-01 | TBCA |
| 729 | SALM protein interactions at the synapse Homo sapiens R-HSA-8849932 | 6.06E-01 | 8.14E-01 | 1.10E+00 | 5.51E-01 | GRIN2B |
| 730 | Ion channel transport Homo sapiens R-HSA-983712 | 6.08E-01 | 8.16E-01 | 9.48E-01 | 4.72E-01 | CLCN5;ATP8B2;FXYD2;WNK4;CLCNKB;ATP2B4;NEDD4L;TRPM3 |
| 731 | Glycerophospholipid biosynthesis Homo sapiens R-HSA-1483206 | 6.15E-01 | 8.19E-01 | 9.44E-01 | 4.59E-01 | MBOAT1;GPCPD1;LPIN1;MGLL |
| 732 | Intra-Golgi and retrograde Golgi-to-ER traffic Homo sapiens R-HSA-6811442 | 6.16E-01 | 8.19E-01 | 9.41E-01 | 4.56E-01 | KIF26A;RAB1B;KIF5A;GOLIM4;DYNLL1;BICD1;DYNLL2 |
| 733 | Laminin interactions Homo sapiens R-HSA-3000157 | 6.23E-01 | 8.19E-01 | 1.05E+00 | 4.98E-01 | COL18A1 |
| 734 | Deadenylation of mRNA Homo sapiens R-HSA-429947 | 6.23E-01 | 8.19E-01 | 1.05E+00 | 4.98E-01 | PABPC1 |

| | | | | | | |
|---|---|---|---|---|---|---|
| 735 | Regulation of TP53 Activity through Acetylation Homo sapiens R-HSA-6804758 | 6.23E-01 | 8.19E-01 | 1.05E+00 | 4.98E-01 | BRPF1 |
| 736 | RNA Polymerase III Transcription Termination Homo sapiens R-HSA-73980 | 6.23E-01 | 8.19E-01 | 1.05E+00 | 4.98E-01 | NFIC |
| 737 | MicroRNA (miRNA) biogenesis Homo sapiens R-HSA-203927 | 6.23E-01 | 8.19E-01 | 1.05E+00 | 4.98E-01 | POLR2A |
| 738 | Abortive elongation of HIV-1 transcript in the absence of Tat Homo sapiens R-HSA-167242 | 6.23E-01 | 8.19E-01 | 1.05E+00 | 4.98E-01 | POLR2A |
| 739 | Gap-filling DNA repair synthesis and ligation in GG-NER Homo sapiens R-HSA-5696397 | 6.23E-01 | 8.19E-01 | 1.05E+00 | 4.98E-01 | RPA1 |
| 740 | Downregulation of SMAD2/3:SMAD4 transcriptional activity Homo sapiens R-HSA-2173795 | 6.23E-01 | 8.19E-01 | 1.05E+00 | 4.98E-01 | NEDD4L |
| 741 | SHC-mediated cascade:FGFR4 Homo sapiens R-HSA-5654719 | 6.23E-01 | 8.19E-01 | 1.05E+00 | 4.98E-01 | FGF9 |
| 742 | Acyl chain remodelling of PE Homo sapiens R-HSA-1482839 | 6.23E-01 | 8.19E-01 | 1.05E+00 | 4.98E-01 | MBOAT1 |
| 743 | Cholesterol biosynthesis Homo sapiens R-HSA-191273 | 6.23E-01 | 8.19E-01 | 1.05E+00 | 4.98E-01 | DHCR24 |
| 744 | Branched-chain amino acid catabolism Homo sapiens R-HSA-70895 | 6.23E-01 | 8.19E-01 | 1.05E+00 | 4.98E-01 | ALDH6A1 |
| 745 | Senescence-Associated Secretory Phenotype (SASP) Homo sapiens R-HSA-2559582 | 6.24E-01 | 8.20E-01 | 9.37E-01 | 4.42E-01 | JUN;CDKN1B;FOS |

| | | | | | | |
|---|---|---|---|---|---|---|
| 746 | G-protein beta:gamma signalling Homo sapiens R-HSA-397795 | 6.30E-01 | 8.27E-01 | 9.44E-01 | 4.36E-01 | PIK3R2;PIK3R1 |
| 747 | Metabolism of lipids and lipoproteins Homo sapiens R-HSA-556833 | 6.31E-01 | 8.28E-01 | 9.48E-01 | 4.36E-01 | SGMS1;HPGD;PIK3R2;PIK3R1;RXRA;SPTLC2;APOE;PPARGC1A;FADS1;DECR1;CERS4;SREBF1;EPHX2;SCD5;ACOT11;MBOAT1;DHCR24;GPCPD1;FABP1;CYP27A1;FABP3;TBL1XR1;EHHADH;ALB;LPIN1;MGLL |
| 748 | COPI-mediated anterograde transport Homo sapiens R-HSA-6807878 | 6.33E-01 | 8.28E-01 | 9.25E-01 | 4.23E-01 | RAB1B;DYNLL1;DYNLL2 |
| 749 | Phosphorylation of CD3 and TCR zeta chains Homo sapiens R-HSA-202427 | 6.38E-01 | 8.28E-01 | 1.01E+00 | 4.52E-01 | PAG1 |
| 750 | Post-Elongation Processing of Intronless pre-mRNA Homo sapiens R-HSA-112297 | 6.38E-01 | 8.28E-01 | 1.01E+00 | 4.52E-01 | NUDT21 |
| 751 | Processing of Capped Intronless Pre-mRNA Homo sapiens R-HSA-75067 | 6.38E-01 | 8.28E-01 | 1.01E+00 | 4.52E-01 | NUDT21 |
| 752 | DARPP-32 events Homo sapiens R-HSA-180024 | 6.38E-01 | 8.28E-01 | 1.01E+00 | 4.52E-01 | PRKAR1B |
| 753 | Nuclear Events (kinase and transcription factor activation) Homo sapiens R-HSA-198725 | 6.38E-01 | 8.28E-01 | 1.01E+00 | 4.52E-01 | MEF2A |
| 754 | Sema4D induced cell migration and growth-cone collapse Homo sapiens R-HSA-416572 | 6.38E-01 | 8.28E-01 | 1.01E+00 | 4.52E-01 | RHOB |
| 755 | Pyrimidine metabolism Homo sapiens R-HSA-73848 | 6.38E-01 | 8.28E-01 | 1.01E+00 | 4.52E-01 | CDA |

| | | | | | | |
|---|---|---|---|---|---|---|
| 756 | Signal Transduction Homo sapiens R-HSA-162582 | 6.50E-01 | 8.32E-01 | 9.63E-01 | 4.15E-01 | CYFIP2;AHCYL1;MAML2;WWC1;ACTB;ACTG1;ARHGAP44;CCND1;FGF9;KIF5A;JUNB;SOX4;MEF2A;TLE3;MAP2K1;SOX13;PRKAB1;OBSCN;PRKAR1B;MAPKAPK3;COL4A2;COL4A1;TBL1XR1;CSNK2B;RRAGD;PTGFR;NOTCH1;CRABP2;SDC4;HPN;PDGFA;NEDD4L;PIK3R2;PIK3R1;FOXO3;FOXO1;KNG1;SOCS1;FLRT1;ST3GAL4;APOE;PAG1;FZD1;SMAD1;JUN;JAG1;ARHGAP29;GRIN2B;ARHGAP24;SMAD7;BCL9;VANGL2;BAMBI;CCDC88C;PDE3A;AMOTL2;TAB2;COL9A2;AMOTL1;FGFR3;FGFR2;GPR27;CDKN1B;PTH1R;GHR;GRM3;NEFL;DUSP2;WNT9B;IL17RD;DUSP9;RHOB;HNRNPH1;COL6A1;GAS1;MGLL;PPP1R15A;LRP1;CAB39;ADH1C;PTGER3;LRP5;FGD3;ZNRF3;RXRA;POLR2A;ERBB4;GPC1;SPP1;FLNA;TCF7L2;EGF;PDE6G;GCGR;MIB1;TNRC6C;CTNNB1;SPRY1;METAP2 |
| 757 | XBP1(S) activates chaperone genes Homo sapiens R-HSA-381038 | 6.51E-01 | 8.32E-01 | 9.07E-01 | 3.89E-01 | SEC61G;CTDSP2 |
| 758 | Arachidonic acid metabolism Homo sapiens R-HSA-2142753 | 6.51E-01 | 8.32E-01 | 9.07E-01 | 3.89E-01 | HPGD;EPHX2 |
| 759 | Mitotic Prophase Homo sapiens R-HSA-68875 | 6.53E-01 | 8.32E-01 | 8.98E-01 | 3.83E-01 | RAB1B;BANF1;LPIN1;EMD |
| 760 | Post-translational modification: synthesis of GPI-anchored proteins Homo sapiens R-HSA-163125 | 6.53E-01 | 8.32E-01 | 9.64E-01 | 4.10E-01 | PIGQ |
| 761 | Antigen Presentation: Folding, assembly and peptide loading of class I MHC Homo sapiens R-HSA-983170 | 6.53E-01 | 8.32E-01 | 9.64E-01 | 4.10E-01 | HLA-B |
| 762 | HDMs demethylate histones Homo sapiens R-HSA-3214842 | 6.53E-01 | 8.32E-01 | 9.64E-01 | 4.10E-01 | ARID5B |

| | | | | | | |
|---|---|---|---|---|---|---|
| 763 | Regulation of IFNA signaling Homo sapiens R-HSA-912694 | 6.53E-01 | 8.32E-01 | 9.64E-01 | 4.10E-01 | SOCS1 |
| 764 | Budding and maturation of HIV virion Homo sapiens R-HSA-162588 | 6.53E-01 | 8.32E-01 | 9.64E-01 | 4.10E-01 | NEDD4L |
| 765 | RAF activation Homo sapiens R-HSA-5673000 | 6.53E-01 | 8.32E-01 | 9.64E-01 | 4.10E-01 | MAP2K1 |
| 766 | TRP channels Homo sapiens R-HSA-3295583 | 6.53E-01 | 8.32E-01 | 9.64E-01 | 4.10E-01 | TRPM3 |
| 767 | Sulfur amino acid metabolism Homo sapiens R-HSA-1614635 | 6.53E-01 | 8.32E-01 | 9.64E-01 | 4.10E-01 | CDO1 |
| 768 | Synthesis and interconversion of nucleotide di- and triphosphates Homo sapiens R-HSA-499943 | 6.53E-01 | 8.32E-01 | 9.64E-01 | 4.10E-01 | GLRX |
| 769 | Glyoxylate metabolism and glycine degradation Homo sapiens R-HSA-389661 | 6.53E-01 | 8.32E-01 | 9.64E-01 | 4.10E-01 | GLDC |
| 770 | COPI-dependent Golgi-to-ER retrograde traffic Homo sapiens R-HSA-6811434 | 6.58E-01 | 8.34E-01 | 8.89E-01 | 3.72E-01 | KIF26A;RAB1B;KIF5A |
| 771 | Heparan sulfate/heparin (HS-GAG) metabolism Homo sapiens R-HSA-1638091 | 6.61E-01 | 8.34E-01 | 8.89E-01 | 3.68E-01 | SDC4;GPC1 |
| 772 | AUF1 (hnRNP D0) binds and destabilizes mRNA Homo sapiens R-HSA-450408 | 6.61E-01 | 8.34E-01 | 8.89E-01 | 3.68E-01 | HSPA8;PABPC1 |
| 773 | Phospholipid metabolism Homo sapiens R-HSA-1483257 | 6.62E-01 | 8.34E-01 | 8.94E-01 | 3.70E-01 | MBOAT1;PIK3R2;PIK3R1;GPCPD1;LPIN1;MGLL |

| | | | | | | |
|---|---|---|---|---|---|---|
| 774 | Signaling by Rho GTPases Homo sapiens R-HSA-194315 | 6.63E-01 | 8.34E-01 | 9.16E-01 | 3.76E-01 | CYFIP2;CDKN1B;ARHGAP29;PIK3R2;ACTB;ARHGAP24;RHOB;ACTG1;FGD3;ARHGAP44;OBSCN;KIF5A;FLNA;CTNNB1 |
| 775 | Cytokine Signaling in Immune system Homo sapiens R-HSA-1280215 | 6.65E-01 | 8.34E-01 | 9.29E-01 | 3.79E-01 | EGR1;DUSP2;MAP2K1;CISH;EGF;PELI2;HLA-B;PDGFA;PIK3R2;PIK3R1;IL17RD;GRIN2B;DUSP9;SOCS2;GHR;MT2A;SOCS1;FGF9;ERBB4;TRIM2;NEFL;TAB2;FGFR3;FGFR2 |
| 776 | Degradation of beta-catenin by the destruction complex Homo sapiens R-HSA-195253 | 6.66E-01 | 8.34E-01 | 8.78E-01 | 3.56E-01 | TLE3;TCF7L2;CTNNB1 |
| 777 | Transport to the Golgi and subsequent modification Homo sapiens R-HSA-948021 | 6.67E-01 | 8.34E-01 | 8.89E-01 | 3.59E-01 | RAB1B;CHST10;ST3GAL4;DYNLL1;DYNLL2;CTSC |
| 778 | Regulation of Hypoxia-inducible Factor (HIF) by oxygen Homo sapiens R-HSA-1234174 | 6.68E-01 | 8.34E-01 | 9.25E-01 | 3.74E-01 | CITED2 |
| 779 | Cellular response to hypoxia Homo sapiens R-HSA-2262749 | 6.68E-01 | 8.34E-01 | 9.25E-01 | 3.74E-01 | CITED2 |
| 780 | TAK1 activates NFkB by phosphorylation and activation of IKKs complex Homo sapiens R-HSA-445989 | 6.68E-01 | 8.34E-01 | 9.25E-01 | 3.74E-01 | TAB2 |
| 781 | TNFR1-induced NFkappaB signaling pathway Homo sapiens R-HSA-5357956 | 6.68E-01 | 8.34E-01 | 9.25E-01 | 3.74E-01 | TAB2 |
| 782 | WNT ligand biogenesis and trafficking Homo sapiens R-HSA-3238698 | 6.68E-01 | 8.34E-01 | 9.25E-01 | 3.74E-01 | WNT9B |
| 783 | CaM pathway Homo sapiens R-HSA-111997 | 6.68E-01 | 8.34E-01 | 9.25E-01 | 3.74E-01 | PRKAR1B |

| | | | | | | |
|---|---|---|---|---|---|---|
| 784 | Calmodulin induced events Homo sapiens R-HSA-111933 | 6.68E-01 | 8.34E-01 | 9.25E-01 | 3.74E-01 | PRKAR1B |
| 785 | IRE1alpha activates chaperones Homo sapiens R-HSA-381070 | 6.71E-01 | 8.36E-01 | 8.72E-01 | 3.48E-01 | SEC61G;CTDSP2 |
| 786 | GABA receptor activation Homo sapiens R-HSA-977443 | 6.71E-01 | 8.36E-01 | 8.72E-01 | 3.48E-01 | KCNJ10;KCNJ15 |
| 787 | VEGFR2 mediated vascular permeability Homo sapiens R-HSA-5218920 | 6.81E-01 | 8.41E-01 | 8.89E-01 | 3.41E-01 | CTNNB1 |
| 788 | Sema4D in semaphorin signaling Homo sapiens R-HSA-400685 | 6.81E-01 | 8.41E-01 | 8.89E-01 | 3.41E-01 | RHOB |
| 789 | Prefoldin mediated transfer of substrate to CCT/TriC Homo sapiens R-HSA-389957 | 6.81E-01 | 8.41E-01 | 8.89E-01 | 3.41E-01 | ACTB |
| 790 | Platelet calcium homeostasis Homo sapiens R-HSA-418360 | 6.81E-01 | 8.41E-01 | 8.89E-01 | 3.41E-01 | ATP2B4 |
| 791 | Phase 1 - inactivation of fast Na+ channels Homo sapiens R-HSA-5576894 | 6.81E-01 | 8.41E-01 | 8.89E-01 | 3.41E-01 | CACNA2D1 |
| 792 | Effects of PIP2 hydrolysis Homo sapiens R-HSA-114508 | 6.81E-01 | 8.41E-01 | 8.89E-01 | 3.41E-01 | MGLL |
| 793 | Peroxisomal lipid metabolism Homo sapiens R-HSA-390918 | 6.81E-01 | 8.41E-01 | 8.89E-01 | 3.41E-01 | EHHADH |
| 794 | G1/S Transition Homo sapiens R-HSA-69206 | 6.87E-01 | 8.48E-01 | 8.56E-01 | 3.21E-01 | WEE1;CDKN1B;CCND1;RPA1 |

| | | | | | | |
|---|---|---|---|---|---|---|
| 795 | RNA Polymerase I Chain Elongation Homo sapiens R-HSA-73777 | 6.90E-01 | 8.50E-01 | 8.41E-01 | 3.12E-01 | GTF2H2;GTF2H5 |
| 796 | Recognition of DNA damage by PCNA-containing replication complex Homo sapiens R-HSA-110314 | 6.95E-01 | 8.52E-01 | 8.56E-01 | 3.12E-01 | RPA1 |
| 797 | Extension of Telomeres Homo sapiens R-HSA-180786 | 6.95E-01 | 8.52E-01 | 8.56E-01 | 3.12E-01 | RPA1 |
| 798 | Ca-dependent events Homo sapiens R-HSA-111996 | 6.95E-01 | 8.52E-01 | 8.56E-01 | 3.12E-01 | PRKAR1B |
| 799 | Regulation of mRNA stability by proteins that bind AU-rich elements Homo sapiens R-HSA-450531 | 6.98E-01 | 8.55E-01 | 8.35E-01 | 3.01E-01 | HSPA8;PABPC1;ZFP36L1 |
| 800 | Organelle biogenesis and maintenance Homo sapiens R-HSA-1852241 | 7.04E-01 | 8.59E-01 | 8.82E-01 | 3.10E-01 | RXRA;TBL1XR1;CNTRL;CEP131;CYCS;CEP41;CYS1;DYNLL1;PRKAB1;PPARGC1A;DYNLL2;MRPL33 |
| 801 | Platelet homeostasis Homo sapiens R-HSA-418346 | 7.05E-01 | 8.59E-01 | 8.25E-01 | 2.89E-01 | KCNMA1;ATP2B4;PDE3A |
| 802 | Neurotransmitter Receptor Binding And Downstream Transmission In The Postsynaptic Cell Homo sapiens R-HSA-112314 | 7.06E-01 | 8.59E-01 | 8.43E-01 | 2.94E-01 | KCNJ10;EPB41L1;KCNJ15;NEFL;GRIN2B |
| 803 | Surfactant metabolism Homo sapiens R-HSA-5683826 | 7.07E-01 | 8.59E-01 | 8.26E-01 | 2.86E-01 | LMCD1 |
| 804 | Interaction between L1 and Ankyrins Homo sapiens R-HSA-445095 | 7.07E-01 | 8.59E-01 | 8.26E-01 | 2.86E-01 | SCN3B |
| 805 | PIWI-interacting RNA (piRNA) biogenesis Homo sapiens R-HSA-5601884 | 7.07E-01 | 8.59E-01 | 8.26E-01 | 2.86E-01 | POLR2A |

| | | | | | | |
|---|---|---|---|---|---|---|
| 806 | DNA Damage/Telomere Stress Induced Senescence Homo sapiens R-HSA-2559586 | 7.08E-01 | 8.59E-01 | 8.11E-01 | 2.80E-01 | CDKN1B;HMGA1 |
| 807 | SCF(Skp2)-mediated degradation of p27/p21 Homo sapiens R-HSA-187577 | 7.08E-01 | 8.59E-01 | 8.11E-01 | 2.80E-01 | CDKN1B;CCND1 |
| 808 | PI Metabolism Homo sapiens R-HSA-1483255 | 7.17E-01 | 8.66E-01 | 7.97E-01 | 2.65E-01 | PIK3R2;PIK3R1 |
| 809 | Regulation of TP53 Activity through Phosphorylation Homo sapiens R-HSA-6804756 | 7.19E-01 | 8.66E-01 | 8.06E-01 | 2.65E-01 | CSNK2B;RPA1;PRKAB1 |
| 810 | Amine compound SLC transporters Homo sapiens R-HSA-425428 | 7.19E-01 | 8.66E-01 | 7.97E-01 | 2.63E-01 | SLC22A2 |
| 811 | Termination of translesion DNA synthesis Homo sapiens R-HSA-5656169 | 7.19E-01 | 8.66E-01 | 7.97E-01 | 2.63E-01 | RPA1 |
| 812 | Activation of the pre-replicative complex Homo sapiens R-HSA-68962 | 7.19E-01 | 8.66E-01 | 7.97E-01 | 2.63E-01 | RPA1 |
| 813 | Glucagon-type ligand receptors Homo sapiens R-HSA-420092 | 7.19E-01 | 8.66E-01 | 7.97E-01 | 2.63E-01 | GCGR |
| 814 | Cell death signalling via NRAGE, NRIF and NADE Homo sapiens R-HSA-204998 | 7.25E-01 | 8.69E-01 | 7.83E-01 | 2.51E-01 | FGD3;OBSCN |
| 815 | TCR signaling Homo sapiens R-HSA-202403 | 7.26E-01 | 8.69E-01 | 8.11E-01 | 2.60E-01 | PIK3R2;TAB2;PIK3R1;PAG1 |

| | | | | | | |
|---|---|---|---|---|---|---|
| 816 | Immune System Homo sapiens R-HSA-168256 | 7.27E-01 | 8.69E-01 | 9.28E-01 | 2.96E-01 | CYFIP2;AHCYL1;CDKN1B;UBE3D;DEFB1;ACTB;ACTG1;GHR;LGALS3;MT2A;FGF9;TRIM2;SEC61G;KIF5A;NEFL;CTSF;CTSD;CTSC;MEF2A;DUSP2;MAP2K1;WSB1;CISH;HLA-B;FOS;DYNLL1;IL17RD;DUSP9;DYNLL2;PJA1;PRKAR1B;MAPKAPK3;TXNIP;CFD;PELI2;PDGFA;NEDD4L;PIK3R2;PIK3R1;FOXO3;FOXO1;SOCS2;SOCS1;ERBB4;PCBP2;PLCG2;PAG1;RAP1GAP2;EGR1;JUN;EGF;GRIN2B;TNRC6C;KIF26A;CTNNB1;TAB2;FBXL4;FGFR3;FGFR2;BCL2L1 |
| 817 | G2/M Transition Homo sapiens R-HSA-69275 | 7.27E-01 | 8.69E-01 | 8.30E-01 | 2.64E-01 | WEE1;CEP131;CNTRL;CEP41;DYNLL1;PHLDA1 |
| 818 | Amino acid transport across the plasma membrane Homo sapiens R-HSA-352230 | 7.31E-01 | 8.69E-01 | 7.71E-01 | 2.41E-01 | SLC3A1 |
| 819 | Trafficking of AMPA receptors Homo sapiens R-HSA-399719 | 7.31E-01 | 8.69E-01 | 7.71E-01 | 2.41E-01 | EPB41L1 |
| 820 | Glutamate Binding, Activation of AMPA Receptors and Synaptic Plasticity Homo sapiens R-HSA-399721 | 7.31E-01 | 8.69E-01 | 7.71E-01 | 2.41E-01 | EPB41L1 |
| 821 | Pausing and recovery of Tat-mediated HIV elongation Homo sapiens R-HSA-167238 | 7.31E-01 | 8.69E-01 | 7.71E-01 | 2.41E-01 | POLR2A |
| 822 | Tat-mediated HIV elongation arrest and recovery Homo sapiens R-HSA-167243 | 7.31E-01 | 8.69E-01 | 7.71E-01 | 2.41E-01 | POLR2A |
| 823 | Negative regulation of FGFR4 signaling Homo sapiens R-HSA-5654733 | 7.31E-01 | 8.69E-01 | 7.71E-01 | 2.41E-01 | FGF9 |
| 824 | NOD1/2 Signaling Pathway Homo sapiens R-HSA-168638 | 7.31E-01 | 8.69E-01 | 7.71E-01 | 2.41E-01 | TAB2 |

| | | | | | | |
|---|---|---|---|---|---|---|
| 825 | FCERI mediated Ca+2 mobilization Homo sapiens R-HSA-2871809 | 7.34E-01 | 8.69E-01 | 7.70E-01 | 2.38E-01 | AHCYL1;PLCG2 |
| 826 | Cytochrome P450 - arranged by substrate type Homo sapiens R-HSA-211897 | 7.34E-01 | 8.69E-01 | 7.70E-01 | 2.38E-01 | CYP27A1;RXRA |
| 827 | Mitotic G2-G2/M phases Homo sapiens R-HSA-453274 | 7.37E-01 | 8.69E-01 | 8.20E-01 | 2.50E-01 | WEE1;CEP131;CNTRL;CEP41;DYNLL1;PHLDA1 |
| 828 | p53-Dependent G1/S DNA damage checkpoint Homo sapiens R-HSA-69580 | 7.42E-01 | 8.69E-01 | 7.58E-01 | 2.26E-01 | CDKN1B;PCBP4 |
| 829 | p53-Dependent G1 DNA Damage Response Homo sapiens R-HSA-69563 | 7.42E-01 | 8.69E-01 | 7.58E-01 | 2.26E-01 | CDKN1B;PCBP4 |
| 830 | Asymmetric localization of PCP proteins Homo sapiens R-HSA-4608870 | 7.42E-01 | 8.69E-01 | 7.58E-01 | 2.26E-01 | FZD1;VANGL2 |
| 831 | HIV elongation arrest and recovery Homo sapiens R-HSA-167287 | 7.42E-01 | 8.69E-01 | 7.46E-01 | 2.22E-01 | POLR2A |
| 832 | Pausing and recovery of HIV elongation Homo sapiens R-HSA-167290 | 7.42E-01 | 8.69E-01 | 7.46E-01 | 2.22E-01 | POLR2A |
| 833 | Elongation arrest and recovery Homo sapiens R-HSA-112387 | 7.42E-01 | 8.69E-01 | 7.46E-01 | 2.22E-01 | POLR2A |
| 834 | DNA strand elongation Homo sapiens R-HSA-69190 | 7.42E-01 | 8.69E-01 | 7.46E-01 | 2.22E-01 | RPA1 |
| 835 | Negative regulation of FGFR1 signaling Homo sapiens R-HSA-5654726 | 7.42E-01 | 8.69E-01 | 7.46E-01 | 2.22E-01 | FGF9 |

| | | | | | | |
|---|---|---|---|---|---|---|
| 836 | Cooperation of Prefoldin and TriC/CCT in actin and tubulin folding Homo sapiens R-HSA-389958 | 7.42E-01 | 8.69E-01 | 7.46E-01 | 2.22E-01 | ACTB |
| 837 | Asparagine N-linked glycosylation Homo sapiens R-HSA-446203 | 7.51E-01 | 8.78E-01 | 8.31E-01 | 2.38E-01 | NUS1;PMM1;RAB1B;CHST10;ST3GAL4;ST3GAL5;DYNLL1;DYNLL2;CTSC |
| 838 | Cargo concentration in the ER Homo sapiens R-HSA-5694530 | 7.53E-01 | 8.79E-01 | 7.22E-01 | 2.05E-01 | CTSC |
| 839 | Smooth Muscle Contraction Homo sapiens R-HSA-445355 | 7.53E-01 | 8.79E-01 | 7.22E-01 | 2.05E-01 | SORBS3 |
| 840 | ER-Phagosome pathway Homo sapiens R-HSA-1236974 | 7.57E-01 | 8.82E-01 | 7.33E-01 | 2.04E-01 | SEC61G;HLA-B |
| 841 | G1/S DNA Damage Checkpoints Homo sapiens R-HSA-69615 | 7.57E-01 | 8.82E-01 | 7.33E-01 | 2.04E-01 | CDKN1B;PCBP4 |
| 842 | S Phase Homo sapiens R-HSA-69242 | 7.61E-01 | 8.83E-01 | 7.70E-01 | 2.10E-01 | WEE1;CDKN1B;CCND1;RPA1 |
| 843 | Negative regulators of RIG-I/MDA5 signaling Homo sapiens R-HSA-936440 | 7.63E-01 | 8.83E-01 | 7.00E-01 | 1.89E-01 | PCBP2 |
| 844 | EPHA-mediated growth cone collapse Homo sapiens R-HSA-3928663 | 7.63E-01 | 8.83E-01 | 7.00E-01 | 1.89E-01 | EFNA5 |
| 845 | Striated Muscle Contraction Homo sapiens R-HSA-390522 | 7.63E-01 | 8.83E-01 | 7.00E-01 | 1.89E-01 | TNNC1 |
| 846 | Purine metabolism Homo sapiens R-HSA-73847 | 7.63E-01 | 8.83E-01 | 7.00E-01 | 1.89E-01 | ADAL |
| 847 | Regulation of actin dynamics for phagocytic cup formation Homo sapiens R-HSA-2029482 | 7.72E-01 | 8.90E-01 | 7.37E-01 | 1.91E-01 | CYFIP2;ACTB;ACTG1 |

| | | | | | | |
|---|---|---|---|---|---|---|
| 848 | Base Excision Repair Homo sapiens R-HSA-73884 | 7.73E-01 | 8.90E-01 | 6.80E-01 | 1.75E-01 | RPA1 |
| 849 | Resolution of Abasic Sites (AP sites) Homo sapiens R-HSA-73933 | 7.73E-01 | 8.90E-01 | 6.80E-01 | 1.75E-01 | RPA1 |
| 850 | Fanconi Anemia Pathway Homo sapiens R-HSA-6783310 | 7.73E-01 | 8.90E-01 | 6.80E-01 | 1.75E-01 | RPA1 |
| 851 | COPII (Coat Protein 2) Mediated Vesicle Transport Homo sapiens R-HSA-204005 | 7.79E-01 | 8.96E-01 | 7.00E-01 | 1.75E-01 | RAB1B;CTSC |
| 852 | Late Phase of HIV Life Cycle Homo sapiens R-HSA-162599 | 7.82E-01 | 8.97E-01 | 7.45E-01 | 1.83E-01 | POLR2A;NEDD4L;GTF2H2;GTF2H5 |
| 853 | Transport of inorganic cations/anions and amino acids/oligopeptides Homo sapiens R-HSA-425393 | 7.84E-01 | 8.97E-01 | 7.22E-01 | 1.76E-01 | SLC20A1;SLC12A1;SLC3A1 |
| 854 | CLEC7A (Dectin-1) signaling Homo sapiens R-HSA-5607764 | 7.84E-01 | 8.97E-01 | 7.22E-01 | 1.76E-01 | AHCYL1;PLCG2;TAB2 |
| 855 | Downstream TCR signaling Homo sapiens R-HSA-202424 | 7.84E-01 | 8.97E-01 | 7.22E-01 | 1.76E-01 | PIK3R2;TAB2;PIK3R1 |
| 856 | HDR through Single Strand Annealing (SSA) Homo sapiens R-HSA-5685938 | 7.91E-01 | 9.03E-01 | 6.42E-01 | 1.50E-01 | RPA1 |
| 857 | Translesion synthesis by Y family DNA polymerases bypasses lesions on DNA template Homo sapiens R-HSA-110313 | 7.91E-01 | 9.03E-01 | 6.42E-01 | 1.50E-01 | RPA1 |
| 858 | Activation of ATR in response to replication stress Homo sapiens R-HSA-176187 | 7.91E-01 | 9.03E-01 | 6.42E-01 | 1.50E-01 | RPA1 |

| | | | | | | |
|---|---|---|---|---|---|---|
| 859 | Assembly of the primary cilium Homo sapiens R-HSA-5617833 | 7.92E-01 | 9.03E-01 | 7.65E-01 | 1.78E-01 | CEP131;CNTRL;CYS1;CEP41;DYNLL1;DYNLL2 |
| 860 | Cell surface interactions at the vascular wall Homo sapiens R-HSA-202733 | 7.95E-01 | 9.05E-01 | 7.07E-01 | 1.62E-01 | PIK3R2;PIK3R1;JAM3 |
| 861 | ER to Golgi Anterograde Transport Homo sapiens R-HSA-199977 | 7.97E-01 | 9.06E-01 | 7.27E-01 | 1.65E-01 | RAB1B;DYNLL1;DYNLL2;CTSC |
| 862 | Beta defensins Homo sapiens R-HSA-1461957 | 8.00E-01 | 9.08E-01 | 6.25E-01 | 1.39E-01 | DEFB1 |
| 863 | DNA Damage Recognition in GG-NER Homo sapiens R-HSA-5696394 | 8.00E-01 | 9.08E-01 | 6.25E-01 | 1.39E-01 | ACTB |
| 864 | Costimulation by the CD28 family Homo sapiens R-HSA-388841 | 8.05E-01 | 9.12E-01 | 6.60E-01 | 1.43E-01 | PIK3R2;PIK3R1 |
| 865 | G alpha (q) signalling events Homo sapiens R-HSA-416476 | 8.08E-01 | 9.12E-01 | 7.48E-01 | 1.59E-01 | PTGFR;GCGR;PIK3R2;PIK3R1;KNG1;MGLL |
| 866 | Formation of Fibrin Clot (Clotting Cascade) Homo sapiens R-HSA-140877 | 8.08E-01 | 9.12E-01 | 6.08E-01 | 1.29E-01 | KNG1 |
| 867 | Presynaptic phase of homologous DNA pairing and strand exchange Homo sapiens R-HSA-5693616 | 8.08E-01 | 9.12E-01 | 6.08E-01 | 1.29E-01 | RPA1 |
| 868 | tRNA modification in the nucleus and cytosol Homo sapiens R-HSA-6782315 | 8.08E-01 | 9.12E-01 | 6.08E-01 | 1.29E-01 | LAGE3 |
| 869 | NoRC negatively regulates rRNA expression Homo sapiens R-HSA-427413 | 8.11E-01 | 9.14E-01 | 6.51E-01 | 1.36E-01 | GTF2H2;GTF2H5 |

| | | | | | | |
|---|---|---|---|---|---|---|
| 870 | Class C/3 (Metabotropic glutamate/pheromone receptors) Homo sapiens R-HSA-420499 | 8.16E-01 | 9.19E-01 | 5.92E-01 | 1.20E-01 | GRM3 |
| 871 | Post-translational protein modification Homo sapiens R-HSA-597592 | 8.18E-01 | 9.19E-01 | 8.24E-01 | 1.66E-01 | NUS1;PMM1;PIGQ;RAB1B;RPA1;DYNLL1;DYNLL2;ADAMTS16;ADAMTS15;ADAMTS5;B3GNT9;ADAMTSL3;CHST10;ST3GAL4;ST3GAL5;GCNT4;MUC20;CTSC |
| 872 | Mitotic G1-G1/S phases Homo sapiens R-HSA-453279 | 8.20E-01 | 9.21E-01 | 6.99E-01 | 1.39E-01 | WEE1;CDKN1B;CCND1;RPA1 |
| 873 | RNA Polymerase III Abortive And Retractive Initiation Homo sapiens R-HSA-749476 | 8.24E-01 | 9.22E-01 | 5.78E-01 | 1.12E-01 | NFIC |
| 874 | RNA Polymerase III Transcription Homo sapiens R-HSA-74158 | 8.24E-01 | 9.22E-01 | 5.78E-01 | 1.12E-01 | NFIC |
| 875 | TNF signaling Homo sapiens R-HSA-75893 | 8.24E-01 | 9.22E-01 | 5.78E-01 | 1.12E-01 | TAB2 |
| 876 | HATs acetylate histones Homo sapiens R-HSA-3214847 | 8.26E-01 | 9.22E-01 | 6.66E-01 | 1.28E-01 | BRPF1;JADE1;ACTB |
| 877 | Negative epigenetic regulation of rRNA expression Homo sapiens R-HSA-5250941 | 8.29E-01 | 9.22E-01 | 6.24E-01 | 1.17E-01 | GTF2H2;GTF2H5 |
| 878 | RNA Polymerase I Promoter Clearance Homo sapiens R-HSA-73854 | 8.29E-01 | 9.22E-01 | 6.24E-01 | 1.17E-01 | GTF2H2;GTF2H5 |
| 879 | Viral Messenger RNA Synthesis Homo sapiens R-HSA-168325 | 8.31E-01 | 9.22E-01 | 5.64E-01 | 1.04E-01 | POLR2A |
| 880 | Homologous DNA Pairing and Strand Exchange Homo sapiens R-HSA-5693579 | 8.31E-01 | 9.22E-01 | 5.64E-01 | 1.04E-01 | RPA1 |
| 881 | Netrin-1 signaling Homo sapiens R-HSA-373752 | 8.31E-01 | 9.22E-01 | 5.64E-01 | 1.04E-01 | ABLIM1 |

| | | | | | | |
|---|---|---|---|---|---|---|
| 882 | Cooperation of PDCL (PhLP1) and TRiC/CCT in G-protein beta folding Homo sapiens R-HSA-6814122 | 8.31E-01 | 9.22E-01 | 5.64E-01 | 1.04E-01 | CSNK2B |
| 883 | Glucagon-like Peptide-1 (GLP1) regulates insulin secretion Homo sapiens R-HSA-381676 | 8.31E-01 | 9.22E-01 | 5.64E-01 | 1.04E-01 | PRKAR1B |
| 884 | RHO GTPase Effectors Homo sapiens R-HSA-195258 | 8.34E-01 | 9.24E-01 | 7.47E-01 | 1.35E-01 | CYFIP2;CDKN1B;KIF5A;FLNA;CTNNB1;ACTB;RHOB;ACTG1 |
| 885 | TP53 Regulates Transcription of Cell Death Genes Homo sapiens R-HSA-5633008 | 8.38E-01 | 9.26E-01 | 5.50E-01 | 9.70E-02 | NDRG1 |
| 886 | Vasopressin regulates renal water homeostasis via Aquaporins Homo sapiens R-HSA-432040 | 8.38E-01 | 9.26E-01 | 5.50E-01 | 9.70E-02 | PRKAR1B |
| 887 | G2/M DNA damage checkpoint Homo sapiens R-HSA-69473 | 8.40E-01 | 9.27E-01 | 6.08E-01 | 1.06E-01 | WEE1;RPA1 |
| 888 | RNA Polymerase I Transcription Homo sapiens R-HSA-73864 | 8.45E-01 | 9.30E-01 | 6.00E-01 | 1.01E-01 | GTF2H2;GTF2H5 |
| 889 | G alpha (s) signalling events Homo sapiens R-HSA-418555 | 8.45E-01 | 9.30E-01 | 6.69E-01 | 1.13E-01 | GPR27;GCGR;PDE3A;PTH1R |
| 890 | Intra-Golgi traffic Homo sapiens R-HSA-6811438 | 8.45E-01 | 9.30E-01 | 5.37E-01 | 9.04E-02 | GOLIM4 |
| 891 | Opioid Signalling Homo sapiens R-HSA-111885 | 8.55E-01 | 9.39E-01 | 5.84E-01 | 9.18E-02 | AHCYL1;PRKAR1B |
| 892 | RHO GTPases Activate Formins Homo sapiens R-HSA-5663220 | 8.57E-01 | 9.40E-01 | 6.24E-01 | 9.65E-02 | ACTB;ACTG1;RHOB |
| 893 | DNA Damage Bypass Homo sapiens R-HSA-73893 | 8.58E-01 | 9.40E-01 | 5.13E-01 | 7.88E-02 | RPA1 |

| | | | | | | |
|---|---|---|---|---|---|---|
| 894 | Antigen processing-Cross presentation Homo sapiens R-HSA-1236975 | 8.59E-01 | 9.41E-01 | 5.77E-01 | 8.75E-02 | SEC61G;HLA-B |
| 895 | Epigenetic regulation of gene expression Homo sapiens R-HSA-212165 | 8.61E-01 | 9.41E-01 | 6.18E-01 | 9.28E-02 | GTF2H2;ACTB;GTF2H5 |
| 896 | Defensins Homo sapiens R-HSA-1461973 | 8.64E-01 | 9.43E-01 | 5.02E-01 | 7.36E-02 | DEFB1 |
| 897 | p75 NTR receptor-mediated signalling Homo sapiens R-HSA-193704 | 8.64E-01 | 9.43E-01 | 5.70E-01 | 8.34E-02 | FGD3;OBSCN |
| 898 | RNA Polymerase I, RNA Polymerase III, and Mitochondrial Transcription Homo sapiens R-HSA-504046 | 8.65E-01 | 9.43E-01 | 6.13E-01 | 8.92E-02 | NFIC;GTF2H2;GTF2H5 |
| 899 | Death Receptor Signalling Homo sapiens R-HSA-73887 | 8.69E-01 | 9.46E-01 | 4.91E-01 | 6.89E-02 | TAB2 |
| 900 | EPH-ephrin mediated repulsion of cells Homo sapiens R-HSA-3928665 | 8.69E-01 | 9.46E-01 | 4.91E-01 | 6.89E-02 | EFNA5 |
| 901 | SLC-mediated transmembrane transport Homo sapiens R-HSA-425407 | 8.71E-01 | 9.46E-01 | 7.09E-01 | 9.81E-02 | SLC35B4;SLC47A1;SLC20A1;SLC22A2;SLCO2A1;ALB;SLC12A1;SLC3A1 |
| 902 | Meiosis Homo sapiens R-HSA-1500620 | 8.73E-01 | 9.47E-01 | 5.56E-01 | 7.58E-02 | RPA1;REC8 |
| 903 | Amino acid and oligopeptide SLC transporters Homo sapiens R-HSA-425374 | 8.75E-01 | 9.48E-01 | 4.81E-01 | 6.44E-02 | SLC3A1 |
| 904 | Regulation of TP53 Activity Homo sapiens R-HSA-5633007 | 8.77E-01 | 9.48E-01 | 6.27E-01 | 8.25E-02 | BRPF1;CSNK2B;RPA1;PRKAB1 |
| 905 | Unfolded Protein Response (UPR) Homo sapiens R-HSA-381119 | 8.77E-01 | 9.48E-01 | 5.50E-01 | 7.23E-02 | SEC61G;CTDSP2 |

| | | | | | | |
|---|---|---|---|---|---|---|
| 906 | Deadenylation-dependent mRNA decay Homo sapiens R-HSA-429914 | 8.80E-01 | 9.49E-01 | 4.71E-01 | 6.03E-02 | PABPC1 |
| 907 | Ubiquitin-dependent degradation of Cyclin D1 Homo sapiens R-HSA-69229 | 8.80E-01 | 9.49E-01 | 4.71E-01 | 6.03E-02 | CCND1 |
| 908 | Ubiquitin-dependent degradation of Cyclin D Homo sapiens R-HSA-75815 | 8.80E-01 | 9.49E-01 | 4.71E-01 | 6.03E-02 | CCND1 |
| 909 | Cargo trafficking to the periciliary membrane Homo sapiens R-HSA-5620920 | 8.85E-01 | 9.52E-01 | 4.62E-01 | 5.65E-02 | CYS1 |
| 910 | Aquaporin-mediated transport Homo sapiens R-HSA-445717 | 8.85E-01 | 9.52E-01 | 4.62E-01 | 5.65E-02 | PRKAR1B |
| 911 | C-type lectin receptors (CLRs) Homo sapiens R-HSA-5621481 | 8.89E-01 | 9.56E-01 | 5.77E-01 | 6.77E-02 | AHCYL1;PLCG2;TAB2 |
| 912 | PCP/CE pathway Homo sapiens R-HSA-4086400 | 8.96E-01 | 9.62E-01 | 5.19E-01 | 5.71E-02 | FZD1;VANGL2 |
| 913 | Meiotic recombination Homo sapiens R-HSA-912446 | 8.99E-01 | 9.64E-01 | 4.36E-01 | 4.66E-02 | RPA1 |
| 914 | HIV Infection Homo sapiens R-HSA-162906 | 9.02E-01 | 9.66E-01 | 6.40E-01 | 6.59E-02 | POLR2A;HMGA1;NEDD4L;BANF1;GTF2H2;GTF2H5 |
| 915 | ECM proteoglycans Homo sapiens R-HSA-3000178 | 9.03E-01 | 9.66E-01 | 4.28E-01 | 4.37E-02 | COL9A2 |
| 916 | Meiotic synapsis Homo sapiens R-HSA-1221632 | 9.11E-01 | 9.70E-01 | 4.12E-01 | 3.85E-02 | REC8 |
| 917 | Interferon Signaling Homo sapiens R-HSA-913531 | 9.13E-01 | 9.70E-01 | 6.03E-01 | 5.49E-02 | EGR1;MT2A;SOCS1;TRIM2;HLA-B |
| 918 | Cell Cycle, Mitotic Homo sapiens R-HSA-69278 | 9.14E-01 | 9.70E-01 | 7.18E-01 | 6.43E-02 | CDKN1B;RAB1B;CEP131;RPA1;DYNLL1;WEE1;CCND1;CSNK2B;CNTRL;BANF1;CEP41;LPIN1;EMD;PHLDA1 |

| | | | | | | |
|---|---|---|---|---|---|---|
| 919 | rRNA modification in the nucleus Homo sapiens R-HSA-6790901 | 9.14E-01 | 9.70E-01 | 4.05E-01 | 3.62E-02 | RPS14 |
| 920 | B-WICH complex positively regulates rRNA expression Homo sapiens R-HSA-5250924 | 9.14E-01 | 9.70E-01 | 4.05E-01 | 3.62E-02 | ACTB |
| 921 | Recruitment and ATM-mediated phosphorylation of repair and signaling proteins at DNA double strand breaks Homo sapiens R-HSA-5693565 | 9.14E-01 | 9.70E-01 | 4.05E-01 | 3.62E-02 | EYA3 |
| 922 | Initial triggering of complement Homo sapiens R-HSA-166663 | 9.14E-01 | 9.70E-01 | 4.05E-01 | 3.62E-02 | CFD |
| 923 | Synthesis of DNA Homo sapiens R-HSA-69239 | 9.15E-01 | 9.70E-01 | 4.86E-01 | 4.32E-02 | CDKN1B;RPA1 |
| 924 | Telomere Maintenance Homo sapiens R-HSA-157579 | 9.18E-01 | 9.72E-01 | 3.98E-01 | 3.40E-02 | RPA1 |
| 925 | DNA Double Strand Break Response Homo sapiens R-HSA-5693606 | 9.18E-01 | 9.72E-01 | 3.98E-01 | 3.40E-02 | EYA3 |
| 926 | RHO GTPases activate PKNs Homo sapiens R-HSA-5625740 | 9.21E-01 | 9.73E-01 | 3.91E-01 | 3.20E-02 | RHOB |
| 927 | HDACs deacetylate histones Homo sapiens R-HSA-3214815 | 9.21E-01 | 9.73E-01 | 3.91E-01 | 3.20E-02 | TBL1XR1 |
| 928 | Transport of glucose and other sugars, bile salts and organic acids, metal ions and amine compounds Homo sapiens R-HSA-425366 | 9.26E-01 | 9.76E-01 | 4.66E-01 | 3.59E-02 | SLC47A1;SLC22A2 |

| | | | | | | |
|---|---|---|---|---|---|---|
| 929 | Gene Silencing by RNA Homo sapiens R-HSA-211000 | 9.26E-01 | 9.76E-01 | 4.66E-01 | 3.59E-02 | TNRC6C;POLR2A |
| 930 | HDR through Homologous Recombination (HRR) Homo sapiens R-HSA-5685942 | 9.34E-01 | 9.83E-01 | 3.66E-01 | 2.51E-02 | RPA1 |
| 931 | DNA Replication Homo sapiens R-HSA-69306 | 9.35E-01 | 9.84E-01 | 4.48E-01 | 2.99E-02 | CDKN1B;RPA1 |
| 932 | Cytosolic sensors of pathogen-associated DNA Homo sapiens R-HSA-1834949 | 9.39E-01 | 9.86E-01 | 3.55E-01 | 2.23E-02 | CTNNB1 |
| 933 | Class I MHC mediated antigen processing & presentation Homo sapiens R-HSA-983169 | 9.40E-01 | 9.87E-01 | 6.19E-01 | 3.81E-02 | WSB1;SOCS1;UBE3D;SEC61G;HLA-B;NEDD4L;FBXL4;PJA1 |
| 934 | Semaphorin interactions Homo sapiens R-HSA-373755 | 9.42E-01 | 9.87E-01 | 3.50E-01 | 2.10E-02 | RHOB |
| 935 | Cell Cycle Checkpoints Homo sapiens R-HSA-69620 | 9.47E-01 | 9.91E-01 | 5.17E-01 | 2.82E-02 | WEE1;CDKN1B;PCBP4;RPA1 |
| 936 | Switching of origins to a post-replicative state Homo sapiens R-HSA-69052 | 9.49E-01 | 9.91E-01 | 3.34E-01 | 1.76E-02 | CDKN1B |
| 937 | Orc1 removal from chromatin Homo sapiens R-HSA-68949 | 9.49E-01 | 9.91E-01 | 3.34E-01 | 1.76E-02 | CDKN1B |
| 938 | Transcriptional regulation by small RNAs Homo sapiens R-HSA-5578749 | 9.53E-01 | 9.93E-01 | 3.25E-01 | 1.57E-02 | POLR2A |
| 939 | Removal of licensing factors from origins Homo sapiens R-HSA-69300 | 9.53E-01 | 9.93E-01 | 3.25E-01 | 1.57E-02 | CDKN1B |
| 940 | DNA Repair Homo sapiens R-HSA-73894 | 9.54E-01 | 9.93E-01 | 5.79E-01 | 2.75E-02 | POLR2A;EYA3;XPA;RPA1;GTF2H2;ACTB;GTF2H5 |
| 941 | Positive epigenetic regulation of rRNA expression Homo sapiens R-HSA-5250913 | 9.55E-01 | 9.93E-01 | 3.20E-01 | 1.48E-02 | ACTB |

| | | | | | | |
|---|---|---|---|---|---|---|
| 942 | SUMOylation of DNA damage response and repair proteins Homo sapiens R-HSA-3108214 | 9.58E-01 | 9.95E-01 | 3.12E-01 | 1.32E-02 | RPA1 |
| 943 | Regulation of DNA replication Homo sapiens R-HSA-69304 | 9.58E-01 | 9.95E-01 | 3.12E-01 | 1.32E-02 | CDKN1B |
| 944 | Antigen processing: Ubiquitination & Proteasome degradation Homo sapiens R-HSA-983168 | 9.61E-01 | 9.96E-01 | 5.43E-01 | 2.15E-02 | WSB1;SOCS1;UBE3D;NEDD4L;FBXL4;PJA1 |
| 945 | RNA polymerase II transcribes snRNA genes Homo sapiens R-HSA-6807505 | 9.62E-01 | 9.96E-01 | 3.03E-01 | 1.18E-02 | POLR2A |
| 946 | Cell Cycle Homo sapiens R-HSA-1640170 | 9.62E-01 | 9.96E-01 | 6.66E-01 | 2.58E-02 | CDKN1B;RAB1B;CEP131;RPA1;DYNLL1;REC8;WEE1;CCND1;PCBP4;CSNK2B;CNTRL;BANF1;CEP41;LPIN1;EMD;PHLDA1 |
| 947 | RIG-I/MDA5 mediated induction of IFN-alpha/beta pathways Homo sapiens R-HSA-168928 | 9.65E-01 | 9.96E-01 | 2.96E-01 | 1.05E-02 | PCBP2 |
| 948 | Complement cascade Homo sapiens R-HSA-166658 | 9.66E-01 | 9.96E-01 | 2.92E-01 | 9.97E-03 | CFD |
| 949 | Processing of DNA double-strand break ends Homo sapiens R-HSA-5693607 | 9.68E-01 | 9.96E-01 | 2.88E-01 | 9.43E-03 | RPA1 |
| 950 | DNA Replication Pre-Initiation Homo sapiens R-HSA-69002 | 9.69E-01 | 9.96E-01 | 2.85E-01 | 8.92E-03 | RPA1 |
| 951 | M/G1 Transition Homo sapiens R-HSA-68874 | 9.69E-01 | 9.96E-01 | 2.85E-01 | 8.92E-03 | RPA1 |
| 952 | Mitochondrial translation initiation Homo sapiens R-HSA-5368286 | 9.72E-01 | 9.96E-01 | 2.78E-01 | 7.98E-03 | MRPL33 |
| 953 | Mitochondrial translation termination Homo sapiens R-HSA-5419276 | 9.72E-01 | 9.96E-01 | 2.78E-01 | 7.98E-03 | MRPL33 |

| | | | | | | |
|---|---|---|---|---|---|---|
| 954 | Mitochondrial translation elongation Homo sapiens R-HSA-5389840 | 9.72E-01 | 9.96E-01 | 2.78E-01 | 7.98E-03 | MRPL33 |
| 955 | Host Interactions of HIV factors Homo sapiens R-HSA-162909 | 9.72E-01 | 9.96E-01 | 3.63E-01 | 1.02E-02 | HMGA1;BANF1 |
| 956 | Hedgehog 'on' state Homo sapiens R-HSA-5632684 | 9.73E-01 | 9.96E-01 | 2.74E-01 | 7.56E-03 | GAS1 |
| 957 | Chromosome Maintenance Homo sapiens R-HSA-73886 | 9.74E-01 | 9.96E-01 | 2.71E-01 | 7.15E-03 | RPA1 |
| 958 | Mitochondrial translation Homo sapiens R-HSA-5368287 | 9.78E-01 | 9.99E-01 | 2.59E-01 | 5.75E-03 | MRPL33 |
| 959 | Signaling by Hedgehog Homo sapiens R-HSA-5358351 | 9.79E-01 | 9.99E-01 | 3.44E-01 | 7.45E-03 | PRKAR1B;GAS1 |
| 960 | SUMO E3 ligases SUMOylate target proteins Homo sapiens R-HSA-3108232 | 9.83E-01 | 1.00E+00 | 2.43E-01 | 4.16E-03 | RPA1 |
| 961 | DNA Double-Strand Break Repair Homo sapiens R-HSA-5693532 | 9.85E-01 | 1.00E+00 | 3.22E-01 | 5.01E-03 | EYA3;RPA1 |
| 962 | Hedgehog 'off' state Homo sapiens R-HSA-5610787 | 9.85E-01 | 1.00E+00 | 2.35E-01 | 3.55E-03 | PRKAR1B |
| 963 | SUMOylation Homo sapiens R-HSA-2990846 | 9.86E-01 | 1.00E+00 | 2.30E-01 | 3.19E-03 | RPA1 |
| 964 | G2/M Checkpoints Homo sapiens R-HSA-69481 | 9.87E-01 | 1.00E+00 | 3.11E-01 | 4.02E-03 | WEE1;RPA1 |
| 965 | tRNA processing Homo sapiens R-HSA-72306 | 9.87E-01 | 1.00E+00 | 2.26E-01 | 2.87E-03 | LAGE3 |
| 966 | M Phase Homo sapiens R-HSA-68886 | 9.88E-01 | 1.00E+00 | 4.36E-01 | 5.37E-03 | RAB1B;CSNK2B;BANF1;LPIN1;EMD |
| 967 | Mitotic Prometaphase Homo sapiens R-HSA-68877 | 9.89E-01 | 1.00E+00 | 2.17E-01 | 2.33E-03 | CSNK2B |

| | | | | | | |
|---|---|---|---|---|---|---|
| 968 | FCERI mediated NF-kB activation Homo sapiens R-HSA-2871837 | 9.91E-01 | 1.00E+00 | 2.09E-01 | 1.89E-03 | TAB2 |
| 969 | HDR through Homologous Recombination (HR) or Single Strand Annealing (SSA) Homo sapiens R-HSA-5693567 | 9.91E-01 | 1.00E+00 | 2.07E-01 | 1.79E-03 | RPA1 |
| 970 | Homology Directed Repair Homo sapiens R-HSA-5693538 | 9.93E-01 | 1.00E+00 | 1.97E-01 | 1.32E-03 | RPA1 |
| 971 | Mitotic Anaphase Homo sapiens R-HSA-68882 | 9.95E-01 | 1.00E+00 | 2.69E-01 | 1.47E-03 | BANF1;EMD |
| 972 | Mitotic Metaphase and Anaphase Homo sapiens R-HSA-2555396 | 9.95E-01 | 1.00E+00 | 2.67E-01 | 1.41E-03 | BANF1;EMD |
| 973 | G alpha (i) signalling events Homo sapiens R-HSA-418594 | 9.98E-01 | 1.00E+00 | 2.90E-01 | 7.02E-04 | GRM3;PTGER3;KNG1 |
| 974 | GPCR ligand binding Homo sapiens R-HSA-500792 | 9.98E-01 | 1.00E+00 | 4.16E-01 | 6.70E-04 | FZD1;GRM3;PTGFR;GCGR;PTGER3;WNT9B;PTH1R;KNG1 |
| 975 | Immunoregulatory interactions between a Lymphoid and a non-Lymphoid cell Homo sapiens R-HSA-198933 | 9.99E-01 | 1.00E+00 | 1.47E-01 | 1.87E-04 | HLA-B |
| 976 | Peptide ligand-binding receptors Homo sapiens R-HSA-375276 | 1.00E+00 | 1.00E+00 | 1.19E-01 | 3.28E-05 | KNG1 |
| 977 | Class A/1 (Rhodopsin-like receptors) Homo sapiens R-HSA-373076 | 1.00E+00 | 1.00E+00 | 2.14E-01 | 2.62E-05 | PTGFR;PTGER3;KNG1 |
| 978 | Signaling by GPCR Homo sapiens R-HSA-372790 | 1.00E+00 | 1.00E+00 | 5.32E-01 | 5.06E-05 | GPR27;PTGFR;AHCYL1;PTGER3;PDGFA;PIK3R2;PTH1R;PIK3R1;KNG1;GRM3;FGD3;FGF9;ERBB4;NEFL;FZD1;DUSP2;MAP2K1;EGF;GCGR;WNT9B;IL17RD;GRIN2B;DUSP9;RHOB;OBSCN;PRKAR1B;PDE3A;FGFR3;MGLL;FGFR2 |

| | | | | | | |
|---|---|---|---|---|---|---|
| 979 | GPCR downstream signaling Homo sapiens R-HSA-388396 | 1.00E+00 | 1.00E+00 | 3.23E-01 | 1.73E-06 | GPR27;PTGFR;GCGR;PTGER3;PIK3R2;PTH1R;PIK3R1;KNG1;RHOB;GRM3;FGD3;OBSCN;PDE3A;MGLL |

**Pathways enriched by genes suppressed with SGLT2 inhibitor in intercalated cells**

| | Term | P value | Adjusted. P value | Odds Ratio | Combined Score | Genes |
|---|---|---|---|---|---|---|
| 1 | Membrane Trafficking Homo sapiens R-HSA-199991 | 7.94E-21 | 1.13E-17 | 2.640887 | 122.22753 | APP;SYS1;TFRC;KDELR1;GOLIM4;GCC2;GOLGA2;GOLGA4;GOLGA5;TFG;AKT2;KIF5B;AKT1;ARFIP2;KIF21A;KIFAP3;VPS36;AP2M1;NSF;COG6;RALGAPA2;COG2;COG1;ANK2;OCRL;ANK3;RINT1;DYNC1LI2;RALGAPB;TBC1D20;KIF16B;CHMP4C;CHMP4B;KDELR2;STAM2;VPS28;FTL;COPB2;SPTBN5;COPA;RAB5C;TMED10;TSG101;STXBP3;SAR1B;COPB1;ARL1;GNS;DYNC2LI1;TGOLN2;SCFD1;KLC2;SNX2;MAN2A2;UBB;KIF3A;MAN2A1;MYO6;UBC;SNX9;AP1M2;SNX5;SPTBN1;SPTBN2;HSPA8;RAB4A;TRAPPC1;TRAPPC4;SURF4;M6PR;SNF8;PUM1;IGF2R;DAB2;GJB1;TMEM115;GOLGB1;CHMP2B;CALM1;COPE;YWHAE;ARF4;MCFD2;C2CD5;SERPINA1;YWHAB;CLTC;CLTB;AP4E1;PRKAG2;ARRB1;KIF12;MIA3;CLINT1;AP1G2;AP1G1;PPP6R3;TMED3;KIF13B;TMED2;KIF1B;MAN1A1;RAC1;TMED7;SPTAN1;RAB8A;YWHAH;TMED9;EXOC7;KIF25;COPZ2;VPS37D;AP3B1;KIF23;VPS37B;ARFGAP3;COPZ1;DNM2;VAMP8;KIF2A;EXOC4;CHMP3;EXOC6;NAA38;DNASE2;EXOC5;PAFAH1B3;EXOC2;ARF5;RHOQ;VAMP3;PAFAH1B1;CHMP5;DYNC1I2;NBAS;RALA;TMF1;PRKAA2;STX17;DCTN2;SRC;STX16;VPS4B;USO1;VPS4A;STX18;NAPG;AGPAT3;STX10;LMAN1;CYTH2;CUX1;LMAN2;VPS53;STX6;MAN1C1;STX5;STX4;CD59;CD55;DYNC1H1;SORT1;TRAPPC10;GALNT1;HIP1R;DTNBP1;TPD52L1;ARCN1;RAB10;TJP1;RAB14;RAB13;YIPF6;TRIP11; [illegible] |
| 2 | Respiratory electron transport Homo sapiens R-HSA-611105 | 2.80E-18 | 1.99E-15 | 6.778261 | 273.96708 | NDUFA11;NDUFA12;NDUFA10;ETFB;ECSIT;COX6A1;COX7C;UQCRFS1;COX8A;ACAD9;NDUFC2;NDUFC1;SDHD;SDHA;SDHB;COX6B1;NDUFS8;NDUFS7;SCO2;NDUFS5;NDUFS4;NDUFS3;UQCRC1;NDUFS2;NDUFS1;UQCRC2;NDUFB9;COX19;NDUFB8;NDUFB7;COX16;NDUFB11;NDUFB4;ETFDH;NDUFB2;UQCR11;UQCR10;COX5A;UQCRH;COX14;NDUFV3;CYC1;NDUFV2;NDUFV1;NDUFA9;NDUFA8;SURF1;NDUFA6;NDUFA3;NDUFA1;COX6C;LRPPRC;NDUFAF6;UQCRQ;NDUFAF7;NDUFAF4;NDUFAB1;COX20 |

| | | | | | | |
|---|---|---|---|---|---|---|
| 3 | The citric acid (TCA) cycle and respiratory electron transport Homo sapiens R-HSA-1428517 | 5.73E-16 | 2.72E-13 | 3.850695 | 135.14197 | NDUFA11;NDUFA12;NDUFA10;ETFB;COX6A1;ECSIT;COX7C;NNT;IDH3B;UQCRFS1;PDK2;COX8A;PDHX;ACAD9;NDUFC2;NDUFC1;SDHD;SDHA;SDHB;COX6B1;NDUFS8;NDUFS7;SCO2;NDUFS5;NDUFS4;UQCRC1;NDUFS3;SUCLG2;NDUFS2;SUCLG1;NDUFS1;UQCRC2;HAGH;DLD;COX19;NDUFB9;NDUFB8;FH;NDUFB7;COX16;NDUFB11;GLO1;NDUFB4;ETFDH;NDUFB2;DLST;UQCR11;UQCR10;PDHB;COX5A;UQCRH;LDHB;LDHA;UCP2;COX14;NDUFV3;CYC1;NDUFV2;NDUFV1;NDUFA9;NDUFA8;SURF1;PDHA1;NDUFA6;MDH2;IDH3G;NDUFA3;NDUFA1;COX6C;LRPPRC;PDP1;SUCLA2;NDUFAF6;UQCRQ;NDUFAF7;NDUFAF4;NDUFAB1;OGDH;ACO2;COX20 |
| 4 | Hh mutants that don't undergo autocatalytic processing are degraded by ERAD Homo sapiens R-HSA-5362768 | 2.65E-15 | 6.53E-13 | 9.320412 | 312.82221 | PSMD12;VCP;PSMD11;PSMD13;SEL1L;DERL2;PSMB10;PSMA7;PSMD8;PSMB6;ERLEC1;SHH;PSMD6;PSMB4;PSMD7;PSMB5;PSMD4;OS9;UBB;PSMB2;PSMB3;PSMD2;PSMD3;UBC;PSMB1;PSMD1;PSMF1;SYVN1;PSMB8;PSMB9;PSMA5;PSMA6;PSMA3;PSMC5;PSMA1;PSMA2;PSMC4;PSMC2;PSME1;PSME2 |
| 5 | Infectious disease Homo sapiens R-HSA-5663205 | 2.66E-15 | 6.53E-13 | 2.465445 | 82.74367 | RPL5;RPL30;CCNT2;RPL31;GTF2B;RPL6;IPO5;PSMD8;RPS15;PSMD6;PSMD7;PSMD4;RPS19;PSMD2;PSMD3;RPL36;PSMD1;KPNA4;RPL37;KPNA2;B2M;RPS10;KPNA3;AP2M1;KPNA1;NUP214;SUPT16H;RPS9;MAP2K1;MAP2K2;RPS7;RPS8;RPS5;GTF2F2;PSME1;CHMP4C;PSME2;CHMP4B;NUP98;RPL29;PPIA;VPS28;NUP205;PSMD12;TSG101;PSMD11;CUL5;PSMD13;NEDD4L;RPS4Y1;AAAS;POM121C;UBB;TPR;UBC;PSMF1;AP1M2;RPL41;XRCC6;TGFB1;XRCC5;EIF2AK2;ISG15;EEF2;CDK9;MSRA;RPS25;RPS28;CDK7;RPS27;RPS29;CHMP2B;NMT1;CALR;CALM1;GRSF1;RPS21;RPS24;HSP90AB1;RPLP1;PSIP1;AP2A1;RPL7A;XPO1;AP1G1;RPLP2;NELFB;RAC1;HSP90AA1;PDCD6IP;VPS37D;VPS37B;PSMA5;DNAJC3;PSMA6;PSMA3;PSMA1;PSMA2;NUP50;RPL37A;VAMP1;CHMP3;SLC25A4;CHMP5;GTF2A1;GTF2A2;RNMT;VPS4B;VPS4A;FURIN;PSMA7;PSMB10;PSMB6;PSMB4;PSMB5;PSMB2;PSMB3;POLR2B;POLR2C;PSMB1;POLR2E;POLR2H;PAK2;POLR2J;RPL19;RANBP2;NPM1;TXNRD1;TAF11;RPL35A;SSRP1;PSMB8;PSMB9;PSMC5;PSMC4;PSMC2;TCEA1;CD9;TAF5;TAF3;RAN;TAF2;HSPA1A |

| | | | | | | |
|---|---|---|---|---|---|---|
| 6 | Metabolism Homo sapiens R-HSA-1430728 | 2.75E-15 | 6.53E-13 | 1.53361 | 51.415221 | PNMT;RPL5;RPL30;RPL31;SLC4A1;ENO1;EPRS;RPL6;GLS;PSMD8;PSMD6;PSMD7;LIPC;SCP2;PSMD4;PSMD2;LIPH;PSMD3;LIPG;RPL36;AKT1;PSMD1;PIP4K2C;RPL37;NUP214;GSTK1;ACOT9;SECISBP2;MED8;HSPG2;MTAP;ADCY9;SCO2;FDX1;UQCRC1;PSME1;ACOT2;PLBD1;PSME2;NUP98;UQCRC2;RPL29;ACOT4;IDO1;MTMR1;NUP205;SAR1B;ABCB7;SHMT1;IQGAP1;RPS4Y1;AAAS;GNS;SLC5A3;OAZ1;ADH5;MTMR6;LDHB;CYB5R1;UGT8;LDHA;CYB5R4;POM121C;HLCS;RDH11;CHST12;PRKAR2A;PTPMT1;AFMID;CHST15;ST3GAL6;LTA4H;PSMF1;PLCG1;FDFT1;CDK19;RPL41;CHKB;CHKA;MTHFS;NR1D1;GLUD1;QARS;ADI1;CYP1A2;CYP1A1;OGDH;LGMN;RPLP1;HEXB;HEXA;QPRT;ADK;MED19;PRKAG2;ECSIT;MCEE;MED13;MED10;SMPD1;RARS;RPLP2;TK2;TK1;TEAD1;TEAD2;ELOVL1;ARG2;TPI1;GSTO2;GSTO1;PGAM1;ITPK1;PTGR1;SUMF2;SUMF1;HADHB;GCHFR;PSMA5;ITPKB;HADHA;PSMA6;PSMA3;NCOR1;BDH2;PSMA1;PSMA2;MED21;IVD;PPARG;PLIN2;TKT;DLD;RAPGEF3;MGLL;FH;ASAH1;SGMS1;GSTP1;DLST;ADCY6;PAPSS2;PSMA7;PTGS1;PSMB6;PSMB4;PSMB5;PSMB2;PSMB3;PSMB1;UCP2;CEPT1;RPL19;AOC1;MDH1;MDH2;AGK;AGL;GSR;PSMB8;LRPPRC;PSMB9;PSMC5;LYRM4;NMNAT1;PSMC4;ETNK1;LPCAT3;PSMC2;CIAPIN1;AGRN;CRAT;VKORC1L1;RORA;SAT1;NAGLU;OSBP;NUDT15;RPS9;PCYT2;DGAT1;[illegible] |
| 7 | Apoptosis Homo sapiens R-HSA-109581 | 1.61E-14 | 3.02E-12 | 3.469676 | 110.20591 | YWHAE;PPP1R13B;YWHAB;HMGB1;PSMD8;PSMD6;CASP7;PSMD7;PSMD4;FNTA;CDH1;AKT2;PSMD2;PSMD3;AKT1;PSMD1;HIST1H1E;DNM1L;SPTAN1;KPNA1;YWHAH;HIST1H1C;APPL1;DSP;DBNL;DAPK1;PRKCD;DAPK3;TICAM1;PSMA5;PSMA6;PSMA3;PSMA1;PSMA2;PSME1;PSME2;BIRC2;PSMD12;PSMD11;ROCK1;PSMD13;XIAP;PSMB10;PSMA7;ADD1;PSMB6;PSMB4;PSMB5;UBB;PSMB2;PSMB3;UBC;LMNA;PSMB1;TP53BP2;RIPK1;PSMF1;BAK1;BID;PAK2;BCAP31;GSN;BAD;CFLAR;PTK2;PSMB8;PSMB9;TJP1;OCLN;H1F0;PSMC5;STK24;APC;PSMC4;PSMC2;NMT1;FAS;BAX;ACIN1;CTNNB1;TJP2 |

| | | | | | | |
|---|---|---|---|---|---|---|
| 8 | Vesicle-mediated transport Homo sapiens R-HSA-5653656 | 1.74E-14 | 3.02E-12 | 2.117279 | 67.075649 | APP;SYS1;TFRC;KDELR1;GOLIM4;GCC2;GOLGA2;GOLGA4;GOLGA5;TFG;AKT2;KIF5B;AKT1;ARFIP2;KIF21A;KIFAP3;VPS36;AP2M1;NSF;COG6;RALGAPA2;COG2;COG1;ANK2;OCRL;ANK3;RINT1;DYNC1LI2;RALGAPB;TBC1D20;KIF16B;CHMP4C;CHMP4B;KDELR2;STAM2;VPS28;FTL;COPB2;SPTBN5;COPA;RAB5C;TMED10;TSG101;STXBP3;SAR1B;COPB1;ARL1;GNS;HSP90B1;DYNC2LI1;TGOLN2;SCFD1;KLC2;SNX2;MAN2A2;UBB;KIF3A;MAN2A1;MYO6;UBC;SNX9;AP1M2;SNX5;SPTBN1;SPTBN2;HSPA8;RAB4A;TRAPPC1;TRAPPC4;SURF4;M6PR;SNF8;PUM1;IGF2R;DAB2;GJB1;TMEM115;GOLGB1;CHMP2B;CALR;CALM1;COPE;YWHAE;ARF4;MCFD2;C2CD5;SERPINA1;YWHAB;CLTC;CLTB;AP4E1;PRKAG2;ARRB1;KIF12;MIA3;CLINT1;AP1G2;AP1G1;PPP6R3;TMED3;KIF13B;TMED2;KIF1B;MAN1A1;CD36;RAC1;TMED7;SPTAN1;RAB8A;YWHAH;HSP90AA1;TMED9;EXOC7;KIF25;COPZ2;VPS37D;AP3B1;KIF23;VPS37B;ARFGAP3;COPZ1;DNM2;VAMP8;KIF2A;EXOC4;CHMP3;EXOC6;NAA38;DNASE2;EXOC5;PAFAH1B3;EXOC2;ARF5;RHOQ;VAMP3;PAFAH1B1;CHMP5;DYNC1I2;NBAS;RALA;TMF1;PRKAA2;STX17;DCTN2;SRC;STX16;VPS4B;USO1;VPS4A;STX18;NAPG;AGPAT3;STX10;LMAN1;CYTH2;CUX1;LMAN2;VPS53;STX6;MAN1C1;STX5;STX4;CD59;APOL1;CD55;DYNC1H1;SORT1;TRAPPC10;GALNT1;HIP1R;DTNBP1;TPD52L1;ARCN1;[illegible] |
| 9 | Hh mutants abrogate ligand secretion Homo sapiens R-HSA-5387390 | 1.91E-14 | 3.02E-12 | 8.222832 | 259.74488 | PSMD12;VCP;PSMD11;PSMD13;SEL1L;DERL2;PSMB10;PSMA7;PSMD8;PSMB6;ERLEC1;SHH;PSMD6;PSMB4;PSMD7;PSMB5;PSMD4;OS9;UBB;PSMB2;PSMB3;PSMD2;PSMD3;UBC;PSMB1;PSMD1;PSMF1;SYVN1;PSMB8;PSMB9;PSMA5;PSMA6;PSMA3;PSMC5;PSMA1;PSMA2;PSMC4;PSMC2;PSME1;PSME2 |

| | | | | | | |
|---|---|---|---|---|---|---|
| 10 | Metabolism of proteins Homo sapiens R-HSA-392499 | 2.60E-14 | 3.70E-12 | 1.694404 | 53.002142 | EIF4A1;RPL5;APP;RPL30;PGAP1;CMAS;KDELR1;RPL31;SMC6;SMC3;ZDHHC2;RPL6;RPS15;GOLGA2;PCMT1;C1GALT1C1;HERC2;TFG;RPS19;RPL36;LONP2;THSD7A;RPL37;SEC62;B2M;RPS10;SEC63;NUP214;ARSA;ALG8;CSNK2A1;ALG6;COG6;CSNK2A2;ALG5;ALG2;COG2;COG1;MOGS;ANK2;ANK3;ALG1;HSPG2;DYNC1LI2;XRN2;KDELR2;VDAC1;NUP98;RPL29;TLN1;ATP6V0D1;ATF6;ATF4;GNE;NUP205;SPTBN5;RPN2;SAR1B;RPN1;XPC;AAAS;RPS4Y1;DYNC2LI1;SCFD1;GRPEL1;POM121C;ADAMTS18;RAD21;ARSK;NEU1;SSR1;MLEC;ST3GAL6;TP53BP1;ARSB;SPTBN1;SPTBN2;EDEM3;XBP1;PHC2;RPL41;ICMT;ABCA3;FUCA1;EDEM2;H3F3A;RAB27A;GNG11;MSRA;RPS25;STAG1;RPS28;EIF5;HNRNPK;RPS27;STAG2;RPS29;GOLGB1;CALR;RPS21;ITM2B;RPS24;PIGS;ARF4;TOP2B;TRAM1;SERPINA1;RPLP1;PIGP;SRP54;MIA3;HSCB;PIGV;TUBA1C;TUBA1B;KIF13A;TMED3;RPLP2;CTSH;TMED2;MUC15;NANS;TMED7;CTSD;SPTAN1;CCT3;CCT2;TMED9;RING1;SSR4;EXOC7;SSR2;SSR3;FN3KRP;COPZ2;TIMM22;SUMF2;COPZ1;SUMF1;TUBB2A;TCP1;PIGK;EXOC4;B3GNT3;B3GNT2;EXOC6;PIGL;EXOC5;KIFC3;PIGG;PIGF;ARF5;EXOC2;DCP2;PIGH;DYNC1I2;SP100;STX17;USO1;GNAI3;DERL2;NAPG;GNAI2;GGA2;EXOSC7;GGA1;LMAN1;SAMM50;OS9;LMAN2;EXOSC8;LMNA;SLC17A5;STX5;CD59;CCT8;CCT7;CD55;CCT5;CCT4;RPL19;RANBP[illegible] |
| 11 | ER-Phagosome pathway Homo sapiens R-HSA-1236974 | 5.35E-14 | 6.90E-12 | 6.832975 | 208.81045 | PSMD12;PSMD11;PSMD13;PSMB10;PSMA7;PSMD8;PSMB6;SEC61A1;PSMD6;PSMB4;PSMD7;PSMB5;PSMD4;UBB;PSMB2;PSMB3;PSMD2;PSMD3;UBC;PSMB1;PSMD1;PSMF1;B2M;PDIA3;TAP2;TAP1;HLA-C;HLA-F;PSMB8;TAPBP;HLA-E;PSMB9;PSMA5;PSMA6;PSMA3;PSMC5;PSMA1;PSMA2;PSMC4;PSMC2;PSME1;PSME2;CALR |

| | | | | | | |
|---|---|---|---|---|---|---|
| 12 | Programmed Cell Death Homo sapiens R-HSA-5357801 | 5.82E-14 | 6.90E-12 | 3.346566 | 101.98409 | YWHAE;PPP1R13B;YWHAB;HMGB1;PSMD8;PSMD6;CASP7;PSMD7;PSMD4;FNTA;CDH1;AKT2;PSMD2;PSMD3;AKT1;PSMD1;HIST1H1E;DNM1L;SPTAN1;KPNA1;YWHAH;HIST1H1C;APPL1;DSP;DBNL;DAPK1;PRKCD;DAPK3;TICAM1;PSMA5;PSMA6;PSMA3;PSMA1;PSMA2;PSME1;PSME2;BIRC2;PSMD12;PSMD11;ROCK1;PSMD13;XIAP;PSMB10;PSMA7;ADD1;PSMB6;PSMB4;PSMB5;UBB;PSMB2;PSMB3;UBC;LMNA;PSMB1;TP53BP2;RIPK1;PSMF1;BAK1;BID;PAK2;BCAP31;GSN;BAD;CFLAR;PTK2;PSMB8;PSMB9;TJP1;OCLN;H1F0;PSMC5;STK24;APC;PSMC4;PSMC2;NMT1;FAS;BAX;ACIN1;CTNNB1;TJP2 |
| 13 | AUF1 (hnRNP D0) binds and destabilizes mRNA Homo sapiens R-HSA-450408 | 7.49E-14 | 8.19E-12 | 8.296717 | 250.74914 | PSMD12;PSMD11;PSMD13;HSPB1;PSMB10;PSMA7;PSMD8;PSMB6;PSMD6;PSMB4;PSMD7;PSMB5;PSMD4;UBB;PSMB2;PSMB3;PSMD2;PSMD3;UBC;PSMB1;PSMD1;PSMF1;HSPA8;PSMB8;PSMB9;PSMA5;PSMA6;PSMA3;PSMC5;PSMA1;PSMA2;PSMC4;PSMC2;PSME1;HNRNPD;PSME2;HSPA1A;EIF4G1 |
| 14 | Disease Homo sapiens R-HSA-1643685 | 8.32E-14 | 8.45E-12 | 1.849765 | 55.710532 | RPL5;RPL30;RPL31;CCNC;RBPJ;RPL6;PSMD8;RPS15;PSMD6;C1GALT1C1;PSMD7;PSMD4;RPS19;AKT2;PSMD2;PSMD3;FGFR1OP2;RPL36;AKT1;PSMD1;RPL37;THSD7A;B2M;RPS10;NUP214;MCCC1;HSPG2;BCR;PPP1R3C;CDC37;FDX1;PSME1;PSME2;NUP98;MAML3;RPL29;VPS28;NUP205;CTBP2;SDC4;SDC2;NEDD4L;PIK3R3;RPS4Y1;AAAS;APH1A;APH1B;POM121C;ADAMTS18;PLCG1;PSMF1;RPL41;XRCC6;FZD5;XRCC5;FZD6;EIF2AK2;MSRA;RPS25;RPS28;RPS27;SNW1;RPS29;NMT1;NAPEPLD;SDC1;GRB2;CALR;CALM1;RPS21;RPS24;CD320;CDKN1B;RPLP1;AP2A1;SHH;ZMYM2;XPO1;HEY1;CNTRL;RPLP2;NELFB;CD36;RAC1;MUC15;PSENEN;PSMA5;PSMA6;KAT2A;PSMA3;NCOR1;PSMA1;PSMA2;KIT;VAMP1;TLR5;RNMT;DERL2;PSMA7;EGFR;RELA;PSMB6;NEURL1B;PSMB4;PSMB5;PSMB2;CUX1;OS9;PSMB3;PSMB1;PAK2;RPL19;RANBP2;NPM1;CSNK1A1;STAT1;STAT3;C1GALT1;SYVN1;MIB2;PSMB8;PSMB9;PSMC5;PSMC4;PSMC2;TCEA1;CTNNB1;AGRN;HSPA1A;CCNT2;GTF2B;IRS2;IPO5;EP300;KPNA4;KPNA2;KPNA3;AP2M1;KPNA1;RNF43;SUPT16H;RPS9;MAP2K1;MAP2K2;RPS7;RPS8;RPS5;ADAM10;TICAM1;GTF2F2;ADAM17;TBL1XR1;CHMP4C;CHMP4B;PPIA;VCP;PSMD12;TSG101;PSMD11;CUL5;PSMD13;SEL1L;CUL1;ACACB;ACACA;UBB;TPR;UBC;AP1M2;SMAD2;SMAD4;TGFB1;SMAD3;ISG15;EEF2;NFKB1;CDK9;[illegible] |

| | | | | | |
|---|---|---|---|---|---|
| 15 | Hedgehog ligand biogenesis Homo sapiens R-HSA-5358346 | 1.60E-13 | 1.43E-11 | 6.672563 | 196.61089 | PSMD12;VCP;PSMD11;PSMD13;SEL1L;DERL2;PSMB10;PSMA7;PSMD8;PSMB6;ERLEC1;SHH;PSMD6;PSMB4;PSMD7;PSMB5;PSMD4;OS9;UBB;PSMB2;PSMB3;PSMD2;PSMD3;UBC;PSMB1;PSMD1;PSMF1;SYVN1;PSMB8;PSMB9;PSMA5;PSMA6;ADAM17;PSMA3;PSMC5;PSMA1;PSMA2;PSMC4;PSMC2;PSME1;PSME2;P4HB |
| 16 | Regulation of mRNA stability by proteins that bind AU-rich elements Homo sapiens R-HSA-450531 | 1.60E-13 | 1.43E-11 | 5.099012 | 150.21948 | YWHAB;HSPB1;PSMD8;PSMD6;PSMD7;XPO1;PSMD4;PSMD2;PSMD3;AKT1;PSMD1;TNPO1;NUP214;PRKCD;PSMA5;PSMA6;PSMA3;XRN1;PSMA1;PSMA2;PSME1;PSME2;DCP2;PSMD12;PSMD11;SET;ANP32A;PSMD13;PARN;PSMB10;PSMA7;EXOSC7;PSMB6;PSMB4;PSMB5;UBB;PSMB2;PSMB3;EXOSC8;UBC;PSMB1;PSMF1;HSPA8;PSMB8;PSMB9;PSMC5;PSMC4;PSMC2;HNRNPD;HSPA1A;EIF4G1 |
| 17 | Respiratory electron transport, ATP synthesis by chemiosmotic coupling, and heat production by uncoupling proteins. Homo sapiens R-HSA-163200 | 5.28E-13 | 4.42E-11 | 4.132671 | 116.83 | NDUFA11;NDUFA12;NDUFA10;ETFB;COX6A1;ECSIT;COX7C;UQCRFS1;COX8A;ACAD9;NDUFC2;NDUFC1;SDHD;SDHA;SDHB;COX6B1;NDUFS8;NDUFS7;SCO2;NDUFS5;NDUFS4;UQCRC1;NDUFS3;NDUFS2;NDUFS1;UQCRC2;COX19;NDUFB9;NDUFB8;NDUFB7;COX16;NDUFB11;NDUFB4;ETFDH;NDUFB2;UQCR11;UQCR10;COX5A;UQCRH;UCP2;COX14;NDUFV3;CYC1;NDUFV2;NDUFV1;NDUFA9;NDUFA8;SURF1;NDUFA6;NDUFA3;NDUFA1;COX6C;LRPPRC;NDUFAF6;UQCRQ;NDUFAF7;NDUFAF4;NDUFAB1;COX20 |
| 18 | Regulation of activated PAK-2p34 by proteasome mediated degradation Homo sapiens R-HSA-211733 | 2.95E-12 | 2.33E-10 | 7.911651 | 210.05936 | PSMD12;PSMD11;PSMD13;PSMB10;PSMA7;PSMD8;PSMB6;PSMD6;PSMB4;PSMD7;PSMB5;PSMD4;UBB;PSMB2;PSMB3;PSMD2;PSMD3;UBC;PSMB1;PSMD1;PSMF1;PAK2;PSMB8;PSMB9;PSMA5;PSMA6;PSMA3;PSMC5;PSMA1;PSMA2;PSMC4;PSMC2;PSME1;PSME2 |
| 19 | Activation of NF-kappaB in B cells Homo sapiens R-HSA-1169091 | 4.53E-12 | 3.39E-10 | 5.729648 | 149.66155 | PSMD12;PSMD11;PSMD13;CUL1;BCL10;RELA;PSMB10;PSMA7;MALT1;PSMD8;PSMB6;PSMD6;PSMB4;PSMD7;PSMB5;PSMD4;UBB;PSMB2;PSMB3;PSMD2;PSMD3;UBC;PSMB1;PSMD1;PSMF1;FBXW11;NFKB1;PSMB8;PSMB9;PSMA5;NFKBIA;PSMA6;PSMA3;PSMC5;PSMA1;PSMA2;PSMC4;PSMC2;PSME1;REL;PSME2 |

| | | | | | | |
|---|---|---|---|---|---|---|
| 20 | Cyclin A:Cdk2-associated events at S phase entry Homo sapiens R-HSA-69656 | 6.57E-12 | 4.67E-10 | 5.43515 | 139.94821 | RB1;PSMD12;CDKN1B;PSMD11;PSMD13;MAX;CUL1;PSMB10;PSMA7;CKS1B;PSMD8;PSMB6;PSMD6;FZR1;PSMB4;PSMD7;PSMB5;PSMD4;UBB;PSMB2;PSMB3;PSMD2;PSMD3;UBC;PSMB1;PSMD1;PSMF1;PSMB8;CDC25B;PSMB9;PSMA5;PSMA6;CCNA1;PSMA3;CDK7;PSMC5;PSMA1;PSMA2;PSMC4;PSMC2;PSME1;PSME2 |
| 21 | Regulation of Apoptosis Homo sapiens R-HSA-169911 | 7.22E-12 | 4.89E-10 | 7.416695 | 190.26718 | PSMD12;PSMD11;PSMD13;PSMB10;PSMA7;PSMD8;PSMB6;PSMD6;PSMB4;PSMD7;PSMB5;PSMD4;UBB;PSMB2;PSMB3;PSMD2;PSMD3;UBC;PSMB1;PSMD1;PSMF1;PAK2;PSMB8;PSMB9;PSMA5;PSMA6;PSMA3;PSMC5;PSMA1;PSMA2;PSMC4;PSMC2;PSME1;PSME2 |
| 22 | Diseases of signal transduction Homo sapiens R-HSA-5663202 | 1.28E-11 | 8.29E-10 | 2.339238 | 58.667972 | CCNC;IRS2;RBPJ;PSMD8;PSMD6;PSMD7;PSMD4;AKT2;PSMD2;PSMD3;FGFR1OP2;AKT1;PSMD1;EP300;RNF43;ADAM10;GTF2F2;BCR;ADAM17;TBL1XR1;CDC37;PSME1;PSME2;MAML3;VCP;PSMD12;PSMD11;CTBP2;PSMD13;SEL1L;CUL1;PIK3R3;APH1A;APH1B;UBB;UBC;PLCG1;PSMF1;SMAD2;SMAD4;TGFB1;SMAD3;FZD5;FZD6;SNW1;NAPEPLD;GRB2;GSK3B;CDKN1B;SHH;ZMYM2;HEY1;CNTRL;MLST8;LRRFIP1;PSENEN;HSP90AA1;PPP2R5B;TGFBR1;PSMA5;PSMA6;KAT2A;PSMA3;NCOR1;CREB1;PSMA1;PSMA2;KIT;SOS1;HDAC2;HDAC1;DERL2;CBL;HDAC8;HDAC9;PSMA7;EGFR;PSMB10;HDAC6;LRP6;PSMB6;NEURL1B;ERLEC1;PSMB4;PSMB5;CUX1;OS9;PSMB2;ERBB3;TNKS2;PSMB3;POLR2B;POLR2C;PSMB1;ERBB2;POLR2E;HES1;POLR2H;POLR2J;CSNK1A1;STAT1;BAD;STAT3;SYVN1;MIB2;PTPN11;PSMB8;MTOR;PSMB9;PSMC5;APC;PSMC4;PSMC2;RPS6KB2;CTNNB1 |
| 23 | NIK-->noncanonical NF-kB signaling Homo sapiens R-HSA-5676590 | 1.59E-11 | 9.80E-10 | 6.151587 | 152.9735 | PSMD12;PSMD11;PSMD13;CUL1;PSMB10;PSMA7;PSMD8;PSMB6;PSMD6;PSMB4;PSMD7;PSMB5;PSMD4;UBB;PSMB2;PSMB3;PSMD2;PSMD3;UBC;PSMB1;PSMD1;PSMF1;FBXW11;PSMB8;PSMB9;PSMA5;PSMA6;PSMA3;PSMC5;PSMA1;PSMA2;PSMC4;PSMC2;PSME1;UBA3;PSME2;UBE2M |
| 24 | Autodegradation of the E3 ubiquitin ligase COP1 Homo sapiens R-HSA-349425 | 1.70E-11 | 1.01E-09 | 6.979968 | 173.0888 | PSMD12;PSMD11;PSMD13;PSMB10;PSMA7;PSMD8;PSMB6;PSMD6;PSMB4;PSMD7;PSMB5;PSMD4;UBB;PSMB2;PSMB3;PSMD2;PSMD3;UBC;PSMB1;PSMD1;PSMF1;PSMB8;PSMB9;PSMA5;PSMA6;PSMA3;PSMC5;PSMA1;PSMA2;PSMC4;PSMC2;PSME1;PSME2;ATM |

| | | | | | | |
|---|---|---|---|---|---|---|
| 25 | HIV Infection Homo sapiens R-HSA-162906 | 2.10E-11 | 1.20E-09 | 2.579444 | 63.417438 | CCNT2;GTF2B;PSIP1;AP2A1;PSMD8;PSMD6;PSMD7;XPO1;PSMD4;AP1G1;PSMD2;PSMD3;PSMD1;NELFB;RAC1;B2M;AP2M1;KPNA1;NUP214;SUPT16H;PDCD6IP;VPS37D;VPS37B;GTF2F2;PSMA5;PSMA6;PSMA3;PSMA1;PSMA2;NUP50;PSME1;CHMP4C;PSME2;CHMP4B;CHMP3;NUP98;PPIA;SLC25A4;VPS28;CHMP5;GTF2A1;NUP205;PSMD12;GTF2A2;TSG101;PSMD11;CUL5;RNMT;PSMD13;VPS4B;VPS4A;NEDD4L;FURIN;AAAS;PSMA7;PSMB10;PSMB6;PSMB4;PSMB5;POM121C;PSMB2;UBB;PSMB3;POLR2B;POLR2C;PSMB1;TPR;UBC;POLR2E;PSMF1;POLR2H;PAK2;POLR2J;AP1M2;RANBP2;XRCC6;NPM1;XRCC5;TAF11;SSRP1;PSMB8;PSMB9;CDK9;PSMC5;CDK7;PSMC4;PSMC2;CHMP2B;NMT1;TCEA1;TAF5;TAF3;RAN;TAF2 |
| 26 | Dectin-1 mediated noncanonical NF-kB signaling Homo sapiens R-HSA-5607761 | 2.43E-11 | 1.33E-09 | 5.769024 | 141.0066 | PSMD12;PSMD11;PSMD13;CUL1;RELA;PSMB10;PSMA7;PSMD8;PSMB6;PSMD6;PSMB4;PSMD7;PSMB5;PSMD4;UBB;PSMB2;PSMB3;PSMD2;PSMD3;UBC;PSMB1;PSMD1;PSMF1;FBXW11;PSMB8;PSMB9;PSMA5;PSMA6;PSMA3;PSMC5;PSMA1;PSMA2;PSMC4;PSMC2;PSME1;UBA3;PSME2;UBE2M |
| 27 | G2/M Transition Homo sapiens R-HSA-69275 | 2.67E-11 | 1.41E-09 | 2.879564 | 70.109211 | YWHAE;CEP57;HSP90AB1;NUMA1;PPP2R2A;PSMD8;PSMD6;ACTR1A;PCM1;PSMD7;XPO1;PSMD4;PSMD2;PSMD3;CNTRL;PSMD1;EP300;CEP290;RAB8A;CDK5RAP2;OFD1;HSP90AA1;PPP1R12A;FBXW11;TUBB;HAUS4;HAUS3;TUBG1;TUBA4A;CDC25B;HAUS1;PSMA5;CCNA1;PSMA6;PSMA3;PSMA1;PSMA2;AKAP9;PSME1;PSME2;MAPRE1;OPTN;PAFAH1B1;SDCCAG8;DYNC1I2;PSMD12;PSMD11;DCTN2;TUBGCP2;PSMD13;SSNA1;CUL1;PSMB10;PSMA7;PSMB6;PSMB4;PSMB5;UBB;RBBP4;PSMB2;PSMB3;UBC;CEP70;PSMB1;PSMF1;CEP78;DYNC1H1;CDK11A;CEP152;TUBB4B;CDK11B;PSMB8;PSMB9;CDK7;PSMC5;PSMC4;CENPJ;PSMC2 |
| 28 | SCF-beta-TrCP mediated degradation of Emi1 Homo sapiens R-HSA-174113 | 2.88E-11 | 1.46E-09 | 6.429536 | 156.04096 | PSMD12;PSMD11;PSMD13;CUL1;PSMB10;PSMA7;PSMD8;PSMB6;PSMD6;FZR1;PSMB4;PSMD7;PSMB5;PSMD4;UBB;PSMB2;PSMB3;PSMD2;PSMD3;UBC;PSMB1;PSMD1;PSMF1;PSMB8;PSMB9;PSMA5;PSMA6;PSMA3;PSMC5;PSMA1;PSMA2;PSMC4;PSMC2;PSME1;PSME2 |

| | | | | | | |
|---|---|---|---|---|---|---|
| 29 | SCF(Skp2)-mediated degradation of p27/p21 Homo sapiens R-HSA-187577 | 3.34E-11 | 1.64E-09 | 5.871591 | 141.64547 | PSMD12;CDKN1B;PSMD11;PSMD13;CUL1;PSMB10;PSMA7;CKS1B;PSMD8;PSMB6;PSMD6;PSMB4;PSMD7;PSMB5;PSMD4;UBB;PSMB2;PSMB3;PSMD2;PSMD3;UBC;PSMB1;PSMD1;PSMF1;PSMB8;PSMB9;PSMA5;PSMA6;CCNA1;PSMA3;PSMC5;PSMA1;PSMA2;PSMC4;PSMC2;PSME1;PSME2 |
| 30 | Antigen processing-Cross presentation Homo sapiens R-HSA-1236975 | 3.64E-11 | 1.66E-09 | 4.466022 | 107.34281 | PSMD12;PSMD11;PSMD13;CTSS;PSMB10;PSMA7;PSMD8;PSMB6;SEC61A1;PSMD6;PSMB4;PSMD7;PSMB5;PSMD4;UBB;PSMB2;CTSL;PSMB3;PSMD2;PSMD3;UBC;PSMB1;PSMD1;CD36;PSMF1;B2M;PDIA3;TAP2;TAP1;HLA-C;HLA-F;PSMB8;TAPBP;HLA-E;PSMB9;PSMA5;PSMA6;PSMA3;PSMC5;PSMA1;PSMA2;PSMC4;PSMC2;PSME1;PSME2;CALR |
| 31 | p53-Independent DNA Damage Response Homo sapiens R-HSA-69610 | 3.85E-11 | 1.66E-09 | 6.591767 | 158.06821 | PSMD12;PSMD11;PSMD13;PSMB10;PSMA7;PSMD8;PSMB6;PSMD6;PSMB4;PSMD7;PSMB5;PSMD4;UBB;PSMB2;PSMB3;PSMD2;PSMD3;UBC;PSMB1;PSMD1;PSMF1;PSMB8;PSMB9;PSMA5;PSMA6;PSMA3;PSMC5;PSMA1;PSMA2;PSMC4;PSMC2;PSME1;PSME2;ATM |
| 32 | p53-Independent G1/S DNA damage checkpoint Homo sapiens R-HSA-69613 | 3.85E-11 | 1.66E-09 | 6.591767 | 158.06821 | PSMD12;PSMD11;PSMD13;PSMB10;PSMA7;PSMD8;PSMB6;PSMD6;PSMB4;PSMD7;PSMB5;PSMD4;UBB;PSMB2;PSMB3;PSMD2;PSMD3;UBC;PSMB1;PSMD1;PSMF1;PSMB8;PSMB9;PSMA5;PSMA6;PSMA3;PSMC5;PSMA1;PSMA2;PSMC4;PSMC2;PSME1;PSME2;ATM |
| 33 | Ubiquitin Mediated Degradation of Phosphorylated Cdc25A Homo sapiens R-HSA-69601 | 3.85E-11 | 1.66E-09 | 6.591767 | 158.06821 | PSMD12;PSMD11;PSMD13;PSMB10;PSMA7;PSMD8;PSMB6;PSMD6;PSMB4;PSMD7;PSMB5;PSMD4;UBB;PSMB2;PSMB3;PSMD2;PSMD3;UBC;PSMB1;PSMD1;PSMF1;PSMB8;PSMB9;PSMA5;PSMA6;PSMA3;PSMC5;PSMA1;PSMA2;PSMC4;PSMC2;PSME1;PSME2;ATM |
| 34 | Autodegradation of Cdh1 by Cdh1:APC/C Homo sapiens R-HSA-174084 | 4.92E-11 | 1.91E-09 | 5.528291 | 131.21997 | PSMD12;PSMD11;ANAPC16;PSMD13;PSMB10;PSMA7;ANAPC11;PSMD8;PSMB6;PSMD6;FZR1;PSMB4;PSMD7;PSMB5;PSMD4;UBB;PSMB2;PSMB3;PSMD2;PSMD3;UBC;PSMB1;PSMD1;PSMF1;UBE2E1;PSMB8;PSMB9;PSMA5;PSMA6;PSMA3;PSMC5;PSMA1;PSMA2;PSMC4;PSMC2;PSME1;PSME2;ANAPC5 |
| 35 | CDK-mediated phosphorylation and removal of Cdc6 Homo sapiens R-HSA-69017 | 5.11E-11 | 1.91E-09 | 6.773151 | 160.50378 | PSMD12;PSMD11;PSMD13;PSMB10;PSMA7;PSMD8;PSMB6;PSMD6;PSMB4;PSMD7;PSMB5;PSMD4;UBB;PSMB2;PSMB3;PSMD2;PSMD3;UBC;PSMB1;PSMD1;PSMF1;PSMB8;PSMB9;PSMA5;PSMA6;PSMA3;PSMC5;PSMA1;PSMA2;PSMC4;PSMC2;PSME1;PSME2 |

| | | | | | | |
|---|---|---|---|---|---|---|
| 36 | Ubiquitin-dependent degradation of Cyclin D1 Homo sapiens R-HSA-69229 | 5.11E-11 | 1.91E-09 | 6.773151 | 160.50378 | PSMD12;PSMD11;PSMD13;PSMB10;PSMA7;PSMD8;PSMB6;PSMD6;PSMB4;PSMD7;PSMB5;PSMD4;UBB;PSMB2;PSMB3;PSMD2;PSMD3;UBC;PSMB1;PSMD1;PSMF1;PSMB8;PSMB9;PSMA5;PSMA6;PSMA3;PSMC5;PSMA1;PSMA2;PSMC4;PSMC2;PSME1;PSME2 |
| 37 | Ubiquitin-dependent degradation of Cyclin D Homo sapiens R-HSA-75815 | 5.11E-11 | 1.91E-09 | 6.773151 | 160.50378 | PSMD12;PSMD11;PSMD13;PSMB10;PSMA7;PSMD8;PSMB6;PSMD6;PSMB4;PSMD7;PSMB5;PSMD4;UBB;PSMB2;PSMB3;PSMD2;PSMD3;UBC;PSMB1;PSMD1;PSMF1;PSMB8;PSMB9;PSMA5;PSMA6;PSMA3;PSMC5;PSMA1;PSMA2;PSMC4;PSMC2;PSME1;PSME2 |
| 38 | Regulation of ornithine decarboxylase (ODC) Homo sapiens R-HSA-350562 | 5.11E-11 | 1.91E-09 | 6.773151 | 160.50378 | PSMD12;PSMD11;PSMD13;OAZ1;PSMB10;PSMA7;PSMD8;PSMB6;PSMD6;PSMB4;PSMD7;PSMB5;PSMD4;PSMB2;PSMB3;PSMD2;PSMD3;PSMB1;PSMD1;PSMF1;NQO1;PSMB8;PSMB9;PSMA5;PSMA6;PSMA3;PSMC5;PSMA1;PSMA2;PSMC4;PSMC2;PSME1;PSME2 |
| 39 | Mitotic G2-G2/M phases Homo sapiens R-HSA-453274 | 5.34E-11 | 1.95E-09 | 2.819825 | 66.698227 | YWHAE;CEP57;HSP90AB1;NUMA1;PPP2R2A;PSMD8;PSMD6;ACTR1A;PCM1;PSMD7;XPO1;PSMD4;PSMD2;PSMD3;CNTRL;PSMD1;EP300;CEP290;RAB8A;CDK5RAP2;OFD1;HSP90AA1;PPP1R12A;FBXW11;TUBB;HAUS4;HAUS3;TUBG1;TUBA4A;CDC25B;HAUS1;PSMA5;CCNA1;PSMA6;PSMA3;PSMA1;PSMA2;AKAP9;PSME1;PSME2;MAPRE1;OPTN;PAFAH1B1;SDCCAG8;DYNC1I2;PSMD12;PSMD11;DCTN2;TUBGCP2;PSMD13;SSNA1;CUL1;PSMB10;PSMA7;PSMB6;PSMB4;PSMB5;UBB;RBBP4;PSMB2;PSMB3;UBC;CEP70;PSMB1;PSMF1;CEP78;DYNC1H1;CDK11A;CEP152;TUBB4B;CDK11B;PSMB8;PSMB9;CDK7;PSMC5;PSMC4;CENPJ;PSMC2 |

| | | | | | | |
|---|---|---|---|---|---|---|
| 40 | Asparagine N-linked glycosylation Homo sapiens R-HSA-446203 | 8.23E-11 | 2.92E-09 | 2.357977 | 54.754187 | DPAGT1;CMAS;KDELR1;GOLGA2;TFG;PDIA3;NSF;ALG8;COG6;ALG6;ALG5;COG2;ALG2;ALG14;COG1;MOGS;ANK2;ALG12;ANK3;ALG1;DPM2;DYNC1LI2;DPM3;TBC1D20;KDELR2;GNE;COPB2;SPTBN5;COPA;TMED10;RPN2;SAR1B;COPB1;RPN1;SEL1L;DYNC2LI1;SCFD1;FUT8;MAN2A2;MAN2A1;NEU1;MLEC;ST3GAL6;SPTBN1;SPTBN2;MPDU1;EDEM3;TRAPPC1;FUCA1;TRAPPC4;EDEM2;GFPT1;TMEM115;GOLGB1;CALR;COPE;ARF4;B4GALT3;MCFD2;SERPINA1;B4GALT1;MIA3;GMPPB;PPP6R3;TMED3;PGM3;TMED2;MAN1A1;TMED7;NANS;SPTAN1;TMED9;COPZ2;ARFGAP3;COPZ1;DDOST;DAD1;ARF5;DYNC1I2;STX17;DCTN2;USO1;DERL2;NAPG;LMAN1;GANAB;OS9;LMAN2;SLC17A5;MAN1C1;STX5;CD59;UGGT2;CD55;DYNC1H1;SLC35A1;TRAPPC10;SYVN1;ARCN1;NAGK;DOLPP1;MGAT4B;COPG2;FOLR1 |
| 41 | Vif-mediated degradation of APOBEC3G Homo sapiens R-HSA-180585 | 8.43E-11 | 2.92E-09 | 6.244429 | 144.84765 | PSMD12;PSMD11;CUL5;PSMD13;PSMB10;PSMA7;PSMD8;PSMB6;PSMD6;PSMB4;PSMD7;PSMB5;PSMD4;UBB;PSMB2;PSMB3;PSMD2;PSMD3;UBC;PSMB1;PSMD1;PSMF1;PSMB8;PSMB9;PSMA5;PSMA6;PSMA3;PSMC5;PSMA1;PSMA2;PSMC4;PSMC2;PSME1;PSME2 |
| 42 | Cyclin E associated events during G1/S transition Homo sapiens R-HSA-69202 | 9.36E-11 | 3.17E-09 | 4.988892 | 115.20177 | RB1;PSMD12;CDKN1B;PSMD11;PSMD13;MAX;CUL1;PSMB10;PSMA7;CKS1B;PSMD8;PSMB6;PSMD6;PSMB4;PSMD7;PSMB5;PSMD4;UBB;PSMB2;PSMB3;PSMD2;PSMD3;UBC;PSMB1;PSMD1;PSMF1;PSMB8;PSMB9;PSMA5;PSMA6;CCNA1;PSMA3;CDK7;PSMC5;PSMA1;PSMA2;PSMC4;PSMC2;PSME1;PSME2 |

| | | | | | | |
|---|---|---|---|---|---|---|
| 43 | Gene Expression Homo sapiens R-HSA-74160 | 1.01E-10 | 3.34E-09 | 1.455785 | 33.507857 | ATF2;RPL5;RPL30;RPL31;HNRNPU;EHMT1;HNRNPR;PHAX;EPRS;RBPJ;RPL6;GLS;RRP9;PSMD8;PSMD6;PSMD7;PSMD4;AKT2;PSMD2;PSMD3;RPL36;AKT1;PSMD1;RPL37;NUP214;CSNK2A1;ZNF160;PRKCD;CSNK2A2;MED8;WDR75;DDX39A;DDX39B;SCO2;DDIT4;PSME1;PSME2;NUP98;MAML3;RPL29;SLBP;NUP205;ZNF274;SET;MEAF6;RPN2;RPN1;NEDD4L;RPS4Y1;RNPC3;AAAS;POM121C;PRDX1;SSR1;PSMF1;ATRIP;PRELID1;RPL41;ZNF141;H3F3A;UPF3B;NR1D2;UPF3A;NR1D1;SNUPN;NR2F6;HNRNPM;HNRNPK;EIF5;QARS;AGO3;HNRNPF;HNRNPD;RAD17;CDK12;ZNF138;FARSA;CDK13;ZNF257;ZNF254;BTG2;SMG1;RARG;CDKN1B;PHF20;PHF1;RPLP1;SRP54;PRKAG2;CHM;SMG9;WDR43;SMG5;SMG6;MED13;AIFM2;MED10;RARS;RPLP2;TNKS1BP1;NELFB;TEAD1;TEAD2;ZNF486;UPF2;SSR4;NSUN2;SSR2;SSR3;BAZ1B;SIRT1;PSMA5;PSMA6;KAT2A;PSMA3;NCOR1;PSMA1;PSMA2;PPARG;GARS;ZNF595;SF3B3;RNMT;URM1;PSMA7;PSMB6;PSMB4;PSMB5;PSMB2;PSMB3;PSMB1;HARS;BID;BRD7;ZNF224;SF3B1;SRSF11;PAIP1;RPL19;TRMT10A;UTP6;USP9X;RIOK3;NR1H2;MBD2;GSR;HIPK1;PSMB8;LRPPRC;ZNF33B;PSMB9;ZNF33A;PSMC5;PHF5A;NFIA;NFIB;PSMC4;CENPJ;PSMC2;TRMT6;TSR1;LAMTOR1;ZNF211;HSPA1A;ZNF331;CRCP;CCNT2;GTF2B;RORA;SESN3;PAPOLA;ZNF205;ZNF567;NBN;TXNL4A;SNRNP35;Z[illegible] |
| 44 | Vpu mediated degradation of CD4 Homo sapiens R-HSA-180534 | 1.14E-10 | 3.67E-09 | 6.396452 | 146.46547 | PSMD12;PSMD11;PSMD13;PSMB10;PSMA7;PSMD8;PSMB6;PSMD6;PSMB4;PSMD7;PSMB5;PSMD4;UBB;PSMB2;PSMB3;PSMD2;PSMD3;UBC;PSMB1;PSMD1;PSMF1;PSMB8;PSMB9;PSMA5;PSMA6;PSMA3;PSMC5;PSMA1;PSMA2;PSMC4;PSMC2;PSME1;PSME2 |
| 45 | Degradation of DVL Homo sapiens R-HSA-4641258 | 1.30E-10 | 4.11E-09 | 5.816449 | 132.39575 | PSMD12;PSMD11;PSMD13;CUL3;PSMB10;PSMA7;PSMD8;PSMB6;PSMD6;PSMB4;PSMD7;PSMB5;PSMD4;UBB;PSMB2;PSMB3;PSMD2;PSMD3;UBC;PSMB1;PSMD1;DVL3;PSMF1;PSMB8;PSMB9;PSMA5;PSMA6;PSMA3;PSMC5;PSMA1;PSMA2;PSMC4;PSMC2;PSME1;PSME2 |
| 46 | Complex I biogenesis Homo sapiens R-HSA-6799198 | 1.52E-10 | 4.70E-09 | 6.566426 | 148.44142 | NDUFB9;NDUFB8;NDUFB7;NDUFA11;NDUFA12;NDUFB11;NDUFB4;NDUFA10;NDUFB2;ECSIT;NDUFV3;NDUFV2;NDUFV1;NDUFA9;NDUFA8;NDUFA6;NDUFA3;ACAD9;NDUFA1;NDUFC2;NDUFC1;NDUFS8;NDUFS7;NDUFAF6;NDUFAF7;NDUFS5;NDUFAF4;NDUFS4;NDUFAB1;NDUFS3;NDUFS2;NDUFS1 |

| | | | | | | |
|---|---|---|---|---|---|---|
| 47 | Stabilization of p53 Homo sapiens R-HSA-69541 | 1.79E-10 | 5.20E-09 | 5.931825 | 133.14274 | PSMD12;PSMD11;PSMD13;PSMB10;PSMA7;PSMD8;PSMB6;PSMD6;PSMB4;PSMD7;PSMB5;PSMD4;UBB;PSMB2;PSMB3;PSMD2;PSMD3;UBC;PSMB1;PSMD1;PSMF1;PSMB8;PSMB9;PSMA5;PSMA6;PSMA3;PSMC5;PSMA1;PSMA2;PSMC4;PSMC2;PSME1;PSME2;ATM |
| 48 | Degradation of AXIN Homo sapiens R-HSA-4641257 | 1.79E-10 | 5.20E-09 | 5.931825 | 133.14274 | PSMD12;PSMD11;PSMD13;PSMB10;PSMA7;PSMD8;PSMB6;PSMD6;PSMB4;PSMD7;PSMB5;PSMD4;TNKS2;UBB;PSMB2;PSMB3;PSMD2;PSMD3;UBC;PSMB1;PSMD1;PSMF1;PSMB8;PSMB9;PSMA5;PSMA6;PSMA3;PSMC5;PSMA1;PSMA2;PSMC4;PSMC2;PSME1;PSME2 |
| 49 | Degradation of GLI1 by the proteasome Homo sapiens R-HSA-5610780 | 1.90E-10 | 5.20E-09 | 5.462929 | 122.28214 | PSMD12;PSMD11;PSMD13;CUL1;PSMB10;PSMA7;PSMD8;PSMB6;PSMD6;PSMB4;PSMD7;PSMB5;PSMD4;UBB;PSMB2;PSMB3;PSMD2;PSMD3;UBC;PSMB1;PSMD1;PSMF1;PSMB8;PSMB9;PSMA5;PSMA6;PSMA3;PSMC5;ITCH;PSMA1;PSMA2;PSMC4;PSMC2;NUMB;PSME1;PSME2 |
| 50 | GLI3 is processed to GLI3R by the proteasome Homo sapiens R-HSA-5610785 | 1.90E-10 | 5.20E-09 | 5.462929 | 122.28214 | GSK3B;PSMD12;PSMD11;PSMD13;CUL1;PSMB10;PSMA7;PSMD8;PSMB6;PSMD6;PSMB4;PSMD7;PSMB5;PSMD4;UBB;PSMB2;PSMB3;PSMD2;PSMD3;UBC;PSMB1;PSMD1;PSMF1;CSNK1A1;PSMB8;PSMB9;PSMA5;PSMA6;PSMA3;PSMC5;PSMA1;PSMA2;PSMC4;PSMC2;PSME1;PSME2 |
| 51 | Degradation of GLI2 by the proteasome Homo sapiens R-HSA-5610783 | 1.90E-10 | 5.20E-09 | 5.462929 | 122.28214 | GSK3B;PSMD12;PSMD11;PSMD13;CUL1;PSMB10;PSMA7;PSMD8;PSMB6;PSMD6;PSMB4;PSMD7;PSMB5;PSMD4;UBB;PSMB2;PSMB3;PSMD2;PSMD3;UBC;PSMB1;PSMD1;PSMF1;CSNK1A1;PSMB8;PSMB9;PSMA5;PSMA6;PSMA3;PSMC5;PSMA1;PSMA2;PSMC4;PSMC2;PSME1;PSME2 |
| 52 | The role of GTSE1 in G2/M progression after G2 checkpoint Homo sapiens R-HSA-8852276 | 1.90E-10 | 5.20E-09 | 5.462929 | 122.28214 | PSMD12;HSP90AB1;PSMD11;PSMD13;PSMB10;PSMA7;PSMD8;PSMB6;PSMD6;PSMB4;PSMD7;PSMB5;PSMD4;UBB;PSMB2;PSMB3;PSMD2;PSMD3;UBC;PSMB1;PSMD1;PSMF1;HSP90AA1;PSMB8;PSMB9;PSMA5;PSMA6;PSMA3;PSMC5;PSMA1;PSMA2;PSMC4;PSMC2;PSME1;PSME2;MAPRE1 |
| 53 | CLEC7A (Dectin-1) signaling Homo sapiens R-HSA-5607764 | 2.14E-10 | 5.75E-09 | 3.714909 | 82.704789 | AHCYL1;UBE2D2;PYCARD;PSMD8;PSMD6;PSMD7;PSMD4;PSMD2;PSMD3;PSMD1;FBXW11;PRKCD;PSMA5;PSMA6;PSMA3;PSMA1;PSMA2;PSME1;PSME2;UBE2V1;PSMD12;PSMD11;PSMD13;SRC;CUL1;ITPR2;ITPR3;BCL10;RELA;PSMB10;PSMA7;MALT1;PSMB6;PSMB4;PSMB5;UBB;PSMB2;PSMB3;UBC;PSMB1;PSMF1;NFKB1;PSMB8;PSMB9;NFKBIA;PSMC5;PSMC4;PSMC2;UBA3;CALM1;UBE2M |

| | | | | | | |
|---|---|---|---|---|---|---|
| 54 | Metabolism of amino acids and derivatives Homo sapiens R-HSA-71291 | 6.94E-10 | 1.83E-08 | 2.0672 | 43.595174 | PNMT;RPL5;RPL30;RPL31;EPRS;SAT1;RPL6;GLS;CNDP2;PSMD8;RPS15;PSMD6;PSMD7;PSMD4;RPS19;PSMD2;PSMD3;RPL36;PSMD1;RPL37;GLUL;RPS10;PDHX;RPS9;RPS7;HGD;PHYKPL;RPS8;SECISBP2;RPS5;MCCC1;SLC6A12;SARS;GCAT;MTAP;KARS;PSME1;SMS;PSME2;RPL29;ALDH7A1;IDO1;PSMD12;PSMD11;PSMD13;SHMT1;MRI1;RPS4Y1;OAZ1;ACAT1;LARS;AFMID;PSMF1;DARS;RPL41;PDHA1;GOT1;GOT2;NMRAL1;AIMP1;AIMP2;RPS25;GLUD1;RPS28;GCLC;RPS27;QARS;RPS29;ADI1;OGDH;RPS21;RPS24;RPLP1;APIP;MTR;RPL7A;AUH;RARS;KYNU;RPLP2;HIBCH;ACAD8;BCKDHA;ARG2;BHMT2;AMT;PSMA5;PSMA6;PSMA3;PSMA1;TST;PSMA2;IVD;RPL37A;DLD;ASPA;ALDH9A1;DLST;PDHB;PSMA7;PAPSS2;PSMB10;PSMB6;PSMB4;PSMB5;PSMB2;PSMB3;PSMB1;PCBD1;CRYM;RPL19;NQO1;TXNRD1;GSR;RPL35A;PSMB8;PSMB9;ALDH4A1;PSMC5;TMLHE;PSMC4;MARS;PSMC2;NDUFAB1 |
| 55 | Cross-presentation of soluble exogenous antigens (endosomes) Homo sapiens R-HSA-1236978 | 9.58E-10 | 2.48E-08 | 6.006085 | 124.72159 | PSMD12;PSMD11;PSMD13;PSMB10;PSMA7;PSMD8;PSMB6;PSMD6;PSMB4;PSMD7;PSMB5;PSMD4;PSMB2;PSMB3;PSMD2;PSMD3;PSMB1;PSMD1;PSMF1;PSMB8;PSMB9;PSMA5;PSMA6;PSMA3;PSMC5;PSMA1;PSMA2;PSMC4;PSMC2;PSME1;PSME2 |
| 56 | COPI-mediated anterograde transport Homo sapiens R-HSA-6807878 | 1.42E-09 | 3.61E-08 | 4.073995 | 82.993072 | ARF4;DYNC1I2;COPB2;SPTBN5;COPA;TMED10;DCTN2;KDELR1;COPB1;USO1;NAPG;GOLGA2;DYNC2LI1;TMED3;TMED2;STX5;CD59;TMED7;SPTAN1;SPTBN1;CD55;SPTBN2;NSF;DYNC1H1;TMED9;COG6;COG2;COG1;COPZ2;ANK2;ANK3;ARFGAP3;COPZ1;ARCN1;DYNC1LI2;TMEM115;GOLGB1;KDELR2;COPG2;FOLR1;ARF5;COPE |
| 57 | Intra-Golgi and retrograde Golgi-to-ER traffic Homo sapiens R-HSA-6811442 | 1.46E-09 | 3.64E-08 | 2.585291 | 52.596373 | ARF4;SYS1;KDELR1;GOLIM4;GCC2;KIF12;GOLGA4;GOLGA5;KIF5B;TMED3;KIF13B;ARFIP2;TMED2;KIF21A;KIF1B;MAN1A1;KIFAP3;TMED7;NSF;TMED9;COG6;COG2;KIF25;COG1;COPZ2;KIF23;ARFGAP3;COPZ1;RINT1;DYNC1LI2;KIF2A;KIF16B;KDELR2;NAA38;PAFAH1B3;ARF5;VAMP3;PAFAH1B1;COPB2;DYNC1I2;COPA;NBAS;TMED10;TMF1;DCTN2;STX16;COPB1;STX18;ARL1;NAPG;AGPAT3;STX10;TGOLN2;DYNC2LI1;CYTH2;KLC2;MAN2A2;CUX1;KIF3A;MAN2A1;VPS53;STX6;MAN1C1;STX5;DYNC1H1;GALNT1;SURF4;M6PR;IGF2R;ARCN1;TRIP11;RAB9A;RAB3GAP2;COPG2;COPE;RAB3GAP1 |

| | | | | | | |
|---|---|---|---|---|---|---|
| 58 | Translation Homo sapiens R-HSA-72766 | 1.50E-09 | 3.67E-08 | 2.79239 | 56.741966 | RPL5;EIF4A1;RPL30;TRAM1;RPL31;RPLP1;SRP54;RPL6;EEF1B2;RPS15;SEC61A1;RPL7A;RPS19;RPL36;RPLP2;RPL37;RPS10;EIF5B;RPS9;RPS7;SSR4;RPS8;RPS5;SSR2;SSR3;DDOST;RPL37A;ETF1;RPL29;RPN2;RPN1;RPS4Y1;EIF4H;SSR1;SEC11A;EIF4B;RPL19;SEC11C;EIF2B5;EIF2B4;RPL41;EIF2B2;RPL35A;EEF2;EIF2S2;EIF2S1;RPS25;SPCS3;RPS28;SPCS2;EIF3M;RPS27;EIF5;SPCS1;EIF3K;RPS29;EIF3L;EIF3I;EIF3J;EIF3G;EIF3H;EIF3C;RPS21;EIF3D;RPS24;EIF3A;EIF4G1 |
| 59 | Signaling by Hedgehog Homo sapiens R-HSA-5358351 | 1.64E-09 | 3.94E-08 | 2.931768 | 59.314399 | GSK3B;ARRB1;CDC73;PSMD8;PSMD6;SHH;PSMD7;PSMD4;PSMD2;PSMD3;PSMD1;IFT57;OFD1;PSMA5;PSMA6;IFT88;ADAM17;PSMA3;ADCY9;PSMA1;PSMA2;ULK3;PSME1;NUMB;PSME2;VCP;PSMD12;PSMD11;INTU;PSMD13;CUL3;SEL1L;CUL1;DERL2;PSMB10;ADCY6;PSMA7;PSMB6;ERLEC1;PSMB4;PSMB5;UBB;OS9;PSMB2;KIF3A;PSMB3;TTC21B;PRKAR2A;UBC;PSMB1;SPOP;PSMF1;EVC2;CSNK1A1;SYVN1;PSMB8;PSMB9;PSMC5;ITCH;PSMC4;PSMC2;P4HB |
| 60 | p53-Dependent G1/S DNA damage checkpoint Homo sapiens R-HSA-69580 | 2.44E-09 | 5.70E-08 | 4.652406 | 92.255946 | PSMD12;CDKN1B;PSMD11;PSMD13;PSMB10;PSMA7;PSMD8;PSMB6;PSMD6;PSMB4;PSMD7;PSMB5;PSMD4;UBB;PSMB2;PSMB3;PSMD2;PSMD3;UBC;PSMB1;PSMD1;PSMF1;PSMB8;PSMB9;PSMA5;PSMA6;CCNA1;PSMA3;PSMC5;PSMA1;PSMA2;PSMC4;PSMC2;PSME1;PSME2;ATM |
| 61 | p53-Dependent G1 DNA Damage Response Homo sapiens R-HSA-69563 | 2.44E-09 | 5.70E-08 | 4.652406 | 92.255946 | PSMD12;CDKN1B;PSMD11;PSMD13;PSMB10;PSMA7;PSMD8;PSMB6;PSMD6;PSMB4;PSMD7;PSMB5;PSMD4;UBB;PSMB2;PSMB3;PSMD2;PSMD3;UBC;PSMB1;PSMD1;PSMF1;PSMB8;PSMB9;PSMA5;PSMA6;CCNA1;PSMA3;PSMC5;PSMA1;PSMA2;PSMC4;PSMC2;PSME1;PSME2;ATM |
| 62 | Cdc20:Phospho-APC/C mediated degradation of Cyclin A Homo sapiens R-HSA-174184 | 2.54E-09 | 5.83E-08 | 4.254091 | 84.186643 | PSMD12;PSMD11;ANAPC16;PSMD13;PSMB10;PSMA7;ANAPC11;PSMD8;PSMB6;PSMD6;PSMB4;PSMD7;PSMB5;PSMD4;UBB;PSMB2;PSMB3;PSMD2;PSMD3;UBC;PSMB1;PSMD1;PSMF1;UBE2E1;PSMB8;PSMB9;PSMA5;PSMA6;CCNA1;PSMA3;PSMC5;PSMA1;PSMA2;PSMC4;PSMC2;PSME1;PSME2;ANAPC5;MAD2L1 |

| | | | | | | |
|---|---|---|---|---|---|---|
| 63 | Degradation of beta-catenin by the destruction complex Homo sapiens R-HSA-195253 | 2.70E-09 | 6.01E-08 | 3.850271 | 75.971609 | GSK3B;PSMD12;PSMD11;CTBP2;PSMD13;HDAC1;CUL1;PSMB10;PSMA7;PSMD8;PSMB6;PSMD6;PSMB4;PSMD7;PSMB5;PSMD4;UBB;PSMB2;PSMB3;PSMD2;PSMD3;UBC;PSMB1;PSMD1;PSMF1;TLE3;TLE2;CSNK1A1;PPP2R5B;PSMB8;PSMB9;PSMA5;PSMA6;PSMA3;PSMC5;PSMA1;APC;PSMA2;PSMC4;PSMC2;PSME1;PSME2;CTNNB1 |
| 64 | CDT1 association with the CDC6:ORC:origin complex Homo sapiens R-HSA-68827 | 2.71E-09 | 6.01E-08 | 4.941912 | 97.491598 | PSMD12;PSMD11;PSMD13;GMNN;PSMB10;PSMA7;PSMD8;PSMB6;PSMD6;PSMB4;PSMD7;PSMB5;PSMD4;UBB;PSMB2;PSMB3;PSMD2;PSMD3;UBC;PSMB1;PSMD1;PSMF1;PSMB8;PSMB9;PSMA5;PSMA6;PSMA3;PSMC5;PSMA1;PSMA2;PSMC4;PSMC2;PSME1;PSME2 |
| 65 | Hedgehog 'on' state Homo sapiens R-HSA-5632684 | 3.01E-09 | 6.55E-08 | 3.747988 | 73.544449 | PSMD12;PSMD11;PSMD13;CUL3;ARRB1;CDC73;PSMB10;PSMA7;PSMD8;PSMB6;SHH;PSMD6;PSMB4;PSMD7;PSMB5;PSMD4;UBB;PSMB2;PSMB3;KIF3A;PSMD2;PSMD3;UBC;PSMB1;SPOP;PSMD1;PSMF1;EVC2;CSNK1A1;PSMB8;PSMB9;PSMA5;PSMA6;PSMA3;PSMC5;ITCH;PSMA1;PSMA2;PSMC4;PSMC2;ULK3;NUMB;PSME1;PSME2 |
| 66 | APC/C:Cdc20 mediated degradation of Securin Homo sapiens R-HSA-174154 | 3.04E-09 | 6.55E-08 | 4.452299 | 87.313695 | PSMD12;PSMD11;ANAPC16;PSMD13;PSMB10;PSMA7;ANAPC11;PSMD8;PSMB6;PSMD6;PSMB4;PSMD7;PSMB5;PSMD4;UBB;PSMB2;PSMB3;PSMD2;PSMD3;UBC;PSMB1;PSMD1;PSMF1;UBE2E1;PSMB8;PSMB9;PSMA5;PSMA6;PSMA3;PSMC5;PSMA1;PSMA2;PSMC4;PSMC2;PSME1;PSME2;ANAPC5 |

| | | | | | | |
|---|---|---|---|---|---|---|
| 67 | Organelle biogenesis and maintenance Homo sapiens R-HSA-1852241 | 3.95E-09 | 8.38E-08 | 2.020388 | 39.094973 | ATF2;TRAF3IP1;KIFAP3;CDK5RAP2;MRPS18B;TUBA4A;TUFM;IFT88;TBL1XR1;AKAP9;TFAM;DAP3;MRPL18;ARL3;MRPL19;MRPL14;MRPL10;DYNC2LI1;KIF3A;TTC21B;CEP70;CEP78;BBS2;ESRRA;RAB3IP;MRPL27;BBIP1;NR1D1;IFT20;AHI1;WDR60;DYNLRB1;CEP89;CEP57;YWHAE;ARF4;GFM1;MRPS14;GFM2;CHD9;MRPL38;MRPL39;MRPS10;PRKAG2;TFB1M;MRPL34;PKD1;MRPL32;MRPL33;ACTR1A;PCM1;MRPL2;MRPL40;CNTRL;TNPO1;IFT57;CC2D2A;CEP290;BBS4;RAB8A;NCOA1;OFD1;CCT3;IQCB1;NCOA2;CCT2;TSFM;MRPS26;HSP90AA1;MRPS24;EXOC7;MRPS25;TUBB;MRPS22;HAUS4;HAUS3;TUBG1;MRPS7;WDR34;MRPS6;MRPS5;MRPL43;HAUS1;MRPS9;NCOR1;CREB1;TCP1;EXOC4;EXOC6;EXOC5;MAPRE1;MRRF;EXOC2;PAFAH1B1;SDCCAG8;DYNC1I2;PRKAA2;MRPS35;DCTN2;MRPS36;SSNA1;MRPS34;MTIF2;MTIF3;HDAC6;IFT74;ARL13B;TGS1;PDE6D;CCT8;CCT5;CCT4;PTCD3;DYNC1H1;CEP152;TUBB4B;CENPJ;TRIP11;CLUAP1 |
| 68 | APC:Cdc20 mediated degradation of cell cycle proteins prior to satisfation of the cell cycle checkpoint Homo sapiens R-HSA-179419 | 4.35E-09 | 8.97E-08 | 4.124913 | 79.413894 | PSMD12;PSMD11;ANAPC16;PSMD13;PSMB10;PSMA7;ANAPC11;PSMD8;PSMB6;PSMD6;PSMB4;PSMD7;PSMB5;PSMD4;UBB;PSMB2;PSMB3;PSMD2;PSMD3;UBC;PSMB1;PSMD1;PSMF1;UBE2E1;PSMB8;PSMB9;PSMA5;PSMA6;CCNA1;PSMA3;PSMC5;PSMA1;PSMA2;PSMC4;PSMC2;PSME1;PSME2;ANAPC5;MAD2L1 |
| 69 | Removal of licensing factors from origins Homo sapiens R-HSA-69300 | 4.35E-09 | 8.97E-08 | 4.124913 | 79.413894 | RB1;PSMD12;CDKN1B;PSMD11;PSMD13;GMNN;PSMB10;PSMA7;PSMD8;PSMB6;PSMD6;FZR1;PSMB4;PSMD7;PSMB5;PSMD4;UBB;PSMB2;PSMB3;PSMD2;PSMD3;UBC;PSMB1;PSMD1;PSMF1;PSMB8;PSMB9;PSMA5;PSMA6;CCNA1;PSMA3;PSMC5;PSMA1;PSMA2;PSMC4;PSMC2;MCM3;PSME1;PSME2 |
| 70 | TCR signaling Homo sapiens R-HSA-202403 | 5.01E-09 | 1.02E-07 | 3.052199 | 58.330122 | UBE2D2;PTPRJ;PSMD8;PSMD6;PSMD7;PSMD4;PSMD2;PSMD3;PSMD1;HLA-DPA1;FBXW11;ENAH;PSMA5;PSMA6;PSMA3;PSMA1;PSMA2;PSME1;PSME2;EVL;UBE2V1;HLA-DQB1;PSMD12;PSMD11;PSMD13;CUL1;BCL10;RELA;PSMB10;PSMA7;MALT1;PSMB6;PSMB4;PSMB5;UBB;PSMB2;PSMB3;UBC;PSMB1;CSK;PLCG1;PSMF1;HLA-DQA2;PAK2;VASP;NFKB1;PSMB8;PSMB9;NFKBIA;PSMC5;PSMC4;PSMC2;HLA-DPB1;HLA-DRA;HLA-DRB1 |

| | | | | | | |
|---|---|---|---|---|---|---|
| 71 | Regulation of RAS by GAPs Homo sapiens R-HSA-5658442 | 5.33E-09 | 1.07E-07 | 4.303611 | 81.9837 | PSMD12;PSMD11;PSMD13;CUL3;PSMB10;PSMA7;PSMD8;PSMB6;PSMD6;PSMB4;PSMD7;PSMB5;PSMD4;UBB;PSMB2;PSMB3;PSMD2;PSMD3;UBC;PSMB1;PSMD1;PSMF1;DAB2IP;PSMB8;PSMB9;PSMA5;PSMA6;PSMA3;PSMC5;PSMA1;PSMA2;PSMC4;RASA2;PSMC2;PSME1;NF1;PSME2 |
| 72 | EPH-Ephrin signaling Homo sapiens R-HSA-2682334 | 5.98E-09 | 1.18E-07 | 3.420452 | 64.764497 | EPHB6;ROCK1;ROCK2;SRC;ARPC1B;SDC2;ARHGEF28;ARPC1A;CLTC;ITSN1;CLTB;AP2A1;WASL;SHB;EFNA4;MYL12A;ACTG1;MYL12B;APH1A;EFNB2;CDC42;SDCBP;APH1B;CFL1;MYH14;RAC1;PAK2;MYH10;AP2M1;ACTR3;PSENEN;VAV3;ACTR2;HSP90AA1;YES1;LIMK2;ADAM10;ARPC4;RHOA;PTK2;EFNA1;MYL6;ARPC2;ARPC3;MYH9;EPHA1;EPHA2 |
| 73 | Switching of origins to a post-replicative state Homo sapiens R-HSA-69052 | 6.32E-09 | 1.21E-07 | 4.144078 | 78.238167 | RB1;PSMD12;CDKN1B;PSMD11;PSMD13;PSMB10;PSMA7;PSMD8;PSMB6;PSMD6;FZR1;PSMB4;PSMD7;PSMB5;PSMD4;UBB;PSMB2;PSMB3;PSMD2;PSMD3;UBC;PSMB1;PSMD1;PSMF1;PSMB8;PSMB9;PSMA5;PSMA6;CCNA1;PSMA3;PSMC5;PSMA1;PSMA2;PSMC4;PSMC2;MCM3;PSME1;PSME2 |
| 74 | Orc1 removal from chromatin Homo sapiens R-HSA-68949 | 6.32E-09 | 1.21E-07 | 4.144078 | 78.238167 | RB1;PSMD12;CDKN1B;PSMD11;PSMD13;PSMB10;PSMA7;PSMD8;PSMB6;PSMD6;FZR1;PSMB4;PSMD7;PSMB5;PSMD4;UBB;PSMB2;PSMB3;PSMD2;PSMD3;UBC;PSMB1;PSMD1;PSMF1;PSMB8;PSMB9;PSMA5;PSMA6;CCNA1;PSMA3;PSMC5;PSMA1;PSMA2;PSMC4;PSMC2;MCM3;PSME1;PSME2 |
| 75 | Metabolism of polyamines Homo sapiens R-HSA-351202 | 7.05E-09 | 1.34E-07 | 3.66198 | 68.738508 | PSMD12;PSMD11;PSMD13;APIP;MRI1;SAT1;OAZ1;PSMB10;PSMA7;PSMD8;PSMB6;PSMD6;PSMB4;PSMD7;PSMB5;PSMD4;PSMB2;PSMB3;PSMD2;PSMD3;PSMB1;PSMD1;PSMF1;NQO1;ARG2;GOT1;SLC6A12;NMRAL1;PSMB8;PSMB9;PSMA5;PSMA6;MTAP;PSMA3;PSMC5;PSMA1;PSMA2;PSMC4;ADI1;PSMC2;SMS;PSME1;PSME2 |

| | | | | | | |
|---|---|---|---|---|---|---|
| 76 | Class I MHC mediated antigen processing & presentation Homo sapiens R-HSA-983169 | 7.65E-09 | 1.42E-07 | 2.038068 | 38.088084 | UBE3C;UBE2D2;LTN1;KEAP1;UBE3A;UBE3B;UBE2Z;HERC5;PSMD8;RNF115;HERC3;PSMD6;RNF114;PSMD7;HERC2;PSMD4;PSMD2;RNF19A;PSMD3;FBXO3;PSMD1;B2M;PDIA3;FBXW11;FBXO2;UBE2E3;UBE2E1;TAP2;HLA-C;UBE4A;TAP1;HLA-F;HLA-E;RNF126;PSME1;PSME2;ANAPC5;RBCK1;UBE2V1;PSMD12;PSMD11;CUL5;SAR1B;PSMD13;UBA6;CUL3;CUL1;UBR4;NEDD4L;UBR2;UBR1;FBXO44;ANAPC11;RNF213;UBB;UBC;PSMF1;UBE2H;HSPA5;ZBTB16;UBAC1;MKRN1;UBA3;TPP2;CALR;UBE2K;DZIP3;UBE2M;LRSAM1;RNF14;CTSS;UFL1;SEC61A1;CTSL;UBE2Q2;ARIH2;CD36;PJA2;TAPBP;PSMA5;PSMA6;PSMA3;PSMA1;PSMA2;ASB13;RNF6;PSMA7;PSMB10;PSMB6;ZNRF1;PSMB4;FZR1;ZNRF2;PSMB5;HECTD1;PSMB2;PSMB3;PSMB1;RBBP6;ERAP2;MIB2;HUWE1;WWP1;UBE2G1;PSMB8;PSMB9;PSMC5;ITCH;PSMC4;PSMC2;LNX1;STUB1 |
| 77 | G1/S DNA Damage Checkpoints Homo sapiens R-HSA-69615 | 7.71E-09 | 1.42E-07 | 4.330991 | 80.906124 | PSMD12;CDKN1B;PSMD11;PSMD13;PSMB10;PSMA7;PSMD8;PSMB6;PSMD6;PSMB4;PSMD7;PSMB5;PSMD4;UBB;PSMB2;PSMB3;PSMD2;PSMD3;UBC;PSMB1;PSMD1;PSMF1;PSMB8;PSMB9;PSMA5;PSMA6;CCNA1;PSMA3;PSMC5;PSMA1;PSMA2;PSMC4;PSMC2;PSME1;PSME2;ATM |
| 78 | Regulation of APC/C activators between G1/S and early anaphase Homo sapiens R-HSA-176408 | 9.34E-09 | 1.70E-07 | 3.766344 | 69.637687 | PSMD12;PSMD11;ANAPC16;PSMD13;CUL1;PSMB10;PSMA7;ANAPC11;PSMD8;PSMB6;PSMD6;FZR1;PSMB4;PSMD7;PSMB5;PSMD4;UBB;PSMB2;PSMB3;PSMD2;PSMD3;UBC;PSMB1;PSMD1;PSMF1;UBE2E1;PSMB8;PSMB9;PSMA5;PSMA6;CCNA1;PSMA3;PSMC5;PSMA1;PSMA2;PSMC4;PSMC2;PSME1;PSME2;ANAPC5;MAD2L1 |
| 79 | Downstream TCR signaling Homo sapiens R-HSA-202424 | 9.58E-09 | 1.72E-07 | 3.287849 | 60.706396 | PSMD12;PSMD11;PSMD13;UBE2D2;CUL1;BCL10;RELA;PSMB10;PSMA7;MALT1;PSMD8;PSMB6;PSMD6;PSMB4;PSMD7;PSMB5;PSMD4;UBB;PSMB2;PSMB3;PSMD2;PSMD3;UBC;PSMB1;PSMD1;PSMF1;HLA-DQA2;HLA-DPA1;FBXW11;NFKB1;PSMB8;PSMB9;NFKBIA;PSMA5;PSMA6;PSMA3;PSMC5;PSMA1;PSMA2;PSMC4;PSMC2;PSME1;HLA-DPB1;PSME2;HLA-DRA;UBE2V1;HLA-DRB1;HLA-DQB1 |

| | | | | | | |
|---|---|---|---|---|---|---|
| 80 | APC/C:Cdh1 mediated degradation of Cdc20 and other APC/C:Cdh1 targeted proteins in late mitosis/early G1 Homo sapiens R-HSA-174178 | 1.07E-08 | 1.90E-07 | 4.018241 | 73.758596 | PSMD12;PSMD11;ANAPC16;PSMD13;PSMB10;PSMA7;ANAPC11;PSMD8;PSMB6;PSMD6;FZR1;PSMB4;PSMD7;PSMB5;PSMD4;UBB;PSMB2;PSMB3;PSMD2;PSMD3;UBC;PSMB1;PSMD1;PSMF1;UBE2E1;PSMB8;PSMB9;PSMA5;PSMA6;PSMA3;PSMC5;PSMA1;PSMA2;PSMC4;PSMC2;PSME1;PSME2;ANAPC5 |
| 81 | Asymmetric localization of PCP proteins Homo sapiens R-HSA-4608870 | 1.11E-08 | 1.96E-07 | 4.360367 | 79.846734 | PSMD12;PSMD11;PSMD13;PSMB10;PSMA7;PSMD8;PSMB6;PSMD6;PSMB4;PSMD7;PSMB5;PSMD4;UBB;PSMB2;PSMB3;PSMD2;PSMD3;UBC;PSMB1;PSMD1;PSMF1;FZD5;WNT5A;PSMB8;PSMB9;PSMA5;PSMA6;PSMA3;PSMC5;PSMA1;PSMA2;PSMC4;PSMC2;PSME1;PSME2 |
| 82 | APC/C:Cdc20 mediated degradation of mitotic proteins Homo sapiens R-HSA-176409 | 1.22E-08 | 2.11E-07 | 3.888701 | 70.871392 | PSMD12;PSMD11;ANAPC16;PSMD13;PSMB10;PSMA7;ANAPC11;PSMD8;PSMB6;PSMD6;PSMB4;PSMD7;PSMB5;PSMD4;UBB;PSMB2;PSMB3;PSMD2;PSMD3;UBC;PSMB1;PSMD1;PSMF1;UBE2E1;PSMB8;PSMB9;PSMA5;PSMA6;CCNA1;PSMA3;PSMC5;PSMA1;PSMA2;PSMC4;PSMC2;PSME1;PSME2;ANAPC5;MAD2L1 |
| 83 | PCP/CE pathway Homo sapiens R-HSA-4086400 | 1.27E-08 | 2.18E-07 | 3.416188 | 62.098607 | PSMD12;PSMD11;PSMD13;CLTC;CLTB;AP2A1;PSMB10;PSMA7;PSMD8;PSMB6;PSMD6;PSMB4;PSMD7;PSMB5;PSMD4;UBB;PSMB2;PSMB3;PSMD2;PSMD3;UBC;PSMB1;PSMD1;DVL3;RAC1;PSMF1;AP2M1;FZD5;RYK;WNT5A;FZD6;RHOA;PSMB8;PSMB9;PSMA5;PSMA6;PSMA3;PSMC5;PSMA1;DAAM1;PSMA2;PSMC4;PSMC2;PSME1;PSME2 |

| | | | | | | |
|---|---|---|---|---|---|---|
| 84 | Post-translational protein modification Homo sapiens R-HSA-597592 | 1.93E-08 | 3.26E-07 | 1.721573 | 30.581842 | DPAGT1;GALNT11;PGAP1;CMAS;KDELR1;SMC6;SMC3;GOLGA2;POFUT2;C1GALT1C1;HERC2;TFG;THSD7A;NUP214;PDIA3;NSF;ARSA;ALG8;COG6;ALG6;ALG5;COG2;ALG2;ALG14;COG1;MITF;MOGS;ANK2;ALG12;ANK3;ALG1;PIAS1;DPM2;DYNC1LI2;DPM3;TBC1D20;KDELR2;NUP98;GNE;COPB2;NUP205;SPTBN5;COPA;TMED10;RPN2;SAR1B;COPB1;RPN1;SEL1L;XPC;AAAS;DYNC2LI1;SCFD1;FUT8;POM121C;MAN2A2;MAN2A1;RAD21;ADAMTS18;TPR;ARSK;NEU1;MLEC;ST3GAL6;TP53BP1;ARSB;SPTBN1;MPDU1;SPTBN2;EDEM3;PHC2;ICMT;TRAPPC1;UBE2I;FUCA1;TRAPPC4;EDEM2;GFPT1;EEF2;STAG1;HNRNPK;STAG2;TMEM115;GOLGB1;SP3;UBA2;CALR;MUC20;COPE;PIGS;ARF4;TOP2B;B4GALT3;MCFD2;SERPINA1;B4GALT1;PIGP;MIA3;PIGV;GMPPB;ADAMTS1;SUMO3;PPP6R3;SUMO2;PGM3;TMED3;TMED2;MAN1A1;MUC15;TMED7;NANS;SPTAN1;TMED9;RING1;DHPS;FN3KRP;COPZ2;SMC1A;ARFGAP3;POMGNT1;COPZ1;SUMF2;DDOST;SUMF1;NSMCE4A;DAD1;NUP50;PIGK;B3GNT3;B3GNT2;PIGL;TOP1;PIGG;PIGF;ARF5;PIGH;DYNC1I2;SP100;STX17;DCTN2;USO1;DERL2;FURIN;NAPG;LMAN1;GANAB;MUC1;OS9;LMAN2;SLC17A5;MAN1C1;STX5;CD59;UGGT2;CD55;RANBP2;GALNT7;SLC35A1;DYNC1H1;GALNT4;GALNT3;TRAPPC10;GALNT1;C1GALT1;SYVN1;ARCN1;NAGK;DOLPP1;MGAT4B;COPG2;FOLR1 |
| 85 | Activation of APC/C and APC/C:Cdc20 mediated degradation of mitotic proteins Homo sapiens R-HSA-176814 | 1.99E-08 | 3.29E-07 | 3.780438 | 67.040926 | PSMD12;PSMD11;ANAPC16;PSMD13;PSMB10;PSMA7;ANAPC11;PSMD8;PSMB6;PSMD6;PSMB4;PSMD7;PSMB5;PSMD4;UBB;PSMB2;PSMB3;PSMD2;PSMD3;UBC;PSMB1;PSMD1;PSMF1;UBE2E1;PSMB8;PSMB9;PSMA5;PSMA6;CCNA1;PSMA3;PSMC5;PSMA1;PSMA2;PSMC4;PSMC2;PSME1;PSME2;ANAPC5;MAD2L1 |
| 86 | Regulation of DNA replication Homo sapiens R-HSA-69304 | 1.99E-08 | 3.29E-07 | 3.780438 | 67.040926 | RB1;PSMD12;CDKN1B;PSMD11;PSMD13;GMNN;PSMB10;PSMA7;PSMD8;PSMB6;PSMD6;FZR1;PSMB4;PSMD7;PSMB5;PSMD4;UBB;PSMB2;PSMB3;PSMD2;PSMD3;UBC;PSMB1;PSMD1;PSMF1;PSMB8;PSMB9;PSMA5;PSMA6;CCNA1;PSMA3;PSMC5;PSMA1;PSMA2;PSMC4;PSMC2;MCM3;PSME1;PSME2 |

| | | | | | | |
|---|---|---|---|---|---|---|
| 87 | TNFR2 non-canonical NF-kB pathway Homo sapiens R-HSA-5668541 | 2.04E-08 | 3.33E-07 | 3.278405 | 58.057102 | CD40;PSMD12;EDA;PSMD11;PSMD13;CUL1;TNFRSF11B;PSMB10;PSMA7;PSMD8;PSMB6;PSMD6;PSMB4;PSMD7;PSMB5;PSMD4;UBB;PSMB2;PSMB3;PSMD2;PSMD3;UBC;PSMB1;PSMD1;TNFRSF14;PSMF1;TNFRSF12A;FBXW11;TNFSF12;PSMB8;TNFRSF1A;PSMB9;PSMA5;PSMA6;PSMA3;PSMC5;PSMA1;PSMA2;PSMC4;PSMC2;PSME1;UBA3;PSME2;LTBR;BIRC2;UBE2M |
| 88 | Host Interactions of HIV factors Homo sapiens R-HSA-162909 | 2.92E-08 | 4.71E-07 | 2.76743 | 48.015779 | PSIP1;AP2A1;PSMD8;PSMD6;PSMD7;XPO1;PSMD4;AP1G1;PSMD2;PSMD3;PSMD1;RAC1;B2M;AP2M1;KPNA1;NUP214;PSMA5;PSMA6;PSMA3;PSMA1;PSMA2;NUP50;PSME1;PSME2;NUP98;PPIA;SLC25A4;NUP205;PSMD12;PSMD11;CUL5;PSMD13;AAAS;PSMB10;PSMA7;PSMB6;PSMB4;PSMB5;UBB;POM121C;PSMB2;PSMB3;TPR;UBC;PSMB1;PSMF1;PAK2;AP1M2;RANBP2;NPM1;PSMB8;PSMB9;CDK9;PSMC5;PSMC4;PSMC2;RAN |
| 89 | C-type lectin receptors (CLRs) Homo sapiens R-HSA-5621481 | 3.07E-08 | 4.90E-07 | 2.826858 | 48.905169 | AHCYL1;UBE2D2;PYCARD;PSMD8;PSMD6;PSMD7;RPS6KA5;PSMD4;PSMD2;PSMD3;PSMD1;EP300;FBXW11;PRKCD;PSMA5;PSMA6;PSMA3;PSMA1;PSMA2;PSME1;PSME2;UBE2V1;RAF1;PSMD12;PSMD11;PSMD13;SRC;CUL1;ITPR2;ITPR3;BCL10;RELA;PSMB10;PSMA7;MALT1;PSMB6;PSMB4;PSMB5;UBB;PSMB2;PSMB3;UBC;PSMB1;PSMF1;PAK2;NFKB1;PSMB8;PSMB9;NFKBIA;PSMC5;PSMC4;PSMC2;UBA3;CALM1;UBE2M |
| 90 | Hedgehog 'off' state Homo sapiens R-HSA-5610787 | 3.14E-08 | 4.96E-07 | 3.156525 | 54.533473 | GSK3B;PSMD12;PSMD11;INTU;PSMD13;CUL1;PSMB10;ADCY6;PSMA7;PSMD8;PSMB6;PSMD6;PSMB4;PSMD7;PSMB5;PSMD4;UBB;PSMB2;KIF3A;PSMB3;PSMD2;TTC21B;PRKAR2A;PSMD3;UBC;PSMB1;PSMD1;PSMF1;IFT57;OFD1;CSNK1A1;PSMB8;PSMB9;PSMA5;PSMA6;IFT88;PSMA3;PSMC5;ITCH;ADCY9;PSMA1;PSMA2;PSMC4;PSMC2;PSME1;NUMB;PSME2 |

| | | | | | | |
|---|---|---|---|---|---|---|
| 91 | Antigen processing: Ubiquitination & Proteasome degradation Homo sapiens R-HSA-983168 | 6.46E-08 | 1.01E-06 | 2.052188 | 33.972571 | LRSAM1;UBE3C;RNF14;UBE2D2;LTN1;KEAP1;UBE3A;UBE3B;UBE2Z;HERC5;PSMD8;UFL1;RNF115;HERC3;PSMD6;RNF114;PSMD7;HERC2;PSMD4;UBE2Q2;PSMD2;RNF19A;PSMD3;ARIH2;FBXO3;PSMD1;FBXW11;FBXO2;UBE2E3;UBE2E1;UBE4A;PJA2;PSMA5;PSMA6;RNF126;PSMA3;PSMA1;PSMA2;PSME1;PSME2;ANAPC5;RBCK1;UBE2V1;PSMD12;PSMD11;CUL5;PSMD13;UBA6;CUL3;CUL1;UBR4;ASB13;NEDD4L;UBR2;RNF6;UBR1;FBXO44;PSMA7;PSMB10;ANAPC11;PSMB6;ZNRF1;PSMB4;FZR1;ZNRF2;RNF213;PSMB5;HECTD1;PSMB2;UBB;PSMB3;PSMB1;UBC;RBBP6;PSMF1;UBE2H;ZBTB16;MIB2;HUWE1;WWP1;UBE2G1;UBAC1;PSMB8;PSMB9;PSMC5;ITCH;PSMC4;PSMC2;MKRN1;UBA3;LNX1;TPP2;STUB1;UBE2K;DZIP3;UBE2M |
| 92 | MAPK6/MAPK4 signaling Homo sapiens R-HSA-5687128 | 6.52E-08 | 1.01E-06 | 3.199959 | 52.946943 | PSMD12;PSMD11;PSMD13;HSPB1;PSMB10;PSMA7;CDC14B;PSMD8;CDC42;PSMB6;CCND3;PSMD6;PSMB4;PSMD7;XPO1;PSMB5;PSMD4;UBB;PSMB2;PSMB3;PSMD2;PSMD3;UBC;PSMB1;PSMD1;RAC1;PSMF1;PAK2;NCOA3;PSMB8;PSMB9;PSMA5;PSMA6;PSMA3;PSMC5;PSMA1;AGO3;PSMA2;PSMC4;PSMC2;CDC42EP2;PSME1;PSME2;TNRC6A |
| 93 | Assembly of the pre-replicative complex Homo sapiens R-HSA-68867 | 9.03E-08 | 1.38E-06 | 3.814336 | 61.869946 | PSMD12;PSMD11;PSMD13;GMNN;PSMB10;PSMA7;PSMD8;PSMB6;PSMD6;PSMB4;PSMD7;PSMB5;PSMD4;UBB;PSMB2;PSMB3;PSMD2;PSMD3;UBC;PSMB1;PSMD1;PSMF1;PSMB8;PSMB9;PSMA5;PSMA6;PSMA3;PSMC5;PSMA1;PSMA2;PSMC4;PSMC2;MCM3;PSME1;PSME2 |

| | | | | | | |
|---|---|---|---|---|---|---|
| 94 | Axon guidance Homo sapiens R-HSA-422475 | 1.21E-07 | 1.81E-06 | 1.67419 | 26.672278 | EPHB6;ITSN1;ITGA2B;PEBP1;WDR83;IRS2;ARHGAP35;ACTG1;PSMD8;PSMD6;PSMD7;RPS6KA5;PSMD4;DPYSL2;PSMD2;PSMD3;CFL1;PSMD1;NRCAM;AP2M1;MAP2K1;MAP2K2;CSNK2A1;CSNK2A2;ADAM10;ANK2;ANK3;MYL6;COL4A4;AKAP9;PSME1;PSME2;TLN1;EPHA1;RAF1;VCL;EPHA2;PFN2;SHC2;SPTBN5;PSMD12;PSMD11;PSMD13;CUL3;SDC2;ARHGEF28;IQGAP1;PHB;EFNA4;MYL12A;MYL12B;RAP1B;APH1A;EFNB2;SDCBP;APH1B;RRAS;UBB;UBC;MYH14;PLCG1;PSMF1;SRGAP3;SRGAP1;MYH10;SPTBN1;SPTBN2;VASP;PRNP;HSPA8;YES1;RDX;MSN;BRAF;ARPC4;PTK2;EFNA1;DLG1;ARPC2;CDK5;DLG3;ARPC3;PTPRA;NF1;GRB2;CALM1;CD24;MAP3K11;ITGB1;GSK3B;HSP90AB1;YWHAB;ARPC1B;RASGRF2;ARPC1A;CLTC;CLTB;ARRB1;AP2A1;WASL;LAMC1;SHB;RASGEF1A;RAC1;JAK2;NEO1;SPTAN1;JAK1;ACTR3;PSENEN;VAV3;ACTR2;HSP90AA1;ARHGEF12;PPP2R5B;RHOA;DNM2;PSMA5;ENAH;PSMA6;PSMA3;CREB1;PSMA1;PSMA2;IL3RA;KIT;RASA2;NUMB;MYH9;EVL;SOS1;MET;ITGA9;ROCK1;ROCK2;SRC;PSMA7;EGFR;PSMB10;PSMB6;CDC42;PSMB4;PSMB5;PSMB2;ERBB3;PSMB3;PSMB1;ERBB2;MAPK1;CSK;SLIT3;PAK2;MAPK3;SEMA4D;MYO10;LIMK2;DAB2IP;RANBP9;PTPN11;LAMB1;PSMB8;PSMB9;PSMC5;PSMC4;PSMC2;AGRN |
| 95 | S Phase Homo sapiens R-HSA-69242 | 1.21E-07 | 1.81E-06 | 2.695204 | 42.936825 | RB1;CDKN1B;SMC3;CKS1B;PSMD8;PSMD6;PSMD7;PSMD4;PSMD2;PSMD3;PSMD1;RFC1;SMC1A;CDC25B;PSMA5;CCNA1;PSMA6;PSMA3;PSMA1;PSMA2;PSME1;MCM3;PSME2;PSMD12;PSMD11;MAX;PSMD13;CUL1;PDS5B;PDS5A;PSMB10;PSMA7;PSMB6;POLD4;FZR1;PSMB4;PSMB5;UBB;PSMB2;PSMB3;RAD21;UBC;PSMB1;PSMF1;RPA2;PSMB8;PSMB9;STAG1;CDK7;PSMC5;STAG2;PSMC4;RPA3;PSMC2 |
| 96 | APC/C-mediated degradation of cell cycle proteins Homo sapiens R-HSA-174143 | 1.35E-07 | 1.98E-06 | 3.251491 | 51.437909 | PSMD12;PSMD11;ANAPC16;PSMD13;CUL1;PSMB10;PSMA7;ANAPC11;PSMD8;PSMB6;PSMD6;FZR1;PSMB4;PSMD7;PSMB5;PSMD4;UBB;PSMB2;PSMB3;PSMD2;PSMD3;UBC;PSMB1;PSMD1;PSMF1;UBE2E1;PSMB8;PSMB9;PSMA5;PSMA6;CCNA1;PSMA3;PSMC5;PSMA1;PSMA2;PSMC4;PSMC2;PSME1;PSME2;ANAPC5;MAD2L1 |

| | | | | | | |
|---|---|---|---|---|---|---|
| 97 | Regulation of mitotic cell cycle Homo sapiens R-HSA-453276 | 1.35E-07 | 1.98E-06 | 3.251491 | 51.437909 | PSMD12;PSMD11;ANAPC16;PSMD13;CUL1;PSMB10;PSMA7;ANAPC11;PSMD8;PSMB6;PSMD6;FZR1;PSMB4;PSMD7;PSMB5;PSMD4;UBB;PSMB2;PSMB3;PSMD2;PSMD3;UBC;PSMB1;PSMD1;PSMF1;UBE2E1;PSMB8;PSMB9;PSMA5;PSMA6;CCNA1;PSMA3;PSMC5;PSMA1;PSMA2;PSMC4;PSMC2;PSME1;PSME2;ANAPC5;MAD2L1 |
| 98 | Ribosomal scanning and start codon recognition Homo sapiens R-HSA-72702 | 1.47E-07 | 2.13E-06 | 4.155914 | 65.383559 | EIF4A1;RPS4Y1;RPS15;RPS19;EIF4H;RPS10;EIF4B;RPS9;RPS7;RPS8;RPS5;EIF2S2;EIF2S1;RPS25;RPS28;EIF3M;RPS27;EIF5;EIF3K;RPS29;EIF3L;EIF3I;EIF3J;EIF3G;EIF3H;EIF3C;RPS21;EIF3D;EIF3A;RPS24;EIF4G1 |
| 99 | ER to Golgi Anterograde Transport Homo sapiens R-HSA-199977 | 1.52E-07 | 2.19E-06 | 2.609027 | 40.952105 | ARF4;MCFD2;SERPINA1;KDELR1;MIA3;GOLGA2;TFG;PPP6R3;TMED3;TMED2;TMED7;SPTAN1;NSF;TMED9;COG6;COG2;COG1;COPZ2;ANK2;ANK3;ARFGAP3;COPZ1;DYNC1LI2;TBC1D20;KDELR2;ARF5;COPB2;DYNC1I2;COPA;SPTBN5;TMED10;STX17;DCTN2;SAR1B;COPB1;USO1;NAPG;DYNC2LI1;LMAN1;SCFD1;LMAN2;STX5;CD59;SPTBN1;CD55;SPTBN2;DYNC1H1;TRAPPC1;TRAPPC4;TRAPPC10;ARCN1;TMEM115;GOLGB1;COPG2;FOLR1;COPE |
| 100 | Citric acid cycle (TCA cycle) Homo sapiens R-HSA-71403 | 2.65E-07 | 3.76E-06 | 13.04264 | 197.53436 | FH;MDH2;IDH3G;DLST;SDHD;SDHA;SDHB;SUCLA2;NNT;IDH3B;OGDH;SUCLG2;SUCLG1;ACO2;DLD |
| 101 | Transport to the Golgi and subsequent modification Homo sapiens R-HSA-948021 | 2.78E-07 | 3.90E-06 | 2.342931 | 35.366389 | B4GALT3;ARF4;MCFD2;B4GALT1;SERPINA1;KDELR1;MIA3;GOLGA2;TFG;PPP6R3;TMED3;TMED2;MAN1A1;TMED7;SPTAN1;NSF;TMED9;COG6;COG2;COG1;COPZ2;ANK2;ANK3;ARFGAP3;COPZ1;DYNC1LI2;TBC1D20;KDELR2;ARF5;COPB2;DYNC1I2;COPA;SPTBN5;TMED10;STX17;DCTN2;SAR1B;COPB1;USO1;NAPG;DYNC2LI1;LMAN1;FUT8;SCFD1;MAN2A2;MAN2A1;LMAN2;MAN1C1;STX5;CD59;SPTBN1;CD55;SPTBN2;DYNC1H1;TRAPPC1;TRAPPC4;FUCA1;TRAPPC10;ARCN1;TMEM115;GOLGB1;MGAT4B;COPG2;FOLR1;COPE |

| | | | | | | |
|---|---|---|---|---|---|---|
| 102 | Cell Cycle Homo sapiens R-HSA-1640170 | 2.79E-07 | 3.90E-06 | 1.615114 | 24.37265 | RB1;ANKLE2;GMNN;PPP2R2A;SMC3;SMC2;PSMD8;GOLGA2;PSMD6;CCND3;PSMD7;HERC2;PSMD4;PSMD2;PSMD3;RUVBL1;PSMD1;EP300;NBN;HIST1H2AC;CDK5RAP2;TMPO;NUP214;RAB2A;CSNK2A1;FBXW11;CSNK2A2;UBE2E1;LEMD2;TUBA4A;CDC25B;CLIP1;DKC1;AKAP9;PSME1;PSME2;ANAPC5;NUP98;OPTN;BLZF1;NUP205;PSMD12;SET;PSMD11;ANAPC16;MAX;TUBGCP2;PSMD13;CUL1;PDS5B;AAAS;PDS5A;PMF1;ANAPC11;SKA2;POM121C;UBB;RAD21;CEP70;TPR;UBC;PSMF1;TP53BP1;ATRIP;CEP78;UBE2I;H3F3A;NEK7;SMARCA5;HSPA2;CDK7;STAG1;RPS27;STAG2;RAD17;ATM;ATR;CEP57;YWHAE;AHCTF1;CDKN1B;HSP90AB1;NUMA1;YWHAB;RSF1;SYNE2;CKS1B;BABAM1;NIPBL;ACTR1A;PCM1;XPO1;UIMC1;CNTRL;TK2;TOPBP1;CEP290;RAB8A;YWHAH;OFD1;HSP90AA1;SUN1;PPP1R12A;RFC1;H2AFZ;TUBB;TOP3A;HAUS4;PPP2R5B;HAUS3;KIF23;TERF1;SMC1A;TUBG1;HAUS1;PSMA5;PSMA6;CCNA1;PSMA3;PSMA1;KIF2A;PSMA2;GORASP2;NUP50;MCM3;TERF2IP;MAPRE1;PAFAH1B1;SDCCAG8;MCPH1;DYNC1I2;DCTN2;HDAC1;SSNA1;USO1;RNF8;HDAC8;PSMA7;PSMB10;CENPC;PSMB6;PSMC3IP;POLD4;PSMB4;FZR1;PSMB5;PSMB2;RBBP4;PSMB3;PSMB1;LMNA;MAPK1;RBBP7;MAPK3;CENPT;RANBP2;DYNC1H1;NPM1;CDK11A;CEP152;RPA2;TUBB4B;MLH1;CDK11B;PSMB8;MLH3;PSMB9;PSMC5[illegible] |
| 103 | Apoptotic execution phase Homo sapiens R-HSA-75153 | 4.52E-07 | 6.24E-06 | 4.241305 | 61.965878 | ROCK1;HMGB1;ADD1;CASP7;FNTA;CDH1;LMNA;HIST1H1E;DNM1L;SPTAN1;PAK2;HIST1H1C;KPNA1;DSP;BCAP31;DBNL;GSN;PRKCD;PTK2;TJP1;OCLN;H1F0;APC;STK24;ACIN1;CTNNB1;BIRC2;TJP2 |
| 104 | Cytosolic tRNA aminoacylation Homo sapiens R-HSA-379716 | 5.76E-07 | 7.88E-06 | 8.448815 | 121.38375 | YARS;CARS;DARS;VARS;SARS;EPRS;AIMP1;NARS;AIMP2;KARS;QARS;LARS;RARS;MARS;GARS;HARS;FARSA |
| 105 | Translation initiation complex formation Homo sapiens R-HSA-72649 | 6.01E-07 | 8.14E-06 | 3.871774 | 55.46293 | EIF4A1;RPS4Y1;RPS15;RPS19;EIF4H;RPS10;EIF4B;RPS9;RPS7;RPS8;RPS5;EIF2S2;EIF2S1;RPS25;RPS28;EIF3M;RPS27;EIF3K;RPS29;EIF3L;EIF3I;EIF3J;EIF3G;EIF3H;EIF3C;RPS21;EIF3D;EIF3A;RPS24;EIF4G1 |

| | | | | | | |
|---|---|---|---|---|---|---|
| 106 | Signaling by Insulin receptor Homo sapiens R-HSA-74752 | 7.17E-07 | 9.62E-06 | 1.838665 | 26.012918 | ITGA2B;PEBP1;WDR83;IRS2;PSMD8;PSMD6;PSMD7;PSMD4;AKT2;PSMD2;PSMD3;AKT1;PSMD1;ATP6V1G1;MAP2K1;ATP6V0E1;MAP2K2;RRAGA;AKAP9;PSME1;PSME2;TLN1;ATP6V0D1;RAF1;VCL;ATP6V1A;SPTBN5;PSMD12;SHC2;PSMD11;PSMD13;CUL3;IQGAP1;PHB;RAP1B;UBB;UBC;THEM4;ATP6V0A4;PSMF1;ATP6V1C1;SPTBN1;ATP6V1C2;SPTBN2;INSR;BRAF;PTK2;PTPRA;NF1;GRB2;CALM1;MAP3K11;EIF4G1;CAB39L;YWHAB;RASGRF2;PRKAG2;ARRB1;TCIRG1;RASGEF1A;MLST8;JAK2;SPTAN1;JAK1;ATP6V0B;ATP6AP1;STRADA;STRADB;PPP2R5B;PSMA5;PSMA6;PPM1A;PSMA3;PSMA1;PSMA2;KIT;RASA2;IL3RA;SOS1;ATP6V0C;PRKAA1;PRKAA2;SRC;PSMA7;EGFR;PSMB10;PSMB6;PSMB4;PSMB5;PSMB2;ERBB3;PSMB3;PSMB1;ERBB2;MAPK1;CSK;EIF4B;MAPK3;DAB2IP;PTPN11;PSMB8;MTOR;PSMB9;PSMC5;PSMC4;PSMC2;LAMTOR1 |
| 107 | Cap-dependent Translation Initiation Homo sapiens R-HSA-72737 | 7.31E-07 | 9.63E-06 | 2.631652 | 37.181295 | RPL5;EIF4A1;RPL30;RPL31;RPLP1;RPL6;RPS15;RPL7A;RPS19;RPL36;RPLP2;RPL37;RPS10;EIF5B;RPS9;RPS7;RPS8;RPS5;RPL37A;RPL29;RPS4Y1;EIF4H;EIF4B;RPL19;EIF2B5;EIF2B4;RPL41;EIF2B2;RPL35A;EIF2S2;EIF2S1;RPS25;RPS28;EIF3M;RPS27;EIF5;EIF3K;RPS29;EIF3L;EIF3I;EIF3J;EIF3G;EIF3H;EIF3C;RPS21;EIF3D;RPS24;EIF3A;EIF4G1 |
| 108 | Eukaryotic Translation Initiation Homo sapiens R-HSA-72613 | 7.31E-07 | 9.63E-06 | 2.631652 | 37.181295 | RPL5;EIF4A1;RPL30;RPL31;RPLP1;RPL6;RPS15;RPL7A;RPS19;RPL36;RPLP2;RPL37;RPS10;EIF5B;RPS9;RPS7;RPS8;RPS5;RPL37A;RPL29;RPS4Y1;EIF4H;EIF4B;RPL19;EIF2B5;EIF2B4;RPL41;EIF2B2;RPL35A;EIF2S2;EIF2S1;RPS25;RPS28;EIF3M;RPS27;EIF5;EIF3K;RPS29;EIF3L;EIF3I;EIF3J;EIF3G;EIF3H;EIF3C;RPS21;EIF3D;RPS24;EIF3A;EIF4G1 |

| | | | | | | |
|---|---|---|---|---|---|---|
| 109 | Cell Cycle, Mitotic Homo sapiens R-HSA-69278 | 8.59E-07 | 1.12E-05 | 1.655942 | 23.130149 | RB1;ANKLE2;GMNN;PPP2R2A;SMC3;SMC2;PSMD8;GOLGA2;PSMD6;CCND3;PSMD7;PSMD4;PSMD2;PSMD3;PSMD1;EP300;HIST1H2AC;CDK5RAP2;TMPO;NUP214;RAB2A;CSNK2A1;FBXW11;CSNK2A2;UBE2E1;LEMD2;TUBA4A;CDC25B;CLIP1;AKAP9;PSME1;PSME2;ANAPC5;NUP98;OPTN;BLZF1;NUP205;PSMD12;SET;PSMD11;ANAPC16;MAX;TUBGCP2;PSMD13;CUL1;PDS5B;AAAS;PDS5A;PMF1;ANAPC11;SKA2;POM121C;UBB;RAD21;CEP70;TPR;UBC;PSMF1;CEP78;H3F3A;NEK7;CDK7;STAG1;RPS27;STAG2;CEP57;YWHAE;AHCTF1;CDKN1B;HSP90AB1;NUMA1;CKS1B;NIPBL;ACTR1A;PCM1;XPO1;CNTRL;TK2;CEP290;RAB8A;OFD1;HSP90AA1;PPP1R12A;RFC1;H2AFZ;TUBB;HAUS4;PPP2R5B;HAUS3;KIF23;SMC1A;TUBG1;HAUS1;PSMA5;PSMA6;CCNA1;PSMA3;PSMA1;KIF2A;PSMA2;GORASP2;NUP50;MCM3;MAPRE1;PAFAH1B1;SDCCAG8;MCPH1;DYNC1I2;DCTN2;HDAC1;SSNA1;USO1;HDAC8;PSMA7;PSMB10;CENPC;PSMB6;POLD4;PSMB4;FZR1;PSMB5;PSMB2;RBBP4;PSMB3;PSMB1;LMNA;MAPK1;MAPK3;CENPT;RANBP2;DYNC1H1;CDK11A;CEP152;RPA2;TUBB4B;CDK11B;PSMB8;PSMB9;PSMC5;PSMC4;CENPJ;RPA3;PSMC2;PPP2R2D;NCAPD3;LPIN2;LPIN3;MAD2L1 |
| 110 | Activation of the mRNA upon binding of the cap-binding complex and eIFs, and subsequent binding to 43S Homo sapiens R-HSA-72662 | 9.79E-07 | 1.27E-05 | 3.733255 | 51.657406 | EIF4A1;RPS4Y1;RPS15;RPS19;EIF4H;RPS10;EIF4B;RPS9;RPS7;RPS8;RPS5;EIF2S2;EIF2S1;RPS25;RPS28;EIF3M;RPS27;EIF3K;RPS29;EIF3L;EIF3I;EIF3J;EIF3G;EIF3H;EIF3C;RPS21;EIF3D;EIF3A;RPS24;EIF4G1 |
| 111 | EPHB-mediated forward signaling Homo sapiens R-HSA-3928662 | 1.13E-06 | 1.44E-05 | 4.642563 | 63.580688 | EPHB6;ACTR3;ACTR2;YES1;ROCK1;SRC;ROCK2;SDC2;ARPC1B;ARHGEF28;LIMK2;ARPC1A;ITSN1;ARPC4;WASL;RHOA;PTK2;ACTG1;CDC42;EFNB2;ARPC2;ARPC3;CFL1;RAC1 |

| | | | | | | |
|---|---|---|---|---|---|---|
| 112 | M Phase Homo sapiens R-HSA-68886 | 1.29E-06 | 1.63E-05 | 1.891852 | 25.661623 | RB1;ANKLE2;AHCTF1;PPP2R2A;SMC3;SMC2;PSMD8;GOLGA2;PSMD6;NIPBL;PSMD7;XPO1;PSMD4;PSMD2;PSMD3;PSMD1;HIST1H2AC;TMPO;NUP214;RAB2A;CSNK2A1;H2AFZ;CSNK2A2;UBE2E1;PPP2R5B;KIF23;SMC1A;LEMD2;PSMA5;PSMA6;PSMA3;CLIP1;PSMA1;KIF2A;PSMA2;GORASP2;NUP50;PSME1;PSME2;ANAPC5;NUP98;MAPRE1;BLZF1;PAFAH1B1;MCPH1;NUP205;PSMD12;SET;PSMD11;ANAPC16;PSMD13;USO1;PDS5B;AAAS;PDS5A;HDAC8;PMF1;PSMA7;PSMB10;ANAPC11;SKA2;CENPC;PSMB6;PSMB4;PSMB5;POM121C;PSMB2;UBB;PSMB3;RAD21;PSMB1;LMNA;TPR;UBC;MAPK1;PSMF1;MAPK3;CENPT;RANBP2;H3F3A;NEK7;PSMB8;PSMB9;STAG1;PSMC5;RPS27;STAG2;PSMC4;PSMC2;PPP2R2D;NCAPD3;LPIN2;LPIN3;MAD2L1 |
| 113 | Golgi-to-ER retrograde transport Homo sapiens R-HSA-8856688 | 1.50E-06 | 1.89E-05 | 2.603533 | 34.916203 | COPB2;ARF4;DYNC1I2;COPA;NBAS;TMED10;DCTN2;KDELR1;COPB1;STX18;KIF12;NAPG;AGPAT3;DYNC2LI1;KLC2;KIF3A;KIF5B;TMED3;KIF13B;TMED2;KIF21A;KIF1B;KIFAP3;TMED7;NSF;DYNC1H1;TMED9;GALNT1;SURF4;KIF25;COPZ2;KIF23;ARFGAP3;COPZ1;ARCN1;RINT1;DYNC1LI2;KIF2A;KIF16B;RAB3GAP2;KDELR2;COPG2;PAFAH1B3;COPE;ARF5;RAB3GAP1;PAFAH1B1 |
| 114 | GTP hydrolysis and joining of the 60S ribosomal subunit Homo sapiens R-HSA-72706 | 1.57E-06 | 1.96E-05 | 2.631431 | 35.170442 | RPL5;EIF4A1;RPL30;RPL31;RPLP1;RPS4Y1;RPL6;RPS15;RPL7A;RPS19;RPL36;EIF4H;RPLP2;RPL37;RPS10;EIF4B;RPL19;EIF5B;RPS9;RPL41;RPS7;RPS8;RPS5;RPL35A;EIF2S2;EIF2S1;RPS25;RPS28;EIF3M;RPS27;EIF5;EIF3K;RPS29;EIF3L;EIF3I;RPL37A;EIF3J;EIF3G;EIF3H;RPL29;EIF3C;RPS21;EIF3D;RPS24;EIF3A;EIF4G1 |
| 115 | Synthesis of DNA Homo sapiens R-HSA-69239 | 3.51E-06 | 4.35E-05 | 2.663343 | 33.44804 | RB1;PSMD12;PSMD11;CDKN1B;PSMD13;PSMB10;PSMA7;PSMD8;PSMB6;POLD4;PSMD6;FZR1;PSMB4;PSMD7;PSMB5;PSMD4;UBB;PSMB2;PSMB3;PSMD2;PSMD3;UBC;PSMB1;PSMD1;PSMF1;RFC1;RPA2;PSMB8;PSMB9;PSMA5;CCNA1;PSMA6;PSMA3;PSMC5;PSMA1;PSMA2;PSMC4;RPA3;PSMC2;PSME1;MCM3;PSME2 |
| 116 | DNA Replication Pre-Initiation Homo sapiens R-HSA-69002 | 4.09E-06 | 4.97E-05 | 2.866296 | 35.564305 | PSMD12;PSMD11;PSMD13;GMNN;PSMB10;PSMA7;PSMD8;PSMB6;PSMD6;PSMB4;PSMD7;PSMB5;PSMD4;UBB;PSMB2;PSMB3;PSMD2;PSMD3;UBC;PSMB1;PSMD1;PSMF1;RPA2;PSMB8;PSMB9;PSMA5;PSMA6;PSMA3;PSMC5;PSMA1;PSMA2;PSMC4;RPA3;PSMC2;MCM3;PSME1;PSME2 |

| | | | | | | |
|---|---|---|---|---|---|---|
| 117 | M/G1 Transition Homo sapiens R-HSA-68874 | 4.09E-06 | 4.97E-05 | 2.866296 | 35.564305 | PSMD12;PSMD11;PSMD13;GMNN;PSMB10;PSMA7;PSMD8;PSMB6;PSMD6;PSMB4;PSMD7;PSMB5;PSMD4;UBB;PSMB2;PSMB3;PSMD2;PSMD3;UBC;PSMB1;PSMD1;PSMF1;RPA2;PSMB8;PSMB9;PSMA5;PSMA6;PSMA3;PSMC5;PSMA1;PSMA2;PSMC4;RPA3;PSMC2;MCM3;PSME1;PSME2 |
| 118 | Mitotic Anaphase Homo sapiens R-HSA-68882 | 4.13E-06 | 4.98E-05 | 2.102798 | 26.066707 | ANKLE2;AHCTF1;PPP2R2A;SMC3;PSMD8;PSMD6;PSMD7;XPO1;PSMD4;PSMD2;PSMD3;PSMD1;TMPO;UBE2E1;PPP2R5B;SMC1A;LEMD2;PSMA5;PSMA6;CLIP1;PSMA3;KIF2A;PSMA1;PSMA2;PSME1;PSME2;ANAPC5;NUP98;MAPRE1;PAFAH1B1;PSMD12;PSMD11;ANAPC16;PSMD13;PDS5B;PDS5A;HDAC8;PMF1;PSMB10;PSMA7;ANAPC11;SKA2;CENPC;PSMB6;PSMB4;PSMB5;UBB;PSMB2;PSMB3;RAD21;UBC;LMNA;PSMB1;PSMF1;CENPT;RANBP2;PSMB8;PSMB9;STAG1;PSMC5;RPS27;STAG2;PSMC4;PSMC2;MAD2L1 |
| 119 | Assembly of the primary cilium Homo sapiens R-HSA-5617833 | 4.44E-06 | 5.31E-05 | 2.043559 | 25.184617 | YWHAE;CEP57;ARF4;TRAF3IP1;PKD1;ACTR1A;PCM1;CNTRL;KIFAP3;TNPO1;IFT57;CC2D2A;CEP290;BBS4;RAB8A;CDK5RAP2;IQCB1;OFD1;CCT3;CCT2;HSP90AA1;EXOC7;TUBB;HAUS4;HAUS3;TUBG1;WDR34;TUBA4A;HAUS1;IFT88;AKAP9;TCP1;EXOC4;EXOC6;EXOC5;MAPRE1;EXOC2;PAFAH1B1;SDCCAG8;DYNC1I2;DCTN2;ARL3;SSNA1;HDAC6;DYNC2LI1;IFT74;ARL13B;KIF3A;TTC21B;CEP70;PDE6D;CCT8;CCT5;CEP78;CCT4;BBS2;DYNC1H1;CEP152;RAB3IP;BBIP1;TUBB4B;IFT20;AHI1;WDR60;CENPJ;TRIP11;DYNLRB1;CLUAP1;CEP89 |
| 120 | Formation of the ternary complex, and subsequently, the 43S complex Homo sapiens R-HSA-72695 | 4.51E-06 | 5.34E-05 | 3.772317 | 46.435492 | RPS4Y1;RPS15;RPS19;RPS10;RPS9;RPS7;RPS8;RPS5;EIF2S2;EIF2S1;RPS25;RPS28;EIF3M;RPS27;EIF3K;RPS29;EIF3L;EIF3I;EIF3J;EIF3G;EIF3H;EIF3C;RPS21;EIF3D;EIF3A;RPS24 |
| 121 | Mitotic Metaphase and Anaphase Homo sapiens R-HSA-2555396 | 5.17E-06 | 6.04E-05 | 2.083371 | 25.359751 | ANKLE2;AHCTF1;PPP2R2A;SMC3;PSMD8;PSMD6;PSMD7;XPO1;PSMD4;PSMD2;PSMD3;PSMD1;TMPO;UBE2E1;PPP2R5B;SMC1A;LEMD2;PSMA5;PSMA6;CLIP1;PSMA3;KIF2A;PSMA1;PSMA2;PSME1;PSME2;ANAPC5;NUP98;MAPRE1;PAFAH1B1;PSMD12;PSMD11;ANAPC16;PSMD13;PDS5B;PDS5A;HDAC8;PMF1;PSMB10;PSMA7;ANAPC11;SKA2;CENPC;PSMB6;PSMB4;PSMB5;UBB;PSMB2;PSMB3;RAD21;UBC;LMNA;PSMB1;PSMF1;CENPT;RANBP2;PSMB8;PSMB9;STAG1;PSMC5;RPS27;STAG2;PSMC4;PSMC2;MAD2L1 |

| | | | | | | |
|---|---|---|---|---|---|---|
| 122 | Influenza Infection Homo sapiens R-HSA-168254 | 5.18E-06 | 6.04E-05 | 2.211365 | 26.913959 | RPL5;RPL30;RPL31;RPLP1;RPL6;IPO5;RPS15;RPL7A;XPO1;RPS19;RPL36;RPLP2;KPNA4;RPL37;KPNA2;KPNA3;RPS10;KPNA1;NUP214;RPS9;HSP90AA1;RPS7;RPS8;RPS5;GTF2F2;DNAJC3;NUP50;RPL37A;NUP98;RPL29;NUP205;RPS4Y1;AAAS;POM121C;POLR2B;POLR2C;TPR;POLR2E;POLR2H;POLR2J;RPL19;RANBP2;RPL41;TGFB1;EIF2AK2;RPL35A;ISG15;RPS25;RPS28;RPS27;RPS29;CALR;GRSF1;RPS21;RPS24;RAN;HSPA1A |
| 123 | Apoptotic cleavage of cellular proteins Homo sapiens R-HSA-111465 | 6.13E-06 | 7.09E-05 | 4.567535 | 54.819518 | DSP;BCAP31;GSN;DBNL;ROCK1;PRKCD;ADD1;PTK2;TJP1;CASP7;OCLN;FNTA;STK24;APC;CDH1;LMNA;ACIN1;CTNNB1;SPTAN1;BIRC2;TJP2 |
| 124 | Signaling by FGFR2 Homo sapiens R-HSA-5654738 | 6.33E-06 | 7.25E-05 | 1.680368 | 20.115459 | AHCYL1;ITGA2B;PEBP1;WDR83;IRS2;TIAL1;PSMD8;PSMD6;PSMD7;PSMD4;AKT2;PSMD2;PSMD3;AKT1;PSMD1;PIP4K2C;IER3;MAP2K1;MAP2K2;PRKCD;GTF2F2;ADCY9;AKAP9;PSME1;PSME2;TLN1;RAF1;VCL;SPTBN5;PSMD12;SHC2;PSMD11;PSMD13;CUL3;PIK3R3;ITPR2;ITPR3;IQGAP1;PHB;RAP1B;UBB;PRKAR2A;UBC;THEM4;PLCG1;PSMF1;SPTBN1;SPTBN2;TIA1;INSR;BRAF;PTK2;HNRNPM;AGO3;PTPRA;HNRNPF;NF1;GRB2;CALM1;MAP3K11;GSK3B;CDKN1B;YWHAB;RASGRF2;ARRB1;PTBP1;RASGEF1A;MLST8;JAK2;SPTAN1;JAK1;PPP2R5B;PSMA5;PSMA6;PSMA3;CREB1;PSMA1;HNRNPH1;PSMA2;KIT;RASA2;IL3RA;SOS1;SRC;CBL;ADCY6;PSMA7;EGFR;PSMB10;PSMB6;PSMB4;PSMB5;PSMB2;ERBB3;PSMB3;POLR2B;MKNK1;POLR2C;PSMB1;ERBB2;POLR2E;MAPK1;CSK;POLR2H;POLR2J;MAPK3;BAD;DAB2IP;PTPN11;PSMB8;MTOR;PSMB9;PSMC5;PSMC4;PSMC2;RPS6KB2;TNRC6A |
| 125 | Sema4D in semaphorin signaling Homo sapiens R-HSA-400685 | 6.94E-06 | 7.90E-05 | 5.913027 | 70.23516 | ARHGEF12;ROCK1;SEMA4D;ROCK2;LIMK2;ARHGAP35;RHOA;MYL12B;CDC42;MYL6;RRAS;ERBB2;MYH14;MYH9;RAC1;MYH10;MET |

| | | | | | | |
|---|---|---|---|---|---|---|
| 126 | Signaling by FGFR Homo sapiens R-HSA-190236 | 7.63E-06 | 8.61E-05 | 1.667342 | 19.647146 | AHCYL1;ITGA2B;PEBP1;WDR83;IRS2;TIAL1;PSMD8;PSMD6;PSMD7;PSMD4;AKT2;PSMD2;PSMD3;AKT1;PSMD1;PIP4K2C;IER3;MAP2K1;MAP2K2;PRKCD;GTF2F2;ADCY9;AKAP9;PSME1;PSME2;TLN1;RAF1;VCL;SPTBN5;PSMD12;SHC2;PSMD11;PSMD13;CUL3;PIK3R3;ITPR2;ITPR3;IQGAP1;PHB;RAP1B;UBB;PRKAR2A;UBC;THEM4;PLCG1;PSMF1;SPTBN1;SPTBN2;TIA1;INSR;BRAF;PTK2;HNRNPM;AGO3;PTPRA;HNRNPF;NF1;GRB2;CALM1;MAP3K11;GSK3B;CDKN1B;YWHAB;RASGRF2;ARRB1;PTBP1;RASGEF1A;MLST8;JAK2;SPTAN1;JAK1;PPP2R5B;PSMA5;PSMA6;PSMA3;CREB1;PSMA1;HNRNPH1;PSMA2;KIT;RASA2;IL3RA;SOS1;SRC;CBL;ADCY6;PSMA7;EGFR;PSMB10;PSMB6;PSMB4;PSMB5;PSMB2;ERBB3;PSMB3;POLR2B;MKNK1;POLR2C;PSMB1;ERBB2;POLR2E;MAPK1;CSK;POLR2H;POLR2J;MAPK3;GALNT3;BAD;DAB2IP;PTPN11;PSMB8;MTOR;PSMB9;PSMC5;PSMC4;PSMC2;RPS6KB2;TNRC6A |
| 127 | L13a-mediated translational silencing of Ceruloplasmin expression Homo sapiens R-HSA-156827 | 7.93E-06 | 8.81E-05 | 2.475146 | 29.069202 | RPL5;EIF4A1;RPL30;RPL31;RPLP1;RPS4Y1;RPL6;RPS15;RPL7A;RPS19;RPL36;EIF4H;RPLP2;RPL37;RPS10;EIF4B;RPL19;RPS9;RPL41;RPS7;RPS8;RPS5;RPL35A;EIF2S2;EIF2S1;RPS25;RPS28;EIF3M;RPS27;EIF3K;RPS29;EIF3L;EIF3I;RPL37A;EIF3J;EIF3G;EIF3H;RPL29;EIF3C;RPS21;EIF3D;RPS24;EIF3A;EIF4G1 |
| 128 | 3' -UTR-mediated translational regulation Homo sapiens R-HSA-157279 | 7.93E-06 | 8.81E-05 | 2.475146 | 29.069202 | RPL5;EIF4A1;RPL30;RPL31;RPLP1;RPS4Y1;RPL6;RPS15;RPL7A;RPS19;RPL36;EIF4H;RPLP2;RPL37;RPS10;EIF4B;RPL19;RPS9;RPL41;RPS7;RPS8;RPS5;RPL35A;EIF2S2;EIF2S1;RPS25;RPS28;EIF3M;RPS27;EIF3K;RPS29;EIF3L;EIF3I;RPL37A;EIF3J;EIF3G;EIF3H;RPL29;EIF3C;RPS21;EIF3D;RPS24;EIF3A;EIF4G1 |

| | | | | | | |
|---|---|---|---|---|---|---|
| 129 | Transcriptional Regulation by TP53 Homo sapiens R-HSA-3700989 | 8.20E-06 | 9.04E-05 | 1.684521 | 19.727497 | ATF2;CCNT2;EHMT1;COX6A1;GLS;SESN3;AKT2;AKT1;EP300;NBN;BNIP3L;SUPT16H;USP7;CSNK2A1;CSNK2A2;GTF2F2;COX6B1;RRAGA;SCO2;DDIT4;COX19;MEAF6;COX16;UBB;PRDX1;UBC;TP53BP2;SMYD2;COX14;ATRIP;FANCI;PRELID1;SURF1;FANCC;CNOT4;CDK9;CDK7;TP53I3;RGCC;AGO3;CDK5;BCL6;CNOT1;CNOT2;RAD17;ATM;CDK12;CDK13;CNOT8;ATR;COX20;YWHAE;BTG2;CDKN1B;PPP1R13B;PHF20;YWHAB;CHM;PRKAG2;CHD4;CHD3;COX7C;AIFM2;MLST8;TNKS1BP1;CASP2;NELFB;TOPBP1;YWHAH;COX8A;GPX2;PRMT1;TOP3A;RABGGTA;CCNA1;RRM2B;PRKAA1;HDAC2;PRKAA2;HDAC1;COX5A;RBBP4;POLR2B;PERP;POLR2C;POLR2E;RBBP7;POLR2H;BID;BRD7;POLR2J;NPM1;TXNRD1;GSR;TAF11;RPA2;SSRP1;COX6C;MLH1;HIPK1;LRPPRC;MTOR;RAD50;CENPJ;RPA3;TCEA1;FAS;BAX;LAMTOR1;TAF5;TAF3;TNRC6A;TAF2 |
| 130 | COPI-dependent Golgi-to-ER retrograde traffic Homo sapiens R-HSA-6811434 | 8.38E-06 | 9.17E-05 | 2.788201 | 32.59326 | ARF4;COPB2;NBAS;COPA;TMED10;KDELR1;COPB1;STX18;KIF12;NAPG;KLC2;KIF3A;KIF5B;TMED3;KIF13B;TMED2;KIF1B;KIF21A;TMED7;KIFAP3;NSF;TMED9;SURF4;KIF25;COPZ2;KIF23;ARFGAP3;COPZ1;ARCN1;RINT1;KIF2A;KIF16B;KDELR2;COPG2;ARF5;COPE |
| 131 | Budding and maturation of HIV virion Homo sapiens R-HSA-162588 | 9.61E-06 | 0.000104 | 6.18257 | 71.427879 | TSG101;PDCD6IP;VPS4B;VPS4A;NEDD4L;VPS37D;VPS37B;UBB;UBC;CHMP2B;CHMP4C;CHMP3;CHMP4B;PPIA;VPS28;CHMP5 |
| 132 | Insulin receptor signalling cascade Homo sapiens R-HSA-74751 | 9.98E-06 | 0.000108 | 1.75899 | 20.254339 | CAB39L;YWHAB;RASGRF2;ITGA2B;PRKAG2;PEBP1;WDR83;IRS2;ARRB1;PSMD8;PSMD6;PSMD7;PSMD4;RASGEF1A;AKT2;PSMD2;PSMD3;MLST8;AKT1;PSMD1;JAK2;SPTAN1;JAK1;MAP2K1;MAP2K2;STRADA;STRADB;PPP2R5B;PSMA5;PSMA6;PPM1A;RRAGA;PSMA3;PSMA1;PSMA2;AKAP9;KIT;RASA2;IL3RA;PSME1;PSME2;TLN1;RAF1;SOS1;VCL;SPTBN5;PSMD12;SHC2;PRKAA1;PSMD11;PRKAA2;PSMD13;SRC;CUL3;IQGAP1;PHB;PSMA7;EGFR;PSMB10;PSMB6;RAP1B;PSMB4;PSMB5;PSMB2;UBB;ERBB3;PSMB3;PSMB1;UBC;ERBB2;THEM4;MAPK1;CSK;PSMF1;SPTBN1;EIF4B;SPTBN2;MAPK3;INSR;DAB2IP;BRAF;PTPN11;PTK2;PSMB8;MTOR;PSMB9;PSMC5;PSMC4;PTPRA;PSMC2;NF1;GRB2;LAMTOR1;CALM1;MAP3K11;EIF4G1 |

| | | | | | | |
|---|---|---|---|---|---|---|
| 133 | IRS-mediated signalling Homo sapiens R-HSA-112399 | 1.11E-05 | 0.000118 | 1.758915 | 20.074157 | CAB39L;YWHAB;RASGRF2;ITGA2B;PRKAG2;PEBP1;WDR83;IRS2;ARRB1;PSMD8;PSMD6;PSMD7;PSMD4;RASGEF1A;AKT2;PSMD2;PSMD3;MLST8;AKT1;PSMD1;JAK2;SPTAN1;JAK1;MAP2K1;MAP2K2;STRADA;STRADB;PPP2R5B;PSMA5;PSMA6;PPM1A;RRAGA;PSMA3;PSMA1;PSMA2;AKAP9;KIT;RASA2;IL3RA;PSME1;PSME2;TLN1;RAF1;SOS1;VCL;SPTBN5;PSMD12;SHC2;PRKAA1;PSMD11;PRKAA2;PSMD13;SRC;CUL3;IQGAP1;PHB;PSMA7;EGFR;PSMB10;PSMB6;RAP1B;PSMB4;PSMB5;PSMB2;UBB;ERBB3;PSMB3;PSMB1;UBC;ERBB2;THEM4;MAPK1;CSK;PSMF1;SPTBN1;EIF4B;SPTBN2;MAPK3;DAB2IP;BRAF;PTPN11;PTK2;PSMB8;MTOR;PSMB9;PSMC5;PSMC4;PTPRA;PSMC2;NF1;GRB2;LAMTOR1;CALM1;MAP3K11;EIF4G1 |
| 134 | trans-Golgi Network Vesicle Budding Homo sapiens R-HSA-199992 | 1.29E-05 | 0.000136 | 2.933633 | 33.032906 | APP;RAB5C;TFRC;CLTC;CLTB;AP4E1;ARRB1;CLINT1;GNS;TGOLN2;SNX2;AP1G2;AP1G1;STX4;SNX9;SNX5;AP1M2;HSPA8;SORT1;DTNBP1;HIP1R;M6PR;OCRL;AP3B1;PUM1;TPD52L1;IGF2R;DNM2;VAMP8;YIPF6;DNASE2;FTL |
| 135 | Clathrin derived vesicle budding Homo sapiens R-HSA-421837 | 1.29E-05 | 0.000136 | 2.933633 | 33.032906 | APP;RAB5C;TFRC;CLTC;CLTB;AP4E1;ARRB1;CLINT1;GNS;TGOLN2;SNX2;AP1G2;AP1G1;STX4;SNX9;SNX5;AP1M2;HSPA8;SORT1;DTNBP1;HIP1R;M6PR;OCRL;AP3B1;PUM1;TPD52L1;IGF2R;DNM2;VAMP8;YIPF6;DNASE2;FTL |
| 136 | Signaling by EGFR Homo sapiens R-HSA-177929 | 1.32E-05 | 0.000138 | 1.656848 | 18.617428 | AHCYL1;ITGA2B;PEBP1;WDR83;IRS2;PSMD8;PSMD6;PSMD7;PSMD4;AKT2;PSMD2;PSMD3;AKT1;PSMD1;PIP4K2C;AP2M1;IER3;MAP2K1;MAP2K2;PRKCD;ADAM10;ADAM17;ADCY9;AKAP9;PSME1;PSME2;TLN1;RAF1;STAM2;VCL;SPTBN5;PSMD12;SHC2;PSMD11;PSMD13;CUL3;PIK3R3;ITPR2;ITPR3;IQGAP1;PHB;RAP1B;UBB;PRKAR2A;UBC;THEM4;PLCG1;PSMF1;SPTBN1;SPTBN2;INSR;BRAF;PTK2;AGO3;PTPRA;NF1;GRB2;CALM1;MAP3K11;GSK3B;CDKN1B;YWHAB;RASGRF2;AP2A1;ARRB1;PTPRK;RASGEF1A;MLST8;JAK2;SPTAN1;JAK1;PPP2R5B;EPN1;PSMA5;PSMA6;PSMA3;CREB1;PSMA1;PSMA2;KIT;RASA2;IL3RA;SOS1;SRC;CBL;ADCY6;PSMA7;EGFR;PSMB10;PSMB6;CDC42;PSMB4;PSMB5;PSMB2;ERBB3;PSMB3;PSMB1;ERBB2;MAPK1;CSK;EPS15;MAPK3;BAD;DAB2IP;PTPN11;PSMB8;MTOR;PSMB9;PSMC5;PSMC4;PSMC2;RPS6KB2;SPRY1;TNRC6A |

| | | | | | | |
|---|---|---|---|---|---|---|
| 137 | DNA Replication Homo sapiens R-HSA-69306 | 1.50E-05 | 0.000156 | 2.418347 | 26.864584 | RB1;PSMD12;PSMD11;CDKN1B;PSMD13;GMNN;PSMB10;PSMA7;PSMD8;PSMB6;POLD4;PSMD6;FZR1;PSMB4;PSMD7;PSMB5;PSMD4;UBB;PSMB2;PSMB3;PSMD2;PSMD3;UBC;PSMB1;PSMD1;PSMF1;RFC1;RPA2;PSMB8;PSMB9;PSMA5;CCNA1;PSMA6;PSMA3;PSMC5;PSMA1;PSMA2;PSMC4;RPA3;PSMC2;PSME1;MCM3;PSME2 |
| 138 | Separation of Sister Chromatids Homo sapiens R-HSA-2467813 | 1.59E-05 | 0.000163 | 2.053696 | 22.697527 | AHCTF1;SMC3;PSMD8;PSMD6;PSMD7;XPO1;PSMD4;PSMD2;PSMD3;PSMD1;UBE2E1;PPP2R5B;SMC1A;PSMA5;PSMA6;CLIP1;PSMA3;KIF2A;PSMA1;PSMA2;PSME1;PSME2;ANAPC5;NUP98;MAPRE1;PAFAH1B1;PSMD12;PSMD11;ANAPC16;PSMD13;PDS5B;PDS5A;HDAC8;PMF1;PSMB10;PSMA7;ANAPC11;SKA2;CENPC;PSMB6;PSMB4;PSMB5;UBB;PSMB2;PSMB3;RAD21;UBC;PSMB1;PSMF1;CENPT;RANBP2;PSMB8;PSMB9;STAG1;PSMC5;RPS27;STAG2;PSMC4;PSMC2;MAD2L1 |
| 139 | VEGFA-VEGFR2 Pathway Homo sapiens R-HSA-4420097 | 1.77E-05 | 0.000181 | 1.68535 | 18.444163 | CYFIP1;NCKAP1;AHCYL1;ITGA2B;PEBP1;WDR83;IRS2;PSMD8;PSMD6;PSMD7;PSMD4;AKT2;PSMD2;PSMD3;AKT1;PSMD1;MAP2K1;MAP2K2;PRKCD;AKAP9;PSME1;PSME2;TLN1;RAF1;VCL;SPTBN5;PSMD12;SHC2;PSMD11;PSMD13;CUL3;ITPR2;ITPR3;IQGAP1;PHB;RAP1B;UBB;UBC;THEM4;PLCG1;PSMF1;SPTBN1;SPTBN2;BRAF;PTK2;PTPRA;NF1;GRB2;CALM1;MAP3K11;YWHAB;RASGRF2;HSPB1;ARRB1;SHB;RASGEF1A;MLST8;RAC1;JAK2;SPTAN1;JAK1;VAV3;HSP90AA1;PPP2R5B;RHOA;PSMA5;PSMA6;PSMA3;PSMA1;PSMA2;KIT;RASA2;IL3RA;SOS1;ROCK1;ROCK2;SRC;PSMA7;EGFR;PSMB10;PSMB6;CDC42;PSMB4;PSMB5;PSMB2;ERBB3;PSMB3;PSMB1;ERBB2;MAPK1;CSK;PAK2;MAPK3;DAB2IP;PSMB8;MTOR;PSMB9;MAPK13;PSMC5;ABI2;PSMC4;PSMC2;CTNNB1;BCAR1 |

| | | | | | |
|---|---|---|---|---|---|
| 140 | Developmental Biology Homo sapiens R-HSA-1266738 | 1.82E-05 | 0.000185 | 1.411289 | 15.401176 | EPHB6;ITSN1;CCNC;WDR83;RBPJ;PSMD8;PSMD6;PSMD7;RPS6KA5;PSMD4;DPYSL2;AKT2;PSMD2;PSMD3;AKT1;PSMD1;CSNK2A1;CSNK2A2;MED8;ANK2;ANK3;MYL6;AKAP9;PSME1;PSME2;HOXB2;EPHA1;TLN1;EPHA2;PFN2;SPTBN5;SDC2;IQGAP1;MYL12A;MYL12B;APH1A;APH1B;RRAS;PSMF1;PLCG1;SPTBN1;SPTBN2;VASP;CDK19;YES1;RDX;H3F3A;BRAF;PTK2;MED13L;GRB2;HOXD3;CALM1;CD24;RARG;ARPC1B;ARPC1A;CLTC;CLTB;MED19;AP2A1;WASL;HIF3A;SHB;MED13;MED10;CD36;RAC1;JAK2;SPTAN1;JAK1;NCOA1;PSENEN;VAV3;NCOA2;TSC22D1;NCOA3;POU5F1;SREBF2;RHOA;PSMA5;ENAH;PSMA6;PSMA3;NCOR1;PSMA1;PSMA2;MED21;KIT;PPARG;PSMA7;EGFR;RELA;PSMB6;PSMB4;PSMB5;PSMB2;PSMB3;PSMB1;PAK2;SUZ12;STAT3;DAB2IP;RANBP9;PSMB8;PSMB9;PSMC5;PSMC4;PSMC2;CTNNB1;AGRN;FOXA3;ITGA2B;PEBP1;IRS2;ARHGAP35;ACTG1;YY1;CCND3;CFL1;EP300;NRCAM;AP2M1;HIST1H2AC;MAP2K1;MAP2K2;ADAM10;TBL1XR1;COL4A4;RAF1;VCL;SHC2;PSMD12;PSMD11;PSMD13;KMT2C;ARHGEF28;CUL3;PHB;EFNA4;RAP1B;EFNB2;SDCBP;UBB;UBC;MYH14;SRGAP3;SRGAP1;MYH10;SMAD2;PRNP;HSPA8;SMAD4;TGFB1;SMAD3;MSN;ARPC4;NFKB1;DRAP1;EFNA1;DLG1;ARPC2;CDK5;DLG3;ARPC3;PTPRA;NF1;MAP3K11;ITGB1;GSK3B;HSP90AB1;YWHAB;CHD9;RASGRF2;ARRB1;LA[illegible] |
| 141 | Chromatin organization Homo sapiens R-HSA-4839726 | 1.91E-05 | 0.000191 | 1.842019 | 20.017186 | KDM5A;ATF2;KDM5B;SETD2;SMARCB1;SETD3;KDM5D;KDM1A;TRRAP;PHF20;KDM1B;EHMT1;ARID4A;YEATS4;ARID4B;CHD4;CHD3;ING4;MECOM;RUVBL1;EPC1;KAT8;EP300;HMG20B;UTY;JAK2;ARID2;ATXN7L3;HIST1H2AC;NCOA1;NCOA2;SMARCC2;PRMT7;KDM2A;PRMT1;H2AFZ;MSL3;ARID1B;JMJD6;MSL1;MORF4L1;KAT2A;NCOR1;BRMS1;TBL1XR1;KDM7A;KMT2E;SMARCD1;HDAC2;MEAF6;KMT2A;HDAC1;KMT2C;KMT2B;GPS2;HDAC8;RBBP4;SAP30L;SMYD2;SAP18;SUDS3;DMAP1;RBBP7;SUZ12;KDM4A;KDM4B;MCRS1;SETDB2;ELP2;ELP3;ELP5;SMARCA2;ELP6;COPRS;MRGBP;TADA3;OGT;RCOR1 |

| | | | | | | |
|---|---|---|---|---|---|---|
| 142 | Chromatin modifying enzymes Homo sapiens R-HSA-3247509 | 1.91E-05 | 0.000191 | 1.842019 | 20.017186 | KDM5A;ATF2;KDM5B;SETD2;SMARCB1;SETD3;KDM5D;KDM1A;TRRAP;PHF20;KDM1B;EHMT1;ARID4A;YEATS4;ARID4B;CHD4;CHD3;ING4;MECOM;RUVBL1;EPC1;KAT8;EP300;HMG20B;UTY;JAK2;ARID2;ATXN7L3;HIST1H2AC;NCOA1;NCOA2;SMARCC2;PRMT7;KDM2A;PRMT1;H2AFZ;MSL3;ARID1B;JMJD6;MSL1;MORF4L1;KAT2A;NCOR1;BRMS1;TBL1XR1;KDM7A;KMT2E;SMARCD1;HDAC2;MEAF6;KMT2A;HDAC1;KMT2C;KMT2B;GPS2;HDAC8;RBBP4;SAP30L;SMYD2;SAP18;SUDS3;DMAP1;RBBP7;SUZ12;KDM4A;KDM4B;MCRS1;SETDB2;ELP2;ELP3;ELP5;SMARCA2;ELP6;COPRS;MRGBP;TADA3;OGT;RCOR1 |
| 143 | IGF1R signaling cascade Homo sapiens R-HSA-2428924 | 2.08E-05 | 0.000204 | 1.722012 | 18.564985 | CAB39L;YWHAB;RASGRF2;ITGA2B;PRKAG2;PEBP1;WDR83;IRS2;ARRB1;PSMD8;PSMD6;PSMD7;PSMD4;RASGEF1A;AKT2;PSMD2;PSMD3;MLST8;AKT1;PSMD1;JAK2;SPTAN1;JAK1;MAP2K1;MAP2K2;STRADA;STRADB;PPP2R5B;PSMA5;PSMA6;PPM1A;RRAGA;PSMA3;PSMA1;PSMA2;AKAP9;KIT;RASA2;IL3RA;PSME1;PSME2;TLN1;RAF1;SOS1;VCL;SPTBN5;PSMD12;SHC2;PRKAA1;PSMD11;PRKAA2;PSMD13;SRC;CUL3;IQGAP1;PHB;PSMA7;EGFR;PSMB10;PSMB6;RAP1B;PSMB4;PSMB5;PSMB2;UBB;ERBB3;PSMB3;PSMB1;UBC;ERBB2;THEM4;MAPK1;CSK;PSMF1;SPTBN1;EIF4B;SPTBN2;MAPK3;DAB2IP;BRAF;PTPN11;PTK2;PSMB8;MTOR;PSMB9;PSMC5;PSMC4;PTPRA;PSMC2;NF1;GRB2;LAMTOR1;CALM1;MAP3K11;EIF4G1 |
| 144 | Signaling by Type 1 Insulin-like Growth Factor 1 Receptor (IGF1R) Homo sapiens R-HSA-2404192 | 2.08E-05 | 0.000204 | 1.722012 | 18.564985 | CAB39L;YWHAB;RASGRF2;ITGA2B;PRKAG2;PEBP1;WDR83;IRS2;ARRB1;PSMD8;PSMD6;PSMD7;PSMD4;RASGEF1A;AKT2;PSMD2;PSMD3;MLST8;AKT1;PSMD1;JAK2;SPTAN1;JAK1;MAP2K1;MAP2K2;STRADA;STRADB;PPP2R5B;PSMA5;PSMA6;PPM1A;RRAGA;PSMA3;PSMA1;PSMA2;AKAP9;KIT;RASA2;IL3RA;PSME1;PSME2;TLN1;RAF1;SOS1;VCL;SPTBN5;PSMD12;SHC2;PRKAA1;PSMD11;PRKAA2;PSMD13;SRC;CUL3;IQGAP1;PHB;PSMA7;EGFR;PSMB10;PSMB6;RAP1B;PSMB4;PSMB5;PSMB2;UBB;ERBB3;PSMB3;PSMB1;UBC;ERBB2;THEM4;MAPK1;CSK;PSMF1;SPTBN1;EIF4B;SPTBN2;MAPK3;DAB2IP;BRAF;PTPN11;PTK2;PSMB8;MTOR;PSMB9;PSMC5;PSMC4;PTPRA;PSMC2;NF1;GRB2;LAMTOR1;CALM1;MAP3K11;EIF4G1 |

| | | | | | | |
|---|---|---|---|---|---|---|
| 145 | IRS-related events triggered by IGF1R Homo sapiens R-HSA-2428928 | 2.08E-05 | 0.000204 | 1.722012 | 18.564985 | CAB39L;YWHAB;RASGRF2;ITGA2B;PRKAG2;PEBP1;WDR83;IRS2;ARRB1;PSMD8;PSMD6;PSMD7;PSMD4;RASGEF1A;AKT2;PSMD2;PSMD3;MLST8;AKT1;PSMD1;JAK2;SPTAN1;JAK1;MAP2K1;MAP2K2;STRADA;STRADB;PPP2R5B;PSMA5;PSMA6;PPM1A;RRAGA;PSMA3;PSMA1;PSMA2;AKAP9;KIT;RASA2;IL3RA;PSME1;PSME2;TLN1;RAF1;SOS1;VCL;SPTBN5;PSMD12;SHC2;PRKAA1;PSMD11;PRKAA2;PSMD13;SRC;CUL3;IQGAP1;PHB;PSMA7;EGFR;PSMB10;PSMB6;RAP1B;PSMB4;PSMB5;PSMB2;UBB;ERBB3;PSMB3;PSMB1;UBC;ERBB2;THEM4;MAPK1;CSK;PSMF1;SPTBN1;EIF4B;SPTBN2;MAPK3;DAB2IP;BRAF;PTPN11;PTK2;PSMB8;MTOR;PSMB9;PSMC5;PSMC4;PTPRA;PSMC2;NF1;GRB2;LAMTOR1;CALM1;MAP3K11;EIF4G1 |
| 146 | Signalling by NGF Homo sapiens R-HSA-166520 | 2.41E-05 | 0.000235 | 1.550574 | 16.488584 | AHCYL1;ITSN1;ITGA2B;PEBP1;WDR83;IRS2;PSMD8;PSMD6;PSMD7;RPS6KA5;PSMD4;AKT2;PSMD2;ARHGDIA;PSMD3;AKT1;PSMD1;PIP4K2C;AP2M1;IER3;PRKCI;MAP2K1;MAP2K2;PRKCD;ADAM17;ADCY9;AKAP9;PSME1;PSME2;TLN1;RAF1;SQSTM1;VCL;SHC2;SPTBN5;PSMD12;PSMD11;PSMD13;CUL3;ITPR2;PIK3R3;ITPR3;IQGAP1;PHB;RTN4;RAP1B;APH1A;APH1B;UBB;PRKAR2A;UBC;THEM4;PLCG1;PSMF1;ECT2;MAP2K5;SPTBN1;SPTBN2;INSR;BRAF;NFKB1;PTK2;NFKBIA;AGO3;PTPRA;NF1;GRB2;CALM1;MAP3K11;YWHAE;GSK3B;CDKN1B;YWHAB;RASGRF2;ARRB1;AP2A1;RASGEF1A;MCF2L;MLST8;CASP2;RAC1;JAK2;SPTAN1;JAK1;PSENEN;VAV3;ARHGEF12;ARHGEF16;PPP2R5B;RHOA;DNM2;PSMA5;PSMA6;PSMA3;CREB1;PSMA1;PSMA2;IL3RA;KIT;RASA2;ARHGEF1;ARHGEF2;SOS1;HDAC2;RALA;SRC;HDAC1;FURIN;ADCY6;PSMA7;EGFR;RELA;PSMB10;PSMB6;PSMB4;PSMB5;PSMB2;ERBB3;PSMB3;PSMB1;ERBB2;MAPK1;CSK;MAPK3;KIDINS220;BAD;STAT3;DAB2IP;PTPN11;PSMB8;MTOR;PSMB9;MAPK13;PSMC5;PSMC4;PSMC2;RPS6KB2;TNRC6A |
| 147 | Pyruvate metabolism and Citric Acid (TCA) cycle Homo sapiens R-HSA-71406 | 2.46E-05 | 0.000237 | 3.480575 | 36.931916 | PDHX;FH;PDHA1;MDH2;IDH3G;GLO1;DLST;PDHB;SDHD;SDHA;SDHB;PDP1;LDHB;LDHA;SUCLA2;NNT;IDH3B;OGDH;SUCLG2;SUCLG1;HAGH;ACO2;DLD;PDK2 |

| | | | | | | |
|---|---|---|---|---|---|---|
| 148 | TCF dependent signaling in response to WNT Homo sapiens R-HSA-201681 | 2.46E-05 | 0.000237 | 1.895469 | 20.112554 | GSK3B;TRRAP;USP34;CBY1;CDC73;PSMD8;PSMD6;PSMD7;XPO1;PSMD4;AKT2;PSMD2;PSMD3;RUVBL1;AKT1;PSMD1;RSPO3;EP300;HIST1H2AC;MEN1;RNF43;USP8;TLE3;TLE2;CSNK2A1;H2AFZ;CSNK2A2;WNT5A;PPP2R5B;PSMA5;PYGO1;PSMA6;PSMA3;PSMA1;PSMA2;PSME1;PSME2;CSNK1G2;PSMD12;PSMD11;PSMD13;HDAC1;CUL3;XIAP;PSMA7;PSMB10;LRP6;PSMB6;PSMB4;PSMB5;PSMB2;UBB;TNKS2;PSMB3;PSMB1;UBC;DVL3;PSMF1;FZD5;CSNK1A1;RYK;FZD6;H3F3A;PSMB8;PSMB9;PSMC5;APC;PSMC4;PSMC2;CTNNB1 |
| 149 | Downstream signaling events of B Cell Receptor (BCR) Homo sapiens R-HSA-1168372 | 2.49E-05 | 0.000237 | 1.915313 | 20.305703 | GSK3B;CDKN1B;IRS2;PSMD8;PSMD6;PSMD7;PSMD4;AKT2;PSMD2;PSMD3;MLST8;AKT1;PSMD1;PIP4K2C;IER3;FBXW11;PPP2R5B;PSMA5;PSMA6;PSMA3;CREB1;PSMA1;PSMA2;KIT;PSME1;PSME2;PSMD12;PSMD11;PSMD13;SRC;CUL1;PIK3R3;BCL10;RELA;EGFR;PSMB10;PSMA7;MALT1;PSMB6;PSMB4;PSMB5;UBB;ERBB3;PSMB2;PSMB3;UBC;ERBB2;PSMB1;THEM4;MAPK1;PSMF1;MAPK3;BAD;INSR;PTPN11;NFKB1;MTOR;PSMB8;PSMB9;NFKBIA;PSMC5;AGO3;PSMC4;PSMC2;RPS6KB2;REL;GRB2;TNRC6A |
| 150 | Fatty acid, triacylglycerol, and ketone body metabolism Homo sapiens R-HSA-535734 | 2.58E-05 | 0.000245 | 1.845472 | 19.494891 | ACADVL;CHD9;ECI1;TECR;FHL2;MED19;PRKAG2;CCNC;RORA;MCEE;MED13;ACADL;CPT2;MED10;EP300;CD36;ACADM;HMGCS2;HADH;TEAD1;TEAD2;NCOA1;ACOT9;NCOA2;ELOVL1;CPT1A;DGAT1;NCOA3;MED8;ACOT12;ACSL5;ACOT13;ACSL3;SREBF2;HADHB;HADHA;NCOR1;BDH2;PCCA;TBL1XR1;MED21;GPD2;PCCB;ACOT2;PPARG;PLIN2;ACOT4;YAP1;AGPAT5;PRKAA2;PEX11A;HMGCR;HSD17B12;ACACB;ACACA;NPAS2;AGPAT3;ACAT1;HMGCL;TGS1;THEM4;SLC25A20;FDFT1;DECR1;ESRRA;WWTR1;CDK19;AGK;NFYC;TXNRD1;NR1D1;MED13L;CYP1A1;LPIN2;LPIN3 |
| 151 | SRP-dependent cotranslational protein targeting to membrane Homo sapiens R-HSA-1799339 | 2.60E-05 | 0.000245 | 2.342471 | 24.731734 | RPL5;RPL30;TRAM1;RPN2;RPL31;RPLP1;RPN1;SRP54;RPS4Y1;RPL6;RPS15;SEC61A1;RPL7A;RPS19;RPL36;SSR1;RPLP2;RPL37;RPS10;SEC11A;RPL19;SEC11C;RPS9;RPL41;RPS7;SSR4;RPS8;RPS5;SSR2;SSR3;RPL35A;DDOST;RPS25;SPCS3;RPS28;SPCS2;RPS27;SPCS1;RPS29;RPL37A;RPL29;RPS21;RPS24 |
| 152 | Translocation of GLUT4 to the plasma membrane Homo sapiens R-HSA-1445148 | 2.76E-05 | 0.000258 | 3.046668 | 31.98315 | YWHAE;C2CD5;RALA;PRKAA2;STXBP3;YWHAB;PRKAG2;KIF3A;AKT2;AKT1;STX4;RAC1;KIFAP3;RAB8A;YWHAH;RAB4A;EXOC7;RALGAPA2;RAB10;RALGAPB;RAB14;RAB13;EXOC4;EXOC6;EXOC5;CALM1;RHOQ;EXOC2 |

| | | | | | | |
|---|---|---|---|---|---|---|
| 153 | HIV Life Cycle Homo sapiens R-HSA-162587 | 3.02E-05 | 0.000281 | 2.101284 | 21.869986 | CCNT2;GTF2B;PSIP1;XPO1;NELFB;KPNA1;NUP214;SUPT16H;PDCD6IP;VPS37D;VPS37B;GTF2F2;NUP50;CHMP4C;CHMP4B;CHMP3;NUP98;PPIA;VPS28;CHMP5;GTF2A1;NUP205;GTF2A2;TSG101;RNMT;VPS4B;VPS4A;NEDD4L;FURIN;AAAS;UBB;POM121C;POLR2B;POLR2C;TPR;UBC;POLR2E;POLR2H;POLR2J;RANBP2;XRCC6;XRCC5;TAF11;SSRP1;CDK9;CDK7;CHMP2B;NMT1;TCEA1;TAF5;TAF3;RAN;TAF2 |
| 154 | N-glycan trimming in the ER and Calnexin/Calreticulin cycle Homo sapiens R-HSA-532668 | 3.06E-05 | 0.000283 | 8.341142 | 86.690246 | PDIA3;EDEM3;GANAB;OS9;EDEM2;SEL1L;SYVN1;DERL2;MLEC;MOGS;CALR;UGGT2 |
| 155 | Signaling by VEGF Homo sapiens R-HSA-194138 | 3.27E-05 | 0.0003 | 1.647767 | 17.016689 | CYFIP1;NCKAP1;AHCYL1;ITGA2B;PEBP1;WDR83;IRS2;PSMD8;PSMD6;PSMD7;PSMD4;AKT2;PSMD2;PSMD3;AKT1;PSMD1;MAP2K1;MAP2K2;PRKCD;AKAP9;PSME1;PSME2;TLN1;RAF1;VCL;SPTBN5;PSMD12;SHC2;PSMD11;PSMD13;CUL3;ITPR2;ITPR3;IQGAP1;PHB;RAP1B;UBB;UBC;THEM4;PLCG1;PSMF1;SPTBN1;SPTBN2;BRAF;PTK2;PTPRA;NF1;GRB2;CALM1;MAP3K11;YWHAB;RASGRF2;HSPB1;ARRB1;SHB;RASGEF1A;MLST8;RAC1;JAK2;SPTAN1;JAK1;VAV3;HSP90AA1;PPP2R5B;RHOA;PSMA5;PSMA6;PSMA3;PSMA1;PSMA2;KIT;RASA2;IL3RA;SOS1;ROCK1;ROCK2;SRC;PSMA7;EGFR;PSMB10;PSMB6;CDC42;PSMB4;PSMB5;PSMB2;ERBB3;PSMB3;PSMB1;ERBB2;MAPK1;CSK;PAK2;MAPK3;VEGFB;DAB2IP;PSMB8;MTOR;PSMB9;MAPK13;PSMC5;ABI2;PSMC4;PSMC2;CTNNB1;BCAR1 |
| 156 | Late Phase of HIV Life Cycle Homo sapiens R-HSA-162599 | 3.42E-05 | 0.000311 | 2.163322 | 22.248698 | CCNT2;GTF2B;XPO1;NELFB;NUP214;SUPT16H;PDCD6IP;VPS37D;VPS37B;GTF2F2;NUP50;CHMP4C;CHMP4B;CHMP3;NUP98;PPIA;VPS28;CHMP5;GTF2A1;NUP205;GTF2A2;TSG101;RNMT;VPS4B;VPS4A;NEDD4L;FURIN;AAAS;UBB;POM121C;POLR2B;POLR2C;TPR;UBC;POLR2E;POLR2H;POLR2J;RANBP2;TAF11;SSRP1;CDK9;CDK7;CHMP2B;NMT1;TCEA1;TAF5;TAF3;RAN;TAF2 |

| | | | | | | |
|---|---|---|---|---|---|---|
| 157 | Metabolism of lipids and lipoproteins Homo sapiens R-HSA-556833 | 3.54E-05 | 0.000321 | 1.433561 | 14.690643 | DMGDH;GLTP;FAAH;LPGAT1;ECI1;PITPNB;ECI2;GDE1;RORA;CCNC;MSMO1;ZDHHC8;GBA2;LIPC;SCP2;LIPH;LIPG;OSBP;CPNE1;EP300;CPNE3;PIP4K2C;HADH;ACAA1;ACOT9;ARSA;CPT1A;PCYT2;DGAT1;MED8;ACOT12;ACSL5;ACOT13;OCRL;ACSL3;HSPG2;PISD;CYP39A1;ALDH3A2;TBL1XR1;GPD2;FDX1;ACOT2;PLBD1;ALDH7A1;ACOT4;CSNK1G2;PTGES;CDS1;MTMR1;SAR1B;ABHD3;INSIG2;INSIG1;PEX11A;PIK3R3;ACACB;MTMR6;NPAS2;ACACA;PLD3;ACAT1;UGT8;PTDSS2;PTDSS1;SGPL1;EBP;INPP5F;PTPMT1;PSAP;ARSK;NEU1;INPP5J;THEM4;SCAP;HACL1;LTA4H;ARSB;FDFT1;DECR1;ESRRA;WWTR1;CDK19;CHKB;CHKA;NR1D1;CYP4F11;MED13L;CYP1A2;CYP1A1;SDC1;SLC44A3;ACADVL;SLC44A4;SLC44A1;SLC44A2;CHD9;HEXB;HEXA;TECR;FHL2;MED19;PRKAG2;AKR1B1;PIK3C2G;TAZ;PIK3C2B;MCEE;MED13;SPTLC1;SPTLC2;ACADL;CPT2;MED10;SMPD1;HMGCS2;CD36;ACADM;TEAD1;LBR;TEAD2;NCOA1;ABCC3;ELOVL1;NCOA2;CBR1;CERS5;GPX2;AMN;ABCC1;NCOA3;PTGR1;SUMF2;SREBF2;SUMF1;HADHB;HADHA;NCOR1;BDH2;PCCA;MED21;CPTP;PCCB;PPARG;PLIN2;ACOX3;MGLL;CERS2;YAP1;AGPAT5;ASAH1;PRKAA2;SGMS1;HSD17B4;HSD17B12;HMGCR;HSD17B11;HSD17B7;AGPAT3;PTGS1;LMF1;CYP17A1;HMGCL;TGS1;CEPT1;SLC25A20;CDIPT;MBTPS1;AGK;PNPLA8;TXNRD1;MBOAT7;NFYC;AKR1C1;CYP5[illegible] |
| 158 | VEGFR2 mediated cell proliferation Homo sapiens R-HSA-5218921 | 3.76E-05 | 0.000339 | 1.758202 | 17.912568 | AHCYL1;YWHAB;RASGRF2;ITGA2B;PEBP1;WDR83;IRS2;ARRB1;PSMD8;PSMD6;PSMD7;PSMD4;RASGEF1A;PSMD2;PSMD3;PSMD1;JAK2;SPTAN1;JAK1;MAP2K1;MAP2K2;PRKCD;PPP2R5B;PSMA5;PSMA6;PSMA3;PSMA1;PSMA2;AKAP9;KIT;RASA2;IL3RA;PSME1;PSME2;TLN1;RAF1;SOS1;VCL;SPTBN5;PSMD12;SHC2;PSMD11;PSMD13;SRC;CUL3;ITPR2;ITPR3;IQGAP1;PHB;PSMA7;EGFR;PSMB10;PSMB6;RAP1B;PSMB4;PSMB5;PSMB2;UBB;ERBB3;PSMB3;PSMB1;UBC;ERBB2;MAPK1;CSK;PLCG1;PSMF1;SPTBN1;SPTBN2;MAPK3;DAB2IP;BRAF;PTK2;PSMB8;PSMB9;PSMC5;PSMC4;PTPRA;PSMC2;NF1;GRB2;CALM1;MAP3K11 |

| | | | | | | |
|---|---|---|---|---|---|---|
| 159 | Signaling by FGFR3 Homo sapiens R-HSA-5654741 | 3.91E-05 | 0.000349 | 1.634094 | 16.586812 | AHCYL1;ITGA2B;PEBP1;WDR83;IRS2;PSMD8;PSMD6;PSMD7;PSMD4;AKT2;PSMD2;PSMD3;AKT1;PSMD1;PIP4K2C;IER3;MAP2K1;MAP2K2;PRKCD;ADCY9;AKAP9;PSME1;PSME2;TLN1;RAF1;VCL;SPTBN5;PSMD12;SHC2;PSMD11;PSMD13;CUL3;PIK3R3;ITPR2;ITPR3;IQGAP1;PHB;RAP1B;UBB;PRKAR2A;UBC;THEM4;PLCG1;PSMF1;SPTBN1;SPTBN2;INSR;BRAF;PTK2;AGO3;PTPRA;NF1;GRB2;CALM1;MAP3K11;GSK3B;CDKN1B;YWHAB;RASGRF2;ARRB1;RASGEF1A;MLST8;JAK2;SPTAN1;JAK1;PPP2R5B;PSMA5;PSMA6;PSMA3;CREB1;PSMA1;PSMA2;KIT;RASA2;IL3RA;SOS1;SRC;CBL;ADCY6;PSMA7;EGFR;PSMB10;PSMB6;PSMB4;PSMB5;PSMB2;ERBB3;PSMB3;MKNK1;PSMB1;ERBB2;MAPK1;CSK;MAPK3;GALNT3;BAD;DAB2IP;PTPN11;PSMB8;MTOR;PSMB9;PSMC5;PSMC4;PSMC2;RPS6KB2;TNRC6A |
| 160 | Glycerophospholipid biosynthesis Homo sapiens R-HSA-1483206 | 3.94E-05 | 0.00035 | 2.342762 | 23.761404 | DMGDH;CDS1;AGPAT5;SLC44A3;LPGAT1;SLC44A4;SLC44A1;ABHD3;SLC44A2;PITPNB;TAZ;PLD3;AGPAT3;PTDSS2;PTDSS1;PTPMT1;CPNE1;CPNE3;CEPT1;CDIPT;PCYT2;CHKB;DGAT1;CHKA;PNPLA8;MBOAT7;DDHD2;MBOAT1;MBOAT2;PISD;HADHB;HADHA;LPCAT3;ETNK1;PLBD1;CHPT1;ALDH7A1;LPIN2;MGLL;LPIN3;PNPLA2 |
| 161 | G1/S Transition Homo sapiens R-HSA-69206 | 4.06E-05 | 0.000359 | 2.255875 | 22.81013 | RB1;PSMD12;PSMD11;CDKN1B;MAX;PSMD13;CUL1;PSMB10;PSMA7;CKS1B;PSMD8;PSMB6;PSMD6;PSMB4;PSMD7;PSMB5;PSMD4;UBB;PSMB2;PSMB3;PSMD2;PSMD3;UBC;PSMB1;PSMD1;TK2;PSMF1;RPA2;PSMB8;PSMB9;PSMA5;CCNA1;PSMA6;CDK7;PSMA3;PSMC5;PSMA1;PSMA2;PSMC4;RPA3;PSMC2;PSME1;MCM3;PSME2 |

| | | | | | | |
|---|---|---|---|---|---|---|
| 162 | Downstream signal transduction Homo sapiens R-HSA-186763 | 4.19E-05 | 0.000368 | 1.622158 | 16.34981 | AHCYL1;ITGA2B;PEBP1;WDR83;IRS2;PSMD8;PSMD6;PSMD7;PSMD4;AKT2;PSMD2;PSMD3;AKT1;PSMD1;PIP4K2C;IER3;MAP2K1;MAP2K2;PRKCD;ADCY9;AKAP9;PSME1;PSME2;TLN1;RAF1;VCL;GRB7;SPTBN5;PSMD12;SHC2;PSMD11;PSMD13;CUL3;PIK3R3;ITPR2;ITPR3;IQGAP1;PHB;RAP1B;UBB;PRKAR2A;UBC;THEM4;PLCG1;PSMF1;SPTBN1;SPTBN2;INSR;BRAF;PTK2;AGO3;PTPRA;NF1;GRB2;CALM1;MAP3K11;GSK3B;CDKN1B;YWHAB;RASGRF2;ARRB1;RASGEF1A;MLST8;JAK2;SPTAN1;JAK1;PPP2R5B;PSMA5;PSMA6;PSMA3;CREB1;PSMA1;PSMA2;KIT;RASA2;IL3RA;SOS1;SRC;ADCY6;PSMA7;EGFR;PSMB10;PSMB6;PSMB4;PSMB5;PSMB2;ERBB3;PSMB3;PSMB1;ERBB2;MAPK1;STAT6;CSK;MAPK3;STAT1;BAD;STAT3;DAB2IP;PTPN11;PSMB8;MTOR;PSMB9;PSMC5;PSMC4;PSMC2;RPS6KB2;TNRC6A;BCAR1 |
| 163 | Signaling by ERBB4 Homo sapiens R-HSA-1236394 | 4.32E-05 | 0.000377 | 1.632906 | 16.410277 | ITGA2B;PEBP1;WDR83;IRS2;PSMD8;PSMD6;PSMD7;PSMD4;AKT2;PSMD2;PSMD3;AKT1;PSMD1;PIP4K2C;IER3;MAP2K1;MAP2K2;ADAM17;AKAP9;PSME1;PSME2;TLN1;RAF1;VCL;SPTBN5;PSMD12;SHC2;PSMD11;PSMD13;CUL3;CUL1;PIK3R3;IQGAP1;PHB;APH1A;RAP1B;APH1B;UBB;UBC;THEM4;PSMF1;SPTBN1;SPTBN2;INSR;BRAF;ESR1;PTK2;AGO3;PTPRA;NF1;GRB2;CALM1;MAP3K11;GSK3B;CDKN1B;YWHAB;RASGRF2;ARRB1;RASGEF1A;MLST8;JAK2;SPTAN1;JAK1;PSENEN;PPP2R5B;PSMA5;PSMA6;PSMA3;NCOR1;CREB1;PSMA1;PSMA2;KIT;RASA2;IL3RA;SOS1;YAP1;SRC;PSMA7;EGFR;PSMB10;PSMB6;PSMB4;PSMB5;PSMB2;ERBB3;PSMB3;PSMB1;ERBB2;MAPK1;CSK;MAPK3;BAD;DAB2IP;WWP1;PTPN11;PSMB8;MTOR;PSMB9;PSMC5;ITCH;PSMC4;PSMC2;RPS6KB2;TNRC6A |
| 164 | Cohesin Loading onto Chromatin Homo sapiens R-HSA-2470946 | 4.52E-05 | 0.000391 | 27.78608 | 277.95632 | STAG1;NIPBL;STAG2;RAD21;PDS5B;PDS5A;SMC1A;SMC3 |
| 165 | Influenza Life Cycle Homo sapiens R-HSA-168255 | 4.54E-05 | 0.000391 | 2.092815 | 20.927947 | RPL5;RPL30;RPL31;RPLP1;RPL6;IPO5;RPS15;RPL7A;XPO1;RPS19;RPL36;RPLP2;RPL37;RPS10;KPNA1;NUP214;RPS9;HSP90AA1;RPS7;RPS8;RPS5;GTF2F2;DNAJC3;NUP50;RPL37A;NUP98;RPL29;NUP205;RPS4Y1;AAAS;POM121C;POLR2B;POLR2C;TPR;POLR2E;POLR2H;POLR2J;RPL19;RANBP2;RPL41;RPL35A;RPS25;RPS28;RPS27;RPS29;CALR;GRSF1;RPS21;RPS24;RAN;HSPA1A |

| | | | | | | |
|---|---|---|---|---|---|---|
| 166 | RHO GTPases activate IQGAPs Homo sapiens R-HSA-5626467 | 4.93E-05 | 0.000419 | 15.63216 | 155.04776 | CDC42;CLIP1;CDH1;CTNNB1;RAC1;IQGAP1;IQGAP2;CALM1;MEN1 |
| 167 | SeMet incorporation into proteins Homo sapiens R-HSA-2408517 | 4.93E-05 | 0.000419 | 15.63216 | 155.04776 | AIMP1;AIMP2;DARS;KARS;QARS;LARS;MARS;RARS;EPRS |
| 168 | FRS-mediated FGFR2 signaling Homo sapiens R-HSA-5654700 | 5.66E-05 | 0.000469 | 1.758057 | 17.191933 | YWHAB;RASGRF2;ITGA2B;PEBP1;WDR83;IRS2;ARRB1;PSMD8;PSMD6;PSMD7;PSMD4;RASGEF1A;PSMD2;PSMD3;PSMD1;JAK2;SPTAN1;JAK1;MAP2K1;MAP2K2;PPP2R5B;PSMA5;PSMA6;PSMA3;PSMA1;PSMA2;AKAP9;KIT;RASA2;IL3RA;PSME1;PSME2;TLN1;RAF1;SOS1;VCL;SPTBN5;PSMD12;SHC2;PSMD11;PSMD13;SRC;CUL3;IQGAP1;PHB;PSMA7;EGFR;PSMB10;PSMB6;RAP1B;PSMB4;PSMB5;PSMB2;UBB;ERBB3;PSMB3;PSMB1;UBC;ERBB2;MAPK1;CSK;PSMF1;SPTBN1;SPTBN2;MAPK3;DAB2IP;BRAF;PTPN11;PTK2;PSMB8;PSMB9;PSMC5;PSMC4;PTPRA;PSMC2;NF1;GRB2;CALM1;MAP3K11 |
| 169 | FRS-mediated FGFR4 signaling Homo sapiens R-HSA-5654712 | 5.66E-05 | 0.000469 | 1.758057 | 17.191933 | YWHAB;RASGRF2;ITGA2B;PEBP1;WDR83;IRS2;ARRB1;PSMD8;PSMD6;PSMD7;PSMD4;RASGEF1A;PSMD2;PSMD3;PSMD1;JAK2;SPTAN1;JAK1;MAP2K1;MAP2K2;PPP2R5B;PSMA5;PSMA6;PSMA3;PSMA1;PSMA2;AKAP9;KIT;RASA2;IL3RA;PSME1;PSME2;TLN1;RAF1;SOS1;VCL;SPTBN5;PSMD12;SHC2;PSMD11;PSMD13;SRC;CUL3;IQGAP1;PHB;PSMA7;EGFR;PSMB10;PSMB6;RAP1B;PSMB4;PSMB5;PSMB2;UBB;ERBB3;PSMB3;PSMB1;UBC;ERBB2;MAPK1;CSK;PSMF1;SPTBN1;SPTBN2;MAPK3;DAB2IP;BRAF;PTPN11;PTK2;PSMB8;PSMB9;PSMC5;PSMC4;PTPRA;PSMC2;NF1;GRB2;CALM1;MAP3K11 |
| 170 | FRS-mediated FGFR3 signaling Homo sapiens R-HSA-5654706 | 5.66E-05 | 0.000469 | 1.758057 | 17.191933 | YWHAB;RASGRF2;ITGA2B;PEBP1;WDR83;IRS2;ARRB1;PSMD8;PSMD6;PSMD7;PSMD4;RASGEF1A;PSMD2;PSMD3;PSMD1;JAK2;SPTAN1;JAK1;MAP2K1;MAP2K2;PPP2R5B;PSMA5;PSMA6;PSMA3;PSMA1;PSMA2;AKAP9;KIT;RASA2;IL3RA;PSME1;PSME2;TLN1;RAF1;SOS1;VCL;SPTBN5;PSMD12;SHC2;PSMD11;PSMD13;SRC;CUL3;IQGAP1;PHB;PSMA7;EGFR;PSMB10;PSMB6;RAP1B;PSMB4;PSMB5;PSMB2;UBB;ERBB3;PSMB3;PSMB1;UBC;ERBB2;MAPK1;CSK;PSMF1;SPTBN1;SPTBN2;MAPK3;DAB2IP;BRAF;PTPN11;PTK2;PSMB8;PSMB9;PSMC5;PSMC4;PTPRA;PSMC2;NF1;GRB2;CALM1;MAP3K11 |

| | | | | | | |
|---|---|---|---|---|---|---|
| 171 | FRS-mediated FGFR1 signaling Homo sapiens R-HSA-5654693 | 5.66E-05 | 0.000469 | 1.758057 | 17.191933 | YWHAB;RASGRF2;ITGA2B;PEBP1;WDR83;IRS2;ARRB1;PSMD8;PSMD6;PSMD7;PSMD4;RASGEF1A;PSMD2;PSMD3;PSMD1;JAK2;SPTAN1;JAK1;MAP2K1;MAP2K2;PPP2R5B;PSMA5;PSMA6;PSMA3;PSMA1;PSMA2;AKAP9;KIT;RASA2;IL3RA;PSME1;PSME2;TLN1;RAF1;SOS1;VCL;SPTBN5;PSMD12;SHC2;PSMD11;PSMD13;SRC;CUL3;IQGAP1;PHB;PSMA7;EGFR;PSMB10;PSMB6;RAP1B;PSMB4;PSMB5;PSMB2;UBB;ERBB3;PSMB3;PSMB1;UBC;ERBB2;MAPK1;CSK;PSMF1;SPTBN1;SPTBN2;MAPK3;DAB2IP;BRAF;PTPN11;PTK2;PSMB8;PSMB9;PSMC5;PSMC4;PTPRA;PSMC2;NF1;GRB2;CALM1;MAP3K11 |
| 172 | Signaling by FGFR4 Homo sapiens R-HSA-5654743 | 5.67E-05 | 0.000469 | 1.618308 | 15.823645 | AHCYL1;ITGA2B;PEBP1;WDR83;IRS2;PSMD8;PSMD6;PSMD7;PSMD4;AKT2;PSMD2;PSMD3;AKT1;PSMD1;PIP4K2C;IER3;MAP2K1;MAP2K2;PRKCD;ADCY9;AKAP9;PSME1;PSME2;TLN1;RAF1;VCL;SPTBN5;PSMD12;SHC2;PSMD11;PSMD13;CUL3;PIK3R3;ITPR2;ITPR3;IQGAP1;PHB;RAP1B;UBB;PRKAR2A;UBC;THEM4;PLCG1;PSMF1;SPTBN1;SPTBN2;INSR;BRAF;PTK2;AGO3;PTPRA;NF1;GRB2;CALM1;MAP3K11;GSK3B;CDKN1B;YWHAB;RASGRF2;ARRB1;RASGEF1A;MLST8;JAK2;SPTAN1;JAK1;PPP2R5B;PSMA5;PSMA6;PSMA3;CREB1;PSMA1;PSMA2;KIT;RASA2;IL3RA;SOS1;SRC;CBL;ADCY6;PSMA7;EGFR;PSMB10;PSMB6;PSMB4;PSMB5;PSMB2;ERBB3;PSMB3;MKNK1;PSMB1;ERBB2;MAPK1;CSK;MAPK3;BAD;DAB2IP;PTPN11;PSMB8;MTOR;PSMB9;PSMC5;PSMC4;PSMC2;RPS6KB2;TNRC6A |
| 173 | Beta-catenin independent WNT signaling Homo sapiens R-HSA-3858494 | 5.85E-05 | 0.000481 | 2.031616 | 19.801355 | CLTC;CLTB;AP2A1;PSMD8;PSMD6;PSMD7;PSMD4;PSMD2;PSMD3;PSMD1;RAC1;AP2M1;WNT5A;RHOA;PSMA5;PSMA6;PSMA3;DAAM1;PSMA1;PSMA2;PSME1;PSME2;PSMD12;PSMD11;PSMD13;ITPR2;ITPR3;PSMB10;PSMA7;PSMB6;PSMB4;PSMB5;UBB;GNG5;PSMB2;PSMB3;UBC;PSMB1;DVL3;PSMF1;FZD5;RYK;FZD6;GNG11;PSMB8;PSMB9;PSMC5;AGO3;PSMC4;PSMC2;CTNNB1;CALM1;TNRC6A |
| 174 | TP53 Regulates Metabolic Genes Homo sapiens R-HSA-5628897 | 5.90E-05 | 0.000482 | 2.488261 | 24.232261 | YWHAE;COX19;PRKAA1;PRKAA2;COX16;YWHAB;PRKAG2;COX6A1;COX7C;COX5A;GLS;SESN3;AKT2;PRDX1;MLST8;AKT1;COX14;YWHAH;COX8A;GPX2;SURF1;TXNRD1;GSR;COX6C;LRPPRC;MTOR;COX6B1;RRAGA;RRM2B;AGO3;SCO2;DDIT4;LAMTOR1;TNRC6A;COX20 |

| | | | | | | |
|---|---|---|---|---|---|---|
| 175 | Signaling by the B Cell Receptor (BCR) Homo sapiens R-HSA-983705 | 6.27E-05 | 0.00051 | 1.75803 | 17.011735 | GSK3B;AHCYL1;CDKN1B;IRS2;PSMD8;PSMD6;PSMD7;PSMD4;AKT2;PSMD2;PSMD3;MLST8;BLNK;AKT1;PSMD1;PIP4K2C;IER3;FBXW11;PPP2R5B;PSMA5;PSMA6;PSMA3;CREB1;PSMA1;STIM1;PSMA2;KIT;PSME1;ORAI1;PSME2;SOS1;PSMD12;PSMD11;PSMD13;SRC;CUL1;PIK3R3;ITPR2;ITPR3;BCL10;CBL;PSMA7;RELA;EGFR;PSMB10;MALT1;PSMB6;PSMB4;PSMB5;PSMB2;UBB;ERBB3;PSMB3;PSMB1;UBC;ERBB2;THEM4;MAPK1;PLCG1;PSMF1;MAPK3;BAD;INSR;PTPN11;PSMB8;NFKB1;MTOR;PSMB9;NFKBIA;PSMC5;AGO3;PSMC4;PSMC2;RPS6KB2;REL;GRB2;CALM1;TNRC6A |
| 176 | RHO GTPase Effectors Homo sapiens R-HSA-195258 | 6.39E-05 | 0.000517 | 1.71667 | 16.578861 | YWHAE;ITGB1;CYFIP1;NCKAP1;AHCTF1;CDKN1B;RTKN;KDM1A;YWHAB;ARPC1B;ARPC1A;WASL;ACTG1;XPO1;CDH1;KIF5B;RAC1;HIST1H2AC;MEN1;YWHAH;ACTR3;NCOA2;ACTR2;PPP1R12A;H2AFZ;PPP2R5B;RHOD;RHOA;CIT;MYL6;CLIP1;DAAM1;CTTN;KIF2A;MYH9;TAX1BP3;EVL;NUP98;MAPRE1;RHOQ;PFN2;PAFAH1B1;ROCK1;ROCK2;SRC;NOXO1;IQGAP1;IQGAP2;PMF1;MYL12B;SKA2;CENPC;CDC42;KLC2;MYH14;DVL3;MAPK1;RHPN2;PAK2;MYH10;MAPK3;CENPT;RANBP2;LIMK2;H3F3A;ARPC4;PTK2;KTN1;DIAPH1;DIAPH2;RPS27;FMNL2;ABI2;ARPC2;ARPC3;NOXA1;CTNNB1;GRB2;PKN2;GOPC;PKN1;NF2;CALM1;MAD2L1 |
| 177 | MAPK1/MAPK3 signaling Homo sapiens R-HSA-5684996 | 7.04E-05 | 0.000566 | 1.736023 | 16.597608 | YWHAB;RASGRF2;ITGA2B;PEBP1;WDR83;IRS2;ARRB1;PSMD8;PSMD6;PSMD7;PSMD4;RASGEF1A;PSMD2;PSMD3;PSMD1;JAK2;SPTAN1;JAK1;MAP2K1;MAP2K2;PPP2R5B;PSMA5;PSMA6;PSMA3;PSMA1;PSMA2;AKAP9;KIT;RASA2;IL3RA;PSME1;PSME2;TLN1;IL6ST;RAF1;SOS1;VCL;SPTBN5;PSMD12;SHC2;PSMD11;PSMD13;SRC;CUL3;IQGAP1;PHB;PSMA7;EGFR;PSMB10;PSMB6;RAP1B;PSMB4;PSMB5;PSMB2;UBB;ERBB3;PSMB3;PSMB1;UBC;ERBB2;MAPK1;CSK;PSMF1;SPTBN1;SPTBN2;MAPK3;DAB2IP;BRAF;PTPN11;PTK2;PSMB8;PSMB9;PSMC5;PSMC4;PTPRA;PSMC2;NF1;GRB2;CALM1;MAP3K11 |
| 178 | Endosomal Sorting Complex Required For Transport (ESCRT) Homo sapiens R-HSA-917729 | 7.11E-05 | 0.000568 | 4.636031 | 44.283039 | TSG101;VPS4B;VPS4A;VPS37D;VPS37B;SNF8;UBB;UBC;CHMP2B;CHMP4C;CHMP3;CHMP4B;VPS36;STAM2;VPS28;CHMP5 |

| | | | | | | |
|---|---|---|---|---|---|---|
| 179 | Signaling by NOTCH1 Homo sapiens R-HSA-1980143 | 7.30E-05 | 0.00058 | 2.632907 | 25.077371 | HDAC2;HDAC1;CUL1;CCNC;ARRB1;RBPJ;HDAC8;HDAC9;HIF1A;HDAC6;APH1A;NEURL1B;APH1B;HEY1;UBB;UBC;EP300;HES1;PSENEN;TLE3;TLE2;MIB2;ADAM10;KAT2A;ADAM17;ITCH;NCOR1;SNW1;TBL1XR1;NUMB;MAML3 |
| 180 | tRNA Aminoacylation Homo sapiens R-HSA-379724 | 7.86E-05 | 0.000621 | 3.478905 | 32.877562 | CARS;YARS;DARS;VARS;SARS;EPRS;NARS;AIMP1;AIMP2;PPA2;KARS;QARS;LARS;RARS;MARS;NARS2;IARS2;GARS;HARS;FARSA;CARS2 |
| 181 | Cytosolic sensors of pathogen-associated DNA Homo sapiens R-HSA-1834949 | 7.92E-05 | 0.000623 | 2.728794 | 25.767992 | APP;CRCP;DHX9;PRKDC;HMGB1;DDX41;RELA;TBK1;UBB;DHX36;UBC;POLR2E;EP300;STAT6;RIPK1;LRRFIP1;POLR2H;XRCC6;POLR3GL;XRCC5;TICAM1;NFKB1;NFKBIA;IRF3;POLR3B;IRF7;POLR1D;CTNNB1;TRIM56 |
| 182 | Prolonged ERK activation events Homo sapiens R-HSA-169893 | 8.24E-05 | 0.000644 | 1.725194 | 16.222813 | YWHAB;RASGRF2;ITGA2B;PEBP1;WDR83;IRS2;ARRB1;PSMD8;PSMD6;PSMD7;PSMD4;RASGEF1A;PSMD2;PSMD3;PSMD1;JAK2;SPTAN1;JAK1;MAP2K1;MAP2K2;PPP2R5B;PSMA5;PSMA6;PSMA3;PSMA1;PSMA2;AKAP9;KIT;RASA2;IL3RA;PSME1;PSME2;TLN1;RAF1;SOS1;VCL;SPTBN5;PSMD12;SHC2;PSMD11;PSMD13;SRC;CUL3;IQGAP1;PHB;PSMA7;EGFR;PSMB10;PSMB6;RAP1B;PSMB4;PSMB5;PSMB2;UBB;ERBB3;PSMB3;PSMB1;UBC;ERBB2;MAPK1;CSK;PLCG1;PSMF1;SPTBN1;SPTBN2;MAPK3;KIDINS220;DAB2IP;BRAF;PTK2;PSMB8;PSMB9;PSMC5;PSMC4;PTPRA;PSMC2;NF1;GRB2;CALM1;MAP3K11 |
| 183 | Golgi Associated Vesicle Biogenesis Homo sapiens R-HSA-432722 | 8.63E-05 | 0.000654 | 3.000201 | 28.076486 | RAB5C;TFRC;CLTC;AP4E1;ARRB1;CLINT1;TGOLN2;SNX2;AP1G1;SNX9;SNX5;AP1M2;HSPA8;SORT1;DTNBP1;HIP1R;OCRL;AP3B1;PUM1;TPD52L1;IGF2R;DNM2;VAMP8;YIPF6;FTL |
| 184 | GRB2 events in EGFR signaling Homo sapiens R-HSA-179812 | 8.65E-05 | 0.000654 | 1.735409 | 16.236202 | YWHAB;RASGRF2;ITGA2B;PEBP1;WDR83;IRS2;ARRB1;PSMD8;PSMD6;PSMD7;PSMD4;RASGEF1A;PSMD2;PSMD3;PSMD1;JAK2;SPTAN1;JAK1;MAP2K1;MAP2K2;PPP2R5B;PSMA5;PSMA6;PSMA3;PSMA1;PSMA2;AKAP9;KIT;RASA2;IL3RA;PSME1;PSME2;TLN1;RAF1;SOS1;VCL;SPTBN5;PSMD12;SHC2;PSMD11;PSMD13;SRC;CUL3;IQGAP1;PHB;PSMA7;EGFR;PSMB10;PSMB6;RAP1B;PSMB4;PSMB5;PSMB2;UBB;ERBB3;PSMB3;PSMB1;UBC;ERBB2;MAPK1;CSK;PSMF1;SPTBN1;SPTBN2;MAPK3;DAB2IP;BRAF;PTK2;PSMB8;PSMB9;PSMC5;PSMC4;PTPRA;PSMC2;NF1;GRB2;CALM1;MAP3K11 |

| | | | | | | |
|---|---|---|---|---|---|---|
| 185 | SHC1 events in EGFR signaling Homo sapiens R-HSA-180336 | 8.65E-05 | 0.000654 | 1.735409 | 16.236202 | YWHAB;RASGRF2;ITGA2B;PEBP1;WDR83;IRS2;ARRB1;PSMD8;PSMD6;PSMD7;PSMD4;RASGEF1A;PSMD2;PSMD3;PSMD1;JAK2;SPTAN1;JAK1;MAP2K1;MAP2K2;PPP2R5B;PSMA5;PSMA6;PSMA3;PSMA1;PSMA2;AKAP9;KIT;RASA2;IL3RA;PSME1;PSME2;TLN1;RAF1;SOS1;VCL;SPTBN5;PSMD12;SHC2;PSMD11;PSMD13;SRC;CUL3;IQGAP1;PHB;PSMA7;EGFR;PSMB10;PSMB6;RAP1B;PSMB4;PSMB5;PSMB2;UBB;ERBB3;PSMB3;PSMB1;UBC;ERBB2;MAPK1;CSK;PSMF1;SPTBN1;SPTBN2;MAPK3;DAB2IP;BRAF;PTK2;PSMB8;PSMB9;PSMC5;PSMC4;PTPRA;PSMC2;NF1;GRB2;CALM1;MAP3K11 |
| 186 | SOS-mediated signalling Homo sapiens R-HSA-112412 | 8.65E-05 | 0.000654 | 1.735409 | 16.236202 | YWHAB;RASGRF2;ITGA2B;PEBP1;WDR83;IRS2;ARRB1;PSMD8;PSMD6;PSMD7;PSMD4;RASGEF1A;PSMD2;PSMD3;PSMD1;JAK2;SPTAN1;JAK1;MAP2K1;MAP2K2;PPP2R5B;PSMA5;PSMA6;PSMA3;PSMA1;PSMA2;AKAP9;KIT;RASA2;IL3RA;PSME1;PSME2;TLN1;RAF1;SOS1;VCL;SPTBN5;PSMD12;SHC2;PSMD11;PSMD13;SRC;CUL3;IQGAP1;PHB;PSMA7;EGFR;PSMB10;PSMB6;RAP1B;PSMB4;PSMB5;PSMB2;UBB;ERBB3;PSMB3;PSMB1;UBC;ERBB2;MAPK1;CSK;PSMF1;SPTBN1;SPTBN2;MAPK3;DAB2IP;BRAF;PTK2;PSMB8;PSMB9;PSMC5;PSMC4;PTPRA;PSMC2;NF1;GRB2;CALM1;MAP3K11 |
| 187 | SHC1 events in ERBB4 signaling Homo sapiens R-HSA-1250347 | 8.65E-05 | 0.000654 | 1.735409 | 16.236202 | YWHAB;RASGRF2;ITGA2B;PEBP1;WDR83;IRS2;ARRB1;PSMD8;PSMD6;PSMD7;PSMD4;RASGEF1A;PSMD2;PSMD3;PSMD1;JAK2;SPTAN1;JAK1;MAP2K1;MAP2K2;PPP2R5B;PSMA5;PSMA6;PSMA3;PSMA1;PSMA2;AKAP9;KIT;RASA2;IL3RA;PSME1;PSME2;TLN1;RAF1;SOS1;VCL;SPTBN5;PSMD12;SHC2;PSMD11;PSMD13;SRC;CUL3;IQGAP1;PHB;PSMA7;EGFR;PSMB10;PSMB6;RAP1B;PSMB4;PSMB5;PSMB2;UBB;ERBB3;PSMB3;PSMB1;UBC;ERBB2;MAPK1;CSK;PSMF1;SPTBN1;SPTBN2;MAPK3;DAB2IP;BRAF;PTK2;PSMB8;PSMB9;PSMC5;PSMC4;PTPRA;PSMC2;NF1;GRB2;CALM1;MAP3K11 |

| | | | | | | |
|---|---|---|---|---|---|---|
| 188 | RAF/MAP kinase cascade Homo sapiens R-HSA-5673001 | 8.65E-05 | 0.000654 | 1.735409 | 16.236202 | YWHAB;RASGRF2;ITGA2B;PEBP1;WDR83;IRS2;ARRB1;PSMD8;PSMD6;PSMD7;PSMD4;RASGEF1A;PSMD2;PSMD3;PSMD1;JAK2;SPTAN1;JAK1;MAP2K1;MAP2K2;PPP2R5B;PSMA5;PSMA6;PSMA3;PSMA1;PSMA2;AKAP9;KIT;RASA2;IL3RA;PSME1;PSME2;TLN1;RAF1;SOS1;VCL;SPTBN5;PSMD12;SHC2;PSMD11;PSMD13;SRC;CUL3;IQGAP1;PHB;PSMA7;EGFR;PSMB10;PSMB6;RAP1B;PSMB4;PSMB5;PSMB2;UBB;ERBB3;PSMB3;PSMB1;UBC;ERBB2;MAPK1;CSK;PSMF1;SPTBN1;SPTBN2;MAPK3;DAB2IP;BRAF;PTK2;PSMB8;PSMB9;PSMC5;PSMC4;PTPRA;PSMC2;NF1;GRB2;CALM1;MAP3K11 |
| 189 | ARMS-mediated activation Homo sapiens R-HSA-170984 | 9.13E-05 | 0.000687 | 1.724757 | 16.04203 | YWHAB;RASGRF2;ITGA2B;PEBP1;WDR83;IRS2;ARRB1;PSMD8;PSMD6;PSMD7;PSMD4;RASGEF1A;PSMD2;PSMD3;PSMD1;JAK2;SPTAN1;JAK1;MAP2K1;MAP2K2;PPP2R5B;PSMA5;PSMA6;PSMA3;PSMA1;PSMA2;AKAP9;KIT;RASA2;IL3RA;PSME1;PSME2;TLN1;RAF1;SOS1;VCL;SPTBN5;PSMD12;SHC2;PSMD11;PSMD13;SRC;CUL3;IQGAP1;PHB;PSMA7;EGFR;PSMB10;PSMB6;RAP1B;PSMB4;PSMB5;PSMB2;UBB;ERBB3;PSMB3;PSMB1;UBC;ERBB2;MAPK1;CSK;PSMF1;SPTBN1;SPTBN2;MAPK3;KIDINS220;DAB2IP;BRAF;PTK2;PSMB8;PSMB9;PSMC5;PSMC4;PTPRA;PSMC2;NF1;GRB2;CALM1;MAP3K11 |
| 190 | Signaling by FGFR1 Homo sapiens R-HSA-5654736 | 9.61E-05 | 0.000717 | 1.589869 | 14.707105 | AHCYL1;ITGA2B;PEBP1;WDR83;IRS2;PSMD8;PSMD6;PSMD7;PSMD4;AKT2;PSMD2;PSMD3;AKT1;PSMD1;PIP4K2C;IER3;MAP2K1;MAP2K2;PRKCD;ADCY9;AKAP9;PSME1;PSME2;TLN1;RAF1;VCL;SPTBN5;PSMD12;SHC2;PSMD11;PSMD13;CUL3;PIK3R3;ITPR2;ITPR3;IQGAP1;PHB;RAP1B;UBB;PRKAR2A;UBC;THEM4;PLCG1;PSMF1;SPTBN1;SPTBN2;INSR;BRAF;PTK2;AGO3;PTPRA;NF1;GRB2;CALM1;MAP3K11;GSK3B;CDKN1B;YWHAB;RASGRF2;ARRB1;RASGEF1A;MLST8;JAK2;SPTAN1;JAK1;PPP2R5B;PSMA5;PSMA6;PSMA3;CREB1;PSMA1;PSMA2;KIT;RASA2;IL3RA;SOS1;SRC;CBL;ADCY6;PSMA7;EGFR;PSMB10;PSMB6;PSMB4;PSMB5;PSMB2;ERBB3;PSMB3;MKNK1;PSMB1;ERBB2;MAPK1;CSK;MAPK3;BAD;DAB2IP;PTPN11;PSMB8;MTOR;PSMB9;PSMC5;PSMC4;PSMC2;RPS6KB2;TNRC6A |

| | | | | | | |
|---|---|---|---|---|---|---|
| 191 | Signaling by Leptin Homo sapiens R-HSA-2586552 | 9.63E-05 | 0.000717 | 1.714499 | 15.856068 | YWHAB;RASGRF2;ITGA2B;PEBP1;WDR83;IRS2;ARRB1;PSMD8;PSMD6;PSMD7;PSMD4;RASGEF1A;PSMD2;PSMD3;PSMD1;JAK2;SPTAN1;JAK1;MAP2K1;MAP2K2;PPP2R5B;PSMA5;PSMA6;PSMA3;PSMA1;PSMA2;AKAP9;KIT;RASA2;IL3RA;PSME1;PSME2;TLN1;RAF1;SOS1;VCL;SPTBN5;PSMD12;SHC2;PSMD11;PSMD13;SRC;CUL3;IQGAP1;PHB;PSMA7;EGFR;PSMB10;PSMB6;RAP1B;PSMB4;PSMB5;PSMB2;UBB;ERBB3;PSMB3;PSMB1;UBC;ERBB2;MAPK1;CSK;PSMF1;SPTBN1;SPTBN2;MAPK3;STAT3;DAB2IP;BRAF;PTPN11;PTK2;PSMB8;PSMB9;PSMC5;PSMC4;PTPRA;PSMC2;NF1;GRB2;CALM1;MAP3K11 |
| 192 | Signaling by SCF-KIT Homo sapiens R-HSA-1433557 | 9.99E-05 | 0.00074 | 1.59959 | 14.734983 | ITGA2B;PEBP1;WDR83;IRS2;PSMD8;PSMD6;PSMD7;PSMD4;AKT2;PSMD2;PSMD3;AKT1;PSMD1;PIP4K2C;IER3;MAP2K1;MAP2K2;AKAP9;PSME1;PSME2;TLN1;RAF1;VCL;GRB7;SPTBN5;PSMD12;SHC2;PSMD11;PSMD13;CUL3;PIK3R3;IQGAP1;PHB;RAP1B;UBB;UBC;THEM4;PSMF1;SPTBN1;SPTBN2;YES1;INSR;BRAF;PTK2;FER;AGO3;PTPRA;NF1;GRB2;CALM1;MAP3K11;GSK3B;CDKN1B;YWHAB;RASGRF2;ARRB1;RASGEF1A;MLST8;RAC1;JAK2;SPTAN1;JAK1;PPP2R5B;PSMA5;PSMA6;PSMA3;CREB1;PSMA1;PSMA2;KIT;RASA2;IL3RA;SOS1;SRC;CBL;PSMA7;EGFR;PSMB10;PSMB6;PSMB4;PSMB5;PSMB2;ERBB3;PSMB3;PSMB1;ERBB2;MAPK1;CSK;MAPK3;STAT1;BAD;STAT3;DAB2IP;PTPN11;PSMB8;MTOR;PSMB9;PSMC5;PSMC4;PSMC2;RPS6KB2;TNRC6A |
| 193 | DAP12 signaling Homo sapiens R-HSA-2424491 | 0.000102 | 0.000743 | 1.579239 | 14.517825 | AHCYL1;ITGA2B;PEBP1;WDR83;IRS2;PSMD8;PSMD6;PSMD7;PSMD4;AKT2;PSMD2;PSMD3;AKT1;PSMD1;PIP4K2C;B2M;IER3;MAP2K1;MAP2K2;PRKCD;HLA-E;ADCY9;AKAP9;PSME1;PSME2;TLN1;RAF1;VCL;SPTBN5;PSMD12;SHC2;PSMD11;PSMD13;CUL3;PIK3R3;ITPR2;ITPR3;IQGAP1;PHB;RAP1B;UBB;PRKAR2A;UBC;THEM4;PLCG1;PSMF1;SPTBN1;SPTBN2;INSR;BRAF;PTK2;AGO3;PTPRA;NF1;GRB2;CALM1;MAP3K11;GSK3B;CDKN1B;YWHAB;RASGRF2;ARRB1;RASGEF1A;MLST8;RAC1;JAK2;SPTAN1;JAK1;VAV3;PPP2R5B;PSMA5;PSMA6;PSMA3;CREB1;PSMA1;PSMA2;KIT;RASA2;IL3RA;SOS1;SRC;ADCY6;PSMA7;EGFR;PSMB10;PSMB6;PSMB4;PSMB5;PSMB2;ERBB3;PSMB3;PSMB1;ERBB2;MAPK1;CSK;MAPK3;BAD;DAB2IP;PTPN11;PSMB8;MTOR;PSMB9;PSMC5;PSMC4;PSMC2;RPS6KB2;TNRC6A |

| | | | | | | |
|---|---|---|---|---|---|---|
| 194 | MAPK family signaling cascades Homo sapiens R-HSA-5683057 | 0.000102 | 0.000743 | 1.648002 | 15.147572 | YWHAB;RASGRF2;ITGA2B;PEBP1;HSPB1;WDR83;IRS2;ARRB1;CDC14B;PSMD8;PSMD6;CCND3;PSMD7;XPO1;PSMD4;RASGEF1A;PSMD2;PSMD3;PSMD1;RAC1;JAK2;SPTAN1;JAK1;MAP2K1;MAP2K2;NCOA3;PPP2R5B;PSMA5;PSMA6;PSMA3;PSMA1;PSMA2;AKAP9;KIT;RASA2;IL3RA;PSME1;CDC42EP2;PSME2;TLN1;IL6ST;RAF1;SOS1;VCL;SPTBN5;PSMD12;SHC2;PSMD11;PSMD13;SRC;CUL3;IQGAP1;PHB;PSMA7;EGFR;PSMB10;PSMB6;RAP1B;CDC42;PSMB4;PSMB5;PSMB2;UBB;ERBB3;PSMB3;PSMB1;UBC;ERBB2;MAPK1;CSK;PSMF1;PAK2;SPTBN1;SPTBN2;MAPK3;DAB2IP;BRAF;PTPN11;PTK2;PSMB8;PSMB9;PSMC5;AGO3;PSMC4;PTPRA;PSMC2;NF1;GRB2;CALM1;MAP3K11;TNRC6A |
| 195 | Downstream signaling of activated FGFR2 Homo sapiens R-HSA-5654696 | 0.000103 | 0.000743 | 1.593882 | 14.633288 | AHCYL1;ITGA2B;PEBP1;WDR83;IRS2;PSMD8;PSMD6;PSMD7;PSMD4;AKT2;PSMD2;PSMD3;AKT1;PSMD1;PIP4K2C;IER3;MAP2K1;MAP2K2;PRKCD;ADCY9;AKAP9;PSME1;PSME2;TLN1;RAF1;VCL;SPTBN5;PSMD12;SHC2;PSMD11;PSMD13;CUL3;PIK3R3;ITPR2;ITPR3;IQGAP1;PHB;RAP1B;UBB;PRKAR2A;UBC;THEM4;PLCG1;PSMF1;SPTBN1;SPTBN2;INSR;BRAF;PTK2;AGO3;PTPRA;NF1;GRB2;CALM1;MAP3K11;GSK3B;CDKN1B;YWHAB;RASGRF2;ARRB1;RASGEF1A;MLST8;JAK2;SPTAN1;JAK1;PPP2R5B;PSMA5;PSMA6;PSMA3;CREB1;PSMA1;PSMA2;KIT;RASA2;IL3RA;SOS1;SRC;ADCY6;PSMA7;EGFR;PSMB10;PSMB6;PSMB4;PSMB5;PSMB2;ERBB3;PSMB3;PSMB1;ERBB2;MAPK1;CSK;MAPK3;BAD;DAB2IP;PTPN11;PSMB8;MTOR;PSMB9;PSMC5;PSMC4;PSMC2;RPS6KB2;TNRC6A |

| | | | | | | |
|---|---|---|---|---|---|---|
| 196 | Downstream signaling of activated FGFR4 Homo sapiens R-HSA-5654716 | 0.000103 | 0.000743 | 1.593882 | 14.633288 | AHCYL1;ITGA2B;PEBP1;WDR83;IRS2;PSMD8;PSMD6;PSMD7;PSMD4;AKT2;PSMD2;PSMD3;AKT1;PSMD1;PIP4K2C;IER3;MAP2K1;MAP2K2;PRKCD;ADCY9;AKAP9;PSME1;PSME2;TLN1;RAF1;VCL;SPTBN5;PSMD12;SHC2;PSMD11;PSMD13;CUL3;PIK3R3;ITPR2;ITPR3;IQGAP1;PHB;RAP1B;UBB;PRKAR2A;UBC;THEM4;PLCG1;PSMF1;SPTBN1;SPTBN2;INSR;BRAF;PTK2;AGO3;PTPRA;NF1;GRB2;CALM1;MAP3K11;GSK3B;CDKN1B;YWHAB;RASGRF2;ARRB1;RASGEF1A;MLST8;JAK2;SPTAN1;JAK1;PPP2R5B;PSMA5;PSMA6;PSMA3;CREB1;PSMA1;PSMA2;KIT;RASA2;IL3RA;SOS1;SRC;ADCY6;PSMA7;EGFR;PSMB10;PSMB6;PSMB4;PSMB5;PSMB2;ERBB3;PSMB3;PSMB1;ERBB2;MAPK1;CSK;MAPK3;BAD;DAB2IP;PTPN11;PSMB8;MTOR;PSMB9;PSMC5;PSMC4;PSMC2;RPS6KB2;TNRC6A |
| 197 | Downstream signaling of activated FGFR3 Homo sapiens R-HSA-5654708 | 0.000103 | 0.000743 | 1.593882 | 14.633288 | AHCYL1;ITGA2B;PEBP1;WDR83;IRS2;PSMD8;PSMD6;PSMD7;PSMD4;AKT2;PSMD2;PSMD3;AKT1;PSMD1;PIP4K2C;IER3;MAP2K1;MAP2K2;PRKCD;ADCY9;AKAP9;PSME1;PSME2;TLN1;RAF1;VCL;SPTBN5;PSMD12;SHC2;PSMD11;PSMD13;CUL3;PIK3R3;ITPR2;ITPR3;IQGAP1;PHB;RAP1B;UBB;PRKAR2A;UBC;THEM4;PLCG1;PSMF1;SPTBN1;SPTBN2;INSR;BRAF;PTK2;AGO3;PTPRA;NF1;GRB2;CALM1;MAP3K11;GSK3B;CDKN1B;YWHAB;RASGRF2;ARRB1;RASGEF1A;MLST8;JAK2;SPTAN1;JAK1;PPP2R5B;PSMA5;PSMA6;PSMA3;CREB1;PSMA1;PSMA2;KIT;RASA2;IL3RA;SOS1;SRC;ADCY6;PSMA7;EGFR;PSMB10;PSMB6;PSMB4;PSMB5;PSMB2;ERBB3;PSMB3;PSMB1;ERBB2;MAPK1;CSK;MAPK3;BAD;DAB2IP;PTPN11;PSMB8;MTOR;PSMB9;PSMC5;PSMC4;PSMC2;RPS6KB2;TNRC6A |
| 198 | Frs2-mediated activation Homo sapiens R-HSA-170968 | 0.000107 | 0.000766 | 1.713933 | 15.675287 | YWHAB;RASGRF2;ITGA2B;PEBP1;WDR83;IRS2;ARRB1;PSMD8;PSMD6;PSMD7;PSMD4;RASGEF1A;PSMD2;PSMD3;PSMD1;JAK2;SPTAN1;JAK1;MAP2K1;MAP2K2;PPP2R5B;PSMA5;PSMA6;PSMA3;PSMA1;PSMA2;AKAP9;KIT;RASA2;IL3RA;PSME1;PSME2;TLN1;RAF1;SOS1;VCL;SPTBN5;PSMD12;SHC2;PSMD11;PSMD13;SRC;CUL3;IQGAP1;PHB;PSMA7;EGFR;PSMB10;PSMB6;RAP1B;PSMB4;PSMB5;PSMB2;UBB;ERBB3;PSMB3;PSMB1;UBC;ERBB2;MAPK1;CSK;PLCG1;PSMF1;SPTBN1;SPTBN2;MAPK3;DAB2IP;BRAF;PTK2;PSMB8;PSMB9;PSMC5;PSMC4;PTPRA;PSMC2;NF1;GRB2;CALM1;MAP3K11 |

| | | | | | | |
|---|---|---|---|---|---|---|
| 199 | Cellular response to heat stress Homo sapiens R-HSA-3371556 | 0.000109 | 0.000774 | 2.28255 | 20.817007 | YWHAE;GSK3B;NUP205;VCP;HSP90AB1;RPS19BP1;MRPL18;AAAS;HDAC6;BAG5;DNAJB6;UBB;POM121C;TPR;BAG1;MLST8;EP300;MAPK1;MAPK3;NUP214;HSPA9;RANBP2;HSPA8;HSP90AA1;HSPA5;HSPA4;RPA2;HSPA2;SIRT1;MTOR;DNAJC7;NUP50;RPA3;ATM;NUP98;HSPA1B;HSPA1A;ATR |
| 200 | Formation of a pool of free 40S subunits Homo sapiens R-HSA-72689 | 0.000109 | 0.000774 | 2.28255 | 20.817007 | RPL5;RPL30;RPL31;RPLP1;RPS4Y1;RPL6;RPS15;RPL7A;RPS19;RPL36;RPLP2;RPL37;RPS10;RPL19;RPS9;RPL41;RPS7;RPS8;RPS5;RPL35A;RPS25;RPS28;EIF3M;RPS27;EIF3K;RPS29;EIF3L;EIF3I;RPL37A;EIF3J;EIF3G;EIF3H;RPL29;EIF3C;RPS21;EIF3D;EIF3A;RPS24 |
| 201 | Processing of Capped Intron-Containing Pre-mRNA Homo sapiens R-HSA-72203 | 0.000109 | 0.000774 | 1.81389 | 16.542537 | NXT1;EIF4A3;HNRNPU;HNRNPR;CCAR1;PTBP1;SNRPD2;SNRNP70;PAPOLA;TXNL4A;SNRNP35;CTNNBL1;RBM5;NUP214;THOC2;ZCRB1;GTF2F2;SRRM1;DDX39A;PRPF6;DDX39B;HNRNPH1;NUP50;SRSF2;SRSF4;SNRPA1;NUP98;DNAJC8;SNRNP200;SLU7;SLBP;NUP205;DDX5;SF3B3;DHX9;DDX23;AAAS;RNPC3;PRPF8;POM121C;U2AF2;POLR2B;POLR2C;TPR;POLR2E;POLR2H;SF3B1;POLR2J;SRSF11;HNRNPA0;RANBP2;HSPA8;HNRNPA3;WTAP;FUS;UPF3B;HNRNPM;LSM7;SNRNP40;HNRNPK;PHF5A;HNRNPF;HNRNPA2B1;HNRNPD;SNRNP48;RBMX |
| 202 | DNA Repair Homo sapiens R-HSA-73894 | 0.000117 | 0.000825 | 1.6394 | 14.838654 | DCLRE1B;MUS81;RIF1;MPG;OGG1;BABAM1;YY1;HERC2;PPP4R2;SUMO3;UIMC1;RUVBL1;SUMO2;EP300;POLI;POLL;NBN;TOPBP1;KPNA2;AQR;USP7;RFC1;USP43;TOP3A;USP45;REV1;SIRT6;RAD23A;ACTR8;BAZ1B;RAD23B;SPIDR;PIAS1;DDB1;MSH6;CCNA1;MSH3;PPIE;NEIL1;FAN1;VCP;INO80E;PRKDC;RNF8;XPC;UNG;POLD4;PPP4C;UBB;POLR2B;POLR2C;UBC;USP1;POLR2E;ASCC2;TP53BP1;ASCC3;POLR2H;ATRIP;POLR2J;FANCI;MBD4;KDM4A;XRCC6;KDM4B;UBE2I;XRCC5;FANCL;POLM;MCRS1;EYA4;SMARCA5;RPA2;ISG15;FANCC;INO80;MLH1;COPS3;COPS6;CDK7;RAD50;COPS5;RPA3;APEX1;UBE2T;RAD17;TCEA1;ATM;ERCC5;ATR;CUL4B |

| | | | | | | |
|---|---|---|---|---|---|---|
| 203 | mRNA Splicing Homo sapiens R-HSA-72172 | 0.000121 | 0.000845 | 1.971044 | 17.784432 | EIF4A3;HNRNPU;HNRNPR;CCAR1;PTBP1;SNRPD2;SNRNP70;PAPOLA;TXNL4A;SNRNP35;CTNNBL1;RBM5;ZCRB1;GTF2F2;SRRM1;PRPF6;HNRNPH1;SRSF2;SRSF4;SNRPA1;DNAJC8;SNRNP200;SLU7;DDX5;SF3B3;DHX9;DDX23;RNPC3;PRPF8;U2AF2;POLR2B;POLR2C;POLR2E;POLR2H;SF3B1;POLR2J;SRSF11;HNRNPA0;HSPA8;HNRNPA3;FUS;UPF3B;HNRNPM;LSM7;SNRNP40;HNRNPK;PHF5A;HNRNPF;HNRNPA2B1;HNRNPD;SNRNP48;RBMX |
| 204 | Negative feedback regulation of MAPK pathway Homo sapiens R-HSA-5674499 | 0.000125 | 0.00087 | 93138 | 836687.97 | MAP2K1;MAP2K2;MAPK1;BRAF;RAF1;MAPK3 |
| 205 | mitochondrial fatty acid beta-oxidation of unsaturated fatty acids Homo sapiens R-HSA-77288 | 0.000125 | 0.00087 | 93138 | 836687.97 | HADHB;HADHA;ACADL;ECI1;ACADM;DECR1 |
| 206 | Mitochondrial translation Homo sapiens R-HSA-5368287 | 0.000128 | 0.000882 | 2.32215 | 20.819377 | GFM1;MRPS35;MRPS14;GFM2;MRPS36;MRPL18;MRPL19;MRPS34;MRPL38;MRPL39;MRPS10;MTIF2;MRPL14;MTIF3;MRPL34;MRPL32;MRPL10;MRPL33;MRPL2;MRPL40;PTCD3;TSFM;MRPS26;MRPS24;MRPS25;MRPS22;MRPS18B;MRPL27;MRPS7;MRPS6;MRPS5;MRPL43;TUFM;MRPS9;MRRF;DAP3 |
| 207 | Regulation of PLK1 Activity at G2/M Transition Homo sapiens R-HSA-2565942 | 0.000138 | 0.000946 | 2.344253 | 20.842233 | YWHAE;CEP57;DYNC1I2;DCTN2;SSNA1;CUL1;ACTR1A;PCM1;UBB;CNTRL;UBC;CEP70;CEP290;RAB8A;CDK5RAP2;CEP78;OFD1;DYNC1H1;HSP90AA1;PPP1R12A;FBXW11;TUBB;CEP152;HAUS4;HAUS3;TUBG1;TUBB4B;TUBA4A;HAUS1;CENPJ;AKAP9;MAPRE1;OPTN;PAFAH1B1;SDCCAG8 |
| 208 | Signalling to ERKs Homo sapiens R-HSA-187687 | 0.000143 | 0.000979 | 1.675035 | 14.825681 | YWHAB;RASGRF2;ITGA2B;PEBP1;WDR83;IRS2;ARRB1;PSMD8;PSMD6;PSMD7;PSMD4;RASGEF1A;PSMD2;PSMD3;PSMD1;JAK2;SPTAN1;JAK1;MAP2K1;MAP2K2;PPP2R5B;PSMA5;PSMA6;PSMA3;PSMA1;PSMA2;AKAP9;KIT;RASA2;IL3RA;PSME1;PSME2;TLN1;RAF1;SOS1;VCL;SPTBN5;PSMD12;SHC2;RALA;PSMD11;PSMD13;SRC;CUL3;IQGAP1;PHB;PSMA7;EGFR;PSMB10;PSMB6;RAP1B;PSMB4;PSMB5;PSMB2;UBB;ERBB3;PSMB3;PSMB1;UBC;ERBB2;MAPK1;CSK;PLCG1;PSMF1;SPTBN1;SPTBN2;MAPK3;KIDINS220;DAB2IP;BRAF;PTK2;PSMB8;PSMB9;MAPK13;PSMC5;PSMC4;PTPRA;PSMC2;NF1;GRB2;CALM1;MAP3K11 |

| | | | | | | |
|---|---|---|---|---|---|---|
| 209 | Sema4D induced cell migration and growth-cone collapse Homo sapiens R-HSA-416572 | 0.00015 | 0.001021 | 4.866278 | 42.844623 | ARHGEF12;ROCK1;SEMA4D;ROCK2;LIMK2;RHOA;MYL12B;CDC42;MYL6;ERBB2;MYH14;MYH9;RAC1;MYH10 |
| 210 | Downstream signaling of activated FGFR1 Homo sapiens R-HSA-5654687 | 0.000151 | 0.001022 | 1.572693 | 13.834687 | AHCYL1;ITGA2B;PEBP1;WDR83;IRS2;PSMD8;PSMD6;PSMD7;PSMD4;AKT2;PSMD2;PSMD3;AKT1;PSMD1;PIP4K2C;IER3;MAP2K1;MAP2K2;PRKCD;ADCY9;AKAP9;PSME1;PSME2;TLN1;RAF1;VCL;SPTBN5;PSMD12;SHC2;PSMD11;PSMD13;CUL3;PIK3R3;ITPR2;ITPR3;IQGAP1;PHB;RAP1B;UBB;PRKAR2A;UBC;THEM4;PLCG1;PSMF1;SPTBN1;SPTBN2;INSR;BRAF;PTK2;AGO3;PTPRA;NF1;GRB2;CALM1;MAP3K11;GSK3B;CDKN1B;YWHAB;RASGRF2;ARRB1;RASGEF1A;MLST8;JAK2;SPTAN1;JAK1;PPP2R5B;PSMA5;PSMA6;PSMA3;CREB1;PSMA1;PSMA2;KIT;RASA2;IL3RA;SOS1;SRC;ADCY6;PSMA7;EGFR;PSMB10;PSMB6;PSMB4;PSMB5;PSMB2;ERBB3;PSMB3;PSMB1;ERBB2;MAPK1;CSK;MAPK3;BAD;DAB2IP;PTPN11;PSMB8;MTOR;PSMB9;PSMC5;PSMC4;PSMC2;RPS6KB2;TNRC6A |
| 211 | Signalling to RAS Homo sapiens R-HSA-167044 | 0.000152 | 0.001022 | 1.683185 | 14.802317 | YWHAB;RASGRF2;ITGA2B;PEBP1;WDR83;IRS2;ARRB1;PSMD8;PSMD6;PSMD7;PSMD4;RASGEF1A;PSMD2;PSMD3;PSMD1;JAK2;SPTAN1;JAK1;MAP2K1;MAP2K2;PPP2R5B;PSMA5;PSMA6;PSMA3;PSMA1;PSMA2;AKAP9;KIT;RASA2;IL3RA;PSME1;PSME2;TLN1;RAF1;SOS1;VCL;SPTBN5;PSMD12;SHC2;RALA;PSMD11;PSMD13;SRC;CUL3;IQGAP1;PHB;PSMA7;EGFR;PSMB10;PSMB6;RAP1B;PSMB4;PSMB5;PSMB2;UBB;ERBB3;PSMB3;PSMB1;UBC;ERBB2;MAPK1;CSK;PSMF1;SPTBN1;SPTBN2;MAPK3;DAB2IP;BRAF;PTK2;PSMB8;PSMB9;MAPK13;PSMC5;PSMC4;PTPRA;PSMC2;NF1;GRB2;CALM1;MAP3K11 |

| | | | | | | |
|---|---|---|---|---|---|---|
| 212 | Signalling to p38 via RIT and RIN Homo sapiens R-HSA-187706 | 0.00016 | 0.001075 | 1.691853 | 14.784958 | YWHAB;RASGRF2;ITGA2B;PEBP1;WDR83;IRS2;ARRB1;PSMD8;PSMD6;PSMD7;PSMD4;RASGEF1A;PSMD2;PSMD3;PSMD1;JAK2;SPTAN1;JAK1;MAP2K1;MAP2K2;PPP2R5B;PSMA5;PSMA6;PSMA3;PSMA1;PSMA2;AKAP9;KIT;RASA2;IL3RA;PSME1;PSME2;TLN1;RAF1;SOS1;VCL;SPTBN5;PSMD12;SHC2;PSMD11;PSMD13;SRC;CUL3;IQGAP1;PHB;PSMA7;EGFR;PSMB10;PSMB6;RAP1B;PSMB4;PSMB5;PSMB2;UBB;ERBB3;PSMB3;PSMB1;UBC;ERBB2;MAPK1;CSK;PSMF1;SPTBN1;SPTBN2;MAPK3;DAB2IP;BRAF;PTK2;PSMB8;PSMB9;PSMC5;PSMC4;PTPRA;PSMC2;NF1;GRB2;CALM1;MAP3K11 |
| 213 | Selenoamino acid metabolism Homo sapiens R-HSA-2408522 | 0.000162 | 0.00108 | 2.121033 | 18.506144 | RPL5;RPL30;RPL31;RPLP1;EPRS;RPS4Y1;RPL6;PAPSS2;RPS15;RPL7A;RPS19;LARS;RARS;RPL36;RPLP2;RPL37;RPS10;RPL19;DARS;RPS9;RPL41;RPS7;RPS8;RPS5;SECISBP2;TXNRD1;GSR;SARS;RPL35A;AIMP1;AIMP2;RPS25;RPS28;RPS27;KARS;QARS;RPS29;MARS;RPL37A;RPL29;RPS21;RPS24 |
| 214 | FCERI mediated NF-kB activation Homo sapiens R-HSA-2871837 | 0.000162 | 0.00108 | 2.121033 | 18.506144 | PSMD12;PSMD11;PSMD13;UBE2D2;CUL1;BCL10;RELA;PSMB10;PSMA7;MALT1;PSMD8;PSMB6;PSMD6;PSMB4;PSMD7;PSMB5;PSMD4;UBB;PSMB2;PSMB3;PSMD2;PSMD3;UBC;PSMB1;PSMD1;PSMF1;FBXW11;NFKB1;PSMB8;PSMB9;NFKBIA;PSMA5;PSMA6;PSMA3;PSMC5;PSMA1;PSMA2;PSMC4;PSMC2;PSME1;PSME2;UBE2V1 |
| 215 | Signaling by Rho GTPases Homo sapiens R-HSA-194315 | 0.000166 | 0.0011 | 1.536299 | 13.368689 | CYFIP1;NCKAP1;RTKN;KDM1A;ITSN1;FAM13A;ARHGAP35;ACTG1;OPHN1;CDH1;KIF5B;ARHGDIA;ARHGDIB;HIST1H2AC;MEN1;RALBP1;OCRL;BCR;MYL6;CLIP1;DAAM1;NUP98;PFN2;ARHGAP18;IQGAP1;IQGAP2;PMF1;MYL12B;SKA2;RHOT2;KLC2;MYH14;DVL3;RHPN2;SRGAP3;ECT2;MYH10;SRGAP1;H3F3A;ARHGAP28;ARPC4;PTK2;DIAPH1;DIAPH2;RPS27;ARPC2;ARPC3;NOXA1;GRB2;NF2;CALM1;YWHAE;ITGB1;AHCTF1;CDKN1B;ARHGAP8;YWHAB;ARPC1B;RASGRF2;ARPC1A;GDI2;WASL;ARHGAP6;XPO1;MCF2L;RAC1;YWHAH;VAV3;ACTR3;NCOA2;ACTR2;ARHGEF12;PPP1R12A;H2AFZ;ARHGEF16;PPP2R5B;ARAP1;RHOD;RHOA;CIT;CTTN;KIF2A;MYH9;TAX1BP3;EVL;ARHGEF2;MAPRE1;SOS1;RHOQ;PAFAH1B1;ROCK1;ROCK2;SRC;NOXO1;CENPC;CDC42;MAPK1;PAK2;MAPK3;CENPT;RANBP2;LIMK2;MYO9A;KTN1;FMNL2;ABI2;TRIP10;CTNNB1;PKN2;GOPC;PKN1;MAD2L1 |

| | | | | | | |
|---|---|---|---|---|---|---|
| 216 | Interferon Signaling Homo sapiens R-HSA-913531 | 0.000182 | 0.0012 | 1.771686 | 15.254207 | EIF4A1;EIF4A3;IFI35;IFIT3;IFIT2;HERC5;TRIM8;TRIM5;TRIM2;KPNA4;JAK2;KPNA2;B2M;KPNA3;HLA-DPA1;JAK1;KPNA1;NUP214;IFNAR2;IFNGR1;IFNGR2;PRKCD;UBE2E1;HLA-C;HLA-F;HLA-E;PIAS1;OAS1;IRF3;NUP50;IRF2;IRF7;NUP98;IRF9;HLA-DQB1;IFNAR1;NUP205;SP100;IFI6;AAAS;POM121C;UBB;TPR;UBC;FLNB;PLCG1;HLA-DQA2;IP6K2;GBP3;MAPK3;RANBP2;STAT1;STAT2;MX1;EIF2AK2;ISG15;PTPN11;PSMB8;BST2;HLA-DPB1;HLA-DRA;TRIM38;EIF4G3;HLA-DRB1;EIF4G2;EIF4G1 |
| 217 | ISG15 antiviral mechanism Homo sapiens R-HSA-1169408 | 0.000194 | 0.001263 | 2.486588 | 21.259579 | EIF4A1;NUP205;EIF4A3;AAAS;HERC5;UBB;POM121C;TPR;UBC;KPNA4;FLNB;PLCG1;KPNA2;KPNA3;KPNA1;JAK1;MAPK3;NUP214;RANBP2;STAT1;MX1;UBE2E1;EIF2AK2;ISG15;IRF3;NUP50;NUP98;EIF4G3;EIF4G2;EIF4G1 |
| 218 | Antiviral mechanism by IFN-stimulated genes Homo sapiens R-HSA-1169410 | 0.000194 | 0.001263 | 2.486588 | 21.259579 | EIF4A1;NUP205;EIF4A3;AAAS;HERC5;UBB;POM121C;TPR;UBC;KPNA4;FLNB;PLCG1;KPNA2;KPNA3;KPNA1;JAK1;MAPK3;NUP214;RANBP2;STAT1;MX1;UBE2E1;EIF2AK2;ISG15;IRF3;NUP50;NUP98;EIF4G3;EIF4G2;EIF4G1 |
| 219 | Mitochondrial Fatty Acid Beta-Oxidation Homo sapiens R-HSA-77289 | 0.000221 | 0.001433 | 6.369869 | 53.625052 | HADHB;HADHA;ACADVL;MCEE;ACADL;PCCA;ECI1;PCCB;ACADM;HADH;DECR1 |
| 220 | Interleukin receptor SHC signaling Homo sapiens R-HSA-912526 | 0.000225 | 0.001443 | 1.661767 | 13.957366 | YWHAB;RASGRF2;ITGA2B;PEBP1;WDR83;IRS2;ARRB1;PSMD8;PSMD6;PSMD7;PSMD4;RASGEF1A;PSMD2;PSMD3;PSMD1;JAK2;SPTAN1;JAK1;MAP2K1;MAP2K2;PPP2R5B;PSMA5;PSMA6;PSMA3;PSMA1;PSMA2;AKAP9;KIT;RASA2;IL3RA;PSME1;PSME2;TLN1;RAF1;SOS1;VCL;SPTBN5;PSMD12;SHC2;PSMD11;PSMD13;SRC;CUL3;PIK3R3;IQGAP1;PHB;PSMA7;EGFR;PSMB10;PSMB6;RAP1B;PSMB4;PSMB5;PSMB2;UBB;ERBB3;PSMB3;PSMB1;UBC;ERBB2;MAPK1;CSK;PSMF1;SPTBN1;SPTBN2;MAPK3;DAB2IP;BRAF;PTK2;PSMB8;PSMB9;PSMC5;PSMC4;PTPRA;PSMC2;NF1;GRB2;CALM1;MAP3K11 |
| 221 | Centrosome maturation Homo sapiens R-HSA-380287 | 0.000225 | 0.001443 | 2.370495 | 19.908014 | YWHAE;CEP57;DYNC1I2;NUMA1;DCTN2;TUBGCP2;SSNA1;ACTR1A;PCM1;CNTRL;CEP70;CEP290;CDK5RAP2;CEP78;OFD1;DYNC1H1;HSP90AA1;CDK11A;TUBB;CEP152;HAUS4;HAUS3;TUBG1;TUBB4B;CDK11B;TUBA4A;HAUS1;CENPJ;AKAP9;MAPRE1;PAFAH1B1;SDCCAG8 |

| | | | | | | |
|---|---|---|---|---|---|---|
| 222 | Recruitment of mitotic centrosome proteins and complexes Homo sapiens R-HSA-380270 | 0.000225 | 0.001443 | 2.370495 | 19.908014 | YWHAE;CEP57;DYNC1I2;NUMA1;DCTN2;TUBGCP2;SSNA1;ACTR1A;PCM1;CNTRL;CEP70;CEP290;CDK5RAP2;CEP78;OFD1;DYNC1H1;HSP90AA1;CDK11A;TUBB;CEP152;HAUS4;HAUS3;TUBG1;TUBB4B;CDK11B;TUBA4A;HAUS1;CENPJ;AKAP9;MAPRE1;PAFAH1B1;SDCCAG8 |
| 223 | NGF signalling via TRKA from the plasma membrane Homo sapiens R-HSA-187037 | 0.000241 | 0.001534 | 1.514135 | 12.616852 | AHCYL1;ITGA2B;PEBP1;WDR83;IRS2;PSMD8;PSMD6;PSMD7;RPS6KA5;PSMD4;AKT2;PSMD2;PSMD3;AKT1;PSMD1;PIP4K2C;AP2M1;IER3;MAP2K1;MAP2K2;PRKCD;ADCY9;AKAP9;PSME1;PSME2;TLN1;RAF1;VCL;SPTBN5;PSMD12;SHC2;PSMD11;PSMD13;CUL3;PIK3R3;ITPR2;ITPR3;IQGAP1;PHB;RAP1B;UBB;PRKAR2A;UBC;THEM4;PLCG1;PSMF1;SPTBN1;MAP2K5;SPTBN2;INSR;BRAF;PTK2;AGO3;PTPRA;NF1;GRB2;CALM1;MAP3K11;GSK3B;CDKN1B;YWHAB;RASGRF2;AP2A1;ARRB1;RASGEF1A;MLST8;JAK2;SPTAN1;JAK1;PPP2R5B;RHOA;DNM2;PSMA5;PSMA6;PSMA3;CREB1;PSMA1;PSMA2;KIT;RASA2;IL3RA;SOS1;RALA;SRC;ADCY6;PSMA7;EGFR;PSMB10;PSMB6;PSMB4;PSMB5;PSMB2;ERBB3;PSMB3;PSMB1;ERBB2;MAPK1;CSK;MAPK3;KIDINS220;BAD;STAT3;DAB2IP;PTPN11;PSMB8;MTOR;PSMB9;MAPK13;PSMC5;PSMC4;PSMC2;RPS6KB2;TNRC6A |
| 224 | Constitutive Signaling by NOTCH1 HD+PEST Domain Mutants Homo sapiens R-HSA-2894862 | 0.00025 | 0.001558 | 2.718406 | 22.548396 | HDAC2;HDAC1;CUL1;CCNC;RBPJ;HDAC8;HDAC9;HDAC6;APH1A;NEURL1B;APH1B;HEY1;UBB;UBC;EP300;HES1;PSENEN;MIB2;ADAM10;KAT2A;ADAM17;NCOR1;SNW1;TBL1XR1;MAML3 |
| 225 | Signaling by NOTCH1 in Cancer Homo sapiens R-HSA-2644603 | 0.00025 | 0.001558 | 2.718406 | 22.548396 | HDAC2;HDAC1;CUL1;CCNC;RBPJ;HDAC8;HDAC9;HDAC6;APH1A;NEURL1B;APH1B;HEY1;UBB;UBC;EP300;HES1;PSENEN;MIB2;ADAM10;KAT2A;ADAM17;NCOR1;SNW1;TBL1XR1;MAML3 |
| 226 | Signaling by NOTCH1 PEST Domain Mutants in Cancer Homo sapiens R-HSA-2644602 | 0.00025 | 0.001558 | 2.718406 | 22.548396 | HDAC2;HDAC1;CUL1;CCNC;RBPJ;HDAC8;HDAC9;HDAC6;APH1A;NEURL1B;APH1B;HEY1;UBB;UBC;EP300;HES1;PSENEN;MIB2;ADAM10;KAT2A;ADAM17;NCOR1;SNW1;TBL1XR1;MAML3 |
| 227 | Constitutive Signaling by NOTCH1 PEST Domain Mutants Homo sapiens R-HSA-2644606 | 0.00025 | 0.001558 | 2.718406 | 22.548396 | HDAC2;HDAC1;CUL1;CCNC;RBPJ;HDAC8;HDAC9;HDAC6;APH1A;NEURL1B;APH1B;HEY1;UBB;UBC;EP300;HES1;PSENEN;MIB2;ADAM10;KAT2A;ADAM17;NCOR1;SNW1;TBL1XR1;MAML3 |

| | | | | | | |
|---|---|---|---|---|---|---|
| 228 | Signaling by NOTCH1 HD+PEST Domain Mutants in Cancer Homo sapiens R-HSA-2894858 | 0.00025 | 0.001558 | 2.718406 | 22.548396 | HDAC2;HDAC1;CUL1;CCNC;RBPJ;HDAC8;HDAC9;HDAC6;APH1A;NEURL1B;APH1B;HEY1;UBB;UBC;EP300;HES1;PSENEN;MIB2;ADAM10;KAT2A;ADAM17;NCOR1;SNW1;TBL1XR1;MAML3 |
| 229 | RHO GTPases Activate WASPs and WAVEs Homo sapiens R-HSA-5663213 | 0.000253 | 0.001571 | 3.477237 | 28.798115 | ACTR3;CYFIP1;NCKAP1;ACTR2;ARPC1B;ARPC1A;ARPC4;WASL;PTK2;ACTG1;CDC42;ABI2;ARPC2;ARPC3;MAPK1;GRB2;RAC1;MAPK3 |
| 230 | Signaling by TGF-beta Receptor Complex Homo sapiens R-HSA-170834 | 0.000259 | 0.001601 | 2.428604 | 20.057524 | USP15;CCNT2;HDAC1;NEDD4L;CCNC;FURIN;F11R;CBL;XPO1;UBB;UBC;PMEPA1;MEN1;SMAD2;WWTR1;SMAD4;TGFB1;SMAD3;USP9X;CGN;RHOA;TGFBR1;SMAD7;CDK9;PPM1A;NCOR1;SNW1;STUB1;TRIM33;UBE2M |
| 231 | Interleukin-3, 5 and GM-CSF signaling Homo sapiens R-HSA-512988 | 0.000264 | 0.001622 | 1.628414 | 13.420133 | YWHAB;RASGRF2;ITGA2B;PEBP1;WDR83;IRS2;ARRB1;PSMD8;PSMD6;PSMD7;PSMD4;RASGEF1A;PSMD2;PSMD3;BLNK;PSMD1;JAK2;SPTAN1;JAK1;MAP2K1;MAP2K2;PPP2R5B;PSMA5;PSMA6;PSMA3;PSMA1;PSMA2;AKAP9;KIT;RASA2;IL3RA;PSME1;PSME2;TLN1;RAF1;SOS1;VCL;SPTBN5;PSMD12;SHC2;PSMD11;PSMD13;SRC;CUL3;PIK3R3;IQGAP1;PHB;CBL;PSMA7;EGFR;PSMB10;PSMB6;RAP1B;PSMB4;PSMB5;PSMB2;UBB;ERBB3;PSMB3;PSMB1;UBC;ERBB2;MAPK1;CSK;PSMF1;SPTBN1;SPTBN2;MAPK3;YES1;DAB2IP;BRAF;PTPN11;PTK2;PSMB8;PSMB9;PSMC5;PSMC4;PTPRA;PSMC2;NF1;GRB2;CALM1;MAP3K11 |
| 232 | mRNA Splicing - Major Pathway Homo sapiens R-HSA-72163 | 0.000268 | 0.001643 | 1.94536 | 15.999577 | DDX5;SF3B3;DHX9;EIF4A3;DDX23;HNRNPU;HNRNPR;PRPF8;CCAR1;PTBP1;SNRPD2;SNRNP70;U2AF2;POLR2B;POLR2C;PAPOLA;POLR2E;POLR2H;TXNL4A;SF3B1;POLR2J;CTNNBL1;RBM5;SRSF11;HNRNPA0;HSPA8;HNRNPA3;FUS;UPF3B;GTF2F2;SRRM1;HNRNPM;LSM7;SNRNP40;PRPF6;HNRNPK;PHF5A;HNRNPH1;HNRNPF;HNRNPA2B1;SRSF2;HNRNPD;SRSF4;SNRPA1;DNAJC8;SNRNP200;SLU7;RBMX |

| | | | | | | |
|---|---|---|---|---|---|---|
| 233 | Signaling by PDGF Homo sapiens R-HSA-186797 | 0.000286 | 0.001746 | 1.514211 | 12.354378 | AHCYL1;ITGA2B;PEBP1;WDR83;IRS2;PSMD8;PSMD6;PSMD7;PSMD4;AKT2;PSMD2;PSMD3;AKT1;PSMD1;PIP4K2C;IER3;MAP2K1;MAP2K2;PRKCD;ADCY9;COL4A4;AKAP9;PSME1;PSME2;TLN1;RAF1;VCL;GRB7;SPTBN5;PSMD12;SHC2;PSMD11;PSMD13;CUL3;PIK3R3;ITPR2;ITPR3;IQGAP1;PHB;RAP1B;UBB;PRKAR2A;UBC;THEM4;PLCG1;PSMF1;SPTBN1;SPTBN2;INSR;BRAF;PTK2;AGO3;PTPRA;NF1;GRB2;CALM1;MAP3K11;GSK3B;CDKN1B;YWHAB;RASGRF2;ARRB1;RASGEF1A;MLST8;JAK2;SPTAN1;JAK1;PPP2R5B;PSMA5;PSMA6;PSMA3;CREB1;PSMA1;PSMA2;KIT;RASA2;IL3RA;SOS1;SRC;FURIN;ADCY6;PSMA7;EGFR;PSMB10;PSMB6;PSMB4;PSMB5;PSMB2;ERBB3;PSMB3;PSMB1;ERBB2;MAPK1;STAT6;CSK;MAPK3;STAT1;BAD;STAT3;DAB2IP;PTPN11;PSMB8;MTOR;PSMB9;PSMC5;PSMC4;PSMC2;RPS6KB2;TNRC6A;BCAR1 |
| 234 | Calnexin/calreticulin cycle Homo sapiens R-HSA-901042 | 0.000304 | 0.001848 | 6.94784 | 56.265026 | PDIA3;EDEM3;GANAB;OS9;EDEM2;SEL1L;SYVN1;DERL2;CALR;UGGT2 |
| 235 | Influenza Viral RNA Transcription and Replication Homo sapiens R-HSA-168273 | 0.000327 | 0.001978 | 1.954869 | 15.689589 | RPL5;RPL30;NUP205;RPL31;RPLP1;RPS4Y1;AAAS;RPL6;IPO5;RPS15;RPL7A;RPS19;POM121C;POLR2B;POLR2C;TPR;RPL36;POLR2E;RPLP2;RPL37;POLR2H;RPS10;POLR2J;RPL19;NUP214;RANBP2;RPS9;RPL41;HSP90AA1;RPS7;RPS8;RPS5;RPL35A;GTF2F2;DNAJC3;RPS25;RPS28;RPS27;RPS29;NUP50;RPL37A;NUP98;RPL29;GRSF1;RPS21;RPS24 |
| 236 | EPHA-mediated growth cone collapse Homo sapiens R-HSA-3928663 | 0.000374 | 0.002256 | 3.476682 | 27.430987 | HSP90AA1;YES1;ROCK1;SRC;ROCK2;SHB;EFNA4;RHOA;PTK2;MYL12B;EFNA1;MYL6;MYH14;MYH9;EPHA1;MYH10;EPHA2 |
| 237 | G2/M Checkpoints Homo sapiens R-HSA-69481 | 0.000384 | 0.002302 | 1.849648 | 14.549184 | YWHAE;YWHAB;PSMD8;BABAM1;PSMD6;PSMD7;HERC2;PSMD4;PSMD2;PSMD3;UIMC1;PSMD1;NBN;TOPBP1;YWHAH;TOP3A;PSMA5;PSMA6;PSMA3;PSMA1;PSMA2;PSME1;MCM3;PSME2;PSMD12;PSMD11;PSMD13;RNF8;PSMB10;PSMA7;PSMB6;PSMB4;PSMB5;UBB;PSMB2;PSMB3;UBC;PSMB1;PSMF1;TP53BP1;ATRIP;RPA2;PSMB8;PSMB9;PSMC5;RAD50;PSMC4;RPA3;PSMC2;RAD17;ATM;ATR |

| | | | | | | |
|---|---|---|---|---|---|---|
| 238 | DAP12 interactions Homo sapiens R-HSA-2172127 | 0.000388 | 0.00232 | 1.504056 | 11.812363 | AHCYL1;ITGA2B;PEBP1;WDR83;IRS2;PSMD8;PSMD6;PSMD7;PSMD4;AKT2;PSMD2;PSMD3;AKT1;PSMD1;PIP4K2C;B2M;IER3;MAP2K1;MAP2K2;PRKCD;HLA-C;HLA-E;ADCY9;AKAP9;PSME1;PSME2;TLN1;RAF1;VCL;SPTBN5;PSMD12;SHC2;PSMD11;PSMD13;CUL3;PIK3R3;ITPR2;ITPR3;IQGAP1;PHB;RAP1B;UBB;PRKAR2A;UBC;THEM4;PLCG1;PSMF1;SPTBN1;SPTBN2;INSR;BRAF;PTK2;AGO3;PTPRA;NF1;GRB2;CALM1;MAP3K11;GSK3B;CDKN1B;YWHAB;RASGRF2;ARRB1;RASGEF1A;MLST8;RAC1;JAK2;SPTAN1;JAK1;VAV3;PPP2R5B;PSMA5;PSMA6;PSMA3;CREB1;PSMA1;PSMA2;KIT;RASA2;IL3RA;SOS1;SRC;ADCY6;PSMA7;EGFR;PSMB10;PSMB6;PSMB4;PSMB5;PSMB2;ERBB3;PSMB3;PSMB1;ERBB2;MAPK1;CSK;MAPK3;BAD;DAB2IP;PTPN11;PSMB8;MTOR;PSMB9;PSMC5;PSMC4;PSMC2;RPS6KB2;TNRC6A |
| 239 | Mitotic G1-G1/S phases Homo sapiens R-HSA-453279 | 0.000397 | 0.00236 | 1.900901 | 14.888913 | RB1;PSMD12;PSMD11;CDKN1B;MAX;PSMD13;HDAC1;CUL1;PPP2R2A;PSMB10;PSMA7;CKS1B;PSMD8;PSMB6;CCND3;PSMD6;PSMB4;PSMD7;PSMB5;PSMD4;UBB;RBBP4;PSMB2;PSMB3;PSMD2;PSMD3;UBC;PSMB1;PSMD1;TK2;PSMF1;RPA2;PSMB8;PSMB9;PSMA5;CCNA1;PSMA6;CDK7;PSMA3;PSMC5;PSMA1;PSMA2;PSMC4;RPA3;PSMC2;PSME1;MCM3;PSME2 |
| 240 | Mitotic Telophase/Cytokinesis Homo sapiens R-HSA-68884 | 0.00041 | 0.002429 | 7.815074 | 60.953446 | STAG1;NIPBL;STAG2;RAD21;KIF23;PDS5B;PDS5A;SMC1A;SMC3 |

| | | | | | | |
|---|---|---|---|---|---|---|
| 241 | Cytokine Signaling in Immune system Homo sapiens R-HSA-1280215 | 0.000452 | 0.002667 | 1.366967 | 10.527986 | EIF4A1;ITGA2B;EIF4A3;PEBP1;WDR83;IRS2;IFI35;IFIT3;IL27RA;IFIT2;PSMD8;HERC5;PSMD6;PSMD7;PSMD4;PSMD2;PSMD3;PSMD1;KPNA4;KPNA2;KPNA3;B2M;KPNA1;NUP214;IFNAR2;MAP2K1;MAP2K2;IL1R1;IL11RA;FBXW11;PRKCD;UBE2E1;HLA-C;LIFR;HLA-F;PIAS1;HLA-E;ADAM17;AKAP9;PSME1;PSME2;NUP98;UBE2V1;IL6ST;TLN1;RAF1;SQSTM1;VCL;IFNAR1;NUP205;SHC2;SPTBN5;PSMD12;PSMD11;PSMD13;CUL3;CUL1;PIK3R3;TNFRSF11B;IQGAP1;PHB;AAAS;RAP1B;POM121C;UBB;TPR;UBC;TNFRSF14;PLCG1;PSMF1;HLA-DQA2;SPTBN1;IP6K2;SPTBN2;YES1;TNFSF12;EIF2AK2;BRAF;ISG15;NFKB1;PTK2;BST2;PTPRA;NF1;UBA3;HLA-DRA;GRB2;CALM1;EIF4G3;HLA-DRB1;MAP3K11;EIF4G2;EIF4G1;UBE2M;CD40;EDA;YWHAB;RASGRF2;ARRB1;TRIM8;RASGEF1A;TRIM5;TRIM2;BLNK;JAK2;SPTAN1;HLA-DPA1;JAK1;TNFRSF12A;IFNGR1;IFNGR2;IL18;PPP2R5B;IRAK4;TNFRSF1A;PSMA5;PSMA6;PSMA3;PSMA1;OAS1;IRF3;PSMA2;NUP50;IL3RA;KIT;IRF2;RASA2;PELI1;IRF7;SOS1;BIRC2;IRF9;HLA-DQB1;SP100;SRC;PELI3;IFI6;CBL;PSMA7;EGFR;RELA;PSMB10;PSMB6;PSMB4;PSMB5;PSMB2;ERBB3;PSMB3;PSMB1;ERBB2;MAPK1;FLNB;STX4;CSK;GBP3;MAPK3;RANBP2;MAP3K3;IL10RB;STAT1;STAT2;STAT3;MX1;DAB2IP;PTPN11;PSMB8;PSMB9;PSMC5;PSMC4;TNIP2;PSMC2;HL[illegible] |

| | | | | | | |
|---|---|---|---|---|---|---|
| 242 | Adaptive Immune System Homo sapiens R-HSA-1280218 | 0.000499 | 0.00293 | 1.325618 | 10.079419 | AHCYL1;KEAP1;PSMD8;HERC5;RNF115;PSMD6;HERC3;RNF114;PSMD7;HERC2;PSMD4;AKT2;PSMD2;PSMD3;AKT1;PSMD1;FBXO3;PIP4K2C;KIFAP3;B2M;IER3;CXADR;FBXO2;HLA-C;HLA-F;HLA-E;DYNC1LI2;RNF126;PSME1;ORAI1;PSME2;RBCK1;SAR1B;NEDD4L;PIK3R3;BCL10;ANAPC11;DYNC2LI1;RNF213;PLCG1;PSMF1;HLA-DQA2;SPTBN2;VASP;YES1;INSR;UBAC1;AGO3;REL;GRB2;CALR;CALM1;DZIP3;HLA-DRB1;LGMN;LRSAM1;CD40;CDKN1B;CLTC;PTPRJ;AP2A1;CTSS;ACTR1A;ACTR1B;CTSL;ARIH2;CTSH;CD36;RAC1;CTSD;HLA-DPA1;CTSB;KIF23;PSMA5;ENAH;PSMA6;PSMA3;PSMA1;KIF2A;PSMA2;KIT;RAPGEF3;HLA-DQB1;DYNC1I2;PSMA7;EGFR;RELA;PSMB6;PSMB4;HLA-DMA;PSMB5;HLA-DMB;PSMB2;PSMB3;PSMB1;PAK2;DYNC1H1;CD74;MIB2;HUWE1;WWP1;UBE2G1;PSMB8;PSMB9;PSMC5;ITCH;PSMC4;PSMC2;HLA-DPB1;SIPA1;UBE3C;CD81;UBE2D2;LTN1;IRS2;UBE3A;UBE3B;UBE2Z;CDH1;RNF19A;KIF5B;AP2M1;PDIA3;FBXW11;UBE2E3;TAP2;UBE2E1;TAP1;UBE4A;STIM1;ANAPC5;UBE2V1;RAF1;PSMD12;PSMD11;CUL5;PSMD13;UBA6;CUL3;CUL1;UBR4;ITPR2;ITPR3;UBR2;UBR1;FBXO44;RAP1B;KLC2;UBB;KIF3A;UBC;THEM4;TNFRSF14;AP1M2;UBE2H;HSPA5;ZBTB16;NFKB1;NFKBIA;MK [illegible] |
| 243 | NCAM signaling for neurite out-growth Homo sapiens R-HSA-375165 | 0.000514 | 0.003009 | 1.583406 | 11.990935 | YWHAB;RASGRF2;ITGA2B;PEBP1;WDR83;IRS2;ARRB1;PSMD8;PSMD6;PSMD7;RPS6KA5;PSMD4;RASGEF1A;PSMD2;PSMD3;PSMD1;JAK2;SPTAN1;JAK1;MAP2K1;MAP2K2;PPP2R5B;PSMA5;PSMA6;PSMA3;CREB1;PSMA1;PSMA2;COL4A4;AKAP9;KIT;RASA2;IL3RA;PSME1;PSME2;TLN1;RAF1;SOS1;VCL;SPTBN5;PSMD12;SHC2;PSMD11;PSMD13;SRC;CUL3;IQGAP1;PHB;PSMA7;EGFR;PSMB10;PSMB6;RAP1B;PSMB4;PSMB5;PSMB2;UBB;ERBB3;PSMB3;PSMB1;UBC;ERBB2;MAPK1;CSK;PSMF1;SPTBN1;SPTBN2;MAPK3;PRNP;DAB2IP;BRAF;PTK2;PSMB8;PSMB9;PSMC5;PSMC4;PTPRA;PSMC2;NF1;GRB2;CALM1;AGRN;MAP3K11 |
| 244 | Loss of Nlp from mitotic centrosomes Homo sapiens R-HSA-380259 | 0.000524 | 0.003034 | 2.376506 | 17.953867 | YWHAE;CEP57;DYNC1I2;DCTN2;SSNA1;ACTR1A;PCM1;CNTRL;CEP70;CEP290;CDK5RAP2;CEP78;OFD1;DYNC1H1;HSP90AA1;TUBB;CEP152;HAUS4;HAUS3;TUBG1;TUBB4B;TUBA4A;HAUS1;CENPJ;AKAP9;MAPRE1;PAFAH1B1;SDCCAG8 |

| | | | | | | |
|---|---|---|---|---|---|---|
| 245 | Loss of proteins required for interphase microtubule organization?from the centrosome Homo sapiens R-HSA-380284 | 0.000524 | 0.003034 | 2.376506 | 17.953867 | YWHAE;CEP57;DYNC1I2;DCTN2;SSNA1;ACTR1A;PCM1;CNTRL;CEP70;CEP290;CDK5RAP2;CEP78;OFD1;DYNC1H1;HSP90AA1;TUBB;CEP152;HAUS4;HAUS3;TUBG1;TUBB4B;TUBA4A;HAUS1;CENPJ;AKAP9;MAPRE1;PAFAH1B1;SDCCAG8 |
| 246 | Nonsense-Mediated Decay (NMD) Homo sapiens R-HSA-927802 | 0.000527 | 0.003034 | 2.027214 | 15.302152 | RPL5;RPL30;SMG1;RPL31;RPLP1;EIF4A3;PPP2R2A;SMG9;RPS4Y1;RPL6;SMG5;SMG6;RPS15;RPL7A;RPS19;RPL36;RPLP2;RPL37;RPS10;RPL19;UPF2;RPS9;RPL41;RPS7;RPS8;RPS5;UPF3B;RPL35A;UPF3A;RPS25;RPS28;RPS27;RPS29;RPL37A;ETF1;RPL29;RPS21;RPS24;EIF4G1 |
| 247 | Nonsense Mediated Decay (NMD) enhanced by the Exon Junction Complex (EJC) Homo sapiens R-HSA-975957 | 0.000527 | 0.003034 | 2.027214 | 15.302152 | RPL5;RPL30;SMG1;RPL31;RPLP1;EIF4A3;PPP2R2A;SMG9;RPS4Y1;RPL6;SMG5;SMG6;RPS15;RPL7A;RPS19;RPL36;RPLP2;RPL37;RPS10;RPL19;UPF2;RPS9;RPL41;RPS7;RPS8;RPS5;UPF3B;RPL35A;UPF3A;RPS25;RPS28;RPS27;RPS29;RPL37A;ETF1;RPL29;RPS21;RPS24;EIF4G1 |
| 248 | TGF-beta receptor signaling activates SMADs Homo sapiens R-HSA-2173789 | 0.000555 | 0.003182 | 3.476126 | 26.059372 | SMAD2;USP15;SMAD4;TGFB1;SMAD3;NEDD4L;FURIN;CBL;TGFBR1;SMAD7;XPO1;UBB;UBC;PMEPA1;STUB1;UBE2M |
| 249 | NOTCH1 Intracellular Domain Regulates Transcription Homo sapiens R-HSA-2122947 | 0.000566 | 0.003231 | 2.808978 | 21.003316 | TLE3;HDAC2;TLE2;HDAC1;CUL1;CCNC;RBPJ;HDAC8;HDAC9;HIF1A;HDAC6;KAT2A;NCOR1;SNW1;HEY1;UBB;TBL1XR1;UBC;EP300;HES1;MAML3 |
| 250 | Unfolded Protein Response (UPR) Homo sapiens R-HSA-381119 | 0.000584 | 0.003311 | 2.167476 | 16.138576 | ACADVL;PARN;CXXC1;ADD1;HSP90B1;EXOSC7;SEC61A1;EXOSC8;LMNA;SSR1;DNAJB9;SEC62;SEC63;KLHDC3;ZBTB17;MBTPS1;XBP1;HSPA5;WFS1;NFYC;GFPT1;PPP2R5B;SYVN1;PDIA6;EIF2S1;DNAJC3;DNAJB11;CALR;TLN1;ATP6V0D1;ATF6;DCP2;ATF4 |
| 251 | RHO GTPases activate PAKs Homo sapiens R-HSA-5627123 | 0.000584 | 0.003311 | 4.632773 | 34.490661 | CDC42;MYL6;PPP1R12A;CTTN;MYH14;MYH9;NF2;RAC1;CALM1;PAK2;MYH10;MYL12B |

| | | | | | | |
|---|---|---|---|---|---|---|
| 252 | Interleukin-2 signaling Homo sapiens R-HSA-451927 | 0.000592 | 0.003339 | 1.593801 | 11.846087 | YWHAB;RASGRF2;ITGA2B;PEBP1;WDR83;IRS2;ARRB1;PSMD8;PSMD6;PSMD7;PSMD4;RASGEF1A;PSMD2;PSMD3;PSMD1;JAK2;SPTAN1;JAK1;MAP2K1;MAP2K2;PPP2R5B;PSMA5;PSMA6;PSMA3;PSMA1;PSMA2;AKAP9;KIT;RASA2;IL3RA;PSME1;PSME2;TLN1;RAF1;SOS1;VCL;SPTBN5;PSMD12;SHC2;PSMD11;PSMD13;SRC;CUL3;PIK3R3;IQGAP1;PHB;PSMA7;EGFR;PSMB10;PSMB6;RAP1B;PSMB4;PSMB5;PSMB2;UBB;ERBB3;PSMB3;PSMB1;UBC;ERBB2;MAPK1;CSK;PSMF1;SPTBN1;SPTBN2;MAPK3;DAB2IP;BRAF;PTK2;PSMB8;PSMB9;PSMC5;PSMC4;PTPRA;PSMC2;NF1;GRB2;CALM1;MAP3K11 |
| 253 | Spry regulation of FGF signaling Homo sapiens R-HSA-1295596 | 0.000648 | 0.003642 | 5.789493 | 42.504223 | UBB;SRC;MKNK1;UBC;MAPK1;PTPN11;BRAF;GRB2;CBL;MAPK3 |
| 254 | Signaling by Wnt Homo sapiens R-HSA-195721 | 0.000695 | 0.00388 | 1.532933 | 11.147249 | GSK3B;TRRAP;CLTC;USP34;CLTB;CBY1;AP2A1;CDC73;PSMD8;PSMD6;PSMD7;XPO1;PSMD4;AKT2;PSMD2;PSMD3;RUVBL1;AKT1;PSMD1;RSPO3;EP300;VPS35;RAC1;TMED5;HIST1H2AC;AP2M1;WLS;MEN1;RNF43;USP8;TLE3;TLE2;CSNK2A1;H2AFZ;CSNK2A2;WNT5A;PPP2R5B;RHOA;PSMA5;PYGO1;PSMA6;PSMA3;DAAM1;PSMA1;PSMA2;PSME1;PSME2;CSNK1G2;PSMD12;PSMD11;CTBP2;PSMD13;HDAC1;CUL3;CUL1;XIAP;ITPR2;VPS26A;ITPR3;PSMA7;PSMB10;LRP6;PSMB6;PSMB4;PSMB5;PSMB2;UBB;GNG5;TNKS2;PSMB3;PSMB1;UBC;DVL3;PSMF1;FZD5;CSNK1A1;RYK;FZD6;H3F3A;GNG11;PSMB8;PSMB9;PSMC5;AGO3;APC;PSMC4;PSMC2;CTNNB1;CALM1;TNRC6A |
| 255 | Regulation of HSF1-mediated heat shock response Homo sapiens R-HSA-3371453 | 0.000696 | 0.00388 | 2.201906 | 16.008979 | YWHAE;GSK3B;NUP205;RPS19BP1;MRPL18;AAAS;BAG5;DNAJB6;UBB;POM121C;TPR;BAG1;MAPK1;MAPK3;NUP214;HSPA9;RANBP2;HSPA8;HSPA5;HSPA4;RPA2;HSPA2;SIRT1;DNAJC7;NUP50;RPA3;ATM;NUP98;HSPA1B;HSPA1A;ATR |
| 256 | Prefoldin mediated transfer of substrate to CCT/TriC Homo sapiens R-HSA-389957 | 0.00077 | 0.004277 | 3.742567 | 26.830729 | CCT3;CCT2;VBP1;TUBB4B;TUBA4A;TUBA1C;TUBB2A;TCP1;PFDN1;CCT8;CCT7;PFDN5;CCT5;CCT4 |

| | | | | | | |
|---|---|---|---|---|---|---|
| 257 | Mitochondrial translation elongation Homo sapiens R-HSA-5389840 | 0.000812 | 0.004491 | 2.141871 | 15.242224 | GFM1;MRPS35;MRPS14;MRPS36;MRPL18;MRPL19;MRPS34;MRPL38;MRPL39;MRPS10;MRPL14;MRPL34;MRPL32;MRPL10;MRPL33;MRPL2;MRPL40;PTCD3;TSFM;MRPS26;MRPS24;MRPS25;MRPS22;MRPS18B;MRPL27;MRPS7;MRPS6;MRPS5;MRPL43;TUFM;MRPS9;DAP3 |
| 258 | Signaling by ERBB2 Homo sapiens R-HSA-1227986 | 0.000826 | 0.004553 | 2.781781 | 19.747487 | GRB7;USP8;HSP90AA1;YES1;CUL5;SRC;EGFR;RHOA;DIAPH1;UBB;ERBB3;AKT2;CDC37;UBC;ERBB2;AKT1;GRB2;STUB1;PLCG1;SOS1 |
| 259 | Retrograde transport at the Trans-Golgi-Network Homo sapiens R-HSA-6811440 | 0.001102 | 0.006049 | 2.608 | 17.762736 | NSF;SYS1;TMF1;COG6;STX16;COG2;ARL1;M6PR;COG1;GCC2;NAPG;IGF2R;STX10;TGOLN2;GOLGA4;VPS53;STX6;ARFIP2;RAB9A;NAA38;VAMP3 |
| 260 | Cell Cycle Checkpoints Homo sapiens R-HSA-69620 | 0.001132 | 0.006189 | 1.672022 | 11.34311 | YWHAE;CDKN1B;YWHAB;PSMD8;BABAM1;PSMD6;PSMD7;HERC2;PSMD4;PSMD2;PSMD3;UIMC1;PSMD1;NBN;TOPBP1;YWHAH;TOP3A;UBE2E1;PSMA5;CCNA1;PSMA6;PSMA3;PSMA1;PSMA2;PSME1;MCM3;PSME2;ANAPC5;PSMD12;PSMD11;ANAPC16;PSMD13;RNF8;PSMB10;PSMA7;ANAPC11;PSMB6;PSMB4;PSMB5;UBB;PSMB2;PSMB3;UBC;PSMB1;PSMF1;TP53BP1;ATRIP;RPA2;PSMB8;PSMB9;PSMC5;RAD50;PSMC4;RPA3;PSMC2;RAD17;ATM;ATR;MAD2L1 |
| 261 | AURKA Activation by TPX2 Homo sapiens R-HSA-8854518 | 0.001158 | 0.006308 | 2.214042 | 14.969685 | YWHAE;CEP57;DYNC1I2;DCTN2;SSNA1;ACTR1A;PCM1;CNTRL;CEP70;CEP290;CDK5RAP2;CEP78;OFD1;DYNC1H1;HSP90AA1;TUBB;CEP152;HAUS4;HAUS3;TUBG1;TUBB4B;TUBA4A;HAUS1;CENPJ;AKAP9;MAPRE1;PAFAH1B1;SDCCAG8 |
| 262 | L1CAM interactions Homo sapiens R-HSA-373760 | 0.001195 | 0.006487 | 1.997217 | 13.44021 | ITGB1;SPTBN5;SRC;ITGA2B;AP2A1;LAMC1;EGFR;SDCBP;RPS6KA5;DPYSL2;MAPK1;NRCAM;RAC1;SPTAN1;AP2M1;SPTBN1;SPTBN2;MAPK3;HSPA8;MAP2K1;MAP2K2;CSNK2A1;CSNK2A2;RDX;MSN;RANBP9;ANK2;LAMB1;ANK3;DNM2;DLG1;DLG3;NUMB;CD24;ITGA9 |
| 263 | MHC class II antigen presentation Homo sapiens R-HSA-2132295 | 0.001205 | 0.006513 | 1.951641 | 13.118287 | DYNC1I2;DCTN2;SAR1B;CLTC;AP2A1;CTSS;DYNC2LI1;ACTR1A;HLA-DMA;KLC2;ACTR1B;HLA-DMB;KIF3A;CTSL;AP1G1;KIF5B;CTSH;KIFAP3;HLA-DQA2;CTSD;RILP;AP2M1;AP1M2;SPTBN2;CTSB;HLA-DPA1;CD74;DYNC1H1;KIF23;DNM2;DYNC1LI2;KIF2A;HLA-DPB1;HLA-DRA;HLA-DRB1;LGMN;HLA-DQB1 |

| | | | | | | |
|---|---|---|---|---|---|---|
| 264 | RHO GTPases Activate ROCKs Homo sapiens R-HSA-5627117 | 0.001258 | 0.006778 | 4.962103 | 33.136531 | MYL6;PPP1R12A;ROCK1;ROCK2;LIMK2;MYH14;MYH9;MYH10;RHOA;MYL12B |
| 265 | ATF6-alpha activates chaperones Homo sapiens R-HSA-381033 | 0.001286 | 0.006902 | 6.945178 | 46.226989 | MBTPS1;XBP1;HSPA5;NFYC;CALR;ATF6;HSP90B1;ATF4 |
| 266 | Cellular responses to stress Homo sapiens R-HSA-2262752 | 0.001372 | 0.007332 | 1.437371 | 9.4749028 | RB1;UBE2D2;EHMT1;ETS2;EP300;NBN;HIST1H2AC;NUP214;ATG3;UBE2E1;ATG14;RRAGA;CHMP4C;CHMP4B;ULK1;ANAPC5;NUP98;NUP205;VCP;ANAPC16;RPS19BP1;MRPL18;AAAS;HIF1A;ANAPC11;BAG5;POM121C;UBB;ATG101;PRDX1;TPR;UBC;BAG1;ATG5;HSPA9;HSPA8;PHC2;HSPA5;HSPA4;H3F3A;HSPA2;NFKB1;H1F0;AGO3;CHMP2B;ATM;ATR;YWHAE;GSK3B;CDKN1B;HSP90AB1;PRKAG2;HIF3A;RB1CC1;MLST8;HIST1H1E;ATP7A;HIST1H1C;GPX2;HSP90AA1;RING1;H2AFZ;WIPI2;TERF1;SIRT1;CCNA1;DNAJC7;NUP50;CAT;ATG4C;CHMP3;ATG4B;TERF2IP;ATG4D;PRKAA1;PRKAA2;GSTP1;RELA;HDAC6;FZR1;RBBP4;DNAJB6;UBN1;MAPK1;RBBP7;MAPK3;RANBP2;SUZ12;EGLN3;EGLN2;TXNRD2;TXNRD1;STAT3;GSR;RPA2;SOD2;MTOR;ATOX1;MAPK10;CCS;RAD50;RPA3;LAMTOR1;P4HB;HSPA1B;TNRC6A;HSPA1A |
| 267 | eNOS activation and regulation Homo sapiens R-HSA-203765 | 0.001522 | 0.008047 | 4.245758 | 27.544571 | GCHFR;HSP90AA1;SPR;DDAH2;NOSIP;AKT1;ZDHHC21;WASL;CALM1;PTS;DNM2 |
| 268 | Metabolism of nitric oxide Homo sapiens R-HSA-202131 | 0.001522 | 0.008047 | 4.245758 | 27.544571 | GCHFR;HSP90AA1;SPR;DDAH2;NOSIP;AKT1;ZDHHC21;WASL;CALM1;PTS;DNM2 |
| 269 | Synthesis of PC Homo sapiens R-HSA-1483191 | 0.001522 | 0.008047 | 4.245758 | 27.544571 | SLC44A3;SLC44A4;CHKB;CHKA;SLC44A1;SLC44A2;ABHD3;CHPT1;CEPT1;LPIN2;LPIN3 |
| 270 | Signaling by NOTCH Homo sapiens R-HSA-157118 | 0.001547 | 0.008148 | 1.828089 | 11.830333 | HDAC2;POGLUT1;B4GALT1;HDAC1;SEL1L;CUL1;ATP2A3;ATP2A2;CCNC;ARRB1;FURIN;RBPJ;HDAC8;HIF1A;HDAC9;HDAC6;APH1A;NEURL1B;APH1B;UBB;HEY1;UBC;EP300;TMED2;ST3GAL6;HES1;PSENEN;TLE3;TLE2;MIB2;ADAM10;ADAM17;KAT2A;ITCH;SNW1;CREB1;NCOR1;AGO3;TBL1XR1;NUMB;MAML3;TNRC6A |
| 271 | Establishment of Sister Chromatid Cohesion Homo sapiens R-HSA-2468052 | 0.001757 | 0.009095 | 8.101417 | 51.394554 | STAG1;STAG2;RAD21;PDS5B;PDS5A;SMC1A;SMC3 |

| | | | | | | |
|---|---|---|---|---|---|---|
| 272 | ER Quality Control Compartment (ERQC) Homo sapiens R-HSA-901032 | 0.001757 | 0.009095 | 8.101417 | 51.394554 | EDEM3;OS9;EDEM2;SEL1L;SYVN1;DERL2;UGGT2 |
| 273 | ATF6-alpha activates chaperone genes Homo sapiens R-HSA-381183 | 0.001757 | 0.009095 | 8.101417 | 51.394554 | XBP1;HSPA5;NFYC;CALR;ATF6;HSP90B1;ATF4 |
| 274 | Folding of actin by CCT/TriC Homo sapiens R-HSA-390450 | 0.001757 | 0.009095 | 8.101417 | 51.394554 | CCT3;CCT2;TCP1;CCT8;CCT7;CCT5;CCT4 |
| 275 | Semaphorin interactions Homo sapiens R-HSA-373755 | 0.001771 | 0.009095 | 2.205764 | 13.975769 | ITGB1;GSK3B;HSP90AB1;ROCK1;ROCK2;ARHGAP35;MYL12B;CDC42;RRAS;DPYSL2;CFL1;ERBB2;MYH14;RAC1;PAK2;MYH10;HSP90AA1;ARHGEF12;SEMA4D;LIMK2;RHOA;MYL6;CDK5;MYH9;TLN1;MET |
| 276 | Mitochondrial translation initiation Homo sapiens R-HSA-5368286 | 0.001772 | 0.009095 | 2.035198 | 12.894723 | MRPS35;MRPS14;MRPS36;MRPL18;MRPL19;MRPS34;MRPL38;MRPL39;MRPS10;MTIF2;MRPL14;MTIF3;MRPL34;MRPL32;MRPL10;MRPL33;MRPL2;MRPL40;PTCD3;MRPS26;MRPS24;MRPS25;MRPS22;MRPS18B;MRPL27;MRPS7;MRPS6;MRPS5;MRPL43;MRPS9;DAP3 |
| 277 | Mitochondrial translation termination Homo sapiens R-HSA-5419276 | 0.001772 | 0.009095 | 2.035198 | 12.894723 | MRPS35;MRPS14;GFM2;MRPS36;MRPL18;MRPL19;MRPS34;MRPL38;MRPL39;MRPS10;MRPL14;MRPL34;MRPL32;MRPL10;MRPL33;MRPL2;MRPL40;PTCD3;MRPS26;MRPS24;MRPS25;MRPS22;MRPS18B;MRPL27;MRPS7;MRPS6;MRPS5;MRPL43;MRPS9;MRRF;DAP3 |
| 278 | MAP2K and MAPK activation Homo sapiens R-HSA-5674135 | 0.001827 | 0.009347 | 2.813731 | 17.7402 | MAP2K1;MAP2K2;SRC;YWHAB;ITGA2B;PEBP1;WDR83;ARRB1;BRAF;IQGAP1;RAP1B;MAPK1;CSK;TLN1;RAF1;VCL;MAPK3 |
| 279 | Cooperation of Prefoldin and TriC/CCT in actin and tubulin folding Homo sapiens R-HSA-389958 | 0.001897 | 0.009669 | 3.066285 | 19.21776 | CCT3;CCT2;VBP1;TUBB4B;TUBA4A;TUBA1C;TUBA1B;TUBB2A;TCP1;PFDN1;CCT8;CCT7;PFDN5;CCT5;CCT4 |
| 280 | Cargo trafficking to the periciliary membrane Homo sapiens R-HSA-5620920 | 0.002023 | 0.010275 | 2.43382 | 15.097156 | CCT3;BBS2;ARF4;CCT2;EXOC7;ARL3;RAB3IP;BBIP1;PKD1;ARL13B;TCP1;EXOC4;PDE6D;EXOC6;EXOC5;CCT8;BBS4;RAB8A;EXOC2;CCT5;CCT4 |

| | | | | | | |
|---|---|---|---|---|---|---|
| 281 | Fc epsilon receptor (FCERI) signaling Homo sapiens R-HSA-2454202 | 0.002092 | 0.010588 | 1.399452 | 8.6339248 | AHCYL1;UBE2D2;ITGA2B;PEBP1;WDR83;IRS2;PSMD8;PSMD6;PSMD7;PSMD4;AKT2;PSMD2;PSMD3;AKT1;PSMD1;PIP4K2C;IER3;MAP2K1;MAP2K2;FBXW11;AKAP9;PSME1;PSME2;UBE2V1;TLN1;RAF1;VCL;SHC2;SPTBN5;PSMD12;PSMD11;PSMD13;CUL3;CUL1;ITPR2;PIK3R3;ITPR3;BCL10;IQGAP1;PHB;RAP1B;UBB;UBC;THEM4;PLCG1;PSMF1;SPTBN1;SPTBN2;INSR;BRAF;NFKB1;PTK2;NFKBIA;AGO3;PTPRA;NF1;GRB2;CALM1;MAP3K11;GSK3B;CDKN1B;YWHAB;RASGRF2;ARRB1;RASGEF1A;MLST8;RAC1;JAK2;SPTAN1;JAK1;VAV3;PPP2R5B;PSMA5;PSMA6;PSMA3;CREB1;PSMA1;PSMA2;IL3RA;KIT;RASA2;SOS1;SRC;PSMA7;EGFR;MALT1;RELA;PSMB10;PSMB6;PSMB4;PSMB5;PSMB2;ERBB3;PSMB3;PSMB1;ERBB2;MAPK1;CSK;PAK2;MAPK3;MAP3K1;BAD;DAB2IP;PTPN11;PSMB8;MTOR;PSMB9;MAPK10;PSMC5;PSMC4;PSMC2;RPS6KB2;TNRC6A |
| 282 | PPARA activates gene expression Homo sapiens R-HSA-1989781 | 0.002116 | 0.01067 | 1.834619 | 11.297988 | YAP1;CHD9;PEX11A;FHL2;MED19;CCNC;RORA;HMGCR;NPAS2;MED13;CPT2;MED10;TGS1;EP300;HMGCS2;CD36;ACADM;TEAD1;TEAD2;FDFT1;ESRRA;NCOA1;WWTR1;NCOA2;CPT1A;CDK19;NCOA3;TXNRD1;NFYC;MED8;NR1D1;SREBF2;MED13L;NCOR1;TBL1XR1;MED21;CYP1A1;PPARG;PLIN2 |
| 283 | Sema4D mediated inhibition of cell attachment and migration Homo sapiens R-HSA-416550 | 0.002298 | 0.011509 | 10.41445 | 63.273033 | SEMA4D;RRAS;RAC1;MET;RHOA;ARHGAP35 |
| 284 | mitochondrial fatty acid beta-oxidation of saturated fatty acids Homo sapiens R-HSA-77286 | 0.002298 | 0.011509 | 10.41445 | 63.273033 | HADHB;HADHA;ACADVL;ACADL;ACADM;HADH |
| 285 | Phospholipid metabolism Homo sapiens R-HSA-1483257 | 0.002342 | 0.011683 | 1.661801 | 10.065465 | DMGDH;SLC44A3;LPGAT1;SLC44A4;SLC44A1;SLC44A2;PITPNB;GDE1;PIK3C2G;TAZ;PIK3C2B;CPNE1;CPNE3;PIP4K2C;PCYT2;DGAT1;OCRL;PISD;HADHB;HADHA;PLBD1;ALDH7A1;MGLL;CDS1;AGPAT5;MTMR1;ABHD3;PIK3R3;MTMR6;PLD3;AGPAT3;PTDSS2;PTDSS1;INPP5F;PTPMT1;INPP5J;CEPT1;CDIPT;CHKB;CHKA;PNPLA8;MBOAT7;DDHD2;MBOAT1;MBOAT2;LPCAT3;ETNK1;CHPT1;PNPLA6;LPIN2;LPIN3;PNPLA2 |

| | | | | | | |
|---|---|---|---|---|---|---|
| 286 | IRE1alpha activates chaperones Homo sapiens R-HSA-381070 | 0.002483 | 0.012347 | 2.317995 | 13.903709 | KLHDC3;ZBTB17;ACADVL;XBP1;HSPA5;WFS1;GFPT1;PPP2R5B;CXXC1;SYVN1;PDIA6;ADD1;DNAJC3;SEC61A1;DNAJB11;LMNA;SSR1;DNAJB9;SEC62;TLN1;ATP6V0D1;SEC63 |
| 287 | Signal transduction by L1 Homo sapiens R-HSA-445144 | 0.002554 | 0.012654 | 3.820936 | 22.811441 | ITGB1;MAP2K1;MAP2K2;CSNK2A1;CSNK2A2;ITGA2B;MAPK1;RAC1;EGFR;MAPK3;ITGA9 |
| 288 | Glucose metabolism Homo sapiens R-HSA-70326 | 0.002685 | 0.013183 | 2.017612 | 11.944495 | PFKFB3;ADPGK;ENO1;HK1;UGP2;UBB;UBC;PHKG2;PGK1;TPI1;GOT1;MDH1;MDH2;PGAM1;GAA;AGL;GOT2;PHKB;PFKL;PKM;GNPDA1;PPP1R3C;ALDOA;CALM1;FBP1;SLC25A11;GAPDH;PFKM;PFKP |
| 289 | Assembly Of The HIV Virion Homo sapiens R-HSA-175474 | 0.002688 | 0.013183 | 5.555784 | 32.883481 | TSG101;UBB;UBC;VPS37D;FURIN;VPS37B;PPIA;VPS28 |
| 290 | Synthesis of PE Homo sapiens R-HSA-1483213 | 0.002688 | 0.013183 | 5.555784 | 32.883481 | PCYT2;CHKB;CHKA;ETNK1;CEPT1;LPIN2;LPIN3;PISD |
| 291 | Methionine salvage pathway Homo sapiens R-HSA-1237112 | 0.002739 | 0.013339 | 17.35465 | 102.39504 | MTAP;GOT1;ADI1;APIP;MRI1 |
| 292 | Uptake and function of diphtheria toxin Homo sapiens R-HSA-5336415 | 0.002739 | 0.013339 | 17.35465 | 102.39504 | HSP90AA1;HSP90AB1;TXNRD1;CD9;EEF2 |
| 293 | Toll-Like Receptors Cascades Homo sapiens R-HSA-168898 | 0.002782 | 0.013503 | 1.703986 | 10.02706 | ATF2;APP;DHX9;UNC93B1;UBE2D2;PELI3;CUL1;HMGB1;ECSIT;TANK;CTSS;RELA;HSP90B1;EEA1;TBK1;RPS6KA5;UBB;CTSL;DHX36;UBC;MAPK1;RIPK1;CD36;CTSB;MAPK3;MAP2K1;MAP3K1;FBXW11;PTPN11;CNPY3;IRAK4;TICAM1;NFKB1;DNM2;SIGIRR;NFKBIA;MAPK10;CREB1;IRF3;TNIP2;IRF7;PELI1;UBE2V1;TLR5;BIRC2;LGMN |
| 294 | EGFR downregulation Homo sapiens R-HSA-182971 | 0.002805 | 0.013565 | 3.226078 | 18.958022 | AP2A1;PTPRK;CBL;EGFR;EPN1;CDC42;UBB;UBC;SPRY1;GRB2;EPS15;STAM2;AP2M1 |
| 295 | Anchoring of the basal body to the plasma membrane Homo sapiens R-HSA-5620912 | 0.002962 | 0.014277 | 1.877896 | 10.93298 | YWHAE;CEP57;DYNC1I2;DCTN2;SSNA1;ACTR1A;PCM1;CEP70;CNTRL;CC2D2A;CEP290;RAB8A;CDK5RAP2;CEP78;IQCB1;OFD1;DYNC1H1;HSP90AA1;TUBB;CEP152;RAB3IP;HAUS4;HAUS3;TUBG1;TUBB4B;TUBA4A;HAUS1;AHI1;CENPJ;AKAP9;MAPRE1;CEP89;PAFAH1B1;SDCCAG8 |

| | | | | | | |
|---|---|---|---|---|---|---|
| 296 | p75 NTR receptor-mediated signalling Homo sapiens R-HSA-193704 | 0.003004 | 0.014431 | 1.969104 | 11.436219 | YWHAE;HDAC2;HDAC1;RASGRF2;ITSN1;RTN4;RELA;APH1A;APH1B;UBB;MCF2L;ARHGDIA;UBC;CASP2;RAC1;ECT2;PSENEN;VAV3;PRKCI;ARHGEF12;BAD;ARHGEF16;RHOA;NFKB1;NFKBIA;ADAM17;ARHGEF1;ARHGEF2;SOS1;SQSTM1 |
| 297 | Nucleotide Excision Repair Homo sapiens R-HSA-5696398 | 0.003127 | 0.014972 | 1.813615 | 10.460318 | INO80E;XPC;YY1;POLD4;UBB;POLR2B;SUMO3;POLR2C;RUVBL1;UBC;SUMO2;POLR2E;EP300;POLR2H;POLR2J;USP7;AQR;UBE2I;RFC1;MCRS1;USP45;RPA2;RAD23A;INO80;ACTR8;RAD23B;PIAS1;DDB1;COPS3;COPS6;CDK7;COPS5;RPA3;PPIE;TCEA1;ERCC5;CUL4B |
| 298 | RHO GTPases activate CIT Homo sapiens R-HSA-5625900 | 0.003378 | 0.01612 | 4.464893 | 25.407225 | MYL6;CDKN1B;MYH14;MYH9;RAC1;MYH10;RHOA;CIT;MYL12B |
| 299 | XBP1(S) activates chaperone genes Homo sapiens R-HSA-381038 | 0.003526 | 0.016767 | 2.281411 | 12.884706 | KLHDC3;ZBTB17;ACADVL;XBP1;WFS1;GFPT1;PPP2R5B;CXXC1;SYVN1;PDIA6;ADD1;DNAJC3;SEC61A1;DNAJB11;LMNA;SSR1;DNAJB9;SEC62;TLN1;ATP6V0D1;SEC63 |
| 300 | Regulation of lipid metabolism by Peroxisome proliferator-activated receptor alpha (PPARalpha) Homo sapiens R-HSA-400206 | 0.003582 | 0.016978 | 1.762798 | 9.9278073 | YAP1;CHD9;PEX11A;FHL2;MED19;CCNC;RORA;HMGCR;NPAS2;MED13;CPT2;MED10;TGS1;EP300;HMGCS2;CD36;ACADM;TEAD1;TEAD2;FDFT1;ESRRA;NCOA1;WWTR1;NCOA2;CPT1A;CDK19;NCOA3;TXNRD1;NFYC;MED8;NR1D1;SREBF2;MED13L;NCOR1;TBL1XR1;MED21;CYP1A1;PPARG;PLIN2 |
| 301 | Viral mRNA Translation Homo sapiens R-HSA-192823 | 0.003689 | 0.017426 | 1.932514 | 10.826945 | RPL5;RPL30;RPL31;RPLP1;RPS4Y1;RPL6;RPS15;RPL7A;RPS19;RPL36;RPLP2;RPL37;RPS10;RPL19;RPS9;RPL41;RPS7;RPS8;RPS5;RPL35A;DNAJC3;RPS25;RPS28;RPS27;RPS29;RPL37A;RPL29;GRSF1;RPS21;RPS24 |
| 302 | Import of palmitoyl-CoA into the mitochondrial matrix Homo sapiens R-HSA-200425 | 0.003899 | 0.018237 | 6.075671 | 33.702316 | CPT1A;PRKAA2;CPT2;PRKAG2;SLC25A20;ACACB;ACACA |
| 303 | Interleukin-6 signaling Homo sapiens R-HSA-1059683 | 0.003899 | 0.018237 | 6.075671 | 33.702316 | STAT1;STAT3;PTPN11;IL6ST;CBL;JAK2;JAK1 |
| 304 | Biotin transport and metabolism Homo sapiens R-HSA-196780 | 0.003899 | 0.018237 | 6.075671 | 33.702316 | BTD;PCCA;HLCS;PCCB;MCCC1;ACACB;ACACA |

| | | | | | | |
|---|---|---|---|---|---|---|
| 305 | Antigen Presentation: Folding, assembly and peptide loading of class I MHC Homo sapiens R-HSA-983170 | 0.004185 | 0.019446 | 3.206478 | 17.55972 | PDIA3;HSPA5;ERAP2;SAR1B;TAP2;HLA-C;TAP1;CALR;HLA-F;B2M;TAPBP;HLA-E |
| 306 | Formation of tubulin folding intermediates by CCT/TriC Homo sapiens R-HSA-389960 | 0.004185 | 0.019446 | 3.206478 | 17.55972 | CCT3;TUBA1C;CCT2;TUBA1B;TUBB2A;TCP1;CCT8;CCT7;TUBB4B;CCT5;TUBA4A;CCT4 |
| 307 | Transcriptional activity of SMAD2/SMAD3:SMAD4 heterotrimer Homo sapiens R-HSA-2173793 | 0.004481 | 0.020757 | 2.406076 | 13.011719 | SMAD2;WWTR1;SMAD4;CCNT2;SMAD3;HDAC1;USP9X;NEDD4L;CCNC;SMAD7;CDK9;PPM1A;NCOR1;SNW1;UBB;UBC;TRIM33;MEN1 |
| 308 | Mitochondrial protein import Homo sapiens R-HSA-1268020 | 0.004571 | 0.021088 | 2.212135 | 11.918955 | HSPA9;DNAJC19;COX19;MTX2;IDH3G;COX17;TIMM22;TIMM44;HSCB;TAZ;HSPD1;SAMM50;GRPEL1;PMPCB;TIMM17B;PMPCA;VDAC1;CYC1;ACO2;SLC25A4;TOMM5 |
| 309 | FCERI mediated MAPK activation Homo sapiens R-HSA-2871796 | 0.004582 | 0.021088 | 1.428783 | 7.6947455 | YWHAB;RASGRF2;ITGA2B;PEBP1;WDR83;IRS2;ARRB1;PSMD8;PSMD6;PSMD7;PSMD4;RASGEF1A;PSMD2;PSMD3;PSMD1;RAC1;JAK2;SPTAN1;JAK1;VAV3;MAP2K1;MAP2K2;PPP2R5B;PSMA5;PSMA6;PSMA3;PSMA1;PSMA2;AKAP9;KIT;RASA2;IL3RA;PSME1;PSME2;TLN1;RAF1;SOS1;VCL;SPTBN5;PSMD12;SHC2;PSMD11;PSMD13;SRC;CUL3;IQGAP1;PHB;PSMA7;EGFR;PSMB10;PSMB6;RAP1B;PSMB4;PSMB5;PSMB2;UBB;ERBB3;PSMB3;PSMB1;UBC;ERBB2;MAPK1;CSK;PLCG1;PSMF1;PAK2;SPTBN1;SPTBN2;MAPK3;MAP3K1;DAB2IP;BRAF;PTK2;PSMB8;PSMB9;MAPK10;PSMC5;PSMC4;PTPRA;PSMC2;NF1;GRB2;CALM1;MAP3K11 |
| 310 | DNA Damage Recognition in GG-NER Homo sapiens R-HSA-5696394 | 0.005157 | 0.023656 | 2.527114 | 13.311293 | INO80E;MCRS1;XPC;RAD23A;ACTR8;INO80;RAD23B;YY1;DDB1;COPS3;COPS6;COPS5;UBB;RUVBL1;UBC;CUL4B |
| 311 | MAPK1 (ERK2) activation Homo sapiens R-HSA-112411 | 0.005587 | 0.025464 | 6.942518 | 36.012969 | MAP2K2;MAPK1;PTPN11;IL6ST;JAK2;JAK1 |

| | | | | | | |
|---|---|---|---|---|---|---|
| 312 | PTK6 Regulates RTKs and Their Effectors AKT1 and DOK1 Homo sapiens R-HSA-8849469 | 0.005587 | 0.025464 | 6.942518 | 36.012969 | DOK1;UBB;UBC;AKT1;ARAP1;CBL |
| 313 | Immune System Homo sapiens R-HSA-168256 | 0.005746 | 0.026105 | 1.171588 | 6.0445186 | ATF2;NCKAP1;CYFIP1;WDR83;PSMD8;PSMD6;RNF115;TBK1;PSMD7;RNF114;PSMD4;AKT2;PSMD2;PSMD3;AKT1;PSMD1;PIP4K2C;IER3;NUP214;CXADR;PRKCD;SIGIRR;DYNC1LI2;RNF126;ADCY9;PSME1;ORAI1;PSME2;NUP98;RBCK1;CFB;NUP205;SAR1B;PRKDC;NEDD4L;BCL10;IQGAP1;AAAS;ANAPC11;DYNC2LI1;RNF213;POM121C;C9;PRKAR2A;PSMF1;PLCG1;HLA-DQA2;ATG5;EIF2AK2;BRAF;AGO3;REL;DZIP3;HLA-DRB1;LGMN;LRSAM1;CDKN1B;ARPC1B;ARPC1A;CLTC;ECSIT;PANX1;EEA1;TRIM8;TRIM5;TRIM2;JAK2;JAK1;HLA-DPA1;IFNGR1;IFNGR2;PSMA5;PSMA6;PSMA3;PSMA1;PSMA2;PELI1;RAPGEF3;HLA-DQB1;PELI3;TANK;EGFR;ADCY6;PSMA7;PSMB6;PSMB4;HLA-DMA;PSMB5;HLA-DMB;PSMB2;PSMB3;PSMB1;STX4;STAT6;FLNB;IL10RB;STAT1;STAT2;STAT3;DAB2IP;WWP1;PSMB8;PSMB9;PSMC5;ABI2;PSMC4;PSMC2;HLA-DPB1;CRCP;UBE2D2;ITGA2B;LTN1;IRS2;PYCARD;CDH1;RNF19A;CFL1;KPNA4;KPNA2;KPNA3;AP2M1;KPNA1;IFNAR2;MAP2K1;MAP2K2;UBE2E3;UBE2E1;TAP2;TAP1;CNPY3;TICAM1;PIAS1;STIM1;ANAPC5;IL6ST;SQSTM1;IFNAR1;SHC2;CUL5;CUL3;UBR4;CUL1;ITPR2;UBR2;TNFRSF11B;ITPR3;PHB;UBR1;FBXO44;HSP90B1;RAP1B;KLC2;TPR;THEM4;TNFRSF14;AP1M2;IP6K2;POLR3[illegible] |
| 314 | Transferrin endocytosis and recycling Homo sapiens R-HSA-917977 | 0.006034 | 0.027328 | 2.822455 | 14.423553 | ATP6V1A;ATP6V1G1;ATP6V0B;ATP6V0E1;TFRC;ATP6AP1;TCIRG1;MCOLN1;ATP6V0A4;ATP6V0D1;ATP6V0C;ATP6V1C1;ATP6V1C2 |
| 315 | FGFR2 alternative splicing Homo sapiens R-HSA-6803529 | 0.006207 | 0.027732 | 2.977252 | 15.130501 | TIAL1;PTBP1;HNRNPM;TIA1;POLR2B;HNRNPH1;POLR2C;HNRNPF;POLR2E;POLR2H;POLR2J;GTF2F2 |
| 316 | Downregulation of TGF-beta receptor signaling Homo sapiens R-HSA-2173788 | 0.006207 | 0.027732 | 2.977252 | 15.130501 | SMAD2;USP15;XPO1;TGFB1;SMAD3;UBB;UBC;NEDD4L;PMEPA1;STUB1;TGFBR1;SMAD7 |

| | | | | | | |
|---|---|---|---|---|---|---|
| 317 | Insulin receptor recycling Homo sapiens R-HSA-77387 | 0.006207 | 0.027732 | 2.977252 | 15.130501 | ATP6V1A;ATP6V1G1;ATP6V0B;ATP6V0E1;ATP6AP1;INSR;ATP6V0A4;TCIRG1;ATP6V0D1;ATP6V0C;ATP6V1C1;ATP6V1C2 |
| 318 | Deadenylation of mRNA Homo sapiens R-HSA-429947 | 0.00626 | 0.027732 | 3.183703 | 16.152672 | CNOT4;EIF4A1;CNOT1;CNOT2;EIF4A3;PARN;TNKS1BP1;CNOT8;EIF4B;EIF4G1;PAIP1 |
| 319 | Regulation of TP53 Activity through Acetylation Homo sapiens R-HSA-6804758 | 0.00626 | 0.027732 | 3.183703 | 16.152672 | HDAC2;MEAF6;RBBP4;HDAC1;AKT2;EP300;AKT1;CHD4;CHD3;RBBP7;BRD7 |
| 320 | Downregulation of SMAD2/3:SMAD4 transcriptional activity Homo sapiens R-HSA-2173795 | 0.00626 | 0.027732 | 3.183703 | 16.152672 | SMAD2;PPM1A;SMAD4;SMAD3;NCOR1;UBB;HDAC1;USP9X;UBC;NEDD4L;TRIM33 |
| 321 | Branched-chain amino acid catabolism Homo sapiens R-HSA-70895 | 0.00626 | 0.027732 | 3.183703 | 16.152672 | BCKDHA;ACAD8;TMLHE;AUH;SHMT1;IVD;MCCC1;DLD;HIBCH;ACAT1;ALDH9A1 |
| 322 | mRNA Splicing - Minor Pathway Homo sapiens R-HSA-72165 | 0.006439 | 0.028436 | 2.172285 | 10.959985 | SF3B3;DDX23;ZCRB1;RNPC3;GTF2F2;PRPF8;SNRNP40;PRPF6;SNRPD2;POLR2B;POLR2C;SRSF2;POLR2E;SNRNP48;POLR2H;TXNL4A;SNRNP35;SNRNP200;SF3B1;POLR2J |
| 323 | Selenocysteine synthesis Homo sapiens R-HSA-2408557 | 0.00658 | 0.028969 | 1.830447 | 9.1956257 | RPL5;RPL30;RPL31;RPLP1;RPS4Y1;RPL6;RPS15;RPL7A;RPS19;RPL36;RPLP2;RPL37;RPS10;RPL19;RPS9;RPL41;RPS7;RPS8;RPS5;SECISBP2;SARS;RPL35A;RPS25;RPS28;RPS27;RPS29;RPL37A;RPL29;RPS21;RPS24 |
| 324 | mTOR signalling Homo sapiens R-HSA-165159 | 0.006956 | 0.030528 | 2.417083 | 12.008533 | PRKAA1;PRKAA2;YWHAB;CAB39L;STRADA;STRADB;PRKAG2;MTOR;PPM1A;RRAGA;AKT2;MLST8;AKT1;LAMTOR1;EIF4B;EIF4G1 |
| 325 | Peptide chain elongation Homo sapiens R-HSA-156902 | 0.007325 | 0.032049 | 1.8336 | 9.0148507 | RPL5;RPL30;RPL31;RPLP1;RPS4Y1;RPL6;RPS15;RPL7A;RPS19;RPL36;RPLP2;RPL37;RPS10;RPL19;RPS9;RPL41;RPS7;RPS8;RPS5;RPL35A;EEF2;RPS25;RPS28;RPS27;RPS29;RPL37A;RPL29;RPS21;RPS24 |
| 326 | RNA polymerase II transcribes snRNA genes Homo sapiens R-HSA-6807505 | 0.00751 | 0.032758 | 1.877622 | 9.1844278 | GTF2A1;GTF2A2;CCNT2;ICE1;ICE2;GTF2B;INTS10;PHAX;RPAP2;SNAPC1;SNAPC2;POLR2B;POLR2C;POLR2E;RPRD1A;POLR2H;POLR2J;POU2F1;INTS3;TAF11;RPRD2;GTF2F2;CDK9;CDK7;TAF5;INTS9;INTS8 |
| 327 | Loss of Function of SMAD2/3 in Cancer Homo sapiens R-HSA-3304349 | 0.007812 | 0.033661 | 8.676767 | 42.101017 | SMAD2;SMAD4;TGFB1;SMAD3;TGFBR1 |

| | | | | | | |
|---|---|---|---|---|---|---|
| 328 | SMAD2/3 MH2 Domain Mutants in Cancer Homo sapiens R-HSA-3315487 | 0.007812 | 0.033661 | 8.676767 | 42.101017 | SMAD2;SMAD4;TGFB1;SMAD3;TGFBR1 |
| 329 | Defective HLCS causes multiple carboxylase deficiency Homo sapiens R-HSA-3371599 | 0.007812 | 0.033661 | 8.676767 | 42.101017 | PCCA;PCCB;MCCC1;ACACB;ACACA |
| 330 | Defects in biotin (Btn) metabolism Homo sapiens R-HSA-3323169 | 0.007812 | 0.033661 | 8.676767 | 42.101017 | PCCA;PCCB;MCCC1;ACACB;ACACA |
| 331 | Response to elevated platelet cytosolic Ca2+ Homo sapiens R-HSA-76005 | 0.008051 | 0.034586 | 1.692351 | 8.1605161 | LGALS3BP;APP;CD63;SERPINA1;STXBP3;WDR1;ITGA2B;A1BG;SERPINA4;CYB5R1;LAMP2;CFL1;PSAP;CDC37L1;PHACTR2;QSOX1;STX4;FAM3C;CD36;ABCC4;TGFB1;HSPA5;APLP2;RARRES2;PCDH7;VEGFB;ACTN4;TUBA4A;TMX3;CALU;CD9;TLN1;CALM1;ALDOA;PPIA;VCL |
| 332 | RORA activates gene expression Homo sapiens R-HSA-1368082 | 0.008929 | 0.038246 | 2.778589 | 13.110518 | NCOA1;NCOA2;CPT1A;NCOR1;CHD9;TBL1XR1;TGS1;EP300;RORA;NR1D1;SIRT1;NPAS2 |
| 333 | PKB-mediated events Homo sapiens R-HSA-109703 | 0.009225 | 0.039395 | 2.316222 | 10.853357 | PRKAA1;PRKAA2;YWHAB;CAB39L;STRADA;STRADB;PRKAG2;MTOR;PPM1A;RRAGA;AKT2;MLST8;AKT1;LAMTOR1;EIF4B;EIF4G1 |
| 334 | Signaling by cytosolic FGFR1 fusion mutants Homo sapiens R-HSA-1839117 | 0.00926 | 0.039424 | 3.472247 | 16.257234 | BCR;ZMYM2;CUX1;STAT1;STAT3;FGFR1OP2;CNTRL;GRB2;LRRFIP1 |
| 335 | Eukaryotic Translation Elongation Homo sapiens R-HSA-156842 | 0.009401 | 0.039788 | 1.76817 | 8.2518595 | RPL5;RPL30;RPL31;RPLP1;RPS4Y1;RPL6;EEF1B2;RPS15;RPL7A;RPS19;RPL36;RPLP2;RPL37;RPS10;RPL19;RPS9;RPL41;RPS7;RPS8;RPS5;RPL35A;EEF2;RPS25;RPS28;RPS27;RPS29;RPL37A;RPL29;RPS21;RPS24 |
| 336 | Nonsense Mediated Decay (NMD) independent of the Exon Junction Complex (EJC) Homo sapiens R-HSA-975956 | 0.009401 | 0.039788 | 1.76817 | 8.2518595 | RPL5;RPL30;RPL31;RPLP1;RPS4Y1;RPL6;RPS15;RPL7A;RPS19;RPL36;RPLP2;RPL37;RPS10;RPL19;RPS9;RPL41;RPS7;RPS8;RPS5;RPL35A;RPS25;RPS28;RPS27;RPS29;RPL37A;ETF1;RPL29;RPS21;RPS24;EIF4G1 |

| | | | | | | |
|---|---|---|---|---|---|---|
| 337 | LRR FLII-interacting protein 1 (LRRFIP1) activates type I IFN production Homo sapiens R-HSA-3134973 | 0.010297 | 0.043065 | 13.88062 | 63.516433 | IRF3;CTNNB1;EP300;LRRFIP1 |
| 338 | Beta oxidation of lauroyl-CoA to decanoyl-CoA-CoA Homo sapiens R-HSA-77310 | 0.010297 | 0.043065 | 13.88062 | 63.516433 | HADHB;HADHA;ACADL;HADH |
| 339 | Beta oxidation of octanoyl-CoA to hexanoyl-CoA Homo sapiens R-HSA-77348 | 0.010297 | 0.043065 | 13.88062 | 63.516433 | HADHB;HADHA;ACADM;HADH |
| 340 | Beta oxidation of decanoyl-CoA to octanoyl-CoA-CoA Homo sapiens R-HSA-77346 | 0.010297 | 0.043065 | 13.88062 | 63.516433 | HADHB;HADHA;ACADM;HADH |
| 341 | Metabolism of carbohydrates Homo sapiens R-HSA-71387 | 0.010594 | 0.04418 | 1.379969 | 6.2753117 | SLC35B3;B4GALT3;B4GALT1;ADPGK;HEXB;HEXA;GXYLT1;SLC2A1;AKR1B1;ENO1;HS6ST1;HK1;NAGLU;HYAL2;MAN2C1;PGLS;SLC37A4;GUSB;NUP214;MANBA;TPI1;PGAM1;GAA;ABCC5;AKR1A1;TALDO1;HSPG2;PKM;PPP1R3C;NUP50;B3GNT3;B3GNT2;NUP98;ALDOA;TKT;FBP1;SLC25A11;B4GALT7;GAPDH;NUP205;PFKFB3;SDC4;SDC2;AAAS;SLC5A3;GNS;PAPSS2;SLC9A1;UGP2;POM121C;UBB;CHST12;TPR;UBC;PHKG2;CHST15;PGK1;ST3GAL6;ARSB;RANBP2;MGAM;MDH1;GOT1;MDH2;B3GAT3;AGL;GOT2;PHKB;HS3ST1;DERA;EXT2;PFKL;G6PC3;GNPDA1;SDC1;CRYL1;CALM1;AGRN;PFKM;PFKP |
| 342 | Intra-Golgi traffic Homo sapiens R-HSA-6811438 | 0.010966 | 0.045594 | 2.18761 | 9.8726656 | NSF;COG6;STX16;GOLIM4;COG2;COG1;NAPG;CYTH2;GOLGA5;CUX1;MAN2A2;MAN2A1;STX6;MAN1C1;STX5;TRIP11;MAN1A1 |

| | | | | | | |
|---|---|---|---|---|---|---|
| 343 | Innate Immune System Homo sapiens R-HSA-168249 | 0.01114 | 0.046183 | 1.213935 | 5.4593562 | ATF2;CYFIP1;NCKAP1;APP;AHCYL1;WDR83;PSMD8;HERC5;LGALS3;PSMD6;PSMD7;TBK1;RPS6KA5;PSMD4;AKT2;PSMD2;PSMD3;AKT1;PSMD1;PIP4K2C;B2M;IER3;PRKCD;HLA-C;HLA-E;SIGIRR;ADCY9;AKAP9;PSME1;PSME2;TLN1;CFB;SPTBN5;DHX9;PRKDC;PIK3R3;IQGAP1;BCL10;C9;PRKAR2A;DHX36;PSMF1;PLCG1;SPTBN1;ATG5;SPTBN2;XRCC6;YES1;XRCC5;INSR;BRAF;PTK2;AGO3;GRB2;CALM1;LGMN;CDKN1B;ARPC1B;ARPC1A;WASL;ECSIT;CTSS;PANX1;EEA1;CTSL;CD36;RAC1;JAK2;SPTAN1;JAK1;CTSB;VAV3;PSMA5;PSMA6;PSMA3;PSMA1;PSMA2;KIT;PELI1;TAX1BP1;CD46;TLR5;PELI3;TANK;PSMA7;EGFR;RELA;ADCY6;PSMB6;PSMB4;PSMB5;PSMB2;PSMB3;PSMB1;STAT6;CD59;PAK2;CD55;MAP3K1;DAB2IP;PSMB8;PSMB9;PSMC5;ITCH;ABI2;PSMC4;PSMC2;CAPZA2;CTNNB1;CRCP;UBE2D2;ITGA2B;PEBP1;IRS2;ACTG1;PYCARD;CFL1;EP300;MAP2K1;MAP2K2;FBXW11;CNPY3;TICAM1;UBE2V1;RAF1;VCL;SHC2;PSMD12;PSMD11;PSMD13;CUL3;CUL1;ITPR2;ITPR3;PHB;PLD3;HSP90B1;RAP1B;UBB;UBC;THEM4;RIPK1;POLR3GL;ARPC4;ISG15;NFKB1;NFKBIA;ARPC2;ARPC3;PTPRA;UBA3;NF1;UBE2K;MAP3K11;UBE2M;GSK3B;HSP90AB1;YWHAB;RASGRF2;ARRB1;HMGB1;DDX41;RASGEF1A;CASP4;MLST8;CASP2;LRRFIP1;ACTR3;ACTR2;HSP90AA1;PPP2R5B;IRAK4;DDOST;DNM2;CREB1;IRF3;IRF2;IL3RA;RASA2 [illegible] |
| 344 | Gap junction degradation Homo sapiens R-HSA-190873 | 0.011331 | 0.046303 | 5.206553 | 23.326261 | DAB2;MYO6;CLTC;CLTB;AP2M1;DNM2 |
| 345 | Signal attenuation Homo sapiens R-HSA-74749 | 0.011331 | 0.046303 | 5.206553 | 23.326261 | INSR;MAPK1;IRS2;GRB2;SOS1;MAPK3 |
| 346 | MAPK3 (ERK1) activation Homo sapiens R-HSA-110056 | 0.011331 | 0.046303 | 5.206553 | 23.326261 | MAP2K1;PTPN11;IL6ST;JAK2;MAPK3;JAK1 |
| 347 | Regulated proteolysis of p75NTR Homo sapiens R-HSA-193692 | 0.011331 | 0.046303 | 5.206553 | 23.326261 | APH1A;PSENEN;ADAM17;APH1B;RELA;NFKB1 |
| 348 | Tetrahydrobiopterin (BH4) synthesis, recycling, salvage and regulation Homo sapiens R-HSA-1474151 | 0.011331 | 0.046303 | 5.206553 | 23.326261 | GCHFR;HSP90AA1;SPR;AKT1;CALM1;PTS |

| | | | | | | |
|---|---|---|---|---|---|---|
| 349 | Platelet degranulation Homo sapiens R-HSA-114608 | 0.01166 | 0.047507 | 1.66544 | 7.4139226 | LGALS3BP;APP;CD63;SERPINA1;WDR1;ITGA2B;A1BG;SERPINA4;CYB5R1;LAMP2;CFL1;PSAP;CDC37L1;PHACTR2;QSOX1;FAM3C;CD36;ABCC4;TGFB1;HSPA5;APLP2;RARRES2;PCDH7;VEGFB;ACTN4;TUBA4A;TMX3;CALU;CD9;TLN1;CALM1;ALDOA;PPIA;VCL |
| 350 | Glycosphingolipid metabolism Homo sapiens R-HSA-1660662 | 0.012045 | 0.04866 | 2.22343 | 9.825548 | GLTP;ARSA;ASAH1;HEXB;HEXA;GBA2;SUMF2;SUMF1;UGT8;SMPD1;CPTP;PSAP;ARSK;NEU1;GLA;ARSB |
| 351 | RNA Polymerase III Abortive And Retractive Initiation Homo sapiens R-HSA-749476 | 0.012045 | 0.04866 | 2.22343 | 9.825548 | GTF3C1;GTF3C3;CRCP;SSB;POU2F1;GTF3C6;POLR3GL;SNAPC1;NFIA;POLR3B;SNAPC2;NFIB;POLR2E;POLR1D;POLR2H;BDP1 |
| 352 | RNA Polymerase III Transcription Homo sapiens R-HSA-74158 | 0.012045 | 0.04866 | 2.22343 | 9.825548 | GTF3C1;GTF3C3;CRCP;SSB;POU2F1;GTF3C6;POLR3GL;SNAPC1;NFIA;POLR3B;SNAPC2;NFIB;POLR2E;POLR1D;POLR2H;BDP1 |
| 353 | Eukaryotic Translation Termination Homo sapiens R-HSA-72764 | 0.012432 | 0.05008 | 1.738422 | 7.6273066 | RPL5;RPL30;RPL31;RPLP1;RPS4Y1;RPL6;RPS15;RPL7A;RPS19;RPL36;RPLP2;RPL37;RPS10;RPL19;RPS9;RPL41;RPS7;RPS8;RPS5;RPL35A;RPS25;RPS28;RPS27;RPS29;RPL37A;ETF1;RPL29;RPS21;RPS24 |
| 354 | EPH-ephrin mediated repulsion of cells Homo sapiens R-HSA-3928665 | 0.012677 | 0.050923 | 2.084728 | 9.1060045 | EPHB6;PSENEN;VAV3;YES1;SRC;CLTC;CLTB;ADAM10;AP2A1;EFNA4;APH1A;EFNB2;EFNA1;APH1B;RAC1;EPHA1;AP2M1;EPHA2 |
| 355 | Post-translational modification: synthesis of GPI-anchored proteins Homo sapiens R-HSA-163125 | 0.013231 | 0.05228 | 2.728536 | 11.801432 | PIGS;DPM2;DPM3;PGAP1;PIGP;PIGK;PIGL;PIGG;PIGV;PIGF;PIGH |
| 356 | HDMs demethylate histones Homo sapiens R-HSA-3214842 | 0.013231 | 0.05228 | 2.728536 | 11.801432 | KDM5A;KDM5B;KDM4A;KDM4B;KDM5D;KDM2A;KDM1A;KDM1B;UTY;KDM7A;JMJD6 |
| 357 | Downregulation of ERBB2:ERBB3 signaling Homo sapiens R-HSA-1358803 | 0.013235 | 0.05228 | 4.049925 | 17.515385 | USP8;UBB;ERBB3;AKT2;UBC;ERBB2;AKT1 |

| | | | | | | |
|---|---|---|---|---|---|---|
| 358 | DEx/H-box helicases activate type I IFN and inflammatory cytokines production Homo sapiens R-HSA-3134963 | 0.013235 | 0.05228 | 4.049925 | 17.515385 | APP;DHX9;DHX36;IRF7;HMGB1;RELA;NFKB1 |
| 359 | Advanced glycosylation endproduct receptor signaling Homo sapiens R-HSA-879415 | 0.013235 | 0.05228 | 4.049925 | 17.515385 | APP;LGALS3;CAPZA2;MAPK1;HMGB1;DDOST;MAPK3 |
| 360 | Trafficking and processing of endosomal TLR Homo sapiens R-HSA-1679131 | 0.013235 | 0.05228 | 4.049925 | 17.515385 | UNC93B1;CTSL;CNPY3;CTSS;HSP90B1;CTSB;LGMN |
| 361 | Constitutive Signaling by Ligand-Responsive EGFR Cancer Variants Homo sapiens R-HSA-1236382 | 0.014147 | 0.055421 | 3.124821 | 13.306169 | HSP90AA1;UBB;CDC37;UBC;GRB2;PLCG1;CBL;SOS1;EGFR |
| 362 | Signaling by Ligand-Responsive EGFR Variants in Cancer Homo sapiens R-HSA-5637815 | 0.014147 | 0.055421 | 3.124821 | 13.306169 | HSP90AA1;UBB;CDC37;UBC;GRB2;PLCG1;CBL;SOS1;EGFR |
| 363 | Signaling by EGFR in Cancer Homo sapiens R-HSA-1643713 | 0.014147 | 0.055421 | 3.124821 | 13.306169 | HSP90AA1;UBB;CDC37;UBC;GRB2;PLCG1;CBL;SOS1;EGFR |
| 364 | Lysosome Vesicle Biogenesis Homo sapiens R-HSA-432720 | 0.014436 | 0.056241 | 2.315632 | 9.8137305 | APP;HSPA8;CLTC;CLTB;AP4E1;M6PR;ARRB1;GNS;DNM2;VAMP8;AP1G2;AP1G1;DNASE2;AP1M2 |
| 365 | Fanconi Anemia Pathway Homo sapiens R-HSA-6783310 | 0.014436 | 0.056241 | 2.315632 | 9.8137305 | DCLRE1B;MUS81;FANCI;FANCL;RPA2;FANCC;UBB;RPA3;UBE2T;UBC;USP1;ATRIP;FAN1;ATR |
| 366 | Signaling by NOTCH2 Homo sapiens R-HSA-1980145 | 0.015728 | 0.060942 | 2.376344 | 9.8672739 | PSENEN;MIB2;ADAM10;RBPJ;APH1A;NEURL1B;APH1B;CREB1;UBB;UBC;EP300;HES1;MAML3 |
| 367 | SMAD2/SMAD3:SMAD4 heterotrimer regulates transcription Homo sapiens R-HSA-2173796 | 0.015728 | 0.060942 | 2.376344 | 9.8672739 | SMAD2;WWTR1;SMAD4;CCNT2;SMAD3;HDAC1;CCNC;SMAD7;CDK9;SNW1;UBB;UBC;MEN1 |

| | | | | | | |
|---|---|---|---|---|---|---|
| 368 | Sphingolipid metabolism Homo sapiens R-HSA-428157 | 0.016258 | 0.062822 | 1.773337 | 7.3047102 | GLTP;ASAH1;SGMS1;HEXB;HEXA;GBA2;UGT8;SGPL1;SPTLC1;SPTLC2;SMPD1;OSBP;PSAP;NEU1;ARSK;ARSB;ARSA;CERS5;SUMF2;SUMF1;ALDH3A2;CPTP;GLA;CSNK1G2;CERS2 |
| 369 | Signaling by PTK6 Homo sapiens R-HSA-8848021 | 0.01694 | 0.064984 | 1.816636 | 7.4083926 | KHDRBS1;CDKN1B;LRRK2;STAT3;DAB2IP;ARAP1;CBL;HIF1A;ARHGAP35;RHOA;EGFR;DOK1;SFPQ;ERBB3;UBB;RASA2;ERBB2;UBC;NF1;AKT1;STAP2;RAC1;BCAR1 |
| 370 | HuR (ELAVL1) binds and stabilizes mRNA Homo sapiens R-HSA-450520 | 0.017 | 0.064984 | 5.784138 | 23.567731 | NUP214;XPO1;SET;ANP32A;PRKCD |
| 371 | Recycling of eIF2:GDP Homo sapiens R-HSA-72731 | 0.017 | 0.064984 | 5.784138 | 23.567731 | EIF2B5;EIF2B4;EIF2B2;EIF2S2;EIF2S1 |
| 372 | Signaling by TGF-beta Receptor Complex in Cancer Homo sapiens R-HSA-3304351 | 0.017 | 0.064984 | 5.784138 | 23.567731 | SMAD2;SMAD4;TGFB1;SMAD3;TGFBR1 |
| 373 | HSF1 activation Homo sapiens R-HSA-3371511 | 0.017057 | 0.065028 | 2.45138 | 9.9800055 | YWHAE;VCP;HSP90AA1;HSP90AB1;DNAJB6;UBB;MRPL18;RPA3;RPA2;HDAC6;HSPA1B;HSPA1A |
| 374 | Association of TriC/CCT with target proteins during biosynthesis Homo sapiens R-HSA-390471 | 0.017109 | 0.065051 | 2.170972 | 8.8318395 | CCT3;ARFGEF2;CCT2;FBXW5;STAT3;KIF13A;TCP1;XRN2;LONP2;CCT8;KIFC3;FKBP9;CCT7;CCT5;CCT4 |
| 375 | Global Genome Nucleotide Excision Repair (GG-NER) Homo sapiens R-HSA-5696399 | 0.01828 | 0.069317 | 1.706378 | 6.8288479 | INO80E;XPC;YY1;POLD4;UBB;SUMO3;RUVBL1;SUMO2;UBC;UBE2I;RFC1;MCRS1;USP45;RPA2;RAD23A;ACTR8;INO80;RAD23B;PIAS1;DDB1;COPS3;COPS6;CDK7;COPS5;RPA3;ERCC5;CUL4B |
| 376 | Activated TLR4 signalling Homo sapiens R-HSA-166054 | 0.018676 | 0.07063 | 1.580574 | 6.2915214 | ATF2;APP;DHX9;UBE2D2;PELI3;CUL1;HMGB1;ECSIT;TANK;RELA;TBK1;RPS6KA5;UBB;UBC;MAPK1;RIPK1;CD36;MAPK3;MAP2K1;MAP3K1;FBXW11;PTPN11;IRAK4;TICAM1;NFKB1;SIGIRR;NFKBIA;MAPK10;CREB1;IRF3;TNIP2;IRF7;PELI1;UBE2V1;BIRC2 |
| 377 | BBSome-mediated cargo-targeting to cilium Homo sapiens R-HSA-5620922 | 0.019634 | 0.07399 | 2.670868 | 10.497826 | CCT3;BBS2;CCT2;RAB3IP;TCP1;BBIP1;CCT8;BBS4;CCT5;CCT4 |
| 378 | Formation of Incision Complex in GG-NER Homo sapiens R-HSA-5696395 | 0.019668 | 0.07399 | 2.058466 | 8.0871907 | UBE2I;USP45;RPA2;XPC;RAD23A;RAD23B;PIAS1;DDB1;CDK7;UBB;SUMO3;RPA3;UBC;SUMO2;ERCC5;CUL4B |

| | | | | | | |
|---|---|---|---|---|---|---|
| 379 | Deadenylation-dependent mRNA decay Homo sapiens R-HSA-429914 | 0.01981 | 0.074328 | 1.95418 | 7.66342 | EIF4A1;EIF4A3;PARN;LSM1;EXOSC7;CNOT4;NT5C3B;LSM7;XRN1;CNOT1;CNOT2;EXOSC8;TNKS1BP1;CNOT8;DCP2;EIF4B;PAIP1;EIF4G1 |
| 380 | Membrane binding and targetting of GAG proteins Homo sapiens R-HSA-174490 | 0.020251 | 0.075187 | 4.164974 | 16.241596 | TSG101;UBB;UBC;VPS37D;VPS37B;VPS28 |
| 381 | Synthesis And Processing Of GAG, GAGPOL Polyproteins Homo sapiens R-HSA-174495 | 0.020251 | 0.075187 | 4.164974 | 16.241596 | TSG101;UBB;UBC;VPS37D;VPS37B;VPS28 |
| 382 | RHO GTPases activate KTN1 Homo sapiens R-HSA-5625970 | 0.020251 | 0.075187 | 4.164974 | 16.241596 | CDC42;KLC2;KIF5B;RAC1;RHOA;KTN1 |
| 383 | Apoptotic cleavage of cell adhesion proteins Homo sapiens R-HSA-351906 | 0.020251 | 0.075187 | 4.164974 | 16.241596 | TJP1;DSP;OCLN;CDH1;CTNNB1;TJP2 |
| 384 | Interferon alpha/beta signaling Homo sapiens R-HSA-909733 | 0.020306 | 0.075196 | 1.776151 | 6.9213612 | IFNAR2;STAT1;STAT2;MX1;IFI6;HLA-C;PTPN11;ISG15;IFI35;HLA-F;IFIT3;PSMB8;IFIT2;HLA-E;BST2;OAS1;IRF3;IRF2;IRF7;IRF9;JAK1;IFNAR1;IP6K2 |
| 385 | Regulation of TP53 Activity Homo sapiens R-HSA-5633007 | 0.020409 | 0.075379 | 1.476749 | 5.7472167 | PRKAA1;HDAC2;PPP1R13B;PRKAA2;MEAF6;PHF20;HDAC1;EHMT1;PRKAG2;CHD4;CHD3;UBB;RBBP4;AKT2;UBC;TP53BP2;SMYD2;MLST8;AKT1;EP300;NBN;TOPBP1;RBBP7;BRD7;ATRIP;SUPT16H;USP7;CSNK2A1;CSNK2A2;TOP3A;TAF11;RPA2;SSRP1;HIPK1;MTOR;CCNA1;RAD50;CDK5;RPA3;RAD17;ATM;TAF5;TAF3;TAF2;ATR |
| 386 | TP53 Regulates Transcription of DNA Repair Genes Homo sapiens R-HSA-6796648 | 0.021161 | 0.077955 | 1.824186 | 7.0333481 | FANCI;ATF2;SUPT16H;CCNT2;FANCC;SSRP1;MLH1;GTF2F2;CDK9;CDK7;POLR2B;POLR2C;POLR2E;TCEA1;NELFB;ATM;POLR2H;CDK12;CDK13;POLR2J;ATR |
| 387 | SUMOylation of transcription factors Homo sapiens R-HSA-3232118 | 0.021423 | 0.078113 | 3.471141 | 13.34053 | UBE2I;SUMO3;SP3;SUMO2;MITF;TP53BP1;PIAS1 |

| | | | | | | |
|---|---|---|---|---|---|---|
| 388 | TP53 regulates transcription of several additional cell death genes whose specific roles in p53-dependent apoptosis remain uncertain Homo sapiens R-HSA-6803205 | 0.021423 | 0.078113 | 3.471141 | 13.34053 | PPP1R13B;TP53I3;BCL6;PERP;TP53BP2;CHM;RABGGTA |
| 389 | Regulation of IFNG signaling Homo sapiens R-HSA-877312 | 0.021423 | 0.078113 | 3.471141 | 13.34053 | STAT1;IFNGR1;IFNGR2;PTPN11;JAK2;PIAS1;JAK1 |
| 390 | Golgi Cisternae Pericentriolar Stack Reorganization Homo sapiens R-HSA-162658 | 0.021423 | 0.078113 | 3.471141 | 13.34053 | RAB2A;GOLGA2;GORASP2;USO1;MAPK1;BLZF1;MAPK3 |
| 391 | Activated NOTCH1 Transmits Signal to the Nucleus Homo sapiens R-HSA-2122948 | 0.022763 | 0.082785 | 2.315043 | 8.7569311 | APH1A;PSENEN;NEURL1B;ITCH;ADAM17;APH1B;UBB;UBC;MIB2;NUMB;ADAM10;ARRB1 |
| 392 | Uptake and actions of bacterial toxins Homo sapiens R-HSA-5339562 | 0.02488 | 0.089797 | 2.387161 | 8.8173929 | HSP90AA1;MAP2K1;HSP90AB1;MAP2K2;PDCD6IP;TXNRD1;CD9;VAMP1;FURIN;EEF2;CALM1 |
| 393 | VEGFR2 mediated vascular permeability Homo sapiens R-HSA-5218920 | 0.02488 | 0.089797 | 2.387161 | 8.8173929 | VAV3;HSP90AA1;AKT2;MLST8;THEM4;AKT1;CTNNB1;RAC1;CALM1;PAK2;MTOR |
| 394 | Integrin alphaIIb beta3 signaling Homo sapiens R-HSA-354192 | 0.02488 | 0.089797 | 2.387161 | 8.8173929 | RAP1B;SRC;ITGA2B;AKT1;CSK;GRB2;TLN1;SOS1;PTK2;RAPGEF3;BCAR1 |
| 395 | SUMO is transferred from E1 to E2 (UBE2I, UBC9) Homo sapiens R-HSA-3065678 | 0.025413 | 0.090796 | 6.939861 | 25.486658 | UBE2I;SUMO3;SUMO2;UBA2 |
| 396 | Sensing of DNA Double Strand Breaks Homo sapiens R-HSA-5693548 | 0.025413 | 0.090796 | 6.939861 | 25.486658 | RAD50;ATM;NBN;KPNA2 |

| | | | | | | |
|---|---|---|---|---|---|---|
| 397 | SMAD2/3 Phosphorylation Motif Mutants in Cancer Homo sapiens R-HSA-3304356 | 0.025413 | 0.090796 | 6.939861 | 25.486658 | SMAD2;TGFB1;SMAD3;TGFBR1 |
| 398 | CLEC7A/inflammasome pathway Homo sapiens R-HSA-5660668 | 0.025413 | 0.090796 | 6.939861 | 25.486658 | PYCARD;RELA;NFKB1;MALT1 |
| 399 | Intraflagellar transport Homo sapiens R-HSA-5620924 | 0.02749 | 0.097726 | 2.003715 | 7.2012444 | TRAF3IP1;WDR34;DYNC2LI1;IFT20;IFT74;IFT88;KIF3A;WDR60;TTC21B;TRIP11;DYNLRB1;TNPO1;KIFAP3;IFT57;CLUAP1 |
| 400 | Host Interactions with Influenza Factors Homo sapiens R-HSA-168253 | 0.02749 | 0.097726 | 2.003715 | 7.2012444 | RANBP2;NUP214;NUP205;TGFB1;EIF2AK2;ISG15;AAAS;POM121C;NUP50;TPR;KPNA4;NUP98;KPNA2;KPNA3;KPNA1 |
| 401 | SUMO E3 ligases SUMOylate target proteins Homo sapiens R-HSA-3108232 | 0.02786 | 0.098794 | 1.579921 | 5.6570265 | TOP2B;NUP205;SP100;XPC;SMC6;SMC3;AAAS;HERC2;POM121C;SUMO3;RAD21;SUMO2;TPR;TP53BP1;NUP214;RANBP2;PHC2;UBE2I;RING1;MITF;SMC1A;PIAS1;STAG1;NSMCE4A;HNRNPK;STAG2;NUP50;SP3;NUP98;TOP1 |
| 402 | Signaling by Interleukins Homo sapiens R-HSA-449147 | 0.028658 | 0.101373 | 1.258067 | 4.4690541 | ITGA2B;PEBP1;WDR83;IRS2;IL27RA;PSMD8;PSMD6;PSMD7;PSMD4;PSMD2;PSMD3;PSMD1;MAP2K1;MAP2K2;IL1R1;IL11RA;FBXW11;LIFR;AKAP9;PSME1;PSME2;UBE2V1;IL6ST;TLN1;RAF1;SQSTM1;VCL;SHC2;SPTBN5;PSMD12;PSMD11;PSMD13;CUL3;CUL1;PIK3R3;IQGAP1;PHB;RAP1B;UBB;UBC;PSMF1;SPTBN1;SPTBN2;YES1;BRAF;NFKB1;PTK2;PTPRA;NF1;GRB2;CALM1;MAP3K11;YWHAB;RASGRF2;ARRB1;RASGEF1A;BLNK;JAK2;SPTAN1;JAK1;IL18;PPP2R5B;IRAK4;PSMA5;PSMA6;PSMA3;PSMA1;PSMA2;IL3RA;KIT;RASA2;PELI1;SOS1;SRC;PELI3;CBL;PSMA7;EGFR;RELA;PSMB10;PSMB6;PSMB4;PSMB5;PSMB2;ERBB3;PSMB3;PSMB1;ERBB2;MAPK1;STX4;CSK;MAPK3;MAP3K3;IL10RB;STAT1;STAT3;DAB2IP;PTPN11;PSMB8;PSMB9;PSMC5;PSMC4;TNIP2;PSMC2 |

| | | | | | | |
|---|---|---|---|---|---|---|
| 403 | TP53 regulates transcription of additional cell cycle genes whose exact role in the p53 pathway remain uncertain Homo sapiens R-HSA-6804115 | 0.029183 | 0.102718 | 2.603682 | 9.2018729 | CNOT4;NPM1;BTG2;RGCC;CNOT1;CNOT2;CENPJ;TNKS1BP1;CNOT8 |
| 404 | VxPx cargo-targeting to cilium Homo sapiens R-HSA-5620916 | 0.029183 | 0.102718 | 2.603682 | 9.2018729 | ARF4;EXOC7;RAB3IP;EXOC4;EXOC6;EXOC5;PKD1;RAB8A;EXOC2 |
| 405 | Circadian Clock Homo sapiens R-HSA-400253 | 0.030307 | 0.106411 | 1.737096 | 6.0735413 | NCOA1;ATF2;NCOA2;CPT1A;CHD9;CUL1;RORA;NR1D1;SIRT1;HIF1A;NPAS2;PER2;PER1;DBP;NCOR1;CREB1;UBB;TBL1XR1;TGS1;UBC;EP300 |
| 406 | Interferon gamma signaling Homo sapiens R-HSA-877300 | 0.031059 | 0.108176 | 1.574833 | 5.4676238 | SP100;TRIM8;TRIM5;TRIM2;JAK2;B2M;HLA-DQA2;HLA-DPA1;JAK1;GBP3;IFNGR1;STAT1;IFNGR2;PRKCD;HLA-C;PTPN11;HLA-F;PIAS1;HLA-E;OAS1;IRF3;IRF2;IRF7;HLA-DPB1;HLA-DRA;TRIM38;IRF9;HLA-DRB1;HLA-DQB1 |
| 407 | Activation of IRF3/IRF7 mediated by TBK1/IKK epsilon Homo sapiens R-HSA-936964 | 0.031113 | 0.108176 | 2.776997 | 9.6365241 | TBK1;UBB;IRF3;UBC;IRF7;PTPN11;TICAM1;TANK |
| 408 | Defects in vitamin and cofactor metabolism Homo sapiens R-HSA-3296482 | 0.031113 | 0.108176 | 2.776997 | 9.6365241 | AMN;PCCA;MCCC1;PCCB;MTR;ACACB;ACACA;CD320 |
| 409 | Displacement of DNA glycosylase by APEX1 Homo sapiens R-HSA-110357 | 0.031266 | 0.108176 | 4.337824 | 15.031516 | MBD4;MPG;OGG1;APEX1;UNG |
| 410 | Integration of provirus Homo sapiens R-HSA-162592 | 0.031266 | 0.108176 | 4.337824 | 15.031516 | XRCC6;XRCC5;PSIP1;PPIA;KPNA1 |
| 411 | Formation of annular gap junctions Homo sapiens R-HSA-196025 | 0.031266 | 0.108176 | 4.337824 | 15.031516 | DAB2;CLTC;CLTB;AP2M1;DNM2 |

| | | | | | | |
|---|---|---|---|---|---|---|
| 412 | Activation of BAD and translocation to mitochondria Homo sapiens R-HSA-111447 | 0.032548 | 0.111525 | 3.037053 | 10.402039 | YWHAE;YWHAB;BAD;AKT2;AKT1;BID;YWHAH |
| 413 | Signaling by EGFRvIII in Cancer Homo sapiens R-HSA-5637812 | 0.032548 | 0.111525 | 3.037053 | 10.402039 | HSP90AA1;CDC37;GRB2;PLCG1;CBL;SOS1;EGFR |
| 414 | Constitutive Signaling by EGFRvIII Homo sapiens R-HSA-5637810 | 0.032548 | 0.111525 | 3.037053 | 10.402039 | HSP90AA1;CDC37;GRB2;PLCG1;CBL;SOS1;EGFR |
| 415 | GRB2:SOS provides linkage to MAPK signaling for Integrins Homo sapiens R-HSA-354194 | 0.032548 | 0.111525 | 3.037053 | 10.402039 | RAP1B;SRC;ITGA2B;GRB2;TLN1;SOS1;PTK2 |
| 416 | SUMOylation Homo sapiens R-HSA-2990846 | 0.03274 | 0.111809 | 1.539242 | 5.2628955 | TOP2B;NUP205;SP100;XPC;SMC6;SMC3;AAAS;HERC2;POM121C;RAD21;SUMO3;TPR;SUMO2;TP53BP1;NUP214;RANBP2;PHC2;UBE2I;RING1;MITF;SMC1A;PIAS1;STAG1;HNRNPK;STAG2;NSMCE4A;NUP50;SP3;UBA2;NUP98;TOP1 |
| 417 | Energy dependent regulation of mTOR by LKB1-AMPK Homo sapiens R-HSA-380972 | 0.032894 | 0.111809 | 2.246595 | 7.6709281 | PPM1A;PRKAA1;RRAGA;PRKAA2;CAB39L;STRADA;STRADB;MLST8;PRKAG2;LAMTOR1;MTOR |
| 418 | Endosomal/Vacuolar pathway Homo sapiens R-HSA-1236977 | 0.032945 | 0.111809 | 3.470588 | 11.844811 | CTSL;HLA-C;HLA-F;B2M;CTSS;HLA-E |
| 419 | Lysine catabolism Homo sapiens R-HSA-71064 | 0.032945 | 0.111809 | 3.470588 | 11.844811 | PHYKPL;OGDH;DLST;CRYM;ALDH7A1;DLD |
| 420 | HDACs deacetylate histones Homo sapiens R-HSA-3214815 | 0.033974 | 0.115028 | 1.736931 | 5.8745548 | HDAC2;KDM1A;HDAC1;ARID4A;GPS2;ARID4B;CHD4;CHD3;HDAC8;NCOR1;BRMS1;RBBP4;TBL1XR1;SAP30L;HMG20B;SAP18;SUDS3;RBBP7;HIST1H2AC;RCOR1 |
| 421 | Non-integrin membrane-ECM interactions Homo sapiens R-HSA-3000171 | 0.034152 | 0.11508 | 1.929379 | 6.5154006 | ITGB1;DDR1;TGFB1;SDC4;LAMB2;SDC2;TRAPPC4;LAMA3;LAMB1;LAMC1;HSPG2;SDC1;ITGA6;AGRN;DDR2 |

| | | | | | | |
|---|---|---|---|---|---|---|
| 422 | BMAL1:CLOCK,NPAS2 activates circadian gene expression Homo sapiens R-HSA-1368108 | 0.034152 | 0.11508 | 1.929379 | 6.5154006 | NCOA1;NCOA2;CHD9;RORA;NR1D1;HIF1A;NPAS2;PER2;PER1;DBP;CREB1;NCOR1;TBL1XR1;TGS1;EP300 |
| 423 | SUMOylation of DNA damage response and repair proteins Homo sapiens R-HSA-3108214 | 0.035003 | 0.117668 | 1.635064 | 5.4812841 | NUP214;RANBP2;NUP205;SP100;PHC2;UBE2I;RING1;XPC;SMC6;SMC3;SMC1A;AAAS;PIAS1;STAG1;HERC2;NSMCE4A;STAG2;POM121C;SUMO3;NUP50;RAD21;SUMO2;TPR;NUP98 |
| 424 | Formation of TC-NER Pre-Incision Complex Homo sapiens R-HSA-6781823 | 0.035803 | 0.120075 | 1.786333 | 5.9479949 | USP7;AQR;DDB1;COPS3;COPS6;CDK7;COPS5;UBB;POLR2B;POLR2C;UBC;POLR2E;EP300;PPIE;TCEA1;POLR2H;POLR2J;CUL4B |
| 425 | rRNA processing Homo sapiens R-HSA-72312 | 0.035926 | 0.120205 | 1.375056 | 4.5738336 | RPL5;RPL30;RPL31;RPLP1;THUMPD1;RPL6;WDR43;RRP9;RPS15;RPL7A;RPS19;RPL36;RPLP2;RPL37;RPS10;RPP38;RPS9;RPS7;RPS8;DIMT1;RPS5;ELAC2;WDR75;DDX52;DKC1;XRN2;RPL37A;RPP21;RPP25;RPL29;NOP2;RPS4Y1;EXOSC7;BMS1;EXOSC8;DCAF13;RPL19;RPL41;UTP6;KRR1;RIOK3;RPL35A;RPS25;RPS28;RPS27;RPS29;MPHOSPH10;TSR1;RPS21;RPS24;MPHOSPH6 |
| 426 | RAF activation Homo sapiens R-HSA-5673000 | 0.036264 | 0.120683 | 2.314454 | 7.6768925 | MAP2K1;MAP2K2;SRC;YWHAB;PPP2R5B;BRAF;PHB;JAK2;RAF1;MAP3K11 |
| 427 | Sulfur amino acid metabolism Homo sapiens R-HSA-1614635 | 0.036264 | 0.120683 | 2.314454 | 7.6768925 | MTAP;GCLC;GOT1;TST;ADI1;BHMT2;APIP;MRI1;MTR;CNDP2 |
| 428 | Transcriptional regulation of white adipocyte differentiation Homo sapiens R-HSA-381340 | 0.036324 | 0.120683 | 1.60862 | 5.3330327 | CEBPD;CHD9;MED19;CCNC;RELA;CCND3;MED13;MED10;TGS1;EP300;CD36;NCOA1;NCOA2;CDK19;TGFB1;NCOA3;MED8;SREBF2;NFKB1;MED13L;NCOR1;KLF5;TBL1XR1;MED21;PPARG |
| 429 | COPII (Coat Protein 2) Mediated Vesicle Transport Homo sapiens R-HSA-204005 | 0.037595 | 0.124616 | 1.661514 | 5.4512312 | NSF;MCFD2;SERPINA1;TMED10;TRAPPC1;STX17;SAR1B;TRAPPC10;TRAPPC4;USO1;NAPG;GOLGA2;LMAN1;SCFD1;TFG;TBC1D20;LMAN2;PPP6R3;TMED2;STX5;CD59;FOLR1 |

| | | | | | | |
|---|---|---|---|---|---|---|
| 430 | Platelet activation, signaling and aggregation Homo sapiens R-HSA-76002 | 0.038036 | 0.124793 | 1.304929 | 4.2661106 | LGALS3BP;APP;SERPINA1;WDR1;ITGA2B;ARRB1;SERPINA4;ABHD12;AKT2;LAMP2;CFL1;AKT1;PHACTR2;QSOX1;CD36;RAC1;FAM3C;JAK2;JAK1;VAV3;ABCC4;APLP2;PRKCD;ACTN4;RHOA;TUBA4A;TMX3;IL3RA;TLN1;ALDOA;RAF1;SOS1;PPIA;RAPGEF3;MGLL;VCL;DGKH;CD63;STXBP3;SRC;GNAI3;PIK3R3;ITPR2;ITPR3;A1BG;GNAI2;RAP1B;CDC42;CYB5R1;GNG5;PSAP;CDC37L1;MAPK1;STX4;CSK;MAPK3;TGFB1;HSPA5;RARRES2;PCDH7;VEGFB;PTPN11;GNG11;PTK2;CALU;CD9;GRB2;CALM1;BCAR1 |
| 431 | HIV elongation arrest and recovery Homo sapiens R-HSA-167287 | 0.038175 | 0.124793 | 2.08327 | 6.8030615 | CDK9;SUPT16H;CCNT2;POLR2B;POLR2C;POLR2E;TCEA1;NELFB;POLR2H;SSRP1;POLR2J;GTF2F2 |
| 432 | Pausing and recovery of HIV elongation Homo sapiens R-HSA-167290 | 0.038175 | 0.124793 | 2.08327 | 6.8030615 | CDK9;SUPT16H;CCNT2;POLR2B;POLR2C;POLR2E;TCEA1;NELFB;POLR2H;SSRP1;POLR2J;GTF2F2 |
| 433 | Elongation arrest and recovery Homo sapiens R-HSA-112387 | 0.038175 | 0.124793 | 2.08327 | 6.8030615 | CDK9;SUPT16H;CCNT2;POLR2B;POLR2C;POLR2E;TCEA1;NELFB;POLR2H;SSRP1;POLR2J;GTF2F2 |
| 434 | Gluconeogenesis Homo sapiens R-HSA-70263 | 0.038175 | 0.124793 | 2.08327 | 6.8030615 | TPI1;GOT1;MDH1;MDH2;PGAM1;GOT2;PGK1;ENO1;ALDOA;SLC25A11;FBP1;GAPDH |
| 435 | Glycolysis Homo sapiens R-HSA-70171 | 0.038175 | 0.124793 | 2.08327 | 6.8030615 | PFKL;PFKFB3;TPI1;PKM;PGAM1;PGK1;ENO1;ALDOA;PFKM;GAPDH;PFKP;HK1 |
| 436 | NS1 Mediated Effects on Host Pathways Homo sapiens R-HSA-168276 | 0.038265 | 0.1248 | 1.944629 | 6.3457467 | RANBP2;NUP214;NUP205;EIF2AK2;ISG15;AAAS;POM121C;NUP50;TPR;KPNA4;NUP98;KPNA2;KPNA3;KPNA1 |
| 437 | Toll Like Receptor 4 (TLR4) Cascade Homo sapiens R-HSA-166016 | 0.040261 | 0.131009 | 1.455078 | 4.6742519 | ATF2;APP;DHX9;UBE2D2;PELI3;CUL1;HMGB1;ECSIT;TANK;RELA;TBK1;RPS6KA5;UBB;UBC;MAPK1;RIPK1;CD36;MAPK3;MAP2K1;MAP3K1;FBXW11;PTPN11;IRAK4;TICAM1;NFKB1;DNM2;SIGIRR;NFKBIA;MAPK10;CREB1;IRF3;TNIP2;IRF7;PELI1;UBE2V1;BIRC2 |
| 438 | Chaperonin-mediated protein folding Homo sapiens R-HSA-390466 | 0.040582 | 0.131752 | 1.526914 | 4.8928961 | GNAI3;GNAI2;TUBA1C;TUBA1B;GNG5;KIF13A;LONP2;CCT8;CCT7;CCT5;CCT4;CCT3;CCT2;ARFGEF2;FBXW5;CSNK2A1;VBP1;CSNK2A2;STAT3;TUBB4B;GNG11;TUBA4A;TUBB2A;TCP1;XRN2;PFDN1;FKBP9;KIFC3;PFDN5 |

| | | | | | | |
|---|---|---|---|---|---|---|
| 439 | TP53 Regulates Transcription of Cell Death Genes Homo sapiens R-HSA-5633008 | 0.041911 | 0.135756 | 1.860353 | 5.9014353 | BNIP3L;PRELID1;PPP1R13B;CHM;RABGGTA;TP53I3;BCL6;AIFM2;PERP;TP53BP2;CASP2;FAS;BAX;ATM;BID |
| 440 | COPI-independent Golgi-to-ER retrograde traffic Homo sapiens R-HSA-6811436 | 0.042572 | 0.137584 | 2.121648 | 6.6971129 | DYNC2LI1;DYNC1I2;DYNC1H1;DYNC1LI2;DCTN2;GALNT1;RAB3GAP2;PAFAH1B3;AGPAT3;RAB3GAP1;PAFAH1B1 |
| 441 | RNA Polymerase III Transcription Initiation Homo sapiens R-HSA-76046 | 0.042902 | 0.138338 | 1.96256 | 6.1797738 | GTF3C1;GTF3C3;CRCP;POU2F1;GTF3C6;POLR3GL;SNAPC1;POLR3B;SNAPC2;POLR2E;POLR1D;POLR2H;BDP1 |
| 442 | TP53 Regulates Transcription of Genes Involved in Cytochrome C Release Homo sapiens R-HSA-6803204 | 0.043452 | 0.139793 | 2.52438 | 7.9167121 | BNIP3L;PRELID1;PPP1R13B;AIFM2;TP53BP2;BAX;ATM;BID |
| 443 | Toll Like Receptor 9 (TLR9) Cascade Homo sapiens R-HSA-168138 | 0.044217 | 0.141934 | 1.537941 | 4.7962858 | ATF2;APP;DHX9;PELI3;CUL1;HMGB1;ECSIT;RELA;EEA1;RPS6KA5;UBB;DHX36;UBC;MAPK1;MAPK3;MAP2K1;MAP3K1;FBXW11;IRAK4;NFKB1;NFKBIA;MAPK10;CREB1;TNIP2;IRF7;PELI1;UBE2V1 |
| 444 | Synthesis of glycosylphosphatidylinositol (GPI) Homo sapiens R-HSA-162710 | 0.046942 | 0.148998 | 2.699428 | 8.2571497 | DPM2;PIGP;PIGL;PIGG;PIGV;PIGF;PIGH |
| 445 | STING mediated induction of host immune responses Homo sapiens R-HSA-1834941 | 0.046942 | 0.148998 | 2.699428 | 8.2571497 | XRCC6;TBK1;IRF3;XRCC5;PRKDC;STAT6;DDX41 |
| 446 | p75NTR signals via NF-kB Homo sapiens R-HSA-193639 | 0.046942 | 0.148998 | 2.699428 | 8.2571497 | NFKBIA;PRKCI;UBB;UBC;SQSTM1;RELA;NFKB1 |
| 447 | MAP3K8 (TPL2)-dependent MAPK1/3 activation Homo sapiens R-HSA-5684264 | 0.046942 | 0.148998 | 2.699428 | 8.2571497 | MAP2K1;UBB;FBXW11;TNIP2;UBC;CUL1;NFKB1 |

| | | | | | | |
|---|---|---|---|---|---|---|
| 448 | TGF-beta receptor signaling in EMT (epithelial to mesenchymal transition) Homo sapiens R-HSA-2173791 | 0.046942 | 0.148998 | 2.699428 | 8.2571497 | TGFB1;UBB;UBC;CGN;F11R;RHOA;TGFBR1 |
| 449 | Recycling pathway of L1 Homo sapiens R-HSA-437239 | 0.04744 | 0.149578 | 2.169661 | 6.613751 | RPS6KA5;SRC;DPYSL2;RDX;NUMB;MAPK1;MSN;AP2A1;AP2M1;DNM2 |
| 450 | Attenuation phase Homo sapiens R-HSA-3371568 | 0.04744 | 0.149578 | 2.169661 | 6.613751 | HSPA8;HSP90AA1;HSP90AB1;DNAJB6;UBB;MRPL18;EP300;HSPA2;HSPA1B;HSPA1A |
| 451 | ZBP1(DAI) mediated induction of type I IFNs Homo sapiens R-HSA-1606322 | 0.04744 | 0.149578 | 2.169661 | 6.613751 | NFKBIA;APP;TBK1;IRF3;DHX9;RIPK1;HMGB1;TICAM1;RELA;NFKB1 |
| 452 | CD28 co-stimulation Homo sapiens R-HSA-389356 | 0.048134 | 0.151096 | 1.983939 | 6.018813 | CDC42;YES1;SRC;AKT2;MLST8;PIK3R3;THEM4;AKT1;GRB2;RAC1;PAK2;MTOR |
| 453 | Detoxification of Reactive Oxygen Species Homo sapiens R-HSA-3299685 | 0.048134 | 0.151096 | 1.983939 | 6.018813 | GPX2;CCS;TXNRD2;GSTP1;PRDX1;TXNRD1;GSR;CAT;P4HB;ATP7A;SOD2;ATOX1 |
| 454 | Loss of Function of TGFBR1 in Cancer Homo sapiens R-HSA-3656534 | 0.048881 | 0.152432 | 4.626276 | 13.963778 | SMAD2;TGFB1;SMAD3;TGFBR1 |
| 455 | TGFBR1 KD Mutants in Cancer Homo sapiens R-HSA-3656532 | 0.048881 | 0.152432 | 4.626276 | 13.963778 | SMAD2;TGFB1;SMAD3;TGFBR1 |
| 456 | Synthesis of Ketone Bodies Homo sapiens R-HSA-77111 | 0.048881 | 0.152432 | 4.626276 | 13.963778 | HMGCL;BDH2;HMGCS2;ACAT1 |
| 457 | MyD88 dependent cascade initiated on endosome Homo sapiens R-HSA-975155 | 0.049319 | 0.153127 | 1.531037 | 4.6075647 | ATF2;APP;DHX9;PELI3;CUL1;HMGB1;ECSIT;RELA;RPS6KA5;UBB;DHX36;UBC;MAPK1;MAPK3;MAP2K1;MAP3K1;FBXW11;IRAK4;NFKB1;NFKBIA;MAPK10;CREB1;TNIP2;IRF7;PELI1;UBE2V1 |
| 458 | Toll Like Receptor 7/8 (TLR7/8) Cascade Homo sapiens R-HSA-168181 | 0.049319 | 0.153127 | 1.531037 | 4.6075647 | ATF2;APP;DHX9;PELI3;CUL1;HMGB1;ECSIT;RELA;RPS6KA5;UBB;DHX36;UBC;MAPK1;MAPK3;MAP2K1;MAP3K1;FBXW11;IRAK4;NFKB1;NFKBIA;MAPK10;CREB1;TNIP2;IRF7;PELI1;UBE2V1 |

| | | | | | | |
|---|---|---|---|---|---|---|
| 459 | WNT5A-dependent internalization of FZD2, FZD5 and ROR2 Homo sapiens R-HSA-5140745 | 0.049839 | 0.153401 | 2.974598 | 8.9206766 | FZD5;WNT5A;CLTC;CLTB;AP2A1;AP2M1 |
| 460 | IRF3-mediated induction of type I IFN Homo sapiens R-HSA-3270619 | 0.049839 | 0.153401 | 2.974598 | 8.9206766 | XRCC6;TBK1;IRF3;XRCC5;PRKDC;DDX41 |
| 461 | NF-kB is activated and signals survival Homo sapiens R-HSA-209560 | 0.049839 | 0.153401 | 2.974598 | 8.9206766 | NFKBIA;UBB;UBC;SQSTM1;RELA;NFKB1 |
| 462 | Signaling by Activin Homo sapiens R-HSA-1502540 | 0.049839 | 0.153401 | 2.974598 | 8.9206766 | SMAD2;DRAP1;SMAD4;SMAD3;ACVR1B;ACVR2B |
| 463 | Regulation of cholesterol biosynthesis by SREBP (SREBF) Homo sapiens R-HSA-1655829 | 0.050739 | 0.155832 | 1.689556 | 5.0366849 | NCOA1;NCOA2;MBTPS1;CHD9;SAR1B;INSIG2;INSIG1;NFYC;CYP51A1;HMGCR;ACACB;SREBF2;ACACA;TBL1XR1;TGS1;SCAP;RAN;FDFT1 |
| 464 | Base-Excision Repair, AP Site Formation Homo sapiens R-HSA-73929 | 0.051201 | 0.156575 | 3.470036 | 10.312946 | MBD4;MPG;OGG1;NEIL1;UNG |
| 465 | eNOS activation Homo sapiens R-HSA-203615 | 0.051201 | 0.156575 | 3.470036 | 10.312946 | HSP90AA1;DDAH2;AKT1;ZDHHC21;CALM1 |
| 466 | Metabolism of vitamins and cofactors Homo sapiens R-HSA-196854 | 0.051585 | 0.157413 | 1.377367 | 4.0832286 | PANK4;ABCD4;VKORC1L1;PANK2;UBIAD1;PDXK;SDC4;SDC2;SHMT1;QPRT;SLC2A1;MTR;NADSYN1;ACACB;ACACA;HLCS;RDH11;CYP2R1;NADK;AASDHPPT;CYB5A;ABCC1;AMN;GSTO2;GSTO1;AKR1C1;MCCC1;SLC52A3;AKR1C3;MTHFS;HSPG2;COASY;LMBRD1;BTD;PCCA;NMNAT1;PCCB;SDC1;LRP10;AGRN;CD320;LGMN |
| 467 | RNA Polymerase III Transcription Termination Homo sapiens R-HSA-73980 | 0.052795 | 0.160635 | 2.231439 | 6.5634156 | SSB;CRCP;NFIA;POLR3B;POLR3GL;NFIB;POLR1D;POLR2E;POLR2H |
| 468 | Protein folding Homo sapiens R-HSA-391251 | 0.052867 | 0.160635 | 1.468184 | 4.3164184 | GNAI3;GNAI2;TUBA1C;TUBA1B;GNG5;KIF13A;LONP2;CCT8;CCT7;CCT5;CCT4;CCT3;CCT2;ARFGEF2;FBXW5;CSNK2A1;VBP1;CSNK2A2;STAT3;TBCB;TUBB4B;GNG11;TUBA4A;TUBB2A;XRN2;TCP1;PFDN1;FKBP9;KIFC3;PFDN5 |

| | | | | | | |
|---|---|---|---|---|---|---|
| 469 | Toll Like Receptor 10 (TLR10) Cascade Homo sapiens R-HSA-168142 | 0.055032 | 0.166146 | 1.52366 | 4.4183813 | ATF2;APP;DHX9;PELI3;CUL1;HMGB1;ECSIT;RELA;RPS6KA5;UBB;UBC;MAPK1;MAPK3;MAP2K1;MAP3K1;FBXW11;IRAK4;NFKB1;NFKBIA;MAPK10;CREB1;TNIP2;PELI1;UBE2V1;TLR5 |
| 470 | Toll Like Receptor 5 (TLR5) Cascade Homo sapiens R-HSA-168176 | 0.055032 | 0.166146 | 1.52366 | 4.4183813 | ATF2;APP;DHX9;PELI3;CUL1;HMGB1;ECSIT;RELA;RPS6KA5;UBB;UBC;MAPK1;MAPK3;MAP2K1;MAP3K1;FBXW11;IRAK4;NFKB1;NFKBIA;MAPK10;CREB1;TNIP2;PELI1;UBE2V1;TLR5 |
| 471 | MyD88 cascade initiated on plasma membrane Homo sapiens R-HSA-975871 | 0.055032 | 0.166146 | 1.52366 | 4.4183813 | ATF2;APP;DHX9;PELI3;CUL1;HMGB1;ECSIT;RELA;RPS6KA5;UBB;UBC;MAPK1;MAPK3;MAP2K1;MAP3K1;FBXW11;IRAK4;NFKB1;NFKBIA;MAPK10;CREB1;TNIP2;PELI1;UBE2V1;TLR5 |
| 472 | Dual incision in TC-NER Homo sapiens R-HSA-6782135 | 0.055166 | 0.166198 | 1.615436 | 4.6805904 | USP7;AQR;RFC1;RPA2;DDB1;POLD4;CDK7;UBB;POLR2B;POLR2C;RPA3;UBC;POLR2E;EP300;PPIE;TCEA1;ERCC5;POLR2H;POLR2J;CUL4B |
| 473 | Factors involved in megakaryocyte development and platelet production Homo sapiens R-HSA-983231 | 0.056592 | 0.170136 | 1.376543 | 3.9532678 | DOCK6;HDAC2;KDM1A;HDAC1;DOCK7;JMJD1C;AK3;KIF12;GATA3;GATA2;AKAP1;CDC42;KLC2;KIF3A;KIF5B;PRKAR2A;KIF13B;MFN1;EP300;HMG20B;KIF21A;KIF1B;RAC1;KIFAP3;JAK2;CBX5;H3F3A;ITPK1;KIF25;KIF23;EHD1;CDK5;KIF2A;IRF3;KIF16B;MAFG;IRF2;CAPZA2;IRF7;RCOR1 |
| 474 | Macroautophagy Homo sapiens R-HSA-1632852 | 0.057023 | 0.170708 | 1.585634 | 4.5417376 | ATG3;PRKAA1;PRKAA2;PRKAG2;WIPI2;ATG14;MTOR;RRAGA;ATG101;RB1CC1;CHMP2B;MLST8;CHMP4C;ATG4C;ATG4B;CHMP4B;CHMP3;ULK1;LAMTOR1;ATG4D;ATG5 |
| 475 | Transcription of the HIV genome Homo sapiens R-HSA-167172 | 0.057023 | 0.170708 | 1.585634 | 4.5417376 | GTF2A1;SUPT16H;GTF2A2;CCNT2;RNMT;GTF2B;TAF11;SSRP1;GTF2F2;CDK9;CDK7;POLR2B;POLR2C;POLR2E;TCEA1;NELFB;POLR2H;TAF5;POLR2J;TAF3;TAF2 |
| 476 | Signaling by Hippo Homo sapiens R-HSA-2028269 | 0.058619 | 0.175037 | 2.313866 | 6.5637505 | YAP1;YWHAE;TJP1;WWTR1;MOB1A;YWHAB;STK3;TJP2 |
| 477 | Metabolism of water-soluble vitamins and cofactors Homo sapiens R-HSA-196849 | 0.058715 | 0.175037 | 1.460243 | 4.139875 | PANK4;ABCD4;PANK2;PDXK;SHMT1;QPRT;SLC2A1;MTR;NADSYN1;ACACB;ACACA;HLCS;NADK;AASDHPPT;CYB5A;ABCC1;AMN;GSTO2;GSTO1;MCCC1;SLC52A3;MTHFS;COASY;LMBRD1;BTD;PCCA;NMNAT1;PCCB;CD320 |
| 478 | HSF1-dependent transactivation Homo sapiens R-HSA-3371571 | 0.059731 | 0.177695 | 1.893637 | 5.3360765 | HSPA8;HSP90AA1;HSP90AB1;DNAJB6;UBB;MRPL18;MLST8;EP300;HSPA2;MTOR;HSPA1B;HSPA1A |

| | | | | | | |
|---|---|---|---|---|---|---|
| 479 | RNA Polymerase III Transcription Initiation From Type 2 Promoter Homo sapiens R-HSA-76066 | 0.06071 | 0.179481 | 2.041902 | 5.7206733 | GTF3C1;GTF3C3;CRCP;POLR3B;GTF3C6;POLR3GL;POLR2E;POLR1D;POLR2H;BDP1 |
| 480 | Interleukin-6 family signaling Homo sapiens R-HSA-6783589 | 0.06071 | 0.179481 | 2.041902 | 5.7206733 | STAT1;IL11RA;STAT3;PTPN11;LIFR;CBL;JAK2;IL6ST;IL27RA;JAK1 |
| 481 | Pyruvate metabolism Homo sapiens R-HSA-70268 | 0.06071 | 0.179481 | 2.041902 | 5.7206733 | PDP1;LDHB;PDHX;LDHA;PDHA1;GLO1;PDHB;HAGH;DLD;PDK2 |
| 482 | RIG-I/MDA5 mediated induction of IFN-alpha/beta pathways Homo sapiens R-HSA-168928 | 0.061431 | 0.181235 | 1.515759 | 4.2287177 | APP;MAP3K1;DHX9;UBE2D2;ISG15;HMGB1;TANK;RELA;NFKB1;HERC5;NFKBIA;ITCH;TBK1;IRF3;UBB;IRF2;DHX36;UBC;TAX1BP1;IRF7;EP300;RIPK1;UBE2K;ATG5 |
| 483 | Major pathway of rRNA processing in the nucleolus Homo sapiens R-HSA-6791226 | 0.061973 | 0.182456 | 1.33254 | 3.7058603 | RPL5;RPL30;RPL31;RPLP1;RPS4Y1;RPL6;WDR43;RRP9;RPS15;EXOSC7;RPL7A;RPS19;BMS1;EXOSC8;RPL36;RPLP2;RPL37;RPS10;RPP38;DCAF13;RPL19;RPS9;RPL41;UTP6;RPS7;RPS8;KRR1;RIOK3;RPS5;WDR75;RPL35A;DDX52;RPS25;RPS28;RPS27;RPS29;XRN2;RPL37A;RPP21;MPHOSPH10;TSR1;RPP25;RPL29;RPS21;RPS24;MPHOSPH6 |
| 484 | RNA Polymerase II Pre-transcription Events Homo sapiens R-HSA-674695 | 0.063939 | 0.187854 | 1.57862 | 4.3409307 | GTF2A1;SUPT16H;GTF2A2;CCNT2;GTF2B;TAF11;SSRP1;GTF2F2;CDK9;CDK7;POLR2B;POLR2C;POLR2E;TCEA1;NELFB;POLR2H;TAF5;POLR2J;TAF3;TAF2 |
| 485 | Translesion synthesis by POLI Homo sapiens R-HSA-5656121 | 0.06482 | 0.19005 | 2.429329 | 6.6469768 | RFC1;UBB;RPA3;UBC;REV1;RPA2;POLI |
| 486 | Costimulation by the CD28 family Homo sapiens R-HSA-388841 | 0.067251 | 0.195985 | 1.528198 | 4.1250903 | YES1;SRC;PPP2R5B;PIK3R3;PTPN11;MTOR;CDC42;AKT2;MLST8;HLA-DPB1;AKT1;THEM4;HLA-DRA;TNFRSF14;CSK;GRB2;RAC1;PAK2;HLA-DQA2;HLA-DRB1;HLA-DPA1;HLA-DQB1 |
| 487 | Export of Viral Ribonucleoproteins from Nucleus Homo sapiens R-HSA-168274 | 0.067396 | 0.195985 | 1.909236 | 5.1495476 | RANBP2;NUP214;NUP205;XPO1;POM121C;NUP50;TPR;NUP98;AAAS;RAN;HSPA1A |

| | | | | | | |
|---|---|---|---|---|---|---|
| 488 | Pausing and recovery of Tat-mediated HIV elongation Homo sapiens R-HSA-167238 | 0.067396 | 0.195985 | 1.909236 | 5.1495476 | CDK9;SUPT16H;POLR2B;POLR2C;POLR2E;TCEA1;NELFB;POLR2H;SSRP1;POLR2J;GTF2F2 |
| 489 | Tat-mediated HIV elongation arrest and recovery Homo sapiens R-HSA-167243 | 0.067396 | 0.195985 | 1.909236 | 5.1495476 | CDK9;SUPT16H;POLR2B;POLR2C;POLR2E;TCEA1;NELFB;POLR2H;SSRP1;POLR2J;GTF2F2 |
| 490 | Insulin processing Homo sapiens R-HSA-264876 | 0.068242 | 0.19804 | 2.082543 | 5.5910001 | SLC30A6;EXOC7;SLC30A5;KIF5B;EXOC4;RAB27A;EXOC6;EXOC5;EXOC2 |
| 491 | Transcription-Coupled Nucleotide Excision Repair (TC-NER) Homo sapiens R-HSA-6781827 | 0.068606 | 0.198185 | 1.507274 | 4.038557 | USP7;AQR;RFC1;RPA2;DDB1;POLD4;COPS3;COPS6;CDK7;COPS5;UBB;POLR2B;POLR2C;RPA3;UBC;POLR2E;EP300;PPIE;TCEA1;ERCC5;POLR2H;POLR2J;CUL4B |
| 492 | Intrinsic Pathway for Apoptosis Homo sapiens R-HSA-109606 | 0.068849 | 0.198185 | 1.73594 | 4.6450957 | YWHAE;PPP1R13B;YWHAB;BAD;XIAP;CASP7;AKT2;TP53BP2;NMT1;AKT1;BAX;BAK1;BID;YWHAH |
| 493 | Viral Messenger RNA Synthesis Homo sapiens R-HSA-168325 | 0.068849 | 0.198185 | 1.73594 | 4.6450957 | RANBP2;NUP214;NUP205;AAAS;GTF2F2;POM121C;POLR2B;POLR2C;NUP50;TPR;POLR2E;POLR2H;NUP98;POLR2J |
| 494 | Deactivation of the beta-catenin transactivating complex Homo sapiens R-HSA-3769402 | 0.068849 | 0.198185 | 1.73594 | 4.6450957 | TLE3;TLE2;HDAC1;CBY1;XIAP;PYGO1;XPO1;UBB;APC;AKT2;UBC;AKT1;CTNNB1;MEN1 |
| 495 | Regulation of pyruvate dehydrogenase (PDH) complex Homo sapiens R-HSA-204174 | 0.071151 | 0.204398 | 2.602606 | 6.8785525 | PDP1;PDHX;PDHA1;PDHB;DLD;PDK2 |
| 496 | Cell death signalling via NRAGE, NRIF and NADE Homo sapiens R-HSA-204998 | 0.071752 | 0.20571 | 1.570953 | 4.1387276 | YWHAE;PSENEN;VAV3;ARHGEF12;BAD;RASGRF2;ITSN1;ARHGEF16;APH1A;APH1B;UBB;MCF2L;UBC;CASP2;RAC1;ARHGEF2;SOS1;ECT2;SQSTM1 |
| 497 | RNA Polymerase III Transcription Initiation From Type 1 Promoter Homo sapiens R-HSA-76061 | 0.076164 | 0.217359 | 1.928339 | 4.9652118 | GTF3C1;GTF3C3;CRCP;POLR3B;GTF3C6;POLR3GL;POLR2E;POLR1D;POLR2H;BDP1 |

| | | | | | | |
|---|---|---|---|---|---|---|
| 498 | RNA Polymerase III Transcription Initiation From Type 3 Promoter Homo sapiens R-HSA-76071 | 0.076164 | 0.217359 | 1.928339 | 4.9652118 | CRCP;POU2F1;SNAPC1;POLR3B;SNAPC2;POLR3GL;POLR2E;POLR1D;POLR2H;BDP1 |
| 499 | Positive epigenetic regulation of rRNA expression Homo sapiens R-HSA-5250913 | 0.076654 | 0.217359 | 1.498136 | 3.8478832 | GSK3B;HDAC2;H2AFZ;HDAC1;H3F3A;SMARCA5;DDX21;CHD4;CHD3;DEK;BAZ1B;KAT2A;TAF1D;RBBP4;TAF1B;POLR1D;POLR2E;EP300;RBBP7;POLR2H;SF3B1;HIST1H2AC |
| 500 | NOTCH2 Activation and Transmission of Signal to the Nucleus Homo sapiens R-HSA-2979096 | 0.076733 | 0.217359 | 2.135739 | 5.4833452 | PSENEN;APH1A;NEURL1B;APH1B;UBB;UBC;MIB2;ADAM10 |
| 501 | RIP-mediated NFkB activation via ZBP1 Homo sapiens R-HSA-1810476 | 0.076733 | 0.217359 | 2.135739 | 5.4833452 | NFKBIA;APP;DHX9;RIPK1;HMGB1;TICAM1;RELA;NFKB1 |
| 502 | Regulation of FZD by ubiquitination Homo sapiens R-HSA-4641263 | 0.076733 | 0.217359 | 2.135739 | 5.4833452 | RNF43;USP8;FZD5;UBB;FZD6;UBC;RSPO3;LRP6 |
| 503 | Uptake and function of anthrax toxins Homo sapiens R-HSA-5210891 | 0.076993 | 0.217661 | 2.89151 | 7.4139643 | MAP2K1;MAP2K2;PDCD6IP;FURIN;CALM1 |
| 504 | SUMO is conjugated to E1 (UBA2:SAE1) Homo sapiens R-HSA-3065676 | 0.077852 | 0.217925 | 5.203733 | 13.284833 | SUMO3;SUMO2;UBA2 |
| 505 | GRB7 events in ERBB2 signaling Homo sapiens R-HSA-1306955 | 0.077852 | 0.217925 | 5.203733 | 13.284833 | GRB7;ERBB3;ERBB2 |
| 506 | Beta oxidation of hexanoyl-CoA to butanoyl-CoA Homo sapiens R-HSA-77350 | 0.077852 | 0.217925 | 5.203733 | 13.284833 | HADHB;HADHA;HADH |
| 507 | Propionyl-CoA catabolism Homo sapiens R-HSA-71032 | 0.077852 | 0.217925 | 5.203733 | 13.284833 | MCEE;PCCA;PCCB |
| 508 | Signaling by MST1 Homo sapiens R-HSA-8852405 | 0.077852 | 0.217925 | 5.203733 | 13.284833 | SPINT1;MST1;SPINT2 |

| | | | | | | |
|---|---|---|---|---|---|---|
| 509 | Meiosis Homo sapiens R-HSA-1500620 | 0.079766 | 0.22128 | 1.447196 | 3.6594658 | SMC3;SYNE2;PSMC3IP;RAD21;LMNA;NBN;HIST1H2AC;SUN1;UBE2I;H2AFZ;TOP3A;H3F3A;RPA2;HSPA2;SMC1A;TERF1;MLH1;MLH3;RAD50;STAG1;STAG2;RPA3;TERF2IP;ATM;ATR |
| 510 | Regulation of TP53 Activity through Phosphorylation Homo sapiens R-HSA-6804756 | 0.0805 | 0.22128 | 1.433457 | 3.6115865 | PRKAA1;PRKAA2;PRKAG2;UBB;UBC;NBN;TOPBP1;ATRIP;SUPT16H;CSNK2A1;CSNK2A2;TOP3A;TAF11;RPA2;SSRP1;HIPK1;CCNA1;RAD50;CDK5;RPA3;RAD17;ATM;TAF5;TAF3;TAF2;ATR |
| 511 | Vitamin C (ascorbate) metabolism Homo sapiens R-HSA-196836 | 0.080762 | 0.22128 | 3.469484 | 8.730063 | CYB5A;GSTO2;GSTO1;SLC2A1 |
| 512 | Cleavage of the damaged pyrimidine Homo sapiens R-HSA-110329 | 0.080762 | 0.22128 | 3.469484 | 8.730063 | MBD4;OGG1;NEIL1;UNG |
| 513 | Recognition and association of DNA glycosylase with site containing an affected pyrimidine Homo sapiens R-HSA-110328 | 0.080762 | 0.22128 | 3.469484 | 8.730063 | MBD4;OGG1;NEIL1;UNG |
| 514 | Depyrimidination Homo sapiens R-HSA-73928 | 0.080762 | 0.22128 | 3.469484 | 8.730063 | MBD4;OGG1;NEIL1;UNG |
| 515 | AMPK inhibits chREBP transcriptional activation activity Homo sapiens R-HSA-163680 | 0.080762 | 0.22128 | 3.469484 | 8.730063 | PRKAA2;PRKAG2;ADIPOR1;ADIPOR2 |
| 516 | Downregulation of ERBB4 signaling Homo sapiens R-HSA-1253288 | 0.080762 | 0.22128 | 3.469484 | 8.730063 | ITCH;UBB;UBC;WWP1 |
| 517 | RNF mutants show enhanced WNT signaling and proliferation Homo sapiens R-HSA-5340588 | 0.080762 | 0.22128 | 3.469484 | 8.730063 | RNF43;FZD5;FZD6;LRP6 |
| 518 | Axonal growth inhibition (RHOA activation) Homo sapiens R-HSA-193634 | 0.080762 | 0.22128 | 3.469484 | 8.730063 | ARHGDIA;ARHGEF1;RHOA;RTN4 |

| | | | | | | |
|---|---|---|---|---|---|---|
| 519 | Ketone body metabolism Homo sapiens R-HSA-74182 | 0.080762 | 0.22128 | 3.469484 | 8.730063 | HMGCL;BDH2;HMGCS2;ACAT1 |
| 520 | MyD88-independent TLR3/TLR4 cascade Homo sapiens R-HSA-166166 | 0.081606 | 0.222305 | 1.409573 | 3.5321891 | ATF2;APP;DHX9;UBE2D2;CUL1;HMGB1;TANK;RELA;TBK1;RPS6KA5;UBB;UBC;MAPK1;RIPK1;MAPK3;MAP2K1;FBXW11;PTPN11;TICAM1;NFKB1;NFKBIA;MAPK10;CREB1;IRF3;TNIP2;IRF7;UBE2V1;BIRC2 |
| 521 | Toll Like Receptor 3 (TLR3) Cascade Homo sapiens R-HSA-168164 | 0.081606 | 0.222305 | 1.409573 | 3.5321891 | ATF2;APP;DHX9;UBE2D2;CUL1;HMGB1;TANK;RELA;TBK1;RPS6KA5;UBB;UBC;MAPK1;RIPK1;MAPK3;MAP2K1;FBXW11;PTPN11;TICAM1;NFKB1;NFKBIA;MAPK10;CREB1;IRF3;TNIP2;IRF7;UBE2V1;BIRC2 |
| 522 | TRIF-mediated TLR3/TLR4 signaling Homo sapiens R-HSA-937061 | 0.081606 | 0.222305 | 1.409573 | 3.5321891 | ATF2;APP;DHX9;UBE2D2;CUL1;HMGB1;TANK;RELA;TBK1;RPS6KA5;UBB;UBC;MAPK1;RIPK1;MAPK3;MAP2K1;FBXW11;PTPN11;TICAM1;NFKB1;NFKBIA;MAPK10;CREB1;IRF3;TNIP2;IRF7;UBE2V1;BIRC2 |
| 523 | Iron uptake and transport Homo sapiens R-HSA-917937 | 0.081854 | 0.222555 | 1.675972 | 4.1946575 | ATP6V1A;ATP6V1G1;ATP6V0B;ATP6V0E1;TFRC;ATP6AP1;TCIRG1;MCOLN1;ATP6V0A4;ATP6V0D1;ATP6V0C;ATP6V1C1;ATP6V1C2;FTL |
| 524 | Gap-filling DNA repair synthesis and ligation in TC-NER Homo sapiens R-HSA-6782210 | 0.082569 | 0.22407 | 1.53432 | 3.8267882 | USP7;AQR;RFC1;RPA2;DDB1;POLD4;CDK7;UBB;POLR2B;POLR2C;RPA3;UBC;POLR2E;EP300;PPIE;TCEA1;POLR2H;POLR2J;CUL4B |
| 525 | Constitutive Signaling by AKT1 E17K in Cancer Homo sapiens R-HSA-5674400 | 0.086229 | 0.231902 | 1.952258 | 4.7844864 | GSK3B;CDKN1B;CREB1;BAD;AKT2;RPS6KB2;MLST8;AKT1;MTOR |
| 526 | Synthesis and interconversion of nucleotide di- and triphosphates Homo sapiens R-HSA-499943 | 0.086229 | 0.231902 | 1.952258 | 4.7844864 | DTYMK;RRM2B;TXNRD1;GSR;AK2;NME2;CMPK1;CTPS2;AK9 |
| 527 | Glyoxylate metabolism and glycine degradation Homo sapiens R-HSA-389661 | 0.086229 | 0.231902 | 1.952258 | 4.7844864 | BCKDHA;PDHX;PDHA1;NDUFAB1;AMT;OGDH;DLST;PDHB;DLD |
| 528 | Pre-NOTCH Processing in Golgi Homo sapiens R-HSA-1912420 | 0.08627 | 0.231902 | 2.208338 | 5.4110268 | B4GALT1;SEL1L;ATP2A3;ATP2A2;TMED2;FURIN;ST3GAL6 |

| | | | | | | |
|---|---|---|---|---|---|---|
| 529 | Oxygen-dependent proline hydroxylation of Hypoxia-inducible Factor Alpha Homo sapiens R-HSA-1234176 | 0.08627 | 0.231902 | 2.208338 | 5.4110268 | EGLN3;EGLN2;UBB;UBE2D2;UBC;HIF3A;HIF1A |
| 530 | Biosynthesis of the N-glycan precursor (dolichol lipid-linked oligosaccharide, LLO) and transfer to a nascent protein Homo sapiens R-HSA-446193 | 0.087921 | 0.235893 | 1.452276 | 3.5309456 | DPAGT1;SLC35A1;ALG8;CMAS;ALG6;ALG5;GFPT1;ALG14;ALG2;ALG12;ALG1;GMPPB;DPM2;DPM3;NAGK;DOLPP1;NEU1;PGM3;SLC17A5;ST3GAL6;NANS;GNE;MPDU1 |
| 531 | PKMTs methylate histone lysines Homo sapiens R-HSA-3214841 | 0.092599 | 0.247976 | 1.671379 | 3.9770149 | KMT2E;SUZ12;SETD2;SETD3;KMT2A;KMT2C;KMT2B;SETDB2;EHMT1;MECOM;RBBP4;SMYD2;RBBP7 |
| 532 | Negative regulation of FGFR3 signaling Homo sapiens R-HSA-5654732 | 0.093844 | 0.250369 | 1.826729 | 4.3222585 | UBB;SRC;MKNK1;UBC;MAPK1;PTPN11;BRAF;GRB2;CBL;MAPK3 |
| 533 | YAP1- and WWTR1 (TAZ)-stimulated gene expression Homo sapiens R-HSA-2032785 | 0.093844 | 0.250369 | 1.826729 | 4.3222585 | YAP1;NCOA1;NCOA2;WWTR1;CHD9;TBL1XR1;TGS1;HIPK1;TEAD1;TEAD2 |
| 534 | RNA Polymerase II HIV Promoter Escape Homo sapiens R-HSA-167162 | 0.096325 | 0.252331 | 1.620001 | 3.790842 | GTF2A1;GTF2A2;GTF2B;TAF11;GTF2F2;CDK7;POLR2B;POLR2C;POLR2E;POLR2H;TAF5;POLR2J;TAF3;TAF2 |
| 535 | RNA Polymerase II Promoter Escape Homo sapiens R-HSA-73776 | 0.096325 | 0.252331 | 1.620001 | 3.790842 | GTF2A1;GTF2A2;GTF2B;TAF11;GTF2F2;CDK7;POLR2B;POLR2C;POLR2E;POLR2H;TAF5;POLR2J;TAF3;TAF2 |
| 536 | RNA Polymerase II Transcription Initiation And Promoter Clearance Homo sapiens R-HSA-76042 | 0.096325 | 0.252331 | 1.620001 | 3.790842 | GTF2A1;GTF2A2;GTF2B;TAF11;GTF2F2;CDK7;POLR2B;POLR2C;POLR2E;POLR2H;TAF5;POLR2J;TAF3;TAF2 |
| 537 | RNA Polymerase II Transcription Initiation Homo sapiens R-HSA-75953 | 0.096325 | 0.252331 | 1.620001 | 3.790842 | GTF2A1;GTF2A2;GTF2B;TAF11;GTF2F2;CDK7;POLR2B;POLR2C;POLR2E;POLR2H;TAF5;POLR2J;TAF3;TAF2 |

| | | | | | | |
|---|---|---|---|---|---|---|
| 538 | RNA Polymerase II Transcription Pre-Initiation And Promoter Opening Homo sapiens R-HSA-73779 | 0.096325 | 0.252331 | 1.620001 | 3.790842 | GTF2A1;GTF2A2;GTF2B;TAF11;GTF2F2;CDK7;POLR2B;POLR2C;POLR2E;POLR2H;TAF5;POLR2J;TAF3;TAF2 |
| 539 | HIV Transcription Initiation Homo sapiens R-HSA-167161 | 0.096325 | 0.252331 | 1.620001 | 3.790842 | GTF2A1;GTF2A2;GTF2B;TAF11;GTF2F2;CDK7;POLR2B;POLR2C;POLR2E;POLR2H;TAF5;POLR2J;TAF3;TAF2 |
| 540 | Interleukin-1 signaling Homo sapiens R-HSA-446652 | 0.096325 | 0.252331 | 1.620001 | 3.790842 | MAP3K3;MAP2K1;IL1R1;FBXW11;PELI3;CUL1;IRAK4;NFKB1;UBB;TNIP2;UBC;PELI1;UBE2V1;SQSTM1 |
| 541 | Regulation of innate immune responses to cytosolic DNA Homo sapiens R-HSA-3134975 | 0.096887 | 0.252331 | 2.313278 | 5.3996829 | TBK1;UBB;IRF3;UBC;DDX41;TRIM56 |
| 542 | Mismatch Repair Homo sapiens R-HSA-5358508 | 0.096887 | 0.252331 | 2.313278 | 5.3996829 | MSH6;POLD4;MSH3;RPA3;RPA2;MLH1 |
| 543 | Constitutive Signaling by NOTCH1 HD Domain Mutants Homo sapiens R-HSA-2691232 | 0.096887 | 0.252331 | 2.313278 | 5.3996829 | NEURL1B;ADAM17;UBB;UBC;MIB2;ADAM10 |
| 544 | Signaling by NOTCH1 HD Domain Mutants in Cancer Homo sapiens R-HSA-2691230 | 0.096887 | 0.252331 | 2.313278 | 5.3996829 | NEURL1B;ADAM17;UBB;UBC;MIB2;ADAM10 |
| 545 | NRIF signals cell death from the nucleus Homo sapiens R-HSA-205043 | 0.096887 | 0.252331 | 2.313278 | 5.3996829 | PSENEN;APH1A;APH1B;UBB;UBC;SQSTM1 |
| 546 | p130Cas linkage to MAPK signaling for integrins Homo sapiens R-HSA-372708 | 0.096887 | 0.252331 | 2.313278 | 5.3996829 | RAP1B;SRC;ITGA2B;TLN1;PTK2;BCAR1 |
| 547 | Pink/Parkin Mediated Mitophagy Homo sapiens R-HSA-5205685 | 0.097827 | 0.25385 | 1.983058 | 4.6097324 | PINK1;UBB;UBC;MFN1;VDAC1;SQSTM1;TOMM5;ATG5 |
| 548 | Mitophagy Homo sapiens R-HSA-5205647 | 0.097827 | 0.25385 | 1.983058 | 4.6097324 | PINK1;UBB;UBC;MFN1;VDAC1;SQSTM1;TOMM5;ATG5 |

| | | | | | | |
|---|---|---|---|---|---|---|
| 549 | TRAF6 mediated induction of NFkB and MAP kinases upon TLR7/8 or 9 activation Homo sapiens R-HSA-975138 | 0.099419 | 0.257511 | 1.41263 | 3.2609368 | ATF2;APP;MAP2K1;MAP3K1;FBXW11;DHX9;PELI3;CUL1;HMGB1;IRAK4;ECSIT;RELA;NFKB1;NFKBIA;MAPK10;RPS6KA5;CREB1;UBB;TNIP2;UBC;PELI1;MAPK1;UBE2V1;MAPK3 |
| 550 | Cargo concentration in the ER Homo sapiens R-HSA-5694530 | 0.099985 | 0.258038 | 1.735446 | 3.9962673 | LMAN1;MCFD2;TMED10;SERPINA1;SAR1B;LMAN2;TMED2;STX5;MIA3;CD59;FOLR1 |
| 551 | Smooth Muscle Contraction Homo sapiens R-HSA-445355 | 0.099985 | 0.258038 | 1.735446 | 3.9962673 | MYL6;ANXA2;TPM4;TPM3;TPM1;MYL6B;TLN1;CALM1;MYL12A;VCL;MYL12B |
| 552 | Epigenetic regulation of gene expression Homo sapiens R-HSA-212165 | 0.100238 | 0.258222 | 1.339206 | 3.080449 | GSK3B;DNMT1;HDAC2;HDAC1;PHF1;DDX21;ARID4B;CHD4;CHD3;TAF1D;RBBP4;TAF1B;SAP30L;POLR2E;EP300;SAP18;SUDS3;RBBP7;POLR2H;SF3B1;HIST1H2AC;SUZ12;H2AFZ;H3F3A;MBD2;SMARCA5;DEK;BAZ1B;SIRT1;CDK7;KAT2A;POLR1D |
| 553 | Mitotic Prophase Homo sapiens R-HSA-68875 | 0.100553 | 0.258564 | 1.353254 | 3.108524 | RB1;NUP205;SET;USO1;AAAS;SMC2;GOLGA2;POM121C;TPR;LMNA;MAPK1;HIST1H2AC;TMPO;MAPK3;RAB2A;NUP214;RANBP2;H2AFZ;NEK7;H3F3A;LEMD2;GORASP2;NUP50;PPP2R2D;NUP98;NCAPD3;LPIN2;BLZF1;LPIN3;MCPH1 |
| 554 | RHO GTPases activate PKNs Homo sapiens R-HSA-5625740 | 0.105846 | 0.271685 | 1.487938 | 3.3415623 | YWHAE;NCOA2;PPP1R12A;KDM1A;YWHAB;H2AFZ;H3F3A;RHOA;MYL12B;MYL6;MYH9;MYH14;PKN2;RAC1;PKN1;MYH10;HIST1H2AC;YWHAH |
| 555 | Transport of Mature mRNA derived from an Intron-Containing Transcript Homo sapiens R-HSA-159236 | 0.108404 | 0.274908 | 1.446694 | 3.2144027 | NUP214;RANBP2;NUP205;NXT1;EIF4A3;UPF3B;THOC2;AAAS;SRRM1;DDX39A;DDX39B;POM121C;U2AF2;NUP50;TPR;SRSF2;SRSF4;NUP98;SLU7;SRSF11 |
| 556 | Defective EXT2 causes exostoses 2 Homo sapiens R-HSA-3656237 | 0.108455 | 0.274908 | 2.478278 | 5.5052923 | SDC4;SDC2;SDC1;AGRN;HSPG2 |
| 557 | Defective EXT1 causes exostoses 1, TRPS2 and CHDS Homo sapiens R-HSA-3656253 | 0.108455 | 0.274908 | 2.478278 | 5.5052923 | SDC4;SDC2;SDC1;AGRN;HSPG2 |

| | | | | | | |
|---|---|---|---|---|---|---|
| 558 | Hyaluronan uptake and degradation Homo sapiens R-HSA-2160916 | 0.108455 | 0.274908 | 2.478278 | 5.5052923 | HYAL2;HEXB;HEXA;GUSB;SLC9A1 |
| 559 | NOTCH2 intracellular domain regulates transcription Homo sapiens R-HSA-2197563 | 0.108455 | 0.274908 | 2.478278 | 5.5052923 | CREB1;EP300;HES1;MAML3;RBPJ |
| 560 | Caspase-mediated cleavage of cytoskeletal proteins Homo sapiens R-HSA-264870 | 0.108455 | 0.274908 | 2.478278 | 5.5052923 | CASP7;GSN;DBNL;SPTAN1;ADD1 |
| 561 | The NLRP3 inflammasome Homo sapiens R-HSA-844456 | 0.108455 | 0.274908 | 2.478278 | 5.5052923 | PYCARD;PANX1;APP;HSP90AB1;SUGT1 |
| 562 | RNA Polymerase II Transcription Homo sapiens R-HSA-73857 | 0.108949 | 0.275669 | 1.312231 | 2.9090512 | SLBP;GTF2A1;GTF2A2;CCNT2;RNMT;GTF2B;EIF4A3;U2AF2;POLR2B;POLR2C;PAPOLA;POLR2E;NELFB;POLR2H;POLR2J;SRSF11;SUPT16H;TAF11;UPF3B;THOC2;SSRP1;GTF2F2;SRRM1;CDK9;CDK7;DDX39A;DDX39B;SRSF2;TCEA1;SRSF4;TAF5;SLU7;TAF3;TAF2 |
| 563 | RNA Polymerase I, RNA Polymerase III, and Mitochondrial Transcription Homo sapiens R-HSA-504046 | 0.109876 | 0.27752 | 1.323177 | 2.9221079 | GTF3C1;GTF3C3;HDAC2;CRCP;GTF3C6;HDAC1;CHD4;CHD3;TAF1D;SNAPC1;RBBP4;TAF1B;SNAPC2;POLR2E;RBBP7;POLR2H;HIST1H2AC;MAPK3;POU2F1;SSB;POLR3GL;H2AFZ;H3F3A;MBD2;CDK7;KAT2A;POLR3B;NFIA;NFIB;POLR1D;TFAM;BDP1 |
| 564 | MyD88:Mal cascade initiated on plasma membrane Homo sapiens R-HSA-166058 | 0.111171 | 0.278021 | 1.368034 | 3.0051371 | ATF2;APP;DHX9;PELI3;CUL1;HMGB1;ECSIT;RELA;RPS6KA5;UBB;UBC;MAPK1;CD36;MAPK3;MAP2K1;MAP3K1;FBXW11;IRAK4;NFKB1;SIGIRR;NFKBIA;MAPK10;CREB1;TNIP2;PELI1;UBE2V1 |
| 565 | Toll Like Receptor TLR1:TLR2 Cascade Homo sapiens R-HSA-168179 | 0.111171 | 0.278021 | 1.368034 | 3.0051371 | ATF2;APP;DHX9;PELI3;CUL1;HMGB1;ECSIT;RELA;RPS6KA5;UBB;UBC;MAPK1;CD36;MAPK3;MAP2K1;MAP3K1;FBXW11;IRAK4;NFKB1;SIGIRR;NFKBIA;MAPK10;CREB1;TNIP2;PELI1;UBE2V1 |
| 566 | Toll Like Receptor TLR6:TLR2 Cascade Homo sapiens R-HSA-168188 | 0.111171 | 0.278021 | 1.368034 | 3.0051371 | ATF2;APP;DHX9;PELI3;CUL1;HMGB1;ECSIT;RELA;RPS6KA5;UBB;UBC;MAPK1;CD36;MAPK3;MAP2K1;MAP3K1;FBXW11;IRAK4;NFKB1;SIGIRR;NFKBIA;MAPK10;CREB1;TNIP2;PELI1;UBE2V1 |

| | | | | | | |
|---|---|---|---|---|---|---|
| 567 | Toll Like Receptor 2 (TLR2) Cascade Homo sapiens R-HSA-181438 | 0.111171 | 0.278021 | 1.368034 | 3.0051371 | ATF2;APP;DHX9;PELI3;CUL1;HMGB1;ECSIT;RELA;RPS6KA5;UBB;UBC;MAPK1;CD36;MAPK3;MAP2K1;MAP3K1;FBXW11;IRAK4;NFKB1;SIGIRR;NFKBIA;MAPK10;CREB1;TNIP2;PELI1;UBE2V1 |
| 568 | Ephrin signaling Homo sapiens R-HSA-3928664 | 0.111248 | 0.278021 | 2.02418 | 4.445092 | EPHB6;EFNB2;SDCBP;SRC;RAC1;PAK2;MYL12A |
| 569 | Signaling by NODAL Homo sapiens R-HSA-1181150 | 0.111248 | 0.278021 | 2.02418 | 4.445092 | SMAD2;DRAP1;SMAD4;SMAD3;FURIN;ACVR1B;ACVR2B |
| 570 | Amine compound SLC transporters Homo sapiens R-HSA-425428 | 0.113746 | 0.282775 | 1.735281 | 3.7721269 | SLC6A6;SLC44A3;SLC14A2;SLC44A4;SLC44A1;SLC44A2;SLC6A13;SLC6A12;RHCG;RHBG |
| 571 | NEP/NS2 Interacts with the Cellular Export Machinery Homo sapiens R-HSA-168333 | 0.113746 | 0.282775 | 1.735281 | 3.7721269 | RANBP2;NUP214;NUP205;XPO1;POM121C;NUP50;TPR;NUP98;AAAS;RAN |
| 572 | Termination of translesion DNA synthesis Homo sapiens R-HSA-5656169 | 0.113746 | 0.282775 | 1.735281 | 3.7721269 | POLD4;UBB;RFC1;USP43;RPA3;UBC;REV1;POLI;RPA2;ISG15 |
| 573 | DNA Double-Strand Break Repair Homo sapiens R-HSA-5693532 | 0.114863 | 0.285054 | 1.278119 | 2.7658637 | MUS81;RIF1;PRKDC;RNF8;BABAM1;POLD4;HERC2;PPP4C;PPP4R2;UBB;UBC;UIMC1;SUMO2;POLL;NBN;TP53BP1;TOPBP1;KPNA2;ATRIP;KDM4A;XRCC6;KDM4B;UBE2I;RFC1;XRCC5;POLM;TOP3A;EYA4;SMARCA5;RPA2;SIRT6;BAZ1B;SPIDR;CCNA1;RAD50;RPA3;RAD17;ATM;ATR |
| 574 | tRNA processing in the nucleus Homo sapiens R-HSA-6784531 | 0.118772 | 0.29357 | 1.475398 | 3.1434047 | NUP214;RANBP2;NUP205;ELAC2;DDX1;RTCB;ZBTB8OS;C2ORF49;AAAS;POM121C;NUP50;TPR;RPP21;NUP98;RPP25;RPP38;RAN |
| 575 | Signaling by WNT in cancer Homo sapiens R-HSA-4791275 | 0.119237 | 0.29357 | 1.659884 | 3.5299787 | GSK3B;RNF43;CTBP2;TNKS2;FZD5;APC;CSNK1A1;FZD6;PPP2R5B;CTNNB1;LRP6 |
| 576 | Negative regulators of RIG-I/MDA5 signaling Homo sapiens R-HSA-936440 | 0.119237 | 0.29357 | 1.659884 | 3.5299787 | HERC5;ITCH;TBK1;UBB;IRF3;UBE2D2;UBC;TAX1BP1;ISG15;UBE2K;ATG5 |
| 577 | EGFR interacts with phospholipase C-gamma Homo sapiens R-HSA-212718 | 0.119237 | 0.29357 | 1.659884 | 3.5299787 | AHCYL1;CREB1;ADCY9;PRKAR2A;PRKCD;ITPR2;ITPR3;PLCG1;CALM1;EGFR;ADCY6 |

| | | | | | | |
|---|---|---|---|---|---|---|
| 578 | Processing and activation of SUMO Homo sapiens R-HSA-3215018 | 0.12036 | 0.29357 | 2.775408 | 5.876292 | UBE2I;SUMO3;SUMO2;UBA2 |
| 579 | AKT phosphorylates targets in the nucleus Homo sapiens R-HSA-198693 | 0.12036 | 0.29357 | 2.775408 | 5.876292 | CREB1;AKT2;RPS6KB2;AKT1 |
| 580 | RHO GTPases Activate Rhotekin and Rhophilins Homo sapiens R-HSA-5666185 | 0.12036 | 0.29357 | 2.775408 | 5.876292 | RTKN;TAX1BP3;RHPN2;RHOA |
| 581 | Inactivation of Cdc42 and Rac Homo sapiens R-HSA-428543 | 0.12036 | 0.29357 | 2.775408 | 5.876292 | CDC42;RAC1;SRGAP3;SRGAP1 |
| 582 | p75NTR regulates axonogenesis Homo sapiens R-HSA-193697 | 0.12036 | 0.29357 | 2.775408 | 5.876292 | ARHGDIA;ARHGEF1;RHOA;RTN4 |
| 583 | Pentose phosphate pathway (hexose monophosphate shunt) Homo sapiens R-HSA-71336 | 0.12036 | 0.29357 | 2.775408 | 5.876292 | TALDO1;PGLS;TKT;DERA |
| 584 | Laminin interactions Homo sapiens R-HSA-3000157 | 0.121844 | 0.295165 | 1.850735 | 3.8958283 | ITGB1;ITGA3;LAMB2;LAMA3;LAMB1;ITGA6;LAMC1;HSPG2 |
| 585 | MicroRNA (miRNA) biogenesis Homo sapiens R-HSA-203927 | 0.121844 | 0.295165 | 1.850735 | 3.8958283 | AGO3;POLR2B;POLR2C;POLR2E;DGCR8;POLR2H;POLR2J;RAN |
| 586 | RAF-independent MAPK1/3 activation Homo sapiens R-HSA-112409 | 0.121844 | 0.295165 | 1.850735 | 3.8958283 | MAP2K1;MAP2K2;MAPK1;PTPN11;IL6ST;JAK2;MAPK3;JAK1 |
| 587 | Signaling by BMP Homo sapiens R-HSA-201451 | 0.121844 | 0.295165 | 1.850735 | 3.8958283 | SMAD4;BMPR2;ZFYVE16;SMAD9;SMAD6;BMPR1B;ACVR2B;SMAD7 |
| 588 | Processing of DNA double-strand break ends Homo sapiens R-HSA-5693607 | 0.123045 | 0.297569 | 1.376891 | 2.884865 | UBE2I;TOP3A;RNF8;RPA2;SIRT6;BABAM1;CCNA1;RAD50;HERC2;PPP4C;PPP4R2;UBB;RPA3;UIMC1;SUMO2;UBC;RAD17;ATM;NBN;TOPBP1;TP53BP1;ATRIP;ATR |
| 589 | Glutathione conjugation Homo sapiens R-HSA-156590 | 0.123571 | 0.298332 | 1.601895 | 3.349468 | GGCT;GSTK1;GCLC;GGT6;GSTO2;GSTO1;GSTP1;MGST3;ESD;AKR1A1;MGST2;CNDP2 |

| | | | | | | |
|---|---|---|---|---|---|---|
| 590 | Translesion synthesis by REV1 Homo sapiens R-HSA-110312 | 0.126854 | 0.30401 | 2.081816 | 4.2983666 | RFC1;UBB;RPA3;UBC;REV1;RPA2 |
| 591 | Regulation of KIT signaling Homo sapiens R-HSA-1433559 | 0.126854 | 0.30401 | 2.081816 | 4.2983666 | YES1;SRC;KIT;GRB2;CBL;SOS1 |
| 592 | Signaling by Retinoic Acid Homo sapiens R-HSA-5362517 | 0.126992 | 0.30401 | 1.555911 | 3.2108315 | PDHX;RARG;CPT1A;PDHA1;AKR1C3;PDHB;DHRS3;RDH11;RDH10;RDH14;RDH13;DLD;PDK2 |
| 593 | Cooperation of PDCL (PhLP1) and TRiC/CCT in G-protein beta folding Homo sapiens R-HSA-6814122 | 0.126992 | 0.30401 | 1.555911 | 3.2108315 | CCT3;CCT2;CSNK2A1;CSNK2A2;GNAI3;GNG11;GNAI2;GNG5;TCP1;CCT8;CCT7;CCT5;CCT4 |
| 594 | Fatty Acyl-CoA Biosynthesis Homo sapiens R-HSA-75105 | 0.126992 | 0.30401 | 1.555911 | 3.2108315 | ELOVL1;ACOT9;AGK;TECR;ACOT12;ACOT13;ACSL5;HSD17B12;ACSL3;ACACA;ACOT2;THEM4;ACOT4 |
| 595 | DNA Damage Bypass Homo sapiens R-HSA-73893 | 0.129682 | 0.308277 | 1.518555 | 3.1019125 | VCP;RFC1;USP43;REV1;RPA2;ISG15;DDB1;POLD4;UBB;RPA3;UBC;USP1;POLI;CUL4B |
| 596 | Platelet calcium homeostasis Homo sapiens R-HSA-418360 | 0.129792 | 0.308277 | 1.735116 | 3.5427997 | P2RX4;STIM1;ATP2A3;ITPR2;ATP2A2;ORAI1;ITPR3;SRI;CALM1 |
| 597 | Peroxisomal lipid metabolism Homo sapiens R-HSA-390918 | 0.129792 | 0.308277 | 1.735116 | 3.5427997 | ALDH3A2;NUDT7;SCP2;ECI2;HSD17B4;HACL1;ACOX3;ACAA1;CRAT |
| 598 | Rho GTPase cycle Homo sapiens R-HSA-194840 | 0.129994 | 0.308277 | 1.287741 | 2.6273361 | ARHGAP8;RASGRF2;FAM13A;ITSN1;GDI2;ARHGAP18;ARHGAP6;ARHGAP35;CDC42;RHOT2;OPHN1;ARHGDIA;MCF2L;ARHGDIB;SRGAP3;RAC1;ECT2;SRGAP1;VAV3;RALBP1;ARHGEF12;ARHGEF16;OCRL;ARHGAP28;ARAP1;RHOD;MYO9A;RHOA;BCR;TRIP10;ARHGEF2;SOS1;RHOQ |
| 599 | Choline catabolism Homo sapiens R-HSA-6798163 | 0.130292 | 0.308277 | 3.468932 | 7.0696126 | DMGDH;SLC44A1;ALDH7A1 |
| 600 | Acyl chain remodeling of CL Homo sapiens R-HSA-1482798 | 0.130292 | 0.308277 | 3.468932 | 7.0696126 | HADHB;HADHA;TAZ |
| 601 | Beta-oxidation of very long chain fatty acids Homo sapiens R-HSA-390247 | 0.130292 | 0.308277 | 3.468932 | 7.0696126 | HSD17B4;ACAA1;CRAT |

| | | | | | | |
|---|---|---|---|---|---|---|
| 602 | Negative regulation of FGFR4 signaling Homo sapiens R-HSA-5654733 | 0.135818 | 0.32082 | 1.652542 | 3.2991984 | UBB;SRC;MKNK1;UBC;MAPK1;PTPN11;BRAF;GRB2;CBL;MAPK3 |
| 603 | Transport of Mature Transcript to Cytoplasm Homo sapiens R-HSA-72202 | 0.136589 | 0.322105 | 1.375589 | 2.738496 | NUP214;RANBP2;SLBP;NUP205;NXT1;EIF4A3;UPF3B;THOC2;AAAS;SRRM1;DDX39A;DDX39B;POM121C;U2AF2;NUP50;TPR;SRSF2;SRSF4;NUP98;SLU7;SRSF11 |
| 604 | Syndecan interactions Homo sapiens R-HSA-3000170 | 0.139586 | 0.328629 | 1.868353 | 3.6789218 | ITGB1;TGFB1;SDC4;TRAPPC4;SDC2;SDC1;ITGA6 |
| 605 | Generation of second messenger molecules Homo sapiens R-HSA-202433 | 0.14041 | 0.328934 | 1.59062 | 3.1226864 | VASP;ENAH;HLA-DPB1;HLA-DRA;EVL;PLCG1;PAK2;HLA-DQA2;HLA-DRB1;HLA-DPA1;HLA-DQB1 |
| 606 | Interactions of Rev with host cellular proteins Homo sapiens R-HSA-177243 | 0.14041 | 0.328934 | 1.59062 | 3.1226864 | RANBP2;NUP214;NUP205;NPM1;XPO1;POM121C;NUP50;TPR;NUP98;AAAS;RAN |
| 607 | Interactions of Vpr with host cellular proteins Homo sapiens R-HSA-176033 | 0.14041 | 0.328934 | 1.59062 | 3.1226864 | RANBP2;NUP214;NUP205;POM121C;NUP50;TPR;PSIP1;NUP98;AAAS;SLC25A4;KPNA1 |
| 608 | The activation of arylsulfatases Homo sapiens R-HSA-1663150 | 0.14509 | 0.335476 | 2.168353 | 4.1857905 | ARSA;ARSK;SUMF2;ARSB;SUMF1 |
| 609 | Initiation of Nuclear Envelope Reformation Homo sapiens R-HSA-2995383 | 0.14509 | 0.335476 | 2.168353 | 4.1857905 | ANKLE2;LMNA;PPP2R2A;LEMD2;TMPO |
| 610 | Nuclear Envelope Reassembly Homo sapiens R-HSA-2995410 | 0.14509 | 0.335476 | 2.168353 | 4.1857905 | ANKLE2;LMNA;PPP2R2A;LEMD2;TMPO |
| 611 | AKT phosphorylates targets in the cytosol Homo sapiens R-HSA-198323 | 0.14509 | 0.335476 | 2.168353 | 4.1857905 | GSK3B;CDKN1B;BAD;AKT2;AKT1 |
| 612 | Activation of DNA fragmentation factor Homo sapiens R-HSA-211227 | 0.14509 | 0.335476 | 2.168353 | 4.1857905 | H1F0;HIST1H1E;HMGB1;HIST1H1C;KPNA1 |

| | | | | | | |
|---|---|---|---|---|---|---|
| 613 | Apoptosis induced DNA fragmentation Homo sapiens R-HSA-140342 | 0.14509 | 0.335476 | 2.168353 | 4.1857905 | H1F0;HIST1H1E;HMGB1;HIST1H1C;KPNA1 |
| 614 | Signal regulatory protein (SIRP) family interactions Homo sapiens R-HSA-391160 | 0.14509 | 0.335476 | 2.168353 | 4.1857905 | SRC;PTPN11;GRB2;CD47;PTK2 |
| 615 | Synthesis, secretion, and inactivation of Glucose-dependent Insulinotropic Polypeptide (GIP) Homo sapiens R-HSA-400511 | 0.14509 | 0.335476 | 2.168353 | 4.1857905 | SPCS3;SPCS2;SPCS1;SEC11A;SEC11C |
| 616 | tRNA processing Homo sapiens R-HSA-72306 | 0.14647 | 0.338118 | 1.296303 | 2.4901115 | NUP205;DDX1;URM1;RTCB;ZBTB8OS;C2ORF49;EPRS;AAAS;MTO1;POM121C;TPR;TYW1;RPP38;NUP214;RANBP2;TRMT10A;NSUN2;ELAC2;QTRT1;CTU1;NUP50;TRMT6;RPP21;NUP98;TRMT11;RPP25;RAN;TRMT61B |
| 617 | Phosphorylation of CD3 and TCR zeta chains Homo sapiens R-HSA-202427 | 0.148645 | 0.342581 | 1.734952 | 3.3071612 | HLA-DPB1;HLA-DRA;PTPRJ;CSK;HLA-DQA2;HLA-DRB1;HLA-DPA1;HLA-DQB1 |
| 618 | Metabolism of nucleotides Homo sapiens R-HSA-15869 | 0.14992 | 0.344476 | 1.322597 | 2.5098268 | DTYMK;DUT;GMPR2;TXNRD1;GSR;ADK;NME2;GMPS;AK2;CTPS2;GMPR;AMPD2;NUDT5;NT5C2;AK9;NT5C3A;NUDT9;RRM2B;CAT;CMPK1;TK2;TK1;NUDT15;ADSS |
| 619 | Ion homeostasis Homo sapiens R-HSA-5578775 | 0.149951 | 0.344476 | 1.446194 | 2.7440725 | AHCYL1;ATP2A3;ATP2A2;ITPR2;ATP1B3;ITPR3;ATP1A1;SRI;ATP1B1;STIM1;ASPH;FXYD4;FXYD3;ORAI1;CALM1 |
| 620 | Ion transport by P-type ATPases Homo sapiens R-HSA-936837 | 0.150943 | 0.346194 | 1.423988 | 2.6925556 | ATP13A1;ATP8B1;ATP2A3;ATP2A2;ATP11C;ATP1B3;ATP1A1;SRI;ATP1B1;ATP2C1;FXYD4;FXYD3;ATP13A2;ATP7A;CALM1;ATP9A |
| 621 | DNA Double Strand Break Response Homo sapiens R-HSA-5693606 | 0.151575 | 0.347084 | 1.404959 | 2.6507037 | KDM4A;KDM4B;UBE2I;EYA4;RNF8;SMARCA5;BAZ1B;BABAM1;RAD50;HERC2;UBB;UIMC1;UBC;ATM;NBN;TP53BP1;KPNA2 |

| | | | | | | |
|---|---|---|---|---|---|---|
| 622 | Hemostasis Homo sapiens R-HSA-109582 | 0.151883 | 0.34723 | 1.11496 | 2.1013058 | APP;KDM1A;ITGA2B;JMJD1C;F11R;ABHD12;GRB14;AKT2;KIF5B;LAMP2;CFL1;AKT1;EP300;KIF21A;FAM3C;KIFAP3;CXADR;APLP2;PRKCD;ATP1B3;ACTN4;ATP1B1;TUBA4A;TMX3;STIM1;KIF16B;ORAI1;TLN1;RAF1;PPIA;VCL;DGKH;GRB7;STXBP3;AK3;ITPR2;PIK3R3;ITPR3;A1BG;GATA3;GATA2;RAP1B;CYB5R1;KLC2;KIF3A;PRKAR2A;PSAP;KCNMB4;PLCG1;S100A10;YES1;TGFB1;CBX5;HSPA5;RARRES2;H3F3A;GNG11;PTK2;EHD1;CDK5;MAFG;CALU;GRB2;CALM1;RCOR1;ITGB1;DOCK6;LGALS3BP;SERPINA1;WDR1;DOCK7;ATP2A3;ATP2A2;ARRB1;KIF12;SERPINA4;SERPINA5;KIF13B;PHACTR2;HMG20B;QSOX1;KIF1B;CD36;RAC1;JAK2;JAK1;VAV3;ABCC4;ANXA2;ITGA3;ITPK1;KIF25;PPP2R5B;KIF23;RHOA;SERPINB6;KIF2A;IRF3;IL3RA;IRF2;IRF7;ITGA6;CD47;SOS1;ALDOA;RAPGEF3;MGLL;CD63;HDAC2;SRC;HDAC1;GNAI3;SRI;AKAP1;GNAI2;CDC42;PDE11A;GNG5;MFN1;CDC37L1;MAPK1;STX4;CSK;CD58;MAPK3;CD74;PPIL2;PCDH7;VEGFB;PTPN11;P2RX4;CAPZA2;CD9;BCAR1 |
| 623 | Fcgamma receptor (FCGR) dependent phagocytosis Homo sapiens R-HSA-2029480 | 0.154164 | 0.351881 | 1.262706 | 2.3609258 | CYFIP1;NCKAP1;AHCYL1;HSP90AB1;SRC;ARPC1B;ARPC1A;ITPR2;ITPR3;WASL;PLD3;ACTG1;CDC42;CFL1;MAPK1;PLCG1;RAC1;MAPK3;ACTR3;VAV3;ACTR2;HSP90AA1;YES1;MYO10;PRKCD;ARPC4;PTK2;ARPC2;ABI2;ARPC3;MYH9;GRB2 |
| 624 | PERK regulates gene expression Homo sapiens R-HSA-381042 | 0.155195 | 0.353099 | 1.643688 | 3.0623155 | EXOSC7;HSPA5;NFYC;EXOSC8;PARN;EIF2S1;ATF6;DCP2;ATF4 |
| 625 | Recognition of DNA damage by PCNA-containing replication complex Homo sapiens R-HSA-110314 | 0.155195 | 0.353099 | 1.643688 | 3.0623155 | DDB1;POLD4;UBB;RFC1;RPA3;UBC;USP1;RPA2;CUL4B |
| 626 | Nuclear import of Rev protein Homo sapiens R-HSA-180746 | 0.159962 | 0.359845 | 1.577325 | 2.8909497 | RANBP2;NUP214;NUP205;NPM1;POM121C;NUP50;TPR;NUP98;AAAS;RAN |
| 627 | Vpr-mediated nuclear import of PICs Homo sapiens R-HSA-180910 | 0.159962 | 0.359845 | 1.577325 | 2.8909497 | RANBP2;NUP214;NUP205;POM121C;NUP50;TPR;PSIP1;NUP98;AAAS;KPNA1 |
| 628 | Negative regulation of FGFR1 signaling Homo sapiens R-HSA-5654726 | 0.159962 | 0.359845 | 1.577325 | 2.8909497 | UBB;SRC;MKNK1;UBC;MAPK1;PTPN11;BRAF;GRB2;CBL;MAPK3 |

| | | | | | | |
|---|---|---|---|---|---|---|
| 629 | Signaling by Robo receptor Homo sapiens R-HSA-376176 | 0.159962 | 0.359845 | 1.577325 | 2.8909497 | CDC42;VASP;ENAH;EVL;RAC1;SRGAP3;SOS1;PAK2;SRGAP1;PFN2 |
| 630 | DAG and IP3 signaling Homo sapiens R-HSA-1489509 | 0.159962 | 0.359845 | 1.577325 | 2.8909497 | AHCYL1;CREB1;ADCY9;PRKAR2A;PRKCD;ITPR2;ITPR3;PLCG1;CALM1;ADCY6 |
| 631 | Zinc transporters Homo sapiens R-HSA-435354 | 0.16069 | 0.359845 | 1.892438 | 3.459899 | SLC30A6;SLC30A5;SLC39A7;SLC39A1;SLC39A4;SLC39A3 |
| 632 | Hyaluronan metabolism Homo sapiens R-HSA-2142845 | 0.16069 | 0.359845 | 1.892438 | 3.459899 | ABCC5;HEXB;HYAL2;HEXA;GUSB;SLC9A1 |
| 633 | Tristetraprolin (TTP, ZFP36) binds and destabilizes mRNA Homo sapiens R-HSA-450513 | 0.16069 | 0.359845 | 1.892438 | 3.459899 | EXOSC7;XRN1;YWHAB;EXOSC8;TNPO1;DCP2 |
| 634 | Butyrate Response Factor 1 (BRF1) binds and destabilizes mRNA Homo sapiens R-HSA-450385 | 0.16069 | 0.359845 | 1.892438 | 3.459899 | EXOSC7;XRN1;YWHAB;EXOSC8;AKT1;DCP2 |
| 635 | Translesion synthesis by POLK Homo sapiens R-HSA-5655862 | 0.16069 | 0.359845 | 1.892438 | 3.459899 | RFC1;UBB;RPA3;UBC;REV1;RPA2 |
| 636 | Transcriptional activation of mitochondrial biogenesis Homo sapiens R-HSA-2151201 | 0.165896 | 0.369865 | 1.487282 | 2.6717443 | ESRRA;NCOA1;ATF2;NCOA2;CREB1;NCOR1;CHD9;TBL1XR1;TGS1;TFAM;NR1D1;TFB1M |
| 637 | Zinc influx into cells by the SLC39 gene family Homo sapiens R-HSA-442380 | 0.166465 | 0.369865 | 2.312691 | 4.1465796 | SLC39A7;SLC39A1;SLC39A4;SLC39A3 |
| 638 | Reactions specific to the complex N-glycan synthesis pathway Homo sapiens R-HSA-975578 | 0.166465 | 0.369865 | 2.312691 | 4.1465796 | FUT8;MAN2A2;MAN2A1;FUCA1 |

| | | | | | | |
|---|---|---|---|---|---|---|
| 639 | Activation of the AP-1 family of transcription factors Homo sapiens R-HSA-450341 | 0.166465 | 0.369865 | 2.312691 | 4.1465796 | MAPK10;ATF2;MAPK1;MAPK3 |
| 640 | Beta-oxidation of pristanoyl-CoA Homo sapiens R-HSA-389887 | 0.166465 | 0.369865 | 2.312691 | 4.1465796 | SCP2;HSD17B4;ACOX3;CRAT |
| 641 | SUMOylation of DNA replication proteins Homo sapiens R-HSA-4615885 | 0.167614 | 0.370104 | 1.455342 | 2.5993753 | RANBP2;NUP214;TOP2B;NUP205;UBE2I;AAAS;POM121C;SUMO3;NUP50;TPR;SUMO2;NUP98;TOP1 |
| 642 | Formation of HIV elongation complex in the absence of HIV Tat Homo sapiens R-HSA-167152 | 0.167614 | 0.370104 | 1.455342 | 2.5993753 | SUPT16H;CCNT2;SSRP1;GTF2F2;CDK9;CDK7;POLR2B;POLR2C;POLR2E;TCEA1;NELFB;POLR2H;POLR2J |
| 643 | RNA Polymerase II Transcription Elongation Homo sapiens R-HSA-75955 | 0.167614 | 0.370104 | 1.455342 | 2.5993753 | SUPT16H;CCNT2;SSRP1;GTF2F2;CDK9;CDK7;POLR2B;POLR2C;POLR2E;TCEA1;NELFB;POLR2H;POLR2J |
| 644 | Formation of RNA Pol II elongation complex Homo sapiens R-HSA-112382 | 0.167614 | 0.370104 | 1.455342 | 2.5993753 | SUPT16H;CCNT2;SSRP1;GTF2F2;CDK9;CDK7;POLR2B;POLR2C;POLR2E;TCEA1;NELFB;POLR2H;POLR2J |
| 645 | TP53 Regulates Transcription of Cell Cycle Genes Homo sapiens R-HSA-6791312 | 0.16874 | 0.372012 | 1.429044 | 2.5428369 | BTG2;NPM1;CDKN1B;PRMT1;CNOT4;CCNA1;RGCC;CNOT1;CNOT2;CENPJ;EP300;TNKS1BP1;BAX;CNOT8 |
| 646 | HS-GAG degradation Homo sapiens R-HSA-2024096 | 0.171013 | 0.375859 | 1.734787 | 3.0636618 | SDC4;NAGLU;SDC2;SDC1;GUSB;AGRN;HSPG2 |
| 647 | Mitochondrial tRNA aminoacylation Homo sapiens R-HSA-379726 | 0.171013 | 0.375859 | 1.734787 | 3.0636618 | PPA2;KARS;QARS;NARS2;IARS2;GARS;CARS2 |
| 648 | Activation of BH3-only proteins Homo sapiens R-HSA-114452 | 0.182802 | 0.401149 | 1.561403 | 2.653371 | YWHAE;PPP1R13B;YWHAB;BAD;AKT2;TP53BP2;AKT1;BID;YWHAH |

| | | | | | | |
|---|---|---|---|---|---|---|
| 649 | Rev-mediated nuclear export of HIV RNA Homo sapiens R-HSA-165054 | 0.186039 | 0.402937 | 1.508648 | 2.5372428 | RANBP2;NUP214;NUP205;XPO1;POM121C;NUP50;TPR;NUP98;AAAS;RAN |
| 650 | Early Phase of HIV Life Cycle Homo sapiens R-HSA-162594 | 0.186167 | 0.402937 | 1.927301 | 3.2400049 | XRCC6;XRCC5;PSIP1;PPIA;KPNA1 |
| 651 | Regulation of TP53 Activity through Association with Co-factors Homo sapiens R-HSA-6804759 | 0.186167 | 0.402937 | 1.927301 | 3.2400049 | PPP1R13B;PHF20;AKT2;TP53BP2;AKT1 |
| 652 | Mismatch repair (MMR) directed by MSH2:MSH3 (MutSbeta) Homo sapiens R-HSA-5358606 | 0.186167 | 0.402937 | 1.927301 | 3.2400049 | POLD4;MSH3;RPA3;RPA2;MLH1 |
| 653 | Mismatch repair (MMR) directed by MSH2:MSH6 (MutSalpha) Homo sapiens R-HSA-5358565 | 0.186167 | 0.402937 | 1.927301 | 3.2400049 | MSH6;POLD4;RPA3;RPA2;MLH1 |
| 654 | Regulation of necroptotic cell death Homo sapiens R-HSA-5675482 | 0.186167 | 0.402937 | 1.927301 | 3.2400049 | XIAP;FAS;RIPK1;CFLAR;BIRC2 |
| 655 | DCC mediated attractive signaling Homo sapiens R-HSA-418885 | 0.186167 | 0.402937 | 1.927301 | 3.2400049 | CDC42;SRC;WASL;RAC1;PTK2 |
| 656 | Repression of WNT target genes Homo sapiens R-HSA-4641265 | 0.186167 | 0.402937 | 1.927301 | 3.2400049 | TLE3;TLE2;CTBP2;HDAC1;CTNNB1 |
| 657 | Synthesis of Prostaglandins (PG) and Thromboxanes (TX) Homo sapiens R-HSA-2162123 | 0.186167 | 0.402937 | 1.927301 | 3.2400049 | CBR1;AKR1C3;PTGR1;PTGS1;PTGES |

| | | | | | | |
|---|---|---|---|---|---|---|
| 658 | Translesion synthesis by Y family DNA polymerases bypasses lesions on DNA template Homo sapiens R-HSA-110313 | 0.188156 | 0.404909 | 1.468075 | 2.4523971 | POLD4;VCP;UBB;RFC1;USP43;RPA3;UBC;REV1;POLI;RPA2;ISG15 |
| 659 | Platelet Aggregation (Plug Formation) Homo sapiens R-HSA-76009 | 0.188156 | 0.404909 | 1.468075 | 2.4523971 | RAP1B;SRC;ITGA2B;AKT1;CSK;GRB2;TLN1;SOS1;PTK2;RAPGEF3;BCAR1 |
| 660 | Histidine, lysine, phenylalanine, tyrosine, proline and tryptophan catabolism Homo sapiens R-HSA-6788656 | 0.189441 | 0.404909 | 1.435904 | 2.3888788 | ALDH4A1;HGD;PHYKPL;KYNU;AFMID;OGDH;DLST;PCBD1;CRYM;ALDH7A1;DLD;IDO1 |
| 661 | SUMOylation of RNA binding proteins Homo sapiens R-HSA-4570464 | 0.190096 | 0.404909 | 1.409771 | 2.340536 | RANBP2;NUP214;NUP205;PHC2;UBE2I;RING1;AAAS;HNRNPK;POM121C;NUP50;TPR;SUMO2;NUP98 |
| 662 | RMTs methylate histone arginines Homo sapiens R-HSA-3214858 | 0.190096 | 0.404909 | 1.409771 | 2.340536 | SMARCD1;SMARCB1;PRMT7;SMARCC2;H2AFZ;PRMT1;ARID1B;SMARCA2;COPRS;RBBP7;ARID2;JAK2;HIST1H2AC |
| 663 | Diseases of metabolism Homo sapiens R-HSA-5668914 | 0.190096 | 0.404909 | 1.409771 | 2.340536 | AMN;MCCC1;CCDC59;MTR;ACACB;ACACA;UBB;PCCA;PPP1R3C;UBC;PCCB;FDX1;CD320 |
| 664 | Mitochondrial biogenesis Homo sapiens R-HSA-1592230 | 0.190265 | 0.404909 | 1.388125 | 2.3033644 | NCOA1;ESRRA;ATF2;NCOA2;PRKAA2;CHD9;PRKAG2;NR1D1;TFB1M;NCOR1;CREB1;TBL1XR1;TGS1;TFAM |
| 665 | Attachment of GPI anchor to uPAR Homo sapiens R-HSA-162791 | 0.19135 | 0.404909 | 2.601531 | 4.3020311 | PIGS;PGAP1;PIGK |
| 666 | 2-LTR circle formation Homo sapiens R-HSA-164843 | 0.19135 | 0.404909 | 2.601531 | 4.3020311 | XRCC6;XRCC5;PSIP1 |
| 667 | NrCAM interactions Homo sapiens R-HSA-447038 | 0.19135 | 0.404909 | 2.601531 | 4.3020311 | DLG1;DLG3;NRCAM |
| 668 | PTK6 promotes HIF1A stabilization Homo sapiens R-HSA-8857538 | 0.19135 | 0.404909 | 2.601531 | 4.3020311 | LRRK2;HIF1A;EGFR |

| | | | | | | |
|---|---|---|---|---|---|---|
| 669 | IkBA variant leads to EDA-ID Homo sapiens R-HSA-5603029 | 0.19135 | 0.404909 | 2.601531 | 4.3020311 | NFKBIA;RELA;NFKB1 |
| 670 | Interleukin-1 processing Homo sapiens R-HSA-448706 | 0.19135 | 0.404909 | 2.601531 | 4.3020311 | IL18;RELA;NFKB1 |
| 671 | Acyl chain remodeling of DAG and TAG Homo sapiens R-HSA-1482883 | 0.19135 | 0.404909 | 2.601531 | 4.3020311 | DGAT1;MGLL;PNPLA2 |
| 672 | Phosphate bond hydrolysis by NUDT proteins Homo sapiens R-HSA-2393930 | 0.19135 | 0.404909 | 2.601531 | 4.3020311 | NUDT9;NUDT15;NUDT5 |
| 673 | Miscellaneous transport and binding events Homo sapiens R-HSA-5223345 | 0.1979 | 0.417529 | 1.734623 | 2.8100741 | DMTN;NIPAL1;NIPAL3;CTNS;NIPA2;ADD1 |
| 674 | RNA Polymerase III Chain Elongation Homo sapiens R-HSA-73780 | 0.1979 | 0.417529 | 1.734623 | 2.8100741 | CRCP;POLR3B;POLR3GL;POLR1D;POLR2E;POLR2H |
| 675 | HATs acetylate histones Homo sapiens R-HSA-3214847 | 0.202703 | 0.427028 | 1.230349 | 1.9636512 | ATF2;MEAF6;TRRAP;PHF20;YEATS4;ING4;RUVBL1;EPC1;KAT8;EP300;DMAP1;RBBP7;ATXN7L3;HIST1H2AC;NCOA1;NCOA2;MCRS1;MSL3;ELP2;ELP3;ELP5;ELP6;MSL1;MORF4L1;KAT2A;MRGBP;TADA3;OGT |
| 676 | mTORC1-mediated signalling Homo sapiens R-HSA-166208 | 0.205167 | 0.430941 | 1.619031 | 2.5644356 | RRAGA;YWHAB;MLST8;LAMTOR1;MTOR;EIF4B;EIF4G1 |
| 677 | SALM protein interactions at the synapse Homo sapiens R-HSA-8849932 | 0.205167 | 0.430941 | 1.619031 | 2.5644356 | RTN3;DLG1;LRFN4;DLG3;FLOT1;FLOT2;PTPRF |
| 678 | Triglyceride Biosynthesis Homo sapiens R-HSA-75109 | 0.207265 | 0.434706 | 1.301441 | 2.0481539 | AGPAT5;ACOT9;ELOVL1;DGAT1;AGK;TECR;ACOT12;ACSL5;ACOT13;ACSL3;HSD17B12;AGPAT3;ACACA;ACOT2;THEM4;LPIN2;ACOT4;LPIN3 |

| | | | | | | |
|---|---|---|---|---|---|---|
| 679 | A tetrasaccharide linker sequence is required for GAG synthesis Homo sapiens R-HSA-1971475 | 0.209684 | 0.4372 | 1.541981 | 2.4088122 | SDC4;B3GAT3;SDC2;GXYLT1;SDC1;AGRN;HSPG2;B4GALT7 |
| 680 | PD-1 signaling Homo sapiens R-HSA-389948 | 0.209684 | 0.4372 | 1.541981 | 2.4088122 | HLA-DPB1;HLA-DRA;CSK;PTPN11;HLA-DQA2;HLA-DRB1;HLA-DPA1;HLA-DQB1 |
| 681 | Regulation of Hypoxia-inducible Factor (HIF) by oxygen Homo sapiens R-HSA-1234174 | 0.209684 | 0.4372 | 1.541981 | 2.4088122 | EGLN3;EGLN2;UBB;UBE2D2;UBC;EP300;HIF3A;HIF1A |
| 682 | Cellular response to hypoxia Homo sapiens R-HSA-2262749 | 0.209684 | 0.4372 | 1.541981 | 2.4088122 | EGLN3;EGLN2;UBB;UBE2D2;UBC;EP300;HIF3A;HIF1A |
| 683 | B-WICH complex positively regulates rRNA expression Homo sapiens R-HSA-5250924 | 0.210473 | 0.437561 | 1.322018 | 2.0602336 | GSK3B;H2AFZ;H3F3A;SMARCA5;DDX21;DEK;BAZ1B;KAT2A;TAF1D;TAF1B;POLR1D;POLR2E;EP300;POLR2H;SF3B1;HIST1H2AC |
| 684 | Recruitment and ATM-mediated phosphorylation of repair and signaling proteins at DNA double strand breaks Homo sapiens R-HSA-5693565 | 0.210473 | 0.437561 | 1.322018 | 2.0602336 | KDM4A;KDM4B;UBE2I;EYA4;RNF8;SMARCA5;BAZ1B;BABAM1;RAD50;HERC2;UBB;UIMC1;UBC;ATM;NBN;TP53BP1 |
| 685 | Gap junction trafficking and regulation Homo sapiens R-HSA-157858 | 0.212398 | 0.440078 | 1.486955 | 2.3037316 | TJP1;DAB2;GJB1;SRC;CLTC;MYO6;CLTB;AP2M1;DNM2 |
| 686 | Transport of Ribonucleoproteins into the Host Nucleus Homo sapiens R-HSA-168271 | 0.212398 | 0.440078 | 1.486955 | 2.3037316 | RANBP2;NUP214;NUP205;POM121C;NUP50;TPR;NUP98;AAAS;KPNA1 |
| 687 | Negative regulation of FGFR2 signaling Homo sapiens R-HSA-5654727 | 0.213873 | 0.440078 | 1.445694 | 2.2297992 | UBB;SRC;MKNK1;UBC;MAPK1;PTPN11;BRAF;GRB2;CBL;MAPK3 |

| | | | | | | |
|---|---|---|---|---|---|---|
| 688 | PLC-gamma1 signalling Homo sapiens R-HSA-167021 | 0.213873 | 0.440078 | 1.445694 | 2.2297992 | AHCYL1;CREB1;ADCY9;PRKAR2A;PRKCD;ITPR2;ITPR3;PLCG1;CALM1;ADCY6 |
| 689 | ROS, RNS production in response to bacteria Homo sapiens R-HSA-1222556 | 0.213873 | 0.440078 | 1.445694 | 2.2297992 | ATP6V1A;ATP6V1G1;ATP6V0B;ATP6V0E1;ATP6V0A4;TCIRG1;ATP6V0D1;ATP6V0C;ATP6V1C1;ATP6V1C2 |
| 690 | Glucose transport Homo sapiens R-HSA-70153 | 0.214421 | 0.440078 | 1.387951 | 2.1371847 | RANBP2;NUP214;NUP205;G6PC3;POM121C;NUP50;TPR;SLC2A1;NUP98;AAAS;SLC37A4;HK1 |
| 691 | Transcriptional regulation of pluripotent stem cells Homo sapiens R-HSA-452723 | 0.214421 | 0.440078 | 1.387951 | 2.1371847 | SMAD2;SMAD4;TSC22D1;POLR2B;POLR2C;STAT3;POLR2E;HIF3A;POLR2H;EPHA1;POLR2J;POU5F1 |
| 692 | Activation of gene expression by SREBF (SREBP) Homo sapiens R-HSA-2426168 | 0.214421 | 0.440078 | 1.387951 | 2.1371847 | NCOA1;NCOA2;CHD9;TBL1XR1;TGS1;NFYC;CYP51A1;HMGCR;ACACB;ACACA;SREBF2;FDFT1 |
| 693 | Signaling by FGFR1 in disease Homo sapiens R-HSA-5655302 | 0.214468 | 0.440078 | 1.413611 | 2.176386 | BCR;ZMYM2;CUX1;STAT1;FGFR1OP2;CNTRL;STAT3;GRB2;LRRFIP1;PLCG1;SOS1 |
| 694 | Signaling by NOTCH3 Homo sapiens R-HSA-1980148 | 0.217593 | 0.443291 | 1.982179 | 3.02308 | APH1A;PSENEN;APH1B;ADAM10 |
| 695 | Signaling by NOTCH4 Homo sapiens R-HSA-1980150 | 0.217593 | 0.443291 | 1.982179 | 3.02308 | APH1A;PSENEN;APH1B;ADAM10 |
| 696 | TRAF6 mediated IRF7 activation in TLR7/8 or 9 signaling Homo sapiens R-HSA-975110 | 0.217593 | 0.443291 | 1.982179 | 3.02308 | DHX36;IRF7;UBE2V1;IRAK4 |
| 697 | CLEC7A (Dectin-1) induces NFAT activation Homo sapiens R-HSA-5607763 | 0.217593 | 0.443291 | 1.982179 | 3.02308 | AHCYL1;ITPR2;ITPR3;CALM1 |
| 698 | Synthesis of IP2, IP, and Ins in the cytosol Homo sapiens R-HSA-1855183 | 0.217593 | 0.443291 | 1.982179 | 3.02308 | ISYNA1;INPP1;INPP5J;OCRL |

| | | | | | | |
|---|---|---|---|---|---|---|
| 699 | Signaling by FGFR in disease Homo sapiens R-HSA-1226099 | 0.230006 | 0.462265 | 1.282458 | 1.8847635 | STAT1;STAT3;GTF2F2;BCR;ZMYM2;CUX1;POLR2B;POLR2C;CNTRL;FGFR1OP2;POLR2E;GRB2;LRRFIP1;PLCG1;POLR2H;SOS1;POLR2J |
| 700 | Depolymerisation of the Nuclear Lamina Homo sapiens R-HSA-4419969 | 0.230807 | 0.462265 | 1.734459 | 2.5430144 | LMNA;LEMD2;LPIN2;LPIN3;TMPO |
| 701 | WNT5A-dependent internalization of FZD4 Homo sapiens R-HSA-5099900 | 0.230807 | 0.462265 | 1.734459 | 2.5430144 | WNT5A;CLTC;CLTB;AP2A1;AP2M1 |
| 702 | Glycogen breakdown (glycogenolysis) Homo sapiens R-HSA-70221 | 0.230807 | 0.462265 | 1.734459 | 2.5430144 | AGL;GAA;PHKG2;PHKB;CALM1 |
| 703 | S33 mutants of beta-catenin aren't phosphorylated Homo sapiens R-HSA-5358747 | 0.230807 | 0.462265 | 1.734459 | 2.5430144 | GSK3B;APC;CSNK1A1;PPP2R5B;CTNNB1 |
| 704 | S45 mutants of beta-catenin aren't phosphorylated Homo sapiens R-HSA-5358751 | 0.230807 | 0.462265 | 1.734459 | 2.5430144 | GSK3B;APC;CSNK1A1;PPP2R5B;CTNNB1 |
| 705 | phosphorylation site mutants of CTNNB1 are not targeted to the proteasome by the destruction complex Homo sapiens R-HSA-4839743 | 0.230807 | 0.462265 | 1.734459 | 2.5430144 | GSK3B;APC;CSNK1A1;PPP2R5B;CTNNB1 |
| 706 | T41 mutants of beta-catenin aren't phosphorylated Homo sapiens R-HSA-5358752 | 0.230807 | 0.462265 | 1.734459 | 2.5430144 | GSK3B;APC;CSNK1A1;PPP2R5B;CTNNB1 |
| 707 | S37 mutants of beta-catenin aren't phosphorylated Homo sapiens R-HSA-5358749 | 0.230807 | 0.462265 | 1.734459 | 2.5430144 | GSK3B;APC;CSNK1A1;PPP2R5B;CTNNB1 |
| 708 | Misspliced GSK3beta mutants stabilize beta-catenin Homo sapiens R-HSA-5339716 | 0.230807 | 0.462265 | 1.734459 | 2.5430144 | GSK3B;APC;CSNK1A1;PPP2R5B;CTNNB1 |

| | | | | | | |
|---|---|---|---|---|---|---|
| 709 | ERBB2 Regulates Cell Motility Homo sapiens R-HSA-6785631 | 0.230807 | 0.462265 | 1.734459 | 2.5430144 | DIAPH1;ERBB3;ERBB2;RHOA;EGFR |
| 710 | Elevation of cytosolic Ca2+ levels Homo sapiens R-HSA-139853 | 0.230807 | 0.462265 | 1.734459 | 2.5430144 | P2RX4;STIM1;ITPR2;ORAI1;ITPR3 |
| 711 | Nuclear Envelope Breakdown Homo sapiens R-HSA-2980766 | 0.23687 | 0.47374 | 1.312921 | 1.8909268 | NUP214;RANBP2;NUP205;NEK7;AAAS;LEMD2;POM121C;NUP50;LMNA;TPR;NUP98;LPIN2;LPIN3;TMPO |
| 712 | Signaling by FGFR2 IIIa TM Homo sapiens R-HSA-8851708 | 0.237894 | 0.473789 | 1.601087 | 2.2990491 | POLR2B;POLR2C;POLR2E;POLR2H;GTF2F2;POLR2J |
| 713 | Regulation of TP53 Activity through Methylation Homo sapiens R-HSA-6804760 | 0.237894 | 0.473789 | 1.601087 | 2.2990491 | UBB;UBC;SMYD2;EHMT1;EP300;ATM |
| 714 | Translesion Synthesis by POLH Homo sapiens R-HSA-110320 | 0.237894 | 0.473789 | 1.601087 | 2.2990491 | VCP;RFC1;UBB;RPA3;UBC;RPA2 |
| 715 | RNA Polymerase I Transcription Initiation Homo sapiens R-HSA-73762 | 0.238892 | 0.475112 | 1.326672 | 1.8994527 | HDAC2;HDAC1;CHD4;CHD3;TAF1D;KAT2A;CDK7;RBBP4;TAF1B;POLR2E;POLR1D;RBBP7;POLR2H |
| 716 | HIV Transcription Elongation Homo sapiens R-HSA-167169 | 0.240693 | 0.475416 | 1.343091 | 1.912873 | CDK9;SUPT16H;CDK7;POLR2B;POLR2C;POLR2E;TCEA1;NELFB;POLR2H;SSRP1;POLR2J;GTF2F2 |
| 717 | Tat-mediated elongation of the HIV-1 transcript Homo sapiens R-HSA-167246 | 0.240693 | 0.475416 | 1.343091 | 1.912873 | CDK9;SUPT16H;CDK7;POLR2B;POLR2C;POLR2E;TCEA1;NELFB;POLR2H;SSRP1;POLR2J;GTF2F2 |
| 718 | Formation of HIV-1 elongation complex containing HIV-1 Tat Homo sapiens R-HSA-167200 | 0.240693 | 0.475416 | 1.343091 | 1.912873 | CDK9;SUPT16H;CDK7;POLR2B;POLR2C;POLR2E;TCEA1;NELFB;POLR2H;SSRP1;POLR2J;GTF2F2 |
| 719 | Abortive elongation of HIV-1 transcript in the absence of Tat Homo sapiens R-HSA-167242 | 0.24162 | 0.475416 | 1.517743 | 2.155783 | POLR2B;POLR2C;POLR2E;NELFB;POLR2H;GTF2F2;POLR2J |

| | | | | | | |
|---|---|---|---|---|---|---|
| 720 | Cholesterol biosynthesis Homo sapiens R-HSA-191273 | 0.24162 | 0.475416 | 1.517743 | 2.155783 | EBP;CYP51A1;MSMO1;HMGCR;HSD17B7;LBR;FDFT1 |
| 721 | Collagen degradation Homo sapiens R-HSA-1442490 | 0.242189 | 0.475416 | 1.363037 | 1.9328374 | ADAM17;MMP7;COL14A1;CTSL;PHYKPL;COL12A1;ADAM10;ADAM9;FURIN;CTSD;CTSB |
| 722 | Base Excision Repair Homo sapiens R-HSA-73884 | 0.243258 | 0.475416 | 1.387777 | 1.9618051 | POLD4;MBD4;RFC1;MPG;OGG1;RPA3;APEX1;RPA2;NEIL1;UNG |
| 723 | Resolution of Abasic Sites (AP sites) Homo sapiens R-HSA-73933 | 0.243258 | 0.475416 | 1.387777 | 1.9618051 | POLD4;MBD4;RFC1;MPG;OGG1;RPA3;APEX1;RPA2;NEIL1;UNG |
| 724 | RNA Pol II CTD phosphorylation and interaction with CE Homo sapiens R-HSA-167160 | 0.243322 | 0.475416 | 1.460729 | 2.064548 | CDK7;RNMT;POLR2B;POLR2C;POLR2E;POLR2H;POLR2J;GTF2F2 |
| 725 | RNA Pol II CTD phosphorylation and interaction with CE Homo sapiens R-HSA-77075 | 0.243322 | 0.475416 | 1.460729 | 2.064548 | CDK7;RNMT;POLR2B;POLR2C;POLR2E;POLR2H;POLR2J;GTF2F2 |
| 726 | CREB phosphorylation through the activation of Ras Homo sapiens R-HSA-442742 | 0.243322 | 0.475416 | 1.460729 | 2.064548 | CREB1;RRAS;RASGRF2;AKAP9;MAPK1;BRAF;RAF1;CALM1 |
| 727 | Disassembly of the destruction complex and recruitment of AXIN to the membrane Homo sapiens R-HSA-4641262 | 0.243726 | 0.475416 | 1.419274 | 2.0036067 | GSK3B;FZD5;APC;CSNK1A1;PPP2R5B;DVL3;CTNNB1;CSNK1G2;LRP6 |
| 728 | FGFR1 mutant receptor activation Homo sapiens R-HSA-1839124 | 0.243726 | 0.475416 | 1.419274 | 2.0036067 | BCR;ZMYM2;CUX1;STAT1;FGFR1OP2;CNTRL;STAT3;GRB2;LRRFIP1 |
| 729 | Regulation of TNFR1 signaling Homo sapiens R-HSA-5357905 | 0.243726 | 0.475416 | 1.419274 | 2.0036067 | SHARPIN;UBB;UBC;TAX1BP1;XIAP;RIPK1;RBCK1;BIRC2;TNFRSF1A |

| | | | | | | |
|---|---|---|---|---|---|---|
| 730 | RHO GTPases Activate Formins Homo sapiens R-HSA-5663220 | 0.247341 | 0.481807 | 1.184146 | 1.6542377 | ITGB1;AHCTF1;SRC;PMF1;ACTG1;SKA2;CENPC;CDC42;XPO1;DVL3;RAC1;CENPT;RANBP2;PPP2R5B;RHOD;RHOA;DIAPH1;CLIP1;DIAPH2;RPS27;FMNL2;DAAM1;KIF2A;EVL;NUP98;MAPRE1;PFN2;PAFAH1B1;MAD2L1 |
| 731 | WNT mediated activation of DVL Homo sapiens R-HSA-201688 | 0.257702 | 0.497897 | 2.081091 | 2.82186 | CSNK2A1;CSNK2A2;DVL3 |
| 732 | Synthesis of UDP-N-acetyl-glucosamine Homo sapiens R-HSA-446210 | 0.257702 | 0.497897 | 2.081091 | 2.82186 | NAGK;GFPT1;PGM3 |
| 733 | IKBKG deficiency causes anhidrotic ectodermal dysplasia with immunodeficiency (EDA-ID) (via TLR) Homo sapiens R-HSA-5603027 | 0.257702 | 0.497897 | 2.081091 | 2.82186 | NFKBIA;RELA;NFKB1 |
| 734 | ChREBP activates metabolic gene expression Homo sapiens R-HSA-163765 | 0.257702 | 0.497897 | 2.081091 | 2.82186 | MLX;ACACB;ACACA |
| 735 | Coenzyme A biosynthesis Homo sapiens R-HSA-196783 | 0.257702 | 0.497897 | 2.081091 | 2.82186 | PANK4;PANK2;COASY |
| 736 | NR1D1 (REV-ERBA) represses gene expression Homo sapiens R-HSA-1368071 | 0.257702 | 0.497897 | 2.081091 | 2.82186 | NCOR1;NR1D1;NPAS2 |
| 737 | Nonhomologous End-Joining (NHEJ) Homo sapiens R-HSA-5693571 | 0.261729 | 0.50499 | 1.278288 | 1.7134781 | XRCC6;RIF1;PRKDC;XRCC5;POLM;RNF8;BABAM1;RAD50;HERC2;UIMC1;POLL;ATM;NBN;TP53BP1 |
| 738 | Hexose transport Homo sapiens R-HSA-189200 | 0.264949 | 0.510511 | 1.288684 | 1.7116534 | RANBP2;NUP214;NUP205;SLC2A1;AAAS;SLC5A3;HK1;G6PC3;POM121C;NUP50;TPR;NUP98;SLC37A4 |
| 739 | Kinesins Homo sapiens R-HSA-983189 | 0.268098 | 0.515183 | 1.301036 | 1.7126859 | KLC2;KIF2A;KIF3A;KIF16B;KIF5B;KIF13B;KIF25;KIF12;KIF21A;KIF1B;KIF23;KIFAP3 |
| 740 | PLC beta mediated events Homo sapiens R-HSA-112043 | 0.268098 | 0.515183 | 1.301036 | 1.7126859 | AHCYL1;CREB1;ADCY9;PRKAR2A;PRKCD;GNAI3;ITPR2;MAPK1;ITPR3;CALM1;ADCY6;GNAI2 |

| | | | | | | |
|---|---|---|---|---|---|---|
| 741 | Negative regulation of MAPK pathway Homo sapiens R-HSA-5675221 | 0.271127 | 0.518091 | 1.31595 | 1.7175344 | MAP2K1;MAP2K2;UBB;YWHAB;UBC;PPP2R5B;PEBP1;MAPK1;BRAF;RAF1;MAPK3 |
| 742 | Erythrocytes take up carbon dioxide and release oxygen Homo sapiens R-HSA-1237044 | 0.272162 | 0.518091 | 1.734295 | 2.2569402 | CYB5R1;CYB5R4;CA2;SLC4A1 |
| 743 | O2/CO2 exchange in erythrocytes Homo sapiens R-HSA-1480926 | 0.272162 | 0.518091 | 1.734295 | 2.2569402 | CYB5R1;CYB5R4;CA2;SLC4A1 |
| 744 | CD28 dependent Vav1 pathway Homo sapiens R-HSA-389359 | 0.272162 | 0.518091 | 1.734295 | 2.2569402 | CDC42;GRB2;RAC1;PAK2 |
| 745 | PECAM1 interactions Homo sapiens R-HSA-210990 | 0.272162 | 0.518091 | 1.734295 | 2.2569402 | YES1;SRC;PTPN11;PLCG1 |
| 746 | Reduction of cytosolic Ca++ levels Homo sapiens R-HSA-418359 | 0.272162 | 0.518091 | 1.734295 | 2.2569402 | ATP2A3;ATP2A2;SRI;CALM1 |
| 747 | Vitamin B5 (pantothenate) metabolism Homo sapiens R-HSA-199220 | 0.272162 | 0.518091 | 1.734295 | 2.2569402 | PANK4;PANK2;AASDHPPT;COASY |
| 748 | Regulated Necrosis Homo sapiens R-HSA-5218859 | 0.278059 | 0.524634 | 1.576679 | 2.0180261 | XIAP;FAS;RIPK1;CFLAR;BIRC2 |
| 749 | RIPK1-mediated regulated necrosis Homo sapiens R-HSA-5213460 | 0.278059 | 0.524634 | 1.576679 | 2.0180261 | XIAP;FAS;RIPK1;CFLAR;BIRC2 |
| 750 | Cell-extracellular matrix interactions Homo sapiens R-HSA-446353 | 0.278059 | 0.524634 | 1.576679 | 2.0180261 | ITGB1;VASP;TESK1;ILK;LIMS1 |
| 751 | Rap1 signalling Homo sapiens R-HSA-392517 | 0.278059 | 0.524634 | 1.576679 | 2.0180261 | RAP1B;YWHAB;RAF1;RAPGEF3;SIPA1 |
| 752 | Sema3A PAK dependent Axon repulsion Homo sapiens R-HSA-399954 | 0.278059 | 0.524634 | 1.576679 | 2.0180261 | HSP90AA1;HSP90AB1;CFL1;RAC1;PAK2 |

| | | | | | | |
|---|---|---|---|---|---|---|
| 753 | Growth hormone receptor signaling Homo sapiens R-HSA-982772 | 0.279905 | 0.524634 | 1.428372 | 1.8187551 | ADAM17;STAT1;STAT3;MAPK1;IRS2;JAK2;MAPK3 |
| 754 | Diseases associated with the TLR signaling cascade Homo sapiens R-HSA-5602358 | 0.279905 | 0.524634 | 1.428372 | 1.8187551 | NFKBIA;IRAK4;CD36;TICAM1;TLR5;RELA;NFKB1 |
| 755 | Diseases of Immune System Homo sapiens R-HSA-5260271 | 0.279905 | 0.524634 | 1.428372 | 1.8187551 | NFKBIA;IRAK4;CD36;TICAM1;TLR5;RELA;NFKB1 |
| 756 | TRAF6 mediated NF-kB activation Homo sapiens R-HSA-933542 | 0.279905 | 0.524634 | 1.428372 | 1.8187551 | NFKBIA;APP;MAP3K1;DHX9;HMGB1;RELA;NFKB1 |
| 757 | IKK complex recruitment mediated by RIP1 Homo sapiens R-HSA-937041 | 0.279905 | 0.524634 | 1.428372 | 1.8187551 | UBB;UBE2D2;UBC;RIPK1;UBE2V1;TICAM1;BIRC2 |
| 758 | IL-6-type cytokine receptor ligand interactions Homo sapiens R-HSA-6788467 | 0.280026 | 0.524634 | 1.486628 | 1.8922888 | IL11RA;LIFR;IL6ST;JAK2;IL27RA;JAK1 |
| 759 | Synthesis, secretion, and inactivation of Glucagon-like Peptide-1 (GLP-1) Homo sapiens R-HSA-381771 | 0.280026 | 0.524634 | 1.486628 | 1.8922888 | SPCS3;SPCS2;SPCS1;CTNNB1;SEC11A;SEC11C |
| 760 | Meiotic synapsis Homo sapiens R-HSA-1221632 | 0.283088 | 0.528977 | 1.239114 | 1.5637586 | SUN1;UBE2I;H2AFZ;HSPA2;SMC3;SMC1A;TERF1;SYNE2;STAG1;STAG2;RAD21;LMNA;TERF2IP;HIST1H2AC;ATR |
| 761 | Pre-NOTCH Expression and Processing Homo sapiens R-HSA-1912422 | 0.283088 | 0.528977 | 1.239114 | 1.5637586 | POGLUT1;B4GALT1;SEL1L;ATP2A3;ATP2A2;FURIN;RBPJ;KAT2A;SNW1;AGO3;TMED2;EP300;ST3GAL6;MAML3;TNRC6A |
| 762 | ABC-family proteins mediated transport Homo sapiens R-HSA-382556 | 0.296463 | 0.552516 | 1.261529 | 1.5338104 | ABCC3;ABCC4;ABCC1;ABCB10;ABCB7;PEX3;ABCC5;ABCA3;ABCC10;EIF2S2;EIF2S1;ABCF1 |
| 763 | G-protein mediated events Homo sapiens R-HSA-112040 | 0.296463 | 0.552516 | 1.261529 | 1.5338104 | AHCYL1;CREB1;ADCY9;PRKAR2A;PRKCD;GNAI3;ITPR2;MAPK1;ITPR3;CALM1;ADCY6;GNAI2 |

| | | | | | | |
|---|---|---|---|---|---|---|
| 764 | PI3K Cascade Homo sapiens R-HSA-109704 | 0.305192 | 0.56804 | 1.176135 | 1.3958547 | PRKAA1;PRKAA2;YWHAB;CAB39L;STRADA;STRADB;PRKAG2;IRS2;PTPN11;MTOR;PPM1A;RRAGA;AKT2;MLST8;AKT1;THEM4;GRB2;LAMTOR1;EIF4B;EIF4G1 |
| 765 | HDR through Single Strand Annealing (SSA) Homo sapiens R-HSA-5685938 | 0.305746 | 0.568328 | 1.284813 | 1.5225044 | RAD50;RPA3;TOP3A;RPA2;RAD17;ATM;NBN;TOPBP1;ATRIP;ATR |
| 766 | rRNA modification in the nucleus Homo sapiens R-HSA-6790901 | 0.308419 | 0.569092 | 1.21022 | 1.4235758 | RPS9;UTP6;RPS7;DIMT1;KRR1;NOP2;WDR75;THUMPD1;DDX52;WDR43;RRP9;DKC1;BMS1;MPHOSPH10;DCAF13 |
| 767 | Sphingolipid de novo biosynthesis Homo sapiens R-HSA-1660661 | 0.310427 | 0.569092 | 1.300834 | 1.5217252 | ALDH3A2;SGPL1;CERS5;SPTLC1;SGMS1;SPTLC2;OSBP;CSNK1G2;CERS2 |
| 768 | Reversal of alkylation damage by DNA dioxygenases Homo sapiens R-HSA-73943 | 0.31216 | 0.569092 | 2.312104 | 2.691842 | ASCC2;ASCC3 |
| 769 | Catabolism of glucuronate to xylulose-5-phosphate Homo sapiens R-HSA-5661270 | 0.31216 | 0.569092 | 2.312104 | 2.691842 | AKR1A1;CRYL1 |
| 770 | APOBEC3G mediated resistance to HIV-1 infection Homo sapiens R-HSA-180689 | 0.31216 | 0.569092 | 2.312104 | 2.691842 | PSIP1;PPIA |
| 771 | Disinhibition of SNARE formation Homo sapiens R-HSA-114516 | 0.31216 | 0.569092 | 2.312104 | 2.691842 | STXBP3;STX4 |
| 772 | N-glycan trimming and elongation in the cis-Golgi Homo sapiens R-HSA-964739 | 0.31216 | 0.569092 | 2.312104 | 2.691842 | MAN1C1;MAN1A1 |
| 773 | PTK6 Regulates Proteins Involved in RNA Processing Homo sapiens R-HSA-8849468 | 0.31216 | 0.569092 | 2.312104 | 2.691842 | KHDRBS1;SFPQ |
| 774 | IRS activation Homo sapiens R-HSA-74713 | 0.31216 | 0.569092 | 2.312104 | 2.691842 | INSR;IRS2 |

| | | | | | | |
|---|---|---|---|---|---|---|
| 775 | NOSTRIN mediated eNOS trafficking Homo sapiens R-HSA-203641 | 0.31216 | 0.569092 | 2.312104 | 2.691842 | WASL;DNM2 |
| 776 | SMAC binds to IAPs Homo sapiens R-HSA-111463 | 0.31216 | 0.569092 | 2.312104 | 2.691842 | CASP7;XIAP |
| 777 | SMAC-mediated dissociation of IAP:caspase complexes Homo sapiens R-HSA-111464 | 0.31216 | 0.569092 | 2.312104 | 2.691842 | CASP7;XIAP |
| 778 | SMAC-mediated apoptotic response Homo sapiens R-HSA-111469 | 0.31216 | 0.569092 | 2.312104 | 2.691842 | CASP7;XIAP |
| 779 | Arachidonate production from DAG Homo sapiens R-HSA-426048 | 0.31216 | 0.569092 | 2.312104 | 2.691842 | ABHD12;MGLL |
| 780 | Alpha-oxidation of phytanate Homo sapiens R-HSA-389599 | 0.31216 | 0.569092 | 2.312104 | 2.691842 | ALDH3A2;HACL1 |
| 781 | mRNA Capping Homo sapiens R-HSA-72086 | 0.315058 | 0.573639 | 1.321442 | 1.526264 | CDK7;RNMT;POLR2B;POLR2C;POLR2E;POLR2H;POLR2J;GTF2F2 |
| 782 | Metal ion SLC transporters Homo sapiens R-HSA-425410 | 0.319528 | 0.576942 | 1.348931 | 1.53901 | SLC30A6;SLC31A1;SLC30A5;SLC39A7;SLC39A1;SLC39A4;SLC39A3 |
| 783 | ATF4 activates genes Homo sapiens R-HSA-380994 | 0.319528 | 0.576942 | 1.348931 | 1.53901 | EXOSC7;NFYC;EXOSC8;PARN;ATF6;DCP2;ATF4 |
| 784 | APC-Cdc20 mediated degradation of Nek2A Homo sapiens R-HSA-179409 | 0.319528 | 0.576942 | 1.348931 | 1.53901 | ANAPC16;UBB;UBC;UBE2E1;ANAPC5;ANAPC11;MAD2L1 |
| 785 | Basigin interactions Homo sapiens R-HSA-210991 | 0.319528 | 0.576942 | 1.348931 | 1.53901 | ITGB1;PPIL2;ITGA3;ATP1B3;ITGA6;ATP1B1;PPIA |
| 786 | Chondroitin sulfate/dermatan sulfate metabolism Homo sapiens R-HSA-1793185 | 0.319643 | 0.576942 | 1.218868 | 1.3901825 | SDC4;B3GAT3;HEXB;SDC2;HEXA;GXYLT1;HSPG2;CHST12;CHST15;SDC1;AGRN;B4GALT7;ARSB |

| | | | | | | |
|---|---|---|---|---|---|---|
| 787 | Translocation of ZAP-70 to Immunological synapse Homo sapiens R-HSA-202430 | 0.323631 | 0.576942 | 1.38743 | 1.5652306 | HLA-DPB1;HLA-DRA;HLA-DQA2;HLA-DRB1;HLA-DPA1;HLA-DQB1 |
| 788 | CD209 (DC-SIGN) signaling Homo sapiens R-HSA-5621575 | 0.323631 | 0.576942 | 1.38743 | 1.5652306 | RPS6KA5;EP300;RAF1;PAK2;RELA;NFKB1 |
| 789 | Cobalamin (Cbl, vitamin B12) transport and metabolism Homo sapiens R-HSA-196741 | 0.323631 | 0.576942 | 1.38743 | 1.5652306 | ABCD4;AMN;ABCC1;LMBRD1;MTR;CD320 |
| 790 | Regulation of actin dynamics for phagocytic cup formation Homo sapiens R-HSA-2029482 | 0.324914 | 0.576942 | 1.140681 | 1.282347 | ACTR3;VAV3;CYFIP1;NCKAP1;ACTR2;HSP90AA1;HSP90AB1;MYO10;ARPC1B;ARPC1A;ARPC4;WASL;PTK2;ACTG1;CDC42;ARPC2;ABI2;ARPC3;CFL1;MYH9;MAPK1;GRB2;RAC1;MAPK3 |
| 791 | Organic cation transport Homo sapiens R-HSA-549127 | 0.326373 | 0.576942 | 1.734131 | 1.9417319 | SLC22A5;SLC22A15;SLC22A18 |
| 792 | Transport of nucleotide sugars Homo sapiens R-HSA-727802 | 0.326373 | 0.576942 | 1.734131 | 1.9417319 | SLC35B3;SLC35A1;SLC35A3 |
| 793 | Type I hemidesmosome assembly Homo sapiens R-HSA-446107 | 0.326373 | 0.576942 | 1.734131 | 1.9417319 | CD151;LAMA3;ITGA6 |
| 794 | Sodium/Proton exchangers Homo sapiens R-HSA-425986 | 0.326373 | 0.576942 | 1.734131 | 1.9417319 | SLC9A4;SLC9A6;SLC9A1 |
| 795 | A third proteolytic cleavage releases NICD Homo sapiens R-HSA-157212 | 0.326373 | 0.576942 | 1.734131 | 1.9417319 | APH1A;PSENEN;APH1B |
| 796 | EGFR Transactivation by Gastrin Homo sapiens R-HSA-2179392 | 0.326373 | 0.576942 | 1.734131 | 1.9417319 | GRB2;SOS1;EGFR |

| | | | | | | |
|---|---|---|---|---|---|---|
| 797 | Mitochondrial iron-sulfur cluster biogenesis Homo sapiens R-HSA-1362409 | 0.326373 | 0.576942 | 1.734131 | 1.9417319 | SLC25A28;LYRM4;FDX1 |
| 798 | TNF receptor superfamily (TNFSF) members mediating non-canonical NF-kB pathway Homo sapiens R-HSA-5676594 | 0.326963 | 0.576942 | 1.445196 | 1.6155983 | CD40;TNFRSF12A;TNFSF12;LTBR;BIRC2 |
| 799 | KSRP (KHSRP) binds and destabilizes mRNA Homo sapiens R-HSA-450604 | 0.326963 | 0.576942 | 1.445196 | 1.6155983 | EXOSC7;EXOSC8;PARN;AKT1;DCP2 |
| 800 | Formation of Senescence-Associated Heterochromatin Foci (SAHF) Homo sapiens R-HSA-2559584 | 0.326963 | 0.576942 | 1.445196 | 1.6155983 | RB1;H1F0;UBN1;HIST1H1E;HIST1H1C |
| 801 | Beta-catenin phosphorylation cascade Homo sapiens R-HSA-196299 | 0.326963 | 0.576942 | 1.445196 | 1.6155983 | GSK3B;APC;CSNK1A1;PPP2R5B;CTNNB1 |
| 802 | Synthesis, secretion, and deacylation of Ghrelin Homo sapiens R-HSA-422085 | 0.326963 | 0.576942 | 1.445196 | 1.6155983 | SPCS3;SPCS2;SPCS1;SEC11A;SEC11C |
| 803 | Chylomicron-mediated lipid transport Homo sapiens R-HSA-174800 | 0.326963 | 0.576942 | 1.445196 | 1.6155983 | LIPC;SAR1B;SDC1;P4HB;HSPG2 |
| 804 | Inflammasomes Homo sapiens R-HSA-622312 | 0.326963 | 0.576942 | 1.445196 | 1.6155983 | PYCARD;PANX1;APP;HSP90AB1;SUGT1 |
| 805 | NICD traffics to nucleus Homo sapiens R-HSA-157052 | 0.328638 | 0.576942 | 1.541496 | 1.7153747 | KAT2A;SNW1;MAML3;RBPJ |

| | | | | | | |
|---|---|---|---|---|---|---|
| 806 | Notch-HLH transcription pathway Homo sapiens R-HSA-350054 | 0.328638 | 0.576942 | 1.541496 | 1.7153747 | KAT2A;SNW1;MAML3;RBPJ |
| 807 | p75NTR recruits signalling complexes Homo sapiens R-HSA-209543 | 0.328638 | 0.576942 | 1.541496 | 1.7153747 | PRKCI;UBB;UBC;SQSTM1 |
| 808 | alpha-linolenic (omega3) and linoleic (omega6) acid metabolism Homo sapiens R-HSA-2046104 | 0.328638 | 0.576942 | 1.541496 | 1.7153747 | ELOVL1;SCP2;HSD17B4;ACAA1 |
| 809 | alpha-linolenic acid (ALA) metabolism Homo sapiens R-HSA-2046106 | 0.328638 | 0.576942 | 1.541496 | 1.7153747 | ELOVL1;SCP2;HSD17B4;ACAA1 |
| 810 | Purine salvage Homo sapiens R-HSA-74217 | 0.328638 | 0.576942 | 1.541496 | 1.7153747 | GMPR2;ADK;GMPR;AMPD2 |
| 811 | Negative epigenetic regulation of rRNA expression Homo sapiens R-HSA-5250941 | 0.333329 | 0.584456 | 1.156423 | 1.2704747 | DNMT1;HDAC2;H2AFZ;HDAC1;H3F3A;MBD2;SMARCA5;ARID4B;SIRT1;TAF1D;CDK7;TAF1B;SAP30L;POLR1D;POLR2E;SAP18;SUDS3;POLR2H;HIST1H2AC |
| 812 | Transport of vitamins, nucleosides, and related molecules Homo sapiens R-HSA-425397 | 0.338345 | 0.592521 | 1.238847 | 1.3425246 | SLC35B3;SLC35A1;SLC35A3;SLCO3A1;SLCO4A1;SLC33A1;SLCO4C1;SLC29A4;SLC27A4;SLC29A2 |
| 813 | Transcriptional regulation by small RNAs Homo sapiens R-HSA-5578749 | 0.339706 | 0.593442 | 1.156388 | 1.248523 | NUP214;RANBP2;NUP205;H2AFZ;H3F3A;AAAS;POM121C;POLR2B;NUP50;POLR2C;TPR;POLR2E;NUP98;POLR2H;POLR2J;HIST1H2AC;TNRC6A;RAN |
| 814 | TRAF6 Mediated Induction of proinflammatory cytokines Homo sapiens R-HSA-168180 | 0.339706 | 0.593442 | 1.156388 | 1.248523 | ATF2;APP;MAP2K1;FBXW11;DHX9;CUL1;HMGB1;RELA;NFKB1;NFKBIA;MAPK10;RPS6KA5;CREB1;UBB;TNIP2;UBC;MAPK1;MAPK3 |
| 815 | Transport of the SLBP Dependant Mature mRNA Homo sapiens R-HSA-159230 | 0.345186 | 0.601537 | 1.24872 | 1.3282284 | RANBP2;NUP214;SLBP;NUP205;POM121C;NUP50;TPR;NUP98;AAAS |

| | | | | | | |
|---|---|---|---|---|---|---|
| 816 | Nuclear Pore Complex (NPC) Disassembly Homo sapiens R-HSA-3301854 | 0.345186 | 0.601537 | 1.24872 | 1.3282284 | RANBP2;NUP214;NUP205;POM121C;NUP50;TPR;NEK7;NUP98;AAAS |
| 817 | Metabolism of fat-soluble vitamins Homo sapiens R-HSA-6806667 | 0.347959 | 0.605628 | 1.186716 | 1.2527797 | VKORC1L1;UBIAD1;SDC4;SDC2;AKR1C1;AKR1C3;HSPG2;RDH11;CYP2R1;SDC1;LRP10;AGRN;LGMN |
| 818 | SLC-mediated transmembrane transport Homo sapiens R-HSA-425407 | 0.351466 | 0.610984 | 1.066489 | 1.115165 | SLC35B3;SLC44A3;SLC44A4;SLC44A1;SLC44A2;SLC2A1;SLC4A1;HK1;SLC22A15;SLC22A18;SLC16A7;SLC39A7;SLC12A6;SLC37A4;SLC12A7;SLC39A1;SLC39A4;SLC39A3;NUP214;SLC34A3;SLC30A6;SLC38A1;SLC30A5;SLC6A13;SLC6A12;SLC9A4;SLC9A6;SLCO4A1;NUP50;SLC29A4;NUP98;RHCG;SLC29A2;SLC27A4;NUP205;SLC22A5;SRI;AAAS;SLC5A3;SLC7A1;SLC9A1;SLC25A26;POM121C;TPR;SLC17A5;RHBG;SLC38A2;RANBP2;SLC35A1;SLC14A2;SLC35A3;SLC31A1;SLC33A1;CTNS;SLC16A10;SLC8B1;SLC4A7;SLC6A6;G6PC3;SLCO3A1;SLCO4C1;SLC26A7;CALM1 |
| 819 | Regulation of Glucokinase by Glucokinase Regulatory Protein Homo sapiens R-HSA-170822 | 0.352389 | 0.611523 | 1.261295 | 1.315556 | RANBP2;NUP214;NUP205;POM121C;NUP50;TPR;NUP98;AAAS |
| 820 | Mitotic Prometaphase Homo sapiens R-HSA-68877 | 0.352637 | 0.611523 | 1.113613 | 1.1607383 | AHCTF1;PDS5B;SMC3;PDS5A;HDAC8;PMF1;SMC2;SKA2;CENPC;XPO1;RAD21;CENPT;RANBP2;CSNK2A1;CSNK2A2;PPP2R5B;SMC1A;CLIP1;STAG1;RPS27;STAG2;KIF2A;NUP98;MAPRE1;PAFAH1B1;MAD2L1 |
| 821 | HDR through Homologous Recombination (HRR) Homo sapiens R-HSA-5685942 | 0.353365 | 0.612041 | 1.156318 | 1.2028624 | MUS81;RFC1;TOP3A;RPA2;SPIDR;POLD4;RAD50;UBB;RPA3;UBC;RAD17;ATM;NBN;TOPBP1;ATRIP;ATR |
| 822 | N-glycan antennae elongation in the medial/trans-Golgi Homo sapiens R-HSA-975576 | 0.359997 | 0.622011 | 1.277852 | 1.3055309 | B4GALT3;FUT8;B4GALT1;MAN2A2;MAN2A1;FUCA1;MGAT4B |
| 823 | PTK6 Regulates RHO GTPases, RAS GTPase and MAP kinases Homo sapiens R-HSA-8849471 | 0.359997 | 0.622011 | 1.277852 | 1.3055309 | RASA2;DAB2IP;NF1;RAC1;ARHGAP35;RHOA;BCAR1 |

| | | | | | | |
|---|---|---|---|---|---|---|
| 824 | NoRC negatively regulates rRNA expression Homo sapiens R-HSA-427413 | 0.363648 | 0.627558 | 1.135289 | 1.148423 | DNMT1;HDAC2;H2AFZ;HDAC1;H3F3A;MBD2;SMARCA5;ARID4B;TAF1D;CDK7;TAF1B;SAP30L;POLR1D;POLR2E;SAP18;SUDS3;POLR2H;HIST1H2AC |
| 825 | HDR through Homologous Recombination (HR) or Single Strand Annealing (SSA) Homo sapiens R-HSA-5693567 | 0.365753 | 0.630426 | 1.101985 | 1.108373 | MUS81;RNF8;BABAM1;POLD4;HERC2;PPP4C;PPP4R2;UBB;UBC;UIMC1;SUMO2;NBN;TP53BP1;TOPBP1;ATRIP;UBE2I;RFC1;TOP3A;RPA2;SIRT6;SPIDR;CCNA1;RAD50;RPA3;RAD17;ATM;ATR |
| 826 | RA biosynthesis pathway Homo sapiens R-HSA-5365859 | 0.368055 | 0.630572 | 1.300632 | 1.3000104 | RDH11;RDH10;AKR1C3;RDH14;RDH13;DHRS3 |
| 827 | CD28 dependent PI3K/Akt signaling Homo sapiens R-HSA-389357 | 0.368055 | 0.630572 | 1.300632 | 1.3000104 | AKT2;MLST8;PIK3R3;THEM4;AKT1;MTOR |
| 828 | CTLA4 inhibitory signaling Homo sapiens R-HSA-389513 | 0.368055 | 0.630572 | 1.300632 | 1.3000104 | YES1;SRC;AKT2;PPP2R5B;AKT1;PTPN11 |
| 829 | Nephrin interactions Homo sapiens R-HSA-373753 | 0.368055 | 0.630572 | 1.300632 | 1.3000104 | ACTN4;WASL;IQGAP1;SPTAN1;SPTBN1;CD2AP |
| 830 | Post-chaperonin tubulin folding pathway Homo sapiens R-HSA-389977 | 0.368055 | 0.630572 | 1.300632 | 1.3000104 | TUBA1C;TUBA1B;TUBB2A;TBCB;TUBB4B;TUBA4A |
| 831 | Gamma carboxylation, hypusine formation and arylsulfatase activation Homo sapiens R-HSA-163841 | 0.371502 | 0.633876 | 1.196051 | 1.1843294 | ARSA;ICMT;ARSK;DHPS;FN3KRP;FURIN;EEF2;ARSB;SUMF2;SUMF1 |
| 832 | Presynaptic phase of homologous DNA pairing and strand exchange Homo sapiens R-HSA-5693616 | 0.371502 | 0.633876 | 1.196051 | 1.1843294 | RAD50;RPA3;TOP3A;RPA2;RAD17;ATM;NBN;TOPBP1;ATRIP;ATR |
| 833 | Nuclear signaling by ERBB4 Homo sapiens R-HSA-1251985 | 0.371502 | 0.633876 | 1.196051 | 1.1843294 | YAP1;APH1A;PSENEN;ADAM17;APH1B;NCOR1;CUL1;PTPN11;JAK2;ESR1 |

| | | | | | | |
|---|---|---|---|---|---|---|
| 834 | Glycosaminoglycan metabolism Homo sapiens R-HSA-1630316 | 0.371767 | 0.633876 | 1.093552 | 1.0820562 | SLC35B3;B4GALT3;B4GALT1;SDC4;SDC2;HEXB;HEXA;GXYLT1;HS6ST1;GNS;PAPSS2;SLC9A1;NAGLU;CHST12;HYAL2;CHST15;ST3GAL6;GUSB;ARSB;B3GAT3;ABCC5;HSPG2;HS3ST1;EXT2;B3GNT3;SDC1;B3GNT2;AGRN;B4GALT7 |
| 835 | Activation of SMO Homo sapiens R-HSA-5635838 | 0.376602 | 0.639055 | 1.333941 | 1.3026824 | EVC2;SHH;CSNK1A1;KIF3A;ARRB1 |
| 836 | SHC1 events in ERBB2 signaling Homo sapiens R-HSA-1250196 | 0.376602 | 0.639055 | 1.333941 | 1.3026824 | ERBB3;ERBB2;GRB2;SOS1;EGFR |
| 837 | Tie2 Signaling Homo sapiens R-HSA-210993 | 0.376602 | 0.639055 | 1.333941 | 1.3026824 | GRB7;GRB14;PTPN11;GRB2;SOS1 |
| 838 | Regulation of signaling by CBL Homo sapiens R-HSA-912631 | 0.376602 | 0.639055 | 1.333941 | 1.3026824 | YES1;BLNK;PIK3R3;GRB2;CBL |
| 839 | Regulation of TP53 Degradation Homo sapiens R-HSA-6804757 | 0.380469 | 0.642861 | 1.200615 | 1.1602161 | CCNA1;USP7;UBB;AKT2;UBC;MLST8;AKT1;ATM;MTOR |
| 840 | Death Receptor Signalling Homo sapiens R-HSA-73887 | 0.385461 | 0.642861 | 1.156178 | 1.1022011 | ADAM17;SHARPIN;UBB;UBC;TAX1BP1;XIAP;FAS;RIPK1;CFLAR;RBCK1;BIRC2;TNFRSF1A |
| 841 | Receptor-ligand binding initiates the second proteolytic cleavage of Notch receptor Homo sapiens R-HSA-156988 | 0.385626 | 0.642861 | 1.387257 | 1.3218988 | ADAM17;UBB;UBC;ADAM10 |
| 842 | AMER1 mutants destabilize the destruction complex Homo sapiens R-HSA-4839748 | 0.385626 | 0.642861 | 1.387257 | 1.3218988 | GSK3B;APC;CSNK1A1;PPP2R5B |
| 843 | Truncations of AMER1 destabilize the destruction complex Homo sapiens R-HSA-5467348 | 0.385626 | 0.642861 | 1.387257 | 1.3218988 | GSK3B;APC;CSNK1A1;PPP2R5B |

| | | | | | | |
|---|---|---|---|---|---|---|
| 844 | truncated APC mutants destabilize the destruction complex Homo sapiens R-HSA-4839744 | 0.385626 | 0.642861 | 1.387257 | 1.3218988 | GSK3B;APC;CSNK1A1;PPP2R5B |
| 845 | AXIN missense mutants destabilize the destruction complex Homo sapiens R-HSA-5467340 | 0.385626 | 0.642861 | 1.387257 | 1.3218988 | GSK3B;APC;CSNK1A1;PPP2R5B |
| 846 | AXIN mutants destabilize the destruction complex, activating WNT signaling Homo sapiens R-HSA-4839735 | 0.385626 | 0.642861 | 1.387257 | 1.3218988 | GSK3B;APC;CSNK1A1;PPP2R5B |
| 847 | APC truncation mutants have impaired AXIN binding Homo sapiens R-HSA-5467337 | 0.385626 | 0.642861 | 1.387257 | 1.3218988 | GSK3B;APC;CSNK1A1;PPP2R5B |
| 848 | TRAF3-dependent IRF activation pathway Homo sapiens R-HSA-918233 | 0.385626 | 0.642861 | 1.387257 | 1.3218988 | TBK1;IRF3;IRF7;EP300 |
| 849 | Inhibition of adenylate cyclase pathway Homo sapiens R-HSA-997269 | 0.385626 | 0.642861 | 1.387257 | 1.3218988 | ADCY9;GNAI3;ADCY6;GNAI2 |
| 850 | Adenylate cyclase inhibitory pathway Homo sapiens R-HSA-170670 | 0.385626 | 0.642861 | 1.387257 | 1.3218988 | ADCY9;GNAI3;ADCY6;GNAI2 |
| 851 | Methylation Homo sapiens R-HSA-156581 | 0.385626 | 0.642861 | 1.387257 | 1.3218988 | GSTO1;CYP1A2;MTR;COMT |
| 852 | Nicotinate metabolism Homo sapiens R-HSA-196807 | 0.385626 | 0.642861 | 1.387257 | 1.3218988 | NMNAT1;QPRT;NADSYN1;NADK |

| | | | | | | |
|---|---|---|---|---|---|---|
| 853 | Glutathione synthesis and recycling Homo sapiens R-HSA-174403 | 0.385626 | 0.642861 | 1.387257 | 1.3218988 | GGCT;GCLC;GGT6;CNDP2 |
| 854 | HS-GAG biosynthesis Homo sapiens R-HSA-2022928 | 0.390185 | 0.646952 | 1.206378 | 1.1353648 | EXT2;SDC4;SDC2;SDC1;AGRN;HS6ST1;HSPG2;HS3ST1 |
| 855 | Inositol phosphate metabolism Homo sapiens R-HSA-1483249 | 0.394842 | 0.646952 | 1.156143 | 1.0743678 | ITPKB;MINPP1;ISYNA1;INPP1;PPIP5K2;ITPK1;INPP5J;OCRL;PLCG1;CALM1;IP6K2 |
| 856 | Nef mediated downregulation of MHC class I complex cell surface expression Homo sapiens R-HSA-164940 | 0.394905 | 0.646952 | 1.486302 | 1.3809398 | AP1G1;B2M;AP1M2 |
| 857 | Activation of Ca-permeable Kainate Receptor Homo sapiens R-HSA-451308 | 0.394905 | 0.646952 | 1.486302 | 1.3809398 | DLG1;DLG3;CALM1 |
| 858 | Ionotropic activity of Kainate Receptors Homo sapiens R-HSA-451306 | 0.394905 | 0.646952 | 1.486302 | 1.3809398 | DLG1;DLG3;CALM1 |
| 859 | Removal of the Flap Intermediate from the C-strand Homo sapiens R-HSA-174437 | 0.394905 | 0.646952 | 1.486302 | 1.3809398 | POLD4;RPA3;RPA2 |
| 860 | Regulation of gene expression by Hypoxia-inducible Factor Homo sapiens R-HSA-1234158 | 0.394905 | 0.646952 | 1.486302 | 1.3809398 | EP300;HIF3A;HIF1A |
| 861 | Diseases of carbohydrate metabolism Homo sapiens R-HSA-5663084 | 0.394905 | 0.646952 | 1.486302 | 1.3809398 | UBB;PPP1R3C;UBC |
| 862 | Myoclonic epilepsy of Lafora Homo sapiens R-HSA-3785653 | 0.394905 | 0.646952 | 1.486302 | 1.3809398 | UBB;PPP1R3C;UBC |

| | | | | | | |
|---|---|---|---|---|---|---|
| 863 | Glycogen storage diseases Homo sapiens R-HSA-3229121 | 0.394905 | 0.646952 | 1.486302 | 1.3809398 | UBB;PPP1R3C;UBC |
| 864 | IRAK1 recruits IKK complex Homo sapiens R-HSA-937039 | 0.394905 | 0.646952 | 1.486302 | 1.3809398 | PELI3;PELI1;UBE2V1 |
| 865 | IRAK1 recruits IKK complex upon TLR7/8 or 9 stimulation Homo sapiens R-HSA-975144 | 0.394905 | 0.646952 | 1.486302 | 1.3809398 | PELI3;PELI1;UBE2V1 |
| 866 | Netrin mediated repulsion signals Homo sapiens R-HSA-418886 | 0.394905 | 0.646952 | 1.486302 | 1.3809398 | SRC;PTPN11;PTK2 |
| 867 | Ligand-independent caspase activation via DCC Homo sapiens R-HSA-418889 | 0.394905 | 0.646952 | 1.486302 | 1.3809398 | DAPK1;DAPK3;APPL1 |
| 868 | Calcitonin-like ligand receptors Homo sapiens R-HSA-419812 | 0.394905 | 0.646952 | 1.486302 | 1.3809398 | CALCA;CALCR;CALCB |
| 869 | Cell-Cell communication Homo sapiens R-HSA-1500931 | 0.395485 | 0.647158 | 1.075378 | 0.9975654 | ITGB1;CD151;SRC;LAMA3;TESK1;ILK;IQGAP1;WASL;F11R;PARD6B;CDH1;RAC1;SPTAN1;SPTBN1;VASP;PRKCI;PTPN11;ACTN4;PTK2;CD2AP;MAPK13;CLDN4;CLDN8;CLDN7;CTNNB1;GRB2;ITGA6;ANG;CD47;LIMS1;CRB3 |
| 870 | Caspase activation via extrinsic apoptotic signalig pathway Homo sapiens R-HSA-5357769 | 0.400833 | 0.650624 | 1.213881 | 1.1097437 | DAPK1;DAPK3;FAS;RIPK1;CFLAR;TICAM1;APPL1 |
| 871 | Synthesis of bile acids and bile salts Homo sapiens R-HSA-192105 | 0.400833 | 0.650624 | 1.213881 | 1.1097437 | NCOA1;CYP39A1;NCOA2;SCP2;AKR1C1;AKR1C3;HSD17B4 |
| 872 | DNA Damage Reversal Homo sapiens R-HSA-73942 | 0.403094 | 0.650624 | 1.733966 | 1.5754554 | ASCC2;ASCC3 |

| | | | | | | |
|---|---|---|---|---|---|---|
| 873 | SUMO is proteolytically processed Homo sapiens R-HSA-3065679 | 0.403094 | 0.650624 | 1.733966 | 1.5754554 | SUMO3;SUMO2 |
| 874 | IRF3 mediated activation of type 1 IFN Homo sapiens R-HSA-1606341 | 0.403094 | 0.650624 | 1.733966 | 1.5754554 | TBK1;IRF3 |
| 875 | Telomere Extension By Telomerase Homo sapiens R-HSA-171319 | 0.403094 | 0.650624 | 1.733966 | 1.5754554 | DKC1;RUVBL1 |
| 876 | Scavenging by Class F Receptors Homo sapiens R-HSA-3000484 | 0.403094 | 0.650624 | 1.733966 | 1.5754554 | HSP90AA1;CALR |
| 877 | NADE modulates death signalling Homo sapiens R-HSA-205025 | 0.403094 | 0.650624 | 1.733966 | 1.5754554 | YWHAE;CASP2 |
| 878 | p75NTR negatively regulates cell cycle via SC1 Homo sapiens R-HSA-193670 | 0.403094 | 0.650624 | 1.733966 | 1.5754554 | HDAC2;HDAC1 |
| 879 | Transport and synthesis of PAPS Homo sapiens R-HSA-174362 | 0.403094 | 0.650624 | 1.733966 | 1.5754554 | SLC35B3;PAPSS2 |
| 880 | Heme degradation Homo sapiens R-HSA-189483 | 0.403094 | 0.650624 | 1.733966 | 1.5754554 | BLVRB;BLVRA |
| 881 | Protein repair Homo sapiens R-HSA-5676934 | 0.403094 | 0.650624 | 1.733966 | 1.5754554 | PCMT1;MSRA |
| 882 | G alpha (12/13) signalling events Homo sapiens R-HSA-416482 | 0.412288 | 0.663085 | 1.095313 | 0.9704828 | VAV3;ARHGEF12;ROCK1;ROCK2;RASGRF2;ITSN1;ARHGEF16;PIK3R3;GNG11;RHOA;CDC42;GNG5;MCF2L;ARHGEF1;RAC1;ARHGEF2;SOS1;ECT2 |
| 883 | Gap-filling DNA repair synthesis and ligation in GG-NER Homo sapiens R-HSA-5696397 | 0.412679 | 0.663085 | 1.224045 | 1.0833834 | POLD4;RFC1;UBB;RPA3;UBC;RPA2 |

| | | | | | | |
|---|---|---|---|---|---|---|
| 884 | APC/C:Cdc20 mediated degradation of Cyclin B Homo sapiens R-HSA-174048 | 0.412679 | 0.663085 | 1.224045 | 1.0833834 | ANAPC16;UBB;UBC;UBE2E1;ANAPC5;ANAPC11 |
| 885 | Incretin synthesis, secretion, and inactivation Homo sapiens R-HSA-400508 | 0.412679 | 0.663085 | 1.224045 | 1.0833834 | SPCS3;SPCS2;SPCS1;CTNNB1;SEC11A;SEC11C |
| 886 | Regulation of TP53 Expression and Degradation Homo sapiens R-HSA-6806003 | 0.41597 | 0.667617 | 1.156073 | 1.0140408 | CCNA1;USP7;UBB;AKT2;UBC;MLST8;AKT1;ATM;MTOR |
| 887 | Degradation of the extracellular matrix Homo sapiens R-HSA-1474228 | 0.420406 | 0.673977 | 1.070541 | 0.9276603 | COL14A1;COL12A1;LAMA3;FURIN;LAMC1;CAPN15;CTSS;CAPNS1;CAPN7;CDH1;CTSL;ADAMTS1;ADAMTS18;CAPN2;CAPN1;CTSD;CTSB;CAST;MMP7;PHYKPL;ADAM10;LAMB1;HSPG2;ADAM17;ADAM9 |
| 888 | Defective B3GALT6 causes EDSP2 and SEMDJL1 Homo sapiens R-HSA-4420332 | 0.42614 | 0.67934 | 1.23858 | 1.0564917 | SDC4;SDC2;SDC1;AGRN;HSPG2 |
| 889 | Defective B4GALT7 causes EDS, progeroid type Homo sapiens R-HSA-3560783 | 0.42614 | 0.67934 | 1.23858 | 1.0564917 | SDC4;SDC2;SDC1;AGRN;HSPG2 |
| 890 | Defective B3GAT3 causes JDSSDHD Homo sapiens R-HSA-3560801 | 0.42614 | 0.67934 | 1.23858 | 1.0564917 | SDC4;SDC2;SDC1;AGRN;HSPG2 |
| 891 | PCNA-Dependent Long Patch Base Excision Repair Homo sapiens R-HSA-5651801 | 0.42614 | 0.67934 | 1.23858 | 1.0564917 | POLD4;RFC1;RPA3;APEX1;RPA2 |
| 892 | Conversion from APC/C:Cdc20 to APC/C:Cdh1 in late anaphase Homo sapiens R-HSA-176407 | 0.42614 | 0.67934 | 1.23858 | 1.0564917 | FZR1;ANAPC16;UBE2E1;ANAPC5;ANAPC11 |
| 893 | NRAGE signals death through JNK Homo sapiens R-HSA-193648 | 0.426671 | 0.679424 | 1.122066 | 0.9557113 | VAV3;ARHGEF12;BAD;RASGRF2;MCF2L;ITSN1;ARHGEF16;RAC1;ARHGEF2;ECT2;SOS1 |

| | | | | | | |
|---|---|---|---|---|---|---|
| 894 | Keratan sulfate/keratin metabolism Homo sapiens R-HSA-1638074 | 0.428079 | 0.680904 | 1.156038 | 0.9808383 | B4GALT3;B4GALT1;HEXB;HEXA;B3GNT3;B3GNT2;ST3GAL6;GNS |
| 895 | Heparan sulfate/heparin (HS-GAG) metabolism Homo sapiens R-HSA-1638091 | 0.434811 | 0.690839 | 1.09967 | 0.9158538 | SDC4;B3GAT3;SDC2;GXYLT1;HS6ST1;HSPG2;HS3ST1;EXT2;NAGLU;SDC1;GUSB;AGRN;B4GALT7 |
| 896 | RNA Polymerase I Promoter Clearance Homo sapiens R-HSA-73854 | 0.436793 | 0.693214 | 1.076359 | 0.8915438 | HDAC2;H2AFZ;HDAC1;H3F3A;MBD2;CHD4;CHD3;KAT2A;TAF1D;CDK7;RBBP4;TAF1B;POLR1D;POLR2E;RBBP7;POLR2H;HIST1H2AC;MAPK3 |
| 897 | TNF signaling Homo sapiens R-HSA-75893 | 0.438471 | 0.695101 | 1.118742 | 0.9223597 | ADAM17;SHARPIN;UBB;UBC;TAX1BP1;XIAP;RIPK1;RBCK1;BIRC2;TNFRSF1A |
| 898 | Synthesis of substrates in N-glycan biosythesis Homo sapiens R-HSA-446219 | 0.441311 | 0.69515 | 1.083805 | 0.8865591 | SLC35A1;CMAS;ALG5;GFPT1;GMPPB;DPM2;DPM3;NAGK;DOLPP1;NEU1;PGM3;SLC17A5;ST3GAL6;NANS;GNE |
| 899 | Gap junction trafficking Homo sapiens R-HSA-190828 | 0.441586 | 0.69515 | 1.156003 | 0.9448971 | DAB2;GJB1;CLTC;MYO6;CLTB;AP2M1;DNM2 |
| 900 | The role of Nef in HIV-1 replication and disease pathogenesis Homo sapiens R-HSA-164952 | 0.441586 | 0.69515 | 1.156003 | 0.9448971 | AP1G1;AP2A1;RAC1;PAK2;B2M;AP1M2;AP2M1 |
| 901 | Ca-dependent events Homo sapiens R-HSA-111996 | 0.441586 | 0.69515 | 1.156003 | 0.9448971 | CREB1;ADCY9;PRKAR2A;PRKCD;MAPK1;CALM1;ADCY6 |
| 902 | mRNA decay by 5' to 3' exoribonuclease Homo sapiens R-HSA-430039 | 0.441924 | 0.69515 | 1.261061 | 1.0298049 | LSM7;XRN1;LSM1;DCP2 |
| 903 | Glycogen synthesis Homo sapiens R-HSA-3322077 | 0.441924 | 0.69515 | 1.261061 | 1.0298049 | UGP2;UBB;PPP1R3C;UBC |
| 904 | Synthesis of bile acids and bile salts via 27-hydroxycholesterol Homo sapiens R-HSA-193807 | 0.441924 | 0.69515 | 1.261061 | 1.0298049 | NCOA1;NCOA2;AKR1C1;AKR1C3 |

| | | | | | | |
|---|---|---|---|---|---|---|
| 905 | Metabolism of non-coding RNA Homo sapiens R-HSA-194441 | 0.446165 | 0.700272 | 1.095185 | 0.883887 | NUP214;RANBP2;NUP205;SNRPD2;POM121C;NUP50;TGS1;TPR;PHAX;NUP98;AAAS;SNUPN |
| 906 | snRNP Assembly Homo sapiens R-HSA-191859 | 0.446165 | 0.700272 | 1.095185 | 0.883887 | NUP214;RANBP2;NUP205;SNRPD2;POM121C;NUP50;TGS1;TPR;PHAX;NUP98;AAAS;SNUPN |
| 907 | DNA Damage/Telomere Stress Induced Senescence Homo sapiens R-HSA-2559586 | 0.452204 | 0.708969 | 1.078955 | 0.856282 | RB1;CDKN1B;H2AFZ;TERF1;CCNA1;RAD50;H1F0;UBN1;HIST1H1E;TERF2IP;ATM;NBN;HIST1H2AC;HIST1H1C |
| 908 | Synthesis of bile acids and bile salts via 7alpha-hydroxycholesterol Homo sapiens R-HSA-193368 | 0.456938 | 0.710071 | 1.155968 | 0.9053642 | NCOA1;NCOA2;SCP2;AKR1C1;AKR1C3;HSD17B4 |
| 909 | Endogenous sterols Homo sapiens R-HSA-211976 | 0.456938 | 0.710071 | 1.155968 | 0.9053642 | NCOA1;CYP39A1;NCOA2;CYP51A1;FDX1;CYP17A1 |
| 910 | Amyloid fiber formation Homo sapiens R-HSA-977225 | 0.45714 | 0.710071 | 1.067094 | 0.8352853 | APP;CALCA;GSN;H2AFZ;USP9X;ODAM;H3F3A;FURIN;HSPG2;GGA2;GGA1;UBB;UBC;B2M;HIST1H2AC;ITM2B |
| 911 | Cellular Senescence Homo sapiens R-HSA-2559583 | 0.458417 | 0.710071 | 1.034879 | 0.8071809 | RB1;CDKN1B;ANAPC16;EHMT1;ETS2;RELA;ANAPC11;FZR1;UBB;RBBP4;UBC;UBN1;HIST1H1E;MAPK1;NBN;RBBP7;HIST1H2AC;HIST1H1C;MAPK3;SUZ12;PHC2;RING1;H2AFZ;H3F3A;STAT3;UBE2E1;TERF1;NFKB1;MAPK10;CCNA1;H1F0;RAD50;AGO3;TERF2IP;ANAPC5;ATM;TNRC6A |
| 912 | Resolution of Sister Chromatid Cohesion Homo sapiens R-HSA-2500257 | 0.458653 | 0.710071 | 1.049562 | 0.8180937 | CENPT;RANBP2;AHCTF1;PPP2R5B;PDS5B;SMC3;PDS5A;SMC1A;HDAC8;PMF1;SKA2;CENPC;CLIP1;STAG1;RPS27;STAG2;XPO1;KIF2A;RAD21;NUP98;MAPRE1;PAFAH1B1;MAD2L1 |
| 913 | Senescence-Associated Secretory Phenotype (SASP) Homo sapiens R-HSA-2559582 | 0.461294 | 0.710071 | 1.058047 | 0.8186324 | CDKN1B;ANAPC16;H2AFZ;H3F3A;STAT3;UBE2E1;EHMT1;RELA;NFKB1;ANAPC11;CCNA1;FZR1;UBB;UBC;MAPK1;ANAPC5;HIST1H2AC;MAPK3 |
| 914 | Transport of organic anions Homo sapiens R-HSA-879518 | 0.461396 | 0.710071 | 1.30043 | 1.0058799 | SLCO3A1;SLCO4A1;SLCO4C1 |

| | | | | | | |
|---|---|---|---|---|---|---|
| 915 | Processive synthesis on the C-strand of the telomere Homo sapiens R-HSA-174414 | 0.461396 | 0.710071 | 1.30043 | 1.0058799 | POLD4;RPA3;RPA2 |
| 916 | Signaling by FGFR4 in disease Homo sapiens R-HSA-5655291 | 0.461396 | 0.710071 | 1.30043 | 1.0058799 | GRB2;PLCG1;SOS1 |
| 917 | CASP8 activity is inhibited Homo sapiens R-HSA-5218900 | 0.461396 | 0.710071 | 1.30043 | 1.0058799 | FAS;RIPK1;CFLAR |
| 918 | Regulation by c-FLIP Homo sapiens R-HSA-3371378 | 0.461396 | 0.710071 | 1.30043 | 1.0058799 | FAS;RIPK1;CFLAR |
| 919 | Dimerization of procaspase-8 Homo sapiens R-HSA-69416 | 0.461396 | 0.710071 | 1.30043 | 1.0058799 | FAS;RIPK1;CFLAR |
| 920 | Recruitment of NuMA to mitotic centrosomes Homo sapiens R-HSA-380320 | 0.461396 | 0.710071 | 1.30043 | 1.0058799 | NUMA1;TUBGCP2;TUBG1 |
| 921 | Condensation of Prometaphase Chromosomes Homo sapiens R-HSA-2514853 | 0.461396 | 0.710071 | 1.30043 | 1.0058799 | CSNK2A1;CSNK2A2;SMC2 |
| 922 | Sodium/Calcium exchangers Homo sapiens R-HSA-425561 | 0.461396 | 0.710071 | 1.30043 | 1.0058799 | SRI;CALM1;SLC8B1 |
| 923 | Defects in cobalamin (B12) metabolism Homo sapiens R-HSA-3296469 | 0.461396 | 0.710071 | 1.30043 | 1.0058799 | AMN;MTR;CD320 |
| 924 | Tryptophan catabolism Homo sapiens R-HSA-71240 | 0.461396 | 0.710071 | 1.30043 | 1.0058799 | KYNU;AFMID;IDO1 |
| 925 | Transport of the SLBP independent Mature mRNA Homo sapiens R-HSA-159227 | 0.465727 | 0.714417 | 1.109725 | 0.8480017 | RANBP2;NUP214;NUP205;POM121C;NUP50;TPR;NUP98;AAAS |

| | | | | | | |
|---|---|---|---|---|---|---|
| 926 | Formation of the Early Elongation Complex Homo sapiens R-HSA-113418 | 0.465727 | 0.714417 | 1.109725 | 0.8480017 | CDK7;POLR2B;POLR2C;POLR2E;NELFB;POLR2H;POLR2J;GTF2F2 |
| 927 | Formation of the HIV-1 Early Elongation Complex Homo sapiens R-HSA-167158 | 0.465727 | 0.714417 | 1.109725 | 0.8480017 | CDK7;POLR2B;POLR2C;POLR2E;NELFB;POLR2H;POLR2J;GTF2F2 |
| 928 | Homologous DNA Pairing and Strand Exchange Homo sapiens R-HSA-5693579 | 0.471797 | 0.722169 | 1.083711 | 0.8140914 | RAD50;RPA3;TOP3A;RPA2;RAD17;ATM;NBN;TOPBP1;ATRIP;ATR |
| 929 | Netrin-1 signaling Homo sapiens R-HSA-373752 | 0.471797 | 0.722169 | 1.083711 | 0.8140914 | CDC42;MYO10;SRC;SLIT3;PTPN11;WASL;PLCG1;RAC1;NEO1;PTK2 |
| 930 | Defective C1GALT1C1 causes Tn polyagglutination syndrome (TNPS) Homo sapiens R-HSA-5083632 | 0.474845 | 0.724496 | 1.155933 | 0.8609003 | MUC1;C1GALT1C1;C1GALT1;MUC15;MUC20 |
| 931 | Inhibition of the proteolytic activity of APC/C required for the onset of anaphase by mitotic spindle checkpoint components Homo sapiens R-HSA-141405 | 0.474845 | 0.724496 | 1.155933 | 0.8609003 | ANAPC16;UBE2E1;ANAPC5;ANAPC11;MAD2L1 |
| 932 | Inactivation of APC/C via direct inhibition of the APC/C complex Homo sapiens R-HSA-141430 | 0.474845 | 0.724496 | 1.155933 | 0.8609003 | ANAPC16;UBE2E1;ANAPC5;ANAPC11;MAD2L1 |
| 933 | mRNA 3'-end processing Homo sapiens R-HSA-72187 | 0.476382 | 0.724508 | 1.067034 | 0.7912434 | DDX39A;DDX39B;U2AF2;EIF4A3;PAPOLA;SRSF2;UPF3B;SRSF4;THOC2;SLU7;SRSF11;SRRM1 |
| 934 | Post-Elongation Processing of Intron-Containing pre-mRNA Homo sapiens R-HSA-112296 | 0.476382 | 0.724508 | 1.067034 | 0.7912434 | DDX39A;DDX39B;U2AF2;EIF4A3;PAPOLA;SRSF2;UPF3B;SRSF4;THOC2;SLU7;SRSF11;SRRM1 |

| | | | | | | |
|---|---|---|---|---|---|---|
| 935 | Nuclear Receptor transcription pathway Homo sapiens R-HSA-383280 | 0.476382 | 0.724508 | 1.067034 | 0.7912434 | ESRRA;RARG;THRB;THRA;NR1H2;RORA;PPARG;NR1D2;NR1D1;ESR1;NRBP1;NR2F6 |
| 936 | Interaction between L1 and Ankyrins Homo sapiens R-HSA-445095 | 0.481845 | 0.724807 | 1.103386 | 0.8056191 | SPTBN5;NRCAM;ANK2;ANK3;SPTAN1;SPTBN1;SPTBN2 |
| 937 | Homology Directed Repair Homo sapiens R-HSA-5693538 | 0.484357 | 0.724807 | 1.028927 | 0.7459032 | MUS81;RNF8;BABAM1;POLD4;HERC2;PPP4C;PPP4R2;UBB;UBC;UIMC1;SUMO2;NBN;TP53BP1;TOPBP1;ATRIP;UBE2I;RFC1;TOP3A;RPA2;SIRT6;SPIDR;CCNA1;RAD50;RPA3;RAD17;ATM;ATR |
| 938 | G2/M DNA damage checkpoint Homo sapiens R-HSA-69473 | 0.485699 | 0.724807 | 1.040345 | 0.7513019 | YWHAE;YWHAB;TOP3A;RNF8;RPA2;BABAM1;RAD50;HERC2;RPA3;UIMC1;RAD17;ATM;NBN;TOPBP1;TP53BP1;ATRIP;YWHAH;ATR |
| 939 | Transport of Mature mRNAs Derived from Intronless Transcripts Homo sapiens R-HSA-159234 | 0.486485 | 0.724807 | 1.076205 | 0.7754595 | RANBP2;NUP214;SLBP;NUP205;POM121C;NUP50;TPR;NUP98;AAAS |
| 940 | G1 Phase Homo sapiens R-HSA-69236 | 0.486485 | 0.724807 | 1.076205 | 0.7754595 | RB1;CCND3;CDK7;CDKN1B;UBB;UBC;CUL1;PPP2R2A;CKS1B |
| 941 | Cyclin D associated events in G1 Homo sapiens R-HSA-69231 | 0.486485 | 0.724807 | 1.076205 | 0.7754595 | RB1;CCND3;CDK7;CDKN1B;UBB;UBC;CUL1;PPP2R2A;CKS1B |
| 942 | Zinc efflux and compartmentalization by the SLC30 family Homo sapiens R-HSA-435368 | 0.487792 | 0.724807 | 1.387084 | 0.99574 | SLC30A6;SLC30A5 |
| 943 | Cation-coupled Chloride cotransporters Homo sapiens R-HSA-426117 | 0.487792 | 0.724807 | 1.387084 | 0.99574 | SLC12A6;SLC12A7 |
| 944 | Transport of nucleosides and free purine and pyrimidine bases across the plasma membrane Homo sapiens R-HSA-83936 | 0.487792 | 0.724807 | 1.387084 | 0.99574 | SLC29A4;SLC29A2 |

| | | | | | | |
|---|---|---|---|---|---|---|
| 945 | APEX1-Independent Resolution of AP Sites via the Single Nucleotide Replacement Pathway Homo sapiens R-HSA-5649702 | 0.487792 | 0.724807 | 1.387084 | 0.99574 | OGG1;NEIL1 |
| 946 | Vitamin D (calciferol) metabolism Homo sapiens R-HSA-196791 | 0.487792 | 0.724807 | 1.387084 | 0.99574 | CYP2R1;LGMN |
| 947 | Clearance of Nuclear Envelope Membranes from Chromatin Homo sapiens R-HSA-2993913 | 0.487792 | 0.724807 | 1.387084 | 0.99574 | LEMD2;TMPO |
| 948 | Nef Mediated CD8 Down-regulation Homo sapiens R-HSA-182218 | 0.487792 | 0.724807 | 1.387084 | 0.99574 | AP2A1;AP2M1 |
| 949 | Neurofascin interactions Homo sapiens R-HSA-447043 | 0.487792 | 0.724807 | 1.387084 | 0.99574 | SDCBP;NRCAM |
| 950 | Regulation of gene expression in late stage (branching morphogenesis) pancreatic bud precursor cells Homo sapiens R-HSA-210744 | 0.487792 | 0.724807 | 1.387084 | 0.99574 | HES1;RBPJ |
| 951 | Signaling by NOTCH1 t(7;9)(NOTCH1:M1580 K2555) Translocation Mutant Homo sapiens R-HSA-2660825 | 0.487792 | 0.724807 | 1.387084 | 0.99574 | ADAM17;ADAM10 |
| 952 | Constitutive Signaling by NOTCH1 t(7;9)(NOTCH1:M1580 K2555) Translocation Mutant Homo sapiens R-HSA-2660826 | 0.487792 | 0.724807 | 1.387084 | 0.99574 | ADAM17;ADAM10 |

| | | | | | | |
|---|---|---|---|---|---|---|
| 953 | Post-transcriptional silencing by small RNAs Homo sapiens R-HSA-426496 | 0.487792 | 0.724807 | 1.387084 | 0.99574 | AGO3;TNRC6A |
| 954 | CREB phosphorylation Homo sapiens R-HSA-199920 | 0.487792 | 0.724807 | 1.387084 | 0.99574 | RPS6KA5;CREB1 |
| 955 | Phase 3 - rapid repolarisation Homo sapiens R-HSA-5576890 | 0.487792 | 0.724807 | 1.387084 | 0.99574 | KCNQ1;AKAP9 |
| 956 | Glycerophospholipid catabolism Homo sapiens R-HSA-6814848 | 0.487792 | 0.724807 | 1.387084 | 0.99574 | GDE1;PNPLA6 |
| 957 | Apoptotic factor-mediated response Homo sapiens R-HSA-111471 | 0.487792 | 0.724807 | 1.387084 | 0.99574 | CASP7;XIAP |
| 958 | Nucleotide-binding domain, leucine rich repeat containing receptor (NLR) signaling pathways Homo sapiens R-HSA-168643 | 0.489938 | 0.726478 | 1.059592 | 0.7559939 | PYCARD;PANX1;APP;ITCH;HSP90AB1;CASP4;CASP2;SUGT1;UBE2V1;BIRC2;MAPK13 |
| 959 | Antigen activates B Cell Receptor (BCR) leading to generation of second messengers Homo sapiens R-HSA-983695 | 0.489938 | 0.726478 | 1.059592 | 0.7559939 | AHCYL1;STIM1;BLNK;ITPR2;ORAI1;ITPR3;GRB2;PLCG1;CBL;CALM1;SOS1 |
| 960 | PI3K events in ERBB2 signaling Homo sapiens R-HSA-1963642 | 0.496545 | 0.734743 | 1.155898 | 0.8092216 | ERBB3;ERBB2;GRB2;EGFR |
| 961 | GRB2 events in ERBB2 signaling Homo sapiens R-HSA-1963640 | 0.496545 | 0.734743 | 1.155898 | 0.8092216 | ERBB2;GRB2;SOS1;EGFR |
| 962 | Amino acid synthesis and interconversion (transamination) Homo sapiens R-HSA-70614 | 0.50033 | 0.738039 | 1.095057 | 0.7583132 | GLUD1;GOT1;GOT2;GLUL;ASPA;GLS |

| | | | | | | |
|---|---|---|---|---|---|---|
| 963 | Regulation of IFNA signaling Homo sapiens R-HSA-912694 | 0.50033 | 0.738039 | 1.095057 | 0.7583132 | IFNAR2;STAT1;STAT2;PTPN11;JAK1;IFNAR1 |
| 964 | Synthesis of IP3 and IP4 in the cytosol Homo sapiens R-HSA-1855204 | 0.50033 | 0.738039 | 1.095057 | 0.7583132 | ITPKB;ITPK1;INPP5J;OCRL;PLCG1;CALM1 |
| 965 | Transport of glucose and other sugars, bile salts and organic acids, metal ions and amine compounds Homo sapiens R-HSA-425366 | 0.501626 | 0.739184 | 1.022518 | 0.7054351 | SLC30A6;SLC14A2;SLC44A3;SLC22A5;SLC44A4;SLC44A1;SLC44A2;SLC31A1;SLC30A5;SLC6A13;SLC2A1;SLC6A12;SLC5A3;SLC6A6;SLC22A15;SLC22A18;SLC16A7;SLC39A7;RHCG;SLC39A1;RHBG;SLC39A4;SLC39A3 |
| 966 | Purine metabolism Homo sapiens R-HSA-73847 | 0.502817 | 0.740172 | 1.066974 | 0.7335758 | GMPR2;CAT;ADK;GMPS;GMPR;AMPD2;NT5C2;ADSS |
| 967 | RNA Polymerase I Transcription Homo sapiens R-HSA-73864 | 0.509921 | 0.749853 | 1.023224 | 0.6891403 | HDAC2;H2AFZ;HDAC1;H3F3A;MBD2;CHD4;CHD3;KAT2A;TAF1D;CDK7;RBBP4;TAF1B;POLR1D;POLR2E;RBBP7;POLR2H;HIST1H2AC;MAPK3 |
| 968 | Transmembrane transport of small molecules Homo sapiens R-HSA-382551 | 0.515155 | 0.756766 | 1.000331 | 0.6635076 | SLC35B3;TFRC;AQP6;SLC4A1;AQP2;AQP3;NIPAL1;NIPAL3;SLC16A7;SLC39A7;SLC12A6;SLC39A1;SLC12A7;SLC39A4;SLC39A3;NUP214;SLC34A3;SLC30A6;SLC38A1;ATP6V1G1;ATP6V0E1;SLC30A5;SLC6A13;ATP11C;SLC6A12;ATP1B3;ATP1B1;SLC9A4;ADCY9;SLC9A6;SCNN1A;NUP98;RHCG;ATP6V0D1;RAF1;SLC27A4;FTL;ATP6V1A;NUP205;SLC22A5;ABCB7;NEDD4L;CLCNKA;AAAS;SLC5A3;SLC9A1;POM121C;UBB;PRKAR2A;TPR;UBC;ATP6V0A4;TRPM7;SLC38A2;ATP6V1C1;ATP6V1C2;SLC14A2;ATP8B1;ABCA3;SLC33A1;SLC16A10;EIF2S2;GNG11;EIF2S1;SLC6A6;G6PC3;FXYD4;TRPV6;FXYD3;MYO5B;SLC26A7;CALM1;SLC44A3;SLC44A4;SLC44A1;SLC44A2;SLC2A1;ATP2A3;ATP2A2;TCIRG1;ABCC10;HK1;SLC22A15;SLC22A18;SLC37A4;ATP7A;ABCC3;ABCC4;ATP6V0B;ABCC1;ATP6AP1;ABCC5;ANO6;NIPA2;ASPH;SLCO4A1;PEX3;NUP50;SLC29A4;SGK2;ATP6V0C;SLC29A2;SRI;ATP1A1;ATP2C1;SLC7A1;ADCY6;ADD1;SLC25A26;GNG5;SLC17A5;RHBG;ATP9A;ABCF1;RANBP2;SLC35A1;DMTN;SLC35A3;SLC31A1;ATP13A1;WWP1;CTNS;MCOLN1;SLC8B1;TPCN1;CLCN7;SLC4A7;ABCB10;CLCN4;OSTM1;SLCO3A1;SLCO4C1;ATP13A2 |

| | | | | | | |
|---|---|---|---|---|---|---|
| 969 | Dual Incision in GG-NER Homo sapiens R-HSA-5696400 | 0.520979 | 0.760248 | 1.040264 | 0.6782993 | DDB1;POLD4;UBB;RFC1;RPA3;UBC;RPA2;ERCC5;CUL4B |
| 970 | tRNA modification in the nucleus and cytosol Homo sapiens R-HSA-6782315 | 0.520979 | 0.760248 | 1.040264 | 0.6782993 | QTRT1;TRMT10A;CTU1;URM1;NSUN2;TRMT6;TYW1;TRMT11;EPRS |
| 971 | Molecules associated with elastic fibres Homo sapiens R-HSA-2129379 | 0.521244 | 0.760248 | 1.055345 | 0.6875956 | ITGB1;EFEMP1;TGFB1;MFAP1;LTBP3;ITGB6;FBLN5 |
| 972 | Tight junction interactions Homo sapiens R-HSA-420029 | 0.521244 | 0.760248 | 1.055345 | 0.6875956 | PARD6B;PRKCI;CLDN4;CLDN8;CLDN7;F11R;CRB3 |
| 973 | Nef-mediates down modulation of cell surface receptors by recruiting them to clathrin adapters Homo sapiens R-HSA-164938 | 0.5221 | 0.760248 | 1.083618 | 0.7042398 | AP1G1;AP2A1;B2M;AP1M2;AP2M1 |
| 974 | Mitotic Spindle Checkpoint Homo sapiens R-HSA-69618 | 0.5221 | 0.760248 | 1.083618 | 0.7042398 | ANAPC16;UBE2E1;ANAPC5;ANAPC11;MAD2L1 |
| 975 | Synthesis of very long-chain fatty acyl-CoAs Homo sapiens R-HSA-75876 | 0.5221 | 0.760248 | 1.083618 | 0.7042398 | ELOVL1;TECR;ACSL5;ACSL3;HSD17B12 |
| 976 | Passive transport by Aquaporins Homo sapiens R-HSA-432047 | 0.524475 | 0.760248 | 1.155864 | 0.7459461 | AQP6;AQP2;AQP3 |
| 977 | Keratan sulfate degradation Homo sapiens R-HSA-2022857 | 0.524475 | 0.760248 | 1.155864 | 0.7459461 | HEXB;HEXA;GNS |
| 978 | mRNA decay by 3' to 5' exoribonuclease Homo sapiens R-HSA-429958 | 0.524475 | 0.760248 | 1.155864 | 0.7459461 | EXOSC7;NT5C3B;EXOSC8 |
| 979 | TP53 Regulates Transcription of Death Receptors and Ligands Homo sapiens R-HSA-6803211 | 0.524475 | 0.760248 | 1.155864 | 0.7459461 | PPP1R13B;TP53BP2;FAS |

| | | | | | | |
|---|---|---|---|---|---|---|
| 980 | Chk1/Chk2(Cds1) mediated inactivation of Cyclin B:Cdk1 complex Homo sapiens R-HSA-75035 | 0.524475 | 0.760248 | 1.155864 | 0.7459461 | YWHAE;YWHAB;YWHAH |
| 981 | Physiological factors Homo sapiens R-HSA-5578768 | 0.524475 | 0.760248 | 1.155864 | 0.7459461 | WWTR1;HIPK1;CORIN |
| 982 | GAB1 signalosome Homo sapiens R-HSA-180292 | 0.533478 | 0.771725 | 1.000869 | 0.6288829 | GSK3B;CDKN1B;SRC;PIK3R3;IRS2;EGFR;ERBB3;AKT2;ERBB2;MLST8;AKT1;THEM4;MAPK1;CSK;PIP4K2C;IER3;MAPK3;BAD;INSR;PPP2R5B;PTPN11;MTOR;CREB1;AGO3;KIT;RPS6KB2;GRB2;TNRC6A |
| 983 | PI3K/AKT activation Homo sapiens R-HSA-198203 | 0.533478 | 0.771725 | 1.000869 | 0.6288829 | GSK3B;CDKN1B;SRC;PIK3R3;IRS2;EGFR;ERBB3;AKT2;ERBB2;MLST8;AKT1;THEM4;MAPK1;PIP4K2C;IER3;MAPK3;BAD;INSR;PPP2R5B;PTPN11;RHOA;MTOR;CREB1;AGO3;KIT;RPS6KB2;GRB2;TNRC6A |
| 984 | GPVI-mediated activation cascade Homo sapiens R-HSA-114604 | 0.535723 | 0.773399 | 1.014853 | 0.6334079 | VAV3;CDC42;AKT2;IL3RA;AKT1;PIK3R3;PTPN11;GRB2;RAC1;JAK2;RHOA;JAK1 |
| 985 | Arachidonic acid metabolism Homo sapiens R-HSA-2142753 | 0.535723 | 0.773399 | 1.014853 | 0.6334079 | CBR1;FAAH;GPX2;ABCC1;CYP1A2;AKR1C3;CYP1A1;LTA4H;PTGR1;CYP4F11;PTGES;PTGS1 |
| 986 | Phospholipase C-mediated cascade; FGFR3 Homo sapiens R-HSA-5654227 | 0.537035 | 0.774506 | 1.019832 | 0.6340221 | AHCYL1;CREB1;ADCY9;PRKAR2A;PRKCD;ITPR2;ITPR3;PLCG1;CALM1;ADCY6 |
| 987 | Post NMDA receptor activation events Homo sapiens R-HSA-438064 | 0.539068 | 0.775866 | 1.02739 | 0.6348378 | CREB1;RRAS;RASGRF2;AKAP9;MAPK1;BRAF;RAF1;CALM1 |
| 988 | Bile acid and bile salt metabolism Homo sapiens R-HSA-194068 | 0.539068 | 0.775866 | 1.02739 | 0.6348378 | ABCC3;NCOA1;CYP39A1;NCOA2;SCP2;AKR1C1;AKR1C3;HSD17B4 |
| 989 | TAK1 activates NFkB by phosphorylation and activation of IKKs complex Homo sapiens R-HSA-445989 | 0.54243 | 0.777556 | 1.040237 | 0.6363092 | NFKBIA;APP;DHX9;HMGB1;RELA;NFKB1 |

| | | | | | | |
|---|---|---|---|---|---|---|
| 990 | TNFR1-induced NFkappaB signaling pathway Homo sapiens R-HSA-5357956 | 0.54243 | 0.777556 | 1.040237 | 0.6363092 | SHARPIN;XIAP;RIPK1;RBCK1;BIRC2;TNFRSF1A |
| 991 | CaM pathway Homo sapiens R-HSA-111997 | 0.54243 | 0.777556 | 1.040237 | 0.6363092 | CREB1;ADCY9;PRKAR2A;PRKCD;CALM1;ADCY6 |
| 992 | Calmodulin induced events Homo sapiens R-HSA-111933 | 0.54243 | 0.777556 | 1.040237 | 0.6363092 | CREB1;ADCY9;PRKAR2A;PRKCD;CALM1;ADCY6 |
| 993 | Trafficking of GluR2-containing AMPA receptors Homo sapiens R-HSA-416993 | 0.548725 | 0.783281 | 1.066914 | 0.6403174 | NSF;PICK1;AP2A1;AP2M1 |
| 994 | Ligand-dependent caspase activation Homo sapiens R-HSA-140534 | 0.548725 | 0.783281 | 1.066914 | 0.6403174 | FAS;RIPK1;CFLAR;TICAM1 |
| 995 | PKA-mediated phosphorylation of CREB Homo sapiens R-HSA-111931 | 0.548725 | 0.783281 | 1.066914 | 0.6403174 | ADCY9;CREB1;PRKAR2A;ADCY6 |
| 996 | Digestion of dietary lipid Homo sapiens R-HSA-192456 | 0.548725 | 0.783281 | 1.066914 | 0.6403174 | LIPC;LIPH;LIPG;LMF1 |
| 997 | Formation of the beta-catenin:TCF transactivating complex Homo sapiens R-HSA-201722 | 0.549178 | 0.783281 | 1.001663 | 0.6003297 | TLE3;TLE2;TRRAP;H2AFZ;HDAC1;H3F3A;CDC73;PYGO1;RUVBL1;EP300;CTNNB1;HIST1H2AC;MEN1 |
| 998 | PI-3K cascade:FGFR1 Homo sapiens R-HSA-5654689 | 0.562344 | 0.790933 | 0.985348 | 0.5672069 | GSK3B;CDKN1B;SRC;PIK3R3;IRS2;EGFR;ERBB3;AKT2;ERBB2;MLST8;AKT1;THEM4;MAPK1;PIP4K2C;IER3;MAPK3;BAD;INSR;PPP2R5B;PTPN11;MTOR;CREB1;AGO3;KIT;RPS6KB2;GRB2;TNRC6A |
| 999 | PI-3K cascade:FGFR3 Homo sapiens R-HSA-5654710 | 0.562344 | 0.790933 | 0.985348 | 0.5672069 | GSK3B;CDKN1B;SRC;PIK3R3;IRS2;EGFR;ERBB3;AKT2;ERBB2;MLST8;AKT1;THEM4;MAPK1;PIP4K2C;IER3;MAPK3;BAD;INSR;PPP2R5B;PTPN11;MTOR;CREB1;AGO3;KIT;RPS6KB2;GRB2;TNRC6A |
| 1000 | PI3K events in ERBB4 signaling Homo sapiens R-HSA-1250342 | 0.562344 | 0.790933 | 0.985348 | 0.5672069 | GSK3B;CDKN1B;SRC;PIK3R3;IRS2;EGFR;ERBB3;AKT2;ERBB2;MLST8;AKT1;THEM4;MAPK1;PIP4K2C;IER3;MAPK3;BAD;INSR;PPP2R5B;PTPN11;MTOR;CREB1;AGO3;KIT;RPS6KB2;GRB2;TNRC6A |

| | | | | | | |
|---|---|---|---|---|---|---|
| 1001 | PIP3 activates AKT signaling Homo sapiens R-HSA-1257604 | 0.562344 | 0.790933 | 0.985348 | 0.5672069 | GSK3B;CDKN1B;SRC;PIK3R3;IRS2;EGFR;ERBB3;AKT2;ERBB2;MLST8;AKT1;THEM4;MAPK1;PIP4K2C;IER3;MAPK3;BAD;INSR;PPP2R5B;PTPN11;MTOR;CREB1;AGO3;KIT;RPS6KB2;GRB2;TNRC6A |
| 1002 | PI-3K cascade:FGFR4 Homo sapiens R-HSA-5654720 | 0.562344 | 0.790933 | 0.985348 | 0.5672069 | GSK3B;CDKN1B;SRC;PIK3R3;IRS2;EGFR;ERBB3;AKT2;ERBB2;MLST8;AKT1;THEM4;MAPK1;PIP4K2C;IER3;MAPK3;BAD;INSR;PPP2R5B;PTPN11;MTOR;CREB1;AGO3;KIT;RPS6KB2;GRB2;TNRC6A |
| 1003 | PI-3K cascade:FGFR2 Homo sapiens R-HSA-5654695 | 0.562344 | 0.790933 | 0.985348 | 0.5672069 | GSK3B;CDKN1B;SRC;PIK3R3;IRS2;EGFR;ERBB3;AKT2;ERBB2;MLST8;AKT1;THEM4;MAPK1;PIP4K2C;IER3;MAPK3;BAD;INSR;PPP2R5B;PTPN11;MTOR;CREB1;AGO3;KIT;RPS6KB2;GRB2;TNRC6A |
| 1004 | Erythrocytes take up oxygen and release carbon dioxide Homo sapiens R-HSA-1247673 | 0.564489 | 0.790933 | 1.155829 | 0.6609426 | CA2;SLC4A1 |
| 1005 | Neurotransmitter Clearance In The Synaptic Cleft Homo sapiens R-HSA-112311 | 0.564489 | 0.790933 | 1.155829 | 0.6609426 | MAOA;COMT |
| 1006 | Release of Hh-Np from the secreting cell Homo sapiens R-HSA-5362798 | 0.564489 | 0.790933 | 1.155829 | 0.6609426 | SHH;ADAM17 |
| 1007 | Activation of PUMA and translocation to mitochondria Homo sapiens R-HSA-139915 | 0.564489 | 0.790933 | 1.155829 | 0.6609426 | PPP1R13B;TP53BP2 |
| 1008 | BH3-only proteins associate with and inactivate anti-apoptotic BCL-2 members Homo sapiens R-HSA-111453 | 0.564489 | 0.790933 | 1.155829 | 0.6609426 | BAD;BID |
| 1009 | Nef and signal transduction Homo sapiens R-HSA-164944 | 0.564489 | 0.790933 | 1.155829 | 0.6609426 | RAC1;PAK2 |

| | | | | | | |
|---|---|---|---|---|---|---|
| 1010 | Formyl peptide receptors bind formyl peptides and many other ligands Homo sapiens R-HSA-444473 | 0.564489 | 0.790933 | 1.155829 | 0.6609426 | HEBP1;APP |
| 1011 | Regulation of cytoskeletal remodeling and cell spreading by IPP complex components Homo sapiens R-HSA-446388 | 0.564489 | 0.790933 | 1.155829 | 0.6609426 | TESK1;LIMS1 |
| 1012 | Synthesis of PG Homo sapiens R-HSA-1483148 | 0.564489 | 0.790933 | 1.155829 | 0.6609426 | PTPMT1;PLD3 |
| 1013 | Synthesis of epoxy (EET) and dihydroxyeicosatrienoic acids (DHET) Homo sapiens R-HSA-2142670 | 0.564489 | 0.790933 | 1.155829 | 0.6609426 | CYP1A2;CYP1A1 |
| 1014 | tRNA modification in the mitochondrion Homo sapiens R-HSA-6787450 | 0.564489 | 0.790933 | 1.155829 | 0.6609426 | MTO1;TRMT61B |
| 1015 | Meiotic recombination Homo sapiens R-HSA-912446 | 0.564555 | 0.790933 | 0.990625 | 0.5663576 | PSMC3IP;RAD50;H2AFZ;RPA3;TOP3A;H3F3A;RPA2;ATM;NBN;MLH1;HIST1H2AC;MLH3 |
| 1016 | Resolution of AP sites via the multiple-nucleotide patch replacement pathway Homo sapiens R-HSA-110373 | 0.567407 | 0.793365 | 1.01981 | 0.5779047 | POLD4;RFC1;RPA3;APEX1;RPA2 |
| 1017 | Cyclin A/B1 associated events during G2/M transition Homo sapiens R-HSA-69273 | 0.567407 | 0.793365 | 1.01981 | 0.5779047 | CCNA1;CDK7;XPO1;PPP2R2A;CDC25B |

| | | | | | | |
|---|---|---|---|---|---|---|
| 1018 | Generic Transcription Pathway Homo sapiens R-HSA-212436 | 0.574464 | 0.802444 | 0.986993 | 0.5471069 | ZNF331;ATF2;CCNT2;EHMT1;RORA;CCNC;RBPJ;COX6A1;GLS;SESN3;AKT2;ZNF726;AKT1;EP300;ZNF205;ZNF567;NBN;ZNF720;ZNF563;MEN1;BNIP3L;SUPT16H;USP7;ZNF680;CSNK2A1;ZNF160;CSNK2A2;MED8;GTF2F2;NRBP1;COX6B1;RRAGA;SCO2;TBL1XR1;DDIT4;MAML3;ZNF552;ZNF430;ZNF793;COX19;ZNF274;MEAF6;COX16;NEDD4L;UBB;ZNF704;PRDX1;UBC;SMYD2;TP53BP2;COX14;ATRIP;FANCI;ESRRA;ZNF540;SMAD2;WWTR1;SURF1;PRELID1;SMAD4;ZNF141;SMAD3;NR1D2;FANCC;NR1D1;ESR1;NR2F6;SMAD7;CNOT4;CDK9;CDK7;RGCC;SNW1;TP53I3;AGO3;CDK5;BCL6;CNOT1;CNOT2;RAD17;ATM;CDK12;ZNF138;CDK13;CNOT8;ZNF257;COX20;ATR;ZNF254;YWHAE;ZNF770;BTG2;RARG;PPP1R13B;CDKN1B;THRB;THRA;YWHAB;CHD9;PHF20;PRKAG2;CHM;CHD4;CHD3;COX7C;MED13;AIFM2;MED10;MLST8;TNKS1BP1;CASP2;NELFB;TOPBP1;TEAD1;TEAD2;ZNF486;YWHAH;COX8A;NCOA1;NCOA2;GPX2;PRMT1;TOP3A;RABGGTA;PPM1A;CCNA1;KAT2A;NCOR1;RRM2B;PPARG;YAP1;ZNF595;PRKAA1;HDAC2;PRKAA2;HDAC1;COX5A;ZFP14;RBBP4;POLR2B;PERP;TGS1;POLR2C;POLR2E;RBBP7;POLR2H;BID;BRD7;ZNF224;POLR2J;NPM1;USP9X;NR1H2;TXNRD1;TAF11;GSR;RPA2;SSRP1;COX6C;HIPK1;MLH1;LRPPRC;MTOR;ZNF33B;ZNF33A;RAD50;CENPJ;RPA3;ZNF737;TCEA1;FAS;BAX;TAF5;LAMTOR1;Z[illegible] |
| 1019 | Effects of PIP2 hydrolysis Homo sapiens R-HSA-114508 | 0.582886 | 0.80833 | 0.990638 | 0.53471 | ABHD12;PRKCD;ITPR2;ITPR3;MGLL;DGKH |
| 1020 | Cytosolic iron-sulfur cluster assembly Homo sapiens R-HSA-2564830 | 0.583226 | 0.80833 | 1.04021 | 0.5608618 | ABCB7;CIAPIN1;NUBP2 |
| 1021 | Dissolution of Fibrin Clot Homo sapiens R-HSA-75205 | 0.583226 | 0.80833 | 1.04021 | 0.5608618 | ANXA2;SERPINB6;S100A10 |
| 1022 | Retrograde neurotrophin signalling Homo sapiens R-HSA-177504 | 0.583226 | 0.80833 | 1.04021 | 0.5608618 | AP2A1;AP2M1;DNM2 |

| | | | | | | |
|---|---|---|---|---|---|---|
| 1023 | POU5F1 (OCT4), SOX2, NANOG activate genes related to proliferation Homo sapiens R-HSA-2892247 | 0.583226 | 0.80833 | 1.04021 | 0.5608618 | STAT3;EPHA1;POU5F1 |
| 1024 | ERBB2 Activates PTK6 Signaling Homo sapiens R-HSA-8847993 | 0.583226 | 0.80833 | 1.04021 | 0.5608618 | ERBB3;ERBB2;EGFR |
| 1025 | p38MAPK events Homo sapiens R-HSA-171007 | 0.583226 | 0.80833 | 1.04021 | 0.5608618 | RALA;SRC;MAPK13 |
| 1026 | RHO GTPases Activate NADPH Oxidases Homo sapiens R-HSA-5668599 | 0.583226 | 0.80833 | 1.04021 | 0.5608618 | NOXA1;NOXO1;RAC1 |
| 1027 | Oncogene Induced Senescence Homo sapiens R-HSA-2559585 | 0.59626 | 0.824497 | 0.970792 | 0.5019764 | UBB;AGO3;UBC;MAPK1;ETS2;TNRC6A;MAPK3 |
| 1028 | TRAF6 mediated IRF7 activation Homo sapiens R-HSA-933541 | 0.59626 | 0.824497 | 0.970792 | 0.5019764 | TBK1;IRF3;DHX36;IRF2;IRF7;EP300;TANK |
| 1029 | Gene Silencing by RNA Homo sapiens R-HSA-211000 | 0.596629 | 0.824497 | 0.965401 | 0.4985901 | NUP214;TSNAX;RANBP2;NUP205;HSP90AA1;H2AFZ;H3F3A;AAAS;POM121C;AGO3;POLR2B;NUP50;POLR2C;TPR;POLR2E;DGCR8;NUP98;POLR2H;POLR2J;HIST1H2AC;RAN;TNRC6A |
| 1030 | Cytosolic sulfonation of small molecules Homo sapiens R-HSA-156584 | 0.597903 | 0.825454 | 0.990642 | 0.5095143 | SLC35B3;ABHD14B;BPNT1;PAPSS2 |
| 1031 | Phospholipase C-mediated cascade; FGFR4 Homo sapiens R-HSA-5654228 | 0.599159 | 0.826387 | 0.96305 | 0.493301 | AHCYL1;CREB1;ADCY9;PRKAR2A;PRKCD;ITPR2;ITPR3;PLCG1;CALM1;ADCY6 |
| 1032 | Cleavage of Growing Transcript in the Termination Region Homo sapiens R-HSA-109688 | 0.603285 | 0.828861 | 0.958915 | 0.4846032 | SLBP;EIF4A3;UPF3B;THOC2;SRRM1;DDX39A;DDX39B;U2AF2;PAPOLA;SRSF2;SRSF4;SLU7;SRSF11 |

| | | | | | | |
|---|---|---|---|---|---|---|
| 1033 | RNA Polymerase II Transcription Termination Homo sapiens R-HSA-73856 | 0.603285 | 0.828861 | 0.958915 | 0.4846032 | SLBP;EIF4A3;UPF3B;THOC2;SRRM1;DDX39A;DDX39B;U2AF2;PAPOLA;SRSF2;SRSF4;SLU7;SRSF11 |
| 1034 | Post-Elongation Processing of the Transcript Homo sapiens R-HSA-76044 | 0.603285 | 0.828861 | 0.958915 | 0.4846032 | SLBP;EIF4A3;UPF3B;THOC2;SRRM1;DDX39A;DDX39B;U2AF2;PAPOLA;SRSF2;SRSF4;SLU7;SRSF11 |
| 1035 | MAP kinase activation in TLR cascade Homo sapiens R-HSA-450294 | 0.603285 | 0.828861 | 0.958915 | 0.4846032 | ATF2;MAP2K1;FBXW11;CUL1;NFKB1;MAPK10;RPS6KA5;CREB1;UBB;TNIP2;UBC;MAPK1;MAPK3 |
| 1036 | Transport of Mature mRNA Derived from an Intronless Transcript Homo sapiens R-HSA-159231 | 0.608126 | 0.834706 | 0.956412 | 0.4756933 | RANBP2;NUP214;NUP205;POM121C;NUP50;TPR;NUP98;AAAS |
| 1037 | Acyl chain remodelling of PE Homo sapiens R-HSA-1482839 | 0.610386 | 0.837001 | 0.963091 | 0.4754426 | PNPLA8;LPCAT3;MBOAT1;MBOAT2;PLBD1 |
| 1038 | Retinoid metabolism and transport Homo sapiens R-HSA-975634 | 0.618852 | 0.847792 | 0.945512 | 0.45374 | SDC4;RDH11;SDC2;AKR1C1;AKR1C3;SDC1;LRP10;AGRN;HSPG2 |
| 1039 | Extension of Telomeres Homo sapiens R-HSA-180786 | 0.621423 | 0.850494 | 0.945548 | 0.4498386 | POLD4;RFC1;DKC1;RPA3;RUVBL1;RPA2 |
| 1040 | Ion channel transport Homo sapiens R-HSA-983712 | 0.623522 | 0.852546 | 0.959096 | 0.4530497 | ATP6V1A;ATP2A3;NEDD4L;CLCNKA;ATP2A2;TCIRG1;ATP1A1;SRI;ATP2C1;UBB;UBC;ATP6V0A4;TRPM7;ATP7A;ATP6V1C1;ATP9A;ATP6V1C2;ATP6V1G1;ATP6V0B;ATP6V0E1;ATP6AP1;ATP8B1;ATP13A1;WWP1;ATP11C;ANO6;ATP1B3;ATP1B1;MCOLN1;TPCN1;CLCN7;CLCN4;FXYD4;OSTM1;TRPV6;ASPH;FXYD3;SCNN1A;ATP13A2;ATP6V0D1;CALM1;RAF1;SGK2;ATP6V0C |
| 1041 | Phospholipase C-mediated cascade: FGFR1 Homo sapiens R-HSA-5654219 | 0.628679 | 0.856313 | 0.936961 | 0.4348767 | AHCYL1;CREB1;ADCY9;PRKAR2A;PRKCD;ITPR2;ITPR3;PLCG1;CALM1;ADCY6 |

| | | | | | | |
|---|---|---|---|---|---|---|
| 1042 | Sialic acid metabolism Homo sapiens R-HSA-4085001 | 0.631403 | 0.856313 | 0.933394 | 0.4291849 | SLC35A1;CMAS;NEU1;SLC17A5;ST3GAL6;NANS;GNE |
| 1043 | Bicarbonate transporters Homo sapiens R-HSA-425381 | 0.632522 | 0.856313 | 0.990646 | 0.4537558 | SLC4A7;SLC4A1 |
| 1044 | Nef Mediated CD4 Down-regulation Homo sapiens R-HSA-167590 | 0.632522 | 0.856313 | 0.990646 | 0.4537558 | AP2A1;AP2M1 |
| 1045 | SHC-related events triggered by IGF1R Homo sapiens R-HSA-2428933 | 0.632522 | 0.856313 | 0.990646 | 0.4537558 | GRB2;SOS1 |
| 1046 | Small interfering RNA (siRNA) biogenesis Homo sapiens R-HSA-426486 | 0.632522 | 0.856313 | 0.990646 | 0.4537558 | TSNAX;AGO3 |
| 1047 | CHL1 interactions Homo sapiens R-HSA-447041 | 0.632522 | 0.856313 | 0.990646 | 0.4537558 | ITGB1;HSPA8 |
| 1048 | Pregnenolone biosynthesis Homo sapiens R-HSA-196108 | 0.632522 | 0.856313 | 0.990646 | 0.4537558 | FDX1;AKR1B1 |
| 1049 | Synthesis of (16-20)-hydroxyeicosatetraenoic acids (HETE) Homo sapiens R-HSA-2142816 | 0.632522 | 0.856313 | 0.990646 | 0.4537558 | CYP1A2;CYP1A1 |
| 1050 | Pyrimidine salvage reactions Homo sapiens R-HSA-73614 | 0.632522 | 0.856313 | 0.990646 | 0.4537558 | TK2;TK1 |
| 1051 | Activation of anterior HOX genes in hindbrain development during early embryogenesis Homo sapiens R-HSA-5617472 | 0.633736 | 0.856313 | 0.940867 | 0.4291508 | SUZ12;RARG;H2AFZ;NCOA3;KMT2C;H3F3A;YY1;NCOR1;RBBP4;POLR2B;POLR2C;POLR2E;EP300;HOXB2;RBBP7;POLR2H;HOXD3;POLR2J;HIST1H2AC |
| 1052 | Activation of HOX genes during differentiation Homo sapiens R-HSA-5619507 | 0.633736 | 0.856313 | 0.940867 | 0.4291508 | SUZ12;RARG;H2AFZ;NCOA3;KMT2C;H3F3A;YY1;NCOR1;RBBP4;POLR2B;POLR2C;POLR2E;EP300;HOXB2;RBBP7;POLR2H;HOXD3;POLR2J;HIST1H2AC |

| | | | | | | |
|---|---|---|---|---|---|---|
| 1053 | Organic cation/anion/zwitterion transport Homo sapiens R-HSA-549132 | 0.637116 | 0.856313 | 0.945585 | 0.4262725 | SLC22A5;SLC22A15;SLC22A18 |
| 1054 | CS/DS degradation Homo sapiens R-HSA-2024101 | 0.637116 | 0.856313 | 0.945585 | 0.4262725 | HEXB;HEXA;ARSB |
| 1055 | Removal of the Flap Intermediate Homo sapiens R-HSA-69166 | 0.637116 | 0.856313 | 0.945585 | 0.4262725 | POLD4;RPA3;RPA2 |
| 1056 | SEMA3A-Plexin repulsion signaling by inhibiting Integrin adhesion Homo sapiens R-HSA-399955 | 0.637116 | 0.856313 | 0.945585 | 0.4262725 | RRAS;RAC1;TLN1 |
| 1057 | Activation of Rac Homo sapiens R-HSA-428540 | 0.637116 | 0.856313 | 0.945585 | 0.4262725 | RAC1;SOS1;PAK2 |
| 1058 | Synthesis of bile acids and bile salts via 24-hydroxycholesterol Homo sapiens R-HSA-193775 | 0.637116 | 0.856313 | 0.945585 | 0.4262725 | CYP39A1;AKR1C1;AKR1C3 |
| 1059 | Gastrin-CREB signalling pathway via PKC and MAPK Homo sapiens R-HSA-881907 | 0.642384 | 0.862577 | 0.963506 | 0.4264186 | APP;YWHAB;RASGRF2;ITGA2B;PEBP1;WDR83;IRS2;ARRB1;PSMD8;ABHD12;PSMD6;PSMD7;PSMD4;RASGEF1A;PSMD2;PSMD3;PSMD1;JAK2;SPTAN1;JAK1;MAP2K1;MAP2K2;PRKCD;PPP2R5B;PSMA5;PSMA6;PSMA3;CREB1;PSMA1;PSMA2;IL3RA;AKAP9;KIT;RASA2;AGTR1;PSME1;PSME2;SOS1;TLN1;RAF1;MGLL;VCL;DGKH;SHC2;SPTBN5;PSMD12;PSMD11;RGS19;SRC;PSMD13;CUL3;ITPR2;LPAR2;PIK3R3;ITPR3;IQGAP1;PHB;NTS;PSMA7;EGFR;PSMB10;PSMB6;RAP1B;PSMB4;PSMB5;PSMB2;UBB;GNG5;ERBB3;PSMB3;PSMB1;UBC;ERBB2;MAPK1;CSK;PSMF1;SPTBN1;SPTBN2;MAPK3;DAB2IP;BRAF;GNG11;PSMB8;PTK2;PSMB9;PSMC5;PSMC4;PTPRA;PSMC2;NF1;F2RL1;GRB2;CALM1;MAP3K11 |
| 1060 | GABA synthesis, release, reuptake and degradation Homo sapiens R-HSA-888590 | 0.643706 | 0.862724 | 0.92454 | 0.407272 | HSPA8;ALDH5A1;SLC6A13;SLC6A12 |

| | | | | | | |
|---|---|---|---|---|---|---|
| 1061 | Phosphorylation of the APC/C Homo sapiens R-HSA-176412 | 0.643706 | 0.862724 | 0.92454 | 0.407272 | ANAPC16;UBE2E1;ANAPC5;ANAPC11 |
| 1062 | Collagen formation Homo sapiens R-HSA-1474290 | 0.647118 | 0.86648 | 0.931231 | 0.4052963 | CRTAP;MMP7;CD151;COL24A1;COL14A1;COL12A1;LAMA3;PLOD2;CTSS;P4HA1;CTSL;P4HA2;COL4A4;COL5A2;ITGA6;P4HB;PPIB;CTSB |
| 1063 | Transport of inorganic cations/anions and amino acids/oligopeptides Homo sapiens R-HSA-425393 | 0.648944 | 0.866638 | 0.933184 | 0.4035167 | SLC34A3;SLC38A1;SLC6A12;CTNS;SLC4A1;SLC16A10;SRI;SLC7A1;SLC8B1;SLC9A1;SLC4A7;SLC6A6;SLC25A26;SLC9A4;SLC9A6;SLC17A5;SLC26A7;SLC12A6;CALM1;SLC12A7;SLC38A2 |
| 1064 | ERCC6 (CSB) and EHMT2 (G9a) positively regulate rRNA expression Homo sapiens R-HSA-427389 | 0.649064 | 0.866638 | 0.917643 | 0.3966266 | HDAC2;RBBP4;HDAC1;H2AFZ;H3F3A;CHD4;CHD3;RBBP7;HIST1H2AC |
| 1065 | Signaling by FGFR2 in disease Homo sapiens R-HSA-5655253 | 0.649064 | 0.866638 | 0.917643 | 0.3966266 | POLR2B;POLR2C;POLR2E;GRB2;PLCG1;POLR2H;SOS1;POLR2J;GTF2F2 |
| 1066 | G beta:gamma signalling through PI3Kgamma Homo sapiens R-HSA-392451 | 0.657015 | 0.876431 | 0.912245 | 0.3831868 | GNG5;AKT2;IL3RA;PIK3R3;AKT1;GRB2;JAK2;GNG11;RHOA;JAK1 |
| 1067 | PIWI-interacting RNA (piRNA) biogenesis Homo sapiens R-HSA-5601884 | 0.657834 | 0.876701 | 0.904379 | 0.3787559 | HSP90AA1;POLR2B;POLR2C;POLR2E;POLR2H;POLR2J |
| 1068 | Lipoprotein metabolism Homo sapiens R-HSA-174824 | 0.66474 | 0.884247 | 0.898765 | 0.3670195 | AMN;LIPC;SAR1B;SDC1;ZDHHC8;P4HB;HSPG2 |
| 1069 | Synthesis of PIPs at the plasma membrane Homo sapiens R-HSA-1660499 | 0.66474 | 0.884247 | 0.898765 | 0.3670195 | MTMR1;INPP5J;PIK3R3;PIK3C2G;PIP4K2C;MTMR6;PIK3C2B |
| 1070 | Cell junction organization Homo sapiens R-HSA-446728 | 0.66804 | 0.887807 | 0.917477 | 0.3701167 | ITGB1;VASP;PRKCI;CD151;LAMA3;TESK1;ILK;F11R;CLDN4;PARD6B;CDH1;CLDN8;CLDN7;CTNNB1;ITGA6;ANG;LIMS1;CRB3 |

| | | | | | | |
|---|---|---|---|---|---|---|
| 1071 | GABA B receptor activation Homo sapiens R-HSA-977444 | 0.671425 | 0.88981 | 0.894593 | 0.356364 | GABBR1;ADCY9;GNG5;GNAI3;KCNJ15;ADCY6;GNAI2;KCNJ3 |
| 1072 | Activation of GABAB receptors Homo sapiens R-HSA-991365 | 0.671425 | 0.88981 | 0.894593 | 0.356364 | GABBR1;ADCY9;GNG5;GNAI3;KCNJ15;ADCY6;GNAI2;KCNJ3 |
| 1073 | Activation of NMDA receptor upon glutamate binding and postsynaptic events Homo sapiens R-HSA-442755 | 0.671425 | 0.88981 | 0.894593 | 0.356364 | CREB1;RRAS;RASGRF2;AKAP9;MAPK1;BRAF;RAF1;CALM1 |
| 1074 | Integration of energy metabolism Homo sapiens R-HSA-163685 | 0.681047 | 0.89699 | 0.916206 | 0.3519369 | AHCYL1;PRKAA2;SLC2A1;TALDO1;ITPR2;PRKAG2;ITPR3;IQGAP1;ADIPOR1;ADIPOR2;ACACB;GNG11;ACACA;ADCY6;GNAI2;MARCKS;ADCY9;GNG5;PRKAR2A;MLX;TKT;SLC25A4;RAPGEF3 |
| 1075 | Phospholipase C-mediated cascade; FGFR2 Homo sapiens R-HSA-5654221 | 0.684083 | 0.89699 | 0.888797 | 0.3374549 | AHCYL1;ADCY9;CREB1;PRKAR2A;PRKCD;ITPR2;ITPR3;PLCG1;CALM1;ADCY6 |
| 1076 | N-Glycan antennae elongation Homo sapiens R-HSA-975577 | 0.685914 | 0.89699 | 0.86673 | 0.3267594 | B4GALT3;B4GALT1;MGAT4B |
| 1077 | Processive synthesis on the lagging strand Homo sapiens R-HSA-69183 | 0.685914 | 0.89699 | 0.86673 | 0.3267594 | POLD4;RPA3;RPA2 |
| 1078 | Association of licensing factors with the pre-replicative complex Homo sapiens R-HSA-69298 | 0.685914 | 0.89699 | 0.86673 | 0.3267594 | UBB;GMNN;UBC |
| 1079 | Prolactin receptor signaling Homo sapiens R-HSA-1170546 | 0.685914 | 0.89699 | 0.86673 | 0.3267594 | CUL1;PTPN11;JAK2 |
| 1080 | CREB phosphorylation through the activation of CaMKII Homo sapiens R-HSA-442729 | 0.685914 | 0.89699 | 0.86673 | 0.3267594 | CREB1;AKAP9;CALM1 |

| | | | | | | |
|---|---|---|---|---|---|---|
| 1081 | Recycling of bile acids and salts Homo sapiens R-HSA-159418 | 0.685914 | 0.89699 | 0.86673 | 0.3267594 | ABCC3;NCOA1;NCOA2 |
| 1082 | Lagging Strand Synthesis Homo sapiens R-HSA-69186 | 0.685921 | 0.89699 | 0.8667 | 0.3267396 | POLD4;RFC1;RPA3;RPA2 |
| 1083 | Activation of PPARGC1A (PGC-1alpha) by phosphorylation Homo sapiens R-HSA-2151209 | 0.691926 | 0.89699 | 0.86676 | 0.3192065 | PRKAA2;PRKAG2 |
| 1084 | POU5F1 (OCT4), SOX2, NANOG repress genes related to differentiation Homo sapiens R-HSA-2892245 | 0.691926 | 0.89699 | 0.86676 | 0.3192065 | TSC22D1;POU5F1 |
| 1085 | HDR through MMEJ (alt-NHEJ) Homo sapiens R-HSA-5685939 | 0.691926 | 0.89699 | 0.86676 | 0.3192065 | RAD50;NBN |
| 1086 | IRAK2 mediated activation of TAK1 complex upon TLR7/8 or 9 stimulation Homo sapiens R-HSA-975163 | 0.691926 | 0.89699 | 0.86676 | 0.3192065 | UBB;UBC |
| 1087 | IRAK2 mediated activation of TAK1 complex Homo sapiens R-HSA-937042 | 0.691926 | 0.89699 | 0.86676 | 0.3192065 | UBB;UBC |
| 1088 | Signaling by FGFR3 fusions in cancer Homo sapiens R-HSA-8853334 | 0.691926 | 0.89699 | 0.86676 | 0.3192065 | GRB2;SOS1 |
| 1089 | TRIF-mediated programmed cell death Homo sapiens R-HSA-2562578 | 0.691926 | 0.89699 | 0.86676 | 0.3192065 | RIPK1;TICAM1 |
| 1090 | GP1b-IX-V activation signalling Homo sapiens R-HSA-430116 | 0.691926 | 0.89699 | 0.86676 | 0.3192065 | SRC;RAF1 |

| | | | | | | |
|---|---|---|---|---|---|---|
| 1091 | Adenylate cyclase activating pathway Homo sapiens R-HSA-170660 | 0.691926 | 0.89699 | 0.86676 | 0.3192065 | ADCY9;ADCY6 |
| 1092 | Synthesis of pyrophosphates in the cytosol Homo sapiens R-HSA-1855167 | 0.691926 | 0.89699 | 0.86676 | 0.3192065 | PPIP5K2;ITPK1 |
| 1093 | Regulation of signaling by NODAL Homo sapiens R-HSA-1433617 | 0.691926 | 0.89699 | 0.86676 | 0.3192065 | ACVR1B;ACVR2B |
| 1094 | Purine catabolism Homo sapiens R-HSA-74259 | 0.691926 | 0.89699 | 0.86676 | 0.3192065 | CAT;NT5C2 |
| 1095 | Urea cycle Homo sapiens R-HSA-70635 | 0.691926 | 0.89699 | 0.86676 | 0.3192065 | ARG2;NMRAL1 |
| 1096 | RNA Polymerase I Promoter Escape Homo sapiens R-HSA-73772 | 0.691982 | 0.89699 | 0.866641 | 0.3190933 | TAF1D;CDK7;TAF1B;POLR2E;POLR1D;POLR2H |
| 1097 | MAPK targets/ Nuclear events mediated by MAP kinases Homo sapiens R-HSA-450282 | 0.691982 | 0.89699 | 0.866641 | 0.3190933 | MAPK10;ATF2;RPS6KA5;CREB1;MAPK1;MAPK3 |
| 1098 | Phase 1 - Functionalization of compounds Homo sapiens R-HSA-211945 | 0.694209 | 0.899058 | 0.902026 | 0.3292232 | NCOA1;NCOA2;AOC1;MAOA;EPHX1;CYP51A1;CMBL;CYP4F11;BPHL;ADH5;PTGS1;CYP17A1;CYP39A1;POR;CYP2R1;FDX1;CYP1A2;CYP1A1;CES2 |
| 1099 | PRC2 methylates histones and DNA Homo sapiens R-HSA-212300 | 0.700615 | 0.905266 | 0.866581 | 0.3083261 | SUZ12;DNMT1;RBBP4;H2AFZ;PHF1;H3F3A;RBBP7;HIST1H2AC |
| 1100 | ECM proteoglycans Homo sapiens R-HSA-3000178 | 0.714418 | 0.905266 | 0.866491 | 0.2913895 | ITGB1;TGFB1;LAMB2;ITGA2B;LAMA3;LAMB1;LAMC1;ITGB6;AGRN;MATN3;ITGA9 |
| 1101 | Sperm:Oocyte Membrane Binding Homo sapiens R-HSA-1300652 | 0.718378 | 0.905266 | 0.86679 | 0.286699 | CD9 |
| 1102 | Toxicity of botulinum toxin type D (BoNT/D) Homo sapiens R-HSA-5250955 | 0.718378 | 0.905266 | 0.86679 | 0.286699 | VAMP1 |

| | | | | | | |
|---|---|---|---|---|---|---|
| 1103 | Toxicity of botulinum toxin type F (BoNT/F) Homo sapiens R-HSA-5250981 | 0.718378 | 0.905266 | 0.86679 | 0.286699 | VAMP1 |
| 1104 | Abacavir metabolism Homo sapiens R-HSA-2161541 | 0.718378 | 0.905266 | 0.86679 | 0.286699 | NT5C2 |
| 1105 | Na+-dependent glucose transporters Homo sapiens R-HSA-428808 | 0.718378 | 0.905266 | 0.86679 | 0.286699 | SLC5A3 |
| 1106 | Sodium-coupled phosphate cotransporters Homo sapiens R-HSA-427652 | 0.718378 | 0.905266 | 0.86679 | 0.286699 | SLC34A3 |
| 1107 | Hyaluronan biosynthesis and export Homo sapiens R-HSA-2142850 | 0.718378 | 0.905266 | 0.86679 | 0.286699 | ABCC5 |
| 1108 | Negative regulation of TCF-dependent signaling by DVL-interacting proteins Homo sapiens R-HSA-5368598 | 0.718378 | 0.905266 | 0.86679 | 0.286699 | DVL3 |
| 1109 | Electric Transmission Across Gap Junctions Homo sapiens R-HSA-112303 | 0.718378 | 0.905266 | 0.86679 | 0.286699 | PANX1 |
| 1110 | Transmission across Electrical Synapses Homo sapiens R-HSA-112307 | 0.718378 | 0.905266 | 0.86679 | 0.286699 | PANX1 |
| 1111 | Synthesis of GDP-mannose Homo sapiens R-HSA-446205 | 0.718378 | 0.905266 | 0.86679 | 0.286699 | GMPPB |
| 1112 | G2 Phase Homo sapiens R-HSA-68911 | 0.718378 | 0.905266 | 0.86679 | 0.286699 | CCNA1 |

| | | | | | | |
|---|---|---|---|---|---|---|
| 1113 | Loss of Function of FBXW7 in Cancer and NOTCH1 Signaling Homo sapiens R-HSA-2644607 | 0.718378 | 0.905266 | 0.86679 | 0.286699 | CUL1 |
| 1114 | FBXW7 Mutants and NOTCH1 in Cancer Homo sapiens R-HSA-2644605 | 0.718378 | 0.905266 | 0.86679 | 0.286699 | CUL1 |
| 1115 | PTK6 Expression Homo sapiens R-HSA-8849473 | 0.718378 | 0.905266 | 0.86679 | 0.286699 | HIF1A |
| 1116 | FasL/ CD95L signaling Homo sapiens R-HSA-75157 | 0.718378 | 0.905266 | 0.86679 | 0.286699 | FAS |
| 1117 | Cyclin B2 mediated events Homo sapiens R-HSA-157881 | 0.718378 | 0.905266 | 0.86679 | 0.286699 | CDC25B |
| 1118 | Scavenging by Class B Receptors Homo sapiens R-HSA-3000471 | 0.718378 | 0.905266 | 0.86679 | 0.286699 | CD36 |
| 1119 | Cytochrome c-mediated apoptotic response Homo sapiens R-HSA-111461 | 0.718378 | 0.905266 | 0.86679 | 0.286699 | CASP7 |
| 1120 | Activation of caspases through apoptosome-mediated cleavage Homo sapiens R-HSA-111459 | 0.718378 | 0.905266 | 0.86679 | 0.286699 | CASP7 |
| 1121 | Vitamin B2 (riboflavin) metabolism Homo sapiens R-HSA-196843 | 0.718378 | 0.905266 | 0.86679 | 0.286699 | SLC52A3 |
| 1122 | Sulfide oxidation to sulfate Homo sapiens R-HSA-1614517 | 0.718378 | 0.905266 | 0.86679 | 0.286699 | TST |
| 1123 | Estrogen biosynthesis Homo sapiens R-HSA-193144 | 0.718378 | 0.905266 | 0.86679 | 0.286699 | HSD17B11 |

| | | | | | | |
|---|---|---|---|---|---|---|
| 1124 | The fatty acid cycling model Homo sapiens R-HSA-167826 | 0.718378 | 0.905266 | 0.86679 | 0.286699 | UCP2 |
| 1125 | The proton buffering model Homo sapiens R-HSA-167827 | 0.718378 | 0.905266 | 0.86679 | 0.286699 | UCP2 |
| 1126 | Mitochondrial Uncoupling Proteins Homo sapiens R-HSA-166187 | 0.718378 | 0.905266 | 0.86679 | 0.286699 | UCP2 |
| 1127 | tRNA processing in the mitochondrion Homo sapiens R-HSA-6785470 | 0.718378 | 0.905266 | 0.86679 | 0.286699 | ELAC2 |
| 1128 | Trafficking of myristoylated proteins to the cilium Homo sapiens R-HSA-5624138 | 0.718378 | 0.905266 | 0.86679 | 0.286699 | ARL3 |
| 1129 | PI Metabolism Homo sapiens R-HSA-1483255 | 0.71897 | 0.905266 | 0.866461 | 0.2858762 | MTMR1;INPP5F;PITPNB;GDE1;INPP5J;PIK3R3;PIK3C2G;OCRL;PIP4K2C;PNPLA6;MTMR6;PIK3C2B |
| 1130 | Diseases associated with glycosaminoglycan metabolism Homo sapiens R-HSA-3560782 | 0.723126 | 0.905266 | 0.825347 | 0.2675538 | SDC4;SDC2;SDC1;AGRN;HSPG2 |
| 1131 | WNT ligand biogenesis and trafficking Homo sapiens R-HSA-3238698 | 0.723126 | 0.905266 | 0.825347 | 0.2675538 | WNT5A;VPS26A;VPS35;TMED5;WLS |
| 1132 | Regulation of Complement cascade Homo sapiens R-HSA-977606 | 0.723126 | 0.905266 | 0.825347 | 0.2675538 | C9;CD59;CD46;CD55;CFB |
| 1133 | Amino acid transport across the plasma membrane Homo sapiens R-HSA-352230 | 0.723785 | 0.905266 | 0.831921 | 0.2689281 | SLC6A6;SLC38A1;SLC6A12;SLC16A10;SLC7A1;SLC38A2 |
| 1134 | Trafficking of AMPA receptors Homo sapiens R-HSA-399719 | 0.723785 | 0.905266 | 0.831921 | 0.2689281 | NSF;DLG1;MYO6;PICK1;AP2A1;AP2M1 |

| | | | | | | |
|---|---|---|---|---|---|---|
| 1135 | Glutamate Binding, Activation of AMPA Receptors and Synaptic Plasticity Homo sapiens R-HSA-399721 | 0.723785 | 0.905266 | 0.831921 | 0.2689281 | NSF;DLG1;MYO6;PICK1;AP2A1;AP2M1 |
| 1136 | NOD1/2 Signaling Pathway Homo sapiens R-HSA-168638 | 0.723785 | 0.905266 | 0.831921 | 0.2689281 | ITCH;CASP4;CASP2;UBE2V1;BIRC2;MAPK13 |
| 1137 | Synthesis of Leukotrienes (LT) and Eoxins (EX) Homo sapiens R-HSA-2142691 | 0.724467 | 0.905266 | 0.815665 | 0.262904 | ABCC1;LTA4H;CYP4F11;PTGR1 |
| 1138 | Aflatoxin activation and detoxification Homo sapiens R-HSA-5423646 | 0.724467 | 0.905266 | 0.815665 | 0.262904 | GGT6;MGST3;CYP1A2;MGST2 |
| 1139 | Elastic fibre formation Homo sapiens R-HSA-1566948 | 0.728076 | 0.90771 | 0.840267 | 0.2666585 | ITGB1;EFEMP1;TGFB1;MFAP1;FURIN;LTBP3;ITGB6;FBLN5 |
| 1140 | Condensation of Prophase Chromosomes Homo sapiens R-HSA-2299718 | 0.728076 | 0.90771 | 0.840267 | 0.2666585 | RB1;SET;H2AFZ;H3F3A;NCAPD3;HIST1H2AC;SMC2;MCPH1 |
| 1141 | TRAF6 mediated induction of TAK1 complex Homo sapiens R-HSA-937072 | 0.729615 | 0.90771 | 0.800007 | 0.2521931 | UBB;UBC;TICAM1 |
| 1142 | CRMPs in Sema3A signaling Homo sapiens R-HSA-399956 | 0.729615 | 0.90771 | 0.800007 | 0.2521931 | GSK3B;CDK5;DPYSL2 |
| 1143 | PKA activation Homo sapiens R-HSA-163615 | 0.729615 | 0.90771 | 0.800007 | 0.2521931 | ADCY9;PRKAR2A;ADCY6 |
| 1144 | Neurotransmitter Release Cycle Homo sapiens R-HSA-112310 | 0.734173 | 0.911785 | 0.845332 | 0.2612166 | HSPA8;ALDH5A1;MAOA;PPFIA1;ARL6IP5;SLC6A13;SLC6A12;LIN7A;SLC38A2;GLS |
| 1145 | G-protein beta:gamma signalling Homo sapiens R-HSA-397795 | 0.734173 | 0.911785 | 0.845332 | 0.2612166 | GNG5;AKT2;IL3RA;AKT1;PIK3R3;GRB2;JAK2;GNG11;RHOA;JAK1 |

| | | | | | | |
|---|---|---|---|---|---|---|
| 1146 | DSCAM interactions Homo sapiens R-HSA-376172 | 0.743155 | 0.918129 | 0.770403 | 0.2286945 | RAC1;MAPK13 |
| 1147 | MyD88 deficiency (TLR2/4) Homo sapiens R-HSA-5602498 | 0.743155 | 0.918129 | 0.770403 | 0.2286945 | IRAK4;CD36 |
| 1148 | IRAK4 deficiency (TLR2/4) Homo sapiens R-HSA-5603041 | 0.743155 | 0.918129 | 0.770403 | 0.2286945 | IRAK4;CD36 |
| 1149 | Interleukin-7 signaling Homo sapiens R-HSA-1266695 | 0.743155 | 0.918129 | 0.770403 | 0.2286945 | PIK3R3;JAK1 |
| 1150 | Androgen biosynthesis Homo sapiens R-HSA-193048 | 0.743155 | 0.918129 | 0.770403 | 0.2286945 | HSD17B12;CYP17A1 |
| 1151 | Phenylalanine and tyrosine catabolism Homo sapiens R-HSA-71182 | 0.743155 | 0.918129 | 0.770403 | 0.2286945 | HGD;PCBD1 |
| 1152 | Negative regulation of the PI3K/AKT network Homo sapiens R-HSA-199418 | 0.744586 | 0.919099 | 0.866282 | 0.2554895 | SRC;INSR;PPP2R5B;PIK3R3;IRS2;PTPN11;EGFR;ERBB3;AKT2;KIT;ERBB2;AKT1;THEM4;MAPK1;GRB2;PIP4K2C;IER3;MAPK3 |
| 1153 | Phase II conjugation Homo sapiens R-HSA-156580 | 0.752412 | 0.925525 | 0.866222 | 0.2464147 | SLC35B3;GSTK1;GGT6;GSTO2;GSTO1;MGST3;GSTP1;AKR1A1;MGST2;ABHD14B;MTR;COMT;PAPSS2;CNDP2;GGCT;GCLC;UGP2;ESD;CYP1A2;BPNT1 |
| 1154 | Steroid hormones Homo sapiens R-HSA-209943 | 0.752993 | 0.925525 | 0.808732 | 0.2294363 | CYP2R1;FDX1;AKR1B1;HSD17B12;HSD17B11;CYP17A1;LGMN |
| 1155 | RNA Polymerase I Transcription Termination Homo sapiens R-HSA-73863 | 0.753213 | 0.925525 | 0.799873 | 0.2266892 | TAF1D;CDK7;TAF1B;POLR2E;POLR1D;POLR2H |
| 1156 | Resolution of D-loop Structures through Holliday Junction Intermediates Homo sapiens R-HSA-5693568 | 0.753213 | 0.925525 | 0.799873 | 0.2266892 | MUS81;RAD50;TOP3A;ATM;NBN;SPIDR |

| | | | | | | |
|---|---|---|---|---|---|---|
| 1157 | Thrombin signalling through proteinase activated receptors (PARs) Homo sapiens R-HSA-456926 | 0.753213 | 0.925525 | 0.799873 | 0.2266892 | GNG5;SRC;MAPK1;ARRB1;GNG11;MAPK3 |
| 1158 | Glucagon-like Peptide-1 (GLP1) regulates insulin secretion Homo sapiens R-HSA-381676 | 0.753776 | 0.925525 | 0.8155 | 0.2305097 | GNG5;PRKAR2A;ITPR2;ITPR3;IQGAP1;GNG11;ADCY6;RAPGEF3 |
| 1159 | Peptide hormone metabolism Homo sapiens R-HSA-2980736 | 0.754947 | 0.925525 | 0.852958 | 0.2397736 | SLC30A6;EXOC7;SLC30A5;RAB27A;SPCS3;SPCS2;SPCS1;KIF5B;EXOC4;CTNNB1;EXOC6;EXOC5;CTSD;SEC11A;EXOC2;SEC11C |
| 1160 | Keratan sulfate biosynthesis Homo sapiens R-HSA-2022854 | 0.754999 | 0.925525 | 0.787781 | 0.2213968 | B4GALT3;B4GALT1;B3GNT3;B3GNT2;ST3GAL6 |
| 1161 | Deposition of new CENPA-containing nucleosomes at the centromere Homo sapiens R-HSA-606279 | 0.757125 | 0.926092 | 0.825152 | 0.2295789 | CENPT;NPM1;RBBP4;H2AFZ;RUVBL1;SMARCA5;RSF1;RBBP7;HIST1H2AC;CENPC |
| 1162 | Nucleosome assembly Homo sapiens R-HSA-774815 | 0.757125 | 0.926092 | 0.825152 | 0.2295789 | CENPT;NPM1;RBBP4;H2AFZ;RUVBL1;SMARCA5;RSF1;RBBP7;HIST1H2AC;CENPC |
| 1163 | Chromosome Maintenance Homo sapiens R-HSA-73886 | 0.758402 | 0.926092 | 0.853701 | 0.2360839 | CENPT;NPM1;RFC1;H2AFZ;SMARCA5;RPA2;RSF1;TERF1;CENPC;POLD4;RBBP4;DKC1;RPA3;RUVBL1;TERF2IP;RBBP7;HIST1H2AC |
| 1164 | PI3K/AKT Signaling in Cancer Homo sapiens R-HSA-2219528 | 0.758402 | 0.926092 | 0.853701 | 0.2360839 | GSK3B;CDKN1B;BAD;PIK3R3;IRS2;PTPN11;EGFR;MTOR;CREB1;ERBB3;AKT2;KIT;ERBB2;RPS6KB2;MLST8;AKT1;GRB2 |
| 1165 | Telomere C-strand (Lagging Strand) Synthesis Homo sapiens R-HSA-174417 | 0.75937 | 0.926092 | 0.770301 | 0.212038 | POLD4;RFC1;RPA3;RPA2 |
| 1166 | ADP signalling through P2Y purinoceptor 12 Homo sapiens R-HSA-392170 | 0.75937 | 0.926092 | 0.770301 | 0.212038 | GNG5;GNAI3;GNG11;GNAI2 |

| | | | | | | |
|---|---|---|---|---|---|---|
| 1167 | Integrin cell surface interactions Homo sapiens R-HSA-216083 | 0.76436 | 0.930653 | 0.834234 | 0.2241717 | ITGB1;ITGA3;ITGA2B;ITGAE;F11R;HSPG2;CDH1;COL4A4;ITGA6;ITGB6;CD47;AGRN;ITGA9 |
| 1168 | Ras activation uopn Ca2+ infux through NMDA receptor Homo sapiens R-HSA-442982 | 0.768377 | 0.930653 | 0.742816 | 0.1957133 | RASGRF2;AKAP9;CALM1 |
| 1169 | PKA activation in glucagon signalling Homo sapiens R-HSA-164378 | 0.768377 | 0.930653 | 0.742816 | 0.1957133 | ADCY9;PRKAR2A;ADCY6 |
| 1170 | Metabolism of porphyrins Homo sapiens R-HSA-189445 | 0.768377 | 0.930653 | 0.742816 | 0.1957133 | UROS;BLVRB;BLVRA |
| 1171 | Cell surface interactions at the vascular wall Homo sapiens R-HSA-202733 | 0.768519 | 0.930653 | 0.855472 | 0.2252367 | ITGB1;GRB7;CD74;CXADR;YES1;PPIL2;ITGA3;SRC;ATP1B3;PTPN11;F11R;ATP1B1;GRB14;GRB2;ITGA6;PLCG1;CD47;CD58;SOS1;PPIA |
| 1172 | Vasopressin regulates renal water homeostasis via Aquaporins Homo sapiens R-HSA-432040 | 0.77771 | 0.930653 | 0.792149 | 0.199148 | ADCY9;GNG5;PRKAR2A;MYO5B;AQP2;AQP3;GNG11;ADCY6 |
| 1173 | Resolution of D-Loop Structures Homo sapiens R-HSA-5693537 | 0.780282 | 0.930653 | 0.770198 | 0.1910856 | MUS81;RAD50;TOP3A;ATM;NBN;SPIDR |
| 1174 | FGFR2 mutant receptor activation Homo sapiens R-HSA-1839126 | 0.780282 | 0.930653 | 0.770198 | 0.1910856 | POLR2B;POLR2C;POLR2E;POLR2H;POLR2J;GTF2F2 |
| 1175 | Inactivation, recovery and regulation of the phototransduction cascade Homo sapiens R-HSA-2514859 | 0.780282 | 0.930653 | 0.770198 | 0.1910856 | FNTA;CNGA1;NMT1;CALM1;CAMKMT;METAP2 |
| 1176 | Fructose metabolism Homo sapiens R-HSA-5652084 | 0.781434 | 0.930653 | 0.693387 | 0.1710059 | AKR1B1 |
| 1177 | Vitamins Homo sapiens R-HSA-211916 | 0.781434 | 0.930653 | 0.693387 | 0.1710059 | CYP2R1 |

| | | | | | | |
|---|---|---|---|---|---|---|
| 1178 | The retinoid cycle in cones (daylight vision) Homo sapiens R-HSA-2187335 | 0.781434 | 0.930653 | 0.693387 | 0.1710059 | DHRS3 |
| 1179 | Synthesis of Dolichyl-phosphate Homo sapiens R-HSA-446199 | 0.781434 | 0.930653 | 0.693387 | 0.1710059 | DOLPP1 |
| 1180 | PTK6 Regulates Cell Cycle Homo sapiens R-HSA-8849470 | 0.781434 | 0.930653 | 0.693387 | 0.1710059 | CDKN1B |
| 1181 | Pre-NOTCH Processing in the Endoplasmic Reticulum Homo sapiens R-HSA-1912399 | 0.781434 | 0.930653 | 0.693387 | 0.1710059 | POGLUT1 |
| 1182 | TNFR1-mediated ceramide production Homo sapiens R-HSA-5626978 | 0.781434 | 0.930653 | 0.693387 | 0.1710059 | TNFRSF1A |
| 1183 | Phosphorylation of Emi1 Homo sapiens R-HSA-176417 | 0.781434 | 0.930653 | 0.693387 | 0.1710059 | FZR1 |
| 1184 | Fibronectin matrix formation Homo sapiens R-HSA-1566977 | 0.781434 | 0.930653 | 0.693387 | 0.1710059 | ITGB1 |
| 1185 | Proton-coupled monocarboxylate transport Homo sapiens R-HSA-433692 | 0.781434 | 0.930653 | 0.693387 | 0.1710059 | SLC16A7 |
| 1186 | Activation of C3 and C5 Homo sapiens R-HSA-174577 | 0.781434 | 0.930653 | 0.693387 | 0.1710059 | CFB |
| 1187 | RSK activation Homo sapiens R-HSA-444257 | 0.781434 | 0.930653 | 0.693387 | 0.1710059 | MAPK1 |
| 1188 | Metabolic disorders of biological oxidation enzymes Homo sapiens R-HSA-5579029 | 0.781434 | 0.930653 | 0.693387 | 0.1710059 | FDX1 |
| 1189 | Synthesis of Lipoxins (LX) Homo sapiens R-HSA-2142700 | 0.781434 | 0.930653 | 0.693387 | 0.1710059 | PTGR1 |

| | | | | | | |
|---|---|---|---|---|---|---|
| 1190 | Synthesis of 5-eicosatetraenoic acids Homo sapiens R-HSA-2142688 | 0.781434 | 0.930653 | 0.693387 | 0.1710059 | GPX2 |
| 1191 | Synthesis of 15-eicosatetraenoic acid derivatives Homo sapiens R-HSA-2142770 | 0.781434 | 0.930653 | 0.693387 | 0.1710059 | GPX2 |
| 1192 | Synthesis of 12-eicosatetraenoic acid derivatives Homo sapiens R-HSA-2142712 | 0.781434 | 0.930653 | 0.693387 | 0.1710059 | GPX2 |
| 1193 | Pyrimidine biosynthesis Homo sapiens R-HSA-500753 | 0.781434 | 0.930653 | 0.693387 | 0.1710059 | DUT |
| 1194 | Synthesis of wybutosine at G37 of tRNA(Phe) Homo sapiens R-HSA-6782861 | 0.781434 | 0.930653 | 0.693387 | 0.1710059 | TYW1 |
| 1195 | Reversible hydration of carbon dioxide Homo sapiens R-HSA-1475029 | 0.786891 | 0.934023 | 0.693318 | 0.1661642 | CA5B;CA2 |
| 1196 | TP53 Regulates Transcription of Caspase Activators and Caspases Homo sapiens R-HSA-6803207 | 0.786891 | 0.934023 | 0.693318 | 0.1661642 | CASP2;ATM |
| 1197 | ERKs are inactivated Homo sapiens R-HSA-202670 | 0.786891 | 0.934023 | 0.693318 | 0.1661642 | MAPK1;MAPK3 |
| 1198 | Purine ribonucleoside monophosphate biosynthesis Homo sapiens R-HSA-73817 | 0.786891 | 0.934023 | 0.693318 | 0.1661642 | GMPS;ADSS |
| 1199 | PI5P, PP2A and IER3 Regulate PI3K/AKT Signaling Homo sapiens R-HSA-6811558 | 0.789382 | 0.936198 | 0.82739 | 0.1956813 | SRC;INSR;PPP2R5B;PIK3R3;IRS2;PTPN11;EGFR;ERBB3;KIT;ERBB2;AKT1;MAPK1;GRB2;PIP4K2C;IER3;MAPK3 |

| | | | | | | |
|---|---|---|---|---|---|---|
| 1200 | Extracellular matrix organization Homo sapiens R-HSA-1474244 | 0.798533 | 0.945793 | 0.892393 | 0.2007691 | DDR1;ITGB1;COL14A1;COL12A1;ITGA2B;PLOD2;ITGAE;LAMC1;F11R;CAPN15;CTSS;EFEMP1;CAPNS1;CTSL;CAPN7;CDH1;ADAMTS1;CAPN2;CAPN1;ITGB6;CTSD;CTSB;CAST;MMP7;PHYKPL;ITGA3;ADAM10;HSPG2;ADAM17;COL4A4;ADAM9;ITGA6;CD47;PPIB;MATN3;DDR2;ITGA9;CD151;SDC4;SDC2;LAMA3;FURIN;LTBP3;FBLN5;ADAMTS18;CRTAP;TGFB1;COL24A1;LAMB2;TRAPPC4;LAMB1;P4HA1;MFAP1;P4HA2;COL5A2;SDC1;P4HB;AGRN |
| 1201 | Assembly of collagen fibrils and other multimeric structures Homo sapiens R-HSA-2022090 | 0.798803 | 0.945793 | 0.787543 | 0.1769145 | MMP7;CD151;COL24A1;CTSL;COL4A4;LAMA3;COL5A2;ITGA6;CTSS;CTSB |
| 1202 | Defective GALNT12 causes colorectal cancer 1 (CRCS1) Homo sapiens R-HSA-5083636 | 0.802472 | 0.94777 | 0.69325 | 0.1525557 | MUC1;MUC15;MUC20 |
| 1203 | Defective GALNT3 causes familial hyperphosphatemic tumoral calcinosis (HFTC) Homo sapiens R-HSA-5083625 | 0.802472 | 0.94777 | 0.69325 | 0.1525557 | MUC1;MUC15;MUC20 |
| 1204 | TP53 Regulates Transcription of Genes Involved in G2 Cell Cycle Arrest Homo sapiens R-HSA-6804114 | 0.802472 | 0.94777 | 0.69325 | 0.1525557 | PRMT1;EP300;BAX |
| 1205 | Activated PKN1 stimulates transcription of AR (androgen receptor) regulated genes KLK2 and KLK3 Homo sapiens R-HSA-5625886 | 0.805042 | 0.948442 | 0.742643 | 0.1610506 | NCOA2;KDM1A;H2AFZ;H3F3A;PKN1;HIST1H2AC |
| 1206 | The phototransduction cascade Homo sapiens R-HSA-2514856 | 0.805042 | 0.948442 | 0.742643 | 0.1610506 | FNTA;CNGA1;NMT1;CALM1;CAMKMT;METAP2 |

| | | | | | | |
|---|---|---|---|---|---|---|
| 1207 | Striated Muscle Contraction Homo sapiens R-HSA-390522 | 0.805042 | 0.948442 | 0.742643 | 0.1610506 | DES;TPM4;TPM3;TMOD3;TPM1;TCAP |
| 1208 | Glutamate Neurotransmitter Release Cycle Homo sapiens R-HSA-210500 | 0.818728 | 0.96138 | 0.693181 | 0.1386387 | ARL6IP5;PPFIA1;SLC38A2;GLS |
| 1209 | TNFs bind their physiological receptors Homo sapiens R-HSA-5669034 | 0.818728 | 0.96138 | 0.693181 | 0.1386387 | EDA;TNFRSF11B;TNFRSF14;TNFRSF1A |
| 1210 | Nuclear Events (kinase and transcription factor activation) Homo sapiens R-HSA-198725 | 0.818728 | 0.96138 | 0.693181 | 0.1386387 | RPS6KA5;CREB1;MAPK1;MAPK3 |
| 1211 | Pyrimidine metabolism Homo sapiens R-HSA-73848 | 0.818728 | 0.96138 | 0.693181 | 0.1386387 | NT5C3A;DUT;TK2;TK1 |
| 1212 | TP53 Regulates Transcription of Genes Involved in G1 Cell Cycle Arrest Homo sapiens R-HSA-6804116 | 0.823921 | 0.962619 | 0.630249 | 0.1220667 | CCNA1;CDKN1B |
| 1213 | TNFR1-induced proapoptotic signaling Homo sapiens R-HSA-5357786 | 0.823921 | 0.962619 | 0.630249 | 0.1220667 | RIPK1;TNFRSF1A |
| 1214 | Stimuli-sensing channels Homo sapiens R-HSA-2672351 | 0.824313 | 0.962619 | 0.812516 | 0.1569826 | NEDD4L;CLCNKA;WWP1;ANO6;SRI;MCOLN1;TPCN1;CLCN7;CLCN4;UBB;TRPV6;ASPH;OSTM1;SCNN1A;UBC;TRPM7;RAF1;CALM1;SGK2 |
| 1215 | SIRT1 negatively regulates rRNA Expression Homo sapiens R-HSA-427359 | 0.827569 | 0.962619 | 0.716988 | 0.1356991 | TAF1D;TAF1B;H2AFZ;H3F3A;SIRT1;HIST1H2AC |
| 1216 | Inositol transporters Homo sapiens R-HSA-429593 | 0.830375 | 0.962619 | 0.577785 | 0.1073977 | SLC5A3 |

| | | | | | | |
|---|---|---|---|---|---|---|
| 1217 | POLB-Dependent Long Patch Base Excision Repair Homo sapiens R-HSA-110362 | 0.830375 | 0.962619 | 0.577785 | 0.1073977 | APEX1 |
| 1218 | Anchoring fibril formation Homo sapiens R-HSA-2214320 | 0.830375 | 0.962619 | 0.577785 | 0.1073977 | LAMA3 |
| 1219 | TRAIL signaling Homo sapiens R-HSA-75158 | 0.830375 | 0.962619 | 0.577785 | 0.1073977 | CFLAR |
| 1220 | Activation of NIMA Kinases NEK9, NEK6, NEK7 Homo sapiens R-HSA-2980767 | 0.830375 | 0.962619 | 0.577785 | 0.1073977 | NEK7 |
| 1221 | Binding of TCF/LEF:CTNNB1 to target gene promoters Homo sapiens R-HSA-4411364 | 0.830375 | 0.962619 | 0.577785 | 0.1073977 | CTNNB1 |
| 1222 | CREB phosphorylation through the activation of Adenylate Cyclase Homo sapiens R-HSA-442720 | 0.830375 | 0.962619 | 0.577785 | 0.1073977 | CREB1 |
| 1223 | Multifunctional anion exchangers Homo sapiens R-HSA-427601 | 0.830375 | 0.962619 | 0.577785 | 0.1073977 | SLC26A7 |
| 1224 | Highly sodium permeable acetylcholine nicotinic receptors Homo sapiens R-HSA-629587 | 0.830375 | 0.962619 | 0.577785 | 0.1073977 | CHRNA3 |
| 1225 | Na+/Cl- dependent neurotransmitter transporters Homo sapiens R-HSA-442660 | 0.83224 | 0.962619 | 0.64988 | 0.1193402 | SLC6A6;SLC6A13;SLC6A12 |
| 1226 | Scavenging by Class A Receptors Homo sapiens R-HSA-3000480 | 0.83224 | 0.962619 | 0.64988 | 0.1193402 | CALR;HSP90B1;FTL |

| | | | | | | |
|---|---|---|---|---|---|---|
| 1227 | O-linked glycosylation of mucins Homo sapiens R-HSA-913709 | 0.833349 | 0.962619 | 0.769889 | 0.1403531 | GALNT7;GALNT11;C1GALT1C1;MUC1;GALNT4;GALNT3;GALNT1;C1GALT1;B3GNT3;B3GNT2;MUC15;MUC20 |
| 1228 | Opioid Signalling Homo sapiens R-HSA-111885 | 0.83379 | 0.962619 | 0.787305 | 0.1431113 | AHCYL1;PRKCD;GNAI3;ITPR2;ITPR3;GNG11;ADCY6;GNAI2;ADCY9;CREB1;GNG5;CDK5;PRKAR2A;MAPK1;CALM1 |
| 1229 | Ca2+ pathway Homo sapiens R-HSA-4086398 | 0.8339 | 0.962619 | 0.762217 | 0.1384506 | GNG5;FZD5;AGO3;WNT5A;FZD6;ITPR2;CTNNB1;ITPR3;CALM1;GNG11;TNRC6A |
| 1230 | RNA Polymerase I Promoter Opening Homo sapiens R-HSA-73728 | 0.833999 | 0.962619 | 0.693113 | 0.1258157 | H2AFZ;H3F3A;MBD2;HIST1H2AC;MAPK3 |
| 1231 | Activation of Kainate Receptors upon glutamate binding Homo sapiens R-HSA-451326 | 0.833999 | 0.962619 | 0.693113 | 0.1258157 | DLG1;GNG5;DLG3;CALM1;GNG11 |
| 1232 | Metabolism of steroid hormones Homo sapiens R-HSA-196071 | 0.833999 | 0.962619 | 0.693113 | 0.1258157 | FDX1;AKR1B1;HSD17B12;HSD17B11;CYP17A1 |
| 1233 | Aquaporin-mediated transport Homo sapiens R-HSA-445717 | 0.836653 | 0.964899 | 0.74247 | 0.1324165 | ADCY9;GNG5;PRKAR2A;MYO5B;AQP6;AQP2;AQP3;GNG11;ADCY6 |
| 1234 | G alpha (z) signalling events Homo sapiens R-HSA-418597 | 0.839187 | 0.967038 | 0.72947 | 0.1278916 | RGS19;ADCY9;GNG5;PRKCD;GNAI3;GNG11;ADCY6;GNAI2 |
| 1235 | Pre-NOTCH Transcription and Translation Homo sapiens R-HSA-1912408 | 0.842827 | 0.96815 | 0.713403 | 0.1219869 | KAT2A;SNW1;AGO3;EP300;MAML3;RBPJ;TNRC6A |
| 1236 | Activation of G protein gated Potassium channels Homo sapiens R-HSA-1296041 | 0.843557 | 0.96815 | 0.66013 | 0.1123068 | GABBR1;GNG5;KCNJ15;KCNJ3 |
| 1237 | Inhibition of voltage gated Ca2+ channels via Gbeta/gamma subunits Homo sapiens R-HSA-997272 | 0.843557 | 0.96815 | 0.66013 | 0.1123068 | GABBR1;GNG5;KCNJ15;KCNJ3 |

| | | | | | | |
|---|---|---|---|---|---|---|
| 1238 | G protein gated Potassium channels Homo sapiens R-HSA-1296059 | 0.843557 | 0.96815 | 0.66013 | 0.1123068 | GABBR1;GNG5;KCNJ15;KCNJ3 |
| 1239 | Acyl chain remodelling of PC Homo sapiens R-HSA-1482788 | 0.843557 | 0.96815 | 0.66013 | 0.1123068 | PNPLA8;LPCAT3;MBOAT2;PLBD1 |
| 1240 | Visual phototransduction Homo sapiens R-HSA-2187338 | 0.848729 | 0.9733 | 0.789165 | 0.1294356 | SDC4;SDC2;AKR1C1;AKR1C3;HSPG2;DHRS3;CAMKMT;FNTA;RDH11;RDH10;NMT1;NAPEPLD;CNGA1;SDC1;LRP10;CALM1;AGRN;METAP2 |
| 1241 | Cytochrome P450 - arranged by substrate type Homo sapiens R-HSA-211897 | 0.849456 | 0.973349 | 0.747223 | 0.1219161 | NCOA1;CYP39A1;NCOA2;POR;CYP2R1;FDX1;CYP51A1;CYP1A2;CYP1A1;CYP4F11;CYP17A1 |
| 1242 | Polymerase switching on the C-strand of the telomere Homo sapiens R-HSA-174411 | 0.855056 | 0.977403 | 0.577691 | 0.0904599 | POLD4;RFC1 |
| 1243 | Polymerase switching Homo sapiens R-HSA-69091 | 0.855056 | 0.977403 | 0.577691 | 0.0904599 | POLD4;RFC1 |
| 1244 | Leading Strand Synthesis Homo sapiens R-HSA-69109 | 0.855056 | 0.977403 | 0.577691 | 0.0904599 | POLD4;RFC1 |
| 1245 | Regulation of gene expression in beta cells Homo sapiens R-HSA-210745 | 0.858061 | 0.980004 | 0.611612 | 0.0936256 | AKT2;AKT1;FOXA3 |
| 1246 | Collagen biosynthesis and modifying enzymes Homo sapiens R-HSA-1650814 | 0.863849 | 0.980004 | 0.732806 | 0.1072512 | CRTAP;COL24A1;P4HA1;COL14A1;P4HA2;COL4A4;COL12A1;COL5A2;PLOD2;P4HB;PPIB |
| 1247 | Resolution of D-loop Structures through Synthesis-Dependent Strand Annealing (SDSA) Homo sapiens R-HSA-5693554 | 0.865453 | 0.980004 | 0.630084 | 0.0910485 | RAD50;TOP3A;ATM;NBN |

| | | | | | | |
|---|---|---|---|---|---|---|
| 1248 | Activation of ATR in response to replication stress Homo sapiens R-HSA-176187 | 0.866347 | 0.980004 | 0.670645 | 0.0962169 | RPA3;MCM3;RPA2;RAD17;ATRIP;ATR |
| 1249 | Organic anion transporters Homo sapiens R-HSA-428643 | 0.868358 | 0.980004 | 0.495213 | 0.0698997 | SLC17A5 |
| 1250 | Transport of fatty acids Homo sapiens R-HSA-804914 | 0.868358 | 0.980004 | 0.495213 | 0.0698997 | SLC27A4 |
| 1251 | Insulin-like Growth Factor-2 mRNA Binding Proteins (IGF2BPs/IMPs/VICKZs) bind RNA Homo sapiens R-HSA-428359 | 0.868358 | 0.980004 | 0.495213 | 0.0698997 | IGF2BP2 |
| 1252 | Ligand-receptor interactions Homo sapiens R-HSA-5632681 | 0.868358 | 0.980004 | 0.495213 | 0.0698997 | SHH |
| 1253 | Digestion of dietary carbohydrate Homo sapiens R-HSA-189085 | 0.868358 | 0.980004 | 0.495213 | 0.0698997 | MGAM |
| 1254 | Synthesis of diphthamide-EEF2 Homo sapiens R-HSA-5358493 | 0.868358 | 0.980004 | 0.495213 | 0.0698997 | EEF2 |
| 1255 | VEGF ligand-receptor interactions Homo sapiens R-HSA-194313 | 0.868358 | 0.980004 | 0.495213 | 0.0698997 | VEGFB |
| 1256 | VEGF binds to VEGFR leading to receptor dimerization Homo sapiens R-HSA-195399 | 0.868358 | 0.980004 | 0.495213 | 0.0698997 | VEGFB |
| 1257 | Terminal pathway of complement Homo sapiens R-HSA-166665 | 0.868358 | 0.980004 | 0.495213 | 0.0698997 | C9 |

| | | | | | | |
|---|---|---|---|---|---|---|
| 1258 | Cross-presentation of particulate exogenous antigens (phagosomes) Homo sapiens R-HSA-1236973 | 0.868358 | 0.980004 | 0.495213 | 0.0698997 | CD36 |
| 1259 | Hydrolysis of LPC Homo sapiens R-HSA-1483115 | 0.868358 | 0.980004 | 0.495213 | 0.0698997 | PLBD1 |
| 1260 | Linoleic acid (LA) metabolism Homo sapiens R-HSA-2046105 | 0.868358 | 0.980004 | 0.495213 | 0.0698997 | ELOVL1 |
| 1261 | Regulation of beta-cell development Homo sapiens R-HSA-186712 | 0.87408 | 0.984118 | 0.641688 | 0.0863608 | AKT2;AKT1;HES1;RBPJ;FOXA3 |
| 1262 | DNA strand elongation Homo sapiens R-HSA-69190 | 0.87408 | 0.984118 | 0.641688 | 0.0863608 | POLD4;RFC1;RPA3;MCM3;RPA2 |
| 1263 | Signal amplification Homo sapiens R-HSA-392518 | 0.87408 | 0.984118 | 0.641688 | 0.0863608 | GNG5;SRC;GNAI3;GNG11;GNAI2 |
| 1264 | Telomere Maintenance Homo sapiens R-HSA-157579 | 0.879485 | 0.989421 | 0.706953 | 0.0907859 | POLD4;RFC1;H2AFZ;DKC1;RPA3;RUVBL1;TERF2IP;RPA2;TERF1;HIST1H2AC |
| 1265 | ERK/MAPK targets Homo sapiens R-HSA-198753 | 0.880327 | 0.989585 | 0.577596 | 0.0736218 | RPS6KA5;MAPK1;MAPK3 |
| 1266 | Negative regulation of TCF-dependent signaling by WNT ligand antagonists Homo sapiens R-HSA-3772470 | 0.881077 | 0.989646 | 0.533219 | 0.0675108 | WNT5A;LRP6 |
| 1267 | Biological oxidations Homo sapiens R-HSA-211859 | 0.887298 | 0.995847 | 0.816809 | 0.0976695 | SLC35B3;MAOA;MGST3;GSTP1;MGST2;MTR;COMT;BPHL;ADH5;PAPSS2;PTGS1;CYP17A1;CNDP2;UGP2;ESD;CYP2R1;BPNT1;NCOA1;GSTK1;NCOA2;AOC1;GGT6;GSTO2;GSTO1;EPHX1;CYP51A1;AKR1A1;ABHD14B;CMBL;CYP4F11;CYP39A1;GGCT;POR;GCLC;FDX1;CYP1A2;CYP1A1;CES2 |
| 1268 | Glucagon signaling in metabolic regulation Homo sapiens R-HSA-163359 | 0.890829 | 0.998881 | 0.618731 | 0.0715269 | ADCY9;GNG5;PRKAR2A;GNG11;ADCY6 |

| | | | | | | |
|---|---|---|---|---|---|---|
| 1269 | Retinoid cycle disease events Homo sapiens R-HSA-2453864 | 0.897838 | 0.998881 | 0.433283 | 0.0466931 | NAPEPLD |
| 1270 | Diseases associated with visual transduction Homo sapiens R-HSA-2474795 | 0.897838 | 0.998881 | 0.433283 | 0.0466931 | NAPEPLD |
| 1271 | Ca2+ activated K+ channels Homo sapiens R-HSA-1296052 | 0.897838 | 0.998881 | 0.433283 | 0.0466931 | KCNMB4 |
| 1272 | Degradation of cysteine and homocysteine Homo sapiens R-HSA-1614558 | 0.897838 | 0.998881 | 0.433283 | 0.0466931 | TST |
| 1273 | Glucocorticoid biosynthesis Homo sapiens R-HSA-194002 | 0.897838 | 0.998881 | 0.433283 | 0.0466931 | CYP17A1 |
| 1274 | Highly calcium permeable nicotinic acetylcholine receptors Homo sapiens R-HSA-629597 | 0.897838 | 0.998881 | 0.433283 | 0.0466931 | CHRNA3 |
| 1275 | Interleukin-19, 20, 22, 24 Homo sapiens R-HSA-8854691 | 0.897838 | 0.998881 | 0.433283 | 0.0466931 | IL10RB |
| 1276 | Signaling by FGFR3 point mutants in cancer Homo sapiens R-HSA-8853338 | 0.899425 | 0.998881 | 0.547161 | 0.0579989 | GRB2;PLCG1;SOS1 |
| 1277 | Signaling by FGFR3 in disease Homo sapiens R-HSA-5655332 | 0.899425 | 0.998881 | 0.547161 | 0.0579989 | GRB2;PLCG1;SOS1 |
| 1278 | Neurotransmitter Receptor Binding And Downstream Transmission In The Postsynaptic Cell Homo sapiens R-HSA-112314 | 0.900311 | 0.998881 | 0.775847 | 0.0814755 | CHRNA3;RASGRF2;GNAI3;AP2A1;ADCY6;GNAI2;RRAS;GNG5;MYO6;NPTN;MAPK1;AP2M1;KCNJ3;NSF;GABBR1;KCNJ15;BRAF;GNG11;DLG1;ADCY9;CREB1;DLG3;AKAP9;PICK1;RAF1;CALM1 |
| 1279 | G-protein activation Homo sapiens R-HSA-202040 | 0.901433 | 0.998881 | 0.577502 | 0.0599272 | GNG5;GNAI3;GNG11;GNAI2 |

| | | | | | | |
|---|---|---|---|---|---|---|
| 1280 | Adrenaline,noradrenaline inhibits insulin secretion Homo sapiens R-HSA-400042 | 0.901433 | 0.998881 | 0.577502 | 0.0599272 | GNG5;GNG11;ADCY6;GNAI2 |
| 1281 | Transmission across Chemical Synapses Homo sapiens R-HSA-112315 | 0.902315 | 0.998881 | 0.809356 | 0.0831948 | CHRNA3;MAOA;RASGRF2;GNAI3;AP2A1;LIN7A;COMT;ADCY6;GLS;GNAI2;RRAS;GNG5;MYO6;ARL6IP5;PPFIA1;NPTN;MAPK1;GLUL;SLC38A2;AP2M1;KCNJ3;NSF;HSPA8;GABBR1;SLC38A1;CACNA2D1;SLC6A13;KCNJ15;SLC6A12;BRAF;GNG11;DLG1;ADCY9;ALDH5A1;CREB1;DLG3;AKAP9;PICK1;CALM1;RAF1 |
| 1282 | Polo-like kinase mediated events Homo sapiens R-HSA-156711 | 0.902715 | 0.998881 | 0.4951 | 0.0506729 | RBBP4;EP300 |
| 1283 | Acyl chain remodelling of PI Homo sapiens R-HSA-1482922 | 0.902715 | 0.998881 | 0.4951 | 0.0506729 | MBOAT7;PLBD1 |
| 1284 | Metabolism of folate and pterines Homo sapiens R-HSA-196757 | 0.902715 | 0.998881 | 0.4951 | 0.0506729 | SHMT1;MTHFS |
| 1285 | Cell-cell junction organization Homo sapiens R-HSA-421270 | 0.903337 | 0.998881 | 0.679142 | 0.069041 | PRKCI;CLDN4;PARD6B;CDH1;CLDN8;CLDN7;CTNNB1;ANG;F11R;CRB3 |
| 1286 | Platelet homeostasis Homo sapiens R-HSA-418346 | 0.903348 | 0.998881 | 0.721416 | 0.0733298 | ATP2A3;PPP2R5B;ATP2A2;ITPR2;ITPR3;PTPN11;SRI;GNG11;PDE11A;P2RX4;STIM1;GNG5;ORAI1;KCNMB4;CALM1 |
| 1287 | The canonical retinoid cycle in rods (twilight vision) Homo sapiens R-HSA-2453902 | 0.915729 | 0.999996 | 0.51977 | 0.0457581 | RDH11;RDH10;NAPEPLD |
| 1288 | SHC-mediated cascade:FGFR4 Homo sapiens R-HSA-5654719 | 0.915729 | 0.999996 | 0.51977 | 0.0457581 | PTPN11;GRB2;SOS1 |
| 1289 | Surfactant metabolism Homo sapiens R-HSA-5683826 | 0.916009 | 0.999996 | 0.554366 | 0.0486338 | ABCA3;CCDC59;CTSH;ZDHHC2 |
| 1290 | RNA Polymerase I Chain Elongation Homo sapiens R-HSA-73777 | 0.917215 | 0.999996 | 0.64941 | 0.0561175 | TAF1D;CDK7;TAF1B;H2AFZ;H3F3A;POLR1D;POLR2E;POLR2H;HIST1H2AC |

| | | | | | | |
|---|---|---|---|---|---|---|
| 1291 | ABC transporters in lipid homeostasis Homo sapiens R-HSA-1369062 | 0.920626 | 0.999996 | 0.462063 | 0.0382131 | PEX3;ABCA3 |
| 1292 | G1/S-Specific Transcription Homo sapiens R-HSA-69205 | 0.920626 | 0.999996 | 0.462063 | 0.0382131 | CCNA1;TK2 |
| 1293 | Platelet sensitization by LDL Homo sapiens R-HSA-432142 | 0.920626 | 0.999996 | 0.462063 | 0.0382131 | PPP2R5B;PTPN11 |
| 1294 | Acyl chain remodelling of PS Homo sapiens R-HSA-1482801 | 0.920626 | 0.999996 | 0.462063 | 0.0382131 | LPCAT3;MBOAT1 |
| 1295 | Interaction With The Zona Pellucida Homo sapiens R-HSA-1300644 | 0.920717 | 0.999996 | 0.385116 | 0.0318117 | B4GALT1 |
| 1296 | Neurotoxicity of clostridium toxins Homo sapiens R-HSA-168799 | 0.920717 | 0.999996 | 0.385116 | 0.0318117 | VAMP1 |
| 1297 | Creatine metabolism Homo sapiens R-HSA-71288 | 0.920717 | 0.999996 | 0.385116 | 0.0318117 | SLC6A12 |
| 1298 | Abacavir transport and metabolism Homo sapiens R-HSA-2161522 | 0.920717 | 0.999996 | 0.385116 | 0.0318117 | NT5C2 |
| 1299 | Diseases associated with surfactant metabolism Homo sapiens R-HSA-5687613 | 0.920717 | 0.999996 | 0.385116 | 0.0318117 | CCDC59 |
| 1300 | Removal of aminoterminal propeptides from gamma-carboxylated proteins Homo sapiens R-HSA-159782 | 0.920717 | 0.999996 | 0.385116 | 0.0318117 | FURIN |
| 1301 | MASTL Facilitates Mitotic Progression Homo sapiens R-HSA-2465910 | 0.920717 | 0.999996 | 0.385116 | 0.0318117 | PPP2R2D |

| | | | | | | |
|---|---|---|---|---|---|---|
| 1302 | Role of second messengers in netrin-1 signaling Homo sapiens R-HSA-418890 | 0.920717 | 0.999996 | 0.385116 | 0.0318117 | PLCG1 |
| 1303 | Acetylcholine regulates insulin secretion Homo sapiens R-HSA-399997 | 0.920717 | 0.999996 | 0.385116 | 0.0318117 | MARCKS |
| 1304 | Glucagon-type ligand receptors Homo sapiens R-HSA-420092 | 0.928626 | 0.999996 | 0.53301 | 0.0394688 | GNG5;GIPR;SCT;GNG11 |
| 1305 | DARPP-32 events Homo sapiens R-HSA-180024 | 0.929586 | 0.999996 | 0.494987 | 0.0361421 | CDK5;PRKAR2A;CALM1 |
| 1306 | Regulation of insulin secretion Homo sapiens R-HSA-422356 | 0.931508 | 0.999996 | 0.671802 | 0.0476648 | AHCYL1;SLC2A1;ITPR2;ITPR3;IQGAP1;GNG11;ADCY6;GNAI2;MARCKS;GNG5;PRKAR2A;SLC25A4;RAPGEF3 |
| 1307 | Oxidative Stress Induced Senescence Homo sapiens R-HSA-2559580 | 0.93486 | 0.999996 | 0.68327 | 0.046024 | SUZ12;PHC2;RING1;H2AFZ;H3F3A;MAPK10;RBBP4;UBB;AGO3;UBC;MAPK1;RBBP7;HIST1H2AC;TNRC6A;MAPK3 |
| 1308 | Norepinephrine Neurotransmitter Release Cycle Homo sapiens R-HSA-181430 | 0.935396 | 0.999996 | 0.433156 | 0.0289287 | MAOA;PPFIA1 |
| 1309 | SHC-mediated cascade:FGFR3 Homo sapiens R-HSA-5654704 | 0.935396 | 0.999996 | 0.433156 | 0.0289287 | GRB2;SOS1 |
| 1310 | cGMP effects Homo sapiens R-HSA-418457 | 0.935396 | 0.999996 | 0.433156 | 0.0289287 | PDE11A;KCNMB4 |
| 1311 | Synthesis of PIPs at the Golgi membrane Homo sapiens R-HSA-1660514 | 0.935396 | 0.999996 | 0.433156 | 0.0289287 | OCRL;PIK3C2G |
| 1312 | Gamma-carboxylation, transport, and amino-terminal cleavage of proteins Homo sapiens R-HSA-159854 | 0.938473 | 0.999996 | 0.346582 | 0.0220084 | FURIN |

| | | | | | | |
|---|---|---|---|---|---|---|
| 1313 | SLBP Dependent Processing of Replication-Dependent Histone Pre-mRNAs Homo sapiens R-HSA-77588 | 0.938473 | 0.999996 | 0.346582 | 0.0220084 | SLBP |
| 1314 | Activation of the phototransduction cascade Homo sapiens R-HSA-2485179 | 0.938473 | 0.999996 | 0.346582 | 0.0220084 | CNGA1 |
| 1315 | Heme biosynthesis Homo sapiens R-HSA-189451 | 0.938473 | 0.999996 | 0.346582 | 0.0220084 | UROS |
| 1316 | Highly calcium permeable postsynaptic nicotinic acetylcholine receptors Homo sapiens R-HSA-629594 | 0.938473 | 0.999996 | 0.346582 | 0.0220084 | CHRNA3 |
| 1317 | Packaging Of Telomere Ends Homo sapiens R-HSA-171306 | 0.939505 | 0.999996 | 0.513236 | 0.032027 | H2AFZ;TERF2IP;TERF1;HIST1H2AC |
| 1318 | Inwardly rectifying K+ channels Homo sapiens R-HSA-1296065 | 0.939505 | 0.999996 | 0.513236 | 0.032027 | GABBR1;GNG5;KCNJ15;KCNJ3 |
| 1319 | G0 and Early G1 Homo sapiens R-HSA-1538133 | 0.941317 | 0.999996 | 0.472457 | 0.028572 | CCNA1;RBBP4;HDAC1 |
| 1320 | ADP signalling through P2Y purinoceptor 1 Homo sapiens R-HSA-418592 | 0.941317 | 0.999996 | 0.472457 | 0.028572 | GNG5;SRC;GNG11 |
| 1321 | TRP channels Homo sapiens R-HSA-3295583 | 0.941317 | 0.999996 | 0.472457 | 0.028572 | TRPV6;TRPM7;MCOLN1 |
| 1322 | Lipid digestion, mobilization, and transport Homo sapiens R-HSA-73923 | 0.943301 | 0.999996 | 0.63477 | 0.0370518 | AMN;LIPC;SAR1B;LIPH;LIPG;SDC1;ZDHHC8;P4HB;HSPG2;MGLL;LMF1 |

| | | | | | | |
|---|---|---|---|---|---|---|
| 1323 | Amino acid and oligopeptide SLC transporters Homo sapiens R-HSA-425374 | 0.944036 | 0.999996 | 0.577218 | 0.0332427 | SLC6A6;SLC38A1;SLC17A5;SLC6A12;SLC16A10;SLC7A1;SLC38A2 |
| 1324 | GABA receptor activation Homo sapiens R-HSA-977443 | 0.946734 | 0.999996 | 0.589441 | 0.0322643 | GABBR1;ADCY9;GNG5;GNAI3;KCNJ15;ADCY6;GNAI2;KCNJ3 |
| 1325 | Other semaphorin interactions Homo sapiens R-HSA-416700 | 0.947531 | 0.999996 | 0.40765 | 0.0219704 | ITGB1;SEMA4D |
| 1326 | Prostacyclin signalling through prostacyclin receptor Homo sapiens R-HSA-392851 | 0.947531 | 0.999996 | 0.40765 | 0.0219704 | GNG5;GNG11 |
| 1327 | HDL-mediated lipid transport Homo sapiens R-HSA-194223 | 0.947531 | 0.999996 | 0.40765 | 0.0219704 | AMN;ZDHHC8 |
| 1328 | DNA methylation Homo sapiens R-HSA-5334118 | 0.948851 | 0.999996 | 0.494874 | 0.0259824 | DNMT1;H2AFZ;H3F3A;HIST1H2AC |
| 1329 | Muscle contraction Homo sapiens R-HSA-397014 | 0.949806 | 0.999996 | 0.751816 | 0.0387165 | KCNK5;AHCYL1;ATP2A3;ITPR2;ATP2A2;MYL6B;ITPR3;ATP1A1;SRI;MYL12A;MYL12B;CORIN;WWTR1;ANXA2;TPM4;TPM3;CACNA2D1;TMOD3;TPM1;ATP1B3;HIPK1;ATP1B1;MYL6;DES;FXYD4;STIM1;ASPH;FXYD3;KCNQ1;AKAP9;ORAI1;TCAP;TLN1;CALM1;VCL |
| 1330 | Termination of O-glycan biosynthesis Homo sapiens R-HSA-977068 | 0.951212 | 0.999996 | 0.451886 | 0.0226025 | MUC1;MUC15;MUC20 |
| 1331 | SHC-mediated cascade:FGFR2 Homo sapiens R-HSA-5654699 | 0.951212 | 0.999996 | 0.451886 | 0.0226025 | PTPN11;GRB2;SOS1 |
| 1332 | Phase 2 - plateau phase Homo sapiens R-HSA-5576893 | 0.951212 | 0.999996 | 0.451886 | 0.0226025 | CACNA2D1;KCNQ1;AKAP9 |
| 1333 | Ethanol oxidation Homo sapiens R-HSA-71384 | 0.952253 | 0.999996 | 0.315054 | 0.0154139 | ADH5 |

| | | | | | | |
|---|---|---|---|---|---|---|
| 1334 | Inhibition of replication initiation of damaged DNA by RB1/E2F1 Homo sapiens R-HSA-113501 | 0.952253 | 0.999996 | 0.315054 | 0.0154139 | RB1 |
| 1335 | Unwinding of DNA Homo sapiens R-HSA-176974 | 0.952253 | 0.999996 | 0.315054 | 0.0154139 | MCM3 |
| 1336 | NF-kB activation through FADD/RIP-1 pathway mediated by caspase-8 and -10 Homo sapiens R-HSA-933543 | 0.952253 | 0.999996 | 0.315054 | 0.0154139 | RIPK1 |
| 1337 | Miscellaneous substrates Homo sapiens R-HSA-211958 | 0.952253 | 0.999996 | 0.315054 | 0.0154139 | CYP4F11 |
| 1338 | Eicosanoids Homo sapiens R-HSA-211979 | 0.952253 | 0.999996 | 0.315054 | 0.0154139 | CYP4F11 |
| 1339 | Pyrimidine catabolism Homo sapiens R-HSA-73621 | 0.952253 | 0.999996 | 0.315054 | 0.0154139 | NT5C3A |
| 1340 | Presynaptic nicotinic acetylcholine receptors Homo sapiens R-HSA-622323 | 0.952253 | 0.999996 | 0.315054 | 0.0154139 | CHRNA3 |
| 1341 | Role of LAT2/NTAL/LAB on calcium mobilization Homo sapiens R-HSA-2730905 | 0.954826 | 0.999996 | 0.722772 | 0.0334111 | GSK3B;CDKN1B;SRC;PIK3R3;IRS2;EGFR;ERBB3;AKT2;ERBB2;MLST8;AKT1;THEM4;MAPK1;PIP4K2C;IER3;MAPK3;BAD;INSR;PPP2R5B;PTPN11;MTOR;CREB1;AGO3;KIT;RPS6KB2;GRB2;SOS1;TNRC6A |
| 1342 | Chondroitin sulfate biosynthesis Homo sapiens R-HSA-2022870 | 0.957473 | 0.999996 | 0.384978 | 0.0167305 | CHST12;CHST15 |
| 1343 | G beta:gamma signalling through PLC beta Homo sapiens R-HSA-418217 | 0.957473 | 0.999996 | 0.384978 | 0.0167305 | GNG5;GNG11 |
| 1344 | Xenobiotics Homo sapiens R-HSA-211981 | 0.957473 | 0.999996 | 0.384978 | 0.0167305 | CYP1A2;CYP1A1 |

| | | | | | | |
|---|---|---|---|---|---|---|
| 1345 | Facilitative Na+-independent glucose transporters Homo sapiens R-HSA-428790 | 0.962947 | 0.999996 | 0.288781 | 0.0109035 | SLC2A1 |
| 1346 | FGFR3c ligand binding and activation Homo sapiens R-HSA-190372 | 0.962947 | 0.999996 | 0.288781 | 0.0109035 | GALNT3 |
| 1347 | FGFR3 ligand binding and activation Homo sapiens R-HSA-190239 | 0.962947 | 0.999996 | 0.288781 | 0.0109035 | GALNT3 |
| 1348 | Platelet Adhesion to exposed collagen Homo sapiens R-HSA-75892 | 0.962947 | 0.999996 | 0.288781 | 0.0109035 | ITGB1 |
| 1349 | Depolarization of the Presynaptic Terminal Triggers the Opening of Calcium Channels Homo sapiens R-HSA-112308 | 0.962947 | 0.999996 | 0.288781 | 0.0109035 | CACNA2D1 |
| 1350 | SHC-mediated cascade:FGFR1 Homo sapiens R-HSA-5654688 | 0.965593 | 0.999996 | 0.364693 | 0.0127689 | GRB2;SOS1 |
| 1351 | Presynaptic function of Kainate receptors Homo sapiens R-HSA-500657 | 0.965593 | 0.999996 | 0.364693 | 0.0127689 | GNG5;GNG11 |
| 1352 | Synthesis of PA Homo sapiens R-HSA-1483166 | 0.966503 | 0.999996 | 0.415682 | 0.0141628 | AGPAT5;DDHD2;AGPAT3 |
| 1353 | Cardiac conduction Homo sapiens R-HSA-5576891 | 0.968381 | 0.999996 | 0.67344 | 0.0216373 | KCNK5;WWTR1;AHCYL1;CACNA2D1;ATP2A3;ITPR2;ATP2A2;ATP1B3;ITPR3;ATP1A1;SRI;ATP1B1;HIPK1;STIM1;FXYD4;ASPH;FXYD3;KCNQ1;AKAP9;ORAI1;CALM1;CORIN |
| 1354 | LGI-ADAM interactions Homo sapiens R-HSA-5682910 | 0.971246 | 0.999996 | 0.26655 | 0.0077766 | LGI2 |
| 1355 | Bile salt and organic anion SLC transporters Homo sapiens R-HSA-425471 | 0.971246 | 0.999996 | 0.26655 | 0.0077766 | SLC16A7 |

| | | | | | | |
|---|---|---|---|---|---|---|
| 1356 | Lysosphingolipid and LPA receptors Homo sapiens R-HSA-419408 | 0.971246 | 0.999996 | 0.26655 | 0.0077766 | LPAR2 |
| 1357 | Synthesis of PIPs at the early endosome membrane Homo sapiens R-HSA-1660516 | 0.971246 | 0.999996 | 0.26655 | 0.0077766 | INPP5F |
| 1358 | Activation of Nicotinic Acetylcholine Receptors Homo sapiens R-HSA-629602 | 0.971246 | 0.999996 | 0.26655 | 0.0077766 | CHRNA3 |
| 1359 | Postsynaptic nicotinic acetylcholine receptors Homo sapiens R-HSA-622327 | 0.971246 | 0.999996 | 0.26655 | 0.0077766 | CHRNA3 |
| 1360 | Acetylcholine Binding And Downstream Events Homo sapiens R-HSA-181431 | 0.971246 | 0.999996 | 0.26655 | 0.0077766 | CHRNA3 |
| 1361 | Diseases of glycosylation Homo sapiens R-HSA-3781865 | 0.972318 | 0.999996 | 0.599833 | 0.0168387 | SDC4;SDC2;C1GALT1;HSPG2;C1GALT1C1;MUC1;ADAMTS1;ADAMTS18;SDC1;MUC15;THSD7A;AGRN;MUC20 |
| 1362 | Myogenesis Homo sapiens R-HSA-525793 | 0.972329 | 0.999996 | 0.399668 | 0.0112153 | CDC42;CTNNB1;NEO1 |
| 1363 | CDO in myogenesis Homo sapiens R-HSA-375170 | 0.972329 | 0.999996 | 0.399668 | 0.0112153 | CDC42;CTNNB1;NEO1 |
| 1364 | O-linked glycosylation Homo sapiens R-HSA-5173105 | 0.972995 | 0.999996 | 0.632408 | 0.0173127 | GALNT7;GALNT11;GALNT4;GALNT3;GALNT1;C1GALT1;POMGNT1;C1GALT1C1;POFUT2;MUC1;ADAMTS1;ADAMTS18;B3GNT3;B3GNT2;THSD7A;MUC15;MUC20 |
| 1365 | Constitutive Signaling by Aberrant PI3K in Cancer Homo sapiens R-HSA-2219530 | 0.976761 | 0.999996 | 0.522509 | 0.0122859 | ERBB3;KIT;ERBB2;PIK3R3;IRS2;PTPN11;GRB2;EGFR |
| 1366 | Class B/2 (Secretin family receptors) Homo sapiens R-HSA-373080 | 0.977117 | 0.999996 | 0.576745 | 0.0133512 | SHH;CALCA;CALCB;CALCR;GNG5;FZD5;SCT;GIPR;WNT5A;FZD6;GNG11;CD55 |

| | | | | | | |
|---|---|---|---|---|---|---|
| 1367 | Activation of the pre-replicative complex Homo sapiens R-HSA-68962 | 0.977184 | 0.999996 | 0.384841 | 0.0088821 | RPA3;MCM3;RPA2 |
| 1368 | Dopamine Neurotransmitter Release Cycle Homo sapiens R-HSA-212676 | 0.977589 | 0.999996 | 0.329918 | 0.0074779 | PPFIA1;LIN7A |
| 1369 | Thromboxane signalling through TP receptor Homo sapiens R-HSA-428930 | 0.977589 | 0.999996 | 0.329918 | 0.0074779 | GNG5;GNG11 |
| 1370 | Processing of Intronless Pre-mRNAs Homo sapiens R-HSA-77595 | 0.977687 | 0.999996 | 0.247495 | 0.0055849 | PAPOLA |
| 1371 | Fatty acids Homo sapiens R-HSA-211935 | 0.977687 | 0.999996 | 0.247495 | 0.0055849 | CYP4F11 |
| 1372 | Diseases associated with O-glycosylation of proteins Homo sapiens R-HSA-3906995 | 0.979886 | 0.999996 | 0.5128 | 0.0104198 | C1GALT1C1;MUC1;ADAMTS1;ADAMTS18;C1GALT1;MUC15;THSD7A;MUC20 |
| 1373 | FCERI mediated Ca+2 mobilization Homo sapiens R-HSA-2871809 | 0.979886 | 0.999996 | 0.5128 | 0.0104198 | VAV3;AHCYL1;ITPR2;ITPR3;GRB2;PLCG1;SOS1;CALM1 |
| 1374 | Adherens junctions interactions Homo sapiens R-HSA-418990 | 0.981221 | 0.999996 | 0.371073 | 0.0070345 | CDH1;CTNNB1;ANG |
| 1375 | Post-Elongation Processing of Intronless pre-mRNA Homo sapiens R-HSA-112297 | 0.981953 | 0.999996 | 0.314901 | 0.005735 | SLBP;PAPOLA |
| 1376 | Processing of Capped Intronless Pre-mRNA Homo sapiens R-HSA-75067 | 0.981953 | 0.999996 | 0.314901 | 0.005735 | SLBP;PAPOLA |

| | | | | | | |
|---|---|---|---|---|---|---|
| 1377 | O-glycosylation of TSR domain-containing proteins Homo sapiens R-HSA-5173214 | 0.982137 | 0.999996 | 0.407385 | 0.0073428 | POFUT2;ADAMTS1;ADAMTS18;THSD7A |
| 1378 | JNK (c-Jun kinases) phosphorylation and activation mediated by activated human TAK1 Homo sapiens R-HSA-450321 | 0.982685 | 0.999996 | 0.23098 | 0.0040345 | MAPK10 |
| 1379 | Metabolism of Angiotensinogen to Angiotensins Homo sapiens R-HSA-2022377 | 0.982685 | 0.999996 | 0.23098 | 0.0040345 | CTSD |
| 1380 | Nitric oxide stimulates guanylate cyclase Homo sapiens R-HSA-392154 | 0.985486 | 0.999996 | 0.30119 | 0.0044034 | PDE11A;KCNMB4 |
| 1381 | Acetylcholine Neurotransmitter Release Cycle Homo sapiens R-HSA-264642 | 0.986564 | 0.999996 | 0.21653 | 0.0029291 | PPFIA1 |
| 1382 | Unblocking of NMDA receptor, glutamate binding and activation Homo sapiens R-HSA-438066 | 0.986564 | 0.999996 | 0.21653 | 0.0029291 | AKAP9 |
| 1383 | Acyl chain remodelling of PG Homo sapiens R-HSA-1482925 | 0.986564 | 0.999996 | 0.21653 | 0.0029291 | LPGAT1 |
| 1384 | E2F mediated regulation of DNA replication Homo sapiens R-HSA-113510 | 0.987342 | 0.999996 | 0.34629 | 0.0044115 | RB1;CCNA1;TK2 |
| 1385 | Fertilization Homo sapiens R-HSA-1187000 | 0.988342 | 0.999996 | 0.288622 | 0.0033844 | B4GALT1;CD9 |
| 1386 | Reproduction Homo sapiens R-HSA-1474165 | 0.988342 | 0.999996 | 0.288622 | 0.0033844 | B4GALT1;CD9 |

| | | | | | | |
|---|---|---|---|---|---|---|
| 1387 | Serotonin Neurotransmitter Release Cycle Homo sapiens R-HSA-181429 | 0.989573 | 0.999996 | 0.20378 | 0.0021359 | PPFIA1 |
| 1388 | Amine-derived hormones Homo sapiens R-HSA-209776 | 0.989573 | 0.999996 | 0.20378 | 0.0021359 | PNMT |
| 1389 | Gap junction assembly Homo sapiens R-HSA-190861 | 0.991909 | 0.999996 | 0.192446 | 0.0015634 | GJB1 |
| 1390 | Phase 4 - resting membrane potential Homo sapiens R-HSA-5576886 | 0.991909 | 0.999996 | 0.192446 | 0.0015634 | KCNK5 |
| 1391 | Hormone-sensitive lipase (HSL)-mediated triacylglycerol hydrolysis Homo sapiens R-HSA-163560 | 0.993721 | 0.999996 | 0.182306 | 0.0011482 | MGLL |
| 1392 | Glucuronidation Homo sapiens R-HSA-156588 | 0.993721 | 0.999996 | 0.182306 | 0.0011482 | UGP2 |
| 1393 | Defective B3GALTL causes Peters-plus syndrome (PpS) Homo sapiens R-HSA-5083635 | 0.994347 | 0.999996 | 0.305471 | 0.0017317 | ADAMTS1;ADAMTS18;THSD7A |
| 1394 | NCAM1 interactions Homo sapiens R-HSA-419037 | 0.994347 | 0.999996 | 0.305471 | 0.0017317 | PRNP;COL4A4;AGRN |
| 1395 | Regulation of Insulin-like Growth Factor (IGF) transport and uptake by Insulin-like Growth Factor Binding Proteins (IGFBPs) Homo sapiens R-HSA-381426 | 0.995128 | 0.999996 | 0.173179 | 0.0008458 | IGFBP5 |
| 1396 | Common Pathway of Fibrin Clot Formation Homo sapiens R-HSA-140875 | 0.996219 | 0.999996 | 0.164922 | 0.0006248 | SERPINA5 |

| | | | | | | |
|---|---|---|---|---|---|---|
| 1397 | Intrinsic Pathway of Fibrin Clot Formation Homo sapiens R-HSA-140837 | 0.996219 | 0.999996 | 0.164922 | 0.0006248 | SERPINA5 |
| 1398 | Role of phospholipids in phagocytosis Homo sapiens R-HSA-2029485 | 0.996232 | 0.999996 | 0.384428 | 0.0014513 | AHCYL1;PRKCD;ITPR2;ITPR3;PLCG1;PLD3 |
| 1399 | Neuronal System Homo sapiens R-HSA-112316 | 0.996701 | 0.999996 | 0.670587 | 0.0022158 | KCNG1;RASGRF2;AP2A1;COMT;PTPRF;GLS;PANX1;ARL6IP5;PPFIA1;FLOT1;NPTN;FLOT2;GLUL;AP2M1;NSF;SLC38A1;CACNA2D1;SLC6A13;SLC6A12;ADCY9;ALDH5A1;CREB1;KCNQ1;AKAP9;PICK1;RAF1;RTN3;CHRNA3;MAOA;GNAI3;LIN7A;ADCY6;GNAI2;RRAS;GNG5;MYO6;KCNMB4;MAPK1;SLC38A2;KCNJ3;HSPA8;GABBR1;KCNJ15;BRAF;GNG11;DLG1;LRFN4;DLG3;CALM1 |
| 1400 | Activation of Matrix Metalloproteinases Homo sapiens R-HSA-1592389 | 0.996937 | 0.999996 | 0.230808 | 0.0007081 | MMP7;FURIN |
| 1401 | Chemokine receptors bind chemokines Homo sapiens R-HSA-380108 | 0.997718 | 0.999996 | 0.339191 | 0.0007748 | ACKR4;ACKR3;CCL28;CX3CL1;CXCL16 |
| 1402 | Phase 1 - inactivation of fast Na+ channels Homo sapiens R-HSA-5576894 | 0.998935 | 0.999996 | 0.133163 | 0.0001419 | CACNA2D1 |
| 1403 | Binding and Uptake of Ligands by Scavenger Receptors Homo sapiens R-HSA-2173782 | 0.999701 | 0.999996 | 0.305005 | 9.11E-05 | HSP90AA1;CD36;CALR;APOL1;FTL;HSP90B1 |
| 1404 | Voltage gated Potassium channels Homo sapiens R-HSA-1296072 | 0.999751 | 0.999996 | 0.168764 | 4.20E-05 | KCNG1;KCNQ1 |
| 1405 | Phase 0 - rapid depolarisation Homo sapiens R-HSA-5576892 | 0.999802 | 0.999996 | 0.164735 | 3.26E-05 | CACNA2D1;CALM1 |
| 1406 | FCGR activation Homo sapiens R-HSA-2029481 | 0.999936 | 0.999996 | 0.147163 | 9.36E-06 | YES1;SRC |

| | | | | | | |
|---|---|---|---|---|---|---|
| 1407 | Formation of Fibrin Clot (Clotting Cascade) Homo sapiens R-HSA-140877 | 0.999947 | 0.999996 | 0.091041 | 4.86E-06 | SERPINA5 |
| 1408 | Class C/3 (Metabotropic glutamate/pheromone receptors) Homo sapiens R-HSA-420499 | 0.999958 | 0.999996 | 0.088701 | 3.73E-06 | GABBR1 |
| 1409 | Signal Transduction Homo sapiens R-HSA-162582 | 0.999974 | 0.999996 | 0.802431 | 2.07E-05 | NCKAP1;CYFIP1;SCT;TRRAP;ITSN1;WDR83;SPINT2;F11R;RBPJ;PSMD8;PSMD6;PSMD7;PSMD4;AKT2;PSMD2;PSMD3;AKT1;PSMD1;VPS35;PIP4K2C;WLS;IER3;PRKCI;CSNK2A1;PRKCD;CSNK2A2;HSPG2;BCR;MYL6;IFT88;ADCY9;ZFYVE16;SPINT1;DAAM1;AGTR1;PSME1;PSME2;NUP98;RBCK1;MAML3;CTBP2;NEDD4L;VPS26A;IQGAP1;IQGAP2;HIF1A;SKA2;RHOT2;RDH11;RDH10;PRKAR2A;RDH14;ST3GAL6;RDH13;PSMF1;PLCG1;FZD5;FZD6;H3F3A;BRAF;CGN;HNRNPM;AGO3;HNRNPF;NMT1;NAPEPLD;RARG;ARHGAP8;CDKN1B;ARPC1B;ARPC1A;CLTC;CLTB;PRKAG2;SHB;ARHGAP6;SHH;HEY1;RSPO3;JAK2;JAK1;OFD1;CIT;PSMA5;PSMA6;KAT2A;PSMA3;NCOR1;PSMA1;PSMA2;CDC42EP2;DLD;RAPGEF3;MGLL;PAFAH1B1;EGFR;ADCY6;PSMA7;CENPC;PSMB6;NEURL1B;PSMB4;PSMB5;PSMB2;PSMB3;PSMB1;SPOP;STAT6;CENPT;EVC2;CSNK1A1;KIDINS220;USP9X;STAT1;STAT3;DAB2IP;SYVN1;WWP1;DHRS3;PSMB8;PSMB9;KTN1;PSMC5;ABI2;PSMC4;PSMC2;PKN2;GOPC;PKN1;LAMTOR1;AGRN;CCL28;POGLUT1;CCNT2;FAM13A;ITGA2B;IRS2;CDC14B;CXCL16;ABHD12;FNTA;CDH1;RUVBL1;AP2M1;MEN1;USP8;TLE3;RNF43;TLE2;MAP2K1;MAP2K2;GTF2F2;PYGO1;CLIP1;IL6ST;SQSTM1;CSNK1G2;DGKH;KHDRBS1;SHC2;CUL5;CUL3;CUL1;ITPR2;ITPR3;PHB;RAP1B;KLC2;TTC21B;PSAP;DVL3;THEM4;SRGAP3;SRGAP1;MAP2K5;TI [illegible] |
| 1410 | Complement cascade Homo sapiens R-HSA-166658 | 0.999976 | 0.999996 | 0.230292 | 5.51E-06 | C9;CD59;CD46;CFB;CD55 |
| 1411 | Potassium Channels Homo sapiens R-HSA-1296071 | 0.999986 | 0.999996 | 0.262662 | 3.70E-06 | KCNG1;GABBR1;GNG5;KCNQ1;KCNJ15;KCNMB4;KCNJ3 |
| 1412 | G alpha (s) signalling events Homo sapiens R-HSA-418555 | 0.999986 | 0.999996 | 0.347521 | 4.80E-06 | CALCA;CALCB;SCT;GIPR;GNAI3;ARRB1;GNG11;ADCY6;GNAI2;PDE11A;ADCY9;CALCR;GNG5 |

| | | | | | | |
|---|---|---|---|---|---|---|
| 1413 | Scavenging of heme from plasma Homo sapiens R-HSA-2168880 | 0.999988 | 0.999996 | 0.076844 | 8.94E-07 | APOL1 |
| 1414 | Signaling by GPCR Homo sapiens R-HSA-372790 | 0.999992 | 0.999996 | 0.412379 | 3.39E-06 | APP;AHCYL1;SCT;ITSN1;ITGA2B;PEBP1;WDR83;IRS2;CXCL16;PSMD8;ABHD12;RGS5;PSMD6;PSMD7;PSMD4;AKT2;PSMD2;PSMD3;AKT1;PSMD1;MAP2K1;MAP2K2;PRKCD;WNT5A;ADCY9;AKAP9;AGTR1;PSME1;PSME2;RAF1;TLN1;VCL;DGKH;SHC2;CALCA;PSMD12;SPTBN5;CALCB;PSMD11;GIPR;PSMD13;CUL3;LPAR2;PIK3R3;ITPR2;ITPR3;IQGAP1;PHB;NTS;RAP1B;HCAR2;HCAR3;CALCR;HCAR1;UBB;PRKAR2A;PSAP;UBC;PSMF1;ECT2;SPTBN1;SPTBN2;FZD5;FZD6;BRAF;GNG11;PTK2;CDK5;PTPRA;NF1;GRB2;CALM1;MAP3K11;YWHAB;RASGRF2;ARRB1;CX3CL1;HEBP1;SHH;RASGEF1A;GPER1;MCF2L;RAC1;JAK2;SPTAN1;JAK1;OXGR1;VAV3;ARHGEF12;ARHGEF16;PPP2R5B;SSTR2;RHOA;PSMA5;PSMA6;PSMA3;CREB1;PSMA1;PSMA2;KIT;IL3RA;RASA2;ARHGEF1;ARHGEF2;SOS1;MGLL;NPB;RGS19;ROCK1;ROCK2;SRC;GNAI3;PSMA7;EGFR;PSMB10;ADCY6;GNAI2;PSMB6;CDC42;PSMB4;PDE11A;PSMB5;PSMB2;GNG5;ERBB3;PSMB3;PSMB1;ERBB2;MAPK1;CSK;CD55;MAPK3;GABBR1;DAB2IP;PSMB8;PSMB9;PSMC5;PSMC4;PSMC2;ACKR4;ACKR3;F2RL1;CCL28 |
| 1415 | GPCR downstream signaling Homo sapiens R-HSA-388396 | 0.999992 | 0.999996 | 0.21487 | 1.66E-06 | APP;SCT;RASGRF2;ITSN1;ARRB1;CXCL16;ABHD12;HEBP1;RGS5;AKT2;GPER1;MCF2L;AKT1;RAC1;JAK2;JAK1;OXGR1;VAV3;ARHGEF12;ARHGEF16;PRKCD;SSTR2;RHOA;ADCY9;IL3RA;AGTR1;ARHGEF1;ARHGEF2;SOS1;MGLL;DGKH;NPB;CALCA;CALCB;RGS19;ROCK1;ROCK2;GIPR;GNAI3;LPAR2;PIK3R3;ITPR2;ITPR3;NTS;ADCY6;GNAI2;CDC42;HCAR2;HCAR3;PDE11A;CALCR;HCAR1;GNG5;ECT2;GABBR1;GNG11;ACKR3;F2RL1;GRB2;CCL28 |
| 1416 | GPCR ligand binding Homo sapiens R-HSA-500792 | 0.999994 | 0.999996 | 0.262082 | 1.67E-06 | APP;NPB;CALCA;CALCB;SCT;GIPR;LPAR2;NTS;CX3CL1;CXCL16;HCAR2;HEBP1;HCAR3;SHH;CALCR;HCAR1;GNG5;GPER1;PSAP;CD55;OXGR1;GABBR1;FZD5;FZD6;WNT5A;SSTR2;GNG11;AGTR1;ACKR4;F2RL1;ACKR3;CCL28 |
| 1417 | Class A/1 (Rhodopsin-like receptors) Homo sapiens R-HSA-373076 | 0.999994 | 0.999996 | 0.213366 | 1.24E-06 | APP;NPB;LPAR2;NTS;SSTR2;CX3CL1;CXCL16;HCAR2;HEBP1;HCAR3;HCAR1;GPER1;PSAP;AGTR1;ACKR4;F2RL1;ACKR3;CCL28;OXGR1 |

| | | | | | | |
|---|---|---|---|---|---|---|
| 1418 | G alpha (i) signalling events Homo sapiens R-HSA-418594 | 0.999995 | 0.999996 | 0.346698 | 1.86E-06 | APP;NPB;GABBR1;RGS19;GNAI3;LPAR2;SSTR2;GNG11;ADCY6;GNAI2;CXCL16;HCAR2;HEBP1;RGS5;HCAR3;ADCY9;HCAR1;GNG5;GPER1;ACKR3;CCL28;OXGR1 |
| 1419 | G alpha (q) signalling events Homo sapiens R-HSA-416476 | 0.999995 | 0.999996 | 0.293138 | 1.47E-06 | APP;RGS19;PRKCD;LPAR2;ITPR2;PIK3R3;ITPR3;NTS;GNG11;ABHD12;GNG5;AGTR1;F2RL1;MGLL;DGKH |
| 1420 | Peptide ligand-binding receptors Homo sapiens R-HSA-375276 | 0.999995 | 0.999996 | 0.268897 | 1.35E-06 | APP;NPB;NTS;SSTR2;CX3CL1;CXCL16;HEBP1;GPER1;PSAP;AGTR1;ACKR4;F2RL1;ACKR3;CCL28 |
| 1421 | Immunoregulatory interactions between a Lymphoid and a non-Lymphoid cell Homo sapiens R-HSA-198933 | 0.999995 | 0.999996 | 0.209258 | 9.89E-07 | ITGB1;CD40;CXADR;CD81;CDH1;HLA-C;HLA-F;B2M;HLA-E |
| 1422 | Initial triggering of complement Homo sapiens R-HSA-166663 | 0.999996 | 0.999996 | 0.06062 | 2.30E-07 | CFB |

**Pathways enriched by genes suppressed with SGLT2 inhibitor in intercalated cells**

| | Term | P value | Adjusted. P value | Odds Ratio | Combined Score | Genes |
|---|---|---|---|---|---|---|
| 1 | Response to metal ions Homo sapiens R-HSA-5660526 | 1.41E-09 | 2.27E-07 | 170.8935 | 3482.8981 | MT2A;MT1F;MT1G;MT1H;MT1E |
| 2 | Metallothioneins bind metals Homo sapiens R-HSA-5661231 | 1.41E-09 | 2.27E-07 | 170.8935 | 3482.8981 | MT2A;MT1F;MT1G;MT1H;MT1E |
| 3 | Constitutive Signaling by Aberrant PI3K in Cancer Homo sapiens R-HSA-2219530 | 1.48E-05 | 0.001591 | 18.26399 | 203.08221 | FGF9;ERBB4;EGF;PDGFA;FGFR3 |
| 4 | NCAM signaling for neurite out-growth Homo sapiens R-HSA-375165 | 6.57E-05 | 0.00251 | 6.478641 | 62.390117 | FGF9;ERBB4;EGF;COL6A2;PDGFA;NEFL;FGFR3;DUSP9 |
| 5 | PI5P, PP2A and IER3 Regulate PI3K/AKT Signaling Homo sapiens R-HSA-6811558 | 6.62E-05 | 0.00251 | 13.09807 | 126.03856 | FGF9;ERBB4;EGF;PDGFA;FGFR3 |
| 6 | Gastrin-CREB signalling pathway via PKC and MAPK Homo sapiens R-HSA-881907 | 7.22E-05 | 0.00251 | 5.016485 | 47.834607 | CASR;FGF9;ERBB4;EGF;PDGFA;NEFL;AVPR1A;FGFR3;DUSP9;KNG1 |
| 7 | PI3K/AKT Signaling in Cancer Homo sapiens R-HSA-2219528 | 7.85E-05 | 0.00251 | 12.61105 | 119.21055 | FGF9;ERBB4;EGF;PDGFA;FGFR3 |
| 8 | Negative regulation of the PI3K/AKT network Homo sapiens R-HSA-199418 | 9.74E-05 | 0.00251 | 12.01516 | 110.97563 | FGF9;ERBB4;EGF;PDGFA;FGFR3 |
| 9 | Signaling by PDGF Homo sapiens R-HSA-186797 | 0.000103 | 0.00251 | 5.327488 | 48.929275 | FGF9;ERBB4;EGF;COL6A2;SPP1;PDGFA;NEFL;FGFR3;DUSP9 |
| 10 | IRS-mediated signalling Homo sapiens R-HSA-112399 | 0.000104 | 0.00251 | 6.05057 | 55.511626 | FGF9;ERBB4;EGF;RRAGD;PDGFA;NEFL;FGFR3;DUSP9 |
| 11 | Insulin receptor signalling cascade Homo sapiens R-HSA-74751 | 0.000111 | 0.00251 | 5.984595 | 54.471861 | FGF9;ERBB4;EGF;RRAGD;PDGFA;NEFL;FGFR3;DUSP9 |
| 12 | IGF1R signaling cascade Homo sapiens R-HSA-2428924 | 0.000114 | 0.00251 | 5.962918 | 54.131446 | FGF9;ERBB4;EGF;RRAGD;PDGFA;NEFL;FGFR3;DUSP9 |

| | | | | | | |
|---|---|---|---|---|---|---|
| 13 | Signaling by Type 1 Insulin-like Growth Factor 1 Receptor (IGF1R) Homo sapiens R-HSA-2404192 | 0.000114 | 0.00251 | 5.962918 | 54.131446 | FGF9;ERBB4;EGF;RRAGD;PDGFA;NEFL;FGFR3;DUSP9 |
| 14 | IRS-related events triggered by IGF1R Homo sapiens R-HSA-2428928 | 0.000114 | 0.00251 | 5.962918 | 54.131446 | FGF9;ERBB4;EGF;RRAGD;PDGFA;NEFL;FGFR3;DUSP9 |
| 15 | FCERI mediated MAPK activation Homo sapiens R-HSA-2871796 | 0.000117 | 0.00251 | 5.941395 | 53.794054 | FGF9;ERBB4;EGF;PDGFA;NEFL;FOS;FGFR3;DUSP9 |
| 16 | Signaling by Insulin receptor Homo sapiens R-HSA-74752 | 0.000193 | 0.002581 | 5.503827 | 47.068498 | FGF9;ERBB4;EGF;RRAGD;PDGFA;NEFL;FGFR3;DUSP9 |
| 17 | GRB2 events in EGFR signaling Homo sapiens R-HSA-179812 | 0.000203 | 0.002581 | 6.356879 | 54.060893 | FGF9;ERBB4;EGF;PDGFA;NEFL;FGFR3;DUSP9 |
| 18 | SHC1 events in EGFR signaling Homo sapiens R-HSA-180336 | 0.000203 | 0.002581 | 6.356879 | 54.060893 | FGF9;ERBB4;EGF;PDGFA;NEFL;FGFR3;DUSP9 |
| 19 | SOS-mediated signalling Homo sapiens R-HSA-112412 | 0.000203 | 0.002581 | 6.356879 | 54.060893 | FGF9;ERBB4;EGF;PDGFA;NEFL;FGFR3;DUSP9 |
| 20 | SHC1 events in ERBB4 signaling Homo sapiens R-HSA-1250347 | 0.000203 | 0.002581 | 6.356879 | 54.060893 | FGF9;ERBB4;EGF;PDGFA;NEFL;FGFR3;DUSP9 |
| 21 | RAF/MAP kinase cascade Homo sapiens R-HSA-5673001 | 0.000203 | 0.002581 | 6.356879 | 54.060893 | FGF9;ERBB4;EGF;PDGFA;NEFL;FGFR3;DUSP9 |
| 22 | FRS-mediated FGFR2 signaling Homo sapiens R-HSA-5654700 | 0.000208 | 0.002581 | 6.328798 | 53.657715 | FGF9;ERBB4;EGF;PDGFA;NEFL;FGFR3;DUSP9 |
| 23 | FRS-mediated FGFR4 signaling Homo sapiens R-HSA-5654712 | 0.000208 | 0.002581 | 6.328798 | 53.657715 | FGF9;ERBB4;EGF;PDGFA;NEFL;FGFR3;DUSP9 |
| 24 | FRS-mediated FGFR3 signaling Homo sapiens R-HSA-5654706 | 0.000208 | 0.002581 | 6.328798 | 53.657715 | FGF9;ERBB4;EGF;PDGFA;NEFL;FGFR3;DUSP9 |
| 25 | FRS-mediated FGFR1 signaling Homo sapiens R-HSA-5654693 | 0.000208 | 0.002581 | 6.328798 | 53.657715 | FGF9;ERBB4;EGF;PDGFA;NEFL;FGFR3;DUSP9 |
| 26 | ARMS-mediated activation Homo sapiens R-HSA-170984 | 0.000225 | 0.002581 | 6.246007 | 52.473936 | FGF9;ERBB4;EGF;PDGFA;NEFL;FGFR3;DUSP9 |
| 27 | Signalling to p38 via RIT and RIN Homo sapiens R-HSA-187706 | 0.000225 | 0.002581 | 6.246007 | 52.473936 | FGF9;ERBB4;EGF;PDGFA;NEFL;FGFR3;DUSP9 |

| | | | | | | |
|---|---|---|---|---|---|---|
| 28 | Frs2-mediated activation Homo sapiens R-HSA-170968 | 0.00023 | 0.002581 | 6.218884 | 52.08772 | FGF9;ERBB4;EGF;PDGFA;NEFL;FGFR3;DUSP9 |
| 29 | MAPK1/MAPK3 signaling Homo sapiens R-HSA-5684996 | 0.000236 | 0.002581 | 6.191993 | 51.705592 | FGF9;ERBB4;EGF;PDGFA;NEFL;FGFR3;DUSP9 |
| 30 | Prolonged ERK activation events Homo sapiens R-HSA-169893 | 0.000242 | 0.002581 | 6.16533 | 51.327493 | FGF9;ERBB4;EGF;PDGFA;NEFL;FGFR3;DUSP9 |
| 31 | Signaling by Leptin Homo sapiens R-HSA-2586552 | 0.000248 | 0.002581 | 6.138894 | 50.953366 | FGF9;ERBB4;EGF;PDGFA;NEFL;FGFR3;DUSP9 |
| 32 | Interleukin receptor SHC signaling Homo sapiens R-HSA-912526 | 0.000261 | 0.002612 | 6.086687 | 50.2168 | FGF9;ERBB4;EGF;PDGFA;NEFL;FGFR3;DUSP9 |
| 33 | Signalling to RAS Homo sapiens R-HSA-167044 | 0.000268 | 0.002612 | 6.060912 | 49.854251 | FGF9;ERBB4;EGF;PDGFA;NEFL;FGFR3;DUSP9 |
| 34 | VEGFR2 mediated cell proliferation Homo sapiens R-HSA-5218921 | 0.000281 | 0.002652 | 6.010002 | 49.140355 | FGF9;ERBB4;EGF;PDGFA;NEFL;FGFR3;DUSP9 |
| 35 | Signaling by ERBB4 Homo sapiens R-HSA-1236394 | 0.000288 | 0.002652 | 5.174045 | 42.176191 | CXCL12;FGF9;ERBB4;EGF;PDGFA;NEFL;FGFR3;DUSP9 |
| 36 | Axon guidance Homo sapiens R-HSA-422475 | 0.000301 | 0.002689 | 4.174128 | 33.850847 | FGF9;ERBB4;EGF;COL6A2;PLXNA2;PDGFA;NEFL;SLIT2;FGFR3;DUSP9 |
| 37 | Interleukin-2 signaling Homo sapiens R-HSA-451927 | 0.00031 | 0.002689 | 5.910677 | 47.755924 | FGF9;ERBB4;EGF;PDGFA;NEFL;FGFR3;DUSP9 |
| 38 | Signalling to ERKs Homo sapiens R-HSA-187687 | 0.000317 | 0.002689 | 5.88635 | 47.41856 | FGF9;ERBB4;EGF;PDGFA;NEFL;FGFR3;DUSP9 |
| 39 | Interleukin-3, 5 and GM-CSF signaling Homo sapiens R-HSA-512988 | 0.000383 | 0.002892 | 5.698632 | 44.838308 | FGF9;ERBB4;EGF;PDGFA;NEFL;FGFR3;DUSP9 |
| 40 | PI-3K cascade:FGFR1 Homo sapiens R-HSA-5654689 | 0.000404 | 0.002892 | 8.714865 | 68.0963 | FGF9;ERBB4;EGF;PDGFA;FGFR3 |
| 41 | PI-3K cascade:FGFR3 Homo sapiens R-HSA-5654710 | 0.000404 | 0.002892 | 8.714865 | 68.0963 | FGF9;ERBB4;EGF;PDGFA;FGFR3 |
| 42 | PI3K events in ERBB4 signaling Homo sapiens R-HSA-1250342 | 0.000404 | 0.002892 | 8.714865 | 68.0963 | FGF9;ERBB4;EGF;PDGFA;FGFR3 |
| 43 | PIP3 activates AKT signaling Homo sapiens R-HSA-1257604 | 0.000404 | 0.002892 | 8.714865 | 68.0963 | FGF9;ERBB4;EGF;PDGFA;FGFR3 |
| 44 | PI-3K cascade:FGFR4 Homo sapiens R-HSA-5654720 | 0.000404 | 0.002892 | 8.714865 | 68.0963 | FGF9;ERBB4;EGF;PDGFA;FGFR3 |

| | | | | | | |
|---|---|---|---|---|---|---|
| 45 | PI-3K cascade:FGFR2 Homo sapiens R-HSA-5654695 | 0.000404 | 0.002892 | 8.714865 | 68.0963 | FGF9;ERBB4;EGF;PDGFA;FGFR3 |
| 46 | GAB1 signalosome Homo sapiens R-HSA-180292 | 0.000452 | 0.003095 | 8.495704 | 65.437034 | FGF9;ERBB4;EGF;PDGFA;FGFR3 |
| 47 | PI3K/AKT activation Homo sapiens R-HSA-198203 | 0.000452 | 0.003095 | 8.495704 | 65.437034 | FGF9;ERBB4;EGF;PDGFA;FGFR3 |
| 48 | Cation-coupled Chloride cotransporters Homo sapiens R-HSA-426117 | 0.000532 | 0.003495 | 79.572 | 599.89492 | SLC12A3;SLC12A1 |
| 49 | FGFR3b ligand binding and activation Homo sapiens R-HSA-190371 | 0.000532 | 0.003495 | 79.572 | 599.89492 | FGF9;FGFR3 |
| 50 | MAPK family signaling cascades Homo sapiens R-HSA-5683057 | 0.000633 | 0.004075 | 5.219343 | 38.443112 | FGF9;ERBB4;EGF;PDGFA;NEFL;FGFR3;DUSP9 |
| 51 | Developmental Biology Homo sapiens R-HSA-1266738 | 0.000659 | 0.004087 | 3.294401 | 24.130445 | FGF9;ERBB4;EGF;COL6A2;PLXNA2;LPL;PDGFA;NEFL;SLIT2;TDGF1;FGFR3;DUSP9 |
| 52 | NGF signalling via TRKA from the plasma membrane Homo sapiens R-HSA-187037 | 0.00066 | 0.004087 | 4.541797 | 33.261062 | NTRK2;FGF9;ERBB4;EGF;PDGFA;NEFL;FGFR3;DUSP9 |
| 53 | Fc epsilon receptor (FCERI) signaling Homo sapiens R-HSA-2454202 | 0.00094 | 0.005711 | 4.290725 | 29.904835 | FGF9;ERBB4;EGF;PDGFA;NEFL;FOS;FGFR3;DUSP9 |
| 54 | VEGFA-VEGFR2 Pathway Homo sapiens R-HSA-4420097 | 0.001268 | 0.007492 | 4.61056 | 30.753508 | FGF9;ERBB4;EGF;PDGFA;NEFL;FGFR3;DUSP9 |
| 55 | Retinoid metabolism and transport Homo sapiens R-HSA-975634 | 0.00128 | 0.007492 | 15.43046 | 102.78544 | GPC3;LPL;APOE |
| 56 | Signaling by SCF-KIT Homo sapiens R-HSA-1433557 | 0.001386 | 0.007846 | 4.536908 | 29.858753 | FGF9;ERBB4;EGF;PDGFA;NEFL;FGFR3;DUSP9 |
| 57 | Role of LAT2/NTAL/LAB on calcium mobilization Homo sapiens R-HSA-2730905 | 0.001451 | 0.007846 | 6.481384 | 42.358891 | FGF9;ERBB4;EGF;PDGFA;FGFR3 |
| 58 | Signaling by VEGF Homo sapiens R-HSA-194138 | 0.001461 | 0.007846 | 4.493819 | 29.339024 | FGF9;ERBB4;EGF;PDGFA;NEFL;FGFR3;DUSP9 |
| 59 | Downstream signaling of activated FGFR2 Homo sapiens R-HSA-5654696 | 0.001486 | 0.007846 | 4.479634 | 29.168544 | FGF9;ERBB4;EGF;PDGFA;NEFL;FGFR3;DUSP9 |

| | | | | | | |
|---|---|---|---|---|---|---|
| 60 | Downstream signaling of activated FGFR4 Homo sapiens R-HSA-5654716 | 0.001486 | 0.007846 | 4.479634 | 29.168544 | FGF9;ERBB4;EGF;PDGFA;NEFL;FGFR3;DUSP9 |
| 61 | Downstream signaling of activated FGFR3 Homo sapiens R-HSA-5654708 | 0.001486 | 0.007846 | 4.479634 | 29.168544 | FGF9;ERBB4;EGF;PDGFA;NEFL;FGFR3;DUSP9 |
| 62 | Signaling by FGFR4 Homo sapiens R-HSA-5654743 | 0.001565 | 0.008001 | 4.437603 | 28.665188 | FGF9;ERBB4;EGF;PDGFA;NEFL;FGFR3;DUSP9 |
| 63 | Downstream signaling of activated FGFR1 Homo sapiens R-HSA-5654687 | 0.001565 | 0.008001 | 4.437603 | 28.665188 | FGF9;ERBB4;EGF;PDGFA;NEFL;FGFR3;DUSP9 |
| 64 | Signaling by FGFR3 Homo sapiens R-HSA-5654741 | 0.001592 | 0.008012 | 4.423765 | 28.50005 | FGF9;ERBB4;EGF;PDGFA;NEFL;FGFR3;DUSP9 |
| 65 | FGFR3 mutant receptor activation Homo sapiens R-HSA-2033514 | 0.001644 | 0.008021 | 39.776 | 254.98533 | FGF9;FGFR3 |
| 66 | Signaling by activated point mutants of FGFR3 Homo sapiens R-HSA-1839130 | 0.001644 | 0.008021 | 39.776 | 254.98533 | FGF9;FGFR3 |
| 67 | Signaling by FGFR1 Homo sapiens R-HSA-5654736 | 0.001676 | 0.008054 | 4.382755 | 28.012383 | FGF9;ERBB4;EGF;PDGFA;NEFL;FGFR3;DUSP9 |
| 68 | Downstream signal transduction Homo sapiens R-HSA-186763 | 0.001822 | 0.008628 | 4.316042 | 27.224635 | FGF9;ERBB4;EGF;PDGFA;NEFL;FGFR3;DUSP9 |
| 69 | DAP12 signaling Homo sapiens R-HSA-2424491 | 0.001915 | 0.008661 | 4.276964 | 26.766442 | FGF9;ERBB4;EGF;PDGFA;NEFL;FGFR3;DUSP9 |
| 70 | FGFR3c ligand binding and activation Homo sapiens R-HSA-190372 | 0.001937 | 0.008661 | 36.15818 | 225.87306 | FGF9;FGFR3 |
| 71 | FGFR3 ligand binding and activation Homo sapiens R-HSA-190239 | 0.001937 | 0.008661 | 36.15818 | 225.87306 | FGF9;FGFR3 |
| 72 | ERBB2 Activates PTK6 Signaling Homo sapiens R-HSA-8847993 | 0.001937 | 0.008661 | 36.15818 | 225.87306 | EGF;ERBB4 |
| 73 | Signalling by NGF Homo sapiens R-HSA-166520 | 0.002141 | 0.009443 | 3.746221 | 23.026588 | NTRK2;FGF9;ERBB4;EGF;PDGFA;NEFL;FGFR3;DUSP9 |
| 74 | Metabolism of fat-soluble vitamins Homo sapiens R-HSA-6806667 | 0.002246 | 0.009771 | 12.53157 | 76.427822 | GPC3;LPL;APOE |

| | | | | | | |
|---|---|---|---|---|---|---|
| 75 | Signaling by EGFR Homo sapiens R-HSA-177929 | 0.002285 | 0.009811 | 4.139443 | 25.173365 | FGF9;ERBB4;EGF;PDGFA;NEFL;FGFR3;DUSP9 |
| 76 | DAP12 interactions Homo sapiens R-HSA-2172127 | 0.002433 | 0.010307 | 4.091567 | 24.625959 | FGF9;ERBB4;EGF;PDGFA;NEFL;FGFR3;DUSP9 |
| 77 | Signaling by FGFR2 Homo sapiens R-HSA-5654738 | 0.002509 | 0.010494 | 4.068034 | 24.358283 | FGF9;ERBB4;EGF;PDGFA;NEFL;FGFR3;DUSP9 |
| 78 | ERBB2 Regulates Cell Motility Homo sapiens R-HSA-6785631 | 0.00259 | 0.010691 | 30.59231 | 182.21389 | EGF;ERBB4 |
| 79 | Signaling by FGFR Homo sapiens R-HSA-190236 | 0.002709 | 0.011041 | 4.01035 | 23.70605 | FGF9;ERBB4;EGF;PDGFA;NEFL;FGFR3;DUSP9 |
| 80 | PI3K events in ERBB2 signaling Homo sapiens R-HSA-1963642 | 0.00295 | 0.011727 | 28.40571 | 165.49141 | EGF;ERBB4 |
| 81 | GRB2 events in ERBB2 signaling Homo sapiens R-HSA-1963640 | 0.00295 | 0.011727 | 28.40571 | 165.49141 | EGF;ERBB4 |
| 82 | Downstream signaling events of B Cell Receptor (BCR) Homo sapiens R-HSA-1168372 | 0.00304 | 0.011937 | 5.43332 | 31.491186 | FGF9;ERBB4;EGF;PDGFA;FGFR3 |
| 83 | Extracellular matrix organization Homo sapiens R-HSA-1474244 | 0.003298 | 0.012773 | 4.427121 | 25.298525 | DST;COL6A2;SPP1;PDGFA;LTBP2;BMP7 |
| 84 | Chylomicron-mediated lipid transport Homo sapiens R-HSA-174800 | 0.003332 | 0.012773 | 26.51067 | 151.22075 | LPL;APOE |
| 85 | SHC-mediated cascade:FGFR3 Homo sapiens R-HSA-5654704 | 0.003736 | 0.013989 | 24.8525 | 138.91762 | FGF9;FGFR3 |
| 86 | SHC1 events in ERBB2 signaling Homo sapiens R-HSA-1250196 | 0.003736 | 0.013989 | 24.8525 | 138.91762 | EGF;ERBB4 |
| 87 | Signaling by Interleukins Homo sapiens R-HSA-449147 | 0.003951 | 0.014621 | 3.734545 | 20.666631 | FGF9;ERBB4;EGF;PDGFA;NEFL;FGFR3;DUSP9 |
| 88 | Cytokine Signaling in Immune system Homo sapiens R-HSA-1280215 | 0.004425 | 0.016109 | 3.054802 | 16.558443 | EGR1;MT2A;FGF9;ERBB4;EGF;PDGFA;NEFL;FGFR3;DUSP9 |
| 89 | Neuronal System Homo sapiens R-HSA-112316 | 0.004452 | 0.016109 | 4.153178 | 22.486595 | KCNJ6;SLC22A2;KCNMA1;NEFL;KCNJ1;NCALD |
| 90 | Signaling by FGFR3 point mutants in cancer Homo sapiens R-HSA-8853338 | 0.005567 | 0.019697 | 19.878 | 103.18609 | FGF9;FGFR3 |

| | | | | | | |
|---|---|---|---|---|---|---|
| 91 | Signaling by FGFR3 in disease Homo sapiens R-HSA-5655332 | 0.005567 | 0.019697 | 19.878 | 103.18609 | FGF9;FGFR3 |
| 92 | Lipid digestion, mobilization, and transport Homo sapiens R-HSA-73923 | 0.005726 | 0.020042 | 8.836898 | 45.622096 | FABP3;LPL;APOE |
| 93 | Signaling by the B Cell Receptor (BCR) Homo sapiens R-HSA-983705 | 0.00686 | 0.023752 | 4.447007 | 22.155191 | FGF9;ERBB4;EGF;PDGFA;FGFR3 |
| 94 | PI3K Cascade Homo sapiens R-HSA-109704 | 0.007694 | 0.026357 | 7.903509 | 38.468719 | FGF9;RRAGD;FGFR3 |
| 95 | Negative regulation of FGFR3 signaling Homo sapiens R-HSA-5654732 | 0.00956 | 0.032402 | 14.71926 | 68.447485 | FGF9;FGFR3 |
| 96 | Cell junction organization Homo sapiens R-HSA-446728 | 0.009706 | 0.032555 | 7.234392 | 33.531636 | CLDN10;DST;CLDN16 |
| 97 | Tight junction interactions Homo sapiens R-HSA-420029 | 0.010209 | 0.033543 | 14.19286 | 65.067301 | CLDN10;CLDN16 |
| 98 | Molecules associated with elastic fibres Homo sapiens R-HSA-2129379 | 0.010209 | 0.033543 | 14.19286 | 65.067301 | LTBP2;BMP7 |
| 99 | HS-GAG biosynthesis Homo sapiens R-HSA-2022928 | 0.010877 | 0.035023 | 13.70276 | 61.951842 | GPC3;HS6ST2 |
| 100 | Inwardly rectifying K+ channels Homo sapiens R-HSA-1296065 | 0.010877 | 0.035023 | 13.70276 | 61.951842 | KCNJ6;KCNJ1 |
| 101 | Signal Transduction Homo sapiens R-HSA-162582 | 0.011996 | 0.038244 | 1.851518 | 8.1896583 | NTRK2;CASR;EGF;PTGER3;LPL;PDGFA;AVPR1A;DUSP9;KNG1;SFRP1;CXCL12;FGF9;ERBB4;COL6A2;RRAGD;GPC3;SPP1;NEFL;APOE;FGFR3;SOX4 |
| 102 | Lipoprotein metabolism Homo sapiens R-HSA-174824 | 0.012993 | 0.041017 | 12.41625 | 53.927986 | LPL;APOE |
| 103 | Visual phototransduction Homo sapiens R-HSA-2187338 | 0.013433 | 0.041995 | 6.384268 | 27.516433 | GPC3;LPL;APOE |
| 104 | Transport of inorganic cations/anions and amino acids/oligopeptides Homo sapiens R-HSA-425393 | 0.014187 | 0.043505 | 6.250631 | 26.599308 | SLC12A3;SLC12A1;SLC8A1 |

| | | | | | | |
|---|---|---|---|---|---|---|
| 105 | Potassium Channels Homo sapiens R-HSA-1296071 | 0.014187 | 0.043505 | 6.250631 | 26.599308 | KCNJ6;KCNMA1;KCNJ1 |
| 106 | Diseases of signal transduction Homo sapiens R-HSA-5663202 | 0.016053 | 0.048765 | 3.57273 | 14.762025 | FGF9;ERBB4;EGF;PDGFA;FGFR3 |
| 107 | Platelet degranulation Homo sapiens R-HSA-114608 | 0.016589 | 0.049922 | 5.881165 | 24.106953 | EGF;PDGFA;KNG1 |
| 108 | Nuclear signaling by ERBB4 Homo sapiens R-HSA-1251985 | 0.016882 | 0.050332 | 10.73568 | 43.818036 | CXCL12;ERBB4 |
| 109 | Elastic fibre formation Homo sapiens R-HSA-1566948 | 0.018559 | 0.054825 | 10.1841 | 40.602106 | LTBP2;BMP7 |
| 110 | Response to elevated platelet cytosolic Ca2+ Homo sapiens R-HSA-76005 | 0.018755 | 0.054901 | 5.604928 | 22.286879 | EGF;PDGFA;KNG1 |
| 111 | TP53 Regulates Transcription of Cell Death Genes Homo sapiens R-HSA-5633008 | 0.020303 | 0.058898 | 9.686341 | 37.747381 | IGFBP3;NDRG1 |
| 112 | Phospholipase C-mediated cascade; FGFR3 Homo sapiens R-HSA-5654227 | 0.0212 | 0.060951 | 9.455238 | 36.43799 | FGF9;FGFR3 |
| 113 | Innate Immune System Homo sapiens R-HSA-168249 | 0.022094 | 0.062461 | 2.316275 | 8.8306413 | FGF9;ERBB4;EGF;PDGFA;NEFL;DEFB1;FOS;FGFR3;DUSP9 |
| 114 | Signaling by ERBB2 Homo sapiens R-HSA-1227986 | 0.022114 | 0.062461 | 9.234884 | 35.199296 | ERBB4;EGF |
| 115 | Transmission across Chemical Synapses Homo sapiens R-HSA-112315 | 0.022951 | 0.064262 | 3.882678 | 14.654818 | KCNJ6;SLC22A2;NEFL;NCALD |
| 116 | Abacavir transmembrane transport Homo sapiens R-HSA-2161517 | 0.025244 | 0.068885 | 49.24257 | 181.17252 | SLC22A2 |
| 117 | NGF-independant TRKA activation Homo sapiens R-HSA-187024 | 0.025244 | 0.068885 | 49.24257 | 181.17252 | NTRK2 |
| 118 | Trafficking of myristoylated proteins to the cilium Homo sapiens R-HSA-5624138 | 0.025244 | 0.068885 | 49.24257 | 181.17252 | CYS1 |
| 119 | Ion homeostasis Homo sapiens R-HSA-5578775 | 0.027925 | 0.075561 | 8.101633 | 28.989599 | FXYD2;SLC8A1 |

| | | | | | | |
|---|---|---|---|---|---|---|
| 120 | Cell-Cell communication Homo sapiens R-HSA-1500931 | 0.029487 | 0.079125 | 4.680398 | 16.492735 | CLDN10;DST;CLDN16 |
| 121 | Vasopressin-like receptors Homo sapiens R-HSA-388479 | 0.030216 | 0.079751 | 39.39208 | 137.84795 | AVPR1A |
| 122 | Activation of TRKA receptors Homo sapiens R-HSA-187015 | 0.030216 | 0.079751 | 39.39208 | 137.84795 | NTRK2 |
| 123 | Heparan sulfate/heparin (HS-GAG) metabolism Homo sapiens R-HSA-1638091 | 0.031035 | 0.080591 | 7.633077 | 26.506901 | GPC3;HS6ST2 |
| 124 | Assembly of collagen fibrils and other multimeric structures Homo sapiens R-HSA-2022090 | 0.031035 | 0.080591 | 7.633077 | 26.506901 | DST;COL6A2 |
| 125 | Signaling by GPCR Homo sapiens R-HSA-372790 | 0.031684 | 0.081617 | 1.937757 | 6.6890502 | CASR;CXCL12;FGF9;ERBB4;EGF;PTGER3;PDGFA;NEFL;AVPR1A;FGFR3;DUSP9;KNG1 |
| 126 | G alpha (i) signalling events Homo sapiens R-HSA-418594 | 0.034522 | 0.088223 | 3.400553 | 11.446794 | CASR;CXCL12;PTGER3;KNG1 |
| 127 | SLC transporter disorders Homo sapiens R-HSA-5619102 | 0.035163 | 0.088458 | 32.82508 | 109.89015 | AVPR1A |
| 128 | Phosphate bond hydrolysis by NUDT proteins Homo sapiens R-HSA-2393930 | 0.035163 | 0.088458 | 32.82508 | 109.89015 | NUDT16 |
| 129 | Neurotransmitter Receptor Binding And Downstream Transmission In The Postsynaptic Cell Homo sapiens R-HSA-112314 | 0.03616 | 0.090261 | 4.307608 | 14.300374 | KCNJ6;NEFL;NCALD |
| 130 | Cell-cell junction organization Homo sapiens R-HSA-421270 | 0.038791 | 0.096082 | 6.725085 | 21.853605 | CLDN10;CLDN16 |
| 131 | Neurotransmitter Clearance In The Synaptic Cleft Homo sapiens R-HSA-112311 | 0.040086 | 0.097085 | 28.13437 | 90.500778 | SLC22A2 |
| 132 | Downregulation of ERBB4 signaling Homo sapiens R-HSA-1253288 | 0.040086 | 0.097085 | 28.13437 | 90.500778 | ERBB4 |

| | | | | | | |
|---|---|---|---|---|---|---|
| 133 | Metabolism of vitamins and cofactors Homo sapiens R-HSA-196854 | 0.0401 | 0.097085 | 4.128109 | 13.277547 | GPC3;LPL;APOE |
| 134 | Signaling by FGFR in disease Homo sapiens R-HSA-1226099 | 0.041129 | 0.098833 | 6.503934 | 20.754301 | FGF9;FGFR3 |
| 135 | Organic cation transport Homo sapiens R-HSA-549127 | 0.044983 | 0.103462 | 24.61634 | 76.346689 | SLC22A2 |
| 136 | Role of Abl in Robo-Slit signaling Homo sapiens R-HSA-428890 | 0.044983 | 0.103462 | 24.61634 | 76.346689 | SLIT2 |
| 137 | Inactivation of Cdc42 and Rac Homo sapiens R-HSA-428543 | 0.044983 | 0.103462 | 24.61634 | 76.346689 | SLIT2 |
| 138 | Type I hemidesmosome assembly Homo sapiens R-HSA-446107 | 0.044983 | 0.103462 | 24.61634 | 76.346689 | DST |
| 139 | Ca2+ activated K+ channels Homo sapiens R-HSA-1296052 | 0.044983 | 0.103462 | 24.61634 | 76.346689 | KCNMA1 |
| 140 | Prostanoid ligand receptors Homo sapiens R-HSA-391908 | 0.044983 | 0.103462 | 24.61634 | 76.346689 | PTGER3 |
| 141 | Signaling by PTK6 Homo sapiens R-HSA-8848021 | 0.04596 | 0.104958 | 6.102462 | 18.795515 | ERBB4;EGF |
| 142 | SLC-mediated transmembrane transport Homo sapiens R-HSA-425407 | 0.048405 | 0.108471 | 3.03556 | 9.1921152 | SLC12A3;SLC22A2;SLC12A1;SLC8A1 |
| 143 | Abacavir transport and metabolism Homo sapiens R-HSA-2161522 | 0.049856 | 0.108471 | 21.88009 | 65.609969 | SLC22A2 |
| 144 | Activation of Ca-permeable Kainate Receptor Homo sapiens R-HSA-451308 | 0.049856 | 0.108471 | 21.88009 | 65.609969 | NCALD |
| 145 | Ionotropic activity of Kainate Receptors Homo sapiens R-HSA-451306 | 0.049856 | 0.108471 | 21.88009 | 65.609969 | NCALD |
| 146 | Regulation of signaling by NODAL Homo sapiens R-HSA-1433617 | 0.049856 | 0.108471 | 21.88009 | 65.609969 | TDGF1 |
| 147 | Signaling by FGFR3 fusions in cancer Homo sapiens R-HSA-8853334 | 0.049856 | 0.108471 | 21.88009 | 65.609969 | FGFR3 |

| | | | | | | |
|---|---|---|---|---|---|---|
| 148 | Activation of the AP-1 family of transcription factors Homo sapiens R-HSA-450341 | 0.049856 | 0.108471 | 21.88009 | 65.609969 | FOS |
| 149 | Sodium/Calcium exchangers Homo sapiens R-HSA-425561 | 0.054704 | 0.116654 | 19.69109 | 57.218659 | SLC8A1 |
| 150 | FGFR1c ligand binding and activation Homo sapiens R-HSA-190373 | 0.054704 | 0.116654 | 19.69109 | 57.218659 | FGF9 |
| 151 | Signaling by activated point mutants of FGFR1 Homo sapiens R-HSA-1839122 | 0.054704 | 0.116654 | 19.69109 | 57.218659 | FGF9 |
| 152 | Defective EXT2 causes exostoses 2 Homo sapiens R-HSA-3656237 | 0.059528 | 0.123664 | 17.90009 | 50.501721 | GPC3 |
| 153 | Defective EXT1 causes exostoses 1, TRPS2 and CHDS Homo sapiens R-HSA-3656253 | 0.059528 | 0.123664 | 17.90009 | 50.501721 | GPC3 |
| 154 | Reduction of cytosolic Ca++ levels Homo sapiens R-HSA-418359 | 0.059528 | 0.123664 | 17.90009 | 50.501721 | SLC8A1 |
| 155 | TP53 Regulates Transcription of Death Receptors and Ligands Homo sapiens R-HSA-6803211 | 0.059528 | 0.123664 | 17.90009 | 50.501721 | IGFBP3 |
| 156 | Hemostasis Homo sapiens R-HSA-109582 | 0.063255 | 0.130273 | 2.215201 | 6.1152433 | PLAU;EGF;KCNMA1;PDGFA;SLC8A1;KNG1 |
| 157 | Dissolution of Fibrin Clot Homo sapiens R-HSA-75205 | 0.064327 | 0.130273 | 16.40759 | 45.018719 | PLAU |
| 158 | FGFR2c ligand binding and activation Homo sapiens R-HSA-190375 | 0.064327 | 0.130273 | 16.40759 | 45.018719 | FGF9 |
| 159 | POU5F1 (OCT4), SOX2, NANOG activate genes related to proliferation Homo sapiens R-HSA-2892247 | 0.064327 | 0.130273 | 16.40759 | 45.018719 | TDGF1 |
| 160 | Organic cation/anion/zwitterion transport Homo sapiens R-HSA-549132 | 0.069102 | 0.135676 | 15.14471 | 40.469222 | SLC22A2 |

| | | | | | | |
|---|---|---|---|---|---|---|
| 161 | SEMA3A-Plexin repulsion signaling by inhibiting Integrin adhesion Homo sapiens R-HSA-399955 | 0.069102 | 0.135676 | 15.14471 | 40.469222 | PLXNA2 |
| 162 | Activation of Rac Homo sapiens R-HSA-428540 | 0.069102 | 0.135676 | 15.14471 | 40.469222 | SLIT2 |
| 163 | FGFR4 ligand binding and activation Homo sapiens R-HSA-190322 | 0.069102 | 0.135676 | 15.14471 | 40.469222 | FGF9 |
| 164 | TP53 regulates transcription of several additional cell death genes whose specific roles in p53-dependent apoptosis remain uncertain Homo sapiens R-HSA-6803205 | 0.069102 | 0.135676 | 15.14471 | 40.469222 | NDRG1 |
| 165 | Collagen formation Homo sapiens R-HSA-1474290 | 0.070002 | 0.13661 | 4.774699 | 12.69704 | DST;COL6A2 |
| 166 | Platelet homeostasis Homo sapiens R-HSA-418346 | 0.072883 | 0.13739 | 4.661882 | 12.209024 | KCNMA1;SLC8A1 |
| 167 | Negative regulation of TCF-dependent signaling by WNT ligand antagonists Homo sapiens R-HSA-3772470 | 0.073853 | 0.13739 | 14.06223 | 36.641656 | SFRP1 |
| 168 | Disorders of transmembrane transporters Homo sapiens R-HSA-5619115 | 0.073853 | 0.13739 | 14.06223 | 36.641656 | AVPR1A |
| 169 | CREB phosphorylation through the activation of CaMKII Homo sapiens R-HSA-442729 | 0.073853 | 0.13739 | 14.06223 | 36.641656 | NEFL |
| 170 | FGFR1 ligand binding and activation Homo sapiens R-HSA-190242 | 0.073853 | 0.13739 | 14.06223 | 36.641656 | FGF9 |
| 171 | Eicosanoid ligand-binding receptors Homo sapiens R-HSA-391903 | 0.073853 | 0.13739 | 14.06223 | 36.641656 | PTGER3 |
| 172 | Signaling by EGFRvIII in Cancer Homo sapiens R-HSA-5637812 | 0.073853 | 0.13739 | 14.06223 | 36.641656 | EGF |

| | | | | | | |
|---|---|---|---|---|---|---|
| 173 | Constitutive Signaling by EGFRvIII Homo sapiens R-HSA-5637810 | 0.073853 | 0.13739 | 14.06223 | 36.641656 | EGF |
| 174 | G alpha (q) signalling events Homo sapiens R-HSA-416476 | 0.074242 | 0.13739 | 3.176983 | 8.2615179 | CASR;AVPR1A;KNG1 |
| 175 | Peptide ligand-binding receptors Homo sapiens R-HSA-375276 | 0.076068 | 0.139965 | 3.143222 | 8.097341 | CXCL12;AVPR1A;KNG1 |
| 176 | Disease Homo sapiens R-HSA-1643685 | 0.077327 | 0.141467 | 1.968333 | 5.038378 | FGF9;ERBB4;EGF;GPC3;PDGFA;AVPR1A;FGFR3 |
| 177 | CRMPs in Sema3A signaling Homo sapiens R-HSA-399956 | 0.07858 | 0.141467 | 13.12409 | 33.382952 | PLXNA2 |
| 178 | Sema3A PAK dependent Axon repulsion Homo sapiens R-HSA-399954 | 0.07858 | 0.141467 | 13.12409 | 33.382952 | PLXNA2 |
| 179 | GPCR ligand binding Homo sapiens R-HSA-500792 | 0.078641 | 0.141467 | 2.268974 | 5.7696756 | CASR;CXCL12;PTGER3;AVPR1A;KNG1 |
| 180 | Ras activation uopn Ca2+ infux through NMDA receptor Homo sapiens R-HSA-442982 | 0.083283 | 0.146714 | 12.30322 | 30.579792 | NEFL |
| 181 | Unblocking of NMDA receptor, glutamate binding and activation Homo sapiens R-HSA-438066 | 0.083283 | 0.146714 | 12.30322 | 30.579792 | NEFL |
| 182 | Activated point mutants of FGFR2 Homo sapiens R-HSA-2033519 | 0.083283 | 0.146714 | 12.30322 | 30.579792 | FGF9 |
| 183 | Class A/1 (Rhodopsin-like receptors) Homo sapiens R-HSA-373076 | 0.083381 | 0.146714 | 2.50515 | 6.2236389 | CXCL12;PTGER3;AVPR1A;KNG1 |
| 184 | Norepinephrine Neurotransmitter Release Cycle Homo sapiens R-HSA-181430 | 0.087962 | 0.152158 | 11.57892 | 28.146594 | SLC22A2 |
| 185 | cGMP effects Homo sapiens R-HSA-418457 | 0.087962 | 0.152158 | 11.57892 | 28.146594 | KCNMA1 |
| 186 | Na+/Cl- dependent neurotransmitter transporters Homo sapiens R-HSA-442660 | 0.092618 | 0.152158 | 10.93509 | 26.017591 | SLC22A2 |
| 187 | Other semaphorin interactions Homo sapiens R-HSA-416700 | 0.092618 | 0.152158 | 10.93509 | 26.017591 | PLXNA2 |

| | | | | | | |
|---|---|---|---|---|---|---|
| 188 | Defective B3GALT6 causes EDSP2 and SEMDJL1 Homo sapiens R-HSA-4420332 | 0.092618 | 0.152158 | 10.93509 | 26.017591 | GPC3 |
| 189 | Defective B4GALT7 causes EDS, progeroid type Homo sapiens R-HSA-3560783 | 0.092618 | 0.152158 | 10.93509 | 26.017591 | GPC3 |
| 190 | Defective B3GAT3 causes JDSSDHD Homo sapiens R-HSA-3560801 | 0.092618 | 0.152158 | 10.93509 | 26.017591 | GPC3 |
| 191 | Scavenging by Class A Receptors Homo sapiens R-HSA-3000480 | 0.092618 | 0.152158 | 10.93509 | 26.017591 | APOE |
| 192 | HDL-mediated lipid transport Homo sapiens R-HSA-194223 | 0.092618 | 0.152158 | 10.93509 | 26.017591 | APOE |
| 193 | Signaling by NODAL Homo sapiens R-HSA-1181150 | 0.092618 | 0.152158 | 10.93509 | 26.017591 | TDGF1 |
| 194 | Constitutive Signaling by Ligand-Responsive EGFR Cancer Variants Homo sapiens R-HSA-1236382 | 0.092618 | 0.152158 | 10.93509 | 26.017591 | EGF |
| 195 | Signaling by Ligand-Responsive EGFR Variants in Cancer Homo sapiens R-HSA-5637815 | 0.092618 | 0.152158 | 10.93509 | 26.017591 | EGF |
| 196 | Signaling by EGFR in Cancer Homo sapiens R-HSA-1643713 | 0.092618 | 0.152158 | 10.93509 | 26.017591 | EGF |
| 197 | FGFR2 ligand binding and activation Homo sapiens R-HSA-190241 | 0.09725 | 0.158154 | 10.35904 | 24.141462 | FGF9 |
| 198 | Hormone-sensitive lipase (HSL)-mediated triacylglycerol hydrolysis Homo sapiens R-HSA-163560 | 0.09725 | 0.158154 | 10.35904 | 24.141462 | FABP3 |
| 199 | Regulation of Insulin-like Growth Factor (IGF) transport and uptake by Insulin-like Growth Factor Binding Proteins (IGFBPs) Homo sapiens R-HSA-381426 | 0.101858 | 0.163176 | 9.840594 | 22.477606 | IGFBP3 |

| | | | | | | |
|---|---|---|---|---|---|---|
| 200 | HS-GAG degradation Homo sapiens R-HSA-2024096 | 0.101858 | 0.163176 | 9.840594 | 22.477606 | GPC3 |
| 201 | SHC-mediated cascade:FGFR1 Homo sapiens R-HSA-5654688 | 0.101858 | 0.163176 | 9.840594 | 22.477606 | FGF9 |
| 202 | Transcriptional Regulation by TP53 Homo sapiens R-HSA-3700989 | 0.102485 | 0.163367 | 2.320123 | 5.2853306 | IGFBP3;RRAGD;FOS;NDRG1 |
| 203 | Post-chaperonin tubulin folding pathway Homo sapiens R-HSA-389977 | 0.106444 | 0.167195 | 9.371523 | 20.993512 | TUBB2B |
| 204 | Intrinsic Pathway of Fibrin Clot Formation Homo sapiens R-HSA-140837 | 0.106444 | 0.167195 | 9.371523 | 20.993512 | KNG1 |
| 205 | mTORC1-mediated signalling Homo sapiens R-HSA-166208 | 0.106444 | 0.167195 | 9.371523 | 20.993512 | RRAGD |
| 206 | RAF-independent MAPK1/3 activation Homo sapiens R-HSA-112409 | 0.111006 | 0.172676 | 8.945095 | 19.662858 | DUSP9 |
| 207 | SHC-mediated cascade:FGFR4 Homo sapiens R-HSA-5654719 | 0.111006 | 0.172676 | 8.945095 | 19.662858 | FGF9 |
| 208 | Formation of tubulin folding intermediates by CCT/TriC Homo sapiens R-HSA-389960 | 0.120061 | 0.182357 | 8.198845 | 17.379539 | TUBB2B |
| 209 | Activation of G protein gated Potassium channels Homo sapiens R-HSA-1296041 | 0.120061 | 0.182357 | 8.198845 | 17.379539 | KCNJ6 |
| 210 | Inhibition of voltage gated Ca2+ channels via Gbeta/gamma subunits Homo sapiens R-HSA-997272 | 0.120061 | 0.182357 | 8.198845 | 17.379539 | KCNJ6 |
| 211 | G protein gated Potassium channels Homo sapiens R-HSA-1296059 | 0.120061 | 0.182357 | 8.198845 | 17.379539 | KCNJ6 |
| 212 | Nitric oxide stimulates guanylate cyclase Homo sapiens R-HSA-392154 | 0.120061 | 0.182357 | 8.198845 | 17.379539 | KCNMA1 |

| | | | | | | |
|---|---|---|---|---|---|---|
| 213 | Diseases associated with glycosaminoglycan metabolism Homo sapiens R-HSA-3560782 | 0.124554 | 0.186542 | 7.870495 | 16.394342 | GPC3 |
| 214 | A tetrasaccharide linker sequence is required for GAG synthesis Homo sapiens R-HSA-1971475 | 0.124554 | 0.186542 | 7.870495 | 16.394342 | GPC3 |
| 215 | SHC-mediated cascade:FGFR2 Homo sapiens R-HSA-5654699 | 0.124554 | 0.186542 | 7.870495 | 16.394342 | FGF9 |
| 216 | Glycosaminoglycan metabolism Homo sapiens R-HSA-1630316 | 0.126841 | 0.188846 | 3.324202 | 6.8638704 | GPC3;HS6ST2 |
| 217 | Prefoldin mediated transfer of substrate to CCT/TriC Homo sapiens R-HSA-389957 | 0.129025 | 0.188846 | 7.567403 | 15.496144 | TUBB2B |
| 218 | CREB phosphorylation through the activation of Ras Homo sapiens R-HSA-442742 | 0.129025 | 0.188846 | 7.567403 | 15.496144 | NEFL |
| 219 | Platelet calcium homeostasis Homo sapiens R-HSA-418360 | 0.129025 | 0.188846 | 7.567403 | 15.496144 | SLC8A1 |
| 220 | EGFR downregulation Homo sapiens R-HSA-182971 | 0.129025 | 0.188846 | 7.567403 | 15.496144 | EGF |
| 221 | Energy dependent regulation of mTOR by LKB1-AMPK Homo sapiens R-HSA-380972 | 0.133473 | 0.194472 | 7.286762 | 14.674493 | RRAGD |
| 222 | Platelet activation, signaling and aggregation Homo sapiens R-HSA-76002 | 0.139192 | 0.201892 | 2.381576 | 4.6962246 | EGF;PDGFA;KNG1 |
| 223 | Amine compound SLC transporters Homo sapiens R-HSA-425428 | 0.142302 | 0.203649 | 6.783544 | 13.226602 | SLC22A2 |
| 224 | Activation of Kainate Receptors upon glutamate binding Homo sapiens R-HSA-451326 | 0.142302 | 0.203649 | 6.783544 | 13.226602 | NCALD |
| 225 | MAPK targets/ Nuclear events mediated by MAP kinases Homo sapiens R-HSA-450282 | 0.142302 | 0.203649 | 6.783544 | 13.226602 | FOS |

| | | | | | | |
|---|---|---|---|---|---|---|
| 226 | Negative regulation of FGFR4 signaling Homo sapiens R-HSA-5654733 | 0.146682 | 0.208069 | 6.557096 | 12.586253 | FGF9 |
| 227 | FGFR1 mutant receptor activation Homo sapiens R-HSA-1839124 | 0.146682 | 0.208069 | 6.557096 | 12.586253 | FGF9 |
| 228 | Signaling by Robo receptor Homo sapiens R-HSA-376176 | 0.151041 | 0.210717 | 6.345257 | 11.99383 | SLIT2 |
| 229 | Cooperation of Prefoldin and TriC/CCT in actin and tubulin folding Homo sapiens R-HSA-389958 | 0.151041 | 0.210717 | 6.345257 | 11.99383 | TUBB2B |
| 230 | Negative regulation of FGFR1 signaling Homo sapiens R-HSA-5654726 | 0.151041 | 0.210717 | 6.345257 | 11.99383 | FGF9 |
| 231 | Cardiac conduction Homo sapiens R-HSA-5576891 | 0.151166 | 0.210717 | 2.97218 | 5.6155608 | FXYD2;SLC8A1 |
| 232 | FGFR2 mutant receptor activation Homo sapiens R-HSA-1839126 | 0.155378 | 0.214728 | 6.146658 | 11.444444 | FGF9 |
| 233 | Detoxification of Reactive Oxygen Species Homo sapiens R-HSA-3299685 | 0.155378 | 0.214728 | 6.146658 | 11.444444 | GPX3 |
| 234 | EGFR interacts with phospholipase C-gamma Homo sapiens R-HSA-212718 | 0.159692 | 0.218812 | 5.960096 | 10.933834 | EGF |
| 235 | Negative regulation of FGFR2 signaling Homo sapiens R-HSA-5654727 | 0.159692 | 0.218812 | 5.960096 | 10.933834 | FGF9 |
| 236 | Post NMDA receptor activation events Homo sapiens R-HSA-438064 | 0.163985 | 0.223403 | 5.784508 | 10.45827 | NEFL |
| 237 | Immune System Homo sapiens R-HSA-168256 | 0.16443 | 0.223403 | 1.445041 | 2.6086877 | EGR1;MT2A;FGF9;ERBB4;EGF;PDGFA;NEFL;DEFB1;FOS;FGFR3;DUSP9 |
| 238 | NCAM1 interactions Homo sapiens R-HSA-419037 | 0.172506 | 0.232665 | 5.462596 | 9.599552 | COL6A2 |
| 239 | Metabolism Homo sapiens R-HSA-1430728 | 0.172692 | 0.232665 | 1.387679 | 2.437103 | SLC22A2;SCD5;LPL;HS6ST2;MT2A;FABP3;MT1F;GPC3;MT1G;MT1H;NUDT16;APOE;MT1E |
| 240 | Beta defensins Homo sapiens R-HSA-1461957 | 0.176734 | 0.236134 | 5.314691 | 9.2109445 | DEFB1 |
| 241 | Signaling by FGFR1 in disease Homo sapiens R-HSA-5655302 | 0.176734 | 0.236134 | 5.314691 | 9.2109445 | FGF9 |

| | | | | | | |
|---|---|---|---|---|---|---|
| 242 | GABA B receptor activation Homo sapiens R-HSA-977444 | 0.180941 | 0.236841 | 5.17457 | 8.8463769 | KCNJ6 |
| 243 | Activation of GABAB receptors Homo sapiens R-HSA-991365 | 0.180941 | 0.236841 | 5.17457 | 8.8463769 | KCNJ6 |
| 244 | Activation of NMDA receptor upon glutamate binding and postsynaptic events Homo sapiens R-HSA-442755 | 0.180941 | 0.236841 | 5.17457 | 8.8463769 | NEFL |
| 245 | Formation of Fibrin Clot (Clotting Cascade) Homo sapiens R-HSA-140877 | 0.180941 | 0.236841 | 5.17457 | 8.8463769 | KNG1 |
| 246 | mTOR signalling Homo sapiens R-HSA-165159 | 0.180941 | 0.236841 | 5.17457 | 8.8463769 | RRAGD |
| 247 | Negative regulation of MAPK pathway Homo sapiens R-HSA-5675221 | 0.185126 | 0.2394 | 5.041635 | 8.5038223 | DUSP9 |
| 248 | Class C/3 (Metabotropic glutamate/pheromone receptors) Homo sapiens R-HSA-420499 | 0.185126 | 0.2394 | 5.041635 | 8.5038223 | CASR |
| 249 | PKB-mediated events Homo sapiens R-HSA-109703 | 0.185126 | 0.2394 | 5.041635 | 8.5038223 | RRAGD |
| 250 | Transmembrane transport of small molecules Homo sapiens R-HSA-382551 | 0.186749 | 0.240532 | 1.689829 | 2.8355178 | SLC12A3;SLC22A2;FXYD2;SLC12A1;SLC8A1 |
| 251 | Netrin-1 signaling Homo sapiens R-HSA-373752 | 0.193433 | 0.244257 | 4.795219 | 7.8776959 | SLIT2 |
| 252 | Non-integrin membrane-ECM interactions Homo sapiens R-HSA-3000171 | 0.193433 | 0.244257 | 4.795219 | 7.8776959 | PDGFA |
| 253 | Deactivation of the beta-catenin transactivating complex Homo sapiens R-HSA-3769402 | 0.193433 | 0.244257 | 4.795219 | 7.8776959 | SOX4 |
| 254 | Fatty Acyl-CoA Biosynthesis Homo sapiens R-HSA-75105 | 0.193433 | 0.244257 | 4.795219 | 7.8776959 | SCD5 |

| | | | | | | |
|---|---|---|---|---|---|---|
| 255 | Transcriptional regulation of pluripotent stem cells Homo sapiens R-HSA-452723 | 0.193433 | 0.244257 | 4.795219 | 7.8776959 | TDGF1 |
| 256 | Signaling by FGFR2 in disease Homo sapiens R-HSA-5655253 | 0.197555 | 0.248488 | 4.680811 | 7.5910424 | FGF9 |
| 257 | Phospholipase C-mediated cascade; FGFR4 Homo sapiens R-HSA-5654228 | 0.209797 | 0.262858 | 4.368097 | 6.821285 | FGF9 |
| 258 | Phospholipase C-mediated cascade: FGFR1 Homo sapiens R-HSA-5654219 | 0.213836 | 0.265851 | 4.272923 | 6.5911735 | FGF9 |
| 259 | Defensins Homo sapiens R-HSA-1461973 | 0.213836 | 0.265851 | 4.272923 | 6.5911735 | DEFB1 |
| 260 | Phospholipase C-mediated cascade; FGFR2 Homo sapiens R-HSA-5654221 | 0.221854 | 0.274757 | 4.094472 | 6.165198 | FGF9 |
| 261 | Chondroitin sulfate/dermatan sulfate metabolism Homo sapiens R-HSA-1793185 | 0.225832 | 0.278613 | 4.010709 | 5.9677905 | GPC3 |
| 262 | Neurotransmitter Release Cycle Homo sapiens R-HSA-112310 | 0.22979 | 0.28134 | 3.930297 | 5.7798506 | SLC22A2 |
| 263 | Cargo trafficking to the periciliary membrane Homo sapiens R-HSA-5620920 | 0.22979 | 0.28134 | 3.930297 | 5.7798506 | CYS1 |
| 264 | GABA receptor activation Homo sapiens R-HSA-977443 | 0.245423 | 0.298213 | 3.638431 | 5.1111603 | KCNJ6 |
| 265 | Ion transport by P-type ATPases Homo sapiens R-HSA-936837 | 0.245423 | 0.298213 | 3.638431 | 5.1111603 | FXYD2 |
| 266 | Chemokine receptors bind chemokines Homo sapiens R-HSA-380108 | 0.249282 | 0.301763 | 3.572097 | 4.9622479 | CXCL12 |
| 267 | Interferon Signaling Homo sapiens R-HSA-913531 | 0.264079 | 0.316642 | 2.03134 | 2.7047425 | EGR1;MT2A |
| 268 | Muscle contraction Homo sapiens R-HSA-397014 | 0.264079 | 0.316642 | 2.03134 | 2.7047425 | FXYD2;SLC8A1 |

| | | | | | | |
|---|---|---|---|---|---|---|
| 269 | MAP kinase activation in TLR cascade Homo sapiens R-HSA-450294 | 0.264524 | 0.316642 | 3.32925 | 4.4273186 | FOS |
| 270 | TP53 Regulates Transcription of DNA Repair Genes Homo sapiens R-HSA-6796648 | 0.268286 | 0.319956 | 3.273597 | 4.3070795 | FOS |
| 271 | TCF dependent signaling in response to WNT Homo sapiens R-HSA-201681 | 0.26975 | 0.320515 | 2.000102 | 2.6206523 | SFRP1;SOX4 |
| 272 | Collagen biosynthesis and modifying enzymes Homo sapiens R-HSA-1650814 | 0.275753 | 0.326443 | 3.167678 | 4.0807583 | COL6A2 |
| 273 | Triglyceride Biosynthesis Homo sapiens R-HSA-75109 | 0.286813 | 0.337649 | 3.021021 | 3.7730282 | SCD5 |
| 274 | Cellular responses to stress Homo sapiens R-HSA-2262752 | 0.288212 | 0.337649 | 1.626207 | 2.0230958 | GPX3;RRAGD;FOS |
| 275 | Semaphorin interactions Homo sapiens R-HSA-373755 | 0.290462 | 0.337649 | 2.975098 | 3.6780587 | PLXNA2 |
| 276 | Integrin cell surface interactions Homo sapiens R-HSA-216083 | 0.290462 | 0.337649 | 2.975098 | 3.6780587 | SPP1 |
| 277 | Macroautophagy Homo sapiens R-HSA-1632852 | 0.290462 | 0.337649 | 2.975098 | 3.6780587 | RRAGD |
| 278 | Interferon alpha/beta signaling Homo sapiens R-HSA-909733 | 0.294093 | 0.34064 | 2.930545 | 3.5865746 | EGR1 |
| 279 | TRAF6 Mediated Induction of proinflammatory cytokines Homo sapiens R-HSA-168180 | 0.308433 | 0.355969 | 2.764886 | 3.2521978 | FOS |
| 280 | Binding and Uptake of Ligands by Scavenger Receptors Homo sapiens R-HSA-2173782 | 0.315495 | 0.362819 | 2.688865 | 3.1019105 | APOE |
| 281 | Senescence-Associated Secretory Phenotype (SASP) Homo sapiens R-HSA-2559582 | 0.325953 | 0.373512 | 2.582335 | 2.8948 | FOS |

| | | | | | | |
|---|---|---|---|---|---|---|
| 282 | Transcriptional regulation of white adipocyte differentiation Homo sapiens R-HSA-381340 | 0.332838 | 0.380049 | 2.515867 | 2.7677064 | LPL |
| 283 | Toll Like Receptor 10 (TLR10) Cascade Homo sapiens R-HSA-168142 | 0.343034 | 0.387568 | 2.42232 | 2.5917037 | FOS |
| 284 | Toll Like Receptor 5 (TLR5) Cascade Homo sapiens R-HSA-168176 | 0.343034 | 0.387568 | 2.42232 | 2.5917037 | FOS |
| 285 | MyD88 cascade initiated on plasma membrane Homo sapiens R-HSA-975871 | 0.343034 | 0.387568 | 2.42232 | 2.5917037 | FOS |
| 286 | TRAF6 mediated induction of NFkB and MAP kinases upon TLR7/8 or 9 activation Homo sapiens R-HSA-975138 | 0.346398 | 0.390001 | 2.392659 | 2.5366167 | FOS |
| 287 | Adaptive Immune System Homo sapiens R-HSA-1280218 | 0.348403 | 0.390891 | 1.303368 | 1.3742647 | FGF9;ERBB4;EGF;PDGFA;FGFR3 |
| 288 | TP53 Regulates Metabolic Genes Homo sapiens R-HSA-5628897 | 0.349745 | 0.391035 | 2.363712 | 2.4831976 | RRAGD |
| 289 | MyD88 dependent cascade initiated on endosome Homo sapiens R-HSA-975155 | 0.353076 | 0.392036 | 2.335455 | 2.4313789 | FOS |
| 290 | Toll Like Receptor 7/8 (TLR7/8) Cascade Homo sapiens R-HSA-168181 | 0.353076 | 0.392036 | 2.335455 | 2.4313789 | FOS |
| 291 | Metabolism of nucleotides Homo sapiens R-HSA-15869 | 0.359686 | 0.398003 | 2.280912 | 2.332289 | NUDT16 |
| 292 | Diseases of glycosylation Homo sapiens R-HSA-3781865 | 0.362966 | 0.39889 | 2.254581 | 2.284899 | GPC3 |
| 293 | Toll Like Receptor 9 (TLR9) Cascade Homo sapiens R-HSA-168138 | 0.362966 | 0.39889 | 2.254581 | 2.284899 | FOS |
| 294 | Oxidative Stress Induced Senescence Homo sapiens R-HSA-2559580 | 0.372706 | 0.406195 | 2.179098 | 2.1506964 | FOS |

| | | | | | | |
|---|---|---|---|---|---|---|
| 295 | MyD88:Mal cascade initiated on plasma membrane Homo sapiens R-HSA-166058 | 0.375919 | 0.406195 | 2.155043 | 2.1084519 | FOS |
| 296 | Toll Like Receptor TLR1:TLR2 Cascade Homo sapiens R-HSA-168179 | 0.375919 | 0.406195 | 2.155043 | 2.1084519 | FOS |
| 297 | Toll Like Receptor TLR6:TLR2 Cascade Homo sapiens R-HSA-168188 | 0.375919 | 0.406195 | 2.155043 | 2.1084519 | FOS |
| 298 | Toll Like Receptor 2 (TLR2) Cascade Homo sapiens R-HSA-181438 | 0.375919 | 0.406195 | 2.155043 | 2.1084519 | FOS |
| 299 | Interferon gamma signaling Homo sapiens R-HSA-877300 | 0.379117 | 0.40828 | 2.131511 | 2.067375 | MT2A |
| 300 | Chaperonin-mediated protein folding Homo sapiens R-HSA-390466 | 0.385463 | 0.413731 | 2.085949 | 1.9885541 | TUBB2B |
| 301 | MyD88-independent TLR3/TLR4 cascade Homo sapiens R-HSA-166166 | 0.391746 | 0.41631 | 2.042285 | 1.9139128 | FOS |
| 302 | Toll Like Receptor 3 (TLR3) Cascade Homo sapiens R-HSA-168164 | 0.391746 | 0.41631 | 2.042285 | 1.9139128 | FOS |
| 303 | TRIF-mediated TLR3/TLR4 signaling Homo sapiens R-HSA-937061 | 0.391746 | 0.41631 | 2.042285 | 1.9139128 | FOS |
| 304 | Transport of glucose and other sugars, bile salts and organic acids, metal ions and amine compounds Homo sapiens R-HSA-425366 | 0.40412 | 0.426645 | 1.960198 | 1.7760253 | SLC22A2 |
| 305 | Protein folding Homo sapiens R-HSA-391251 | 0.40412 | 0.426645 | 1.960198 | 1.7760253 | TUBB2B |
| 306 | Degradation of the extracellular matrix Homo sapiens R-HSA-1474228 | 0.419238 | 0.441159 | 1.866384 | 1.62248 | SPP1 |
| 307 | Metabolism of carbohydrates Homo sapiens R-HSA-71387 | 0.422715 | 0.443369 | 1.401286 | 1.2065864 | GPC3;HS6ST2 |
| 308 | Metabolism of lipids and lipoproteins Homo sapiens R-HSA-556833 | 0.434582 | 0.454335 | 1.199128 | 0.9993188 | FABP3;SCD5;LPL;APOE |

| | | | | | | |
|---|---|---|---|---|---|---|
| 309 | Activated TLR4 signalling Homo sapiens R-HSA-166054 | 0.436878 | 0.455258 | 1.764963 | 1.4615668 | FOS |
| 310 | Signaling by Wnt Homo sapiens R-HSA-195721 | 0.445353 | 0.462592 | 1.338225 | 1.0824755 | SFRP1;SOX4 |
| 311 | Toll Like Receptor 4 (TLR4) Cascade Homo sapiens R-HSA-166016 | 0.465108 | 0.481559 | 1.61828 | 1.2387703 | FOS |
| 312 | Toll-Like Receptors Cascades Homo sapiens R-HSA-168898 | 0.512435 | 0.528859 | 1.407436 | 0.9409867 | FOS |
| 313 | Cellular Senescence Homo sapiens R-HSA-2559583 | 0.56243 | 0.578602 | 1.221411 | 0.7029083 | FOS |
| 314 | GPCR downstream signaling Homo sapiens R-HSA-388396 | 0.566758 | 0.581198 | 0.997196 | 0.5662299 | CASR;CXCL12;PTGER3;AVPR1A;KNG1 |
| 315 | Generic Transcription Pathway Homo sapiens R-HSA-212436 | 0.598617 | 0.61192 | 0.964336 | 0.4948331 | IGFBP3;RRAGD;FOS;NDRG1 |
| 316 | Assembly of the primary cilium Homo sapiens R-HSA-5617833 | 0.617346 | 0.629068 | 1.049292 | 0.5061008 | CYS1 |
| 317 | Ion channel transport Homo sapiens R-HSA-983712 | 0.647688 | 0.657904 | 0.965396 | 0.4193162 | FXYD2 |
| 318 | Fatty acid, triacylglycerol, and ketone body metabolism Homo sapiens R-HSA-535734 | 0.672275 | 0.680731 | 0.902182 | 0.3582458 | SCD5 |
| 319 | Organelle biogenesis and maintenance Homo sapiens R-HSA-1852241 | 0.813733 | 0.821386 | 0.596283 | 0.1229075 | CYS1 |
| 320 | Vesicle-mediated transport Homo sapiens R-HSA-5653656 | 0.92169 | 0.92745 | 0.391341 | 0.0319125 | APOE |
| 321 | Gene Expression Homo sapiens R-HSA-74160 | 0.971159 | 0.974184 | 0.458362 | 0.0134141 | IGFBP3;RRAGD;FOS;NDRG1 |
| 322 | Metabolism of proteins Homo sapiens R-HSA-392499 | 0.975881 | 0.975881 | 0.351231 | 0.0085751 | TUBB2B;IGFBP3 |

**Supplemental Table 4**: List of reversed genes in mouse model experiment in response to SGLT2i treatment.

| | gene_mouse | gene_human | logFC_dbdb_dbm | logFC_SGLT2i_dbdb |
|---|---|---|---|---|
| 1 | Igf2 | IGF2 | -1.02E+00 | -8.78E-02 |
| 2 | Igfals | IGFALS | -4.10E-01 | -2.63E-02 |
| 3 | Slc9a8 | SLC9A8 | -3.24E-01 | -8.20E-02 |
| 4 | Spats2l | SPATS2L | -2.76E-01 | -2.21E-02 |
| 5 | Fkbp5 | FKBP5 | 3.80E-01 | 1.64E-02 |
| 6 | 1110032F04Rik | C3orf80 | -5.85E-01 | -4.10E-02 |
| 7 | Hapln1 | HAPLN1 | -6.08E-01 | -7.95E-02 |
| 8 | Sec14l3 | SEC14L3 | -9.38E-01 | -1.71E-01 |
| 9 | Cacna1i | CACNA1I | -4.06E-01 | -1.73E-02 |
| 10 | Trim46 | TRIM46 | -4.31E-01 | 6.75E-02 |
| 11 | Tgm1 | TGM1 | -3.36E-01 | 9.53E-02 |
| 12 | Slc5a6 | SLC5A6 | -2.00E-01 | -5.93E-02 |
| 13 | Prok1 | PROK1 | -4.57E-01 | -1.11E-01 |
| 14 | Cyp51 | CYP51A1 | -2.45E-01 | -5.39E-02 |
| 15 | Gdf11 | GDF11 | 2.68E-01 | -2.68E-02 |
| 16 | Pcbp3 | PCBP3 | 1.36E-01 | 1.75E-03 |
| 17 | Armc12 | ARMC12 | 6.76E-01 | -6.54E-02 |
| 18 | Spidr | SPIDR | -3.33E-01 | -4.33E-02 |
| 19 | Acot7 | ACOT7 | -2.36E-01 | -4.05E-02 |
| 20 | Ugt1a10 | UGT1A10 | 5.62E-01 | -6.68E-02 |
| 21 | Ugt1a10 | UGT1A8 | 5.62E-01 | -6.68E-02 |
| 22 | Ugt1a10 | UGT1A7 | 5.62E-01 | -6.68E-02 |
| 23 | Ugt1a10 | UGT1A9 | 5.62E-01 | -6.68E-02 |
| 24 | Maob | MAOB | 3.44E-01 | 6.64E-03 |
| 25 | Hmgcr | HMGCR | -3.14E-01 | -2.55E-02 |
| 26 | Nceh1 | NCEH1 | -1.40E-01 | -3.72E-02 |
| 27 | Mmp13 | MMP13 | -4.77E-01 | 1.88E-02 |
| 28 | Creb3l1 | CREB3L1 | -3.29E-01 | -9.08E-02 |
| 29 | Ttpa | TTPA | 2.80E-01 | 5.89E-02 |
| 30 | Sytl3 | SYTL3 | -3.54E-01 | -2.33E-02 |
| 31 | Hif3a | HIF3A | 7.03E-01 | -1.31E-02 |
| 32 | Hdc | HDC | -8.54E-01 | -1.24E-01 |
| 33 | Cd79a | CD79A | -2.92E-01 | 3.92E-02 |
| 34 | Baat | BAAT | -4.30E-01 | -2.16E-02 |
| 35 | Cited1 | CITED1 | -5.25E-01 | -1.48E-01 |
| 36 | Alox15 | ALOX15 | -7.13E-01 | 5.43E-02 |
| 37 | Slco4c1 | SLCO4C1 | 5.78E-01 | -9.97E-02 |
| 38 | Svopl | SVOPL | 1.97E-01 | 2.22E-03 |
| 39 | Sost | SOST | 7.11E-01 | -7.22E-02 |
| 40 | Pop1 | POP1 | -1.92E-01 | 2.80E-02 |
| 41 | Asb11 | ASB11 | 4.49E-01 | -6.89E-03 |
| 42 | Hunk | HUNK | 2.38E-01 | -7.59E-04 |
| 43 | Lpl | LPL | -2.34E-01 | 4.26E-03 |
| 44 | Ggt6 | GGT6 | -3.22E-01 | -8.33E-02 |
| 45 | Il10rb | IL10RB | -1.18E-01 | 2.14E-03 |
| 46 | Pdk1 | PDK1 | 1.40E-01 | -1.73E-02 |
| 47 | Rnd2 | RND2 | 2.21E-01 | -2.38E-02 |
| 48 | Gbp3 | GBP4 | -3.02E-01 | 2.14E-02 |
| 49 | Metrnl | METRNL | -1.47E-01 | 3.58E-02 |
| 50 | Ccdc170 | CCDC170 | -7.09E-01 | -1.81E-01 |

| | | | | |
|---|---|---|---|---|
| 51 | Kcng1 | KCNG1 | -3.27E-01 | 1.74E-02 |
| 52 | Scg5 | SCG5 | 1.72E-01 | -8.10E-03 |
| 53 | Hprt | HPRT1 | 8.13E-02 | 8.73E-03 |
| 54 | Gabpb1 | GABPB1 | -1.55E-01 | 3.07E-03 |
| 55 | Snta1 | SNTA1 | 1.12E-01 | -2.50E-02 |
| 56 | Socs2 | SOCS2 | -3.15E-01 | 1.21E-01 |
| 57 | Myo5a | MYO5A | -2.68E-01 | -7.03E-02 |
| 58 | Ptpn5 | PTPN5 | -6.39E-01 | 2.31E-02 |
| 59 | Syp | SYP | -2.14E-01 | -1.08E-02 |
| 60 | Elovl6 | ELOVL6 | -1.30E-01 | -4.17E-02 |
| 61 | Galr2 | GALR2 | -3.60E-01 | -1.40E-01 |
| 62 | Hsd3b1 | HSD3B2 | 4.20E-01 | -9.09E-02 |
| 63 | Cyp4f14 | CYP4F12 | -3.00E-01 | -1.29E-01 |
| 64 | Prdm1 | PRDM1 | -3.79E-01 | 9.95E-02 |
| 65 | Gimap8 | GIMAP8 | -1.89E-01 | -6.78E-03 |
| 66 | Ankrd31 | ANKRD31 | -2.77E-01 | 1.76E-03 |
| 67 | Adamts8 | ADAMTS8 | -4.53E-01 | 5.19E-02 |
| 68 | Vsnl1 | VSNL1 | -3.72E-01 | -1.71E-02 |
| 69 | Ston2 | STON2 | -1.57E-01 | -3.46E-02 |
| 70 | Rpf2 | RPF2 | 1.07E-01 | -1.64E-02 |
| 71 | Ldha | LDHA | 9.65E-02 | -1.15E-02 |
| 72 | A2m | A2M | -3.74E-01 | 6.06E-02 |
| 73 | Mpst | MPST | -1.32E-01 | -2.13E-02 |
| 74 | Enpp2 | ENPP2 | 1.21E-01 | -1.44E-02 |
| 75 | Slc22a27 | SLC22A25 | -2.99E-01 | -6.92E-02 |
| 76 | Npr2 | NPR2 | -1.40E-01 | -5.87E-02 |
| 77 | Serpine2 | SERPINE2 | 1.54E-01 | 5.17E-02 |
| 78 | Abca7 | ABCA7 | 8.67E-02 | -2.06E-02 |
| 79 | Ms4a1 | MS4A1 | -2.32E-01 | 2.51E-02 |
| 80 | Plekhf1 | PLEKHF1 | 2.24E-01 | -5.43E-02 |
| 81 | Atg4a | ATG4A | 1.09E-01 | -2.06E-02 |
| 82 | Utp20 | UTP20 | 2.00E-01 | -9.01E-03 |
| 83 | Gna14 | GNA14 | 2.71E-01 | -3.62E-02 |
| 84 | Tbc1d14 | TBC1D14 | 1.16E-01 | -5.73E-04 |
| 85 | Igf1 | IGF1 | -2.69E-01 | 4.97E-02 |
| 86 | Slc22a17 | SLC22A17 | -1.85E-01 | -2.89E-02 |
| 87 | Nkx6-2 | NKX6-2 | 5.53E-01 | 1.66E-01 |
| 88 | Slc5a8 | SLC5A8 | 1.50E-01 | -2.58E-02 |
| 89 | Ttyh1 | TTYH1 | -3.57E-01 | 2.94E-02 |
| 90 | Rpgrip1 | RPGRIP1 | -2.19E-01 | -8.42E-03 |
| 91 | Zbtb16 | ZBTB16 | 4.31E-01 | 4.15E-02 |
| 92 | Ncoa4 | NCOA4 | 1.30E-01 | 1.26E-03 |
| 93 | Apln | APLN | -2.33E-01 | -5.67E-02 |
| 94 | Egln3 | EGLN3 | 2.70E-01 | 9.76E-03 |
| 95 | Tspan3 | TSPAN3 | 9.88E-02 | -2.47E-02 |
| 96 | Csmd1 | CSMD1 | -3.62E-01 | -7.62E-03 |
| 97 | Ebp | EBP | -1.01E-01 | -1.02E-02 |
| 98 | Clec9a | CLEC9A | -3.95E-01 | 5.65E-02 |
| 99 | Cdr2 | CDR2 | 1.64E-01 | -2.45E-02 |
| 100 | Qrfpr | QRFPR | 2.59E-01 | -1.56E-02 |
| 101 | Myh1 | MYH1 | -4.65E-01 | -1.84E-01 |
| 102 | Zbtb44 | ZBTB44 | -1.14E-01 | -1.51E-02 |
| 103 | Pde11a | PDE11A | -2.52E-01 | 3.31E-02 |

| | | | | |
|---|---|---|---|---|
| 104 | Aars | AARS | 7.17E-02 | -1.29E-02 |
| 105 | Plcxd2 | PLCXD2 | -2.87E-01 | 8.75E-03 |
| 106 | Slco3a1 | SLCO3A1 | 1.02E-01 | 2.04E-02 |
| 107 | Ggct | GGCT | -1.75E-01 | -4.88E-02 |
| 108 | Ifi203 | IFI16 | -2.62E-01 | 3.70E-02 |
| 109 | Ccl28 | CCL28 | -5.65E-01 | -8.03E-02 |
| 110 | Fbln7 | FBLN7 | -2.79E-01 | 1.18E-02 |
| 111 | Plcb2 | PLCB2 | -2.69E-01 | 7.79E-02 |
| 112 | Mndal | IFI16 | -2.67E-01 | 6.04E-02 |
| 113 | Lad1 | LAD1 | 8.20E-02 | 3.67E-02 |
| 114 | Plek2 | PLEK2 | -2.52E-01 | -1.36E-02 |
| 115 | Lrrc55 | LRRC55 | -1.65E-01 | -7.38E-03 |
| 116 | Aplnr | APLNR | -3.98E-01 | 6.83E-02 |
| 117 | Rgcc | RGCC | 1.89E-01 | -2.93E-02 |
| 118 | Inpp5b | INPP5B | 1.42E-01 | -1.05E-02 |
| 119 | Osbpl10 | OSBPL10 | 1.61E-01 | -5.47E-03 |
| 120 | Osbpl9 | OSBPL9 | 9.50E-02 | -8.88E-03 |
| 121 | Dll4 | DLL4 | -2.71E-01 | 6.64E-02 |
| 122 | Hemk1 | HEMK1 | -1.03E-01 | -3.04E-02 |
| 123 | Peg3 | PEG3 | 1.39E-01 | -6.48E-02 |
| 124 | Ror2 | ROR2 | -2.15E-01 | -4.06E-02 |
| 125 | Tmprss6 | TMPRSS6 | -5.53E-01 | -9.24E-02 |
| 126 | Calb1 | CALB1 | 1.46E-01 | 1.07E-03 |
| 127 | Ltc4s | LTC4S | 2.61E-01 | 1.40E-01 |
| 128 | Rab4a | RAB4A | 8.83E-02 | -2.47E-02 |
| 129 | Mad2l2 | MAD2L2 | 1.76E-01 | 5.56E-02 |
| 130 | Slc16a4 | SLC16A4 | -1.49E-01 | -6.83E-02 |
| 131 | Get4 | GET4 | 8.37E-02 | -1.04E-02 |
| 132 | Pxk | PXK | 5.55E-02 | 1.45E-02 |
| 133 | Acat3 | ACAT2 | 2.24E-01 | 3.90E-02 |
| 134 | Ppp1r26 | PPP1R26 | -1.78E-01 | 1.05E-02 |
| 135 | Nt5c3b | NT5C3B | -1.49E-01 | 1.94E-03 |
| 136 | Vwa1 | VWA1 | -1.39E-01 | -3.91E-02 |
| 137 | Prkag3 | PRKAG3 | 2.88E-01 | -6.71E-02 |
| 138 | Xpnpep2 | XPNPEP2 | 2.73E-01 | -9.86E-02 |
| 139 | Sdc2 | SDC2 | 9.11E-02 | -3.39E-03 |
| 140 | Fndc3a | FNDC3A | -8.92E-02 | -3.98E-03 |
| 141 | Apobec2 | APOBEC2 | -3.38E-01 | -8.64E-02 |
| 142 | Vwa2 | VWA2 | -1.68E-01 | -9.93E-02 |
| 143 | Ftcd | FTCD | 2.88E-01 | 8.12E-03 |
| 144 | Entpd1 | ENTPD1 | 1.64E-01 | -2.52E-02 |
| 145 | Il17f | IL17F | -2.26E-01 | -3.71E-02 |
| 146 | Mpped1 | MPPED1 | 2.63E-01 | -1.44E-02 |
| 147 | Cd19 | CD19 | -2.47E-01 | -1.77E-02 |
| 148 | Upp2 | UPP2 | 2.77E-01 | -9.03E-03 |
| 149 | Cep95 | CEP95 | -1.23E-01 | 4.45E-03 |
| 150 | Glrx | GLRX | 1.49E-01 | 1.74E-03 |
| 151 | Ank | ANKH | -1.25E-01 | 5.27E-02 |
| 152 | Pcolce2 | PCOLCE2 | -2.31E-01 | -7.60E-02 |
| 153 | Aacs | AACS | -1.47E-01 | 2.83E-02 |
| 154 | Hsbp1l1 | HSBP1L1 | -3.02E-01 | -1.84E-02 |
| 155 | Repin1 | REPIN1 | -7.16E-02 | 9.71E-03 |
| 156 | Slfn8 | SLFN13 | -2.86E-01 | 7.19E-02 |

| | | | | |
|---|---|---|---|---|
| 157 | Asph | ASPH | 8.21E-02 | -6.46E-02 |
| 158 | Tbl1x | TBL1X | 5.74E-02 | -1.26E-02 |
| 159 | Hspb8 | HSPB8 | 1.58E-01 | 9.03E-02 |
| 160 | Scd1 | SCD | -2.60E-01 | -5.84E-02 |
| 161 | Ext1 | EXT1 | 6.71E-02 | -8.65E-03 |
| 162 | Oas3 | OAS3 | -2.16E-01 | 1.03E-01 |
| 163 | B4galt5 | B4GALT5 | -1.64E-01 | 2.25E-03 |
| 164 | Cd22 | CD22 | -2.34E-01 | 1.63E-02 |
| 165 | Mast3 | MAST3 | 2.22E-01 | -3.00E-02 |
| 166 | Bcap29 | BCAP29 | 8.42E-02 | -3.04E-04 |
| 167 | Cys1 | CYS1 | -1.27E-01 | -6.95E-03 |
| 168 | Lgals12 | LGALS12 | -3.20E-01 | 4.92E-02 |
| 169 | Grem1 | GREM1 | 7.94E-01 | -5.21E-02 |
| 170 | Pcyox1l | PCYOX1L | -1.54E-01 | 6.63E-02 |
| 171 | Slc19a1 | SLC19A1 | 1.41E-01 | -1.96E-04 |
| 172 | Nup210 | NUP210 | 8.60E-02 | -2.26E-02 |
| 173 | Tgfb2 | TGFB2 | -2.62E-01 | 2.82E-02 |
| 174 | Mme | MME | -1.73E-01 | -4.95E-02 |
| 175 | Fam133b | FAM133B | 9.49E-02 | 5.27E-03 |
| 176 | Rgs5 | RGS5 | 1.26E-01 | 1.04E-02 |
| 177 | Sntg2 | SNTG2 | -3.43E-01 | 5.96E-02 |
| 178 | Fgl1 | FGL1 | -2.30E-01 | 1.05E-02 |
| 179 | Clcn7 | CLCN7 | 6.68E-02 | -2.23E-02 |
| 180 | Tmem169 | TMEM169 | -2.05E-01 | -8.88E-03 |
| 181 | Irf5 | IRF5 | -1.56E-01 | -3.80E-02 |
| 182 | Ptpn1 | PTPN1 | 9.78E-02 | 4.34E-03 |
| 183 | Esm1 | ESM1 | -4.37E-01 | 6.09E-02 |
| 184 | Fbxo40 | FBXO40 | 2.02E-01 | -2.82E-02 |
| 185 | Dclk2 | DCLK2 | -2.64E-01 | -8.37E-02 |
| 186 | Fkbp9 | FKBP9 | -6.50E-02 | -4.36E-03 |
| 187 | Fgf1 | FGF1 | -1.38E-01 | -4.75E-02 |
| 188 | Kcnh1 | KCNH1 | -4.47E-01 | 1.95E-01 |
| 189 | Ccnyl1 | CCNYL1 | 1.03E-01 | -1.26E-02 |
| 190 | Ufsp1 | UFSP1 | 1.29E-01 | -1.42E-02 |
| 191 | Dnase1l3 | DNASE1L3 | -2.92E-01 | 7.54E-03 |
| 192 | Kctd9 | KCTD9 | 7.06E-02 | -8.18E-03 |
| 193 | Acy3 | ACY3 | -9.19E-02 | -1.98E-02 |
| 194 | Ephb6 | EPHB6 | -2.47E-01 | 1.07E-01 |
| 195 | Cdh6 | CDH6 | -2.07E-01 | 1.14E-02 |
| 196 | Dnaaf1 | DNAAF1 | -1.97E-01 | -8.50E-04 |
| 197 | Rhov | RHOV | -3.20E-01 | -3.14E-02 |
| 198 | Acy1 | ACY1 | -1.40E-01 | -6.81E-02 |
| 199 | Mgat5b | MGAT5B | -2.71E-01 | -6.09E-02 |
| 200 | Mmp19 | MMP19 | 3.15E-01 | 6.46E-02 |
| 201 | Ubiad1 | UBIAD1 | -1.09E-01 | -2.04E-02 |
| 202 | Fbxw9 | FBXW9 | 9.46E-02 | 2.02E-02 |
| 203 | Vat1 | VAT1 | 9.21E-02 | 1.44E-02 |
| 204 | Gfra1 | GFRA1 | -2.95E-01 | -1.36E-01 |
| 205 | Col13a1 | COL13A1 | 3.22E-01 | 3.93E-02 |
| 206 | Mab21l3 | MAB21L3 | 3.76E-01 | -1.68E-01 |
| 207 | Fam214b | FAM214B | 8.10E-02 | -1.45E-02 |
| 208 | Ano6 | ANO6 | 5.54E-02 | -2.03E-02 |
| 209 | Ankrd37 | ANKRD37 | 3.10E-01 | 1.82E-02 |

| | | | | |
|---|---|---|---|---|
| 210 | Gzma | GZMA | -1.81E-01 | 2.60E-02 |
| 211 | Masp2 | MASP2 | -2.82E-01 | 2.09E-03 |
| 212 | Chrdl2 | CHRDL2 | 4.07E-01 | -2.33E-02 |
| 213 | Vps36 | VPS36 | 5.57E-02 | -2.28E-02 |
| 214 | Smap1 | SMAP1 | 6.56E-02 | 9.03E-03 |
| 215 | Elmod1 | ELMOD1 | 4.22E-01 | -1.30E-01 |
| 216 | B4galt1 | B4GALT1 | 1.42E-01 | 2.65E-02 |
| 217 | Dpp7 | DPP7 | -9.58E-02 | -6.58E-03 |
| 218 | Sdhaf1 | SDHAF1 | 9.45E-02 | -8.08E-03 |
| 219 | Wdfy4 | WDFY4 | -2.89E-01 | 1.20E-01 |
| 220 | Ciapin1 | CIAPIN1 | 1.06E-01 | -1.10E-03 |
| 221 | Jak2 | JAK2 | -8.14E-02 | -4.97E-03 |
| 222 | Ifih1 | IFIH1 | -1.28E-01 | 8.21E-03 |
| 223 | Acvr1b | ACVR1B | 6.40E-02 | -3.38E-02 |
| 224 | Stat3 | STAT3 | 1.02E-01 | 4.34E-02 |
| 225 | Dqx1 | DQX1 | -1.62E-01 | -7.81E-02 |
| 226 | Slc22a13 | SLC22A13 | -1.65E-01 | -7.69E-02 |
| 227 | Gorasp2 | GORASP2 | 5.47E-02 | -8.73E-03 |
| 228 | Gab2 | GAB2 | 9.06E-02 | -1.68E-02 |
| 229 | Cdkn1a | CDKN1A | 4.22E-01 | 5.15E-02 |
| 230 | Sntb1 | SNTB1 | -1.89E-01 | -4.79E-02 |
| 231 | Map3k5 | MAP3K5 | 8.95E-02 | 1.57E-02 |
| 232 | Mb21d2 | MB21D2 | 1.13E-01 | 4.78E-03 |
| 233 | Iqca | IQCA1 | -1.86E-01 | -4.06E-02 |
| 234 | Lypd6 | LYPD6 | 1.45E-01 | -3.91E-02 |
| 235 | Pcyt1a | PCYT1A | 9.93E-02 | 1.19E-02 |
| 236 | Gbp7 | GBP7 | -3.33E-01 | 3.40E-02 |
| 237 | 1700037H04Rik | C20orf27 | 1.43E-01 | -1.35E-02 |
| 238 | Ptch1 | PTCH1 | -6.88E-02 | -4.07E-02 |
| 239 | Slitrk6 | SLITRK6 | -4.81E-01 | -2.52E-02 |
| 240 | Ret | RET | 3.54E-01 | -1.13E-01 |
| 241 | Sema7a | SEMA7A | 1.65E-01 | 2.04E-03 |
| 242 | Pgp | PGP | 1.06E-01 | 1.23E-02 |
| 243 | Prdm5 | PRDM5 | 1.89E-01 | -7.13E-02 |
| 244 | Vmp1 | VMP1 | 1.34E-01 | 9.01E-03 |
| 245 | Rpp40 | RPP40 | 1.32E-01 | -1.26E-02 |
| 246 | Fbxw8 | FBXW8 | 6.74E-02 | -2.33E-02 |
| 247 | Acsm3 | ACSM3 | 4.03E-01 | -5.94E-02 |
| 248 | Sec14l1 | SEC14L1 | 1.09E-01 | 1.93E-03 |
| 249 | Dclre1b | DCLRE1B | 1.73E-01 | -4.92E-02 |
| 250 | Xaf1 | XAF1 | -1.47E-01 | 3.30E-02 |
| 251 | Gltpd2 | GLTPD2 | -1.94E-01 | 1.34E-02 |
| 252 | Tspan1 | TSPAN1 | 1.91E-01 | 8.73E-03 |
| 253 | Arhgef3 | ARHGEF3 | 8.17E-02 | -1.97E-02 |
| 254 | Clstn1 | CLSTN1 | -4.08E-02 | -1.27E-02 |
| 255 | Uhrf2 | UHRF2 | -7.72E-02 | 1.34E-02 |
| 256 | Plekha1 | PLEKHA1 | 8.42E-02 | -1.12E-04 |
| 257 | Itgb3 | ITGB3 | 2.25E-01 | 8.85E-02 |
| 258 | Frmd4b | FRMD4B | 1.51E-01 | -1.42E-02 |
| 259 | Akr1c18 | AKR1C3 | -8.79E-01 | -2.86E-01 |
| 260 | Actl6a | ACTL6A | 5.64E-02 | -9.67E-03 |
| 261 | Mmp11 | MMP11 | -2.32E-01 | 1.28E-02 |
| 262 | Slamf9 | SLAMF9 | -2.68E-01 | 7.88E-02 |

| | | | | |
|---|---|---|---|---|
| 263 | Chrdl1 | CHRDL1 | -4.15E-01 | -2.06E-01 |
| 264 | Nubp2 | NUBP2 | 6.88E-02 | 5.26E-03 |
| 265 | Ncs1 | NCS1 | 1.39E-01 | -1.07E-02 |
| 266 | Phlda2 | PHLDA2 | -1.73E-01 | 2.06E-02 |
| 267 | Dhrs11 | DHRS11 | 8.23E-02 | -1.49E-02 |
| 268 | 1700019D03Rik | C2orf88 | -2.80E-01 | -1.69E-02 |
| 269 | Sipa1l2 | SIPA1L2 | 1.43E-01 | 4.46E-03 |
| 270 | Pbx4 | PBX4 | 2.89E-01 | -1.04E-02 |
| 271 | Bag2 | BAG2 | 1.45E-01 | 2.44E-02 |
| 272 | Plce1 | PLCE1 | -1.06E-01 | -1.20E-02 |
| 273 | E2f3 | E2F3 | 1.52E-01 | 1.97E-02 |
| 274 | Efna1 | EFNA1 | -8.10E-02 | 2.20E-02 |
| 275 | Dio1 | DIO1 | -2.34E-01 | -1.08E-01 |
| 276 | Tmed6 | TMED6 | 1.41E-01 | -5.00E-02 |
| 277 | Ankrd13a | ANKRD13A | 4.64E-02 | -8.28E-03 |
| 278 | Glt8d1 | GLT8D1 | -6.08E-02 | -1.49E-03 |
| 279 | Mcam | MCAM | 1.03E-01 | 3.99E-02 |
| 280 | Anapc16 | ANAPC16 | 7.50E-02 | -6.67E-03 |
| 281 | Nfix | NFIX | -6.97E-02 | -1.14E-03 |
| 282 | Slc22a4 | SLC22A4 | -9.01E-02 | -5.08E-02 |
| 283 | Tubb4b | TUBB4B | 9.10E-02 | 4.74E-03 |
| 284 | Cand2 | CAND2 | -1.76E-01 | -5.56E-02 |
| 285 | Arhgap4 | ARHGAP4 | -1.86E-01 | -1.56E-03 |
| 286 | Setdb1 | SETDB1 | -5.67E-02 | 1.87E-02 |
| 287 | Aldh1b1 | ALDH1B1 | -2.35E-01 | -3.42E-02 |
| 288 | Adamts15 | ADAMTS15 | 1.77E-01 | 1.75E-02 |
| 289 | Cdc37l1 | CDC37L1 | 8.29E-02 | -2.01E-02 |
| 290 | Ykt6 | YKT6 | 5.68E-02 | -1.79E-02 |
| 291 | Tshr | TSHR | 2.61E-01 | 1.46E-02 |
| 292 | Usp18 | USP18 | -2.01E-01 | 8.26E-02 |
| 293 | Fbxo44 | FBXO44 | 8.38E-02 | -9.95E-03 |
| 294 | Twf1 | TWF1 | 6.47E-02 | 1.60E-03 |
| 295 | Cyp4a12b | CYP4A22 | 4.53E-01 | 4.65E-02 |
| 296 | Depdc1b | DEPDC1B | 1.50E-01 | -3.97E-02 |
| 297 | Kcnab2 | KCNAB2 | 2.57E-01 | -9.50E-02 |
| 298 | Plin4 | PLIN4 | 4.21E-01 | -3.30E-02 |
| 299 | Zmat4 | ZMAT4 | -1.53E-01 | -3.58E-02 |
| 300 | Josd2 | JOSD2 | -9.83E-02 | 8.63E-03 |
| 301 | Slc25a45 | SLC25A45 | 9.89E-02 | 2.21E-02 |
| 302 | Csf1r | CSF1R | -1.57E-01 | -2.22E-02 |
| 303 | Steap3 | STEAP3 | -2.70E-01 | -1.14E-01 |
| 304 | Nxpe3 | NXPE3 | -1.71E-01 | -6.68E-02 |
| 305 | Tifab | TIFAB | -2.59E-01 | 1.58E-01 |
| 306 | Sdcbp | SDCBP | 5.00E-02 | -3.43E-03 |
| 307 | Tppp | TPPP | -2.31E-01 | -5.85E-02 |
| 308 | Lpin1 | LPIN1 | 1.40E-01 | -1.26E-02 |
| 309 | Ssu72 | SSU72 | 5.34E-02 | -8.43E-03 |
| 310 | Col8a1 | COL8A1 | 3.49E-01 | -3.35E-03 |
| 311 | Map3k6 | MAP3K6 | 2.28E-01 | 4.41E-03 |
| 312 | Pkm | PKM | 8.76E-02 | -1.09E-02 |
| 313 | Slc38a3 | SLC38A3 | 1.96E-01 | -2.15E-02 |
| 314 | Klk1b27 | KLK3 | 2.24E-01 | 2.54E-03 |
| 315 | Rnf170 | RNF170 | 6.34E-02 | -4.51E-03 |

| | | | | |
|---|---|---|---|---|
| 316 | Dtx1 | DTX1 | -1.69E-01 | -6.22E-03 |
| 317 | Polr3e | POLR3E | 1.74E-01 | -2.65E-02 |
| 318 | Rhoc | RHOC | 1.13E-01 | 3.06E-02 |
| 319 | H2-Ob | HLA-DOB | -2.28E-01 | 1.11E-02 |
| 320 | Ccnd1 | CCND1 | 1.28E-01 | -6.39E-02 |
| 321 | Wbp1l | WBP1L | 6.76E-02 | -1.12E-02 |
| 322 | Serpina1a | SERPINA1 | 4.57E-01 | 2.14E-03 |
| 323 | Guca2b | GUCA2B | 8.04E-02 | 6.60E-03 |
| 324 | Spata13 | SPATA13 | -6.07E-02 | -2.10E-04 |
| 325 | Camk1g | CAMK1G | 2.48E-01 | -1.08E-01 |
| 326 | Rbm34 | RBM34 | 6.47E-02 | -1.40E-02 |
| 327 | Rnf24 | RNF24 | -2.29E-01 | -6.59E-02 |
| 328 | Pigz | PIGZ | 2.39E-01 | -5.67E-02 |
| 329 | Optn | OPTN | 9.31E-02 | -1.40E-02 |
| 330 | Tesk2 | TESK2 | 9.34E-02 | -1.86E-02 |
| 331 | Msi2 | MSI2 | 5.85E-02 | -2.49E-02 |
| 332 | Bspry | BSPRY | 1.35E-01 | 2.04E-02 |
| 333 | Smim1 | SMIM1 | 9.73E-02 | -2.50E-02 |
| 334 | Gatad2a | GATAD2A | 7.57E-02 | -8.92E-03 |
| 335 | Kcnk5 | KCNK5 | 1.50E-01 | -4.75E-02 |
| 336 | Kansl1l | KANSL1L | -9.81E-02 | 1.82E-02 |
| 337 | Pfkl | PFKL | 7.84E-02 | -2.66E-02 |
| 338 | Zscan26 | ZSCAN26 | -6.90E-02 | 2.77E-03 |
| 339 | Asb2 | ASB2 | -2.92E-01 | 6.05E-02 |
| 340 | Vwa3a | VWA3A | 3.27E-01 | -1.37E-01 |
| 341 | Ulk1 | ULK1 | 7.63E-02 | -1.95E-02 |
| 342 | Phf11b | PHF11 | -2.40E-01 | 4.58E-02 |
| 343 | Ankrd24 | ANKRD24 | -7.23E-02 | -3.61E-02 |
| 344 | Pi4k2a | PI4K2A | 5.78E-02 | -1.01E-02 |
| 345 | Elmo2 | ELMO2 | 5.86E-02 | -1.42E-02 |
| 346 | Ggcx | GGCX | -8.21E-02 | -3.14E-02 |
| 347 | Cd55 | CD55 | -1.64E-01 | 4.86E-02 |
| 348 | Rbpms | RBPMS | 6.11E-02 | 1.92E-02 |
| 349 | Gimap3 | GIMAP1-GIMAP5 | -3.56E-01 | -1.21E-02 |
| 350 | Herc6 | HERC6 | -1.70E-01 | 3.74E-02 |
| 351 | Atp12a | ATP12A | 3.95E-01 | -4.50E-02 |
| 352 | Psat1 | PSAT1 | 2.27E-01 | 4.81E-02 |
| 353 | Cdc42se2 | CDC42SE2 | 5.38E-02 | 7.30E-03 |
| 354 | Actr1a | ACTR1A | 4.40E-02 | 3.85E-03 |
| 355 | Slc39a5 | SLC39A5 | -1.38E-01 | 2.55E-02 |
| 356 | Eno1 | ENO1 | 8.25E-02 | -1.21E-02 |
| 357 | Sspo | SSPO | -2.01E-01 | -9.44E-02 |
| 358 | Tsc22d1 | TSC22D1 | 1.61E-01 | 3.11E-02 |
| 359 | Serinc3 | SERINC3 | 6.12E-02 | 6.70E-03 |
| 360 | Sv2c | SV2C | -2.24E-01 | 3.49E-02 |
| 361 | Csrp1 | CSRP1 | 1.03E-01 | 6.63E-03 |
| 362 | Spon2 | SPON2 | 2.32E-01 | -4.02E-02 |
| 363 | Rad54l2 | RAD54L2 | 9.06E-02 | -1.16E-02 |
| 364 | H2-Q7 | HLA-A | -3.79E-01 | 6.72E-02 |
| 365 | Arsi | ARSI | -2.50E-01 | 4.12E-02 |
| 366 | Vps37c | VPS37C | 5.24E-02 | -4.33E-03 |
| 367 | Acox3 | ACOX3 | 1.19E-01 | -1.73E-03 |
| 368 | Ergic1 | ERGIC1 | 5.56E-02 | 4.29E-03 |

| | | | | |
|---|---|---|---|---|
| 369 | Sar1b | SAR1B | 7.28E-02 | -4.77E-04 |
| 370 | Sap30 | SAP30 | -1.07E-01 | -4.04E-03 |
| 371 | Spice1 | SPICE1 | -1.11E-01 | 1.30E-02 |
| 372 | Slc22a3 | SLC22A3 | 2.40E-01 | -3.03E-02 |
| 373 | Sugp1 | SUGP1 | 5.79E-02 | -2.32E-02 |
| 374 | Med14 | MED14 | -9.56E-02 | -2.26E-02 |
| 375 | P2rx1 | P2RX1 | 1.62E-01 | -9.28E-03 |
| 376 | Ect2 | ECT2 | -3.61E-01 | 9.97E-02 |
| 377 | Lama5 | LAMA5 | 6.56E-02 | -6.40E-03 |
| 378 | Npm3 | NPM3 | -1.01E-01 | 2.18E-02 |
| 379 | Dusp3 | DUSP3 | 5.95E-02 | -5.41E-04 |
| 380 | Gamt | GAMT | -1.29E-01 | -4.91E-03 |
| 381 | Pgbd1 | PGBD1 | -1.39E-01 | -5.04E-02 |
| 382 | Hltf | HLTF | -6.91E-02 | -5.67E-03 |
| 383 | Slc17a3 | SLC17A3 | -1.04E-01 | 5.25E-03 |
| 384 | Cadm2 | CADM2 | -1.89E-01 | 3.45E-02 |
| 385 | Cyp2j6 | CYP2J2 | 6.41E-02 | 7.85E-03 |
| 386 | Chmp6 | CHMP6 | 8.91E-02 | -1.72E-02 |
| 387 | Stim1 | STIM1 | 8.02E-02 | -2.47E-02 |
| 388 | Klhdc8b | KLHDC8B | -8.52E-02 | 3.89E-02 |
| 389 | Arnt2 | ARNT2 | 1.27E-01 | -2.14E-02 |
| 390 | Fbxo36 | FBXO36 | 8.22E-02 | -2.31E-02 |
| 391 | Sdk1 | SDK1 | 1.53E-01 | 4.79E-02 |
| 392 | 6330403K07Rik | LOC728392 | -1.33E-01 | -8.82E-04 |
| 393 | Tchhl1 | TCHHL1 | 4.19E-01 | 4.21E-02 |
| 394 | Nfib | NFIB | -5.02E-02 | -1.26E-02 |
| 395 | Apom | APOM | -1.51E-01 | 1.54E-02 |
| 396 | Prob1 | PROB1 | 1.43E-01 | -5.82E-02 |
| 397 | Fgd2 | FGD2 | -1.85E-01 | 7.13E-02 |
| 398 | Gcgr | GCGR | -1.06E-01 | -7.50E-03 |
| 399 | Slc15a2 | SLC15A2 | -1.24E-01 | -5.63E-02 |
| 400 | Eif5 | EIF5 | 7.31E-02 | -2.16E-02 |
| 401 | Pax5 | PAX5 | -1.86E-01 | 2.77E-02 |
| 402 | Bbs12 | BBS12 | -1.78E-01 | 5.52E-02 |
| 403 | Ppp5c | PPP5C | 5.31E-02 | -1.21E-02 |
| 404 | Gls | GLS | 6.24E-02 | 2.97E-03 |
| 405 | Lpin2 | LPIN2 | 9.64E-02 | -2.87E-02 |
| 406 | Nmrk1 | NMRK1 | -1.03E-01 | 2.83E-02 |
| 407 | Sulf2 | SULF2 | 1.29E-01 | 3.90E-02 |
| 408 | Dusp9 | DUSP9 | -2.74E-01 | -4.99E-02 |
| 409 | Chl1 | CHL1 | -2.31E-01 | 1.65E-01 |
| 410 | Arl2bp | ARL2BP | 6.37E-02 | -1.24E-02 |
| 411 | Metap1 | METAP1 | 4.42E-02 | -4.44E-03 |
| 412 | Ube2e2 | UBE2E2 | 8.27E-02 | -2.07E-02 |
| 413 | Add2 | ADD2 | 2.58E-01 | -8.89E-02 |
| 414 | Tmem109 | TMEM109 | -5.51E-02 | -4.68E-03 |
| 415 | Bdkrb2 | BDKRB2 | 3.25E-01 | 3.27E-02 |
| 416 | Rnf213 | RNF213 | -7.21E-02 | 2.34E-02 |
| 417 | Papss2 | PAPSS2 | -9.04E-02 | -2.56E-02 |
| 418 | Setd4 | SETD4 | -1.41E-01 | 1.01E-02 |
| 419 | Sult2a3 | SULT2A1 | -2.72E-01 | 1.23E-01 |
| 420 | Cyb5rl | CYB5RL | 1.27E-01 | 2.62E-02 |
| 421 | Map2k1 | MAP2K1 | 4.66E-02 | 1.26E-03 |

| | | | | |
|---|---|---|---|---|
| 422 | Cxcr5 | CXCR5 | -1.01E-01 | 3.09E-03 |
| 423 | Spock1 | SPOCK1 | 2.36E-01 | 1.40E-01 |
| 424 | Tmprss2 | TMPRSS2 | 5.35E-02 | 1.89E-03 |
| 425 | Hipk1 | HIPK1 | 7.08E-02 | -2.61E-02 |
| 426 | Mrpl54 | MRPL54 | 5.95E-02 | -4.90E-03 |
| 427 | 4931414P19Rik | C14orf93 | -1.22E-01 | 7.51E-03 |
| 428 | Ttc19 | TTC19 | 5.27E-02 | -1.30E-02 |
| 429 | Treh | TREH | 9.87E-02 | -1.20E-02 |
| 430 | Syt7 | SYT7 | -1.79E-01 | 8.55E-02 |
| 431 | Gldc | GLDC | 1.35E-01 | 3.62E-02 |
| 432 | Gpt | GPT | 1.66E-01 | 2.15E-03 |
| 433 | Poli | POLI | -1.02E-01 | 1.13E-02 |
| 434 | Wdr62 | WDR62 | 2.59E-01 | 3.36E-02 |
| 435 | Zfp91 | ZFP91 | 4.62E-02 | -2.07E-02 |
| 436 | Mettl3 | METTL3 | -6.74E-02 | 2.62E-02 |
| 437 | Ngef | NGEF | -1.35E-01 | -4.10E-02 |
| 438 | Nagpa | NAGPA | -7.26E-02 | 1.93E-02 |
| 439 | Trappc10 | TRAPPC10 | 4.78E-02 | -2.30E-02 |
| 440 | Vgll4 | VGLL4 | -8.94E-02 | 4.41E-04 |
| 441 | Ift27 | IFT27 | -7.94E-02 | 8.90E-03 |
| 442 | Irf9 | IRF9 | -9.02E-02 | 4.38E-02 |
| 443 | Xcr1 | XCR1 | -2.70E-01 | 5.30E-02 |
| 444 | Ccdc66 | CCDC66 | -9.47E-02 | -3.44E-03 |
| 445 | Fgf13 | FGF13 | -1.48E-01 | 7.59E-02 |
| 446 | Ncoa5 | NCOA5 | -6.26E-02 | 1.78E-02 |
| 447 | Zmat3 | ZMAT3 | 8.18E-02 | -1.80E-02 |
| 448 | 1700028P14Rik | C9orf135 | 2.55E-01 | -5.48E-02 |
| 449 | Cd248 | CD248 | 1.91E-01 | 2.80E-02 |
| 450 | Pklr | PKLR | 2.03E-01 | 2.57E-02 |
| 451 | Abhd15 | ABHD15 | 2.19E-01 | 1.98E-02 |
| 452 | Itgb7 | ITGB7 | -2.38E-01 | 6.21E-02 |
| 453 | Hoxc6 | HOXC6 | -1.36E-01 | -4.04E-03 |
| 454 | Slc30a2 | SLC30A2 | 1.91E-01 | -4.00E-03 |
| 455 | Fhit | FHIT | 1.00E-01 | -5.36E-03 |
| 456 | Cdkl4 | CDKL4 | 1.62E-01 | -1.57E-02 |
| 457 | Syt4 | SYT4 | 1.13E-01 | -5.91E-02 |
| 458 | Slc7a13 | SLC7A13 | 5.52E-01 | 8.57E-02 |
| 459 | Glb1l2 | GLB1L2 | 7.05E-02 | -2.58E-02 |
| 460 | Hpgd | HPGD | -1.83E-01 | -9.14E-02 |
| 461 | Kel | KEL | 1.62E-01 | 3.86E-02 |
| 462 | Fbxo34 | FBXO34 | 9.34E-02 | -2.50E-02 |
| 463 | Smad3 | SMAD3 | -6.78E-02 | 2.39E-02 |
| 464 | Bdh1 | BDH1 | -1.37E-01 | -7.26E-02 |
| 465 | Dtnbp1 | DTNBP1 | 6.43E-02 | -1.67E-02 |
| 466 | Per3 | PER3 | -1.01E-01 | -3.47E-02 |
| 467 | H2-Q6 | HLA-A | -3.23E-01 | -1.92E-03 |
| 468 | Mtmr10 | MTMR10 | 5.74E-02 | -2.70E-02 |
| 469 | Reep3 | REEP3 | 5.78E-02 | -1.90E-02 |
| 470 | Mfn1 | MFN1 | 5.29E-02 | -9.08E-03 |
| 471 | Mogs | MOGS | -6.21E-02 | -3.45E-03 |
| 472 | Phf11d | PHF11 | -2.01E-01 | 1.12E-02 |
| 473 | Susd5 | SUSD5 | 1.94E-01 | 1.40E-01 |
| 474 | Rps6kc1 | RPS6KC1 | -6.55E-02 | -6.42E-03 |

| | | | | |
|---|---|---|---|---|
| 475 | Ralyl | RALYL | -2.42E-01 | -2.96E-02 |
| 476 | Nat2 | NAT1 | 1.45E-01 | -2.28E-03 |
| 477 | Ifi204 | IFI16 | -2.28E-01 | 1.42E-01 |
| 478 | Trpv5 | TRPV5 | 1.59E-01 | 4.99E-02 |
| 479 | Tgfb3 | TGFB3 | 1.39E-01 | 6.96E-02 |
| 480 | Mogat1 | MOGAT1 | 1.85E-01 | 7.06E-03 |
| 481 | Oas1g | OAS1 | -2.19E-01 | 9.55E-02 |
| 482 | Rfxank | RFXANK | -5.62E-02 | -7.19E-03 |
| 483 | Chrna2 | CHRNA2 | 1.39E-01 | 3.30E-02 |
| 484 | Wdr17 | WDR17 | -2.37E-01 | -4.58E-02 |
| 485 | Eif3b | EIF3B | 4.40E-02 | -1.01E-02 |
| 486 | Hexa | HEXA | 7.43E-02 | 2.85E-03 |
| 487 | Cd209a | CLEC4M | -2.36E-01 | 1.17E-01 |
| 488 | Srsf4 | SRSF4 | 6.98E-02 | -1.71E-02 |
| 489 | Efr3b | EFR3B | -1.72E-01 | 1.32E-02 |
| 490 | Mtmr2 | MTMR2 | 4.53E-02 | -2.35E-02 |
| 491 | Phc3 | PHC3 | -6.15E-02 | -9.64E-03 |
| 492 | Ntng2 | NTNG2 | -1.72E-01 | 9.01E-03 |
| 493 | Dhdh | DHDH | -1.18E-01 | -4.95E-02 |
| 494 | Arglu1 | ARGLU1 | 4.52E-02 | -5.26E-03 |
| 495 | Gtf2f2 | GTF2F2 | 7.22E-02 | 2.32E-02 |
| 496 | Pdk3 | PDK3 | -7.73E-02 | -4.28E-03 |
| 497 | Fam76a | FAM76A | 7.36E-02 | -1.59E-02 |
| 498 | Lilra5 | LILRA5 | -2.82E-01 | 7.01E-03 |
| 499 | Zfp189 | ZNF189 | -1.30E-01 | 2.34E-02 |
| 500 | Mmp17 | MMP17 | 1.66E-01 | 5.08E-03 |
| 501 | R3hdml | R3HDML | 2.17E-01 | 7.79E-02 |
| 502 | Srxn1 | SRXN1 | 9.23E-02 | 1.74E-02 |
| 503 | Ghr | GHR | -7.81E-02 | -4.04E-02 |
| 504 | Plau | PLAU | -1.35E-01 | -3.53E-02 |
| 505 | Zhx3 | ZHX3 | 7.42E-02 | -1.08E-02 |
| 506 | Tfpi2 | TFPI2 | 1.78E-01 | -2.94E-02 |
| 507 | Fndc4 | FNDC4 | -1.52E-01 | 7.63E-02 |
| 508 | Tmem87a | TMEM87A | 5.69E-02 | -1.10E-02 |
| 509 | Arhgap20 | ARHGAP20 | -1.60E-01 | 3.14E-02 |
| 510 | Atp2b4 | ATP2B4 | 1.30E-01 | 1.98E-02 |
| 511 | Sgtb | SGTB | 1.85E-01 | -3.65E-02 |
| 512 | Ly9 | LY9 | -2.06E-01 | 7.06E-02 |
| 513 | Ghdc | GHDC | 5.93E-02 | -8.91E-03 |
| 514 | Mdp1 | MDP1 | 5.88E-02 | -1.08E-02 |
| 515 | Mfng | MFNG | -1.80E-01 | 1.05E-02 |
| 516 | Prss53 | PRSS53 | -2.21E-01 | -6.62E-04 |
| 517 | Myh14 | MYH14 | 8.44E-02 | -2.84E-02 |
| 518 | Tmem63b | TMEM63B | 5.55E-02 | -3.55E-03 |
| 519 | Tmco3 | TMCO3 | -5.35E-02 | -2.59E-02 |
| 520 | Gbe1 | GBE1 | 8.29E-02 | -9.67E-03 |
| 521 | Rap1b | RAP1B | 4.88E-02 | 2.17E-02 |
| 522 | Bcl6b | BCL6B | -1.41E-01 | 5.54E-02 |
| 523 | Zscan22 | ZSCAN22 | -9.97E-02 | -3.77E-04 |
| 524 | Aif1 | AIF1 | -2.32E-01 | 1.42E-01 |
| 525 | Usp5 | USP5 | 4.15E-02 | -1.92E-02 |
| 526 | Nudt11 | NUDT11 | -1.86E-01 | 7.60E-02 |
| 527 | Gpi1 | GPI | 6.38E-02 | -1.01E-02 |

| | | | | |
|---|---|---|---|---|
| 528 | Cd36 | CD36 | -2.37E-01 | -3.39E-02 |
| 529 | Zfp385a | ZNF385A | 1.52E-01 | -5.52E-02 |
| 530 | Mapkapk3 | MAPKAPK3 | -5.77E-02 | 1.76E-02 |
| 531 | Pno1 | PNO1 | 6.77E-02 | -1.49E-02 |
| 532 | Sytl4 | SYTL4 | -1.34E-01 | 1.66E-03 |
| 533 | Napg | NAPG | 4.11E-02 | -1.75E-02 |
| 534 | Asb13 | ASB13 | 8.85E-02 | -2.37E-02 |
| 535 | Slc37a4 | SLC37A4 | 9.90E-02 | -8.58E-03 |
| 536 | Rimklb | RIMKLB | -2.17E-01 | 1.71E-02 |
| 537 | Cyp2a4 | CYP2A6 | -2.46E-01 | -7.91E-02 |
| 538 | Cyp2a4 | CYP2A7 | -2.46E-01 | -7.91E-02 |
| 539 | Cyp2a4 | CYP2A13 | -2.46E-01 | -7.91E-02 |
| 540 | Proca1 | PROCA1 | -1.79E-01 | 8.50E-04 |
| 541 | Nprl3 | NPRL3 | 4.88E-02 | -1.59E-02 |
| 542 | Gpd2 | GPD2 | 1.10E-01 | -7.47E-03 |
| 543 | Fam53a | FAM53A | 6.17E-02 | -5.32E-03 |
| 544 | Champ1 | CHAMP1 | -5.19E-02 | -8.47E-03 |
| 545 | Angptl4 | ANGPTL4 | 3.58E-01 | 3.79E-03 |
| 546 | Gpr37 | GPR37 | -1.66E-01 | 1.66E-02 |
| 547 | Rpusd3 | RPUSD3 | 1.27E-01 | -3.48E-02 |
| 548 | Il16 | IL16 | -2.08E-01 | 6.71E-02 |
| 549 | Lta | LTA | 2.26E-01 | -4.17E-02 |
| 550 | Sema4g | SEMA4G | -1.08E-01 | -4.44E-02 |
| 551 | Ip6k2 | IP6K2 | 1.31E-01 | -1.30E-02 |
| 552 | Rps6ka1 | RPS6KA1 | 6.15E-02 | 1.63E-02 |
| 553 | Abcf2 | ABCF2 | 4.65E-02 | -7.01E-03 |
| 554 | Klf11 | KLF11 | 1.99E-01 | -5.19E-02 |
| 555 | Lrig1 | LRIG1 | 1.03E-01 | -2.45E-02 |
| 556 | Tmem25 | TMEM25 | -1.14E-01 | -1.35E-02 |
| 557 | Gbp2 | GBP2 | -2.85E-01 | 1.68E-01 |
| 558 | Slc10a2 | SLC10A2 | -1.38E-01 | -6.57E-02 |
| 559 | Gtf2h5 | GTF2H5 | 5.24E-02 | -1.23E-02 |
| 560 | Nubp1 | NUBP1 | 7.19E-02 | 5.09E-03 |
| 561 | Cpq | CPQ | 5.79E-02 | -5.76E-03 |
| 562 | Slc8a1 | SLC8A1 | 1.44E-01 | -3.46E-02 |
| 563 | Mdh1b | MDH1B | -9.54E-02 | 1.60E-02 |
| 564 | 1700016C15Rik | C1orf100 | 1.91E-01 | -1.02E-02 |
| 565 | Mga | MGA | -5.55E-02 | -3.35E-03 |
| 566 | Rasl11a | RASL11A | 1.58E-01 | -2.83E-02 |
| 567 | Kcmf1 | KCMF1 | 4.40E-02 | -1.14E-02 |
| 568 | Susd3 | SUSD3 | 1.05E-01 | -2.26E-03 |
| 569 | Pcgf6 | PCGF6 | 7.93E-02 | 2.75E-02 |
| 570 | Cdc14a | CDC14A | -9.23E-02 | 9.16E-03 |
| 571 | Shkbp1 | SHKBP1 | 8.40E-02 | -3.92E-04 |
| 572 | Dnajc16 | DNAJC16 | 4.32E-02 | -2.20E-02 |
| 573 | Alms1 | ALMS1 | -1.21E-01 | 2.48E-02 |
| 574 | Cited4 | CITED4 | 2.59E-01 | -1.07E-02 |
| 575 | Rnf31 | RNF31 | -5.00E-02 | 1.07E-02 |
| 576 | Mylk3 | MYLK3 | -2.01E-01 | 4.54E-02 |
| 577 | Shroom1 | SHROOM1 | 2.08E-01 | -3.73E-02 |
| 578 | Scnn1a | SCNN1A | 6.02E-02 | 1.64E-02 |
| 579 | Arhgap29 | ARHGAP29 | -7.38E-02 | -1.95E-02 |
| 580 | Gosr2 | GOSR2 | 4.16E-02 | -5.50E-03 |

| | | | | |
|---|---|---|---|---|
| 581 | Pitpna | PITPNA | 4.22E-02 | 1.03E-02 |
| 582 | Ccr1 | CCR1 | -3.69E-01 | 7.57E-02 |
| 583 | C77080 | KIAA1522 | 3.66E-02 | 9.63E-03 |
| 584 | Ace | ACE | -1.41E-01 | -1.02E-01 |
| 585 | Tmem106c | TMEM106C | -4.90E-02 | -8.44E-03 |
| 586 | Wdr26 | WDR26 | 4.08E-02 | -1.09E-02 |
| 587 | Cnpy1 | CNPY1 | -1.84E-01 | 1.93E-02 |
| 588 | Naglu | NAGLU | -6.92E-02 | 4.31E-03 |
| 589 | Mafa | MAFA | -6.42E-02 | 1.13E-02 |
| 590 | Ptpn11 | PTPN11 | 4.02E-02 | -1.30E-02 |
| 591 | Trim67 | TRIM67 | 1.61E-01 | 3.48E-02 |
| 592 | Zgpat | ZGPAT | -6.26E-02 | 1.27E-02 |
| 593 | Antxr1 | ANTXR1 | 1.44E-01 | 5.76E-02 |
| 594 | Psmd13 | PSMD13 | 5.19E-02 | -9.51E-03 |
| 595 | Ece1 | ECE1 | -7.43E-02 | -2.99E-02 |
| 596 | Oas1a | OAS1 | -2.18E-01 | 8.76E-02 |
| 597 | Hs6st2 | HS6ST2 | -1.72E-01 | -8.82E-02 |
| 598 | Trim21 | TRIM21 | -1.03E-01 | 6.45E-04 |
| 599 | Cdc42ep4 | CDC42EP4 | 8.96E-02 | 8.51E-03 |
| 600 | Rbm4b | RBM4B | -7.27E-02 | 1.28E-02 |
| 601 | Uqcrq | UQCRQ | 5.72E-02 | -1.42E-02 |
| 602 | H2-M3 | HLA-G | -2.02E-01 | 4.95E-02 |
| 603 | Tra2a | TRA2A | -1.10E-01 | 5.02E-02 |
| 604 | Hpca | HPCA | -1.31E-01 | 2.15E-02 |
| 605 | Ctnnbip1 | CTNNBIP1 | 1.08E-01 | -2.27E-02 |
| 606 | Tpgs1 | TPGS1 | 6.72E-02 | 3.41E-03 |
| 607 | Smurf2 | SMURF2 | -5.63E-02 | 5.55E-04 |
| 608 | Irx3 | IRX3 | -8.88E-02 | -8.30E-04 |
| 609 | Ppp2r3a | PPP2R3A | 6.41E-02 | -1.23E-02 |
| 610 | Arhgap22 | ARHGAP22 | -1.40E-01 | 9.50E-03 |
| 611 | Morn2 | MORN2 | -6.61E-02 | -1.16E-02 |
| 612 | Ypel5 | YPEL5 | 7.03E-02 | -1.33E-02 |
| 613 | Ica1l | ICA1L | 1.31E-01 | -3.17E-02 |
| 614 | Trim15 | TRIM15 | -1.50E-01 | 8.87E-03 |
| 615 | Trpm3 | TRPM3 | 1.60E-01 | -7.18E-02 |
| 616 | Reep6 | REEP6 | 2.05E-01 | 8.65E-03 |
| 617 | Tagln | TAGLN | 1.72E-01 | 4.54E-02 |
| 618 | Fam189a2 | FAM189A2 | -2.12E-01 | -3.72E-02 |
| 619 | Cox6a2 | COX6A2 | 4.00E-01 | -2.01E-01 |
| 620 | Psme3 | PSME3 | 4.28E-02 | -5.44E-03 |
| 621 | Nlrc4 | NLRC4 | -2.21E-01 | 6.07E-02 |
| 622 | Ttr | TTR | 3.22E-01 | 1.75E-01 |
| 623 | Sox12 | SOX12 | 7.03E-02 | -4.68E-02 |
| 624 | Cd37 | CD37 | -2.03E-01 | 6.73E-02 |
| 625 | Plekhb1 | PLEKHB1 | -1.29E-01 | -1.88E-02 |
| 626 | Aldoa | ALDOA | 5.30E-02 | -1.70E-02 |
| 627 | Npr3 | NPR3 | -2.33E-01 | 7.45E-04 |
| 628 | Satb2 | SATB2 | -1.47E-01 | -6.17E-02 |
| 629 | Gpr162 | GPR162 | -1.55E-01 | 4.08E-02 |
| 630 | Camk1 | CAMK1 | 6.42E-02 | -2.06E-02 |
| 631 | Zfp62 | ZFP62 | -6.90E-02 | 1.22E-02 |
| 632 | Flrt2 | FLRT2 | -1.56E-01 | 1.34E-02 |
| 633 | Ifi44l | IFI44L | -1.86E-01 | -2.12E-02 |

| | | | | |
|---|---|---|---|---|
| 634 | Tacc2 | TACC2 | 6.76E-02 | -3.72E-03 |
| 635 | Wdr1 | WDR1 | 4.73E-02 | -1.70E-03 |
| 636 | Fbxo10 | FBXO10 | 1.18E-01 | -9.59E-03 |
| 637 | 1810030O07Rik | CXorf38 | -5.51E-02 | -5.40E-03 |
| 638 | Dhx9 | DHX9 | 6.46E-02 | -1.50E-02 |
| 639 | Tbcb | TBCB | 6.00E-02 | -9.75E-03 |
| 640 | Spib | SPIB | -1.75E-01 | 3.39E-02 |
| 641 | Rraga | RRAGA | 3.93E-02 | -1.02E-02 |
| 642 | Igsf5 | IGSF5 | 5.44E-02 | 2.61E-03 |
| 643 | Slc17a2 | SLC17A2 | -5.74E-01 | -2.59E-01 |
| 644 | Aifm3 | AIFM3 | -9.17E-02 | -5.03E-02 |
| 645 | Crabp1 | CRABP1 | 1.10E-01 | -2.78E-02 |
| 646 | Rsph9 | RSPH9 | -7.41E-02 | 5.23E-03 |
| 647 | Poll | POLL | -6.92E-02 | -8.08E-03 |
| 648 | Samhd1 | SAMHD1 | -6.49E-02 | 1.32E-02 |
| 649 | Slc2a12 | SLC2A12 | -1.18E-01 | -8.18E-02 |
| 650 | Spock3 | SPOCK3 | -1.60E-01 | -1.52E-02 |
| 651 | Prss12 | PRSS12 | -1.64E-01 | -7.44E-03 |
| 652 | Fam169a | FAM169A | -1.98E-01 | -1.03E-01 |
| 653 | Vps11 | VPS11 | 4.60E-02 | -1.98E-02 |
| 654 | Kif16b | KIF16B | 4.58E-02 | -3.40E-02 |
| 655 | Rdh1 | RDH16 | -1.66E-01 | -3.07E-02 |
| 656 | Ltv1 | LTV1 | 5.17E-02 | 2.61E-03 |
| 657 | Camk2d | CAMK2D | 5.64E-02 | 1.54E-02 |
| 658 | Ubox5 | UBOX5 | 7.44E-02 | -4.54E-02 |
| 659 | Nt5dc2 | NT5DC2 | 1.20E-01 | -1.35E-02 |
| 660 | Gimap4 | GIMAP4 | -1.61E-01 | 7.57E-03 |
| 661 | Lin7a | LIN7A | -2.43E-01 | -8.04E-02 |
| 662 | Lyrm4 | LYRM4 | 8.13E-02 | -2.61E-02 |
| 663 | Atg7 | ATG7 | 5.76E-02 | -1.77E-02 |
| 664 | Adh1 | ADH1C | 7.32E-02 | -1.39E-02 |
| 665 | Dnajc5 | DNAJC5 | 3.71E-02 | -2.13E-04 |
| 666 | Macrod1 | MACROD1 | -8.42E-02 | -2.63E-02 |
| 667 | Tbl3 | TBL3 | 5.07E-02 | -9.50E-03 |
| 668 | Fcer2a | FCER2 | -1.55E-01 | -1.73E-02 |
| 669 | Klk1b24 | KLK3 | 1.51E-01 | 8.55E-02 |
| 670 | Arap3 | ARAP3 | -7.55E-02 | -6.71E-03 |
| 671 | Gramd3 | GRAMD3 | 6.87E-02 | -2.58E-03 |
| 672 | Acsbg1 | ACSBG1 | 2.63E-01 | 8.76E-02 |
| 673 | Acap1 | ACAP1 | -2.22E-01 | 1.55E-01 |
| 674 | Mageh1 | MAGEH1 | -8.44E-02 | -1.64E-02 |
| 675 | Mthfd1l | MTHFD1L | 2.72E-01 | 6.91E-02 |
| 676 | Kpna1 | KPNA1 | 6.04E-02 | -1.88E-03 |
| 677 | Adm | ADM | 1.46E-01 | 3.30E-02 |
| 678 | Nos2 | NOS2 | -1.78E-01 | 8.44E-02 |
| 679 | Spock2 | SPOCK2 | 1.72E-01 | 1.27E-02 |
| 680 | Gna12 | GNA12 | 8.58E-02 | -3.50E-02 |
| 681 | Hps1 | HPS1 | 8.93E-02 | 1.05E-02 |
| 682 | Pmm1 | PMM1 | 8.46E-02 | 1.07E-03 |
| 683 | Lpcat1 | LPCAT1 | -1.09E-01 | -2.95E-02 |
| 684 | Dusp19 | DUSP19 | -1.12E-01 | -1.80E-02 |
| 685 | Spryd7 | SPRYD7 | 5.71E-02 | -1.03E-02 |
| 686 | Gsr | GSR | 1.01E-01 | 1.12E-02 |

| | | | | |
|---|---|---|---|---|
| 687 | Arhgef5 | ARHGEF5 | 4.67E-02 | -1.00E-02 |
| 688 | Tmem38b | TMEM38B | 4.14E-02 | -2.06E-02 |
| 689 | Zmynd15 | ZMYND15 | -1.67E-01 | 5.05E-02 |
| 690 | Pnkd | PNKD | 5.06E-02 | -1.78E-02 |
| 691 | Dennd1c | DENND1C | -1.82E-01 | 1.53E-01 |
| 692 | Nek7 | NEK7 | 4.01E-02 | -1.63E-02 |
| 693 | Trak1 | TRAK1 | 5.89E-02 | -2.15E-02 |
| 694 | Lrg1 | LRG1 | 2.72E-01 | 1.64E-01 |
| 695 | Slc25a46 | SLC25A46 | 4.61E-02 | -8.16E-03 |
| 696 | Slc35b1 | SLC35B1 | 8.22E-02 | -5.16E-03 |
| 697 | Il10ra | IL10RA | -1.73E-01 | 1.15E-01 |
| 698 | Ikbkb | IKBKB | -5.24E-02 | -4.50E-03 |
| 699 | Galns | GALNS | 8.38E-02 | -1.44E-02 |
| 700 | Zkscan3 | ZKSCAN3 | -6.20E-02 | -8.88E-03 |
| 701 | Adm2 | ADM2 | 1.95E-01 | 4.49E-02 |
| 702 | Gtf3c6 | GTF3C6 | 4.26E-02 | -2.50E-02 |
| 703 | Syde1 | SYDE1 | -5.36E-02 | -7.20E-04 |
| 704 | Uhrf1bp1l | UHRF1BP1L | 4.23E-02 | -9.68E-03 |
| 705 | Grb2 | GRB2 | 2.94E-02 | -1.61E-03 |
| 706 | Slamf8 | SLAMF8 | -2.40E-01 | 5.86E-02 |
| 707 | Pear1 | PEAR1 | 1.40E-01 | -2.66E-02 |
| 708 | Pacsin1 | PACSIN1 | 1.77E-01 | 1.84E-03 |
| 709 | Krt8 | KRT8 | 7.89E-02 | 3.21E-02 |
| 710 | Zfp385b | ZNF385B | -1.87E-01 | -1.91E-03 |
| 711 | Fgg | FGG | 2.38E-01 | 3.14E-01 |
| 712 | Cryz | CRYZ | 7.76E-02 | -2.37E-02 |
| 713 | Slc37a3 | SLC37A3 | -4.31E-02 | -1.17E-02 |
| 714 | Zfp704 | ZNF704 | -6.22E-02 | -1.61E-02 |
| 715 | Dmrta1 | DMRTA1 | -8.71E-02 | 9.49E-03 |
| 716 | Parp14 | PARP14 | -1.37E-01 | 7.68E-02 |
| 717 | Rmnd1 | RMND1 | -6.03E-02 | -8.42E-03 |
| 718 | Rnase6 | RNASE6 | -1.84E-01 | 6.09E-02 |
| 719 | Cacna2d2 | CACNA2D2 | -1.83E-01 | -2.19E-02 |
| 720 | Ptprt | PTPRT | 1.42E-01 | 7.70E-02 |
| 721 | Piga | PIGA | 1.16E-01 | -3.65E-02 |
| 722 | Ercc3 | ERCC3 | 4.10E-02 | -5.15E-03 |
| 723 | Col12a1 | COL12A1 | 2.10E-01 | 4.90E-02 |
| 724 | Tmprss12 | TMPRSS12 | 2.18E-01 | -8.91E-02 |
| 725 | Edil3 | EDIL3 | -1.85E-01 | -4.44E-02 |
| 726 | Ogdhl | OGDHL | -1.87E-01 | -1.17E-02 |
| 727 | Fbrsl1 | FBRSL1 | 4.92E-02 | -3.28E-03 |
| 728 | Aadat | AADAT | -1.88E-01 | -1.11E-01 |
| 729 | Swsap1 | SWSAP1 | 7.31E-02 | -2.01E-02 |
| 730 | Slit2 | SLIT2 | -1.00E-01 | -1.66E-02 |
| 731 | Ssbp4 | SSBP4 | 9.28E-02 | 4.10E-02 |
| 732 | Zfp239 | ZNF239 | -2.76E-01 | 6.41E-02 |
| 733 | Adamts6 | ADAMTS6 | 1.94E-01 | -2.48E-04 |
| 734 | Klk1b3 | KLK3 | 1.76E-01 | 1.31E-01 |
| 735 | Exosc9 | EXOSC9 | 5.92E-02 | -1.10E-04 |
| 736 | Rnf103 | RNF103 | 4.18E-02 | -6.81E-03 |
| 737 | Ptp4a2 | PTP4A2 | 4.51E-02 | 5.06E-03 |
| 738 | Tsg101 | TSG101 | 3.69E-02 | -7.74E-03 |
| 739 | Slc6a20a | SLC6A20 | -8.50E-02 | -3.12E-02 |

| | | | | |
|---|---|---|---|---|
| 740 | Wsb1 | WSB1 | 8.10E-02 | 2.79E-02 |
| 741 | Uros | UROS | -7.71E-02 | -2.45E-02 |
| 742 | Dock11 | DOCK11 | -1.31E-01 | 4.48E-02 |
| 743 | Rbm14 | RBM14 | -8.61E-02 | 3.58E-02 |
| 744 | Trp53bp2 | TP53BP2 | 6.46E-02 | -1.34E-02 |
| 745 | Trp53inp1 | TP53INP1 | 1.55E-01 | -5.70E-02 |
| 746 | Cnot6l | CNOT6L | -5.72E-02 | -1.43E-02 |
| 747 | Cmpk2 | CMPK2 | -1.01E-01 | 1.69E-02 |
| 748 | Zfp467 | ZNF467 | -8.56E-02 | -1.42E-02 |
| 749 | Mxd4 | MXD4 | 4.11E-02 | -1.87E-02 |
| 750 | Tppp3 | TPPP3 | -1.46E-01 | 7.68E-02 |
| 751 | Ramp3 | RAMP3 | 1.16E-01 | -1.16E-02 |
| 752 | Msmp | MSMP | -1.35E-01 | 7.33E-04 |
| 753 | Adamtsl3 | ADAMTSL3 | 2.87E-01 | -1.09E-01 |
| 754 | Ppp1r2 | PPP1R2 | 5.09E-02 | -5.56E-03 |
| 755 | Eif2s2 | EIF2S2 | 4.33E-02 | 9.06E-03 |
| 756 | Arpc2 | ARPC2 | 4.49E-02 | 4.67E-03 |
| 757 | Odf4 | ODF4 | 2.01E-01 | -4.18E-02 |
| 758 | Mrps18b | MRPS18B | 5.87E-02 | -7.15E-03 |
| 759 | Ccdc71l | CCDC71L | 8.09E-02 | 1.92E-04 |
| 760 | Aars2 | AARS2 | 6.21E-02 | -2.13E-02 |
| 761 | Abhd14a | ABHD14A | -8.49E-02 | -6.41E-03 |
| 762 | Plscr1 | PLSCR1 | -6.82E-02 | 1.80E-02 |
| 763 | Spata24 | SPATA24 | 1.26E-01 | 8.46E-03 |
| 764 | Flna | FLNA | 7.28E-02 | 2.58E-02 |
| 765 | Cmpk1 | CMPK1 | 4.90E-02 | -1.07E-02 |
| 766 | Astn2 | ASTN2 | -2.21E-01 | 7.79E-03 |
| 767 | Slc39a14 | SLC39A14 | 1.05E-01 | 9.32E-03 |
| 768 | Nucb2 | NUCB2 | 8.39E-02 | 1.38E-02 |
| 769 | Extl1 | EXTL1 | 2.27E-01 | -8.17E-03 |
| 770 | Ube2d1 | UBE2D1 | 5.67E-02 | -1.57E-02 |
| 771 | Rasa4 | RASA4 | -1.87E-01 | 1.07E-01 |
| 772 | Pik3r6 | PIK3R6 | -1.47E-01 | 6.82E-02 |
| 773 | Wdfy1 | WDFY1 | 4.90E-02 | -1.08E-02 |
| 774 | Fyb | FYB | -2.71E-01 | 1.90E-01 |
| 775 | Slc5a10 | SLC5A10 | 9.48E-02 | -2.75E-02 |
| 776 | Limk1 | LIMK1 | 1.11E-01 | -6.37E-03 |
| 777 | Ap5b1 | AP5B1 | 1.02E-01 | -3.68E-02 |
| 778 | Acta2 | ACTA2 | 1.76E-01 | 4.66E-02 |
| 779 | Ntrk1 | NTRK1 | -1.85E-01 | 1.36E-01 |
| 780 | Mocs2 | MOCS2 | 8.33E-02 | -1.89E-02 |
| 781 | Gbp5 | GBP5 | -3.12E-01 | 7.85E-02 |
| 782 | Sra1 | SRA1 | 5.23E-02 | -5.54E-04 |
| 783 | Cntn5 | CNTN5 | 1.38E-01 | -1.36E-01 |
| 784 | Tbxas1 | TBXAS1 | -2.45E-01 | -8.71E-02 |
| 785 | Bcl2l1 | BCL2L1 | 7.28E-02 | 6.43E-03 |
| 786 | Tcta | TCTA | 4.82E-02 | -1.53E-03 |
| 787 | Rab11b | RAB11B | 4.74E-02 | -8.33E-03 |
| 788 | Dbp | DBP | -1.64E-01 | -1.58E-01 |
| 789 | Gria3 | GRIA3 | 1.91E-01 | -4.33E-02 |
| 790 | Mettl7b | METTL7B | -9.46E-02 | -2.38E-02 |
| 791 | Alox8 | ALOX15B | -1.64E-01 | 1.57E-02 |
| 792 | Ido2 | IDO2 | -2.06E-01 | -8.47E-02 |

| | | | | |
|---|---|---|---|---|
| 793 | Dclk3 | DCLK3 | -8.94E-02 | -4.45E-03 |
| 794 | Pgam1 | PGAM1 | 5.64E-02 | -1.94E-02 |
| 795 | Adamtsl2 | ADAMTSL2 | 1.53E-01 | 5.90E-03 |
| 796 | Thsd7a | THSD7A | -1.45E-01 | 3.29E-02 |
| 797 | Mrps34 | MRPS34 | 5.58E-02 | -1.75E-02 |
| 798 | Mill2 | MICB | -1.60E-01 | -5.34E-03 |
| 799 | Arpp19 | ARPP19 | 4.95E-02 | -1.12E-02 |
| 800 | Mccc2 | MCCC2 | -6.45E-02 | -4.07E-02 |
| 801 | Ubp1 | UBP1 | 3.39E-02 | 5.73E-03 |
| 802 | Atp6v1g1 | ATP6V1G1 | 6.15E-02 | -1.67E-02 |
| 803 | Manbal | MANBAL | 6.77E-02 | -1.94E-02 |
| 804 | Timm17a | TIMM17A | 5.38E-02 | -1.82E-02 |
| 805 | Rtn4 | RTN4 | 3.68E-02 | 1.24E-02 |
| 806 | Rpia | RPIA | -9.51E-02 | 3.52E-02 |
| 807 | Trim40 | TRIM40 | -1.80E-01 | 2.75E-02 |
| 808 | St8sia1 | ST8SIA1 | -1.05E-01 | -5.92E-02 |
| 809 | Atxn7l1 | ATXN7L1 | 6.55E-02 | -1.67E-02 |
| 810 | Ethe1 | ETHE1 | 1.04E-01 | -2.11E-02 |
| 811 | Gprin3 | GPRIN3 | 1.76E-01 | -6.26E-02 |
| 812 | Uchl5 | UCHL5 | 6.15E-02 | -1.66E-03 |
| 813 | Trappc6a | TRAPPC6A | 8.22E-02 | -1.07E-02 |
| 814 | Sharpin | SHARPIN | 4.25E-02 | -1.06E-02 |
| 815 | Tbc1d20 | TBC1D20 | 3.74E-02 | -9.94E-03 |
| 816 | Pdap1 | PDAP1 | 4.03E-02 | -1.35E-02 |
| 817 | Arpc4 | ARPC4 | 4.44E-02 | -7.14E-03 |
| 818 | Nabp1 | NABP1 | 1.00E-01 | 8.46E-03 |
| 819 | Mark2 | MARK2 | 3.66E-02 | -1.37E-02 |
| 820 | Slc7a4 | SLC7A4 | -1.27E-01 | 5.52E-02 |
| 821 | Pdlim2 | PDLIM2 | 1.42E-01 | -1.39E-03 |
| 822 | Sel1l3 | SEL1L3 | -6.44E-02 | -2.81E-02 |
| 823 | R3hdm4 | R3HDM4 | 3.30E-02 | -5.10E-03 |
| 824 | Morc3 | MORC3 | -4.46E-02 | 2.44E-02 |
| 825 | Etf1 | ETF1 | 4.96E-02 | -1.35E-03 |
| 826 | Trim63 | TRIM63 | 2.02E-01 | -1.14E-02 |
| 827 | Rbm15b | RBM15B | -7.12E-02 | 4.15E-02 |
| 828 | Btla | BTLA | -2.07E-01 | 6.38E-02 |
| 829 | Mapre3 | MAPRE3 | 5.56E-02 | 1.48E-02 |
| 830 | Atp7b | ATP7B | -1.27E-01 | -6.12E-02 |
| 831 | Gak | GAK | 4.21E-02 | -1.38E-02 |
| 832 | Fabp3 | FABP3 | 6.78E-02 | -5.22E-02 |
| 833 | Cd4 | CD4 | -2.54E-01 | 6.54E-02 |
| 834 | Lrp11 | LRP11 | 8.07E-02 | 6.38E-04 |
| 835 | Tmem106b | TMEM106B | 5.60E-02 | -2.59E-02 |
| 836 | Sec24b | SEC24B | -3.71E-02 | 5.11E-03 |
| 837 | Helz | HELZ | -4.67E-02 | -1.73E-02 |
| 838 | Galnt14 | GALNT14 | 8.88E-02 | 1.16E-02 |
| 839 | Lpar2 | LPAR2 | 1.59E-01 | -6.44E-03 |
| 840 | Pdcd5 | PDCD5 | 4.42E-02 | 3.31E-03 |
| 841 | Ltb | LTB | -1.90E-01 | 6.99E-02 |
| 842 | Acvr2b | ACVR2B | -1.65E-01 | -6.38E-02 |
| 843 | Pdia5 | PDIA5 | -7.83E-02 | -6.78E-03 |
| 844 | Tuft1 | TUFT1 | 7.74E-02 | 2.05E-02 |
| 845 | Lias | LIAS | 5.32E-02 | -2.97E-02 |

| | | | | |
|---|---|---|---|---|
| 846 | Mapkapk2 | MAPKAPK2 | 4.76E-02 | 1.59E-02 |
| 847 | Aifm2 | AIFM2 | -5.73E-02 | -1.81E-02 |
| 848 | Plbd1 | PLBD1 | -1.34E-01 | 5.15E-03 |
| 849 | Phldb2 | PHLDB2 | -4.73E-02 | -1.42E-02 |
| 850 | Arhgap11a | ARHGAP11A | 2.11E-01 | 7.07E-02 |
| 851 | B3gat2 | B3GAT2 | -1.20E-01 | -2.94E-02 |
| 852 | Dstn | DSTN | 3.95E-02 | -6.83E-03 |
| 853 | Chfr | CHFR | 5.19E-02 | -4.39E-03 |
| 854 | Pus3 | PUS3 | -7.56E-02 | 2.48E-02 |
| 855 | Gars | GARS | 3.98E-02 | 2.51E-03 |
| 856 | Klf4 | KLF4 | 1.05E-01 | 4.65E-02 |
| 857 | Plcg1 | PLCG1 | 4.26E-02 | -1.65E-02 |
| 858 | Rdm1 | RDM1 | -6.27E-02 | -2.81E-02 |
| 859 | Phf11a | PHF11 | -1.57E-01 | 1.83E-02 |
| 860 | Atp2c2 | ATP2C2 | 2.61E-01 | -3.97E-02 |
| 861 | Lipc | LIPC | -1.39E-01 | 1.40E-02 |
| 862 | Zbp1 | ZBP1 | -3.12E-01 | 1.03E-01 |
| 863 | Cnnm1 | CNNM1 | 1.09E-01 | -1.45E-02 |
| 864 | Lrat | LRAT | -1.80E-01 | -2.26E-02 |
| 865 | Arhgap12 | ARHGAP12 | -4.53E-02 | -8.65E-03 |
| 866 | Rtp4 | RTP4 | -1.50E-01 | 4.73E-02 |
| 867 | Wdr25 | WDR25 | -8.49E-02 | -4.88E-02 |
| 868 | Exoc5 | EXOC5 | 4.62E-02 | 1.03E-02 |
| 869 | Mapk13 | MAPK13 | 1.19E-01 | -3.44E-02 |
| 870 | St8sia4 | ST8SIA4 | -2.10E-01 | 3.82E-02 |
| 871 | Ankrd29 | ANKRD29 | 9.50E-02 | 3.85E-02 |
| 872 | Ugt3a1 | UGT3A1 | -1.37E-01 | -6.72E-02 |
| 873 | Ccng1 | CCNG1 | 6.70E-02 | -4.82E-02 |
| 874 | Dkk1 | DKK1 | -1.14E-01 | 4.53E-02 |
| 875 | Hcn3 | HCN3 | -1.38E-01 | 1.91E-02 |
| 876 | Eif6 | EIF6 | 6.34E-02 | 1.59E-02 |
| 877 | Slc22a29 | SLC22A25 | -2.41E-01 | -7.26E-02 |
| 878 | Tmcc3 | TMCC3 | 7.37E-02 | 3.62E-02 |
| 879 | D930020B18Rik | C12orf56 | -9.86E-02 | -7.55E-03 |
| 880 | Dusp12 | DUSP12 | -7.66E-02 | -1.11E-03 |
| 881 | Iqgap1 | IQGAP1 | 4.07E-02 | -2.73E-03 |
| 882 | Cyp17a1 | CYP17A1 | 2.92E-01 | -2.65E-02 |
| 883 | Sema5a | SEMA5A | -7.36E-02 | -1.01E-02 |
| 884 | Hspb6 | HSPB6 | 1.32E-01 | 2.18E-02 |
| 885 | Rbm11 | RBM11 | 1.54E-01 | -4.16E-02 |
| 886 | Kcng4 | KCNG4 | 1.46E-01 | -1.15E-01 |
| 887 | Acadm | ACADM | -6.27E-02 | -4.49E-02 |
| 888 | Myo1d | MYO1D | 4.33E-02 | -1.97E-02 |
| 889 | Ube2d2a | UBE2D2 | 3.46E-02 | 1.31E-03 |
| 890 | Dagla | DAGLA | -1.09E-01 | 4.46E-02 |
| 891 | Zfp687 | ZNF687 | 5.06E-02 | -1.30E-02 |
| 892 | Slc28a2 | SLC28A2 | -2.12E-01 | 6.39E-02 |
| 893 | Chrnb2 | CHRNB2 | 9.59E-02 | 7.18E-03 |
| 894 | Dnah8 | DNAH8 | -1.81E-01 | 8.31E-02 |
| 895 | Pdhx | PDHX | 4.41E-02 | -1.22E-02 |
| 896 | Tasp1 | TASP1 | -6.71E-02 | -2.15E-02 |
| 897 | Nr1h4 | NR1H4 | 1.46E-01 | -4.92E-03 |
| 898 | Hnrnpab | HNRNPAB | 3.52E-02 | 1.30E-02 |

| | | | | |
|---|---|---|---|---|
| 899 | Nynrin | NYNRIN | -1.02E-01 | -2.64E-02 |
| 900 | Nagk | NAGK | -5.15E-02 | -1.23E-02 |
| 901 | Timm23 | TIMM23 | 5.00E-02 | -1.69E-02 |
| 902 | Map3k4 | MAP3K4 | -5.44E-02 | -1.53E-03 |
| 903 | Nmd3 | NMD3 | 4.07E-02 | -1.13E-02 |
| 904 | Osbp | OSBP | 4.96E-02 | -3.01E-02 |
| 905 | Spsb3 | SPSB3 | 5.62E-02 | 1.62E-02 |
| 906 | Tmc6 | TMC6 | 5.75E-02 | -1.62E-02 |
| 907 | Slamf7 | SLAMF7 | -3.07E-01 | 7.45E-02 |
| 908 | Plaa | PLAA | 4.54E-02 | 2.65E-03 |
| 909 | Mcl1 | MCL1 | 5.44E-02 | -4.25E-03 |
| 910 | Amz2 | AMZ2 | -3.60E-02 | -4.64E-03 |
| 911 | Rad18 | RAD18 | 2.03E-01 | 5.51E-02 |
| 912 | Psmd6 | PSMD6 | 4.48E-02 | -1.13E-02 |
| 913 | Sh2d4b | SH2D4B | -1.52E-01 | 7.48E-02 |
| 914 | Nucks1 | NUCKS1 | 4.57E-02 | -7.28E-03 |
| 915 | Plxnd1 | PLXND1 | -5.77E-02 | 8.76E-03 |
| 916 | Uxt | UXT | 5.54E-02 | -1.31E-02 |
| 917 | Pfkfb4 | PFKFB4 | 9.25E-02 | 7.09E-03 |
| 918 | Sema4d | SEMA4D | -7.93E-02 | -9.01E-03 |
| 919 | Pdcd10 | PDCD10 | 4.44E-02 | -2.94E-03 |
| 920 | Lmo2 | LMO2 | -1.41E-01 | -4.19E-03 |
| 921 | Zbtb22 | ZBTB22 | 6.74E-02 | -4.58E-02 |
| 922 | Tmem63a | TMEM63A | 5.96E-02 | -1.75E-02 |
| 923 | Psmd8 | PSMD8 | 3.89E-02 | -4.47E-04 |
| 924 | Fcrla | FCRLA | -1.27E-01 | -1.39E-02 |
| 925 | Hhipl1 | HHIPL1 | 1.77E-01 | -7.31E-03 |
| 926 | Cuedc1 | CUEDC1 | 8.69E-02 | -1.25E-02 |
| 927 | Nckap1l | NCKAP1L | -1.78E-01 | 1.85E-01 |
| 928 | Rap1gds1 | RAP1GDS1 | 4.44E-02 | -2.03E-02 |
| 929 | Rp1 | RP1 | -1.74E-01 | 9.82E-02 |
| 930 | Pcgf5 | PCGF5 | -5.94E-02 | -1.71E-02 |
| 931 | Agfg2 | AGFG2 | 7.84E-02 | -3.09E-03 |
| 932 | Prep | PREP | 4.98E-02 | -1.30E-03 |
| 933 | Lox | LOX | 2.68E-01 | 1.12E-01 |
| 934 | Hmg20b | HMG20B | 4.96E-02 | -5.28E-04 |
| 935 | Higd1a | HIGD1A | 5.22E-02 | 1.63E-03 |
| 936 | Tlr9 | TLR9 | -2.43E-01 | 4.09E-02 |
| 937 | Sfxn4 | SFXN4 | -9.48E-02 | -1.35E-02 |
| 938 | Mapk4 | MAPK4 | 9.05E-02 | 1.38E-02 |
| 939 | Arf3 | ARF3 | 3.02E-02 | -8.05E-03 |
| 940 | Grin3a | GRIN3A | -1.81E-01 | 3.77E-02 |
| 941 | Ubl3 | UBL3 | 3.87E-02 | -8.59E-03 |
| 942 | Angptl3 | ANGPTL3 | -2.05E-01 | -9.20E-02 |
| 943 | Eps8l3 | EPS8L3 | 2.31E-01 | 5.69E-02 |
| 944 | Psmc4 | PSMC4 | 4.49E-02 | -9.49E-03 |
| 945 | Mkks | MKKS | -1.08E-01 | 2.48E-02 |
| 946 | Edem2 | EDEM2 | -4.50E-02 | -4.71E-03 |
| 947 | Ptprc | PTPRC | -1.85E-01 | 1.25E-01 |
| 948 | Ncald | NCALD | -7.29E-02 | 1.54E-02 |
| 949 | Sec14l4 | SEC14L4 | -1.48E-01 | 1.18E-02 |
| 950 | Akap1 | AKAP1 | 7.79E-02 | -3.55E-03 |
| 951 | Ifi44 | IFI44 | -2.09E-01 | 2.30E-02 |

| | | | | |
|---|---|---|---|---|
| 952 | Ppcs | PPCS | 7.25E-02 | 4.60E-03 |
| 953 | Jmjd8 | JMJD8 | -3.83E-02 | -3.31E-03 |
| 954 | Irf7 | IRF7 | -1.65E-01 | 1.36E-01 |
| 955 | Dync1h1 | DYNC1H1 | 3.90E-02 | -1.53E-02 |
| 956 | Serpinh1 | SERPINH1 | -1.06E-01 | 2.93E-02 |
| 957 | Kcna3 | KCNA3 | -1.41E-01 | -1.48E-02 |
| 958 | Thap2 | THAP2 | -9.40E-02 | 2.89E-02 |
| 959 | Naaa | NAAA | -6.84E-02 | 5.36E-02 |
| 960 | Ddx54 | DDX54 | 3.94E-02 | -1.92E-03 |
| 961 | Pptc7 | PPTC7 | 4.65E-02 | -1.64E-02 |
| 962 | Mark3 | MARK3 | 3.88E-02 | -5.17E-03 |
| 963 | Brcc3 | BRCC3 | 5.39E-02 | -1.53E-02 |
| 964 | Gm867 | C22orf15 | -1.84E-01 | -1.82E-02 |
| 965 | Ddx60 | DDX60 | -1.62E-01 | 2.06E-02 |
| 966 | Ube2m | UBE2M | 4.29E-02 | -7.99E-03 |
| 967 | Dph3 | DPH3 | 3.86E-02 | -7.24E-03 |
| 968 | Uggt2 | UGGT2 | -1.13E-01 | -2.00E-02 |
| 969 | Exosc3 | EXOSC3 | -5.61E-02 | 1.07E-02 |
| 970 | Klf10 | KLF10 | 1.49E-01 | 3.65E-02 |
| 971 | Rhbdl3 | RHBDL3 | -1.22E-01 | 4.58E-02 |
| 972 | Dffa | DFFA | 5.24E-02 | -4.51E-03 |
| 973 | Plekhn1 | PLEKHN1 | -4.20E-02 | -2.12E-02 |
| 974 | Cbr1 | CBR1 | -1.21E-01 | -6.72E-02 |
| 975 | Arhgap30 | ARHGAP30 | -1.68E-01 | 1.29E-01 |
| 976 | Tmem161b | TMEM161B | 5.29E-02 | -2.50E-02 |
| 977 | Amph | AMPH | -2.23E-01 | 1.12E-01 |
| 978 | Man2a1 | MAN2A1 | 6.98E-02 | -1.32E-02 |
| 979 | Hspbap1 | HSPBAP1 | 1.01E-01 | -2.47E-02 |
| 980 | Uso1 | USO1 | 4.43E-02 | -1.32E-02 |
| 981 | Cxcl10 | CXCL10 | 2.77E-01 | 1.04E-01 |
| 982 | Actr1b | ACTR1B | 4.10E-02 | -9.67E-03 |
| 983 | Cstf2t | CSTF2T | -4.73E-02 | -7.90E-03 |
| 984 | Med24 | MED24 | 3.94E-02 | -1.60E-02 |
| 985 | Sfswap | SFSWAP | 3.88E-02 | -2.21E-02 |
| 986 | F2r | F2R | 1.72E-01 | 3.44E-02 |
| 987 | Gjb2 | GJB2 | -1.26E-01 | -6.88E-02 |
| 988 | Endou | ENDOU | 1.70E-01 | -4.47E-02 |
| 989 | Gdf15 | GDF15 | 3.04E-01 | 3.53E-02 |
| 990 | Pde10a | PDE10A | 2.97E-01 | -6.01E-03 |
| 991 | Tle2 | TLE2 | -8.35E-02 | -2.71E-02 |
| 992 | Flt3l | FLT3LG | -1.88E-01 | 3.15E-02 |
| 993 | Herpud1 | HERPUD1 | 7.87E-02 | -4.41E-02 |
| 994 | Serpinc1 | SERPINC1 | 1.99E-01 | 9.30E-02 |
| 995 | Klc2 | KLC2 | 3.29E-02 | -2.92E-03 |
| 996 | Gypc | GYPC | -2.24E-01 | -9.17E-02 |
| 997 | Tagln2 | TAGLN2 | 6.90E-02 | 3.72E-02 |
| 998 | Serpina10 | SERPINA10 | -4.25E-01 | 3.68E-01 |
| 999 | Npepps | NPEPPS | 4.03E-02 | -7.40E-03 |
| 1000 | Cysltr2 | CYSLTR2 | -1.88E-01 | 1.71E-03 |
| 1001 | Rbm3 | RBM3 | 9.22E-02 | 2.78E-02 |
| 1002 | Apbb1 | APBB1 | -8.27E-02 | -9.34E-03 |
| 1003 | Odf3b | ODF3B | -1.33E-01 | 3.85E-02 |
| 1004 | Pid1 | PID1 | -1.59E-01 | 5.95E-02 |

| | | | | |
|---|---|---|---|---|
| 1005 | S100g | S100G | 1.06E-01 | -1.53E-02 |
| 1006 | Klk1b5 | KLK3 | 1.59E-01 | 8.16E-02 |
| 1007 | Cyp4a12a | CYP4A22 | 4.41E-01 | 2.64E-03 |
| 1008 | Capns1 | CAPNS1 | 4.02E-02 | 8.72E-03 |
| 1009 | Syt1 | SYT1 | -8.52E-02 | 5.01E-02 |
| 1010 | Cyp2e1 | CYP2E1 | 1.10E-01 | 2.17E-02 |
| 1011 | Ikzf3 | IKZF3 | -1.62E-01 | 3.64E-02 |
| 1012 | Bcl7b | BCL7B | 3.35E-02 | 4.52E-03 |
| 1013 | Piezo1 | PIEZO1 | 6.78E-02 | 6.03E-03 |
| 1014 | Abcc3 | ABCC3 | -2.12E-01 | -1.33E-01 |
| 1015 | Mettl8 | METTL8 | -5.75E-02 | -1.06E-02 |
| 1016 | Cpeb3 | CPEB3 | 1.36E-01 | -5.93E-02 |
| 1017 | Rbm12b1 | RBM12B | -1.58E-01 | 6.12E-02 |
| 1018 | Col7a1 | COL7A1 | 1.51E-01 | 1.40E-02 |
| 1019 | Dpp6 | DPP6 | -3.18E-01 | 3.74E-02 |
| 1020 | Wdr6 | WDR6 | 5.30E-02 | -2.14E-02 |
| 1021 | Pdzd7 | PDZD7 | -1.60E-01 | 3.07E-03 |
| 1022 | Stk10 | STK10 | -1.01E-01 | 3.63E-02 |
| 1023 | Cckar | CCKAR | -7.61E-02 | -4.32E-02 |
| 1024 | Heg1 | HEG1 | 1.23E-01 | -1.11E-02 |
| 1025 | Pura | PURA | -5.03E-02 | 3.68E-02 |
| 1026 | F8 | F8 | -1.60E-01 | -3.37E-02 |
| 1027 | Ovol1 | OVOL1 | 1.31E-01 | -1.85E-02 |
| 1028 | Itgax | ITGAX | -2.39E-01 | 2.54E-01 |
| 1029 | Iffo1 | IFFO1 | -9.10E-02 | 1.20E-02 |
| 1030 | Slc43a2 | SLC43A2 | 7.19E-02 | -1.56E-02 |
| 1031 | Vapa | VAPA | 3.77E-02 | 1.65E-03 |
| 1032 | Ints8 | INTS8 | -5.70E-02 | 8.37E-03 |
| 1033 | Myrip | MYRIP | 1.45E-01 | -2.57E-02 |
| 1034 | Pdcd11 | PDCD11 | -3.87E-02 | 7.10E-03 |
| 1035 | Mvb12a | MVB12A | 3.91E-02 | 2.24E-04 |
| 1036 | Snai3 | SNAI3 | -1.44E-01 | -1.98E-02 |
| 1037 | Adh5 | ADH5 | 5.25E-02 | -2.40E-02 |
| 1038 | Galnt15 | GALNT15 | -1.76E-01 | -1.39E-02 |
| 1039 | Trappc9 | TRAPPC9 | 6.19E-02 | -1.91E-02 |
| 1040 | Rac2 | RAC2 | -2.27E-01 | 2.02E-01 |
| 1041 | Cwh43 | CWH43 | 8.62E-02 | -7.56E-03 |
| 1042 | Hyal1 | HYAL1 | 1.26E-01 | 4.47E-03 |
| 1043 | Tmem14a | TMEM14A | -8.48E-02 | -1.22E-03 |
| 1044 | Zcrb1 | ZCRB1 | 3.76E-02 | -5.20E-03 |
| 1045 | Mfge8 | MFGE8 | 7.80E-02 | 1.39E-02 |
| 1046 | Ppfibp1 | PPFIBP1 | 4.76E-02 | -1.01E-02 |
| 1047 | Grpel1 | GRPEL1 | 4.97E-02 | -2.58E-02 |
| 1048 | Rnf115 | RNF115 | 3.87E-02 | -6.74E-03 |
| 1049 | St3gal5 | ST3GAL5 | 1.72E-01 | -4.00E-02 |
| 1050 | Mrps10 | MRPS10 | 4.01E-02 | -8.23E-03 |
| 1051 | Baz1b | BAZ1B | -4.94E-02 | 1.08E-02 |
| 1052 | Phlpp2 | PHLPP2 | -6.47E-02 | -2.93E-02 |
| 1053 | Itgae | ITGAE | -1.76E-01 | 3.92E-02 |
| 1054 | Dao | DAO | 8.01E-02 | -1.87E-02 |
| 1055 | Slc16a1 | SLC16A1 | 2.09E-01 | 1.08E-01 |
| 1056 | Ppp4r4 | PPP4R4 | 1.16E-01 | -4.80E-02 |
| 1057 | Zfp691 | ZNF691 | -1.30E-01 | 2.94E-02 |

| | | | | |
|---|---|---|---|---|
| 1058 | Gbp9 | GBP6 | -1.45E-01 | 1.30E-02 |
| 1059 | Acad9 | ACAD9 | -5.39E-02 | -3.30E-02 |
| 1060 | Grb7 | GRB7 | -7.24E-02 | -1.23E-02 |
| 1061 | Hgsnat | HGSNAT | 3.01E-02 | -1.15E-02 |
| 1062 | Mrpl20 | MRPL20 | 5.34E-02 | -2.67E-03 |
| 1063 | Ift46 | IFT46 | 4.30E-02 | -3.21E-02 |
| 1064 | Trim12c | TRIM5 | -1.11E-01 | 4.16E-02 |
| 1065 | Katnal1 | KATNAL1 | -5.68E-02 | -2.29E-02 |
| 1066 | Smim22 | SMIM22 | -8.71E-02 | 3.13E-03 |
| 1067 | Slc22a30 | SLC22A25 | 1.62E-01 | -8.75E-04 |
| 1068 | Ctsw | CTSW | -1.88E-01 | 2.48E-02 |
| 1069 | Smpd1 | SMPD1 | -3.99E-02 | -1.97E-02 |
| 1070 | Mast1 | MAST1 | -1.24E-01 | 7.90E-03 |
| 1071 | Tbce | TBCE | 3.60E-02 | -5.09E-03 |
| 1072 | Arrb1 | ARRB1 | -4.65E-02 | -3.69E-02 |
| 1073 | Fam207a | FAM207A | 5.79E-02 | 8.36E-03 |
| 1074 | Nedd9 | NEDD9 | 7.96E-02 | -2.24E-02 |
| 1075 | Hnrnpl | HNRNPL | 4.07E-02 | -1.71E-02 |
| 1076 | Cxcl14 | CXCL14 | -2.59E-01 | 4.60E-02 |
| 1077 | Themis2 | THEMIS2 | -2.23E-01 | 1.47E-01 |
| 1078 | Rbbp7 | RBBP7 | 3.69E-02 | 4.34E-03 |
| 1079 | Dctn2 | DCTN2 | 3.64E-02 | -9.88E-03 |
| 1080 | Wdr55 | WDR55 | -4.54E-02 | -1.41E-02 |
| 1081 | Msl2 | MSL2 | -5.14E-02 | -6.64E-04 |
| 1082 | Hdhd3 | HDHD3 | 9.58E-02 | -3.58E-02 |
| 1083 | Psmb9 | PSMB9 | -2.62E-01 | 7.78E-02 |
| 1084 | Epc2 | EPC2 | 4.24E-02 | 1.02E-03 |
| 1085 | Sh3rf1 | SH3RF1 | 7.88E-02 | -6.28E-03 |
| 1086 | Ccdc85c | CCDC85C | 5.78E-02 | -3.45E-02 |
| 1087 | Vegfc | VEGFC | -1.18E-01 | 2.81E-02 |
| 1088 | Kcns1 | KCNS1 | -1.84E-01 | -3.41E-02 |
| 1089 | Mpv17l | MPV17L | -1.22E-01 | -6.82E-02 |
| 1090 | Dnm2 | DNM2 | 3.15E-02 | -1.13E-02 |
| 1091 | Rorb | RORB | -1.22E-01 | -2.78E-02 |
| 1092 | Mrpl16 | MRPL16 | 5.70E-02 | -1.66E-02 |
| 1093 | Ifnar2 | IFNAR2 | -5.23E-02 | 1.43E-02 |
| 1094 | Ell2 | ELL2 | 6.81E-02 | 2.41E-02 |
| 1095 | Bambi | BAMBI | -8.60E-02 | -2.00E-02 |
| 1096 | Olfm4 | OLFM4 | 9.94E-02 | -9.99E-03 |
| 1097 | Brd7 | BRD7 | 3.23E-02 | -9.67E-03 |
| 1098 | Adrm1 | ADRM1 | 5.22E-02 | -5.38E-03 |
| 1099 | Mapre1 | MAPRE1 | 3.34E-02 | 5.08E-03 |
| 1100 | Sptlc2 | SPTLC2 | 4.53E-02 | 2.01E-02 |
| 1101 | Map1lc3a | MAP1LC3A | 6.12E-02 | -8.59E-03 |
| 1102 | Ly86 | LY86 | -1.94E-01 | 1.78E-01 |
| 1103 | Ola1 | OLA1 | 4.59E-02 | -1.73E-02 |
| 1104 | Atox1 | ATOX1 | 6.38E-02 | 1.81E-02 |
| 1105 | Zfp296 | ZNF296 | -1.68E-01 | 4.89E-02 |
| 1106 | Erlin1 | ERLIN1 | -7.26E-02 | 1.30E-02 |
| 1107 | Rcan1 | RCAN1 | 1.96E-01 | -4.56E-02 |
| 1108 | Arid3b | ARID3B | -1.04E-01 | 3.01E-02 |
| 1109 | Tbc1d1 | TBC1D1 | 6.50E-02 | -1.25E-02 |
| 1110 | Acot9 | ACOT9 | 7.38E-02 | 2.33E-02 |

| | | | | |
|---|---|---|---|---|
| 1111 | Pi4k2b | PI4K2B | 4.38E-02 | -1.62E-02 |
| 1112 | Ppm1m | PPM1M | -6.44E-02 | 1.52E-03 |
| 1113 | Sfrp4 | SFRP4 | -2.54E-01 | 5.56E-02 |
| 1114 | Phyh | PHYH | 6.63E-02 | -1.18E-02 |
| 1115 | Guk1 | GUK1 | 4.50E-02 | -1.51E-02 |
| 1116 | Antxr2 | ANTXR2 | -8.64E-02 | -1.01E-02 |
| 1117 | Mrap | MRAP | 2.07E-01 | 6.43E-02 |
| 1118 | Rnf152 | RNF152 | -8.72E-02 | 1.09E-02 |
| 1119 | Rhbdf2 | RHBDF2 | 1.17E-01 | -7.86E-03 |
| 1120 | Stat2 | STAT2 | -6.40E-02 | 3.07E-03 |
| 1121 | Lama4 | LAMA4 | -1.77E-01 | -4.40E-03 |
| 1122 | Ltbr | LTBR | 4.33E-02 | -3.95E-04 |
| 1123 | Atoh7 | ATOH7 | 1.96E-01 | -1.10E-01 |
| 1124 | Ube2i | UBE2I | 4.29E-02 | -5.68E-03 |
| 1125 | Ppard | PPARD | -8.07E-02 | 3.17E-02 |
| 1126 | P2ry10 | P2RY10 | -1.18E-01 | 3.98E-02 |
| 1127 | Arpc5 | ARPC5 | 3.72E-02 | 1.11E-02 |
| 1128 | Npc1 | NPC1 | 4.00E-02 | -4.07E-03 |
| 1129 | Filip1l | FILIP1L | 9.26E-02 | -1.92E-03 |
| 1130 | Slc22a19 | SLC22A24 | -3.12E-01 | -1.56E-01 |
| 1131 | Trim34a | TRIM6-TRIM34 | -1.17E-01 | 3.67E-02 |
| 1132 | Strap | STRAP | 4.13E-02 | 2.98E-03 |
| 1133 | Ndufv3 | NDUFV3 | 4.93E-02 | -2.21E-02 |
| 1134 | Zbtb38 | ZBTB38 | 6.20E-02 | 1.63E-02 |
| 1135 | Tceal3 | TCEAL6 | 2.01E-01 | -1.09E-01 |
| 1136 | Grem2 | GREM2 | 4.02E-01 | -4.18E-02 |
| 1137 | Palld | PALLD | -1.10E-01 | 1.80E-02 |
| 1138 | Bcl2 | BCL2 | -8.59E-02 | 1.62E-03 |
| 1139 | Erlin2 | ERLIN2 | -3.56E-02 | -1.93E-02 |
| 1140 | Serpina1d | SERPINA1 | 2.03E-01 | -7.58E-02 |
| 1141 | Eva1a | EVA1A | 5.08E-02 | -7.39E-04 |
| 1142 | Arrdc4 | ARRDC4 | 8.58E-02 | -3.96E-03 |
| 1143 | Micall1 | MICALL1 | 6.18E-02 | 1.05E-02 |
| 1144 | Sema6b | SEMA6B | 1.63E-01 | -1.28E-02 |
| 1145 | Gdnf | GDNF | 1.58E-01 | -7.06E-03 |
| 1146 | Tpst2 | TPST2 | 4.90E-02 | -2.43E-04 |
| 1147 | Car5a | CA5A | -1.73E-01 | -1.19E-01 |
| 1148 | Pdlim5 | PDLIM5 | 4.58E-02 | -1.18E-02 |
| 1149 | Nacc1 | NACC1 | 3.30E-02 | -2.15E-03 |
| 1150 | Ccdc124 | CCDC124 | 5.50E-02 | -1.95E-02 |
| 1151 | Tspo | TSPO | -5.95E-02 | 1.63E-02 |
| 1152 | Sec16b | SEC16B | 2.52E-01 | -8.34E-02 |
| 1153 | Fcgr1 | FCGR1A | -1.96E-01 | 1.48E-01 |
| 1154 | Ctsd | CTSD | 4.29E-02 | 2.78E-02 |
| 1155 | C1qtnf3 | C1QTNF3 | -1.08E-01 | -9.37E-03 |
| 1156 | Stard3 | STARD3 | -3.67E-02 | -9.46E-03 |
| 1157 | Sirt5 | SIRT5 | -5.50E-02 | -2.88E-02 |
| 1158 | Col26a1 | COL26A1 | -1.27E-01 | -5.48E-02 |
| 1159 | Galnt1 | GALNT1 | 5.83E-02 | -3.61E-03 |
| 1160 | Timm21 | TIMM21 | -4.84E-02 | -2.41E-02 |
| 1161 | Ppme1 | PPME1 | 3.64E-02 | -8.62E-03 |
| 1162 | Kat6a | KAT6A | -4.30E-02 | -8.37E-03 |
| 1163 | Sidt2 | SIDT2 | 4.43E-02 | -1.03E-02 |

| | | | | |
|---|---|---|---|---|
| 1164 | Ndufb9 | NDUFB9 | 4.98E-02 | -1.80E-02 |
| 1165 | Ppan | PPAN | -6.06E-02 | 6.34E-02 |
| 1166 | Wipf1 | WIPF1 | -1.10E-01 | 6.74E-02 |
| 1167 | Inpp5a | INPP5A | 5.62E-02 | 4.59E-03 |
| 1168 | Spata2l | SPATA2L | 7.76E-02 | -1.36E-02 |
| 1169 | Pgd | PGD | 7.58E-02 | 1.86E-02 |
| 1170 | Rbm10 | RBM10 | -4.81E-02 | 7.90E-03 |
| 1171 | Mtch1 | MTCH1 | 3.50E-02 | -3.48E-03 |
| 1172 | Riok3 | RIOK3 | 3.84E-02 | -8.53E-03 |
| 1173 | Git1 | GIT1 | 3.88E-02 | -9.65E-03 |
| 1174 | Spry2 | SPRY2 | -6.95E-02 | -1.27E-03 |
| 1175 | Six5 | SIX5 | -1.14E-01 | 2.27E-02 |
| 1176 | Pithd1 | PITHD1 | 4.22E-02 | 8.53E-03 |
| 1177 | Pappa | PAPPA | 2.04E-01 | -5.22E-02 |
| 1178 | Tcf23 | TCF23 | 2.24E-01 | -4.01E-02 |
| 1179 | Psmd4 | PSMD4 | 3.70E-02 | -3.91E-03 |
| 1180 | Cab39 | CAB39 | 3.19E-02 | -1.07E-02 |
| 1181 | Mpv17l2 | MPV17L2 | 9.59E-02 | -1.92E-02 |
| 1182 | Cycs | CYCS | 5.66E-02 | -1.24E-02 |
| 1183 | Uxs1 | UXS1 | -5.53E-02 | -6.06E-03 |
| 1184 | Dusp26 | DUSP26 | -1.67E-01 | -5.99E-02 |
| 1185 | Atp6v0e | ATP6V0E1 | 4.47E-02 | 2.08E-03 |
| 1186 | Unc13b | UNC13B | 7.05E-02 | 3.81E-02 |
| 1187 | Gdpd3 | GDPD3 | -1.72E-01 | -5.74E-02 |
| 1188 | Man1b1 | MAN1B1 | -2.78E-02 | -1.27E-02 |
| 1189 | Spred3 | SPRED3 | 1.35E-01 | 3.32E-02 |
| 1190 | Siah1a | SIAH1 | 5.72E-02 | 4.58E-03 |
| 1191 | Rrp12 | RRP12 | 7.06E-02 | 3.90E-02 |
| 1192 | Erbb2 | ERBB2 | -5.91E-02 | -3.36E-03 |
| 1193 | Txndc5 | TXNDC5 | -4.64E-02 | -2.16E-02 |
| 1194 | Chst3 | CHST3 | -1.62E-01 | -8.19E-02 |
| 1195 | Sp2 | SP2 | 7.11E-02 | -2.62E-02 |
| 1196 | 2310033P09Rik | C1orf35 | -5.52E-02 | 1.13E-02 |
| 1197 | Zmym6 | ZMYM6 | -6.99E-02 | -2.24E-02 |
| 1198 | Amigo1 | AMIGO1 | -8.69E-02 | -2.82E-02 |
| 1199 | Dusp8 | DUSP8 | 2.06E-01 | 1.03E-01 |
| 1200 | Cdv3 | CDV3 | 3.59E-02 | -3.98E-03 |
| 1201 | Trps1 | TRPS1 | -1.12E-01 | 3.85E-03 |
| 1202 | Acsl6 | ACSL6 | -1.68E-01 | 5.85E-02 |
| 1203 | Tsen2 | TSEN2 | 7.30E-02 | -3.16E-02 |
| 1204 | Capg | CAPG | -1.79E-01 | 7.86E-03 |
| 1205 | Galnt18 | GALNT18 | 6.43E-02 | -1.60E-02 |
| 1206 | Ipcef1 | IPCEF1 | -1.16E-01 | -1.40E-03 |
| 1207 | Ptdss2 | PTDSS2 | 3.61E-02 | -1.11E-02 |
| 1208 | Ttll4 | TTLL4 | 6.45E-02 | -2.13E-02 |
| 1209 | Folr2 | FOLR2 | -1.80E-01 | -6.12E-02 |
| 1210 | Lpin3 | LPIN3 | 8.11E-02 | -3.93E-02 |
| 1211 | Fbln1 | FBLN1 | -2.11E-01 | 9.78E-02 |
| 1212 | Cldn5 | CLDN5 | -8.52E-02 | -3.16E-02 |
| 1213 | Sephs1 | SEPHS1 | 4.70E-02 | -1.13E-02 |
| 1214 | Mcu | MCU | 4.72E-02 | -9.98E-04 |
| 1215 | Golga3 | GOLGA3 | -3.53E-02 | -6.23E-03 |
| 1216 | Zbtb10 | ZBTB10 | -7.75E-02 | 2.55E-02 |

| | | | | |
|---|---|---|---|---|
| 1217 | Baiap3 | BAIAP3 | -2.02E-01 | -1.24E-01 |
| 1218 | Snx20 | SNX20 | -1.39E-01 | 6.38E-02 |
| 1219 | Col4a2 | COL4A2 | 8.92E-02 | 1.57E-02 |
| 1220 | Pop4 | POP4 | 6.03E-02 | -1.58E-02 |
| 1221 | Ap2a1 | AP2A1 | 4.45E-02 | -6.44E-03 |
| 1222 | Sec24a | SEC24A | 6.12E-02 | -1.25E-03 |
| 1223 | Tmed3 | TMED3 | -5.50E-02 | 1.74E-02 |
| 1224 | Kcna5 | KCNA5 | 1.59E-01 | 4.40E-03 |
| 1225 | Slc10a5 | SLC10A5 | -1.32E-01 | -8.77E-03 |
| 1226 | Tuba4a | TUBA4A | 5.40E-02 | -9.06E-03 |
| 1227 | 2010003K11Rik | C11orf86 | 2.16E-01 | 1.68E-01 |
| 1228 | Ap5z1 | AP5Z1 | -4.12E-02 | -1.19E-02 |
| 1229 | Ing2 | ING2 | 5.99E-02 | 7.10E-04 |
| 1230 | Bfar | BFAR | -3.25E-02 | 4.41E-03 |
| 1231 | Kit | KIT | -5.89E-02 | -1.07E-02 |
| 1232 | Prkra | PRKRA | -4.93E-02 | -8.91E-03 |
| 1233 | Csnk1a1 | CSNK1A1 | 3.38E-02 | 5.03E-03 |
| 1234 | Klf13 | KLF13 | 9.43E-02 | -2.18E-02 |
| 1235 | Chkb | CHKB | 4.13E-02 | -1.67E-02 |
| 1236 | Pgm2 | PGM1 | 5.34E-02 | 3.48E-02 |
| 1237 | Sars | SARS | 4.61E-02 | 4.67E-03 |
| 1238 | Fbxl16 | FBXL16 | -1.43E-01 | -1.46E-02 |
| 1239 | Ylpm1 | YLPM1 | -3.72E-02 | 8.44E-03 |
| 1240 | Bend7 | BEND7 | 8.45E-02 | -2.60E-02 |
| 1241 | Reck | RECK | -9.59E-02 | -5.26E-02 |
| 1242 | Ldlrap1 | LDLRAP1 | 5.17E-02 | -1.97E-02 |
| 1243 | Dtx3l | DTX3L | -1.10E-01 | 6.14E-02 |
| 1244 | Clec10a | CLEC10A | -2.43E-01 | 1.65E-01 |
| 1245 | Crtac1 | CRTAC1 | -1.22E-01 | 1.81E-02 |
| 1246 | Pkd2l2 | PKD2L2 | -1.31E-01 | -3.82E-02 |
| 1247 | Man1c1 | MAN1C1 | 1.32E-01 | -3.46E-03 |
| 1248 | Nat8 | NAT8 | 1.48E-01 | -5.65E-03 |
| 1249 | Tbcd | TBCD | 4.18E-02 | -1.11E-02 |
| 1250 | Tubg1 | TUBG1 | 6.07E-02 | 3.82E-03 |
| 1251 | Ptk2 | PTK2 | 3.45E-02 | -4.85E-03 |
| 1252 | Nudt10 | NUDT11 | -1.33E-01 | -4.33E-03 |
| 1253 | Adamts14 | ADAMTS14 | 1.92E-01 | -1.41E-01 |
| 1254 | Wfdc1 | WFDC1 | 1.54E-01 | 8.75E-04 |
| 1255 | Aen | AEN | 8.17E-02 | -7.62E-03 |
| 1256 | Disp1 | DISP1 | -1.05E-01 | 6.05E-03 |
| 1257 | Tcf25 | TCF25 | 3.58E-02 | -1.34E-02 |
| 1258 | Polr3d | POLR3D | 9.45E-02 | -2.42E-03 |
| 1259 | Cd320 | CD320 | -4.99E-02 | 5.68E-03 |
| 1260 | 2210016L21Rik | C12orf43 | 3.57E-02 | -1.41E-02 |
| 1261 | Mark4 | MARK4 | 3.83E-02 | -8.41E-03 |
| 1262 | Ivd | IVD | -6.14E-02 | -2.08E-02 |
| 1263 | Siglecg | SIGLEC10 | -1.52E-01 | -5.73E-03 |
| 1264 | Dync1li1 | DYNC1LI1 | 3.52E-02 | -1.55E-03 |
| 1265 | Bank1 | BANK1 | -1.13E-01 | -7.57E-03 |
| 1266 | Mapk8ip1 | MAPK8IP1 | 5.00E-02 | -3.28E-02 |
| 1267 | Tmem201 | TMEM201 | -4.35E-02 | -5.74E-03 |
| 1268 | Tpi1 | TPI1 | 5.15E-02 | -2.16E-02 |
| 1269 | Stard8 | STARD8 | -4.49E-02 | -2.11E-02 |

| | | | | |
|---|---|---|---|---|
| 1270 | Mgme1 | MGME1 | -7.34E-02 | -8.21E-03 |
| 1271 | Mms22l | MMS22L | 1.49E-01 | 4.02E-02 |
| 1272 | Dag1 | DAG1 | 3.38E-02 | -9.32E-03 |
| 1273 | Podnl1 | PODNL1 | -1.42E-01 | 6.82E-02 |
| 1274 | Dmtf1 | DMTF1 | -4.59E-02 | 1.04E-03 |
| 1275 | Rmnd5b | RMND5B | 4.42E-02 | 9.94E-03 |
| 1276 | Adam11 | ADAM11 | 1.45E-01 | -5.02E-02 |
| 1277 | Sp1 | SP1 | -3.74E-02 | -5.95E-03 |
| 1278 | Arsk | ARSK | -4.34E-02 | -4.31E-03 |
| 1279 | Thrsp | THRSP | 2.18E-01 | 8.45E-03 |
| 1280 | Eif4a2 | EIF4A2 | -4.07E-02 | -1.03E-03 |
| 1281 | Anxa3 | ANXA3 | 1.05E-01 | 5.32E-02 |
| 1282 | Atp4a | ATP4A | 1.04E-01 | -2.92E-02 |
| 1283 | Ddhd2 | DDHD2 | 6.45E-02 | 8.45E-03 |
| 1284 | Ints10 | INTS10 | -4.69E-02 | -6.86E-03 |
| 1285 | Rangrf | RANGRF | -7.20E-02 | 4.41E-03 |
| 1286 | Cbfa2t3 | CBFA2T3 | -1.03E-01 | 5.30E-03 |
| 1287 | Gimap7 | GIMAP7 | -9.64E-02 | 2.12E-02 |
| 1288 | Atf7ip | ATF7IP | -4.43E-02 | 2.25E-03 |
| 1289 | Ppp2ca | PPP2CA | 3.46E-02 | -7.65E-03 |
| 1290 | Tspan2 | TSPAN2 | 1.43E-01 | 4.02E-02 |
| 1291 | Foxred1 | FOXRED1 | 4.08E-02 | -1.08E-02 |
| 1292 | Intu | INTU | -6.49E-02 | -4.04E-02 |
| 1293 | Acr | ACR | 1.45E-01 | -1.18E-01 |
| 1294 | Cstb | CSTB | 4.46E-02 | 2.63E-02 |
| 1295 | Flad1 | FLAD1 | -4.13E-02 | -2.21E-02 |
| 1296 | Snx5 | SNX5 | 5.05E-02 | 6.06E-03 |
| 1297 | Pacsin2 | PACSIN2 | 3.63E-02 | -2.67E-02 |
| 1298 | Usp4 | USP4 | 3.57E-02 | -1.54E-02 |
| 1299 | Map3k9 | MAP3K9 | 9.07E-02 | -1.95E-02 |
| 1300 | Gcsam | GCSAM | -1.08E-01 | 1.83E-02 |
| 1301 | Hsd17b1 | HSD17B1 | -1.56E-01 | 2.95E-02 |
| 1302 | Utp18 | UTP18 | 4.74E-02 | -4.17E-03 |
| 1303 | Zdhhc4 | ZDHHC4 | 3.70E-02 | -1.60E-02 |
| 1304 | Cxcl1 | CXCL2 | 4.38E-01 | 3.47E-01 |
| 1305 | Ywhaq | YWHAQ | 3.18E-02 | 5.87E-03 |
| 1306 | Kif1a | KIF1A | 1.67E-01 | 1.16E-01 |
| 1307 | Cspg4 | CSPG4 | 1.01E-01 | -1.62E-02 |
| 1308 | Abca2 | ABCA2 | 3.83E-02 | -4.06E-02 |
| 1309 | Adss | ADSS | 3.92E-02 | 2.57E-03 |
| 1310 | Zc3h6 | ZC3H6 | 1.76E-01 | -8.36E-02 |
| 1311 | Nkx3-1 | NKX3-1 | 2.00E-01 | -1.61E-01 |
| 1312 | Got1 | GOT1 | 5.86E-02 | -9.49E-03 |
| 1313 | Gla | GLA | 7.00E-02 | 2.50E-02 |
| 1314 | Sergef | SERGEF | 5.51E-02 | 1.96E-02 |
| 1315 | Pnpla6 | PNPLA6 | 3.66E-02 | -2.47E-02 |
| 1316 | Glrx5 | GLRX5 | 5.02E-02 | 5.31E-04 |
| 1317 | Eya4 | EYA4 | -1.51E-01 | 4.52E-02 |
| 1318 | Slc36a4 | SLC36A4 | 7.18E-02 | -1.94E-02 |
| 1319 | Prkar1a | PRKAR1A | 2.80E-02 | -2.20E-03 |
| 1320 | Ephb3 | EPHB3 | -1.88E-01 | 5.61E-02 |
| 1321 | Klhl21 | KLHL21 | -4.73E-02 | -1.70E-02 |
| 1322 | Aldh3b1 | ALDH3B1 | 1.42E-01 | 1.95E-02 |

| | | | | |
|---|---|---|---|---|
| 1323 | Wnt10a | WNT10A | 3.06E-01 | 1.20E-01 |
| 1324 | Kcnq1 | KCNQ1 | 5.71E-02 | 2.70E-02 |
| 1325 | Chmp1b | CHMP1B | 3.54E-02 | 1.99E-03 |
| 1326 | Ppm1l | PPM1L | -6.72E-02 | -1.72E-02 |
| 1327 | Itpk1 | ITPK1 | -4.21E-02 | -1.22E-03 |
| 1328 | Unc119b | UNC119B | 3.55E-02 | -1.55E-02 |
| 1329 | Nelfcd | NELFCD | 4.18E-02 | -5.14E-03 |
| 1330 | Rnf185 | RNF185 | 3.07E-02 | -1.23E-02 |
| 1331 | Rab5c | RAB5C | 4.90E-02 | -1.20E-02 |
| 1332 | Tceal8 | TCEAL8 | 5.33E-02 | 4.76E-04 |
| 1333 | Nsg1 | NSG1 | -1.90E-01 | -1.13E-02 |
| 1334 | Dapk2 | DAPK2 | -7.76E-02 | -5.77E-02 |
| 1335 | C1qa | C1QA | -1.19E-01 | 1.26E-01 |
| 1336 | Nrip2 | NRIP2 | -1.51E-01 | 4.78E-02 |
| 1337 | Mettl4 | METTL4 | -9.01E-02 | -1.50E-02 |
| 1338 | Cirbp | CIRBP | 1.08E-01 | -5.74E-02 |
| 1339 | S100a1 | S100A1 | 4.46E-02 | 2.24E-02 |
| 1340 | Cyp7b1 | CYP7B1 | -2.44E-01 | 2.29E-02 |
| 1341 | Sgk1 | SGK1 | 1.30E-01 | 2.49E-02 |
| 1342 | Pgs1 | PGS1 | 5.55E-02 | 4.75E-03 |
| 1343 | Bcl11a | BCL11A | -9.40E-02 | 2.15E-02 |
| 1344 | Pianp | PIANP | -8.73E-02 | 4.02E-02 |
| 1345 | Phlda3 | PHLDA3 | 1.53E-01 | 3.60E-02 |
| 1346 | Dok7 | DOK7 | -2.58E-01 | -1.91E-02 |
| 1347 | Itga1 | ITGA1 | -6.02E-02 | -2.34E-02 |
| 1348 | Kras | KRAS | 4.59E-02 | 1.03E-02 |
| 1349 | Cdc42bpb | CDC42BPB | 3.39E-02 | -8.00E-03 |
| 1350 | Fuz | FUZ | -5.72E-02 | -3.66E-02 |
| 1351 | Agxt2 | AGXT2 | 8.25E-02 | -1.51E-02 |
| 1352 | Ctso | CTSO | -5.14E-02 | -3.21E-03 |
| 1353 | Ubtf | UBTF | -3.42E-02 | 1.14E-03 |
| 1354 | Rdh5 | RDH5 | -1.26E-01 | -8.07E-02 |
| 1355 | Psd3 | PSD3 | -7.45E-02 | 3.58E-02 |
| 1356 | Mgarp | MGARP | 1.54E-01 | 1.37E-02 |
| 1357 | Prr5l | PRR5L | -1.58E-01 | 2.33E-02 |
| 1358 | Tab1 | TAB1 | 5.69E-02 | -4.69E-02 |
| 1359 | Rhob | RHOB | 1.03E-01 | 1.66E-02 |
| 1360 | Bpnt1 | BPNT1 | 4.39E-02 | -2.75E-02 |
| 1361 | Dusp15 | DUSP15 | -2.21E-01 | -1.35E-01 |
| 1362 | Evi2a | EVI2A | -1.73E-01 | 1.32E-01 |
| 1363 | Nusap1 | NUSAP1 | 3.50E-01 | 1.26E-01 |
| 1364 | Plcg2 | PLCG2 | -5.03E-02 | 6.85E-02 |
| 1365 | Nrp1 | NRP1 | -9.23E-02 | 2.35E-02 |
| 1366 | Ttc39b | TTC39B | 6.88E-02 | -1.22E-02 |
| 1367 | Ctsc | CTSC | -6.79E-02 | 8.94E-02 |
| 1368 | Nek6 | NEK6 | 8.88E-02 | 5.51E-02 |
| 1369 | Zmynd12 | ZMYND12 | 1.53E-01 | -8.67E-02 |
| 1370 | Chd4 | CHD4 | 3.44E-02 | 2.00E-04 |
| 1371 | Pknox1 | PKNOX1 | -6.48E-02 | -2.21E-02 |
| 1372 | Fkbp14 | FKBP14 | 7.68E-02 | -9.23E-04 |
| 1373 | Erp44 | ERP44 | -3.88E-02 | -8.92E-04 |
| 1374 | Akap10 | AKAP10 | -4.21E-02 | -3.54E-04 |
| 1375 | Sec22a | SEC22A | 3.97E-02 | -8.37E-03 |

| | | | | |
|---|---|---|---|---|
| 1376 | Ttc13 | TTC13 | 3.74E-02 | 3.09E-03 |
| 1377 | Slc6a6 | SLC6A6 | 8.03E-02 | -3.01E-02 |
| 1378 | Emg1 | EMG1 | -4.24E-02 | 5.37E-03 |
| 1379 | Lrpap1 | LRPAP1 | -5.10E-02 | -2.75E-02 |
| 1380 | Akirin1 | AKIRIN1 | 7.11E-02 | 4.89E-03 |
| 1381 | Angpt4 | ANGPT4 | 1.80E-01 | -8.45E-02 |
| 1382 | Abhd6 | ABHD6 | 6.12E-02 | 1.10E-02 |
| 1383 | Pdilt | PDILT | -1.63E-01 | 5.99E-03 |
| 1384 | Cish | CISH | -1.82E-01 | 3.78E-02 |
| 1385 | Thbs4 | THBS4 | -4.84E-01 | -6.45E-05 |
| 1386 | Ppp1r32 | PPP1R32 | 1.26E-01 | -6.13E-02 |
| 1387 | Xkr8 | XKR8 | -9.58E-02 | 2.75E-02 |
| 1388 | Anapc7 | ANAPC7 | 4.51E-02 | -4.93E-03 |
| 1389 | Cd93 | CD93 | 7.15E-02 | -1.09E-02 |
| 1390 | Sh3pxd2a | SH3PXD2A | 8.30E-02 | -3.19E-02 |
| 1391 | Upf2 | UPF2 | -4.96E-02 | 8.48E-03 |
| 1392 | Sgpl1 | SGPL1 | 3.61E-02 | -1.08E-02 |
| 1393 | Flrt3 | FLRT3 | -1.72E-01 | -1.66E-02 |
| 1394 | Cars | CARS | 5.47E-02 | 2.30E-02 |
| 1395 | Tmem126b | TMEM126B | -5.34E-02 | 3.43E-04 |
| 1396 | Sh3rf3 | SH3RF3 | 1.37E-01 | 5.66E-02 |
| 1397 | Sele | SELE | 2.29E-01 | 3.38E-02 |
| 1398 | Rgs3 | RGS3 | -8.96E-02 | -2.74E-02 |
| 1399 | Hdac6 | HDAC6 | 4.75E-02 | -2.05E-02 |
| 1400 | Dap | DAP | -5.18E-02 | 3.93E-02 |
| 1401 | Rheb | RHEB | 3.32E-02 | 1.62E-04 |
| 1402 | Nsf | NSF | 4.86E-02 | -2.39E-02 |
| 1403 | Mthfr | MTHFR | 8.40E-02 | -2.30E-02 |
| 1404 | Pknox2 | PKNOX2 | -1.52E-01 | 1.76E-04 |
| 1405 | Srd5a3 | SRD5A3 | 5.99E-02 | -7.47E-05 |
| 1406 | Itch | ITCH | 6.22E-02 | -1.10E-02 |
| 1407 | Mtrf1l | MTRF1L | -7.22E-02 | -2.56E-02 |
| 1408 | Cyth4 | CYTH4 | -1.21E-01 | 1.25E-01 |
| 1409 | Snx4 | SNX4 | 3.75E-02 | -1.52E-02 |
| 1410 | Zfp830 | ZNF830 | -6.54E-02 | -4.51E-03 |
| 1411 | Rnaseh2b | RNASEH2B | 7.99E-02 | 1.83E-02 |
| 1412 | Plekha5 | PLEKHA5 | 3.96E-02 | -8.17E-03 |
| 1413 | Rac1 | RAC1 | 2.87E-02 | -4.10E-03 |
| 1414 | Capza2 | CAPZA2 | 3.51E-02 | -6.18E-03 |
| 1415 | Rangap1 | RANGAP1 | 4.84E-02 | 1.24E-02 |
| 1416 | Cldnd2 | CLDND2 | -8.52E-02 | 1.14E-02 |
| 1417 | Chid1 | CHID1 | -4.85E-02 | -1.55E-02 |
| 1418 | Ywhaz | YWHAZ | 3.99E-02 | 1.04E-02 |
| 1419 | Rexo2 | REXO2 | 3.58E-02 | -3.64E-03 |
| 1420 | Etnk2 | ETNK2 | 1.38E-01 | -5.35E-02 |
| 1421 | Qsox1 | QSOX1 | 5.17E-02 | 8.49E-06 |
| 1422 | Acer2 | ACER2 | 1.15E-01 | -5.08E-04 |
| 1423 | Sh2d2a | SH2D2A | -1.21E-01 | 5.13E-02 |
| 1424 | Mtfp1 | MTFP1 | -6.29E-02 | 2.27E-02 |
| 1425 | Dennd4b | DENND4B | -5.95E-02 | -1.85E-02 |
| 1426 | Scyl3 | SCYL3 | 4.07E-02 | -2.04E-02 |
| 1427 | Pbxip1 | PBXIP1 | 2.59E-02 | 7.16E-03 |
| 1428 | Ppm1e | PPM1E | -1.41E-01 | 5.20E-03 |

| | | | | |
|---|---|---|---|---|
| 1429 | Zfp426 | ZNF426 | -4.02E-02 | 7.85E-03 |
| 1430 | Spryd3 | SPRYD3 | 3.81E-02 | -1.65E-02 |
| 1431 | Psmc5 | PSMC5 | 3.38E-02 | -9.93E-03 |
| 1432 | Habp4 | HABP4 | 3.92E-02 | -1.56E-02 |
| 1433 | Coro1a | CORO1A | -1.54E-01 | 1.70E-01 |
| 1434 | Pogz | POGZ | -4.30E-02 | 2.60E-03 |
| 1435 | Ugt2b37 | UGT2B28 | 3.10E-01 | 2.98E-01 |
| 1436 | Nln | NLN | 5.50E-02 | -1.62E-02 |
| 1437 | Glt1d1 | GLT1D1 | 1.59E-01 | -4.75E-02 |
| 1438 | Tmed5 | TMED5 | 5.00E-02 | 1.58E-02 |
| 1439 | Cep68 | CEP68 | -5.23E-02 | -1.30E-02 |
| 1440 | Traf3ip3 | TRAF3IP3 | -1.53E-01 | 1.24E-01 |
| 1441 | Rnf149 | RNF149 | 3.93E-02 | -3.13E-03 |
| 1442 | Pigv | PIGV | -5.09E-02 | -2.36E-02 |
| 1443 | Usp21 | USP21 | -4.72E-02 | -3.57E-03 |
| 1444 | Cebpb | CEBPB | 2.85E-01 | 3.31E-02 |
| 1445 | Abcb9 | ABCB9 | 5.24E-02 | -3.23E-02 |
| 1446 | Dpp3 | DPP3 | 4.47E-02 | -9.02E-03 |
| 1447 | Pclo | PCLO | -1.08E-01 | -3.58E-02 |
| 1448 | Slc39a8 | SLC39A8 | -1.18E-01 | -8.57E-02 |
| 1449 | Rnf220 | RNF220 | 2.75E-02 | -1.24E-02 |
| 1450 | Cplx3 | CPLX3 | -1.41E-01 | -2.72E-02 |
| 1451 | Nup93 | NUP93 | 5.61E-02 | 1.96E-02 |
| 1452 | Tpt1 | TPT1 | 4.76E-02 | 1.99E-02 |
| 1453 | Slc22a7 | SLC22A7 | -4.22E-01 | -2.33E-01 |
| 1454 | Nqo1 | NQO1 | 1.24E-01 | -4.02E-02 |
| 1455 | Setdb2 | SETDB2 | 1.03E-01 | -3.98E-02 |
| 1456 | Dync2h1 | DYNC2H1 | -7.36E-02 | -1.04E-02 |
| 1457 | Tnrc18 | TNRC18 | -4.73E-02 | -4.41E-03 |
| 1458 | Tnfrsf18 | TNFRSF18 | -9.16E-02 | 5.14E-02 |
| 1459 | Rab31 | RAB31 | 7.97E-02 | 3.45E-02 |
| 1460 | Ing1 | ING1 | -3.81E-02 | 5.49E-03 |
| 1461 | Angptl2 | ANGPTL2 | -1.42E-01 | -4.05E-02 |
| 1462 | Fnbp4 | FNBP4 | 4.06E-02 | -1.18E-02 |
| 1463 | Hsf1 | HSF1 | -4.19E-02 | 4.34E-03 |
| 1464 | Zfp128 | ZNF8 | -1.23E-01 | 2.35E-02 |
| 1465 | Slc27a2 | SLC27A2 | -5.50E-02 | -1.47E-02 |
| 1466 | Ppp2r5b | PPP2R5B | 7.22E-02 | 3.13E-02 |
| 1467 | Lgr4 | LGR4 | 5.96E-02 | -3.08E-02 |
| 1468 | Pcdhga5 | PCDHGA5 | -1.36E-01 | 2.02E-02 |
| 1469 | Zdhhc3 | ZDHHC3 | 2.78E-02 | 1.90E-04 |
| 1470 | Dhx32 | DHX32 | 4.74E-02 | -1.48E-02 |
| 1471 | Asah1 | ASAH1 | -4.10E-02 | 7.75E-03 |
| 1472 | Tbc1d7 | TBC1D7 | 5.58E-02 | -2.28E-02 |
| 1473 | Slc12a5 | SLC12A5 | -1.31E-01 | 4.09E-02 |
| 1474 | Nap1l3 | NAP1L3 | -1.39E-01 | 1.98E-02 |
| 1475 | Sh3bgrl2 | SH3BGRL2 | 5.71E-02 | 8.76E-03 |
| 1476 | Ubr3 | UBR3 | 4.45E-02 | -2.28E-02 |
| 1477 | Gja3 | GJA3 | -1.19E-01 | 4.09E-02 |
| 1478 | Cnot4 | CNOT4 | -3.78E-02 | 1.36E-02 |
| 1479 | Ltk | LTK | -1.76E-01 | 5.05E-02 |
| 1480 | Inf2 | INF2 | 7.25E-02 | 1.18E-02 |
| 1481 | Krt7 | KRT7 | 6.55E-02 | 3.20E-02 |

| | | | | |
|---|---|---|---|---|
| 1482 | Il13ra1 | IL13RA1 | -5.79E-02 | 1.58E-02 |
| 1483 | Tars | TARS | 3.60E-02 | -2.33E-03 |
| 1484 | Gapdh | GAPDH | 5.71E-02 | -6.37E-03 |
| 1485 | Galnt3 | GALNT3 | 6.71E-02 | -3.08E-02 |
| 1486 | Eif2ak1 | EIF2AK1 | 3.25E-02 | -1.29E-02 |
| 1487 | Nhsl1 | NHSL1 | -7.33E-02 | -4.83E-02 |
| 1488 | Chrm3 | CHRM3 | -1.81E-01 | -8.87E-02 |
| 1489 | Dkk2 | DKK2 | -8.53E-02 | -1.05E-03 |
| 1490 | Emx2 | EMX2 | -5.58E-02 | 1.58E-02 |
| 1491 | Cebpa | CEBPA | -1.19E-01 | 5.07E-02 |
| 1492 | Avpr1a | AVPR1A | -7.52E-02 | 1.27E-02 |
| 1493 | Pitpnb | PITPNB | 2.48E-02 | -6.54E-03 |
| 1494 | Psmd14 | PSMD14 | 4.53E-02 | -1.62E-03 |
| 1495 | Dnajb9 | DNAJB9 | 5.30E-02 | 2.89E-03 |
| 1496 | Gnb1 | GNB1 | 2.79E-02 | 1.76E-03 |
| 1497 | Nrep | NREP | -1.36E-01 | 1.51E-02 |
| 1498 | 2310022B05Rik | C1orf198 | 6.06E-02 | -5.80E-03 |
| 1499 | Rnf14 | RNF14 | 3.16E-02 | -1.90E-02 |
| 1500 | Mia3 | MIA3 | 3.58E-02 | -1.50E-02 |
| 1501 | Rasa2 | RASA2 | 6.13E-02 | 1.90E-02 |
| 1502 | Slc9a2 | SLC9A2 | -6.67E-02 | -8.18E-03 |
| 1503 | Rasgrp3 | RASGRP3 | -9.94E-02 | -1.07E-02 |
| 1504 | Usp15 | USP15 | 3.59E-02 | -2.00E-03 |
| 1505 | Mtf1 | MTF1 | -4.16E-02 | 1.30E-02 |
| 1506 | Trim35 | TRIM35 | -7.04E-02 | 5.92E-04 |
| 1507 | Vsig10l | VSIG10L | 9.33E-02 | -3.19E-02 |
| 1508 | Nae1 | NAE1 | 3.26E-02 | -7.16E-03 |
| 1509 | Hnrnpll | HNRNPLL | 3.84E-02 | -1.58E-02 |
| 1510 | Tenm4 | TENM4 | -1.25E-01 | -1.44E-01 |
| 1511 | Marveld1 | MARVELD1 | 6.63E-02 | 5.85E-04 |
| 1512 | Itga7 | ITGA7 | 8.31E-02 | -2.27E-02 |
| 1513 | Arhgef37 | ARHGEF37 | 2.87E-01 | -5.26E-02 |
| 1514 | Tmc5 | TMC5 | -2.15E-01 | -2.44E-01 |
| 1515 | Gc | GC | 1.44E-01 | 1.85E-02 |
| 1516 | Rif1 | RIF1 | -5.77E-02 | 6.10E-03 |
| 1517 | Hs3st3a1 | HS3ST3A1 | -1.52E-01 | -3.77E-02 |
| 1518 | Slc35f6 | SLC35F6 | 3.61E-02 | -4.35E-03 |
| 1519 | Mgl2 | CLEC10A | -2.12E-01 | 1.12E-01 |
| 1520 | Myl9 | MYL9 | 9.27E-02 | -2.68E-03 |
| 1521 | Gpr135 | GPR135 | 8.75E-02 | 4.79E-02 |
| 1522 | Pla2r1 | PLA2R1 | -1.53E-01 | -9.44E-03 |
| 1523 | Kdm4b | KDM4B | 3.76E-02 | -4.40E-03 |
| 1524 | Meis2 | MEIS2 | -7.41E-02 | -2.24E-02 |
| 1525 | Arhgef19 | ARHGEF19 | -1.19E-01 | 4.63E-02 |
| 1526 | Lars | LARS | 5.47E-02 | -1.68E-02 |
| 1527 | Snap29 | SNAP29 | -3.90E-02 | -3.26E-03 |
| 1528 | Eif4e | EIF4E | 3.66E-02 | 1.90E-03 |
| 1529 | Crocc | CROCC | -6.27E-02 | -2.59E-02 |
| 1530 | Il20rb | IL20RB | 1.84E-01 | 4.51E-03 |
| 1531 | Echdc3 | ECHDC3 | -4.52E-02 | -2.85E-02 |
| 1532 | Tespa1 | TESPA1 | -8.23E-02 | 4.60E-02 |
| 1533 | Lrp2 | LRP2 | -6.57E-02 | -3.15E-02 |
| 1534 | Lpcat3 | LPCAT3 | 3.32E-02 | -4.98E-03 |

| | | | | |
|---|---|---|---|---|
| 1535 | Pfn2 | PFN2 | 6.76E-02 | -2.43E-02 |
| 1536 | Sh3glb1 | SH3GLB1 | 2.46E-02 | 5.09E-03 |
| 1537 | Bdp1 | BDP1 | -5.89E-02 | 1.28E-02 |
| 1538 | Npl | NPL | 1.72E-01 | -4.19E-02 |
| 1539 | Ppp1r12c | PPP1R12C | 3.76E-02 | -2.82E-03 |
| 1540 | Tipin | TIPIN | 9.54E-02 | -3.11E-03 |
| 1541 | Cc2d1b | CC2D1B | 3.97E-02 | -6.90E-03 |
| 1542 | Hfe | HFE | 7.20E-02 | 3.72E-03 |
| 1543 | Dsc2 | DSC2 | -1.42E-01 | 1.08E-01 |
| 1544 | Cmtm4 | CMTM4 | 3.84E-02 | -2.37E-02 |
| 1545 | Pik3r3 | PIK3R3 | 1.22E-01 | 1.63E-02 |
| 1546 | Dnali1 | DNALI1 | -1.08E-01 | -1.33E-03 |
| 1547 | H2-Aa | HLA-DQA1 | -1.45E-01 | 1.03E-01 |
| 1548 | Chml | CHML | -9.31E-02 | 6.89E-02 |
| 1549 | Stk35 | STK35 | -5.54E-02 | -5.59E-03 |
| 1550 | Dcun1d4 | DCUN1D4 | 4.34E-02 | -2.37E-02 |
| 1551 | Atp6v1h | ATP6V1H | 4.19E-02 | -4.33E-06 |
| 1552 | Vav1 | VAV1 | -1.34E-01 | 1.41E-01 |
| 1553 | Gpr17 | GPR17 | 1.60E-01 | -1.38E-01 |
| 1554 | Nckipsd | NCKIPSD | 7.20E-02 | -3.92E-02 |
| 1555 | Mprip | MPRIP | 3.18E-02 | -1.17E-02 |
| 1556 | Slc4a7 | SLC4A7 | 1.63E-01 | -2.92E-02 |
| 1557 | Phf20 | PHF20 | 5.67E-02 | -1.66E-02 |
| 1558 | Cers4 | CERS4 | -1.10E-01 | -3.41E-02 |
| 1559 | Lyrm1 | LYRM1 | 1.03E-01 | -2.38E-03 |
| 1560 | Tmco4 | TMCO4 | 5.68E-02 | -6.81E-03 |
| 1561 | Cog2 | COG2 | 3.49E-02 | -1.83E-02 |
| 1562 | Farp2 | FARP2 | -4.59E-02 | -1.79E-02 |
| 1563 | Stxbp5 | STXBP5 | 4.01E-02 | -7.56E-03 |
| 1564 | Aif1l | AIF1L | 5.20E-02 | -1.69E-02 |
| 1565 | Fchsd2 | FCHSD2 | -4.93E-02 | -2.68E-02 |
| 1566 | Fam117b | FAM117B | 4.50E-02 | -6.42E-03 |
| 1567 | Bmp1 | BMP1 | -8.16E-02 | 2.96E-02 |
| 1568 | Gchfr | GCHFR | -7.57E-02 | -2.28E-02 |
| 1569 | Dtnb | DTNB | 5.25E-02 | -2.15E-02 |
| 1570 | Prkcq | PRKCQ | -1.52E-01 | -6.05E-02 |
| 1571 | Slc1a5 | SLC1A5 | -7.94E-02 | 3.30E-02 |
| 1572 | Lipg | LIPG | -2.65E-01 | -1.28E-01 |
| 1573 | Fam120c | FAM120C | -7.89E-02 | -1.86E-02 |
| 1574 | C2cd2 | C2CD2 | 4.36E-02 | -4.68E-03 |
| 1575 | Rffl | RFFL | -3.78E-02 | -7.70E-03 |
| 1576 | Pgpep1l | PGPEP1L | -1.38E-01 | 1.02E-02 |
| 1577 | Ttc9c | TTC9C | -6.02E-02 | 1.86E-02 |
| 1578 | Catsperg1 | CATSPERG | -1.43E-01 | -6.10E-03 |
| 1579 | Gtpbp3 | GTPBP3 | -3.77E-02 | -2.42E-02 |
| 1580 | Brap | BRAP | 3.71E-02 | -2.34E-02 |
| 1581 | Ubqln1 | UBQLN1 | 3.56E-02 | -5.59E-03 |
| 1582 | Lpgat1 | LPGAT1 | -4.50E-02 | -2.65E-02 |
| 1583 | Coprs | COPRS | -8.07E-02 | -8.74E-03 |
| 1584 | Hspb11 | HSPB11 | -7.46E-02 | 2.57E-02 |
| 1585 | Nr2c2ap | NR2C2AP | 6.07E-02 | 2.74E-03 |
| 1586 | Il2rb | IL2RB | -1.77E-01 | -1.16E-02 |
| 1587 | Htt | HTT | 4.13E-02 | -1.99E-02 |

| | | | | |
|---|---|---|---|---|
| 1588 | Rps27l | RPS27L | 4.23E-02 | -1.05E-02 |
| 1589 | Lcn2 | LCN2 | 4.39E-01 | 3.46E-01 |
| 1590 | Ache | ACHE | 1.68E-01 | -7.36E-02 |
| 1591 | Atp8b2 | ATP8B2 | -5.96E-02 | 3.06E-02 |
| 1592 | Ntmt1 | NTMT1 | 5.29E-02 | 1.84E-03 |
| 1593 | Slc10a3 | SLC10A3 | 5.57E-02 | 4.53E-03 |
| 1594 | Fam81a | FAM81A | -6.36E-02 | -3.42E-02 |
| 1595 | C7 | C7 | 2.54E-01 | 5.45E-02 |
| 1596 | Tmx2 | TMX2 | 3.63E-02 | -1.87E-02 |
| 1597 | Taf1c | TAF1C | -5.81E-02 | 2.97E-02 |
| 1598 | Rhobtb2 | RHOBTB2 | -6.43E-02 | -1.35E-02 |
| 1599 | Etnk1 | ETNK1 | -3.99E-02 | -2.58E-02 |
| 1600 | Asap3 | ASAP3 | -7.63E-02 | -6.44E-03 |
| 1601 | Drosha | DROSHA | -3.48E-02 | -9.06E-03 |
| 1602 | Pja1 | PJA1 | -2.93E-02 | -8.20E-03 |
| 1603 | Ptp4a1 | PTP4A1 | 7.88E-02 | 6.92E-02 |
| 1604 | Sec63 | SEC63 | 2.86E-02 | -1.60E-02 |
| 1605 | Arf1 | ARF1 | 3.27E-02 | -3.88E-03 |
| 1606 | Tfr2 | TFR2 | 2.04E-01 | 2.11E-01 |
| 1607 | Tmem98 | TMEM98 | -5.80E-02 | 3.05E-02 |
| 1608 | Tsku | TSKU | 2.13E-01 | -3.07E-02 |
| 1609 | Adora2b | ADORA2B | -1.48E-01 | 1.02E-01 |
| 1610 | Tnfaip1 | TNFAIP1 | 2.59E-02 | 3.17E-03 |
| 1611 | 1810010H24Rik | C17orf58 | 1.39E-01 | -7.12E-02 |
| 1612 | Cenpb | CENPB | 4.59E-02 | -6.52E-03 |
| 1613 | Pdcd6ip | PDCD6IP | 2.79E-02 | -2.18E-03 |
| 1614 | Ccl12 | CCL2 | -1.64E-01 | 1.27E-01 |
| 1615 | Zfp799 | ZNF799 | -6.72E-02 | -4.86E-02 |
| 1616 | Cdk18 | CDK18 | 6.11E-02 | -6.03E-03 |
| 1617 | Cdk8 | CDK8 | 5.91E-02 | -7.75E-03 |
| 1618 | Mllt3 | MLLT3 | -8.02E-02 | -1.71E-03 |
| 1619 | Srsf2 | SRSF2 | 3.16E-02 | -4.66E-03 |
| 1620 | Pon3 | PON3 | -7.74E-02 | 5.60E-02 |
| 1621 | Oxct1 | OXCT1 | -5.66E-02 | -3.05E-02 |
| 1622 | Arfgef2 | ARFGEF2 | 3.58E-02 | -1.88E-02 |
| 1623 | Dgke | DGKE | 8.46E-02 | -3.83E-02 |
| 1624 | Cep63 | CEP63 | -2.92E-02 | 4.42E-03 |
| 1625 | Mfap1a | MFAP1 | -4.39E-02 | 8.75E-03 |
| 1626 | Fap | FAP | -1.21E-01 | 2.43E-02 |
| 1627 | Rad23b | RAD23B | 3.20E-02 | 4.98E-04 |
| 1628 | Zfp653 | ZNF653 | 5.44E-02 | -1.33E-02 |
| 1629 | Faf1 | FAF1 | 3.89E-02 | -1.40E-02 |
| 1630 | Cib1 | CIB1 | 4.92E-02 | -3.88E-03 |
| 1631 | Lrp6 | LRP6 | -3.75E-02 | -2.48E-02 |
| 1632 | Nsun7 | NSUN7 | 1.19E-01 | 5.40E-02 |
| 1633 | Gnai1 | GNAI1 | 5.84E-02 | -1.80E-02 |
| 1634 | Actr2 | ACTR2 | 3.23E-02 | -6.63E-03 |
| 1635 | Brk1 | BRK1 | 3.51E-02 | -9.87E-03 |
| 1636 | Hoxa6 | HOXA6 | -1.35E-01 | 5.20E-04 |
| 1637 | Pex14 | PEX14 | 4.40E-02 | -1.58E-02 |
| 1638 | Hoxd8 | HOXD8 | 4.95E-02 | 7.04E-03 |
| 1639 | Dusp14 | DUSP14 | -1.02E-01 | 2.91E-02 |
| 1640 | Sipa1 | SIPA1 | -6.17E-02 | 1.59E-02 |

| | | | | |
|---|---|---|---|---|
| 1641 | Mdm1 | MDM1 | 8.87E-02 | 1.19E-02 |
| 1642 | Gcm1 | GCM1 | -2.28E-01 | -1.40E-01 |
| 1643 | Ndufa11 | NDUFA11 | 4.23E-02 | -2.09E-02 |
| 1644 | Atp8b4 | ATP8B4 | -1.50E-01 | -2.06E-02 |
| 1645 | Myh11 | MYH11 | 1.32E-01 | -9.20E-03 |
| 1646 | Plac8 | PLAC8 | -1.50E-01 | 1.89E-01 |
| 1647 | Sirpa | SIRPA | -7.85E-02 | -1.68E-02 |
| 1648 | Rad21 | RAD21 | 2.98E-02 | -1.32E-03 |
| 1649 | Fut9 | FUT9 | -8.03E-02 | -8.99E-03 |
| 1650 | Trim56 | TRIM56 | -3.65E-02 | -3.68E-02 |
| 1651 | Lpar1 | LPAR1 | 1.54E-01 | -1.71E-02 |
| 1652 | Fn3k | FN3K | -7.39E-02 | -3.10E-02 |
| 1653 | Rgs14 | RGS14 | -1.34E-01 | 2.59E-02 |
| 1654 | Maneal | MANEAL | -1.00E-01 | 5.53E-03 |
| 1655 | BC034090 | KIAA1614 | -4.23E-02 | -3.00E-02 |
| 1656 | Ap1s1 | AP1S1 | 4.30E-02 | -1.01E-03 |
| 1657 | Zfp142 | ZNF142 | -5.79E-02 | 1.53E-02 |
| 1658 | Arrdc3 | ARRDC3 | 6.87E-02 | -2.21E-02 |
| 1659 | Spata20 | SPATA20 | -1.28E-01 | 1.64E-03 |
| 1660 | Fcf1 | FCF1 | 4.48E-02 | -1.14E-02 |
| 1661 | Ube2g1 | UBE2G1 | 3.30E-02 | 9.86E-04 |
| 1662 | Ppm1g | PPM1G | 2.96E-02 | 7.38E-03 |
| 1663 | Ankrd28 | ANKRD28 | 5.34E-02 | -8.01E-03 |
| 1664 | Sf3a3 | SF3A3 | -3.03E-02 | -2.33E-03 |
| 1665 | Serpine1 | SERPINE1 | 2.75E-01 | 2.11E-01 |
| 1666 | Rab2b | RAB2B | 4.60E-02 | 9.11E-03 |
| 1667 | P4ha2 | P4HA2 | 1.09E-01 | 6.61E-03 |
| 1668 | Cd3d | CD3D | -1.20E-01 | 2.15E-02 |
| 1669 | Snx22 | SNX22 | -1.73E-01 | 3.77E-02 |
| 1670 | Zfp709 | ZNF443 | -9.38E-02 | 7.28E-03 |
| 1671 | Rasl12 | RASL12 | 1.35E-01 | 7.63E-02 |
| 1672 | Evpl | EVPL | -6.55E-02 | -1.18E-02 |
| 1673 | Iqcc | IQCC | -4.94E-02 | -2.31E-03 |
| 1674 | Lfng | LFNG | 1.31E-01 | -9.44E-02 |
| 1675 | Pxmp2 | PXMP2 | -7.56E-02 | -1.44E-02 |
| 1676 | Hddc2 | HDDC2 | 8.28E-02 | 1.01E-02 |
| 1677 | Exd1 | EXD1 | 1.52E-01 | -1.85E-02 |
| 1678 | Pde5a | PDE5A | 8.96E-02 | 2.52E-02 |
| 1679 | Ankrd55 | ANKRD55 | -6.33E-02 | 2.88E-02 |
| 1680 | Slc20a1 | SLC20A1 | 1.00E-01 | 1.53E-02 |
| 1681 | F13a1 | F13A1 | 2.93E-01 | 1.83E-01 |
| 1682 | Syap1 | SYAP1 | 3.61E-02 | -1.10E-02 |
| 1683 | Fem1c | FEM1C | -5.06E-02 | -4.17E-03 |
| 1684 | Tgfbrap1 | TGFBRAP1 | 4.54E-02 | -1.90E-02 |
| 1685 | Ppp2r1b | PPP2R1B | 3.98E-02 | 2.38E-02 |
| 1686 | Trmt6 | TRMT6 | 4.12E-02 | 6.14E-03 |
| 1687 | Rara | RARA | 7.07E-02 | -1.11E-02 |
| 1688 | Klrd1 | KLRD1 | -1.20E-01 | 2.26E-02 |
| 1689 | Slc25a21 | SLC25A21 | -1.03E-01 | -8.81E-02 |
| 1690 | Zfp688 | ZNF688 | 5.35E-02 | -2.93E-02 |
| 1691 | Mmaa | MMAA | -5.13E-02 | -4.30E-03 |
| 1692 | Camk2n2 | CAMK2N2 | 1.13E-01 | 8.13E-02 |
| 1693 | Map2k4 | MAP2K4 | 3.75E-02 | -1.30E-02 |

| | | | | |
|---|---|---|---|---|
| 1694 | Unc45a | UNC45A | 4.21E-02 | -1.32E-02 |
| 1695 | Acot3 | ACOT1 | -1.58E-01 | 5.30E-02 |
| 1696 | Ilk | ILK | 2.31E-02 | -8.99E-04 |
| 1697 | Vps33b | VPS33B | 4.79E-02 | -5.01E-03 |
| 1698 | Eif3m | EIF3M | 3.75E-02 | 4.54E-05 |
| 1699 | Pou2af1 | POU2AF1 | -8.33E-02 | -1.65E-02 |
| 1700 | Vps37a | VPS37A | 3.46E-02 | -1.09E-02 |
| 1701 | Tmem229b | TMEM229B | -1.35E-01 | 4.14E-02 |
| 1702 | Cbwd1 | CBWD1 | 5.27E-02 | -2.33E-02 |
| 1703 | Cul4a | CUL4A | 3.52E-02 | -1.06E-02 |
| 1704 | Sc5d | SC5D | -5.14E-02 | -8.73E-03 |
| 1705 | Fads1 | FADS1 | -6.33E-02 | -1.48E-02 |
| 1706 | Atp6v0e2 | ATP6V0E2 | -6.37E-02 | -3.60E-02 |
| 1707 | Tmem160 | TMEM160 | 5.42E-02 | -4.88E-03 |
| 1708 | Cd48 | CD48 | -1.46E-01 | 1.38E-01 |
| 1709 | Efnb2 | EFNB2 | -1.07E-01 | 5.18E-02 |
| 1710 | Mphosph9 | MPHOSPH9 | -5.01E-02 | 2.75E-02 |
| 1711 | Ube2e3 | UBE2E3 | 3.16E-02 | -1.80E-02 |
| 1712 | Bhlha15 | BHLHA15 | -1.32E-01 | 2.78E-02 |
| 1713 | Ube2a | UBE2A | 3.24E-02 | 6.89E-04 |
| 1714 | Il17rb | IL17RB | 1.09E-01 | -1.19E-03 |
| 1715 | Cd2 | CD2 | -1.49E-01 | 2.22E-02 |
| 1716 | Usp31 | USP31 | -5.96E-02 | 2.01E-02 |
| 1717 | Dlg5 | DLG5 | 5.05E-02 | -8.99E-03 |
| 1718 | Mlph | MLPH | 7.35E-02 | 5.44E-02 |
| 1719 | Zswim8 | ZSWIM8 | 2.78E-02 | -8.19E-03 |
| 1720 | Itgb8 | ITGB8 | -7.32E-02 | -2.69E-02 |
| 1721 | Blmh | BLMH | 3.65E-02 | -1.90E-02 |
| 1722 | Arhgap9 | ARHGAP9 | -1.23E-01 | 8.03E-02 |
| 1723 | Tmem64 | TMEM64 | -5.68E-02 | -2.20E-02 |
| 1724 | Vasn | VASN | 6.95E-02 | -4.37E-02 |
| 1725 | Rab11fip5 | RAB11FIP5 | 5.42E-02 | -1.55E-02 |
| 1726 | Lrrc34 | LRRC34 | -1.20E-01 | 1.33E-03 |
| 1727 | Sncaip | SNCAIP | -1.13E-01 | -6.08E-02 |
| 1728 | Commd3 | COMMD3 | 4.11E-02 | -1.59E-02 |
| 1729 | Pygo1 | PYGO1 | -1.05E-01 | -2.88E-03 |
| 1730 | Unkl | UNKL | -7.93E-02 | -2.34E-02 |
| 1731 | Eif3f | EIF3F | -3.60E-02 | 3.30E-02 |
| 1732 | D630003M21Rik | KIAA1755 | -1.84E-01 | 9.49E-02 |
| 1733 | Gna15 | GNA15 | -1.45E-01 | 6.59E-02 |
| 1734 | Bag1 | BAG1 | 4.07E-02 | -1.89E-02 |
| 1735 | Zcchc17 | ZCCHC17 | -3.86E-02 | 1.65E-02 |
| 1736 | Nlrc5 | NLRC5 | -2.34E-01 | 5.43E-02 |
| 1737 | Plec | PLEC | 4.04E-02 | 3.05E-02 |
| 1738 | Slc18a2 | SLC18A2 | -1.19E-01 | -2.01E-02 |
| 1739 | Rtca | RTCA | 3.61E-02 | -1.62E-03 |
| 1740 | Kif26b | KIF26B | 1.34E-01 | -1.21E-02 |
| 1741 | Mlf2 | MLF2 | 3.71E-02 | -4.10E-03 |
| 1742 | Actr6 | ACTR6 | 7.03E-02 | -6.48E-03 |
| 1743 | Rbm12b2 | RBM12B | -1.24E-01 | 8.24E-02 |
| 1744 | Utp14b | UTP14C | -1.13E-01 | -1.08E-02 |
| 1745 | Cast | CAST | 2.30E-02 | -4.45E-03 |
| 1746 | Hmbs | HMBS | 4.73E-02 | -6.33E-03 |

| | | | | |
|---|---|---|---|---|
| 1747 | Wdr70 | WDR70 | 4.61E-02 | 4.35E-03 |
| 1748 | Lman2l | LMAN2L | -4.99E-02 | -2.31E-02 |
| 1749 | Ttll11 | TTLL11 | 1.15E-01 | 8.87E-03 |
| 1750 | Ddi2 | DDI2 | -3.91E-02 | -2.25E-03 |
| 1751 | Uri1 | URI1 | 4.58E-02 | 1.98E-03 |
| 1752 | Lrp8 | LRP8 | -1.50E-01 | -4.14E-02 |
| 1753 | Efcab12 | EFCAB12 | -1.10E-01 | -4.01E-02 |
| 1754 | Eif2b1 | EIF2B1 | 3.66E-02 | 1.09E-02 |
| 1755 | Gpm6a | GPM6A | -8.42E-02 | -7.70E-02 |
| 1756 | Rimkla | RIMKLA | 1.07E-01 | 3.81E-02 |
| 1757 | Slc45a3 | SLC45A3 | 1.63E-01 | -1.06E-02 |
| 1758 | Ephx1 | EPHX1 | 8.76E-02 | -3.79E-02 |
| 1759 | Coro1c | CORO1C | 4.83E-02 | 2.69E-02 |
| 1760 | Usp22 | USP22 | 3.22E-02 | -1.93E-04 |
| 1761 | Sash3 | SASH3 | -1.49E-01 | 1.36E-01 |
| 1762 | Tacc1 | TACC1 | 3.98E-02 | -1.16E-02 |
| 1763 | Il11ra1 | IL11RA | -4.44E-02 | -2.92E-02 |
| 1764 | Sec23a | SEC23A | 3.78E-02 | -2.26E-02 |
| 1765 | Ankrd16 | ANKRD16 | 5.46E-02 | -7.71E-03 |
| 1766 | Atp9b | ATP9B | 2.41E-02 | -5.69E-03 |
| 1767 | Siva1 | SIVA1 | -8.40E-02 | 9.24E-03 |
| 1768 | Rab30 | RAB30 | 1.24E-01 | 4.49E-02 |
| 1769 | Mapk9 | MAPK9 | 2.69E-02 | -2.79E-02 |
| 1770 | Lrrc8c | LRRC8C | 5.65E-02 | -3.79E-03 |
| 1771 | Siae | SIAE | -4.32E-02 | -3.21E-02 |
| 1772 | Xpot | XPOT | 3.92E-02 | -2.00E-02 |
| 1773 | Nipa2 | NIPA2 | 3.42E-02 | 8.83E-03 |
| 1774 | Sae1 | SAE1 | 4.28E-02 | 6.73E-03 |
| 1775 | Cap1 | CAP1 | 2.72E-02 | 6.32E-03 |
| 1776 | Ensa | ENSA | 2.80E-02 | 6.54E-04 |
| 1777 | H2-Eb1 | HLA-DRB5 | -1.50E-01 | 1.03E-01 |
| 1778 | Cdh20 | CDH20 | -6.44E-02 | -6.37E-03 |
| 1779 | Hnf4g | HNF4G | -1.66E-01 | 5.96E-03 |
| 1780 | Wdr3 | WDR3 | -4.74E-02 | 2.40E-02 |
| 1781 | Icmt | ICMT | 3.39E-02 | 2.07E-03 |
| 1782 | Trim6 | TRIM6 | -1.23E-01 | 9.95E-02 |
| 1783 | Chpf2 | CHPF2 | 4.61E-02 | -8.22E-03 |
| 1784 | Insrr | INSRR | -9.87E-02 | 5.50E-02 |
| 1785 | Arhgap21 | ARHGAP21 | -3.15E-02 | -1.43E-02 |
| 1786 | Ppp4r1 | PPP4R1 | 3.53E-02 | -4.78E-03 |
| 1787 | Fgd4 | FGD4 | -7.71E-02 | -1.79E-02 |
| 1788 | Zbtb40 | ZBTB40 | -8.49E-02 | 2.61E-02 |
| 1789 | Ptpn9 | PTPN9 | 2.82E-02 | -2.27E-03 |
| 1790 | Mroh2a | MROH2A | 1.37E-01 | 1.03E-01 |
| 1791 | Slfn3 | SLFN12L | -1.85E-01 | 1.27E-02 |
| 1792 | Cyp27a1 | CYP27A1 | 8.80E-02 | -4.76E-02 |
| 1793 | Gsdmd | GSDMD | -7.83E-02 | 5.00E-02 |
| 1794 | Ndnf | NDNF | 1.34E-01 | -8.74E-03 |
| 1795 | Mtmr11 | MTMR11 | 6.26E-02 | -4.84E-03 |
| 1796 | Csdc2 | CSDC2 | 1.33E-01 | -6.49E-03 |
| 1797 | Pex6 | PEX6 | -4.72E-02 | -2.44E-02 |
| 1798 | Asb6 | ASB6 | 4.13E-02 | -9.22E-03 |
| 1799 | Fam219b | FAM219B | -5.29E-02 | -4.16E-03 |

| | | | | |
|---|---|---|---|---|
| 1800 | Agmat | AGMAT | -1.43E-01 | -2.25E-02 |
| 1801 | Slc17a4 | SLC17A4 | -2.02E-01 | -7.03E-03 |
| 1802 | Smarcd1 | SMARCD1 | 4.12E-02 | -2.20E-02 |
| 1803 | Vps53 | VPS53 | 3.24E-02 | -1.75E-02 |
| 1804 | H2-Oa | HLA-DOA | -9.37E-02 | 1.35E-02 |
| 1805 | Cask | CASK | 3.98E-02 | -4.45E-03 |
| 1806 | Gpt2 | GPT2 | -9.34E-02 | -2.62E-02 |
| 1807 | Dennd1b | DENND1B | 4.19E-02 | -1.33E-02 |
| 1808 | Tnfsf12 | TNFSF12 | -1.10E-01 | 4.19E-02 |
| 1809 | Gem | GEM | 3.39E-01 | -6.99E-03 |
| 1810 | Wdr93 | WDR93 | -1.18E-01 | -3.54E-02 |
| 1811 | Zfp317 | ZNF317 | -3.57E-02 | 9.91E-03 |
| 1812 | Slc25a30 | SLC25A30 | 1.10E-01 | 2.56E-02 |
| 1813 | Lrrc59 | LRRC59 | 3.42E-02 | 1.11E-02 |
| 1814 | P2rx3 | P2RX3 | -1.29E-01 | -3.65E-02 |
| 1815 | Herc2 | HERC2 | 2.87E-02 | -1.14E-02 |
| 1816 | Stx12 | STX12 | 3.11E-02 | 1.94E-03 |
| 1817 | Steap4 | STEAP4 | 1.31E-01 | 3.84E-02 |
| 1818 | Aasdh | AASDH | -4.93E-02 | -4.11E-03 |
| 1819 | Sp110 | SP110 | -9.30E-02 | 2.37E-03 |
| 1820 | Guca1b | GUCA1B | -1.55E-01 | 3.18E-02 |
| 1821 | Lgals3 | LGALS3 | 9.45E-02 | 9.48E-02 |
| 1822 | Clec4b1 | CLEC4C | -1.09E-01 | 7.68E-02 |
| 1823 | Sptan1 | SPTAN1 | 2.99E-02 | -2.22E-02 |
| 1824 | Trp53inp2 | TP53INP2 | 3.74E-02 | -5.09E-03 |
| 1825 | Tns1 | TNS1 | 4.64E-02 | -1.95E-02 |
| 1826 | Iscu | ISCU | 4.25E-02 | -2.70E-02 |
| 1827 | Phkb | PHKB | -3.86E-02 | -3.32E-02 |
| 1828 | Csrnp3 | CSRNP3 | -1.83E-01 | 1.39E-02 |
| 1829 | Arih2 | ARIH2 | 3.40E-02 | -1.06E-02 |
| 1830 | Ffar1 | FFAR1 | -5.51E-02 | 7.54E-03 |
| 1831 | Usp49 | USP49 | 8.47E-02 | -2.65E-02 |
| 1832 | Pou3f3 | POU3F3 | -9.25E-02 | -2.95E-02 |
| 1833 | Pde6g | PDE6G | -1.70E-01 | -2.78E-02 |
| 1834 | Ercc2 | ERCC2 | -2.68E-02 | -2.29E-03 |
| 1835 | Gipc1 | GIPC1 | 4.58E-02 | -3.82E-03 |
| 1836 | Slc25a12 | SLC25A12 | 3.55E-02 | -3.80E-02 |
| 1837 | Zfp800 | ZNF800 | -7.05E-02 | 1.22E-02 |
| 1838 | Klhl9 | KLHL9 | -4.51E-02 | 1.22E-02 |
| 1839 | Nr2f2 | NR2F2 | -4.53E-02 | -4.75E-03 |
| 1840 | Pnrc2 | PNRC2 | 3.54E-02 | -3.58E-05 |
| 1841 | Fyttd1 | FYTTD1 | 3.32E-02 | -6.08E-03 |
| 1842 | Dkc1 | DKC1 | -4.35E-02 | 8.01E-03 |
| 1843 | Isg20 | ISG20 | -1.92E-01 | 8.17E-02 |
| 1844 | Mgst3 | MGST3 | 4.02E-02 | -9.20E-03 |
| 1845 | Il2rg | IL2RG | -1.43E-01 | 9.87E-02 |
| 1846 | Ddx50 | DDX50 | 3.15E-02 | -6.77E-03 |
| 1847 | Med16 | MED16 | 4.21E-02 | -1.33E-02 |
| 1848 | Trim65 | TRIM65 | -1.11E-01 | 6.17E-02 |
| 1849 | Gpr173 | GPR173 | 1.23E-01 | -4.04E-02 |
| 1850 | Immp1l | IMMP1L | -4.59E-02 | -5.04E-03 |
| 1851 | Cfl1 | CFL1 | 3.61E-02 | 9.53E-03 |
| 1852 | Fas | FAS | 7.31E-02 | -1.86E-02 |

| | | | | |
|---|---|---|---|---|
| 1853 | Arhgdig | ARHGDIG | 2.07E-01 | 9.93E-02 |
| 1854 | Rfx4 | RFX4 | -1.22E-01 | -3.36E-02 |
| 1855 | Gas2l3 | GAS2L3 | 2.49E-01 | 7.34E-02 |
| 1856 | Ralgps2 | RALGPS2 | -4.62E-02 | -3.22E-02 |
| 1857 | Igsf11 | IGSF11 | 8.66E-02 | 4.16E-02 |
| 1858 | Hmx2 | HMX2 | -1.55E-01 | 1.10E-01 |
| 1859 | Sctr | SCTR | -1.31E-01 | 1.05E-01 |
| 1860 | Gm128 | C1orf56 | 2.03E-01 | -7.52E-02 |
| 1861 | Cog7 | COG7 | 3.58E-02 | -1.35E-02 |
| 1862 | Kif23 | KIF23 | 2.49E-01 | 4.60E-02 |
| 1863 | Amer2 | AMER2 | -9.73E-02 | -1.83E-02 |
| 1864 | Dock2 | DOCK2 | -1.75E-01 | 1.48E-01 |
| 1865 | Eef1d | EEF1D | -3.16E-02 | 8.56E-03 |
| 1866 | Efhd1 | EFHD1 | 5.92E-02 | -2.56E-02 |
| 1867 | Psmb8 | PSMB8 | -1.78E-01 | 1.21E-01 |
| 1868 | Kifc5b | KIFC1 | 1.65E-01 | 2.20E-02 |
| 1869 | Btbd1 | BTBD1 | 3.58E-02 | 4.24E-03 |
| 1870 | Kat2b | KAT2B | 3.60E-02 | -2.71E-02 |
| 1871 | Derl1 | DERL1 | 3.19E-02 | -1.69E-02 |
| 1872 | Ptafr | PTAFR | -1.80E-01 | 1.46E-01 |
| 1873 | Mep1b | MEP1B | -1.77E-01 | -1.12E-01 |
| 1874 | Id4 | ID4 | -1.41E-01 | -9.99E-03 |
| 1875 | Mapk12 | MAPK12 | -8.91E-02 | -4.71E-05 |
| 1876 | Bbox1 | BBOX1 | 1.97E-01 | -8.72E-02 |
| 1877 | Mbtps2 | MBTPS2 | 4.54E-02 | -9.30E-03 |
| 1878 | Cdk5rap3 | CDK5RAP3 | 4.56E-02 | -2.15E-02 |
| 1879 | Robo3 | ROBO3 | -2.16E-01 | -1.16E-01 |
| 1880 | Tesk1 | TESK1 | 2.75E-02 | -4.77E-03 |
| 1881 | Kcnj11 | KCNJ11 | -1.38E-01 | 2.29E-02 |
| 1882 | Ift80 | IFT80 | -4.68E-02 | 2.65E-03 |
| 1883 | Cd151 | CD151 | 2.76E-02 | 3.57E-03 |
| 1884 | Mbd1 | MBD1 | 4.30E-02 | 4.44E-03 |
| 1885 | Nt5c | NT5C | 3.83E-02 | 1.38E-02 |
| 1886 | Golga5 | GOLGA5 | 2.90E-02 | -1.26E-02 |
| 1887 | Atp2b2 | ATP2B2 | -9.89E-02 | 9.60E-03 |
| 1888 | Cabyr | CABYR | 7.66E-02 | 6.88E-03 |
| 1889 | Cxcr6 | CXCR6 | -2.13E-01 | 4.05E-02 |
| 1890 | Slc7a5 | SLC7A5 | 1.55E-01 | -2.69E-02 |
| 1891 | Dnajc24 | DNAJC24 | -5.66E-02 | 2.52E-02 |
| 1892 | Tmem53 | TMEM53 | 7.52E-02 | -2.30E-02 |
| 1893 | Ndrg1 | NDRG1 | 9.41E-02 | 1.50E-02 |
| 1894 | Rab18 | RAB18 | 3.20E-02 | -6.60E-03 |
| 1895 | Eaf1 | EAF1 | 2.73E-02 | -1.04E-02 |
| 1896 | Ankrd26 | ANKRD26 | -8.17E-02 | 3.71E-03 |
| 1897 | Ikzf1 | IKZF1 | -1.88E-01 | 9.53E-02 |
| 1898 | Tnfsf10 | TNFSF10 | -1.28E-01 | -1.92E-02 |
| 1899 | Smco3 | SMCO3 | -7.25E-02 | 1.18E-02 |
| 1900 | Necap2 | NECAP2 | 2.82E-02 | -5.59E-04 |
| 1901 | Sec24d | SEC24D | 4.68E-02 | -2.08E-02 |
| 1902 | Rundc3a | RUNDC3A | 1.89E-01 | 9.25E-02 |
| 1903 | Tamm41 | TAMM41 | 5.89E-02 | 3.67E-05 |
| 1904 | Plekha8 | PLEKHA8 | 4.50E-02 | 1.15E-02 |
| 1905 | Bpgm | BPGM | 3.93E-02 | 4.53E-03 |

| | | | | |
|---|---|---|---|---|
| 1906 | C1ql4 | C1QL4 | 1.03E-01 | -4.24E-02 |
| 1907 | Slc27a1 | SLC27A1 | -5.79E-02 | -8.43E-03 |
| 1908 | Unc13c | UNC13C | -1.46E-01 | -5.68E-02 |
| 1909 | Ubr4 | UBR4 | 2.77E-02 | -1.38E-02 |
| 1910 | Ndufb6 | NDUFB6 | 4.20E-02 | -4.37E-03 |
| 1911 | Bop1 | BOP1 | 3.61E-02 | -4.97E-03 |
| 1912 | Mmp25 | MMP25 | -7.59E-02 | 2.55E-02 |
| 1913 | Akna | AKNA | -9.59E-02 | 5.78E-02 |
| 1914 | G0s2 | G0S2 | 1.60E-01 | 6.56E-02 |
| 1915 | Rnf126 | RNF126 | 4.08E-02 | -3.77E-04 |
| 1916 | Foxo3 | FOXO3 | 7.24E-02 | -3.44E-02 |
| 1917 | Zfp39 | ZNF717 | -8.21E-02 | -2.47E-02 |
| 1918 | Gipc3 | GIPC3 | -1.29E-01 | 3.08E-03 |
| 1919 | Cyp2c38 | CYP2C8 | 9.85E-02 | -1.76E-02 |
| 1920 | Slc30a8 | SLC30A8 | 5.24E-02 | -7.19E-03 |
| 1921 | Eno2 | ENO2 | 1.45E-01 | 1.47E-01 |
| 1922 | Prpf3 | PRPF3 | -4.55E-02 | 1.40E-02 |
| 1923 | Psmd1 | PSMD1 | 3.96E-02 | -1.83E-02 |
| 1924 | Pced1a | PCED1A | -4.44E-02 | -7.17E-03 |
| 1925 | Adamts4 | ADAMTS4 | 2.31E-01 | 1.33E-01 |
| 1926 | Slc22a6 | SLC22A6 | 7.88E-02 | -7.06E-03 |
| 1927 | N4bp1 | N4BP1 | -3.09E-02 | 7.61E-03 |
| 1928 | Esrp1 | ESRP1 | -1.06E-01 | -1.54E-03 |
| 1929 | St6galnac2 | ST6GALNAC2 | -5.52E-02 | 2.03E-02 |
| 1930 | Fbxl4 | FBXL4 | 3.77E-02 | -1.87E-02 |
| 1931 | Ddit4 | DDIT4 | 2.63E-01 | -4.21E-02 |
| 1932 | Ptpro | PTPRO | -1.30E-01 | -9.22E-03 |
| 1933 | Prkab1 | PRKAB1 | 4.02E-02 | 3.13E-04 |
| 1934 | Cd274 | CD274 | -1.65E-01 | -1.09E-02 |
| 1935 | Dlgap4 | DLGAP4 | 3.46E-02 | 6.62E-03 |
| 1936 | Fgf9 | FGF9 | -8.50E-02 | -1.17E-02 |
| 1937 | Mtdh | MTDH | 3.90E-02 | 2.42E-04 |
| 1938 | Ufc1 | UFC1 | 3.27E-02 | -4.65E-03 |
| 1939 | Edn1 | EDN1 | 1.40E-01 | 1.27E-01 |
| 1940 | Slc17a5 | SLC17A5 | 5.21E-02 | -2.25E-02 |
| 1941 | Ubxn2a | UBXN2A | 4.24E-02 | -1.73E-02 |
| 1942 | Itm2b | ITM2B | 2.87E-02 | -1.34E-02 |
| 1943 | Utp14a | UTP14A | -4.83E-02 | 5.40E-02 |
| 1944 | Synpo2l | SYNPO2L | 1.42E-01 | -2.35E-02 |
| 1945 | Lgals4 | LGALS4 | 1.20E-01 | -2.14E-02 |
| 1946 | Rragc | RRAGC | 2.99E-02 | -4.93E-03 |
| 1947 | Endov | ENDOV | 4.08E-02 | -2.64E-02 |
| 1948 | Slc2a5 | SLC2A5 | 9.27E-02 | -3.59E-02 |
| 1949 | Arrb2 | ARRB2 | -5.09E-02 | 4.48E-02 |
| 1950 | Ndufb4 | NDUFB4 | 4.40E-02 | -2.18E-02 |
| 1951 | Cltb | CLTB | 4.39E-02 | -5.62E-03 |
| 1952 | Pltp | PLTP | -1.41E-01 | 2.80E-02 |
| 1953 | St6galnac3 | ST6GALNAC3 | -1.04E-01 | -6.49E-02 |
| 1954 | Nfya | NFYA | -6.68E-02 | 3.44E-02 |
| 1955 | Birc3 | BIRC3 | 8.12E-02 | 5.73E-02 |
| 1956 | Dnase1l1 | DNASE1L1 | -4.93E-02 | -7.83E-03 |
| 1957 | Pik3cd | PIK3CD | -1.19E-01 | 7.57E-02 |
| 1958 | Atp5l | ATP5L | 3.94E-02 | -1.01E-02 |

| | | | | |
|---|---|---|---|---|
| 1959 | Ankrd40 | ANKRD40 | 2.64E-02 | -2.67E-02 |
| 1960 | Lcp2 | LCP2 | -2.22E-01 | 2.32E-01 |
| 1961 | Ppp3ca | PPP3CA | 3.14E-02 | -1.41E-02 |
| 1962 | Tnfaip8l2 | TNFAIP8L2 | -1.61E-01 | 5.20E-02 |
| 1963 | Lamc2 | LAMC2 | 6.57E-02 | 3.22E-02 |
| 1964 | Zfp445 | ZNF445 | -3.49E-02 | -6.35E-03 |
| 1965 | Lmx1b | LMX1B | -1.22E-01 | -5.16E-02 |
| 1966 | Fbxo6 | FBXO6 | 5.42E-02 | 9.77E-05 |
| 1967 | Msrb2 | MSRB2 | -3.86E-02 | -2.26E-02 |
| 1968 | Mlxip | MLXIP | 4.55E-02 | -1.60E-02 |
| 1969 | Gm14305 | ZNF442 | -1.04E-01 | 3.81E-02 |
| 1970 | Pde4dip | PDE4DIP | 4.21E-02 | -1.68E-02 |
| 1971 | Cep85 | CEP85 | 7.38E-02 | 2.38E-03 |
| 1972 | Rasgrp4 | RASGRP4 | -1.58E-01 | 9.43E-02 |
| 1973 | Ctdnep1 | CTDNEP1 | -2.81E-02 | 1.33E-02 |
| 1974 | Cox6a1 | COX6A1 | 3.95E-02 | -1.22E-02 |
| 1975 | Tspan4 | TSPAN4 | -6.27E-02 | 3.83E-02 |
| 1976 | Sfmbt1 | SFMBT1 | 4.62E-02 | -2.45E-02 |
| 1977 | Rad1 | RAD1 | -5.61E-02 | 5.72E-03 |
| 1978 | Lyst | LYST | -4.15E-02 | -5.00E-03 |
| 1979 | Prkd3 | PRKD3 | -4.13E-02 | -3.42E-03 |
| 1980 | Faah | FAAH | 4.23E-02 | -3.44E-03 |
| 1981 | Gabrr2 | GABRR2 | -1.57E-01 | 5.79E-03 |
| 1982 | Tmem97 | TMEM97 | -6.40E-02 | -2.33E-02 |
| 1983 | 4933434E20Rik | C1orf43 | -3.09E-02 | 2.13E-03 |
| 1984 | Edar | EDAR | -1.62E-01 | -9.70E-02 |
| 1985 | Mxi1 | MXI1 | 3.49E-02 | -2.46E-02 |
| 1986 | Tnfsf13 | TNFSF13 | -1.26E-01 | 1.80E-02 |
| 1987 | Trp53bp1 | TP53BP1 | -4.49E-02 | 3.16E-03 |
| 1988 | Ppp2r5e | PPP2R5E | 6.43E-02 | -2.21E-02 |
| 1989 | Kdm3a | KDM3A | 4.59E-02 | -1.77E-02 |
| 1990 | Tdg | TDG | 5.10E-02 | 2.95E-03 |
| 1991 | Adarb1 | ADARB1 | 7.47E-02 | -6.45E-03 |
| 1992 | Hook1 | HOOK1 | 5.78E-02 | -2.28E-02 |
| 1993 | Eif2b2 | EIF2B2 | 3.72E-02 | -5.32E-03 |
| 1994 | Rbp2 | RBP2 | 7.89E-02 | -1.56E-02 |
| 1995 | Kpnb1 | KPNB1 | 2.80E-02 | 2.31E-03 |
| 1996 | Lzts3 | LZTS3 | -4.37E-02 | -3.95E-02 |
| 1997 | Arhgap25 | ARHGAP25 | -9.73E-02 | 1.42E-02 |
| 1998 | Lrp2bp | LRP2BP | 1.23E-01 | -2.51E-02 |
| 1999 | Ndfip2 | NDFIP2 | 3.38E-02 | -1.16E-02 |
| 2000 | Lrrc28 | LRRC28 | 6.29E-02 | 9.68E-03 |
| 2001 | Klhdc8a | KLHDC8A | 9.97E-02 | -1.38E-02 |
| 2002 | Prmt1 | PRMT1 | 2.99E-02 | 1.14E-02 |
| 2003 | Cep76 | CEP76 | 6.44E-02 | 2.19E-02 |
| 2004 | Sgpp1 | SGPP1 | 6.59E-02 | -2.64E-02 |
| 2005 | Lyplal1 | LYPLAL1 | -5.17E-02 | -4.33E-02 |
| 2006 | Gmppa | GMPPA | 3.29E-02 | -1.58E-02 |
| 2007 | Emc3 | EMC3 | 2.87E-02 | 6.17E-04 |
| 2008 | Selplg | SELPLG | -1.41E-01 | 1.18E-01 |
| 2009 | Magi1 | MAGI1 | -3.93E-02 | -9.20E-03 |
| 2010 | Xcl1 | XCL1 | -7.22E-02 | 2.94E-02 |
| 2011 | Tfcp2l1 | TFCP2L1 | 5.56E-02 | 6.37E-04 |

| | | | | |
|---|---|---|---|---|
| 2012 | Aebp1 | AEBP1 | -9.82E-02 | 3.77E-02 |
| 2013 | Casr | CASR | -8.55E-02 | -3.07E-02 |
| 2014 | Chmp4b | CHMP4B | 2.83E-02 | -4.62E-03 |
| 2015 | Zbtb26 | ZBTB26 | -1.35E-01 | 2.33E-02 |
| 2016 | Bcar3 | BCAR3 | 3.79E-02 | -8.22E-03 |
| 2017 | Lcor | LCOR | -1.19E-01 | -1.70E-02 |
| 2018 | Zfp346 | ZNF346 | 3.81E-02 | -1.36E-02 |
| 2019 | Srsf7 | SRSF7 | -4.88E-02 | 2.14E-02 |
| 2020 | Efcab5 | EFCAB5 | 1.30E-01 | -3.96E-02 |
| 2021 | Vstm5 | VSTM5 | -6.01E-02 | -8.67E-02 |
| 2022 | Epsti1 | EPSTI1 | -1.53E-01 | 4.64E-02 |
| 2023 | Cnpy3 | CNPY3 | -2.92E-02 | 1.81E-04 |
| 2024 | Rrnad1 | RRNAD1 | -5.28E-02 | 5.57E-03 |
| 2025 | Parl | PARL | 3.40E-02 | -9.13E-03 |
| 2026 | Tlr1 | TLR1 | -1.46E-01 | 1.66E-01 |
| 2027 | Eif5a2 | EIF5A2 | 7.70E-02 | 3.07E-02 |
| 2028 | Dlec1 | DLEC1 | -4.15E-02 | -2.16E-02 |
| 2029 | Anapc4 | ANAPC4 | 3.01E-02 | -1.86E-03 |
| 2030 | Nupl2 | NUPL2 | 6.57E-02 | -1.37E-02 |
| 2031 | Lnpep | LNPEP | -9.69E-02 | -6.12E-03 |
| 2032 | Mppe1 | MPPE1 | -4.10E-02 | -1.47E-02 |
| 2033 | Prickle1 | PRICKLE1 | -9.71E-02 | 3.67E-02 |
| 2034 | Hipk4 | HIPK4 | 1.26E-01 | -7.82E-02 |
| 2035 | Tmem200b | TMEM200B | 1.34E-01 | 1.28E-02 |
| 2036 | Ankrd9 | ANKRD9 | 1.48E-01 | 1.24E-02 |
| 2037 | Scarf1 | SCARF1 | -6.76E-02 | 2.06E-03 |
| 2038 | Nphp4 | NPHP4 | 9.29E-02 | 2.75E-02 |
| 2039 | Mdm2 | MDM2 | 3.98E-02 | -1.26E-02 |
| 2040 | Sfxn1 | SFXN1 | 6.01E-02 | -1.73E-02 |
| 2041 | Fcho1 | FCHO1 | -1.11E-01 | -2.11E-02 |
| 2042 | Wdr45b | WDR45B | 2.31E-02 | -8.91E-03 |
| 2043 | Cfp | CFP | -1.10E-01 | 1.13E-01 |
| 2044 | Exoc3l | EXOC3L1 | -1.02E-01 | 1.63E-02 |
| 2045 | Lrrc8a | LRRC8A | 7.89E-02 | -3.50E-03 |
| 2046 | Fubp1 | FUBP1 | -4.62E-02 | 1.96E-02 |
| 2047 | Anks1b | ANKS1B | -2.74E-01 | 5.52E-02 |
| 2048 | Atp6v1d | ATP6V1D | 3.09E-02 | -3.41E-03 |
| 2049 | Ahr | AHR | -9.52E-02 | 3.95E-02 |
| 2050 | Casd1 | CASD1 | -4.08E-02 | -1.67E-02 |
| 2051 | Uhrf1bp1 | UHRF1BP1 | 3.13E-02 | -8.08E-03 |
| 2052 | Tigd3 | TIGD3 | 1.20E-01 | -4.52E-02 |
| 2053 | Lpar5 | LPAR5 | -6.25E-02 | 3.52E-02 |
| 2054 | Kcna7 | KCNA7 | 5.18E-02 | -2.22E-02 |
| 2055 | Aspdh | ASPDH | -9.00E-02 | -2.01E-02 |
| 2056 | Asb3 | GPR75-ASB3 | -6.77E-02 | 1.20E-02 |
| 2057 | Asb3 | ASB3 | -6.77E-02 | 1.20E-02 |
| 2058 | Slc25a16 | SLC25A16 | 4.96E-02 | -2.78E-02 |
| 2059 | Nars | NARS | 3.31E-02 | -6.95E-03 |
| 2060 | Serp1 | SERP1 | 3.47E-02 | -1.35E-02 |
| 2061 | Dnajc18 | DNAJC18 | 3.48E-02 | -1.50E-02 |
| 2062 | Tubb2b | TUBB2B | 1.44E-01 | 1.11E-01 |
| 2063 | Plagl1 | PLAGL1 | -6.01E-02 | -4.84E-02 |
| 2064 | Armc6 | ARMC6 | -5.09E-02 | 1.59E-02 |

| | | | | |
|---|---|---|---|---|
| 2065 | Invs | INVS | -6.43E-02 | -1.92E-03 |
| 2066 | Fam78a | FAM78A | 1.41E-01 | 3.79E-03 |
| 2067 | Endog | ENDOG | 5.46E-02 | 5.89E-03 |
| 2068 | Csde1 | CSDE1 | 2.44E-02 | -7.88E-03 |
| 2069 | Cdk11b | CDK11A | 2.50E-02 | 1.02E-03 |
| 2070 | Rttn | RTTN | -7.94E-02 | 1.39E-03 |
| 2071 | Megf9 | MEGF9 | 1.06E-01 | -1.44E-02 |
| 2072 | Spink8 | SPINK8 | 1.54E-01 | 5.02E-02 |
| 2073 | Pak1ip1 | PAK1IP1 | 3.10E-02 | 2.07E-03 |
| 2074 | Rergl | RERGL | 1.56E-01 | 9.61E-02 |
| 2075 | Mroh1 | MROH1 | 3.28E-02 | -1.69E-02 |
| 2076 | Adap1 | ADAP1 | 8.20E-02 | -2.69E-02 |
| 2077 | Banp | BANP | -1.82E-01 | 7.51E-02 |
| 2078 | Vwa5a | VWA5A | -3.95E-02 | 1.42E-02 |
| 2079 | Pafah2 | PAFAH2 | 4.76E-02 | -8.65E-03 |
| 2080 | Krtcap2 | KRTCAP2 | -3.39E-02 | 1.03E-02 |
| 2081 | Cox7a2 | COX7A2 | 3.62E-02 | -1.38E-02 |
| 2082 | Ube2j1 | UBE2J1 | 3.58E-02 | 8.77E-03 |
| 2083 | Harbi1 | HARBI1 | -4.84E-02 | -5.06E-04 |
| 2084 | Gys1 | GYS1 | 4.43E-02 | 7.17E-04 |
| 2085 | Rxrb | RXRB | -2.90E-02 | -5.58E-03 |
| 2086 | Hpcal4 | HPCAL4 | -1.10E-01 | 1.26E-01 |
| 2087 | Eif4e3 | EIF4E3 | -8.00E-02 | 5.12E-02 |
| 2088 | Pus10 | PUS10 | 3.82E-02 | -5.96E-03 |
| 2089 | Pcdhgb7 | PCDHGB7 | -1.28E-01 | 5.21E-02 |
| 2090 | Prodh | LOC102724788 | 7.65E-02 | -1.62E-02 |
| 2091 | Prodh | PRODH | 7.65E-02 | -1.62E-02 |
| 2092 | Enpp3 | ENPP3 | -6.23E-02 | 2.48E-02 |
| 2093 | Ptbp2 | PTBP2 | 4.43E-02 | -2.24E-02 |
| 2094 | Papln | PAPLN | 1.49E-01 | 2.35E-02 |
| 2095 | Avpi1 | AVPI1 | 5.85E-02 | 1.30E-02 |
| 2096 | Trmt12 | TRMT12 | -6.67E-02 | -1.54E-02 |
| 2097 | Slc16a3 | SLC16A3 | 3.28E-01 | 1.31E-01 |
| 2098 | Slc6a15 | SLC6A15 | -8.58E-02 | -3.41E-02 |
| 2099 | Nek2 | NEK2 | 2.49E-01 | 5.89E-02 |
| 2100 | Gde1 | GDE1 | 3.19E-02 | -4.05E-03 |
| 2101 | Napsa | NAPSA | -9.09E-02 | -7.73E-02 |
| 2102 | Rcc2 | RCC2 | 3.33E-02 | 6.10E-03 |
| 2103 | Rhod | RHOD | 5.36E-02 | 1.43E-02 |
| 2104 | Wars | WARS | 3.95E-02 | -8.77E-03 |
| 2105 | Hoxb4 | HOXB4 | -9.86E-02 | 1.05E-02 |
| 2106 | Zfp180 | ZNF180 | -3.92E-02 | 3.01E-03 |
| 2107 | Prkx | PRKX | 3.85E-02 | 1.20E-02 |
| 2108 | Pigt | PIGT | 2.89E-02 | -3.04E-03 |
| 2109 | Igsf8 | IGSF8 | -5.83E-02 | 1.68E-02 |
| 2110 | Mcm10 | MCM10 | 1.50E-01 | 4.32E-03 |
| 2111 | Nup160 | NUP160 | -4.78E-02 | 1.10E-02 |
| 2112 | Eci1 | ECI1 | 7.24E-02 | -2.89E-03 |
| 2113 | Siah2 | SIAH2 | -5.18E-02 | -1.31E-02 |
| 2114 | Arl6ip1 | ARL6IP1 | 4.38E-02 | -1.22E-02 |
| 2115 | Cinp | CINP | 4.29E-02 | 1.02E-02 |
| 2116 | Sema6a | SEMA6A | -8.39E-02 | 6.48E-02 |
| 2117 | Prkcd | PRKCD | 2.67E-02 | -1.35E-02 |

| | | | | |
|---|---|---|---|---|
| 2118 | Gabarapl2 | GABARAPL2 | 2.68E-02 | -8.17E-03 |
| 2119 | Vcp | VCP | 3.54E-02 | -1.19E-02 |
| 2120 | Nudt17 | NUDT17 | -1.43E-01 | -3.24E-02 |
| 2121 | Cyp27b1 | CYP27B1 | 3.63E-01 | 8.22E-02 |
| 2122 | Zpld1 | ZPLD1 | -1.35E-01 | 8.78E-03 |
| 2123 | Rbbp9 | RBBP9 | -3.79E-02 | -1.26E-02 |
| 2124 | Synj1 | SYNJ1 | 3.61E-02 | -9.62E-03 |
| 2125 | Stard3nl | STARD3NL | -3.06E-02 | 2.59E-04 |
| 2126 | Kctd15 | KCTD15 | 7.35E-02 | 2.26E-02 |
| 2127 | Trip4 | TRIP4 | 3.21E-02 | -1.53E-02 |
| 2128 | Leng8 | LENG8 | 3.01E-02 | -1.14E-02 |
| 2129 | Dera | DERA | 6.19E-02 | -2.37E-02 |
| 2130 | Mgrn1 | MGRN1 | 4.15E-02 | -1.13E-02 |
| 2131 | Slc35d2 | SLC35D2 | 7.58E-02 | -1.71E-02 |
| 2132 | Tkt | TKT | 6.85E-02 | -1.80E-02 |
| 2133 | Rdh10 | RDH10 | 4.17E-02 | 1.60E-02 |
| 2134 | Casp2 | CASP2 | -3.63E-02 | -6.70E-03 |
| 2135 | Zfand4 | ZFAND4 | 1.32E-01 | 2.40E-02 |
| 2136 | Nploc4 | NPLOC4 | 3.06E-02 | -9.37E-03 |
| 2137 | Atp6v1c1 | ATP6V1C1 | 3.49E-02 | -9.51E-04 |
| 2138 | Dupd1 | DUPD1 | 6.30E-02 | -2.48E-02 |
| 2139 | Bod1l | BOD1L1 | -3.34E-02 | -1.22E-02 |
| 2140 | Tmeff2 | TMEFF2 | 1.11E-01 | 3.87E-02 |
| 2141 | Upp1 | UPP1 | -6.86E-02 | 5.34E-02 |
| 2142 | Pag1 | PAG1 | -7.70E-02 | 2.03E-02 |
| 2143 | Phgdh | PHGDH | 1.03E-01 | -1.78E-02 |
| 2144 | Cd99l2 | CD99L2 | -2.93E-02 | -1.12E-02 |
| 2145 | Col16a1 | COL16A1 | -9.49E-02 | -1.56E-02 |
| 2146 | Napb | NAPB | 6.03E-02 | -3.94E-02 |
| 2147 | Ptchd1 | PTCHD1 | -1.12E-01 | 2.72E-02 |
| 2148 | Actr10 | ACTR10 | 2.96E-02 | -1.93E-02 |
| 2149 | Zfp251 | ZNF251 | -5.53E-02 | 7.58E-05 |
| 2150 | Entpd8 | ENTPD8 | -8.00E-02 | -3.68E-02 |
| 2151 | Gramd4 | GRAMD4 | -4.25E-02 | -2.76E-02 |
| 2152 | Eif1ad | EIF1AD | 3.82E-02 | -6.79E-04 |
| 2153 | Cubn | CUBN | -6.81E-02 | -2.35E-02 |
| 2154 | Plcl2 | PLCL2 | 1.40E-01 | -4.76E-03 |
| 2155 | St3gal6 | ST3GAL6 | -6.03E-02 | -4.23E-03 |
| 2156 | Ubr5 | UBR5 | 2.93E-02 | -1.84E-02 |
| 2157 | Itsn1 | ITSN1 | -3.88E-02 | 1.66E-02 |
| 2158 | Cenpw | CENPW | 1.28E-01 | 1.21E-01 |
| 2159 | Egf | EGF | -5.84E-02 | -5.33E-02 |
| 2160 | Rasl10a | RASL10A | -1.23E-01 | 6.29E-02 |
| 2161 | Raver2 | RAVER2 | -1.01E-01 | -5.36E-02 |
| 2162 | Slc14a1 | SLC14A1 | 3.69E-01 | -2.14E-01 |
| 2163 | Gpx3 | GPX3 | 6.10E-02 | -7.77E-03 |
| 2164 | Ppp3r1 | PPP3R1 | 3.12E-02 | 1.72E-03 |
| 2165 | Dnah7b | DNAH7 | -9.46E-02 | -4.43E-02 |
| 2166 | Letm1 | LETM1 | 3.67E-02 | -2.14E-02 |
| 2167 | Atp1a1 | ATP1A1 | 3.56E-02 | -9.92E-03 |
| 2168 | Rfc3 | RFC3 | 7.77E-02 | 1.56E-03 |
| 2169 | Thsd4 | THSD4 | 7.26E-02 | -4.06E-02 |
| 2170 | Rbms1 | RBMS1 | -4.22E-02 | -1.30E-02 |

| | | | | |
|---|---|---|---|---|
| 2171 | Khsrp | KHSRP | -2.13E-02 | 1.51E-03 |
| 2172 | Ears2 | EARS2 | -7.61E-02 | -1.03E-02 |
| 2173 | Med28 | MED28 | -4.68E-02 | -1.11E-02 |
| 2174 | Tor1a | TOR1A | -3.11E-02 | -8.45E-03 |
| 2175 | Pdk4 | PDK4 | 2.45E-01 | -1.32E-02 |
| 2176 | Car5b | CA5B | 5.50E-02 | -8.07E-03 |
| 2177 | Thrb | THRB | -6.39E-02 | -4.51E-02 |
| 2178 | Dym | DYM | -4.39E-02 | -2.80E-03 |
| 2179 | Hacl1 | HACL1 | 1.05E-01 | 5.59E-03 |
| 2180 | Ppcdc | PPCDC | -4.66E-02 | -7.07E-03 |
| 2181 | Jmjd4 | JMJD4 | -5.88E-02 | 5.22E-03 |
| 2182 | Stam2 | STAM2 | 2.70E-02 | 3.26E-03 |
| 2183 | Dazap2 | DAZAP2 | 2.68E-02 | -4.34E-03 |
| 2184 | Eng | ENG | 5.71E-02 | -2.97E-02 |
| 2185 | Tead3 | TEAD3 | 3.81E-02 | -1.24E-02 |
| 2186 | Ptges2 | PTGES2 | 3.98E-02 | -1.10E-02 |
| 2187 | Bin3 | BIN3 | 3.53E-02 | -2.17E-03 |
| 2188 | Nme1 | NME1 | 5.48E-02 | 9.51E-03 |
| 2189 | Cmtm6 | CMTM6 | -2.82E-02 | -1.07E-02 |
| 2190 | Blk | BLK | -8.88E-02 | 1.78E-02 |
| 2191 | Cd8b1 | CD8B | -1.11E-01 | 6.33E-02 |
| 2192 | Rpusd4 | RPUSD4 | -4.32E-02 | -3.43E-03 |
| 2193 | Tmtc1 | TMTC1 | 8.57E-02 | -4.13E-02 |
| 2194 | Zscan12 | ZSCAN12 | -4.18E-02 | 1.88E-04 |
| 2195 | Map10 | MAP10 | -7.52E-02 | -1.08E-02 |
| 2196 | Smim15 | SMIM15 | -3.19E-02 | 2.90E-03 |
| 2197 | Mcm9 | MCM9 | 5.01E-02 | -2.75E-03 |
| 2198 | St6galnac4 | ST6GALNAC4 | 9.03E-02 | -1.23E-02 |
| 2199 | Nfatc2 | NFATC2 | -8.46E-02 | -1.71E-02 |
| 2200 | Rsph4a | RSPH4A | 1.21E-01 | 5.84E-02 |
| 2201 | Nt5c3 | NT5C3A | -3.59E-02 | -7.53E-03 |
| 2202 | Marf1 | KIAA0430 | -3.93E-02 | -1.79E-02 |
| 2203 | Pcdhga11 | PCDHGA11 | -1.08E-01 | 1.51E-02 |
| 2204 | Srrd | SRRD | 4.45E-02 | -3.05E-02 |
| 2205 | Ppwd1 | PPWD1 | -3.90E-02 | 4.43E-03 |
| 2206 | Tox | TOX | -1.07E-01 | 3.90E-02 |
| 2207 | Adat1 | ADAT1 | -6.39E-02 | 1.25E-02 |
| 2208 | 4932438H23Rik | C21orf62 | 1.15E-01 | 2.20E-03 |
| 2209 | Actr5 | ACTR5 | -4.57E-02 | -6.23E-03 |
| 2210 | Arsg | ARSG | -7.69E-02 | -7.79E-03 |
| 2211 | Rasal3 | RASAL3 | -1.58E-01 | 7.99E-02 |
| 2212 | Rtkn | RTKN | -6.68E-02 | 1.67E-02 |
| 2213 | Nop2 | NOP2 | -4.21E-02 | 2.16E-02 |
| 2214 | Arsb | ARSB | 5.90E-02 | -6.20E-03 |
| 2215 | Phip | PHIP | -5.00E-02 | 1.36E-03 |
| 2216 | Cpox | CPOX | 7.51E-02 | 8.86E-03 |
| 2217 | Gys2 | GYS2 | -1.36E-01 | -4.70E-02 |
| 2218 | Zfp777 | ZNF777 | 4.23E-02 | -1.94E-02 |
| 2219 | Hspb7 | HSPB7 | 7.76E-02 | 9.14E-02 |
| 2220 | Snrpb2 | SNRPB2 | -4.14E-02 | 2.14E-02 |
| 2221 | Afm | AFM | -7.01E-02 | -3.27E-02 |
| 2222 | Pip5k1a | PIP5K1A | 3.84E-02 | -3.27E-03 |
| 2223 | Dlg3 | DLG3 | -3.35E-02 | -1.58E-02 |

| | | | | |
|---|---|---|---|---|
| 2224 | Nckap1 | NCKAP1 | 2.79E-02 | -9.32E-03 |
| 2225 | Pik3c2a | PIK3C2A | 4.15E-02 | -1.79E-02 |
| 2226 | Ank3 | ANK3 | 3.87E-02 | -3.64E-02 |
| 2227 | Phf7 | PHF7 | 4.82E-02 | -2.48E-02 |
| 2228 | Mtm1 | MTM1 | 4.34E-02 | -1.80E-02 |
| 2229 | Atp5o | ATP5O | 3.62E-02 | -1.20E-02 |
| 2230 | Kpna2 | KPNA2 | 8.40E-02 | 4.01E-02 |
| 2231 | Eif1b | EIF1B | -5.27E-02 | 1.19E-02 |
| 2232 | Prpsap2 | PRPSAP2 | -3.96E-02 | 1.05E-03 |
| 2233 | Ephb1 | EPHB1 | -1.42E-01 | -3.27E-02 |
| 2234 | Ppia | PPIA | 3.11E-02 | -1.02E-02 |
| 2235 | Prss36 | PRSS36 | -4.62E-02 | -9.55E-03 |
| 2236 | Ncbp2 | NCBP2 | -3.65E-02 | 2.39E-02 |
| 2237 | Cadps2 | CADPS2 | 6.37E-02 | 1.53E-03 |
| 2238 | Tbx10 | TBX10 | 1.58E-01 | -1.88E-02 |
| 2239 | Rab5a | RAB5A | 3.29E-02 | -8.69E-03 |
| 2240 | Arl15 | ARL15 | 7.81E-02 | -9.36E-03 |
| 2241 | Socs1 | SOCS1 | -1.15E-01 | 3.90E-02 |
| 2242 | Prr12 | PRR12 | -4.83E-02 | -6.96E-03 |
| 2243 | Map3k3 | MAP3K3 | 3.87E-02 | -7.14E-03 |
| 2244 | Zfp157 | ZNF613 | -4.91E-02 | -7.43E-03 |
| 2245 | Sell | SELL | -1.67E-01 | 8.88E-02 |
| 2246 | Scnn1g | SCNN1G | 5.46E-02 | 5.96E-03 |
| 2247 | Bcl7c | BCL7C | 5.40E-02 | 2.86E-02 |
| 2248 | Sparcl1 | SPARCL1 | 7.36E-02 | 1.88E-02 |
| 2249 | Gna13 | GNA13 | 3.10E-02 | -1.82E-03 |
| 2250 | Acyp2 | ACYP2 | -4.32E-02 | -8.79E-03 |
| 2251 | Golph3 | GOLPH3 | 2.96E-02 | -2.80E-03 |
| 2252 | Fez2 | FEZ2 | 3.16E-02 | -1.07E-02 |
| 2253 | Zbed3 | ZBED3 | -4.17E-02 | 3.19E-02 |
| 2254 | Mef2c | MEF2C | -1.04E-01 | 1.97E-02 |
| 2255 | Spred1 | SPRED1 | -4.58E-02 | 1.69E-02 |
| 2256 | Ptprn2 | PTPRN2 | 4.04E-01 | -1.58E-01 |
| 2257 | Igf2bp2 | IGF2BP2 | 9.53E-02 | 7.49E-02 |
| 2258 | Sumo3 | SUMO3 | 3.60E-02 | -1.72E-03 |
| 2259 | Osbpl11 | OSBPL11 | 3.52E-02 | -1.58E-02 |
| 2260 | Dnpep | DNPEP | 3.08E-02 | -4.52E-03 |
| 2261 | Grin2c | GRIN2C | -1.32E-01 | -2.13E-02 |
| 2262 | Gbp4 | GBP6 | -1.95E-01 | -9.79E-03 |
| 2263 | Dll1 | DLL1 | -6.99E-02 | 3.98E-02 |
| 2264 | Slc2a2 | SLC2A2 | 9.13E-02 | -3.59E-02 |
| 2265 | S1pr4 | S1PR4 | -1.09E-01 | -2.89E-02 |
| 2266 | Slc28a1 | SLC28A1 | 9.04E-02 | -3.52E-02 |
| 2267 | Stard9 | STARD9 | -9.86E-02 | 1.65E-02 |
| 2268 | Lhfpl4 | LHFPL4 | -7.14E-02 | 4.63E-02 |
| 2269 | Tnfrsf14 | TNFRSF14 | -1.64E-01 | 2.86E-02 |
| 2270 | Gcc1 | GCC1 | -4.44E-02 | 3.84E-03 |
| 2271 | Zfp383 | ZNF383 | -7.15E-02 | -4.18E-02 |
| 2272 | Nab2 | NAB2 | -6.88E-02 | 2.23E-02 |
| 2273 | Acsf2 | ACSF2 | 1.16E-01 | 3.57E-03 |
| 2274 | Zfp111 | ZNF226 | -8.12E-02 | 5.10E-04 |
| 2275 | Adcy2 | ADCY2 | 1.12E-01 | -5.69E-02 |
| 2276 | Smpd2 | SMPD2 | 4.48E-02 | -2.37E-02 |

| | | | | |
|---|---|---|---|---|
| 2277 | Kif26a | KIF26A | 1.37E-01 | 9.04E-03 |
| 2278 | Cops5 | COPS5 | 2.72E-02 | -1.01E-02 |
| 2279 | Rpa1 | RPA1 | 2.76E-02 | -1.59E-02 |
| 2280 | Slc52a3 | SLC52A3 | 9.79E-02 | 2.27E-02 |
| 2281 | S100a10 | S100A10 | 5.66E-02 | 3.58E-02 |
| 2282 | Lmo4 | LMO4 | -4.66E-02 | -2.37E-02 |
| 2283 | Drap1 | DRAP1 | 3.12E-02 | -1.49E-02 |
| 2284 | Zwint | ZWINT | 3.16E-02 | -1.74E-04 |
| 2285 | Hsd17b2 | HSD17B2 | -1.39E-01 | -5.60E-02 |
| 2286 | Cdkl3 | CDKL3 | 9.53E-02 | -1.56E-02 |
| 2287 | Slc6a7 | SLC6A7 | -1.67E-01 | -3.82E-02 |
| 2288 | Snx6 | SNX6 | -3.39E-02 | -3.70E-04 |
| 2289 | Dus1l | DUS1L | 3.35E-02 | 5.61E-03 |
| 2290 | Dkk3 | DKK3 | -5.36E-02 | 4.18E-02 |
| 2291 | Sdk2 | SDK2 | 1.17E-01 | -1.20E-02 |
| 2292 | Tmem39a | TMEM39A | -4.81E-02 | 1.05E-02 |
| 2293 | Pyroxd2 | PYROXD2 | 7.11E-02 | -2.00E-02 |
| 2294 | Atg2b | ATG2B | 3.02E-02 | -6.94E-03 |
| 2295 | Rhoa | RHOA | 2.41E-02 | -3.23E-03 |
| 2296 | Zfp449 | ZNF449 | -9.30E-02 | 1.79E-02 |
| 2297 | Hpcal1 | HPCAL1 | 4.47E-02 | -2.09E-03 |
| 2298 | Shank3 | SHANK3 | -5.03E-02 | -2.04E-02 |
| 2299 | Tmem132e | TMEM132E | -1.09E-01 | 2.18E-03 |
| 2300 | Ctbp2 | CTBP2 | -2.77E-02 | -5.11E-03 |
| 2301 | Erf | ERF | 4.61E-02 | -1.15E-02 |
| 2302 | Lipt1 | LIPT1 | -7.24E-02 | 9.18E-03 |
| 2303 | Ndufa3 | NDUFA3 | 3.34E-02 | -1.47E-02 |
| 2304 | Il18 | IL18 | 1.14E-01 | 7.81E-03 |
| 2305 | Samd4 | SAMD4A | 8.57E-02 | 4.40E-02 |
| 2306 | Syt15 | SYT15 | -9.88E-02 | -1.06E-01 |
| 2307 | Ubxn8 | UBXN8 | 4.04E-02 | -6.17E-03 |
| 2308 | Itga5 | ITGA5 | 7.07E-02 | 5.29E-02 |
| 2309 | Bora | BORA | 1.38E-01 | 3.13E-03 |
| 2310 | Nr3c1 | NR3C1 | -5.93E-02 | -1.54E-02 |
| 2311 | Cenpt | CENPT | 8.08E-02 | 4.67E-02 |
| 2312 | Txndc16 | TXNDC16 | -4.91E-02 | -2.84E-02 |
| 2313 | Arl14ep | ARL14EP | -4.57E-02 | -1.30E-02 |
| 2314 | Fibin | FIBIN | 1.63E-01 | -2.17E-02 |
| 2315 | Jag1 | JAG1 | -8.50E-02 | 3.42E-02 |
| 2316 | Cd3g | CD3G | -1.69E-01 | 4.99E-02 |
| 2317 | Ccrl2 | CCRL2 | -1.19E-01 | 3.61E-02 |
| 2318 | Rab7 | RAB7A | 3.66E-02 | -1.30E-02 |
| 2319 | Tubb2a | TUBB2A | 9.25E-02 | 8.28E-02 |
| 2320 | Frmd8 | FRMD8 | 2.90E-02 | 2.14E-02 |
| 2321 | Papolg | PAPOLG | -7.04E-02 | -3.97E-03 |
| 2322 | Ptgs1 | PTGS1 | 8.49E-02 | -3.13E-02 |
| 2323 | Cntn6 | CNTN6 | 1.14E-01 | 4.52E-04 |
| 2324 | Sorbs2 | SORBS2 | 5.20E-02 | -2.05E-04 |
| 2325 | Slamf6 | SLAMF6 | -6.20E-02 | 4.77E-02 |
| 2326 | Mtf2 | MTF2 | 4.45E-02 | 2.00E-03 |
| 2327 | Anapc10 | ANAPC10 | -6.41E-02 | 2.77E-02 |
| 2328 | Adcy10 | ADCY10 | -1.01E-01 | -9.13E-02 |
| 2329 | Mrap2 | MRAP2 | 1.14E-01 | -3.44E-04 |

| | | | | |
|---|---|---|---|---|
| 2330 | Tnfrsf13c | TNFRSF13C | -4.72E-02 | -3.44E-03 |
| 2331 | Pdzrn4 | PDZRN4 | -1.85E-01 | -5.09E-02 |
| 2332 | Psmd11 | PSMD11 | 2.70E-02 | 7.24E-04 |
| 2333 | Aqp3 | AQP3 | 6.87E-02 | -5.23E-02 |
| 2334 | Mocos | MOCOS | -7.95E-02 | -3.79E-02 |
| 2335 | Ogg1 | OGG1 | 5.93E-02 | -3.71E-02 |
| 2336 | Agap2 | AGAP2 | 1.21E-01 | 6.62E-02 |
| 2337 | Bri3bp | BRI3BP | -3.31E-02 | -8.44E-03 |
| 2338 | Dcaf10 | DCAF10 | 4.55E-02 | -1.06E-02 |
| 2339 | Coa5 | COA5 | 2.91E-02 | -2.26E-02 |
| 2340 | Fgd3 | FGD3 | -1.41E-01 | 4.04E-02 |
| 2341 | Polr1d | POLR1D | -2.65E-02 | -1.11E-02 |
| 2342 | Ranbp9 | RANBP9 | 3.18E-02 | -3.34E-03 |
| 2343 | Six4 | SIX4 | -8.82E-02 | 3.78E-03 |
| 2344 | G3bp2 | G3BP2 | 2.46E-02 | -2.58E-03 |
| 2345 | Eftud2 | EFTUD2 | 2.53E-02 | 1.42E-03 |
| 2346 | Cox8a | COX8A | 3.48E-02 | -1.37E-02 |
| 2347 | Tmod4 | TMOD4 | -1.52E-01 | 2.74E-02 |
| 2348 | Rab11a | RAB11A | 2.93E-02 | 8.33E-03 |
| 2349 | Gpcpd1 | GPCPD1 | -3.77E-02 | 7.08E-03 |
| 2350 | Mios | MIOS | 4.30E-02 | -1.61E-02 |
| 2351 | Ppp6r2 | PPP6R2 | 2.66E-02 | -1.41E-02 |
| 2352 | Icosl | ICOSLG | -1.12E-01 | 3.57E-04 |
| 2353 | Icosl | LOC102723996 | -1.12E-01 | 3.57E-04 |
| 2354 | Timm8b | TIMM8B | 3.73E-02 | -4.46E-03 |
| 2355 | Edf1 | EDF1 | 3.42E-02 | -1.52E-02 |
| 2356 | Zfp524 | ZNF524 | -9.58E-02 | 2.81E-02 |
| 2357 | G6pc3 | G6PC3 | 2.66E-02 | -5.48E-03 |
| 2358 | Khdrbs3 | KHDRBS3 | -5.28E-02 | -3.04E-02 |
| 2359 | Coro1b | CORO1B | -3.55E-02 | 1.69E-02 |
| 2360 | Flnb | FLNB | 3.35E-02 | 9.27E-03 |
| 2361 | Memo1 | MEMO1 | 4.76E-02 | 1.88E-03 |
| 2362 | Ppic | PPIC | -1.23E-01 | -1.01E-01 |
| 2363 | Gpr182 | GPR182 | 9.73E-02 | -2.64E-02 |
| 2364 | Pex5 | PEX5 | 4.22E-02 | -1.44E-02 |
| 2365 | Stub1 | STUB1 | 2.87E-02 | -1.06E-02 |
| 2366 | Ccdc15 | CCDC15 | -1.02E-01 | -9.57E-03 |
| 2367 | Tm9sf2 | TM9SF2 | -3.13E-02 | -7.21E-03 |
| 2368 | Specc1 | SPECC1 | 5.80E-02 | 1.09E-03 |
| 2369 | C1qb | C1QB | -9.42E-02 | 1.45E-01 |
| 2370 | Glud1 | GLUD1 | 5.25E-02 | -1.64E-03 |
| 2371 | Glmn | GLMN | 6.27E-02 | -1.31E-03 |
| 2372 | Pou6f1 | POU6F1 | 3.45E-02 | -3.24E-02 |
| 2373 | Ube2k | UBE2K | 2.62E-02 | -8.86E-03 |
| 2374 | Kcnj8 | KCNJ8 | -1.27E-01 | 2.15E-02 |
| 2375 | Zhx1 | ZHX1 | -3.42E-02 | -8.28E-03 |
| 2376 | Alx4 | ALX4 | -6.73E-02 | 9.18E-03 |
| 2377 | Fgfbp1 | FGFBP1 | -1.12E-01 | -3.79E-02 |
| 2378 | Pigp | PIGP | -3.10E-02 | 2.90E-03 |
| 2379 | Clec1a | CLEC1A | -8.87E-02 | 1.21E-02 |
| 2380 | Znhit2 | ZNHIT2 | 4.64E-02 | -1.20E-03 |
| 2381 | Eml1 | EML1 | 5.43E-02 | -1.15E-02 |
| 2382 | Mei4 | MEI4 | -8.86E-02 | 1.96E-02 |

| | | | | |
|---|---|---|---|---|
| 2383 | Syne2 | SYNE2 | 4.05E-02 | -1.40E-02 |
| 2384 | Nkg7 | NKG7 | -1.15E-01 | 3.23E-02 |
| 2385 | Tada2b | TADA2B | 3.88E-02 | -9.29E-03 |
| 2386 | Fbxl14 | FBXL14 | -3.51E-02 | 8.52E-03 |
| 2387 | Sh2b1 | SH2B1 | -2.44E-02 | -8.32E-03 |
| 2388 | Adck5 | ADCK5 | 5.15E-02 | -1.90E-02 |
| 2389 | Col4a4 | COL4A4 | 3.13E-02 | -2.67E-02 |
| 2390 | C2cd5 | C2CD5 | -3.47E-02 | -2.70E-02 |
| 2391 | Ushbp1 | USHBP1 | -5.75E-02 | -1.30E-02 |
| 2392 | Arfrp1 | ARFRP1 | -4.16E-02 | -1.08E-03 |
| 2393 | Opcml | OPCML | 1.49E-01 | -1.76E-03 |
| 2394 | Psmb2 | PSMB2 | 3.02E-02 | -7.33E-03 |
| 2395 | Ogfr | OGFR | -3.33E-02 | 1.90E-02 |
| 2396 | Slirp | SLIRP | 4.43E-02 | -9.27E-03 |
| 2397 | Mmadhc | MMADHC | 3.24E-02 | 1.25E-03 |
| 2398 | Bfsp2 | BFSP2 | 1.03E-01 | -1.05E-02 |
| 2399 | Srp9 | SRP9 | 2.97E-02 | -7.39E-03 |
| 2400 | Btd | BTD | -4.04E-02 | 4.71E-03 |
| 2401 | Slain2 | SLAIN2 | 4.07E-02 | -1.12E-02 |
| 2402 | Rsph1 | RSPH1 | 1.12E-01 | 3.19E-02 |
| 2403 | Birc2 | BIRC2 | -3.42E-02 | 3.68E-02 |
| 2404 | Sema4b | SEMA4B | 7.64E-02 | 2.11E-03 |
| 2405 | Pla2g7 | PLA2G7 | 1.14E-01 | 8.40E-03 |
| 2406 | Gsk3a | GSK3A | 2.68E-02 | -7.59E-03 |
| 2407 | Tap2 | TAP2 | -9.54E-02 | 3.80E-02 |
| 2408 | Zmym3 | ZMYM3 | 4.30E-02 | -3.63E-02 |
| 2409 | Siglece | SIGLEC9 | -1.49E-01 | 8.11E-02 |
| 2410 | Akt3 | AKT3 | -7.45E-02 | 3.04E-02 |
| 2411 | Uqcr10 | UQCR10 | 3.73E-02 | -1.53E-02 |
| 2412 | Narf | NARF | 4.24E-02 | -4.31E-02 |
| 2413 | Eva1b | EVA1B | 5.87E-02 | 3.55E-02 |
| 2414 | Pdgfb | PDGFB | -6.72E-02 | 6.63E-02 |
| 2415 | Zfp738 | ZNF729 | -6.41E-02 | 1.46E-02 |
| 2416 | Mvb12b | MVB12B | 4.52E-02 | -4.00E-02 |
| 2417 | Copz1 | COPZ1 | 2.87E-02 | -1.27E-02 |
| 2418 | Arrdc2 | ARRDC2 | 1.46E-01 | -2.36E-02 |
| 2419 | Otoa | OTOA | -1.66E-01 | -1.57E-02 |
| 2420 | Irs1 | IRS1 | -6.18E-02 | 2.41E-03 |
| 2421 | Eci2 | ECI2 | 4.60E-02 | -5.87E-03 |
| 2422 | Pxn | PXN | 4.13E-02 | -5.89E-03 |
| 2423 | Bap1 | BAP1 | 3.51E-02 | -1.89E-02 |
| 2424 | Myo19 | MYO19 | 3.72E-02 | 9.82E-04 |
| 2425 | Morf4l2 | MORF4L2 | 4.03E-02 | 1.80E-02 |
| 2426 | Cd27 | CD27 | -7.12E-02 | 1.92E-02 |
| 2427 | Cxcl13 | CXCL13 | 2.23E-01 | 1.31E-01 |
| 2428 | Ttyh2 | TTYH2 | 8.17E-02 | 1.35E-03 |
| 2429 | Slc22a14 | SLC22A14 | -1.64E-01 | -7.37E-02 |
| 2430 | Hoxc8 | HOXC8 | -6.54E-02 | 1.59E-02 |
| 2431 | Tbc1d9b | TBC1D9B | 2.89E-02 | -2.55E-02 |
| 2432 | Rab38 | RAB38 | -9.38E-02 | -6.62E-02 |
| 2433 | Ufl1 | UFL1 | 2.60E-02 | -1.78E-02 |
| 2434 | Pxmp4 | PXMP4 | 6.13E-02 | 2.34E-02 |
| 2435 | Ccng2 | CCNG2 | -4.49E-02 | -4.03E-02 |

| | | | | |
|---|---|---|---|---|
| 2436 | Comtd1 | COMTD1 | 8.32E-02 | 6.36E-03 |
| 2437 | Fcamr | FCAMR | -2.16E-01 | -2.20E-02 |
| 2438 | Lbp | LBP | 1.03E-01 | 6.64E-02 |
| 2439 | Plxdc1 | PLXDC1 | -1.16E-01 | 4.65E-02 |
| 2440 | Car2 | CA2 | 4.30E-02 | -1.49E-02 |
| 2441 | Atp5g3 | ATP5G3 | 3.59E-02 | -9.97E-03 |
| 2442 | Pla2g12a | PLA2G12A | 4.31E-02 | -7.59E-03 |
| 2443 | Lrmp | LRMP | -1.25E-01 | 1.12E-01 |
| 2444 | Ppox | PPOX | -3.06E-02 | -1.49E-02 |
| 2445 | Notch3 | NOTCH3 | 5.29E-02 | 3.40E-02 |
| 2446 | Exd2 | EXD2 | -4.72E-02 | -1.43E-02 |
| 2447 | Shisa5 | SHISA5 | -3.05E-02 | -5.49E-04 |
| 2448 | Pik3ap1 | PIK3AP1 | -1.95E-01 | 1.29E-01 |
| 2449 | Gm5566 | NME1-NME2 | 1.43E-01 | -5.22E-03 |
| 2450 | Smarca2 | SMARCA2 | -5.03E-02 | -2.00E-02 |
| 2451 | Inca1 | INCA1 | 6.91E-02 | -5.54E-02 |
| 2452 | Slc38a1 | SLC38A1 | -1.92E-01 | 5.03E-02 |
| 2453 | Msrb1 | MSRB1 | 5.27E-02 | -1.33E-02 |
| 2454 | Mical2 | MICAL2 | 8.75E-02 | 2.20E-02 |
| 2455 | Pou2f2 | POU2F2 | -1.79E-01 | 7.02E-02 |
| 2456 | Pdcl3 | PDCL3 | 2.54E-02 | 5.25E-03 |
| 2457 | Kif1c | KIF1C | 2.63E-02 | -2.20E-02 |
| 2458 | Zfp418 | ZNF416 | -1.05E-01 | 4.71E-02 |
| 2459 | Rragb | RRAGB | -8.04E-02 | -5.02E-02 |
| 2460 | L2hgdh | L2HGDH | -5.51E-02 | -3.68E-02 |
| 2461 | Nek11 | NEK11 | 8.83E-02 | -1.29E-02 |
| 2462 | C1qc | C1QC | -1.03E-01 | 1.33E-01 |
| 2463 | Sntg1 | SNTG1 | -1.38E-01 | 2.23E-02 |
| 2464 | Zfx | ZFX | -3.51E-02 | 3.71E-03 |
| 2465 | Lats2 | LATS2 | -6.50E-02 | 2.38E-02 |
| 2466 | Fzd6 | FZD6 | 1.10E-01 | -4.29E-02 |
| 2467 | D11Wsu47e | C17orf80 | -5.19E-02 | 3.57E-02 |
| 2468 | Ano8 | ANO8 | -4.19E-02 | -2.99E-03 |
| 2469 | Tnfsf14 | TNFSF14 | -8.92E-02 | 4.85E-02 |
| 2470 | Mid1 | MID1 | -1.33E-01 | 3.16E-02 |
| 2471 | H2-Q4 | HLA-A | -1.08E-01 | 3.81E-02 |
| 2472 | Ism1 | ISM1 | -1.48E-01 | 4.68E-02 |
| 2473 | Bahd1 | BAHD1 | 2.77E-02 | -6.49E-03 |
| 2474 | Kti12 | KTI12 | -3.31E-02 | 3.22E-02 |
| 2475 | Ankhd1 | ANKHD1 | -2.75E-02 | -3.91E-03 |
| 2476 | Ankhd1 | ANKHD1-EIF4EBP3 | -2.75E-02 | -3.91E-03 |
| 2477 | Mrpl14 | MRPL14 | 4.10E-02 | -9.91E-03 |
| 2478 | Api5 | API5 | 2.56E-02 | 5.34E-03 |
| 2479 | Pofut1 | POFUT1 | -3.31E-02 | -3.94E-03 |
| 2480 | 3830406C13Rik | C3orf14 | -5.54E-02 | 9.79E-03 |
| 2481 | A4gnt | A4GNT | 1.14E-01 | -9.57E-02 |
| 2482 | Mphosph6 | MPHOSPH6 | 6.76E-02 | -2.74E-03 |
| 2483 | Nlrc3 | NLRC3 | -1.14E-01 | -6.30E-03 |
| 2484 | Dmd | DMD | 5.75E-02 | -8.86E-03 |
| 2485 | Usb1 | USB1 | 5.09E-02 | -7.92E-05 |
| 2486 | Cdk1 | CDK1 | 2.90E-01 | 1.99E-01 |
| 2487 | Dcaf15 | DCAF15 | 5.57E-02 | 7.77E-03 |
| 2488 | Ran | RAN | 2.96E-02 | 7.27E-03 |

| | | | | |
|---|---|---|---|---|
| 2489 | Mif | MIF | 6.06E-02 | -2.10E-02 |
| 2490 | C2 | C2 | -1.01E-01 | 1.48E-02 |
| 2491 | Paqr9 | PAQR9 | -5.51E-02 | -1.07E-02 |
| 2492 | P2rx2 | P2RX2 | -9.31E-02 | 1.00E-03 |
| 2493 | Heph | HEPH | 4.20E-02 | 8.98E-03 |
| 2494 | Mlx | MLX | -3.33E-02 | -6.76E-03 |
| 2495 | Col27a1 | COL27A1 | 1.67E-01 | -4.61E-03 |
| 2496 | Unc93b1 | UNC93B1 | -6.19E-02 | 5.00E-02 |
| 2497 | Cdhr4 | CDHR4 | 8.25E-02 | -2.99E-02 |
| 2498 | Cox6b1 | COX6B1 | 3.01E-02 | -1.18E-02 |
| 2499 | Ndufa12 | NDUFA12 | 3.14E-02 | -1.79E-02 |
| 2500 | Qtrt1 | QTRT1 | -5.39E-02 | 2.65E-04 |
| 2501 | Itgb1 | ITGB1 | 2.37E-02 | 6.77E-03 |
| 2502 | Xpnpep1 | XPNPEP1 | -5.51E-02 | 7.26E-03 |
| 2503 | Scimp | SCIMP | -1.20E-01 | 4.55E-02 |
| 2504 | Sfrp2 | SFRP2 | -1.71E-01 | 1.12E-01 |
| 2505 | Prkg2 | PRKG2 | 1.31E-01 | -3.54E-04 |
| 2506 | Scyl2 | SCYL2 | 2.84E-02 | 1.81E-03 |
| 2507 | Atp6v1e1 | ATP6V1E1 | 3.49E-02 | -3.43E-03 |
| 2508 | Il27ra | IL27RA | -1.09E-01 | 5.56E-02 |
| 2509 | Ugt3a2 | UGT3A1 | -5.24E-02 | -1.92E-02 |
| 2510 | Ccdc9 | CCDC9 | 3.74E-02 | -1.46E-02 |
| 2511 | Fndc5 | FNDC5 | -1.02E-01 | -5.40E-02 |
| 2512 | Rab33a | RAB33A | 6.50E-02 | -3.44E-02 |
| 2513 | Gse1 | GSE1 | 4.48E-02 | 7.88E-03 |
| 2514 | Rapgef4 | RAPGEF4 | -7.47E-02 | -5.17E-02 |
| 2515 | Gsto2 | GSTO2 | -1.34E-01 | -5.42E-02 |
| 2516 | Nipbl | NIPBL | -3.05E-02 | -1.04E-02 |
| 2517 | Sh3tc2 | SH3TC2 | 6.27E-02 | 2.12E-02 |
| 2518 | Dusp11 | DUSP11 | -4.38E-02 | 1.16E-02 |
| 2519 | Zfp839 | ZNF839 | 3.38E-02 | -2.26E-02 |
| 2520 | Nfatc2ip | NFATC2IP | 8.46E-02 | 4.59E-04 |
| 2521 | Mfsd2a | MFSD2A | -2.12E-01 | -2.11E-01 |
| 2522 | Ric8b | RIC8B | -4.30E-02 | 9.88E-03 |
| 2523 | Dnajc28 | DNAJC28 | -6.00E-02 | -3.33E-02 |
| 2524 | Ezh1 | EZH1 | -2.59E-02 | -2.78E-02 |
| 2525 | Slc25a10 | SLC25A10 | 6.49E-02 | -3.04E-03 |
| 2526 | Sars2 | SARS2 | 4.38E-02 | -3.58E-02 |
| 2527 | Zfp322a | ZNF322 | -7.03E-02 | 2.20E-02 |
| 2528 | Slc39a3 | SLC39A3 | -2.91E-02 | -8.80E-03 |
| 2529 | Ttll9 | TTLL9 | -1.29E-01 | -1.18E-02 |
| 2530 | Hesx1 | HESX1 | 1.69E-01 | 8.82E-02 |
| 2531 | BC004004 | C6orf89 | 3.42E-02 | -1.37E-02 |
| 2532 | Lrrc29 | LRRC29 | 9.59E-02 | -7.07E-02 |
| 2533 | Psme1 | PSME1 | -3.53E-02 | 2.85E-02 |
| 2534 | Fgf18 | FGF18 | 1.69E-01 | 1.55E-01 |
| 2535 | Nod1 | NOD1 | -5.10E-02 | -2.09E-02 |
| 2536 | Zfp563 | ZNF563 | -8.76E-02 | -1.87E-02 |
| 2537 | Mrpl17 | MRPL17 | 3.04E-02 | -1.72E-02 |
| 2538 | Tmem54 | TMEM54 | 1.45E-01 | -4.69E-02 |
| 2539 | Ptgis | PTGIS | 1.13E-01 | 1.75E-02 |
| 2540 | Inhbb | INHBB | -1.74E-01 | 1.62E-01 |
| 2541 | C3 | C3 | -8.87E-02 | 1.81E-01 |

| | | | | |
|---|---|---|---|---|
| 2542 | Acrbp | ACRBP | -1.15E-01 | 9.43E-03 |
| 2543 | Nup35 | NUP35 | -6.17E-02 | 5.43E-02 |
| 2544 | Higd2a | HIGD2A | 3.78E-02 | -7.34E-03 |
| 2545 | Usp39 | USP39 | 3.70E-02 | 8.00E-04 |
| 2546 | Fam171a1 | FAM171A1 | 3.28E-02 | -2.40E-02 |
| 2547 | Cnot8 | CNOT8 | -3.15E-02 | 1.16E-03 |
| 2548 | Wnt2 | WNT2 | -8.96E-02 | -1.05E-02 |
| 2549 | Maea | MAEA | 2.32E-02 | -9.79E-03 |
| 2550 | Lman1 | LMAN1 | -2.58E-02 | -1.71E-02 |
| 2551 | Slc6a4 | SLC6A4 | -1.21E-01 | -6.75E-02 |
| 2552 | Gmcl1 | GMCL1 | -3.76E-02 | -1.69E-02 |
| 2553 | Thsd1 | THSD1 | 9.60E-02 | -3.83E-02 |
| 2554 | Dzank1 | DZANK1 | -7.00E-02 | -3.45E-02 |
| 2555 | Zfp553 | ZNF48 | -4.96E-02 | 1.80E-02 |
| 2556 | Zfand3 | ZFAND3 | 3.07E-02 | -5.95E-04 |
| 2557 | Cep78 | CEP78 | 6.24E-02 | 3.65E-02 |
| 2558 | Wscd1 | WSCD1 | -1.41E-01 | -3.26E-02 |
| 2559 | Zbtb11 | ZBTB11 | -6.45E-02 | 2.04E-02 |
| 2560 | Xpc | XPC | 2.61E-02 | -3.54E-02 |
| 2561 | Rbm43 | RBM43 | -5.25E-02 | 4.17E-02 |
| 2562 | Htatip2 | HTATIP2 | -5.84E-02 | -2.44E-02 |
| 2563 | Trmt1l | TRMT1L | -5.13E-02 | 1.89E-02 |
| 2564 | Jazf1 | JAZF1 | 1.54E-01 | 2.19E-02 |
| 2565 | Lrrc4 | LRRC4 | -7.75E-02 | 1.85E-02 |
| 2566 | Rnd1 | RND1 | -1.69E-01 | 2.08E-01 |
| 2567 | Agt | AGT | 8.39E-02 | -1.17E-02 |
| 2568 | Iyd | IYD | 7.33E-02 | -2.53E-02 |
| 2569 | Atp11a | ATP11A | -5.88E-02 | -1.99E-02 |
| 2570 | Gpbp1 | GPBP1 | -3.09E-02 | -6.98E-04 |
| 2571 | Btg1 | BTG1 | 6.77E-02 | 1.23E-02 |
| 2572 | Acat2 | ACAT2 | 5.27E-02 | -1.62E-03 |
| 2573 | Tgfa | TGFA | -1.18E-01 | -6.11E-03 |
| 2574 | Bzw2 | BZW2 | 3.12E-02 | 4.44E-02 |
| 2575 | Zfp74 | ZNF569 | -6.41E-02 | 1.71E-04 |
| 2576 | Ttc4 | TTC4 | -2.65E-02 | -7.99E-03 |
| 2577 | Zfp398 | ZNF398 | -5.10E-02 | -1.11E-02 |
| 2578 | Htra1 | HTRA1 | 5.73E-02 | -1.11E-02 |
| 2579 | Tlk2 | TLK2 | 2.54E-02 | 1.26E-02 |
| 2580 | Tacstd2 | TACSTD2 | 1.08E-01 | 6.17E-02 |
| 2581 | Myom1 | MYOM1 | 9.14E-02 | 4.61E-03 |
| 2582 | Ucp2 | UCP2 | 6.94E-02 | 5.34E-02 |
| 2583 | Gdf5 | GDF5 | -9.08E-02 | 2.13E-02 |
| 2584 | Casp6 | CASP6 | -4.49E-02 | -9.04E-03 |
| 2585 | Rnf114 | RNF114 | -2.40E-02 | 8.71E-04 |
| 2586 | Pdcd4 | PDCD4 | 5.21E-02 | -9.78E-03 |
| 2587 | Mtg2 | MTG2 | -4.06E-02 | -2.89E-02 |
| 2588 | Prpf40a | PRPF40A | 2.73E-02 | 2.22E-04 |
| 2589 | Zfp521 | ZNF521 | -1.05E-01 | -9.59E-03 |
| 2590 | Spns3 | SPNS3 | 1.74E-01 | 2.36E-02 |
| 2591 | Pfdn1 | PFDN1 | 3.41E-02 | -2.58E-03 |
| 2592 | C1qtnf6 | C1QTNF6 | 1.62E-01 | 6.70E-02 |
| 2593 | Gpr65 | GPR65 | -1.42E-01 | 7.65E-02 |
| 2594 | Med12l | MED12L | 9.13E-02 | -6.63E-03 |

| | | | | |
|---|---|---|---|---|
| 2595 | Pex11a | PEX11A | 8.50E-02 | 1.80E-02 |
| 2596 | Clic6 | CLIC6 | -1.18E-01 | 8.81E-02 |
| 2597 | Amigo2 | AMIGO2 | 6.26E-02 | 2.32E-02 |
| 2598 | Opa3 | OPA3 | 3.02E-02 | -6.40E-04 |
| 2599 | Phf13 | PHF13 | 6.28E-02 | 1.57E-02 |
| 2600 | Zfp623 | ZNF623 | -4.61E-02 | 1.80E-02 |
| 2601 | Kcna1 | KCNA1 | -5.08E-02 | -1.79E-02 |
| 2602 | Rcl1 | RCL1 | -6.21E-02 | 2.16E-03 |
| 2603 | Gps1 | GPS1 | 2.70E-02 | -1.18E-02 |
| 2604 | Gpr137c | GPR137C | 8.26E-02 | -2.00E-02 |
| 2605 | Trim24 | TRIM24 | 2.95E-02 | -1.30E-02 |
| 2606 | Robo4 | ROBO4 | -5.97E-02 | -6.14E-03 |
| 2607 | Ovgp1 | OVGP1 | 1.12E-01 | -2.23E-02 |
| 2608 | Bmp2k | BMP2K | 4.25E-02 | 3.73E-03 |
| 2609 | Fam214a | FAM214A | 8.10E-02 | -2.87E-02 |
| 2610 | Igfbp3 | IGFBP3 | -1.13E-01 | -5.32E-02 |
| 2611 | Tmem43 | TMEM43 | 3.90E-02 | 5.22E-03 |
| 2612 | Tnfrsf12a | TNFRSF12A | 1.11E-01 | 1.35E-01 |
| 2613 | Sumf1 | SUMF1 | 2.59E-02 | -2.08E-02 |
| 2614 | Focad | FOCAD | 4.63E-02 | -2.27E-02 |
| 2615 | Tcea1 | TCEA1 | 2.71E-02 | -1.48E-02 |
| 2616 | Znhit3 | ZNHIT3 | 4.78E-02 | -3.20E-02 |
| 2617 | Mamdc4 | MAMDC4 | -9.89E-02 | 8.69E-03 |
| 2618 | Clu | CLU | 6.18E-02 | 8.08E-02 |
| 2619 | 6430548M08Rik | KIAA0513 | 4.75E-02 | 2.00E-03 |
| 2620 | Dusp5 | DUSP5 | 2.14E-01 | 7.51E-02 |
| 2621 | Gm5617 | C11orf71 | -6.11E-02 | 4.41E-03 |
| 2622 | Mybl2 | MYBL2 | -1.01E-01 | 3.07E-03 |
| 2623 | Unc5d | UNC5D | -5.63E-02 | 9.09E-03 |
| 2624 | Ncf2 | NCF2 | -1.03E-01 | 1.53E-01 |
| 2625 | Cdh3 | CDH3 | 7.90E-02 | 5.97E-02 |
| 2626 | Ddx3x | DDX3X | 4.61E-02 | -1.25E-02 |
| 2627 | Psme4 | PSME4 | 3.67E-02 | -1.29E-02 |
| 2628 | Cldn12 | CLDN12 | -4.72E-02 | -1.94E-02 |
| 2629 | Rgl2 | RGL2 | -3.46E-02 | 1.61E-02 |
| 2630 | Slc29a1 | SLC29A1 | 4.86E-02 | 2.05E-02 |
| 2631 | Sox10 | SOX10 | -8.46E-02 | 3.61E-02 |
| 2632 | Slc25a4 | SLC25A4 | 4.41E-02 | -1.00E-02 |
| 2633 | Slc9a9 | SLC9A9 | -1.17E-01 | 1.07E-01 |
| 2634 | Palm3 | PALM3 | 7.27E-02 | -3.68E-02 |
| 2635 | Cyp2s1 | CYP2S1 | -6.58E-02 | 2.33E-02 |
| 2636 | Slc25a22 | SLC25A22 | 6.94E-02 | 1.14E-02 |
| 2637 | Pgk1 | PGK1 | 3.61E-02 | -1.56E-02 |
| 2638 | Chordc1 | CHORDC1 | -9.55E-02 | 3.03E-02 |
| 2639 | Msc | MSC | 4.00E-02 | -1.46E-02 |
| 2640 | Zfp787 | ZNF787 | 3.30E-02 | -1.43E-02 |
| 2641 | Fkbp11 | FKBP11 | -1.16E-01 | 3.36E-02 |
| 2642 | Sil1 | SIL1 | 2.96E-02 | -1.45E-02 |
| 2643 | Chchd4 | CHCHD4 | 3.70E-02 | -7.79E-03 |
| 2644 | Lgr5 | LGR5 | -1.43E-01 | -9.00E-03 |
| 2645 | Angptl1 | ANGPTL1 | 1.41E-01 | -4.33E-02 |
| 2646 | Loxl2 | LOXL2 | 1.13E-01 | 7.29E-02 |
| 2647 | Elovl1 | ELOVL1 | 3.35E-02 | -8.87E-03 |

| | | | | |
|---|---|---|---|---|
| 2648 | Ccdc61 | CCDC61 | -5.59E-02 | 4.32E-02 |
| 2649 | Wdr41 | WDR41 | 2.85E-02 | -1.61E-02 |
| 2650 | Gorasp1 | GORASP1 | -3.31E-02 | -1.64E-02 |
| 2651 | Gm12407 | CCL19 | -6.81E-02 | -1.17E-02 |
| 2652 | Tmem139 | TMEM139 | -5.64E-02 | -2.98E-02 |
| 2653 | Ifitm3 | IFITM3 | -6.46E-02 | 4.03E-02 |
| 2654 | Aoc1 | AOC1 | 2.45E-01 | 1.47E-01 |
| 2655 | Il18bp | IL18BP | -1.09E-01 | 7.24E-02 |
| 2656 | Rbm15 | RBM15 | -6.65E-02 | 3.81E-02 |
| 2657 | Slc51a | SLC51A | -8.92E-02 | 3.49E-02 |
| 2658 | Ddx19b | DDX19B | 4.47E-02 | -1.07E-02 |
| 2659 | Timp4 | TIMP4 | 2.51E-01 | 8.59E-03 |
| 2660 | Lekr1 | LEKR1 | -9.28E-02 | 6.75E-02 |
| 2661 | Cd69 | CD69 | -8.89E-02 | 7.45E-02 |
| 2662 | Fads6 | FADS6 | -5.62E-02 | -1.35E-02 |
| 2663 | Cyp24a1 | CYP24A1 | 4.40E-01 | -9.04E-02 |
| 2664 | Zfpm1 | ZFPM1 | 7.32E-02 | 2.31E-02 |
| 2665 | B3galt1 | B3GALT1 | 1.57E-01 | 4.20E-02 |
| 2666 | Dgat1 | DGAT1 | 3.08E-02 | 1.89E-03 |
| 2667 | Psmd12 | PSMD12 | 2.84E-02 | 1.98E-03 |
| 2668 | Polr3a | POLR3A | 3.38E-02 | -1.93E-02 |
| 2669 | Tmem82 | TMEM82 | 5.58E-02 | -2.84E-03 |
| 2670 | Lrfn2 | LRFN2 | -8.93E-02 | 2.12E-03 |
| 2671 | Ttll7 | TTLL7 | 1.40E-01 | 3.19E-02 |
| 2672 | Tspan18 | TSPAN18 | -6.64E-02 | -4.77E-03 |
| 2673 | Enpp5 | ENPP5 | 3.60E-02 | -1.23E-02 |
| 2674 | Acbd5 | ACBD5 | 3.35E-02 | -1.40E-02 |
| 2675 | Mtch2 | MTCH2 | 2.90E-02 | -6.24E-03 |
| 2676 | Wdtc1 | WDTC1 | 3.16E-02 | -1.12E-02 |
| 2677 | Vav2 | VAV2 | -3.73E-02 | -6.63E-03 |
| 2678 | Nkx2-2 | NKX2-2 | 4.99E-02 | -3.78E-02 |
| 2679 | Atp6v1f | ATP6V1F | 3.03E-02 | -3.06E-03 |
| 2680 | Nr1d2 | NR1D2 | -5.37E-02 | -4.07E-02 |
| 2681 | Aldh1a1 | ALDH1A1 | 8.97E-02 | -1.20E-02 |
| 2682 | Plbd2 | PLBD2 | 3.26E-02 | -2.12E-03 |
| 2683 | Aldob | ALDOB | 5.49E-02 | -5.58E-03 |
| 2684 | Cd180 | CD180 | -1.74E-01 | 5.25E-02 |
| 2685 | Stk39 | STK39 | 7.93E-02 | -3.11E-02 |
| 2686 | Tbc1d25 | TBC1D25 | -5.56E-02 | 2.38E-02 |
| 2687 | Vps37b | VPS37B | 4.84E-02 | 3.40E-02 |
| 2688 | Klf12 | KLF12 | -8.74E-02 | -5.66E-02 |
| 2689 | Gpm6b | GPM6B | -7.08E-02 | 1.34E-02 |
| 2690 | Dclre1a | DCLRE1A | -4.65E-02 | -2.77E-02 |
| 2691 | Spc24 | SPC24 | 1.65E-01 | 7.91E-02 |
| 2692 | Tfdp2 | TFDP2 | 4.50E-02 | -3.76E-02 |
| 2693 | Ccdc39 | CCDC39 | -9.91E-02 | -9.10E-03 |
| 2694 | Nppc | NPPC | 1.73E-01 | -3.00E-02 |
| 2695 | Bace1 | BACE1 | -3.81E-02 | 2.05E-03 |
| 2696 | Pgam2 | PGAM2 | 8.87E-02 | 1.53E-02 |
| 2697 | Pygl | PYGL | -8.07E-02 | -3.76E-02 |
| 2698 | Trib3 | TRIB3 | -1.26E-01 | 8.56E-02 |
| 2699 | Orai2 | ORAI2 | 1.16E-01 | -6.06E-04 |
| 2700 | Ly96 | LY96 | -1.03E-01 | 3.53E-02 |

| | | | | |
|---|---|---|---|---|
| 2701 | Prom1 | PROM1 | -3.95E-02 | -2.21E-02 |
| 2702 | Suco | SUCO | 3.51E-02 | -3.66E-03 |
| 2703 | Aard | AARD | 1.40E-01 | 9.51E-02 |
| 2704 | Dcst2 | DCST2 | 9.89E-02 | -1.58E-02 |
| 2705 | Nabp2 | NABP2 | 3.31E-02 | -1.25E-02 |
| 2706 | Svip | SVIP | -3.91E-02 | -3.13E-02 |
| 2707 | Trpm6 | TRPM6 | 7.99E-02 | -1.65E-02 |
| 2708 | Fth1 | FTH1 | 4.70E-02 | 3.18E-03 |
| 2709 | Vash1 | VASH1 | -1.33E-01 | 8.23E-02 |
| 2710 | Nthl1 | NTHL1 | -6.32E-02 | 5.52E-03 |
| 2711 | Sdad1 | SDAD1 | 3.30E-02 | 1.25E-02 |
| 2712 | Usp32 | USP32 | 3.18E-02 | -2.41E-02 |
| 2713 | G6pc | G6PC | 1.20E-01 | 2.01E-02 |
| 2714 | Il17re | IL17RE | -9.78E-02 | -1.72E-02 |
| 2715 | Kcnq3 | KCNQ3 | -1.24E-01 | 1.10E-01 |
| 2716 | Cd1d1 | CD1D | -1.16E-01 | 4.19E-02 |
| 2717 | Sf3a1 | SF3A1 | -2.24E-02 | 9.28E-03 |
| 2718 | Qpctl | QPCTL | 3.01E-02 | -3.57E-03 |
| 2719 | Spp1 | SPP1 | 5.62E-02 | 9.59E-02 |
| 2720 | Ppp2r5d | PPP2R5D | 2.31E-02 | -8.81E-03 |
| 2721 | Ccdc126 | CCDC126 | -6.85E-02 | 1.49E-02 |
| 2722 | Hdlbp | HDLBP | 2.81E-02 | -8.27E-03 |
| 2723 | Abra | ABRA | 8.51E-02 | 4.65E-03 |
| 2724 | Ring1 | RING1 | -3.40E-02 | -1.82E-02 |
| 2725 | Vps13d | VPS13D | 2.59E-02 | -1.08E-02 |
| 2726 | Actn4 | ACTN4 | 4.01E-02 | -9.08E-03 |
| 2727 | Gcat | GCAT | -6.07E-02 | -1.82E-02 |
| 2728 | D7Ertd443e | C10orf90 | -1.47E-01 | -2.33E-02 |
| 2729 | Misp | MISP | -1.79E-01 | -1.93E-02 |
| 2730 | Mas1 | MAS1 | 2.03E-01 | -1.30E-02 |
| 2731 | Dnaja1 | DNAJA1 | -6.42E-02 | 2.39E-03 |
| 2732 | Syt5 | SYT5 | 1.16E-01 | -1.74E-02 |
| 2733 | Nap1l1 | NAP1L1 | -3.46E-02 | 1.05E-02 |
| 2734 | Tmem67 | TMEM67 | -5.01E-02 | 4.53E-03 |
| 2735 | Spty2d1 | SPTY2D1 | -3.30E-02 | 3.50E-03 |
| 2736 | Chmp3 | CHMP3 | 2.35E-02 | -1.56E-02 |
| 2737 | Gid4 | GID4 | -4.74E-02 | -2.10E-02 |
| 2738 | Bgn | BGN | 7.17E-02 | 2.34E-02 |
| 2739 | Ascc1 | ASCC1 | 5.50E-02 | -2.47E-02 |
| 2740 | Akt1 | AKT1 | 2.17E-02 | 8.58E-03 |
| 2741 | Hadh | HADH | 5.12E-02 | -2.05E-02 |
| 2742 | Smdt1 | SMDT1 | 3.22E-02 | -1.40E-02 |
| 2743 | Usp2 | USP2 | 6.19E-02 | -3.53E-02 |
| 2744 | Ndufa7 | NDUFA7 | 3.08E-02 | -6.11E-03 |
| 2745 | Erap1 | ERAP1 | -3.35E-02 | 2.00E-02 |
| 2746 | Mybpc3 | MYBPC3 | -1.36E-01 | -1.75E-03 |
| 2747 | Tmf1 | TMF1 | 3.13E-02 | -8.49E-03 |
| 2748 | Poc5 | POC5 | -3.40E-02 | -1.04E-02 |
| 2749 | Rnf4 | RNF4 | 2.35E-02 | 1.13E-03 |
| 2750 | Tmed2 | TMED2 | 2.68E-02 | -8.36E-03 |
| 2751 | Bmp4 | BMP4 | 5.58E-02 | -2.94E-02 |
| 2752 | Ltbp3 | LTBP3 | 3.52E-02 | 1.39E-05 |
| 2753 | Aspscr1 | ASPSCR1 | 4.07E-02 | -1.22E-02 |

| | | | | |
|---|---|---|---|---|
| 2754 | Samd5 | SAMD5 | 1.28E-01 | 1.45E-01 |
| 2755 | Laptm4b | LAPTM4B | -5.93E-02 | -3.65E-02 |
| 2756 | Klhl32 | KLHL32 | -9.84E-02 | -2.70E-02 |
| 2757 | Marveld3 | MARVELD3 | -4.97E-02 | -1.80E-02 |
| 2758 | BC049715 | C12orf60 | 9.09E-02 | 6.47E-02 |
| 2759 | Ufm1 | UFM1 | 2.80E-02 | -9.97E-03 |
| 2760 | Apoo | APOO | 3.18E-02 | -2.25E-02 |
| 2761 | Erh | ERH | 3.73E-02 | 6.88E-03 |
| 2762 | Alg2 | ALG2 | 3.29E-02 | -1.48E-02 |
| 2763 | Dennd4c | DENND4C | -4.70E-02 | -5.33E-03 |
| 2764 | Slc15a1 | SLC15A1 | 1.21E-01 | -4.08E-02 |
| 2765 | Pcnp | PCNP | 2.95E-02 | -1.74E-02 |
| 2766 | Ppp2r2d | PPP2R2D | 2.57E-02 | 3.86E-03 |
| 2767 | Frmd3 | FRMD3 | 8.83E-02 | 2.46E-02 |
| 2768 | Ikzf5 | IKZF5 | -3.70E-02 | 1.37E-02 |
| 2769 | Sec23ip | SEC23IP | -2.65E-02 | 6.49E-03 |
| 2770 | Tnk2 | TNK2 | 4.73E-02 | -2.35E-02 |
| 2771 | Klhl31 | KLHL31 | 1.00E-01 | -3.37E-02 |
| 2772 | Ppp1r14a | PPP1R14A | 1.29E-01 | -4.24E-02 |
| 2773 | Smagp | SMAGP | -4.54E-02 | -1.11E-02 |
| 2774 | Plscr3 | PLSCR3 | 3.96E-02 | 1.51E-02 |
| 2775 | Trnp1 | TRNP1 | 1.63E-01 | -5.47E-02 |
| 2776 | Rgs11 | RGS11 | 9.22E-02 | 6.37E-03 |
| 2777 | Pcyox1 | PCYOX1 | 2.61E-02 | -8.38E-03 |
| 2778 | Ccnd2 | CCND2 | -5.92E-02 | 3.03E-02 |
| 2779 | Gcnt4 | GCNT4 | 8.29E-02 | 1.35E-02 |
| 2780 | Arl4a | ARL4A | 5.04E-02 | -3.51E-03 |
| 2781 | Sgsm3 | SGSM3 | 4.68E-02 | 1.20E-02 |
| 2782 | Gpr174 | GPR174 | -7.55E-02 | 2.77E-02 |
| 2783 | Il23a | IL23A | -6.45E-02 | 3.56E-02 |
| 2784 | Eml3 | EML3 | 3.39E-02 | -4.07E-03 |
| 2785 | Ift140 | IFT140 | -3.28E-02 | 7.89E-03 |
| 2786 | Creb1 | CREB1 | -4.57E-02 | 1.50E-02 |
| 2787 | Gng7 | GNG7 | -1.27E-01 | -2.92E-02 |
| 2788 | Hmgcs1 | HMGCS1 | 5.16E-02 | 6.78E-03 |
| 2789 | Acsm1 | ACSM1 | -4.35E-02 | -2.63E-02 |
| 2790 | Tcap | TCAP | 7.11E-02 | -6.97E-02 |
| 2791 | Glyctk | GLYCTK | -5.09E-02 | -2.75E-02 |
| 2792 | Fam117a | FAM117A | 1.03E-01 | -3.67E-02 |
| 2793 | Def8 | DEF8 | 4.43E-02 | 1.04E-03 |
| 2794 | Prkd1 | PRKD1 | -6.33E-02 | 1.46E-03 |
| 2795 | Epn2 | EPN2 | -3.55E-02 | -4.80E-03 |
| 2796 | Tmem107 | TMEM107 | -5.57E-02 | 6.70E-03 |
| 2797 | Spdl1 | SPDL1 | 1.71E-01 | 6.66E-02 |
| 2798 | Ndn | NDN | -1.35E-01 | 6.65E-02 |
| 2799 | Chchd7 | CHCHD7 | -2.98E-02 | 2.17E-04 |
| 2800 | 2610528J11Rik | C1orf210 | -4.53E-02 | -1.94E-02 |
| 2801 | AW209491 | C7orf25 | -4.50E-02 | 3.28E-02 |
| 2802 | Armcx3 | ARMCX3 | -5.37E-02 | 5.82E-02 |
| 2803 | 2610301B20Rik | C8orf37 | 4.42E-02 | 6.21E-03 |
| 2804 | Gtf2h4 | GTF2H4 | -4.47E-02 | 7.21E-03 |
| 2805 | Eda2r | EDA2R | 1.72E-01 | 7.17E-03 |
| 2806 | 1700102P08Rik | C3orf62 | -1.19E-01 | 4.73E-02 |

| | | | | |
|---|---|---|---|---|
| 2807 | Phyhd1 | PHYHD1 | 7.67E-02 | 1.63E-02 |
| 2808 | Tnnc1 | TNNC1 | -1.42E-01 | 3.10E-02 |
| 2809 | Psmd10 | PSMD10 | 3.34E-02 | 2.61E-02 |
| 2810 | Cpt1c | CPT1C | 1.28E-01 | -4.31E-03 |
| 2811 | Slc35f3 | SLC35F3 | 9.56E-02 | -5.86E-02 |
| 2812 | Dmwd | DMWD | -2.47E-02 | 8.06E-04 |
| 2813 | Csgalnact1 | CSGALNACT1 | 8.98E-02 | -1.77E-02 |
| 2814 | Prc1 | PRC1 | 2.75E-01 | 2.07E-01 |
| 2815 | Lrp12 | LRP12 | 5.42E-02 | 8.81E-03 |
| 2816 | Tmem41a | TMEM41A | 6.87E-02 | -2.10E-03 |
| 2817 | Atp6v0d1 | ATP6V0D1 | 3.33E-02 | -3.05E-03 |
| 2818 | Dnal4 | DNAL4 | -2.94E-02 | -3.21E-03 |
| 2819 | Vegfb | VEGFB | -3.07E-02 | -5.15E-03 |
| 2820 | Adpgk | ADPGK | 2.78E-02 | 1.81E-02 |
| 2821 | Lrrc20 | LRRC20 | 1.06E-01 | 4.52E-03 |
| 2822 | Llph | LLPH | -4.23E-02 | 2.78E-02 |
| 2823 | Klkb1 | KLKB1 | -1.25E-01 | 8.52E-02 |
| 2824 | Larp4b | LARP4B | 2.75E-02 | -8.27E-03 |
| 2825 | Nags | NAGS | -6.33E-02 | 8.20E-03 |
| 2826 | Nol6 | NOL6 | 3.39E-02 | -4.72E-03 |
| 2827 | Adat2 | ADAT2 | -8.39E-02 | 2.05E-02 |
| 2828 | Rpa3 | RPA3 | -7.55E-02 | 4.76E-02 |
| 2829 | Ngb | NGB | -9.75E-02 | -3.99E-02 |
| 2830 | Suox | SUOX | 3.93E-02 | -1.80E-02 |
| 2831 | Gpr132 | GPR132 | -1.55E-01 | 1.42E-01 |
| 2832 | Fam216a | FAM216A | 4.18E-02 | 4.20E-03 |
| 2833 | Haus3 | HAUS3 | 5.96E-02 | 1.79E-02 |
| 2834 | Bex4 | BEX4 | -9.78E-02 | 5.97E-02 |
| 2835 | Pla2g2c | PLA2G2C | 1.76E-01 | -9.15E-02 |
| 2836 | Kcnu1 | KCNU1 | -1.06E-01 | 5.91E-03 |
| 2837 | Rpgrip1l | RPGRIP1L | -5.31E-02 | -4.12E-04 |
| 2838 | Gabpa | GABPA | -4.02E-02 | -1.68E-02 |
| 2839 | Fndc7 | FNDC7 | -5.44E-02 | -2.63E-03 |
| 2840 | Gprasp1 | GPRASP1 | -6.33E-02 | -3.56E-02 |
| 2841 | Slc38a7 | SLC38A7 | -3.64E-02 | 9.08E-03 |
| 2842 | Dhps | DHPS | 3.64E-02 | 5.84E-03 |
| 2843 | Myo1g | MYO1G | -1.99E-01 | 1.84E-01 |
| 2844 | Tmem178b | TMEM178B | 1.26E-01 | -3.65E-02 |
| 2845 | Slc35b3 | SLC35B3 | 3.12E-02 | -5.84E-03 |
| 2846 | Creb3 | CREB3 | 2.45E-02 | 1.95E-03 |
| 2847 | Otud7b | OTUD7B | 2.88E-02 | -2.19E-02 |
| 2848 | Krt20 | KRT20 | 3.08E-01 | 5.24E-01 |
| 2849 | Il33 | IL33 | 1.53E-01 | -4.24E-02 |
| 2850 | Hsd17b14 | HSD17B14 | -1.52E-01 | -1.48E-01 |
| 2851 | Akap7 | AKAP7 | -5.76E-02 | -1.88E-02 |
| 2852 | Atp13a1 | ATP13A1 | 2.77E-02 | -1.18E-02 |
| 2853 | Pcdhb22 | PCDHB15 | -8.81E-02 | 6.44E-02 |
| 2854 | Atp1b1 | ATP1B1 | 2.85E-02 | -1.04E-02 |
| 2855 | Sox5 | SOX5 | -1.36E-01 | -5.85E-02 |
| 2856 | G6pdx | G6PD | 4.26E-02 | 1.76E-02 |
| 2857 | Pask | PASK | 1.05E-01 | 3.94E-02 |
| 2858 | Zfp879 | ZNF879 | -1.07E-01 | -2.07E-03 |
| 2859 | Grap2 | GRAP2 | -1.59E-01 | 2.12E-02 |

| | | | | |
|---|---|---|---|---|
| 2860 | Dbnl | DBNL | 2.56E-02 | -8.59E-03 |
| 2861 | Med31 | MED31 | -6.60E-02 | 5.80E-03 |
| 2862 | Arl5b | ARL5B | -5.64E-02 | -3.42E-03 |
| 2863 | Gpbar1 | GPBAR1 | 6.93E-02 | 9.11E-02 |
| 2864 | Zswim1 | ZSWIM1 | 3.85E-02 | -1.01E-02 |
| 2865 | Psma6 | PSMA6 | 3.07E-02 | -9.39E-03 |
| 2866 | Ovca2 | OVCA2 | -5.57E-02 | -1.70E-03 |
| 2867 | Skap1 | SKAP1 | -1.04E-01 | 1.12E-02 |
| 2868 | Ubfd1 | UBFD1 | -4.41E-02 | 1.35E-03 |
| 2869 | Lif | LIF | 1.53E-01 | 2.42E-01 |
| 2870 | Psap | PSAP | 4.54E-02 | -2.79E-03 |
| 2871 | Fbxl2 | FBXL2 | 1.42E-01 | 3.22E-02 |
| 2872 | Taf4b | TAF4B | 6.81E-02 | 3.73E-02 |
| 2873 | Cblb | CBLB | 4.07E-02 | 1.72E-04 |
| 2874 | Elp5 | ELP5 | 2.56E-02 | -8.32E-04 |
| 2875 | Stk17b | STK17B | 5.36E-02 | 5.72E-02 |
| 2876 | Amot | AMOT | -6.22E-02 | 9.84E-03 |
| 2877 | Thnsl2 | THNSL2 | 5.52E-02 | -2.05E-02 |
| 2878 | Ppp1r13b | PPP1R13B | 2.81E-02 | -1.18E-02 |
| 2879 | Srbd1 | SRBD1 | -5.14E-02 | 9.71E-03 |
| 2880 | Ndufa6 | NDUFA6 | 3.43E-02 | -1.04E-02 |
| 2881 | Nup98 | NUP98 | 2.30E-02 | -5.53E-03 |
| 2882 | Cops3 | COPS3 | 2.78E-02 | -5.00E-03 |
| 2883 | Msl1 | MSL1 | -3.07E-02 | 1.14E-02 |
| 2884 | Thyn1 | THYN1 | 4.11E-02 | -2.37E-03 |
| 2885 | Slc38a10 | SLC38A10 | -2.78E-02 | -2.14E-02 |
| 2886 | Tapbpl | TAPBPL | -7.27E-02 | 5.40E-02 |
| 2887 | Oas2 | OAS2 | -2.32E-01 | 1.35E-01 |
| 2888 | Fen1 | FEN1 | 7.98E-02 | 2.89E-02 |
| 2889 | Fabp4 | FABP4 | 8.02E-02 | 2.03E-02 |
| 2890 | Mcc | MCC | -7.57E-02 | -2.74E-02 |
| 2891 | Vstm2b | VSTM2B | -8.66E-02 | 4.02E-02 |
| 2892 | Zc3h12a | ZC3H12A | 8.89E-02 | 3.64E-03 |
| 2893 | Slc26a4 | SLC26A4 | -7.46E-02 | 8.30E-02 |
| 2894 | Traf3ip1 | TRAF3IP1 | 3.72E-02 | -7.89E-03 |
| 2895 | Stox2 | STOX2 | -3.96E-02 | 1.06E-02 |
| 2896 | Cyth1 | CYTH1 | 2.85E-02 | -3.39E-03 |
| 2897 | Srl | SRL | 8.66E-02 | 1.37E-02 |
| 2898 | Cyb5d2 | CYB5D2 | -3.39E-02 | -2.57E-03 |
| 2899 | Cd8a | CD8A | -1.57E-01 | 9.54E-02 |
| 2900 | Il1rl1 | IL1RL1 | -1.34E-01 | 2.56E-02 |
| 2901 | Trappc3 | TRAPPC3 | 2.63E-02 | -9.37E-03 |
| 2902 | Rnft2 | RNFT2 | 1.18E-01 | -1.99E-02 |
| 2903 | Myl4 | MYL4 | 1.08E-01 | -1.60E-02 |
| 2904 | Pld4 | PLD4 | -1.11E-01 | 1.29E-01 |
| 2905 | Txnip | TXNIP | 5.54E-02 | -2.99E-02 |
| 2906 | Braf | BRAF | 2.76E-02 | -2.02E-02 |
| 2907 | Polr3gl | POLR3GL | -3.88E-02 | -2.23E-02 |
| 2908 | Slc27a4 | SLC27A4 | 2.41E-02 | -1.09E-02 |
| 2909 | Nfkbie | NFKBIE | -7.09E-02 | 1.62E-01 |
| 2910 | Slc10a6 | SLC10A6 | 1.80E-01 | -2.39E-02 |
| 2911 | Ubn1 | UBN1 | -2.58E-02 | -2.03E-03 |
| 2912 | Zfp931 | ZNF559-ZNF177 | -6.59E-02 | 4.88E-03 |

| | | | | |
|---|---|---|---|---|
| 2913 | Pex7 | PEX7 | 2.44E-02 | -1.57E-02 |
| 2914 | Dhrs3 | DHRS3 | -4.86E-02 | -9.21E-03 |
| 2915 | Zc3hav1l | ZC3HAV1L | -1.24E-01 | 2.66E-02 |
| 2916 | Plp1 | PLP1 | -7.26E-02 | -1.27E-02 |
| 2917 | C130074G19Rik | C1orf115 | 4.71E-02 | 1.87E-03 |
| 2918 | Cox7b | COX7B | 2.92E-02 | -1.24E-02 |
| 2919 | H2-T23 | HLA-E | -7.33E-02 | -2.50E-02 |
| 2920 | Ndufs6 | NDUFS6 | 2.98E-02 | -2.38E-02 |
| 2921 | Mfap5 | MFAP5 | 1.41E-01 | 1.62E-01 |
| 2922 | Igsf3 | IGSF3 | 6.61E-02 | -1.61E-02 |
| 2923 | Lat | LAT | -1.04E-01 | 3.08E-02 |
| 2924 | Fzd1 | FZD1 | -9.07E-02 | 2.13E-02 |
| 2925 | Pomk | POMK | -4.53E-02 | -3.90E-03 |
| 2926 | Dzip3 | DZIP3 | -7.82E-02 | -1.75E-02 |
| 2927 | Tstd2 | TSTD2 | -4.05E-02 | 3.70E-03 |
| 2928 | Gbp6 | GBP6 | -3.02E-01 | 1.20E-01 |
| 2929 | Cav1 | CAV1 | -6.24E-02 | 4.72E-02 |
| 2930 | Sh3bp2 | SH3BP2 | -1.35E-01 | 2.84E-02 |
| 2931 | Sla2 | SLA2 | -5.23E-02 | 2.49E-02 |
| 2932 | Inpp5e | INPP5E | -2.86E-02 | -1.23E-02 |
| 2933 | Zbtb25 | ZBTB25 | 9.87E-02 | -3.25E-02 |
| 2934 | Wnt5b | WNT5B | -1.22E-01 | 1.80E-03 |
| 2935 | Mast4 | MAST4 | 5.07E-02 | -7.77E-03 |
| 2936 | Bmper | BMPER | 8.03E-02 | 4.25E-02 |
| 2937 | Hira | HIRA | 6.08E-02 | -7.01E-03 |
| 2938 | Ncoa7 | NCOA7 | -6.15E-02 | 1.52E-03 |
| 2939 | Zfp598 | ZNF598 | 2.65E-02 | 5.30E-04 |
| 2940 | Rab2a | RAB2A | 2.56E-02 | -1.10E-02 |
| 2941 | Tnnt1 | TNNT1 | -9.80E-02 | 6.04E-02 |
| 2942 | Ms4a7 | MS4A7 | -1.97E-01 | 1.56E-01 |
| 2943 | Clhc1 | AUNIP | -7.78E-02 | 1.84E-02 |
| 2944 | Edc3 | EDC3 | 4.18E-02 | -3.05E-02 |
| 2945 | Stk32b | STK32B | -5.47E-02 | -2.57E-02 |
| 2946 | Sh3kbp1 | SH3KBP1 | -5.93E-02 | 1.76E-02 |
| 2947 | Il5ra | IL5RA | 1.71E-01 | -1.61E-02 |
| 2948 | Setd7 | SETD7 | 3.60E-02 | 2.47E-03 |
| 2949 | Lpo | LPO | 1.13E-01 | 5.22E-02 |
| 2950 | Ino80 | INO80 | 2.32E-02 | -1.86E-02 |
| 2951 | Ibtk | IBTK | 2.22E-02 | -4.56E-03 |
| 2952 | Ifrd1 | IFRD1 | 9.98E-02 | 7.84E-03 |
| 2953 | Ebag9 | EBAG9 | 2.89E-02 | -1.90E-02 |
| 2954 | Plod1 | PLOD1 | 3.13E-02 | -1.18E-03 |
| 2955 | Entpd2 | ENTPD2 | -8.94E-02 | 2.91E-02 |
| 2956 | Cnpy2 | CNPY2 | -3.26E-02 | -2.84E-03 |
| 2957 | Ppil6 | PPIL6 | 6.26E-02 | -5.34E-02 |
| 2958 | Bbs7 | BBS7 | 5.60E-02 | -2.19E-02 |
| 2959 | Ezh2 | EZH2 | 8.13E-02 | 8.05E-02 |
| 2960 | Cope | COPE | 2.47E-02 | -1.61E-02 |
| 2961 | Ndrg3 | NDRG3 | 3.09E-02 | -2.65E-02 |
| 2962 | Pdlim7 | PDLIM7 | 7.09E-02 | 6.16E-02 |
| 2963 | Slc16a6 | SLC16A6 | 1.37E-01 | 6.63E-02 |
| 2964 | Tekt4 | TEKT4 | 6.11E-02 | -3.53E-02 |
| 2965 | Txndc12 | TXNDC12 | -3.12E-02 | -5.98E-03 |

| | | | | |
|---|---|---|---|---|
| 2966 | Uchl3 | UCHL3 | 4.79E-02 | 8.18E-04 |
| 2967 | Otud1 | OTUD1 | -7.71E-02 | -3.26E-02 |
| 2968 | Eif2b3 | EIF2B3 | 4.41E-02 | 1.34E-02 |
| 2969 | Sparc | SPARC | 6.57E-02 | 6.22E-02 |
| 2970 | Chaf1b | CHAF1B | 1.57E-01 | 1.18E-01 |
| 2971 | Dalrd3 | DALRD3 | -3.02E-02 | -7.45E-03 |
| 2972 | Il21r | IL21R | -1.28E-01 | 5.84E-02 |
| 2973 | Pdcd2l | PDCD2L | 3.86E-02 | -2.45E-02 |
| 2974 | Men1 | MEN1 | -3.80E-02 | 6.63E-03 |
| 2975 | Rabgap1l | RABGAP1L | 3.11E-02 | -2.09E-02 |
| 2976 | Nhp2 | NHP2 | 5.23E-02 | -5.62E-04 |
| 2977 | Arhgdia | ARHGDIA | 2.50E-02 | 1.24E-03 |
| 2978 | Vdr | VDR | 7.55E-02 | -3.24E-02 |
| 2979 | Irf3 | IRF3 | -2.87E-02 | 3.43E-03 |
| 2980 | Chaf1a | CHAF1A | 1.42E-01 | 4.42E-02 |
| 2981 | Rgs4 | RGS4 | 1.41E-01 | 7.07E-02 |
| 2982 | Psma3 | PSMA3 | 2.56E-02 | -1.63E-03 |
| 2983 | Pls3 | PLS3 | 4.19E-02 | 1.90E-03 |
| 2984 | Dcaf13 | DCAF13 | 2.89E-02 | -3.68E-04 |
| 2985 | Ash1l | ASH1L | -2.53E-02 | -7.01E-03 |
| 2986 | C1ra | C1R | -1.02E-01 | 4.18E-02 |
| 2987 | Atp6ap1 | ATP6AP1 | 2.44E-02 | -5.36E-03 |
| 2988 | Dnaja4 | DNAJA4 | 4.96E-02 | -1.29E-02 |
| 2989 | 1110004F10Rik | C11orf58 | 2.57E-02 | -2.70E-03 |
| 2990 | Gbp10 | GBP6 | -2.55E-01 | 6.38E-02 |
| 2991 | Frs3 | FRS3 | -7.58E-02 | -4.12E-02 |
| 2992 | Irf8 | IRF8 | -9.11E-02 | 5.02E-02 |
| 2993 | Gabrg1 | GABRG1 | 7.13E-02 | -7.98E-02 |
| 2994 | Syk | SYK | -1.81E-01 | 2.31E-01 |
| 2995 | Pex11g | PEX11G | 5.05E-02 | -2.68E-02 |
| 2996 | Tspan14 | TSPAN14 | -2.76E-02 | 4.58E-03 |
| 2997 | Fermt1 | FERMT1 | 6.63E-02 | -2.67E-02 |
| 2998 | Pck1 | PCK1 | 7.90E-02 | 8.39E-03 |
| 2999 | Nbas | NBAS | -3.15E-02 | -2.88E-02 |
| 3000 | Ints6 | INTS6 | -5.46E-02 | 3.61E-02 |
| 3001 | Enox2 | ENOX2 | -3.91E-02 | -1.34E-02 |
| 3002 | Tead1 | TEAD1 | 4.06E-02 | -1.31E-02 |
| 3003 | Zbed6 | ZBED6 | -8.82E-02 | 5.12E-03 |
| 3004 | Mrpl27 | MRPL27 | 3.32E-02 | -1.27E-02 |
| 3005 | Elac2 | ELAC2 | 3.15E-02 | -1.36E-02 |
| 3006 | Reep5 | REEP5 | -3.86E-02 | -1.06E-02 |
| 3007 | Tspan33 | TSPAN33 | 2.78E-02 | -4.36E-03 |
| 3008 | Dyrk1b | DYRK1B | 4.17E-02 | -5.24E-02 |
| 3009 | Eif2b5 | EIF2B5 | 2.59E-02 | -6.10E-03 |
| 3010 | Cdca7l | CDCA7L | -8.68E-02 | -7.98E-02 |
| 3011 | Pik3c2g | PIK3C2G | -1.85E-01 | -3.94E-02 |
| 3012 | Ccdc84 | CCDC84 | 2.94E-02 | -1.97E-02 |
| 3013 | Jam3 | JAM3 | 6.06E-02 | -2.06E-03 |
| 3014 | Zfp518a | ZNF518A | -4.34E-02 | -8.49E-03 |
| 3015 | Cttnbp2 | CTTNBP2 | -7.72E-02 | -1.09E-02 |
| 3016 | Gga3 | GGA3 | -4.03E-02 | -6.82E-03 |
| 3017 | Ildr1 | ILDR1 | 6.52E-02 | -1.82E-02 |
| 3018 | Pibf1 | PIBF1 | -5.09E-02 | 5.31E-03 |

| | | | | |
|---|---|---|---|---|
| 3019 | Hmmr | HMMR | 2.91E-01 | 1.76E-01 |
| 3020 | Polr2i | POLR2I | 3.86E-02 | -2.11E-02 |
| 3021 | Ltbp2 | LTBP2 | 1.57E-01 | 3.49E-01 |
| 3022 | Kcnab1 | KCNAB1 | 1.10E-01 | -8.60E-03 |
| 3023 | Pebp1 | PEBP1 | 3.12E-02 | -6.74E-03 |
| 3024 | Cyb5d1 | CYB5D1 | -5.30E-02 | -1.94E-02 |
| 3025 | Becn1 | BECN1 | 2.30E-02 | -1.69E-03 |
| 3026 | Serpinb6a | SERPINB6 | 4.31E-02 | 3.72E-02 |
| 3027 | Gpx4 | GPX4 | 3.70E-02 | -1.44E-02 |
| 3028 | Vill | VILL | -6.27E-02 | -1.68E-02 |
| 3029 | Dnah2 | DNAH2 | -1.01E-01 | -2.48E-02 |
| 3030 | Dnajc10 | DNAJC10 | 2.34E-02 | 1.73E-03 |
| 3031 | Lrrc8b | LRRC8B | -5.35E-02 | -2.75E-02 |
| 3032 | Mrps23 | MRPS23 | 3.09E-02 | -1.93E-02 |
| 3033 | Pi4kb | PI4KB | -2.19E-02 | 2.29E-03 |
| 3034 | Ifrd2 | IFRD2 | 5.03E-02 | -7.94E-03 |
| 3035 | Hypk | HYPK | 9.34E-02 | -6.50E-02 |
| 3036 | Kcnk2 | KCNK2 | -2.33E-01 | 1.22E-03 |
| 3037 | Zfp579 | ZNF579 | -3.17E-02 | -3.43E-05 |
| 3038 | Zfand6 | ZFAND6 | 2.50E-02 | -7.47E-04 |
| 3039 | Kif22 | KIF22 | 2.13E-01 | 1.99E-01 |
| 3040 | Psmb10 | PSMB10 | -1.00E-01 | 7.00E-02 |
| 3041 | Cenpv | CENPV | 4.80E-02 | -2.52E-03 |
| 3042 | Gpat2 | GPAT2 | 1.07E-01 | -5.32E-02 |
| 3043 | Usp46 | USP46 | 2.86E-02 | -6.04E-03 |
| 3044 | Suclg2 | SUCLG2 | 5.11E-02 | -6.67E-03 |
| 3045 | Sbsn | SBSN | 9.70E-02 | -3.15E-02 |
| 3046 | Fxyd6 | FXYD6 | -7.95E-02 | 4.27E-02 |
| 3047 | Vdac2 | VDAC2 | 2.54E-02 | -1.31E-02 |
| 3048 | Capzb | CAPZB | 2.83E-02 | 4.64E-03 |
| 3049 | Il6ra | IL6R | -8.77E-02 | 5.65E-02 |
| 3050 | Slc19a2 | SLC19A2 | 4.84E-02 | -1.37E-02 |
| 3051 | Fgd6 | FGD6 | -7.17E-02 | 2.61E-02 |
| 3052 | Dusp27 | DUSP27 | 4.50E-02 | -1.51E-02 |
| 3053 | Olfr78 | OR51E2 | 1.16E-01 | 5.13E-02 |
| 3054 | Srd5a2 | SRD5A2 | -1.27E-01 | -9.84E-02 |
| 3055 | Pcm1 | PCM1 | -2.74E-02 | -1.01E-02 |
| 3056 | Adtrp | ADTRP | -6.69E-02 | 1.13E-02 |
| 3057 | Sptbn5 | SPTBN5 | 1.52E-01 | 5.13E-02 |
| 3058 | Snd1 | SND1 | 3.29E-02 | -1.10E-02 |
| 3059 | Klhl29 | KLHL29 | -1.19E-01 | -6.86E-02 |
| 3060 | Strada | STRADA | 3.25E-02 | 7.89E-03 |
| 3061 | Pon2 | PON2 | -4.25E-02 | 9.75E-04 |
| 3062 | Mrpl2 | MRPL2 | 3.12E-02 | -2.48E-02 |
| 3063 | Usp6nl | USP6NL | 2.41E-02 | -6.05E-03 |
| 3064 | Thoc3 | THOC3 | 3.11E-02 | -2.19E-02 |
| 3065 | Tcof1 | TCOF1 | -4.13E-02 | 1.00E-02 |
| 3066 | Ufsp2 | UFSP2 | 3.13E-02 | -7.75E-03 |
| 3067 | Cyb561d1 | CYB561D1 | 3.34E-02 | -1.40E-02 |
| 3068 | Tomm20 | TOMM20 | 3.37E-02 | -8.83E-03 |
| 3069 | Golm1 | GOLM1 | 8.93E-02 | 3.64E-02 |
| 3070 | Ndufs3 | NDUFS3 | 3.24E-02 | -1.73E-02 |
| 3071 | Ddn | DDN | -1.18E-01 | -1.92E-02 |

| | | | | |
|---|---|---|---|---|
| 3072 | Stbd1 | STBD1 | -1.19E-01 | 3.00E-02 |
| 3073 | Utrn | UTRN | -6.41E-02 | -4.16E-02 |
| 3074 | Nr1d1 | NR1D1 | -1.20E-01 | -1.85E-01 |
| 3075 | Tnks1bp1 | TNKS1BP1 | 3.15E-02 | -2.17E-03 |
| 3076 | Pafah1b2 | PAFAH1B2 | 2.11E-02 | 4.29E-03 |
| 3077 | Mgst1 | MGST1 | 6.49E-02 | 9.95E-03 |
| 3078 | Ndufv2 | NDUFV2 | 2.91E-02 | -1.30E-02 |
| 3079 | Tbc1d16 | TBC1D16 | 3.28E-02 | -6.72E-02 |
| 3080 | Rbbp5 | RBBP5 | -3.10E-02 | 2.29E-03 |
| 3081 | Cdc34 | CDC34 | 3.49E-02 | -4.13E-04 |
| 3082 | Cyp1b1 | CYP1B1 | 1.05E-01 | -5.34E-02 |
| 3083 | Bcat2 | BCAT2 | -4.33E-02 | -2.56E-02 |
| 3084 | Hif1a | HIF1A | 4.58E-02 | 7.28E-03 |
| 3085 | Wnt9a | WNT9A | 1.18E-01 | -9.55E-03 |
| 3086 | Cwc27 | CWC27 | 3.73E-02 | -8.80E-03 |
| 3087 | Klhdc3 | KLHDC3 | 2.36E-02 | -1.06E-02 |
| 3088 | Crot | CROT | -3.14E-02 | -1.42E-02 |
| 3089 | Psip1 | PSIP1 | -3.57E-02 | 2.36E-02 |
| 3090 | Srek1 | SREK1 | 2.24E-02 | 7.57E-03 |
| 3091 | Dda1 | DDA1 | 2.01E-02 | 9.77E-03 |
| 3092 | Ranbp17 | RANBP17 | 1.05E-01 | 2.17E-02 |
| 3093 | Hoxa10 | HOXA10 | -3.22E-02 | -2.01E-02 |
| 3094 | Slc20a2 | SLC20A2 | 4.85E-02 | -1.07E-02 |
| 3095 | Stau2 | STAU2 | 2.74E-02 | -3.74E-02 |
| 3096 | Prickle4 | PRICKLE4 | -9.21E-02 | 3.56E-02 |
| 3097 | Fgl2 | FGL2 | -1.27E-01 | 1.44E-01 |
| 3098 | Exoc3l4 | EXOC3L4 | -5.99E-02 | 3.07E-02 |
| 3099 | Cbr4 | CBR4 | 3.28E-02 | -2.84E-02 |
| 3100 | Alg5 | ALG5 | 2.38E-02 | 6.79E-03 |
| 3101 | Hs3st3b1 | HS3ST3B1 | -1.34E-01 | 8.24E-03 |
| 3102 | Wdr61 | WDR61 | 3.21E-02 | -6.76E-03 |
| 3103 | Scnn1b | SCNN1B | 3.70E-02 | 8.08E-03 |
| 3104 | Arhgap39 | ARHGAP39 | -3.85E-02 | -1.64E-02 |
| 3105 | Furin | FURIN | 3.70E-02 | -1.11E-02 |
| 3106 | Gbp2b | GBP3 | -6.14E-02 | -4.97E-03 |
| 3107 | Efna2 | EFNA2 | -8.33E-02 | -1.19E-02 |
| 3108 | Trim45 | TRIM45 | -9.47E-02 | 1.59E-02 |
| 3109 | Psrc1 | PSRC1 | 1.73E-01 | 5.36E-02 |
| 3110 | Atp5d | ATP5D | 3.20E-02 | -1.54E-02 |
| 3111 | Ccr5 | CCR5 | -1.73E-01 | 1.75E-01 |
| 3112 | Mmp10 | MMP10 | 9.60E-02 | 1.56E-01 |
| 3113 | Ggact | GGACT | -3.46E-02 | -2.09E-02 |
| 3114 | Mkrn2 | MKRN2 | 2.90E-02 | -1.58E-02 |
| 3115 | Ifnar1 | IFNAR1 | -2.43E-02 | 2.25E-03 |
| 3116 | Tnfrsf13b | TNFRSF13B | -1.09E-01 | 1.04E-01 |
| 3117 | Ccdc32 | C15orf57 | 3.55E-02 | -1.72E-02 |
| 3118 | Mtpn | MTPN | 2.21E-02 | -5.63E-03 |
| 3119 | Dnah1 | DNAH1 | -9.30E-02 | 4.36E-02 |
| 3120 | Itgbl1 | ITGBL1 | 1.58E-01 | 1.12E-01 |
| 3121 | Trim66 | TRIM66 | 6.55E-02 | -6.62E-03 |
| 3122 | Ccdc88b | CCDC88B | -1.32E-01 | 1.64E-01 |
| 3123 | Net1 | NET1 | 6.71E-02 | 1.65E-02 |
| 3124 | Ppfia1 | PPFIA1 | 2.94E-02 | -1.56E-02 |

| | | | | |
|---|---|---|---|---|
| 3125 | Impact | IMPACT | 6.15E-02 | 6.67E-04 |
| 3126 | Zbtb12 | ZBTB12 | -6.80E-02 | 4.67E-02 |
| 3127 | Sez6 | SEZ6 | 1.34E-01 | -5.56E-02 |
| 3128 | Nptx1 | NPTX1 | 5.40E-01 | -1.67E-01 |
| 3129 | Morf4l1 | MORF4L1 | 1.95E-02 | -9.34E-03 |
| 3130 | Pde3b | PDE3B | 5.87E-02 | 1.59E-02 |
| 3131 | Yae1d1 | YAE1D1 | -5.91E-02 | -1.28E-03 |
| 3132 | Tst | TST | -6.20E-02 | 1.46E-02 |
| 3133 | Ech1 | ECH1 | 4.33E-02 | -2.46E-02 |
| 3134 | Lamp1 | LAMP1 | 3.34E-02 | -1.03E-02 |
| 3135 | Zdhhc21 | ZDHHC21 | 3.49E-02 | 3.57E-03 |
| 3136 | 4921507P07Rik | C7orf31 | 1.23E-01 | -4.35E-02 |
| 3137 | Slc44a2 | SLC44A2 | 2.72E-02 | 1.95E-02 |
| 3138 | Kcnk1 | KCNK1 | -8.20E-02 | -1.64E-02 |
| 3139 | Rfc5 | RFC5 | 6.03E-02 | 1.85E-02 |
| 3140 | Isy1 | ISY1-RAB43 | -3.89E-02 | 1.93E-02 |
| 3141 | Ptpn13 | PTPN13 | -4.22E-02 | -2.55E-02 |
| 3142 | Rab27a | RAB27A | 5.81E-02 | 7.02E-03 |
| 3143 | Ccdc93 | CCDC93 | -3.97E-02 | -2.03E-02 |
| 3144 | Plk5 | PLK5 | -1.79E-01 | -1.08E-01 |
| 3145 | Anln | ANLN | 2.40E-01 | 1.81E-01 |
| 3146 | Itih1 | ITIH1 | -1.31E-01 | 1.37E-01 |
| 3147 | Gpr137b | GPR137B | -4.66E-02 | -2.18E-02 |
| 3148 | Hint1 | HINT1 | 3.66E-02 | -1.63E-02 |
| 3149 | Msh3 | MSH3 | 4.45E-02 | -2.55E-02 |
| 3150 | Scai | SCAI | -6.47E-02 | -8.19E-03 |
| 3151 | Gclc | GCLC | -7.12E-02 | 4.19E-03 |
| 3152 | Tmem233 | TMEM233 | 6.66E-02 | -2.40E-02 |
| 3153 | Fam163b | FAM163B | 1.11E-01 | -2.79E-02 |
| 3154 | Fgf16 | FGF16 | -7.17E-02 | 8.58E-02 |
| 3155 | Higd1b | HIGD1B | 1.20E-01 | -4.67E-02 |
| 3156 | C1ql1 | C1QL1 | -5.21E-02 | -5.90E-03 |
| 3157 | Slc6a18 | SLC6A18 | -4.68E-02 | -4.89E-02 |
| 3158 | Spsb4 | SPSB4 | -5.83E-02 | 3.34E-02 |
| 3159 | Fblim1 | FBLIM1 | 8.60E-02 | 3.09E-02 |
| 3160 | 1700066M21Rik | C2orf69 | 4.49E-02 | 6.15E-02 |
| 3161 | Rhoh | RHOH | -8.14E-02 | 3.37E-02 |
| 3162 | Ndufa5 | NDUFA5 | 2.76E-02 | -2.01E-02 |
| 3163 | Mgmt | MGMT | 1.05E-01 | -8.55E-02 |
| 3164 | Ermp1 | ERMP1 | 5.67E-02 | -2.92E-02 |
| 3165 | Wdhd1 | WDHD1 | 1.21E-01 | 6.69E-02 |
| 3166 | Pfn1 | PFN1 | 2.39E-02 | 1.47E-02 |
| 3167 | Mapk8 | MAPK8 | 3.36E-02 | -2.35E-02 |
| 3168 | Psmb5 | PSMB5 | 3.71E-02 | -1.05E-02 |
| 3169 | Bmp8a | BMP8A | -7.33E-02 | 1.47E-02 |
| 3170 | Tcf4 | TCF4 | -3.54E-02 | -2.19E-02 |
| 3171 | Ehhadh | EHHADH | -6.51E-02 | -2.34E-02 |
| 3172 | Xrn2 | XRN2 | 4.25E-02 | -7.23E-03 |
| 3173 | Kat7 | KAT7 | 2.07E-02 | -7.87E-03 |
| 3174 | Frk | FRK | 4.11E-02 | 5.26E-02 |
| 3175 | Lrrc25 | LRRC25 | -1.23E-01 | 1.39E-01 |
| 3176 | Ksr1 | KSR1 | -6.96E-02 | 2.65E-02 |
| 3177 | Cul5 | CUL5 | 2.74E-02 | -1.15E-02 |

| | | | | |
|---|---|---|---|---|
| 3178 | Bola3 | BOLA3 | 3.13E-02 | -4.16E-03 |
| 3179 | Astn1 | ASTN1 | -9.82E-02 | 4.04E-02 |
| 3180 | Csrnp2 | CSRNP2 | 6.65E-02 | -2.61E-03 |
| 3181 | Card11 | CARD11 | -1.27E-01 | 2.63E-02 |
| 3182 | Prkci | PRKCI | -2.60E-02 | 2.75E-03 |
| 3183 | Gna11 | GNA11 | 1.96E-02 | -9.87E-03 |
| 3184 | Dnajc25 | DNAJC25 | 3.82E-02 | -9.67E-03 |
| 3185 | Cldn15 | CLDN15 | 9.67E-02 | -2.69E-02 |
| 3186 | Hnrnpdl | HNRNPDL | 3.01E-02 | -1.79E-02 |
| 3187 | Rnf144b | RNF144B | 1.18E-01 | -2.60E-02 |
| 3188 | Tex9 | TEX9 | 5.31E-02 | -5.80E-02 |
| 3189 | Uba7 | UBA7 | -8.65E-02 | 2.50E-02 |
| 3190 | Sat1 | SAT1 | 3.57E-02 | 3.03E-02 |
| 3191 | Tapt1 | TAPT1 | 2.35E-02 | -3.12E-04 |
| 3192 | Aplf | APLF | 5.87E-02 | 1.67E-02 |
| 3193 | Rab37 | RAB37 | -1.03E-01 | -6.43E-02 |
| 3194 | Tbca | TBCA | 2.84E-02 | -1.84E-04 |
| 3195 | Tfap2b | TFAP2B | -6.95E-02 | -5.37E-03 |
| 3196 | Nudt9 | NUDT9 | 2.27E-02 | -1.47E-02 |
| 3197 | Zfp324 | ZNF324 | -5.51E-02 | -3.84E-02 |
| 3198 | Mypop | MYPOP | -6.23E-02 | 3.94E-02 |
| 3199 | Scnm1 | SCNM1 | 3.40E-02 | -1.14E-02 |
| 3200 | Oxgr1 | OXGR1 | -5.74E-02 | -3.74E-03 |
| 3201 | Ogdh | OGDH | 3.47E-02 | -1.17E-02 |
| 3202 | Osbpl3 | OSBPL3 | -5.41E-02 | -3.54E-02 |
| 3203 | Clip4 | CLIP4 | 1.37E-01 | -8.11E-03 |
| 3204 | Lhx6 | LHX6 | -1.21E-01 | 2.38E-02 |
| 3205 | Bcl2l2 | BCL2L2 | -2.14E-02 | -8.06E-03 |
| 3206 | Brox | BROX | 2.38E-02 | -1.12E-02 |
| 3207 | Trmu | TRMU | 3.10E-02 | -9.04E-03 |
| 3208 | Epm2aip1 | EPM2AIP1 | -3.46E-02 | 7.86E-03 |
| 3209 | Tom1l1 | TOM1L1 | -3.13E-02 | 6.35E-03 |
| 3210 | Copg2 | COPG2 | 1.96E-02 | -1.56E-02 |
| 3211 | Dgkg | DGKG | -1.16E-01 | -3.67E-02 |
| 3212 | Ankrd65 | ANKRD65 | 1.09E-01 | -8.30E-02 |
| 3213 | Ptger2 | PTGER2 | -1.19E-01 | 3.34E-02 |
| 3214 | Slc24a3 | SLC24A3 | -1.31E-01 | 6.91E-02 |
| 3215 | Hdac4 | HDAC4 | 4.60E-02 | -2.18E-02 |
| 3216 | Stard7 | STARD7 | 2.72E-02 | -2.36E-02 |
| 3217 | Ptrhd1 | PTRHD1 | 4.74E-02 | -1.62E-03 |
| 3218 | Aqp11 | AQP11 | -6.19E-02 | -2.69E-02 |
| 3219 | Tmem179 | TMEM179 | 7.04E-02 | 1.17E-02 |
| 3220 | Kcnj4 | KCNJ4 | 7.25E-02 | -1.05E-02 |
| 3221 | Lamc3 | LAMC3 | -9.44E-02 | -8.47E-02 |
| 3222 | E2f5 | E2F5 | 3.35E-02 | 1.51E-02 |
| 3223 | Shisa2 | SHISA2 | 1.55E-01 | -6.82E-02 |
| 3224 | Setd3 | SETD3 | 2.48E-02 | -2.00E-02 |
| 3225 | Stat6 | STAT6 | -2.09E-02 | 5.85E-03 |
| 3226 | Spag8 | SPAG8 | -1.06E-01 | 1.47E-03 |
| 3227 | Syt2 | SYT2 | 1.37E-01 | 1.36E-01 |
| 3228 | Stil | STIL | 1.57E-01 | 1.20E-01 |
| 3229 | Atp5h | ATP5H | 2.57E-02 | -1.24E-02 |
| 3230 | Kifc2 | KIFC2 | -4.28E-02 | -5.63E-03 |

| | | | | |
|---|---|---|---|---|
| 3231 | Ccdc116 | CCDC116 | 1.32E-01 | -6.63E-02 |
| 3232 | Tnfrsf21 | TNFRSF21 | 4.27E-02 | -9.84E-03 |
| 3233 | Fam122a | FAM122A | 4.13E-02 | -1.75E-02 |
| 3234 | Clstn2 | CLSTN2 | -1.52E-01 | 7.58E-02 |
| 3235 | Ruvbl2 | RUVBL2 | -3.07E-02 | 1.00E-02 |
| 3236 | Syngr2 | SYNGR2 | 3.19E-02 | 1.36E-02 |
| 3237 | Hoxa5 | HOXA5 | -6.33E-02 | 2.83E-02 |
| 3238 | Eri3 | ERI3 | 3.21E-02 | 5.89E-04 |
| 3239 | Uspl1 | USPL1 | -4.85E-02 | 1.47E-02 |
| 3240 | Ppp2r1a | PPP2R1A | 2.08E-02 | -1.24E-02 |
| 3241 | Ndufa1 | NDUFA1 | 3.52E-02 | -1.87E-02 |
| 3242 | Fpgt | FPGT | -3.61E-02 | 1.54E-02 |
| 3243 | Hcrtr1 | HCRTR1 | 6.03E-02 | -3.15E-02 |
| 3244 | Ptp4a3 | PTP4A3 | -5.51E-02 | 2.27E-02 |
| 3245 | Piwil4 | PIWIL4 | 1.26E-01 | -6.12E-02 |
| 3246 | Parp2 | PARP2 | 2.49E-02 | -5.51E-03 |
| 3247 | Vps41 | VPS41 | 2.69E-02 | -1.49E-02 |
| 3248 | Pygo2 | PYGO2 | -3.45E-02 | 2.33E-02 |
| 3249 | Prr15 | PRR15 | 7.26E-02 | 3.05E-02 |
| 3250 | Engase | ENGASE | 4.63E-02 | -1.96E-02 |
| 3251 | Pcdh7 | PCDH7 | -1.87E-01 | 6.63E-02 |
| 3252 | Dlg2 | DLG2 | -1.52E-01 | -4.24E-02 |
| 3253 | Cilp | CILP | 6.90E-02 | -4.94E-02 |
| 3254 | Foxk1 | FOXK1 | -3.41E-02 | -1.08E-02 |
| 3255 | Bean1 | BEAN1 | 5.84E-02 | 2.17E-02 |
| 3256 | Trim13 | TRIM13 | 5.60E-02 | 3.60E-02 |
| 3257 | Nsun5 | NSUN5 | -3.59E-02 | 4.01E-03 |
| 3258 | Rufy1 | RUFY1 | 2.33E-02 | -1.68E-02 |
| 3259 | Cd40lg | CD40LG | -3.52E-02 | 2.47E-02 |
| 3260 | Chd1l | CHD1L | 3.72E-02 | 1.55E-03 |
| 3261 | Pdp2 | PDP2 | -6.15E-02 | -1.12E-02 |
| 3262 | Cnn1 | CNN1 | 2.05E-01 | 8.34E-02 |
| 3263 | Cldn16 | CLDN16 | 5.42E-02 | -6.47E-02 |
| 3264 | Ankrd12 | ANKRD12 | -4.01E-02 | -1.96E-02 |
| 3265 | Smoc1 | SMOC1 | 1.43E-01 | 1.02E-01 |
| 3266 | Whamm | WHAMM | 3.70E-02 | -2.50E-03 |
| 3267 | Fbxo25 | FBXO25 | -2.72E-02 | -4.74E-02 |
| 3268 | Prkar2a | PRKAR2A | 3.18E-02 | -6.49E-03 |
| 3269 | Nr3c2 | NR3C2 | -4.44E-02 | -3.62E-02 |
| 3270 | Tmem145 | TMEM145 | 1.16E-01 | 6.85E-02 |
| 3271 | Tro | TRO | 8.95E-02 | -1.53E-02 |
| 3272 | Oit3 | OIT3 | 6.68E-02 | -4.50E-02 |
| 3273 | Spata33 | SPATA33 | 6.85E-02 | -2.61E-02 |
| 3274 | Slc35e1 | SLC35E1 | -2.16E-02 | -4.07E-03 |
| 3275 | Cxcr3 | CXCR3 | -1.53E-01 | 4.74E-02 |
| 3276 | Klhdc7a | KLHDC7A | -3.91E-02 | -8.70E-03 |
| 3277 | Atf7ip2 | ATF7IP2 | -1.12E-01 | 3.20E-02 |
| 3278 | Map7d2 | MAP7D2 | 8.27E-02 | 2.81E-02 |
| 3279 | Galk2 | GALK2 | 3.55E-02 | -9.45E-03 |
| 3280 | Nme7 | NME7 | -3.31E-02 | 4.73E-03 |
| 3281 | U2surp | U2SURP | -2.29E-02 | 1.73E-03 |
| 3282 | Mfsd11 | MFSD11 | 3.62E-02 | -3.27E-03 |
| 3283 | Usp40 | USP40 | 2.42E-02 | -1.26E-02 |

| | | | | |
|---|---|---|---|---|
| 3284 | Atxn7l2 | ATXN7L2 | 6.07E-02 | 8.69E-03 |
| 3285 | Pcx | PC | 6.63E-02 | -6.07E-03 |
| 3286 | Grhpr | GRHPR | 4.83E-02 | -2.49E-02 |
| 3287 | Pkhd1 | PKHD1 | -3.42E-02 | -1.67E-02 |
| 3288 | Mib2 | MIB2 | -3.02E-02 | -1.22E-02 |
| 3289 | Clip1 | CLIP1 | 2.82E-02 | -2.45E-02 |
| 3290 | Nr2c1 | NR2C1 | -3.99E-02 | 5.94E-03 |
| 3291 | Cnr2 | CNR2 | -1.57E-01 | 4.94E-02 |
| 3292 | Zfp65 | ZNF253 | -4.78E-02 | 1.56E-03 |
| 3293 | Tap1 | TAP1 | -1.41E-01 | 3.22E-02 |
| 3294 | Slc10a4 | SLC10A4 | -4.23E-02 | 1.69E-02 |
| 3295 | Zcchc8 | ZCCHC8 | -2.27E-02 | -9.68E-03 |
| 3296 | Psmd7 | PSMD7 | 2.35E-02 | -1.64E-03 |
| 3297 | Asxl1 | ASXL1 | 2.75E-02 | -1.92E-02 |
| 3298 | Rdh14 | RDH14 | 2.70E-02 | -1.66E-02 |
| 3299 | Kctd21 | KCTD21 | -8.58E-02 | 3.72E-02 |
| 3300 | Got2 | GOT2 | 3.75E-02 | -1.79E-02 |
| 3301 | Kctd1 | KCTD1 | 4.53E-02 | 3.77E-02 |
| 3302 | Hrh1 | HRH1 | -6.27E-02 | 1.29E-02 |
| 3303 | Rab26 | RAB26 | -6.20E-02 | 1.16E-02 |
| 3304 | Mafk | MAFK | 6.05E-02 | -2.64E-02 |
| 3305 | Dhx29 | DHX29 | 3.40E-02 | -1.21E-02 |
| 3306 | Cpne4 | CPNE4 | -1.35E-01 | 3.36E-02 |
| 3307 | Eif4g3 | EIF4G3 | 3.01E-02 | -2.42E-02 |
| 3308 | Nsun2 | NSUN2 | 2.71E-02 | -1.24E-02 |
| 3309 | Sumf2 | SUMF2 | -3.07E-02 | 5.24E-03 |
| 3310 | Sltm | SLTM | 2.40E-02 | -9.39E-03 |
| 3311 | Tmem45a | TMEM45A | 1.60E-01 | 6.93E-02 |
| 3312 | Prss8 | PRSS8 | -3.58E-02 | -2.31E-02 |
| 3313 | Batf2 | BATF2 | -1.06E-01 | -1.23E-02 |
| 3314 | Clic3 | CLIC3 | -9.48E-02 | -5.72E-02 |
| 3315 | Jph2 | JPH2 | 1.20E-01 | 1.41E-02 |
| 3316 | Tnrc6b | TNRC6B | -2.26E-02 | -8.85E-03 |
| 3317 | N6amt1 | N6AMT1 | 4.33E-02 | -1.30E-02 |
| 3318 | Znhit1 | ZNHIT1 | 2.91E-02 | -2.09E-02 |
| 3319 | Myh2 | MYH2 | -4.51E-02 | -1.23E-02 |
| 3320 | Adamts3 | ADAMTS3 | 1.13E-01 | -3.51E-03 |
| 3321 | Dbi | DBI | 4.20E-02 | 7.43E-03 |
| 3322 | Cgref1 | CGREF1 | -3.95E-02 | -1.98E-02 |
| 3323 | Smim5 | SMIM5 | -7.51E-02 | -8.66E-03 |
| 3324 | Usp11 | USP11 | -5.43E-02 | -2.62E-02 |
| 3325 | Ecm2 | ECM2 | -1.19E-01 | -2.48E-02 |
| 3326 | Spryd4 | SPRYD4 | 4.07E-02 | -8.51E-03 |
| 3327 | H2-Q10 | HLA-A | -1.16E-01 | 1.12E-01 |
| 3328 | Nudt21 | NUDT21 | -3.29E-02 | 3.63E-03 |
| 3329 | Rnpepl1 | RNPEPL1 | -2.34E-02 | -1.72E-02 |
| 3330 | Acp6 | ACP6 | 3.82E-02 | -1.39E-02 |
| 3331 | Emp1 | EMP1 | -5.31E-02 | 2.83E-02 |
| 3332 | Arhgef1 | ARHGEF1 | 2.01E-02 | -5.69E-03 |
| 3333 | Fam20c | FAM20C | -4.63E-02 | -5.31E-03 |
| 3334 | Zbtb14 | ZBTB14 | -4.47E-02 | 4.61E-03 |
| 3335 | Gata6 | GATA6 | 1.08E-01 | 2.45E-02 |
| 3336 | Ankib1 | ANKIB1 | -4.44E-02 | -2.57E-02 |

| | | | | |
|---|---|---|---|---|
| 3337 | Gabpb2 | GABPB2 | -2.75E-02 | -9.55E-03 |
| 3338 | Cox4i1 | COX4I1 | 3.02E-02 | -1.57E-02 |
| 3339 | Chuk | CHUK | 3.34E-02 | -4.37E-03 |
| 3340 | Sh3bp1 | SH3BP1 | -4.85E-02 | 7.30E-02 |
| 3341 | Pllp | PLLP | 8.22E-02 | -3.59E-02 |
| 3342 | Sp140 | SP140 | -8.94E-02 | 3.58E-02 |
| 3343 | Cab39l | CAB39L | 5.16E-02 | -1.86E-02 |
| 3344 | Ifit3 | IFIT3 | -1.43E-01 | 4.64E-02 |
| 3345 | D5Ertd579e | KIAA0232 | 2.49E-02 | -3.05E-03 |
| 3346 | Nsmf | NSMF | 2.71E-02 | 9.60E-03 |
| 3347 | Cst3 | CST3 | -2.08E-02 | 8.73E-03 |
| 3348 | Smarca4 | SMARCA4 | 2.38E-02 | -3.63E-03 |
| 3349 | Kmt2c | KMT2C | -2.99E-02 | -5.57E-03 |
| 3350 | Capn12 | CAPN12 | -6.86E-02 | -8.39E-03 |
| 3351 | Pnrc1 | PNRC1 | 7.76E-02 | 3.73E-02 |
| 3352 | Scamp5 | SCAMP5 | 3.28E-02 | -2.99E-03 |
| 3353 | Nostrin | NOSTRIN | 8.90E-02 | 6.36E-02 |
| 3354 | Gkap1 | GKAP1 | 3.57E-02 | -2.31E-02 |
| 3355 | Dnaaf3 | DNAAF3 | -9.57E-02 | 3.38E-02 |
| 3356 | Tfb1m | TFB1M | -5.10E-02 | 1.09E-02 |
| 3357 | Eif4h | EIF4H | 2.28E-02 | -1.12E-02 |
| 3358 | Limch1 | LIMCH1 | -5.72E-02 | -3.22E-02 |
| 3359 | Cftr | CFTR | 1.21E-01 | 3.63E-02 |
| 3360 | Spata17 | SPATA17 | -7.24E-02 | -4.64E-02 |
| 3361 | Kif1b | KIF1B | 2.57E-02 | -2.18E-02 |
| 3362 | Cul3 | CUL3 | 2.58E-02 | -9.76E-03 |
| 3363 | Zscan29 | ZSCAN29 | -3.21E-02 | -1.42E-02 |
| 3364 | Ska2 | SKA2 | -6.67E-02 | 1.19E-02 |
| 3365 | Sptlc1 | SPTLC1 | -2.27E-02 | -1.38E-02 |
| 3366 | Ngly1 | NGLY1 | 3.32E-02 | -4.33E-03 |
| 3367 | Flot2 | FLOT2 | 2.86E-02 | 8.56E-03 |
| 3368 | Arcn1 | ARCN1 | 2.45E-02 | -1.41E-02 |
| 3369 | Phyhipl | PHYHIPL | -3.26E-02 | -2.87E-02 |
| 3370 | Akap3 | AKAP3 | 2.76E-02 | -2.60E-02 |
| 3371 | Ccdc92 | CCDC92 | -6.25E-02 | 2.12E-02 |
| 3372 | Btg3 | BTG3 | 6.08E-02 | 4.20E-02 |
| 3373 | Slc25a19 | SLC25A19 | -3.41E-02 | -1.06E-02 |
| 3374 | Btc | BTC | 1.12E-01 | -6.92E-02 |
| 3375 | Ciita | CIITA | -1.82E-01 | 1.22E-01 |
| 3376 | Dhrs7 | DHRS7 | -6.13E-02 | 7.81E-03 |
| 3377 | Slc40a1 | SLC40A1 | -5.65E-02 | 8.32E-02 |
| 3378 | Chchd2 | CHCHD2 | 3.19E-02 | -5.58E-03 |
| 3379 | Man1a2 | MAN1A2 | -2.49E-02 | 1.09E-03 |
| 3380 | Prdm15 | PRDM15 | -5.37E-02 | 1.26E-02 |
| 3381 | Pglyrp2 | PGLYRP2 | -8.93E-02 | 1.05E-01 |
| 3382 | Ei24 | EI24 | 2.08E-02 | -1.82E-02 |
| 3383 | Ifi205 | IFI16 | -1.13E-01 | 7.01E-02 |
| 3384 | Snx25 | SNX25 | -4.94E-02 | -2.91E-02 |
| 3385 | Rrp7a | RRP7A | 2.06E-02 | -7.85E-04 |
| 3386 | Trappc6b | TRAPPC6B | 1.82E-02 | -1.52E-02 |
| 3387 | Etfdh | ETFDH | 3.95E-02 | -1.91E-02 |
| 3388 | Igsf9 | IGSF9 | -8.52E-02 | -3.14E-02 |
| 3389 | Ehbp1 | EHBP1 | 3.53E-02 | -2.86E-02 |

| | | | | |
|---|---|---|---|---|
| 3390 | Pcdh19 | PCDH19 | -1.50E-01 | -2.79E-02 |
| 3391 | Tra2b | TRA2B | -2.90E-02 | 2.67E-02 |
| 3392 | Glis2 | GLIS2 | -3.04E-02 | -2.73E-02 |
| 3393 | Syce1l | SYCE1L | 6.96E-02 | -5.03E-02 |
| 3394 | Rnf167 | RNF167 | -3.93E-02 | -3.21E-02 |
| 3395 | Ube2l6 | UBE2L6 | -1.10E-01 | 5.92E-02 |
| 3396 | BC048403 | C12orf66 | 4.30E-02 | -1.30E-03 |
| 3397 | Coq7 | COQ7 | 2.95E-02 | -1.39E-02 |
| 3398 | Maff | MAFF | 1.81E-01 | 1.58E-01 |
| 3399 | Myo1f | MYO1F | -1.66E-01 | 1.83E-01 |
| 3400 | Cbll1 | CBLL1 | -5.24E-02 | 1.52E-02 |
| 3401 | Mbtps1 | MBTPS1 | -2.40E-02 | -1.76E-02 |
| 3402 | Nop10 | NOP10 | 3.71E-02 | -1.70E-02 |
| 3403 | Rbm12 | RBM12 | -1.00E-01 | 7.13E-02 |
| 3404 | Oaz2 | OAZ2 | 3.02E-02 | -1.17E-02 |
| 3405 | Il4ra | IL4R | 4.27E-02 | 2.69E-02 |
| 3406 | C1qtnf7 | C1QTNF7 | -1.16E-01 | 2.65E-02 |
| 3407 | 1700017B05Rik | C15orf39 | -5.97E-02 | 4.68E-02 |
| 3408 | Car11 | CA11 | -1.19E-01 | 3.47E-02 |
| 3409 | Emc10 | EMC10 | -2.14E-02 | 2.17E-03 |
| 3410 | Ica1 | ICA1 | 6.30E-02 | 9.64E-04 |
| 3411 | Smurf1 | SMURF1 | 3.24E-02 | -8.09E-03 |
| 3412 | Paip1 | PAIP1 | 2.47E-02 | -4.68E-03 |
| 3413 | Dvl3 | DVL3 | 2.93E-02 | -1.28E-02 |
| 3414 | Mmd2 | MMD2 | -1.23E-01 | 9.81E-02 |
| 3415 | Gtf2f1 | GTF2F1 | 2.51E-02 | 6.28E-03 |
| 3416 | Fkbp3 | FKBP3 | 2.02E-02 | -5.92E-03 |
| 3417 | Itga10 | ITGA10 | -1.58E-01 | 5.91E-02 |
| 3418 | Deaf1 | DEAF1 | 3.56E-02 | -2.27E-02 |
| 3419 | Meaf6 | MEAF6 | 2.87E-02 | 1.60E-02 |
| 3420 | Git2 | GIT2 | -3.04E-02 | 9.58E-03 |
| 3421 | Galnt7 | GALNT7 | -5.56E-02 | -2.25E-03 |
| 3422 | Spta1 | SPTA1 | -7.89E-02 | -3.79E-03 |
| 3423 | Laptm5 | LAPTM5 | -1.04E-01 | 1.22E-01 |
| 3424 | Rev3l | REV3L | 2.76E-02 | -2.82E-03 |
| 3425 | Ttc36 | TTC36 | -3.83E-02 | -1.07E-02 |
| 3426 | Tanc2 | TANC2 | 5.65E-02 | -2.41E-02 |
| 3427 | Myrfl | MYRFL | -8.61E-02 | -1.44E-02 |
| 3428 | Dhx16 | DHX16 | -2.21E-02 | 7.28E-03 |
| 3429 | Gpr68 | GPR68 | -8.00E-02 | 4.68E-03 |
| 3430 | Lef1 | LEF1 | -1.35E-01 | -2.74E-02 |
| 3431 | Xylt2 | XYLT2 | -3.06E-02 | -4.26E-02 |
| 3432 | Stk11ip | STK11IP | 2.63E-02 | -1.70E-02 |
| 3433 | Rcan3 | RCAN3 | 3.12E-02 | -1.15E-02 |
| 3434 | Mki67 | MKI67 | 2.56E-01 | 1.80E-01 |
| 3435 | Tctn3 | TCTN3 | 3.47E-02 | -1.50E-02 |
| 3436 | Slc31a1 | SLC31A1 | 3.50E-02 | -7.95E-03 |
| 3437 | Ttc1 | TTC1 | 2.19E-02 | 5.82E-04 |
| 3438 | Cd74 | CD74 | -1.10E-01 | 1.01E-01 |
| 3439 | Inpp1 | INPP1 | -3.56E-02 | 6.86E-03 |
| 3440 | Ptpn22 | PTPN22 | 1.17E-01 | 5.11E-02 |
| 3441 | Bub3 | BUB3 | 2.25E-02 | 4.70E-03 |
| 3442 | Psmb1 | PSMB1 | 2.24E-02 | -3.47E-03 |

| | | | | |
|---|---|---|---|---|
| 3443 | L3mbtl2 | L3MBTL2 | 3.77E-02 | -2.96E-04 |
| 3444 | Ctu2 | CTU2 | 5.13E-02 | -2.55E-02 |
| 3445 | Coasy | COASY | -3.17E-02 | -3.83E-02 |
| 3446 | Slc25a53 | SLC25A53 | 7.04E-02 | -6.14E-02 |
| 3447 | Zfand2b | ZFAND2B | 3.63E-02 | -2.01E-02 |
| 3448 | Nop9 | NOP9 | 2.77E-02 | 1.19E-02 |
| 3449 | Mgam | MGAM | -4.96E-02 | -2.69E-02 |
| 3450 | Atp2a2 | ATP2A2 | 3.54E-02 | -1.63E-02 |
| 3451 | Rabep2 | RABEP2 | 4.37E-02 | 4.51E-03 |
| 3452 | Zfp667 | ZNF667 | -6.34E-02 | -9.73E-03 |
| 3453 | Proc | PROC | -4.59E-02 | -2.56E-02 |
| 3454 | Ralgapb | RALGAPB | 2.58E-02 | -1.81E-02 |
| 3455 | Ube2w | UBE2W | -2.64E-02 | 1.32E-02 |
| 3456 | Cyc1 | CYC1 | 3.09E-02 | -1.66E-02 |
| 3457 | Endod1 | ENDOD1 | 1.17E-01 | 1.88E-02 |
| 3458 | Gtf2ird1 | GTF2IRD1 | 3.09E-02 | -2.91E-02 |
| 3459 | Lamb1 | LAMB1 | -2.89E-02 | -6.26E-03 |
| 3460 | Smyd5 | SMYD5 | -4.43E-02 | -6.20E-03 |
| 3461 | Ccdc90b | CCDC90B | -2.86E-02 | -1.51E-02 |
| 3462 | Stat1 | STAT1 | -8.80E-02 | 1.75E-02 |
| 3463 | Mrps11 | MRPS11 | -2.88E-02 | -1.28E-02 |
| 3464 | Herc4 | HERC4 | -2.98E-02 | -1.70E-02 |
| 3465 | Tes | TES | 6.26E-02 | 3.32E-02 |
| 3466 | 0610030E20Rik | C2orf68 | -7.16E-02 | 2.53E-03 |
| 3467 | Hnrnph1 | HNRNPH1 | -2.90E-02 | -1.94E-03 |
| 3468 | Cyp1a1 | CYP1A1 | -1.59E-01 | -4.86E-03 |
| 3469 | Nudt18 | NUDT18 | 4.58E-02 | -5.06E-03 |
| 3470 | Dab1 | DAB1 | 1.91E-01 | -8.83E-02 |
| 3471 | Dpyd | DPYD | 7.99E-02 | -2.91E-02 |
| 3472 | Tmie | TMIE | 6.76E-02 | -2.54E-02 |
| 3473 | Prkd2 | PRKD2 | -5.51E-02 | 1.64E-02 |
| 3474 | Slc38a2 | SLC38A2 | 7.13E-02 | 9.45E-03 |
| 3475 | Car9 | CA9 | 1.02E-01 | -1.81E-02 |
| 3476 | Notum | NOTUM | -6.69E-02 | 1.15E-02 |
| 3477 | Dusp7 | DUSP7 | 3.68E-02 | -2.70E-02 |
| 3478 | Ddx21 | DDX21 | 2.41E-02 | 2.54E-02 |
| 3479 | Rab35 | RAB35 | 2.02E-02 | -2.94E-03 |
| 3480 | Plin5 | PLIN5 | 1.59E-01 | 4.54E-02 |
| 3481 | Pcsk9 | PCSK9 | -1.56E-01 | 7.31E-02 |
| 3482 | Scamp3 | SCAMP3 | 2.26E-02 | -1.95E-03 |
| 3483 | Eif4enif1 | EIF4ENIF1 | 1.89E-02 | -9.49E-04 |
| 3484 | Cyb561a3 | CYB561A3 | -3.20E-02 | -1.23E-02 |
| 3485 | Btf3l4 | BTF3L4 | 2.51E-02 | 8.76E-04 |
| 3486 | Impdh1 | IMPDH1 | 4.63E-02 | 1.67E-02 |
| 3487 | Tulp1 | TULP1 | 3.66E-02 | 6.64E-03 |
| 3488 | Aco2 | ACO2 | 3.45E-02 | -1.64E-02 |
| 3489 | Zc3hav1 | ZC3HAV1 | -3.89E-02 | 2.29E-02 |
| 3490 | Scrib | SCRIB | 2.43E-02 | -7.61E-03 |
| 3491 | Rbfox1 | RBFOX1 | -1.07E-01 | -3.48E-02 |
| 3492 | Ptbp1 | PTBP1 | -2.61E-02 | 5.79E-03 |
| 3493 | Akr1a1 | AKR1A1 | 4.83E-02 | -1.51E-02 |
| 3494 | Sfpq | SFPQ | -3.73E-02 | 3.05E-02 |
| 3495 | Cript | CRIPT | 2.81E-02 | 4.51E-03 |

| | | | | |
|---|---|---|---|---|
| 3496 | Slc7a9 | SLC7A9 | -6.84E-02 | -1.12E-02 |
| 3497 | Ascc3 | ASCC3 | 3.04E-02 | -6.64E-03 |
| 3498 | Cry2 | CRY2 | 7.50E-02 | -1.87E-02 |
| 3499 | Klk1b16 | KLK3 | 7.82E-02 | 6.11E-02 |
| 3500 | Adcy9 | ADCY9 | -5.68E-02 | -4.24E-02 |
| 3501 | Micu3 | MICU3 | 3.48E-02 | -8.57E-03 |
| 3502 | Arhgef16 | ARHGEF16 | 2.31E-02 | -2.23E-02 |
| 3503 | Idh3a | IDH3A | 3.10E-02 | -2.83E-03 |
| 3504 | Phtf2 | PHTF2 | 5.91E-02 | -9.04E-03 |
| 3505 | Zfp12 | ZNF12 | -3.48E-02 | -5.81E-03 |
| 3506 | Gpr156 | GPR156 | 7.62E-02 | -3.42E-02 |
| 3507 | Aatk | AATK | -8.06E-02 | -6.17E-02 |
| 3508 | Zbtb48 | ZBTB48 | -4.92E-02 | 5.24E-03 |
| 3509 | Bag5 | BAG5 | 2.84E-02 | 7.82E-03 |
| 3510 | Jam2 | JAM2 | -8.92E-02 | 9.81E-02 |
| 3511 | Ctc1 | CTC1 | 2.95E-02 | -1.49E-02 |
| 3512 | Apc2 | APC2 | -1.00E-01 | 4.19E-02 |
| 3513 | Scrn2 | SCRN2 | 4.63E-02 | -1.90E-02 |
| 3514 | Zfp84 | ZNF565 | -6.80E-02 | -5.13E-03 |
| 3515 | Gnl2 | GNL2 | 2.37E-02 | -4.83E-03 |
| 3516 | Exoc4 | EXOC4 | 3.64E-02 | -4.73E-02 |
| 3517 | Pola1 | POLA1 | -8.26E-02 | 1.03E-01 |
| 3518 | Pigu | PIGU | 2.00E-02 | -6.06E-04 |
| 3519 | Slc30a9 | SLC30A9 | 2.58E-02 | -1.13E-02 |
| 3520 | Zfp366 | ZNF366 | -8.90E-02 | -2.17E-02 |
| 3521 | Gbgt1 | GBGT1 | -8.53E-02 | -1.87E-03 |
| 3522 | Tmem255b | TMEM255B | -7.86E-02 | 2.62E-02 |
| 3523 | Apod | APOD | 1.70E-01 | 5.08E-02 |
| 3524 | Paip2b | PAIP2B | -3.71E-02 | -5.31E-03 |
| 3525 | Qpct | QPCT | -8.24E-02 | -5.15E-03 |
| 3526 | Zmym2 | ZMYM2 | -2.72E-02 | 7.81E-03 |
| 3527 | Ky | KY | -8.28E-02 | -2.92E-02 |
| 3528 | Tmem215 | TMEM215 | -8.17E-02 | -8.87E-03 |
| 3529 | Psmb7 | PSMB7 | 3.07E-02 | -1.02E-02 |
| 3530 | Steap1 | STEAP1 | 8.13E-02 | 6.14E-02 |
| 3531 | Mdh2 | MDH2 | 2.92E-02 | -1.49E-02 |
| 3532 | Ccdc181 | CCDC181 | -2.88E-02 | -8.59E-03 |
| 3533 | Mafb | MAFB | -7.49E-02 | 4.94E-02 |
| 3534 | Uqcrfs1 | UQCRFS1 | 3.03E-02 | -1.69E-02 |
| 3535 | Agmo | AGMO | 7.42E-02 | -2.89E-03 |
| 3536 | Med17 | MED17 | -3.03E-02 | 4.22E-05 |
| 3537 | Slc39a1 | SLC39A1 | 2.46E-02 | 1.48E-02 |
| 3538 | Stx3 | STX3 | 3.88E-02 | -7.42E-03 |
| 3539 | Spatc1l | SPATC1L | -5.26E-02 | 1.31E-02 |
| 3540 | Spats2 | SPATS2 | 4.71E-02 | 5.35E-03 |
| 3541 | Trabd | TRABD | 2.50E-02 | -4.46E-03 |
| 3542 | Flrt1 | FLRT1 | -7.06E-02 | -1.23E-02 |
| 3543 | Erbb4 | ERBB4 | -1.20E-01 | -5.56E-02 |
| 3544 | Slc13a1 | SLC13A1 | 7.20E-02 | 4.52E-02 |
| 3545 | Scaper | SCAPER | -3.80E-02 | -1.76E-02 |
| 3546 | Bcdin3d | BCDIN3D | -5.95E-02 | 2.80E-02 |
| 3547 | Otud6b | OTUD6B | 2.72E-02 | -1.22E-02 |
| 3548 | Il18rap | IL18RAP | 8.82E-02 | 4.82E-02 |

| | | | | |
|---|---|---|---|---|
| 3549 | Pappa2 | PAPPA2 | -1.65E-01 | -4.64E-02 |
| 3550 | Mob3b | MOB3B | -4.31E-02 | -3.04E-02 |
| 3551 | Dnttip2 | DNTTIP2 | -2.32E-02 | 1.05E-02 |
| 3552 | Cfdp1 | CFDP1 | -2.14E-02 | -5.93E-03 |
| 3553 | Runx3 | RUNX3 | -1.01E-01 | 2.71E-02 |
| 3554 | Cmtm7 | CMTM7 | -5.75E-02 | 2.38E-02 |
| 3555 | Dnajc11 | DNAJC11 | 2.47E-02 | -4.88E-03 |
| 3556 | Gja4 | GJA4 | 6.31E-02 | -1.15E-02 |
| 3557 | Msi1 | MSI1 | 8.29E-02 | -3.65E-02 |
| 3558 | Slc35e4 | SLC35E4 | 6.29E-02 | 1.02E-02 |
| 3559 | Nudt5 | NUDT5 | 2.74E-02 | -1.34E-02 |
| 3560 | Itpr1 | ITPR1 | 3.46E-02 | -1.72E-02 |
| 3561 | Gimap1 | GIMAP1 | -6.10E-02 | 1.40E-02 |
| 3562 | Slc43a1 | SLC43A1 | 1.21E-01 | -1.07E-01 |
| 3563 | Dcaf5 | DCAF5 | 1.97E-02 | -7.82E-03 |
| 3564 | Klhl30 | KLHL30 | 1.05E-01 | 4.12E-02 |
| 3565 | Slc3a1 | SLC3A1 | -3.64E-02 | -2.95E-02 |
| 3566 | Smad9 | SMAD9 | -1.20E-01 | 9.10E-02 |
| 3567 | Rccd1 | RCCD1 | 6.47E-02 | -1.74E-02 |
| 3568 | Rtcb | RTCB | 2.32E-02 | -8.96E-03 |
| 3569 | Tbrg1 | TBRG1 | 2.68E-02 | 4.85E-03 |
| 3570 | Ndufa8 | NDUFA8 | 3.04E-02 | -1.75E-02 |
| 3571 | Tfrc | TFRC | 8.93E-02 | -2.91E-02 |
| 3572 | Zcchc9 | ZCCHC9 | 2.78E-02 | 1.50E-02 |
| 3573 | Crhr2 | CRHR2 | -5.91E-02 | -1.47E-03 |
| 3574 | Gpd1 | GPD1 | 6.08E-02 | -6.22E-05 |
| 3575 | Psmb6 | PSMB6 | 2.77E-02 | -1.54E-02 |
| 3576 | Cyfip2 | CYFIP2 | 3.56E-02 | -6.43E-02 |
| 3577 | Hcfc1r1 | HCFC1R1 | 2.50E-02 | 6.07E-03 |
| 3578 | Cobll1 | COBLL1 | 2.90E-02 | -2.76E-02 |
| 3579 | Zfp512 | ZNF512 | -2.54E-02 | -1.01E-02 |
| 3580 | Psmc1 | PSMC1 | 2.31E-02 | -1.21E-02 |
| 3581 | Jade2 | JADE2 | 4.04E-02 | -4.29E-04 |
| 3582 | Dnajc8 | DNAJC8 | 2.32E-02 | 3.34E-03 |
| 3583 | Rnf214 | RNF214 | -3.09E-02 | -7.78E-03 |
| 3584 | Cerk | CERK | 2.50E-02 | -2.84E-03 |
| 3585 | Cox10 | COX10 | 3.28E-02 | -8.30E-03 |
| 3586 | Itpka | ITPKA | -6.94E-02 | 3.56E-02 |
| 3587 | Zfp780b | ZNF780B | -6.88E-02 | -1.29E-03 |
| 3588 | E2f6 | E2F6 | -2.67E-02 | 1.36E-02 |
| 3589 | Shox2 | SHOX2 | 4.94E-02 | -2.68E-02 |
| 3590 | Ints7 | INTS7 | 2.74E-02 | 4.08E-03 |
| 3591 | Fancb | FANCB | -1.08E-01 | 9.62E-02 |
| 3592 | Specc1l | SPECC1L | 2.17E-02 | -1.09E-02 |
| 3593 | Apba3 | APBA3 | -2.77E-02 | 1.38E-02 |
| 3594 | Slfn5 | SLFN5 | -5.92E-02 | -3.79E-02 |
| 3595 | Arel1 | AREL1 | 1.75E-02 | -1.60E-02 |
| 3596 | Carns1 | CARNS1 | -4.93E-02 | 2.25E-02 |
| 3597 | Bcl2l11 | BCL2L11 | 7.38E-02 | -2.72E-02 |
| 3598 | Commd2 | COMMD2 | 2.96E-02 | -9.06E-04 |
| 3599 | Dlg4 | DLG4 | -7.21E-02 | 6.04E-02 |
| 3600 | Sh3bp5 | SH3BP5 | 5.06E-02 | -1.52E-02 |
| 3601 | Fndc8 | FNDC8 | 5.33E-02 | -1.54E-02 |

| | | | | |
|---|---|---|---|---|
| 3602 | Rad51ap2 | RAD51AP2 | -1.48E-01 | -1.49E-01 |
| 3603 | Adhfe1 | ADHFE1 | -4.24E-02 | -9.65E-03 |
| 3604 | Adam15 | ADAM15 | 3.00E-02 | -1.18E-02 |
| 3605 | Phc1 | PHC1 | 2.30E-02 | -2.49E-02 |
| 3606 | Nampt | NAMPT | 3.92E-02 | 6.05E-03 |
| 3607 | Sod1 | SOD1 | -3.50E-02 | -5.27E-03 |
| 3608 | Cdc5l | CDC5L | -2.34E-02 | 8.13E-03 |
| 3609 | Dock9 | DOCK9 | -5.07E-02 | -4.59E-02 |
| 3610 | Ndufv1 | NDUFV1 | 2.97E-02 | -2.41E-02 |
| 3611 | Nek4 | NEK4 | -3.68E-02 | -1.50E-02 |
| 3612 | Hebp1 | HEBP1 | 5.90E-02 | -7.67E-03 |
| 3613 | Rapsn | RAPSN | -8.05E-02 | 5.46E-02 |
| 3614 | Nmt2 | NMT2 | 3.61E-02 | 2.02E-03 |
| 3615 | Lmod1 | LMOD1 | 1.23E-01 | -1.46E-02 |
| 3616 | Dnajc17 | DNAJC17 | 5.71E-02 | -2.39E-02 |
| 3617 | Prr15l | PRR15L | 2.81E-02 | -8.76E-03 |
| 3618 | B3galnt2 | B3GALNT2 | -3.93E-02 | 3.48E-02 |
| 3619 | Klhdc10 | KLHDC10 | 2.39E-02 | -1.32E-02 |
| 3620 | Ccz1 | CCZ1 | 2.34E-02 | 5.24E-03 |
| 3621 | Ccz1 | CCZ1B | 2.34E-02 | 5.24E-03 |
| 3622 | Tnni3 | TNNI3 | -1.04E-01 | 2.53E-02 |
| 3623 | Ctnna1 | CTNNA1 | 1.73E-02 | -1.83E-03 |
| 3624 | Dgkd | DGKD | 3.51E-02 | -2.19E-02 |
| 3625 | Cadm4 | CADM4 | 2.94E-02 | -2.51E-02 |
| 3626 | Slc39a2 | SLC39A2 | -1.14E-01 | 2.25E-02 |
| 3627 | Uimc1 | UIMC1 | 2.29E-02 | -6.65E-03 |
| 3628 | Slc29a2 | SLC29A2 | -6.72E-02 | 2.53E-02 |
| 3629 | Rxra | RXRA | 4.07E-02 | -7.81E-03 |
| 3630 | I830077J02Rik | C1orf162 | -9.08E-02 | 4.38E-02 |
| 3631 | 2700049A03Rik | KIAA0586 | -5.31E-02 | -6.37E-03 |
| 3632 | Dctd | DCTD | -1.22E-01 | 1.08E-01 |
| 3633 | Nr5a2 | NR5A2 | -1.03E-01 | 1.91E-02 |
| 3634 | Plekhm2 | PLEKHM2 | 5.65E-02 | -2.94E-02 |
| 3635 | Gpr18 | GPR18 | -7.83E-02 | 4.41E-02 |
| 3636 | Mtmr12 | MTMR12 | 3.77E-02 | -1.92E-02 |
| 3637 | Pkia | PKIA | -4.50E-02 | -4.55E-03 |
| 3638 | Armcx4 | ARMCX4 | 6.10E-02 | 3.82E-02 |
| 3639 | Zfp78 | ZNF471 | -8.86E-02 | -4.09E-02 |
| 3640 | Mzb1 | MZB1 | 1.16E-01 | -3.96E-02 |
| 3641 | Alg11 | ALG11 | -2.65E-02 | -4.27E-03 |
| 3642 | Arid1a | ARID1A | -2.06E-02 | 9.26E-04 |
| 3643 | Gpn2 | GPN2 | 4.22E-02 | -1.37E-02 |
| 3644 | Mcmdc2 | MCMDC2 | -8.96E-02 | -2.08E-02 |
| 3645 | Arhgap44 | ARHGAP44 | 3.14E-02 | -1.86E-02 |
| 3646 | Mtss1 | MTSS1 | -4.77E-02 | -2.13E-02 |
| 3647 | Nsdhl | NSDHL | 6.00E-02 | -1.85E-02 |
| 3648 | 0610040J01Rik | C4orf19 | 3.30E-02 | -1.24E-02 |
| 3649 | Pop7 | POP7 | 4.39E-02 | -8.87E-03 |
| 3650 | H2-K1 | HLA-A | -6.57E-02 | 5.36E-02 |
| 3651 | Sirpb1a | SIRPB1 | -9.64E-02 | 7.45E-02 |
| 3652 | Usp10 | USP10 | 3.51E-02 | -3.46E-03 |
| 3653 | Smad6 | SMAD6 | 4.60E-02 | 2.50E-02 |
| 3654 | Fgfrl1 | FGFRL1 | 4.29E-02 | -2.66E-02 |

| | | | | |
|---|---|---|---|---|
| 3655 | Gpr157 | GPR157 | 1.16E-01 | -5.59E-02 |
| 3656 | Ttc8 | TTC8 | -3.99E-02 | -1.11E-02 |
| 3657 | Parp12 | PARP12 | -3.81E-02 | 1.08E-03 |
| 3658 | Enpp6 | ENPP6 | 7.71E-02 | -4.02E-03 |
| 3659 | Plekhg5 | PLEKHG5 | 4.69E-02 | 2.44E-02 |
| 3660 | Chdh | CHDH | 3.53E-02 | -2.56E-02 |
| 3661 | Pipox | PIPOX | 4.78E-02 | -2.32E-02 |
| 3662 | Ppid | PPID | -3.63E-02 | -5.96E-03 |
| 3663 | Nosip | NOSIP | 2.12E-02 | -4.30E-03 |
| 3664 | Tyw3 | TYW3 | -1.05E-01 | -6.28E-02 |
| 3665 | Lipt2 | LIPT2 | 7.44E-02 | -1.75E-02 |
| 3666 | Mbp | MBP | 8.73E-02 | 1.50E-02 |
| 3667 | Vps37d | VPS37D | 7.05E-02 | -2.05E-02 |
| 3668 | Gabrg3 | GABRG3 | -4.25E-02 | -4.48E-03 |
| 3669 | Slc26a8 | SLC26A8 | 1.04E-01 | 2.66E-03 |
| 3670 | Epha6 | EPHA6 | 6.62E-02 | 1.23E-02 |
| 3671 | Pbk | PBK | 2.41E-01 | 2.11E-01 |
| 3672 | Zmym1 | ZMYM1 | -6.73E-02 | 7.41E-03 |
| 3673 | Bcl2l13 | BCL2L13 | 3.39E-02 | -8.67E-03 |
| 3674 | Clec7a | CLEC7A | -1.35E-01 | 1.46E-01 |
| 3675 | Vars2 | VARS2 | -3.14E-02 | -1.87E-02 |
| 3676 | Rpa2 | RPA2 | 5.53E-02 | 2.71E-02 |
| 3677 | Xpo6 | XPO6 | 1.88E-02 | -4.74E-03 |
| 3678 | Vti1a | VTI1A | 2.82E-02 | -1.41E-02 |
| 3679 | Erich2 | ERICH2 | -7.66E-02 | -2.06E-02 |
| 3680 | Khnyn | KHNYN | -3.06E-02 | -2.09E-02 |
| 3681 | Pmp22 | PMP22 | -3.65E-02 | 5.71E-02 |
| 3682 | Rab1b | RAB1B | 2.39E-02 | -1.63E-02 |
| 3683 | Fbxo46 | FBXO46 | -4.84E-02 | -1.36E-02 |
| 3684 | Izumo4 | IZUMO4 | -4.51E-02 | 7.61E-03 |
| 3685 | Bend6 | BEND6 | -5.51E-02 | -1.55E-02 |
| 3686 | Rims2 | RIMS2 | -1.21E-01 | 2.09E-02 |
| 3687 | Txnrd2 | TXNRD2 | -4.12E-02 | -4.22E-02 |
| 3688 | Psen1 | PSEN1 | -2.42E-02 | -6.84E-03 |
| 3689 | Foxs1 | FOXS1 | 1.19E-01 | -1.81E-02 |
| 3690 | Dek | DEK | 2.48E-02 | 8.63E-03 |
| 3691 | Hoga1 | HOGA1 | 4.17E-02 | -1.03E-02 |
| 3692 | Prdx5 | PRDX5 | 2.81E-02 | -1.63E-02 |
| 3693 | Uba2 | UBA2 | 1.72E-02 | -6.11E-03 |
| 3694 | Pnpla7 | PNPLA7 | -4.95E-02 | -6.28E-02 |
| 3695 | Ssr3 | SSR3 | 2.38E-02 | -2.36E-03 |
| 3696 | Nek1 | NEK1 | -4.43E-02 | -6.42E-03 |
| 3697 | Ndufa10 | NDUFA10 | 2.98E-02 | -1.80E-02 |
| 3698 | Apex1 | APEX1 | 4.42E-02 | 1.85E-02 |
| 3699 | Srebf2 | SREBF2 | -3.66E-02 | -2.56E-02 |
| 3700 | Nhlrc2 | NHLRC2 | -3.21E-02 | -1.69E-02 |
| 3701 | Prokr1 | PROKR1 | -3.85E-02 | -2.75E-04 |
| 3702 | Ep300 | EP300 | -2.77E-02 | -6.30E-03 |
| 3703 | Col20a1 | COL20A1 | -8.30E-02 | -1.06E-02 |
| 3704 | Dpm3 | DPM3 | -3.21E-02 | -7.85E-03 |
| 3705 | Ube2f | UBE2F | 3.81E-02 | 1.00E-02 |
| 3706 | Prex2 | PREX2 | -9.90E-02 | -3.27E-02 |
| 3707 | Atp6ap2 | ATP6AP2 | 2.34E-02 | -2.49E-03 |

| | | | | |
|---|---|---|---|---|
| 3708 | Nup54 | NUP54 | -3.02E-02 | 8.15E-03 |
| 3709 | Esrrb | ESRRB | 8.40E-02 | -6.37E-02 |
| 3710 | Cnnm2 | CNNM2 | 3.17E-02 | 8.58E-03 |
| 3711 | Map4k2 | MAP4K2 | -2.76E-02 | -2.13E-02 |
| 3712 | Aimp2 | AIMP2 | 3.37E-02 | 2.86E-03 |
| 3713 | Dok3 | DOK3 | -1.08E-01 | 5.96E-02 |
| 3714 | Ube2v2 | UBE2V2 | -4.05E-02 | -1.69E-02 |
| 3715 | Crcp | CRCP | 2.97E-02 | -9.44E-03 |
| 3716 | Plag1 | PLAG1 | -8.93E-02 | -6.15E-02 |
| 3717 | Lair1 | LAIR1 | -1.35E-01 | 1.25E-01 |
| 3718 | Stxbp2 | STXBP2 | 2.34E-02 | -6.25E-03 |
| 3719 | Thy1 | THY1 | -1.50E-01 | 1.70E-01 |
| 3720 | Zfp617 | ZNF878 | 3.49E-02 | -1.11E-02 |
| 3721 | Clec4a2 | CLEC4A | -9.21E-02 | 2.05E-01 |
| 3722 | Slc6a5 | SLC6A5 | 4.89E-02 | -1.89E-02 |
| 3723 | S1pr3 | S1PR3 | 6.88E-02 | -1.77E-02 |
| 3724 | Spata7 | SPATA7 | 5.25E-02 | 2.06E-03 |
| 3725 | Tmem14c | TMEM14C | 2.64E-02 | -1.61E-03 |
| 3726 | Ncr1 | NCR1 | -6.38E-02 | -2.07E-02 |
| 3727 | Trpc6 | TRPC6 | 7.54E-02 | 1.18E-02 |
| 3728 | Cd7 | CD7 | -5.97E-02 | -5.20E-03 |
| 3729 | Aph1b | APH1B | 3.35E-02 | -2.37E-02 |
| 3730 | Pign | PIGN | -2.91E-02 | 3.26E-03 |
| 3731 | Fbf1 | FBF1 | 3.81E-02 | -6.95E-03 |
| 3732 | Ift74 | IFT74 | 3.51E-02 | 1.01E-03 |
| 3733 | Gimap5 | GIMAP1-GIMAP5 | -6.80E-02 | -2.76E-02 |
| 3734 | Snx3 | SNX3 | 2.22E-02 | -1.05E-02 |
| 3735 | Spon1 | SPON1 | 1.54E-01 | 5.79E-03 |
| 3736 | 1700067K01Rik | C19orf67 | 1.16E-01 | 4.79E-02 |
| 3737 | Pole2 | POLE2 | -8.75E-02 | 4.04E-02 |
| 3738 | Cradd | CRADD | -4.23E-02 | -1.36E-02 |
| 3739 | Hbp1 | HBP1 | 3.83E-02 | -2.38E-02 |
| 3740 | Fut4 | FUT4 | -1.13E-01 | 1.67E-02 |
| 3741 | Ceacam2 | CEACAM1 | -1.56E-01 | -1.86E-01 |
| 3742 | Casp14 | CASP14 | 5.09E-02 | -1.20E-04 |
| 3743 | Bak1 | BAK1 | 5.48E-02 | 5.21E-02 |
| 3744 | Fcrl1 | FCRL1 | -6.75E-02 | 1.23E-02 |
| 3745 | Dhx58 | DHX58 | -8.93E-02 | 1.87E-02 |
| 3746 | Wdr91 | WDR91 | 6.00E-02 | -9.76E-03 |
| 3747 | Pcgf2 | PCGF2 | 2.78E-02 | -9.37E-03 |
| 3748 | Slc25a38 | SLC25A38 | -3.44E-02 | -3.64E-03 |
| 3749 | Immt | IMMT | 2.74E-02 | -2.31E-02 |
| 3750 | Tmprss4 | TMPRSS4 | 1.10E-01 | 2.79E-02 |
| 3751 | Shisa9 | SHISA9 | 8.25E-02 | -4.78E-02 |
| 3752 | Ndufa2 | NDUFA2 | 2.98E-02 | -1.14E-02 |
| 3753 | Cspp1 | CSPP1 | -2.80E-02 | 1.78E-02 |
| 3754 | Zfp369 | ZNF274 | -4.17E-02 | 1.88E-02 |
| 3755 | Mpp7 | MPP7 | 5.01E-02 | -4.77E-02 |
| 3756 | Taf7 | TAF7 | -6.34E-02 | -1.16E-02 |
| 3757 | Vps29 | VPS29 | 2.97E-02 | 1.71E-03 |
| 3758 | Mansc4 | MANSC4 | 1.28E-01 | -5.70E-02 |
| 3759 | Col4a1 | COL4A1 | 5.44E-02 | 2.35E-02 |
| 3760 | Glipr1 | GLIPR1 | -9.56E-02 | 4.32E-02 |

| | | | | |
|---|---|---|---|---|
| 3761 | Tll1 | TLL1 | -1.14E-01 | -3.86E-02 |
| 3762 | Dph6 | DPH6 | 3.81E-02 | -5.55E-03 |
| 3763 | Capn3 | CAPN3 | 1.00E-01 | -3.44E-02 |
| 3764 | Mbnl1 | MBNL1 | 3.49E-02 | -1.11E-02 |
| 3765 | Kcnk6 | KCNK6 | -7.77E-02 | 9.22E-02 |
| 3766 | Hes6 | HES6 | 5.09E-02 | -3.48E-02 |
| 3767 | Ssh2 | SSH2 | 4.03E-02 | 2.13E-02 |
| 3768 | Zfp420 | ZNF420 | -5.59E-02 | -9.48E-03 |
| 3769 | Fads2 | FADS2 | -4.70E-02 | -1.60E-02 |
| 3770 | Angel2 | ANGEL2 | -2.51E-02 | -6.34E-03 |
| 3771 | Tob2 | TOB2 | 1.00E-01 | -1.99E-02 |
| 3772 | Kif5b | KIF5B | 2.30E-02 | -9.26E-03 |
| 3773 | Mmab | MMAB | -3.34E-02 | -2.87E-02 |
| 3774 | Zfp92 | ZFP92 | 4.52E-02 | 4.98E-02 |
| 3775 | Fermt3 | FERMT3 | -1.13E-01 | 1.41E-01 |
| 3776 | Ndufs4 | NDUFS4 | 2.52E-02 | -9.58E-03 |
| 3777 | Sorbs3 | SORBS3 | 3.12E-02 | -8.23E-03 |
| 3778 | Pycr2 | PYCR2 | 3.47E-02 | -1.82E-03 |
| 3779 | Aagab | AAGAB | 2.50E-02 | -7.68E-03 |
| 3780 | Slc9a3 | SLC9A3 | 3.76E-02 | -5.28E-02 |
| 3781 | Ythdc1 | YTHDC1 | -2.69E-02 | -2.20E-03 |
| 3782 | Pde6a | PDE6A | -1.73E-01 | -1.21E-01 |
| 3783 | Copb2 | COPB2 | 2.30E-02 | -1.66E-02 |
| 3784 | Obp2a | OBP2B | 5.64E-02 | 1.53E-02 |
| 3785 | Ppp1r16b | PPP1R16B | 7.85E-02 | -2.67E-02 |
| 3786 | Stat5a | STAT5A | 4.88E-02 | -6.22E-03 |
| 3787 | Rpl22 | RPL22 | -2.21E-02 | 1.48E-02 |
| 3788 | Eif5a | EIF5A | 2.75E-02 | 2.07E-03 |
| 3789 | Il15 | IL15 | -5.97E-02 | -3.98E-02 |
| 3790 | Adam9 | ADAM9 | 2.81E-02 | -9.51E-03 |
| 3791 | Synpo2 | SYNPO2 | -1.02E-01 | -4.86E-02 |
| 3792 | Atp6v0a4 | ATP6V0A4 | 3.43E-02 | -8.92E-03 |
| 3793 | Emc8 | EMC8 | 2.31E-02 | -2.93E-02 |
| 3794 | Usp7 | USP7 | 1.88E-02 | -1.07E-02 |
| 3795 | Denr | DENR | 2.54E-02 | -9.96E-03 |
| 3796 | Amdhd1 | AMDHD1 | -1.61E-01 | -8.05E-02 |
| 3797 | Clpx | CLPX | 4.15E-02 | -1.04E-02 |
| 3798 | Cntn2 | CNTN2 | 5.59E-02 | -1.73E-02 |
| 3799 | Zfp131 | ZNF131 | 4.56E-02 | 7.72E-03 |
| 3800 | Pnmal2 | PNMAL2 | -7.81E-02 | -3.65E-02 |
| 3801 | Pcdh9 | PCDH9 | 1.30E-01 | 3.82E-02 |
| 3802 | Mrpl40 | MRPL40 | 2.52E-02 | -8.46E-04 |
| 3803 | Snrnp48 | SNRNP48 | -3.08E-02 | 5.25E-03 |
| 3804 | Tet2 | TET2 | -4.38E-02 | -1.24E-02 |
| 3805 | Zswim4 | ZSWIM4 | 5.93E-02 | -1.91E-02 |
| 3806 | Cdk20 | CDK20 | -6.46E-02 | 3.66E-02 |
| 3807 | Atp5g1 | ATP5G1 | 2.53E-02 | -1.23E-02 |
| 3808 | Ntan1 | NTAN1 | 2.61E-02 | -6.14E-03 |
| 3809 | Prelp | PRELP | -1.24E-01 | -1.66E-02 |
| 3810 | Pex1 | PEX1 | 3.49E-02 | -1.11E-02 |
| 3811 | Omd | OMD | 1.23E-01 | -6.70E-02 |
| 3812 | Ggh | GGH | 2.19E-02 | 3.68E-03 |
| 3813 | Mchr1 | MCHR1 | 2.22E-01 | 3.04E-02 |

| | | | | |
|---|---|---|---|---|
| 3814 | Slc2a13 | SLC2A13 | -1.05E-01 | -7.31E-02 |
| 3815 | Fbxw2 | FBXW2 | 1.80E-02 | 6.36E-03 |
| 3816 | Ttc3 | TTC3 | -2.22E-02 | -1.25E-02 |
| 3817 | Afap1l1 | AFAP1L1 | -4.21E-02 | -3.32E-02 |
| 3818 | Brpf3 | BRPF3 | 2.85E-02 | -1.35E-02 |
| 3819 | 4931406C07Rik | C11orf54 | 4.23E-02 | -1.59E-02 |
| 3820 | Cntnap1 | CNTNAP1 | -7.38E-02 | 3.86E-03 |
| 3821 | Dpagt1 | DPAGT1 | 2.27E-02 | -1.15E-02 |
| 3822 | Vps45 | VPS45 | 3.13E-02 | -5.79E-04 |
| 3823 | C4b | C4B | 1.96E-01 | 2.18E-01 |
| 3824 | C4b | C4A | 1.96E-01 | 2.18E-01 |
| 3825 | D130043K22Rik | KIAA0319 | -8.83E-02 | 2.73E-02 |
| 3826 | Tefm | TEFM | -3.28E-02 | -4.34E-03 |
| 3827 | Ccdc81 | CCDC81 | 3.44E-02 | -2.36E-02 |
| 3828 | Pold2 | POLD2 | -4.18E-02 | 9.96E-03 |
| 3829 | Otud3 | OTUD3 | 4.18E-02 | 8.22E-03 |
| 3830 | Adamts13 | ADAMTS13 | -1.42E-01 | -1.44E-02 |
| 3831 | Ttc32 | TTC32 | -3.39E-02 | -8.51E-03 |
| 3832 | Cd79b | CD79B | -1.77E-01 | -5.55E-03 |
| 3833 | Pole3 | POLE3 | 2.33E-02 | 9.14E-04 |
| 3834 | Adamts9 | ADAMTS9 | 4.56E-02 | -1.37E-02 |
| 3835 | Rassf1 | RASSF1 | 2.90E-02 | 1.51E-03 |
| 3836 | Pms1 | PMS1 | -5.45E-02 | 2.41E-02 |
| 3837 | Sfi1 | SFI1 | -4.49E-02 | 1.65E-02 |
| 3838 | Pdcd6 | PDCD6 | 2.03E-02 | -1.98E-03 |
| 3839 | Gdi2 | GDI2 | 2.54E-02 | -4.41E-03 |
| 3840 | Pdzd11 | PDZD11 | 3.16E-02 | -2.11E-02 |
| 3841 | C1d | C1D | 2.84E-02 | -1.52E-02 |
| 3842 | Chchd1 | CHCHD1 | 2.73E-02 | -1.66E-02 |
| 3843 | Ntn1 | NTN1 | -2.92E-02 | -1.47E-02 |
| 3844 | Slc5a2 | SLC5A2 | 6.38E-02 | -3.91E-02 |
| 3845 | Acp1 | ACP1 | 2.19E-02 | -1.57E-02 |
| 3846 | Pip5k1b | PIP5K1B | 7.95E-02 | 5.50E-03 |
| 3847 | Fbxl17 | FBXL17 | 2.95E-02 | -2.33E-02 |
| 3848 | Wbp11 | WBP11 | 1.79E-02 | -2.61E-03 |
| 3849 | Cr1l | CR1L | -1.78E-02 | 2.49E-03 |
| 3850 | Cnnm3 | CNNM3 | -2.38E-02 | -1.55E-02 |
| 3851 | Slc1a4 | SLC1A4 | -6.11E-02 | 2.37E-02 |
| 3852 | Dtx4 | DTX4 | 8.83E-02 | 5.46E-02 |
| 3853 | Nphp3 | NPHP3 | 4.64E-02 | -2.98E-02 |
| 3854 | Rhot1 | RHOT1 | 2.35E-02 | -2.95E-03 |
| 3855 | Ptprj | PTPRJ | 2.40E-02 | -3.88E-03 |
| 3856 | Ephx2 | EPHX2 | 6.18E-02 | -1.20E-02 |
| 3857 | Kank1 | KANK1 | -2.92E-02 | -5.92E-03 |
| 3858 | Slc25a25 | SLC25A25 | 2.50E-01 | 1.76E-02 |
| 3859 | Dgkk | DGKK | 3.78E-02 | -1.85E-02 |
| 3860 | Sp4 | SP4 | -7.16E-02 | -1.49E-02 |
| 3861 | Cd109 | CD109 | -1.10E-01 | 1.11E-01 |
| 3862 | Nufip1 | NUFIP1 | 3.76E-02 | 1.99E-02 |
| 3863 | Pdcl | PDCL | 2.42E-02 | -1.44E-02 |
| 3864 | Zfp275 | ZNF275 | -5.38E-02 | -7.98E-03 |
| 3865 | Phf10 | PHF10 | 3.13E-02 | -4.35E-03 |
| 3866 | P2ry14 | P2RY14 | 1.93E-01 | 4.18E-02 |

| | | | | |
|---|---|---|---|---|
| 3867 | Degs1 | DEGS1 | 3.14E-02 | -5.57E-03 |
| 3868 | Gdf3 | GDF3 | -5.62E-02 | -1.19E-02 |
| 3869 | Adam23 | ADAM23 | 1.25E-01 | 1.59E-01 |
| 3870 | Fer1l5 | FER1L5 | -3.59E-02 | 6.72E-03 |
| 3871 | Wdr45 | WDR45 | -3.55E-02 | -1.66E-02 |
| 3872 | Nqo2 | NQO2 | -3.15E-02 | -9.96E-03 |
| 3873 | Helb | HELB | 4.90E-02 | -1.55E-02 |
| 3874 | Cfl2 | CFL2 | 2.77E-02 | 5.50E-04 |
| 3875 | Lyve1 | LYVE1 | 1.08E-01 | 5.82E-02 |
| 3876 | Nek5 | NEK5 | -1.15E-01 | -2.01E-02 |
| 3877 | Zfp583 | ZNF583 | -1.03E-01 | -1.82E-02 |
| 3878 | Nubpl | NUBPL | 4.09E-02 | -1.61E-02 |
| 3879 | Tbc1d9 | TBC1D9 | 1.39E-01 | -1.09E-02 |
| 3880 | Ywhae | YWHAE | 2.12E-02 | -4.39E-03 |
| 3881 | Timeless | TIMELESS | -6.81E-02 | 7.82E-02 |
| 3882 | Pik3cb | PIK3CB | -2.94E-02 | -2.87E-02 |
| 3883 | Rae1 | RAE1 | 2.10E-02 | -4.02E-03 |
| 3884 | Rtn3 | RTN3 | 1.95E-02 | -6.66E-03 |
| 3885 | Wt1 | WT1 | -6.91E-02 | -4.85E-02 |
| 3886 | Ide | IDE | 3.78E-02 | -1.95E-02 |
| 3887 | Lgi2 | LGI2 | 1.69E-01 | 3.12E-01 |
| 3888 | Fads3 | FADS3 | -3.49E-02 | -2.27E-02 |
| 3889 | 4930550C14Rik | C11orf65 | 7.77E-02 | 2.43E-02 |
| 3890 | Psmd2 | PSMD2 | 2.24E-02 | -6.40E-03 |
| 3891 | Fahd2a | FAHD2B | 4.11E-02 | -9.82E-03 |
| 3892 | Arl1 | ARL1 | 1.96E-02 | -3.42E-03 |
| 3893 | Oxr1 | OXR1 | -3.27E-02 | 5.52E-03 |
| 3894 | Tsnaxip1 | TSNAXIP1 | 4.24E-02 | -1.29E-02 |
| 3895 | Capn5 | CAPN5 | 6.88E-02 | 3.39E-02 |
| 3896 | Cog6 | COG6 | 2.08E-02 | -1.59E-02 |
| 3897 | Lrrk2 | LRRK2 | -3.67E-02 | -2.88E-02 |
| 3898 | Nsun3 | NSUN3 | 3.02E-02 | -2.64E-02 |
| 3899 | Ccdc73 | CCDC73 | -7.83E-02 | 3.14E-02 |
| 3900 | Saysd1 | SAYSD1 | 4.56E-02 | -3.47E-03 |
| 3901 | Rabgef1 | RABGEF1 | 4.04E-02 | 1.24E-02 |
| 3902 | Mdc1 | MDC1 | -2.75E-02 | 1.45E-02 |
| 3903 | Foxq1 | FOXQ1 | -1.37E-01 | -1.82E-02 |
| 3904 | Snx17 | SNX17 | 2.28E-02 | -9.83E-03 |
| 3905 | Slc25a14 | SLC25A14 | -5.62E-02 | 1.69E-02 |
| 3906 | Slc45a2 | SLC45A2 | -5.97E-02 | 5.33E-03 |
| 3907 | Ndor1 | NDOR1 | 3.97E-02 | -3.41E-02 |
| 3908 | Oaz1 | OAZ1 | 2.48E-02 | -1.51E-02 |
| 3909 | Cdc23 | CDC23 | -2.46E-02 | 3.94E-03 |
| 3910 | Chmp1a | CHMP1A | 2.41E-02 | -4.50E-03 |
| 3911 | Cox5a | COX5A | 2.33E-02 | 2.04E-03 |
| 3912 | Poldip2 | POLDIP2 | 2.69E-02 | -2.02E-02 |
| 3913 | Rhoj | RHOJ | 6.83E-02 | -2.57E-02 |
| 3914 | Abcb1a | ABCB1 | -1.24E-01 | -4.65E-02 |
| 3915 | Itga4 | ITGA4 | -1.55E-01 | 1.25E-01 |
| 3916 | Serac1 | SERAC1 | -4.18E-02 | -1.19E-02 |
| 3917 | Lrp3 | LRP3 | -5.39E-02 | 1.34E-02 |
| 3918 | Ccr7 | CCR7 | -6.78E-02 | -8.24E-03 |
| 3919 | Neurl4 | NEURL4 | -2.54E-02 | -5.79E-03 |

| | | | | |
|---|---|---|---|---|
| 3920 | Itgal | ITGAL | -2.08E-01 | 1.50E-01 |
| 3921 | St7 | ST7 | 3.65E-02 | -3.12E-02 |
| 3922 | Tmtc4 | TMTC4 | -3.94E-02 | 3.97E-03 |
| 3923 | Acacb | ACACB | 4.90E-02 | -3.28E-02 |
| 3924 | Cdc42ep3 | CDC42EP3 | -9.35E-02 | -1.35E-02 |
| 3925 | Cbx6 | CBX6 | -1.91E-02 | -1.09E-02 |
| 3926 | Slc46a3 | SLC46A3 | -7.57E-02 | 1.11E-02 |
| 3927 | Mrrf | MRRF | -3.05E-02 | -1.70E-02 |
| 3928 | Il6st | IL6ST | -2.06E-02 | 1.69E-03 |
| 3929 | Gbp11 | GBP6 | -9.73E-02 | 1.75E-02 |
| 3930 | Rnf32 | RNF32 | -6.63E-02 | 4.31E-02 |
| 3931 | Cdc16 | CDC16 | 2.29E-02 | 3.67E-04 |
| 3932 | Ube2q2 | UBE2Q2 | 2.29E-02 | -2.78E-02 |
| 3933 | Tsr1 | TSR1 | 3.26E-02 | 1.55E-02 |
| 3934 | Lcat | LCAT | -1.12E-01 | 1.04E-01 |
| 3935 | Pomgnt2 | POMGNT2 | -3.93E-02 | 1.83E-02 |
| 3936 | Zfp277 | ZNF277 | 4.20E-02 | 2.07E-03 |
| 3937 | 1700029I15Rik | C11orf94 | 7.70E-02 | -4.03E-02 |
| 3938 | Il3ra | IL3RA | -9.57E-02 | 8.51E-02 |
| 3939 | Hey2 | HEY2 | -8.86E-02 | 2.01E-02 |
| 3940 | Uqcc2 | UQCC2 | 3.68E-02 | -6.65E-03 |
| 3941 | Lhpp | LHPP | -6.25E-02 | 3.10E-02 |
| 3942 | Oma1 | OMA1 | -3.04E-02 | -1.51E-02 |
| 3943 | Tdrp | TDRP | -2.84E-02 | 1.54E-02 |
| 3944 | Irgm2 | IRGM | -1.69E-01 | 2.62E-02 |
| 3945 | Hdac9 | HDAC9 | -8.68E-02 | 7.18E-02 |
| 3946 | Sdhd | SDHD | 2.73E-02 | -1.27E-02 |
| 3947 | Tinag | TINAG | -4.19E-02 | -2.24E-02 |
| 3948 | Tgfbr3 | TGFBR3 | -6.46E-02 | -6.07E-02 |
| 3949 | Hsdl1 | HSDL1 | -2.51E-02 | -1.33E-03 |
| 3950 | Irak1 | IRAK1 | 2.92E-02 | -1.33E-02 |
| 3951 | 4930503L19Rik | C18orf54 | -6.35E-02 | 4.95E-02 |
| 3952 | Ercc1 | ERCC1 | 4.50E-02 | -4.22E-03 |
| 3953 | Grina | GRINA | 4.06E-02 | -7.31E-04 |
| 3954 | Stpg1 | STPG1 | -7.19E-02 | -7.59E-02 |
| 3955 | Notch1 | NOTCH1 | -3.49E-02 | -1.85E-03 |
| 3956 | Zfp639 | ZNF639 | 2.62E-02 | -8.55E-03 |
| 3957 | Tbx21 | TBX21 | -6.74E-02 | 4.35E-03 |
| 3958 | Btk | BTK | -8.47E-02 | 9.28E-02 |
| 3959 | Mepce | MEPCE | -3.05E-02 | 4.16E-04 |
| 3960 | Spg7 | SPG7 | 2.61E-02 | -1.28E-02 |
| 3961 | Wdr43 | WDR43 | 2.62E-02 | 1.36E-02 |
| 3962 | Mblac2 | MBLAC2 | -5.52E-02 | -9.88E-03 |
| 3963 | 4930579G24Rik | C4orf46 | 9.67E-02 | 9.29E-02 |
| 3964 | Snf8 | SNF8 | 2.28E-02 | -5.59E-03 |
| 3965 | Idi1 | IDI1 | -1.07E-01 | 2.59E-02 |
| 3966 | Tjp1 | TJP1 | -2.89E-02 | 1.39E-04 |
| 3967 | Plekhh1 | PLEKHH1 | -2.97E-02 | 1.84E-02 |
| 3968 | Zfp612 | ZNF23 | -4.47E-02 | -5.34E-03 |
| 3969 | Col15a1 | COL15A1 | -1.43E-01 | 1.35E-01 |
| 3970 | Pusl1 | PUSL1 | 4.34E-02 | 6.75E-03 |
| 3971 | Ccnb2 | CCNB2 | 2.68E-01 | 1.77E-01 |
| 3972 | Npm1 | NPM1 | -3.19E-02 | 3.40E-02 |

| | | | | |
|---|---|---|---|---|
| 3973 | Sh2d5 | SH2D5 | 9.85E-02 | 2.96E-02 |
| 3974 | Foxo1 | FOXO1 | 4.16E-02 | -6.80E-03 |
| 3975 | Kank3 | KANK3 | 5.09E-02 | 2.94E-03 |
| 3976 | Tomm70a | TOMM70A | 2.75E-02 | 6.55E-03 |
| 3977 | Ptch2 | PTCH2 | 8.81E-02 | 4.94E-04 |
| 3978 | Agpat5 | AGPAT5 | 2.74E-02 | -1.34E-02 |
| 3979 | Myof | MYOF | 4.12E-02 | 1.40E-02 |
| 3980 | Clcn6 | CLCN6 | 4.68E-02 | -3.41E-02 |
| 3981 | Pik3cg | PIK3CG | -1.03E-01 | 7.55E-02 |
| 3982 | Asb8 | ASB8 | 2.30E-02 | -1.12E-02 |
| 3983 | Kifc1 | KIFC1 | 1.67E-01 | 9.06E-02 |
| 3984 | Vps35 | VPS35 | 2.83E-02 | -1.40E-02 |
| 3985 | Ankrd46 | ANKRD46 | -2.09E-02 | -1.59E-03 |
| 3986 | N4bp2l2 | N4BP2L2 | -2.52E-02 | 3.29E-03 |
| 3987 | Dlat | DLAT | 2.33E-02 | -1.31E-02 |
| 3988 | Zmat1 | ZMAT1 | -4.96E-02 | -2.46E-02 |
| 3989 | Aamdc | AAMDC | -3.31E-02 | -1.24E-02 |
| 3990 | H3f3b | H3F3A | 4.55E-02 | -1.04E-02 |
| 3991 | H3f3b | H3F3B | 4.55E-02 | -1.04E-02 |
| 3992 | Atp5a1 | ATP5A1 | 2.85E-02 | -2.07E-02 |
| 3993 | Romo1 | ROMO1 | 2.98E-02 | -8.92E-03 |
| 3994 | Vmac | VMAC | -3.04E-02 | -2.14E-02 |
| 3995 | Yars | YARS | 2.57E-02 | -9.14E-03 |
| 3996 | Slco2a1 | SLCO2A1 | -4.48E-02 | -2.23E-02 |
| 3997 | Auts2 | AUTS2 | 1.15E-01 | -2.46E-02 |
| 3998 | Rabepk | RABEPK | 2.64E-02 | -1.94E-02 |
| 3999 | Stat4 | STAT4 | -5.09E-02 | 1.36E-02 |
| 4000 | Ppp2r2c | PPP2R2C | -1.02E-01 | 8.88E-02 |
| 4001 | Fgf12 | FGF12 | 1.39E-01 | 1.88E-02 |
| 4002 | Clec16a | CLEC16A | 2.57E-02 | -2.85E-02 |
| 4003 | Adgb | ADGB | -3.66E-02 | 4.61E-03 |
| 4004 | Dynlt3 | DYNLT3 | 2.16E-02 | -1.27E-02 |
| 4005 | Ppp1r13l | PPP1R13L | 6.36E-02 | 2.65E-02 |
| 4006 | Zfp526 | ZNF526 | -6.50E-02 | -3.54E-02 |
| 4007 | Traf6 | TRAF6 | 3.07E-02 | -2.00E-02 |
| 4008 | Porcn | PORCN | -4.00E-02 | -9.81E-03 |
| 4009 | Tmc3 | TMC3 | -1.00E-01 | -1.51E-01 |
| 4010 | Tnxb | TNXB | -8.34E-02 | -9.63E-03 |
| 4011 | Lsm12 | LSM12 | 2.95E-02 | 6.40E-03 |
| 4012 | Jup | JUP | 1.88E-02 | -2.46E-03 |
| 4013 | Pex16 | PEX16 | 3.02E-02 | -1.92E-02 |
| 4014 | Rsad2 | RSAD2 | -5.19E-02 | -2.88E-02 |
| 4015 | Fyn | FYN | -4.31E-02 | 5.07E-03 |
| 4016 | Zcwpw1 | ZCWPW1 | 5.10E-02 | -2.10E-03 |
| 4017 | Rnf169 | RNF169 | 4.95E-02 | -2.15E-02 |
| 4018 | Rnf145 | RNF145 | -3.47E-02 | -1.41E-03 |
| 4019 | Tpd52l1 | TPD52L1 | 6.21E-02 | -1.01E-02 |
| 4020 | Btnl2 | BTNL2 | -2.50E-01 | 1.05E-01 |
| 4021 | Eif4g1 | EIF4G1 | 2.46E-02 | -8.37E-03 |
| 4022 | Ahnak | AHNAK | -4.29E-02 | 2.34E-02 |
| 4023 | Nkain1 | NKAIN1 | 1.85E-01 | 4.79E-02 |
| 4024 | Odf2 | ODF2 | 2.73E-02 | -9.67E-03 |
| 4025 | Nudt8 | NUDT8 | 7.41E-02 | -3.56E-03 |

| | | | | |
|---|---|---|---|---|
| 4026 | Tsacc | TSACC | -7.27E-02 | 2.87E-02 |
| 4027 | Tmem61 | TMEM61 | -7.83E-02 | -6.38E-02 |
| 4028 | Pctp | PCTP | 4.82E-02 | -3.77E-02 |
| 4029 | Dock8 | DOCK8 | 2.83E-02 | -1.34E-02 |
| 4030 | Mrpl50 | MRPL50 | -2.90E-02 | -9.63E-03 |
| 4031 | Npb | NPB | -1.29E-01 | -5.47E-03 |
| 4032 | Etnppl | ETNPPL | 1.17E-01 | -5.48E-03 |
| 4033 | Phf11c | PHF11 | 8.22E-02 | 1.67E-02 |
| 4034 | Trim14 | TRIM14 | -9.13E-02 | 5.13E-02 |
| 4035 | Pyroxd1 | PYROXD1 | 3.11E-02 | -1.98E-02 |
| 4036 | Cd96 | CD96 | -4.62E-02 | 3.44E-02 |
| 4037 | Slc6a8 | SLC6A8 | 4.42E-02 | -1.27E-02 |
| 4038 | Zdhhc16 | ZDHHC16 | 2.30E-02 | 3.32E-03 |
| 4039 | Atp13a2 | ATP13A2 | 2.80E-02 | -8.08E-03 |
| 4040 | Aoc3 | AOC3 | 1.44E-01 | 5.10E-02 |
| 4041 | Spsb1 | SPSB1 | 1.03E-01 | -3.00E-02 |
| 4042 | Unc5cl | UNC5CL | 8.40E-02 | 7.31E-02 |
| 4043 | Aida | AIDA | -2.15E-02 | 1.28E-02 |
| 4044 | Tmem185b | TMEM185B | -2.60E-02 | -8.56E-03 |
| 4045 | Rgs9bp | RGS9BP | -7.98E-02 | 2.00E-02 |
| 4046 | Papss1 | PAPSS1 | 2.18E-02 | -1.61E-02 |
| 4047 | Rnf157 | RNF157 | 7.44E-02 | 8.36E-02 |
| 4048 | Ptpn3 | PTPN3 | -2.29E-02 | -2.35E-02 |
| 4049 | 1700030K09Rik | C19orf44 | -4.20E-02 | -1.84E-02 |
| 4050 | Csnk1d | CSNK1D | 1.60E-02 | 4.92E-03 |
| 4051 | Asb9 | ASB9 | 5.25E-02 | -3.84E-02 |
| 4052 | Ppp2r2a | PPP2R2A | 2.42E-02 | 2.12E-02 |
| 4053 | Frzb | FRZB | 1.50E-01 | -7.58E-02 |
| 4054 | Kirrel3 | KIRREL3 | -1.28E-01 | 2.77E-02 |
| 4055 | Rcbtb2 | RCBTB2 | -2.11E-02 | -1.34E-02 |
| 4056 | Hip1 | HIP1 | 4.59E-02 | 1.78E-02 |
| 4057 | Zscan30 | ZSCAN30 | -7.13E-02 | -2.64E-02 |
| 4058 | Lca5 | LCA5 | -8.97E-02 | 3.91E-02 |
| 4059 | Haus1 | HAUS1 | 4.03E-02 | -8.69E-03 |
| 4060 | Cdc20 | CDC20 | 2.13E-01 | 1.39E-01 |
| 4061 | Mutyh | MUTYH | -6.76E-02 | 1.40E-02 |
| 4062 | Rnasel | RNASEL | -5.33E-02 | -3.30E-02 |
| 4063 | Adck1 | ADCK1 | 2.94E-02 | -1.54E-02 |
| 4064 | Clec4a1 | ZNF705A | -1.54E-01 | 1.84E-01 |
| 4065 | Pycard | PYCARD | -5.40E-02 | 7.58E-02 |
| 4066 | Noxred1 | NOXRED1 | -1.29E-01 | -7.76E-03 |
| 4067 | Dnajb1 | DNAJB1 | -7.36E-02 | 2.33E-02 |
| 4068 | Ildr2 | ILDR2 | 6.63E-02 | -1.39E-02 |
| 4069 | Bysl | BYSL | 3.82E-02 | 2.62E-02 |
| 4070 | Arpc5l | ARPC5L | 2.29E-02 | 4.84E-03 |
| 4071 | Liph | LIPH | 1.27E-01 | 4.85E-02 |
| 4072 | Wasf2 | WASF2 | -1.76E-02 | -6.24E-03 |
| 4073 | Mtus1 | MTUS1 | -4.22E-02 | -1.84E-02 |
| 4074 | Tgm7 | TGM7 | -7.94E-02 | 7.79E-02 |
| 4075 | Btf3 | BTF3 | -2.06E-02 | 4.73E-03 |
| 4076 | Trim16 | TRIM16 | 1.43E-01 | -1.92E-02 |
| 4077 | Eif3c | EIF3C | 2.51E-02 | -8.18E-03 |
| 4078 | Dimt1 | DIMT1 | -5.18E-02 | 3.54E-02 |

| | | | | |
|---|---|---|---|---|
| 4079 | Aire | AIRE | 1.21E-01 | -2.45E-02 |
| 4080 | Slc16a14 | SLC16A14 | 8.61E-02 | 1.43E-03 |
| 4081 | Kif20a | KIF20A | 2.12E-01 | 1.10E-01 |
| 4082 | Tmem150c | TMEM150C | -4.30E-02 | -3.63E-02 |
| 4083 | Usp28 | USP28 | 4.17E-02 | 4.36E-03 |
| 4084 | Unc45b | UNC45B | -7.50E-02 | -5.83E-03 |
| 4085 | Rab34 | RAB34 | 3.76E-02 | 1.13E-02 |
| 4086 | Svil | SVIL | -3.54E-02 | -3.65E-02 |
| 4087 | Knstrn | KNSTRN | 2.32E-01 | 1.31E-01 |
| 4088 | H13 | HM13 | 2.52E-02 | -6.81E-03 |
| 4089 | Top2a | TOP2A | 2.22E-01 | 2.14E-01 |
| 4090 | Dab2ip | DAB2IP | 2.49E-02 | -6.75E-03 |
| 4091 | Ldlr | LDLR | -1.20E-01 | -1.77E-02 |
| 4092 | Pla2g6 | PLA2G6 | -3.92E-02 | 8.97E-03 |
| 4093 | Etaa1 | ETAA1 | -5.75E-02 | -1.74E-02 |
| 4094 | Cactin | CACTIN | -2.98E-02 | -6.09E-03 |
| 4095 | Tyrobp | TYROBP | -9.69E-02 | 1.54E-01 |
| 4096 | Mfsd5 | MFSD5 | 3.09E-02 | -7.83E-03 |
| 4097 | Itpkc | ITPKC | -3.79E-02 | 3.10E-02 |
| 4098 | Ckap2 | CKAP2 | 1.94E-01 | 1.19E-01 |
| 4099 | Odf2l | ODF2L | -2.30E-02 | -1.46E-02 |
| 4100 | Rnmt | RNMT | 3.07E-02 | -3.85E-03 |
| 4101 | D2hgdh | D2HGDH | -3.39E-02 | -3.72E-03 |
| 4102 | Sema3e | SEMA3E | -1.00E-01 | 1.65E-02 |
| 4103 | Rnf39 | RNF39 | -1.45E-01 | 3.04E-02 |
| 4104 | Hdac3 | HDAC3 | 1.83E-02 | -2.40E-03 |
| 4105 | Rpl3 | RPL3 | -2.36E-02 | 3.70E-02 |
| 4106 | Rrp9 | RRP9 | -3.71E-02 | 7.17E-04 |
| 4107 | Rab21 | RAB21 | 1.98E-02 | -8.77E-03 |
| 4108 | Notch2 | NOTCH2 | -2.44E-02 | -1.51E-02 |
| 4109 | Eif4ebp3 | EIF4EBP3 | 1.02E-01 | 2.52E-02 |
| 4110 | Fam120b | FAM120B | -3.05E-02 | 2.90E-03 |
| 4111 | Kif2c | KIF2C | 2.10E-01 | 9.23E-02 |
| 4112 | Irf6 | IRF6 | 4.48E-02 | -1.23E-02 |
| 4113 | Eprs | EPRS | 2.45E-02 | -5.38E-03 |
| 4114 | Slc33a1 | SLC33A1 | -2.78E-02 | -4.26E-02 |
| 4115 | Cblc | CBLC | 8.21E-02 | -5.16E-03 |
| 4116 | Pafah1b1 | PAFAH1B1 | 1.96E-02 | -1.11E-02 |
| 4117 | Ubn2 | UBN2 | -2.76E-02 | 7.83E-04 |
| 4118 | Mthfd2l | MTHFD2L | 5.25E-02 | 3.54E-02 |
| 4119 | Tom1 | TOM1 | 2.88E-02 | -1.34E-02 |
| 4120 | Lclat1 | LCLAT1 | 2.92E-02 | -2.90E-02 |
| 4121 | Hnrnpd | HNRNPD | 2.43E-02 | -1.51E-02 |
| 4122 | Stac | STAC | 9.14E-02 | -4.50E-03 |
| 4123 | Noc3l | NOC3L | -3.10E-02 | 2.33E-02 |
| 4124 | Dgkq | DGKQ | 2.65E-02 | -1.94E-02 |
| 4125 | Trim44 | TRIM44 | 2.24E-02 | -1.56E-02 |
| 4126 | Eid2b | EID2B | 4.20E-02 | -2.82E-02 |
| 4127 | Mrpl33 | MRPL33 | 4.42E-02 | 1.44E-02 |
| 4128 | Isyna1 | ISYNA1 | 4.51E-02 | -9.46E-03 |
| 4129 | Psma2 | PSMA2 | 2.10E-02 | -6.39E-03 |
| 4130 | Gstm7 | GSTM2 | -5.49E-02 | -5.27E-02 |
| 4131 | Zfp655 | ZNF655 | -3.81E-02 | 1.46E-03 |

| | | | | |
|---|---|---|---|---|
| 4132 | Nudt19 | NUDT19 | -4.17E-02 | 1.61E-02 |
| 4133 | Trem2 | TREM2 | 1.35E-01 | 1.66E-01 |
| 4134 | Zfp595 | ZNF595 | -4.88E-02 | -1.90E-02 |
| 4135 | Swap70 | SWAP70 | -3.20E-02 | -1.25E-03 |
| 4136 | Mgat2 | MGAT2 | 2.46E-02 | -1.88E-02 |
| 4137 | Brpf1 | BRPF1 | 2.79E-02 | 1.80E-03 |
| 4138 | Mmp16 | MMP16 | -7.31E-02 | 7.36E-02 |
| 4139 | D6Wsu163e | C12orf4 | -3.74E-02 | 4.57E-03 |
| 4140 | Zfp11 | ZNF546 | -6.16E-02 | -2.52E-02 |
| 4141 | Man1a | MAN1A1 | 4.00E-02 | -1.03E-02 |
| 4142 | Hexim2 | HEXIM2 | 5.77E-02 | -2.25E-02 |
| 4143 | Rab3a | RAB3A | -5.39E-02 | 8.93E-03 |
| 4144 | Bod1 | BOD1 | 2.15E-02 | -1.76E-02 |
| 4145 | Mmgt1 | MMGT1 | 2.40E-02 | -1.89E-02 |
| 4146 | Rhoq | RHOQ | 2.49E-02 | -1.33E-02 |
| 4147 | Cysltr1 | CYSLTR1 | -1.08E-01 | 1.24E-01 |
| 4148 | Tmem147 | TMEM147 | 2.35E-02 | -1.94E-03 |
| 4149 | Padi4 | PADI4 | -1.08E-01 | 4.73E-02 |
| 4150 | Trim72 | TRIM72 | 8.60E-02 | -1.31E-02 |
| 4151 | Naaladl1 | NAALADL1 | -7.10E-02 | -2.33E-02 |
| 4152 | Map2k2 | MAP2K2 | 2.24E-02 | -2.61E-04 |
| 4153 | Pcgf3 | PCGF3 | 2.65E-02 | 2.69E-03 |
| 4154 | Ctr9 | CTR9 | -1.83E-02 | 3.81E-03 |
| 4155 | Tnfsf11 | TNFSF11 | -2.96E-02 | -2.68E-03 |
| 4156 | Creg2 | CREG2 | -1.54E-01 | 3.29E-02 |
| 4157 | Tor2a | TOR2A | 2.54E-02 | 6.24E-04 |
| 4158 | Tmem203 | TMEM203 | 4.62E-02 | -1.61E-02 |
| 4159 | Psma7 | PSMA7 | 2.39E-02 | 3.62E-03 |
| 4160 | Cry1 | CRY1 | 1.60E-01 | 1.65E-01 |
| 4161 | Ccr2 | CCR2 | -1.87E-01 | 2.15E-01 |
| 4162 | Arid2 | ARID2 | -2.54E-02 | -1.19E-03 |
| 4163 | Rpl35a | RPL35A | -2.28E-02 | 2.23E-03 |
| 4164 | Fam189b | FAM189B | -6.12E-02 | 3.27E-02 |
| 4165 | Klhl11 | KLHL11 | -8.57E-02 | -5.12E-02 |
| 4166 | Igfn1 | IGFN1 | 9.71E-02 | -9.65E-02 |
| 4167 | Nrde2 | NRDE2 | 4.51E-02 | -5.07E-02 |
| 4168 | Bivm | BIVM | -2.52E-02 | -1.88E-03 |
| 4169 | Ank1 | ANK1 | 9.96E-02 | -6.42E-03 |
| 4170 | Iqgap3 | IQGAP3 | 2.27E-01 | 2.48E-01 |
| 4171 | Jakmip1 | JAKMIP1 | -4.91E-02 | 3.37E-02 |
| 4172 | Arhgap18 | ARHGAP18 | -3.64E-02 | -1.84E-02 |
| 4173 | Rilp | RILP | 3.51E-02 | -4.04E-03 |
| 4174 | Ptrh1 | PTRH1 | 6.84E-02 | 6.24E-02 |
| 4175 | Noc4l | NOC4L | 3.20E-02 | 1.90E-02 |
| 4176 | Kcnmb4 | KCNMB4 | 5.75E-02 | -1.80E-02 |
| 4177 | Usp19 | USP19 | -1.96E-02 | -9.30E-03 |
| 4178 | Nedd8 | NEDD8 | 2.15E-02 | -1.51E-03 |
| 4179 | Cep57 | CEP57 | 2.42E-02 | -5.90E-03 |
| 4180 | Fbxo2 | FBXO2 | 1.22E-01 | -1.93E-02 |
| 4181 | Dctn3 | DCTN3 | 2.09E-02 | 4.21E-03 |
| 4182 | Dnajc19-ps | DNAJC19 | 6.69E-02 | -2.54E-02 |
| 4183 | Bmp3 | BMP3 | 2.12E-01 | -1.72E-02 |
| 4184 | Tsc22d3 | TSC22D3 | 6.95E-02 | -1.03E-03 |

| | | | | |
|---|---|---|---|---|
| 4185 | Tatdn2 | TATDN2 | 2.59E-02 | -1.07E-03 |
| 4186 | Slc25a17 | SLC25A17 | -2.91E-02 | 9.66E-03 |
| 4187 | Pfkp | PFKP | 7.27E-02 | -1.84E-02 |
| 4188 | Icam1 | ICAM1 | 9.08E-02 | 9.00E-02 |
| 4189 | Bend4 | BEND4 | -8.23E-02 | -4.14E-03 |
| 4190 | Cntrob | CNTROB | -4.27E-02 | 2.62E-02 |
| 4191 | Gper1 | GPER1 | 9.22E-02 | -6.83E-02 |
| 4192 | Syn3 | SYN3 | -7.37E-02 | -8.29E-02 |
| 4193 | Ncaph | NCAPH | 1.64E-01 | 2.16E-01 |
| 4194 | Avil | AVIL | -1.16E-01 | 3.59E-02 |
| 4195 | Alkbh3 | ALKBH3 | 3.27E-02 | -2.48E-02 |
| 4196 | Stab2 | STAB2 | -1.13E-01 | -1.61E-03 |
| 4197 | Col25a1 | COL25A1 | -6.60E-02 | 1.64E-01 |
| 4198 | Il27 | IL27 | -3.68E-02 | 1.61E-02 |
| 4199 | Ankrd53 | ANKRD53 | 9.52E-02 | 5.18E-02 |
| 4200 | Txnl1 | TXNL1 | 2.49E-02 | -4.61E-03 |
| 4201 | Mrpl4 | MRPL4 | 2.80E-02 | -9.58E-03 |
| 4202 | Cecr2 | CECR2 | -6.73E-02 | -8.51E-02 |
| 4203 | Atp5c1 | ATP5C1 | 2.53E-02 | -1.17E-02 |
| 4204 | Aqp2 | AQP2 | 9.82E-02 | -5.14E-03 |
| 4205 | Bmp5 | BMP5 | -1.76E-01 | -5.62E-02 |
| 4206 | Pmm2 | PMM2 | 2.18E-02 | -5.51E-03 |
| 4207 | Spata1 | SPATA1 | -7.41E-02 | 4.02E-02 |
| 4208 | Il1b | IL1B | -3.12E-01 | 2.06E-01 |
| 4209 | Bhmt2 | BHMT2 | 4.27E-02 | -2.83E-02 |
| 4210 | Kcnj3 | KCNJ3 | -5.15E-02 | -1.88E-02 |
| 4211 | Nudcd2 | NUDCD2 | 2.92E-02 | 7.63E-03 |
| 4212 | Fanca | FANCA | 1.38E-01 | 6.92E-03 |
| 4213 | Tpsb2 | TPSB2 | -8.60E-02 | -6.17E-03 |
| 4214 | Tpsb2 | TPSAB1 | -8.60E-02 | -6.17E-03 |
| 4215 | Nfkbia | NFKBIA | 1.06E-01 | -4.88E-03 |
| 4216 | Vps13a | VPS13A | -3.23E-02 | -1.08E-02 |
| 4217 | Ugt2b38 | UGT2B28 | 8.43E-02 | 6.15E-02 |
| 4218 | Fhod3 | FHOD3 | 4.50E-02 | -5.51E-02 |
| 4219 | Psmb3 | PSMB3 | 2.46E-02 | -7.54E-03 |
| 4220 | Cryl1 | CRYL1 | 4.90E-02 | -5.24E-03 |
| 4221 | Ppp1r3d | PPP1R3D | -7.30E-02 | 9.96E-02 |
| 4222 | Apcdd1 | APCDD1 | 1.25E-01 | -9.70E-02 |
| 4223 | Yipf1 | YIPF1 | 2.04E-02 | -1.43E-02 |
| 4224 | Olfml1 | OLFML1 | -1.41E-01 | 2.14E-02 |
| 4225 | Dtl | DTL | 1.94E-01 | 1.77E-01 |
| 4226 | Wipf2 | WIPF2 | -2.21E-02 | -1.88E-03 |
| 4227 | Klhdc4 | KLHDC4 | 2.61E-02 | -1.76E-02 |
| 4228 | Vps16 | VPS16 | 2.60E-02 | -1.69E-02 |
| 4229 | B2m | B2M | -5.97E-02 | 5.38E-02 |
| 4230 | Kifc3 | KIFC3 | 2.32E-02 | -2.39E-02 |
| 4231 | Gpr108 | GPR108 | 3.44E-02 | -1.02E-02 |
| 4232 | Ctcf | CTCF | -1.66E-02 | 6.55E-04 |
| 4233 | Trappc11 | TRAPPC11 | 2.28E-02 | -1.32E-02 |
| 4234 | Ppp1r9b | PPP1R9B | -2.53E-02 | 1.23E-02 |
| 4235 | Olfr558 | OR51E1 | 9.54E-02 | 1.62E-02 |
| 4236 | Crem | CREM | 8.70E-02 | 5.34E-02 |
| 4237 | Rock2 | ROCK2 | 2.36E-02 | 2.63E-02 |

| | | | | |
|---|---|---|---|---|
| 4238 | Abcb8 | ABCB8 | 2.65E-02 | -2.41E-02 |
| 4239 | Cebpz | CEBPZ | -2.86E-02 | 2.93E-03 |
| 4240 | Bnip3 | BNIP3 | 3.77E-02 | -3.60E-03 |
| 4241 | B3gnt5 | B3GNT5 | -4.80E-02 | -3.15E-02 |
| 4242 | Cnih2 | CNIH2 | 6.48E-02 | 6.12E-03 |
| 4243 | Phf14 | PHF14 | 2.60E-02 | -2.56E-02 |
| 4244 | Sgms1 | SGMS1 | -3.40E-02 | 1.08E-02 |
| 4245 | Hecw2 | HECW2 | 6.80E-02 | -5.45E-02 |
| 4246 | Zfp764 | ZNF764 | -4.25E-02 | -3.87E-03 |
| 4247 | Rpap2 | RPAP2 | 2.77E-02 | -2.48E-02 |
| 4248 | Map4k4 | MAP4K4 | 2.50E-02 | 2.35E-02 |
| 4249 | Ccdc117 | CCDC117 | -6.60E-02 | -1.68E-02 |
| 4250 | Adra2b | ADRA2B | 5.39E-02 | -1.47E-02 |
| 4251 | Mau2 | MAU2 | 1.79E-02 | -6.63E-03 |
| 4252 | Ecd | ECD | 2.19E-02 | -9.57E-03 |
| 4253 | Slc16a10 | SLC16A10 | 5.42E-02 | -2.13E-02 |
| 4254 | Clcn1 | CLCN1 | 5.21E-02 | -2.80E-02 |
| 4255 | Psmf1 | PSMF1 | 2.44E-02 | -1.24E-02 |
| 4256 | Lmf1 | LMF1 | 2.94E-02 | -2.00E-02 |
| 4257 | Rpp14 | RPP14 | -2.28E-02 | 1.38E-02 |
| 4258 | Jmjd1c | JMJD1C | -3.45E-02 | -7.46E-03 |
| 4259 | Iqgap2 | IQGAP2 | 4.34E-02 | -1.86E-02 |
| 4260 | Chn1 | CHN1 | 3.62E-02 | -6.77E-03 |
| 4261 | Ankrd61 | ANKRD61 | 4.77E-02 | -5.37E-02 |
| 4262 | Ptprs | PTPRS | -4.46E-02 | 6.52E-03 |
| 4263 | Zbtb37 | ZBTB37 | -3.53E-02 | -2.85E-02 |
| 4264 | Pirb | LILRB3 | -1.49E-01 | 1.80E-01 |
| 4265 | Tmem259 | TMEM259 | 2.18E-02 | -1.40E-02 |
| 4266 | Gale | GALE | 7.09E-02 | 6.78E-02 |
| 4267 | Uqcr11 | UQCR11 | 2.61E-02 | -1.32E-02 |
| 4268 | Lmcd1 | LMCD1 | 5.91E-02 | 1.15E-02 |
| 4269 | Plekhd1 | PLEKHD1 | -4.43E-02 | 4.34E-02 |
| 4270 | Tmem199 | TMEM199 | 2.85E-02 | 6.15E-03 |
| 4271 | Abhd14b | ABHD14B | -2.93E-02 | -3.15E-02 |
| 4272 | Sypl | SYPL1 | 2.04E-02 | -1.53E-02 |
| 4273 | Csnk2b | CSNK2B | 2.42E-02 | -9.03E-03 |
| 4274 | F10 | F10 | 1.93E-01 | 4.83E-02 |
| 4275 | Hddc3 | HDDC3 | -4.49E-02 | -2.37E-02 |
| 4276 | Epha2 | EPHA2 | 1.10E-01 | 5.49E-02 |
| 4277 | Stc1 | STC1 | 1.30E-01 | -2.11E-02 |
| 4278 | Exosc7 | EXOSC7 | 2.93E-02 | -1.92E-02 |
| 4279 | Naa10 | NAA10 | -4.52E-02 | 2.14E-02 |
| 4280 | Fgfbp3 | FGFBP3 | -7.10E-02 | -5.17E-03 |
| 4281 | Slc25a34 | SLC25A34 | -5.04E-02 | 1.13E-02 |
| 4282 | Acaa2 | ACAA2 | 4.90E-02 | -1.55E-02 |
| 4283 | Pcmtd2 | PCMTD2 | -3.29E-02 | -2.90E-02 |
| 4284 | Wnt11 | WNT11 | -2.42E-01 | -3.33E-02 |
| 4285 | Tacc3 | TACC3 | 1.34E-01 | 1.08E-01 |
| 4286 | Colq | COLQ | 1.24E-01 | -7.50E-03 |
| 4287 | Fam110c | FAM110C | -6.58E-02 | 1.89E-02 |
| 4288 | Dynlrb2 | DYNLRB2 | 6.05E-02 | -2.14E-02 |
| 4289 | Cyp4a10 | CYP4A11 | 6.76E-02 | -2.54E-02 |
| 4290 | Abce1 | ABCE1 | 2.03E-02 | 2.90E-03 |

| | | | | |
|---|---|---|---|---|
| 4291 | 4930505A04Rik | C2orf73 | -5.46E-02 | -2.11E-04 |
| 4292 | Eif2a | EIF2A | -2.75E-02 | 9.67E-03 |
| 4293 | Ctps2 | CTPS2 | -2.33E-02 | -5.00E-03 |
| 4294 | Fut11 | FUT11 | -5.30E-02 | 3.44E-02 |
| 4295 | Hk3 | HK3 | -1.28E-01 | 8.49E-02 |
| 4296 | Mex3a | MEX3A | 9.15E-02 | 6.54E-02 |
| 4297 | Unc13d | UNC13D | -1.01E-01 | 6.96E-02 |
| 4298 | Camk2a | CAMK2A | -8.41E-02 | 3.56E-02 |
| 4299 | Atf6b | ATF6B | -2.19E-02 | -2.98E-04 |
| 4300 | Ryr3 | RYR3 | -4.96E-02 | 1.19E-02 |
| 4301 | Tpgs2 | TPGS2 | 3.80E-02 | -8.83E-03 |
| 4302 | Thoc5 | THOC5 | 2.44E-02 | 9.32E-03 |
| 4303 | Atg9b | ATG9B | -7.87E-02 | 1.01E-01 |
| 4304 | Gimap6 | GIMAP6 | -3.77E-02 | -1.64E-02 |
| 4305 | Zfpm2 | ZFPM2 | -1.06E-01 | -5.79E-02 |
| 4306 | Ptgir | PTGIR | -1.11E-01 | 3.50E-02 |
| 4307 | Gab3 | GAB3 | 1.02E-01 | 5.70E-03 |
| 4308 | Kansl2 | KANSL2 | 2.93E-02 | 9.92E-04 |
| 4309 | Slc3a2 | SLC3A2 | 4.45E-02 | -2.33E-02 |
| 4310 | Tuba1c | TUBA1C | 5.10E-02 | 4.73E-02 |
| 4311 | Osbpl2 | OSBPL2 | 1.81E-02 | -7.45E-03 |
| 4312 | Nfam1 | NFAM1 | -1.08E-01 | 1.89E-01 |
| 4313 | Zfp276 | ZNF276 | -3.57E-02 | -1.08E-04 |
| 4314 | Tax1bp3 | TAX1BP3 | 3.66E-02 | 2.51E-02 |
| 4315 | Sfxn3 | SFXN3 | 3.67E-02 | 1.63E-02 |
| 4316 | Nrbp1 | NRBP1 | 1.62E-02 | -4.74E-03 |
| 4317 | Pdzrn3 | PDZRN3 | 5.63E-02 | -4.40E-02 |
| 4318 | Cd72 | CD72 | -1.12E-01 | 8.98E-02 |
| 4319 | Fam174a | FAM174A | 2.77E-02 | -1.81E-02 |
| 4320 | Yipf2 | YIPF2 | -2.99E-02 | -1.12E-02 |
| 4321 | Cox19 | COX19 | 2.71E-02 | -2.30E-03 |
| 4322 | Lmbrd2 | LMBRD2 | -9.54E-02 | -6.80E-02 |
| 4323 | Copb1 | COPB1 | 2.08E-02 | -9.59E-03 |
| 4324 | Hnrnph2 | HNRNPH2 | 1.99E-02 | -6.30E-03 |
| 4325 | Zfand5 | ZFAND5 | 2.95E-02 | 2.83E-02 |
| 4326 | Wdfy2 | WDFY2 | 2.82E-02 | -2.24E-02 |
| 4327 | Itgb2 | ITGB2 | -1.53E-01 | 2.11E-01 |
| 4328 | Dand5 | DAND5 | -4.71E-02 | 8.38E-03 |
| 4329 | Adam21 | ADAM21 | 4.03E-02 | -3.11E-02 |
| 4330 | Ak5 | AK5 | -1.56E-01 | -2.64E-02 |
| 4331 | Ogfrl1 | OGFRL1 | -5.56E-02 | 5.88E-03 |
| 4332 | Mospd2 | MOSPD2 | -3.64E-02 | 2.41E-02 |
| 4333 | Ccdc86 | CCDC86 | 4.53E-02 | 2.99E-02 |
| 4334 | Dnajb11 | DNAJB11 | 4.31E-02 | -4.31E-03 |
| 4335 | Pgls | PGLS | 2.94E-02 | -5.34E-03 |
| 4336 | Klrg1 | KLRG1 | -3.28E-02 | 4.65E-03 |
| 4337 | Emc6 | EMC6 | 2.87E-02 | -1.44E-03 |
| 4338 | Zfp770 | ZNF770 | -6.11E-02 | -1.18E-02 |
| 4339 | L3mbtl3 | L3MBTL3 | -5.01E-02 | 1.17E-02 |
| 4340 | S100a14 | S100A14 | 1.74E-01 | -6.50E-04 |
| 4341 | Fbp1 | FBP1 | 4.46E-02 | -3.89E-03 |
| 4342 | Casq1 | CASQ1 | 3.57E-02 | 1.99E-03 |
| 4343 | Trpm2 | TRPM2 | -1.15E-01 | 8.56E-02 |

| | | | | |
|---|---|---|---|---|
| 4344 | Adamts16 | ADAMTS16 | 8.45E-02 | 6.27E-02 |
| 4345 | B4galnt2 | B4GALNT2 | 7.45E-02 | -4.42E-02 |
| 4346 | Myct1 | MYCT1 | -8.89E-02 | 7.01E-03 |
| 4347 | Clec1b | CLEC1B | 7.89E-02 | -4.08E-02 |
| 4348 | Gm3055 | ZNF624 | -7.14E-02 | 2.82E-02 |
| 4349 | Mbd4 | MBD4 | -6.24E-02 | -1.47E-02 |
| 4350 | Gpsm2 | GPSM2 | 9.61E-02 | 1.90E-02 |
| 4351 | Serhl | SERHL2 | 2.26E-02 | -1.44E-02 |
| 4352 | Tssk6 | TSSK6 | 6.85E-02 | 1.38E-02 |
| 4353 | Slc46a1 | SLC46A1 | -3.09E-02 | -4.12E-02 |
| 4354 | Armcx6 | ARMCX6 | -7.70E-02 | 3.04E-02 |
| 4355 | Lrch3 | LRCH3 | -2.10E-02 | 2.49E-03 |
| 4356 | Polr3c | POLR3C | 2.18E-02 | -8.16E-03 |
| 4357 | Ccdc149 | CCDC149 | 5.91E-02 | -3.48E-02 |
| 4358 | Gm3776 | GSTA5 | 2.33E-01 | 1.32E-01 |
| 4359 | Gmds | GMDS | 3.76E-02 | 1.78E-02 |
| 4360 | Trpc5 | TRPC5 | -6.07E-02 | 5.14E-02 |
| 4361 | P2ry6 | P2RY6 | -8.96E-02 | 1.16E-01 |
| 4362 | Cacng4 | CACNG4 | -6.63E-02 | 3.13E-02 |
| 4363 | Arfgap3 | ARFGAP3 | 2.80E-02 | 2.26E-03 |
| 4364 | Ube3c | UBE3C | 2.65E-02 | -4.98E-03 |
| 4365 | Socs7 | SOCS7 | 2.57E-02 | -1.87E-02 |
| 4366 | Lmtk3 | LMTK3 | -6.60E-02 | 3.25E-02 |
| 4367 | Ap5s1 | AP5S1 | 2.55E-02 | 6.21E-03 |
| 4368 | Nup188 | NUP188 | -2.60E-02 | 2.12E-02 |
| 4369 | Zfhx3 | ZFHX3 | -6.12E-02 | 3.13E-02 |
| 4370 | Imp3 | IMP3 | -1.91E-02 | -5.12E-03 |
| 4371 | Nipal2 | NIPAL2 | 5.81E-02 | 3.02E-02 |
| 4372 | Cep350 | CEP350 | -2.89E-02 | 1.18E-02 |
| 4373 | Igf1r | IGF1R | 2.38E-02 | -1.87E-02 |
| 4374 | Meox1 | MEOX1 | -8.82E-02 | -3.37E-02 |
| 4375 | Gcnt1 | GCNT1 | -3.95E-02 | -3.88E-02 |
| 4376 | Gnai3 | GNAI3 | 2.29E-02 | -5.08E-03 |
| 4377 | Szrd1 | SZRD1 | 2.35E-02 | -2.07E-02 |
| 4378 | Ppm1k | PPM1K | -4.43E-02 | -5.19E-02 |
| 4379 | Pcf11 | PCF11 | -2.46E-02 | 3.55E-03 |
| 4380 | Trmt1 | TRMT1 | -2.52E-02 | 1.61E-04 |
| 4381 | Urm1 | URM1 | 2.49E-02 | -4.14E-03 |
| 4382 | Galm | GALM | 2.84E-02 | -1.92E-02 |
| 4383 | Taz | TAZ | 1.88E-02 | -8.58E-03 |
| 4384 | Prr7 | PRR7 | 5.52E-02 | -2.92E-02 |
| 4385 | Cpped1 | CPPED1 | 2.53E-02 | -1.53E-02 |
| 4386 | Mag | MAG | -5.92E-02 | 6.97E-04 |
| 4387 | Mlh1 | MLH1 | 2.70E-02 | -1.31E-02 |
| 4388 | Kitl | KITLG | -5.19E-02 | 3.07E-02 |
| 4389 | Zbtb39 | ZBTB39 | -3.38E-02 | 1.04E-03 |
| 4390 | Asb15 | ASB15 | -5.95E-02 | -2.48E-02 |
| 4391 | Topbp1 | TOPBP1 | 3.52E-02 | 3.35E-02 |
| 4392 | Dlgap1 | DLGAP1 | -1.06E-01 | 7.97E-03 |
| 4393 | Rps6kb2 | RPS6KB2 | 2.79E-02 | 7.66E-03 |
| 4394 | Scn2b | SCN2B | -1.06E-01 | 6.33E-03 |
| 4395 | Acadvl | ACADVL | 2.90E-02 | -1.92E-02 |
| 4396 | Vwf | VWF | 1.51E-01 | 5.26E-02 |

| | | | | |
|---|---|---|---|---|
| 4397 | Mzt1 | MZT1 | 2.18E-02 | -5.75E-03 |
| 4398 | Mtif2 | MTIF2 | -2.24E-02 | -7.33E-03 |
| 4399 | Fxn | FXN | 3.56E-02 | -1.22E-02 |
| 4400 | Vrk2 | VRK2 | 4.49E-02 | -2.81E-02 |
| 4401 | Kat8 | KAT8 | 2.89E-02 | -6.41E-03 |
| 4402 | Fech | FECH | 3.67E-02 | -2.08E-02 |
| 4403 | Cops6 | COPS6 | 2.31E-02 | -7.13E-03 |
| 4404 | Snrnp27 | SNRNP27 | -2.62E-02 | 5.53E-03 |
| 4405 | Klhl5 | KLHL5 | 2.50E-02 | -3.41E-02 |
| 4406 | Card6 | CARD6 | -4.89E-02 | -1.58E-02 |
| 4407 | Mroh6 | MROH6 | -8.76E-02 | 3.09E-02 |
| 4408 | Tbc1d32 | TBC1D32 | -3.74E-02 | -2.11E-02 |
| 4409 | Pwp1 | PWP1 | 2.15E-02 | 2.46E-03 |
| 4410 | Surf4 | SURF4 | 2.07E-02 | 1.76E-03 |
| 4411 | Myo16 | MYO16 | 4.34E-02 | -2.01E-02 |
| 4412 | Pi4ka | PI4KA | 2.30E-02 | -2.46E-02 |
| 4413 | Hnrnpc | HNRNPC | -2.48E-02 | 9.53E-03 |
| 4414 | Efna3 | EFNA3 | 8.90E-02 | -3.00E-02 |
| 4415 | Olfm2 | OLFM2 | 1.62E-01 | -1.57E-02 |
| 4416 | Nav2 | NAV2 | 4.18E-02 | -1.14E-02 |
| 4417 | Pcdh1 | PCDH1 | 4.91E-02 | -1.19E-02 |
| 4418 | D630023F18Rik | C2orf80 | 8.14E-02 | 1.49E-03 |
| 4419 | Serp2 | SERP2 | 7.87E-02 | 1.55E-02 |
| 4420 | Hopx | HOPX | -6.89E-02 | -1.63E-02 |
| 4421 | Rragd | RRAGD | 3.19E-02 | -9.45E-04 |
| 4422 | Cdca3 | CDCA3 | 1.76E-01 | 1.12E-01 |
| 4423 | Rexo1 | REXO1 | 3.05E-02 | 3.60E-03 |
| 4424 | Man2a2 | MAN2A2 | 2.45E-02 | -2.59E-02 |
| 4425 | Ywhah | YWHAH | 3.02E-02 | 3.68E-02 |
| 4426 | Rfesd | RFESD | 3.75E-02 | -4.10E-02 |
| 4427 | Slc22a23 | SLC22A23 | -5.45E-02 | -4.55E-03 |
| 4428 | Ube2d3 | UBE2D3 | 2.08E-02 | -1.16E-02 |
| 4429 | Lmbrd1 | LMBRD1 | 1.95E-02 | -1.86E-02 |
| 4430 | Cers5 | CERS5 | -2.01E-02 | -6.62E-03 |
| 4431 | Tram2 | TRAM2 | -5.11E-02 | -6.21E-03 |
| 4432 | Med13 | MED13 | -2.27E-02 | 7.90E-03 |
| 4433 | Fam167b | FAM167B | 8.50E-02 | -2.10E-02 |
| 4434 | Prdx6 | PRDX6 | 2.75E-02 | -7.79E-03 |
| 4435 | Tmem74b | TMEM74B | -5.96E-02 | 5.23E-02 |
| 4436 | Tmem170 | TMEM170A | -3.89E-02 | 1.33E-02 |
| 4437 | Mrpl1 | MRPL1 | -2.95E-02 | -2.02E-02 |
| 4438 | Zfp3 | ZFP3 | -6.04E-02 | -1.12E-03 |
| 4439 | Lmx1a | LMX1A | 5.58E-02 | -3.47E-02 |
| 4440 | Lepr | LEPR | -2.14E-01 | -6.43E-02 |
| 4441 | Mkrn1 | MKRN1 | 1.98E-02 | -4.76E-03 |
| 4442 | Ppargc1b | PPARGC1B | -5.16E-02 | -3.27E-02 |
| 4443 | Abhd4 | ABHD4 | -1.88E-02 | -1.91E-02 |
| 4444 | Aig1 | AIG1 | -3.68E-02 | -4.41E-04 |
| 4445 | Mef2d | MEF2D | 2.92E-02 | 1.19E-02 |
| 4446 | Npdc1 | NPDC1 | -4.86E-02 | 2.87E-02 |
| 4447 | Pam16 | PAM16 | 3.15E-02 | -1.42E-02 |
| 4448 | Suz12 | SUZ12 | 2.12E-02 | 9.28E-06 |
| 4449 | Pigm | PIGM | 2.29E-02 | -1.21E-03 |

| | | | | |
|---|---|---|---|---|
| 4450 | Lctl | LCTL | 1.17E-01 | -6.34E-02 |
| 4451 | Haghl | HAGHL | 3.02E-02 | -5.69E-03 |
| 4452 | Med27 | MED27 | 3.06E-02 | -1.10E-03 |
| 4453 | Zbtb7b | ZBTB7B | 1.83E-02 | -1.20E-03 |
| 4454 | Acox2 | ACOX2 | -6.80E-02 | -1.13E-01 |
| 4455 | Smim8 | SMIM8 | -3.04E-02 | 6.30E-03 |
| 4456 | Msl3 | MSL3 | 1.73E-02 | -1.94E-02 |
| 4457 | Tmem141 | TMEM141 | 4.09E-02 | -2.09E-02 |
| 4458 | Lancl2 | LANCL2 | 3.06E-02 | 3.92E-03 |
| 4459 | Capn11 | CAPN11 | 2.53E-02 | 2.26E-03 |
| 4460 | Pcdhgb8 | PCDHGB3 | -5.31E-02 | 1.72E-02 |
| 4461 | Prkcb | PRKCB | -9.41E-02 | 3.90E-03 |
| 4462 | Rbm20 | RBM20 | 6.35E-02 | -1.46E-02 |
| 4463 | Cbr3 | CBR3 | 1.70E-01 | 7.27E-02 |
| 4464 | Zfp266 | ZNF266 | -2.68E-02 | 2.32E-03 |
| 4465 | Ddx11 | DDX11 | -4.95E-02 | 4.68E-02 |
| 4466 | Vamp2 | VAMP2 | 2.35E-02 | 7.27E-03 |
| 4467 | Serpinb8 | SERPINB8 | -8.69E-02 | 5.80E-02 |
| 4468 | Rgp1 | RGP1 | 1.99E-02 | -2.27E-02 |
| 4469 | Cacna1s | CACNA1S | 3.74E-02 | -3.49E-02 |
| 4470 | Synj2bp | SYNJ2BP | -2.17E-02 | -4.01E-03 |
| 4471 | Trim2 | TRIM2 | 2.16E-02 | -2.98E-02 |
| 4472 | Syngr3 | SYNGR3 | -4.82E-02 | 4.99E-02 |
| 4473 | Setd1a | SETD1A | 2.50E-02 | -1.20E-02 |
| 4474 | Ell3 | ELL3 | -8.31E-02 | -6.62E-02 |
| 4475 | Gabra3 | GABRA3 | 1.63E-01 | -3.34E-02 |
| 4476 | Kcnj15 | KCNJ15 | 4.45E-02 | -1.10E-02 |
| 4477 | Pla2g4b | PLA2G4B | -1.11E-01 | 2.40E-02 |
| 4478 | Fktn | FKTN | -2.79E-02 | -1.64E-02 |
| 4479 | Slc4a4 | SLC4A4 | 5.20E-02 | -2.39E-02 |
| 4480 | Zfp446 | ZNF446 | -3.08E-02 | -2.90E-02 |
| 4481 | Ctsl | CTSV | 3.26E-02 | -2.33E-03 |
| 4482 | Tubb6 | TUBB6 | 9.68E-02 | 1.08E-01 |
| 4483 | Nxpe4 | NXPE4 | 9.03E-02 | -1.96E-02 |
| 4484 | Adsl | ADSL | -2.24E-02 | -1.20E-02 |
| 4485 | Msh6 | MSH6 | -2.21E-02 | 7.60E-04 |
| 4486 | Kif5c | KIF5C | 1.70E-01 | -5.57E-02 |
| 4487 | Frem2 | FREM2 | -8.27E-02 | -3.10E-02 |
| 4488 | Pcna | PCNA | 3.06E-02 | 4.27E-02 |
| 4489 | Lrrn4 | LRRN4 | -4.78E-02 | 2.76E-02 |
| 4490 | Nus1 | NUS1 | 3.72E-02 | -1.15E-02 |
| 4491 | Pstk | PSTK | -3.46E-02 | -2.52E-03 |
| 4492 | Gnb4 | GNB4 | -6.69E-02 | 6.27E-02 |
| 4493 | Etv6 | ETV6 | -3.02E-02 | 1.22E-04 |
| 4494 | Ckap2l | CKAP2L | 2.16E-01 | 1.64E-01 |
| 4495 | Acmsd | ACMSD | 7.75E-02 | -4.82E-02 |
| 4496 | Igsf1 | IGSF1 | -4.16E-02 | 2.59E-02 |
| 4497 | Vcl | VCL | 2.53E-02 | 1.82E-02 |
| 4498 | Arl8a | ARL8A | 1.96E-02 | -1.07E-02 |
| 4499 | Tmem231 | TMEM231 | -3.01E-02 | -1.11E-02 |
| 4500 | Ckmt1 | CKMT1B | 3.07E-02 | 4.18E-02 |
| 4501 | Ckmt1 | CKMT1A | 3.07E-02 | 4.18E-02 |
| 4502 | Ccnb1 | CCNB1 | 2.43E-01 | 1.75E-01 |

| | | | | |
|---|---|---|---|---|
| 4503 | Apc | APC | 1.92E-02 | -1.65E-02 |
| 4504 | Dusp6 | DUSP6 | -4.25E-02 | 5.95E-03 |
| 4505 | Mfap1b | MFAP1 | -3.28E-02 | 4.29E-03 |
| 4506 | Rspry1 | RSPRY1 | -2.43E-02 | 2.53E-03 |
| 4507 | Lmod3 | LMOD3 | 7.46E-02 | 2.08E-02 |
| 4508 | Gapt | GAPT | 5.05E-02 | -5.06E-02 |
| 4509 | Pes1 | PES1 | 2.21E-02 | 6.90E-03 |
| 4510 | Prickle3 | PRICKLE3 | -4.52E-02 | 3.49E-02 |
| 4511 | Zfp280c | ZNF280C | 5.03E-02 | -2.97E-04 |
| 4512 | Ube3a | UBE3A | -2.37E-02 | 6.04E-04 |
| 4513 | Arpc3 | ARPC3 | 2.11E-02 | 1.30E-02 |
| 4514 | Pdha1 | PDHA1 | 2.59E-02 | -1.47E-02 |
| 4515 | Rbm22 | RBM22 | 1.89E-02 | -5.59E-03 |
| 4516 | Nxph4 | NXPH4 | 1.05E-01 | -1.27E-02 |
| 4517 | Akap12 | AKAP12 | 1.16E-01 | 8.73E-02 |
| 4518 | Zfp365 | ZNF365 | 6.22E-02 | 3.10E-03 |
| 4519 | Map4k1 | MAP4K1 | -7.50E-02 | 4.62E-02 |
| 4520 | Prim2 | PRIM2 | -5.56E-02 | 1.37E-02 |
| 4521 | Cenpe | CENPE | 1.68E-01 | 1.38E-01 |
| 4522 | Rnf183 | RNF183 | 5.84E-02 | 3.72E-03 |
| 4523 | Mitf | MITF | -3.31E-02 | -1.91E-02 |
| 4524 | Ubtd1 | UBTD1 | 4.00E-02 | 1.84E-02 |
| 4525 | Hic2 | HIC2 | -5.62E-02 | 4.74E-02 |
| 4526 | Zfp566 | ZNF566 | -7.36E-02 | 3.57E-02 |
| 4527 | Zfp629 | ZNF629 | -4.29E-02 | -3.73E-03 |
| 4528 | Spink4 | SPINK4 | 7.41E-02 | -2.58E-02 |
| 4529 | Hnmt | HNMT | -3.99E-02 | -1.32E-02 |
| 4530 | Ywhab | YWHAB | 1.82E-02 | 2.25E-05 |
| 4531 | Sash1 | SASH1 | 3.44E-02 | 1.42E-03 |
| 4532 | P2rx4 | P2RX4 | 2.85E-02 | 2.22E-05 |
| 4533 | Ralgps1 | RALGPS1 | 3.64E-02 | -3.62E-02 |
| 4534 | Esf1 | ESF1 | -2.42E-02 | 4.88E-03 |
| 4535 | 4833439L19Rik | KIAA1191 | 3.67E-02 | -7.80E-03 |
| 4536 | C6 | C6 | 1.01E-01 | 1.10E-01 |
| 4537 | Tmem42 | TMEM42 | 4.36E-02 | 7.59E-03 |
| 4538 | Acad10 | ACAD10 | -3.46E-02 | -1.30E-02 |
| 4539 | Trim31 | TRIM31 | -2.81E-02 | 1.67E-04 |
| 4540 | Ddx19a | DDX19A | 2.03E-02 | 5.35E-03 |
| 4541 | Tmem260 | TMEM260 | -2.54E-02 | -1.33E-03 |
| 4542 | Mras | MRAS | 5.14E-02 | -1.31E-02 |
| 4543 | Tomm34 | TOMM34 | -2.74E-02 | -1.77E-02 |
| 4544 | Ndufs2 | NDUFS2 | 2.35E-02 | -1.91E-02 |
| 4545 | Vwa5b1 | VWA5B1 | 3.05E-02 | -1.76E-02 |
| 4546 | Fgf2 | FGF2 | 1.11E-01 | 7.98E-02 |
| 4547 | Necab1 | NECAB1 | 3.93E-02 | -7.75E-03 |
| 4548 | Rassf2 | RASSF2 | 7.77E-02 | 5.63E-02 |
| 4549 | Them4 | THEM4 | 3.91E-02 | 1.03E-03 |
| 4550 | Ddo | DDO | -4.43E-02 | -1.59E-02 |
| 4551 | Atp6v1g2 | ATP6V1G2 | -5.13E-02 | 6.83E-02 |
| 4552 | St3gal3 | ST3GAL3 | 2.73E-02 | -9.37E-03 |
| 4553 | Scfd1 | SCFD1 | 2.26E-02 | -1.76E-02 |
| 4554 | Lcmt1 | LCMT1 | 2.27E-02 | -5.00E-03 |
| 4555 | Emc2 | EMC2 | 2.11E-02 | -1.08E-02 |

| | | | | |
|---|---|---|---|---|
| 4556 | Sgk2 | SGK2 | -2.77E-02 | -2.59E-02 |
| 4557 | Ostm1 | OSTM1 | 2.43E-02 | -3.45E-03 |
| 4558 | Gm527 | C14orf28 | -7.58E-02 | 3.87E-02 |
| 4559 | Kctd12 | KCTD12 | -2.90E-02 | 7.86E-03 |
| 4560 | Srrt | SRRT | 1.53E-02 | -2.80E-03 |
| 4561 | Mapk1 | MAPK1 | 2.08E-02 | -8.42E-03 |
| 4562 | Ccny | CCNY | 1.80E-02 | -6.61E-03 |
| 4563 | Wif1 | WIF1 | -1.20E-01 | 6.33E-02 |
| 4564 | Plk3 | PLK3 | 2.63E-01 | 7.20E-03 |
| 4565 | Elp3 | ELP3 | -1.88E-02 | -1.37E-03 |
| 4566 | Kcnn1 | KCNN1 | -6.94E-02 | -7.18E-02 |
| 4567 | Pias4 | PIAS4 | -2.19E-02 | -1.39E-02 |
| 4568 | Rsu1 | RSU1 | 1.62E-02 | -7.25E-03 |
| 4569 | Naa20 | NAA20 | -1.88E-02 | 3.22E-03 |
| 4570 | Prima1 | PRIMA1 | 9.21E-02 | -3.42E-02 |
| 4571 | Tbc1d2b | TBC1D2B | -5.27E-02 | 2.64E-02 |
| 4572 | Zkscan8 | ZKSCAN8 | -2.91E-02 | -9.56E-03 |
| 4573 | Capn2 | CAPN2 | 3.47E-02 | -5.57E-03 |
| 4574 | Grid1 | GRID1 | -1.23E-01 | -9.45E-02 |
| 4575 | Kdm6a | KDM6A | -2.54E-02 | -6.88E-03 |
| 4576 | Tuba1a | TUBA1A | 4.94E-02 | 1.16E-01 |
| 4577 | Tspan9 | TSPAN9 | 2.15E-02 | -1.72E-02 |
| 4578 | Extl3 | EXTL3 | 1.70E-02 | -1.60E-02 |
| 4579 | Igsf9b | IGSF9B | -1.06E-01 | -4.32E-02 |
| 4580 | Rcor1 | RCOR1 | -2.42E-02 | 1.53E-03 |
| 4581 | Agfg1 | AGFG1 | 1.96E-02 | 1.19E-02 |
| 4582 | Cnst | CNST | -3.01E-02 | -2.71E-03 |
| 4583 | Sh3bp5l | SH3BP5L | -2.35E-02 | -2.81E-03 |
| 4584 | Slit3 | SLIT3 | -8.91E-02 | 5.31E-02 |
| 4585 | AB124611 | C19orf38 | -1.05E-01 | 1.27E-01 |
| 4586 | Hax1 | HAX1 | -3.01E-02 | -1.06E-02 |
| 4587 | Purg | PURG | -8.15E-02 | 4.62E-02 |
| 4588 | Tldc2 | TLDC2 | -1.07E-01 | 1.47E-01 |
| 4589 | Qdpr | QDPR | 3.49E-02 | -1.76E-02 |
| 4590 | Lcmt2 | LCMT2 | 3.53E-02 | -8.26E-03 |
| 4591 | Dnah7c | DNAH7 | -7.38E-02 | -5.30E-02 |
| 4592 | Klf9 | KLF9 | 4.61E-02 | -3.81E-02 |
| 4593 | Rassf8 | RASSF8 | 1.78E-02 | -1.31E-02 |
| 4594 | Mapk6 | MAPK6 | 3.48E-02 | 2.74E-02 |
| 4595 | Hsd3b3 | HSD3B1 | 9.83E-02 | 1.12E-02 |
| 4596 | Mad1l1 | MAD1L1 | 4.61E-02 | 8.62E-03 |
| 4597 | Comt | COMT | 6.05E-02 | -1.86E-02 |
| 4598 | Cc2d1a | CC2D1A | 2.29E-02 | -9.58E-03 |
| 4599 | Fras1 | FRAS1 | -4.24E-02 | -4.13E-02 |
| 4600 | Nsmaf | NSMAF | 1.82E-02 | -1.38E-02 |
| 4601 | Jrkl | JRKL | -3.98E-02 | 6.62E-03 |
| 4602 | Atp8a2 | ATP8A2 | -7.91E-02 | 3.44E-02 |
| 4603 | Soat2 | SOAT2 | 1.00E-01 | -3.02E-02 |
| 4604 | Sgip1 | SGIP1 | -1.14E-01 | -3.76E-02 |
| 4605 | Snx16 | SNX16 | -5.09E-02 | 3.89E-02 |
| 4606 | Lax1 | LAX1 | 1.06E-01 | 2.29E-02 |
| 4607 | Rnd3 | RND3 | 4.68E-02 | 5.64E-02 |
| 4608 | Sap30bp | SAP30BP | 1.84E-02 | -5.06E-03 |

| | | | | |
|---|---|---|---|---|
| 4609 | Zik1 | ZIK1 | -8.80E-02 | 9.19E-03 |
| 4610 | Esrp2 | ESRP2 | 2.36E-02 | -1.46E-02 |
| 4611 | Polr2k | POLR2K | -3.43E-02 | -8.24E-03 |
| 4612 | Gclm | GCLM | -5.45E-02 | -1.69E-02 |
| 4613 | Nxn | NXN | -4.05E-02 | -1.07E-02 |
| 4614 | Dbt | DBT | -3.54E-02 | -2.41E-02 |
| 4615 | Nrros | NRROS | -7.45E-02 | 6.58E-02 |
| 4616 | Snrpb | SNRPB | 1.94E-02 | -1.05E-02 |
| 4617 | Rsf1 | RSF1 | -2.16E-02 | -2.07E-03 |
| 4618 | Smad2 | SMAD2 | 2.27E-02 | -1.08E-03 |
| 4619 | Scn4a | SCN4A | -7.71E-02 | -3.65E-02 |
| 4620 | Fbxo7 | FBXO7 | 2.91E-02 | -1.86E-02 |
| 4621 | Rhbdf1 | RHBDF1 | 3.59E-02 | 7.15E-03 |
| 4622 | Acot12 | ACOT12 | 5.26E-02 | -9.75E-03 |
| 4623 | Wscd2 | WSCD2 | -7.88E-02 | -1.75E-02 |
| 4624 | Polg2 | POLG2 | -3.98E-02 | -5.35E-03 |
| 4625 | H6pd | H6PD | 4.39E-02 | 1.41E-02 |
| 4626 | E2f4 | E2F4 | -1.93E-02 | 4.39E-03 |
| 4627 | Apold1 | APOLD1 | 1.91E-01 | 9.26E-02 |
| 4628 | Tmem234 | TMEM234 | 1.62E-02 | -7.75E-03 |
| 4629 | Htr7 | HTR7 | -2.83E-02 | 3.45E-02 |
| 4630 | Brip1 | BRIP1 | 8.30E-02 | -1.31E-02 |
| 4631 | Pdlim4 | PDLIM4 | -7.76E-02 | 6.97E-03 |
| 4632 | Pygm | PYGM | -9.38E-02 | -3.41E-02 |
| 4633 | Nhlrc1 | NHLRC1 | -4.74E-02 | 1.12E-03 |
| 4634 | Haus2 | HAUS2 | 3.72E-02 | 1.05E-03 |
| 4635 | Dusp16 | DUSP16 | 3.38E-02 | 9.28E-03 |
| 4636 | Ttc39a | TTC39A | 9.87E-02 | -1.76E-02 |
| 4637 | Itga2b | ITGA2B | -4.87E-02 | -2.14E-02 |
| 4638 | E2f2 | E2F2 | 1.02E-01 | 1.99E-02 |
| 4639 | Lrtm2 | LRTM2 | -9.05E-02 | 7.88E-02 |
| 4640 | Tnfrsf11b | TNFRSF11B | -9.03E-02 | 7.69E-02 |
| 4641 | Orai1 | ORAI1 | -3.15E-02 | -2.13E-02 |
| 4642 | Eri2 | ERI2 | 5.99E-02 | 1.70E-02 |
| 4643 | Stxbp6 | STXBP6 | -3.83E-02 | -4.00E-02 |
| 4644 | Klk1b11 | KLK3 | 7.98E-02 | 9.28E-02 |
| 4645 | Pgm3 | PGM3 | -3.34E-02 | -2.51E-02 |
| 4646 | Rbck1 | RBCK1 | -1.80E-02 | -1.45E-02 |
| 4647 | Slc6a12 | SLC6A12 | 3.85E-01 | -1.13E-01 |
| 4648 | Cmtr1 | CMTR1 | -4.88E-02 | 7.86E-03 |
| 4649 | Kcnab3 | KCNAB3 | 7.67E-02 | -3.86E-02 |
| 4650 | 1700093K21Rik | C2orf74 | 4.89E-02 | -2.27E-02 |
| 4651 | Snx2 | SNX2 | 1.83E-02 | -5.68E-03 |
| 4652 | Aspn | ASPN | 1.78E-01 | -5.25E-02 |
| 4653 | Tprgl | TPRG1L | 1.88E-02 | -1.76E-02 |
| 4654 | Tpst1 | TPST1 | -6.31E-02 | -8.15E-03 |
| 4655 | Gdpd2 | GDPD2 | -7.36E-02 | -6.98E-02 |
| 4656 | Aggf1 | AGGF1 | -2.29E-02 | 5.76E-03 |
| 4657 | C87436 | C2orf42 | 2.62E-02 | -1.82E-02 |
| 4658 | Mbd3 | MBD3 | 1.88E-02 | -4.59E-03 |
| 4659 | Cdk2 | CDK2 | 3.64E-02 | 1.29E-02 |
| 4660 | Slc12a4 | SLC12A4 | 3.22E-02 | 1.18E-02 |
| 4661 | Shpk | SHPK | 2.92E-02 | -1.33E-02 |

| | | | | |
|---|---|---|---|---|
| 4662 | Bcl2l14 | BCL2L14 | 1.13E-01 | 2.81E-02 |
| 4663 | Tsn | TSN | 1.71E-02 | -4.91E-03 |
| 4664 | Trpc4 | TRPC4 | -1.19E-01 | 1.56E-02 |
| 4665 | Crygs | CRYGS | -5.19E-02 | -2.26E-02 |
| 4666 | Notch4 | NOTCH4 | 2.79E-02 | 3.04E-03 |
| 4667 | Pds5b | PDS5B | -2.71E-02 | -2.27E-03 |
| 4668 | Ctdspl | CTDSPL | -3.24E-02 | -4.66E-02 |
| 4669 | Dync1li2 | DYNC1LI2 | 1.53E-02 | -8.04E-03 |
| 4670 | Otud4 | OTUD4 | -2.43E-02 | 1.89E-04 |
| 4671 | Fam160a1 | FAM160A1 | 4.13E-02 | -2.63E-02 |
| 4672 | Galnt12 | GALNT12 | -6.06E-02 | -6.47E-03 |
| 4673 | Nrap | NRAP | 1.34E-01 | -2.27E-01 |
| 4674 | Klrc1 | KLRC1 | -5.34E-02 | 2.32E-02 |
| 4675 | Spag1 | SPAG1 | -7.88E-02 | 3.15E-02 |
| 4676 | Nlrx1 | NLRX1 | 4.12E-02 | -2.58E-02 |
| 4677 | Cd226 | CD226 | -8.32E-02 | 4.35E-02 |
| 4678 | Plekhf2 | PLEKHF2 | -2.94E-02 | 4.67E-03 |
| 4679 | Rcn1 | RCN1 | 4.60E-02 | 2.91E-02 |
| 4680 | Kif6 | KIF6 | 7.75E-02 | 6.05E-02 |
| 4681 | Zc2hc1c | ZC2HC1C | 4.33E-02 | -3.69E-02 |
| 4682 | Ruvbl1 | RUVBL1 | -2.38E-02 | 6.51E-03 |
| 4683 | Dusp18 | DUSP18 | 4.40E-02 | -2.01E-02 |
| 4684 | Ugt1a6b | UGT1A6 | 8.44E-02 | 2.44E-02 |
| 4685 | Prelid1 | PRELID1 | 2.69E-02 | -1.12E-02 |
| 4686 | Ambra1 | AMBRA1 | -1.93E-02 | -1.21E-02 |
| 4687 | Cxcl17 | CXCL17 | 5.83E-02 | 1.72E-01 |
| 4688 | Picalm | PICALM | 2.82E-02 | -6.37E-03 |
| 4689 | Hoxa3 | HOXA3 | -3.68E-02 | -1.39E-02 |
| 4690 | Ctsa | CTSA | 3.36E-02 | -2.81E-03 |
| 4691 | D3Ertd751e | C4orf33 | -3.25E-02 | -2.02E-02 |
| 4692 | Chchd10 | CHCHD10 | 2.62E-02 | -1.65E-02 |
| 4693 | Sbf1 | SBF1 | 2.62E-02 | -5.05E-03 |
| 4694 | Slc4a2 | SLC4A2 | 1.55E-02 | -9.01E-03 |
| 4695 | Igf2r | IGF2R | 3.85E-02 | -2.23E-02 |
| 4696 | Spast | SPAST | 2.66E-02 | 8.15E-03 |
| 4697 | 1700001C19Rik | LOC102723903 | 5.17E-02 | -2.22E-02 |
| 4698 | Adcy3 | ADCY3 | -1.14E-01 | -1.33E-02 |
| 4699 | Prpf38b | PRPF38B | 1.90E-02 | -4.92E-03 |
| 4700 | Slc38a9 | SLC38A9 | 2.33E-02 | 5.97E-03 |
| 4701 | Fbrs | FBRS | -1.78E-02 | 5.97E-03 |
| 4702 | Pla2g4c | PLA2G4C | -8.61E-02 | 6.07E-02 |
| 4703 | Rai2 | RAI2 | 5.04E-02 | -2.20E-02 |
| 4704 | Naa25 | NAA25 | 2.08E-02 | 3.50E-03 |
| 4705 | Tpk1 | TPK1 | -2.34E-02 | -2.58E-02 |
| 4706 | Phldb1 | PHLDB1 | -1.95E-02 | 1.43E-02 |
| 4707 | Zfp1 | ZFP1 | 3.86E-02 | 3.51E-03 |
| 4708 | Ak6 | AK6 | 3.31E-02 | 2.93E-02 |
| 4709 | Creb3l3 | CREB3L3 | 1.98E-01 | 5.49E-02 |
| 4710 | Mpzl1 | MPZL1 | 6.64E-02 | -1.50E-02 |
| 4711 | Zranb3 | ZRANB3 | -5.03E-02 | 4.48E-02 |
| 4712 | Atg9a | ATG9A | 3.57E-02 | -3.39E-02 |
| 4713 | Atrn | ATRN | 1.95E-02 | -2.42E-02 |
| 4714 | Ccdc25 | CCDC25 | 2.87E-02 | 5.28E-03 |

| | | | | |
|---|---|---|---|---|
| 4715 | Kctd17 | KCTD17 | 4.71E-02 | 1.06E-02 |
| 4716 | Wtip | WTIP | -3.98E-02 | -2.54E-05 |
| 4717 | Zbtb42 | ZBTB42 | 7.18E-02 | 2.59E-02 |
| 4718 | Kdm2b | KDM2B | -3.32E-02 | 1.26E-02 |
| 4719 | Trim62 | TRIM62 | 4.55E-02 | 1.36E-02 |
| 4720 | Mertk | MERTK | 3.91E-02 | -2.64E-02 |
| 4721 | Twf2 | TWF2 | -3.65E-02 | 1.72E-02 |
| 4722 | Dmpk | DMPK | 7.52E-02 | 7.01E-02 |
| 4723 | Nfrkb | NFRKB | 2.22E-02 | -1.50E-02 |
| 4724 | Bsn | BSN | 6.91E-02 | -6.70E-03 |
| 4725 | Arhgef10 | ARHGEF10 | -3.71E-02 | 3.04E-02 |
| 4726 | Dnajc4 | DNAJC4 | 3.22E-02 | -2.14E-02 |
| 4727 | Mrps28 | MRPS28 | 3.10E-02 | -1.64E-02 |
| 4728 | Eif4b | EIF4B | -2.93E-02 | 1.59E-02 |
| 4729 | Abhd5 | ABHD5 | 1.95E-02 | 5.61E-03 |
| 4730 | Zzef1 | ZZEF1 | 2.01E-02 | -1.14E-02 |
| 4731 | Hexb | HEXB | -2.77E-02 | -1.27E-02 |
| 4732 | Ldhb | LDHB | 2.40E-02 | -1.18E-02 |
| 4733 | Acox1 | ACOX1 | 5.42E-02 | 7.62E-03 |
| 4734 | Pcgf1 | PCGF1 | -3.46E-02 | -2.26E-02 |
| 4735 | Smu1 | SMU1 | 1.71E-02 | -1.40E-02 |
| 4736 | Epdr1 | EPDR1 | 5.85E-02 | -2.00E-02 |
| 4737 | Pxdc1 | PXDC1 | 8.28E-02 | 7.48E-02 |
| 4738 | Pi16 | PI16 | 4.98E-02 | 1.82E-02 |
| 4739 | Ptk2b | PTK2B | -3.54E-02 | -3.89E-02 |
| 4740 | Rnf7 | RNF7 | 1.74E-02 | -1.46E-02 |
| 4741 | Crybb1 | CRYBB1 | 3.66E-02 | -1.27E-02 |
| 4742 | Bcl7a | BCL7A | 2.66E-02 | -1.04E-02 |
| 4743 | Atl2 | ATL2 | 2.24E-02 | -3.49E-04 |
| 4744 | Zfp710 | ZNF710 | 3.32E-02 | -8.99E-04 |
| 4745 | Dtd2 | DTD2 | 2.85E-02 | -1.14E-02 |
| 4746 | Tfec | TFEC | -5.80E-02 | 8.86E-02 |
| 4747 | Ash2l | ASH2L | 1.79E-02 | -4.59E-03 |
| 4748 | Arhgef15 | ARHGEF15 | -5.87E-02 | -2.04E-02 |
| 4749 | Mmd | MMD | -6.36E-02 | 4.22E-02 |
| 4750 | E2f1 | E2F1 | 9.75E-02 | 1.41E-01 |
| 4751 | Cflar | CFLAR | 2.57E-02 | -1.59E-02 |
| 4752 | 3110040N11Rik | C15orf40 | -2.99E-02 | 1.17E-02 |
| 4753 | Mfhas1 | MFHAS1 | -2.27E-02 | 1.20E-02 |
| 4754 | Adam33 | ADAM33 | -6.97E-02 | -2.93E-02 |
| 4755 | Tmco6 | TMCO6 | -4.20E-02 | 2.52E-03 |
| 4756 | Skap2 | SKAP2 | 1.93E-02 | -4.94E-03 |
| 4757 | Mtmr9 | MTMR9 | 2.95E-02 | -2.20E-02 |
| 4758 | Prx | PRX | 7.07E-02 | -6.31E-02 |
| 4759 | Thumpd1 | THUMPD1 | 2.35E-02 | 8.07E-03 |
| 4760 | Ppp1r21 | PPP1R21 | 2.83E-02 | -1.85E-02 |
| 4761 | Rap2c | RAP2C | 3.15E-02 | 1.38E-02 |
| 4762 | Dpp4 | DPP4 | -3.69E-02 | -4.78E-02 |
| 4763 | Chrna4 | CHRNA4 | 6.44E-02 | -5.33E-02 |
| 4764 | Tk1 | TK1 | 1.32E-01 | 8.19E-02 |
| 4765 | Dhx34 | DHX34 | -3.69E-02 | -1.15E-02 |
| 4766 | F7 | F7 | -4.65E-02 | 2.72E-02 |
| 4767 | Mier2 | MIER2 | -2.08E-02 | -6.80E-03 |

| | | | | |
|---|---|---|---|---|
| 4768 | Rwdd1 | RWDD1 | 2.37E-02 | 1.27E-02 |
| 4769 | Zfp523 | ZNF76 | 2.29E-02 | -1.52E-02 |
| 4770 | Btbd17 | BTBD17 | 3.14E-02 | 9.76E-03 |
| 4771 | Hcn2 | HCN2 | 1.13E-01 | -2.55E-02 |
| 4772 | Phactr2 | PHACTR2 | -2.24E-02 | -1.15E-02 |
| 4773 | Adcy7 | ADCY7 | -6.66E-02 | 1.33E-01 |
| 4774 | Ercc5 | ERCC5 | -2.46E-02 | 7.88E-03 |
| 4775 | Calu | CALU | 1.72E-02 | 5.74E-03 |
| 4776 | Tmem178 | TMEM178A | 4.74E-02 | 4.39E-03 |
| 4777 | Cmas | CMAS | 2.13E-02 | -3.55E-03 |
| 4778 | Prkcg | PRKCG | 6.83E-02 | -9.20E-03 |
| 4779 | Txndc17 | TXNDC17 | 2.96E-02 | -3.58E-03 |
| 4780 | Gng12 | GNG12 | 2.12E-02 | -5.22E-03 |
| 4781 | Dcaf7 | DCAF7 | 1.53E-02 | -1.70E-02 |
| 4782 | Cd38 | CD38 | -5.54E-02 | 8.88E-03 |
| 4783 | Pik3r5 | PIK3R5 | -1.19E-01 | 1.88E-01 |
| 4784 | Nat14 | NAT14 | -4.84E-02 | 7.46E-03 |
| 4785 | Arf4 | ARF4 | 1.71E-02 | -4.37E-04 |
| 4786 | Usp14 | USP14 | 1.84E-02 | 9.40E-03 |
| 4787 | Rala | RALA | 2.22E-02 | -8.05E-03 |
| 4788 | Mllt1 | MLLT1 | -1.84E-02 | 7.54E-03 |
| 4789 | Btbd2 | BTBD2 | -2.03E-02 | -1.53E-02 |
| 4790 | Cybb | CYBB | -1.05E-01 | 1.46E-01 |
| 4791 | Slc35f1 | SLC35F1 | -8.26E-02 | -5.10E-02 |
| 4792 | Pcsk7 | PCSK7 | 2.16E-02 | -1.57E-02 |
| 4793 | Mrpl38 | MRPL38 | 2.51E-02 | -8.28E-03 |
| 4794 | Eif5b | EIF5B | 1.73E-02 | 5.25E-03 |
| 4795 | Per1 | PER1 | 9.11E-02 | -4.62E-02 |
| 4796 | Stambpl1 | STAMBPL1 | -6.07E-02 | 7.02E-02 |
| 4797 | Cnbd1 | CNBD1 | -3.84E-02 | -1.19E-03 |
| 4798 | Myo1e | MYO1E | -2.57E-02 | 1.82E-02 |
| 4799 | Cndp2 | CNDP2 | -3.37E-02 | 3.14E-03 |
| 4800 | Dcaf17 | DCAF17 | -2.92E-02 | 6.72E-03 |
| 4801 | Lrrc58 | LRRC58 | 2.95E-02 | -2.01E-02 |
| 4802 | Rad51 | RAD51 | 1.43E-01 | 1.41E-01 |
| 4803 | Naa40 | NAA40 | 2.63E-02 | -4.19E-03 |
| 4804 | Serpind1 | SERPIND1 | -8.80E-02 | -4.71E-02 |
| 4805 | Fkbp1b | FKBP1B | 8.83E-02 | 5.53E-02 |
| 4806 | Keap1 | KEAP1 | 1.81E-02 | 1.02E-03 |
| 4807 | Agbl4 | AGBL4 | 4.65E-02 | 1.81E-02 |
| 4808 | Gtf2e2 | GTF2E2 | -3.02E-02 | 1.74E-02 |
| 4809 | Letm2 | LETM2 | -3.72E-02 | -1.89E-02 |
| 4810 | Lsp1 | LSP1 | -6.10E-02 | 1.46E-01 |
| 4811 | Sarm1 | SARM1 | -7.51E-02 | 9.28E-03 |
| 4812 | Tprg | TPRG1 | -3.17E-02 | 1.06E-02 |
| 4813 | Nipsnap1 | NIPSNAP1 | 3.53E-02 | -1.60E-02 |
| 4814 | Inpp4b | INPP4B | -8.48E-02 | 3.00E-02 |
| 4815 | Ndufb8 | NDUFB8 | 1.87E-02 | -2.25E-02 |
| 4816 | Trmt112 | TRMT112 | -3.00E-02 | -1.30E-02 |
| 4817 | Mfsd9 | MFSD9 | 3.61E-02 | -3.59E-02 |
| 4818 | Cd59b | CD59 | -1.34E-01 | -1.44E-02 |
| 4819 | Enpp1 | ENPP1 | -3.72E-02 | -5.18E-02 |
| 4820 | Sass6 | SASS6 | 6.20E-02 | 3.42E-02 |

| | | | | |
|---|---|---|---|---|
| 4821 | Cep97 | CEP97 | -3.06E-02 | 8.41E-03 |
| 4822 | Eml2 | EML2 | -3.18E-02 | 3.10E-03 |
| 4823 | Tcirg1 | TCIRG1 | 3.50E-02 | 2.46E-03 |
| 4824 | Tekt5 | TEKT5 | -5.17E-02 | -1.10E-02 |
| 4825 | Nktr | NKTR | 2.06E-02 | -1.27E-02 |
| 4826 | Mea1 | MEA1 | 1.79E-02 | -1.16E-02 |
| 4827 | Tln2 | TLN2 | -4.87E-02 | -6.12E-02 |
| 4828 | Sdhc | SDHC | 2.70E-02 | -1.44E-02 |
| 4829 | Selp | SELP | -1.02E-01 | 8.29E-02 |
| 4830 | Dpysl5 | DPYSL5 | -1.04E-01 | 4.38E-02 |
| 4831 | Acyp1 | ACYP1 | -3.02E-02 | -1.72E-02 |
| 4832 | Nfatc1 | NFATC1 | 3.17E-02 | -8.81E-03 |
| 4833 | Hmcn2 | HMCN2 | -1.01E-01 | -6.77E-02 |
| 4834 | Sesn3 | SESN3 | 2.89E-02 | -1.95E-02 |
| 4835 | Cntnap2 | CNTNAP2 | 1.10E-01 | 2.63E-03 |
| 4836 | Cdipt | CDIPT | 2.03E-02 | -7.55E-03 |
| 4837 | Rec8 | REC8 | -1.48E-01 | 1.66E-01 |
| 4838 | Strip1 | STRIP1 | 2.47E-02 | 1.44E-02 |
| 4839 | Lrp10 | LRP10 | 1.72E-02 | -3.49E-03 |
| 4840 | AW554918 | KIAA1328 | 3.36E-02 | -3.63E-02 |
| 4841 | Hspa9 | HSPA9 | 2.52E-02 | -2.10E-02 |
| 4842 | Zfp28 | ZFP28 | -3.61E-02 | -7.15E-03 |
| 4843 | Ccdc97 | CCDC97 | -1.76E-02 | -7.04E-03 |
| 4844 | Dcun1d2 | DCUN1D2 | -3.66E-02 | 1.49E-02 |
| 4845 | Suclg1 | SUCLG1 | 3.03E-02 | -2.06E-02 |
| 4846 | Galnt9 | GALNT9 | -7.98E-02 | -3.41E-02 |
| 4847 | Ing4 | ING4 | 7.59E-02 | -4.08E-02 |
| 4848 | Efr3a | EFR3A | 2.20E-02 | -1.30E-02 |
| 4849 | Evc | EVC | -3.28E-02 | 3.48E-03 |
| 4850 | Arpc1b | ARPC1B | 3.46E-02 | 5.70E-02 |
| 4851 | Brca2 | BRCA2 | 7.89E-02 | 2.86E-03 |
| 4852 | Pde9a | PDE9A | 5.29E-02 | -6.07E-02 |
| 4853 | Oxtr | OXTR | 7.74E-02 | 2.53E-02 |
| 4854 | Nkain4 | NKAIN4 | 5.37E-02 | -3.58E-02 |
| 4855 | Stx2 | STX2 | 2.07E-02 | 3.01E-03 |
| 4856 | Atp6v0b | ATP6V0B | 1.93E-02 | -4.93E-03 |
| 4857 | Srsf11 | SRSF11 | 2.12E-02 | 9.88E-03 |
| 4858 | Ltbp4 | LTBP4 | -7.01E-02 | -2.38E-03 |
| 4859 | Gjb3 | GJB3 | 1.34E-01 | 5.79E-03 |
| 4860 | Thop1 | THOP1 | 2.21E-02 | 7.11E-03 |
| 4861 | Kcnj16 | KCNJ16 | 3.34E-02 | -1.87E-02 |
| 4862 | Timm10b | TIMM10B | -6.90E-02 | 3.73E-02 |
| 4863 | Trp53i11 | TP53I11 | -1.39E-01 | 8.78E-02 |
| 4864 | Nfkbib | NFKBIB | 3.12E-02 | -3.05E-03 |
| 4865 | Zfp182 | ZNF182 | -6.38E-02 | 9.08E-03 |
| 4866 | Gdi1 | GDI1 | 1.66E-02 | -8.05E-03 |
| 4867 | Cadm3 | CADM3 | -4.90E-02 | 5.90E-02 |
| 4868 | Pard6b | PARD6B | -2.43E-02 | -2.32E-02 |
| 4869 | Plk1 | PLK1 | 1.91E-01 | 1.43E-01 |
| 4870 | Grsf1 | GRSF1 | 2.06E-02 | -2.32E-02 |
| 4871 | Agbl3 | AGBL3 | -2.40E-02 | -7.04E-03 |
| 4872 | Zfp511 | ZNF511 | -3.07E-02 | 9.13E-03 |
| 4873 | Pde7b | PDE7B | -5.02E-02 | -6.22E-02 |

| | | | | |
|---|---|---|---|---|
| 4874 | Mapkap1 | MAPKAP1 | 1.80E-02 | -2.51E-02 |
| 4875 | Nrf1 | NRF1 | -2.20E-02 | 1.35E-02 |
| 4876 | Camk1d | CAMK1D | 1.35E-01 | -3.15E-02 |
| 4877 | Rab5b | RAB5B | 2.01E-02 | -2.23E-02 |
| 4878 | Chgb | CHGB | 3.32E-02 | -1.38E-02 |
| 4879 | Alpk3 | ALPK3 | 7.00E-02 | -3.24E-02 |
| 4880 | Ajap1 | AJAP1 | 6.37E-02 | 2.60E-03 |
| 4881 | Larp1 | LARP1 | 1.91E-02 | -1.68E-03 |
| 4882 | Sycp2 | SYCP2 | -6.41E-02 | -2.29E-02 |
| 4883 | Txndc15 | TXNDC15 | -1.81E-02 | -2.63E-03 |
| 4884 | Uchl1 | UCHL1 | 1.12E-01 | 9.61E-02 |
| 4885 | Foxc2 | FOXC2 | -5.73E-02 | 1.29E-02 |
| 4886 | Fbxw11 | FBXW11 | 2.11E-02 | -1.40E-02 |
| 4887 | Stmnd1 | STMND1 | -5.14E-02 | 7.65E-03 |
| 4888 | Car7 | CA7 | 5.95E-02 | 1.46E-02 |
| 4889 | Pcdhgb6 | PCDHGB6 | -7.27E-02 | -2.01E-02 |
| 4890 | Fga | FGA | 7.85E-02 | 2.40E-01 |
| 4891 | Rhog | RHOG | 2.63E-02 | 2.04E-02 |
| 4892 | Cfb | CFB | -9.79E-02 | 7.64E-03 |
| 4893 | S100a9 | S100A9 | 2.30E-01 | 1.22E-01 |
| 4894 | Tbkbp1 | TBKBP1 | -5.43E-02 | -2.17E-02 |
| 4895 | Wasf1 | WASF1 | 6.40E-02 | 8.56E-03 |
| 4896 | Msto1 | MSTO1 | 3.17E-02 | -8.72E-03 |
| 4897 | Ctcfl | CTCFL | 8.61E-02 | -7.33E-03 |
| 4898 | Cd34 | CD34 | 4.00E-02 | -2.14E-02 |
| 4899 | Clec12b | CLEC12B | -6.76E-02 | 1.49E-02 |
| 4900 | Snapc1 | SNAPC1 | 4.24E-02 | -1.42E-02 |
| 4901 | Med7 | MED7 | 2.50E-02 | 2.76E-02 |
| 4902 | Supt7l | SUPT7L | -2.21E-02 | -1.42E-02 |
| 4903 | Zswim6 | ZSWIM6 | -3.81E-02 | 3.17E-02 |
| 4904 | Tmem87b | TMEM87B | 1.79E-02 | -1.36E-02 |
| 4905 | Slc16a8 | SLC16A8 | -4.41E-02 | 1.41E-02 |
| 4906 | Mrpl3 | MRPL3 | -2.40E-02 | 2.92E-03 |
| 4907 | Rab6a | RAB6A | 1.74E-02 | 7.39E-03 |
| 4908 | Sdf2 | SDF2 | -1.70E-02 | 1.00E-02 |
| 4909 | Ctdsp2 | CTDSP2 | -2.95E-02 | -1.31E-02 |
| 4910 | Gca | GCA | -3.25E-02 | 4.74E-02 |
| 4911 | Glg1 | GLG1 | 2.24E-02 | -5.84E-03 |
| 4912 | Eefsec | EEFSEC | 3.71E-02 | -1.90E-02 |
| 4913 | Hook2 | HOOK2 | 3.08E-02 | -2.60E-02 |
| 4914 | Ippk | IPPK | 2.93E-02 | 1.24E-02 |
| 4915 | Clk3 | CLK3 | 2.51E-02 | 8.52E-03 |
| 4916 | Ssbp3 | SSBP3 | -2.28E-02 | -1.13E-02 |
| 4917 | Creg1 | CREG1 | 4.75E-02 | -4.01E-03 |
| 4918 | 9130401M01Rik | C8orf76 | 6.03E-02 | 1.62E-02 |
| 4919 | AI837181 | C11orf68 | 2.08E-02 | 4.78E-03 |
| 4920 | Gabrd | GABRD | -7.42E-02 | 5.45E-02 |
| 4921 | Dot1l | DOT1L | 2.15E-02 | 1.24E-02 |
| 4922 | Phospho1 | PHOSPHO1 | 7.01E-02 | -3.66E-02 |
| 4923 | Calml3 | CALML3 | 1.86E-01 | 1.07E-01 |
| 4924 | Sema4a | SEMA4A | -3.65E-02 | -2.04E-02 |
| 4925 | Foxi1 | FOXI1 | -8.65E-02 | 1.37E-02 |
| 4926 | Ptx3 | PTX3 | -5.21E-02 | 7.59E-03 |

| | | | | |
|---|---|---|---|---|
| 4927 | Wbp2 | WBP2 | 2.62E-02 | -7.84E-03 |
| 4928 | Cdh13 | CDH13 | -8.41E-02 | -2.05E-02 |
| 4929 | Sik2 | SIK2 | 2.73E-02 | -1.50E-03 |
| 4930 | Bet1l | BET1L | -3.45E-02 | -6.53E-03 |
| 4931 | Abcg5 | ABCG5 | 3.87E-02 | -1.93E-02 |
| 4932 | Fuca1 | FUCA1 | 3.31E-02 | -4.24E-03 |
| 4933 | Ercc4 | ERCC4 | 2.28E-02 | -1.90E-02 |
| 4934 | Scamp1 | SCAMP1 | 2.38E-02 | -2.44E-02 |
| 4935 | Alg3 | ALG3 | 2.76E-02 | 9.29E-03 |
| 4936 | Primpol | PRIMPOL | -3.79E-02 | -3.84E-02 |
| 4937 | Map7d1 | MAP7D1 | 2.41E-02 | 3.62E-02 |
| 4938 | Sgsm1 | SGSM1 | -5.60E-02 | 5.15E-03 |
| 4939 | Prcc | PRCC | -1.57E-02 | -8.17E-03 |
| 4940 | Slc13a5 | SLC13A5 | -7.26E-02 | -8.32E-02 |
| 4941 | Tagln3 | TAGLN3 | -8.28E-02 | 3.97E-02 |
| 4942 | Mtx2 | MTX2 | 1.97E-02 | -6.95E-03 |
| 4943 | Tnrc6c | TNRC6C | -2.27E-02 | -7.08E-03 |
| 4944 | Grip1 | GRIP1 | 7.10E-02 | -9.09E-02 |
| 4945 | Park7 | PARK7 | 2.63E-02 | -1.22E-02 |
| 4946 | Chst1 | CHST1 | -7.25E-02 | 2.95E-02 |
| 4947 | Kmo | KMO | -5.05E-02 | -1.92E-02 |
| 4948 | Lcp1 | LCP1 | -7.39E-02 | 1.67E-02 |
| 4949 | Abca8b | ABCA8 | 8.41E-02 | -3.47E-03 |
| 4950 | Emc4 | EMC4 | 1.95E-02 | -6.63E-03 |
| 4951 | Nup214 | NUP214 | 2.27E-02 | -2.75E-02 |
| 4952 | Procr | PROCR | -7.28E-02 | 9.26E-02 |
| 4953 | Rai14 | RAI14 | -2.07E-02 | -1.13E-02 |
| 4954 | Calcr | CALCR | -8.66E-02 | 9.28E-02 |
| 4955 | Sgk3 | C8orf44-SGK3 | -2.55E-02 | 9.93E-03 |
| 4956 | Sgk3 | SGK3 | -2.55E-02 | 9.93E-03 |
| 4957 | Pdlim1 | PDLIM1 | 7.69E-02 | 9.59E-02 |
| 4958 | Elac1 | ELAC1 | -2.58E-02 | -3.07E-02 |
| 4959 | C8a | C8A | -2.67E-01 | -3.03E-01 |
| 4960 | Zfp750 | ZNF750 | -7.04E-02 | -2.77E-02 |
| 4961 | Bhmt | BHMT | 1.12E-01 | -8.91E-02 |
| 4962 | Ikzf2 | IKZF2 | -7.36E-02 | -5.04E-03 |
| 4963 | Bglap3 | BGLAP | -1.18E-01 | -6.89E-02 |
| 4964 | Psmb4 | PSMB4 | 2.09E-02 | -2.40E-03 |
| 4965 | Nsun4 | NSUN4 | 2.80E-02 | -1.34E-02 |
| 4966 | Siglec1 | SIGLEC1 | -1.20E-01 | 1.85E-01 |
| 4967 | Phb | PHB | 1.89E-02 | -2.36E-02 |
| 4968 | Shmt1 | SHMT1 | -2.55E-02 | -9.00E-03 |
| 4969 | Ccdc138 | CCDC138 | -4.57E-02 | 2.80E-02 |
| 4970 | Celsr1 | CELSR1 | -2.60E-02 | -2.44E-02 |
| 4971 | Cd247 | CD247 | -7.64E-02 | 1.70E-02 |
| 4972 | Alkbh2 | ALKBH2 | -5.08E-02 | 3.40E-02 |
| 4973 | Iqch | IQCH | -2.74E-02 | 2.61E-02 |
| 4974 | Glb1 | GLB1 | -3.28E-02 | -6.32E-03 |
| 4975 | Rtf1 | RTF1 | 1.55E-02 | -9.18E-03 |
| 4976 | Tor1b | TOR1B | 1.72E-02 | -1.09E-02 |
| 4977 | Adipor2 | ADIPOR2 | 1.72E-02 | -5.45E-03 |
| 4978 | Ttc9 | TTC9 | 9.10E-02 | -3.05E-02 |
| 4979 | Pphln1 | PPHLN1 | 1.99E-02 | -2.34E-03 |

| | | | | |
|---|---|---|---|---|
| 4980 | Strbp | STRBP | 1.97E-02 | -3.87E-03 |
| 4981 | Per2 | PER2 | 5.92E-02 | 8.42E-03 |
| 4982 | Dennd5a | DENND5A | 1.86E-02 | 9.89E-04 |
| 4983 | Ccdc22 | CCDC22 | 7.76E-02 | -5.81E-03 |
| 4984 | Rrm2 | RRM2 | 1.52E-01 | 1.83E-01 |
| 4985 | Psmc3ip | PSMC3IP | -6.26E-02 | 7.01E-02 |
| 4986 | Col6a5 | COL6A5 | 4.56E-02 | -4.14E-02 |
| 4987 | Mettl18 | METTL18 | -4.31E-02 | 2.84E-03 |
| 4988 | Syngr1 | SYNGR1 | -2.18E-02 | -6.89E-02 |
| 4989 | Slc25a15 | SLC25A15 | 3.78E-02 | -1.72E-02 |
| 4990 | Ak4 | AK4 | -5.30E-02 | -1.40E-02 |
| 4991 | Kxd1 | KXD1 | -2.40E-02 | 4.75E-03 |
| 4992 | Dclk1 | DCLK1 | 1.07E-01 | 8.52E-02 |
| 4993 | Birc5 | BIRC5 | 1.84E-01 | 1.90E-01 |
| 4994 | Ube2z | UBE2Z | 2.16E-02 | -2.08E-03 |
| 4995 | Fis1 | FIS1 | 1.82E-02 | -1.37E-02 |
| 4996 | Bhlhb9 | BHLHB9 | -3.98E-02 | 6.08E-03 |
| 4997 | Dcp1b | DCP1B | -3.28E-02 | -1.96E-02 |
| 4998 | Kcnc3 | KCNC3 | -4.79E-02 | -5.32E-02 |
| 4999 | Sspn | SSPN | -7.34E-02 | 3.77E-02 |
| 5000 | Uqcrh | UQCRHL | 2.13E-02 | -1.22E-02 |
| 5001 | Spata5 | SPATA5 | -3.82E-02 | 3.41E-02 |
| 5002 | Kif4 | KIF4A | 1.76E-01 | 1.99E-01 |
| 5003 | Aurka | AURKA | 1.70E-01 | 1.48E-01 |
| 5004 | Polr1e | POLR1E | 3.47E-02 | -4.69E-03 |
| 5005 | Egr2 | EGR2 | 1.92E-01 | 7.29E-02 |
| 5006 | Slc12a2 | SLC12A2 | 6.90E-02 | 2.48E-02 |
| 5007 | Nrarp | NRARP | -1.00E-01 | 4.79E-02 |
| 5008 | Zfp715 | ZNF175 | -3.39E-02 | 1.17E-02 |
| 5009 | Dnajb2 | DNAJB2 | -3.63E-02 | 1.09E-02 |
| 5010 | Crtc2 | CRTC2 | 2.05E-02 | -1.19E-02 |
| 5011 | Idh1 | IDH1 | 3.72E-02 | 1.29E-03 |
| 5012 | Insl6 | INSL6 | 6.91E-02 | 3.75E-02 |
| 5013 | Afap1l2 | AFAP1L2 | -4.92E-02 | 3.23E-02 |
| 5014 | Rbm46 | RBM46 | -4.84E-02 | 2.02E-02 |
| 5015 | Rsbn1l | RSBN1L | -2.67E-02 | -1.08E-02 |
| 5016 | Cd46 | CD46 | 5.67E-02 | -6.39E-02 |
| 5017 | Leprot | LEPROT | 1.87E-02 | 7.75E-03 |
| 5018 | Zfp395 | ZNF395 | -4.91E-02 | -4.38E-03 |
| 5019 | Kcnj10 | KCNJ10 | 2.95E-02 | 5.45E-03 |
| 5020 | Hmx3 | HMX3 | -8.07E-02 | 7.93E-02 |
| 5021 | Fignl2 | FIGNL2 | 3.32E-02 | 1.89E-03 |
| 5022 | Hoxc11 | HOXC11 | 1.01E-01 | 7.94E-02 |
| 5023 | Rad9a | RAD9A | -5.56E-02 | 5.31E-03 |
| 5024 | Nmnat2 | NMNAT2 | -5.52E-02 | 1.63E-02 |
| 5025 | Cx3cr1 | CX3CR1 | -1.15E-01 | 1.11E-01 |
| 5026 | Rabgap1 | RABGAP1 | 1.81E-02 | -1.92E-03 |
| 5027 | Itfg2 | ITFG2 | 2.43E-02 | -2.01E-02 |
| 5028 | Tmpo | TMPO | -2.25E-02 | 1.83E-02 |
| 5029 | Leo1 | LEO1 | 3.64E-02 | 4.26E-02 |
| 5030 | Gpatch4 | GPATCH4 | -2.95E-02 | 3.05E-02 |
| 5031 | Ldlrad1 | LDLRAD1 | 2.42E-02 | -6.66E-03 |
| 5032 | Tut1 | TUT1 | -2.97E-02 | 3.71E-03 |

| | | | | |
|---|---|---|---|---|
| 5033 | Taf6l | TAF6L | -1.98E-02 | 2.39E-03 |
| 5034 | Casp1 | CASP1 | -9.92E-02 | 6.01E-02 |
| 5035 | Fam107a | FAM107A | -5.07E-02 | -9.13E-02 |
| 5036 | Mrpl11 | MRPL11 | 1.96E-02 | -1.70E-02 |
| 5037 | Cep72 | CEP72 | 5.62E-02 | -6.48E-02 |
| 5038 | Alox5 | ALOX5 | 9.85E-02 | -2.13E-02 |
| 5039 | Adra1a | ADRA1A | 8.24E-02 | -8.63E-03 |
| 5040 | Cyp2r1 | CYP2R1 | -4.25E-02 | 4.20E-02 |
| 5041 | Lhfp | LHFP | 7.16E-02 | 1.63E-02 |
| 5042 | Nuf2 | NUF2 | 1.65E-01 | 1.84E-01 |
| 5043 | Pbdc1 | PBDC1 | 2.70E-02 | 8.04E-03 |
| 5044 | Col5a3 | COL5A3 | 9.71E-02 | 8.68E-02 |
| 5045 | Slc12a9 | SLC12A9 | -2.15E-02 | -2.14E-02 |
| 5046 | Tspan32 | TSPAN32 | -7.62E-02 | 3.15E-02 |
| 5047 | Phkg1 | PHKG1 | -7.82E-02 | -7.14E-03 |
| 5048 | Tmem200a | TMEM200A | -6.10E-02 | 1.29E-02 |
| 5049 | Tmem248 | TMEM248 | 1.90E-02 | -6.87E-03 |
| 5050 | Tma7 | TMA7 | -2.01E-02 | 1.09E-02 |
| 5051 | Prss16 | PRSS16 | -4.30E-02 | 3.01E-02 |
| 5052 | Mon2 | MON2 | 2.64E-02 | -4.17E-03 |
| 5053 | Adrb1 | ADRB1 | -7.41E-02 | -1.91E-03 |
| 5054 | Mettl17 | METTL17 | -2.41E-02 | -1.65E-02 |
| 5055 | Fibp | FIBP | 1.99E-02 | -1.37E-02 |
| 5056 | Pim3 | PIM3 | 7.14E-02 | -2.38E-02 |
| 5057 | Apba1 | APBA1 | -7.88E-02 | 1.12E-01 |
| 5058 | Nobox | NOBOX | 4.14E-02 | -4.99E-02 |
| 5059 | Mgat5 | MGAT5 | 3.26E-02 | -4.59E-02 |
| 5060 | Slc4a9 | SLC4A9 | -4.19E-02 | 1.95E-03 |
| 5061 | Ankrd44 | ANKRD44 | 4.30E-02 | -4.74E-02 |
| 5062 | Bax | BAX | 3.51E-02 | 3.49E-03 |
| 5063 | Wwtr1 | WWTR1 | 2.58E-02 | 1.90E-02 |
| 5064 | Trub2 | TRUB2 | 2.12E-02 | -3.11E-02 |
| 5065 | Pabpc4 | PABPC4 | -2.70E-02 | 1.70E-02 |
| 5066 | Gabarap | GABARAP | 1.80E-02 | 3.76E-04 |
| 5067 | Hoxa11 | HOXA11 | 9.79E-02 | -6.52E-02 |
| 5068 | Churc1 | CHURC1 | 3.85E-02 | 1.34E-02 |
| 5069 | Kif18a | KIF18A | 9.00E-02 | 3.93E-02 |
| 5070 | Ifnlr1 | IFNLR1 | 7.28E-02 | -4.31E-02 |
| 5071 | Ap2a2 | AP2A2 | 2.88E-02 | -1.81E-02 |
| 5072 | Kcng2 | KCNG2 | -6.45E-02 | -4.97E-02 |
| 5073 | 1700025G04Rik | C1orf21 | -4.83E-02 | 3.44E-02 |
| 5074 | Arhgap26 | ARHGAP26 | -3.94E-02 | -1.98E-02 |
| 5075 | Cmtr2 | CMTR2 | -4.98E-02 | 2.86E-02 |
| 5076 | Kbtbd7 | KBTBD7 | -3.56E-02 | 1.14E-02 |
| 5077 | Spi1 | SPI1 | -1.11E-01 | 1.81E-01 |
| 5078 | Eif4a1 | EIF4A1 | 1.81E-02 | 1.81E-02 |
| 5079 | Krit1 | KRIT1 | -2.87E-02 | 3.24E-03 |
| 5080 | Lims1 | LIMS1 | 2.00E-02 | -4.87E-03 |
| 5081 | Phf5a | PHF5A | 2.70E-02 | 1.11E-02 |
| 5082 | Etv3 | ETV3 | -2.61E-02 | 4.45E-03 |
| 5083 | Clcnka | CLCNKB | 8.97E-02 | -6.60E-02 |
| 5084 | Cpz | CPZ | 8.00E-02 | -1.12E-02 |
| 5085 | Polr1b | POLR1B | -1.80E-02 | 7.21E-03 |

| | | | | |
|---|---|---|---|---|
| 5086 | Zfp358 | ZNF358 | 2.59E-02 | -8.66E-03 |
| 5087 | Rarg | RARG | -6.13E-02 | 1.01E-02 |
| 5088 | Ahcyl1 | AHCYL1 | 1.94E-02 | -6.14E-03 |
| 5089 | Slc37a2 | SLC37A2 | -8.24E-02 | -3.58E-02 |
| 5090 | Hspa13 | HSPA13 | 2.50E-02 | -8.90E-03 |
| 5091 | Mrm1 | MRM1 | 3.37E-02 | -2.14E-02 |
| 5092 | Cwf19l1 | CWF19L1 | -2.75E-02 | -4.30E-03 |
| 5093 | Map2k6 | MAP2K6 | 5.28E-02 | -8.54E-02 |
| 5094 | Pcdhga4 | PCDHGA4 | -7.89E-02 | 5.41E-02 |
| 5095 | Igf2bp1 | IGF2BP1 | -1.08E-01 | -2.14E-02 |
| 5096 | Zc3h11a | ZC3H11A | -6.04E-02 | -1.36E-02 |
| 5097 | Fgd5 | FGD5 | -3.16E-02 | -1.03E-02 |
| 5098 | Lrpprc | LRPPRC | 2.88E-02 | -2.58E-02 |
| 5099 | Chka | CHKA | 3.73E-02 | 1.49E-02 |
| 5100 | Shisa7 | SHISA7 | -8.03E-02 | -7.79E-02 |
| 5101 | Tbx2 | TBX2 | 2.90E-02 | 6.23E-03 |
| 5102 | Kcna6 | KCNA6 | -9.30E-02 | 4.85E-02 |
| 5103 | Tec | TEC | -2.57E-02 | -2.56E-02 |
| 5104 | Cd300ld | CD300LD | -1.21E-01 | 1.41E-01 |
| 5105 | Pigh | PIGH | -3.51E-02 | 1.10E-02 |
| 5106 | Crispld2 | CRISPLD2 | 8.18E-02 | 2.25E-03 |
| 5107 | Ebf3 | EBF3 | 8.31E-02 | -5.13E-03 |
| 5108 | Dcun1d3 | DCUN1D3 | 3.60E-02 | 8.53E-03 |
| 5109 | Henmt1 | HENMT1 | 5.46E-02 | 1.57E-02 |
| 5110 | Snx1 | SNX1 | -2.06E-02 | -1.47E-02 |
| 5111 | Cpxm1 | CPXM1 | -1.41E-01 | 1.13E-01 |
| 5112 | Gpr160 | GPR160 | -3.43E-02 | 1.02E-02 |
| 5113 | Ager | AGER | 6.55E-02 | 3.94E-02 |
| 5114 | Sned1 | SNED1 | -9.60E-02 | -2.32E-02 |
| 5115 | Nim1k | NIM1K | 6.41E-02 | -5.93E-02 |
| 5116 | Ccdc127 | CCDC127 | -1.86E-02 | -2.92E-03 |
| 5117 | Maz | MAZ | -1.72E-02 | 1.38E-02 |
| 5118 | Gnl1 | GNL1 | 2.28E-02 | -6.86E-03 |
| 5119 | Gcsh | GCSH | -2.66E-02 | -1.50E-02 |
| 5120 | Gpc3 | GPC3 | 6.84E-02 | 4.34E-02 |
| 5121 | Gpd1l | GPD1L | 1.94E-02 | -1.64E-02 |
| 5122 | Rita1 | RITA1 | -2.79E-02 | -2.52E-03 |
| 5123 | Lck | LCK | -8.51E-02 | 6.76E-02 |
| 5124 | Dusp1 | DUSP1 | 1.21E-01 | -4.16E-02 |
| 5125 | Sec23b | SEC23B | 2.13E-02 | -5.37E-03 |
| 5126 | Trabd2b | TRABD2B | -4.15E-02 | 1.45E-02 |
| 5127 | Pm20d1 | PM20D1 | -3.44E-02 | -2.69E-02 |
| 5128 | Eif3k | EIF3K | 2.05E-02 | -2.31E-03 |
| 5129 | Magoh | MAGOH | 2.46E-02 | -1.14E-02 |
| 5130 | Crybg3 | CRYBG3 | 2.69E-02 | -2.98E-02 |
| 5131 | Aco1 | ACO1 | 3.43E-02 | -1.12E-02 |
| 5132 | Tceal1 | TCEAL1 | -6.00E-02 | -3.09E-03 |
| 5133 | Spag9 | SPAG9 | 2.72E-02 | -2.30E-02 |
| 5134 | Hpse2 | HPSE2 | 3.98E-02 | 2.81E-03 |
| 5135 | Ptpdc1 | PTPDC1 | -2.53E-02 | -4.22E-02 |
| 5136 | Catsperd | CATSPERD | -8.61E-02 | -1.29E-02 |
| 5137 | Pou4f1 | POU4F1 | -5.09E-02 | 5.23E-02 |
| 5138 | Mc2r | MC2R | 7.21E-02 | -4.67E-02 |

| | | | | |
|---|---|---|---|---|
| 5139 | Gabrp | GABRP | -1.31E-01 | 3.11E-02 |
| 5140 | Dhrs4 | DHRS4 | 3.85E-02 | -1.64E-02 |
| 5141 | Spag5 | SPAG5 | 2.40E-01 | -5.14E-02 |
| 5142 | Gm10499 | HLA-A | -6.66E-02 | 8.08E-02 |
| 5143 | Kpna6 | KPNA6 | 1.85E-02 | -8.88E-03 |
| 5144 | Zfp281 | ZNF281 | -3.47E-02 | 4.91E-03 |
| 5145 | Taldo1 | TALDO1 | 2.19E-02 | -4.74E-03 |
| 5146 | Ncf4 | NCF4 | -1.40E-01 | 2.18E-01 |
| 5147 | Apob | APOB | 7.00E-02 | -4.35E-02 |
| 5148 | Kcnn4 | KCNN4 | -7.39E-02 | 4.08E-02 |
| 5149 | Fcgr4 | FCGR3A | -1.35E-01 | 9.26E-02 |
| 5150 | Fcgr4 | FCGR3B | -1.35E-01 | 9.26E-02 |
| 5151 | Lonrf1 | LONRF1 | -5.00E-02 | -1.74E-02 |
| 5152 | Chac2 | CHAC2 | 3.14E-02 | 6.26E-03 |
| 5153 | Hscb | HSCB | 2.92E-02 | 3.56E-03 |
| 5154 | Zswim7 | ZSWIM7 | 3.35E-02 | 3.69E-03 |
| 5155 | Veph1 | VEPH1 | -6.66E-02 | -7.25E-02 |
| 5156 | Cxcl12 | CXCL12 | -3.90E-02 | -4.79E-02 |
| 5157 | Pnpla3 | PNPLA3 | -1.42E-01 | 3.31E-02 |
| 5158 | Pip5k1c | PIP5K1C | 3.00E-02 | -1.39E-02 |
| 5159 | Shisa3 | SHISA3 | -6.37E-02 | -7.85E-02 |
| 5160 | Ndufc2 | NDUFC2 | 2.16E-02 | -1.13E-02 |
| 5161 | Wnk2 | WNK2 | 6.69E-02 | -1.23E-01 |
| 5162 | Gtse1 | GTSE1 | 1.42E-01 | 6.65E-02 |
| 5163 | Snx14 | SNX14 | -2.15E-02 | -4.49E-03 |
| 5164 | 2210016F16Rik | C9orf64 | 2.75E-02 | -1.02E-02 |
| 5165 | Atp6v0a1 | ATP6V0A1 | 1.63E-02 | -6.37E-03 |
| 5166 | Ulk4 | ULK4 | -7.87E-02 | 1.29E-01 |
| 5167 | Mdn1 | MDN1 | -3.46E-02 | -2.36E-02 |
| 5168 | Bmi1 | COMMD3-BMI1 | 2.20E-02 | -4.38E-03 |
| 5169 | Sema6c | SEMA6C | -6.52E-02 | -9.43E-02 |
| 5170 | Ptges3l | PTGES3L | 5.48E-02 | 1.19E-02 |
| 5171 | Gemin4 | GEMIN4 | 4.41E-02 | -1.87E-02 |
| 5172 | Tmem9b | TMEM9B | 1.61E-02 | -3.60E-03 |
| 5173 | Renbp | RENBP | 4.21E-02 | -1.33E-02 |
| 5174 | Aox1 | AOX1 | 1.19E-01 | 3.48E-02 |
| 5175 | Hoxa4 | HOXA4 | 5.62E-02 | 1.21E-02 |
| 5176 | Cdk9 | CDK9 | 1.95E-02 | -6.32E-05 |
| 5177 | Arhgef28 | ARHGEF28 | -2.76E-02 | -6.83E-03 |
| 5178 | Pcsk1 | PCSK1 | 4.42E-02 | 5.77E-02 |
| 5179 | Ints3 | INTS3 | -2.80E-02 | 1.09E-02 |
| 5180 | Timm13 | TIMM13 | 2.32E-02 | -1.15E-02 |
| 5181 | Cenpf | CENPF | 2.14E-01 | 1.82E-01 |
| 5182 | Oscar | OSCAR | 5.12E-02 | -1.42E-02 |
| 5183 | Atp8a1 | ATP8A1 | -5.70E-02 | 1.52E-02 |
| 5184 | Kcnip3 | KCNIP3 | 1.35E-01 | -1.02E-01 |
| 5185 | Scara5 | SCARA5 | -7.01E-02 | 3.90E-02 |
| 5186 | Hnf1b | HNF1B | -2.84E-02 | -1.01E-03 |
| 5187 | Anxa1 | ANXA1 | 4.09E-02 | 7.02E-02 |
| 5188 | Usp20 | USP20 | 2.59E-02 | -2.29E-02 |
| 5189 | Brms1 | BRMS1 | 2.25E-02 | 3.56E-03 |
| 5190 | Gm973 | LOC100652824 | -7.87E-02 | 6.89E-02 |
| 5191 | Cd3eap | CD3EAP | 3.15E-02 | 8.48E-03 |

| | | | | |
|---|---|---|---|---|
| 5192 | Zfp580 | ZNF580 | 6.49E-02 | 1.55E-03 |
| 5193 | Nlk | NLK | 2.27E-02 | 1.38E-03 |
| 5194 | Adra1b | ADRA1B | 8.26E-02 | -6.78E-02 |
| 5195 | Zfp831 | ZNF831 | -4.45E-02 | -2.17E-02 |
| 5196 | Slc13a4 | SLC13A4 | 1.04E-01 | -3.95E-02 |
| 5197 | Dcp2 | DCP2 | -2.37E-02 | -4.08E-03 |
| 5198 | Chsy1 | CHSY1 | 4.71E-02 | 1.93E-02 |
| 5199 | Igsf6 | IGSF6 | -1.11E-01 | 2.02E-01 |
| 5200 | Fcer1g | FCER1G | -9.45E-02 | 1.42E-01 |
| 5201 | Tmem17 | TMEM17 | 5.40E-02 | 1.00E-02 |
| 5202 | Ifngr1 | IFNGR1 | 3.01E-02 | 4.96E-02 |
| 5203 | Cpne7 | CPNE7 | 5.59E-02 | 3.42E-02 |
| 5204 | Pnn | PNN | -1.47E-02 | 1.65E-02 |
| 5205 | Glp1r | GLP1R | 1.21E-01 | -8.21E-02 |
| 5206 | Trim60 | TRIM60 | -1.20E-01 | -1.00E-01 |
| 5207 | Zbtb8b | ZBTB8B | 1.31E-01 | -3.09E-02 |
| 5208 | Ralb | RALB | 2.69E-02 | 2.88E-03 |
| 5209 | Tmem255a | TMEM255A | -3.44E-02 | 1.85E-02 |
| 5210 | Wnk1 | WNK1 | 3.21E-02 | -1.12E-02 |
| 5211 | Plcl1 | PLCL1 | -4.56E-02 | -5.78E-02 |
| 5212 | Trpm7 | TRPM7 | 3.30E-02 | -1.09E-02 |
| 5213 | Ap3m2 | AP3M2 | -6.17E-02 | 1.86E-02 |
| 5214 | Ssu2 | SSUH2 | -7.75E-02 | 3.71E-02 |
| 5215 | Upf3b | UPF3B | 1.99E-02 | 1.56E-02 |
| 5216 | Rnf166 | RNF166 | 2.02E-02 | -1.03E-02 |
| 5217 | Nras | NRAS | 1.79E-02 | 1.18E-02 |
| 5218 | Ugt1a5 | UGT1A4 | 3.57E-02 | 1.28E-02 |
| 5219 | Usp16 | USP16 | -1.93E-02 | 5.77E-03 |
| 5220 | Tsc22d4 | TSC22D4 | 2.65E-02 | -1.66E-03 |
| 5221 | Nek3 | NEK3 | -5.18E-02 | 6.82E-02 |
| 5222 | Pigc | PIGC | -2.47E-02 | 5.32E-03 |
| 5223 | Smg1 | SMG1 | -2.25E-02 | -1.10E-03 |
| 5224 | Trpv1 | TRPV1 | -4.20E-02 | 1.07E-02 |
| 5225 | Gfer | GFER | 2.71E-02 | 6.46E-03 |
| 5226 | Gpr171 | GPR171 | -5.50E-02 | 7.43E-02 |
| 5227 | Zan | ZAN | 2.13E-01 | -8.36E-03 |
| 5228 | Mtmr4 | MTMR4 | -3.09E-02 | -8.22E-03 |
| 5229 | Rap2b | RAP2B | 5.96E-02 | 4.15E-02 |
| 5230 | Tab2 | TAB2 | 1.78E-02 | 4.89E-03 |
| 5231 | Rpe | RPE | 2.40E-02 | -2.07E-02 |
| 5232 | Rgs7 | RGS7 | 2.06E-01 | -1.35E-01 |
| 5233 | Bphl | BPHL | -2.83E-02 | -2.21E-02 |
| 5234 | Slc17a1 | SLC17A1 | 3.56E-02 | 9.63E-03 |
| 5235 | Cyfip1 | CYFIP1 | 2.02E-02 | -3.68E-03 |
| 5236 | Asrgl1 | ASRGL1 | 3.17E-02 | 9.57E-04 |
| 5237 | Zfp703 | ZNF703 | 3.80E-02 | 1.63E-03 |
| 5238 | Psmd9 | PSMD9 | 2.64E-02 | -1.31E-02 |
| 5239 | Rbm17 | RBM17 | -1.56E-02 | 4.31E-03 |
| 5240 | Rell1 | RELL1 | -3.35E-02 | 5.83E-02 |
| 5241 | Hspa8 | HSPA8 | -3.47E-02 | 2.16E-02 |
| 5242 | Tmem186 | TMEM186 | -2.41E-02 | 1.39E-02 |
| 5243 | Slc29a3 | SLC29A3 | 4.21E-02 | -4.16E-02 |
| 5244 | Pcdhga3 | PCDHGA3 | -7.83E-02 | 2.49E-02 |

| | | | | |
|---|---|---|---|---|
| 5245 | Nkpd1 | NKPD1 | 4.29E-02 | 5.37E-03 |
| 5246 | Aim2 | AIM2 | -5.55E-02 | 8.91E-02 |
| 5247 | Lmbr1l | LMBR1L | 1.93E-02 | -1.90E-02 |
| 5248 | Iars | IARS | 2.06E-02 | 1.12E-02 |
| 5249 | Gcfc2 | GCFC2 | 3.94E-02 | -2.66E-02 |
| 5250 | Ube2j2 | UBE2J2 | 2.14E-02 | 1.86E-04 |
| 5251 | Pcdhb1 | PCDHB1 | -5.82E-02 | 2.63E-02 |
| 5252 | Nxf1 | NXF1 | 1.68E-02 | 2.97E-03 |
| 5253 | C1rb | C1R | -7.15E-02 | 3.11E-02 |
| 5254 | Fignl1 | FIGNL1 | 1.29E-01 | 1.64E-01 |
| 5255 | Parva | PARVA | 1.82E-02 | -1.18E-02 |
| 5256 | Vkorc1l1 | VKORC1L1 | 1.63E-02 | -7.18E-03 |
| 5257 | Prmt7 | PRMT7 | 2.57E-02 | 7.41E-03 |
| 5258 | Myo1b | MYO1B | 6.85E-02 | -2.64E-03 |
| 5259 | Exosc5 | EXOSC5 | 2.17E-02 | -1.03E-02 |
| 5260 | Zfyve19 | ZFYVE19 | 1.75E-02 | -1.82E-02 |
| 5261 | Taf1a | TAF1A | -3.81E-02 | 4.80E-03 |
| 5262 | Slc41a1 | SLC41A1 | 2.47E-02 | -1.69E-03 |
| 5263 | Eps8l2 | EPS8L2 | 2.64E-02 | -3.50E-02 |
| 5264 | Ino80c | INO80C | 2.32E-02 | -4.34E-03 |
| 5265 | Oard1 | OARD1 | -1.98E-02 | -6.91E-03 |
| 5266 | Farp1 | FARP1 | -2.15E-02 | -3.79E-02 |
| 5267 | Edaradd | EDARADD | 8.75E-02 | 8.02E-02 |
| 5268 | Prf1 | PRF1 | -5.57E-02 | -4.30E-02 |
| 5269 | Arhgef33 | ARHGEF33 | 8.71E-02 | -5.08E-02 |
| 5270 | Apex2 | APEX2 | -3.21E-02 | 4.92E-02 |
| 5271 | Hpd | HPD | -8.89E-02 | -6.05E-02 |
| 5272 | Rabif | RABIF | 1.98E-02 | 1.54E-02 |
| 5273 | Car13 | CA13 | 8.22E-02 | 7.05E-02 |
| 5274 | Rxfp1 | RXFP1 | -2.55E-02 | -2.73E-03 |
| 5275 | Lysmd2 | LYSMD2 | -7.40E-02 | 4.60E-02 |
| 5276 | Cdk13 | CDK13 | -1.73E-02 | 7.85E-03 |
| 5277 | Epc1 | EPC1 | 2.34E-02 | -1.03E-02 |
| 5278 | Cln5 | CLN5 | -2.15E-02 | 2.16E-02 |
| 5279 | Atxn1 | ATXN1 | -4.83E-02 | -2.73E-02 |
| 5280 | BC003965 | C16orf91 | -2.19E-02 | -6.10E-03 |
| 5281 | Krt14 | KRT14 | -2.17E-01 | 7.47E-02 |
| 5282 | Chst10 | CHST10 | -7.40E-02 | 2.49E-02 |
| 5283 | Jrk | JRK | -4.97E-02 | -2.98E-02 |
| 5284 | Pgpep1 | PGPEP1 | 2.34E-02 | -3.30E-02 |
| 5285 | Krt18 | KRT18 | 1.19E-01 | 7.87E-02 |
| 5286 | Zfp507 | ZNF507 | -2.40E-02 | -5.32E-03 |
| 5287 | Csgalnact2 | CSGALNACT2 | 2.39E-02 | -1.62E-02 |
| 5288 | Col5a2 | COL5A2 | 1.15E-01 | 1.29E-01 |
| 5289 | Cited2 | CITED2 | -3.41E-02 | -2.52E-02 |
| 5290 | Gpx2 | GPX2 | 2.95E-02 | -1.89E-03 |
| 5291 | Xylt1 | XYLT1 | -7.38E-02 | -5.76E-02 |
| 5292 | Tmem102 | TMEM102 | 2.51E-02 | 3.25E-03 |
| 5293 | Fosl2 | FOSL2 | 9.46E-02 | 7.91E-02 |
| 5294 | Gpam | GPAM | 7.29E-02 | -5.08E-02 |
| 5295 | Ate1 | ATE1 | 2.31E-02 | -7.60E-03 |
| 5296 | Vti1b | VTI1B | 1.66E-02 | -2.43E-03 |
| 5297 | En2 | EN2 | -2.87E-02 | -1.84E-02 |

| | | | | |
|---|---|---|---|---|
| 5298 | Slc35g1 | SLC35G1 | 4.53E-02 | -1.35E-03 |
| 5299 | Fam160a2 | FAM160A2 | -1.95E-02 | -6.57E-03 |
| 5300 | Kcne1 | LOC102723475 | 4.86E-02 | 1.49E-02 |
| 5301 | Ppip5k1 | PPIP5K1 | 2.43E-02 | -2.72E-02 |
| 5302 | Lsmem2 | LSMEM2 | 6.05E-02 | -3.80E-02 |
| 5303 | Slc29a4 | SLC29A4 | 7.19E-02 | 8.05E-03 |
| 5304 | Sema3f | SEMA3F | 4.45E-02 | 2.77E-02 |
| 5305 | Fbxo8 | FBXO8 | 2.76E-02 | -1.85E-02 |
| 5306 | Atraid | ATRAID | -1.89E-02 | -7.46E-03 |
| 5307 | Arhgef26 | ARHGEF26 | 5.22E-02 | -3.39E-02 |
| 5308 | Mylip | MYLIP | 5.88E-02 | -1.15E-02 |
| 5309 | Rph3a | RPH3A | 2.87E-01 | -2.31E-01 |
| 5310 | Parp9 | PARP9 | -4.14E-02 | 3.78E-02 |
| 5311 | Nup43 | NUP43 | -3.25E-02 | 2.16E-02 |
| 5312 | 1700030J22Rik | C16orf46 | -7.17E-02 | 1.30E-03 |
| 5313 | Zfp36 | ZFP36 | 9.97E-02 | -2.13E-02 |
| 5314 | Dcdc2b | DCDC2B | -6.53E-02 | -1.54E-02 |
| 5315 | Slain1 | SLAIN1 | -6.17E-02 | 2.42E-02 |
| 5316 | Rlf | RLF | 3.07E-02 | -9.94E-03 |
| 5317 | Wdr86 | WDR86 | -9.54E-02 | 2.06E-02 |
| 5318 | Abhd13 | ABHD13 | 1.88E-02 | -1.73E-03 |
| 5319 | Uba3 | UBA3 | 1.83E-02 | -4.84E-03 |
| 5320 | Tbxa2r | TBXA2R | -6.38E-02 | 2.37E-02 |
| 5321 | Pkib | PKIB | 5.94E-02 | 3.85E-02 |
| 5322 | Clptm1 | CLPTM1 | 2.20E-02 | -1.29E-02 |
| 5323 | Kif13b | KIF13B | 1.32E-02 | -7.47E-03 |
| 5324 | Cep44 | CEP44 | -2.56E-02 | 5.22E-03 |
| 5325 | Msra | MSRA | 2.92E-02 | -1.92E-02 |
| 5326 | Khdrbs1 | KHDRBS1 | -1.61E-02 | -6.03E-03 |
| 5327 | Serpina1c | SERPINA1 | -1.15E-01 | 5.25E-02 |
| 5328 | Mettl1 | METTL1 | 3.84E-02 | -1.01E-02 |
| 5329 | Meox2 | MEOX2 | -3.68E-02 | 2.28E-02 |
| 5330 | Btrc | BTRC | -1.97E-02 | -2.06E-02 |
| 5331 | Bcl2l12 | BCL2L12 | -4.11E-02 | 3.06E-02 |
| 5332 | Zfp846 | ZNF846 | -5.77E-02 | -2.75E-02 |
| 5333 | Gga2 | GGA2 | 2.55E-02 | 6.52E-03 |
| 5334 | Shbg | SHBG | 5.95E-02 | 7.10E-02 |
| 5335 | Scarb1 | SCARB1 | -4.44E-02 | 3.51E-02 |
| 5336 | Itk | ITK | -8.50E-02 | 6.23E-02 |
| 5337 | Uck2 | UCK2 | -2.05E-02 | 7.63E-03 |
| 5338 | Tmem52b | TMEM52B | -4.13E-02 | 3.13E-02 |
| 5339 | 9930021J03Rik | KIAA2026 | -3.33E-02 | -8.81E-03 |
| 5340 | Atxn10 | ATXN10 | 1.57E-02 | -1.58E-02 |
| 5341 | Ambp | AMBP | 6.99E-02 | -2.57E-02 |
| 5342 | Fam186b | FAM186B | 7.47E-02 | -2.91E-02 |
| 5343 | Kbtbd3 | KBTBD3 | -4.83E-02 | -2.21E-03 |
| 5344 | Lpcat2 | LPCAT2 | -7.62E-02 | 1.66E-01 |
| 5345 | Myh6 | MYH6 | -7.79E-02 | -1.60E-01 |
| 5346 | Ace2 | ACE2 | 4.76E-02 | -1.52E-02 |
| 5347 | Ctbp1 | CTBP1 | 1.38E-02 | -1.36E-02 |
| 5348 | Lgi3 | LGI3 | 1.26E-01 | -3.82E-02 |
| 5349 | Rras | RRAS | -2.74E-02 | 4.89E-02 |
| 5350 | Chd5 | CHD5 | -8.78E-02 | -4.13E-03 |

| | | | | |
|---|---|---|---|---|
| 5351 | Seh1l | SEH1L | 1.75E-02 | -9.61E-03 |
| 5352 | Tnrc6a | TNRC6A | 1.76E-02 | -2.00E-02 |
| 5353 | Magi3 | MAGI3 | -4.46E-02 | -1.31E-02 |
| 5354 | Blm | BLM | 1.11E-01 | -2.06E-02 |
| 5355 | Cage1 | CAGE1 | 7.37E-02 | -5.45E-02 |
| 5356 | Thap11 | THAP11 | -3.05E-02 | 4.65E-03 |
| 5357 | Vps4a | VPS4A | 1.55E-02 | -1.51E-03 |
| 5358 | Gnai2 | GNAI2 | 2.32E-02 | 1.95E-02 |
| 5359 | Syne4 | SYNE4 | -3.44E-02 | -6.24E-03 |
| 5360 | Serinc5 | SERINC5 | 3.19E-02 | -7.17E-03 |
| 5361 | Bmp15 | BMP15 | 9.04E-02 | 6.72E-03 |
| 5362 | Otub2 | OTUB2 | -5.89E-02 | 2.68E-02 |
| 5363 | Necab3 | NECAB3 | 8.87E-02 | 6.31E-02 |
| 5364 | 4931428F04Rik | KIAA0895L | -5.15E-02 | 2.42E-02 |
| 5365 | Ccdc57 | CCDC57 | 3.60E-02 | -3.06E-02 |
| 5366 | Cldn2 | CLDN2 | 3.79E-02 | -2.41E-02 |
| 5367 | Mrpl21 | MRPL21 | 2.43E-02 | -2.30E-02 |
| 5368 | Slit1 | SLIT1 | -6.31E-02 | 8.68E-03 |
| 5369 | Pank2 | PANK2 | 1.42E-02 | 3.71E-03 |
| 5370 | Ddit4l | DDIT4L | 1.10E-01 | 2.21E-02 |
| 5371 | Vsig1 | VSIG1 | 7.55E-02 | 6.95E-02 |
| 5372 | Atp6v1b2 | ATP6V1B2 | 2.95E-02 | -2.64E-02 |
| 5373 | Fam76b | FAM76B | 3.24E-02 | 1.66E-03 |
| 5374 | Plcb1 | PLCB1 | 4.30E-02 | -7.31E-02 |
| 5375 | Gtf2h3 | GTF2H3 | -2.14E-02 | 3.58E-03 |
| 5376 | Nova1 | NOVA1 | -8.90E-02 | 8.30E-02 |
| 5377 | Tbrg4 | TBRG4 | 1.68E-02 | -1.01E-02 |
| 5378 | Zcchc2 | ZCCHC2 | -2.83E-02 | 3.11E-03 |
| 5379 | Rtn4r | RTN4R | 5.71E-02 | -4.55E-02 |
| 5380 | Ms4a2 | MS4A2 | -7.54E-02 | -8.57E-02 |
| 5381 | Smg9 | SMG9 | 2.23E-02 | 2.67E-03 |
| 5382 | Afg3l2 | AFG3L2 | 2.25E-02 | -1.56E-02 |
| 5383 | Angptl7 | ANGPTL7 | -9.34E-02 | 1.13E-02 |
| 5384 | Dennd4a | DENND4A | 3.43E-02 | -1.54E-02 |
| 5385 | Tmem213 | TMEM213 | -2.47E-02 | -1.49E-03 |
| 5386 | Myl12b | MYL12A | -1.50E-02 | 3.94E-03 |
| 5387 | Dpysl2 | DPYSL2 | -6.40E-02 | 1.89E-02 |
| 5388 | Klf6 | KLF6 | 5.06E-02 | 5.21E-02 |
| 5389 | Bcs1l | BCS1L | -2.74E-02 | -1.77E-02 |
| 5390 | Lca5l | LCA5L | 8.45E-02 | -5.82E-02 |
| 5391 | Map1s | MAP1S | -2.61E-02 | 3.32E-02 |
| 5392 | Nt5e | NT5E | -3.50E-02 | 2.39E-02 |
| 5393 | Dok4 | DOK4 | 2.73E-02 | 9.52E-03 |
| 5394 | 2610507B11Rik | KIAA0100 | 2.02E-02 | -2.66E-02 |
| 5395 | Slc25a27 | SLC25A27 | -6.67E-02 | -4.66E-03 |
| 5396 | Hsph1 | HSPH1 | -7.73E-02 | 9.31E-03 |
| 5397 | Cux2 | CUX2 | -6.58E-02 | -9.30E-02 |
| 5398 | Ccnl2 | CCNL2 | -2.00E-02 | 8.21E-03 |
| 5399 | Tmem140 | TMEM140 | -5.63E-02 | -1.34E-02 |
| 5400 | P4ha3 | P4HA3 | 2.72E-02 | 9.42E-03 |
| 5401 | Qser1 | QSER1 | -2.16E-02 | 2.83E-03 |
| 5402 | Bfsp1 | BFSP1 | -6.12E-02 | -3.57E-02 |
| 5403 | Ptdss1 | PTDSS1 | 1.55E-02 | -1.08E-03 |

| | | | | |
|---|---|---|---|---|
| 5404 | Mgat3 | MGAT3 | -3.99E-02 | -3.78E-02 |
| 5405 | Lgr6 | LGR6 | 2.01E-01 | -1.57E-01 |
| 5406 | Bcas1 | BCAS1 | -5.85E-02 | 4.33E-02 |
| 5407 | Xab2 | XAB2 | -1.48E-02 | -8.24E-03 |
| 5408 | Pla2g4f | PLA2G4F | -3.82E-02 | 1.67E-02 |
| 5409 | Timp1 | TIMP1 | 1.55E-01 | 4.13E-01 |
| 5410 | Gpx1 | GPX1 | 3.09E-02 | -1.13E-02 |
| 5411 | Tmem163 | TMEM163 | -6.41E-02 | 5.98E-02 |
| 5412 | Calr | CALR | 3.05E-02 | -6.75E-03 |
| 5413 | Drg1 | DRG1 | 1.56E-02 | -6.51E-03 |
| 5414 | Fam104a | FAM104A | 1.93E-02 | 1.34E-02 |
| 5415 | Cd47 | CD47 | 1.64E-02 | 1.70E-02 |
| 5416 | Tspyl2 | TSPYL2 | 3.11E-02 | -2.38E-02 |
| 5417 | Cct6b | CCT6B | -4.89E-02 | -6.29E-02 |
| 5418 | Col23a1 | COL23A1 | -5.66E-02 | -1.05E-01 |
| 5419 | Trim68 | TRIM68 | -3.94E-02 | 2.94E-02 |
| 5420 | Bsg | BSG | 1.69E-02 | -2.55E-03 |
| 5421 | Tmppe | TMPPE | -5.29E-02 | -3.02E-02 |
| 5422 | Llgl2 | LLGL2 | 2.18E-02 | 9.28E-03 |
| 5423 | Dhrs1 | DHRS1 | 2.35E-02 | -1.85E-02 |
| 5424 | Lrrc45 | LRRC45 | 2.32E-02 | 3.09E-03 |
| 5425 | Babam1 | BABAM1 | 1.78E-02 | -1.86E-02 |
| 5426 | Gmnc | GMNC | 2.96E-02 | -1.20E-02 |
| 5427 | Gdf10 | GDF10 | -9.99E-02 | 3.83E-02 |
| 5428 | Slc35e2 | SLC35E2B | -3.70E-02 | -6.76E-03 |
| 5429 | Agpat3 | AGPAT3 | 1.99E-02 | -2.13E-02 |
| 5430 | Ginm1 | GINM1 | 1.47E-02 | -4.60E-03 |
| 5431 | Loxl4 | LOXL4 | 1.09E-01 | -1.26E-01 |
| 5432 | Icam2 | ICAM2 | -2.96E-02 | -2.79E-03 |
| 5433 | Rbm6 | RBM6 | -2.25E-02 | 5.32E-03 |
| 5434 | AW549877 | C5orf51 | -2.56E-02 | -2.47E-02 |
| 5435 | Dnah5 | DNAH5 | -1.00E-01 | 2.05E-02 |
| 5436 | Acly | ACLY | 2.90E-02 | 2.09E-02 |
| 5437 | Kbtbd2 | KBTBD2 | -3.05E-02 | 2.50E-03 |
| 5438 | Slc27a3 | SLC27A3 | -5.23E-02 | 6.27E-03 |
| 5439 | Nasp | NASP | 2.03E-02 | 3.19E-02 |
| 5440 | Nup205 | NUP205 | 2.47E-02 | -1.06E-02 |
| 5441 | Adssl1 | ADSSL1 | -7.02E-02 | -2.94E-02 |
| 5442 | Naa50 | NAA50 | 2.20E-02 | -1.56E-03 |
| 5443 | Zfp367 | ZNF367 | 2.11E-02 | -9.81E-03 |
| 5444 | Rbm7 | RBM7 | -1.80E-02 | 1.15E-02 |
| 5445 | Mrps18c | MRPS18C | 3.21E-02 | 7.75E-03 |
| 5446 | Cstf2 | CSTF2 | 1.73E-02 | -7.94E-04 |
| 5447 | Matk | MATK | -7.28E-02 | 4.07E-02 |
| 5448 | Fbxo27 | FBXO27 | -6.46E-02 | -2.55E-02 |
| 5449 | Mdm4 | MDM4 | -1.45E-02 | 1.31E-02 |
| 5450 | Lima1 | LIMA1 | -2.30E-02 | 1.96E-03 |
| 5451 | Slc39a7 | SLC39A7 | 6.02E-02 | -3.64E-02 |
| 5452 | Zpbp2 | ZPBP2 | -4.41E-02 | -1.99E-02 |
| 5453 | Hnrnpa0 | HNRNPA0 | 2.31E-02 | 1.44E-02 |
| 5454 | Spata6 | SPATA6 | 3.35E-02 | 1.07E-02 |
| 5455 | Trmt5 | TRMT5 | -2.76E-02 | 5.53E-03 |
| 5456 | Olfm1 | OLFM1 | 2.86E-02 | 1.19E-02 |

| | | | | |
|---|---|---|---|---|
| 5457 | Telo2 | TELO2 | -2.69E-02 | 5.43E-03 |
| 5458 | Mpp3 | MPP3 | 6.33E-02 | -5.38E-03 |
| 5459 | Wdr7 | WDR7 | 3.86E-02 | -1.02E-02 |
| 5460 | Trip12 | TRIP12 | 1.67E-02 | -6.80E-03 |
| 5461 | Cldn9 | CLDN9 | 5.32E-02 | 1.86E-02 |
| 5462 | Ythdf2 | YTHDF2 | -1.98E-02 | 1.91E-02 |
| 5463 | Homez | HOMEZ | 2.58E-02 | 1.04E-04 |
| 5464 | Tmem184a | TMEM184A | -2.35E-02 | -1.78E-02 |
| 5465 | Lypla1 | LYPLA1 | -3.11E-02 | -1.52E-02 |
| 5466 | Prkag1 | PRKAG1 | 1.39E-02 | -5.31E-03 |
| 5467 | Fam83b | FAM83B | -6.74E-02 | 2.82E-03 |
| 5468 | Hoxb8 | HOXB8 | -4.44E-02 | 2.20E-02 |
| 5469 | Klhl2 | KLHL2 | 2.29E-02 | 6.97E-04 |
| 5470 | Fzr1 | FZR1 | 1.95E-02 | 1.01E-02 |
| 5471 | Cd200r4 | CD200R1L | -3.09E-02 | 9.51E-03 |
| 5472 | Vapb | VAPB | 1.55E-02 | -1.37E-02 |
| 5473 | Pole4 | POLE4 | -3.03E-02 | 1.15E-02 |
| 5474 | Zfp280d | ZNF280D | -1.91E-02 | -7.03E-04 |
| 5475 | Aak1 | AAK1 | 2.48E-02 | -2.26E-02 |
| 5476 | Lipe | LIPE | 5.46E-02 | 3.94E-03 |
| 5477 | Nck1 | NCK1 | -3.87E-02 | 1.91E-02 |
| 5478 | Pdxdc1 | PDXDC1 | 1.44E-02 | 5.34E-05 |
| 5479 | Hcst | HCST | -7.10E-02 | 7.50E-02 |
| 5480 | Rerg | RERG | 8.31E-02 | 6.43E-03 |
| 5481 | Adam2 | ADAM2 | 3.30E-02 | -1.99E-02 |
| 5482 | Foxk2 | FOXK2 | 2.14E-02 | 8.36E-03 |
| 5483 | Nap1l5 | NAP1L5 | -9.42E-02 | 1.75E-02 |
| 5484 | Chadl | CHADL | -6.40E-02 | -3.17E-02 |
| 5485 | Acvrl1 | ACVRL1 | 2.56E-02 | -3.68E-02 |
| 5486 | Lrch1 | LRCH1 | -2.11E-02 | -7.28E-03 |
| 5487 | Gm7030 | HLA-F | -8.15E-02 | -5.28E-02 |
| 5488 | Znrf2 | ZNRF2 | -1.96E-02 | -2.16E-02 |
| 5489 | Syt14 | SYT14 | 6.56E-02 | 2.51E-02 |
| 5490 | Slc25a24 | SLC25A24 | 7.30E-02 | 1.23E-01 |
| 5491 | Sav1 | SAV1 | -1.64E-02 | 2.08E-02 |
| 5492 | Ndufa13 | NDUFA13 | 2.38E-02 | -1.74E-02 |
| 5493 | Gpr137 | GPR137 | 2.18E-02 | -2.13E-02 |
| 5494 | Txlng | TXLNG | -2.89E-02 | 6.58E-03 |
| 5495 | Tdrd12 | LOC101929327 | -8.16E-02 | 5.20E-03 |
| 5496 | Ik | IK | 1.47E-02 | -9.89E-03 |
| 5497 | Emilin1 | EMILIN1 | 6.94E-02 | 2.70E-02 |
| 5498 | Mmp2 | MMP2 | 8.84E-02 | 1.37E-01 |
| 5499 | Arsa | ARSA | -3.44E-02 | -3.62E-02 |
| 5500 | Card14 | CARD14 | 8.23E-02 | 1.24E-02 |
| 5501 | Pax8 | PAX8 | 4.22E-02 | 5.06E-03 |
| 5502 | Brd8 | BRD8 | -1.23E-02 | -1.23E-02 |
| 5503 | Zyg11a | ZYG11A | 3.01E-02 | -2.11E-02 |
| 5504 | Rps6ka5 | RPS6KA5 | -6.94E-02 | 1.68E-03 |
| 5505 | Col6a2 | COL6A2 | 1.16E-01 | 6.11E-02 |
| 5506 | Hif1an | HIF1AN | 1.39E-02 | -1.75E-02 |
| 5507 | Esd | ESD | 2.46E-02 | -2.13E-02 |
| 5508 | Kctd18 | KCTD18 | 3.02E-02 | -1.80E-02 |
| 5509 | Emc9 | EMC9 | -2.62E-02 | -2.27E-02 |

| | | | | |
|---|---|---|---|---|
| 5510 | Rad51d | RAD51D | -2.71E-02 | -2.13E-02 |
| 5511 | Cep120 | CEP120 | -1.64E-02 | -4.92E-03 |
| 5512 | Zfp648 | ZNF648 | 9.31E-02 | -5.40E-03 |
| 5513 | Zdhhc20 | ZDHHC20 | 2.65E-02 | -3.18E-03 |
| 5514 | Vars | VARS | 2.26E-02 | 2.81E-02 |
| 5515 | Pcdh8 | PCDH8 | 6.01E-02 | 1.04E-01 |
| 5516 | Ppp6r3 | PPP6R3 | 1.94E-02 | -1.33E-02 |
| 5517 | Xpo4 | XPO4 | -2.61E-02 | -1.37E-02 |
| 5518 | Flnc | FLNC | 1.16E-01 | 2.50E-01 |
| 5519 | Tubg2 | TUBG2 | 8.21E-02 | -4.41E-02 |
| 5520 | Gas8 | GAS8 | 2.24E-02 | 7.51E-03 |
| 5521 | Pcdhga12 | PCDHGA12 | -4.78E-02 | -3.36E-02 |
| 5522 | Rpl27 | RPL27 | 1.95E-02 | 6.79E-03 |
| 5523 | Scyl1 | SCYL1 | 1.54E-02 | -1.79E-02 |
| 5524 | Lingo2 | LINGO2 | -9.13E-02 | 8.85E-03 |
| 5525 | Cep170 | CEP170 | -5.42E-02 | 6.00E-02 |
| 5526 | Atp6v1b1 | ATP6V1B1 | -3.12E-02 | -7.75E-03 |
| 5527 | Ppp1r37 | PPP1R37 | -1.56E-02 | -7.07E-03 |
| 5528 | Lyrm2 | LYRM2 | -2.42E-02 | -1.04E-02 |
| 5529 | Abcc8 | ABCC8 | -1.51E-01 | 5.31E-02 |
| 5530 | S1pr1 | S1PR1 | 6.63E-02 | -1.52E-03 |
| 5531 | Ptpn2 | PTPN2 | 2.29E-02 | 2.28E-02 |
| 5532 | Dbf4 | DBF4 | 7.03E-02 | 1.20E-01 |
| 5533 | Apbb1ip | APBB1IP | -1.03E-01 | 1.57E-01 |
| 5534 | Pbx3 | PBX3 | 3.40E-02 | 1.23E-02 |
| 5535 | Bad | BAD | -2.41E-02 | -1.64E-02 |
| 5536 | Snapc4 | SNAPC4 | -1.94E-02 | 1.57E-03 |
| 5537 | Ap4s1 | AP4S1 | -1.98E-02 | 1.52E-02 |
| 5538 | Nf1 | NF1 | 1.71E-02 | -8.98E-03 |
| 5539 | Cdyl2 | CDYL2 | -5.73E-02 | -1.72E-02 |
| 5540 | Wdr27 | WDR27 | 6.89E-02 | -1.38E-02 |
| 5541 | Mdk | MDK | -6.20E-02 | 2.84E-02 |
| 5542 | Tmed10 | TMED10 | 1.56E-02 | -1.60E-03 |
| 5543 | Rbm41 | RBM41 | -3.89E-02 | -5.91E-03 |
| 5544 | Rab40b | RAB40B | 8.42E-02 | 7.83E-03 |
| 5545 | Tep1 | TEP1 | -1.74E-02 | -1.24E-02 |
| 5546 | Usp53 | USP53 | 5.53E-02 | -1.28E-02 |
| 5547 | Mlst8 | MLST8 | 3.34E-02 | 3.16E-03 |
| 5548 | Trmt44 | TRMT44 | -4.41E-02 | 1.30E-02 |
| 5549 | Cdo1 | CDO1 | 3.73E-02 | -3.46E-02 |
| 5550 | Dsg2 | DSG2 | -2.61E-02 | 2.72E-03 |
| 5551 | Haus7 | HAUS7 | 3.12E-02 | 6.87E-03 |
| 5552 | Pcdhga10 | PCDHGA10 | -7.55E-02 | -2.72E-03 |
| 5553 | Myg1 | C12orf10 | -1.76E-02 | 3.94E-03 |
| 5554 | Ctif | CTIF | 3.87E-02 | -1.67E-02 |
| 5555 | Atp6v0d2 | ATP6V0D2 | -2.41E-02 | 3.92E-02 |
| 5556 | Zcchc24 | ZCCHC24 | 5.17E-02 | -6.00E-03 |
| 5557 | Wdr48 | WDR48 | -1.65E-02 | -1.53E-02 |
| 5558 | Ndufab1 | NDUFAB1 | 1.94E-02 | -1.11E-02 |
| 5559 | Ogfod2 | OGFOD2 | 2.04E-02 | -1.44E-02 |
| 5560 | Anks1 | ANKS1A | -1.97E-02 | -1.02E-02 |
| 5561 | Tnip3 | TNIP3 | 7.22E-02 | 8.19E-02 |
| 5562 | Cxcr2 | CXCR2 | 6.45E-02 | 4.03E-02 |

| | | | | |
|---|---|---|---|---|
| 5563 | Col18a1 | COL18A1 | 2.44E-02 | 3.21E-02 |
| 5564 | Pin1 | PIN1 | 2.08E-02 | -4.87E-03 |
| 5565 | Ripk4 | RIPK4 | 2.91E-02 | 1.79E-03 |
| 5566 | Zpbp | ZPBP | 7.30E-02 | -2.60E-02 |
| 5567 | Secisbp2l | SECISBP2L | 1.63E-02 | -1.85E-02 |
| 5568 | Usp43 | USP43 | 7.28E-02 | 5.22E-02 |
| 5569 | Psmc3 | PSMC3 | 1.59E-02 | -1.34E-02 |
| 5570 | Cfi | CFI | -1.10E-01 | 3.04E-01 |
| 5571 | Cachd1 | CACHD1 | -2.72E-02 | -1.94E-02 |
| 5572 | Usp37 | USP37 | -3.02E-02 | 7.12E-03 |
| 5573 | Napa | NAPA | 1.91E-02 | 5.03E-04 |
| 5574 | Matn2 | MATN2 | -5.94E-02 | -4.55E-02 |
| 5575 | Cdkal1 | CDKAL1 | 3.56E-02 | -2.30E-02 |
| 5576 | Zfp213 | ZNF213 | -2.83E-02 | -1.91E-02 |
| 5577 | Mustn1 | MUSTN1 | 5.56E-02 | 9.67E-02 |
| 5578 | Fig4 | FIG4 | 2.33E-02 | -2.49E-02 |
| 5579 | Asf1a | ASF1A | 2.46E-02 | 9.89E-03 |
| 5580 | Polh | POLH | -4.46E-02 | 3.86E-02 |
| 5581 | Mrps14 | MRPS14 | 2.13E-02 | -1.22E-02 |
| 5582 | Ddb1 | DDB1 | 2.21E-02 | -1.35E-02 |
| 5583 | Fitm2 | FITM2 | 2.80E-02 | -1.03E-02 |
| 5584 | Gfm1 | GFM1 | 2.35E-02 | -2.30E-02 |
| 5585 | Ofd1 | OFD1 | 3.06E-02 | -2.16E-02 |
| 5586 | Scn4b | SCN4B | -4.32E-02 | 1.81E-02 |
| 5587 | Cox17 | COX17 | -6.26E-02 | 2.43E-02 |
| 5588 | Tmem205 | TMEM205 | 3.02E-02 | -5.51E-03 |
| 5589 | Rpp21 | RPP21 | 1.99E-02 | -1.17E-02 |
| 5590 | Eif4a3 | EIF4A3 | 1.52E-02 | 9.04E-04 |
| 5591 | Hyls1 | HYLS1 | -7.83E-02 | 7.54E-02 |
| 5592 | Mr1 | MR1 | 5.36E-02 | -1.01E-03 |
| 5593 | Ang | ANG | -7.31E-02 | 1.14E-01 |
| 5594 | Rhbdl1 | RHBDL1 | -6.00E-02 | 2.45E-02 |
| 5595 | Fam171b | FAM171B | -5.41E-02 | 1.29E-01 |
| 5596 | Oxld1 | OXLD1 | -3.19E-02 | -1.16E-02 |
| 5597 | Mis18bp1 | MIS18BP1 | 1.32E-01 | 1.88E-01 |
| 5598 | Hhip | HHIP | -8.76E-02 | -1.29E-02 |
| 5599 | 2310009B15Rik | C1orf53 | 4.21E-02 | 2.47E-02 |
| 5600 | Prkar1b | PRKAR1B | 3.95E-02 | 3.30E-02 |
| 5601 | Lzts2 | LZTS2 | 1.55E-02 | -7.24E-03 |
| 5602 | Atl3 | ATL3 | 1.65E-02 | -3.62E-03 |
| 5603 | Sqstm1 | SQSTM1 | 3.48E-02 | 9.69E-03 |
| 5604 | Tor3a | TOR3A | -2.66E-02 | -9.84E-03 |
| 5605 | Zfp672 | ZNF672 | -3.48E-02 | 9.48E-03 |
| 5606 | Pofut2 | POFUT2 | 1.84E-02 | 2.33E-02 |
| 5607 | Artn | ARTN | -9.33E-02 | 7.01E-02 |
| 5608 | Npepl1 | NPEPL1 | 1.97E-02 | -1.94E-02 |
| 5609 | Crat | CRAT | 2.28E-02 | -1.46E-02 |
| 5610 | Paqr5 | PAQR5 | 4.16E-02 | -2.50E-02 |
| 5611 | Cnksr3 | CNKSR3 | 2.37E-02 | 3.33E-03 |
| 5612 | Cpsf4l | CPSF4L | -6.54E-02 | -4.76E-02 |
| 5613 | Nod2 | NOD2 | -8.07E-02 | 5.83E-02 |
| 5614 | Arid4a | ARID4A | -2.64E-02 | -2.88E-03 |
| 5615 | Add1 | ADD1 | 1.72E-02 | -2.23E-02 |

| | | | | |
|---|---|---|---|---|
| 5616 | Ccdc107 | CCDC107 | 2.68E-02 | -1.64E-02 |
| 5617 | Stard5 | STARD5 | -2.57E-02 | -1.51E-02 |
| 5618 | Psen2 | PSEN2 | 1.93E-02 | 6.49E-03 |
| 5619 | Gtf2b | GTF2B | 1.77E-02 | -8.12E-03 |
| 5620 | Mid1ip1 | MID1IP1 | 5.28E-02 | 5.07E-02 |
| 5621 | Cyp4x1 | CYP4X1 | -2.88E-02 | 2.51E-02 |
| 5622 | Ubxn6 | UBXN6 | 1.62E-02 | -1.31E-02 |
| 5623 | Stx1b | STX1B | 7.86E-02 | 2.72E-02 |
| 5624 | Efcab9 | EFCAB9 | 3.82E-02 | -4.20E-02 |
| 5625 | Sypl2 | SYPL2 | -4.07E-02 | 1.56E-02 |
| 5626 | Abcc2 | ABCC2 | 5.62E-02 | -6.64E-03 |
| 5627 | Kif18b | KIF18B | 1.71E-01 | 1.34E-01 |
| 5628 | Ccl22 | CCL22 | -5.93E-02 | 1.67E-02 |
| 5629 | Tspan5 | TSPAN5 | -3.44E-02 | -4.15E-02 |
| 5630 | Bnip2 | BNIP2 | 1.96E-02 | 9.81E-03 |
| 5631 | Qk | QKI | -2.00E-02 | -2.58E-03 |
| 5632 | Robo1 | ROBO1 | -9.21E-02 | 2.91E-02 |
| 5633 | Fam71e1 | FAM71E1 | 5.07E-02 | -2.78E-02 |
| 5634 | Hs6st1 | HS6ST1 | 2.89E-02 | -2.96E-03 |
| 5635 | Mark1 | MARK1 | 7.09E-02 | 2.36E-03 |
| 5636 | Eif2ak4 | EIF2AK4 | 3.03E-02 | -4.65E-02 |
| 5637 | Ccpg1os | C15orf65 | -3.50E-02 | -2.60E-02 |
| 5638 | Ephb4 | EPHB4 | 2.00E-02 | -1.44E-02 |
| 5639 | Timm9 | TIMM9 | 2.24E-02 | 1.97E-02 |
| 5640 | Rasip1 | RASIP1 | -3.53E-02 | 1.05E-02 |
| 5641 | Otof | OTOF | -2.96E-02 | -1.68E-02 |
| 5642 | Rgma | RGMA | -5.38E-02 | 1.40E-02 |
| 5643 | Mtrf1 | MTRF1 | -3.39E-02 | -2.27E-02 |
| 5644 | Aebp2 | AEBP2 | -2.02E-02 | -5.91E-03 |
| 5645 | Nhsl2 | NHSL2 | -2.66E-02 | -2.00E-02 |
| 5646 | Bmp2 | BMP2 | 8.70E-02 | 5.13E-02 |
| 5647 | Fxr1 | FXR1 | -1.68E-02 | 1.70E-02 |
| 5648 | Sulf1 | SULF1 | -1.02E-01 | 1.38E-01 |
| 5649 | Chrac1 | CHRAC1 | 2.44E-02 | -9.85E-03 |
| 5650 | Pwwp2a | PWWP2A | -2.06E-02 | 2.30E-02 |
| 5651 | Dph5 | DPH5 | 3.14E-02 | -4.57E-05 |
| 5652 | Sytl5 | SYTL5 | 6.07E-02 | -2.98E-02 |
| 5653 | Ndufb10 | NDUFB10 | 1.96E-02 | -2.04E-02 |
| 5654 | Fbxw5 | FBXW5 | 1.90E-02 | -9.45E-03 |
| 5655 | Sertad2 | SERTAD2 | -1.85E-02 | -1.87E-03 |
| 5656 | Clmn | CLMN | 2.16E-02 | -1.47E-02 |
| 5657 | Slc41a3 | SLC41A3 | 3.83E-02 | -1.47E-02 |
| 5658 | Slc35b4 | SLC35B4 | 1.98E-02 | -5.16E-03 |
| 5659 | Mcoln2 | MCOLN2 | -1.22E-01 | -1.80E-01 |
| 5660 | Oip5 | OIP5 | 1.32E-01 | 1.19E-01 |
| 5661 | Ybey | YBEY | -3.78E-02 | -2.65E-02 |
| 5662 | Trmt10a | TRMT10A | -3.30E-02 | -6.07E-03 |
| 5663 | Gaa | GAA | -2.72E-02 | -1.48E-02 |
| 5664 | Hoxc5 | HOXC5 | -3.75E-02 | -2.63E-02 |
| 5665 | Shroom2 | SHROOM2 | 2.52E-02 | -1.85E-02 |
| 5666 | Bmp6 | BMP6 | 3.96E-02 | -4.73E-03 |
| 5667 | Zfp444 | ZNF444 | -2.55E-02 | -2.32E-02 |
| 5668 | Decr1 | DECR1 | 3.50E-02 | -2.38E-02 |

| | | | | |
|---|---|---|---|---|
| 5669 | Slitrk4 | SLITRK4 | 1.30E-01 | 1.47E-01 |
| 5670 | Wee1 | WEE1 | 4.58E-02 | 2.62E-02 |
| 5671 | Zswim3 | ZSWIM3 | -7.96E-02 | -9.80E-03 |
| 5672 | Zbtb43 | ZBTB43 | -2.53E-02 | -8.05E-03 |
| 5673 | Grcc10 | C12orf57 | -4.88E-02 | 1.13E-02 |
| 5674 | Ctf1 | CTF1 | 4.99E-02 | 2.95E-03 |
| 5675 | Thg1l | THG1L | 3.40E-02 | 4.34E-03 |
| 5676 | Spata18 | SPATA18 | 1.95E-02 | -1.38E-02 |
| 5677 | Vps8 | VPS8 | -2.66E-02 | -4.24E-02 |
| 5678 | Pcid2 | PCID2 | -1.93E-02 | 1.53E-03 |
| 5679 | Camta2 | CAMTA2 | 1.73E-02 | -9.48E-03 |
| 5680 | Pigg | PIGG | -2.37E-02 | -3.91E-02 |
| 5681 | Elmo1 | ELMO1 | -5.64E-02 | 1.81E-02 |
| 5682 | Tmem50a | TMEM50A | -1.45E-02 | 2.94E-03 |
| 5683 | Pcif1 | PCIF1 | 1.71E-02 | -2.67E-02 |
| 5684 | Tex2 | TEX2 | 2.59E-02 | -1.61E-02 |
| 5685 | Hspg2 | HSPG2 | 3.77E-02 | -9.24E-03 |
| 5686 | Cdk16 | CDK16 | 1.37E-02 | -7.87E-03 |
| 5687 | Tnks | TNKS | 1.86E-02 | -2.16E-02 |
| 5688 | Rab19 | RAB19 | -6.18E-02 | -2.27E-02 |
| 5689 | Ndufa9 | NDUFA9 | 2.28E-02 | -2.19E-02 |
| 5690 | Taf2 | TAF2 | 1.89E-02 | -1.50E-03 |
| 5691 | Glul | GLUL | -3.07E-02 | -2.22E-02 |
| 5692 | Ctnna3 | CTNNA3 | 3.86E-02 | -1.59E-02 |
| 5693 | Pak1 | PAK1 | 3.85E-02 | 8.63E-03 |
| 5694 | Ndufc1 | NDUFC1 | 2.01E-02 | -1.14E-02 |
| 5695 | Sfrp5 | SFRP5 | 8.98E-02 | 1.18E-01 |
| 5696 | Zscan20 | ZSCAN20 | -5.47E-02 | 1.50E-02 |
| 5697 | Lsm5 | LSM5 | 3.76E-02 | 2.00E-02 |
| 5698 | Fgf17 | FGF17 | -5.08E-02 | 1.86E-02 |
| 5699 | Fkbp1a | FKBP1A | 1.45E-02 | 1.67E-02 |
| 5700 | Abcd1 | ABCD1 | -1.89E-02 | 5.25E-03 |
| 5701 | Smn1 | SMN1 | 2.73E-02 | 6.34E-03 |
| 5702 | Smn1 | SMN2 | 2.73E-02 | 6.34E-03 |
| 5703 | Epn3 | EPN3 | 7.08E-02 | 9.49E-04 |
| 5704 | Gnl3l | GNL3L | 1.67E-02 | -9.19E-03 |
| 5705 | Igip | IGIP | -4.27E-02 | 3.45E-02 |
| 5706 | Dcst1 | DCST1 | 7.35E-02 | -3.86E-02 |
| 5707 | Tmcc2 | TMCC2 | -4.55E-02 | 4.89E-03 |
| 5708 | Lpar3 | LPAR3 | 6.51E-02 | -3.40E-02 |
| 5709 | N4bp2 | N4BP2 | -2.89E-02 | -1.93E-02 |
| 5710 | Safb2 | SAFB2 | -1.44E-02 | 2.91E-03 |
| 5711 | Kbtbd12 | KBTBD12 | 3.81E-02 | -1.84E-02 |
| 5712 | Ipo5 | IPO5 | 1.81E-02 | 4.63E-03 |
| 5713 | Nup62 | NUP62 | -3.26E-02 | 3.77E-02 |
| 5714 | Kat2a | KAT2A | 1.75E-02 | 1.16E-02 |
| 5715 | Ddx55 | DDX55 | -2.59E-02 | 2.90E-03 |
| 5716 | Btbd9 | BTBD9 | 1.99E-02 | -1.55E-02 |
| 5717 | Uba1 | UBA1 | 1.94E-02 | -1.10E-02 |
| 5718 | Tex30 | TEX30 | 4.95E-02 | 3.83E-02 |
| 5719 | Hsbp1 | HSBP1 | 1.56E-02 | 1.45E-03 |
| 5720 | Tbc1d15 | TBC1D15 | 2.27E-02 | -3.40E-03 |
| 5721 | Mc4r | MC4R | 5.31E-02 | -7.21E-02 |

| | | | | |
|---|---|---|---|---|
| 5722 | Adh7 | ADH7 | 2.75E-01 | 1.35E-01 |
| 5723 | Bmpr1a | BMPR1A | -1.82E-02 | -4.63E-03 |
| 5724 | Tmem69 | TMEM69 | -2.68E-02 | 2.34E-03 |
| 5725 | Crtap | CRTAP | 2.40E-02 | 4.22E-03 |
| 5726 | Cbx7 | CBX7 | -2.53E-02 | -3.58E-02 |
| 5727 | Capn7 | CAPN7 | 1.28E-02 | -1.01E-02 |
| 5728 | Ncl | NCL | 1.99E-02 | 4.91E-03 |
| 5729 | Ndufb3 | NDUFB3 | 2.09E-02 | -6.16E-03 |
| 5730 | Ldb2 | LDB2 | -7.08E-02 | -1.63E-02 |
| 5731 | Vkorc1 | VKORC1 | -5.71E-02 | 2.76E-02 |
| 5732 | Kcnma1 | KCNMA1 | -5.78E-02 | -1.66E-03 |
| 5733 | Sh3bp4 | SH3BP4 | 2.67E-02 | 4.39E-03 |
| 5734 | Grhl1 | GRHL1 | 1.39E-01 | 6.19E-02 |
| 5735 | Ndufb7 | NDUFB7 | 1.96E-02 | -1.30E-02 |
| 5736 | Tmem116 | TMEM116 | 1.80E-02 | -1.33E-02 |
| 5737 | Idh2 | IDH2 | 1.92E-02 | -1.52E-02 |
| 5738 | Hip1r | HIP1R | 2.17E-02 | 2.25E-03 |
| 5739 | Lrrc42 | LRRC42 | 2.16E-02 | -1.26E-02 |
| 5740 | Camkk2 | CAMKK2 | 6.27E-02 | 8.04E-03 |
| 5741 | Smarcd2 | SMARCD2 | -2.28E-02 | 8.91E-03 |
| 5742 | Gnal | GNAL | 7.80E-02 | -9.67E-03 |
| 5743 | B9d2 | B9D2 | 2.61E-02 | 5.11E-03 |
| 5744 | Fntb | FNTB | 2.67E-02 | -1.50E-02 |
| 5745 | Cd164 | CD164 | -1.56E-02 | -1.11E-02 |
| 5746 | Ackr3 | ACKR3 | -5.13E-02 | 7.83E-02 |
| 5747 | Gm14326 | ZNF442 | -5.02E-02 | 6.62E-03 |
| 5748 | Kctd11 | KCTD11 | -5.46E-02 | 2.54E-02 |
| 5749 | Nr0b2 | NR0B2 | -9.86E-02 | 2.31E-02 |
| 5750 | Ipo13 | IPO13 | -1.59E-02 | -2.70E-03 |
| 5751 | Herc3 | HERC3 | 3.07E-02 | -4.20E-02 |
| 5752 | Rundc1 | RUNDC1 | 2.02E-02 | -3.52E-03 |
| 5753 | Efnb1 | EFNB1 | -2.32E-02 | 2.77E-02 |
| 5754 | Pmpca | PMPCA | 1.79E-02 | -1.54E-02 |
| 5755 | Atp1a2 | ATP1A2 | -1.23E-01 | 3.99E-02 |
| 5756 | Ublcp1 | UBLCP1 | 1.83E-02 | -1.63E-04 |
| 5757 | Prr18 | PRR18 | -1.05E-01 | 4.90E-03 |
| 5758 | Numbl | NUMBL | 6.85E-02 | 3.89E-02 |
| 5759 | Rasgef1a | RASGEF1A | -8.34E-02 | 8.83E-03 |
| 5760 | Saa4 | SAA2-SAA4 | 4.83E-02 | 7.00E-02 |
| 5761 | Plekhg6 | PLEKHG6 | 4.42E-02 | -3.54E-02 |
| 5762 | Itfg1 | ITFG1 | -1.90E-02 | -2.28E-02 |
| 5763 | Prtg | PRTG | -5.54E-02 | -7.19E-02 |
| 5764 | Serpinb1a | SERPINB1 | 5.15E-02 | 8.25E-02 |
| 5765 | Has3 | HAS3 | -9.08E-02 | -2.83E-04 |
| 5766 | Abcb4 | ABCB4 | -7.38E-02 | 6.90E-02 |
| 5767 | Tm4sf19 | TM4SF19 | 2.71E-02 | 8.01E-03 |
| 5768 | Vdac1 | VDAC1 | 2.12E-02 | -1.15E-02 |
| 5769 | Pcyt1b | PCYT1B | -3.76E-02 | 4.08E-02 |
| 5770 | 2410004B18Rik | C1orf52 | -3.54E-02 | 2.04E-02 |
| 5771 | Otog | OTOG | -2.63E-02 | 3.41E-03 |
| 5772 | Dpysl3 | DPYSL3 | -5.68E-02 | 1.69E-01 |
| 5773 | Ogfod3 | OGFOD3 | -2.43E-02 | -1.95E-02 |
| 5774 | Pgam5 | PGAM5 | 2.29E-02 | 2.10E-03 |

| | | | | |
|---|---|---|---|---|
| 5775 | Fam111a | FAM111A | -9.18E-02 | 1.37E-01 |
| 5776 | Emcn | EMCN | -3.92E-02 | -6.32E-03 |
| 5777 | Hsd11b1 | HSD11B1 | 6.06E-02 | -2.19E-02 |
| 5778 | Tmem100 | TMEM100 | 1.80E-01 | -5.91E-02 |
| 5779 | Cks1b | CKS1B | 8.29E-02 | 1.43E-01 |
| 5780 | Trip11 | TRIP11 | -1.74E-02 | -1.56E-03 |
| 5781 | Nop14 | NOP14 | 1.83E-02 | 5.18E-03 |
| 5782 | Anpep | ANPEP | -3.34E-02 | -3.09E-02 |
| 5783 | Xpa | XPA | -2.42E-02 | 1.00E-02 |
| 5784 | Slc48a1 | SLC48A1 | 3.15E-02 | -5.16E-03 |
| 5785 | Lyz1 | LYZ | 8.20E-02 | 7.31E-02 |
| 5786 | Fabp5 | FABP5 | -6.57E-02 | 6.70E-04 |
| 5787 | Zdhhc19 | ZDHHC19 | -6.17E-02 | 2.65E-02 |
| 5788 | Hivep1 | HIVEP1 | -1.84E-02 | -2.12E-02 |
| 5789 | Rmi1 | RMI1 | -2.26E-02 | -5.66E-03 |
| 5790 | Kcnh7 | KCNH7 | -3.64E-02 | 1.09E-02 |
| 5791 | Gpn1 | GPN1 | -2.19E-02 | -1.15E-02 |
| 5792 | Cul1 | CUL1 | 1.50E-02 | 1.70E-03 |
| 5793 | Riok1 | RIOK1 | 2.52E-02 | 9.37E-03 |
| 5794 | Itpr2 | ITPR2 | -2.55E-02 | -2.46E-02 |
| 5795 | Bend5 | BEND5 | -4.36E-02 | -1.36E-03 |
| 5796 | Synj2 | SYNJ2 | 5.62E-02 | 1.69E-03 |
| 5797 | Atg4b | ATG4B | 1.63E-02 | -3.48E-04 |
| 5798 | Efhc1 | EFHC1 | -6.44E-02 | 2.60E-02 |
| 5799 | Dock4 | DOCK4 | 5.70E-02 | -1.58E-02 |
| 5800 | Rptor | RPTOR | 1.67E-02 | -1.65E-02 |
| 5801 | Purb | PURB | 2.18E-02 | -3.81E-03 |
| 5802 | Gadd45g | GADD45G | 1.29E-01 | -9.97E-03 |
| 5803 | Exosc2 | EXOSC2 | 1.81E-02 | 3.77E-03 |
| 5804 | Cebpg | CEBPG | -2.43E-02 | 2.42E-02 |
| 5805 | Zfp574 | ZNF574 | -2.02E-02 | 3.53E-03 |
| 5806 | Gas1 | GAS1 | -3.39E-02 | -1.80E-02 |
| 5807 | Dpt | DPT | -1.48E-01 | 3.71E-02 |
| 5808 | Map1b | MAP1B | 1.37E-01 | -8.27E-02 |
| 5809 | Loxl3 | LOXL3 | 7.13E-02 | 1.85E-02 |
| 5810 | Zfp397 | ZNF397 | -2.16E-02 | 6.91E-05 |
| 5811 | Bptf | BPTF | -1.51E-02 | -5.36E-03 |
| 5812 | Trit1 | TRIT1 | 2.70E-02 | -9.52E-03 |
| 5813 | Snrnp40 | SNRNP40 | 2.68E-02 | -9.61E-03 |
| 5814 | Foxp3 | FOXP3 | -3.32E-02 | 4.20E-03 |
| 5815 | Oprl1 | OPRL1 | 2.87E-02 | -2.28E-02 |
| 5816 | Gpr34 | GPR34 | -7.38E-02 | 1.36E-01 |
| 5817 | Sh2b2 | SH2B2 | -7.84E-02 | 1.42E-01 |
| 5818 | Tmem126a | TMEM126A | 2.37E-02 | 9.68E-04 |
| 5819 | Tmem144 | TMEM144 | 4.62E-02 | -4.54E-02 |
| 5820 | Dgkz | DGKZ | -2.10E-02 | -1.12E-02 |
| 5821 | Ush1c | USH1C | 3.27E-02 | -2.20E-02 |
| 5822 | B3galt5 | B3GALT5 | -3.26E-02 | -1.21E-02 |
| 5823 | Pa2g4 | PA2G4 | 1.50E-02 | 5.71E-03 |
| 5824 | Scube2 | SCUBE2 | 2.12E-01 | 1.36E-02 |
| 5825 | Stambp | STAMBP | 1.89E-02 | -1.51E-02 |
| 5826 | Myo10 | MYO10 | 2.58E-02 | -2.34E-04 |
| 5827 | Gspt1 | GSPT1 | 1.50E-02 | 8.36E-03 |

| | | | | |
|---|---|---|---|---|
| 5828 | M6pr | M6PR | -1.37E-02 | -1.01E-02 |
| 5829 | Lrrc3b | LRRC3B | -5.68E-02 | -2.27E-02 |
| 5830 | Thrap3 | THRAP3 | 1.37E-02 | 2.20E-03 |
| 5831 | Mis12 | MIS12 | -3.52E-02 | 9.06E-03 |
| 5832 | Rab3gap2 | RAB3GAP2 | 1.41E-02 | -3.56E-03 |
| 5833 | Ereg | EREG | 3.60E-02 | -2.59E-02 |
| 5834 | Insc | INSC | -1.01E-01 | 3.08E-02 |
| 5835 | Rfwd3 | RFWD3 | -1.89E-02 | -6.52E-03 |
| 5836 | Kynu | KYNU | -5.51E-02 | -6.12E-03 |
| 5837 | Umod | UMOD | -3.18E-02 | -1.69E-02 |
| 5838 | Cd9 | CD9 | 2.55E-02 | 2.86E-02 |
| 5839 | Prkar2b | PRKAR2B | -1.80E-01 | 1.33E-01 |
| 5840 | Pcdhgc4 | PCDHGC4 | -4.80E-02 | 9.51E-02 |
| 5841 | Ube2cbp | UBE3D | 5.59E-02 | 9.78E-03 |
| 5842 | Lurap1 | LURAP1 | 5.94E-02 | -4.27E-03 |
| 5843 | Dmxl1 | DMXL1 | -2.58E-02 | -8.03E-04 |
| 5844 | Clpp | CLPP | 1.98E-02 | -1.64E-02 |
| 5845 | Ccna2 | CCNA2 | 1.61E-01 | 1.75E-01 |
| 5846 | Tgfbi | TGFBI | -8.49E-02 | 1.68E-01 |
| 5847 | C1qbp | C1QBP | 2.20E-02 | -1.23E-02 |
| 5848 | Bzw1 | BZW1 | 1.72E-02 | -1.26E-03 |
| 5849 | Ahcyl2 | AHCYL2 | 2.89E-02 | -3.88E-02 |
| 5850 | Cluap1 | CLUAP1 | -2.59E-02 | -6.27E-03 |
| 5851 | Mynn | MYNN | -2.27E-02 | -2.62E-02 |
| 5852 | Oscp1 | OSCP1 | -4.25E-02 | 5.34E-03 |
| 5853 | Crkl | CRKL | -2.52E-02 | 1.07E-03 |
| 5854 | Ints12 | INTS12 | -2.31E-02 | 1.83E-02 |
| 5855 | C330018D20Rik | C5orf63 | -2.88E-02 | -8.04E-03 |
| 5856 | Mogat2 | MOGAT2 | 9.05E-02 | 2.50E-02 |
| 5857 | Snx13 | SNX13 | 1.49E-02 | -1.34E-02 |
| 5858 | Cyb561d2 | CYB561D2 | 2.02E-02 | -6.93E-03 |
| 5859 | BC055324 | C1orf112 | -6.53E-02 | 9.06E-02 |
| 5860 | Arid3a | ARID3A | 3.83E-02 | -1.39E-02 |
| 5861 | Anapc1 | ANAPC1 | -1.56E-02 | -9.78E-03 |
| 5862 | Dcaf6 | DCAF6 | 1.27E-02 | -2.99E-02 |
| 5863 | Atp1b3 | ATP1B3 | 3.23E-02 | -1.41E-02 |
| 5864 | Zfp146 | ZNF146 | -2.05E-02 | 2.73E-03 |
| 5865 | Rnf123 | RNF123 | 2.11E-02 | -2.06E-02 |
| 5866 | Xirp1 | XIRP1 | 5.63E-02 | 2.54E-02 |
| 5867 | Slc6a14 | SLC6A14 | -6.27E-02 | 1.03E-02 |
| 5868 | Gm8909 | HLA-A | -7.89E-02 | 2.94E-02 |
| 5869 | Kdm5c | KDM5C | -2.05E-02 | 1.20E-03 |
| 5870 | Pecam1 | PECAM1 | 2.63E-02 | -1.59E-02 |
| 5871 | Anapc5 | ANAPC5 | 1.30E-02 | -3.48E-03 |
| 5872 | Bicd1 | BICD1 | 4.37E-02 | -8.46E-03 |
| 5873 | Hs1bp3 | HS1BP3 | -2.79E-02 | -1.29E-02 |
| 5874 | Rbm42 | RBM42 | -2.64E-02 | 2.83E-02 |
| 5875 | Rdh9 | RDH16 | -2.71E-02 | 6.94E-03 |
| 5876 | Tmem196 | TMEM196 | 4.15E-02 | 9.82E-03 |
| 5877 | Cd3e | CD3E | -7.53E-02 | 6.49E-03 |
| 5878 | Cdkn2c | CDKN2C | 5.25E-02 | 4.40E-02 |
| 5879 | Zmynd19 | ZMYND19 | 2.91E-02 | 1.00E-02 |
| 5880 | Fndc3b | FNDC3B | 2.02E-02 | -1.89E-02 |

| | | | | |
|---|---|---|---|---|
| 5881 | Bag4 | BAG4 | 2.90E-02 | -2.84E-02 |
| 5882 | Msh2 | MSH2 | -2.21E-02 | -3.42E-03 |
| 5883 | Dis3 | DIS3 | -2.86E-02 | -2.37E-02 |
| 5884 | Copa | COPA | 1.61E-02 | -1.81E-02 |
| 5885 | Prap1 | PRAP1 | -5.99E-02 | -5.06E-02 |
| 5886 | Nfasc | NFASC | -3.58E-02 | 7.06E-02 |
| 5887 | Erlec1 | ERLEC1 | 1.56E-02 | 6.79E-03 |
| 5888 | Atp1b2 | ATP1B2 | -4.79E-02 | 3.05E-02 |
| 5889 | Sh2d1b1 | SH2D1B | -6.91E-02 | 1.26E-01 |
| 5890 | Zfp286 | ZNF286A | 9.29E-02 | -1.16E-01 |
| 5891 | Neurl3 | NEURL3 | -5.16E-02 | 5.34E-02 |
| 5892 | Ubxn10 | UBXN10 | 7.85E-02 | 3.26E-02 |
| 5893 | Zfp775 | ZNF775 | -4.12E-02 | 2.13E-02 |
| 5894 | Ppm1a | PPM1A | 1.52E-02 | -1.40E-02 |
| 5895 | Psd4 | PSD4 | -8.75E-02 | 1.08E-01 |
| 5896 | Cherp | CHERP | -1.30E-02 | 2.01E-03 |
| 5897 | Bcl2a1d | BCL2A1 | -5.04E-02 | 9.60E-02 |
| 5898 | Fryl | FRYL | 1.36E-02 | -1.60E-02 |
| 5899 | Olfr460 | OR9A4 | 5.85E-02 | -8.51E-02 |
| 5900 | Wars2 | WARS2 | -2.15E-02 | 8.33E-03 |
| 5901 | Ubr1 | UBR1 | 1.70E-02 | -1.56E-02 |
| 5902 | Ube2t | UBE2T | 1.09E-01 | 1.10E-01 |
| 5903 | Ppdpf | PPDPF | -1.89E-02 | 1.80E-03 |
| 5904 | Ercc6l2 | ERCC6L2 | 1.70E-02 | 1.95E-03 |
| 5905 | Robo2 | ROBO2 | -8.57E-02 | -5.22E-02 |
| 5906 | Manea | MANEA | -1.93E-02 | -1.37E-02 |
| 5907 | Gng10 | GNG10 | -3.00E-02 | 7.30E-03 |
| 5908 | C1galt1c1 | C1GALT1C1 | -1.81E-02 | -2.30E-03 |
| 5909 | Lin9 | LIN9 | 3.76E-02 | -9.94E-03 |
| 5910 | Dact1 | DACT1 | 8.76E-02 | -4.71E-02 |
| 5911 | Chd7 | CHD7 | -2.50E-02 | -2.79E-03 |
| 5912 | Gad1 | GAD1 | -7.85E-02 | -6.81E-02 |
| 5913 | Atp6v1c2 | ATP6V1C2 | -3.38E-02 | -1.26E-03 |
| 5914 | Ptpn4 | PTPN4 | 2.58E-02 | -2.10E-02 |
| 5915 | Cgnl1 | CGNL1 | 1.73E-02 | -3.50E-02 |
| 5916 | Ankrd54 | ANKRD54 | 3.27E-02 | -9.61E-03 |
| 5917 | Zbtb8a | ZBTB8A | -4.70E-02 | 3.03E-02 |
| 5918 | Mrs2 | MRS2 | 1.74E-02 | -2.08E-03 |
| 5919 | Gnb5 | GNB5 | 4.66E-02 | 7.72E-03 |
| 5920 | Cars2 | CARS2 | -1.97E-02 | -1.63E-02 |
| 5921 | F2rl3 | F2RL3 | -6.59E-02 | 4.87E-02 |
| 5922 | Cs | CS | 1.88E-02 | -1.19E-02 |
| 5923 | Myo9a | MYO9A | -2.69E-02 | -1.54E-02 |
| 5924 | Il9r | IL9R | -2.93E-02 | 1.49E-02 |
| 5925 | Ccdc65 | CCDC65 | -7.12E-02 | 3.23E-02 |
| 5926 | Cyb5b | CYB5B | 2.46E-02 | -2.64E-02 |
| 5927 | Acot2 | ACOT2 | 6.74E-02 | 1.96E-02 |
| 5928 | Rnf146 | RNF146 | -2.20E-02 | -2.03E-03 |
| 5929 | Micall2 | MICALL2 | 7.35E-02 | 2.01E-02 |
| 5930 | Txnrd3 | TXNRD3 | -2.76E-02 | -1.24E-02 |
| 5931 | Adamts12 | ADAMTS12 | 1.02E-01 | 2.90E-02 |
| 5932 | Ndufb5 | NDUFB5 | 1.92E-02 | -1.08E-02 |
| 5933 | Akap13 | AKAP13 | 2.31E-02 | -8.51E-03 |

| | | | | |
|---|---|---|---|---|
| 5934 | Cdk15 | CDK15 | -3.61E-02 | 4.45E-02 |
| 5935 | Hdac11 | HDAC11 | -1.75E-02 | -5.14E-02 |
| 5936 | Hoxb6 | HOXB6 | -5.70E-02 | 2.18E-02 |
| 5937 | Hlx | HLX | -9.74E-02 | 7.27E-02 |
| 5938 | Phf12 | PHF12 | -1.35E-02 | 1.58E-03 |
| 5939 | P2rx6 | P2RX6 | -6.29E-02 | 5.91E-02 |
| 5940 | Lrrc41 | LRRC41 | 1.38E-02 | 6.10E-03 |
| 5941 | Hells | HELLS | 8.44E-02 | 1.73E-01 |
| 5942 | Canx | CANX | 2.01E-02 | -1.35E-02 |
| 5943 | Tmem222 | TMEM222 | 1.79E-02 | -1.80E-02 |
| 5944 | Zfp790 | ZNF790 | -6.19E-02 | 1.05E-03 |
| 5945 | Fbxl7 | FBXL7 | -5.74E-02 | -1.62E-02 |
| 5946 | Chrnb4 | CHRNB4 | 5.13E-02 | 9.61E-03 |
| 5947 | Gucy1a2 | GUCY1A2 | -4.90E-02 | 2.45E-02 |
| 5948 | Kalrn | KALRN | 2.15E-02 | -2.64E-02 |
| 5949 | Tube1 | TUBE1 | 7.76E-02 | 4.84E-02 |
| 5950 | Wdr24 | WDR24 | -1.64E-02 | -8.36E-03 |
| 5951 | Lrrc49 | LRRC49 | -3.84E-02 | 5.72E-03 |
| 5952 | Serpina3n | SERPINA3 | 1.32E-01 | 5.04E-01 |
| 5953 | Ccl5 | CCL5 | -9.07E-02 | 2.25E-01 |
| 5954 | Adamtsl4 | ADAMTSL4 | 5.16E-02 | 7.78E-02 |
| 5955 | Epha4 | EPHA4 | -7.81E-02 | -1.06E-02 |
| 5956 | Arhgap6 | ARHGAP6 | -2.82E-02 | -2.78E-02 |
| 5957 | Pcdhga8 | PCDHGA8 | -3.44E-02 | -9.84E-03 |
| 5958 | F12 | F12 | -3.04E-02 | 2.41E-02 |
| 5959 | Irf4 | IRF4 | -9.80E-02 | 1.68E-01 |
| 5960 | Atad1 | ATAD1 | 1.37E-02 | -2.57E-03 |
| 5961 | Zkscan7 | ZKSCAN7 | -3.49E-02 | -1.28E-02 |
| 5962 | Rem2 | REM2 | 5.43E-02 | 5.00E-02 |
| 5963 | Zmiz1 | ZMIZ1 | 2.29E-02 | 4.54E-04 |
| 5964 | Cdc25a | CDC25A | 2.27E-02 | 1.97E-02 |
| 5965 | Smpd4 | SMPD4 | 1.63E-02 | -1.21E-02 |
| 5966 | Crispld1 | CRISPLD1 | 6.57E-02 | -1.73E-02 |
| 5967 | Acsl3 | ACSL3 | -2.87E-02 | 6.56E-03 |
| 5968 | Mtcp1 | CMC4 | -3.91E-02 | 9.52E-03 |
| 5969 | Ankmy1 | ANKMY1 | -8.25E-02 | -5.45E-02 |
| 5970 | Nme5 | NME5 | -5.04E-02 | -1.49E-02 |
| 5971 | Ptpn7 | PTPN7 | -6.24E-02 | 9.68E-02 |
| 5972 | Lypd6b | LYPD6B | 1.00E-01 | 4.94E-02 |
| 5973 | Zkscan2 | ZKSCAN2 | -7.44E-02 | -4.87E-03 |
| 5974 | Dnase2a | DNASE2 | -2.46E-02 | -7.39E-03 |
| 5975 | Tmem192 | TMEM192 | 2.05E-02 | -8.42E-03 |
| 5976 | Rapgef5 | RAPGEF5 | 4.42E-02 | -1.95E-02 |
| 5977 | Elk1 | ELK1 | -2.35E-02 | -1.90E-02 |
| 5978 | Hps4 | HPS4 | -2.81E-02 | -2.70E-02 |
| 5979 | Ttc7 | TTC7A | 2.30E-02 | -3.77E-02 |
| 5980 | Slc12a6 | SLC12A6 | 3.43E-02 | -7.19E-03 |
| 5981 | Cyb561 | CYB561 | 2.85E-02 | 1.20E-02 |
| 5982 | Rchy1 | RCHY1 | 1.86E-02 | -6.79E-03 |
| 5983 | Set | SET | 1.38E-02 | 3.49E-03 |
| 5984 | Rnpc3 | RNPC3 | -2.97E-02 | 1.87E-02 |
| 5985 | Spopl | SPOPL | 2.94E-02 | -3.23E-02 |
| 5986 | Alk | ALK | -4.90E-02 | 7.97E-03 |

| | | | | |
|---|---|---|---|---|
| 5987 | Fes | FES | -3.27E-02 | 7.64E-02 |
| 5988 | Chd9 | CHD9 | -1.67E-02 | -1.53E-03 |
| 5989 | Id3 | ID3 | -7.21E-02 | 8.60E-02 |
| 5990 | Srms | SRMS | 7.18E-02 | -7.26E-02 |
| 5991 | Ccl4 | CCL4 | -9.98E-02 | 3.89E-02 |
| 5992 | Tpx2 | TPX2 | 1.67E-01 | 2.43E-01 |
| 5993 | Grm7 | GRM7 | 7.66E-02 | -1.01E-01 |
| 5994 | Magi2 | MAGI2 | -6.63E-02 | -3.43E-02 |
| 5995 | Acad8 | ACAD8 | 1.85E-02 | -2.83E-02 |
| 5996 | Vamp4 | VAMP4 | -2.82E-02 | -2.13E-02 |
| 5997 | Calhm2 | CALHM2 | -8.03E-02 | 2.26E-02 |
| 5998 | Mrc1 | MRC1 | -8.68E-02 | 1.58E-01 |
| 5999 | Cldn3 | CLDN3 | -7.02E-02 | 4.95E-02 |
| 6000 | Zwilch | ZWILCH | 1.04E-01 | 1.20E-01 |
| 6001 | Polq | POLQ | 9.56E-02 | 7.11E-02 |
| 6002 | Pah | PAH | -3.71E-02 | -2.00E-02 |
| 6003 | Ndufs8 | NDUFS8 | 1.68E-02 | -2.01E-02 |
| 6004 | Sec14l2 | SEC14L2 | -4.91E-02 | 7.42E-02 |
| 6005 | Srpk1 | SRPK1 | 1.39E-02 | 3.99E-03 |
| 6006 | Nemf | NEMF | 1.68E-02 | -1.27E-02 |
| 6007 | Itpr3 | ITPR3 | -3.68E-02 | 4.10E-02 |
| 6008 | C3ar1 | C3AR1 | -1.41E-01 | 1.65E-01 |
| 6009 | Il1rn | IL1RN | 8.54E-02 | 3.38E-01 |
| 6010 | Lancl1 | LANCL1 | 2.27E-02 | -1.46E-02 |
| 6011 | Uqcrb | UQCRB | 1.71E-02 | -1.32E-02 |
| 6012 | Ankrd13b | ANKRD13B | -2.83E-02 | 1.64E-02 |
| 6013 | Was | WAS | -6.90E-02 | 8.77E-02 |
| 6014 | Trap1 | TRAP1 | -1.97E-02 | -1.66E-02 |
| 6015 | Traip | TRAIP | 9.03E-02 | 5.29E-02 |
| 6016 | H2-D1 | HLA-A | -3.80E-02 | 3.24E-02 |
| 6017 | Flywch1 | FLYWCH1 | 1.41E-02 | -1.24E-02 |
| 6018 | Smpd3 | SMPD3 | 1.45E-01 | -2.31E-02 |
| 6019 | Syt17 | SYT17 | -4.11E-02 | 3.77E-02 |
| 6020 | Tmem174 | TMEM174 | -3.60E-02 | -2.47E-02 |
| 6021 | Top1mt | TOP1MT | 3.28E-02 | -1.62E-02 |
| 6022 | Slc11a1 | SLC11A1 | -9.51E-02 | 1.24E-01 |
| 6023 | Pick1 | PICK1 | 1.49E-02 | -1.17E-02 |
| 6024 | Phf8 | PHF8 | -2.35E-02 | 1.03E-02 |
| 6025 | Adamts5 | ADAMTS5 | 8.07E-02 | -3.61E-02 |
| 6026 | Kif27 | KIF27 | 6.49E-02 | -2.48E-02 |
| 6027 | St3gal1 | ST3GAL1 | 3.98E-02 | 9.57E-03 |
| 6028 | Srp72 | SRP72 | 2.00E-02 | -2.22E-03 |
| 6029 | Nos3 | NOS3 | 3.91E-02 | 5.70E-02 |
| 6030 | Ccdc120 | CCDC120 | 6.43E-02 | 2.17E-02 |
| 6031 | Tmcc1 | TMCC1 | -1.86E-02 | 1.28E-02 |
| 6032 | Mrpl43 | MRPL43 | 1.70E-02 | -2.00E-03 |
| 6033 | Zfp410 | ZNF410 | 2.28E-02 | 1.02E-02 |
| 6034 | Rbak | RBAK | 3.34E-02 | -1.25E-02 |
| 6035 | Smcr8 | SMCR8 | -2.13E-02 | -6.64E-03 |
| 6036 | Gspt2 | GSPT2 | -5.81E-02 | 6.09E-02 |
| 6037 | Fzd2 | FZD2 | -7.11E-02 | 6.36E-02 |
| 6038 | Mrpl12 | MRPL12 | 2.22E-02 | -2.13E-02 |
| 6039 | Exoc3 | EXOC3 | -1.62E-02 | -8.24E-03 |

| | | | | |
|---|---|---|---|---|
| 6040 | Card9 | CARD9 | -7.45E-02 | 7.72E-02 |
| 6041 | B4galnt4 | B4GALNT4 | 5.08E-02 | -2.77E-02 |
| 6042 | Crym | CRYM | 3.75E-02 | -3.43E-02 |
| 6043 | Tnfaip8l3 | TNFAIP8L3 | -1.04E-01 | 2.69E-02 |
| 6044 | Hp | HP | 2.75E-01 | 3.69E-01 |
| 6045 | Atg14 | ATG14 | -2.89E-02 | -2.96E-02 |
| 6046 | Lamp2 | LAMP2 | 2.17E-02 | -9.39E-03 |
| 6047 | Daxx | DAXX | -1.76E-02 | -2.22E-04 |
| 6048 | Lsg1 | LSG1 | 1.46E-02 | -4.13E-03 |
| 6049 | Slbp | SLBP | -2.17E-02 | 1.21E-02 |
| 6050 | Rnf135 | RNF135 | 2.83E-02 | 7.19E-03 |
| 6051 | Prpf19 | PRPF19 | 1.37E-02 | -2.54E-03 |
| 6052 | Mdh1 | MDH1 | 2.25E-02 | -1.77E-02 |
| 6053 | Tph1 | TPH1 | 4.13E-02 | 1.35E-03 |
| 6054 | Sall1 | SALL1 | -2.16E-02 | 2.21E-04 |
| 6055 | Rnps1 | RNPS1 | 2.17E-02 | 6.72E-03 |
| 6056 | Tnik | TNIK | -5.84E-02 | 2.66E-02 |
| 6057 | Naca | NACA | -1.54E-02 | 1.50E-02 |
| 6058 | Pfdn2 | PFDN2 | 1.90E-02 | -1.00E-02 |
| 6059 | Slc16a5 | SLC16A5 | 3.39E-02 | -5.52E-04 |
| 6060 | Olfr267 | OR2K2 | -3.85E-02 | 1.36E-02 |
| 6061 | Fasl | FASLG | -3.88E-02 | -1.69E-02 |
| 6062 | Racgap1 | RACGAP1 | 1.32E-01 | 1.34E-01 |
| 6063 | Unc119 | UNC119 | 5.09E-02 | -6.61E-03 |
| 6064 | Csk | CSK | -1.85E-02 | -9.65E-04 |
| 6065 | Saal1 | SAAL1 | -3.18E-02 | -8.91E-03 |
| 6066 | AA467197 | C15orf48 | 7.43E-02 | 5.09E-02 |
| 6067 | Gmfg | GMFG | -5.30E-02 | 3.71E-02 |
| 6068 | Begain | BEGAIN | -9.90E-02 | 1.85E-02 |
| 6069 | Ccl3 | CCL3L3 | -1.08E-01 | 3.80E-02 |
| 6070 | Tm2d1 | TM2D1 | 1.59E-02 | -9.35E-03 |
| 6071 | Rb1cc1 | RB1CC1 | -2.03E-02 | -4.45E-03 |
| 6072 | Asprv1 | ASPRV1 | 8.07E-02 | -6.44E-03 |
| 6073 | Manba | MANBA | -2.17E-02 | 3.49E-03 |
| 6074 | Tia1 | TIA1 | 2.22E-02 | 2.68E-03 |
| 6075 | Atg4c | ATG4C | 2.16E-02 | -1.43E-03 |
| 6076 | Themis | THEMIS | -5.34E-02 | 1.41E-02 |
| 6077 | Slc35a1 | SLC35A1 | -1.64E-02 | -1.50E-02 |
| 6078 | Nptxr | NPTXR | -8.09E-02 | -6.66E-02 |
| 6079 | Cyld | CYLD | -1.99E-02 | -2.48E-02 |
| 6080 | Pcdha12 | PCDHA13 | -3.29E-02 | -8.62E-03 |
| 6081 | Cdkn2d | CDKN2D | 3.68E-02 | 4.02E-02 |
| 6082 | Plod2 | PLOD2 | 3.73E-02 | 7.85E-03 |
| 6083 | Ino80e | INO80E | -1.56E-02 | -4.61E-03 |
| 6084 | Slc7a3 | SLC7A3 | -7.58E-02 | 4.27E-02 |
| 6085 | Elovl2 | ELOVL2 | -3.62E-02 | -2.66E-02 |
| 6086 | Hoxc4 | HOXC4 | 4.19E-02 | -1.35E-02 |
| 6087 | Nlrp3 | NLRP3 | -8.99E-02 | 1.44E-01 |
| 6088 | Ccdc137 | CCDC137 | -2.63E-02 | 1.42E-02 |
| 6089 | Bhlhe40 | BHLHE40 | -4.90E-02 | 2.26E-02 |
| 6090 | Xpo1 | XPO1 | -2.69E-02 | 1.47E-02 |
| 6091 | Uba6 | UBA6 | -3.57E-02 | 1.79E-03 |
| 6092 | Tnnt3 | TNNT3 | 3.04E-02 | -2.10E-02 |

| | | | | |
|---|---|---|---|---|
| 6093 | Met | MET | -2.59E-02 | -1.67E-02 |
| 6094 | Snap91 | SNAP91 | 7.43E-02 | -1.77E-02 |
| 6095 | Sox13 | SOX13 | 1.89E-02 | -2.33E-02 |
| 6096 | Hck | HCK | -1.35E-01 | 1.80E-01 |
| 6097 | Efhd2 | EFHD2 | 3.65E-02 | 6.96E-02 |
| 6098 | Ctxn2 | CTXN2 | -3.29E-02 | -2.22E-03 |
| 6099 | Smarcal1 | SMARCAL1 | -1.66E-02 | -1.34E-02 |
| 6100 | Grtp1 | GRTP1 | -3.38E-02 | -4.65E-02 |
| 6101 | Ano4 | ANO4 | -1.54E-01 | -1.14E-01 |
| 6102 | Taf12 | TAF12 | -2.21E-02 | 8.71E-03 |
| 6103 | Zfp7 | ZNF7 | -4.62E-02 | -1.58E-02 |
| 6104 | Kir3dl1 | KIR3DL3 | 1.66E-02 | -1.37E-02 |
| 6105 | Arl5c | ARL5C | -6.85E-02 | 8.04E-02 |
| 6106 | Ly6g5b | LY6G5B | 6.05E-02 | 5.34E-03 |
| 6107 | Ap2s1 | AP2S1 | 1.78E-02 | 2.31E-03 |
| 6108 | Tmeff1 | TMEFF1 | -7.87E-02 | 6.48E-02 |
| 6109 | Plg | PLG | 1.10E-01 | 8.67E-02 |
| 6110 | Gnb2 | GNB2 | 1.63E-02 | 7.92E-05 |
| 6111 | Sirt2 | SIRT2 | 1.71E-02 | -1.10E-02 |
| 6112 | Fank1 | FANK1 | 2.99E-02 | 7.06E-04 |
| 6113 | Fstl1 | FSTL1 | 3.97E-02 | 2.40E-02 |
| 6114 | Tmub2 | TMUB2 | 1.21E-02 | -4.98E-03 |
| 6115 | Rdh12 | RDH12 | -8.73E-02 | 9.95E-02 |
| 6116 | Scp2 | SCP2 | 2.96E-02 | -2.42E-02 |
| 6117 | Snx7 | SNX7 | -2.46E-02 | -1.43E-02 |
| 6118 | Fbn2 | FBN2 | 6.64E-02 | 1.40E-01 |
| 6119 | Cotl1 | COTL1 | 3.48E-02 | -1.33E-02 |
| 6120 | Irf1 | IRF1 | -9.23E-02 | 6.94E-02 |
| 6121 | Ctnnal1 | CTNNAL1 | 5.02E-02 | -1.61E-02 |
| 6122 | Kmt2d | KMT2D | -1.79E-02 | -8.51E-03 |
| 6123 | Zfp2 | ZFP2 | 2.58E-02 | -8.35E-03 |
| 6124 | Ube2e1 | UBE2E1 | -1.57E-02 | -2.20E-02 |
| 6125 | Rbm45 | RBM45 | 2.22E-02 | 1.15E-02 |
| 6126 | Ccdc82 | CCDC82 | -2.06E-02 | 8.66E-03 |
| 6127 | Anxa5 | ANXA5 | 2.23E-02 | 1.98E-02 |
| 6128 | Cfh | CFH | -6.60E-02 | 1.82E-02 |
| 6129 | Myh7 | MYH7 | -2.86E-02 | -1.70E-02 |
| 6130 | Sik3 | SIK3 | 3.19E-02 | -6.53E-03 |
| 6131 | Gstm1 | GSTM5 | -4.05E-02 | -4.11E-03 |
| 6132 | Fam20b | FAM20B | -1.42E-02 | -2.87E-02 |
| 6133 | Chchd6 | CHCHD6 | 2.35E-02 | -3.50E-02 |
| 6134 | 5730409E04Rik | C1orf216 | -3.51E-02 | -4.36E-02 |
| 6135 | Atxn2l | ATXN2L | 1.32E-02 | -4.51E-03 |
| 6136 | Atpaf1 | ATPAF1 | 2.73E-02 | -2.30E-02 |
| 6137 | Stx16 | STX16 | -1.61E-02 | 1.58E-03 |
| 6138 | Ntf3 | NTF3 | -9.46E-02 | -1.12E-01 |
| 6139 | Ak9 | AK9 | -5.62E-02 | 5.12E-03 |
| 6140 | Iglon5 | IGLON5 | -7.73E-02 | 1.34E-01 |
| 6141 | Cul2 | CUL2 | 1.40E-02 | -9.41E-03 |
| 6142 | Mapk3 | MAPK3 | -1.30E-02 | -5.92E-03 |
| 6143 | Nop58 | NOP58 | -4.75E-02 | 4.41E-02 |
| 6144 | Taco1 | TACO1 | 2.68E-02 | -2.97E-02 |
| 6145 | Dnajc21 | DNAJC21 | 1.69E-02 | -2.08E-03 |

| | | | | |
|---|---|---|---|---|
| 6146 | Zfhx4 | ZFHX4 | -9.32E-02 | -2.01E-02 |
| 6147 | Cbfb | CBFB | -2.63E-02 | 5.23E-02 |
| 6148 | Snph | SNPH | 4.15E-02 | 1.71E-02 |
| 6149 | Vstm2l | VSTM2L | 2.29E-02 | -2.06E-02 |
| 6150 | Neu1 | NEU1 | 2.92E-02 | -3.40E-02 |
| 6151 | Ankmy2 | ANKMY2 | 1.61E-02 | -1.24E-02 |
| 6152 | Hs3st5 | HS3ST5 | -3.55E-02 | 7.18E-03 |
| 6153 | Fah | FAH | -2.48E-02 | -1.68E-02 |
| 6154 | Sumo1 | SUMO1 | -1.79E-02 | -5.76E-04 |
| 6155 | Camsap3 | CAMSAP3 | -1.68E-02 | 6.00E-03 |
| 6156 | Nmi | NMI | -3.11E-02 | 2.09E-02 |
| 6157 | Eef2k | EEF2K | -4.55E-02 | -2.30E-02 |
| 6158 | Ppfia3 | PPFIA3 | 5.94E-02 | 7.71E-02 |
| 6159 | Tma16 | TMA16 | 2.80E-02 | -2.32E-03 |
| 6160 | Cntln | CNTLN | -6.49E-02 | 9.31E-03 |
| 6161 | Trim3 | TRIM3 | 1.56E-02 | -1.35E-02 |
| 6162 | Bdh2 | BDH2 | 3.75E-02 | -1.79E-02 |
| 6163 | Med9 | MED9 | 1.99E-02 | 3.61E-03 |
| 6164 | Rinl | RINL | -5.29E-02 | -3.45E-04 |
| 6165 | Mpz | MPZ | -6.67E-02 | 1.69E-02 |
| 6166 | Rilpl2 | RILPL2 | 2.59E-02 | 1.87E-02 |
| 6167 | Hoxb2 | HOXB2 | -3.33E-02 | 2.13E-02 |
| 6168 | Mtbp | MTBP | 4.15E-02 | -1.92E-03 |
| 6169 | Aurkb | AURKB | 1.64E-01 | 2.18E-01 |
| 6170 | Efs | EFS | -1.19E-01 | 4.74E-02 |
| 6171 | Rapgef1 | RAPGEF1 | 1.86E-02 | -2.01E-02 |
| 6172 | Tmem221 | TMEM221 | -5.53E-02 | 4.49E-03 |
| 6173 | Fbxo22 | FBXO22 | 2.00E-02 | -1.31E-02 |
| 6174 | Psmg1 | PSMG1 | -2.10E-02 | -1.48E-02 |
| 6175 | Polr3h | POLR3H | -2.62E-02 | -1.99E-02 |
| 6176 | Stim2 | STIM2 | 2.60E-02 | 1.89E-02 |
| 6177 | Ldb1 | LDB1 | 1.81E-02 | -7.78E-03 |
| 6178 | Lrrc39 | LRRC39 | 4.66E-02 | 1.30E-02 |
| 6179 | Dguok | DGUOK | -1.91E-02 | -4.08E-03 |
| 6180 | Pilrb2 | PILRB | -7.18E-02 | 9.66E-02 |
| 6181 | Hgf | HGF | -1.30E-01 | 8.39E-02 |
| 6182 | 2210408I21Rik | KIAA0825 | 3.70E-02 | -3.91E-02 |
| 6183 | Eid1 | EID1 | 1.79E-02 | 3.60E-03 |
| 6184 | Kmt2a | KMT2A | -1.82E-02 | -1.26E-02 |
| 6185 | Usp48 | USP48 | 1.26E-02 | -7.75E-03 |
| 6186 | Tank | TANK | -2.01E-02 | 3.05E-02 |
| 6187 | Rbfox2 | RBFOX2 | 1.56E-02 | -1.49E-03 |
| 6188 | Ccnk | CCNK | -1.62E-02 | -3.58E-03 |
| 6189 | Lmnb2 | LMNB2 | 2.00E-02 | 1.80E-02 |
| 6190 | Haus4 | HAUS4 | 5.55E-02 | 4.11E-03 |
| 6191 | Cnn3 | CNN3 | -1.95E-02 | -1.03E-02 |
| 6192 | Csrp2 | CSRP2 | -2.41E-02 | -1.94E-03 |
| 6193 | Vdac3 | VDAC3 | 1.78E-02 | -9.46E-03 |
| 6194 | Gpx8 | GPX8 | -3.95E-02 | 7.23E-02 |
| 6195 | Ntn4 | NTN4 | 2.35E-02 | -1.03E-02 |
| 6196 | Atxn7l3 | ATXN7L3 | 1.50E-02 | 5.82E-03 |
| 6197 | Dlx4 | DLX4 | -2.92E-02 | 2.42E-02 |
| 6198 | Efcab14 | EFCAB14 | -1.48E-02 | -1.77E-02 |

| | | | | |
|---|---|---|---|---|
| 6199 | Dock10 | DOCK10 | -6.57E-02 | 1.04E-01 |
| 6200 | Them6 | THEM6 | 3.02E-02 | 1.89E-02 |
| 6201 | Wdr19 | WDR19 | 2.29E-02 | -3.17E-02 |
| 6202 | Tshz2 | TSHZ2 | -5.83E-02 | -5.12E-02 |
| 6203 | Ehf | EHF | 9.51E-02 | -2.81E-02 |
| 6204 | Tspyl4 | TSPYL4 | -3.00E-02 | -2.14E-02 |
| 6205 | Serinc1 | SERINC1 | 1.80E-02 | -1.10E-02 |
| 6206 | Ap1b1 | AP1B1 | 2.22E-02 | -1.45E-02 |
| 6207 | Uhrf1 | UHRF1 | 1.11E-01 | 2.31E-01 |
| 6208 | Gemin8 | GEMIN8 | -3.02E-02 | -1.77E-02 |
| 6209 | Pcdha2 | PCDHA2 | -3.86E-02 | 2.19E-02 |
| 6210 | Deptor | DEPTOR | -2.30E-02 | -2.76E-02 |
| 6211 | Stard6 | STARD6 | -5.69E-02 | 5.32E-02 |
| 6212 | Ncam1 | NCAM1 | -7.10E-02 | -4.95E-04 |
| 6213 | Ak1 | AK1 | -4.35E-02 | 3.99E-03 |
| 6214 | Cep128 | CEP128 | 5.36E-02 | 5.08E-02 |
| 6215 | F13b | F13B | -5.27E-02 | -4.27E-02 |
| 6216 | Map3k8 | MAP3K8 | 6.36E-02 | 4.15E-03 |
| 6217 | Aqp7 | AQP7 | -3.13E-02 | -3.60E-02 |
| 6218 | Gm525 | C17orf67 | -5.20E-02 | -8.20E-03 |
| 6219 | Lama1 | LAMA1 | 4.82E-02 | -7.00E-02 |
| 6220 | Fam217b | FAM217B | -4.84E-02 | 2.81E-02 |
| 6221 | G3bp1 | G3BP1 | -1.54E-02 | 3.79E-03 |
| 6222 | Nat1 | NAT2 | -6.00E-02 | 6.79E-02 |
| 6223 | Lrrc8d | LRRC8D | 2.68E-02 | -1.94E-02 |
| 6224 | Ptgs2 | PTGS2 | 1.80E-01 | -1.61E-02 |
| 6225 | Armcx5 | ARMCX5 | 4.92E-02 | -2.25E-02 |
| 6226 | Zfp273 | ZNF676 | -5.22E-02 | 6.33E-04 |
| 6227 | Cenpq | CENPQ | -4.25E-02 | 5.02E-02 |
| 6228 | Zmym5 | ZMYM5 | -2.00E-02 | -8.35E-03 |
| 6229 | Timd4 | TIMD4 | -1.67E-02 | 9.05E-03 |
| 6230 | Inmt | INMT | -7.04E-02 | -1.44E-02 |
| 6231 | Cacna1f | CACNA1F | -1.92E-02 | 2.18E-03 |
| 6232 | Kbtbd13 | KBTBD13 | 7.79E-02 | 1.05E-02 |
| 6233 | Gpsm3 | GPSM3 | -4.78E-02 | 2.91E-02 |
| 6234 | Cx3cl1 | CX3CL1 | -2.74E-02 | 5.48E-02 |
| 6235 | Ttyh3 | TTYH3 | 2.98E-02 | 1.55E-02 |
| 6236 | Snx11 | SNX11 | 2.89E-02 | -3.05E-03 |
| 6237 | Wdr12 | WDR12 | 2.18E-02 | 2.19E-02 |
| 6238 | Rft1 | RFT1 | 2.66E-02 | -2.32E-02 |
| 6239 | Zranb1 | ZRANB1 | 1.92E-02 | -2.21E-02 |
| 6240 | Kif11 | KIF11 | 1.44E-01 | 1.80E-01 |
| 6241 | Wdr59 | WDR59 | 2.09E-02 | -5.55E-03 |
| 6242 | Myo18a | MYO18A | 1.32E-02 | -1.30E-02 |
| 6243 | Nom1 | NOM1 | 2.17E-02 | -7.14E-03 |
| 6244 | 1700001L19Rik | C5orf49 | -4.82E-02 | -3.28E-03 |
| 6245 | Mlh3 | MLH3 | -1.65E-02 | -2.37E-02 |
| 6246 | Metap2 | METAP2 | 1.30E-02 | -1.07E-02 |
| 6247 | Tmem164 | TMEM164 | 2.24E-02 | -1.63E-02 |
| 6248 | Extl2 | EXTL2 | 2.42E-02 | -2.57E-02 |
| 6249 | Mcm2 | MCM2 | 4.29E-02 | 6.26E-02 |
| 6250 | Dars2 | DARS2 | -1.97E-02 | -1.67E-02 |
| 6251 | Kdm4a | KDM4A | 1.75E-02 | -7.06E-03 |

| | | | | |
|---|---|---|---|---|
| 6252 | Pank1 | PANK1 | 2.59E-02 | -2.02E-02 |
| 6253 | Rb1 | RB1 | 1.89E-02 | -2.01E-02 |
| 6254 | Ltb4r2 | LTB4R2 | -5.67E-02 | 2.86E-02 |
| 6255 | Mmp3 | MMP3 | 5.09E-02 | 1.75E-02 |
| 6256 | Alkbh5 | ALKBH5 | 1.63E-02 | -7.04E-03 |
| 6257 | Cpsf7 | CPSF7 | 1.15E-02 | 2.56E-03 |
| 6258 | Cryga | CRYGA | 2.03E-02 | -8.12E-04 |
| 6259 | Trpc4ap | TRPC4AP | 1.38E-02 | -1.17E-02 |
| 6260 | Pdgfc | PDGFC | -5.68E-02 | -7.09E-02 |
| 6261 | Sgca | SGCA | 4.36E-02 | 5.12E-02 |
| 6262 | Muc13 | MUC13 | 6.62E-02 | 3.67E-02 |
| 6263 | Dlgap5 | DLGAP5 | 1.68E-01 | 1.90E-01 |
| 6264 | Ccdc106 | CCDC106 | -6.11E-02 | -2.55E-02 |
| 6265 | Gdpd1 | GDPD1 | 4.86E-02 | 4.82E-02 |
| 6266 | Sectm1a | SECTM1 | -9.99E-02 | -3.52E-02 |
| 6267 | Tmem230 | TMEM230 | -1.74E-02 | -8.57E-03 |
| 6268 | Col6a1 | COL6A1 | 8.16E-02 | 4.99E-02 |
| 6269 | Nrxn2 | NRXN2 | -6.97E-02 | -7.17E-04 |
| 6270 | Cct4 | CCT4 | -1.58E-02 | 1.25E-02 |
| 6271 | Tigd4 | TIGD4 | 4.45E-02 | -2.06E-02 |
| 6272 | Ccl20 | CCL20 | 3.76E-02 | 5.92E-02 |
| 6273 | Cdc26 | CDC26 | -1.80E-02 | -9.57E-03 |
| 6274 | Arl4c | ARL4C | 4.91E-02 | 1.29E-01 |
| 6275 | Asap2 | ASAP2 | -2.43E-02 | 1.10E-02 |
| 6276 | Zbtb33 | ZBTB33 | -2.36E-02 | 6.16E-03 |
| 6277 | B4galt2 | B4GALT2 | -4.37E-02 | 2.97E-02 |
| 6278 | Ampd3 | AMPD3 | 5.93E-02 | 2.51E-02 |
| 6279 | Tcf3 | TCF3 | -1.87E-02 | 1.32E-02 |
| 6280 | Rph3al | RPH3AL | 2.17E-02 | -1.56E-02 |
| 6281 | Kl | KL | 3.31E-02 | -2.91E-02 |
| 6282 | Tbc1d10b | TBC1D10B | -1.46E-02 | 6.69E-03 |
| 6283 | Snrnp35 | SNRNP35 | 2.59E-02 | -1.73E-02 |
| 6284 | Rnase4 | RNASE4 | 5.60E-02 | 6.14E-02 |
| 6285 | Lamtor1 | LAMTOR1 | 1.96E-02 | -1.45E-02 |
| 6286 | Ptprq | PTPRQ | 2.04E-02 | 5.55E-03 |
| 6287 | Ppil1 | PPIL1 | 2.50E-02 | 2.61E-02 |
| 6288 | C2cd4a | C2CD4A | -3.95E-02 | 1.42E-03 |
| 6289 | Nrcam | NRCAM | 8.85E-02 | -4.96E-02 |
| 6290 | Kremen1 | KREMEN1 | 2.23E-02 | -1.99E-02 |
| 6291 | Atat1 | ATAT1 | -2.14E-02 | 7.46E-03 |
| 6292 | Espn | ESPN | -2.48E-02 | -2.71E-02 |
| 6293 | H2-DMa | HLA-DMA | -9.13E-02 | 1.07E-01 |
| 6294 | Bloc1s6 | BLOC1S6 | 1.58E-02 | -1.76E-02 |
| 6295 | Ccdc88a | CCDC88A | -3.77E-02 | 2.53E-03 |
| 6296 | Osbp2 | OSBP2 | 7.18E-02 | -6.22E-02 |
| 6297 | Cd163 | CD163 | 7.75E-02 | 1.85E-01 |
| 6298 | Gpx6 | GPX6 | -7.38E-02 | 2.74E-02 |
| 6299 | Ttc23 | TTC23 | 2.91E-02 | 2.15E-02 |
| 6300 | Slc22a18 | SLC22A18 | -2.86E-02 | -2.86E-02 |
| 6301 | Itpa | ITPA | 1.76E-02 | 5.65E-03 |
| 6302 | Srcin1 | SRCIN1 | -3.81E-02 | -3.33E-02 |
| 6303 | Abca3 | ABCA3 | 2.46E-02 | -1.50E-02 |
| 6304 | Prrt4 | PRRT4 | 3.62E-02 | -1.39E-02 |

| | | | | |
|---|---|---|---|---|
| 6305 | Nrg2 | NRG2 | -4.45E-02 | 5.14E-03 |
| 6306 | Emd | EMD | 2.69E-02 | 1.12E-02 |
| 6307 | Rftn1 | RFTN1 | -5.22E-02 | 3.58E-02 |
| 6308 | Pold1 | POLD1 | -4.34E-02 | 4.55E-02 |
| 6309 | Cbl | CBL | -1.90E-02 | -7.46E-04 |
| 6310 | Ska1 | SKA1 | 9.82E-02 | 7.53E-02 |
| 6311 | Dach2 | DACH2 | -2.01E-02 | 3.68E-04 |
| 6312 | Sybu | SYBU | 4.89E-02 | 2.95E-03 |
| 6313 | Sec61g | SEC61G | 2.12E-02 | 1.73E-03 |
| 6314 | Ptrh2 | PTRH2 | -2.12E-02 | 1.54E-02 |
| 6315 | Gnptab | GNPTAB | 2.33E-02 | -8.62E-03 |
| 6316 | Trmt10b | TRMT10B | 2.27E-02 | -2.51E-02 |
| 6317 | Mms19 | MMS19 | 1.45E-02 | -1.47E-02 |
| 6318 | Rprd1b | RPRD1B | -1.77E-02 | -6.98E-03 |
| 6319 | Adal | ADAL | -2.98E-02 | 1.26E-02 |
| 6320 | Senp8 | SENP8 | -4.72E-02 | -1.99E-02 |
| 6321 | Lrrc2 | LRRC2 | -2.70E-02 | -4.97E-03 |
| 6322 | Trio | TRIO | -2.81E-02 | -3.26E-02 |
| 6323 | Lgals8 | LGALS8 | 1.69E-02 | -6.51E-03 |
| 6324 | Rnf182 | RNF182 | 1.12E-01 | -1.03E-01 |
| 6325 | Ap1m1 | AP1M1 | 1.94E-02 | -2.39E-03 |
| 6326 | Rxrg | RXRG | 9.46E-02 | -5.90E-02 |
| 6327 | Sfxn2 | SFXN2 | -2.99E-02 | -1.23E-02 |
| 6328 | Zfp30 | ZFP30 | -3.76E-02 | -1.43E-02 |
| 6329 | Actr3 | ACTR3 | 1.25E-02 | -3.43E-04 |
| 6330 | Cbx2 | CBX2 | -5.52E-02 | -9.20E-03 |
| 6331 | Rps2 | RPS2 | 1.92E-02 | 2.09E-02 |
| 6332 | Sec14l5 | SEC14L5 | -2.28E-02 | -1.41E-02 |
| 6333 | Ncln | NCLN | -1.44E-02 | 7.89E-03 |
| 6334 | Chchd3 | CHCHD3 | 1.97E-02 | -1.47E-02 |
| 6335 | Ifi35 | IFI35 | -3.98E-02 | 2.97E-02 |
| 6336 | Rsph3a | RSPH3 | -1.67E-02 | -6.84E-03 |
| 6337 | Cenpm | CENPM | 1.30E-01 | 7.07E-02 |
| 6338 | Vcpip1 | VCPIP1 | -1.97E-02 | 6.68E-03 |
| 6339 | Lmln | LMLN | -3.06E-02 | -1.33E-02 |
| 6340 | Fam3b | FAM3B | -2.28E-02 | 2.10E-02 |
| 6341 | Cdk5 | CDK5 | 1.68E-02 | -8.01E-03 |
| 6342 | Plk4 | PLK4 | 6.45E-02 | 6.41E-02 |
| 6343 | Pccb | PCCB | 2.58E-02 | -3.30E-02 |
| 6344 | Arhgef17 | ARHGEF17 | -2.27E-02 | -1.86E-02 |
| 6345 | Maml1 | MAML1 | -1.83E-02 | 2.12E-03 |
| 6346 | Styxl1 | STYXL1 | 5.83E-02 | 7.56E-03 |
| 6347 | Adora3 | ADORA3 | -2.70E-02 | 3.97E-02 |
| 6348 | Ydjc | YDJC | -4.83E-02 | -1.22E-02 |
| 6349 | Zfp386 | ZNF519 | -1.92E-02 | 1.47E-02 |
| 6350 | Lgals9 | LGALS9B | -3.73E-02 | 2.96E-02 |
| 6351 | Tcf24 | TCF24 | -4.96E-02 | 1.79E-02 |
| 6352 | Klhl4 | KLHL4 | -7.69E-02 | -1.02E-02 |
| 6353 | Isoc1 | ISOC1 | 2.11E-02 | -9.38E-03 |
| 6354 | Zdhhc18 | ZDHHC18 | -2.65E-02 | 1.31E-02 |
| 6355 | Col9a3 | COL9A3 | -4.59E-02 | 4.63E-03 |
| 6356 | Cd86 | CD86 | -6.21E-02 | 6.89E-02 |
| 6357 | R3hcc1l | R3HCC1L | -1.92E-02 | 7.63E-03 |

| | | | | |
|---|---|---|---|---|
| 6358 | Rrs1 | RRS1 | -2.87E-02 | 3.28E-02 |
| 6359 | Lhfpl3 | LHFPL3 | -6.23E-02 | -1.34E-02 |
| 6360 | Fpgs | FPGS | 2.30E-02 | -8.00E-03 |
| 6361 | F2rl1 | F2RL1 | 5.30E-02 | -8.98E-03 |
| 6362 | Oxa1l | OXA1L | -1.89E-02 | -6.93E-03 |
| 6363 | Sult1b1 | SULT1B1 | 3.92E-02 | -2.13E-02 |
| 6364 | Hgs | HGS | -1.18E-02 | 5.72E-03 |
| 6365 | Espnl | ESPNL | 6.77E-02 | -2.51E-02 |
| 6366 | Gan | GAN | 2.65E-02 | -4.01E-03 |
| 6367 | Tmem181a | TMEM181 | 5.00E-02 | -1.27E-02 |
| 6368 | Srp54a | SRP54 | 3.69E-02 | -4.17E-02 |
| 6369 | Mob2 | MOB2 | 1.61E-02 | -3.09E-03 |
| 6370 | Polr3k | POLR3K | 2.07E-02 | -2.88E-03 |
| 6371 | Fam161a | FAM161A | -5.84E-02 | 1.14E-02 |
| 6372 | Agtr2 | AGTR2 | 2.81E-02 | -2.64E-02 |
| 6373 | Zmynd10 | ZMYND10 | -3.31E-02 | -4.81E-02 |
| 6374 | Hras | HRAS | 1.91E-02 | 3.04E-03 |
| 6375 | Sec61a1 | SEC61A1 | 1.30E-02 | -3.69E-03 |
| 6376 | Lrrc52 | LRRC52 | 6.92E-02 | 2.14E-02 |
| 6377 | Dse | DSE | -6.79E-02 | 1.34E-02 |
| 6378 | Lrrc66 | LRRC66 | -5.47E-02 | -1.01E-01 |
| 6379 | Wrn | WRN | -2.28E-02 | -1.95E-02 |
| 6380 | Il34 | IL34 | 8.76E-02 | 6.59E-02 |
| 6381 | Nkain2 | NKAIN2 | -3.47E-02 | 3.65E-04 |
| 6382 | Taf1d | TAF1D | -2.09E-02 | 1.19E-02 |
| 6383 | Chst13 | CHST13 | -6.84E-02 | 3.61E-02 |
| 6384 | Lpar6 | LPAR6 | -3.49E-02 | 3.04E-02 |
| 6385 | Nmt1 | NMT1 | 1.44E-02 | -1.03E-02 |
| 6386 | C1ql3 | C1QL3 | 8.54E-02 | 4.48E-02 |
| 6387 | Kcnb2 | KCNB2 | -3.39E-02 | 1.57E-03 |
| 6388 | Nfkbiz | NFKBIZ | 6.98E-02 | 8.05E-02 |
| 6389 | Clcn2 | CLCN2 | -2.95E-02 | -1.55E-02 |
| 6390 | Zc3h7b | ZC3H7B | 2.45E-02 | -2.42E-02 |
| 6391 | Ovol2 | OVOL2 | -7.39E-02 | 1.66E-02 |
| 6392 | Ggt1 | GGT1 | -2.34E-02 | -1.39E-02 |
| 6393 | Slc25a44 | SLC25A44 | 2.00E-02 | -7.91E-03 |
| 6394 | Fnbp1 | FNBP1 | -1.43E-02 | -1.51E-02 |
| 6395 | Rell2 | RELL2 | 5.72E-02 | -3.80E-02 |
| 6396 | Depdc1a | DEPDC1 | 1.33E-01 | 1.19E-01 |
| 6397 | Qrsl1 | QRSL1 | 2.56E-02 | -1.01E-02 |
| 6398 | Rps9 | RPS9 | -1.63E-02 | 1.75E-02 |
| 6399 | Zfp112 | ZNF112 | 4.51E-02 | -4.49E-02 |
| 6400 | Abcb7 | ABCB7 | 2.36E-02 | 1.10E-04 |
| 6401 | Fam83f | FAM83F | -7.24E-02 | 1.20E-02 |
| 6402 | Farsa | FARSA | 1.78E-02 | 6.27E-03 |
| 6403 | Tomm22 | TOMM22 | -1.24E-02 | 2.54E-04 |
| 6404 | Brd3 | BRD3 | -1.97E-02 | 4.22E-03 |
| 6405 | Clcn5 | CLCN5 | 2.02E-02 | -2.66E-04 |
| 6406 | Cthrc1 | CTHRC1 | -4.06E-02 | 2.90E-02 |
| 6407 | Kcnip4 | KCNIP4 | -2.03E-02 | 1.00E-03 |
| 6408 | Stx8 | STX8 | 1.84E-02 | -2.05E-02 |
| 6409 | Ar | AR | -3.22E-02 | 9.58E-03 |
| 6410 | Trappc1 | TRAPPC1 | 1.99E-02 | 6.41E-03 |

| | | | | |
|---|---|---|---|---|
| 6411 | Entpd5 | ENTPD5 | -2.60E-02 | -1.51E-02 |
| 6412 | Eif2ak3 | EIF2AK3 | 1.73E-02 | -4.14E-03 |
| 6413 | Ppp6c | PPP6C | 1.29E-02 | -4.79E-03 |
| 6414 | Arl9 | ARL9 | -6.10E-02 | -6.88E-02 |
| 6415 | Slc2a9 | SLC2A9 | 4.33E-02 | -1.33E-02 |
| 6416 | Fam210b | FAM210B | -2.15E-02 | -3.13E-02 |
| 6417 | Zap70 | ZAP70 | -6.01E-02 | 5.09E-02 |
| 6418 | Ttn | TTN | -8.02E-02 | 3.62E-02 |
| 6419 | Ttc30a1 | TTC30A | -3.77E-02 | 1.78E-02 |
| 6420 | Jph1 | JPH1 | -6.40E-02 | -4.48E-02 |
| 6421 | Tmem143 | TMEM143 | -2.71E-02 | -8.91E-03 |
| 6422 | Grwd1 | GRWD1 | -2.31E-02 | 3.82E-02 |
| 6423 | Slc25a5 | SLC25A5 | 2.06E-02 | -5.04E-03 |
| 6424 | Ddit3 | DDIT3 | -6.68E-02 | 5.87E-03 |
| 6425 | Exoc8 | EXOC8 | -2.57E-02 | -1.01E-04 |
| 6426 | Cd160 | CD160 | -4.15E-02 | -1.99E-02 |
| 6427 | Slc4a3 | SLC4A3 | 2.06E-02 | 3.60E-02 |
| 6428 | Ctsb | CTSB | 2.02E-02 | 1.41E-03 |
| 6429 | Dnm1l | DNM1L | 1.48E-02 | -3.65E-03 |
| 6430 | Acsl4 | ACSL4 | -2.26E-02 | 7.88E-02 |
| 6431 | Gstcd | GSTCD | -2.71E-02 | 2.85E-03 |
| 6432 | Msx1 | MSX1 | 6.39E-02 | -5.70E-02 |
| 6433 | Vta1 | VTA1 | 1.54E-02 | 4.55E-03 |
| 6434 | Anxa2 | ANXA2 | 6.07E-02 | 8.32E-02 |
| 6435 | Zdhhc24 | ZDHHC24 | 1.55E-02 | -7.44E-03 |
| 6436 | Dnhd1 | DNHD1 | -3.81E-02 | -5.49E-02 |
| 6437 | Prrx2 | PRRX2 | -4.72E-02 | 1.71E-02 |
| 6438 | Polr2g | POLR2G | 1.86E-02 | -1.64E-02 |
| 6439 | Fam162a | FAM162A | 1.89E-02 | -6.64E-03 |
| 6440 | Dnajb4 | DNAJB4 | 3.60E-02 | 6.71E-03 |
| 6441 | Kif2a | KIF2A | 2.45E-02 | 2.03E-02 |
| 6442 | Sh3yl1 | SH3YL1 | 1.53E-02 | -1.98E-02 |
| 6443 | Uroc1 | UROC1 | 2.80E-02 | -2.25E-02 |
| 6444 | B3galnt1 | B3GALNT1 | -6.25E-02 | -8.20E-02 |
| 6445 | Zfp597 | ZNF597 | -4.49E-02 | -7.28E-03 |
| 6446 | Tob1 | TOB1 | 4.22E-02 | -2.38E-02 |
| 6447 | Lars2 | LARS2 | 4.23E-02 | 1.98E-02 |
| 6448 | Jak3 | JAK3 | 5.21E-02 | -2.70E-02 |
| 6449 | Tmem95 | TMEM95 | -3.35E-02 | 1.10E-02 |
| 6450 | Wdr46 | WDR46 | 2.07E-02 | -4.52E-03 |
| 6451 | Gngt2 | GNGT2 | -7.35E-02 | 2.82E-02 |
| 6452 | Dnajb14 | DNAJB14 | 1.69E-02 | -1.07E-02 |
| 6453 | Glrx3 | GLRX3 | 1.63E-02 | -1.33E-02 |
| 6454 | Samm50 | SAMM50 | 1.89E-02 | -1.81E-02 |
| 6455 | Lyn | LYN | -1.93E-02 | 3.54E-02 |
| 6456 | Commd7 | COMMD7 | -1.87E-02 | 1.96E-02 |
| 6457 | Ppp1cb | PPP1CB | 1.59E-02 | -6.22E-03 |
| 6458 | Cacna1g | CACNA1G | -1.10E-01 | -5.49E-03 |
| 6459 | A4galt | A4GALT | -3.37E-02 | 3.74E-03 |
| 6460 | Stxbp4 | STXBP4 | -3.98E-02 | -3.32E-02 |
| 6461 | Nalcn | NALCN | -4.24E-02 | 1.10E-02 |
| 6462 | Serpinf2 | SERPINF2 | 3.14E-02 | 5.38E-02 |
| 6463 | Fbxl3 | FBXL3 | -1.89E-02 | -1.95E-02 |

| | | | | |
|---|---|---|---|---|
| 6464 | Lrfn3 | LRFN3 | -4.19E-02 | 3.77E-03 |
| 6465 | Eva1c | EVA1C | -8.16E-02 | -1.33E-01 |
| 6466 | Rabac1 | RABAC1 | 1.54E-02 | -4.41E-03 |
| 6467 | Agbl5 | AGBL5 | -2.03E-02 | -2.75E-02 |
| 6468 | Pigf | PIGF | -2.50E-02 | 2.50E-02 |
| 6469 | Mrpl32 | MRPL32 | 1.82E-02 | -1.38E-02 |
| 6470 | Cenpj | CENPJ | -3.94E-02 | -1.02E-02 |
| 6471 | Trpm5 | TRPM5 | 2.32E-02 | -1.66E-03 |
| 6472 | Zfp319 | ZNF319 | -2.96E-02 | -1.55E-03 |
| 6473 | Zfp72 | ZNF502 | -4.45E-02 | -8.06E-03 |
| 6474 | Nkrf | NKRF | 4.09E-02 | 5.92E-03 |
| 6475 | App | APP | 1.85E-02 | -1.48E-02 |
| 6476 | Syt9 | SYT9 | -6.21E-02 | 1.07E-01 |
| 6477 | Ralgapa2 | RALGAPA2 | -3.37E-02 | -1.90E-03 |
| 6478 | Cds1 | CDS1 | 1.67E-02 | -1.73E-02 |
| 6479 | Fbxo28 | FBXO28 | 1.92E-02 | -1.09E-02 |
| 6480 | Sdc1 | SDC1 | 5.39E-02 | 4.92E-02 |
| 6481 | Rgs2 | RGS2 | -5.25E-02 | 2.02E-02 |
| 6482 | Tnnt2 | TNNT2 | 8.34E-02 | 1.05E-01 |
| 6483 | Hnrnpm | HNRNPM | -1.68E-02 | -3.72E-03 |
| 6484 | Fcgr2b | FCGR2B | -3.57E-02 | -1.82E-02 |
| 6485 | Ube2o | UBE2O | 2.02E-02 | 1.58E-02 |
| 6486 | Prpf4 | PRPF4 | 1.56E-02 | -7.55E-03 |
| 6487 | Hars | HARS | 1.46E-02 | -7.86E-03 |
| 6488 | Asap1 | ASAP1 | -3.10E-02 | -1.75E-02 |
| 6489 | Ddx49 | DDX49 | 1.63E-02 | 1.27E-03 |
| 6490 | Nol3 | NOL3 | -3.66E-02 | 2.24E-02 |
| 6491 | Rnf11 | RNF11 | 1.81E-02 | -1.26E-02 |
| 6492 | Hs6st3 | HS6ST3 | -3.41E-02 | 2.19E-02 |
| 6493 | Mecr | MECR | 2.35E-02 | -4.16E-02 |
| 6494 | Bdnf | BDNF | -7.27E-02 | 8.02E-02 |
| 6495 | Thoc7 | THOC7 | 1.49E-02 | -6.88E-04 |
| 6496 | Ciz1 | CIZ1 | -1.15E-02 | 6.26E-04 |
| 6497 | Tcf15 | TCF15 | -5.02E-02 | -1.95E-02 |
| 6498 | Ptpn23 | PTPN23 | 1.92E-02 | 9.33E-03 |
| 6499 | Ddah1 | DDAH1 | 2.68E-02 | -7.39E-03 |
| 6500 | Stx7 | STX7 | 1.29E-02 | -2.18E-03 |
| 6501 | Ormdl1 | ORMDL1 | 2.67E-02 | -1.09E-03 |
| 6502 | Mcm3 | MCM3 | 8.67E-02 | 1.28E-01 |
| 6503 | Zdhhc6 | ZDHHC6 | -1.42E-02 | 3.15E-03 |
| 6504 | Trnt1 | TRNT1 | 2.10E-02 | 6.85E-03 |
| 6505 | Cops4 | COPS4 | 1.48E-02 | -1.11E-02 |
| 6506 | Trpc3 | TRPC3 | -6.91E-02 | 2.50E-03 |
| 6507 | Xpo5 | XPO5 | -1.40E-02 | -1.59E-03 |
| 6508 | Gpr107 | GPR107 | 1.30E-02 | -5.36E-03 |
| 6509 | Mapk10 | MAPK10 | 5.57E-02 | -1.01E-01 |
| 6510 | Corin | CORIN | -2.73E-01 | -5.02E-02 |
| 6511 | Ss18l1 | SS18L1 | 2.88E-02 | -1.99E-02 |
| 6512 | Pstpip1 | PSTPIP1 | -7.88E-02 | 1.00E-01 |
| 6513 | C2cd2l | C2CD2L | 2.24E-02 | -1.42E-02 |
| 6514 | Mtfr1l | MTFR1L | 1.56E-02 | -1.15E-02 |
| 6515 | Upk1a | UPK1A | 2.76E-01 | 8.28E-02 |
| 6516 | Emilin3 | EMILIN3 | 2.30E-02 | -2.11E-02 |

| | | | | |
|---|---|---|---|---|
| 6517 | Fam185a | FAM185A | 2.72E-02 | 3.87E-03 |
| 6518 | Prpf38a | PRPF38A | 1.88E-02 | 2.00E-02 |
| 6519 | Ryr1 | RYR1 | -2.68E-02 | 3.55E-02 |
| 6520 | Dpp10 | DPP10 | -2.72E-02 | -7.50E-03 |
| 6521 | Mast2 | MAST2 | 2.08E-02 | -6.95E-03 |
| 6522 | Proser1 | PROSER1 | 1.92E-02 | -8.42E-03 |
| 6523 | Hgfac | HGFAC | -4.03E-02 | -3.15E-02 |
| 6524 | Ulk2 | ULK2 | 1.41E-02 | -2.42E-03 |
| 6525 | Osgin1 | OSGIN1 | -5.97E-02 | 3.97E-02 |
| 6526 | Fbxl13 | FBXL13 | 2.71E-02 | 3.05E-03 |
| 6527 | Nrbf2 | NRBF2 | 2.21E-02 | 2.26E-02 |
| 6528 | Clk2 | CLK2 | -1.46E-02 | -2.15E-03 |
| 6529 | Hspbp1 | HSPBP1 | 1.61E-02 | -8.24E-03 |
| 6530 | Sun2 | SUN2 | 1.92E-02 | -3.69E-03 |
| 6531 | Fnta | FNTA | 1.11E-02 | -3.38E-03 |
| 6532 | Sgta | SGTA | 1.47E-02 | -7.47E-03 |
| 6533 | Clock | CLOCK | 1.96E-02 | -1.16E-03 |
| 6534 | Ubtd2 | UBTD2 | 4.93E-02 | 1.28E-02 |
| 6535 | Rrp8 | RRP8 | -2.43E-02 | 1.43E-02 |
| 6536 | Nipsnap3b | NIPSNAP3A | 2.28E-02 | 1.93E-02 |
| 6537 | Arhgef4 | ARHGEF4 | 6.29E-02 | -2.97E-02 |
| 6538 | Stk11 | STK11 | -1.51E-02 | -5.51E-03 |
| 6539 | Lzic | LZIC | -2.49E-02 | -4.11E-03 |
| 6540 | Noc2l | NOC2L | -1.32E-02 | 4.24E-03 |
| 6541 | Hmox2 | HMOX2 | 1.35E-02 | -1.88E-02 |
| 6542 | 2810459M11Rik | C2orf72 | 7.34E-02 | 6.87E-02 |
| 6543 | Ndc80 | NDC80 | 1.26E-01 | 1.63E-01 |
| 6544 | Krt19 | KRT19 | 2.29E-01 | 1.36E-01 |
| 6545 | Prmt6 | PRMT6 | 2.54E-02 | 1.56E-02 |
| 6546 | Haus8 | HAUS8 | -5.71E-02 | 8.28E-02 |
| 6547 | Rab32 | RAB32 | -3.18E-02 | 3.25E-02 |
| 6548 | Fitm1 | FITM1 | 6.04E-02 | -8.07E-03 |
| 6549 | Nr2f1 | NR2F1 | -5.32E-02 | 8.78E-02 |
| 6550 | Eps15l1 | EPS15L1 | 1.25E-02 | -1.06E-02 |
| 6551 | Ubxn4 | UBXN4 | 1.66E-02 | -2.38E-03 |
| 6552 | Marveld2 | MARVELD2 | 2.87E-02 | -3.77E-02 |
| 6553 | Slc11a2 | SLC11A2 | 2.37E-02 | -2.11E-02 |
| 6554 | Aasdhppt | AASDHPPT | 1.80E-02 | -7.46E-03 |
| 6555 | Ireb2 | IREB2 | 1.90E-02 | -9.67E-03 |
| 6556 | Fscn2 | FSCN2 | 3.59E-02 | -7.82E-03 |
| 6557 | Ghitm | GHITM | 1.84E-02 | -1.87E-02 |
| 6558 | Hmgcs2 | HMGCS2 | 9.69E-02 | -2.23E-02 |
| 6559 | Arl3 | ARL3 | 1.31E-02 | -7.09E-03 |
| 6560 | Tfap4 | TFAP4 | -7.26E-02 | -5.56E-03 |
| 6561 | Rer1 | RER1 | 1.35E-02 | -1.38E-02 |
| 6562 | Mrfap1 | MRFAP1 | 1.34E-02 | -7.60E-03 |
| 6563 | Hoxd9 | HOXD9 | 2.82E-02 | 3.16E-02 |
| 6564 | Uprt | UPRT | -6.58E-02 | -3.35E-02 |
| 6565 | Dnaja3 | DNAJA3 | 1.72E-02 | -1.90E-02 |
| 6566 | Agpat4 | AGPAT4 | -4.18E-02 | 3.49E-02 |
| 6567 | Upf3a | UPF3A | 1.45E-02 | -1.08E-02 |
| 6568 | 4921524J17Rik | C16orf87 | -3.03E-02 | 2.08E-03 |
| 6569 | Kdm2a | KDM2A | -1.27E-02 | -6.25E-03 |

| | | | | |
|---|---|---|---|---|
| 6570 | Pard3 | PARD3 | -2.06E-02 | -1.82E-02 |
| 6571 | Cdc42se1 | CDC42SE1 | 2.11E-02 | 1.09E-02 |
| 6572 | Ninj1 | NINJ1 | 1.78E-02 | 6.68E-03 |
| 6573 | Lum | LUM | -9.27E-02 | 1.07E-01 |
| 6574 | Lpxn | LPXN | -5.26E-02 | 6.04E-02 |
| 6575 | Map3k15 | MAP3K15 | -2.34E-02 | -2.64E-02 |
| 6576 | Kri1 | KRI1 | -1.46E-02 | -2.09E-03 |
| 6577 | Gria2 | GRIA2 | -2.48E-02 | -2.01E-02 |
| 6578 | Rbl1 | RBL1 | -4.05E-02 | 2.77E-02 |
| 6579 | Kcnk10 | KCNK10 | -2.09E-02 | -5.55E-03 |
| 6580 | Phlda1 | PHLDA1 | -3.65E-02 | 1.58E-02 |
| 6581 | Sco2 | SCO2 | -5.50E-02 | 8.19E-02 |
| 6582 | Foxo6 | FOXO6 | -3.83E-02 | 9.86E-03 |
| 6583 | Uevld | UEVLD | 2.27E-02 | 5.37E-03 |
| 6584 | Galk1 | GALK1 | 2.72E-02 | -1.72E-02 |
| 6585 | Gpkow | GPKOW | 1.27E-02 | 4.12E-03 |
| 6586 | Slc26a11 | SLC26A11 | 2.56E-02 | -2.32E-02 |
| 6587 | Eya1 | EYA1 | -4.14E-02 | 7.68E-03 |
| 6588 | Rap1a | RAP1A | 1.62E-02 | 9.08E-03 |
| 6589 | Trmt10c | TRMT10C | -2.03E-02 | 2.78E-03 |
| 6590 | Usp12 | USP12 | -1.59E-02 | -1.54E-02 |
| 6591 | St8sia2 | ST8SIA2 | -4.91E-02 | 1.22E-01 |
| 6592 | Sgcb | SGCB | -3.33E-02 | -4.43E-03 |
| 6593 | Xrcc1 | XRCC1 | 1.53E-02 | -1.34E-03 |
| 6594 | Gtf3a | GTF3A | -1.75E-02 | 8.91E-03 |
| 6595 | Ccdc47 | CCDC47 | -1.37E-02 | -1.19E-02 |
| 6596 | Aar2 | AAR2 | 1.94E-02 | -5.04E-03 |
| 6597 | Efcc1 | EFCC1 | -7.09E-02 | -3.10E-02 |
| 6598 | Prmt5 | PRMT5 | 1.60E-02 | -3.01E-03 |
| 6599 | Aanat | AANAT | 2.73E-02 | -2.83E-02 |
| 6600 | Trim9 | TRIM9 | 2.99E-02 | 1.59E-02 |
| 6601 | Nsg2 | HMP19 | -6.44E-02 | -2.66E-02 |
| 6602 | Apoa2 | APOA2 | 7.35E-02 | -1.77E-02 |
| 6603 | Bub1b | BUB1B | 1.32E-01 | 1.06E-01 |
| 6604 | Kpna4 | KPNA4 | -1.58E-02 | 1.94E-02 |
| 6605 | Hnrnpa2b1 | HNRNPA2B1 | 1.24E-02 | -8.87E-03 |
| 6606 | Slitrk1 | SLITRK1 | 9.17E-02 | -4.45E-02 |
| 6607 | Coq3 | COQ3 | 2.44E-02 | -2.36E-02 |
| 6608 | Slc25a3 | SLC25A3 | 1.66E-02 | -9.77E-03 |
| 6609 | Slc25a40 | SLC25A40 | -1.85E-02 | -1.81E-03 |
| 6610 | Glce | GLCE | 2.64E-02 | 3.22E-03 |
| 6611 | Mospd3 | MOSPD3 | 1.82E-02 | -1.79E-03 |
| 6612 | Adra1d | ADRA1D | 4.89E-02 | -1.30E-01 |
| 6613 | Zfp772 | ZNF773 | 3.38E-02 | -1.46E-02 |
| 6614 | Ano10 | ANO10 | 1.87E-02 | 1.07E-03 |
| 6615 | Nox1 | NOX1 | -3.20E-02 | 2.27E-02 |
| 6616 | Strn4 | STRN4 | 1.06E-02 | -1.12E-02 |
| 6617 | Pcdha1 | PCDHA1 | -3.04E-02 | 9.07E-03 |
| 6618 | Eif3l | EIF3L | 1.61E-02 | 2.99E-03 |
| 6619 | Stau1 | STAU1 | -1.20E-02 | 1.12E-03 |
| 6620 | Eapp | EAPP | 1.58E-02 | -3.86E-03 |
| 6621 | Fry | FRY | 6.60E-02 | -2.61E-02 |
| 6622 | Wdr90 | WDR90 | -2.47E-02 | 2.92E-02 |

| | | | | |
|---|---|---|---|---|
| 6623 | Prps2 | PRPS2 | -1.81E-02 | -3.36E-02 |
| 6624 | Rfx1 | RFX1 | -2.76E-02 | -7.93E-03 |
| 6625 | Adamts20 | ADAMTS20 | 4.29E-02 | -6.76E-02 |
| 6626 | Nkx6-1 | NKX6-1 | -3.39E-02 | 6.30E-02 |
| 6627 | Fbn1 | FBN1 | 9.49E-02 | 3.48E-02 |
| 6628 | Ehd1 | EHD1 | 2.75E-02 | -5.47E-03 |
| 6629 | Rbbp4 | RBBP4 | -1.43E-02 | 2.55E-04 |
| 6630 | Snx8 | SNX8 | -2.39E-02 | -3.38E-02 |
| 6631 | Wdr5b | WDR5B | -3.29E-02 | -1.35E-02 |
| 6632 | Hoxb5 | HOXB5 | -6.56E-02 | 3.51E-04 |
| 6633 | Nup85 | NUP85 | -1.55E-02 | 8.50E-03 |
| 6634 | Gdpd5 | GDPD5 | -3.70E-02 | -2.50E-02 |
| 6635 | Szt2 | SZT2 | 1.88E-02 | -2.64E-02 |
| 6636 | Zbtb45 | ZBTB45 | -2.64E-02 | 1.02E-02 |
| 6637 | Ipmk | IPMK | -3.53E-02 | -3.64E-02 |
| 6638 | Vgll3 | VGLL3 | 9.20E-02 | -2.38E-03 |
| 6639 | Traf5 | TRAF5 | -7.06E-02 | 2.21E-02 |
| 6640 | Malsu1 | MALSU1 | -1.86E-02 | 7.66E-03 |
| 6641 | Clec12a | CLEC12A | -9.43E-02 | 1.57E-01 |
| 6642 | Tdrd3 | TDRD3 | 1.52E-02 | 1.46E-02 |
| 6643 | Hibch | HIBCH | -1.87E-02 | -2.89E-02 |
| 6644 | Gtpbp4 | GTPBP4 | -1.94E-02 | 2.12E-02 |
| 6645 | Arfgap2 | ARFGAP2 | 1.03E-02 | -8.62E-03 |
| 6646 | Tyms | TYMS | 3.89E-02 | 4.30E-03 |
| 6647 | Gps2 | GPS2 | -1.27E-02 | 1.02E-02 |
| 6648 | Yrdc | YRDC | 3.50E-02 | 1.44E-02 |
| 6649 | Gins2 | GINS2 | 9.85E-02 | 1.13E-01 |
| 6650 | Ctsh | CTSH | -1.40E-02 | -1.46E-02 |
| 6651 | Plxdc2 | PLXDC2 | 2.18E-02 | -2.65E-02 |
| 6652 | Nox4 | NOX4 | -5.10E-02 | 1.09E-02 |
| 6653 | Usp47 | USP47 | 1.42E-02 | -1.69E-03 |
| 6654 | Gstt2 | GSTT2 | -3.35E-02 | -3.50E-02 |
| 6655 | Gstt2 | GSTT2B | -3.35E-02 | -3.50E-02 |
| 6656 | Aldoc | ALDOC | 9.69E-02 | -8.31E-02 |
| 6657 | Prrg4 | PRRG4 | 6.84E-02 | 7.11E-02 |
| 6658 | Samd3 | SAMD3 | -3.66E-02 | 7.34E-03 |
| 6659 | Nfatc4 | NFATC4 | -6.84E-02 | 5.53E-02 |
| 6660 | Slu7 | SLU7 | -1.32E-02 | -1.38E-03 |
| 6661 | Rab11fip3 | RAB11FIP3 | -3.11E-02 | -1.11E-02 |
| 6662 | H2-Ab1 | HLA-DQB1 | -6.94E-02 | 1.01E-01 |
| 6663 | Ranbp1 | RANBP1 | 1.80E-02 | 2.28E-02 |
| 6664 | Svop | SVOP | 4.48E-02 | -9.28E-02 |
| 6665 | Tubd1 | TUBD1 | 4.08E-02 | -1.19E-03 |
| 6666 | Rab28 | RAB28 | -1.49E-02 | -2.02E-02 |
| 6667 | Irf2bp1 | IRF2BP1 | -1.63E-02 | 1.02E-02 |
| 6668 | Inpp5j | INPP5J | -2.59E-02 | 1.81E-02 |
| 6669 | Mrps35 | MRPS35 | 1.73E-02 | -2.78E-02 |
| 6670 | Syn2 | SYN2 | -4.12E-02 | 3.53E-02 |
| 6671 | Ssna1 | SSNA1 | 1.89E-02 | 6.76E-03 |
| 6672 | Cdk7 | CDK7 | -1.39E-02 | 8.56E-03 |
| 6673 | Rsl24d1 | RSL24D1 | -2.87E-02 | 3.11E-02 |
| 6674 | Cacul1 | CACUL1 | -2.11E-02 | -4.73E-03 |
| 6675 | Dync1i1 | DYNC1I1 | -6.35E-02 | -1.19E-02 |

| | | | | |
|---|---|---|---|---|
| 6676 | Nipal1 | NIPAL1 | 2.01E-01 | -9.60E-02 |
| 6677 | Fam98b | FAM98B | -1.30E-02 | 9.66E-03 |
| 6678 | Frmpd1 | FRMPD1 | -4.34E-02 | -1.79E-02 |
| 6679 | Gtpbp1 | GTPBP1 | 1.33E-02 | -2.11E-02 |
| 6680 | Zfp64 | ZFP64 | -2.74E-02 | 9.37E-03 |
| 6681 | Rassf7 | RASSF7 | -2.38E-02 | -3.91E-03 |
| 6682 | Fjx1 | FJX1 | 5.77E-02 | 5.67E-02 |
| 6683 | Kcna2 | KCNA2 | -5.06E-02 | -1.09E-02 |
| 6684 | Cpeb2 | CPEB2 | 2.10E-02 | -5.67E-04 |
| 6685 | Ubash3a | UBASH3A | -5.14E-02 | 1.96E-02 |
| 6686 | Ecm1 | ECM1 | 4.04E-02 | -1.14E-01 |
| 6687 | Ankle2 | ANKLE2 | -1.35E-02 | -1.02E-02 |
| 6688 | Cyp26c1 | CYP26C1 | -3.30E-02 | 2.00E-02 |
| 6689 | Smtnl2 | SMTNL2 | 3.48E-02 | 1.93E-03 |
| 6690 | Mpc2 | MPC2 | 1.39E-02 | -3.61E-03 |
| 6691 | Slc35a4 | SLC35A4 | 1.44E-02 | -2.88E-03 |
| 6692 | Litaf | LITAF | 7.59E-02 | 4.00E-02 |
| 6693 | Zfp27 | ZNF585A | -2.65E-02 | -3.70E-03 |
| 6694 | Trpm4 | TRPM4 | 3.57E-02 | 8.09E-03 |
| 6695 | Rabl3 | RABL3 | 2.74E-02 | -2.96E-02 |
| 6696 | Rnf17 | RNF17 | -3.87E-02 | 1.07E-02 |
| 6697 | Plxna4 | PLXNA4 | 4.58E-02 | -1.77E-02 |
| 6698 | Id2 | ID2 | -3.31E-02 | 7.79E-02 |
| 6699 | Sod3 | SOD3 | -2.50E-02 | 1.69E-03 |
| 6700 | Rsrc1 | RSRC1 | 1.50E-02 | -2.11E-02 |
| 6701 | Ankrd42 | ANKRD42 | -3.30E-02 | -3.37E-02 |
| 6702 | Ubac2 | UBAC2 | 1.70E-02 | -1.93E-02 |
| 6703 | Bnip3l | BNIP3L | -1.62E-02 | -2.53E-02 |
| 6704 | Dynll2 | DYNLL2 | -2.04E-02 | -1.68E-02 |
| 6705 | L3hypdh | L3HYPDH | -2.77E-02 | 4.08E-03 |
| 6706 | Rassf5 | RASSF5 | -3.50E-02 | 7.18E-02 |
| 6707 | Clec3b | CLEC3B | -1.01E-01 | -2.15E-02 |
| 6708 | Shisa4 | SHISA4 | 3.35E-02 | 7.72E-02 |
| 6709 | Calca | CALCB | 8.15E-02 | 1.59E-02 |
| 6710 | Rmi2 | RMI2 | -5.48E-02 | 1.34E-01 |
| 6711 | Slc25a36 | SLC25A36 | -2.01E-02 | -3.21E-02 |
| 6712 | Plin3 | PLIN3 | -1.54E-02 | 2.33E-03 |
| 6713 | Cdk5rap2 | CDK5RAP2 | -2.31E-02 | -6.94E-03 |
| 6714 | Sh3gl2 | SH3GL2 | -2.58E-02 | -5.23E-03 |
| 6715 | Mtus2 | MTUS2 | 1.02E-01 | 6.85E-02 |
| 6716 | Cdh26 | CDH26 | -2.37E-02 | 1.52E-02 |
| 6717 | Pigl | PIGL | 2.89E-02 | 2.39E-02 |
| 6718 | Ppef1 | PPEF1 | -2.23E-02 | 4.47E-03 |
| 6719 | Ncmap | NCMAP | 7.46E-02 | 1.42E-02 |
| 6720 | Copg1 | COPG1 | 1.23E-02 | -1.24E-02 |
| 6721 | Cdt1 | CDT1 | 8.48E-02 | 2.32E-02 |
| 6722 | Ppm1h | PPM1H | 2.29E-02 | -3.30E-02 |
| 6723 | Atxn7l3b | ATXN7L3B | -1.33E-02 | -6.53E-03 |
| 6724 | Trim59 | TRIM59 | 9.73E-02 | 1.51E-01 |
| 6725 | Rnaseh2c | RNASEH2C | -2.30E-02 | 2.32E-02 |
| 6726 | Spag7 | SPAG7 | 1.43E-02 | -6.85E-03 |
| 6727 | Ubr7 | UBR7 | 1.22E-02 | -4.69E-03 |
| 6728 | Khk | KHK | 2.62E-02 | -1.90E-02 |

| | | | | |
|---|---|---|---|---|
| 6729 | Slc4a1ap | SLC4A1AP | -1.43E-02 | 6.07E-03 |
| 6730 | Slc13a2 | SLC13A2 | 4.47E-02 | -1.68E-02 |
| 6731 | Gins1 | GINS1 | 5.69E-02 | 9.81E-02 |
| 6732 | Fam151b | FAM151B | -3.19E-02 | -4.10E-02 |
| 6733 | Zfp593 | ZNF593 | 3.68E-02 | -7.53E-03 |
| 6734 | Fastkd2 | FASTKD2 | -2.01E-02 | -1.86E-02 |
| 6735 | Dtwd2 | DTWD2 | -3.46E-02 | 6.92E-03 |
| 6736 | Igsf10 | IGSF10 | -8.44E-02 | 4.90E-02 |
| 6737 | Nr6a1 | NR6A1 | 2.92E-02 | -1.21E-02 |
| 6738 | Triap1 | TRIAP1 | 2.07E-02 | -6.45E-03 |
| 6739 | Chp2 | CHP2 | -3.58E-02 | -1.22E-02 |
| 6740 | Bcl11b | BCL11B | -6.65E-02 | 1.51E-02 |
| 6741 | Fam167a | FAM167A | 1.18E-01 | 1.27E-01 |
| 6742 | Bckdha | BCKDHA | 2.48E-02 | -2.73E-02 |
| 6743 | Tbc1d12 | TBC1D12 | 2.55E-02 | 1.08E-03 |
| 6744 | Mcm6 | MCM6 | -4.75E-02 | 4.76E-02 |
| 6745 | Atp5j2 | ATP5J2 | 1.61E-02 | -1.26E-02 |
| 6746 | Kcnc2 | KCNC2 | -4.75E-02 | -5.15E-02 |
| 6747 | Stmn1 | STMN1 | -5.63E-02 | 8.78E-02 |
| 6748 | Kank4 | KANK4 | -4.78E-02 | -2.83E-02 |
| 6749 | Exosc4 | EXOSC4 | 1.71E-02 | -7.11E-03 |
| 6750 | Jun | JUN | 7.14E-02 | 6.40E-03 |
| 6751 | Cers2 | CERS2 | 1.36E-02 | -1.70E-03 |
| 6752 | Gskip | GSKIP | 2.76E-02 | -8.53E-03 |
| 6753 | Vps26a | VPS26A | 1.81E-02 | -8.50E-03 |
| 6754 | Acads | ACADS | 2.36E-02 | -2.48E-02 |
| 6755 | Baz2b | BAZ2B | -2.05E-02 | -8.99E-03 |
| 6756 | Ghrl | GHRL | 2.92E-02 | -2.88E-02 |
| 6757 | Parp3 | PARP3 | 3.66E-02 | -3.01E-03 |
| 6758 | Cytip | CYTIP | -4.09E-02 | -1.20E-02 |
| 6759 | Tm9sf3 | TM9SF3 | -1.34E-02 | -9.66E-03 |
| 6760 | Casq2 | CASQ2 | 4.93E-02 | 7.62E-04 |
| 6761 | Rnf38 | RNF38 | 1.48E-02 | 2.94E-03 |
| 6762 | Fastkd5 | FASTKD5 | -4.39E-02 | 4.37E-02 |
| 6763 | Atic | ATIC | -1.47E-02 | -4.54E-04 |
| 6764 | Slmap | SLMAP | 1.44E-02 | -1.52E-02 |
| 6765 | Sema4c | SEMA4C | 4.53E-02 | 8.40E-03 |
| 6766 | Bin2 | BIN2 | -8.38E-02 | 1.43E-01 |
| 6767 | Nt5dc3 | NT5DC3 | 3.19E-02 | 4.58E-02 |
| 6768 | Cep19 | CEP19 | -1.31E-02 | -1.22E-02 |
| 6769 | Dut | DUT | -3.01E-02 | 1.63E-02 |
| 6770 | Tecta | TECTA | 4.34E-02 | -1.93E-03 |
| 6771 | Fxyd1 | FXYD1 | -5.13E-02 | 4.39E-02 |
| 6772 | Mrpl30 | MRPL30 | 1.57E-02 | 1.63E-03 |
| 6773 | Abcb10 | ABCB10 | -1.91E-02 | -6.58E-03 |
| 6774 | Slc46a2 | SLC46A2 | 4.13E-02 | -2.09E-02 |
| 6775 | Rin1 | RIN1 | 6.86E-02 | 1.04E-01 |
| 6776 | Fastkd3 | FASTKD3 | 2.06E-02 | -2.37E-02 |
| 6777 | Ccdc150 | CCDC150 | 4.70E-02 | -6.24E-02 |
| 6778 | Brca1 | BRCA1 | 1.13E-01 | 1.73E-01 |
| 6779 | Il4i1 | IL4I1 | -5.28E-02 | 1.82E-02 |
| 6780 | Nt5m | NT5M | 2.04E-02 | -1.40E-02 |
| 6781 | Rhbdd1 | RHBDD1 | 1.75E-02 | -7.05E-03 |

| | | | | |
|---|---|---|---|---|
| 6782 | Ctbs | CTBS | -2.95E-02 | -2.36E-02 |
| 6783 | Mef2a | MEF2A | 1.46E-02 | -9.67E-03 |
| 6784 | Vps39 | VPS39 | 1.31E-02 | -1.79E-02 |
| 6785 | Rnf128 | RNF128 | 1.69E-02 | -1.03E-02 |
| 6786 | Orc1 | ORC1 | 5.98E-02 | 1.17E-01 |
| 6787 | Chac1 | CHAC1 | -1.48E-01 | 6.99E-02 |
| 6788 | Chek2 | CHEK2 | 4.29E-02 | 5.65E-02 |
| 6789 | Map4 | MAP4 | 1.46E-02 | 4.73E-03 |
| 6790 | Gns | GNS | 2.23E-02 | -1.38E-02 |
| 6791 | Sox8 | SOX8 | -5.87E-02 | 1.61E-02 |
| 6792 | Arl6ip6 | ARL6IP6 | -2.18E-02 | 1.72E-02 |
| 6793 | Gprin2 | GPRIN2 | 8.17E-02 | -5.10E-02 |
| 6794 | Slc15a3 | SLC15A3 | -7.27E-02 | 5.08E-02 |
| 6795 | Sub1 | SUB1 | 1.38E-02 | 3.12E-03 |
| 6796 | Rassf9 | RASSF9 | -4.39E-02 | -1.08E-02 |
| 6797 | Ripk1 | RIPK1 | 1.63E-02 | 1.54E-02 |
| 6798 | Mrpl55 | MRPL55 | 1.68E-02 | -1.61E-02 |
| 6799 | Cggbp1 | CGGBP1 | -1.72E-02 | -1.57E-04 |
| 6800 | Bbs1 | BBS1 | -1.76E-02 | -3.13E-02 |
| 6801 | Eif4g2 | EIF4G2 | 1.66E-02 | -1.15E-02 |
| 6802 | Dmap1 | DMAP1 | 1.68E-02 | -3.26E-02 |
| 6803 | Zfp592 | ZNF592 | -1.23E-02 | 5.64E-03 |
| 6804 | Stc2 | STC2 | 5.09E-02 | 2.46E-02 |
| 6805 | Dapk1 | DAPK1 | -2.45E-02 | 4.60E-03 |
| 6806 | Supt5 | SUPT5H | 1.20E-02 | -1.07E-02 |
| 6807 | Tmc4 | TMC4 | -3.91E-02 | 1.53E-03 |
| 6808 | Mtmr1 | MTMR1 | 1.50E-02 | 1.44E-03 |
| 6809 | Ostf1 | OSTF1 | 1.63E-02 | -2.59E-03 |
| 6810 | Cyp20a1 | CYP20A1 | -2.01E-02 | -1.09E-02 |
| 6811 | Zfp82 | ZFP82 | -3.63E-02 | -7.30E-03 |
| 6812 | Xiap | XIAP | -1.33E-02 | -5.02E-03 |
| 6813 | Pabpc1 | PABPC1 | -2.13E-02 | 2.45E-02 |
| 6814 | Sdha | SDHA | 2.13E-02 | -2.48E-02 |
| 6815 | Arhgap23 | ARHGAP23 | 4.03E-02 | -1.01E-02 |
| 6816 | Mmp23 | MMP23B | -4.51E-02 | 1.85E-02 |
| 6817 | Ankrd6 | ANKRD6 | 7.69E-02 | 6.69E-02 |
| 6818 | Nfe2l2 | NFE2L2 | -1.54E-02 | 3.07E-02 |
| 6819 | Aff4 | AFF4 | 1.71E-02 | -1.11E-02 |
| 6820 | Fmnl3 | FMNL3 | 3.74E-02 | -6.93E-03 |
| 6821 | Ska3 | SKA3 | 1.26E-01 | 1.40E-01 |
| 6822 | Sec31b | SEC31B | 3.36E-02 | -3.24E-02 |
| 6823 | Msh5 | MSH5 | -5.06E-02 | -1.51E-02 |
| 6824 | Nup153 | NUP153 | -1.72E-02 | 9.95E-03 |
| 6825 | Ccdc171 | CCDC171 | -4.91E-02 | -7.10E-02 |
| 6826 | Eaf2 | EAF2 | -3.42E-02 | -7.44E-03 |
| 6827 | Mus81 | MUS81 | -2.82E-02 | 1.57E-02 |
| 6828 | Smarca1 | SMARCA1 | -8.52E-02 | -3.81E-02 |
| 6829 | Cd300lf | CD300LF | 1.01E-01 | 1.40E-01 |
| 6830 | Ccser2 | CCSER2 | -2.79E-02 | -2.50E-02 |
| 6831 | Crls1 | CRLS1 | -1.57E-02 | 6.51E-03 |
| 6832 | Zfp385c | LOC100507656 | 4.41E-02 | -2.61E-02 |
| 6833 | Spns1 | SPNS1 | -1.50E-02 | -6.06E-03 |
| 6834 | Rc3h2 | RC3H2 | 1.81E-02 | 4.91E-03 |

| | | | | |
|---|---|---|---|---|
| 6835 | Mta3 | MTA3 | 3.65E-02 | -3.54E-02 |
| 6836 | Foxd2 | FOXD2 | -5.66E-02 | -5.96E-02 |
| 6837 | Ell | ELL | 2.40E-02 | -1.41E-02 |
| 6838 | Sox17 | SOX17 | -3.82E-02 | -7.40E-03 |
| 6839 | Slc39a6 | SLC39A6 | 4.16E-02 | 1.92E-02 |
| 6840 | Mb | MB | 2.27E-02 | 7.99E-04 |
| 6841 | Ccdc28b | CCDC28B | 2.39E-02 | 1.59E-02 |
| 6842 | Tprkb | TPRKB | -2.84E-02 | -9.58E-03 |
| 6843 | Usp33 | USP33 | -1.60E-02 | -2.82E-03 |
| 6844 | Llph-ps2 | LLPH | 4.71E-02 | 1.58E-02 |
| 6845 | Erc1 | ERC1 | 2.05E-02 | 8.85E-03 |
| 6846 | Cacna2d3 | CACNA2D3 | -3.15E-02 | 1.47E-02 |
| 6847 | Ano7 | ANO7 | -2.26E-02 | 1.20E-02 |
| 6848 | Atp13a4 | ATP13A4 | -7.08E-02 | -7.56E-03 |
| 6849 | Ubxn2b | UBXN2B | -1.92E-02 | 3.21E-03 |
| 6850 | Mccc1 | MCCC1 | -2.35E-02 | -3.51E-02 |
| 6851 | Zfp105 | ZNF35 | -4.96E-02 | 3.03E-02 |
| 6852 | Fabp1 | FABP1 | -1.23E-01 | 7.94E-02 |
| 6853 | Trappc2 | TRAPPC2 | 2.46E-02 | 6.19E-03 |
| 6854 | Zbtb4 | ZBTB4 | -1.27E-02 | -2.73E-02 |
| 6855 | Irx4 | IRX4 | -4.32E-02 | -1.24E-02 |
| 6856 | Bcorl1 | BCORL1 | -2.89E-02 | 3.39E-03 |
| 6857 | Kdelr1 | KDELR1 | -1.17E-02 | -1.90E-03 |
| 6858 | Syvn1 | SYVN1 | -2.56E-02 | -1.46E-03 |
| 6859 | Bckdk | BCKDK | -1.48E-02 | -1.17E-02 |
| 6860 | Slc44a5 | SLC44A5 | 3.05E-02 | 7.43E-03 |
| 6861 | Zcchc18 | ZCCHC18 | -7.07E-02 | 5.46E-02 |
| 6862 | Eif3i | EIF3I | 1.41E-02 | -2.25E-03 |
| 6863 | Pnliprp1 | PNLIPRP1 | -4.83E-02 | -5.99E-02 |
| 6864 | Car8 | CA8 | 5.69E-02 | -8.65E-02 |
| 6865 | A130010J15Rik | C1orf74 | 4.16E-02 | 9.48E-03 |
| 6866 | Sepsecs | SEPSECS | -2.37E-02 | -1.24E-02 |
| 6867 | Zcchc14 | ZCCHC14 | -2.33E-02 | -9.23E-03 |
| 6868 | Cdk19 | CDK19 | 2.07E-02 | -1.79E-02 |
| 6869 | Stip1 | STIP1 | -2.68E-02 | 2.12E-03 |
| 6870 | Ttl | TTL | -1.46E-02 | -2.23E-02 |
| 6871 | Smo | SMO | -2.00E-02 | -3.85E-02 |
| 6872 | Evl | EVL | -2.64E-02 | 4.73E-02 |
| 6873 | Rbm48 | RBM48 | 2.78E-02 | -3.24E-02 |
| 6874 | Ankrd1 | ANKRD1 | 1.12E-01 | 2.08E-01 |
| 6875 | Gm11127 | HLA-F | -6.81E-02 | -5.00E-02 |
| 6876 | Stx6 | STX6 | 1.70E-02 | -1.48E-02 |
| 6877 | 1700088E04Rik | C22orf23 | 2.32E-02 | -2.46E-02 |
| 6878 | Lpcat4 | LPCAT4 | 4.45E-02 | 2.15E-02 |
| 6879 | Atm | ATM | -3.13E-02 | -2.25E-02 |
| 6880 | Gabrb1 | GABRB1 | 7.27E-02 | -7.01E-02 |
| 6881 | Mpdu1 | MPDU1 | 1.43E-02 | 4.71E-03 |
| 6882 | Akap8 | AKAP8 | -1.10E-02 | -3.39E-03 |
| 6883 | Mtrr | MTRR | -1.54E-02 | -4.21E-03 |
| 6884 | Bcar1 | BCAR1 | 1.92E-02 | 5.63E-04 |
| 6885 | Stab1 | STAB1 | 2.57E-02 | 3.61E-03 |
| 6886 | Trim32 | TRIM32 | -1.94E-02 | 4.19E-03 |
| 6887 | Pak3 | PAK3 | -6.64E-02 | 1.26E-01 |

| | | | | |
|---|---|---|---|---|
| 6888 | Chpt1 | CHPT1 | -2.01E-02 | -1.69E-02 |
| 6889 | Shank2 | SHANK2 | -3.54E-02 | -6.18E-02 |
| 6890 | Ssb | SSB | 1.14E-02 | 3.11E-03 |
| 6891 | Sec16a | SEC16A | 1.70E-02 | -1.66E-02 |
| 6892 | Camk4 | CAMK4 | -3.97E-02 | 1.58E-02 |
| 6893 | Wdr36 | WDR36 | -1.96E-02 | 1.19E-02 |
| 6894 | Sorcs1 | SORCS1 | 7.61E-02 | 1.14E-01 |
| 6895 | Atxn3 | ATXN3 | -1.47E-02 | -4.20E-05 |
| 6896 | Snx19 | SNX19 | 1.08E-02 | -1.28E-02 |
| 6897 | 1190007I07Rik | C12orf73 | -2.82E-02 | -2.92E-02 |
| 6898 | Abl1 | ABL1 | 1.71E-02 | -1.49E-02 |
| 6899 | Hnrnpk | HNRNPK | 1.37E-02 | -8.09E-03 |
| 6900 | Prdx3 | PRDX3 | 1.66E-02 | -1.52E-02 |
| 6901 | Gtf3c4 | GTF3C4 | -2.47E-02 | 1.84E-02 |
| 6902 | Snx12 | SNX12 | 1.53E-02 | -1.25E-03 |
| 6903 | Kcnk4 | KCNK4 | -2.50E-02 | 1.01E-02 |
| 6904 | Speg | SPEG | -3.71E-02 | 1.53E-02 |
| 6905 | Timm50 | TIMM50 | 1.71E-02 | -1.52E-02 |
| 6906 | Mpg | MPG | 2.74E-02 | -1.54E-02 |
| 6907 | Ceacam1 | CEACAM1 | -3.79E-02 | -6.97E-02 |
| 6908 | Efcab11 | EFCAB11 | 6.27E-02 | 8.41E-02 |
| 6909 | Tuba1b | TUBA1B | 3.41E-02 | 3.12E-02 |
| 6910 | Samd1 | SAMD1 | 2.31E-02 | 3.73E-02 |
| 6911 | Gpr33 | GPR33 | -2.28E-02 | 1.99E-03 |
| 6912 | Slk | SLK | -1.24E-02 | -1.23E-02 |
| 6913 | Ift57 | IFT57 | 1.36E-02 | -9.22E-03 |
| 6914 | Mtpap | MTPAP | -1.66E-02 | -2.81E-03 |
| 6915 | A930018P22Rik | C11orf91 | 2.11E-02 | -2.58E-03 |
| 6916 | Alox12b | ALOX12B | 1.19E-01 | -4.40E-02 |
| 6917 | Lbh | LBH | 4.60E-02 | -1.44E-02 |
| 6918 | Ocrl | OCRL | 1.88E-02 | -1.70E-02 |
| 6919 | Mvk | MVK | -4.16E-02 | -3.20E-02 |
| 6920 | Tinagl1 | TINAGL1 | 6.03E-02 | -2.13E-02 |
| 6921 | Oxnad1 | OXNAD1 | -1.48E-02 | -8.03E-03 |
| 6922 | Hoxc10 | HOXC10 | -2.56E-02 | 3.79E-02 |
| 6923 | Rcn3 | RCN3 | -5.53E-02 | 3.57E-02 |
| 6924 | Fdxacb1 | FDXACB1 | 2.44E-02 | -4.01E-03 |
| 6925 | Zfp646 | ZNF646 | -2.07E-02 | -5.36E-04 |
| 6926 | Ypel2 | YPEL2 | 5.33E-02 | 1.29E-01 |
| 6927 | Baiap2l2 | BAIAP2L2 | 1.81E-02 | -2.82E-04 |
| 6928 | Fam135a | FAM135A | 2.02E-02 | -9.71E-03 |
| 6929 | Uck1 | UCK1 | 2.02E-02 | -2.27E-03 |
| 6930 | Cenpn | CENPN | 6.69E-02 | 5.60E-02 |
| 6931 | Wwc2 | WWC2 | -1.68E-02 | -3.13E-02 |
| 6932 | Grin1 | GRIN1 | -3.78E-02 | -2.65E-03 |
| 6933 | Thap1 | THAP1 | -3.35E-02 | -4.45E-04 |
| 6934 | Zfp248 | ZNF248 | -4.98E-02 | -1.09E-02 |
| 6935 | Enkur | ENKUR | 6.06E-02 | 4.10E-02 |
| 6936 | Proz | PROZ | -8.89E-02 | -8.71E-02 |
| 6937 | Zfp335 | ZNF335 | -2.27E-02 | -2.04E-03 |
| 6938 | Loxl1 | LOXL1 | 7.31E-02 | 1.37E-02 |
| 6939 | Gria4 | GRIA4 | -7.35E-02 | 1.77E-02 |
| 6940 | Eef1g | EEF1G | -1.29E-02 | 8.20E-04 |

| | | | | |
|---|---|---|---|---|
| 6941 | Cckbr | CCKBR | 3.91E-02 | -1.31E-02 |
| 6942 | Nlgn1 | NLGN1 | -3.54E-02 | 1.85E-02 |
| 6943 | Sec22b | SEC22B | 1.28E-02 | -5.24E-03 |
| 6944 | Zc2hc1a | ZC2HC1A | -2.60E-02 | 9.48E-03 |
| 6945 | Rlim | RLIM | 1.75E-02 | 1.05E-02 |
| 6946 | Dcp1a | DCP1A | 1.66E-02 | -9.21E-03 |
| 6947 | Rfxap | RFXAP | 2.39E-02 | -4.44E-03 |
| 6948 | Macrod2 | MACROD2 | 3.66E-02 | -7.40E-03 |
| 6949 | Hsd17b4 | HSD17B4 | 2.50E-02 | -9.16E-03 |
| 6950 | Runx1 | RUNX1 | -8.23E-02 | 2.07E-01 |
| 6951 | Nmbr | NMBR | -5.03E-02 | -1.49E-02 |
| 6952 | Dld | DLD | 1.86E-02 | -2.10E-02 |
| 6953 | Arl6ip4 | ARL6IP4 | 1.55E-02 | -8.66E-04 |
| 6954 | Ror1 | ROR1 | -5.16E-02 | -1.32E-02 |
| 6955 | Cct6a | CCT6A | 1.53E-02 | -6.57E-03 |
| 6956 | Rap1gap2 | RAP1GAP2 | 2.03E-02 | -2.28E-02 |
| 6957 | Zfp202 | ZNF202 | -4.30E-02 | 2.22E-02 |
| 6958 | Cpeb4 | CPEB4 | 3.47E-02 | -2.51E-02 |
| 6959 | Rorc | RORC | 6.72E-02 | 8.03E-02 |
| 6960 | Casc3 | CASC3 | -1.23E-02 | 1.19E-03 |
| 6961 | Rnf122 | RNF122 | -4.06E-02 | 6.34E-02 |
| 6962 | Rsl1d1 | RSL1D1 | -1.24E-02 | 1.29E-02 |
| 6963 | Phf20l1 | PHF20L1 | 1.73E-02 | 2.48E-03 |
| 6964 | Tada1 | TADA1 | 2.03E-02 | 5.37E-03 |
| 6965 | Rpp25 | RPP25 | 6.56E-02 | 5.23E-02 |
| 6966 | Ggnbp2 | GGNBP2 | 1.43E-02 | -1.06E-03 |
| 6967 | Bcr | BCR | -1.45E-02 | -2.10E-02 |
| 6968 | Cenpi | CENPI | 1.11E-01 | 1.61E-01 |
| 6969 | Adcyap1r1 | ADCYAP1R1 | -8.44E-02 | -7.16E-02 |
| 6970 | Foxo4 | FOXO4 | -1.71E-02 | -2.41E-02 |
| 6971 | Eepd1 | EEPD1 | 5.30E-02 | -3.72E-02 |
| 6972 | Nono | NONO | -1.31E-02 | 6.31E-03 |
| 6973 | Cdc25c | CDC25C | 1.07E-01 | 1.14E-01 |
| 6974 | Pdhb | PDHB | 1.55E-02 | -1.21E-02 |
| 6975 | Slc35c1 | SLC35C1 | -1.72E-02 | 3.25E-02 |
| 6976 | Tm7sf2 | TM7SF2 | -2.48E-02 | -9.03E-03 |
| 6977 | Nipal4 | NIPAL4 | 1.12E-01 | 5.24E-02 |
| 6978 | Trappc12 | TRAPPC12 | -1.60E-02 | -1.73E-02 |
| 6979 | Mrps26 | MRPS26 | 1.63E-02 | -2.75E-03 |
| 6980 | Ints2 | INTS2 | 3.02E-02 | -2.12E-02 |
| 6981 | Abhd10 | ABHD10 | 2.05E-02 | -6.08E-03 |
| 6982 | Ccdc3 | CCDC3 | 1.80E-01 | -4.40E-02 |
| 6983 | Zmat2 | ZMAT2 | 1.18E-02 | -3.08E-04 |
| 6984 | Tubb4a | TUBB4A | -5.19E-02 | 5.43E-02 |
| 6985 | Glipr2 | GLIPR2 | -7.41E-02 | 1.98E-01 |
| 6986 | Skiv2l | SKIV2L | 1.36E-02 | -1.26E-02 |
| 6987 | Cyp4a31 | CYP4A11 | 3.62E-02 | 6.60E-03 |
| 6988 | Smim12 | SMIM12 | 1.70E-02 | -1.40E-02 |
| 6989 | Fam114a2 | FAM114A2 | -1.28E-02 | -1.17E-02 |
| 6990 | Snx9 | SNX9 | 1.61E-02 | -2.45E-02 |
| 6991 | Tfap2a | TFAP2A | 5.29E-02 | 5.45E-02 |
| 6992 | Wipi1 | WIPI1 | 2.34E-02 | -6.02E-02 |
| 6993 | Slc24a4 | SLC24A4 | 2.58E-02 | -2.48E-02 |

| | | | | |
|---|---|---|---|---|
| 6994 | Zdhhc14 | ZDHHC14 | 3.22E-02 | 2.11E-02 |
| 6995 | Cd276 | CD276 | 6.88E-02 | 1.15E-01 |
| 6996 | Mettl2 | METTL2A | -2.14E-02 | 1.03E-02 |
| 6997 | Rad52 | RAD52 | -2.51E-02 | -1.85E-03 |
| 6998 | Ppp1r15a | PPP1R15A | 6.33E-02 | -2.44E-02 |
| 6999 | Pip4k2c | PIP4K2C | 1.15E-02 | -8.37E-03 |
| 7000 | Ddx18 | DDX18 | -1.54E-02 | 1.78E-03 |
| 7001 | Phf1 | PHF1 | -1.97E-02 | -8.67E-03 |
| 7002 | Sart3 | SART3 | -1.18E-02 | -6.30E-03 |
| 7003 | Celf5 | CELF5 | -3.38E-02 | -3.43E-03 |
| 7004 | Phtf1 | PHTF1 | -1.68E-02 | 1.19E-02 |
| 7005 | Col4a5 | COL4A5 | 2.82E-02 | -1.47E-02 |
| 7006 | Aoc2 | AOC2 | 4.33E-02 | -1.04E-03 |
| 7007 | Osgep | OSGEP | -1.49E-02 | -1.32E-02 |
| 7008 | Fgfr4 | FGFR4 | -2.25E-02 | -7.95E-03 |
| 7009 | Ddr1 | DDR1 | 1.46E-02 | 1.77E-02 |
| 7010 | Mrpl44 | MRPL44 | -1.73E-02 | -9.12E-03 |
| 7011 | Atg2a | ATG2A | 1.88E-02 | -1.96E-02 |
| 7012 | Syce2 | SYCE2 | 7.25E-02 | 8.60E-02 |
| 7013 | Ins2 | INS | -2.68E-02 | -1.30E-02 |
| 7014 | Sfr1 | SFR1 | 1.49E-02 | 7.78E-03 |
| 7015 | Hpn | HPN | 1.99E-02 | -1.41E-02 |
| 7016 | Mpc1 | MPC1 | 1.43E-02 | -2.36E-02 |
| 7017 | Tmem204 | TMEM204 | -3.71E-02 | -4.00E-02 |
| 7018 | Golgb1 | GOLGB1 | 1.14E-02 | -1.96E-02 |
| 7019 | Atf2 | ATF2 | -1.64E-02 | -1.02E-02 |
| 7020 | Col11a1 | COL11A1 | -1.47E-01 | 5.23E-02 |
| 7021 | Rpusd2 | RPUSD2 | -4.56E-02 | -8.95E-03 |
| 7022 | Frmd4a | FRMD4A | -4.02E-02 | -5.73E-02 |
| 7023 | Nt5c2 | NT5C2 | 1.88E-02 | -2.53E-02 |
| 7024 | Sufu | SUFU | -2.32E-02 | -2.09E-02 |
| 7025 | Pde12 | PDE12 | 2.18E-02 | 1.96E-02 |
| 7026 | Sh3d21 | SH3D21 | -2.95E-02 | 2.12E-03 |
| 7027 | Ammecr1 | AMMECR1 | -3.96E-02 | 1.10E-01 |
| 7028 | D16Ertd472e | C21orf91 | -5.52E-02 | 3.34E-02 |
| 7029 | Aste1 | ASTE1 | 2.42E-02 | -5.05E-03 |
| 7030 | Golga2 | GOLGA2 | 1.23E-02 | -1.82E-02 |
| 7031 | Tmem19 | TMEM19 | -1.74E-02 | -9.41E-03 |
| 7032 | Tmtc2 | TMTC2 | -9.99E-02 | -1.57E-02 |
| 7033 | Klk8 | KLK8 | -6.36E-02 | 8.39E-03 |
| 7034 | Brwd3 | BRWD3 | 2.86E-02 | -6.63E-03 |
| 7035 | Ccdc122 | CCDC122 | 5.74E-02 | -2.11E-02 |
| 7036 | Trim69 | TRIM69 | -3.26E-02 | 3.24E-02 |
| 7037 | Zbtb24 | ZBTB24 | -1.82E-02 | -2.78E-02 |
| 7038 | Sin3a | SIN3A | -1.16E-02 | 2.68E-03 |
| 7039 | Stx4a | STX4 | -1.62E-02 | -1.98E-02 |
| 7040 | Dok1 | DOK1 | 4.77E-02 | 3.75E-02 |
| 7041 | Bcat1 | BCAT1 | 2.63E-01 | 3.58E-03 |
| 7042 | Cd83 | CD83 | -6.52E-02 | 2.36E-02 |
| 7043 | Epha1 | EPHA1 | -1.76E-02 | -2.47E-02 |
| 7044 | Klf8 | KLF8 | -5.69E-02 | 3.07E-02 |
| 7045 | Coa4 | COA4 | -4.04E-02 | 2.08E-02 |
| 7046 | Slc30a5 | SLC30A5 | 1.30E-02 | -2.68E-04 |

| | | | | |
|---|---|---|---|---|
| 7047 | Vgll1 | VGLL1 | 3.00E-02 | -5.13E-03 |
| 7048 | Phyhip | PHYHIP | -6.33E-02 | 1.85E-02 |
| 7049 | Dennd2c | DENND2C | -6.05E-02 | -1.51E-02 |
| 7050 | Prkaa2 | PRKAA2 | 2.44E-02 | -2.06E-02 |
| 7051 | Mfn2 | MFN2 | 1.42E-02 | -2.12E-02 |
| 7052 | Galnt11 | GALNT11 | -2.62E-02 | -2.92E-02 |
| 7053 | Mep1a | MEP1A | -2.07E-02 | -5.13E-02 |
| 7054 | Sycn | SYCN | 2.70E-01 | -2.10E-01 |
| 7055 | Tlr7 | TLR7 | -6.37E-02 | 8.50E-03 |
| 7056 | Ahsa1 | AHSA1 | -2.73E-02 | 2.43E-03 |
| 7057 | Ppp2r2b | PPP2R2B | -4.32E-02 | -2.36E-03 |
| 7058 | Retsat | RETSAT | 3.25E-02 | -3.84E-02 |
| 7059 | Klc1 | KLC1 | 1.09E-02 | -3.22E-03 |
| 7060 | Serbp1 | SERBP1 | -9.59E-03 | 4.29E-03 |
| 7061 | Micu2 | MICU2 | 1.43E-02 | -5.51E-03 |
| 7062 | Cox18 | COX18 | 2.18E-02 | 9.47E-03 |
| 7063 | Usp34 | USP34 | 1.23E-02 | -1.19E-02 |
| 7064 | Hsh2d | HSH2D | -2.15E-02 | 1.75E-03 |
| 7065 | Suds3 | SUDS3 | 1.20E-02 | -1.82E-02 |
| 7066 | Psph | PSPH | 3.24E-02 | -3.97E-02 |
| 7067 | Rab9b | RAB9B | 3.21E-02 | 3.60E-03 |
| 7068 | Nifk | NIFK | 1.63E-02 | 8.19E-03 |
| 7069 | Nol8 | NOL8 | -2.12E-02 | 1.49E-02 |
| 7070 | Vps4b | VPS4B | 1.24E-02 | -1.52E-02 |
| 7071 | Daam2 | DAAM2 | 3.58E-02 | -5.71E-02 |
| 7072 | Fzd5 | FZD5 | -2.34E-02 | -3.93E-03 |
| 7073 | Tie1 | TIE1 | -1.98E-02 | -1.51E-02 |
| 7074 | Atp5j | ATP5J | 1.43E-02 | -5.87E-03 |
| 7075 | Tgs1 | TGS1 | -2.55E-02 | 3.58E-03 |
| 7076 | Ctss | CTSS | -5.21E-02 | 1.57E-01 |
| 7077 | Gsdma | GSDMA | 1.63E-02 | -1.12E-03 |
| 7078 | Zfp518b | ZNF518B | -2.97E-02 | 3.35E-02 |
| 7079 | Ddx52 | DDX52 | 1.41E-02 | 3.04E-03 |
| 7080 | Als2cl | ALS2CL | 1.71E-02 | 5.55E-03 |
| 7081 | Aadac | AADAC | -3.23E-02 | -1.27E-02 |
| 7082 | Mcoln1 | MCOLN1 | 1.79E-02 | -6.86E-03 |
| 7083 | Zfyve1 | ZFYVE1 | 1.96E-02 | -1.82E-02 |
| 7084 | Efna4 | EFNA4 | 7.31E-02 | 2.22E-02 |
| 7085 | Abca5 | ABCA5 | 2.17E-02 | -1.99E-02 |
| 7086 | Apoh | APOH | 9.90E-02 | 1.46E-01 |
| 7087 | Vangl2 | VANGL2 | 5.25E-02 | -2.90E-02 |
| 7088 | Rgs10 | RGS10 | -5.14E-02 | 8.06E-02 |
| 7089 | Cops7a | COPS7A | 1.39E-02 | -1.05E-02 |
| 7090 | Efna5 | EFNA5 | 6.05E-02 | -3.51E-02 |
| 7091 | Cacnb1 | CACNB1 | -6.48E-02 | 2.62E-02 |
| 7092 | Hoxa7 | HOXA7 | 1.08E-02 | -2.51E-02 |
| 7093 | Rho | RHO | 5.16E-02 | 3.41E-03 |
| 7094 | Tsen54 | TSEN54 | -2.36E-02 | -1.72E-03 |
| 7095 | Pde1b | PDE1B | -6.97E-02 | 2.57E-02 |
| 7096 | Dgcr2 | DGCR2 | -1.18E-02 | -6.52E-03 |
| 7097 | Cntnap5a | CNTNAP5 | 1.89E-02 | -5.37E-03 |
| 7098 | Clic1 | CLIC1 | 2.08E-02 | 3.02E-02 |
| 7099 | Tas1r1 | TAS1R1 | 3.83E-02 | 3.17E-03 |

| | | | | |
|---|---|---|---|---|
| 7100 | Hinfp | HINFP | -2.49E-02 | -9.68E-03 |
| 7101 | Gid8 | GID8 | 1.18E-02 | -4.62E-03 |
| 7102 | Crb2 | CRB2 | -4.95E-02 | -2.87E-02 |
| 7103 | Mapk8ip2 | MAPK8IP2 | -9.58E-02 | -7.16E-02 |
| 7104 | Il15ra | IL15RA | -2.34E-02 | -1.09E-02 |
| 7105 | Dlc1 | DLC1 | -3.21E-02 | -1.48E-02 |
| 7106 | Psmc2 | PSMC2 | 1.50E-02 | -7.73E-03 |
| 7107 | Ngrn | NGRN | 1.72E-02 | -1.19E-02 |
| 7108 | Tbc1d22a | TBC1D22A | 1.30E-02 | -1.97E-02 |
| 7109 | Prdx2 | PRDX2 | 1.32E-02 | -5.72E-03 |
| 7110 | Klhl10 | KLHL10 | -2.69E-02 | -6.24E-03 |
| 7111 | Rbbp8nl | RBBP8NL | -6.88E-02 | -1.52E-02 |
| 7112 | Mpnd | MPND | -2.48E-02 | 3.07E-03 |
| 7113 | Incenp | INCENP | 6.01E-02 | 1.00E-01 |
| 7114 | Srebf1 | SREBF1 | -2.94E-02 | -8.34E-03 |
| 7115 | Slc44a3 | SLC44A3 | 2.93E-02 | 5.36E-03 |
| 7116 | Acsm2 | ACSM2A | 2.85E-02 | 3.46E-03 |
| 7117 | Tnfaip3 | TNFAIP3 | 7.73E-02 | 5.65E-02 |
| 7118 | Klk14 | KLK14 | -2.18E-02 | 4.75E-03 |
| 7119 | Zfp217 | ZNF217 | 2.67E-02 | 1.01E-02 |
| 7120 | Ngdn | NGDN | -1.48E-02 | -1.03E-03 |
| 7121 | Il11 | IL11 | 6.62E-02 | 6.12E-02 |
| 7122 | Il12rb2 | IL12RB2 | -3.58E-02 | -5.40E-04 |
| 7123 | Pank3 | PANK3 | 2.12E-02 | -6.87E-03 |
| 7124 | Mnt | MNT | 4.85E-02 | -3.02E-03 |
| 7125 | Tmem238 | TMEM238 | 3.13E-02 | 6.05E-03 |
| 7126 | Spin4 | SPIN4 | -5.27E-02 | 6.36E-03 |
| 7127 | Ust | UST | -4.83E-02 | 4.18E-02 |
| 7128 | Fgfr1op2 | FGFR1OP2 | -1.27E-02 | -1.51E-03 |
| 7129 | Pomp | POMP | 1.44E-02 | -4.74E-03 |
| 7130 | Ctla4 | CTLA4 | -1.54E-02 | 3.46E-03 |
| 7131 | Mad2l1 | MAD2L1 | 7.07E-02 | 1.11E-01 |
| 7132 | Txnl4a | TXNL4A | -2.67E-02 | 1.25E-02 |
| 7133 | Sdf4 | SDF4 | 9.94E-03 | -4.98E-03 |
| 7134 | Disp2 | DISP2 | 4.71E-02 | 5.36E-02 |
| 7135 | Arhgef6 | ARHGEF6 | -3.29E-02 | 6.66E-03 |
| 7136 | Fam83g | FAM83G | -5.38E-02 | -4.65E-02 |
| 7137 | Cox11 | COX11 | 2.32E-02 | -8.72E-03 |
| 7138 | Gipc2 | GIPC2 | 1.60E-02 | -1.54E-02 |
| 7139 | Dhh | DHH | 5.94E-02 | 2.06E-02 |
| 7140 | Lrit1 | LRIT1 | 4.78E-02 | -6.36E-02 |
| 7141 | Ada | ADA | 5.46E-02 | 2.95E-02 |
| 7142 | Tom1l2 | TOM1L2 | 1.45E-02 | -3.57E-02 |
| 7143 | Kcnc4 | KCNC4 | 5.52E-02 | -9.91E-03 |
| 7144 | Jarid2 | JARID2 | -2.64E-02 | -4.01E-04 |
| 7145 | Mctp2 | MCTP2 | 5.53E-02 | -3.67E-02 |
| 7146 | Mettl15 | METTL15 | -2.38E-02 | 2.71E-03 |
| 7147 | 4932438A13Rik | KIAA1109 | 1.54E-02 | -1.83E-02 |
| 7148 | Fubp3 | FUBP3 | 1.71E-02 | -1.54E-02 |
| 7149 | Exosc10 | EXOSC10 | 1.36E-02 | -1.01E-02 |
| 7150 | Gnptg | GNPTG | 1.61E-02 | -1.40E-02 |
| 7151 | Ndst1 | NDST1 | 1.87E-02 | -4.06E-02 |
| 7152 | Cpt2 | CPT2 | 3.01E-02 | -3.82E-03 |

| | | | | |
|---|---|---|---|---|
| 7153 | Ptgr1 | PTGR1 | 4.35E-02 | 3.32E-02 |
| 7154 | Tmbim4 | TMBIM4 | 1.86E-02 | -1.13E-02 |
| 7155 | Kif3b | KIF3B | 1.39E-02 | -5.94E-03 |
| 7156 | Kctd7 | KCTD7 | -5.35E-02 | 1.52E-02 |
| 7157 | Malt1 | MALT1 | 2.63E-02 | -1.74E-02 |
| 7158 | Sfn | SFN | 9.15E-02 | 9.58E-02 |
| 7159 | Fam43b | FAM43B | 2.20E-02 | -9.70E-03 |
| 7160 | Nelfe | NELFE | 1.81E-02 | -1.84E-02 |
| 7161 | Hsf5 | HSF5 | 3.38E-02 | -1.52E-02 |
| 7162 | Fchsd1 | FCHSD1 | 3.35E-02 | -2.72E-02 |
| 7163 | Pthlh | PTHLH | -3.10E-02 | 3.52E-02 |
| 7164 | Stk19 | STK19 | 2.00E-02 | 4.51E-03 |
| 7165 | Hlf | HLF | 3.48E-02 | -1.56E-02 |
| 7166 | Camk2b | CAMK2B | -3.54E-02 | 2.10E-02 |
| 7167 | Gpatch11 | GPATCH11 | -1.80E-02 | -1.28E-03 |
| 7168 | Hnrnph3 | HNRNPH3 | -4.20E-02 | -9.55E-03 |
| 7169 | Smyd2 | SMYD2 | -1.85E-02 | -1.40E-02 |
| 7170 | Rpl15 | RPL15 | -1.40E-02 | 1.03E-02 |
| 7171 | Fbxo4 | FBXO4 | 2.83E-02 | 2.64E-02 |
| 7172 | Ipo4 | IPO4 | 1.59E-02 | 3.12E-03 |
| 7173 | Sowahc | SOWAHC | 2.95E-02 | 2.62E-02 |
| 7174 | Mapk7 | MAPK7 | 2.39E-02 | 2.45E-03 |
| 7175 | Rrp36 | RRP36 | -1.37E-02 | -4.98E-03 |
| 7176 | Kremen2 | KREMEN2 | -2.26E-02 | 1.02E-02 |
| 7177 | Tbpl1 | TBPL1 | -2.64E-02 | 9.62E-03 |
| 7178 | Fam78b | FAM78B | -9.52E-02 | 1.18E-02 |
| 7179 | Fam163a | FAM163A | 6.98E-02 | 3.70E-02 |
| 7180 | Smim7 | SMIM7 | 1.03E-02 | -1.25E-02 |
| 7181 | Abhd17c | ABHD17C | -1.95E-02 | 7.42E-03 |
| 7182 | Ccne1 | CCNE1 | 7.07E-02 | 1.31E-01 |
| 7183 | Mrps27 | MRPS27 | 1.72E-02 | -1.52E-02 |
| 7184 | Tgfb1i1 | TGFB1I1 | 4.23E-02 | 5.30E-02 |
| 7185 | Vps26b | VPS26B | 1.11E-02 | -1.30E-02 |
| 7186 | Slc39a10 | SLC39A10 | 2.53E-02 | -1.58E-02 |
| 7187 | Elk3 | ELK3 | -2.78E-02 | 2.13E-02 |
| 7188 | Ren1 | REN | -8.21E-02 | -2.95E-01 |
| 7189 | Slfn9 | SLFN13 | -9.86E-02 | 2.15E-01 |
| 7190 | Ctsk | CTSK | 7.22E-02 | 9.63E-02 |
| 7191 | Ugt2a3 | UGT2A3 | -5.08E-02 | 7.91E-02 |
| 7192 | Pgr | PGR | -7.34E-02 | -7.35E-02 |
| 7193 | Myl12a | MYL12B | 1.29E-02 | 5.40E-03 |
| 7194 | Bag6 | BAG6 | 1.21E-02 | -1.58E-02 |
| 7195 | Gpr155 | GPR155 | -3.91E-02 | -5.90E-02 |
| 7196 | Sppl3 | SPPL3 | -1.26E-02 | 2.06E-03 |
| 7197 | Bloc1s2 | BLOC1S2 | -1.74E-02 | 2.14E-02 |
| 7198 | Me2 | ME2 | 3.62E-02 | 3.07E-02 |
| 7199 | Trub1 | TRUB1 | 1.81E-02 | -1.79E-02 |
| 7200 | Arl13b | ARL13B | -3.04E-02 | 8.49E-03 |
| 7201 | 1700007K13Rik | C9orf116 | 4.23E-02 | -2.40E-02 |
| 7202 | Dnah10 | DNAH10 | 6.22E-02 | -1.81E-02 |
| 7203 | Rpsa | RPSA | -1.62E-02 | 1.46E-02 |
| 7204 | Metap1d | METAP1D | -1.47E-02 | -1.51E-02 |
| 7205 | Wsb2 | WSB2 | 1.40E-02 | -1.28E-02 |

| | | | | |
|---|---|---|---|---|
| 7206 | Slc37a1 | SLC37A1 | 5.39E-02 | -1.65E-02 |
| 7207 | Pik3r1 | PIK3R1 | 1.50E-02 | 4.34E-03 |
| 7208 | Ebna1bp2 | EBNA1BP2 | -1.37E-02 | 4.64E-03 |
| 7209 | Zmynd11 | ZMYND11 | 8.87E-03 | -1.25E-02 |
| 7210 | Gstm5 | GSTM3 | 2.53E-02 | 9.49E-04 |
| 7211 | Edem1 | EDEM1 | -1.88E-02 | -8.65E-03 |
| 7212 | Cd68 | CD68 | -6.23E-02 | 1.71E-01 |
| 7213 | Col11a2 | COL11A2 | 4.80E-02 | 3.55E-02 |
| 7214 | Tnfrsf25 | TNFRSF25 | -1.08E-01 | 7.21E-02 |
| 7215 | Tatdn3 | TATDN3 | -2.37E-02 | -4.97E-02 |
| 7216 | Tubgcp2 | TUBGCP2 | 1.97E-02 | -1.84E-02 |
| 7217 | Tinf2 | TINF2 | 1.68E-02 | 7.96E-03 |
| 7218 | Cdnf | CDNF | -4.50E-02 | -3.59E-02 |
| 7219 | Rprd2 | RPRD2 | -1.33E-02 | -9.89E-03 |
| 7220 | Atg3 | ATG3 | 1.62E-02 | 6.46E-03 |
| 7221 | Klk1b9 | KLK3 | 4.83E-02 | 1.33E-01 |
| 7222 | Ppih | PPIH | 1.82E-02 | 1.89E-02 |
| 7223 | Aamp | AAMP | 1.55E-02 | -1.39E-02 |
| 7224 | Gstk1 | GSTK1 | -2.47E-02 | -2.09E-02 |
| 7225 | Cntn3 | CNTN3 | -4.11E-02 | 3.00E-03 |
| 7226 | Mul1 | MUL1 | 1.32E-02 | -1.51E-02 |
| 7227 | Pkp3 | PKP3 | 2.20E-02 | -6.58E-03 |
| 7228 | Atg16l2 | ATG16L2 | -5.36E-02 | 2.57E-02 |
| 7229 | Clip3 | CLIP3 | 6.34E-02 | 5.47E-02 |
| 7230 | Coro6 | CORO6 | -5.16E-02 | -7.57E-04 |
| 7231 | Batf | BATF | -4.53E-02 | 5.01E-02 |
| 7232 | Rps6ka6 | RPS6KA6 | -5.17E-02 | -2.27E-02 |
| 7233 | Csnk2a2 | CSNK2A2 | 1.20E-02 | -1.60E-03 |
| 7234 | Art5 | ART5 | 3.71E-02 | -3.45E-03 |
| 7235 | Rhobtb3 | RHOBTB3 | 3.31E-02 | 1.71E-02 |
| 7236 | Ggt5 | GGT5 | 4.96E-02 | 5.36E-03 |
| 7237 | 1600012H06Rik | C6orf120 | 1.13E-02 | 9.41E-04 |
| 7238 | Cd5 | CD5 | -5.21E-02 | 4.59E-02 |
| 7239 | Eps15 | EPS15 | 1.09E-02 | -1.09E-02 |
| 7240 | Pnldc1 | PNLDC1 | 4.49E-02 | -1.55E-02 |
| 7241 | Fgfr2 | FGFR2 | 4.67E-02 | 1.13E-02 |
| 7242 | Pip4k2b | PIP4K2B | -1.99E-02 | -1.17E-02 |
| 7243 | Zfp558 | ZNF558 | -4.94E-02 | -3.21E-03 |
| 7244 | Lrrc69 | LRRC69 | -2.85E-02 | 1.21E-02 |
| 7245 | Acvr1 | ACVR1 | -1.88E-02 | 2.30E-02 |
| 7246 | Mboat1 | MBOAT1 | -3.26E-02 | 3.17E-02 |
| 7247 | Wfikkn1 | WFIKKN1 | 6.39E-02 | -1.30E-03 |
| 7248 | Plaur | PLAUR | 1.11E-01 | 9.42E-02 |
| 7249 | Nr4a3 | NR4A3 | 1.33E-01 | 6.46E-02 |
| 7250 | Rwdd2a | RWDD2A | -3.94E-02 | -3.19E-02 |
| 7251 | C8g | C8G | -3.89E-02 | 9.72E-03 |
| 7252 | Mttp | MTTP | -5.08E-02 | 2.87E-02 |
| 7253 | Alad | ALAD | -1.92E-02 | -1.35E-02 |
| 7254 | B4galnt3 | B4GALNT3 | -2.87E-02 | -3.81E-02 |
| 7255 | Fars2 | FARS2 | 1.56E-02 | -2.91E-02 |
| 7256 | Senp7 | SENP7 | -1.93E-02 | -9.36E-03 |
| 7257 | Tal2 | TAL2 | 1.28E-01 | -7.51E-02 |
| 7258 | Trf | TF | -5.36E-02 | 6.58E-02 |

| | | | | |
|---|---|---|---|---|
| 7259 | Dpep2 | DPEP2 | 2.98E-02 | 3.41E-02 |
| 7260 | Creb3l4 | CREB3L4 | -8.31E-02 | -4.45E-02 |
| 7261 | Galnt13 | GALNT13 | 2.07E-02 | 6.99E-03 |
| 7262 | Rnf121 | RNF121 | 1.86E-02 | 4.65E-03 |
| 7263 | Gpr153 | GPR153 | 5.44E-02 | 5.03E-02 |
| 7264 | Armc8 | ARMC8 | 1.03E-02 | -1.39E-02 |
| 7265 | Fam222a | FAM222A | -3.71E-02 | 5.10E-02 |
| 7266 | 1700001O22Rik | C9orf50 | -6.54E-02 | -1.15E-02 |
| 7267 | Dpys | DPYS | 8.22E-02 | -1.02E-01 |
| 7268 | Plcb4 | PLCB4 | 4.91E-02 | -3.32E-03 |
| 7269 | Tpd52 | TPD52 | 1.56E-02 | -6.19E-03 |
| 7270 | Dapp1 | DAPP1 | -7.21E-02 | 1.03E-01 |
| 7271 | Elf4 | ELF4 | -4.10E-02 | 7.65E-02 |
| 7272 | Top1 | TOP1 | -1.96E-02 | 2.71E-02 |
| 7273 | Nt5dc1 | NT5DC1 | 1.88E-02 | -3.05E-02 |
| 7274 | Fstl3 | FSTL3 | 7.27E-02 | -2.64E-02 |
| 7275 | Tctn1 | TCTN1 | 1.81E-02 | -1.47E-02 |
| 7276 | Zeb1 | ZEB1 | -3.81E-02 | 1.05E-02 |
| 7277 | Armcx2 | ARMCX2 | -2.90E-02 | 2.00E-02 |
| 7278 | Ccr4 | CCR4 | -1.97E-02 | 1.41E-02 |
| 7279 | Ilf3 | ILF3 | -1.16E-02 | 3.59E-03 |
| 7280 | Adprm | ADPRM | 2.32E-02 | -2.28E-03 |
| 7281 | Tspan15 | TSPAN15 | 3.21E-02 | 2.30E-02 |
| 7282 | Fbxl20 | FBXL20 | 1.55E-02 | -2.58E-02 |
| 7283 | Ralbp1 | RALBP1 | -1.15E-02 | -7.92E-03 |
| 7284 | Ttc27 | TTC27 | -2.13E-02 | -7.07E-03 |
| 7285 | Rps6ka2 | RPS6KA2 | -4.34E-02 | 5.62E-03 |
| 7286 | Wnt5a | WNT5A | -7.19E-02 | -1.60E-02 |
| 7287 | Glcci1 | GLCCI1 | -5.29E-02 | -9.09E-03 |
| 7288 | Lhfpl1 | LHFPL1 | -5.11E-02 | 2.12E-03 |
| 7289 | Stpg2 | STPG2 | -2.57E-02 | -1.61E-02 |
| 7290 | Klhl33 | KLHL33 | -4.47E-02 | 2.67E-02 |
| 7291 | Fez1 | FEZ1 | 4.17E-02 | 2.55E-03 |
| 7292 | Csad | CSAD | -2.79E-02 | -5.18E-02 |
| 7293 | Lhx1 | LHX1 | -3.46E-02 | -1.80E-02 |
| 7294 | Rrp15 | RRP15 | -1.85E-02 | -1.27E-03 |
| 7295 | Fam221a | FAM221A | -6.50E-02 | 1.89E-02 |
| 7296 | Csnk2a1 | CSNK2A1 | 1.19E-02 | -8.59E-03 |
| 7297 | Mrps30 | MRPS30 | 1.50E-02 | -1.25E-02 |
| 7298 | Mcat | MCAT | -1.73E-02 | -2.33E-02 |
| 7299 | Slc7a1 | SLC7A1 | 6.39E-02 | 1.65E-02 |
| 7300 | Amfr | AMFR | -1.28E-02 | -1.90E-02 |
| 7301 | Klrb1b | KLRB1 | 2.41E-02 | 3.28E-02 |
| 7302 | Pdzd2 | PDZD2 | 3.54E-02 | -4.88E-02 |
| 7303 | Nme2 | NME2 | 4.99E-02 | -1.24E-02 |
| 7304 | Myoc | MYOC | -3.10E-02 | 2.01E-02 |
| 7305 | Jph4 | JPH4 | 2.18E-02 | -1.02E-02 |
| 7306 | Rpl7 | RPL7 | -1.22E-02 | 1.82E-02 |
| 7307 | Cercam | CERCAM | 6.73E-02 | 5.37E-02 |
| 7308 | Cdc37 | CDC37 | 1.27E-02 | -9.19E-03 |
| 7309 | Nutf2 | NUTF2 | 1.99E-02 | -1.73E-02 |
| 7310 | Krr1 | KRR1 | -1.55E-02 | 2.62E-03 |
| 7311 | Syt13 | SYT13 | -8.05E-02 | -1.71E-02 |

| | | | | |
|---|---|---|---|---|
| 7312 | D8Ertd738e | C19orf53 | 1.35E-02 | -6.01E-04 |
| 7313 | Homer3 | HOMER3 | 2.50E-02 | 2.32E-02 |
| 7314 | 6430550D23Rik | C20orf173 | -3.45E-02 | -3.90E-02 |
| 7315 | Cp | CP | -4.51E-02 | -2.12E-02 |
| 7316 | Kpna3 | KPNA3 | 1.46E-02 | 1.65E-02 |
| 7317 | Gigyf2 | GIGYF2 | -2.03E-02 | 2.28E-03 |
| 7318 | Sdccag8 | SDCCAG8 | 1.88E-02 | -4.20E-03 |
| 7319 | Dynlt1a | DYNLT1 | 4.81E-02 | 9.67E-03 |
| 7320 | Ltb4r1 | LTB4R | -3.23E-02 | 7.62E-02 |
| 7321 | Nap1l2 | NAP1L2 | -1.55E-02 | 1.21E-02 |
| 7322 | Nccrp1 | NCCRP1 | 1.72E-01 | -1.64E-01 |
| 7323 | Sgpp2 | SGPP2 | -3.90E-02 | -1.62E-02 |
| 7324 | Muc2 | MUC2 | 3.43E-02 | -1.87E-03 |
| 7325 | Tyw1 | TYW1 | 1.92E-02 | -2.46E-03 |
| 7326 | Shprh | SHPRH | -1.93E-02 | 5.68E-03 |
| 7327 | Dip2a | DIP2A | 1.50E-02 | -2.09E-02 |
| 7328 | Mecom | MECOM | 2.86E-02 | 1.33E-02 |
| 7329 | Nol10 | NOL10 | 1.67E-02 | -3.24E-04 |
| 7330 | Trhr | TRHR | 5.17E-02 | -1.98E-02 |
| 7331 | Treml2 | TREML2 | -3.68E-02 | 7.46E-02 |
| 7332 | Tmem120b | TMEM120B | 3.36E-02 | -6.71E-03 |
| 7333 | Rasd2 | RASD2 | 4.50E-02 | 9.05E-03 |
| 7334 | 1700029J07Rik | C4orf47 | -2.78E-02 | -9.38E-03 |
| 7335 | Zfp408 | ZNF408 | 1.62E-02 | -4.10E-03 |
| 7336 | Gpalpp1 | GPALPP1 | 1.66E-02 | -1.31E-02 |
| 7337 | Mob3a | MOB3A | 2.76E-02 | 2.37E-02 |
| 7338 | Sec11a | SEC11A | -1.33E-02 | 2.45E-02 |
| 7339 | Gp2 | GP2 | -6.83E-02 | -6.38E-02 |
| 7340 | E2f7 | E2F7 | 8.99E-02 | 1.53E-01 |
| 7341 | Rps6kl1 | RPS6KL1 | 6.77E-02 | 2.07E-03 |
| 7342 | Ptcd2 | PTCD2 | -1.74E-02 | -6.74E-03 |
| 7343 | Zfp644 | ZNF644 | -2.32E-02 | 1.66E-03 |
| 7344 | Arhgap27 | ARHGAP27 | 2.22E-02 | -3.48E-03 |
| 7345 | Oplah | OPLAH | 3.07E-02 | -1.38E-02 |
| 7346 | Taf13 | TAF13 | 1.94E-02 | 1.54E-02 |
| 7347 | Socs3 | SOCS3 | 1.20E-01 | 2.73E-01 |
| 7348 | Il7 | IL7 | -6.33E-02 | -4.09E-02 |
| 7349 | Dgki | DGKI | 2.53E-02 | -3.13E-02 |
| 7350 | Pnpt1 | PNPT1 | -1.69E-02 | -5.08E-03 |
| 7351 | S100a8 | S100A8 | 1.64E-01 | 1.48E-01 |
| 7352 | Paqr4 | PAQR4 | -6.98E-02 | 1.12E-01 |
| 7353 | Tshz1 | TSHZ1 | 1.80E-02 | 1.46E-06 |
| 7354 | Dmxl2 | DMXL2 | 2.21E-02 | -2.55E-02 |
| 7355 | Srprb | SRPRB | 2.27E-02 | 1.29E-02 |
| 7356 | Chmp7 | CHMP7 | 1.08E-02 | -1.01E-02 |
| 7357 | Prkrip1 | PRKRIP1 | 2.40E-02 | 1.82E-03 |
| 7358 | Fbxo9 | FBXO9 | 1.46E-02 | -8.00E-03 |
| 7359 | Creb3l2 | CREB3L2 | -1.23E-02 | -2.47E-02 |
| 7360 | Ap1s2 | AP1S2 | 2.70E-02 | 8.71E-02 |
| 7361 | Cdca8 | CDCA8 | 1.00E-01 | 1.75E-01 |
| 7362 | Gatad2b | GATAD2B | 1.28E-02 | -1.03E-02 |
| 7363 | Bclaf1 | BCLAF1 | 1.23E-02 | 9.65E-03 |
| 7364 | Banf1 | BANF1 | 1.75E-02 | 5.49E-03 |

| | | | | |
|---|---|---|---|---|
| 7365 | Cdh5 | CDH5 | 2.48E-02 | -1.76E-02 |
| 7366 | Gpatch1 | GPATCH1 | -1.48E-02 | -9.79E-03 |
| 7367 | Sphkap | SPHKAP | 2.58E-02 | 1.30E-02 |
| 7368 | Pcdhb15 | PCDHB7 | -3.19E-02 | 4.32E-02 |
| 7369 | Ptger4 | PTGER4 | -3.90E-02 | 3.69E-02 |
| 7370 | Caps2 | CAPS2 | 4.31E-02 | 3.62E-02 |
| 7371 | Gli3 | GLI3 | -4.78E-02 | 2.66E-02 |
| 7372 | Melk | MELK | 1.13E-01 | 2.07E-01 |
| 7373 | Atad2 | ATAD2 | -6.51E-02 | 1.21E-01 |
| 7374 | Fut10 | FUT10 | -5.23E-02 | 1.63E-02 |
| 7375 | Alg14 | ALG14 | 2.52E-02 | -1.84E-02 |
| 7376 | Gm94 | C5orf46 | 2.53E-02 | -3.36E-02 |
| 7377 | Btbd7 | BTBD7 | -1.77E-02 | -1.62E-03 |
| 7378 | Cct3 | CCT3 | 1.44E-02 | -2.95E-03 |
| 7379 | Ddx43 | DDX43 | -2.55E-02 | 3.51E-03 |
| 7380 | Bok | BOK | 9.15E-02 | 2.64E-02 |
| 7381 | Adipor1 | ADIPOR1 | 9.70E-03 | -4.46E-04 |
| 7382 | Gja1 | GJA1 | -5.10E-02 | 1.04E-01 |
| 7383 | Zfp354b | ZNF354B | -3.02E-02 | -2.49E-02 |
| 7384 | Cwc15 | CWC15 | 1.04E-02 | -1.99E-02 |
| 7385 | Eomes | EOMES | -4.66E-02 | 4.45E-02 |
| 7386 | Ucn2 | UCN2 | 2.51E-02 | 2.05E-02 |
| 7387 | Sstr2 | SSTR2 | 3.95E-02 | -4.84E-02 |
| 7388 | Map6d1 | MAP6D1 | -2.06E-02 | 3.26E-02 |
| 7389 | Ypel3 | YPEL3 | -1.92E-02 | -2.41E-02 |
| 7390 | Lrit3 | LRIT3 | -4.23E-02 | -4.06E-02 |
| 7391 | Chst15 | CHST15 | -2.43E-02 | -5.04E-02 |
| 7392 | Shh | SHH | -1.03E-01 | 5.37E-02 |
| 7393 | Tjp3 | TJP3 | -4.10E-02 | 2.93E-02 |
| 7394 | Coq9 | COQ9 | 1.67E-02 | -2.69E-02 |
| 7395 | Gabarapl1 | GABARAPL1 | -2.19E-02 | -2.41E-02 |
| 7396 | Scamp4 | SCAMP4 | -3.32E-02 | 5.40E-02 |
| 7397 | Ndp | NDP | -2.49E-02 | 1.20E-03 |
| 7398 | Cables1 | CABLES1 | -3.91E-02 | -2.33E-03 |
| 7399 | Nudcd3 | NUDCD3 | 1.21E-02 | -1.15E-02 |
| 7400 | Zdhhc15 | ZDHHC15 | 5.37E-02 | 7.03E-03 |
| 7401 | Huwe1 | HUWE1 | 1.17E-02 | -1.02E-02 |
| 7402 | Idh3g | IDH3G | 1.74E-02 | -1.96E-02 |
| 7403 | Cuedc2 | CUEDC2 | 1.42E-02 | 5.26E-03 |
| 7404 | Gdf6 | GDF6 | 3.53E-02 | -1.43E-02 |
| 7405 | Spg21 | SPG21 | 1.27E-02 | -9.61E-03 |
| 7406 | Sort1 | SORT1 | 4.07E-02 | -2.24E-02 |
| 7407 | Tm2d3 | TM2D3 | -1.47E-02 | -1.03E-02 |
| 7408 | Tssk4 | TSSK4 | 5.40E-02 | 3.85E-03 |
| 7409 | Gm5741 | LOC648044 | -3.48E-02 | 5.57E-02 |
| 7410 | Hspe1 | HSPE1 | -2.03E-02 | -1.12E-02 |
| 7411 | Mmp15 | MMP15 | -6.74E-02 | 2.63E-02 |
| 7412 | Yeats2 | YEATS2 | 1.76E-02 | -1.60E-02 |
| 7413 | Sugp2 | SUGP2 | 1.62E-02 | -1.18E-02 |
| 7414 | Arntl | ARNTL | 8.43E-02 | 1.47E-01 |
| 7415 | Ap3s2 | AP3S2 | 1.19E-02 | -1.45E-02 |
| 7416 | Sqle | SQLE | 5.11E-02 | -2.90E-03 |
| 7417 | Tfpi | TFPI | -3.70E-02 | 2.44E-02 |

| | | | | |
|---|---|---|---|---|
| 7418 | Cltc | CLTC | 1.45E-02 | -7.83E-03 |
| 7419 | Gas2l1 | GAS2L1 | 1.36E-02 | 2.62E-03 |
| 7420 | Slco4a1 | SLCO4A1 | 1.36E-01 | -1.80E-02 |
| 7421 | Lrig2 | LRIG2 | 2.39E-02 | 2.98E-02 |
| 7422 | Hmgxb4 | HMGXB4 | -1.88E-02 | -1.87E-02 |
| 7423 | Lingo4 | LINGO4 | 9.52E-02 | 1.51E-01 |
| 7424 | Gpatch3 | GPATCH3 | 2.32E-02 | -3.89E-02 |
| 7425 | Mtfr2 | MTFR2 | 6.63E-02 | 6.50E-02 |
| 7426 | Tchp | TCHP | -2.17E-02 | -6.07E-03 |
| 7427 | Rbbp8 | RBBP8 | 1.54E-02 | 2.23E-02 |
| 7428 | Nab1 | NAB1 | -1.35E-02 | -1.05E-02 |
| 7429 | Mcf2 | MCF2 | -3.08E-02 | -1.34E-03 |
| 7430 | Syt3 | SYT3 | 2.76E-02 | -1.61E-02 |
| 7431 | Pus1 | PUS1 | 1.78E-02 | -4.40E-03 |
| 7432 | H2-Q1 | HLA-A | -7.07E-02 | 3.53E-02 |
| 7433 | Pigo | PIGO | 1.45E-02 | -1.19E-02 |
| 7434 | Emid1 | EMID1 | 4.37E-02 | -4.21E-02 |
| 7435 | Ube2q1 | UBE2Q1 | -1.00E-02 | -6.87E-03 |
| 7436 | Shmt2 | SHMT2 | 2.72E-02 | -2.19E-02 |
| 7437 | Map2k3 | MAP2K3 | -1.96E-02 | 2.49E-02 |
| 7438 | Nek8 | NEK8 | 1.39E-02 | -2.70E-02 |
| 7439 | Alg12 | ALG12 | 2.25E-02 | -4.15E-03 |
| 7440 | Pard3b | PARD3B | -2.06E-02 | -1.38E-02 |
| 7441 | Vps18 | VPS18 | 1.86E-02 | 6.42E-03 |
| 7442 | Ldb3 | LDB3 | -4.46E-02 | 2.76E-03 |
| 7443 | Gli2 | GLI2 | 3.82E-02 | 2.09E-02 |
| 7444 | Zdhhc23 | ZDHHC23 | -5.28E-02 | -2.52E-04 |
| 7445 | Prim1 | PRIM1 | -3.43E-02 | 5.54E-02 |
| 7446 | Cnksr1 | CNKSR1 | -4.99E-02 | 3.04E-05 |
| 7447 | Caprin1 | CAPRIN1 | -1.48E-02 | 1.18E-02 |
| 7448 | Hao1 | HAO1 | 2.89E-02 | 4.51E-03 |
| 7449 | Nip7 | NIP7 | 1.67E-02 | 5.05E-03 |
| 7450 | Dctpp1 | DCTPP1 | -2.33E-02 | 6.25E-03 |
| 7451 | Ptprr | PTPRR | 6.37E-02 | -3.24E-02 |
| 7452 | Zfp334 | ZNF334 | -4.31E-02 | -1.26E-03 |
| 7453 | F2rl2 | F2RL2 | -2.55E-02 | 6.11E-03 |
| 7454 | Rnf26 | RNF26 | 3.70E-02 | -1.32E-02 |
| 7455 | Cyb5r4 | CYB5R4 | -1.26E-02 | 5.41E-03 |
| 7456 | B4galt4 | B4GALT4 | 2.28E-02 | -8.42E-03 |
| 7457 | Por | POR | -2.31E-02 | -1.47E-02 |
| 7458 | Fgd1 | FGD1 | -2.97E-02 | -8.51E-03 |
| 7459 | Mcoln3 | MCOLN3 | 4.87E-02 | -8.04E-02 |
| 7460 | Oprd1 | OPRD1 | 2.03E-02 | -8.31E-03 |
| 7461 | Zfp560 | ZNF560 | -2.81E-02 | -1.03E-02 |
| 7462 | Tceal5 | TCEAL5 | 4.49E-02 | -3.22E-02 |
| 7463 | Cers1 | CERS1 | -2.85E-02 | 3.78E-03 |
| 7464 | Ckap5 | CKAP5 | -1.41E-02 | 9.20E-03 |
| 7465 | Dhcr7 | DHCR7 | -3.24E-02 | -3.25E-02 |
| 7466 | Rbks | RBKS | 2.22E-02 | 1.19E-02 |
| 7467 | Cpa3 | CPA3 | -2.55E-02 | 1.44E-02 |
| 7468 | Pacsin3 | PACSIN3 | 3.52E-02 | -1.97E-02 |
| 7469 | Mfsd6l | MFSD6L | -5.35E-02 | 1.57E-02 |
| 7470 | Cers3 | CERS3 | 1.32E-01 | 8.97E-02 |

| | | | | |
|---|---|---|---|---|
| 7471 | Prepl | PREPL | 1.13E-02 | 3.24E-03 |
| 7472 | Mtfr1 | MTFR1 | 1.89E-02 | -1.08E-02 |
| 7473 | S100a6 | S100A6 | 1.26E-01 | 1.63E-01 |
| 7474 | Aldh6a1 | ALDH6A1 | 2.52E-02 | -2.84E-02 |
| 7475 | 1810013L24Rik | C16orf72 | -2.37E-02 | -8.88E-03 |
| 7476 | Pcdhac2 | PCDHAC2 | 1.26E-02 | -2.58E-03 |
| 7477 | Ubd | UBD | -9.34E-02 | 3.12E-01 |
| 7478 | Plcxd3 | PLCXD3 | 3.31E-02 | -3.88E-02 |
| 7479 | Mblac1 | MBLAC1 | 4.42E-02 | -1.88E-02 |
| 7480 | Mettl16 | METTL16 | -1.87E-02 | 4.23E-03 |
| 7481 | Ttc30b | TTC30B | -2.79E-02 | -3.14E-02 |
| 7482 | Kifap3 | KIFAP3 | 1.39E-02 | -1.33E-02 |
| 7483 | Kcnh2 | KCNH2 | 8.12E-02 | 4.43E-02 |
| 7484 | Vrk1 | VRK1 | -1.99E-02 | 3.73E-02 |
| 7485 | Mre11a | MRE11A | -1.83E-02 | -8.10E-03 |
| 7486 | Nfe2l1 | NFE2L1 | 1.44E-02 | -2.52E-02 |
| 7487 | Prkca | PRKCA | 1.91E-02 | 6.26E-03 |
| 7488 | Stap1 | STAP1 | -5.31E-02 | 1.43E-01 |
| 7489 | Pcdha3 | PCDHA3 | -1.52E-02 | 1.23E-02 |
| 7490 | Pdss1 | PDSS1 | 2.24E-02 | -7.90E-03 |
| 7491 | Tmem171 | TMEM171 | 5.21E-02 | 2.26E-02 |
| 7492 | Cd300lb | CD300LB | 4.90E-02 | 1.05E-01 |
| 7493 | Elof1 | ELOF1 | 1.33E-02 | -2.48E-03 |
| 7494 | Spin1 | SPIN1 | -1.12E-02 | -8.41E-03 |
| 7495 | Cyp2u1 | CYP2U1 | -3.36E-02 | 3.26E-02 |
| 7496 | Yipf4 | YIPF4 | -1.29E-02 | -1.21E-03 |
| 7497 | Heatr6 | HEATR6 | -1.44E-02 | -5.51E-04 |
| 7498 | Gas2 | GAS2 | 2.49E-02 | -1.84E-03 |
| 7499 | Ptger3 | PTGER3 | 7.29E-02 | -1.09E-01 |
| 7500 | Aph1a | APH1A | 1.64E-02 | -1.04E-03 |
| 7501 | Eea1 | EEA1 | 1.58E-02 | -1.50E-02 |
| 7502 | Atg10 | ATG10 | -1.77E-02 | -5.09E-02 |
| 7503 | Troap | TROAP | 1.06E-01 | 1.49E-01 |
| 7504 | Ccdc102a | CCDC102A | 5.25E-02 | 2.61E-02 |
| 7505 | Nol7 | NOL7 | 1.26E-02 | -1.09E-02 |
| 7506 | Snw1 | SNW1 | 1.53E-02 | -1.37E-02 |
| 7507 | Brf1 | BRF1 | 1.26E-02 | -5.68E-03 |
| 7508 | Spry1 | SPRY1 | -3.59E-02 | -2.91E-02 |
| 7509 | Prox2 | PROX2 | -4.37E-02 | -2.02E-02 |
| 7510 | Trim58 | TRIM58 | 1.78E-02 | 1.04E-02 |
| 7511 | Shc1 | SHC1 | 1.15E-02 | -2.36E-03 |
| 7512 | Morn5 | MORN5 | 4.35E-02 | -2.08E-02 |
| 7513 | Abca13 | ABCA13 | 2.69E-02 | -3.37E-02 |
| 7514 | Stk3 | STK3 | 1.54E-02 | -1.19E-02 |
| 7515 | Ttc37 | TTC37 | -1.75E-02 | 1.87E-02 |
| 7516 | Snn | SNN | 1.54E-02 | -3.81E-02 |
| 7517 | Fnip1 | FNIP1 | -2.65E-02 | -9.92E-03 |
| 7518 | Cct2 | CCT2 | 1.17E-02 | 2.93E-03 |
| 7519 | Tomm40 | TOMM40 | 1.55E-02 | 7.25E-03 |
| 7520 | H2-DMb2 | HLA-DMB | -1.01E-01 | 2.52E-01 |
| 7521 | Bsdc1 | BSDC1 | 1.33E-02 | -8.10E-03 |
| 7522 | Ankfy1 | ANKFY1 | 1.15E-02 | -8.26E-03 |
| 7523 | Mrpl37 | MRPL37 | 1.33E-02 | -6.03E-03 |

| | | | | |
|---|---|---|---|---|
| 7524 | Rnf43 | RNF43 | -4.45E-02 | -2.42E-02 |
| 7525 | Ube2l3 | UBE2L3 | 1.23E-02 | -1.12E-02 |
| 7526 | Sap18 | SAP18 | 1.10E-02 | -1.09E-02 |
| 7527 | Ebi3 | EBI3 | -4.68E-02 | 1.79E-02 |
| 7528 | Chic2 | CHIC2 | 2.61E-02 | -3.16E-03 |
| 7529 | Srgap3 | SRGAP3 | 6.63E-02 | -2.68E-02 |
| 7530 | Odc1 | ODC1 | -2.89E-02 | -9.32E-03 |
| 7531 | Zfp784 | ZNF784 | -3.13E-02 | 2.10E-02 |
| 7532 | Cers6 | CERS6 | -2.20E-02 | -1.87E-02 |
| 7533 | Necap1 | NECAP1 | 1.24E-02 | -1.33E-03 |
| 7534 | Alg1 | ALG1 | 1.23E-02 | -9.14E-03 |
| 7535 | Kif9 | KIF9 | -2.83E-02 | 1.76E-02 |
| 7536 | Tstd1 | TSTD1 | 3.91E-02 | 3.22E-02 |
| 7537 | Cep104 | CEP104 | 1.18E-02 | -6.19E-03 |
| 7538 | Fkbp10 | FKBP10 | 5.21E-02 | 5.08E-02 |
| 7539 | Tagap | TAGAP | 2.95E-02 | -2.47E-02 |
| 7540 | Nwd1 | NWD1 | -3.38E-02 | 1.88E-02 |
| 7541 | Tmem11 | TMEM11 | 1.54E-02 | 2.28E-04 |
| 7542 | Ppil2 | PPIL2 | 9.97E-03 | -1.72E-03 |
| 7543 | Oat | OAT | -2.05E-02 | -1.16E-02 |
| 7544 | Exoc3l2 | EXOC3L2 | 3.04E-02 | -2.94E-02 |
| 7545 | Spata9 | SPATA9 | 2.45E-02 | -4.68E-03 |
| 7546 | Tenm2 | TENM2 | -1.05E-01 | -1.64E-01 |
| 7547 | Hace1 | HACE1 | -2.37E-02 | -2.10E-04 |
| 7548 | Csnk1e | CSNK1E | 2.78E-02 | 2.60E-02 |
| 7549 | Dynlrb1 | DYNLRB1 | 1.12E-02 | -7.55E-03 |
| 7550 | Zfp608 | ZNF608 | -2.43E-02 | -4.05E-03 |
| 7551 | Htr2b | HTR2B | 1.13E-01 | -7.26E-02 |
| 7552 | Btbd6 | BTBD6 | 1.29E-02 | -1.82E-02 |
| 7553 | Tysnd1 | TYSND1 | -1.80E-02 | 6.98E-03 |
| 7554 | Sephs2 | SEPHS2 | 2.41E-02 | -1.37E-02 |
| 7555 | Mrps18a | MRPS18A | 1.38E-02 | -2.54E-02 |
| 7556 | Panx1 | PANX1 | -6.36E-02 | 1.65E-01 |
| 7557 | Pisd | PISD | 1.50E-02 | -7.45E-03 |
| 7558 | Hs3st1 | HS3ST1 | -9.17E-02 | 8.74E-02 |
| 7559 | Mettl9 | METTL9 | 1.29E-02 | 1.41E-03 |
| 7560 | Magt1 | MAGT1 | 1.76E-02 | -2.53E-02 |
| 7561 | Cpd | CPD | 1.10E-02 | -1.33E-02 |
| 7562 | Sp5 | SP5 | 4.07E-02 | 9.37E-03 |
| 7563 | Snap47 | SNAP47 | 1.44E-02 | -1.70E-02 |
| 7564 | Slc7a6 | SLC7A6 | 6.37E-02 | 5.48E-02 |
| 7565 | Lrrc4c | LRRC4C | 2.52E-02 | -2.35E-02 |
| 7566 | Arfip2 | ARFIP2 | 1.05E-02 | -1.82E-02 |
| 7567 | Ier3 | IER3 | 6.13E-02 | 3.25E-02 |
| 7568 | Ganc | GANC | -1.36E-02 | -2.67E-02 |
| 7569 | Fsd2 | FSD2 | -2.85E-02 | 2.66E-02 |
| 7570 | Rps20 | RPS20 | -1.50E-02 | 2.84E-02 |
| 7571 | Arhgef12 | ARHGEF12 | 1.14E-02 | -2.10E-02 |
| 7572 | Tmem253 | TMEM253 | 6.21E-02 | 2.70E-02 |
| 7573 | Gata5 | GATA5 | -5.46E-02 | 1.60E-02 |
| 7574 | Uggt1 | UGGT1 | 1.42E-02 | -5.67E-03 |
| 7575 | Fundc1 | FUNDC1 | 1.59E-02 | -9.14E-04 |
| 7576 | Ddx58 | DDX58 | -1.81E-02 | 8.58E-03 |

| | | | | |
|---|---|---|---|---|
| 7577 | Ddx25 | DDX25 | -2.97E-02 | -6.13E-03 |
| 7578 | Pnkp | PNKP | 1.55E-02 | -1.08E-02 |
| 7579 | Krcc1 | KRCC1 | 1.10E-02 | -1.31E-02 |
| 7580 | Dhodh | DHODH | 1.68E-02 | -2.70E-02 |
| 7581 | Pex12 | PEX12 | 1.54E-02 | -1.27E-02 |
| 7582 | Sstr3 | SSTR3 | -3.46E-02 | -2.61E-02 |
| 7583 | Zfp106 | ZNF106 | 1.33E-02 | -2.51E-02 |
| 7584 | Wdr38 | WDR38 | -5.27E-02 | 2.24E-02 |
| 7585 | Cdh22 | CDH22 | -1.63E-02 | 1.90E-02 |
| 7586 | Mmp28 | MMP28 | -4.75E-02 | 5.91E-04 |
| 7587 | Plch1 | PLCH1 | 4.79E-02 | 3.96E-02 |
| 7588 | Crmp1 | CRMP1 | 3.99E-02 | 1.38E-02 |
| 7589 | Ntrk2 | NTRK2 | 8.07E-02 | 5.18E-02 |
| 7590 | Ccm2l | CCM2L | -2.49E-02 | 2.85E-03 |
| 7591 | Dnah11 | DNAH11 | 1.16E-01 | -1.31E-01 |
| 7592 | Cyb5r1 | CYB5R1 | -2.32E-02 | 7.68E-04 |
| 7593 | Tmem237 | TMEM237 | -1.88E-02 | -4.07E-03 |
| 7594 | Cdc42ep2 | CDC42EP2 | -4.75E-02 | 2.46E-02 |
| 7595 | Heyl | HEYL | -4.25E-02 | 9.91E-02 |
| 7596 | Hspd1 | HSPD1 | -1.86E-02 | -1.85E-02 |
| 7597 | Lamtor4 | LAMTOR4 | 1.79E-02 | -1.69E-02 |
| 7598 | Lingo1 | LINGO1 | -4.86E-02 | 2.32E-02 |
| 7599 | Asb4 | ASB4 | 4.86E-02 | -8.23E-03 |
| 7600 | Kdm4c | KDM4C | -1.46E-02 | -4.39E-03 |
| 7601 | Irf2bpl | IRF2BPL | 3.40E-02 | 5.56E-03 |
| 7602 | Clec4d | CLEC4D | 1.12E-01 | 1.28E-01 |
| 7603 | Tbccd1 | TBCCD1 | -1.55E-02 | -6.93E-03 |
| 7604 | Adora1 | ADORA1 | 1.35E-01 | -2.91E-02 |
| 7605 | Trp53i13 | TP53I13 | -2.78E-02 | 1.15E-03 |
| 7606 | Tpd52l2 | TPD52L2 | -1.12E-02 | 8.41E-03 |
| 7607 | Dock7 | DOCK7 | 1.47E-02 | 2.39E-03 |
| 7608 | Rad51c | RAD51C | -4.87E-02 | 8.67E-02 |
| 7609 | Dhx40 | DHX40 | 1.40E-02 | -3.81E-03 |
| 7610 | Fsd1l | FSD1L | -4.26E-02 | 4.87E-02 |
| 7611 | Rhbdl2 | RHBDL2 | 7.00E-02 | 1.34E-02 |
| 7612 | Acaa1a | ACAA1 | -2.54E-02 | -2.16E-02 |
| 7613 | Lama2 | LAMA2 | -7.17E-02 | -3.96E-02 |
| 7614 | Lrrc23 | LRRC23 | -4.41E-02 | 2.40E-02 |
| 7615 | Apoe | APOE | 2.84E-02 | 5.96E-02 |
| 7616 | Pdcd7 | PDCD7 | -2.36E-02 | 1.41E-02 |
| 7617 | Ccdc96 | CCDC96 | 4.55E-02 | -2.05E-04 |
| 7618 | Hk1 | HK1 | 2.62E-02 | -1.14E-02 |
| 7619 | Elp4 | ELP4 | 2.76E-02 | -1.76E-02 |
| 7620 | Pla2g15 | PLA2G15 | -2.89E-02 | 5.64E-02 |
| 7621 | Gas7 | GAS7 | 5.15E-02 | 4.13E-03 |
| 7622 | Uqcrc2 | UQCRC2 | 1.69E-02 | -2.22E-02 |
| 7623 | Ppp1r7 | PPP1R7 | -1.25E-02 | 9.51E-04 |
| 7624 | Ano5 | ANO5 | -7.51E-02 | 1.11E-03 |
| 7625 | Pcbp1 | PCBP1 | 1.20E-02 | -5.39E-03 |
| 7626 | Arhgap1 | ARHGAP1 | 1.61E-02 | 6.06E-03 |
| 7627 | Gap43 | GAP43 | -1.96E-02 | 5.01E-03 |
| 7628 | Zfand1 | ZFAND1 | -1.30E-02 | -2.30E-02 |
| 7629 | Ndufb11 | NDUFB11 | 1.29E-02 | -1.35E-02 |

| | | | | |
|---|---|---|---|---|
| 7630 | Foxm1 | FOXM1 | 9.08E-02 | 1.04E-01 |
| 7631 | Dnajb5 | DNAJB5 | 2.42E-02 | -1.02E-02 |
| 7632 | Mastl | MASTL | 7.70E-02 | 4.18E-02 |
| 7633 | Arl10 | ARL10 | -2.17E-02 | 8.26E-03 |
| 7634 | Nicn1 | NICN1 | 1.54E-02 | -1.26E-02 |
| 7635 | Pif1 | PIF1 | 1.07E-01 | 1.49E-01 |
| 7636 | Fam210a | FAM210A | -1.96E-02 | -1.76E-02 |
| 7637 | Slc6a17 | SLC6A17 | -4.72E-02 | -5.89E-02 |
| 7638 | Tmem128 | TMEM128 | -1.60E-02 | 1.12E-02 |
| 7639 | Trpv6 | TRPV6 | 6.88E-02 | 9.54E-02 |
| 7640 | Arid5b | ARID5B | -2.52E-02 | 3.04E-02 |
| 7641 | Scand1 | SCAND1 | 2.93E-02 | 2.58E-02 |
| 7642 | Tmem176b | TMEM176B | -1.68E-02 | 3.29E-02 |
| 7643 | Ncoa2 | NCOA2 | -1.92E-02 | -3.06E-02 |
| 7644 | Dennd3 | DENND3 | 3.28E-02 | -2.38E-02 |
| 7645 | Zmiz2 | ZMIZ2 | 1.04E-02 | -8.30E-03 |
| 7646 | Nup133 | NUP133 | -2.12E-02 | 2.22E-03 |
| 7647 | Ccdc34 | CCDC34 | 4.53E-02 | 5.05E-02 |
| 7648 | Ntpcr | NTPCR | 2.48E-02 | -1.11E-02 |
| 7649 | Spire1 | SPIRE1 | 1.61E-02 | -1.37E-02 |
| 7650 | Akr1e1 | AKR1E2 | 1.76E-02 | -2.32E-02 |
| 7651 | Bik | BIK | -5.27E-02 | -2.47E-02 |
| 7652 | Slc26a2 | SLC26A2 | 3.22E-02 | -5.10E-02 |
| 7653 | Serpina6 | SERPINA6 | -2.06E-01 | -2.89E-01 |
| 7654 | Hpdl | HPDL | 6.27E-02 | 4.33E-02 |
| 7655 | Arhgap8 | PRR5-ARHGAP8 | 7.43E-02 | 2.66E-02 |
| 7656 | Arhgap8 | ARHGAP8 | 7.43E-02 | 2.66E-02 |
| 7657 | Arhgef9 | ARHGEF9 | 4.41E-02 | -4.62E-02 |
| 7658 | Ube3b | UBE3B | 1.24E-02 | -1.77E-02 |
| 7659 | Nrg4 | NRG4 | -5.96E-02 | -2.86E-02 |
| 7660 | Dido1 | DIDO1 | 1.49E-02 | -3.90E-03 |
| 7661 | Usf1 | USF1 | -1.55E-02 | -1.18E-03 |
| 7662 | Ramp2 | RAMP2 | 2.91E-02 | 2.63E-02 |
| 7663 | Atp10a | ATP10A | -5.33E-02 | 2.58E-02 |
| 7664 | Prnp | PRNP | -2.94E-02 | -4.32E-03 |
| 7665 | Ppp4c | PPP4C | -1.25E-02 | 1.44E-02 |
| 7666 | Stag2 | STAG2 | -1.44E-02 | -9.37E-03 |
| 7667 | Zfp768 | ZNF768 | -1.64E-02 | -1.26E-02 |
| 7668 | Dock3 | DOCK3 | -3.25E-02 | 3.22E-03 |
| 7669 | Exosc8 | EXOSC8 | -1.99E-02 | 3.63E-02 |
| 7670 | Arhgef7 | ARHGEF7 | -1.58E-02 | -1.75E-02 |
| 7671 | Mpeg1 | MPEG1 | -5.24E-02 | 1.03E-01 |
| 7672 | Timm44 | TIMM44 | 1.22E-02 | -1.55E-02 |
| 7673 | Mkrn3 | MKRN3 | -4.63E-02 | -1.87E-03 |
| 7674 | Cep89 | CEP89 | -1.75E-02 | -2.94E-02 |
| 7675 | Sucla2 | SUCLA2 | 1.60E-02 | -2.46E-02 |
| 7676 | Tmem134 | TMEM134 | 2.61E-02 | 2.67E-03 |
| 7677 | Ttc39c | TTC39C | 2.77E-02 | -1.72E-02 |
| 7678 | Rrp1b | RRP1B | 1.45E-02 | -8.34E-04 |
| 7679 | Slc15a4 | SLC15A4 | -1.82E-02 | 1.73E-02 |
| 7680 | Pank4 | PANK4 | -1.30E-02 | -2.72E-03 |
| 7681 | S1pr5 | S1PR5 | -5.01E-02 | 3.81E-02 |
| 7682 | Il17rd | IL17RD | 3.31E-02 | -2.13E-02 |

| | | | | |
|---|---|---|---|---|
| 7683 | Ddx27 | DDX27 | 9.92E-03 | 9.80E-03 |
| 7684 | Myef2 | MYEF2 | 4.86E-02 | 1.54E-02 |
| 7685 | Siah3 | SIAH3 | 2.08E-02 | -1.24E-02 |
| 7686 | Dpm2 | DPM2 | 1.20E-02 | -3.44E-03 |
| 7687 | Rps6 | RPS6 | -1.20E-02 | 9.23E-03 |
| 7688 | Col24a1 | COL24A1 | 5.61E-02 | 1.62E-01 |
| 7689 | Lhx5 | LHX5 | -1.34E-02 | 7.85E-03 |
| 7690 | Nfyb | NFYB | -1.72E-02 | -1.44E-04 |
| 7691 | Ddrgk1 | DDRGK1 | 1.51E-02 | -1.71E-02 |
| 7692 | Ppip5k2 | PPIP5K2 | -2.02E-02 | -5.59E-04 |
| 7693 | Mrps25 | MRPS25 | 1.50E-02 | 2.45E-03 |
| 7694 | Rps5 | RPS5 | -1.26E-02 | 1.83E-02 |
| 7695 | Slpi | SLPI | 6.15E-02 | 2.55E-01 |
| 7696 | BC005537 | C6orf62 | -1.42E-02 | -2.61E-03 |
| 7697 | Zfp14 | ZFP14 | -4.54E-02 | -4.64E-04 |
| 7698 | Slc25a23 | SLC25A23 | -1.59E-02 | -1.64E-02 |
| 7699 | Rce1 | RCE1 | 1.86E-02 | 4.80E-03 |
| 7700 | Htra2 | HTRA2 | -1.36E-02 | 5.82E-03 |
| 7701 | Atp6v1g3 | ATP6V1G3 | -1.83E-02 | 3.69E-02 |
| 7702 | Zer1 | ZER1 | -1.46E-02 | -2.65E-02 |
| 7703 | Rtp1 | RTP1 | -2.61E-02 | 2.87E-02 |
| 7704 | Cenpk | CENPK | 6.05E-02 | 1.34E-01 |
| 7705 | Fam83e | FAM83E | -5.21E-02 | 4.18E-02 |
| 7706 | Orc2 | ORC2 | 1.30E-02 | 5.28E-03 |
| 7707 | Osbpl6 | OSBPL6 | 2.26E-02 | -2.96E-02 |
| 7708 | Smc3 | SMC3 | -1.36E-02 | -1.05E-02 |
| 7709 | Gpr176 | GPR176 | 8.30E-02 | 1.17E-01 |
| 7710 | Rom1 | ROM1 | 2.52E-02 | -3.84E-03 |
| 7711 | Cisd1 | CISD1 | 1.32E-02 | -1.89E-02 |
| 7712 | Il17ra | IL17RA | 2.09E-02 | -2.79E-02 |
| 7713 | Yif1a | YIF1A | -1.50E-02 | -1.74E-02 |
| 7714 | Scaf4 | SCAF4 | 1.21E-02 | -1.64E-04 |
| 7715 | Cd40 | CD40 | 3.73E-02 | 5.45E-02 |
| 7716 | Psme2 | PSME2 | -1.98E-02 | 2.26E-02 |
| 7717 | Mtg1 | MTG1 | 1.65E-02 | -1.15E-02 |
| 7718 | Fermt2 | FERMT2 | -9.52E-03 | -4.74E-03 |
| 7719 | Slc16a9 | SLC16A9 | 3.56E-02 | -2.83E-02 |
| 7720 | Aprt | APRT | 2.27E-02 | -1.28E-02 |
| 7721 | Mov10 | MOV10 | 2.67E-02 | -7.27E-03 |
| 7722 | Acsm5 | ACSM5 | -2.91E-02 | -4.92E-02 |
| 7723 | Ppp1r35 | PPP1R35 | -2.48E-02 | 1.14E-02 |
| 7724 | Rbpj | RBPJ | -1.27E-02 | -9.31E-03 |
| 7725 | Prpf40b | PRPF40B | 1.67E-02 | -1.50E-02 |
| 7726 | Tmem167b | TMEM167B | 1.05E-02 | -1.69E-02 |
| 7727 | Pawr | PAWR | 2.30E-02 | -8.86E-03 |
| 7728 | Kbtbd8 | KBTBD8 | -6.71E-02 | 4.14E-02 |
| 7729 | Cdkn3 | CDKN3 | 1.36E-01 | 1.84E-01 |
| 7730 | Pex11b | PEX11B | 1.30E-02 | -4.02E-03 |
| 7731 | Lgalsl | LGALSL | 2.96E-02 | 3.32E-02 |
| 7732 | Dpep3 | DPEP3 | 2.98E-02 | 3.89E-02 |
| 7733 | Sec11c | SEC11C | 1.28E-02 | 3.25E-03 |
| 7734 | Zfp292 | ZNF292 | -1.64E-02 | -6.13E-03 |
| 7735 | Med12 | MED12 | 1.55E-02 | -1.06E-02 |

| | | | | |
|---|---|---|---|---|
| 7736 | Pde1c | PDE1C | 1.05E-01 | 2.31E-02 |
| 7737 | Gpr50 | GPR50 | 1.73E-02 | -3.81E-03 |
| 7738 | Arhgap40 | ARHGAP40 | 1.04E-01 | 5.07E-02 |
| 7739 | Eef1b2 | EEF1B2 | -1.15E-02 | 2.07E-02 |
| 7740 | Sacm1l | SACM1L | 1.35E-02 | -1.57E-02 |
| 7741 | Appl2 | APPL2 | -1.75E-02 | -3.79E-02 |
| 7742 | Tgfbr1 | TGFBR1 | 1.40E-02 | 1.98E-02 |
| 7743 | Klhl14 | KLHL14 | -5.95E-02 | -3.39E-02 |
| 7744 | Slc9a7 | SLC9A7 | -3.48E-02 | 3.09E-02 |
| 7745 | Cox16 | COX16 | -3.87E-02 | -1.53E-02 |
| 7746 | Lig3 | LIG3 | -1.15E-02 | -5.28E-03 |
| 7747 | Dnmt3b | DNMT3B | 3.49E-02 | -2.46E-02 |
| 7748 | Zfp341 | ZNF341 | -3.02E-02 | 8.47E-03 |
| 7749 | Top3b | TOP3B | 2.38E-02 | -1.60E-02 |
| 7750 | Adamts10 | ADAMTS10 | -3.13E-02 | 1.60E-02 |
| 7751 | Pigb | PIGB | 2.11E-02 | -2.10E-02 |
| 7752 | St3gal4 | ST3GAL4 | -2.80E-02 | 2.40E-02 |
| 7753 | Ccdc58 | CCDC58 | -1.58E-02 | 8.74E-03 |
| 7754 | C1qtnf9 | C1QTNF9B | -6.34E-02 | -6.89E-02 |
| 7755 | Rgsl1 | RGSL1 | -2.40E-02 | 2.74E-03 |
| 7756 | Cdca5 | CDCA5 | 7.65E-02 | 2.22E-01 |
| 7757 | Alkbh4 | ALKBH4 | -2.61E-02 | 1.41E-03 |
| 7758 | Ncoa3 | NCOA3 | -1.13E-02 | -1.36E-02 |
| 7759 | Zfp422 | ZNF22 | -1.22E-02 | -1.06E-02 |
| 7760 | Atad5 | ATAD5 | 5.38E-02 | 6.46E-03 |
| 7761 | Mettl22 | METTL22 | 2.24E-02 | -8.90E-03 |
| 7762 | C2cd4d | C2CD4D | 3.84E-02 | 4.81E-04 |
| 7763 | Usp54 | USP54 | 2.70E-02 | -2.94E-02 |
| 7764 | Ppp1r36 | PPP1R36 | 9.32E-02 | 9.30E-03 |
| 7765 | Adam10 | ADAM10 | -1.24E-02 | 8.66E-03 |
| 7766 | Jdp2 | JDP2 | -3.77E-02 | 8.32E-03 |
| 7767 | Lrrc47 | LRRC47 | 1.17E-02 | -6.49E-03 |
| 7768 | Gpatch2 | GPATCH2 | 1.26E-02 | -1.15E-02 |
| 7769 | Polm | POLM | 2.26E-02 | -2.08E-02 |
| 7770 | Tpcn2 | TPCN2 | 4.72E-02 | 2.32E-02 |
| 7771 | L1cam | L1CAM | 4.42E-02 | -5.89E-03 |
| 7772 | Miip | MIIP | 1.78E-02 | -1.26E-03 |
| 7773 | Golga1 | GOLGA1 | -1.18E-02 | -4.33E-03 |
| 7774 | Rep15 | REP15 | 5.98E-02 | -1.69E-02 |
| 7775 | Ndufa4 | NDUFA4 | 1.34E-02 | -1.21E-02 |
| 7776 | Lsm1 | LSM1 | 1.73E-02 | 3.01E-03 |
| 7777 | Tex26 | TEX26 | -2.07E-02 | 1.57E-02 |
| 7778 | Agrn | AGRN | -1.73E-02 | 1.76E-02 |
| 7779 | Fam114a1 | FAM114A1 | -3.37E-02 | -2.89E-02 |
| 7780 | Rprd1a | RPRD1A | -2.22E-02 | -2.24E-02 |
| 7781 | Frmd7 | FRMD7 | 3.70E-02 | 1.06E-03 |
| 7782 | Slc12a7 | SLC12A7 | 2.13E-02 | -3.32E-02 |
| 7783 | Hmox1 | HMOX1 | 1.38E-01 | 2.92E-01 |
| 7784 | Slc13a3 | SLC13A3 | 3.30E-02 | 2.23E-02 |
| 7785 | Ankrd22 | ANKRD22 | 2.74E-02 | -8.70E-03 |
| 7786 | Dtna | DTNA | -3.94E-02 | -2.60E-02 |
| 7787 | Satb1 | SATB1 | 4.20E-02 | 6.18E-02 |
| 7788 | Stmn4 | STMN4 | -1.81E-02 | 2.77E-02 |

| | | | | |
|---|---|---|---|---|
| 7789 | Tmem108 | TMEM108 | 6.91E-02 | -6.18E-03 |
| 7790 | Neb | NEB | -4.34E-02 | 1.88E-02 |
| 7791 | Pygb | PYGB | 2.20E-02 | 1.74E-02 |
| 7792 | Rfc4 | RFC4 | 3.30E-02 | 3.85E-02 |
| 7793 | Zfp329 | ZNF329 | -2.40E-02 | 2.12E-02 |
| 7794 | Klhl8 | KLHL8 | -2.94E-02 | 3.57E-03 |
| 7795 | Kcnmb1 | KCNMB1 | 4.49E-02 | 1.93E-02 |
| 7796 | Trp53 | TP53 | 1.61E-02 | 2.47E-02 |
| 7797 | Pdk2 | PDK2 | 2.04E-02 | -3.23E-02 |
| 7798 | Tmem117 | TMEM117 | -2.85E-02 | 3.03E-03 |
| 7799 | Usp8 | USP8 | -1.42E-02 | -1.27E-02 |
| 7800 | Vwa3b | VWA3B | -1.79E-02 | -1.36E-03 |
| 7801 | Zc3h8 | ZC3H8 | 2.80E-02 | 1.72E-02 |
| 7802 | Mtor | MTOR | -1.98E-02 | -3.28E-02 |
| 7803 | Pnp2 | PNP | 3.46E-02 | -4.62E-03 |
| 7804 | Mfap2 | MFAP2 | 6.93E-02 | 5.41E-02 |
| 7805 | Pilrb1 | PILRB | -5.82E-02 | 7.92E-02 |
| 7806 | Egln2 | EGLN2 | 1.19E-02 | -8.12E-03 |
| 7807 | Lrrn1 | LRRN1 | 4.29E-02 | 7.51E-02 |
| 7808 | Atp5b | ATP5B | 1.56E-02 | -1.97E-02 |
| 7809 | Nnmt | NNMT | 6.90E-02 | 4.62E-02 |
| 7810 | Trip13 | TRIP13 | 6.76E-02 | 9.53E-02 |
| 7811 | Asb10 | ASB10 | -2.40E-02 | 2.07E-02 |
| 7812 | Izumo1 | IZUMO1 | 2.58E-02 | -3.78E-02 |
| 7813 | Ccdc6 | CCDC6 | -1.83E-02 | 4.13E-03 |
| 7814 | Ccno | CCNO | 4.55E-02 | 4.42E-02 |
| 7815 | Chtf8 | CHTF8 | -1.18E-02 | 7.34E-03 |
| 7816 | Clec14a | CLEC14A | 4.62E-02 | -6.24E-02 |
| 7817 | Gle1 | GLE1 | -1.11E-02 | -1.51E-02 |
| 7818 | Ankrd13d | ANKRD13D | 5.49E-02 | 2.70E-02 |
| 7819 | Gpr150 | GPR150 | -3.35E-02 | 9.26E-03 |
| 7820 | Igfbp1 | IGFBP1 | -9.00E-02 | -1.29E-01 |
| 7821 | Slc6a13 | SLC6A13 | 2.29E-02 | -3.09E-02 |
| 7822 | Nsa2 | NSA2 | -1.29E-02 | 2.07E-02 |
| 7823 | Tmem121 | TMEM121 | 1.44E-02 | -1.63E-02 |
| 7824 | Tmem28 | FAM155B | -3.92E-02 | -1.53E-02 |
| 7825 | Gnpda1 | GNPDA1 | -2.29E-02 | -1.68E-02 |
| 7826 | Gsto1 | GSTO1 | 1.34E-02 | 1.58E-02 |
| 7827 | Ddr2 | DDR2 | -4.92E-02 | -1.33E-03 |
| 7828 | Ergic2 | ERGIC2 | 1.02E-02 | 1.15E-02 |
| 7829 | Cln3 | CLN3 | 1.40E-02 | -2.21E-02 |
| 7830 | Cntd1 | CNTD1 | -3.73E-02 | -6.34E-02 |
| 7831 | Clint1 | CLINT1 | -1.21E-02 | 4.34E-03 |
| 7832 | Slc22a12 | SLC22A12 | 3.17E-02 | -1.50E-02 |
| 7833 | Rpap1 | RPAP1 | -1.47E-02 | -2.65E-02 |
| 7834 | Clip2 | CLIP2 | 2.37E-02 | -1.77E-02 |
| 7835 | Scn3b | SCN3B | -2.33E-02 | 8.62E-03 |
| 7836 | Fam13b | FAM13B | -2.16E-02 | 2.61E-03 |
| 7837 | Gata2 | GATA2 | -3.35E-02 | 2.65E-03 |
| 7838 | Ttll5 | TTLL5 | -2.07E-02 | -2.05E-02 |
| 7839 | Ccnd3 | CCND3 | 1.40E-02 | -3.54E-03 |
| 7840 | Prdm11 | PRDM11 | -3.99E-02 | 1.69E-02 |
| 7841 | Sptlc3 | SPTLC3 | -6.46E-02 | 1.46E-02 |

| | | | | |
|---|---|---|---|---|
| 7842 | Ppm1f | PPM1F | 1.17E-02 | -1.44E-02 |
| 7843 | Ugcg | UGCG | 2.03E-02 | 2.09E-02 |
| 7844 | Ankrd49 | ANKRD49 | -2.23E-02 | 2.33E-02 |
| 7845 | Atg5 | ATG5 | 1.29E-02 | -1.05E-02 |
| 7846 | Bckdhb | BCKDHB | -3.00E-02 | -4.84E-02 |
| 7847 | Cyp4b1 | CYP4B1 | 3.97E-02 | 5.06E-03 |
| 7848 | Zfp280b | ZNF280B | -2.98E-02 | -2.24E-02 |
| 7849 | Dnajc2 | DNAJC2 | 1.34E-02 | 1.03E-02 |
| 7850 | Ddb2 | DDB2 | 2.24E-02 | -1.45E-02 |
| 7851 | Mief1 | MIEF1 | -1.33E-02 | -2.73E-03 |
| 7852 | Dennd6b | DENND6B | 1.61E-02 | -1.71E-02 |
| 7853 | Micu1 | MICU1 | 1.44E-02 | -2.45E-02 |
| 7854 | Trmt2a | TRMT2A | -1.46E-02 | 1.28E-02 |
| 7855 | Rit1 | RIT1 | -1.09E-02 | 8.08E-03 |
| 7856 | Cyp8b1 | CYP8B1 | -2.12E-02 | 2.49E-02 |
| 7857 | Sh3bgrl3 | SH3BGRL3 | 1.93E-02 | 8.33E-02 |
| 7858 | Tspan7 | TSPAN7 | 2.43E-02 | 1.01E-02 |
| 7859 | Uap1l1 | UAP1L1 | 2.67E-02 | -6.41E-03 |
| 7860 | Scaf8 | SCAF8 | -1.13E-02 | 4.76E-04 |
| 7861 | Fh1 | FH | 1.44E-02 | 3.45E-03 |
| 7862 | Yap1 | YAP1 | -1.56E-02 | 6.06E-03 |
| 7863 | Bms1 | BMS1 | -8.80E-03 | 4.75E-05 |
| 7864 | Samd14 | SAMD14 | -3.40E-02 | 7.86E-02 |
| 7865 | Nbeal1 | NBEAL1 | 1.63E-02 | -5.50E-04 |
| 7866 | Ern1 | ERN1 | 1.36E-02 | -3.93E-03 |
| 7867 | Asb5 | ASB5 | -4.97E-02 | 3.56E-02 |
| 7868 | Gpc2 | GPC2 | -5.58E-02 | 1.70E-02 |
| 7869 | Slc1a6 | SLC1A6 | 2.83E-02 | 8.23E-03 |
| 7870 | Slc38a6 | SLC38A6 | -2.37E-02 | -2.21E-02 |
| 7871 | Gjb1 | GJB1 | -3.68E-02 | -5.95E-03 |
| 7872 | Cdkl1 | CDKL1 | -2.03E-02 | -6.84E-02 |
| 7873 | Pfkfb1 | PFKFB1 | 4.87E-02 | 3.55E-02 |
| 7874 | Ccdc112 | CCDC112 | 4.58E-02 | -4.09E-02 |
| 7875 | Esco1 | ESCO1 | -1.97E-02 | 7.90E-03 |
| 7876 | Usp29 | USP29 | 3.38E-02 | 3.49E-02 |
| 7877 | Pdrg1 | PDRG1 | -1.33E-02 | -4.08E-03 |
| 7878 | Folr1 | FOLR1 | -1.96E-02 | -2.12E-02 |
| 7879 | Taok3 | TAOK3 | -1.62E-02 | 1.25E-03 |
| 7880 | Sf3b1 | SF3B1 | 1.08E-02 | -7.61E-03 |
| 7881 | Bco2 | BCO2 | -4.18E-02 | -6.85E-02 |
| 7882 | Aldh7a1 | ALDH7A1 | 2.55E-02 | -2.23E-02 |
| 7883 | Hhatl | HHATL | -3.92E-02 | 1.02E-01 |
| 7884 | Slc22a1 | SLC22A1 | 2.28E-02 | -1.48E-02 |
| 7885 | Actr3b | ACTR3B | 3.73E-02 | 2.48E-02 |
| 7886 | Htr1b | HTR1B | 1.71E-01 | -1.19E-01 |
| 7887 | Eme2 | EME2 | -1.95E-02 | -2.32E-02 |
| 7888 | Eif1a | EIF1AX | -2.42E-02 | 3.17E-02 |
| 7889 | Tmem256 | TMEM256 | 1.86E-02 | -7.07E-03 |
| 7890 | Tmem88 | TMEM88 | 3.14E-02 | -2.08E-02 |
| 7891 | Pinx1 | PINX1 | 4.14E-02 | 8.07E-02 |
| 7892 | Adprh | ADPRH | 1.25E-02 | -1.68E-02 |
| 7893 | Ncapd3 | NCAPD3 | -1.37E-02 | 5.95E-03 |
| 7894 | Cyp4v3 | CYP4V2 | 2.88E-02 | 5.30E-02 |

| | | | | |
|---|---|---|---|---|
| 7895 | Hspa12a | HSPA12A | -2.52E-02 | -4.30E-02 |
| 7896 | Trim8 | TRIM8 | -1.12E-02 | 1.96E-02 |
| 7897 | Mrpl45 | MRPL45 | 1.35E-02 | -8.60E-03 |
| 7898 | Nudt14 | NUDT14 | 1.69E-02 | -1.27E-02 |
| 7899 | Meiob | MEIOB | -4.78E-02 | -1.66E-02 |
| 7900 | Cmc1 | CMC1 | -1.68E-02 | 1.44E-02 |
| 7901 | Lhx9 | LHX9 | -1.72E-02 | -7.34E-03 |
| 7902 | Fmn2 | FMN2 | 3.51E-02 | -1.31E-03 |
| 7903 | Ing3 | ING3 | -1.56E-02 | -9.17E-03 |
| 7904 | Ifngr2 | IFNGR2 | 1.97E-02 | 4.20E-02 |
| 7905 | B3gnt3 | B3GNT3 | 4.66E-02 | 6.42E-02 |
| 7906 | Zfp13 | ZNF205 | -3.93E-02 | -3.47E-02 |
| 7907 | Olfr1029 | OR5M11 | -1.57E-02 | 7.53E-03 |
| 7908 | Zfp963 | ZNF669 | -4.14E-02 | -5.49E-03 |
| 7909 | Megf11 | MEGF11 | -1.01E-01 | 3.81E-02 |
| 7910 | Enpp4 | ENPP4 | -1.68E-02 | -1.02E-02 |
| 7911 | Sult2b1 | SULT2B1 | 1.11E-01 | -8.13E-02 |
| 7912 | Ttc21b | TTC21B | -1.81E-02 | -1.99E-02 |
| 7913 | Xrra1 | XRRA1 | 2.82E-02 | -9.96E-03 |
| 7914 | Sec22c | SEC22C | 2.14E-02 | 1.08E-02 |
| 7915 | Ezr | EZR | 1.42E-02 | -4.88E-03 |
| 7916 | Ogt | OGT | -1.17E-02 | -7.90E-03 |
| 7917 | Olfr1034 | OR5M9 | 5.19E-02 | 1.39E-02 |
| 7918 | Elk4 | ELK4 | 1.33E-02 | -7.55E-04 |
| 7919 | Cacna1d | CACNA1D | 4.61E-02 | -1.67E-03 |
| 7920 | Col8a2 | COL8A2 | 3.86E-02 | 4.82E-02 |
| 7921 | Echdc2 | ECHDC2 | -1.60E-02 | -2.18E-02 |
| 7922 | Espl1 | ESPL1 | 9.97E-02 | 1.56E-01 |
| 7923 | Mplkip | MPLKIP | -1.80E-02 | 2.24E-02 |
| 7924 | Mnd1 | MND1 | -4.53E-02 | 7.34E-03 |
| 7925 | Foxj1 | FOXJ1 | 4.93E-02 | 1.48E-01 |
| 7926 | Slc44a1 | SLC44A1 | 2.16E-02 | 2.76E-03 |
| 7927 | Sclt1 | SCLT1 | -1.48E-02 | -3.03E-03 |
| 7928 | Zfyve28 | ZFYVE28 | 4.67E-02 | 2.35E-03 |
| 7929 | Tnfaip2 | TNFAIP2 | 2.72E-02 | 1.22E-02 |
| 7930 | Abca9 | ABCA9 | -6.00E-02 | -1.10E-02 |
| 7931 | Tm4sf20 | TM4SF20 | 1.44E-01 | -3.55E-03 |
| 7932 | Foxred2 | FOXRED2 | 3.42E-02 | 3.58E-02 |
| 7933 | P2ry1 | P2RY1 | 4.07E-02 | 3.32E-02 |
| 7934 | Klrb1a | KLRB1 | 2.31E-02 | 1.07E-02 |
| 7935 | Ccdc125 | CCDC125 | 3.29E-02 | 8.22E-03 |
| 7936 | Tmem232 | TMEM232 | -2.23E-02 | -1.12E-02 |
| 7937 | Zfp438 | ZNF438 | -3.09E-02 | -1.43E-02 |
| 7938 | 4930563E22Rik | C17orf100 | -4.25E-02 | 4.40E-03 |
| 7939 | Cdadc1 | CDADC1 | -1.55E-02 | -1.86E-02 |
| 7940 | Sowaha | SOWAHA | -4.72E-02 | -2.47E-02 |
| 7941 | Neil3 | NEIL3 | 1.10E-01 | 1.05E-01 |
| 7942 | Egfl8 | EGFL8 | -4.74E-02 | -2.17E-02 |
| 7943 | Il23r | IL23R | 2.46E-02 | 1.86E-02 |
| 7944 | Lrrk1 | LRRK1 | -2.92E-02 | -1.56E-02 |
| 7945 | Rcor2 | RCOR2 | 5.69E-02 | 4.16E-02 |
| 7946 | Adck2 | ADCK2 | -2.17E-02 | 8.69E-03 |
| 7947 | Angptl6 | ANGPTL6 | -3.97E-02 | 1.12E-02 |

| | | | | |
|---|---|---|---|---|
| 7948 | Smarca5 | SMARCA5 | 1.07E-02 | 1.71E-02 |
| 7949 | Zfp867 | ZNF627 | 3.72E-02 | -3.50E-02 |
| 7950 | Pla2g5 | PLA2G5 | -3.50E-02 | -3.21E-03 |
| 7951 | Hoxa1 | HOXA1 | -3.68E-02 | 1.27E-02 |
| 7952 | Snca | SNCA | -6.35E-02 | -8.82E-03 |
| 7953 | Sytl1 | SYTL1 | -3.47E-02 | -4.47E-02 |
| 7954 | Rnf34 | RNF34 | -1.19E-02 | -4.88E-03 |
| 7955 | Batf3 | BATF3 | 5.80E-02 | 1.87E-02 |
| 7956 | Sprr1a | SPRR1A | 2.27E-01 | 2.23E-01 |
| 7957 | Wnk3 | WNK3 | 5.98E-02 | -1.67E-02 |
| 7958 | Ribc1 | RIBC1 | -4.12E-02 | -2.18E-02 |
| 7959 | Diablo | DIABLO | 1.10E-02 | -5.73E-03 |
| 7960 | Rps27a | RPS27A | -1.17E-02 | 8.83E-03 |
| 7961 | Mpped2 | MPPED2 | -6.93E-02 | -7.07E-02 |
| 7962 | Alas2 | ALAS2 | -9.23E-02 | 5.11E-02 |
| 7963 | Tspan31 | TSPAN31 | 1.04E-02 | 2.34E-03 |
| 7964 | Tmem45b | TMEM45B | -4.82E-02 | 4.58E-02 |
| 7965 | Wnt6 | WNT6 | 3.61E-02 | -4.86E-03 |
| 7966 | Atxn2 | ATXN2 | -1.29E-02 | 6.14E-04 |
| 7967 | Prrc1 | PRRC1 | 1.07E-02 | -3.38E-02 |
| 7968 | Rnf217 | RNF217 | -2.32E-02 | 1.50E-02 |
| 7969 | Tarm1 | TARM1 | 6.38E-02 | -2.75E-02 |
| 7970 | Xkr9 | XKR9 | -4.97E-02 | 2.25E-02 |
| 7971 | Abat | ABAT | -2.53E-02 | -3.44E-02 |
| 7972 | Mcm5 | MCM5 | 7.01E-02 | 1.59E-01 |
| 7973 | Txndc11 | TXNDC11 | 1.72E-02 | -2.37E-03 |
| 7974 | Zfp69 | ZFP69 | 6.09E-02 | -3.35E-02 |
| 7975 | Lysmd4 | LYSMD4 | 2.10E-02 | -1.93E-02 |
| 7976 | Ndel1 | NDEL1 | 1.80E-02 | -7.88E-03 |
| 7977 | Npas2 | NPAS2 | 8.33E-02 | 4.98E-02 |
| 7978 | Chp1 | CHP1 | 1.65E-02 | 4.31E-03 |
| 7979 | B4galt3 | B4GALT3 | -1.65E-02 | -3.80E-03 |
| 7980 | Lrba | LRBA | 1.27E-02 | -2.55E-02 |
| 7981 | Ugt2b5 | UGT2B28 | 8.11E-02 | 1.83E-01 |
| 7982 | Nell2 | NELL2 | -4.20E-02 | 6.26E-02 |
| 7983 | Mfsd1 | MFSD1 | 1.89E-02 | -1.76E-02 |
| 7984 | Cdr2l | CDR2L | 4.04E-02 | 3.78E-02 |
| 7985 | Carf | CARF | 2.50E-02 | 2.34E-03 |
| 7986 | Toe1 | TOE1 | -1.58E-02 | 7.46E-03 |
| 7987 | Pm20d2 | PM20D2 | 4.45E-02 | 1.52E-02 |
| 7988 | Ctnnbl1 | CTNNBL1 | 1.68E-02 | -6.63E-03 |
| 7989 | Gemin5 | GEMIN5 | -1.20E-02 | 1.74E-02 |
| 7990 | Sema3c | SEMA3C | 2.67E-02 | 6.74E-02 |
| 7991 | Etfa | ETFA | 1.52E-02 | -2.45E-02 |
| 7992 | Ptprz1 | PTPRZ1 | -2.66E-02 | 1.63E-02 |
| 7993 | Ddx41 | DDX41 | -9.76E-03 | 2.02E-03 |
| 7994 | Rfng | RFNG | -1.78E-02 | -4.37E-03 |
| 7995 | Thtpa | THTPA | 2.46E-02 | -2.65E-02 |
| 7996 | Tm4sf1 | TM4SF1 | 1.67E-02 | -1.08E-02 |
| 7997 | Cldn22 | CLDN22 | -2.45E-02 | 8.05E-03 |
| 7998 | Zbtb1 | ZBTB1 | 1.86E-02 | 2.53E-03 |
| 7999 | Mest | MEST | 3.47E-02 | 9.62E-02 |
| 8000 | Mrpl51 | MRPL51 | 1.18E-02 | -4.90E-03 |

| | | | | |
|---|---|---|---|---|
| 8001 | Gfm2 | GFM2 | -1.25E-02 | -1.77E-02 |
| 8002 | Kctd2 | KCTD2 | -1.42E-02 | -1.02E-02 |
| 8003 | Cenpc1 | CENPC | 2.19E-02 | -1.61E-02 |
| 8004 | Fam181b | FAM181B | -6.02E-02 | 3.42E-03 |
| 8005 | Tfb2m | TFB2M | -1.48E-02 | -2.14E-02 |
| 8006 | Sms | SMS | 2.50E-02 | -2.83E-03 |
| 8007 | Pdxk | PDXK | -2.09E-02 | -7.83E-03 |
| 8008 | Unc5b | UNC5B | 2.72E-02 | 3.09E-02 |
| 8009 | P2ry2 | P2RY2 | 6.43E-02 | -8.47E-02 |
| 8010 | Ndufa4l2 | NDUFA4L2 | 4.34E-02 | 4.60E-03 |
| 8011 | Tfe3 | TFE3 | 1.02E-02 | -2.76E-03 |
| 8012 | Gstm4 | GSTM4 | -3.10E-02 | -2.05E-02 |
| 8013 | Thbs3 | THBS3 | 3.74E-02 | 6.25E-02 |
| 8014 | Tigd5 | TIGD5 | -3.69E-02 | -4.56E-02 |
| 8015 | Ttll1 | TTLL1 | -2.22E-02 | 1.93E-02 |
| 8016 | Ssx2ip | SSX2IP | 1.25E-02 | -9.52E-03 |
| 8017 | B3gnt9 | B3GNT9 | -6.74E-02 | 7.46E-02 |
| 8018 | Tnfsf8 | TNFSF8 | 3.34E-02 | 2.31E-02 |
| 8019 | Grik2 | GRIK2 | -2.20E-02 | 1.46E-02 |
| 8020 | Tmem71 | TMEM71 | -3.75E-02 | 5.08E-02 |
| 8021 | Ddx56 | DDX56 | 1.26E-02 | 4.05E-03 |
| 8022 | Dhrs7b | DHRS7B | 1.59E-02 | 2.42E-04 |
| 8023 | Magee1 | MAGEE1 | -3.11E-02 | -1.96E-02 |
| 8024 | Hectd2 | HECTD2 | 4.51E-02 | 2.70E-02 |
| 8025 | Surf2 | SURF2 | -1.34E-02 | -5.48E-03 |
| 8026 | Adamts2 | ADAMTS2 | 8.64E-02 | 1.43E-01 |
| 8027 | Exo5 | EXO5 | -3.26E-02 | 5.66E-03 |
| 8028 | Eya3 | EYA3 | -9.99E-03 | -1.02E-02 |
| 8029 | Ccdc68 | CCDC68 | 2.16E-02 | -2.27E-02 |
| 8030 | Tmem104 | TMEM104 | 1.16E-02 | -2.74E-03 |
| 8031 | Pir | PIR | 5.14E-02 | 1.44E-02 |
| 8032 | Lilra6 | LILRB3 | -1.59E-02 | 3.11E-02 |
| 8033 | Ermard | ERMARD | 1.32E-02 | -2.21E-03 |
| 8034 | N4bp3 | N4BP3 | 2.53E-02 | 4.17E-02 |
| 8035 | Ankrd33b | ANKRD33B | 1.92E-02 | -2.85E-03 |
| 8036 | Gzf1 | GZF1 | -1.16E-02 | -1.69E-02 |
| 8037 | Gpc5 | GPC5 | -7.83E-02 | -3.82E-02 |
| 8038 | Armc9 | ARMC9 | 2.50E-02 | -1.33E-04 |
| 8039 | Atp6v1a | ATP6V1A | 1.45E-02 | -1.49E-02 |
| 8040 | Xkr4 | XKR4 | 2.18E-02 | 3.52E-02 |
| 8041 | Dcbld1 | DCBLD1 | -1.55E-02 | 8.66E-03 |
| 8042 | E130311K13Rik | C3orf33 | -2.46E-02 | -3.03E-02 |
| 8043 | Atp6v0a2 | ATP6V0A2 | 1.05E-02 | -5.94E-03 |
| 8044 | Ankrd66 | ANKRD66 | 1.59E-02 | 1.10E-02 |
| 8045 | Prdm4 | PRDM4 | 1.23E-02 | 4.76E-03 |
| 8046 | Golim4 | GOLIM4 | -2.01E-02 | -9.07E-03 |
| 8047 | Ngf | NGF | 5.29E-02 | 1.38E-01 |
| 8048 | Tmed9 | TMED9 | 1.17E-02 | -5.49E-03 |
| 8049 | Ropn1l | ROPN1L | -3.55E-02 | 3.27E-02 |
| 8050 | Clic4 | CLIC4 | 1.05E-02 | 2.90E-03 |
| 8051 | Hsd11b2 | HSD11B2 | -1.52E-02 | -7.34E-03 |
| 8052 | Eps8 | EPS8 | -2.02E-02 | -1.89E-02 |
| 8053 | Skp2 | SKP2 | -3.95E-02 | 4.24E-02 |

| | | | | |
|---|---|---|---|---|
| 8054 | Ppa1 | PPA1 | -1.83E-02 | -1.91E-02 |
| 8055 | Yars2 | YARS2 | -1.73E-02 | -4.08E-03 |
| 8056 | Klhdc9 | KLHDC9 | -3.47E-02 | -8.34E-02 |
| 8057 | Zfp628 | ZNF628 | 2.29E-02 | -1.37E-02 |
| 8058 | Ly6e | LY6E | 3.01E-02 | 5.40E-02 |
| 8059 | Kcnb1 | KCNB1 | 3.60E-02 | -6.66E-02 |
| 8060 | Klk1b8 | KLK3 | 4.88E-02 | 1.11E-01 |
| 8061 | Hdac10 | HDAC10 | -1.59E-02 | -2.69E-02 |
| 8062 | Nkap | NKAP | 1.54E-02 | -1.88E-02 |
| 8063 | Snx33 | SNX33 | -2.13E-02 | -6.79E-03 |
| 8064 | Wnt16 | WNT16 | -3.89E-02 | 1.96E-02 |
| 8065 | Ndfip1 | NDFIP1 | 1.21E-02 | -1.15E-02 |
| 8066 | Atpif1 | ATPIF1 | -1.52E-02 | -1.26E-02 |
| 8067 | Il20ra | IL20RA | 1.67E-02 | -1.94E-02 |
| 8068 | Plekhb2 | PLEKHB2 | 1.81E-02 | -2.00E-02 |
| 8069 | Stxbp1 | STXBP1 | 4.07E-02 | 1.07E-03 |
| 8070 | Ppp1r1b | PPP1R1B | -8.34E-02 | -1.20E-01 |
| 8071 | Hccs | HCCS | 1.71E-02 | -2.85E-03 |
| 8072 | Zc3h14 | ZC3H14 | 1.10E-02 | -1.51E-02 |
| 8073 | Prr14l | PRR14L | -1.73E-02 | -2.04E-03 |
| 8074 | Pcdhb17 | PCDHB16 | 4.03E-02 | 5.02E-02 |
| 8075 | Hlcs | HLCS | 1.58E-02 | -1.57E-02 |
| 8076 | Rasa3 | RASA3 | 3.35E-02 | -1.27E-02 |
| 8077 | Wbp1 | WBP1 | 9.00E-03 | -7.05E-03 |
| 8078 | Shd | SHD | -3.61E-02 | -1.97E-02 |
| 8079 | Katna1 | KATNA1 | -1.47E-02 | -8.76E-03 |
| 8080 | Puf60 | PUF60 | 8.93E-03 | -9.33E-03 |
| 8081 | Ankrd2 | ANKRD2 | -2.14E-02 | 2.38E-03 |
| 8082 | Ppp1r14b | PPP1R14B | 2.79E-02 | 4.88E-02 |
| 8083 | Snx24 | SNX24 | 1.91E-02 | -1.71E-03 |
| 8084 | Msh4 | MSH4 | 2.40E-02 | 1.38E-02 |
| 8085 | Depdc7 | DEPDC7 | 2.56E-02 | -1.76E-03 |
| 8086 | Foxj2 | FOXJ2 | -1.47E-02 | -6.83E-03 |
| 8087 | Nkd2 | NKD2 | -5.38E-02 | 1.63E-01 |
| 8088 | Cc2d2a | CC2D2A | -2.51E-02 | -1.42E-02 |
| 8089 | Fbxo31 | FBXO31 | 1.59E-02 | -3.63E-02 |
| 8090 | Cd302 | CD302 | -3.51E-02 | 9.32E-03 |
| 8091 | Scoc | SCOC | 2.19E-02 | -2.36E-02 |
| 8092 | Cct8 | CCT8 | 1.13E-02 | -4.59E-03 |
| 8093 | Tmem38a | TMEM38A | 2.19E-02 | -2.54E-02 |
| 8094 | Asb16 | ASB16 | -3.33E-02 | -1.19E-03 |
| 8095 | Rnf8 | RNF8 | -2.81E-02 | 1.93E-02 |
| 8096 | Caskin2 | CASKIN2 | -1.48E-02 | 1.78E-03 |
| 8097 | Slc16a12 | SLC16A12 | -2.32E-02 | -1.75E-02 |
| 8098 | Vipr2 | VIPR2 | 3.51E-02 | -1.78E-02 |
| 8099 | Rad51ap1 | RAD51AP1 | 6.95E-02 | 9.29E-02 |
| 8100 | Cacna2d1 | CACNA2D1 | 5.07E-02 | 6.09E-02 |
| 8101 | Poc1a | POC1A | 3.60E-02 | 7.42E-02 |
| 8102 | Mecp2 | MECP2 | -1.19E-02 | -1.75E-02 |
| 8103 | Best3 | BEST3 | 4.81E-02 | 2.99E-02 |
| 8104 | Mcmbp | MCMBP | -8.71E-03 | 8.30E-03 |
| 8105 | Pnmal1 | PNMAL1 | 1.83E-02 | -1.67E-02 |
| 8106 | Scn7a | SCN7A | -5.70E-02 | 9.22E-03 |

| | | | | |
|---|---|---|---|---|
| 8107 | Bcl6 | BCL6 | 8.76E-02 | 3.07E-02 |
| 8108 | Lrit2 | LRIT2 | 1.24E-02 | -1.78E-03 |
| 8109 | Mbl2 | MBL2 | -4.33E-02 | 3.99E-03 |
| 8110 | Add3 | ADD3 | -2.06E-02 | 4.64E-04 |
| 8111 | Krba1 | KRBA1 | 1.12E-02 | -2.94E-02 |
| 8112 | Mtmr6 | MTMR6 | 1.08E-02 | -1.85E-02 |
| 8113 | Mettl14 | METTL14 | -1.46E-02 | 1.35E-02 |
| 8114 | Tor4a | TOR4A | -2.82E-02 | 1.67E-02 |
| 8115 | Zmpste24 | ZMPSTE24 | 1.26E-02 | -9.11E-03 |
| 8116 | Pkp4 | PKP4 | 1.35E-02 | -3.28E-02 |
| 8117 | Cabin1 | CABIN1 | 9.69E-03 | -8.23E-03 |
| 8118 | Snx15 | SNX15 | -1.76E-02 | 1.39E-02 |
| 8119 | Grik5 | GRIK5 | -1.61E-02 | -3.64E-03 |
| 8120 | Arhgef2 | ARHGEF2 | 1.31E-02 | 1.58E-03 |
| 8121 | H2-DMb1 | HLA-DMB | -8.95E-02 | 1.73E-01 |
| 8122 | Trpa1 | TRPA1 | -1.63E-02 | 6.32E-03 |
| 8123 | Fam20a | FAM20A | -3.90E-02 | 1.14E-02 |
| 8124 | 9330159F19Rik | KIAA0408 | 1.86E-02 | -7.95E-03 |
| 8125 | Tcerg1 | TCERG1 | 1.09E-02 | 5.07E-03 |
| 8126 | Commd5 | COMMD5 | 1.39E-02 | -2.74E-02 |
| 8127 | Slc39a13 | SLC39A13 | 2.17E-02 | -3.19E-02 |
| 8128 | Dvl1 | DVL1 | 1.04E-02 | -6.59E-03 |
| 8129 | Tyw5 | TYW5 | 2.38E-02 | -2.92E-02 |
| 8130 | Styx | STYX | -2.39E-02 | 1.47E-02 |
| 8131 | Klhl26 | KLHL26 | 1.95E-02 | -1.99E-02 |
| 8132 | Fam3a | FAM3A | -1.06E-02 | -9.84E-03 |
| 8133 | Slc25a20 | SLC25A20 | 3.24E-02 | 2.07E-02 |
| 8134 | Scarf2 | SCARF2 | 3.54E-02 | 1.21E-02 |
| 8135 | Galnt10 | GALNT10 | -2.43E-02 | 2.51E-03 |
| 8136 | Sertad3 | SERTAD3 | 2.60E-02 | -3.65E-02 |
| 8137 | Irak3 | IRAK3 | 4.59E-02 | 3.44E-02 |
| 8138 | Cacnb4 | CACNB4 | 4.78E-02 | -4.28E-02 |
| 8139 | Fam193a | FAM193A | -1.09E-02 | 3.53E-03 |
| 8140 | Ctnna2 | CTNNA2 | 6.60E-02 | -1.04E-01 |
| 8141 | Tbc1d10a | TBC1D10A | -1.94E-02 | -3.63E-02 |
| 8142 | Sbno2 | SBNO2 | 1.96E-02 | 4.80E-02 |
| 8143 | Disc1 | DISC1 | -4.77E-02 | 1.31E-02 |
| 8144 | Apmap | APMAP | 1.54E-02 | -1.99E-02 |
| 8145 | Fbxo17 | FBXO17 | -4.00E-02 | 1.08E-03 |
| 8146 | Map4k3 | MAP4K3 | 9.73E-03 | -1.87E-02 |
| 8147 | Tapbp | TAPBP | -2.00E-02 | 2.05E-02 |
| 8148 | Plxnb3 | PLXNB3 | -3.71E-02 | 4.09E-02 |
| 8149 | Tirap | TIRAP | -2.60E-02 | 4.16E-02 |
| 8150 | Nars2 | NARS2 | 1.68E-02 | -1.98E-02 |
| 8151 | Abhd3 | ABHD3 | 2.24E-02 | -2.75E-02 |
| 8152 | Ccr3 | CCR3 | -1.82E-02 | 8.61E-03 |
| 8153 | Myom2 | MYOM2 | -5.46E-02 | -8.49E-03 |
| 8154 | Fam83d | FAM83D | 9.31E-02 | 1.49E-01 |
| 8155 | Iqce | IQCE | -1.55E-02 | -1.87E-02 |
| 8156 | Bsnd | BSND | -3.51E-02 | -3.37E-03 |
| 8157 | Slc35f5 | SLC35F5 | 7.95E-03 | 1.50E-04 |
| 8158 | Gba | GBA | 1.30E-02 | 8.08E-03 |
| 8159 | Ccdc136 | CCDC136 | -4.07E-02 | 4.84E-02 |

| | | | | |
|---|---|---|---|---|
| 8160 | Slc6a16 | SLC6A16 | 2.60E-02 | -1.48E-02 |
| 8161 | Mbd2 | MBD2 | 1.02E-02 | 1.77E-03 |
| 8162 | Abcg2 | ABCG2 | 2.13E-02 | -1.51E-02 |
| 8163 | Lrguk | LRGUK | 4.47E-02 | -1.41E-03 |
| 8164 | Srsf3 | SRSF3 | -1.41E-02 | 6.51E-03 |
| 8165 | St13 | ST13 | 1.54E-02 | -5.15E-03 |
| 8166 | Sh3gl1 | SH3GL1 | -1.34E-02 | 2.17E-02 |
| 8167 | Zdhhc13 | ZDHHC13 | 2.00E-02 | 2.86E-02 |
| 8168 | Vps13c | VPS13C | 1.72E-02 | -3.31E-02 |
| 8169 | Slc4a11 | SLC4A11 | -1.13E-01 | -1.30E-01 |
| 8170 | Hepacam2 | HEPACAM2 | -2.52E-02 | 4.94E-02 |
| 8171 | Sat2 | SAT2 | 2.89E-02 | -3.03E-02 |
| 8172 | Rpn1 | RPN1 | 1.16E-02 | -9.88E-03 |
| 8173 | Bloc1s3 | BLOC1S3 | 1.39E-02 | 3.82E-03 |
| 8174 | Agps | AGPS | -2.20E-02 | -3.72E-02 |
| 8175 | Slc43a3 | SLC43A3 | 2.96E-02 | 1.45E-02 |
| 8176 | Gin1 | GIN1 | -1.59E-02 | -3.31E-03 |
| 8177 | Art4 | ART4 | 6.37E-02 | 8.02E-02 |
| 8178 | Pfkfb2 | PFKFB2 | -1.76E-02 | -2.17E-02 |
| 8179 | Limk2 | LIMK2 | 1.52E-02 | -7.39E-03 |
| 8180 | Kctd20 | KCTD20 | 1.17E-02 | -4.80E-03 |
| 8181 | B3gnt2 | B3GNT2 | 3.20E-02 | -4.31E-03 |
| 8182 | Trim34b | TRIM6-TRIM34 | -3.48E-02 | 5.25E-02 |
| 8183 | Fbxo15 | FBXO15 | 1.65E-02 | -2.85E-02 |
| 8184 | Ndufaf7 | NDUFAF7 | 1.06E-02 | -2.53E-02 |
| 8185 | Nid2 | NID2 | 2.50E-02 | -2.70E-02 |
| 8186 | Shc4 | SHC4 | 4.10E-02 | 1.44E-03 |
| 8187 | Grip2 | GRIP2 | -3.86E-02 | -3.22E-02 |
| 8188 | Opa1 | OPA1 | 1.61E-02 | -2.25E-02 |
| 8189 | Cdk4 | CDK4 | -1.39E-02 | 8.10E-03 |
| 8190 | Cd200 | CD200 | -2.47E-02 | 1.30E-02 |
| 8191 | Sorcs3 | SORCS3 | -1.55E-01 | -7.22E-02 |
| 8192 | Mmp7 | MMP7 | 3.61E-02 | 1.42E-01 |
| 8193 | Pck2 | PCK2 | 1.94E-02 | 1.15E-02 |
| 8194 | Cbx1 | CBX1 | -1.16E-02 | 5.45E-03 |
| 8195 | Bcl9l | BCL9L | 1.66E-02 | 6.22E-03 |
| 8196 | Gnat2 | GNAT2 | -3.19E-02 | -2.96E-02 |
| 8197 | Prdm9 | PRDM9 | -1.74E-02 | -2.75E-03 |
| 8198 | Pak6 | PAK6 | 6.15E-02 | -4.07E-02 |
| 8199 | Pou2f1 | POU2F1 | -2.17E-02 | -3.70E-02 |
| 8200 | Nexn | NEXN | 3.93E-02 | 3.30E-02 |
| 8201 | Ctns | CTNS | -1.18E-02 | -2.89E-03 |
| 8202 | Pcnx | PCNX | 1.00E-02 | -2.23E-02 |
| 8203 | Fam227a | FAM227A | 3.40E-02 | -1.65E-02 |
| 8204 | Snx32 | SNX32 | -1.82E-02 | -9.05E-03 |
| 8205 | Gpsm1 | GPSM1 | 3.47E-02 | -8.74E-03 |
| 8206 | Gjc1 | GJC1 | 3.60E-02 | -7.89E-03 |
| 8207 | Tmprss5 | TMPRSS5 | 2.35E-02 | -4.89E-03 |
| 8208 | Thra | THRA | -1.58E-02 | -5.42E-02 |
| 8209 | Tmem132a | TMEM132A | -2.54E-02 | 5.76E-02 |
| 8210 | Trank1 | TRANK1 | 4.28E-02 | 1.17E-01 |
| 8211 | Znrf1 | ZNRF1 | 1.66E-02 | 2.80E-02 |
| 8212 | Srpx2 | SRPX2 | 7.43E-02 | -1.70E-02 |

| | | | | |
|---|---|---|---|---|
| 8213 | AI429214 | C8orf48 | -3.38E-02 | -2.32E-02 |
| 8214 | Srf | SRF | -1.60E-02 | 6.86E-03 |
| 8215 | Mbtd1 | MBTD1 | 1.89E-02 | -2.84E-02 |
| 8216 | Yif1b | YIF1B | 1.40E-02 | -1.24E-02 |
| 8217 | Gdap2 | GDAP2 | 1.32E-02 | 1.30E-02 |
| 8218 | Egr3 | EGR3 | 7.76E-02 | 9.16E-03 |
| 8219 | Naalad2 | NAALAD2 | -4.95E-02 | 4.85E-02 |
| 8220 | Fam166a | FAM166A | 2.82E-02 | -6.36E-02 |
| 8221 | Egfl7 | EGFL7 | 1.67E-02 | -1.23E-02 |
| 8222 | Pabpc4l | PABPC4L | -2.90E-02 | -5.87E-03 |
| 8223 | BC031181 | C18orf32 | 1.18E-02 | -6.57E-03 |
| 8224 | Gstp2 | GSTP1 | -5.25E-02 | 1.81E-02 |
| 8225 | Ccdc8 | CCDC8 | 5.58E-02 | -8.97E-03 |
| 8226 | Homer2 | HOMER2 | -8.98E-02 | 1.06E-01 |
| 8227 | Cutc | CUTC | -1.68E-02 | 2.16E-03 |
| 8228 | Terf1 | TERF1 | -2.44E-02 | 1.54E-03 |
| 8229 | H3f3a | H3F3A | 9.41E-03 | 5.21E-03 |
| 8230 | H3f3a | H3F3B | 9.41E-03 | 5.21E-03 |
| 8231 | Cav2 | CAV2 | 1.74E-02 | 3.61E-02 |
| 8232 | Tulp3 | TULP3 | -1.59E-02 | -1.46E-02 |
| 8233 | Celf4 | CELF4 | 4.38E-02 | 1.32E-02 |
| 8234 | Kdsr | KDSR | -1.43E-02 | 1.81E-03 |
| 8235 | Sh3tc1 | SH3TC1 | -1.97E-02 | 2.43E-02 |
| 8236 | Tmem242 | TMEM242 | 1.05E-02 | -1.21E-02 |
| 8237 | Cluh | CLUH | 1.56E-02 | -1.14E-02 |
| 8238 | Cyp4f13 | CYP4F3 | -1.71E-02 | -3.55E-02 |
| 8239 | Ankrd52 | ANKRD52 | 1.17E-02 | -1.09E-02 |
| 8240 | Cd2ap | CD2AP | 1.15E-02 | 8.62E-03 |
| 8241 | Hbb-bs | HBB | -6.29E-02 | 2.92E-02 |
| 8242 | Emc7 | EMC7 | 1.03E-02 | -5.78E-03 |
| 8243 | Slc2a6 | SLC2A6 | 1.07E-01 | 1.90E-01 |
| 8244 | Zmynd8 | ZMYND8 | 2.29E-02 | -1.71E-03 |
| 8245 | Ddx6 | DDX6 | -8.53E-03 | -1.26E-02 |
| 8246 | Nmnat1 | NMNAT1 | 1.74E-02 | -2.44E-02 |
| 8247 | Sult1e1 | SULT1E1 | 1.82E-02 | 3.46E-02 |
| 8248 | Ppp2r5a | PPP2R5A | -9.72E-03 | -1.43E-02 |
| 8249 | Commd1 | COMMD1 | 2.03E-02 | 5.91E-04 |
| 8250 | Heatr5b | HEATR5B | 1.23E-02 | 4.19E-03 |
| 8251 | 4930523C07Rik | KIAA0040 | 7.87E-02 | -1.05E-02 |
| 8252 | Casp4 | CASP4 | -5.27E-02 | 1.08E-01 |
| 8253 | Mmp8 | MMP8 | 7.55E-02 | 8.52E-02 |
| 8254 | Rab6b | RAB6B | 3.07E-02 | -3.84E-02 |
| 8255 | Tmem62 | TMEM62 | -2.10E-02 | 8.09E-04 |
| 8256 | Ilkap | ILKAP | 1.14E-02 | -1.95E-02 |
| 8257 | Zfp407 | ZNF407 | 1.87E-02 | -4.30E-02 |
| 8258 | Rac3 | RAC3 | -3.59E-02 | 1.06E-02 |
| 8259 | Mon1b | MON1B | -8.86E-03 | -8.92E-03 |
| 8260 | Arap2 | ARAP2 | 1.55E-02 | -3.47E-02 |
| 8261 | Kif24 | KIF24 | 5.81E-02 | 1.58E-02 |
| 8262 | Timm22 | TIMM22 | 1.33E-02 | 8.17E-03 |
| 8263 | Bcor | BCOR | -1.31E-02 | 2.20E-02 |
| 8264 | Hsp90ab1 | HSP90AB1 | -1.67E-02 | -7.63E-04 |
| 8265 | Aatf | AATF | 1.53E-02 | -1.84E-02 |

| | | | | |
|---|---|---|---|---|
| 8266 | Dnttip1 | DNTTIP1 | 9.97E-03 | -3.80E-03 |
| 8267 | Tmem91 | TMEM91 | 2.25E-02 | -3.06E-02 |
| 8268 | Clec4n | CLEC6A | 5.18E-02 | 2.12E-01 |
| 8269 | Synpo | SYNPO | -2.79E-02 | 5.80E-03 |
| 8270 | Dap3 | DAP3 | -1.03E-02 | -1.06E-02 |
| 8271 | Rhou | RHOU | 9.06E-02 | 1.61E-01 |
| 8272 | Fbxl8 | FBXL8 | -3.88E-02 | 1.26E-02 |
| 8273 | Ypel4 | YPEL4 | 4.17E-02 | 1.06E-01 |
| 8274 | Bbc3 | BBC3 | 4.78E-02 | 3.01E-02 |
| 8275 | Ubqln2 | UBQLN2 | -1.17E-02 | 9.43E-03 |
| 8276 | Nsmce4a | NSMCE4A | 1.02E-02 | -1.08E-02 |
| 8277 | Pnpla8 | PNPLA8 | 1.44E-02 | -1.61E-02 |
| 8278 | Nomo1 | NOMO2 | 1.10E-02 | -1.48E-02 |
| 8279 | Nomo1 | NOMO1 | 1.10E-02 | -1.48E-02 |
| 8280 | Nomo1 | NOMO3 | 1.10E-02 | -1.48E-02 |
| 8281 | Pex10 | PEX10 | 1.65E-02 | -3.27E-02 |
| 8282 | Carm1 | CARM1 | 1.04E-02 | -4.58E-04 |
| 8283 | Ldhd | LDHD | -1.73E-02 | -9.80E-03 |
| 8284 | Sncg | SNCG | -4.26E-02 | 1.06E-02 |
| 8285 | Ube4b | UBE4B | -1.61E-02 | -7.46E-03 |
| 8286 | Prrg1 | PRRG1 | -9.07E-02 | 1.75E-02 |
| 8287 | Pecr | PECR | -1.92E-02 | -7.82E-04 |
| 8288 | Urgcp | URGCP | 1.58E-02 | -1.01E-03 |
| 8289 | Msantd1 | MSANTD1 | -4.04E-02 | 4.94E-02 |
| 8290 | Ago3 | AGO3 | -1.68E-02 | 9.83E-03 |
| 8291 | Mfsd3 | MFSD3 | -2.62E-02 | -2.76E-02 |
| 8292 | Grik4 | GRIK4 | -2.25E-02 | -1.58E-02 |
| 8293 | Mbnl3 | MBNL3 | -4.73E-02 | 5.29E-02 |
| 8294 | Angel1 | ANGEL1 | 2.13E-02 | -1.92E-02 |
| 8295 | Fbxo30 | FBXO30 | -2.40E-02 | 1.41E-03 |
| 8296 | Ndc1 | NDC1 | 2.04E-02 | 1.72E-02 |
| 8297 | Ugt2b34 | UGT2B10 | 3.96E-02 | 3.86E-03 |
| 8298 | Pdf | PDF | 4.19E-02 | -5.36E-02 |
| 8299 | Mrps17 | MRPS17 | 1.09E-02 | -2.60E-03 |
| 8300 | Pogk | POGK | 1.46E-02 | -2.22E-03 |
| 8301 | Cdkl2 | CDKL2 | 2.14E-02 | -1.93E-02 |
| 8302 | Kcns3 | KCNS3 | -6.89E-02 | 5.34E-02 |
| 8303 | Jtb | JTB | 1.07E-02 | -7.41E-03 |
| 8304 | Cndp1 | CNDP1 | -3.87E-02 | -2.11E-02 |
| 8305 | Taok1 | TAOK1 | -1.19E-02 | -4.35E-03 |
| 8306 | Etv4 | ETV4 | 7.81E-02 | 1.13E-01 |
| 8307 | Cdh4 | CDH4 | 5.05E-02 | -1.46E-01 |
| 8308 | Gmip | GMIP | 1.46E-02 | 2.69E-02 |
| 8309 | Eif2d | EIF2D | 1.01E-02 | -2.34E-03 |
| 8310 | Tnfaip8l1 | TNFAIP8L1 | 5.44E-02 | 2.82E-02 |
| 8311 | Tmem216 | TMEM216 | -1.72E-02 | -7.70E-05 |
| 8312 | Actb | ACTB | 1.30E-02 | 5.48E-03 |
| 8313 | Ilf2 | ILF2 | 1.00E-02 | 7.66E-03 |
| 8314 | Ttll8 | TTLL8 | 1.98E-02 | 1.83E-02 |
| 8315 | Fmo3 | FMO3 | 8.61E-02 | -8.21E-03 |
| 8316 | Cnot3 | CNOT3 | -1.10E-02 | -3.05E-03 |
| 8317 | Pgrmc1 | PGRMC1 | 1.47E-02 | -9.65E-03 |
| 8318 | Anxa11 | ANXA11 | 1.07E-02 | -7.54E-03 |

| | | | | |
|---|---|---|---|---|
| 8319 | Ccnf | CCNF | 1.09E-01 | 1.73E-01 |
| 8320 | Dnah9 | DNAH9 | -1.93E-02 | 1.36E-02 |
| 8321 | Strn | STRN | 1.40E-02 | -1.84E-02 |
| 8322 | Sun3 | SUN3 | -3.99E-02 | -3.81E-02 |
| 8323 | Chst9 | CHST9 | -1.02E-01 | -7.24E-02 |
| 8324 | Nudt13 | NUDT13 | 1.19E-02 | -2.01E-02 |
| 8325 | Fancc | FANCC | -1.62E-02 | -2.22E-02 |
| 8326 | Nyx | NYX | -3.73E-02 | 3.03E-02 |
| 8327 | Tcn2 | TCN2 | -1.81E-02 | -1.17E-04 |
| 8328 | F8a | F8A2 | 2.01E-02 | 8.50E-03 |
| 8329 | F8a | F8A1 | 2.01E-02 | 8.50E-03 |
| 8330 | F8a | F8A3 | 2.01E-02 | 8.50E-03 |
| 8331 | Foxf2 | FOXF2 | -1.55E-02 | 1.61E-02 |
| 8332 | Edem3 | EDEM3 | 1.41E-02 | -3.44E-03 |
| 8333 | Hivep2 | HIVEP2 | -2.24E-02 | 6.64E-03 |
| 8334 | Camta1 | CAMTA1 | 2.07E-02 | 3.50E-02 |
| 8335 | Dhdds | DHDDS | 1.32E-02 | 1.95E-03 |
| 8336 | Polrmt | POLRMT | 1.30E-02 | -1.81E-02 |
| 8337 | Nol9 | NOL9 | 1.34E-02 | 8.52E-03 |
| 8338 | Sh2d6 | SH2D6 | 2.80E-02 | -1.21E-02 |
| 8339 | Cetn2 | CETN2 | -1.16E-02 | 3.10E-03 |
| 8340 | Lonrf3 | LONRF3 | -4.33E-02 | 9.76E-02 |
| 8341 | Fancg | FANCG | -2.31E-02 | 1.77E-02 |
| 8342 | Tbc1d10c | TBC1D10C | -3.98E-02 | 2.14E-02 |
| 8343 | Dnah3 | DNAH3 | -4.66E-02 | -1.30E-03 |
| 8344 | Gjb5 | GJB5 | 1.31E-01 | 1.02E-02 |
| 8345 | Abcf3 | ABCF3 | -8.94E-03 | -8.60E-03 |
| 8346 | Nr1h3 | NR1H3 | -1.79E-02 | -1.27E-02 |
| 8347 | Mtmr14 | MTMR14 | -1.30E-02 | -9.42E-03 |
| 8348 | Tmem132c | TMEM132C | -1.98E-02 | -1.15E-02 |
| 8349 | Stk40 | STK40 | 1.17E-02 | 2.18E-02 |
| 8350 | Rhpn1 | RHPN1 | 2.51E-02 | -1.45E-02 |
| 8351 | Mapkapk5 | MAPKAPK5 | 4.09E-02 | 1.49E-02 |
| 8352 | Plekhm3 | PLEKHM3 | 2.23E-02 | -6.43E-03 |
| 8353 | Mpp2 | MPP2 | -3.12E-02 | 1.88E-02 |
| 8354 | Asf1b | ASF1B | 8.53E-02 | 1.95E-01 |
| 8355 | Cldn4 | CLDN4 | 5.35E-02 | 8.70E-02 |
| 8356 | Gpr12 | GPR12 | -2.49E-02 | -1.68E-02 |
| 8357 | Ptprh | PTPRH | 1.31E-01 | -4.05E-02 |
| 8358 | Zfp647 | ZNF250 | -4.63E-02 | 6.52E-03 |
| 8359 | Ccdc85a | CCDC85A | 6.49E-02 | -5.88E-02 |
| 8360 | Lin7b | LIN7B | -4.60E-02 | 2.85E-02 |
| 8361 | Setbp1 | SETBP1 | -2.36E-02 | -3.03E-02 |
| 8362 | Mex3d | MEX3D | -3.70E-02 | 1.59E-02 |
| 8363 | Cyp4f15 | CYP4F8 | 1.31E-01 | 1.56E-01 |
| 8364 | Wnk4 | WNK4 | 1.25E-02 | -1.71E-02 |
| 8365 | Sel1l | SEL1L | 1.21E-02 | -1.61E-02 |
| 8366 | Ankrd33 | ANKRD33 | 1.85E-02 | 4.13E-03 |
| 8367 | Klk10 | KLK10 | 1.93E-02 | 1.55E-02 |
| 8368 | Tcf19 | TCF19 | 6.05E-02 | 1.08E-01 |
| 8369 | Stac3 | STAC3 | -4.00E-02 | 1.15E-03 |
| 8370 | Tnfrsf1a | TNFRSF1A | 1.22E-02 | 2.11E-02 |
| 8371 | Mrpl35 | MRPL35 | 1.49E-02 | -1.93E-02 |

| | | | | |
|---|---|---|---|---|
| 8372 | Drp2 | DRP2 | 2.80E-02 | 2.81E-04 |
| 8373 | Ehd4 | EHD4 | 2.96E-02 | 1.29E-02 |
| 8374 | Cox7a2l | COX7A2L | -1.03E-02 | 4.12E-03 |
| 8375 | Rabggtb | RABGGTB | 1.10E-02 | 8.62E-03 |
| 8376 | Serpini1 | SERPINI1 | -3.49E-02 | 1.09E-01 |
| 8377 | Myl6 | MYL6 | 1.01E-02 | -6.07E-03 |
| 8378 | Fkbp4 | FKBP4 | -1.62E-02 | -1.07E-02 |
| 8379 | Cacna1a | CACNA1A | 3.72E-02 | -2.84E-02 |
| 8380 | Phf21b | PHF21B | 3.25E-02 | -6.22E-03 |
| 8381 | Best1 | BEST1 | 3.96E-02 | -2.39E-02 |
| 8382 | Ahctf1 | AHCTF1 | -1.34E-02 | 9.20E-03 |
| 8383 | Trappc4 | TRAPPC4 | -1.28E-02 | -1.84E-02 |
| 8384 | 1700010I14Rik | C6orf118 | 3.69E-02 | -1.12E-02 |
| 8385 | 2310061I04Rik | C6orf136 | 1.13E-02 | -6.85E-03 |
| 8386 | Blvrb | BLVRB | 2.84E-02 | -8.94E-03 |
| 8387 | Mgst2 | MGST2 | -5.18E-02 | 6.00E-02 |
| 8388 | Tk2 | TK2 | 1.68E-02 | -3.38E-02 |
| 8389 | Rhot2 | RHOT2 | 9.83E-03 | -8.75E-03 |
| 8390 | St8sia5 | ST8SIA5 | -1.66E-02 | 2.38E-03 |
| 8391 | Iars2 | IARS2 | 1.38E-02 | -1.67E-02 |
| 8392 | Ptges3 | PTGES3 | -1.79E-02 | 1.86E-02 |
| 8393 | Tbc1d23 | TBC1D23 | 1.18E-02 | -1.36E-02 |
| 8394 | Dph2 | DPH2 | -2.11E-02 | 2.26E-02 |
| 8395 | Usp30 | USP30 | 1.11E-02 | -2.15E-02 |
| 8396 | Vps33a | VPS33A | 1.13E-02 | -2.37E-02 |
| 8397 | Tlr5 | TLR5 | -4.74E-02 | 5.00E-03 |
| 8398 | Mapt | MAPT | 2.31E-02 | -2.20E-02 |
| 8399 | Nacc2 | NACC2 | 1.47E-02 | -3.01E-02 |
| 8400 | Plxna3 | PLXNA3 | -6.54E-02 | -3.04E-03 |
| 8401 | Slc27a6 | SLC27A6 | -1.72E-02 | 3.69E-03 |
| 8402 | Tulp4 | TULP4 | 1.70E-02 | -2.67E-02 |
| 8403 | Gsn | GSN | 2.04E-02 | 1.49E-02 |
| 8404 | Smc6 | SMC6 | 1.09E-02 | -8.87E-03 |
| 8405 | Gpr21 | GPR21 | -3.04E-02 | 5.56E-03 |
| 8406 | Vps13b | VPS13B | 1.13E-02 | -2.51E-02 |
| 8407 | Sfrp1 | SFRP1 | -2.39E-02 | -9.87E-03 |
| 8408 | Irf2 | IRF2 | -1.39E-02 | 4.75E-03 |
| 8409 | Srfbp1 | SRFBP1 | -1.92E-02 | 1.52E-02 |
| 8410 | Prrc2b | PRRC2B | 1.23E-02 | -1.70E-02 |
| 8411 | Chst2 | CHST2 | -6.00E-02 | -3.52E-02 |
| 8412 | Fam160b1 | FAM160B1 | 1.07E-02 | -8.03E-03 |
| 8413 | Grk1 | GRK1 | 2.46E-02 | -2.13E-02 |
| 8414 | Prrc2c | PRRC2C | -9.68E-03 | -3.50E-03 |
| 8415 | Trmt13 | TRMT13 | -1.81E-02 | 2.53E-02 |
| 8416 | Mat2b | MAT2B | 1.14E-02 | -1.84E-02 |
| 8417 | Chn2 | CHN2 | -2.47E-02 | -4.12E-02 |
| 8418 | Plekhh2 | PLEKHH2 | 2.55E-02 | -4.19E-02 |
| 8419 | Lztr1 | LZTR1 | -1.32E-02 | -7.50E-03 |
| 8420 | Atg16l1 | ATG16L1 | 1.28E-02 | -1.11E-03 |
| 8421 | Epor | EPOR | 9.76E-02 | -5.45E-02 |
| 8422 | Samd12 | SAMD12 | -5.32E-02 | -8.28E-03 |
| 8423 | Pi15 | PI15 | -5.69E-02 | 9.93E-02 |
| 8424 | Lrfn4 | LRFN4 | -4.96E-02 | 6.25E-02 |

| | | | | |
|---|---|---|---|---|
| 8425 | Phkg2 | PHKG2 | 1.08E-02 | -9.08E-03 |
| 8426 | Rasal2 | RASAL2 | -1.95E-02 | -2.48E-02 |
| 8427 | Chrna7 | CHRNA7 | 4.99E-02 | -4.67E-02 |
| 8428 | Ctsf | CTSF | -1.24E-02 | -2.80E-02 |
| 8429 | Echs1 | ECHS1 | -1.37E-02 | -1.36E-02 |
| 8430 | Akr1b8 | AKR1B10 | 9.58E-02 | 2.39E-01 |
| 8431 | P4htm | P4HTM | -3.11E-02 | -2.16E-02 |
| 8432 | Fbxo11 | FBXO11 | 1.18E-02 | -1.40E-02 |
| 8433 | Far2 | FAR2 | -1.59E-02 | 2.42E-03 |
| 8434 | Pom121 | POM121C | 1.24E-02 | 1.45E-03 |
| 8435 | Pom121 | POM121 | 1.24E-02 | 1.45E-03 |
| 8436 | Txnrd1 | TXNRD1 | 2.17E-02 | 1.88E-02 |
| 8437 | 2310011J03Rik | C19orf25 | 1.16E-02 | -2.52E-03 |
| 8438 | Cep164 | CEP164 | 1.05E-02 | -2.25E-02 |
| 8439 | Bbs9 | BBS9 | 1.52E-02 | -2.90E-02 |
| 8440 | Mrps5 | MRPS5 | -1.14E-02 | -2.45E-02 |
| 8441 | Baiap2l1 | BAIAP2L1 | -1.48E-02 | -1.40E-02 |
| 8442 | Abcg8 | ABCG8 | -3.21E-02 | 1.21E-02 |
| 8443 | Dmtn | DMTN | -1.38E-02 | -7.98E-03 |
| 8444 | Zdhhc8 | ZDHHC8 | 1.97E-02 | -1.23E-02 |
| 8445 | Slc7a2 | SLC7A2 | -5.14E-02 | -5.33E-02 |
| 8446 | Iqsec1 | IQSEC1 | -1.37E-02 | -3.11E-02 |
| 8447 | Rusc1 | RUSC1 | 2.60E-02 | 7.20E-02 |
| 8448 | Evc2 | EVC2 | -1.48E-02 | -5.87E-03 |
| 8449 | Aqp4 | AQP4 | -8.12E-02 | -5.18E-02 |
| 8450 | Ano1 | ANO1 | -5.28E-02 | 1.08E-01 |
| 8451 | Zfp551 | ZNF551 | -3.95E-02 | -3.42E-02 |
| 8452 | Casc1 | CASC1 | 3.58E-02 | -4.01E-02 |
| 8453 | Plk2 | PLK2 | 4.98E-02 | -3.22E-02 |
| 8454 | Sim1 | SIM1 | -5.38E-02 | -5.60E-02 |
| 8455 | Tmprss9 | TMPRSS9 | 2.70E-02 | 5.08E-03 |
| 8456 | Wdr77 | WDR77 | -1.47E-02 | 1.54E-02 |
| 8457 | Scfd2 | SCFD2 | 1.84E-02 | 1.09E-02 |
| 8458 | Lgals7 | LGALS7B | 1.62E-02 | -1.34E-02 |
| 8459 | Lgals7 | LGALS7 | 1.62E-02 | -1.34E-02 |
| 8460 | Gpr89 | GPR89A | -1.31E-02 | 3.21E-03 |
| 8461 | Gpr89 | GPR89B | -1.31E-02 | 3.21E-03 |
| 8462 | Fhl3 | FHL3 | 4.49E-02 | -4.26E-03 |
| 8463 | H3f3c | HIST2H3C | -2.00E-02 | 1.02E-02 |
| 8464 | Mterf1a | MTERF | -3.38E-02 | -4.31E-02 |
| 8465 | Tfdp1 | TFDP1 | 1.60E-02 | 1.89E-02 |
| 8466 | Nfil3 | NFIL3 | -5.80E-02 | 1.60E-01 |
| 8467 | Gm2a | GM2A | 1.66E-02 | -8.67E-03 |
| 8468 | Calcrl | CALCRL | -3.69E-02 | 9.43E-03 |
| 8469 | Csf1 | CSF1 | -3.19E-02 | 9.94E-02 |
| 8470 | Rest | REST | 1.40E-02 | -1.33E-02 |
| 8471 | Ppp2r3c | PPP2R3C | 1.35E-02 | 9.02E-03 |
| 8472 | Gm12657 | H3F3C | 3.44E-02 | -1.25E-02 |
| 8473 | Cd80 | CD80 | -2.57E-02 | 3.61E-02 |
| 8474 | Pdgfrb | PDGFRB | -3.61E-02 | 2.39E-02 |
| 8475 | Gk2 | GK2 | -2.44E-02 | 2.84E-02 |
| 8476 | Sart1 | SART1 | -8.74E-03 | -9.57E-03 |
| 8477 | B3galt2 | B3GALT2 | -6.64E-02 | 1.25E-01 |

| | | | | |
|---|---|---|---|---|
| 8478 | Fbxw4 | FBXW4 | 1.92E-02 | 4.14E-03 |
| 8479 | Dcaf4 | DCAF4 | -2.32E-02 | -5.29E-02 |
| 8480 | Cacng5 | CACNG5 | 1.21E-02 | 2.25E-03 |
| 8481 | Pip4k2a | PIP4K2A | -2.06E-02 | 2.23E-02 |
| 8482 | Gtpbp8 | GTPBP8 | 1.23E-02 | -2.71E-02 |
| 8483 | Traf2 | TRAF2 | 1.94E-02 | 2.03E-02 |
| 8484 | Tmem101 | TMEM101 | -1.84E-02 | -2.69E-03 |
| 8485 | Zfhx2 | ZFHX2 | 1.68E-02 | -1.60E-02 |
| 8486 | Fkbp7 | FKBP7 | 3.78E-02 | -2.31E-02 |
| 8487 | Dicer1 | DICER1 | 9.03E-03 | -1.35E-02 |
| 8488 | Ccdc113 | CCDC113 | -2.81E-02 | -2.11E-02 |
| 8489 | Arhgap24 | ARHGAP24 | -1.24E-02 | -2.23E-02 |
| 8490 | Pcdhb21 | PCDHB11 | -2.89E-02 | 3.17E-02 |
| 8491 | Mief2 | MIEF2 | 2.16E-02 | -1.88E-02 |
| 8492 | Hrh3 | HRH3 | 4.03E-02 | 1.14E-02 |
| 8493 | Agtr1a | AGTR1 | -2.88E-02 | -3.75E-02 |
| 8494 | Zc3h12c | ZC3H12C | -4.19E-02 | -1.45E-02 |
| 8495 | Cped1 | CPED1 | -4.49E-02 | -1.62E-02 |
| 8496 | Rimbp3 | RIMBP3C | -2.28E-02 | -4.25E-03 |
| 8497 | Rimbp3 | RIMBP3 | -2.28E-02 | -4.25E-03 |
| 8498 | Rimbp3 | RIMBP3B | -2.28E-02 | -4.25E-03 |
| 8499 | Rbp1 | RBP1 | -3.54E-02 | 3.26E-02 |
| 8500 | Rspo1 | RSPO1 | 2.59E-02 | 6.87E-03 |
| 8501 | Klf7 | KLF7 | -1.99E-02 | -2.00E-02 |
| 8502 | Synrg | SYNRG | 1.31E-02 | -1.10E-02 |
| 8503 | Pts | PTS | 8.80E-03 | -6.37E-03 |
| 8504 | Rpusd1 | RPUSD1 | 2.21E-02 | 3.47E-04 |
| 8505 | Cnih1 | CNIH1 | 1.10E-02 | 7.34E-03 |
| 8506 | Tmem229a | TMEM229A | -1.70E-02 | -1.19E-02 |
| 8507 | Chrne | CHRNE | -3.75E-02 | -3.78E-02 |
| 8508 | Eif2ak2 | EIF2AK2 | 1.43E-02 | -8.79E-03 |
| 8509 | Hvcn1 | HVCN1 | -4.25E-02 | -5.86E-02 |
| 8510 | Lrp1 | LRP1 | 3.90E-02 | 1.91E-03 |
| 8511 | 6430571L13Rik | C3orf18 | -3.86E-02 | -3.87E-02 |
| 8512 | Slc1a3 | SLC1A3 | -5.61E-02 | -3.53E-02 |
| 8513 | Tsen34 | TSEN34 | 9.88E-03 | -8.06E-03 |
| 8514 | Phrf1 | PHRF1 | -1.09E-02 | 1.23E-02 |
| 8515 | Wdr81 | WDR81 | 1.71E-02 | -9.85E-03 |
| 8516 | Lats1 | LATS1 | -1.27E-02 | -5.30E-03 |
| 8517 | Pola2 | POLA2 | 2.52E-02 | 2.03E-02 |
| 8518 | Dusp23 | DUSP23 | -1.89E-02 | -3.19E-02 |
| 8519 | B3gnt7 | B3GNT7 | -6.78E-02 | 3.36E-02 |
| 8520 | Slc26a1 | SLC26A1 | 3.37E-02 | -4.34E-02 |
| 8521 | Scarb2 | SCARB2 | 1.28E-02 | 5.99E-03 |
| 8522 | Lysmd1 | LYSMD1 | 2.90E-02 | -1.97E-02 |
| 8523 | Nsun6 | NSUN6 | -2.21E-02 | -2.40E-02 |
| 8524 | Osr1 | OSR1 | -4.19E-02 | -2.10E-03 |
| 8525 | Traf1 | TRAF1 | -3.46E-02 | 4.13E-02 |
| 8526 | Ctdp1 | CTDP1 | 1.60E-02 | 1.78E-02 |
| 8527 | Tomm7 | TOMM7 | -1.26E-02 | -1.56E-02 |
| 8528 | Chek1 | CHEK1 | 6.04E-02 | 1.89E-01 |
| 8529 | Zxdb | ZXDB | -3.53E-02 | -1.86E-03 |
| 8530 | B3gat1 | B3GAT1 | -1.53E-02 | -1.18E-02 |

| | | | | |
|---|---|---|---|---|
| 8531 | Vcam1 | VCAM1 | -4.92E-02 | 3.03E-01 |
| 8532 | Rrm1 | RRM1 | 2.01E-02 | 3.01E-02 |
| 8533 | Timp3 | TIMP3 | -1.78E-02 | -2.49E-02 |
| 8534 | Lmo7 | LMO7 | -1.50E-02 | -4.04E-02 |
| 8535 | Cpsf3 | CPSF3 | -1.06E-02 | -1.01E-02 |
| 8536 | Pomt1 | POMT1 | -1.44E-02 | -1.15E-02 |
| 8537 | Trerf1 | TRERF1 | -2.59E-02 | 9.64E-04 |
| 8538 | Nlrp6 | NLRP6 | 2.12E-02 | -4.18E-02 |
| 8539 | Tmem165 | TMEM165 | 1.25E-02 | 1.53E-02 |
| 8540 | Bcl3 | BCL3 | -6.23E-02 | 2.09E-01 |
| 8541 | Crk | CRK | 9.77E-03 | -1.24E-03 |
| 8542 | Lrrc73 | LRRC73 | 4.12E-02 | 3.69E-02 |
| 8543 | Ift172 | IFT172 | -1.18E-02 | -1.84E-02 |
| 8544 | Lypd3 | LYPD3 | 5.15E-02 | 8.73E-02 |
| 8545 | Shcbp1l | SHCBP1L | -1.44E-02 | 2.40E-02 |
| 8546 | Aimp1 | AIMP1 | -1.10E-02 | 7.87E-03 |
| 8547 | Galnt4 | GALNT4 | -1.68E-02 | 1.32E-03 |
| 8548 | Mier1 | MIER1 | 1.33E-02 | -7.85E-03 |
| 8549 | Zfp473 | ZNF473 | -4.22E-02 | 9.94E-03 |
| 8550 | Map3k2 | MAP3K2 | -1.08E-02 | 1.73E-02 |
| 8551 | Mrpl10 | MRPL10 | 1.12E-02 | -2.28E-02 |
| 8552 | Nfyc | NFYC | -1.02E-02 | -1.88E-02 |
| 8553 | Ipp | IPP | 1.73E-02 | -1.59E-02 |
| 8554 | Nlgn3 | NLGN3 | -3.12E-02 | 3.64E-02 |
| 8555 | Asb7 | ASB7 | 1.55E-02 | -1.87E-02 |
| 8556 | Iqub | IQUB | -1.93E-02 | -3.85E-03 |
| 8557 | Slc35d1 | SLC35D1 | 1.17E-02 | -1.37E-02 |
| 8558 | Mllt11 | MLLT11 | 1.76E-02 | 1.23E-02 |
| 8559 | Txndc9 | TXNDC9 | 9.63E-03 | -1.18E-02 |
| 8560 | Lsamp | LSAMP | -4.77E-02 | 8.06E-02 |
| 8561 | Ccp110 | CCP110 | -1.55E-02 | 3.15E-04 |
| 8562 | Tmem127 | TMEM127 | 7.18E-03 | -1.37E-02 |
| 8563 | Tmem33 | TMEM33 | -1.23E-02 | -5.38E-03 |
| 8564 | Kctd10 | KCTD10 | 1.05E-02 | 1.70E-02 |
| 8565 | Ccdc146 | CCDC146 | -1.58E-02 | 1.21E-02 |
| 8566 | Lsm14b | LSM14B | 9.48E-03 | -4.68E-03 |
| 8567 | Suv39h1 | SUV39H1 | 2.06E-02 | 2.12E-02 |
| 8568 | Mcf2l | MCF2L | 1.64E-02 | -4.68E-02 |
| 8569 | Parvg | PARVG | -3.85E-02 | 5.52E-02 |
| 8570 | Ddx47 | DDX47 | -6.60E-03 | -8.21E-03 |
| 8571 | Tbc1d22b | TBC1D22B | 1.49E-02 | -1.50E-02 |
| 8572 | Upf1 | UPF1 | -9.16E-03 | -1.28E-03 |
| 8573 | Ppp1r14c | PPP1R14C | -3.24E-02 | 6.83E-02 |
| 8574 | Lsm11 | LSM11 | -2.54E-02 | 2.38E-02 |
| 8575 | Cdk17 | CDK17 | -2.68E-02 | 2.67E-02 |
| 8576 | Ppil4 | PPIL4 | 1.17E-02 | 1.18E-03 |
| 8577 | Usp27x | USP27X | -2.75E-02 | 5.01E-02 |
| 8578 | Fmo4 | FMO4 | 2.86E-02 | -1.28E-02 |
| 8579 | Gosr1 | GOSR1 | 7.83E-03 | -1.77E-02 |
| 8580 | Zfp235 | ZNF235 | -2.58E-02 | -1.17E-02 |
| 8581 | Nufip2 | NUFIP2 | -1.98E-02 | 1.23E-02 |
| 8582 | Tmem65 | TMEM65 | 1.47E-02 | -1.06E-02 |
| 8583 | Sh3rf2 | SH3RF2 | -2.98E-02 | 7.56E-02 |

| | | | | |
|---|---|---|---|---|
| 8584 | Eps8l1 | EPS8L1 | 7.71E-02 | -4.33E-02 |
| 8585 | Gfra2 | GFRA2 | -3.39E-02 | 9.96E-02 |
| 8586 | Slc25a26 | SLC25A26 | -1.59E-02 | -1.91E-02 |
| 8587 | Plekhh3 | PLEKHH3 | 1.82E-02 | 2.17E-03 |
| 8588 | Doc2b | DOC2B | -4.72E-02 | -1.65E-02 |
| 8589 | Dnajc27 | DNAJC27 | 2.20E-02 | 2.55E-02 |
| 8590 | Adcy4 | ADCY4 | -3.21E-02 | -3.42E-03 |
| 8591 | Lactb | LACTB | 1.31E-02 | -6.42E-03 |
| 8592 | Ebf4 | EBF4 | 4.03E-02 | 1.77E-02 |
| 8593 | Bub1 | BUB1 | 9.09E-02 | 1.73E-01 |
| 8594 | Arg2 | ARG2 | -5.26E-02 | -2.45E-02 |
| 8595 | Mylk2 | MYLK2 | 2.53E-02 | 3.69E-02 |
| 8596 | Zfp260 | ZNF260 | -1.27E-02 | 6.14E-03 |
| 8597 | Esr1 | ESR1 | -4.30E-02 | -5.79E-02 |
| 8598 | Srgap1 | SRGAP1 | 3.14E-02 | 2.87E-02 |
| 8599 | Uqcc1 | UQCC1 | -1.29E-02 | -1.91E-02 |
| 8600 | Hykk | HYKK | -2.07E-02 | -6.23E-03 |
| 8601 | Fam71b | FAM71B | -3.29E-02 | 6.49E-02 |
| 8602 | Amz1 | AMZ1 | 4.11E-02 | -7.69E-02 |
| 8603 | Fam53b | FAM53B | 1.56E-02 | -6.42E-03 |
| 8604 | Vrk3 | VRK3 | -1.00E-02 | -3.91E-03 |
| 8605 | C1qtnf2 | C1QTNF2 | -5.52E-02 | -2.08E-02 |
| 8606 | Sstr4 | SSTR4 | -2.13E-02 | 1.49E-02 |
| 8607 | Vash2 | VASH2 | 4.18E-02 | 9.02E-03 |
| 8608 | Csf2ra | CSF2RA | -2.72E-02 | 3.77E-02 |
| 8609 | Mknk1 | MKNK1 | -1.09E-02 | 1.29E-02 |
| 8610 | Padi2 | PADI2 | 3.90E-02 | 5.98E-02 |
| 8611 | Fbxo21 | FBXO21 | 1.97E-02 | -4.44E-02 |
| 8612 | Plagl2 | PLAGL2 | -1.87E-02 | 1.48E-02 |
| 8613 | Ctnnb1 | CTNNB1 | -9.17E-03 | -2.49E-03 |
| 8614 | Pkn3 | PKN3 | -2.83E-02 | 1.11E-01 |
| 8615 | Ranbp3l | RANBP3L | 7.82E-02 | -8.37E-02 |
| 8616 | Ankfn1 | ANKFN1 | 7.38E-02 | 3.18E-02 |
| 8617 | Gsap | GSAP | -1.92E-02 | 1.52E-02 |
| 8618 | Cul4b | CUL4B | 1.15E-02 | -5.95E-03 |
| 8619 | Tmem52 | TMEM52 | 3.96E-02 | 3.31E-02 |
| 8620 | Pgbd5 | PGBD5 | 4.55E-02 | 7.19E-02 |
| 8621 | Gnpnat1 | GNPNAT1 | -1.34E-02 | 1.33E-02 |
| 8622 | Cdc123 | CDC123 | 1.07E-02 | 1.12E-03 |
| 8623 | Pepd | PEPD | 1.54E-02 | -1.66E-02 |
| 8624 | Arid5a | ARID5A | 4.93E-02 | 1.04E-01 |
| 8625 | Pkd1l3 | PKD1L3 | -2.40E-02 | 2.89E-02 |
| 8626 | Klhl20 | KLHL20 | 1.30E-02 | -2.16E-02 |
| 8627 | Uap1 | UAP1 | -1.77E-02 | 1.78E-02 |
| 8628 | Mlkl | MLKL | -4.57E-02 | 7.11E-02 |
| 8629 | Sh3glb2 | SH3GLB2 | 1.21E-02 | -1.39E-02 |
| 8630 | Chrm1 | CHRM1 | -3.16E-02 | 2.12E-04 |
| 8631 | 1110038F14Rik | C8orf33 | -1.72E-02 | 9.86E-03 |
| 8632 | Ccr9 | CCR9 | -2.38E-02 | -2.71E-02 |
| 8633 | Smpdl3a | SMPDL3A | 1.72E-02 | -6.08E-03 |
| 8634 | AI593442 | C11orf87 | -1.95E-02 | 2.34E-02 |
| 8635 | Tmx4 | TMX4 | 1.18E-02 | 2.74E-03 |
| 8636 | Ppp1r42 | PPP1R42 | 2.65E-02 | 2.39E-02 |

| | | | | |
|---|---|---|---|---|
| 8637 | Egflam | EGFLAM | -5.19E-02 | 1.64E-02 |
| 8638 | Sez6l | SEZ6L | -2.85E-02 | 4.67E-03 |
| 8639 | Heatr5a | HEATR5A | 1.74E-02 | -9.17E-03 |
| 8640 | Stxbp5l | STXBP5L | -4.46E-02 | -6.32E-02 |
| 8641 | Ppp1r3b | PPP1R3B | -4.13E-02 | 9.77E-02 |
| 8642 | Zkscan5 | ZKSCAN5 | -1.31E-02 | -1.31E-02 |
| 8643 | Foxd3 | FOXD3 | -1.63E-02 | 1.76E-02 |
| 8644 | H2-Bl | HLA-A | -3.04E-02 | -5.32E-02 |
| 8645 | Leprotl1 | LEPROTL1 | 1.10E-02 | -3.32E-03 |
| 8646 | Ccdc103 | CCDC103 | -3.28E-02 | -1.40E-02 |
| 8647 | Wasf3 | WASF3 | -5.14E-02 | -1.25E-01 |
| 8648 | Fer1l6 | FER1L6 | 4.47E-02 | -2.57E-02 |
| 8649 | C2cd4c | C2CD4C | -1.62E-02 | 1.48E-02 |
| 8650 | Il1r1 | IL1R1 | 2.76E-02 | 1.43E-02 |
| 8651 | Dnajc13 | DNAJC13 | 2.04E-02 | -2.32E-02 |
| 8652 | Mbnl2 | MBNL2 | 1.35E-02 | -4.88E-03 |
| 8653 | 9130008F23Rik | C6orf141 | -7.68E-02 | 5.59E-02 |
| 8654 | Zc3h4 | ZC3H4 | -1.04E-02 | -5.82E-03 |
| 8655 | Ap1s3 | AP1S3 | 2.35E-02 | 6.14E-03 |
| 8656 | Sdsl | SDSL | 2.20E-02 | 1.38E-03 |
| 8657 | Plcd3 | PLCD3 | -2.45E-02 | 1.31E-02 |
| 8658 | Mybph | MYBPH | 2.15E-02 | -1.10E-02 |
| 8659 | Ccdc43 | CCDC43 | -1.29E-02 | -7.41E-03 |
| 8660 | Kcnd1 | KCND1 | -4.05E-02 | 2.79E-02 |
| 8661 | Hspa1l | HSPA1L | 2.98E-02 | -1.28E-02 |
| 8662 | Apobec3 | APOBEC3B | -3.92E-02 | 2.12E-02 |
| 8663 | Taf9 | TAF9 | -1.58E-02 | 9.85E-03 |
| 8664 | Nme6 | NME6 | -1.55E-02 | -9.84E-03 |
| 8665 | Rad23a | RAD23A | 1.23E-02 | -1.31E-02 |
| 8666 | Fam168a | FAM168A | 1.18E-02 | -1.65E-02 |
| 8667 | Fam3c | FAM3C | 2.01E-02 | 2.75E-02 |
| 8668 | Ppara | PPARA | 4.23E-02 | -3.28E-02 |
| 8669 | Ctxn1 | CTXN1 | 3.99E-02 | 1.48E-01 |
| 8670 | Bcl9 | BCL9 | 2.22E-02 | -3.27E-02 |
| 8671 | Dusp22 | DUSP22 | 1.53E-02 | 1.12E-02 |
| 8672 | Man2b1 | MAN2B1 | 1.72E-02 | -4.36E-03 |
| 8673 | Carhsp1 | CARHSP1 | 1.05E-02 | 1.26E-02 |
| 8674 | Zc3hc1 | ZC3HC1 | 1.43E-02 | -2.81E-02 |
| 8675 | Cdkn2aip | CDKN2AIP | 1.55E-02 | -3.43E-02 |
| 8676 | Aqp6 | AQP6 | 5.56E-02 | -4.07E-02 |
| 8677 | Pcdh20 | PCDH20 | 1.35E-02 | 2.55E-02 |
| 8678 | Col22a1 | COL22A1 | -1.43E-02 | 1.11E-02 |
| 8679 | Ifi30 | IFI30 | 2.15E-02 | 1.70E-02 |
| 8680 | Med20 | MED20 | -1.59E-02 | 1.07E-02 |
| 8681 | Nudt15 | NUDT15 | 3.77E-02 | 4.57E-03 |
| 8682 | Cables2 | CABLES2 | -1.50E-02 | 1.00E-02 |
| 8683 | Pde6d | PDE6D | 1.40E-02 | 1.13E-02 |
| 8684 | Cxcl16 | CXCL16 | 2.59E-02 | 5.36E-02 |
| 8685 | Ndrg2 | NDRG2 | -1.78E-02 | -1.76E-03 |
| 8686 | Upk1b | UPK1B | 2.64E-01 | 1.74E-01 |
| 8687 | Tmem80 | TMEM80 | -1.51E-02 | -4.36E-03 |
| 8688 | Dach1 | DACH1 | -2.18E-02 | -2.43E-02 |
| 8689 | Ankrd11 | ANKRD11 | 1.05E-02 | 1.23E-03 |

| | | | | |
|---|---|---|---|---|
| 8690 | Ssr4 | SSR4 | 1.09E-02 | 9.09E-03 |
| 8691 | Myo1a | MYO1A | -2.65E-02 | -9.68E-03 |
| 8692 | Appbp2 | APPBP2 | -8.54E-03 | -4.37E-03 |
| 8693 | Tmc7 | TMC7 | 3.58E-02 | 1.16E-02 |
| 8694 | Tspo2 | TSPO2 | -1.40E-02 | 2.97E-02 |
| 8695 | Ajuba | AJUBA | -3.66E-02 | 3.86E-02 |
| 8696 | Smtn | SMTN | 1.40E-02 | -2.12E-02 |
| 8697 | Kif20b | KIF20B | 5.45E-02 | 1.21E-03 |
| 8698 | Lsm7 | LSM7 | -1.91E-02 | 1.36E-02 |
| 8699 | Col1a2 | COL1A2 | 5.83E-02 | 9.80E-02 |
| 8700 | Mrps33 | MRPS33 | 1.60E-02 | -2.07E-02 |
| 8701 | Zbtb41 | ZBTB41 | -1.53E-02 | 3.35E-03 |
| 8702 | Zyg11b | ZYG11B | 1.11E-02 | -2.32E-02 |
| 8703 | Slc9a3r1 | SLC9A3R1 | -1.54E-02 | 2.37E-03 |
| 8704 | Kbtbd11 | KBTBD11 | 2.55E-02 | 2.36E-04 |
| 8705 | Ctdspl2 | CTDSPL2 | -2.11E-02 | -1.01E-02 |
| 8706 | Lime1 | LIME1 | -2.39E-02 | 1.19E-02 |
| 8707 | Slc4a5 | SLC4A5 | -7.57E-02 | -4.71E-02 |
| 8708 | Zfyve16 | ZFYVE16 | -1.43E-02 | -5.97E-03 |
| 8709 | Dck | DCK | 3.96E-02 | 5.62E-02 |
| 8710 | Rictor | RICTOR | -1.45E-02 | -2.28E-02 |
| 8711 | Ppfia2 | PPFIA2 | -3.07E-02 | 5.36E-03 |
| 8712 | Slc25a32 | SLC25A32 | -2.47E-02 | -2.47E-02 |
| 8713 | Acad11 | ACAD11 | 2.06E-02 | -2.21E-02 |
| 8714 | Inpp5d | INPP5D | -2.37E-02 | 6.44E-02 |
| 8715 | Acoxl | ACOXL | -2.51E-02 | 4.29E-03 |
| 8716 | Ccdc166 | CCDC166 | -3.90E-02 | 8.68E-03 |
| 8717 | Nup107 | NUP107 | 2.19E-02 | -2.09E-02 |
| 8718 | Ercc6 | ERCC6 | -1.32E-02 | -3.76E-03 |
| 8719 | Susd2 | SUSD2 | -2.48E-02 | -3.53E-02 |
| 8720 | Panx2 | PANX2 | -1.57E-02 | 9.64E-03 |
| 8721 | Arntl2 | ARNTL2 | 2.71E-02 | -9.13E-03 |
| 8722 | Gtf2h1 | GTF2H1 | -1.18E-02 | 4.77E-03 |
| 8723 | Unc79 | UNC79 | 2.95E-02 | 3.07E-02 |
| 8724 | Rab3il1 | RAB3IL1 | 2.95E-02 | -4.14E-02 |
| 8725 | Tnni2 | TNNI2 | -3.45E-02 | 6.62E-02 |
| 8726 | Foxj3 | FOXJ3 | 9.79E-03 | -1.17E-02 |
| 8727 | Ybx3 | YBX3 | 9.89E-03 | 2.12E-02 |
| 8728 | Copz2 | COPZ2 | -1.10E-02 | -3.36E-02 |
| 8729 | Itgb3bp | ITGB3BP | 2.23E-02 | 5.09E-03 |
| 8730 | Nfkb1 | NFKB1 | -1.08E-02 | 1.32E-02 |
| 8731 | Psca | PSCA | 2.44E-01 | 1.35E-01 |
| 8732 | 2310022A10Rik | C19orf47 | -1.42E-02 | 7.03E-03 |
| 8733 | Clptm1l | CLPTM1L | 9.03E-03 | -6.34E-03 |
| 8734 | Ggt7 | GGT7 | 3.64E-02 | 2.29E-02 |
| 8735 | Rpl38 | RPL38 | 1.17E-02 | 3.91E-03 |
| 8736 | Scn8a | SCN8A | -4.88E-02 | -3.92E-02 |
| 8737 | Hjurp | HJURP | 2.00E-02 | 1.88E-02 |
| 8738 | Smim20 | SMIM20 | 1.12E-02 | -3.31E-02 |
| 8739 | Kif3c | KIF3C | -2.31E-02 | 2.83E-03 |
| 8740 | P2ry13 | P2RY13 | -3.51E-02 | 1.04E-01 |
| 8741 | Slc35b2 | SLC35B2 | 1.07E-02 | -1.78E-03 |
| 8742 | Radil | RADIL | -3.33E-02 | 2.23E-02 |

| | | | | |
|---|---|---|---|---|
| 8743 | Nanp | NANP | -1.83E-02 | 8.31E-03 |
| 8744 | Lsm10 | LSM10 | -2.35E-02 | 3.58E-02 |
| 8745 | Clnk | CLNK | -4.63E-02 | -1.72E-02 |
| 8746 | Mmachc | MMACHC | -1.20E-02 | -1.73E-02 |
| 8747 | Ebf1 | EBF1 | -3.84E-02 | -3.12E-02 |
| 8748 | Hmgcll1 | HMGCLL1 | -4.41E-02 | 5.62E-02 |
| 8749 | Alkbh7 | ALKBH7 | -1.54E-02 | -9.32E-03 |
| 8750 | Snapin | SNAPIN | 1.08E-02 | -1.03E-02 |
| 8751 | Aff3 | AFF3 | -5.60E-02 | 1.29E-02 |
| 8752 | Pacs1 | PACS1 | 1.39E-02 | 8.67E-03 |
| 8753 | 1810055G02Rik | C11orf24 | 1.84E-02 | 2.61E-02 |
| 8754 | Nkd1 | NKD1 | 4.98E-02 | 1.41E-02 |
| 8755 | Psd | PSD | 2.91E-02 | -3.12E-02 |
| 8756 | Tarbp2 | TARBP2 | -1.23E-02 | -5.98E-04 |
| 8757 | Rwdd3 | RWDD3 | -2.25E-02 | -2.89E-02 |
| 8758 | Chst8 | CHST8 | -4.12E-02 | -1.91E-02 |
| 8759 | Cops8 | COPS8 | 8.03E-03 | -6.45E-03 |
| 8760 | Insr | INSR | -1.33E-02 | -1.16E-02 |
| 8761 | Atg13 | ATG13 | 9.67E-03 | -2.50E-02 |
| 8762 | Smim3 | SMIM3 | 2.83E-02 | 6.97E-03 |
| 8763 | Cd300lg | CD300LG | 2.72E-02 | -5.77E-02 |
| 8764 | Ino80b | INO80B | 1.48E-02 | 7.40E-03 |
| 8765 | Ybx2 | YBX2 | 1.74E-02 | -1.23E-02 |
| 8766 | Mcts1 | MCTS1 | 1.10E-02 | -2.51E-03 |
| 8767 | Eny2 | ENY2 | 9.26E-03 | 5.57E-06 |
| 8768 | Rfx2 | RFX2 | 2.62E-02 | 3.45E-03 |
| 8769 | Zfp771 | ZNF771 | 1.78E-02 | 5.73E-03 |
| 8770 | Klf1 | KLF1 | -4.08E-02 | -1.68E-03 |
| 8771 | Cdk12 | CDK12 | -1.27E-02 | -2.25E-03 |
| 8772 | Arhgap10 | ARHGAP10 | -2.29E-02 | -2.19E-02 |
| 8773 | Entpd6 | ENTPD6 | 1.24E-02 | 5.76E-04 |
| 8774 | Sh2d3c | SH2D3C | 2.03E-02 | -3.39E-02 |
| 8775 | Rpn2 | RPN2 | 9.28E-03 | -1.04E-03 |
| 8776 | Susd1 | SUSD1 | 3.42E-02 | 5.91E-02 |
| 8777 | Ucp3 | UCP3 | 7.49E-02 | 1.07E-01 |
| 8778 | Exoc6b | EXOC6B | 1.32E-02 | 5.31E-03 |
| 8779 | Tbp | TBP | -1.54E-02 | -1.48E-02 |
| 8780 | Pou2f3 | POU2F3 | 6.98E-02 | 2.20E-02 |
| 8781 | Btbd11 | BTBD11 | -3.77E-02 | 3.19E-03 |
| 8782 | Parp8 | PARP8 | -4.38E-02 | 2.61E-03 |
| 8783 | Aldh3a2 | ALDH3A2 | 1.56E-02 | -1.07E-02 |
| 8784 | Myo1c | MYO1C | 7.95E-03 | -6.20E-03 |
| 8785 | Ggps1 | GGPS1 | 1.12E-02 | -3.14E-03 |
| 8786 | Ces1d | CES1 | -4.22E-02 | 9.96E-03 |
| 8787 | Crim1 | CRIM1 | -1.65E-02 | -2.98E-02 |
| 8788 | Naa35 | NAA35 | 9.46E-03 | -7.49E-03 |
| 8789 | Neto2 | NETO2 | 3.28E-02 | 5.43E-02 |
| 8790 | Cyp26a1 | CYP26A1 | -3.93E-02 | -5.15E-03 |
| 8791 | Amh | AMH | 2.31E-02 | -7.17E-03 |
| 8792 | Fxr2 | FXR2 | 1.18E-02 | -3.92E-03 |
| 8793 | Clvs1 | CLVS1 | 1.53E-02 | -7.97E-03 |
| 8794 | Ptn | PTN | -5.70E-02 | 4.66E-02 |
| 8795 | Atp2a3 | ATP2A3 | 2.31E-02 | -1.28E-02 |

| | | | | |
|---|---|---|---|---|
| 8796 | Ywhag | YWHAG | 1.08E-02 | 1.75E-02 |
| 8797 | Crb1 | CRB1 | 2.76E-02 | -3.04E-02 |
| 8798 | Orc6 | ORC6 | 2.44E-02 | 2.44E-02 |
| 8799 | Lmtk2 | LMTK2 | 1.36E-02 | -1.95E-02 |
| 8800 | Ddx42 | DDX42 | 9.31E-03 | 4.97E-04 |
| 8801 | Btbd8 | BTBD8 | -2.63E-02 | -7.02E-03 |
| 8802 | Cept1 | CEPT1 | 9.40E-03 | 1.85E-03 |
| 8803 | Tspyl1 | TSPYL1 | -8.96E-03 | -5.96E-03 |
| 8804 | Catsper3 | CATSPER3 | 2.38E-02 | -2.09E-02 |
| 8805 | Rpl37 | RPL37 | -1.01E-02 | 1.61E-02 |
| 8806 | Ccdc42 | CCDC42 | 5.78E-02 | -3.91E-02 |
| 8807 | Btbd3 | BTBD3 | -1.45E-02 | 6.46E-03 |
| 8808 | Pdia4 | PDIA4 | -2.20E-02 | -6.52E-03 |
| 8809 | Egln1 | EGLN1 | -9.32E-03 | -1.06E-02 |
| 8810 | Zscan21 | ZSCAN21 | 1.42E-02 | 3.62E-07 |
| 8811 | Lmna | LMNA | 1.16E-02 | 3.74E-02 |
| 8812 | Akap6 | AKAP6 | 4.23E-02 | 4.47E-02 |
| 8813 | Gfra3 | GFRA3 | -2.30E-02 | 2.25E-02 |
| 8814 | Rab43 | RAB43 | -3.45E-02 | 2.44E-02 |
| 8815 | Chmp2b | CHMP2B | 8.61E-03 | 1.83E-02 |
| 8816 | Mlec | MLEC | -1.09E-02 | -1.80E-02 |
| 8817 | Fbxo24 | FBXO24 | -3.72E-02 | 1.05E-02 |
| 8818 | Myh7b | MYH7B | 3.67E-02 | -2.43E-02 |
| 8819 | Apba2 | APBA2 | -2.15E-02 | -9.73E-03 |
| 8820 | Prkcz | PRKCZ | -1.35E-02 | -2.99E-02 |
| 8821 | Smpd5 | SMPD5 | -3.41E-02 | 4.06E-02 |
| 8822 | Tmem129 | TMEM129 | 1.04E-02 | -3.08E-03 |
| 8823 | Bex2 | BEX1 | 3.20E-02 | 3.47E-02 |
| 8824 | Dbr1 | DBR1 | 1.57E-02 | -3.57E-03 |
| 8825 | Kat5 | KAT5 | 1.06E-02 | -1.33E-02 |
| 8826 | Lefty1 | LEFTY1 | -4.13E-02 | 1.71E-02 |
| 8827 | Nrp2 | NRP2 | -2.73E-02 | -2.90E-02 |
| 8828 | Zfand2a | ZFAND2A | 1.84E-02 | -1.22E-02 |
| 8829 | Phactr3 | PHACTR3 | -1.65E-02 | 1.88E-03 |
| 8830 | Pan2 | PAN2 | -9.22E-03 | -2.35E-02 |
| 8831 | Ythdc2 | YTHDC2 | 2.07E-02 | -5.05E-03 |
| 8832 | Ccnh | CCNH | -1.16E-02 | -1.66E-02 |
| 8833 | Pter | PTER | 1.80E-02 | -1.76E-02 |
| 8834 | Hat1 | HAT1 | 1.31E-02 | 1.80E-02 |
| 8835 | Magix | MAGIX | -2.55E-02 | -1.76E-02 |
| 8836 | Ankrd27 | ANKRD27 | -9.29E-03 | -8.46E-03 |
| 8837 | Stk32c | STK32C | -6.94E-02 | -8.69E-02 |
| 8838 | Foxd1 | FOXD1 | -5.04E-02 | -2.10E-02 |
| 8839 | Pdik1l | PDIK1L | -1.69E-02 | 6.54E-03 |
| 8840 | BC028528 | C1orf54 | -3.68E-02 | 4.87E-02 |
| 8841 | Fam149b | FAM149B1 | 1.06E-02 | -2.49E-02 |
| 8842 | Supt3 | SUPT3H | -2.27E-02 | -2.18E-02 |
| 8843 | Prph | PRPH | 3.26E-02 | 1.14E-02 |
| 8844 | Penk | PENK | 1.09E-01 | 1.10E-02 |
| 8845 | Vamp5 | VAMP5 | -3.45E-02 | -3.13E-02 |
| 8846 | Nadsyn1 | NADSYN1 | -1.66E-02 | -1.74E-02 |
| 8847 | Camk2g | CAMK2G | 1.52E-02 | -1.39E-02 |
| 8848 | Dzip1l | DZIP1L | -1.49E-02 | -2.83E-02 |

| | | | | |
|---|---|---|---|---|
| 8849 | Apoa4 | APOA4 | 6.74E-02 | -1.49E-02 |
| 8850 | Fbxo43 | FBXO43 | -2.52E-02 | -3.59E-03 |
| 8851 | Acap2 | ACAP2 | 1.02E-02 | -1.67E-02 |
| 8852 | Arhgef40 | ARHGEF40 | -1.91E-02 | 2.12E-02 |
| 8853 | Plxna1 | PLXNA1 | -1.13E-02 | -9.07E-03 |
| 8854 | Ranbp3 | RANBP3 | 8.55E-03 | -8.17E-03 |
| 8855 | Smad1 | SMAD1 | 1.65E-02 | 1.34E-02 |
| 8856 | Dock1 | DOCK1 | -1.14E-02 | -4.14E-02 |
| 8857 | Rad54b | RAD54B | 4.32E-02 | 8.02E-02 |
| 8858 | Tceanc2 | TCEANC2 | 6.31E-02 | -3.20E-02 |
| 8859 | Fli1 | FLI1 | -3.69E-02 | 2.32E-02 |
| 8860 | Fam89a | FAM89A | -3.35E-02 | 3.08E-02 |
| 8861 | Ppp2cb | PPP2CB | 1.00E-02 | -6.07E-04 |
| 8862 | Itgb1bp1 | ITGB1BP1 | 1.21E-02 | -2.45E-02 |
| 8863 | Spint1 | SPINT1 | 1.07E-02 | -7.18E-04 |
| 8864 | Napepld | NAPEPLD | 4.78E-02 | -2.08E-02 |
| 8865 | Rab8b | RAB8B | -1.54E-02 | 9.67E-03 |
| 8866 | Farsb | FARSB | 1.26E-02 | 3.31E-04 |
| 8867 | Clasp1 | CLASP1 | 1.02E-02 | -2.01E-03 |
| 8868 | Cdkl5 | CDKL5 | -2.39E-02 | -4.63E-02 |
| 8869 | Ckap4 | CKAP4 | 2.73E-02 | 5.43E-02 |
| 8870 | Olfml3 | OLFML3 | 3.97E-02 | -4.65E-02 |
| 8871 | Nfia | NFIA | -1.32E-02 | -1.80E-02 |
| 8872 | Abtb1 | ABTB1 | 1.17E-02 | -3.21E-02 |
| 8873 | Morc4 | MORC4 | -1.96E-02 | -4.11E-02 |
| 8874 | Traf3ip2 | TRAF3IP2 | 1.92E-02 | -1.33E-03 |
| 8875 | Acss2 | ACSS2 | 1.85E-02 | -4.11E-02 |
| 8876 | Osr2 | OSR2 | -2.63E-02 | 4.34E-03 |
| 8877 | Wnt8b | WNT8B | 8.11E-02 | -3.83E-02 |
| 8878 | Nup88 | NUP88 | 7.91E-03 | -6.13E-03 |
| 8879 | Nxnl1 | NXNL1 | 3.41E-02 | -4.08E-02 |
| 8880 | Grk6 | GRK6 | 9.74E-03 | -3.99E-03 |
| 8881 | Rnf19a | RNF19A | -9.38E-03 | 1.69E-02 |
| 8882 | Nrn1 | NRN1 | -3.02E-02 | 3.90E-02 |
| 8883 | Pim2 | PIM2 | 1.67E-02 | 1.16E-02 |
| 8884 | Cpne8 | CPNE8 | 6.98E-02 | 6.03E-02 |
| 8885 | Rab9 | RAB9A | 1.26E-02 | 5.52E-03 |
| 8886 | Mrpl13 | MRPL13 | 1.07E-02 | -2.77E-03 |
| 8887 | Irx2 | IRX2 | -5.38E-02 | -6.15E-02 |
| 8888 | 1700011L22Rik | C4orf51 | 1.41E-02 | 6.09E-03 |
| 8889 | Trdn | TRDN | -5.48E-02 | -4.73E-02 |
| 8890 | Pabpn1 | PABPN1 | 1.33E-02 | 2.85E-02 |
| 8891 | Asic2 | ASIC2 | 5.33E-02 | -3.35E-02 |
| 8892 | Fmnl2 | FMNL2 | -2.25E-02 | -2.61E-03 |
| 8893 | Nxnl2 | NXNL2 | -3.60E-02 | 1.21E-02 |
| 8894 | Tmc1 | TMC1 | -1.66E-02 | -1.35E-02 |
| 8895 | Cnbp | CNBP | 8.86E-03 | -4.71E-03 |
| 8896 | Ikbkg | IKBKG | -1.04E-02 | -6.98E-04 |
| 8897 | Trim28 | TRIM28 | -6.69E-03 | 1.36E-02 |
| 8898 | Ppp3r2 | PPP3R2 | -1.62E-02 | 6.19E-03 |
| 8899 | Bmx | BMX | 2.94E-02 | -9.66E-03 |
| 8900 | Ddx1 | DDX1 | 9.72E-03 | -1.62E-02 |
| 8901 | Pcdhgb5 | PCDHGB5 | -4.53E-02 | -4.27E-03 |

| | | | | |
|---|---|---|---|---|
| 8902 | Tbc1d24 | TBC1D24 | -1.96E-02 | -5.66E-03 |
| 8903 | Ncapg2 | NCAPG2 | 5.09E-02 | 1.17E-01 |
| 8904 | Pikfyve | PIKFYVE | 1.41E-02 | -4.03E-03 |
| 8905 | Ap4b1 | AP4B1 | 3.52E-02 | -3.67E-02 |
| 8906 | Slc38a11 | SLC38A11 | 3.82E-02 | 3.12E-02 |
| 8907 | Catsper2 | CATSPER2 | -2.93E-02 | -8.55E-03 |
| 8908 | Zfp451 | ZNF451 | -1.43E-02 | 1.10E-02 |
| 8909 | Jmjd6 | JMJD6 | -1.62E-02 | 1.16E-02 |
| 8910 | Habp2 | HABP2 | 2.71E-02 | 8.10E-03 |
| 8911 | Tmem184c | TMEM184C | 1.09E-02 | 4.65E-03 |
| 8912 | Pin4 | PIN4 | 1.43E-02 | 6.11E-03 |
| 8913 | Slc35d3 | SLC35D3 | 1.49E-02 | -1.56E-02 |
| 8914 | Abcf1 | ABCF1 | 8.21E-03 | -4.06E-03 |
| 8915 | Actg2 | ACTG2 | 1.50E-01 | 1.24E-01 |
| 8916 | Akt1s1 | AKT1S1 | 1.13E-02 | -1.05E-02 |
| 8917 | Zbtb17 | ZBTB17 | 1.16E-02 | -1.20E-02 |
| 8918 | Idnk | IDNK | -1.49E-02 | 1.42E-02 |
| 8919 | Parp11 | PARP11 | -2.08E-02 | 1.02E-02 |
| 8920 | Fancm | FANCM | 1.98E-02 | -1.15E-02 |
| 8921 | Wrap53 | WRAP53 | 1.86E-02 | -3.39E-02 |
| 8922 | Wdr20 | WDR20 | 1.07E-02 | -5.89E-03 |
| 8923 | Als2 | ALS2 | 2.10E-02 | -1.68E-02 |
| 8924 | Eif3g | EIF3G | 7.93E-03 | 5.72E-03 |
| 8925 | Pdgfa | PDGFA | 3.56E-02 | 3.71E-02 |
| 8926 | Dpep1 | DPEP1 | 2.86E-02 | -1.34E-02 |
| 8927 | Fkbp15 | FKBP15 | 1.21E-02 | -2.33E-02 |
| 8928 | Naf1 | NAF1 | 1.93E-02 | 2.49E-02 |
| 8929 | Tifa | TIFA | 2.76E-02 | 9.86E-02 |
| 8930 | Tlr3 | TLR3 | -2.37E-02 | 3.13E-03 |
| 8931 | Tox4 | TOX4 | -7.38E-03 | -8.10E-03 |
| 8932 | Plekhg4 | PLEKHG4 | -2.81E-02 | 5.03E-02 |
| 8933 | Myo3b | MYO3B | 5.17E-02 | -2.60E-02 |
| 8934 | Chst11 | CHST11 | 3.80E-02 | -3.95E-02 |
| 8935 | Slc34a1 | SLC34A1 | 2.05E-02 | -2.05E-02 |
| 8936 | Rufy3 | RUFY3 | 1.65E-02 | -2.18E-02 |
| 8937 | Ociad1 | OCIAD1 | 8.57E-03 | -6.27E-03 |
| 8938 | Lgmn | LGMN | 1.94E-02 | 6.29E-03 |
| 8939 | Gpc1 | GPC1 | -1.49E-02 | -3.31E-02 |
| 8940 | Itgam | ITGAM | 6.27E-02 | 2.15E-01 |
| 8941 | Mospd1 | MOSPD1 | 1.24E-02 | 1.44E-02 |
| 8942 | Chic1 | CHIC1 | 1.72E-02 | 3.92E-02 |
| 8943 | Tmem198 | TMEM198 | 3.65E-02 | -1.09E-02 |
| 8944 | Rab39 | RAB39A | -2.40E-02 | 1.35E-02 |
| 8945 | Npy1r | NPY1R | 1.04E-01 | -5.44E-03 |
| 8946 | Chrm2 | CHRM2 | 1.66E-02 | -8.22E-03 |
| 8947 | Eif3d | EIF3D | 8.38E-03 | 7.55E-04 |
| 8948 | Hoxa9 | HOXA9 | -1.11E-02 | -5.35E-03 |
| 8949 | Snrpn | SNRPN | -2.33E-02 | -6.80E-03 |
| 8950 | Sphk1 | SPHK1 | 6.32E-02 | 4.16E-05 |
| 8951 | Nfkbil1 | NFKBIL1 | 1.85E-02 | 2.04E-02 |
| 8952 | Ikzf4 | IKZF4 | -4.36E-02 | 1.84E-02 |
| 8953 | Chmp2a | CHMP2A | 8.75E-03 | -7.26E-03 |
| 8954 | Pars2 | PARS2 | 1.91E-02 | -6.37E-02 |

| | | | | |
|---|---|---|---|---|
| 8955 | Rab39b | RAB39B | -4.01E-02 | -1.61E-02 |
| 8956 | Nop16 | NOP16 | 1.53E-02 | 2.66E-02 |
| 8957 | Kif15 | KIF15 | 7.25E-02 | 1.45E-01 |
| 8958 | Slfn4 | SLFN12L | -5.70E-02 | 6.09E-02 |
| 8959 | Zfp354c | ZNF354C | -2.87E-02 | -9.17E-03 |
| 8960 | Calr3 | CALR3 | 3.04E-02 | -4.09E-02 |
| 8961 | Mrto4 | MRTO4 | 1.51E-02 | 2.08E-04 |
| 8962 | Smim13 | SMIM13 | 1.81E-02 | 3.98E-03 |
| 8963 | Fosl1 | FOSL1 | 6.04E-02 | 1.75E-01 |
| 8964 | Scml4 | SCML4 | 5.20E-02 | -7.54E-02 |
| 8965 | Gpr55 | GPR55 | -4.85E-02 | 4.88E-02 |
| 8966 | Tfg | TFG | 1.22E-02 | 9.80E-03 |
| 8967 | Upk2 | UPK2 | 1.98E-01 | 1.13E-01 |
| 8968 | Spag17 | SPAG17 | 2.01E-02 | 6.75E-03 |
| 8969 | Slc44a4 | SLC44A4 | -1.26E-02 | -1.56E-02 |
| 8970 | Esco2 | ESCO2 | 9.29E-02 | 1.66E-01 |
| 8971 | Frmd5 | FRMD5 | -1.70E-02 | -1.30E-02 |
| 8972 | Ston1 | STON1 | -2.69E-02 | -5.45E-02 |
| 8973 | Tmem241 | TMEM241 | 2.19E-02 | -1.66E-02 |
| 8974 | Agpat1 | AGPAT1 | -9.24E-03 | 1.80E-03 |
| 8975 | Zyx | ZYX | 1.36E-02 | 2.73E-02 |
| 8976 | Caprin2 | CAPRIN2 | -2.34E-02 | -1.96E-02 |
| 8977 | Rnf150 | RNF150 | 3.92E-02 | 5.20E-02 |
| 8978 | Gopc | GOPC | 1.11E-02 | -5.20E-03 |
| 8979 | Pcdhb20 | PCDHB14 | -3.20E-02 | 6.72E-02 |
| 8980 | Wiz | WIZ | 8.00E-03 | -4.77E-03 |
| 8981 | Ugt1a6a | UGT1A6 | -3.77E-02 | 4.10E-02 |
| 8982 | Peli2 | PELI2 | -1.52E-02 | -1.52E-02 |
| 8983 | Hibadh | HIBADH | -1.52E-02 | -2.37E-02 |
| 8984 | Erp27 | ERP27 | 1.99E-02 | -3.74E-02 |
| 8985 | Abi1 | ABI1 | 7.37E-03 | 3.37E-03 |
| 8986 | Shcbp1 | SHCBP1 | 7.34E-02 | 1.49E-01 |
| 8987 | Tdrd7 | TDRD7 | 8.25E-03 | -3.02E-02 |
| 8988 | Cox20 | COX20 | -1.58E-02 | 3.48E-03 |
| 8989 | Tax1bp1 | TAX1BP1 | 1.14E-02 | -1.51E-02 |
| 8990 | Elavl1 | ELAVL1 | -1.15E-02 | 1.75E-02 |
| 8991 | Prpf4b | PRPF4B | -1.06E-02 | 6.66E-03 |
| 8992 | Wtap | WTAP | 8.15E-03 | -7.01E-03 |
| 8993 | Aqp8 | AQP8 | 4.33E-02 | -6.62E-02 |
| 8994 | Itga9 | ITGA9 | -2.78E-02 | 1.00E-02 |
| 8995 | Gnpat | GNPAT | -8.46E-03 | -2.43E-03 |
| 8996 | Syf2 | SYF2 | 9.03E-03 | -1.45E-02 |
| 8997 | Ppp1r3c | PPP1R3C | 1.02E-01 | 7.99E-02 |
| 8998 | Ccdc157 | CCDC157 | 9.56E-03 | -1.05E-02 |
| 8999 | Bid | BID | -1.68E-02 | -1.92E-02 |
| 9000 | Reps1 | REPS1 | 1.51E-02 | -2.41E-03 |
| 9001 | Megf8 | MEGF8 | 9.18E-03 | -2.14E-02 |
| 9002 | Nudt7 | NUDT7 | 1.26E-02 | -1.21E-03 |
| 9003 | Gli1 | GLI1 | -5.28E-02 | 6.67E-02 |
| 9004 | St8sia6 | ST8SIA6 | -2.36E-02 | 2.98E-04 |
| 9005 | Lrfn1 | LRFN1 | -3.15E-02 | 4.36E-02 |
| 9006 | Pbx2 | PBX2 | -1.06E-02 | -9.89E-03 |
| 9007 | Vbp1 | VBP1 | 9.80E-03 | 1.68E-03 |

| | | | | |
|---|---|---|---|---|
| 9008 | Zfp618 | ZNF618 | -1.29E-02 | -2.86E-02 |
| 9009 | Rbm39 | RBM39 | -8.00E-03 | -2.25E-03 |
| 9010 | Gins3 | GINS3 | 4.12E-02 | 2.18E-02 |
| 9011 | Tmem200c | TMEM200C | -1.93E-02 | 5.44E-04 |
| 9012 | Apobec1 | APOBEC1 | 4.48E-02 | 4.65E-02 |
| 9013 | Snai2 | SNAI2 | -4.49E-02 | -2.45E-02 |
| 9014 | Cdc42bpg | CDC42BPG | 1.16E-02 | -1.65E-02 |
| 9015 | Tssk1 | TSSK1B | -2.47E-02 | -1.49E-02 |
| 9016 | Coq10a | COQ10A | 1.07E-02 | 2.61E-03 |
| 9017 | Mtr | MTR | 9.11E-03 | -4.40E-04 |
| 9018 | Rbp4 | RBP4 | 8.94E-02 | 1.41E-01 |
| 9019 | Opn3 | OPN3 | -3.33E-02 | 4.98E-02 |
| 9020 | Det1 | DET1 | -2.01E-02 | 8.46E-03 |
| 9021 | Plekhg2 | PLEKHG2 | 1.78E-02 | 1.36E-02 |
| 9022 | Fbxo47 | FBXO47 | -2.94E-02 | -3.75E-02 |
| 9023 | Ssbp2 | SSBP2 | -1.17E-02 | 5.95E-03 |
| 9024 | Capn6 | CAPN6 | -3.94E-02 | 4.99E-02 |
| 9025 | Zfp219 | ZNF219 | -1.10E-02 | 7.58E-03 |
| 9026 | Fam222b | FAM222B | -1.36E-02 | 2.38E-03 |
| 9027 | Pitrm1 | PITRM1 | 1.58E-02 | -2.26E-02 |
| 9028 | Adam17 | ADAM17 | 1.08E-02 | -8.81E-04 |
| 9029 | Fam228a | FAM228A | -3.93E-02 | -9.31E-03 |
| 9030 | Tnpo1 | TNPO1 | 1.26E-02 | -3.28E-03 |
| 9031 | Tnip2 | TNIP2 | 1.89E-02 | 2.65E-03 |
| 9032 | Btg2 | BTG2 | 8.07E-02 | -1.27E-02 |
| 9033 | Myb | MYB | -2.03E-02 | 2.59E-02 |
| 9034 | Sde2 | SDE2 | -1.26E-02 | -1.96E-02 |
| 9035 | Hpgds | HPGDS | -4.91E-02 | 9.94E-02 |
| 9036 | Stk24 | STK24 | 9.42E-03 | -1.72E-02 |
| 9037 | Tle4 | TLE4 | -1.64E-02 | -5.59E-03 |
| 9038 | Cnp | CNP | -2.39E-02 | 1.85E-02 |
| 9039 | Atf5 | ATF5 | -2.95E-02 | 7.20E-02 |
| 9040 | Slc8b1 | SLC8B1 | 2.27E-02 | -3.12E-02 |
| 9041 | Ccdc178 | CCDC178 | 2.57E-02 | 5.30E-03 |
| 9042 | Nhlrc4 | NHLRC4 | -5.53E-02 | -2.99E-02 |
| 9043 | Hspb9 | HSPB9 | -1.15E-02 | 2.19E-02 |
| 9044 | Prkch | PRKCH | 2.05E-02 | -9.83E-03 |
| 9045 | Sprtn | SPRTN | 1.53E-02 | -1.99E-02 |
| 9046 | Ccdc51 | CCDC51 | -1.62E-02 | -3.01E-02 |
| 9047 | Myom3 | MYOM3 | 4.57E-02 | -1.68E-03 |
| 9048 | Vav3 | VAV3 | -1.70E-02 | -6.78E-03 |
| 9049 | Fn3krp | FN3KRP | -1.88E-02 | -1.49E-02 |
| 9050 | Clec5a | CLEC5A | -4.36E-02 | 1.30E-01 |
| 9051 | Ppfibp2 | PPFIBP2 | -1.77E-02 | -4.16E-03 |
| 9052 | Hey1 | HEY1 | -6.31E-02 | 1.27E-01 |
| 9053 | Traf4 | TRAF4 | -9.37E-03 | 6.91E-03 |
| 9054 | Rnf40 | RNF40 | -6.89E-03 | -4.33E-03 |
| 9055 | Fau | FAU | 1.12E-02 | 2.09E-03 |
| 9056 | Polr1c | POLR1C | -1.39E-02 | 2.72E-03 |
| 9057 | Slc22a2 | SLC22A2 | -1.36E-02 | -1.04E-02 |
| 9058 | Hoxd10 | HOXD10 | 2.63E-02 | 4.57E-02 |
| 9059 | Wdr47 | WDR47 | -1.75E-02 | -2.67E-03 |
| 9060 | Xkr5 | XKR5 | 2.56E-02 | 2.32E-02 |

| | | | | |
|---|---|---|---|---|
| 9061 | Rab42 | RAB42 | -5.34E-02 | -1.46E-02 |
| 9062 | Usf2 | USF2 | -1.23E-02 | -3.80E-03 |
| 9063 | Chit1 | CHIT1 | 1.90E-02 | 2.03E-02 |
| 9064 | Comp | COMP | -3.32E-02 | -6.15E-02 |
| 9065 | Tmem214 | TMEM214 | 7.94E-03 | 2.36E-04 |
| 9066 | Gng5 | GNG5 | 9.02E-03 | 9.10E-03 |
| 9067 | Flt3 | FLT3 | -4.67E-02 | 5.23E-02 |
| 9068 | Smndc1 | SMNDC1 | 1.31E-02 | -8.29E-04 |
| 9069 | Reg3g | REG3G | -5.23E-02 | 6.18E-02 |
| 9070 | Eif3e | EIF3E | -9.89E-03 | 2.31E-02 |
| 9071 | Sh2d1a | SH2D1A | -1.37E-02 | -6.62E-03 |
| 9072 | Hspa14 | HSPA14 | 1.14E-02 | 1.61E-03 |
| 9073 | Pacrg | PACRG | 3.45E-02 | -4.01E-02 |
| 9074 | Lamtor3 | LAMTOR3 | 7.01E-03 | -2.96E-04 |
| 9075 | B3gntl1 | B3GNTL1 | -2.20E-02 | -3.93E-02 |
| 9076 | Lsm4 | LSM4 | 1.70E-02 | 1.17E-03 |
| 9077 | Olfml2a | OLFML2A | -6.04E-02 | 1.36E-02 |
| 9078 | Lrrc14 | LRRC14 | -1.35E-02 | -1.09E-03 |
| 9079 | F5 | F5 | -9.37E-02 | -6.01E-02 |
| 9080 | Lcn12 | LCN12 | 1.61E-02 | -6.79E-03 |
| 9081 | Erich1 | ERICH1 | 1.82E-02 | -4.00E-02 |
| 9082 | Cnpy4 | CNPY4 | -1.88E-02 | 1.82E-02 |
| 9083 | Evi5 | EVI5 | 1.85E-02 | -2.21E-03 |
| 9084 | Pdia6 | PDIA6 | 1.88E-02 | -1.35E-02 |
| 9085 | Tmem119 | TMEM119 | 3.70E-02 | 8.09E-02 |
| 9086 | Herpud2 | HERPUD2 | -8.99E-03 | -7.96E-04 |
| 9087 | Nfatc3 | NFATC3 | -8.97E-03 | -7.19E-03 |
| 9088 | Xk | XK | 5.77E-02 | 1.01E-03 |
| 9089 | Bglap | BGLAP | 1.51E-02 | -5.48E-03 |
| 9090 | Zfp57 | ZFP57 | -4.37E-02 | 3.63E-02 |
| 9091 | Snrnp70 | SNRNP70 | 1.01E-02 | -3.29E-03 |
| 9092 | Cenpp | CENPP | 5.16E-02 | 8.62E-02 |
| 9093 | Nxpe2 | NXPE2 | -3.40E-02 | -6.20E-02 |
| 9094 | Parp6 | PARP6 | -2.35E-02 | 3.22E-03 |
| 9095 | Dzip1 | DZIP1 | -4.57E-02 | 6.09E-03 |
| 9096 | Mafg | MAFG | -1.34E-02 | -1.51E-03 |
| 9097 | Gpr35 | GPR35 | 4.58E-02 | 5.05E-02 |
| 9098 | Ehd3 | EHD3 | 2.59E-02 | -4.82E-02 |
| 9099 | Ybx1 | YBX1 | 9.88E-03 | 1.78E-02 |
| 9100 | Brdt | BRDT | 3.80E-02 | -3.79E-02 |
| 9101 | Arnt | ARNT | 6.70E-03 | -6.09E-03 |
| 9102 | Alg10b | ALG10 | -1.73E-02 | -2.82E-02 |
| 9103 | Mrps2 | MRPS2 | -1.02E-02 | -1.18E-02 |
| 9104 | Fahd1 | FAHD1 | -1.22E-02 | -2.39E-02 |
| 9105 | Zfp263 | ZNF263 | -1.08E-02 | 7.36E-03 |
| 9106 | Chst14 | CHST14 | -2.82E-02 | -5.92E-03 |
| 9107 | Rcan2 | RCAN2 | 1.58E-02 | 3.50E-02 |
| 9108 | Klhl18 | KLHL18 | -8.29E-03 | 2.18E-03 |
| 9109 | Prkaca | PRKACA | 8.89E-03 | -7.62E-03 |
| 9110 | Tti2 | TTI2 | -1.09E-02 | -2.22E-02 |
| 9111 | Hipk3 | HIPK3 | -1.11E-02 | -2.04E-02 |
| 9112 | Sapcd2 | SAPCD2 | 4.99E-02 | 8.77E-02 |
| 9113 | Dnah7a | DNAH7 | -1.85E-02 | -1.96E-02 |

| | | | | |
|---|---|---|---|---|
| 9114 | Kcnn3 | KCNN3 | -5.65E-02 | -2.48E-02 |
| 9115 | Leng1 | LENG1 | 1.23E-02 | -1.17E-02 |
| 9116 | Kank2 | KANK2 | -1.86E-02 | 3.43E-02 |
| 9117 | Klhl36 | KLHL36 | -1.85E-02 | -2.25E-02 |
| 9118 | Ficd | FICD | -1.33E-02 | -1.64E-02 |
| 9119 | Tmem217 | TMEM217 | -1.89E-02 | -1.14E-03 |
| 9120 | Lsm3 | LSM3 | -1.77E-02 | 1.97E-02 |
| 9121 | Rusc2 | RUSC2 | 1.24E-02 | 5.05E-03 |
| 9122 | Setd6 | SETD6 | 1.32E-02 | 2.27E-03 |
| 9123 | Cep55 | CEP55 | 8.18E-02 | 2.08E-01 |
| 9124 | Atp2b1 | ATP2B1 | -1.16E-02 | 7.24E-03 |
| 9125 | Zmat5 | ZMAT5 | 1.35E-02 | -2.01E-02 |
| 9126 | Bccip | BCCIP | 8.72E-03 | -1.79E-02 |
| 9127 | Mbip | MBIP | -1.48E-02 | 8.29E-03 |
| 9128 | Med10 | MED10 | 1.21E-02 | 6.79E-03 |
| 9129 | Etl4 | KIAA1217 | -1.12E-02 | -2.91E-03 |
| 9130 | Stk38 | STK38 | 8.66E-03 | 3.10E-03 |
| 9131 | Coch | COCH | -9.83E-02 | -6.11E-02 |
| 9132 | Parpbp | PARPBP | 6.78E-02 | 1.86E-01 |
| 9133 | Ctnnd1 | CTNND1 | -1.18E-02 | 5.05E-03 |
| 9134 | Abtb2 | ABTB2 | 2.19E-02 | -1.40E-02 |
| 9135 | Chchd5 | CHCHD5 | -1.42E-02 | -1.85E-02 |
| 9136 | Ptges | PTGES | 2.89E-02 | 5.20E-02 |
| 9137 | Imp4 | IMP4 | -9.55E-03 | -1.47E-02 |
| 9138 | Vstm2a | VSTM2A | -2.74E-02 | -8.75E-03 |
| 9139 | Slc30a1 | SLC30A1 | 4.00E-02 | -1.77E-02 |
| 9140 | Dhx57 | DHX57 | -1.05E-02 | -1.06E-02 |
| 9141 | Hebp2 | HEBP2 | 5.06E-02 | 1.34E-02 |
| 9142 | Efcab2 | EFCAB2 | 1.39E-02 | -3.96E-02 |
| 9143 | B4galnt1 | B4GALNT1 | 3.20E-02 | 2.98E-02 |
| 9144 | Rbm26 | RBM26 | 1.01E-02 | -1.66E-02 |
| 9145 | Glp2r | GLP2R | 1.48E-02 | -7.73E-03 |
| 9146 | Pdzd4 | PDZD4 | 4.36E-02 | 6.38E-02 |
| 9147 | Nfe2l3 | NFE2L3 | 1.78E-01 | 7.52E-02 |
| 9148 | Anxa7 | ANXA7 | 1.09E-02 | -8.32E-03 |
| 9149 | Dynlt1f | DYNLT1 | 2.51E-02 | 4.71E-02 |
| 9150 | Snrpf | SNRPF | -1.32E-02 | 1.14E-02 |
| 9151 | Htr4 | HTR4 | -7.79E-02 | -3.41E-02 |
| 9152 | Cryzl1 | CRYZL1 | -7.37E-03 | 3.33E-04 |
| 9153 | Tpra1 | TPRA1 | -9.68E-03 | 6.18E-03 |
| 9154 | Srd5a1 | SRD5A1 | 2.75E-02 | 3.31E-02 |
| 9155 | Cobl | COBL | -1.39E-02 | -2.91E-03 |
| 9156 | Tmem159 | TMEM159 | 1.26E-02 | -2.57E-02 |
| 9157 | Smad7 | SMAD7 | -1.91E-02 | 1.58E-02 |
| 9158 | Rnf130 | RNF130 | 7.61E-03 | -5.79E-03 |
| 9159 | Swt1 | SWT1 | -1.56E-02 | -7.09E-03 |
| 9160 | Prdm10 | PRDM10 | 1.48E-02 | -5.51E-03 |
| 9161 | Donson | DONSON | -3.44E-02 | 7.84E-02 |
| 9162 | Smg8 | SMG8 | -1.25E-02 | 1.30E-03 |
| 9163 | Muc20 | MUC20 | 1.46E-01 | -2.80E-02 |
| 9164 | Ube4a | UBE4A | -9.32E-03 | -5.51E-03 |
| 9165 | Eif4e2 | EIF4E2 | 9.13E-03 | -1.43E-03 |
| 9166 | Unc13a | UNC13A | 3.22E-02 | 1.13E-03 |

| | | | | |
|---|---|---|---|---|
| 9167 | Cchcr1 | CCHCR1 | -1.82E-02 | 6.21E-03 |
| 9168 | Slc9a4 | SLC9A4 | 5.83E-02 | 8.94E-02 |
| 9169 | Rplp0 | RPLP0 | 1.14E-02 | 1.17E-02 |
| 9170 | St3gal2 | ST3GAL2 | -1.33E-02 | 7.87E-03 |
| 9171 | Sprn | SPRN | -1.55E-02 | 2.48E-02 |
| 9172 | Zzz3 | ZZZ3 | -9.19E-03 | -4.40E-03 |
| 9173 | Hint2 | HINT2 | 1.35E-02 | -2.42E-02 |
| 9174 | Aktip | AKTIP | 1.50E-02 | -2.58E-02 |
| 9175 | Rbp7 | RBP7 | -2.01E-02 | 4.41E-02 |
| 9176 | Smg5 | SMG5 | 7.29E-03 | -4.77E-03 |
| 9177 | Jag2 | JAG2 | 2.95E-02 | 4.24E-02 |
| 9178 | Efhc2 | EFHC2 | 3.04E-02 | 9.28E-02 |
| 9179 | Rgs12 | RGS12 | -2.61E-02 | -5.15E-02 |
| 9180 | Ido1 | IDO1 | -2.55E-02 | 6.25E-04 |
| 9181 | Lemd3 | LEMD3 | -1.10E-02 | -1.08E-02 |
| 9182 | Popdc2 | POPDC2 | 2.60E-02 | 6.87E-03 |
| 9183 | Rnf144a | RNF144A | -2.66E-02 | -5.29E-02 |
| 9184 | Adam32 | ADAM32 | -2.68E-02 | -6.05E-02 |
| 9185 | Klrg2 | KLRG2 | 1.11E-01 | 1.42E-01 |
| 9186 | Tnfrsf1b | TNFRSF1B | -4.02E-02 | 9.32E-02 |
| 9187 | Max | MAX | 1.03E-02 | -4.25E-03 |
| 9188 | Mpzl2 | MPZL2 | -1.70E-02 | -2.49E-02 |
| 9189 | Mmp12 | MMP12 | 5.11E-02 | 1.35E-01 |
| 9190 | Ttc28 | TTC28 | -3.36E-02 | -4.86E-02 |
| 9191 | Dct | DCT | -1.56E-02 | 8.22E-03 |
| 9192 | Dlgap3 | DLGAP3 | 3.16E-02 | 3.06E-02 |
| 9193 | Pbrm1 | PBRM1 | -9.59E-03 | -1.06E-02 |
| 9194 | Dnmbp | DNMBP | -1.46E-02 | -1.03E-02 |
| 9195 | Ttc22 | TTC22 | 3.52E-02 | 6.83E-03 |
| 9196 | Tlr2 | TLR2 | 6.00E-02 | 1.57E-01 |
| 9197 | Mical3 | MICAL3 | 1.08E-02 | 2.92E-03 |
| 9198 | Ephx4 | EPHX4 | -1.78E-02 | -1.21E-02 |
| 9199 | Nbeal2 | NBEAL2 | -1.40E-02 | 3.92E-02 |
| 9200 | Mtap | MTAP | -8.96E-03 | -1.44E-02 |
| 9201 | Ppil3 | PPIL3 | 2.27E-02 | 1.39E-02 |
| 9202 | Xkrx | XKRX | 4.73E-02 | -9.60E-02 |
| 9203 | Vamp8 | VAMP8 | 2.97E-01 | -1.49E-01 |
| 9204 | Map2k5 | MAP2K5 | 1.10E-02 | -1.18E-02 |
| 9205 | Sbds | SBDS | -8.71E-03 | -5.56E-04 |
| 9206 | Tnfrsf4 | TNFRSF4 | 4.13E-02 | -3.03E-02 |
| 9207 | Naif1 | NAIF1 | 1.77E-02 | 9.53E-04 |
| 9208 | Myo9b | MYO9B | -8.81E-03 | 1.37E-02 |
| 9209 | Atp2a1 | ATP2A1 | 2.59E-02 | 9.82E-04 |
| 9210 | Stt3a | STT3A | -7.78E-03 | -8.39E-03 |
| 9211 | Anxa8 | ANXA8L1 | -1.10E-01 | 1.65E-01 |
| 9212 | Anxa8 | ANXA8 | -1.10E-01 | 1.65E-01 |
| 9213 | Samd10 | SAMD10 | 1.08E-02 | -2.13E-03 |
| 9214 | Wdr54 | WDR54 | -2.81E-02 | 2.06E-02 |
| 9215 | Arpp21 | ARPP21 | -2.52E-02 | 5.24E-02 |
| 9216 | Mllt6 | MLLT6 | -9.15E-03 | -2.67E-03 |
| 9217 | Snx21 | SNX21 | -1.39E-02 | -4.02E-02 |
| 9218 | Ccdc134 | CCDC134 | 1.85E-02 | -7.40E-03 |
| 9219 | Ccdc173 | CCDC173 | -2.51E-02 | -7.54E-03 |

| | | | | |
|---|---|---|---|---|
| 9220 | Rtel1 | RTEL1 | 1.54E-02 | 5.43E-03 |
| 9221 | 1810009A15Rik | LOC102288414 | 2.75E-02 | 2.48E-03 |
| 9222 | Pkd1 | PKD1 | -9.84E-03 | -2.83E-03 |
| 9223 | Mfsd6 | MFSD6 | -3.00E-02 | -9.06E-03 |
| 9224 | Zfat | ZFAT | -3.63E-02 | 2.25E-02 |
| 9225 | Ednra | EDNRA | 3.84E-02 | 4.91E-02 |
| 9226 | Sdhaf2 | SDHAF2 | 9.71E-03 | 2.69E-03 |
| 9227 | Ndufaf2 | NDUFAF2 | 1.50E-02 | -7.38E-03 |
| 9228 | Rsph3b | RSPH3 | -1.65E-02 | -2.90E-02 |
| 9229 | S100a4 | S100A4 | 2.08E-02 | 4.87E-02 |
| 9230 | Kcnd3 | KCND3 | -5.75E-02 | -1.82E-02 |
| 9231 | Commd9 | COMMD9 | 1.05E-02 | -8.02E-03 |
| 9232 | Ergic3 | ERGIC3 | -1.03E-02 | -6.78E-04 |
| 9233 | Cd200r1 | CD200R1 | -2.35E-02 | 4.06E-02 |
| 9234 | Rcc1 | RCC1 | 1.69E-02 | 1.57E-02 |
| 9235 | Ercc6l | ERCC6L | 7.66E-02 | 1.97E-01 |
| 9236 | Oaf | OAF | -1.75E-02 | 1.20E-02 |
| 9237 | Mmp20 | MMP20 | 1.11E-02 | 1.68E-03 |
| 9238 | Tmem72 | TMEM72 | 1.76E-02 | -2.75E-02 |
| 9239 | Elavl4 | ELAVL4 | -1.71E-02 | 1.78E-02 |
| 9240 | Lrp1b | LRP1B | -1.96E-02 | -5.91E-04 |
| 9241 | Il2ra | IL2RA | 3.14E-02 | -2.88E-02 |
| 9242 | Casp16 | CASP16 | -1.57E-02 | 1.96E-03 |
| 9243 | A1cf | A1CF | -3.71E-02 | -4.12E-03 |
| 9244 | Ulk3 | ULK3 | -1.35E-02 | 4.20E-03 |
| 9245 | Galc | GALC | 1.38E-02 | -2.45E-02 |
| 9246 | Ankrd34b | ANKRD34B | -4.60E-02 | -8.50E-02 |
| 9247 | 2410004P03Rik | C2orf50 | 1.87E-02 | 1.10E-02 |
| 9248 | Tmem63c | TMEM63C | 1.55E-02 | -2.22E-02 |
| 9249 | Trim39 | TRIM39 | -1.06E-02 | 1.92E-03 |
| 9250 | Rnft1 | RNFT1 | 1.03E-02 | -1.01E-03 |
| 9251 | Ablim3 | ABLIM3 | -2.99E-02 | 3.49E-02 |
| 9252 | Morc2a | MORC2 | 8.40E-03 | -1.89E-03 |
| 9253 | Crlf3 | CRLF3 | -1.77E-02 | -1.01E-02 |
| 9254 | Zbtb34 | ZBTB34 | -1.91E-02 | 1.88E-02 |
| 9255 | Rin3 | RIN3 | 1.37E-02 | 3.82E-03 |
| 9256 | Plat | PLAT | -2.66E-02 | 2.92E-02 |
| 9257 | Mxd3 | MXD3 | 7.14E-02 | 1.57E-01 |
| 9258 | Rreb1 | RREB1 | -1.27E-02 | 1.26E-02 |
| 9259 | Nr1i3 | NR1I3 | 5.82E-02 | 5.25E-02 |
| 9260 | Atp11c | ATP11C | -1.36E-02 | -2.15E-02 |
| 9261 | Mkln1 | MKLN1 | 1.05E-02 | -1.06E-02 |
| 9262 | Itgav | ITGAV | 1.27E-02 | -1.56E-02 |
| 9263 | Sema5b | SEMA5B | 4.62E-02 | -9.18E-02 |
| 9264 | Wfdc2 | WFDC2 | -1.53E-02 | 7.26E-02 |
| 9265 | Slc23a1 | SLC23A1 | -1.42E-02 | -4.39E-02 |
| 9266 | Nid1 | NID1 | 4.16E-02 | 4.28E-03 |
| 9267 | Zfp503 | ZNF503 | -1.29E-02 | -2.67E-03 |
| 9268 | 6030458C11Rik | C5orf22 | -1.01E-02 | 1.92E-03 |
| 9269 | Pex3 | PEX3 | -8.33E-03 | -1.33E-02 |
| 9270 | Ppp1r9a | PPP1R9A | -3.04E-02 | -3.93E-02 |
| 9271 | Axdnd1 | AXDND1 | 2.09E-02 | 1.64E-02 |
| 9272 | 4930453N24Rik | C3orf38 | 1.24E-02 | 4.49E-04 |

| | | | | |
|---|---|---|---|---|
| 9273 | Peak1 | PEAK1 | -1.12E-02 | -1.07E-02 |
| 9274 | Nr4a1 | NR4A1 | 1.25E-01 | 2.16E-02 |
| 9275 | Gsta1 | GSTA5 | 1.01E-01 | 8.81E-02 |
| 9276 | Tmed8 | TMED8 | -1.23E-02 | -2.31E-02 |
| 9277 | Rab12 | RAB12 | 7.41E-03 | 1.75E-02 |
| 9278 | Tspan13 | TSPAN13 | -4.57E-02 | 4.76E-02 |
| 9279 | Igfbp5 | IGFBP5 | 1.41E-02 | -6.16E-02 |
| 9280 | Sgcd | SGCD | -3.30E-02 | 2.93E-02 |
| 9281 | Tmem182 | TMEM182 | -3.97E-02 | -8.50E-03 |
| 9282 | Cth | CTH | -1.27E-02 | -4.65E-02 |
| 9283 | Man2c1 | MAN2C1 | 7.26E-03 | -1.29E-02 |
| 9284 | Fgr | FGR | -6.94E-02 | 1.84E-01 |
| 9285 | Smap2 | SMAP2 | 1.37E-02 | -2.32E-02 |
| 9286 | Dlk1 | DLK1 | -1.02E-02 | 8.41E-04 |
| 9287 | Rabep1 | RABEP1 | -9.55E-03 | -6.21E-03 |
| 9288 | Vil1 | VIL1 | 1.86E-02 | -5.91E-03 |
| 9289 | Fut7 | FUT7 | 1.94E-02 | -1.18E-02 |
| 9290 | Amotl1 | AMOTL1 | -1.99E-02 | -7.90E-03 |
| 9291 | Kazald1 | KAZALD1 | -2.72E-02 | 6.27E-03 |
| 9292 | Mfsd2b | MFSD2B | 3.28E-02 | -3.98E-02 |
| 9293 | Nanos3 | NANOS3 | -1.39E-02 | 2.36E-03 |
| 9294 | Aspg | ASPG | 3.48E-02 | 2.01E-02 |
| 9295 | Mrps31 | MRPS31 | 1.46E-02 | -2.49E-02 |
| 9296 | Ak8 | AK8 | 4.29E-02 | 2.44E-02 |
| 9297 | Sfxn5 | SFXN5 | -1.38E-02 | -1.98E-02 |
| 9298 | Iqsec2 | IQSEC2 | 1.27E-02 | -8.92E-03 |
| 9299 | Poglut1 | POGLUT1 | -9.55E-03 | -7.44E-03 |
| 9300 | Il17d | IL17D | 2.57E-02 | -1.68E-02 |
| 9301 | Tsc1 | TSC1 | 1.34E-02 | -2.65E-02 |
| 9302 | Actr8 | ACTR8 | 8.38E-03 | -7.02E-03 |
| 9303 | Atp1a4 | ATP1A4 | 1.16E-02 | 1.92E-02 |
| 9304 | Bves | BVES | 2.16E-02 | 1.46E-02 |
| 9305 | Ryk | RYK | -7.81E-03 | 3.72E-03 |
| 9306 | Mettl24 | METTL24 | 3.31E-02 | -1.81E-02 |
| 9307 | Arc | ARC | 1.33E-01 | -6.94E-03 |
| 9308 | Nudt2 | NUDT2 | -1.13E-02 | -1.86E-02 |
| 9309 | Ckb | CKB | 3.70E-02 | -3.52E-02 |
| 9310 | Ccdc85b | CCDC85B | -1.53E-02 | -2.09E-02 |
| 9311 | Krt4 | KRT4 | 5.53E-02 | 1.71E-02 |
| 9312 | Mad2l1bp | MAD2L1BP | 1.74E-02 | -1.47E-02 |
| 9313 | Kmt2e | KMT2E | -1.10E-02 | -5.63E-04 |
| 9314 | Pop5 | POP5 | 9.95E-03 | -8.92E-03 |
| 9315 | Sox7 | SOX7 | 3.78E-02 | -2.38E-02 |
| 9316 | Hbb-bt | HBB | -6.72E-02 | 2.42E-02 |
| 9317 | Tbx6 | TBX6 | -3.90E-02 | -8.09E-03 |
| 9318 | Ttbk1 | TTBK1 | 2.96E-02 | -1.28E-02 |
| 9319 | Adcy1 | ADCY1 | -3.85E-02 | 1.82E-02 |
| 9320 | Nudc | NUDC | 9.41E-03 | -5.43E-03 |
| 9321 | Ampd2 | AMPD2 | 1.20E-02 | -1.16E-02 |
| 9322 | Nova2 | NOVA2 | -3.15E-02 | 1.77E-02 |
| 9323 | Vtcn1 | VTCN1 | -8.59E-02 | 1.88E-01 |
| 9324 | Rnf186 | RNF186 | 1.83E-02 | -2.74E-02 |
| 9325 | Mroh8 | MROH8 | 2.71E-02 | 2.10E-02 |

| | | | | |
|---|---|---|---|---|
| 9326 | Sema3g | SEMA3G | -2.19E-02 | 1.45E-02 |
| 9327 | 1700037C18Rik | C16orf90 | -2.00E-02 | -1.48E-02 |
| 9328 | Kcnh5 | KCNH5 | -2.90E-02 | -1.52E-02 |
| 9329 | Pla2g12b | PLA2G12B | 2.73E-02 | 5.10E-02 |
| 9330 | Tet1 | TET1 | -3.18E-02 | -4.69E-02 |
| 9331 | Lrrc26 | LRRC26 | -1.01E-01 | 1.01E-01 |
| 9332 | Bcl10 | BCL10 | 1.16E-02 | 2.50E-02 |
| 9333 | Slc5a1 | SLC5A1 | 1.72E-02 | 1.12E-03 |
| 9334 | Epn1 | EPN1 | 1.21E-02 | -4.15E-03 |
| 9335 | Kif7 | KIF7 | 3.69E-02 | 1.73E-02 |
| 9336 | Krtcap3 | KRTCAP3 | -4.14E-02 | 5.29E-02 |
| 9337 | Ibsp | IBSP | -7.40E-03 | 1.70E-03 |
| 9338 | Isg15 | ISG15 | -4.19E-02 | 1.39E-01 |
| 9339 | Tmem220 | TMEM220 | 1.88E-02 | -2.42E-02 |
| 9340 | Crtc1 | CRTC1 | 1.12E-02 | -9.06E-03 |
| 9341 | Mipol1 | MIPOL1 | -2.16E-02 | 7.51E-03 |
| 9342 | Stox1 | STOX1 | -3.30E-02 | -2.93E-03 |
| 9343 | Mta2 | MTA2 | 6.84E-03 | 4.74E-03 |
| 9344 | Pfn4 | PFN4 | 2.72E-02 | -2.79E-02 |
| 9345 | Thnsl1 | THNSL1 | 1.74E-02 | -1.54E-02 |
| 9346 | D030056L22Rik | C9orf40 | -2.66E-02 | 2.93E-02 |
| 9347 | Zbtb7a | ZBTB7A | -8.09E-03 | 4.95E-03 |
| 9348 | Sac3d1 | SAC3D1 | -1.69E-02 | 1.58E-02 |
| 9349 | Cog4 | COG4 | -8.68E-03 | -2.58E-03 |
| 9350 | Mcm4 | MCM4 | 2.77E-02 | 5.54E-02 |
| 9351 | Ext2 | EXT2 | -8.83E-03 | -6.66E-03 |
| 9352 | Amd1 | AMD1 | -1.65E-02 | 1.27E-02 |
| 9353 | Sec61a2 | SEC61A2 | 1.19E-02 | 3.75E-03 |
| 9354 | Stom | STOM | -1.78E-02 | 1.51E-02 |
| 9355 | Mcph1 | MCPH1 | 2.20E-02 | 2.77E-02 |
| 9356 | 4930590J08Rik | C3orf20 | 2.86E-02 | -1.53E-02 |
| 9357 | Anapc13 | ANAPC13 | -8.81E-03 | -1.19E-02 |
| 9358 | Cd84 | CD84 | -3.20E-02 | 1.62E-01 |
| 9359 | Trim5 | TRIM5 | -4.01E-02 | -4.73E-03 |
| 9360 | Dcbld2 | DCBLD2 | -2.23E-02 | 7.12E-03 |
| 9361 | Mettl5 | METTL5 | 1.81E-02 | -2.11E-02 |
| 9362 | Slc30a6 | SLC30A6 | -9.12E-03 | -1.58E-02 |
| 9363 | Nav1 | NAV1 | 4.56E-02 | 3.79E-02 |
| 9364 | Marcksl1 | MARCKSL1 | 2.16E-02 | 8.21E-02 |
| 9365 | Cox15 | COX15 | -1.28E-02 | -1.35E-02 |
| 9366 | Grin2d | GRIN2D | -2.87E-02 | 3.17E-02 |
| 9367 | Fbxw10 | FBXW10 | -2.47E-02 | -5.05E-02 |
| 9368 | Ccdc148 | CCDC148 | 3.17E-02 | -1.24E-02 |
| 9369 | Pik3ca | PIK3CA | -1.30E-02 | -3.18E-02 |
| 9370 | Nsl1 | NSL1 | 5.04E-02 | 9.13E-02 |
| 9371 | Ltf | LTF | -3.65E-02 | -2.10E-03 |
| 9372 | Jmjd7 | JMJD7 | -2.66E-02 | -9.68E-03 |
| 9373 | Gck | GCK | -1.93E-02 | 3.15E-02 |
| 9374 | Pcmtd1 | PCMTD1 | -8.96E-03 | -2.21E-02 |
| 9375 | Frmd6 | FRMD6 | -2.86E-02 | -2.11E-03 |
| 9376 | Hsd3b7 | HSD3B7 | -3.03E-02 | 5.73E-02 |
| 9377 | Camsap1 | CAMSAP1 | -1.49E-02 | 1.12E-02 |
| 9378 | P4ha1 | P4HA1 | -1.83E-02 | -2.60E-03 |

| | | | | |
|---|---|---|---|---|
| 9379 | Mrpl41 | MRPL41 | 1.14E-02 | -2.22E-02 |
| 9380 | Zadh2 | ZADH2 | 7.99E-03 | -2.84E-02 |
| 9381 | Bhlha9 | BHLHA9 | -8.96E-03 | 1.06E-02 |
| 9382 | Plscr4 | PLSCR4 | 2.53E-02 | -5.11E-03 |
| 9383 | Ppm1d | PPM1D | 1.25E-02 | -1.34E-02 |
| 9384 | Kdm3b | KDM3B | -7.35E-03 | -1.29E-02 |
| 9385 | Chd1 | CHD1 | 8.24E-03 | -1.09E-02 |
| 9386 | Gprin1 | GPRIN1 | -1.11E-02 | 1.59E-02 |
| 9387 | Pms2 | PMS2 | 2.14E-02 | 2.52E-03 |
| 9388 | Rps21 | RPS21 | -9.72E-03 | 3.75E-03 |
| 9389 | Ltbp1 | LTBP1 | -3.23E-02 | -3.04E-02 |
| 9390 | Samd9l | SAMD9L | -1.11E-02 | -2.42E-02 |
| 9391 | Mipep | MIPEP | -1.16E-02 | -1.65E-02 |
| 9392 | Trim41 | TRIM41 | -8.26E-03 | -6.17E-03 |
| 9393 | Snx10 | SNX10 | 2.57E-02 | 7.72E-02 |
| 9394 | Ggn | GGN | -2.88E-02 | 4.63E-02 |
| 9395 | Prss33 | PRSS33 | -8.04E-03 | -4.72E-04 |
| 9396 | Cdc45 | CDC45 | 3.57E-02 | 6.74E-02 |
| 9397 | Dnajc6 | DNAJC6 | -3.35E-02 | -5.88E-02 |
| 9398 | S100pbp | S100PBP | 8.85E-03 | -4.97E-03 |
| 9399 | Plxnb1 | PLXNB1 | -1.17E-02 | 2.05E-04 |
| 9400 | Arhgap33 | ARHGAP33 | 3.00E-02 | 1.49E-02 |
| 9401 | Ddx20 | DDX20 | 1.24E-02 | -7.35E-03 |
| 9402 | Prlr | PRLR | -2.42E-02 | -4.85E-02 |
| 9403 | Mta1 | MTA1 | -1.13E-02 | -1.38E-03 |
| 9404 | Wrap73 | WRAP73 | -1.48E-02 | -1.57E-02 |
| 9405 | Zfp160 | ZNF665 | -1.61E-02 | -8.82E-03 |
| 9406 | Gab1 | GAB1 | -1.32E-02 | -2.13E-02 |
| 9407 | Tas1r2 | TAS1R2 | -1.58E-02 | 6.43E-03 |
| 9408 | Col6a3 | COL6A3 | -4.25E-02 | 1.06E-02 |
| 9409 | Cln6 | CLN6 | -1.18E-02 | 5.61E-03 |
| 9410 | Mrpl19 | MRPL19 | -9.81E-03 | -5.23E-03 |
| 9411 | Gpr27 | GPR27 | -5.11E-02 | 8.23E-02 |
| 9412 | Cntnap3 | CNTNAP3 | -2.50E-02 | 4.01E-02 |
| 9413 | Kctd5 | KCTD5 | -1.89E-02 | 7.60E-03 |
| 9414 | Arfgef1 | ARFGEF1 | -7.57E-03 | -1.48E-02 |
| 9415 | Fnip2 | FNIP2 | -3.18E-02 | -3.60E-02 |
| 9416 | Areg | AREG | 3.08E-02 | 1.28E-01 |
| 9417 | Irx1 | IRX1 | -4.86E-02 | -7.87E-02 |
| 9418 | Gnas | GNAS | 8.04E-03 | -6.68E-03 |
| 9419 | Phf19 | PHF19 | -4.76E-02 | 1.33E-01 |
| 9420 | Phc2 | PHC2 | 1.11E-02 | 2.84E-03 |
| 9421 | Fbxl18 | FBXL18 | 1.05E-02 | -4.69E-03 |
| 9422 | Alg9 | ALG9 | 9.13E-03 | -1.54E-02 |
| 9423 | Zfp606 | ZNF606 | -1.57E-02 | -4.08E-03 |
| 9424 | Abhd17a | ABHD17A | 9.79E-03 | 9.73E-04 |
| 9425 | Rpl27a | RPL27A | -8.38E-03 | 7.13E-03 |
| 9426 | Zfp300 | ZNF567 | -2.61E-02 | -1.65E-02 |
| 9427 | Anxa4 | ANXA4 | -8.62E-03 | 8.80E-03 |
| 9428 | Vamp1 | VAMP1 | 1.43E-02 | -2.39E-03 |
| 9429 | Pcp4 | PCP4 | -1.45E-02 | 1.36E-02 |
| 9430 | Cxadr | CXADR | -1.79E-02 | 4.30E-02 |
| 9431 | Ocln | OCLN | 1.33E-02 | -4.07E-02 |

| | | | | |
|---|---|---|---|---|
| 9432 | P4hb | P4HB | 1.00E-02 | -4.13E-04 |
| 9433 | 2310030G06Rik | C11orf52 | -1.32E-02 | 6.36E-03 |
| 9434 | Zcchc7 | ZCCHC7 | -9.58E-03 | 2.27E-02 |
| 9435 | Lrrc15 | LRRC15 | -1.20E-02 | 1.42E-02 |
| 9436 | Rnf187 | RNF187 | 7.83E-03 | -1.22E-02 |
| 9437 | Cyp2f2 | CYP2F1 | 6.61E-02 | -4.10E-03 |
| 9438 | Cln8 | CLN8 | -9.59E-03 | 4.34E-03 |
| 9439 | Ntf5 | NTF4 | -4.66E-02 | 5.84E-02 |
| 9440 | Wwp2 | WWP2 | 1.15E-02 | -3.74E-02 |
| 9441 | Rnls | RNLS | 3.10E-02 | -5.52E-02 |
| 9442 | Ap3d1 | AP3D1 | 7.21E-03 | -7.23E-03 |
| 9443 | Wwox | WWOX | 1.19E-02 | 6.96E-03 |
| 9444 | Cyhr1 | CYHR1 | -7.83E-03 | -1.18E-02 |
| 9445 | Klhl3 | KLHL3 | 1.96E-02 | -1.92E-02 |
| 9446 | Pot1a | POT1 | -2.10E-02 | 9.05E-03 |
| 9447 | Sptbn2 | SPTBN2 | 3.43E-02 | -1.52E-02 |
| 9448 | Clspn | CLSPN | 8.26E-02 | 2.39E-01 |
| 9449 | Prmt3 | PRMT3 | 1.28E-02 | -4.59E-03 |
| 9450 | Armcx1 | ARMCX1 | -2.15E-02 | 3.60E-02 |
| 9451 | Rab14 | RAB14 | 8.05E-03 | -8.44E-03 |
| 9452 | Socs4 | SOCS4 | -1.28E-02 | 8.19E-03 |
| 9453 | Unk | UNK | 1.06E-02 | -2.13E-02 |
| 9454 | Tdrd9 | TDRD9 | -2.03E-02 | 1.93E-02 |
| 9455 | Lrriq1 | LRRIQ1 | -4.68E-02 | -4.68E-02 |
| 9456 | Ormdl2 | ORMDL2 | -1.35E-02 | 5.59E-03 |
| 9457 | Gga1 | GGA1 | 6.48E-03 | -6.04E-03 |
| 9458 | Tex264 | TEX264 | 7.75E-03 | -1.12E-02 |
| 9459 | Hyal2 | HYAL2 | 1.43E-02 | 8.28E-04 |
| 9460 | Paxbp1 | PAXBP1 | 8.98E-03 | 8.84E-03 |
| 9461 | Gpr161 | GPR161 | 3.95E-02 | -7.80E-03 |
| 9462 | Ahrr | AHRR | -5.54E-02 | -1.36E-03 |
| 9463 | Glrb | GLRB | -1.61E-02 | 1.61E-03 |
| 9464 | Runx2 | RUNX2 | -3.66E-02 | 4.66E-02 |
| 9465 | D430041D05Rik | KIAA1549L | -4.09E-02 | -3.20E-02 |
| 9466 | Phldb3 | PHLDB3 | 1.92E-02 | -1.20E-02 |
| 9467 | Cebpe | CEBPE | 1.10E-02 | -2.05E-02 |
| 9468 | Tada2a | TADA2A | -1.01E-02 | -2.09E-02 |
| 9469 | Wdr35 | WDR35 | 1.11E-02 | -1.78E-02 |
| 9470 | Necab2 | NECAB2 | 1.33E-02 | 2.90E-03 |
| 9471 | Tecr | TECR | -8.99E-03 | -3.61E-03 |
| 9472 | Tial1 | TIAL1 | -8.43E-03 | 8.12E-03 |
| 9473 | Dpy19l3 | DPY19L3 | -2.10E-02 | -3.38E-02 |
| 9474 | Abracl | ABRACL | -1.41E-02 | 3.61E-02 |
| 9475 | Tbc1d8 | TBC1D8 | 2.59E-02 | 9.52E-03 |
| 9476 | Mgat4a | MGAT4A | 2.04E-02 | -3.10E-02 |
| 9477 | Nedd4l | NEDD4L | -1.02E-02 | -1.75E-02 |
| 9478 | Asah2 | ASAH2 | -1.18E-02 | -2.80E-02 |
| 9479 | Zcwpw2 | ZCWPW2 | -2.57E-02 | -3.32E-03 |
| 9480 | Cdc73 | CDC73 | 9.83E-03 | -5.92E-04 |
| 9481 | Mrpl42 | MRPL42 | 9.23E-03 | -1.11E-02 |
| 9482 | Klf3 | KLF3 | 1.42E-02 | 1.12E-02 |
| 9483 | Magohb | MAGOHB | 2.85E-02 | -5.13E-03 |
| 9484 | Myh9 | MYH9 | 8.94E-03 | 1.83E-02 |

| | | | | |
|---|---|---|---|---|
| 9485 | Gm960 | C11orf80 | -5.58E-02 | 6.37E-03 |
| 9486 | Gfi1 | GFI1 | -2.03E-02 | -1.42E-02 |
| 9487 | Sec13 | SEC13 | 1.22E-02 | -1.28E-02 |
| 9488 | Dnlz | DNLZ | -7.37E-03 | -3.04E-03 |
| 9489 | Cdh19 | CDH19 | -2.66E-02 | 3.94E-03 |
| 9490 | Vcpkmt | VCPKMT | 1.45E-02 | 6.22E-03 |
| 9491 | Aplp2 | APLP2 | 6.45E-03 | -1.72E-02 |
| 9492 | Sema3d | SEMA3D | -3.86E-02 | -2.60E-02 |
| 9493 | Med4 | MED4 | 8.65E-03 | 3.22E-04 |
| 9494 | Spo11 | SPO11 | 1.96E-02 | -4.56E-02 |
| 9495 | Hp1bp3 | HP1BP3 | 6.43E-03 | -1.32E-02 |
| 9496 | Fuom | FUOM | 1.22E-02 | -5.41E-04 |
| 9497 | Letmd1 | LETMD1 | -1.15E-02 | -3.96E-03 |
| 9498 | Lrp4 | LRP4 | -1.85E-02 | -4.26E-02 |
| 9499 | Pqlc3 | PQLC3 | 3.27E-02 | 3.67E-02 |
| 9500 | Rai1 | RAI1 | -1.08E-02 | 1.69E-02 |
| 9501 | Tmx1 | TMX1 | 1.04E-02 | 9.32E-03 |
| 9502 | Ptpn6 | PTPN6 | -1.04E-02 | -8.74E-03 |
| 9503 | Itsn2 | ITSN2 | -8.39E-03 | -6.92E-03 |
| 9504 | Jak1 | JAK1 | 1.12E-02 | -4.87E-03 |
| 9505 | Nop56 | NOP56 | 1.18E-02 | 2.82E-02 |
| 9506 | Sdhb | SDHB | 1.10E-02 | -1.91E-02 |
| 9507 | Lhx2 | LHX2 | -1.71E-02 | 3.85E-02 |
| 9508 | Wdyhv1 | WDYHV1 | -2.07E-02 | 2.12E-02 |
| 9509 | Cenpu | CENPU | 4.21E-02 | 1.19E-01 |
| 9510 | Ccne2 | CCNE2 | 6.06E-02 | 8.53E-02 |
| 9511 | Nudt4 | NUDT4 | -1.11E-02 | -9.58E-03 |
| 9512 | Myo3a | MYO3A | -1.39E-02 | -2.24E-02 |
| 9513 | Srp68 | SRP68 | -7.16E-03 | -1.44E-02 |
| 9514 | Unc5c | UNC5C | -4.19E-02 | -9.17E-02 |
| 9515 | Maoa | MAOA | 1.64E-02 | 4.92E-03 |
| 9516 | Arhgap36 | ARHGAP36 | 5.22E-02 | 1.99E-01 |
| 9517 | Fam166b | FAM166B | 1.94E-02 | -3.81E-02 |
| 9518 | Spr | SPR | 1.28E-02 | -8.34E-03 |
| 9519 | Dact3 | DACT3 | 3.51E-02 | 6.91E-02 |
| 9520 | Pcolce | PCOLCE | 1.23E-02 | 4.52E-02 |
| 9521 | Grk4 | GRK4 | 1.95E-02 | 2.10E-02 |
| 9522 | Wdr82 | WDR82 | -1.08E-02 | 4.46E-03 |
| 9523 | Yme1l1 | YME1L1 | 7.64E-03 | -5.41E-03 |
| 9524 | Ubxn1 | UBXN1 | 6.90E-03 | -7.78E-03 |
| 9525 | Frg1 | FRG1 | -1.09E-02 | 1.86E-03 |
| 9526 | Polb | POLB | -1.54E-02 | -3.72E-03 |
| 9527 | Mrpl52 | MRPL52 | 1.09E-02 | -1.05E-02 |
| 9528 | Rbm19 | RBM19 | 1.23E-02 | 5.72E-03 |
| 9529 | Podn | PODN | 1.85E-02 | -3.64E-02 |
| 9530 | Hdgf | HDGF | 8.21E-03 | -3.64E-03 |
| 9531 | Id1 | ID1 | -5.20E-02 | 1.22E-01 |
| 9532 | Zfp605 | ZNF605 | 1.99E-02 | -3.80E-02 |
| 9533 | Pds5a | PDS5A | 8.95E-03 | -2.36E-03 |
| 9534 | Paox | PAOX | 1.55E-02 | -1.25E-02 |
| 9535 | Bcl2a1a | BCL2A1 | -3.01E-02 | 9.50E-02 |
| 9536 | Ugt1a1 | UGT1A1 | -5.63E-02 | 5.34E-02 |
| 9537 | Rpl14 | RPL14 | -8.08E-03 | 1.05E-02 |

| | | | | |
|---|---|---|---|---|
| 9538 | Il1rap | IL1RAP | 1.98E-02 | -1.04E-02 |
| 9539 | Tgds | TGDS | -1.13E-02 | -5.57E-03 |
| 9540 | Sox6 | SOX6 | -1.40E-02 | -3.67E-02 |
| 9541 | Pkhd1l1 | PKHD1L1 | -3.16E-02 | 2.62E-02 |
| 9542 | Kansl1 | KANSL1 | -8.46E-03 | -3.44E-03 |
| 9543 | Tub | TUB | -2.30E-02 | -2.71E-02 |
| 9544 | Bscl2 | BSCL2 | 9.19E-03 | -2.06E-02 |
| 9545 | Smc1a | SMC1A | -7.95E-03 | 2.33E-03 |
| 9546 | Insig2 | INSIG2 | 7.28E-03 | -5.77E-03 |
| 9547 | Sigirr | SIGIRR | 1.26E-02 | -8.38E-03 |
| 9548 | Gucy2f | GUCY2F | -3.65E-02 | -1.47E-02 |
| 9549 | Srp19 | SRP19 | -1.05E-02 | 2.46E-02 |
| 9550 | Tpp2 | TPP2 | -7.66E-03 | 1.53E-02 |
| 9551 | Ccdc59 | CCDC59 | 9.72E-03 | -1.64E-02 |
| 9552 | Lrrc24 | LRRC24 | 1.33E-02 | -2.64E-02 |
| 9553 | Dctn6 | DCTN6 | 7.84E-03 | -2.20E-02 |
| 9554 | Hoxd4 | HOXD4 | -2.91E-02 | -4.33E-03 |
| 9555 | Mael | MAEL | 6.98E-03 | -8.48E-03 |
| 9556 | Mrps12 | MRPS12 | 1.15E-02 | -1.67E-02 |
| 9557 | Klhl6 | KLHL6 | -2.64E-02 | 2.07E-03 |
| 9558 | Rab11fip2 | RAB11FIP2 | 9.68E-03 | -3.06E-02 |
| 9559 | Clcnkb | CLCNKA | 1.36E-02 | 1.80E-02 |
| 9560 | Slc19a3 | SLC19A3 | -1.78E-02 | 1.34E-02 |
| 9561 | Rassf4 | RASSF4 | 2.15E-02 | 9.84E-03 |
| 9562 | Lrrc14b | LRRC14B | -3.89E-02 | 3.16E-03 |
| 9563 | Nnat | NNAT | 1.37E-01 | 1.69E-01 |
| 9564 | Pld3 | PLD3 | 7.08E-03 | -2.11E-03 |
| 9565 | Lig4 | LIG4 | 1.75E-02 | 2.43E-02 |
| 9566 | Pdpn | PDPN | -2.40E-02 | 4.94E-02 |
| 9567 | Cpt1a | CPT1A | 1.71E-02 | -1.12E-03 |
| 9568 | Gnaq | GNAQ | 5.73E-03 | -6.89E-03 |
| 9569 | Zfp869 | ZNF91 | -1.24E-02 | -2.54E-02 |
| 9570 | Ms4a4a | MS4A4A | 5.20E-02 | 1.51E-01 |
| 9571 | Tff2 | TFF2 | -1.18E-02 | 3.16E-02 |
| 9572 | Tubgcp3 | TUBGCP3 | 6.75E-03 | 8.50E-04 |
| 9573 | Tmem168 | TMEM168 | -1.01E-02 | -1.31E-03 |
| 9574 | Rnh1 | RNH1 | 8.16E-03 | 1.04E-02 |
| 9575 | Ermap | ERMAP | 2.39E-02 | 1.03E-02 |
| 9576 | Gpr19 | GPR19 | 2.68E-02 | -1.45E-02 |
| 9577 | Cdkn2b | CDKN2B | 1.17E-01 | -3.10E-02 |
| 9578 | Kiss1r | KISS1R | -2.45E-02 | -8.44E-03 |
| 9579 | Pparg | PPARG | 6.23E-02 | 4.76E-02 |
| 9580 | Rap2a | RAP2A | 1.07E-02 | 1.55E-02 |
| 9581 | Actrt3 | ACTRT3 | 1.77E-02 | -2.61E-02 |
| 9582 | Pcdhga9 | PCDHGA9 | -2.59E-02 | -1.05E-02 |
| 9583 | Smg7 | SMG7 | 1.13E-02 | -7.03E-03 |
| 9584 | Zfp287 | ZNF287 | -3.12E-02 | -2.11E-02 |
| 9585 | Nckap5l | NCKAP5L | 2.16E-02 | 3.68E-02 |
| 9586 | Hoxb7 | HOXB7 | -1.85E-02 | 1.09E-02 |
| 9587 | Fam13c | FAM13C | 2.73E-02 | -1.17E-02 |
| 9588 | Lypd1 | LYPD1 | -1.44E-02 | 2.30E-02 |
| 9589 | Tmem86b | TMEM86B | -1.90E-02 | -2.78E-02 |
| 9590 | Cnot10 | CNOT10 | 6.29E-03 | -7.08E-03 |

| | | | | |
|---|---|---|---|---|
| 9591 | Rint1 | RINT1 | -9.92E-03 | -1.14E-02 |
| 9592 | Dexi | DEXI | 1.40E-02 | -2.81E-03 |
| 9593 | Med8 | MED8 | 9.97E-03 | 6.60E-03 |
| 9594 | Dlst | DLST | 1.12E-02 | -1.76E-02 |
| 9595 | Clec11a | CLEC11A | -2.96E-02 | -8.55E-03 |
| 9596 | Nat8l | NAT8L | -4.63E-02 | 5.47E-02 |
| 9597 | Stmn2 | STMN2 | -3.12E-02 | 7.10E-02 |
| 9598 | Ccnt2 | CCNT2 | -8.05E-03 | -1.51E-02 |
| 9599 | Cyp4a32 | CYP4A11 | -4.93E-02 | 6.15E-03 |
| 9600 | Thumpd2 | THUMPD2 | 1.93E-02 | 1.58E-02 |
| 9601 | Rem1 | REM1 | -5.45E-02 | -3.97E-02 |
| 9602 | Slc45a4 | SLC45A4 | -1.07E-02 | -7.48E-03 |
| 9603 | Wbp4 | WBP4 | 8.96E-03 | -1.08E-02 |
| 9604 | Tjap1 | TJAP1 | 1.03E-02 | -9.45E-03 |
| 9605 | Ganab | GANAB | -7.52E-03 | -6.01E-03 |
| 9606 | Slc9a3r2 | SLC9A3R2 | 1.26E-02 | -6.84E-03 |
| 9607 | Ccdc17 | CCDC17 | -2.10E-02 | -2.78E-02 |
| 9608 | 1600014C10Rik | C19orf12 | 1.53E-02 | -3.53E-02 |
| 9609 | Ankzf1 | ANKZF1 | -1.22E-02 | -2.39E-02 |
| 9610 | Lcorl | LCORL | 1.39E-02 | 8.55E-03 |
| 9611 | Dus4l | DUS4L | -1.81E-02 | 1.58E-02 |
| 9612 | Elovl7 | ELOVL7 | -1.40E-02 | -1.54E-02 |
| 9613 | Psd2 | PSD2 | 2.96E-02 | 1.59E-02 |
| 9614 | Acot10 | ACOT9 | 2.60E-02 | 3.82E-02 |
| 9615 | Slc25a39 | SLC25A39 | 1.07E-02 | -1.09E-02 |
| 9616 | Eif2b4 | EIF2B4 | 1.02E-02 | 3.21E-03 |
| 9617 | Glis3 | GLIS3 | -1.47E-02 | -4.23E-02 |
| 9618 | Mef2b | MEF2B | -1.99E-02 | 3.97E-03 |
| 9619 | Reln | RELN | -7.07E-02 | -4.75E-02 |
| 9620 | Prmt2 | PRMT2 | -2.49E-02 | -1.06E-02 |
| 9621 | Gsta3 | GSTA3 | 2.71E-02 | -1.42E-02 |
| 9622 | Klhl41 | KLHL41 | 2.37E-02 | -1.59E-02 |
| 9623 | Slc5a12 | SLC5A12 | -3.07E-02 | 1.55E-02 |
| 9624 | Sgcg | SGCG | 3.55E-02 | 4.61E-02 |
| 9625 | Slc2a10 | SLC2A10 | 3.83E-02 | 1.14E-02 |
| 9626 | Lamb3 | LAMB3 | 1.27E-01 | 1.03E-01 |
| 9627 | Cerkl | CERKL | -2.38E-02 | 1.75E-02 |
| 9628 | Tollip | TOLLIP | 8.81E-03 | -1.18E-02 |
| 9629 | Tmbim1 | TMBIM1 | 1.85E-02 | 2.59E-02 |
| 9630 | Mroh7 | MROH7 | -2.99E-02 | 7.02E-03 |
| 9631 | Ccdc158 | CCDC158 | 2.81E-02 | -2.60E-02 |
| 9632 | Dpy30 | DPY30 | 8.41E-03 | -7.93E-03 |
| 9633 | Anks4b | ANKS4B | 1.53E-02 | 9.39E-03 |
| 9634 | Tusc1 | TUSC1 | 2.43E-02 | 2.55E-02 |
| 9635 | Fam184a | FAM184A | 2.44E-02 | -2.19E-02 |
| 9636 | Dock5 | DOCK5 | 1.10E-02 | -6.52E-03 |
| 9637 | Hdac2 | HDAC2 | 7.14E-03 | 3.08E-03 |
| 9638 | Clic5 | CLIC5 | -1.92E-02 | -3.94E-02 |
| 9639 | N4bp2l1 | N4BP2L1 | 2.11E-02 | -1.50E-02 |
| 9640 | Rab3d | RAB3D | -2.11E-02 | 2.92E-02 |
| 9641 | Rnaseh1 | RNASEH1 | 1.11E-02 | 8.52E-03 |
| 9642 | Vps9d1 | VPS9D1 | 8.25E-03 | -1.12E-02 |
| 9643 | Gzmm | GZMM | -4.43E-02 | -4.31E-02 |

| | | | | |
|---|---|---|---|---|
| 9644 | Slc25a51 | SLC25A51 | -9.82E-03 | -7.84E-03 |
| 9645 | Pdgfra | PDGFRA | 3.41E-02 | -4.85E-02 |
| 9646 | Dvl2 | DVL2 | -1.69E-02 | 4.55E-03 |
| 9647 | Wnt9b | WNT9B | 1.27E-01 | -9.79E-02 |
| 9648 | Leng9 | LENG9 | -2.31E-02 | 2.63E-02 |
| 9649 | Polr2a | POLR2A | 1.08E-02 | -8.00E-03 |
| 9650 | Cpvl | CPVL | -1.79E-02 | -5.37E-03 |
| 9651 | Ncdn | NCDN | 9.39E-03 | 1.00E-02 |
| 9652 | Rpl19 | RPL19 | 8.41E-03 | 1.79E-03 |
| 9653 | Reep2 | REEP2 | 3.51E-02 | 1.35E-02 |
| 9654 | Tyk2 | TYK2 | -8.64E-03 | 4.01E-03 |
| 9655 | St6galnac5 | ST6GALNAC5 | -4.79E-02 | -1.16E-01 |
| 9656 | Pkd1l2 | PKD1L2 | -2.08E-02 | -3.58E-02 |
| 9657 | Gsta2 | GSTA5 | 2.75E-02 | -7.75E-03 |
| 9658 | Rnf138 | RNF138 | 1.50E-02 | 2.56E-03 |
| 9659 | Lrrc19 | LRRC19 | 1.96E-02 | -1.12E-02 |
| 9660 | Slurp1 | SLURP1 | -1.27E-02 | 1.88E-02 |
| 9661 | Shroom4 | SHROOM4 | 1.21E-02 | -1.68E-02 |
| 9662 | Uvssa | UVSSA | 1.51E-02 | -1.07E-02 |
| 9663 | Mrps21 | MRPS21 | -9.87E-03 | -9.50E-03 |
| 9664 | Cyp2b10 | CYP2B6 | 1.97E-02 | 1.99E-02 |
| 9665 | Car12 | CA12 | 1.77E-02 | -7.16E-03 |
| 9666 | Rftn2 | RFTN2 | -3.41E-02 | -3.14E-02 |
| 9667 | Spc25 | SPC25 | 3.40E-02 | 3.90E-02 |
| 9668 | Ect2l | ECT2L | 1.41E-02 | -6.55E-03 |
| 9669 | Sipa1l3 | SIPA1L3 | -8.38E-03 | -1.58E-02 |
| 9670 | Tbc1d30 | TBC1D30 | 2.46E-02 | -4.62E-02 |
| 9671 | Srgap2 | SRGAP2 | 9.19E-03 | -8.92E-03 |
| 9672 | Zfp457 | ZNF93 | -3.51E-02 | -1.90E-02 |
| 9673 | Lrrc18 | LRRC18 | -2.43E-02 | 3.49E-03 |
| 9674 | Rfx3 | RFX3 | -1.72E-02 | -9.07E-03 |
| 9675 | Eif3j1 | EIF3J | 1.05E-02 | 1.40E-02 |
| 9676 | Cd2bp2 | CD2BP2 | 5.71E-03 | -8.92E-03 |
| 9677 | Lsm14a | LSM14A | -5.60E-03 | -2.48E-03 |
| 9678 | Polr2h | POLR2H | -1.58E-02 | 6.48E-03 |
| 9679 | Elf3 | ELF3 | 5.65E-02 | 8.97E-02 |
| 9680 | Capn9 | CAPN9 | 1.15E-02 | -8.47E-04 |
| 9681 | Zfp90 | ZFP90 | 1.77E-02 | -1.60E-02 |
| 9682 | Ttll6 | TTLL6 | 1.43E-02 | -1.82E-02 |
| 9683 | Foxp4 | FOXP4 | 1.31E-02 | 3.33E-02 |
| 9684 | Akr1b3 | AKR1B1 | 9.93E-02 | 7.93E-02 |
| 9685 | Upk3b | UPK3B | 1.89E-01 | 8.97E-02 |
| 9686 | Smarcc1 | SMARCC1 | 6.68E-03 | 8.21E-04 |
| 9687 | Pitpnm2 | PITPNM2 | 1.62E-02 | 5.27E-03 |
| 9688 | Tspan8 | TSPAN8 | -2.29E-02 | 1.04E-02 |
| 9689 | Cep57l1 | CEP57L1 | 1.75E-02 | -1.98E-02 |
| 9690 | Hsf2bp | HSF2BP | -3.85E-02 | -5.68E-02 |
| 9691 | Trim17 | TRIM17 | -3.73E-02 | -2.13E-02 |
| 9692 | Pfas | PFAS | -1.05E-02 | -3.90E-02 |
| 9693 | Ttk | TTK | 6.28E-02 | 2.11E-01 |
| 9694 | Eif1ax | EIF1AY | 8.22E-03 | -6.80E-03 |
| 9695 | Tgm3 | TGM3 | 1.29E-02 | 4.37E-04 |
| 9696 | Smox | SMOX | 2.09E-02 | 7.13E-02 |

| | | | | |
|---|---|---|---|---|
| 9697 | Btnl9 | BTNL9 | -4.33E-02 | -5.54E-02 |
| 9698 | Tm2d2 | TM2D2 | -8.77E-03 | -1.63E-02 |
| 9699 | Galr3 | GALR3 | 2.51E-02 | -1.92E-03 |
| 9700 | Acin1 | ACIN1 | 5.34E-03 | -1.06E-02 |
| 9701 | Ptprk | PTPRK | 1.13E-02 | -4.75E-02 |
| 9702 | Agpat2 | AGPAT2 | 1.26E-02 | 1.47E-02 |
| 9703 | Wdr18 | WDR18 | -9.02E-03 | -1.07E-02 |
| 9704 | Ush1g | USH1G | -1.40E-02 | 1.56E-02 |
| 9705 | Rnf207 | RNF207 | 2.39E-02 | 2.85E-02 |
| 9706 | Mst1r | MST1R | -4.18E-02 | 2.72E-03 |
| 9707 | Megf10 | MEGF10 | -1.64E-02 | 4.48E-02 |
| 9708 | Baiap2 | BAIAP2 | -1.49E-02 | -3.27E-03 |
| 9709 | Cmss1 | CMSS1 | 1.72E-02 | -3.11E-02 |
| 9710 | Selenbp1 | SELENBP1 | 1.64E-02 | -1.90E-02 |
| 9711 | Rgs1 | RGS1 | -5.53E-02 | 1.17E-01 |
| 9712 | Srpr | SRPR | 7.70E-03 | -1.20E-02 |
| 9713 | Pmepa1 | PMEPA1 | 1.27E-02 | 5.69E-02 |
| 9714 | Kdelr2 | KDELR2 | 1.14E-02 | 2.07E-02 |
| 9715 | Cldn17 | CLDN17 | 1.49E-02 | -2.03E-02 |
| 9716 | Decr2 | DECR2 | -2.19E-02 | -1.12E-02 |
| 9717 | Mmp9 | MMP9 | -4.65E-02 | 4.21E-02 |
| 9718 | Pkn1 | PKN1 | 7.64E-03 | -1.14E-02 |
| 9719 | Cldn7 | CLDN7 | -2.23E-02 | 5.61E-02 |
| 9720 | Taf1b | TAF1B | 1.72E-02 | -2.20E-02 |
| 9721 | Zfp36l2 | ZFP36L2 | -1.40E-02 | 4.27E-03 |
| 9722 | Skil | SKIL | 1.79E-02 | 3.21E-02 |
| 9723 | Naa30 | NAA30 | -1.30E-02 | 2.94E-03 |
| 9724 | Tram1 | TRAM1 | 7.81E-03 | 1.08E-03 |
| 9725 | Tg | TG | 2.12E-02 | -2.55E-03 |
| 9726 | Grhl3 | GRHL3 | 9.95E-02 | 5.81E-02 |
| 9727 | Slc6a3 | SLC6A3 | 1.79E-02 | -2.11E-02 |
| 9728 | Sirpb1b | SIRPB1 | -2.11E-02 | 1.17E-01 |
| 9729 | Rps25 | RPS25 | 8.37E-03 | -6.40E-03 |
| 9730 | Aqp1 | AQP1 | -1.41E-02 | -5.63E-02 |
| 9731 | L3mbtl4 | L3MBTL4 | -1.83E-02 | -3.52E-02 |
| 9732 | Bnc2 | BNC2 | -4.12E-02 | -1.64E-02 |
| 9733 | Trappc2l | TRAPPC2L | 9.34E-03 | 2.42E-03 |
| 9734 | Tbpl2 | TBPL2 | -2.55E-02 | -7.70E-03 |
| 9735 | Ndst2 | NDST2 | 9.60E-03 | -6.63E-03 |
| 9736 | Dnajb7 | DNAJB7 | -2.23E-02 | -7.45E-03 |
| 9737 | Tradd | TRADD | 1.24E-02 | 5.90E-03 |
| 9738 | Cpn1 | CPN1 | -2.13E-02 | 4.63E-02 |
| 9739 | Rps15a | RPS15A | -7.55E-03 | 1.27E-02 |
| 9740 | Plxnc1 | PLXNC1 | -3.17E-02 | -1.62E-02 |
| 9741 | Fkrp | FKRP | 7.58E-03 | 7.45E-03 |
| 9742 | Slc30a7 | SLC30A7 | 7.58E-03 | 2.43E-03 |
| 9743 | Chil1 | CHI3L1 | 3.71E-02 | -1.38E-02 |
| 9744 | Galnt2 | GALNT2 | 2.64E-02 | -3.19E-02 |
| 9745 | Ttc34 | TTC34 | -1.21E-02 | 7.91E-03 |
| 9746 | Mrpl48 | MRPL48 | 8.41E-03 | -2.08E-02 |
| 9747 | Hsd3b2 | HSD3B1 | -1.57E-02 | -2.53E-02 |
| 9748 | Trim27 | TRIM27 | 1.12E-02 | 2.78E-02 |
| 9749 | Eno4 | ENO4 | 2.73E-02 | 1.76E-03 |

| | | | | |
|---|---|---|---|---|
| 9750 | Nupr1 | NUPR1 | 8.43E-02 | 9.07E-02 |
| 9751 | Crebl2 | CREBL2 | 1.02E-02 | -2.27E-02 |
| 9752 | Aptx | APTX | -9.20E-03 | -1.74E-02 |
| 9753 | Coa3 | COA3 | 9.09E-03 | -2.12E-02 |
| 9754 | Foxl1 | FOXL1 | -2.82E-02 | -2.02E-02 |
| 9755 | P2rx7 | P2RX7 | -3.56E-02 | 4.07E-02 |
| 9756 | Plch2 | PLCH2 | 1.06E-01 | 1.89E-02 |
| 9757 | Sh2b3 | SH2B3 | 1.25E-02 | 1.14E-03 |
| 9758 | Sgcz | SGCZ | -4.85E-02 | -3.85E-02 |
| 9759 | Pea15a | PEA15 | 1.17E-02 | 4.28E-02 |
| 9760 | Lat2 | LAT2 | 2.78E-02 | 7.52E-02 |
| 9761 | Ccdc12 | CCDC12 | 9.07E-03 | -9.67E-03 |
| 9762 | Prcp | PRCP | 1.67E-02 | 5.61E-02 |
| 9763 | Dnajb13 | DNAJB13 | 1.42E-02 | 7.07E-02 |
| 9764 | Drg2 | DRG2 | 8.56E-03 | -6.52E-03 |
| 9765 | Pcdha10 | PCDHA12 | -8.99E-03 | -1.32E-03 |
| 9766 | Acbd4 | ACBD4 | 1.42E-02 | -1.72E-02 |
| 9767 | Rnf111 | RNF111 | 8.31E-03 | 3.42E-03 |
| 9768 | Hexdc | HEXDC | -1.29E-02 | -3.15E-02 |
| 9769 | Smarcb1 | SMARCB1 | 9.56E-03 | 1.84E-03 |
| 9770 | Zfp113 | ZNF3 | -1.64E-02 | -3.49E-02 |
| 9771 | Klhl12 | KLHL12 | 9.39E-03 | -1.20E-02 |
| 9772 | Rab33b | RAB33B | 1.00E-02 | 1.76E-02 |
| 9773 | Efcab6 | EFCAB6 | -2.01E-02 | -1.63E-03 |
| 9774 | Casp9 | CASP9 | -8.63E-03 | -1.66E-02 |
| 9775 | Gulp1 | GULP1 | 2.36E-02 | -1.97E-03 |
| 9776 | BC005624 | C9orf78 | -6.93E-03 | -1.71E-02 |
| 9777 | Brix1 | BRIX1 | 8.66E-03 | 7.54E-03 |
| 9778 | Kdm5b | KDM5B | -1.11E-02 | 2.20E-04 |
| 9779 | Tas1r3 | TAS1R3 | -2.83E-02 | -4.81E-02 |
| 9780 | Abca4 | ABCA4 | -4.69E-02 | -1.48E-01 |
| 9781 | Galnt16 | GALNT16 | 7.97E-02 | 7.90E-02 |
| 9782 | Hdx | HDX | 2.96E-02 | -1.38E-02 |
| 9783 | Fam161b | FAM161B | -3.16E-02 | -4.97E-03 |
| 9784 | Ddx5 | DDX5 | -8.43E-03 | -4.14E-03 |
| 9785 | Fam98c | FAM98C | 1.04E-02 | -8.23E-04 |
| 9786 | Dyrk4 | DYRK4 | -1.26E-02 | 9.66E-03 |
| 9787 | Supt16 | SUPT16H | 5.73E-03 | -4.98E-03 |
| 9788 | Tango2 | TANGO2 | -1.67E-02 | -4.25E-02 |
| 9789 | Klrb1c | KLRB1 | -2.17E-02 | -1.09E-02 |
| 9790 | Ccpg1 | CCPG1 | -1.92E-02 | 1.51E-02 |
| 9791 | Dnd1 | DND1 | -4.06E-02 | 2.78E-02 |
| 9792 | Grk5 | GRK5 | 2.36E-02 | -8.84E-03 |
| 9793 | Zbtb5 | ZBTB5 | 1.37E-02 | -1.29E-03 |
| 9794 | Ces2h | CES2 | -4.53E-02 | -1.79E-02 |
| 9795 | Thoc6 | THOC6 | 1.42E-02 | 1.44E-02 |
| 9796 | Smpx | SMPX | -3.05E-02 | -5.88E-02 |
| 9797 | Lypla2 | LYPLA2 | 6.68E-03 | -4.91E-03 |
| 9798 | Proser2 | PROSER2 | 1.20E-01 | -1.35E-02 |
| 9799 | She | SHE | 2.57E-02 | -2.18E-02 |
| 9800 | Zfp609 | ZNF609 | -1.04E-02 | -5.71E-03 |
| 9801 | Pik3r2 | PIK3R2 | 1.28E-02 | -2.42E-02 |
| 9802 | Ino80d | INO80D | -1.04E-02 | 5.21E-03 |

| | | | | |
|---|---|---|---|---|
| 9803 | Rbms3 | RBMS3 | -4.62E-02 | -2.98E-02 |
| 9804 | Ovol3 | OVOL3 | 1.50E-02 | -1.98E-02 |
| 9805 | Fam221b | FAM221B | -1.63E-02 | 8.21E-03 |
| 9806 | Unc50 | UNC50 | 6.43E-03 | 9.55E-04 |
| 9807 | Cd6 | CD6 | -3.37E-02 | 8.82E-03 |
| 9808 | Nuak2 | NUAK2 | 4.14E-02 | 3.67E-02 |
| 9809 | Tbc1d31 | TBC1D31 | 2.17E-02 | 1.66E-02 |
| 9810 | Zcchc12 | ZCCHC12 | -3.16E-02 | -4.41E-02 |
| 9811 | Tsnax | TSNAX | 5.94E-03 | -1.85E-03 |
| 9812 | Serpinb10 | SERPINB10 | 2.96E-02 | 8.08E-03 |
| 9813 | Cmbl | CMBL | 1.80E-02 | -2.40E-02 |
| 9814 | Tdp1 | TDP1 | -1.56E-02 | 1.02E-02 |
| 9815 | Acaca | ACACA | -1.47E-02 | -8.24E-03 |
| 9816 | Ttc12 | TTC12 | -1.35E-02 | 6.89E-03 |
| 9817 | Ihh | IHH | -2.35E-02 | -2.32E-02 |
| 9818 | Ogn | OGN | 2.91E-02 | -7.21E-02 |
| 9819 | Cnih4 | CNIH4 | 7.81E-03 | 2.26E-02 |
| 9820 | Ddc | DDC | 1.99E-02 | -8.67E-02 |
| 9821 | Fhl5 | FHL5 | 1.69E-02 | 3.82E-02 |
| 9822 | Bbx | BBX | 1.01E-02 | -1.02E-02 |
| 9823 | Prdm6 | PRDM6 | -1.33E-02 | 4.67E-03 |
| 9824 | Tm7sf3 | TM7SF3 | -1.12E-02 | -1.77E-02 |
| 9825 | Pced1b | PCED1B | 3.15E-02 | -4.96E-02 |
| 9826 | Rbmx | RBMX | 7.17E-03 | 4.67E-03 |
| 9827 | Wnt7b | WNT7B | 1.14E-01 | -7.40E-02 |
| 9828 | Apbb2 | APBB2 | -1.47E-02 | -3.97E-02 |
| 9829 | Fam110a | FAM110A | 3.07E-02 | 3.97E-02 |
| 9830 | Slc35f2 | SLC35F2 | 1.09E-02 | -1.71E-02 |
| 9831 | Ticrr | TICRR | 6.35E-02 | 1.63E-01 |
| 9832 | Dmp1 | DMP1 | 1.11E-02 | 2.57E-02 |
| 9833 | Cnn2 | CNN2 | 1.90E-02 | 1.88E-02 |
| 9834 | Blvra | BLVRA | 1.57E-02 | -7.60E-03 |
| 9835 | Mgll | MGLL | 1.39E-02 | -4.76E-02 |
| 9836 | Cklf | CKLF | -2.88E-02 | 7.59E-02 |
| 9837 | Zkscan17 | ZNF496 | 1.82E-02 | -5.05E-03 |
| 9838 | Sh3bgrl | SH3BGRL | -8.76E-03 | 3.31E-03 |
| 9839 | Mthfs | MTHFS | -2.59E-02 | 4.58E-02 |
| 9840 | Hagh | HAGH | 9.79E-03 | -9.34E-03 |
| 9841 | Hcn4 | HCN4 | 1.57E-02 | 3.18E-03 |
| 9842 | Synm | SYNM | -4.54E-02 | 3.46E-02 |
| 9843 | Glrx2 | GLRX2 | 7.81E-03 | -2.16E-02 |
| 9844 | Ankle1 | ANKLE1 | 6.55E-02 | 1.99E-01 |
| 9845 | Sv2a | SV2A | 1.22E-02 | -2.17E-02 |
| 9846 | Nsfl1c | NSFL1C | 7.84E-03 | -1.03E-02 |
| 9847 | Xbp1 | XBP1 | -1.93E-02 | 1.82E-02 |
| 9848 | Fam83h | FAM83H | 9.97E-03 | 9.39E-03 |
| 9849 | Stard10 | STARD10 | 1.09E-02 | -1.49E-02 |
| 9850 | Foxa3 | FOXA3 | 1.48E-02 | 8.09E-03 |
| 9851 | Acap3 | ACAP3 | 1.34E-02 | 6.21E-03 |
| 9852 | Flot1 | FLOT1 | 1.04E-02 | 1.26E-02 |
| 9853 | Rock1 | ROCK1 | 7.27E-03 | -4.20E-04 |
| 9854 | Tsen15 | TSEN15 | -1.40E-02 | 1.62E-03 |
| 9855 | Gprc5b | GPRC5B | 4.77E-02 | 1.71E-02 |

| | | | | |
|---|---|---|---|---|
| 9856 | Tslp | TSLP | -3.58E-02 | 2.21E-02 |
| 9857 | Acbd3 | ACBD3 | -1.09E-02 | -2.20E-03 |
| 9858 | Supt6 | SUPT6H | -6.40E-03 | -9.63E-03 |
| 9859 | Mob1a | MOB1A | -8.17E-03 | 1.09E-02 |
| 9860 | Taf6 | TAF6 | -6.31E-03 | -2.85E-04 |
| 9861 | Pkd2 | PKD2 | -9.24E-03 | -4.03E-02 |
| 9862 | Klk1b22 | KLK3 | 2.98E-02 | 9.42E-02 |
| 9863 | Nos1 | NOS1 | -2.77E-02 | 1.65E-02 |
| 9864 | AU022252 | C1orf50 | 7.92E-03 | -2.46E-02 |
| 9865 | Ccdc87 | CCDC87 | 2.07E-02 | 5.10E-02 |
| 9866 | Tspear | TSPEAR | -2.72E-02 | 3.49E-02 |
| 9867 | Rufy2 | RUFY2 | 9.32E-03 | -2.23E-02 |
| 9868 | Hectd3 | HECTD3 | 8.69E-03 | -2.33E-02 |
| 9869 | Spef2 | SPEF2 | 2.98E-02 | 1.18E-02 |
| 9870 | Cdc27 | CDC27 | 8.10E-03 | 3.08E-03 |
| 9871 | Pdzk1ip1 | PDZK1IP1 | -1.10E-02 | -1.64E-02 |
| 9872 | Tcf21 | TCF21 | -5.60E-02 | -2.98E-03 |
| 9873 | Nudt12 | NUDT12 | 1.26E-02 | -3.49E-03 |
| 9874 | Cdk6 | CDK6 | -1.21E-02 | 3.03E-02 |
| 9875 | Clec2l | CLEC2L | 3.42E-02 | 2.66E-02 |
| 9876 | Hyou1 | HYOU1 | 2.22E-02 | -8.53E-03 |
| 9877 | Pcsk6 | PCSK6 | 6.42E-02 | -1.50E-01 |
| 9878 | Nr2f6 | NR2F6 | -8.88E-03 | 9.85E-03 |
| 9879 | Slc17a8 | SLC17A8 | 2.25E-02 | -1.05E-02 |
| 9880 | Plekhm1 | PLEKHM1 | -7.81E-03 | 1.45E-03 |
| 9881 | Ip6k1 | IP6K1 | -6.98E-03 | -1.35E-02 |
| 9882 | Bche | BCHE | 6.19E-02 | 4.56E-02 |
| 9883 | Vldlr | VLDLR | -9.66E-03 | -2.60E-02 |
| 9884 | Ncoa1 | NCOA1 | -8.17E-03 | -3.77E-02 |
| 9885 | Rela | RELA | 1.11E-02 | 1.79E-02 |
| 9886 | Tpbg | TPBG | 4.54E-02 | -1.04E-02 |
| 9887 | Ptpmt1 | PTPMT1 | -1.21E-01 | 1.77E-01 |
| 9888 | Gmps | GMPS | 7.71E-03 | 1.93E-03 |
| 9889 | Ackr2 | ACKR2 | 3.70E-02 | 3.29E-02 |
| 9890 | Cdk2ap1 | CDK2AP1 | 1.19E-02 | 1.84E-02 |
| 9891 | Pilra | PILRA | -6.23E-02 | 1.94E-01 |
| 9892 | Umps | UMPS | -1.37E-02 | -6.98E-03 |
| 9893 | Pprc1 | PPRC1 | -1.98E-02 | 4.87E-02 |
| 9894 | Prr5 | PRR5 | 2.57E-02 | 3.13E-02 |
| 9895 | Enpep | ENPEP | 2.22E-02 | -3.02E-03 |
| 9896 | Gnrh1 | GNRH1 | 3.97E-02 | 3.30E-02 |
| 9897 | Rpl18a | RPL18A | -8.48E-03 | 1.31E-02 |
| 9898 | Slc4a8 | SLC4A8 | -2.97E-02 | 1.74E-03 |
| 9899 | Bach1 | BACH1 | 1.80E-02 | 3.44E-02 |
| 9900 | Utp6 | UTP6 | -6.33E-03 | -1.86E-03 |
| 9901 | Chd6 | CHD6 | -7.03E-03 | -1.79E-02 |
| 9902 | Slc27a5 | SLC27A5 | -4.01E-02 | 5.20E-02 |
| 9903 | Insl5 | INSL5 | 1.10E-02 | 1.16E-04 |
| 9904 | Tac2 | TAC3 | 8.49E-03 | 7.39E-03 |
| 9905 | Vasp | VASP | 1.40E-02 | 3.24E-02 |
| 9906 | Ccr8 | CCR8 | -9.99E-03 | -7.11E-05 |
| 9907 | Apol8 | APOL2 | 2.43E-02 | -4.08E-03 |
| 9908 | Eral1 | ERAL1 | -9.84E-03 | -2.78E-02 |

| | | | | |
|---|---|---|---|---|
| 9909 | Nudt16 | NUDT16 | -2.37E-02 | -6.74E-03 |
| 9910 | Klk1b21 | KLK3 | 2.29E-02 | 1.55E-03 |
| 9911 | Ubald2 | UBALD2 | 1.76E-02 | 6.08E-03 |
| 9912 | Phactr1 | PHACTR1 | 3.32E-02 | 4.51E-02 |
| 9913 | Ccdc91 | CCDC91 | 9.74E-03 | -1.79E-02 |
| 9914 | Relt | RELT | 3.98E-02 | 1.15E-01 |
| 9915 | Usp1 | USP1 | -7.06E-03 | 3.67E-03 |
| 9916 | Pja2 | PJA2 | 6.65E-03 | -1.99E-02 |
| 9917 | Orc3 | ORC3 | 9.44E-03 | -1.99E-02 |
| 9918 | Rplp2 | RPLP2 | -8.15E-03 | -1.26E-03 |
| 9919 | Dennd2d | DENND2D | -3.33E-02 | 1.92E-02 |
| 9920 | Guca2a | GUCA2A | -2.80E-02 | 9.22E-02 |
| 9921 | Prkacb | PRKACB | -9.34E-03 | -1.25E-02 |
| 9922 | Anxa13 | ANXA13 | 4.35E-02 | 8.48E-02 |
| 9923 | Nutm1 | NUTM1 | -1.20E-02 | -7.74E-03 |
| 9924 | Spry4 | SPRY4 | 1.98E-02 | 5.31E-03 |
| 9925 | Pmf1 | PMF1 | 2.10E-02 | 2.69E-02 |
| 9926 | Pmf1 | PMF1-BGLAP | 2.10E-02 | 2.69E-02 |
| 9927 | Tmem26 | TMEM26 | 4.71E-02 | -6.04E-02 |
| 9928 | Rnf222 | RNF222 | 1.32E-02 | -3.09E-02 |
| 9929 | Rps4x | RPS4Y1 | -8.26E-03 | 1.87E-02 |
| 9930 | Obscn | OBSCN | 1.00E-01 | -5.48E-02 |
| 9931 | Gprc5d | GPRC5D | 1.34E-02 | -5.83E-03 |
| 9932 | Hspa4 | HSPA4 | 8.05E-03 | -7.41E-03 |
| 9933 | Rnasek | RNASEK | 2.46E-02 | -1.21E-02 |
| 9934 | Sox4 | SOX4 | 1.19E-02 | 1.05E-01 |
| 9935 | Nphp1 | NPHP1 | 8.30E-03 | -1.78E-02 |
| 9936 | 9530077C05Rik | KIAA0895 | 2.51E-02 | 3.42E-03 |
| 9937 | Fdps | FDPS | 4.26E-02 | 7.18E-02 |
| 9938 | Colec12 | COLEC12 | -3.05E-02 | 1.70E-02 |
| 9939 | Adar | ADAR | -8.44E-03 | -2.72E-03 |
| 9940 | Kctd4 | KCTD4 | -1.44E-02 | -5.99E-03 |
| 9941 | Prpsap1 | PRPSAP1 | 6.22E-03 | -5.78E-03 |
| 9942 | Sap130 | SAP130 | -6.41E-03 | -1.16E-02 |
| 9943 | Zfp207 | ZNF207 | -6.68E-03 | 1.36E-02 |
| 9944 | Rbm8a | RBM8A | -8.27E-03 | -1.88E-03 |
| 9945 | Car3 | CA3 | -8.46E-02 | 5.83E-02 |
| 9946 | Syde2 | SYDE2 | -1.11E-02 | -3.36E-02 |
| 9947 | Sp6 | SP6 | -3.10E-02 | -1.69E-02 |
| 9948 | Fam171a2 | FAM171A2 | 5.11E-02 | 5.20E-02 |
| 9949 | Msr1 | MSR1 | 4.54E-02 | 2.19E-01 |
| 9950 | Med22 | MED22 | 7.03E-03 | -6.24E-03 |
| 9951 | Dnmt1 | DNMT1 | 1.46E-02 | 2.08E-02 |
| 9952 | Knop1 | KNOP1 | 7.56E-03 | -1.04E-02 |
| 9953 | Sf3b4 | SF3B4 | -7.72E-03 | 1.87E-02 |
| 9954 | Med6 | MED6 | -9.93E-03 | 2.98E-03 |
| 9955 | Bmf | BMF | -2.39E-02 | -1.90E-02 |
| 9956 | Fam131b | FAM131B | -8.78E-03 | 1.25E-02 |
| 9957 | Alkbh1 | ALKBH1 | 1.48E-02 | 4.60E-03 |
| 9958 | Klhl28 | KLHL28 | -1.96E-02 | -5.02E-03 |
| 9959 | Plxnb2 | PLXNB2 | 5.55E-03 | -1.12E-02 |
| 9960 | Abcg1 | ABCG1 | 5.85E-02 | 6.98E-02 |
| 9961 | Sft2d2 | SFT2D2 | -6.94E-03 | -2.60E-02 |

| | | | | |
|---|---|---|---|---|
| 9962 | Mrgpre | MRGPRE | 3.63E-02 | -3.59E-02 |
| 9963 | Mrpl36 | MRPL36 | -1.07E-02 | -7.72E-04 |
| 9964 | Klk15 | KLK15 | -1.41E-02 | 2.62E-02 |
| 9965 | Pex2 | PEX2 | 7.28E-03 | -8.66E-05 |
| 9966 | Sema6d | SEMA6D | -2.24E-02 | -9.51E-03 |
| 9967 | Nfx1 | NFX1 | 6.46E-03 | -1.13E-02 |
| 9968 | Eif4ebp2 | EIF4EBP2 | -9.79E-03 | -1.84E-02 |
| 9969 | Paics | PAICS | 7.25E-03 | -1.60E-02 |
| 9970 | Snx29 | SNX29 | -2.73E-02 | -5.46E-02 |
| 9971 | Gm4724 | ZNF442 | -2.01E-02 | -4.99E-02 |
| 9972 | Aplp1 | APLP1 | 2.28E-02 | 6.20E-03 |
| 9973 | Mysm1 | MYSM1 | 9.70E-03 | -7.69E-03 |
| 9974 | Mboat2 | MBOAT2 | 3.76E-02 | -5.84E-02 |
| 9975 | Ralgapa1 | RALGAPA1 | -8.44E-03 | -2.34E-02 |
| 9976 | Lrrtm1 | LRRTM1 | -3.18E-02 | -2.69E-03 |
| 9977 | Slc26a6 | SLC26A6 | 2.76E-02 | -3.45E-02 |
| 9978 | Cpne2 | CPNE2 | -4.38E-02 | 5.27E-02 |
| 9979 | Vstm4 | VSTM4 | -4.58E-02 | 2.65E-02 |
| 9980 | Rel | REL | -3.44E-02 | 7.60E-02 |
| 9981 | Il1r2 | IL1R2 | 5.99E-02 | 6.68E-02 |
| 9982 | Pigyl | PIGY | 1.05E-02 | -4.23E-03 |
| 9983 | Wdfy3 | WDFY3 | -1.36E-02 | -1.55E-02 |
| 9984 | Colgalt2 | COLGALT2 | -1.34E-02 | -2.82E-02 |
| 9985 | Cdh23 | CDH23 | -1.92E-02 | -2.00E-02 |
| 9986 | Agbl2 | AGBL2 | -2.91E-02 | 5.37E-02 |
| 9987 | Trak2 | TRAK2 | 8.27E-03 | -2.22E-02 |
| 9988 | Kazn | KAZN | 1.87E-02 | -1.30E-02 |
| 9989 | Spata2 | SPATA2 | 8.15E-03 | 9.56E-03 |
| 9990 | Ythdf1 | YTHDF1 | -7.88E-03 | 7.90E-03 |
| 9991 | Dab2 | DAB2 | -1.70E-02 | -1.91E-02 |
| 9992 | Elp6 | ELP6 | 1.68E-02 | 1.50E-03 |
| 9993 | Dhtkd1 | DHTKD1 | -2.82E-02 | -3.24E-02 |
| 9994 | Rasef | RASEF | -5.82E-02 | 5.18E-02 |
| 9995 | Syn1 | SYN1 | -1.94E-02 | 4.86E-02 |
| 9996 | Tbl2 | TBL2 | 1.04E-02 | 1.61E-02 |
| 9997 | Shank1 | SHANK1 | -2.34E-02 | -3.80E-02 |
| 9998 | Dmc1 | DMC1 | -1.74E-02 | 1.35E-02 |
| 9999 | Mzf1 | MZF1 | 2.42E-02 | 2.59E-02 |
| 10000 | Tpcn1 | TPCN1 | -1.26E-02 | -2.66E-02 |
| 10001 | Serping1 | SERPING1 | 2.33E-02 | 4.89E-02 |
| 10002 | Cdkn1b | CDKN1B | -7.51E-03 | -4.36E-03 |
| 10003 | B3galt6 | B3GALT6 | -1.79E-02 | -8.13E-03 |
| 10004 | Rbm24 | RBM24 | 3.54E-02 | 5.71E-02 |
| 10005 | Ppp3cc | PPP3CC | 1.11E-02 | -4.61E-03 |
| 10006 | Stat5b | STAT5B | 8.77E-03 | -1.57E-02 |
| 10007 | Vegfa | VEGFA | -9.43E-03 | -8.04E-04 |
| 10008 | Traf3 | TRAF3 | 9.21E-03 | 2.64E-04 |
| 10009 | Psmd3 | PSMD3 | -7.91E-03 | -2.04E-02 |
| 10010 | Gng4 | GNG4 | -2.73E-02 | 8.65E-02 |
| 10011 | Cdyl | CDYL | 1.48E-02 | 2.32E-03 |
| 10012 | Sall2 | SALL2 | -2.34E-02 | 2.60E-03 |
| 10013 | Stx17 | STX17 | -1.02E-02 | -1.46E-02 |
| 10014 | Zfp637 | ZNF32 | -9.75E-03 | 3.69E-03 |

| | | | | |
|---|---|---|---|---|
| 10015 | Junb | JUNB | 5.89E-02 | -2.12E-03 |
| 10016 | Acot4 | ACOT4 | -1.84E-02 | 1.18E-02 |
| 10017 | Stx11 | STX11 | -3.80E-02 | 1.59E-01 |
| 10018 | Sh2d4a | SH2D4A | -3.10E-02 | -6.89E-03 |
| 10019 | Igf2bp3 | IGF2BP3 | -3.00E-02 | 1.42E-02 |
| 10020 | Kif12 | KIF12 | -1.15E-02 | -2.39E-02 |
| 10021 | Nup155 | NUP155 | 9.77E-03 | 7.73E-03 |
| 10022 | Kcne1l | KCNE1L | 3.29E-02 | 3.34E-02 |
| 10023 | Gm13889 | C11orf96 | 2.88E-02 | -2.16E-02 |
| 10024 | Scube1 | SCUBE1 | 1.17E-01 | -6.58E-02 |
| 10025 | Podxl2 | PODXL2 | -2.69E-02 | -9.34E-02 |
| 10026 | Ccni | CCNI | -7.35E-03 | -1.04E-02 |
| 10027 | Akip1 | AKIP1 | -1.31E-02 | -2.42E-02 |
| 10028 | Sh3gl3 | SH3GL3 | -2.42E-02 | 1.06E-02 |
| 10029 | Nde1 | NDE1 | 1.09E-02 | 6.15E-03 |
| 10030 | Mapk14 | MAPK14 | -1.05E-02 | -2.96E-03 |
| 10031 | Agk | AGK | 1.17E-02 | -2.48E-02 |
| 10032 | Praf2 | PRAF2 | -2.01E-02 | 3.56E-02 |
| 10033 | Eqtn | EQTN | -2.30E-02 | 3.61E-02 |
| 10034 | Gprc5a | GPRC5A | 9.71E-02 | 6.38E-02 |
| 10035 | Brms1l | BRMS1L | 9.44E-03 | 1.26E-02 |
| 10036 | Abcd4 | ABCD4 | 1.80E-02 | -2.70E-02 |
| 10037 | Lrrc10b | LRRC10B | 2.83E-02 | -6.29E-02 |
| 10038 | Snrnp200 | SNRNP200 | -8.20E-03 | -1.93E-02 |
| 10039 | Rdh13 | RDH13 | -1.13E-02 | -9.28E-03 |
| 10040 | Clgn | CLGN | 1.11E-02 | -1.15E-02 |
| 10041 | Glo1 | GLO1 | 8.02E-03 | -1.35E-02 |
| 10042 | Baz1a | BAZ1A | -1.68E-02 | 5.01E-02 |
| 10043 | Slc25a47 | SLC25A47 | -3.05E-02 | -3.60E-03 |
| 10044 | Zfp326 | ZNF326 | 7.73E-03 | -7.48E-03 |
| 10045 | R3hdm1 | R3HDM1 | -7.10E-03 | 2.03E-03 |
| 10046 | Ankrd10 | ANKRD10 | -9.07E-03 | 4.87E-03 |
| 10047 | Ubl5 | UBL5 | 7.02E-03 | -1.02E-02 |
| 10048 | Aass | AASS | 1.81E-02 | 5.05E-03 |
| 10049 | Slc25a37 | SLC25A37 | 1.06E-02 | 1.44E-02 |
| 10050 | Polr2f | POLR2F | 8.42E-03 | 1.35E-03 |
| 10051 | Ccna1 | CCNA1 | 1.07E-02 | 7.25E-03 |
| 10052 | Fem1a | FEM1A | 6.03E-03 | -6.31E-03 |
| 10053 | Pla2g4a | PLA2G4A | 2.20E-02 | 2.29E-03 |
| 10054 | Rhbdd2 | RHBDD2 | 8.31E-03 | -6.14E-03 |
| 10055 | Twsg1 | TWSG1 | 1.89E-02 | -3.78E-02 |
| 10056 | Vnn1 | VNN1 | 2.83E-02 | -7.52E-03 |
| 10057 | Ptk6 | PTK6 | -1.62E-02 | 2.34E-02 |
| 10058 | Kcnt2 | KCNT2 | -4.18E-02 | 1.11E-02 |
| 10059 | Cks2 | CKS2 | 1.66E-02 | 1.45E-02 |
| 10060 | Dsn1 | DSN1 | -2.46E-02 | 5.95E-02 |
| 10061 | Gadd45b | GADD45B | 6.95E-02 | -7.87E-03 |
| 10062 | Faim | FAIM | 1.33E-02 | -5.83E-03 |
| 10063 | Cap2 | CAP2 | -3.22E-02 | 1.03E-01 |
| 10064 | Map1a | MAP1A | -4.10E-02 | 6.47E-02 |
| 10065 | Brf2 | BRF2 | 1.43E-02 | -1.55E-02 |
| 10066 | Tor1aip1 | TOR1AIP1 | -7.24E-03 | -2.21E-03 |
| 10067 | Duox1 | DUOX1 | -3.12E-02 | -1.50E-03 |

| | | | | |
|---|---|---|---|---|
| 10068 | Sf3b2 | SF3B2 | -6.22E-03 | -9.55E-03 |
| 10069 | Relb | RELB | 3.16E-02 | 9.93E-02 |
| 10070 | Hnrnpul1 | HNRNPUL1 | 7.98E-03 | -1.52E-02 |
| 10071 | Greb1 | GREB1 | 1.93E-02 | 1.88E-02 |
| 10072 | Wdr72 | WDR72 | 1.30E-02 | 8.06E-03 |
| 10073 | Guf1 | GUF1 | -9.12E-03 | -5.68E-03 |
| 10074 | Gpr63 | GPR63 | 3.28E-02 | -1.40E-02 |
| 10075 | Orc5 | ORC5 | 1.15E-02 | 2.34E-03 |
| 10076 | Nadk2 | NADK2 | 1.72E-02 | -3.73E-02 |
| 10077 | Spdya | SPDYA | 2.12E-02 | 2.46E-02 |
| 10078 | Nat10 | NAT10 | 7.56E-03 | -4.17E-03 |
| 10079 | Gatc | GATC | 8.27E-03 | -1.81E-02 |
| 10080 | Hs3st4 | HS3ST4 | -1.56E-02 | 4.58E-02 |
| 10081 | Ccbe1 | CCBE1 | 2.22E-02 | -1.46E-02 |
| 10082 | Lrch4 | LRCH4 | -2.22E-02 | 4.66E-03 |
| 10083 | Gripap1 | GRIPAP1 | 6.36E-03 | -7.48E-03 |
| 10084 | Tpp1 | TPP1 | 9.68E-03 | -1.05E-02 |
| 10085 | Abca1 | ABCA1 | 2.65E-02 | 4.35E-02 |
| 10086 | Sh3pxd2b | SH3PXD2B | -1.49E-02 | 6.38E-02 |
| 10087 | Rybp | RYBP | -1.36E-02 | -6.13E-03 |
| 10088 | Gpr82 | GPR82 | -8.31E-03 | 3.51E-03 |
| 10089 | 8030462N17Rik | C18orf25 | -1.27E-02 | -1.79E-02 |
| 10090 | Ces3a | CES3 | 2.89E-02 | 1.57E-02 |
| 10091 | Atf4 | ATF4 | 1.19E-02 | 2.10E-02 |
| 10092 | Cd81 | CD81 | -5.43E-03 | -7.00E-03 |
| 10093 | Dedd2 | DEDD2 | -1.27E-02 | -3.79E-02 |
| 10094 | Usp24 | USP24 | 8.61E-03 | -1.38E-02 |
| 10095 | Ppif | PPIF | -8.67E-03 | -1.47E-02 |
| 10096 | Hmgn1 | HMGN1 | -7.16E-03 | 2.50E-02 |
| 10097 | Arid4b | ARID4B | -9.06E-03 | -4.04E-03 |
| 10098 | Htr2a | HTR2A | -2.25E-02 | 1.95E-02 |
| 10099 | Caap1 | CAAP1 | -1.86E-02 | 2.19E-02 |
| 10100 | Adam22 | ADAM22 | 1.80E-02 | -2.17E-02 |
| 10101 | Pmvk | PMVK | 1.48E-02 | 1.95E-02 |
| 10102 | Zhx2 | ZHX2 | -1.23E-02 | 2.58E-02 |
| 10103 | Katnbl1 | KATNBL1 | 1.55E-02 | 1.96E-02 |
| 10104 | Cd300a | CD300A | -3.32E-02 | 1.08E-01 |
| 10105 | Ticam1 | TICAM1 | -1.02E-02 | 1.38E-02 |
| 10106 | Aldh9a1 | ALDH9A1 | 1.57E-02 | -1.36E-02 |
| 10107 | Commd4 | COMMD4 | 8.10E-03 | -1.88E-03 |
| 10108 | Chsy3 | CHSY3 | 5.19E-02 | -3.72E-02 |
| 10109 | Steap2 | STEAP2 | -1.54E-02 | 2.09E-02 |
| 10110 | Zfp654 | ZNF654 | 1.16E-02 | 4.93E-04 |
| 10111 | Zbtb32 | ZBTB32 | -3.10E-02 | 6.30E-02 |
| 10112 | Capza1 | CAPZA1 | 9.17E-03 | 1.38E-02 |
| 10113 | Prkaa1 | PRKAA1 | 9.33E-03 | 1.62E-03 |
| 10114 | Grik3 | GRIK3 | -4.30E-02 | 1.56E-02 |
| 10115 | Pole | POLE | 5.09E-02 | 1.48E-01 |
| 10116 | Rab3c | RAB3C | 1.72E-02 | -4.32E-02 |
| 10117 | Aaas | AAAS | 1.15E-02 | 9.84E-03 |
| 10118 | Ube2b | UBE2B | -7.12E-03 | -4.75E-03 |
| 10119 | Tmprss7 | TMPRSS7 | 2.94E-02 | -3.44E-02 |
| 10120 | Senp5 | SENP5 | -1.10E-02 | 1.55E-03 |

| | | | | |
|---|---|---|---|---|
| 10121 | Hpx | HPX | -6.44E-02 | 1.77E-01 |
| 10122 | Nsd1 | NSD1 | 5.73E-03 | -3.63E-03 |
| 10123 | Gnl3 | GNL3 | 1.07E-02 | 3.15E-02 |
| 10124 | Exosc1 | EXOSC1 | 1.14E-02 | 5.98E-04 |
| 10125 | Zfp316 | ZNF316 | -1.41E-02 | -3.04E-02 |
| 10126 | Srsf6 | SRSF6 | 6.35E-03 | 1.37E-02 |
| 10127 | Rbm38 | RBM38 | 1.93E-02 | 3.87E-02 |
| 10128 | Sugt1 | SUGT1 | -5.85E-03 | 5.44E-03 |
| 10129 | Gm14391 | ZNF442 | -9.38E-03 | 8.77E-03 |
| 10130 | Dusp10 | DUSP10 | -2.54E-02 | 7.03E-02 |
| 10131 | Mllt10 | MLLT10 | 8.94E-03 | 3.19E-03 |
| 10132 | Pbx1 | PBX1 | -9.98E-03 | -1.50E-02 |
| 10133 | Atp11b | ATP11B | 8.38E-03 | -1.38E-04 |
| 10134 | Lrp5 | LRP5 | 1.21E-02 | -4.56E-03 |
| 10135 | Snap23 | SNAP23 | 1.03E-02 | -2.32E-02 |
| 10136 | Fggy | FGGY | -1.66E-02 | -2.58E-02 |
| 10137 | Gng11 | GNG11 | -1.52E-02 | 1.56E-02 |
| 10138 | Taf5l | TAF5L | 1.08E-02 | 5.95E-03 |
| 10139 | Klhl42 | KLHL42 | 1.43E-02 | -5.92E-03 |
| 10140 | Pik3c3 | PIK3C3 | 9.41E-03 | -1.01E-02 |
| 10141 | Haus5 | HAUS5 | 2.11E-02 | 1.46E-02 |
| 10142 | Rpl21 | RPL21 | -7.30E-03 | 2.26E-03 |
| 10143 | Lrrn3 | LRRN3 | -2.30E-02 | -1.13E-02 |
| 10144 | Lpp | LPP | -1.02E-02 | -7.66E-03 |
| 10145 | Vwa8 | VWA8 | 1.20E-02 | -2.56E-02 |
| 10146 | Ceacam16 | CEACAM16 | -7.89E-03 | 1.16E-02 |
| 10147 | Ahi1 | AHI1 | 1.09E-02 | -1.62E-02 |
| 10148 | Mcee | MCEE | -7.85E-03 | -1.74E-02 |
| 10149 | Csf2rb2 | CSF2RB | -6.52E-02 | 3.05E-02 |
| 10150 | Lacc1 | LACC1 | -2.87E-02 | 5.53E-02 |
| 10151 | Ten1 | TEN1 | -1.74E-02 | 3.57E-02 |
| 10152 | Rasgef1c | RASGEF1C | 1.35E-02 | -5.75E-03 |
| 10153 | Gpr146 | GPR146 | 1.10E-02 | -1.12E-02 |
| 10154 | Casp8 | CASP8 | 1.14E-02 | 3.07E-02 |
| 10155 | Kcnk13 | KCNK13 | 1.92E-02 | 2.55E-02 |
| 10156 | Glis1 | GLIS1 | 2.77E-02 | -4.99E-02 |
| 10157 | Apcs | APCS | 1.54E-02 | 4.50E-02 |
| 10158 | Cad | CAD | -9.99E-03 | 3.34E-04 |
| 10159 | Enho | ENHO | 2.83E-02 | -1.93E-02 |
| 10160 | Slc16a11 | SLC16A11 | 1.37E-02 | -1.51E-02 |
| 10161 | Ppp3cb | PPP3CB | -6.75E-03 | -2.87E-03 |
| 10162 | Zfp428 | ZNF428 | 2.32E-02 | 3.00E-02 |
| 10163 | Gtf2a2 | GTF2A2 | 8.01E-03 | 6.16E-03 |
| 10164 | Ubap2 | UBAP2 | -8.77E-03 | 5.92E-03 |
| 10165 | Serpinb9 | SERPINB9 | 2.51E-02 | 1.32E-02 |
| 10166 | Ksr2 | KSR2 | -3.48E-02 | -4.06E-02 |
| 10167 | Myo15 | MYO15A | -2.13E-02 | 3.20E-02 |
| 10168 | Tmem37 | TMEM37 | 1.86E-02 | 2.89E-02 |
| 10169 | Anapc11 | ANAPC11 | 7.25E-03 | -3.45E-03 |
| 10170 | Shb | SHB | -1.11E-02 | 2.68E-02 |
| 10171 | Ndufs1 | NDUFS1 | 1.03E-02 | -2.24E-02 |
| 10172 | Dpf3 | DPF3 | -2.67E-02 | -4.30E-03 |
| 10173 | Cnga4 | CNGA4 | -1.87E-02 | -1.67E-02 |

| | | | | |
|---|---|---|---|---|
| 10174 | Rarb | RARB | -1.71E-02 | -4.99E-02 |
| 10175 | Tmem59l | TMEM59L | -9.85E-03 | 2.13E-02 |
| 10176 | Rab20 | RAB20 | -1.26E-02 | -5.21E-03 |
| 10177 | Dhx35 | DHX35 | 9.04E-03 | -2.45E-02 |
| 10178 | Trpt1 | TRPT1 | -1.16E-02 | -5.23E-02 |
| 10179 | Ranbp10 | RANBP10 | -6.07E-03 | -5.04E-03 |
| 10180 | Lrig3 | LRIG3 | 1.94E-02 | -3.95E-02 |
| 10181 | Larp6 | LARP6 | 3.28E-02 | -5.44E-02 |
| 10182 | Alg6 | ALG6 | -1.48E-02 | -1.46E-02 |
| 10183 | Zcchc3 | ZCCHC3 | -1.96E-02 | -4.30E-02 |
| 10184 | Arhgap32 | ARHGAP32 | -8.40E-03 | -4.37E-02 |
| 10185 | Slx4 | SLX4 | -1.14E-02 | 2.68E-02 |
| 10186 | Lix1l | LIX1L | -2.49E-02 | 1.38E-02 |
| 10187 | Lamc1 | LAMC1 | 9.89E-03 | 7.28E-04 |
| 10188 | Tdrd1 | TDRD1 | 1.21E-02 | -5.68E-03 |
| 10189 | Tcp1 | TCP1 | -7.60E-03 | 4.32E-03 |
| 10190 | Npm2 | NPM2 | 2.82E-02 | -4.41E-02 |
| 10191 | Pde6h | PDE6H | 1.17E-02 | -1.62E-02 |
| 10192 | Sesn1 | SESN1 | 1.00E-02 | -2.94E-02 |
| 10193 | Lyz2 | LYZ | 4.32E-02 | 1.28E-01 |
| 10194 | Ccdc152 | CCDC152 | -2.29E-02 | -2.66E-02 |
| 10195 | Atp13a3 | ATP13A3 | 9.93E-03 | -5.57E-04 |
| 10196 | Kntc1 | KNTC1 | 6.17E-02 | 1.89E-01 |
| 10197 | Eif2s1 | EIF2S1 | 6.86E-03 | 2.87E-03 |
| 10198 | Ccdc60 | CCDC60 | 1.21E-02 | -9.17E-03 |
| 10199 | Sftpd | SFTPD | 1.97E-02 | 6.01E-02 |
| 10200 | Thap3 | THAP3 | -1.10E-02 | -1.17E-02 |
| 10201 | Lactb2 | LACTB2 | 1.19E-02 | -2.28E-02 |
| 10202 | Pkdcc | PKDCC | 1.84E-02 | 8.61E-03 |
| 10203 | Tmem135 | TMEM135 | 7.61E-03 | -1.10E-02 |
| 10204 | Mdfi | MDFI | -2.92E-02 | 7.49E-02 |
| 10205 | Agl | AGL | 9.65E-03 | -1.55E-02 |
| 10206 | Yipf3 | YIPF3 | -6.06E-03 | -2.65E-03 |
| 10207 | P2rx5 | P2RX5 | -3.43E-02 | -4.92E-02 |
| 10208 | Gm136 | C6orf163 | -1.17E-02 | 1.39E-02 |
| 10209 | Yipf5 | YIPF5 | 6.21E-03 | -3.54E-03 |
| 10210 | Creld1 | CRELD1 | -1.31E-02 | 1.09E-02 |
| 10211 | Kcnq5 | KCNQ5 | 2.02E-02 | -1.56E-02 |
| 10212 | Ric3 | RIC3 | -3.85E-02 | -9.69E-02 |
| 10213 | Kptn | KPTN | 1.06E-02 | -2.20E-02 |
| 10214 | Ets2 | ETS2 | 1.13E-02 | 2.85E-02 |
| 10215 | Pald1 | PALD1 | 3.47E-02 | 8.30E-03 |
| 10216 | Ncstn | NCSTN | -6.21E-03 | 3.62E-03 |
| 10217 | Sod2 | SOD2 | 9.92E-03 | -2.30E-02 |
| 10218 | Cipc | CIPC | -8.67E-03 | -9.48E-04 |
| 10219 | Slc38a5 | SLC38A5 | -8.36E-03 | 2.43E-02 |
| 10220 | Cttnbp2nl | CTTNBP2NL | -8.92E-03 | 3.71E-02 |
| 10221 | Hkdc1 | HKDC1 | 2.83E-02 | 1.41E-01 |
| 10222 | Tram1l1 | TRAM1L1 | 1.82E-02 | -1.67E-02 |
| 10223 | Cst6 | CST6 | -1.78E-02 | -1.35E-03 |
| 10224 | Dnal1 | DNAL1 | -1.32E-02 | 1.04E-02 |
| 10225 | Glyat | GLYAT | 2.18E-02 | -2.17E-03 |
| 10226 | Trip6 | TRIP6 | 6.30E-03 | -1.89E-02 |

| | | | | |
|---|---|---|---|---|
| 10227 | Lztfl1 | LZTFL1 | -8.26E-03 | 1.57E-04 |
| 10228 | Cant1 | CANT1 | -6.65E-03 | -9.35E-03 |
| 10229 | Fam136a | FAM136A | -9.97E-03 | 3.24E-02 |
| 10230 | Wfikkn2 | WFIKKN2 | -2.46E-02 | -3.63E-02 |
| 10231 | Cryab | CRYAB | -4.12E-02 | -5.93E-02 |
| 10232 | Ccdc62 | CCDC62 | 1.59E-02 | -8.71E-03 |
| 10233 | Dcn | DCN | -2.86E-02 | 8.21E-04 |
| 10234 | Abt1 | ABT1 | -1.11E-02 | 2.80E-03 |
| 10235 | Acsl5 | ACSL5 | -9.51E-03 | 1.40E-03 |
| 10236 | Exoc7 | EXOC7 | -7.65E-03 | -2.11E-02 |
| 10237 | Serf2 | SERF2 | 6.62E-03 | -9.99E-03 |
| 10238 | Bach2 | BACH2 | -2.54E-02 | -4.14E-02 |
| 10239 | Vps25 | VPS25 | 2.26E-02 | -1.30E-02 |
| 10240 | Pafah1b3 | PAFAH1B3 | 1.19E-02 | 2.32E-02 |
| 10241 | Kng2 | KNG1 | -1.20E-02 | -2.95E-02 |
| 10242 | Zfp641 | ZNF641 | 1.60E-02 | 8.39E-03 |
| 10243 | Tyro3 | TYRO3 | -2.98E-02 | 8.15E-02 |
| 10244 | Chpf | CHPF | 1.31E-02 | 1.84E-02 |
| 10245 | Dok2 | DOK2 | -5.59E-02 | -9.66E-02 |
| 10246 | Nbr1 | NBR1 | -7.56E-03 | -1.36E-02 |
| 10247 | Polg | POLG | -7.10E-03 | -6.70E-04 |
| 10248 | Arfgap1 | ARFGAP1 | 5.16E-03 | -7.63E-03 |
| 10249 | Mkx | MKX | -4.91E-02 | -6.40E-02 |
| 10250 | Riiad1 | RIIAD1 | 2.99E-02 | 1.54E-02 |
| 10251 | Cgn | CGN | -2.74E-02 | -3.59E-03 |
| 10252 | Npnt | NPNT | 1.12E-02 | -1.77E-02 |
| 10253 | Cdc40 | CDC40 | -8.13E-03 | -5.23E-03 |
| 10254 | Ets1 | ETS1 | -1.92E-02 | 6.39E-04 |
| 10255 | Ulbp1 | ULBP1 | -3.80E-02 | 1.78E-01 |
| 10256 | Zar1 | ZAR1 | 1.42E-02 | -1.73E-02 |
| 10257 | Ntng1 | NTNG1 | 4.18E-02 | -4.87E-02 |
| 10258 | Tle1 | TLE1 | -1.08E-02 | 8.43E-03 |
| 10259 | Uckl1 | UCKL1 | 8.18E-03 | -1.48E-03 |
| 10260 | Pex19 | PEX19 | 7.68E-03 | -1.06E-02 |
| 10261 | Zfp433 | ZNF433 | -2.18E-02 | -2.75E-02 |
| 10262 | Hes2 | HES2 | -4.32E-02 | 3.96E-02 |
| 10263 | Dnajc7 | DNAJC7 | 5.10E-03 | -1.10E-02 |
| 10264 | Zfp711 | ZNF711 | -3.07E-02 | -4.43E-02 |
| 10265 | Fanci | FANCI | 6.01E-02 | 1.77E-01 |
| 10266 | Klhl22 | KLHL22 | -1.22E-02 | -1.52E-02 |
| 10267 | Ephx3 | EPHX3 | 2.77E-02 | 1.90E-02 |
| 10268 | Fgf21 | FGF21 | 8.08E-03 | 1.72E-02 |
| 10269 | Mnat1 | MNAT1 | -1.14E-02 | 5.64E-03 |
| 10270 | Efcab1 | EFCAB1 | 1.93E-02 | 1.23E-02 |
| 10271 | Cep135 | CEP135 | 1.41E-02 | -3.28E-02 |
| 10272 | Rps3 | RPS3 | -7.11E-03 | 6.40E-03 |
| 10273 | B3galt4 | B3GALT4 | -2.26E-02 | 7.55E-03 |
| 10274 | Ddx4 | DDX4 | -1.27E-02 | 1.78E-02 |
| 10275 | Gxylt1 | GXYLT1 | -9.74E-03 | -7.35E-03 |
| 10276 | Lbx2 | LBX2 | -2.57E-02 | -4.48E-03 |
| 10277 | Tbc1d13 | TBC1D13 | -1.06E-02 | -2.62E-02 |
| 10278 | Msrb3 | MSRB3 | 1.62E-02 | -9.63E-03 |
| 10279 | Ints5 | INTS5 | -9.78E-03 | -1.24E-03 |

| | | | | |
|---|---|---|---|---|
| 10280 | Dscam | DSCAM | 1.18E-02 | -1.47E-03 |
| 10281 | Slc51b | SLC51B | 1.16E-02 | 1.40E-03 |
| 10282 | Acot11 | ACOT11 | 1.16E-02 | -1.02E-02 |
| 10283 | Gins4 | GINS4 | 8.24E-03 | 2.61E-02 |
| 10284 | Lect2 | LECT2 | -1.74E-02 | 3.70E-02 |
| 10285 | Luc7l3 | LUC7L3 | -6.87E-03 | 1.52E-02 |
| 10286 | Gpank1 | GPANK1 | -1.08E-02 | -1.13E-02 |
| 10287 | Grin3b | GRIN3B | -3.10E-02 | -9.98E-02 |
| 10288 | Dlx3 | DLX3 | -1.92E-02 | 2.09E-02 |
| 10289 | Aqp5 | AQP5 | 7.00E-02 | -5.86E-03 |
| 10290 | Akr1c21 | AKR1C1 | 1.49E-02 | 5.69E-03 |
| 10291 | Akr1c21 | AKR1C2 | 1.49E-02 | 5.69E-03 |
| 10292 | Cep70 | CEP70 | -1.28E-02 | -2.58E-02 |
| 10293 | Sys1 | SYS1 | -8.15E-03 | 2.93E-03 |
| 10294 | Npw | NPW | -1.27E-02 | 1.34E-02 |
| 10295 | Sfmbt2 | SFMBT2 | -2.55E-02 | 8.08E-03 |
| 10296 | Sapcd1 | SAPCD1 | -2.80E-02 | 3.61E-02 |
| 10297 | Pdia3 | PDIA3 | 1.00E-02 | 5.27E-04 |
| 10298 | Shq1 | SHQ1 | -1.80E-02 | 1.69E-02 |
| 10299 | Dcdc5 | DCDC5 | -1.41E-02 | -2.54E-02 |
| 10300 | Dcxr | DCXR | 1.36E-02 | -3.29E-02 |
| 10301 | Gpc6 | GPC6 | -3.03E-02 | -1.74E-03 |
| 10302 | Minpp1 | MINPP1 | 8.08E-03 | 4.58E-04 |
| 10303 | Fam219a | FAM219A | -7.07E-03 | -1.01E-02 |
| 10304 | Rpl10 | RPL10 | -8.08E-03 | 1.05E-02 |
| 10305 | Inppl1 | INPPL1 | -6.34E-03 | -3.73E-03 |
| 10306 | As3mt | AS3MT | -1.09E-02 | -6.99E-03 |
| 10307 | Ddx24 | DDX24 | -5.81E-03 | -5.75E-03 |
| 10308 | Msantd3 | MSANTD3 | 3.50E-02 | -4.45E-02 |
| 10309 | 9930012K11Rik | C8orf58 | -2.90E-02 | -1.67E-02 |
| 10310 | Gfod2 | GFOD2 | -2.00E-02 | 2.71E-03 |
| 10311 | Esyt2 | ESYT2 | -7.45E-03 | 9.85E-03 |
| 10312 | Sap30l | SAP30L | -8.41E-03 | -1.01E-02 |
| 10313 | Padi1 | PADI1 | -5.85E-02 | 9.71E-02 |
| 10314 | Mak16 | MAK16 | -7.70E-03 | 3.67E-02 |
| 10315 | Pycr1 | PYCR1 | -2.36E-02 | 3.25E-02 |
| 10316 | S100z | S100Z | 1.46E-02 | 5.31E-03 |
| 10317 | Gtf2i | GTF2I | 7.94E-03 | -1.72E-02 |
| 10318 | Tmem74 | TMEM74 | -1.16E-02 | 3.01E-02 |
| 10319 | Cdca4 | CDCA4 | -2.36E-02 | 2.67E-02 |
| 10320 | Morn3 | MORN3 | -9.17E-03 | 2.99E-03 |
| 10321 | Phax | PHAX | 6.56E-03 | -2.63E-03 |
| 10322 | Ccdc13 | CCDC13 | -2.04E-02 | 2.86E-02 |
| 10323 | Rapgef3 | RAPGEF3 | 9.87E-03 | -2.93E-03 |
| 10324 | Hsf4 | HSF4 | -2.45E-02 | -2.72E-02 |
| 10325 | Mybbp1a | MYBBP1A | 7.12E-03 | 3.60E-03 |
| 10326 | Gp1bb | GP1BB | 1.21E-02 | 5.98E-02 |
| 10327 | Psma4 | PSMA4 | 8.99E-03 | -1.02E-02 |
| 10328 | Mylk | MYLK | 1.11E-02 | -1.55E-02 |
| 10329 | Msln | MSLN | 1.19E-01 | 1.64E-01 |
| 10330 | Osbpl8 | OSBPL8 | -1.23E-02 | -2.92E-02 |
| 10331 | Ncaph2 | NCAPH2 | 6.72E-03 | -2.95E-03 |
| 10332 | Dcun1d5 | DCUN1D5 | 6.58E-03 | 1.88E-02 |

| | | | | |
|---|---|---|---|---|
| 10333 | 3110082I17Rik | C7orf50 | 2.73E-02 | 5.14E-02 |
| 10334 | Dgat2 | DGAT2 | 1.57E-02 | -9.26E-03 |
| 10335 | Lct | LCT | -1.39E-02 | 2.47E-02 |
| 10336 | Asxl2 | ASXL2 | -6.92E-03 | 2.75E-03 |
| 10337 | Igll1 | IGLL5 | -2.61E-02 | 4.31E-02 |
| 10338 | Igll1 | IGLL1 | -2.61E-02 | 4.31E-02 |
| 10339 | Nhej1 | NHEJ1 | -1.80E-02 | -1.64E-02 |
| 10340 | Mboat4 | MBOAT4 | 1.18E-02 | -3.65E-02 |
| 10341 | Aspm | ASPM | 5.62E-02 | 1.92E-01 |
| 10342 | Me3 | ME3 | 1.19E-02 | -2.76E-02 |
| 10343 | Npsr1 | NPSR1 | 1.53E-02 | -3.08E-02 |
| 10344 | Ankdd1b | ANKDD1B | -1.84E-02 | 1.44E-02 |
| 10345 | Chrm4 | CHRM4 | -1.25E-02 | 1.13E-02 |
| 10346 | Caml | CAMLG | -6.88E-03 | 7.76E-03 |
| 10347 | Abi2 | ABI2 | -1.06E-02 | 1.04E-02 |
| 10348 | Smc5 | SMC5 | 1.00E-02 | -1.80E-03 |
| 10349 | Zfp622 | ZNF622 | -9.61E-03 | 1.56E-02 |
| 10350 | Map2 | MAP2 | -4.34E-02 | -3.12E-02 |
| 10351 | Tmed7 | TMED7 | -6.83E-03 | -1.02E-02 |
| 10352 | Bbs2 | BBS2 | -8.11E-03 | -2.08E-02 |
| 10353 | Preb | PREB | 7.08E-03 | -8.68E-03 |
| 10354 | Spred2 | SPRED2 | -1.73E-02 | 6.02E-03 |
| 10355 | Rad17 | RAD17 | 8.00E-03 | -4.90E-03 |
| 10356 | Skida1 | SKIDA1 | -1.74E-02 | 2.61E-03 |
| 10357 | Pnp | PNP | 1.98E-02 | 3.00E-03 |
| 10358 | Stk32a | STK32A | 2.38E-02 | -3.30E-03 |
| 10359 | Rpl7a | RPL7A | 7.23E-03 | 4.66E-03 |
| 10360 | Fpr2 | FPR2 | -7.65E-02 | 1.09E-01 |
| 10361 | Wdr53 | WDR53 | -1.15E-02 | -8.18E-03 |
| 10362 | Dnph1 | DNPH1 | 1.90E-02 | 2.20E-02 |
| 10363 | Mob3c | MOB3C | 1.29E-02 | -2.06E-02 |
| 10364 | Ptpra | PTPRA | -6.22E-03 | -1.62E-02 |
| 10365 | Mtx3 | MTX3 | -8.81E-03 | -4.37E-02 |
| 10366 | Nupr1l | NUPR1L | -2.71E-02 | -3.50E-02 |
| 10367 | Pan3 | PAN3 | 7.62E-03 | -6.96E-03 |
| 10368 | Tspyl5 | TSPYL5 | 2.18E-02 | -2.28E-03 |
| 10369 | Vps52 | VPS52 | -5.73E-03 | -8.36E-03 |
| 10370 | Zfp454 | ZNF454 | -2.05E-02 | 2.31E-03 |
| 10371 | Eno3 | ENO3 | 1.73E-02 | -2.43E-02 |
| 10372 | Zbtb6 | ZBTB6 | -1.22E-02 | -4.43E-03 |
| 10373 | Ptgr2 | PTGR2 | 8.91E-03 | -1.63E-02 |
| 10374 | Cabp2 | CABP2 | 1.62E-02 | 1.79E-02 |
| 10375 | Lonp1 | LONP1 | 8.11E-03 | -2.14E-02 |
| 10376 | Lrrc27 | LRRC27 | 2.27E-02 | 4.98E-02 |
| 10377 | Il7r | IL7R | -3.60E-02 | 5.46E-02 |
| 10378 | Plcd4 | PLCD4 | -3.01E-02 | 1.46E-02 |
| 10379 | Pate2 | PATE2 | 1.40E-02 | -3.00E-02 |
| 10380 | Fastkd1 | FASTKD1 | -8.25E-03 | 7.72E-04 |
| 10381 | Cd44 | CD44 | 3.35E-02 | 3.47E-01 |
| 10382 | Mst1 | MST1 | -2.19E-02 | 5.30E-03 |
| 10383 | Abhd12 | ABHD12 | -6.66E-03 | 8.86E-03 |
| 10384 | Dhx36 | DHX36 | 6.37E-03 | -1.09E-02 |
| 10385 | 3425401B19Rik | C10orf71 | 5.67E-03 | 5.18E-04 |

| | | | | |
|---|---|---|---|---|
| 10386 | Cd177 | CD177 | -2.37E-02 | 5.67E-02 |
| 10387 | Sox30 | SOX30 | -1.18E-02 | 3.79E-04 |
| 10388 | Tmem183a | TMEM183A | 6.43E-03 | -9.80E-03 |
| 10389 | Agtrap | AGTRAP | 7.74E-03 | -1.50E-02 |
| 10390 | Kcnmb2 | KCNMB2 | 1.18E-02 | -2.42E-02 |
| 10391 | Atad3a | ATAD3A | 7.77E-03 | -1.57E-02 |
| 10392 | Cdhr2 | CDHR2 | 1.57E-02 | -3.51E-02 |
| 10393 | Wdr44 | WDR44 | -1.70E-02 | 8.28E-03 |
| 10394 | Gnaz | GNAZ | -2.74E-02 | -2.05E-02 |
| 10395 | Zc3h18 | ZC3H18 | -6.71E-03 | -1.29E-02 |
| 10396 | Gdpgp1 | GDPGP1 | 1.61E-02 | -2.34E-02 |
| 10397 | Amn1 | AMN1 | 1.48E-02 | 1.85E-02 |
| 10398 | Hdac5 | HDAC5 | -9.00E-03 | -3.10E-02 |
| 10399 | Lrrc7 | LRRC7 | 8.84E-03 | 1.42E-02 |
| 10400 | Cog1 | COG1 | 6.01E-03 | -4.90E-03 |
| 10401 | Maged2 | MAGED2 | -9.35E-03 | 2.13E-02 |
| 10402 | Slc25a11 | SLC25A11 | 8.69E-03 | -8.87E-03 |
| 10403 | Prpf8 | PRPF8 | 8.44E-03 | -2.39E-02 |
| 10404 | Sptbn4 | SPTBN4 | -2.91E-02 | -6.32E-02 |
| 10405 | Rbm5 | RBM5 | -5.63E-03 | -3.63E-03 |
| 10406 | Sult4a1 | SULT4A1 | 3.60E-02 | 2.21E-01 |
| 10407 | Tmem9 | TMEM9 | -6.95E-03 | -1.86E-02 |
| 10408 | Rere | RERE | -9.03E-03 | -9.52E-03 |
| 10409 | Ppt1 | PPT1 | -7.97E-03 | -8.82E-03 |
| 10410 | Tmem208 | TMEM208 | -7.06E-03 | -3.18E-03 |
| 10411 | Lpar4 | LPAR4 | 2.32E-02 | -1.84E-02 |
| 10412 | Scpep1 | SCPEP1 | 1.10E-02 | 8.22E-03 |
| 10413 | Ddx23 | DDX23 | -4.90E-03 | -8.47E-03 |
| 10414 | Fmo2 | FMO2 | -1.72E-02 | -2.50E-04 |
| 10415 | Asic5 | ASIC5 | 9.48E-03 | 5.35E-03 |
| 10416 | Frs2 | FRS2 | -8.49E-03 | -1.07E-02 |
| 10417 | Cog5 | COG5 | -9.10E-03 | -6.91E-03 |
| 10418 | Cnot11 | CNOT11 | 8.03E-03 | -1.63E-02 |
| 10419 | Tnip1 | TNIP1 | -9.60E-03 | 1.97E-02 |
| 10420 | Slc9a6 | SLC9A6 | -8.62E-03 | -3.70E-02 |
| 10421 | Pde8a | PDE8A | 1.01E-02 | -5.93E-03 |
| 10422 | Npas4 | NPAS4 | 1.75E-02 | 6.51E-02 |
| 10423 | Fbxw7 | FBXW7 | -7.91E-03 | 1.15E-02 |
| 10424 | Abhd11 | ABHD11 | 8.71E-03 | -3.90E-03 |
| 10425 | Cetn3 | CETN3 | -6.26E-03 | 5.16E-03 |
| 10426 | Dleu7 | DLEU7 | -2.63E-02 | 2.69E-02 |
| 10427 | Dnmt3l | DNMT3L | 2.70E-02 | 1.95E-02 |
| 10428 | Rpl7l1 | RPL7L1 | 7.31E-03 | -2.19E-03 |
| 10429 | Coq4 | COQ4 | -8.76E-03 | -2.04E-02 |
| 10430 | Il31ra | IL31RA | -1.83E-02 | -3.97E-03 |
| 10431 | Tek | TEK | -1.60E-02 | -2.88E-02 |
| 10432 | Sdr42e1 | SDR42E1 | 1.71E-02 | -6.73E-03 |
| 10433 | Serpinf1 | SERPINF1 | 3.52E-02 | 2.00E-02 |
| 10434 | Gpr22 | GPR22 | 2.50E-02 | 6.52E-02 |
| 10435 | Cd82 | CD82 | 2.38E-02 | -3.57E-02 |
| 10436 | Osgepl1 | OSGEPL1 | -1.10E-02 | -1.72E-02 |
| 10437 | Pemt | PEMT | 2.15E-02 | 3.92E-02 |
| 10438 | Arhgap15 | ARHGAP15 | 3.97E-02 | 2.08E-01 |

| | | | | |
|---|---|---|---|---|
| 10439 | Gda | GDA | -4.76E-02 | 1.74E-01 |
| 10440 | Trim47 | TRIM47 | 1.49E-02 | 8.05E-02 |
| 10441 | Ppig | PPIG | 5.57E-03 | -2.90E-03 |
| 10442 | Ier5l | IER5L | -3.63E-02 | 7.49E-02 |
| 10443 | Glt8d2 | GLT8D2 | -4.02E-02 | 2.29E-02 |
| 10444 | Ceacam20 | CEACAM20 | 7.82E-03 | 6.47E-03 |
| 10445 | Cttn | CTTN | 5.20E-03 | -1.71E-03 |
| 10446 | Urb2 | URB2 | -9.75E-03 | 2.71E-02 |
| 10447 | Hilpda | HILPDA | 2.41E-02 | 2.75E-02 |
| 10448 | Nyap1 | NYAP1 | 5.29E-02 | 5.12E-02 |
| 10449 | Ppp1r8 | PPP1R8 | -6.83E-03 | -2.99E-03 |
| 10450 | Tm4sf4 | TM4SF4 | 1.53E-02 | 5.20E-02 |
| 10451 | 2510039O18Rik | KIAA2013 | 6.52E-03 | 9.42E-03 |
| 10452 | Ftsj3 | FTSJ3 | 5.30E-03 | 1.62E-02 |
| 10453 | Rgs6 | RGS6 | 2.44E-02 | -1.03E-01 |
| 10454 | Rab3b | RAB3B | -1.90E-02 | 5.08E-03 |
| 10455 | Zbtb18 | ZBTB18 | -1.04E-02 | 2.40E-03 |
| 10456 | Ak7 | AK7 | -3.90E-02 | -3.41E-02 |
| 10457 | Ppp1r11 | PPP1R11 | -7.04E-03 | -1.28E-02 |
| 10458 | Brd2 | BRD2 | -5.50E-03 | -3.28E-03 |
| 10459 | Zfp664 | ZNF664 | -5.34E-03 | -7.67E-03 |
| 10460 | Hspa12b | HSPA12B | 1.76E-02 | -4.82E-03 |
| 10461 | Trpc1 | TRPC1 | 1.31E-02 | -2.40E-02 |
| 10462 | Rab24 | RAB24 | 7.49E-03 | -7.50E-03 |
| 10463 | Zfp865 | ZNF865 | -8.15E-03 | 1.08E-02 |
| 10464 | Ist1 | IST1 | 5.45E-03 | -3.28E-03 |
| 10465 | Prkag2 | PRKAG2 | 1.25E-02 | 1.22E-02 |
| 10466 | Cit | CIT | 3.69E-02 | -3.88E-02 |
| 10467 | Arhgap5 | ARHGAP5 | 9.79E-03 | -1.37E-02 |
| 10468 | Mvd | MVD | -1.35E-02 | -1.50E-02 |
| 10469 | Col6a6 | COL6A6 | -1.66E-02 | -1.47E-03 |
| 10470 | Pitpnm1 | PITPNM1 | 1.04E-02 | 4.35E-02 |
| 10471 | Myt1 | MYT1 | 8.46E-03 | -6.18E-03 |
| 10472 | Rab36 | RAB36 | 1.80E-02 | -9.18E-02 |
| 10473 | Slc2a1 | SLC2A1 | 2.05E-02 | -5.97E-03 |
| 10474 | Rnf151 | RNF151 | -8.45E-03 | 4.19E-03 |
| 10475 | Cic | CIC | 6.66E-03 | -1.06E-03 |
| 10476 | Chd2 | CHD2 | 8.53E-03 | -3.22E-02 |
| 10477 | Zc3h10 | ZC3H10 | 8.90E-03 | -5.57E-03 |
| 10478 | Stac2 | STAC2 | 1.12E-02 | -7.34E-03 |
| 10479 | Slc24a5 | SLC24A5 | -4.74E-02 | -6.45E-02 |
| 10480 | Impdh2 | IMPDH2 | -7.27E-03 | 3.16E-02 |
| 10481 | Fdft1 | FDFT1 | -1.40E-02 | -1.24E-02 |
| 10482 | Alkbh8 | ALKBH8 | -8.63E-03 | -1.13E-03 |
| 10483 | Pdzk1 | PDZK1 | -1.17E-02 | -1.82E-02 |
| 10484 | Hsf2 | HSF2 | 1.09E-02 | -3.15E-04 |
| 10485 | Fbxl21 | FBXL21 | 2.20E-02 | -6.23E-02 |
| 10486 | BC024978 | C19orf54 | 1.03E-02 | -3.40E-03 |
| 10487 | Rprm | RPRM | -7.70E-02 | 1.50E-01 |
| 10488 | Entpd7 | ENTPD7 | -1.37E-02 | 1.51E-02 |
| 10489 | Rpl3l | RPL3L | 2.68E-02 | 1.29E-02 |
| 10490 | Xpo7 | XPO7 | -7.86E-03 | -1.86E-02 |
| 10491 | Stk25 | STK25 | -5.06E-03 | 3.49E-04 |

| | | | | |
|---|---|---|---|---|
| 10492 | Mal2 | MAL2 | 3.55E-02 | 2.46E-02 |
| 10493 | Mansc1 | MANSC1 | -1.82E-02 | -3.06E-02 |
| 10494 | Lrrc8e | LRRC8E | 4.82E-02 | 1.87E-02 |
| 10495 | Apeh | APEH | -1.11E-02 | -1.93E-02 |
| 10496 | Thoc2 | THOC2 | 8.92E-03 | -6.43E-03 |
| 10497 | Eif3a | EIF3A | -7.12E-03 | 4.96E-03 |
| 10498 | Fmn1 | FMN1 | -9.41E-03 | -3.17E-02 |
| 10499 | Asnsd1 | ASNSD1 | 1.89E-02 | 4.28E-03 |
| 10500 | Ccdc78 | CCDC78 | -1.66E-02 | 2.97E-02 |
| 10501 | Scly | SCLY | 9.87E-03 | -1.61E-02 |
| 10502 | Ms4a6d | MS4A6A | 4.59E-02 | 2.51E-01 |
| 10503 | Casp7 | CASP7 | -1.74E-02 | -1.68E-02 |
| 10504 | Col4a3 | COL4A3 | 8.00E-03 | -3.49E-02 |
| 10505 | Gmeb2 | GMEB2 | 8.26E-03 | 6.73E-03 |
| 10506 | Trappc13 | TRAPPC13 | 6.70E-03 | -9.54E-04 |
| 10507 | Zfp174 | ZNF174 | -1.75E-02 | -1.12E-02 |
| 10508 | Naaladl2 | NAALADL2 | -3.82E-02 | 5.14E-02 |
| 10509 | Myo7a | MYO7A | 1.61E-02 | -3.31E-02 |
| 10510 | Gal3st3 | GAL3ST3 | 2.14E-02 | -1.64E-02 |
| 10511 | Pcbd1 | PCBD1 | 9.28E-03 | -3.32E-02 |
| 10512 | Gpha2 | GPHA2 | 1.36E-02 | 9.12E-03 |
| 10513 | Adi1 | ADI1 | 8.62E-03 | -2.27E-02 |
| 10514 | Peli3 | PELI3 | -3.05E-02 | -5.06E-02 |
| 10515 | Nlgn2 | NLGN2 | 1.75E-02 | 4.89E-02 |
| 10516 | 4930444P10Rik | LOC100130301 | -2.29E-02 | 4.65E-02 |
| 10517 | Zfp9 | ZNF25 | -2.81E-02 | -2.75E-03 |
| 10518 | Myoz1 | MYOZ1 | 1.59E-02 | 9.13E-02 |
| 10519 | Luzp1 | LUZP1 | -7.43E-03 | -3.76E-03 |
| 10520 | Sall3 | SALL3 | -2.63E-02 | 8.31E-03 |
| 10521 | Ccdc121 | CCDC121 | 1.17E-02 | -2.23E-04 |
| 10522 | Helz2 | HELZ2 | 9.46E-03 | -4.68E-03 |
| 10523 | Inha | INHA | -2.22E-02 | 4.24E-02 |
| 10524 | Slc25a33 | SLC25A33 | 2.04E-02 | -1.17E-02 |
| 10525 | Psmc6 | PSMC6 | 6.12E-03 | -3.70E-03 |
| 10526 | Tnk1 | TNK1 | -2.48E-02 | -9.62E-03 |
| 10527 | Fat1 | FAT1 | 1.06E-02 | 1.77E-03 |
| 10528 | Nxph3 | NXPH3 | 2.24E-02 | -4.60E-03 |
| 10529 | Hmga1 | HMGA1 | 2.64E-02 | 2.93E-02 |
| 10530 | Cd101 | CD101 | -1.67E-02 | 4.18E-02 |
| 10531 | AU021092 | C16orf89 | -4.25E-02 | 4.31E-02 |
| 10532 | Spaca7 | SPACA7 | -2.80E-02 | 1.31E-01 |
| 10533 | Gk5 | GK5 | 1.72E-02 | 1.93E-02 |
| 10534 | Dis3l | DIS3L | -7.64E-03 | -3.14E-02 |
| 10535 | Mpp4 | MPP4 | 4.61E-02 | 1.74E-02 |
| 10536 | Gon4l | GON4L | 5.75E-03 | 4.49E-04 |
| 10537 | Zfp697 | ZNF697 | -1.94E-02 | 3.02E-02 |
| 10538 | Dyrk1a | DYRK1A | -5.88E-03 | -5.99E-04 |
| 10539 | Camsap2 | CAMSAP2 | -8.53E-03 | -1.98E-03 |
| 10540 | Kcnip2 | KCNIP2 | -2.77E-02 | -5.58E-02 |
| 10541 | Gtf3c5 | GTF3C5 | -6.38E-03 | -3.62E-03 |
| 10542 | Itpripl2 | ITPRIPL2 | 1.23E-02 | -8.85E-04 |
| 10543 | Arl6 | ARL6 | -7.04E-03 | -8.68E-03 |
| 10544 | Ntsr1 | NTSR1 | 1.68E-02 | 7.53E-02 |

| | | | | |
|---|---|---|---|---|
| 10545 | Ipo9 | IPO9 | -6.33E-03 | -6.65E-03 |
| 10546 | Pla1a | PLA1A | 4.12E-02 | -8.69E-02 |
| 10547 | Ube2u | UBE2U | 1.69E-02 | -3.70E-03 |
| 10548 | Rsph10b | RSPH10B | 2.24E-02 | -2.31E-03 |
| 10549 | Rsph10b | RSPH10B2 | 2.24E-02 | -2.31E-03 |
| 10550 | Ap3s1 | AP3S1 | 9.17E-03 | 1.47E-02 |
| 10551 | Alpk1 | ALPK1 | -2.14E-02 | 6.67E-02 |
| 10552 | Fam229a | FAM229A | 1.93E-02 | -1.02E-02 |
| 10553 | Fbxo32 | FBXO32 | 2.45E-02 | -4.06E-02 |
| 10554 | Peg10 | PEG10 | -3.99E-02 | 4.89E-03 |
| 10555 | F3 | F3 | -1.56E-02 | 5.81E-02 |
| 10556 | Pskh1 | PSKH1 | 5.80E-03 | -2.06E-02 |
| 10557 | Fam181a | FAM181A | 1.10E-02 | -2.47E-02 |
| 10558 | Adrb3 | ADRB3 | -6.08E-02 | 3.57E-02 |
| 10559 | Lyrm9 | LYRM9 | 7.94E-03 | -3.53E-02 |
| 10560 | Ccdc40 | CCDC40 | 2.00E-02 | 1.41E-02 |
| 10561 | Hgd | HGD | -1.12E-02 | -2.37E-02 |
| 10562 | Naip2 | NAIP | -2.43E-02 | 4.33E-02 |
| 10563 | Ankrd34a | ANKRD34A | 1.57E-02 | 7.79E-03 |
| 10564 | Trim55 | TRIM55 | 5.31E-03 | 7.67E-03 |
| 10565 | Ccdc160 | CCDC160 | 1.69E-02 | 2.80E-02 |
| 10566 | Grb14 | GRB14 | -1.17E-02 | 5.92E-03 |
| 10567 | Sike1 | SIKE1 | -6.49E-03 | -6.36E-03 |
| 10568 | Map3k11 | MAP3K11 | 6.15E-03 | 1.57E-03 |
| 10569 | Fgf11 | FGF11 | 3.18E-02 | -5.30E-02 |
| 10570 | Aldh18a1 | ALDH18A1 | 2.26E-02 | 8.85E-02 |
| 10571 | Sp3 | SP3 | -8.24E-03 | 7.26E-03 |
| 10572 | Neil1 | NEIL1 | 9.81E-03 | -7.83E-03 |
| 10573 | Tnc | TNC | 6.15E-02 | 8.19E-02 |
| 10574 | Stx1a | STX1A | 3.33E-02 | 2.12E-02 |
| 10575 | Xrcc2 | XRCC2 | -2.11E-02 | 2.72E-02 |
| 10576 | Lsm6 | LSM6 | 1.45E-02 | -5.17E-04 |
| 10577 | Prex1 | PREX1 | -1.40E-02 | 5.39E-05 |
| 10578 | Tomm40l | TOMM40L | -6.97E-03 | -6.32E-03 |
| 10579 | Golga7 | GOLGA7 | 6.06E-03 | -9.78E-03 |
| 10580 | Abr | ABR | -5.52E-03 | -1.67E-02 |
| 10581 | Des | DES | -2.03E-02 | -1.18E-02 |
| 10582 | Ostc | OSTC | -1.09E-02 | 2.06E-02 |
| 10583 | Dnajc22 | DNAJC22 | 1.33E-02 | -1.62E-02 |
| 10584 | Neurl2 | NEURL2 | 2.37E-02 | 3.46E-02 |
| 10585 | Prkdc | PRKDC | -8.19E-03 | -2.44E-03 |
| 10586 | Spp2 | SPP2 | -1.84E-02 | -7.25E-03 |
| 10587 | Tarsl2 | TARSL2 | -1.14E-02 | -1.62E-02 |
| 10588 | Fkbp6 | FKBP6 | -9.31E-03 | 1.20E-02 |
| 10589 | Paf1 | PAF1 | 5.13E-03 | 4.29E-04 |
| 10590 | B4galt7 | B4GALT7 | -1.04E-02 | 1.38E-02 |
| 10591 | Zfp791 | ZNF791 | -2.01E-02 | -4.85E-02 |
| 10592 | Lipa | LIPA | -2.19E-02 | -2.93E-02 |
| 10593 | Wipi2 | WIPI2 | -6.22E-03 | -1.58E-02 |
| 10594 | Nipal3 | NIPAL3 | -7.88E-03 | -3.59E-02 |
| 10595 | Rab3gap1 | RAB3GAP1 | 5.07E-03 | -1.51E-02 |
| 10596 | Sptbn1 | SPTBN1 | 6.20E-03 | -1.93E-02 |
| 10597 | Ttf2 | TTF2 | 9.79E-03 | -7.41E-03 |

| | | | | |
|---|---|---|---|---|
| 10598 | Wdr5 | WDR5 | 6.08E-03 | 1.88E-03 |
| 10599 | Gtpbp2 | GTPBP2 | -6.42E-03 | -1.23E-02 |
| 10600 | Ccar1 | CCAR1 | 7.02E-03 | 1.09E-02 |
| 10601 | Katnb1 | KATNB1 | 1.51E-02 | -2.22E-02 |
| 10602 | Pdzd8 | PDZD8 | -5.75E-03 | -4.26E-03 |
| 10603 | Ahcy | AHCY | 1.21E-02 | -1.30E-02 |
| 10604 | Gpc4 | GPC4 | -9.65E-03 | -9.17E-03 |
| 10605 | Pcdhgc3 | PCDHGC3 | -3.15E-02 | -3.34E-03 |
| 10606 | Aff1 | AFF1 | 8.26E-03 | -1.10E-02 |
| 10607 | Mrc2 | MRC2 | 2.16E-02 | 2.84E-02 |
| 10608 | Igdcc3 | IGDCC3 | -4.22E-02 | 2.36E-03 |
| 10609 | Kcnj5 | KCNJ5 | 2.91E-02 | -8.06E-02 |
| 10610 | Ankrd13c | ANKRD13C | -9.79E-03 | -1.77E-02 |
| 10611 | Il12b | IL12B | -1.08E-02 | 2.21E-02 |
| 10612 | Abhd8 | ABHD8 | 7.52E-03 | -1.45E-02 |
| 10613 | 1110017D15Rik | C9orf24 | 1.78E-02 | 7.97E-03 |
| 10614 | Ncor2 | NCOR2 | -5.85E-03 | 7.27E-03 |
| 10615 | Usp13 | USP13 | -2.80E-02 | 4.67E-02 |
| 10616 | Tmem209 | TMEM209 | -7.91E-03 | -6.13E-03 |
| 10617 | Slc5a3 | SLC5A3 | -3.91E-02 | -5.59E-02 |
| 10618 | Armc10 | ARMC10 | -8.81E-03 | 2.62E-02 |
| 10619 | Cnep1r1 | CNEP1R1 | -6.46E-03 | 3.42E-03 |
| 10620 | Rbm4 | RBM4 | -2.09E-02 | -4.19E-03 |
| 10621 | Dars | DARS | 7.14E-03 | -1.77E-02 |
| 10622 | Scgb3a1 | SCGB3A1 | 7.68E-03 | 1.45E-04 |
| 10623 | Pdcd1 | PDCD1 | -1.47E-02 | 1.61E-02 |
| 10624 | Med30 | MED30 | -1.05E-02 | 1.51E-02 |
| 10625 | Tmem123 | TMEM123 | 8.89E-03 | 1.13E-02 |
| 10626 | Phospho2 | PHOSPHO2 | 1.06E-02 | 1.28E-02 |
| 10627 | Tiam1 | TIAM1 | 5.33E-02 | -3.06E-02 |
| 10628 | Prdx1 | PRDX1 | 8.36E-03 | -4.19E-03 |
| 10629 | Rmdn3 | RMDN3 | 7.97E-03 | -1.06E-02 |
| 10630 | 1110059G10Rik | KIAA1143 | 8.34E-03 | 3.43E-03 |
| 10631 | B4galt6 | B4GALT6 | 2.27E-02 | 8.72E-03 |
| 10632 | Rims3 | RIMS3 | -2.11E-02 | 1.03E-02 |
| 10633 | Slc10a1 | SLC10A1 | 3.01E-02 | 3.62E-02 |
| 10634 | Trim37 | TRIM37 | 6.20E-03 | -2.12E-03 |
| 10635 | Hs3st6 | HS3ST6 | -2.20E-02 | -2.81E-02 |
| 10636 | Cstf3 | CSTF3 | -9.97E-03 | 1.10E-02 |
| 10637 | Pex13 | PEX13 | -7.41E-03 | 6.16E-05 |
| 10638 | Nvl | NVL | 6.78E-03 | 6.24E-04 |
| 10639 | Arih1 | ARIH1 | 6.25E-03 | -9.89E-04 |
| 10640 | Rapgef6 | RAPGEF6 | -8.00E-03 | -1.25E-02 |
| 10641 | Ncbp1 | NCBP1 | -6.59E-03 | 1.74E-03 |
| 10642 | Syndig1 | SYNDIG1 | -1.38E-02 | 3.38E-02 |
| 10643 | Draxin | DRAXIN | 8.35E-03 | 1.40E-02 |
| 10644 | Cacybp | CACYBP | -1.47E-02 | -6.97E-03 |
| 10645 | Wwc1 | WWC1 | -7.88E-03 | -1.48E-02 |
| 10646 | Adamts1 | ADAMTS1 | 1.93E-02 | 8.32E-02 |
| 10647 | Tnfsf13b | TNFSF13B | -2.66E-02 | 9.78E-02 |
| 10648 | Mterf1b | MTERF | -2.39E-02 | -2.18E-02 |
| 10649 | Kin | KIN | -1.11E-02 | 1.34E-02 |
| 10650 | Luzp2 | LUZP2 | -3.90E-02 | -1.13E-01 |

| | | | | |
|---|---|---|---|---|
| 10651 | Trib1 | TRIB1 | 3.01E-02 | 3.09E-03 |
| 10652 | Idua | IDUA | -1.16E-02 | -9.25E-03 |
| 10653 | Sit1 | SIT1 | -1.54E-02 | 9.45E-03 |
| 10654 | Spag6 | SPAG6 | 8.88E-03 | -7.00E-03 |
| 10655 | Klf2 | KLF2 | -2.47E-02 | 2.69E-02 |
| 10656 | Amn | AMN | -1.08E-02 | -4.33E-03 |
| 10657 | Cdc42bpa | CDC42BPA | -6.40E-03 | -2.54E-02 |
| 10658 | Pfdn4 | PFDN4 | -1.06E-02 | 1.32E-02 |
| 10659 | Elfn1 | ELFN1 | -6.72E-02 | -1.77E-01 |
| 10660 | 1700001P01Rik | C17orf98 | 1.46E-02 | -1.46E-02 |
| 10661 | Tiparp | TIPARP | -1.77E-02 | 3.50E-02 |
| 10662 | Ppp1r18 | PPP1R18 | 1.37E-02 | 5.35E-02 |
| 10663 | Zfp458 | ZNF43 | -2.25E-02 | 2.20E-02 |
| 10664 | Plrg1 | PLRG1 | -5.84E-03 | -2.04E-03 |
| 10665 | Map9 | MAP9 | 1.81E-02 | 2.03E-02 |
| 10666 | Osmr | OSMR | 1.88E-02 | 6.41E-02 |
| 10667 | Golt1a | GOLT1A | 2.40E-02 | -2.06E-02 |
| 10668 | Zfp110 | ZNF274 | 6.55E-03 | -5.59E-03 |
| 10669 | Prss57 | PRSS57 | -1.10E-02 | 1.26E-02 |
| 10670 | Gm11837 | C8orf88 | -2.49E-02 | -5.91E-02 |
| 10671 | Glb1l | GLB1L | 9.87E-03 | -1.99E-02 |
| 10672 | Cacna1e | CACNA1E | -3.06E-02 | 4.26E-03 |
| 10673 | Slc30a4 | SLC30A4 | 2.41E-02 | -3.74E-02 |
| 10674 | Srsf9 | SRSF9 | 6.79E-03 | -1.77E-03 |
| 10675 | Pdgfrl | PDGFRL | 2.95E-02 | -1.19E-02 |
| 10676 | Ehmt2 | EHMT2 | 5.09E-03 | -8.47E-03 |
| 10677 | Plekhs1 | PLEKHS1 | 3.26E-02 | -2.43E-02 |
| 10678 | Cd28 | CD28 | -9.52E-03 | -5.56E-03 |
| 10679 | Txlna | TXLNA | -4.51E-03 | 3.01E-03 |
| 10680 | Sco1 | SCO1 | -7.35E-03 | -8.56E-03 |
| 10681 | Nol11 | NOL11 | 6.61E-03 | -6.02E-03 |
| 10682 | Trp73 | TP73 | 1.62E-02 | -9.28E-03 |
| 10683 | Derl3 | DERL3 | -6.28E-02 | 7.18E-02 |
| 10684 | Nfat5 | NFAT5 | -1.02E-02 | -7.17E-03 |
| 10685 | Ppp1r3e | PPP1R3E | -1.88E-02 | -1.53E-02 |
| 10686 | Tvp23b | TVP23B | -5.39E-03 | -1.43E-02 |
| 10687 | Rab44 | RAB44 | 1.86E-02 | 3.19E-02 |
| 10688 | Bag3 | BAG3 | -2.19E-02 | 1.82E-02 |
| 10689 | Terf2 | TERF2 | 5.97E-03 | -7.09E-04 |
| 10690 | Bricd5 | BRICD5 | -1.91E-02 | 6.01E-03 |
| 10691 | Kif13a | KIF13A | -6.71E-03 | -3.40E-02 |
| 10692 | Lrrc71 | LRRC71 | 2.43E-02 | 1.24E-02 |
| 10693 | Cldn19 | CLDN19 | -3.68E-02 | -4.45E-02 |
| 10694 | Hirip3 | HIRIP3 | 9.62E-03 | 7.30E-03 |
| 10695 | Zfp148 | ZNF148 | -5.75E-03 | -1.79E-03 |
| 10696 | Slfnl1 | SLFNL1 | 9.46E-03 | -2.71E-02 |
| 10697 | Cwc22 | CWC22 | 9.56E-03 | 3.15E-03 |
| 10698 | Cul7 | CUL7 | 6.70E-03 | -4.60E-03 |
| 10699 | Blzf1 | BLZF1 | 7.22E-03 | 1.79E-03 |
| 10700 | Phf21a | PHF21A | 6.85E-03 | -1.56E-02 |
| 10701 | Fam172a | FAM172A | 8.30E-03 | -7.76E-03 |
| 10702 | Fbp2 | FBP2 | -2.03E-02 | -4.58E-02 |
| 10703 | Trrap | TRRAP | 6.12E-03 | -1.60E-02 |

| | | | | |
|---|---|---|---|---|
| 10704 | Matn3 | MATN3 | 6.82E-03 | 4.54E-03 |
| 10705 | Kctd6 | KCTD6 | 9.04E-03 | -1.86E-03 |
| 10706 | Coro2a | CORO2A | 3.48E-02 | 1.37E-01 |
| 10707 | Gatm | GATM | -1.86E-02 | -5.52E-02 |
| 10708 | Rmnd5a | RMND5A | 6.45E-03 | -2.30E-02 |
| 10709 | Galnt6 | GALNT6 | 3.22E-02 | 9.75E-02 |
| 10710 | Aga | AGA | -7.69E-03 | -1.01E-02 |
| 10711 | Map3k14 | MAP3K14 | 1.68E-02 | 1.12E-02 |
| 10712 | Tsga10 | TSGA10 | 2.57E-02 | -3.43E-02 |
| 10713 | Lmbr1 | LMBR1 | -7.92E-03 | 4.91E-03 |
| 10714 | Ss18 | SS18 | -4.12E-03 | 5.08E-03 |
| 10715 | Klhl15 | KLHL15 | 1.74E-02 | -1.21E-02 |
| 10716 | Pla2g2d | PLA2G2D | -1.13E-02 | -8.65E-03 |
| 10717 | Tgif1 | TGIF1 | -2.00E-02 | 8.45E-02 |
| 10718 | Slc8a3 | SLC8A3 | 3.32E-02 | -1.18E-02 |
| 10719 | Clybl | CLYBL | 8.29E-03 | -2.60E-02 |
| 10720 | Atg12 | ATG12 | -4.70E-03 | -1.08E-05 |
| 10721 | Hhla1 | HHLA1 | -1.02E-02 | 1.83E-02 |
| 10722 | Mall | MALL | 2.25E-02 | 1.34E-01 |
| 10723 | Dph7 | DPH7 | 9.41E-03 | -2.14E-02 |
| 10724 | Dnah6 | DNAH6 | -1.49E-02 | 3.24E-02 |
| 10725 | Lmnb1 | LMNB1 | 2.29E-02 | 6.22E-02 |
| 10726 | Pcdhb3 | PCDHB3 | 1.87E-02 | -3.12E-02 |
| 10727 | Aqp9 | AQP9 | -1.81E-02 | 5.42E-02 |
| 10728 | Mettl6 | METTL6 | 9.76E-03 | 4.30E-03 |
| 10729 | Haus6 | HAUS6 | 1.21E-02 | 1.03E-02 |
| 10730 | Dtd1 | DTD1 | 7.50E-03 | -3.12E-02 |
| 10731 | Txnl4b | TXNL4B | 9.29E-03 | -1.03E-02 |
| 10732 | Dpy19l4 | DPY19L4 | -9.48E-03 | -2.21E-02 |
| 10733 | Tm6sf2 | TM6SF2 | -2.02E-02 | 6.20E-02 |
| 10734 | Adam8 | ADAM8 | 4.09E-02 | 1.16E-01 |
| 10735 | Tacr3 | TACR3 | -1.59E-02 | 5.27E-02 |
| 10736 | Wwp1 | WWP1 | 7.66E-03 | -1.04E-02 |
| 10737 | Ranbp2 | RGPD1 | -5.93E-03 | -7.56E-03 |
| 10738 | Ranbp2 | RGPD2 | -5.93E-03 | -7.56E-03 |
| 10739 | Ranbp2 | RGPD4 | -5.93E-03 | -7.56E-03 |
| 10740 | Ranbp2 | RGPD3 | -5.93E-03 | -7.56E-03 |
| 10741 | Ranbp2 | RGPD5 | -5.93E-03 | -7.56E-03 |
| 10742 | Ranbp2 | RGPD6 | -5.93E-03 | -7.56E-03 |
| 10743 | Ranbp2 | RGPD8 | -5.93E-03 | -7.56E-03 |
| 10744 | Tmem138 | TMEM138 | -1.44E-02 | 1.38E-04 |
| 10745 | Ribc2 | RIBC2 | 1.28E-02 | -3.03E-02 |
| 10746 | Rcbtb1 | RCBTB1 | 7.89E-03 | 3.29E-03 |
| 10747 | Prox1 | PROX1 | -4.56E-02 | -5.16E-02 |
| 10748 | Ly6d | LY6D | -1.16E-01 | 1.96E-01 |
| 10749 | Adk | ADK | 9.23E-03 | -1.37E-02 |
| 10750 | Xkr6 | XKR6 | -1.69E-02 | 3.80E-02 |
| 10751 | Os9 | OS9 | -5.41E-03 | -6.46E-03 |
| 10752 | Pigr | PIGR | -3.87E-02 | 4.02E-02 |
| 10753 | Chst12 | CHST12 | -2.10E-02 | 2.85E-02 |
| 10754 | Atxn7 | ATXN7 | -8.22E-03 | -8.63E-03 |
| 10755 | Urb1 | URB1 | -1.04E-02 | -9.00E-03 |
| 10756 | Zfp423 | ZNF423 | -2.10E-02 | -3.62E-02 |

| | | | | |
|---|---|---|---|---|
| 10757 | Ifit1 | IFIT1B | -2.81E-02 | 1.21E-01 |
| 10758 | Hmgxb3 | HMGXB3 | 5.57E-03 | -1.03E-02 |
| 10759 | Fscn1 | FSCN1 | -1.95E-02 | 3.06E-02 |
| 10760 | Fbxo16 | FBXO16 | 2.77E-02 | -2.65E-02 |
| 10761 | Cdkn2aipnl | CDKN2AIPNL | 7.00E-03 | 3.32E-02 |
| 10762 | Gng2 | GNG2 | -2.46E-02 | -2.52E-02 |
| 10763 | H2-Q2 | HLA-A | -3.31E-02 | 2.22E-02 |
| 10764 | Zfp318 | ZNF318 | -7.72E-03 | 3.06E-03 |
| 10765 | Grhl2 | GRHL2 | -2.66E-02 | 1.11E-02 |
| 10766 | Dennd6a | DENND6A | -7.00E-03 | -1.54E-02 |
| 10767 | Ugdh | UGDH | 1.69E-02 | -2.02E-02 |
| 10768 | Mettl23 | METTL23 | -8.19E-03 | -2.12E-02 |
| 10769 | Fancd2os | FANCD2OS | 3.14E-02 | -5.28E-02 |
| 10770 | Pim1 | PIM1 | -3.46E-02 | 2.48E-02 |
| 10771 | Sec62 | SEC62 | 5.93E-03 | -1.13E-02 |
| 10772 | Rundc3b | RUNDC3B | -3.09E-02 | -5.32E-02 |
| 10773 | Gp5 | GP5 | 1.08E-02 | 4.24E-02 |
| 10774 | Flii | FLII | 4.19E-03 | -9.02E-03 |
| 10775 | Ndufs7 | NDUFS7 | 7.14E-03 | -1.99E-02 |
| 10776 | Kdm4d | KDM4D | 1.28E-02 | -2.71E-03 |
| 10777 | Setd2 | SETD2 | -3.96E-03 | -5.70E-03 |
| 10778 | Tmem245 | TMEM245 | -6.51E-03 | -1.88E-02 |
| 10779 | Cygb | CYGB | -3.43E-02 | 7.01E-02 |
| 10780 | Cyp46a1 | CYP46A1 | -2.03E-02 | -4.63E-02 |
| 10781 | Hes1 | HES1 | -2.70E-02 | -2.04E-02 |
| 10782 | Shc3 | SHC3 | 1.97E-02 | 2.48E-02 |
| 10783 | Anxa9 | ANXA9 | 3.90E-02 | 6.19E-03 |
| 10784 | 1700123O20Rik | C14orf119 | 6.01E-03 | 4.46E-03 |
| 10785 | Lss | LSS | -1.25E-02 | -9.76E-03 |
| 10786 | Slc25a35 | SLC25A35 | -1.39E-02 | -2.10E-02 |
| 10787 | Gabbr1 | GABBR1 | -7.78E-03 | -1.41E-02 |
| 10788 | Tbc1d5 | TBC1D5 | -8.04E-03 | -2.33E-02 |
| 10789 | Dtwd1 | DTWD1 | -1.12E-02 | 2.40E-02 |
| 10790 | Serinc4 | SERINC4 | 2.15E-02 | 4.50E-02 |
| 10791 | Ano2 | ANO2 | -1.63E-02 | -6.25E-03 |
| 10792 | Pf4 | PF4 | -2.96E-02 | 1.14E-01 |
| 10793 | Adora2a | ADORA2A | -3.46E-02 | 1.69E-02 |
| 10794 | Ptk7 | PTK7 | 1.46E-02 | -9.31E-05 |
| 10795 | Rasd1 | RASD1 | 5.77E-02 | -1.47E-02 |
| 10796 | Hint3 | HINT3 | 8.03E-03 | -1.99E-02 |
| 10797 | Icos | ICOS | -1.07E-02 | 1.62E-02 |
| 10798 | Prkab2 | PRKAB2 | 1.14E-02 | 1.10E-02 |
| 10799 | Smim14 | SMIM14 | 6.47E-03 | -4.11E-03 |
| 10800 | Fam47e | FAM47E | -1.38E-02 | -3.05E-02 |
| 10801 | Cse1l | CSE1L | 6.06E-03 | 7.70E-03 |
| 10802 | Snrpd3 | SNRPD3 | -6.61E-03 | -1.80E-02 |
| 10803 | Fam122b | FAM122B | -1.22E-02 | -7.02E-03 |
| 10804 | Nmb | NMB | -2.22E-02 | 2.84E-02 |
| 10805 | Errfi1 | ERRFI1 | 1.64E-02 | 1.29E-02 |
| 10806 | Fgb | FGB | -4.58E-02 | 5.01E-01 |
| 10807 | Kif3a | KIF3A | -8.40E-03 | -2.49E-02 |
| 10808 | Top2b | TOP2B | -5.27E-03 | -7.88E-03 |
| 10809 | Fgf10 | FGF10 | -3.13E-02 | 1.30E-02 |

| | | | | |
|---|---|---|---|---|
| 10810 | Efemp1 | EFEMP1 | -2.43E-02 | 2.40E-02 |
| 10811 | Rab11fip4 | RAB11FIP4 | -8.58E-03 | -1.91E-02 |
| 10812 | Tril | TRIL | -3.15E-02 | -6.45E-02 |
| 10813 | Mapk1ip1l | MAPK1IP1L | 3.74E-03 | 5.73E-03 |
| 10814 | Pcdhgb4 | PCDHGB4 | -2.75E-02 | -4.91E-02 |
| 10815 | Gpr20 | GPR20 | 1.43E-02 | 1.88E-02 |
| 10816 | Sntb2 | SNTB2 | 1.10E-02 | -1.75E-02 |
| 10817 | Cds2 | CDS2 | 4.98E-03 | -2.09E-02 |
| 10818 | Amotl2 | AMOTL2 | -1.85E-02 | -1.37E-02 |
| 10819 | Ptpn21 | PTPN21 | -8.87E-03 | -9.51E-03 |
| 10820 | Clk1 | CLK1 | -7.16E-03 | 2.15E-03 |
| 10821 | Epha3 | EPHA3 | 3.79E-02 | 7.43E-02 |
| 10822 | Zmym4 | ZMYM4 | -6.29E-03 | -1.85E-02 |
| 10823 | Eif2s3x | EIF2S3 | -9.60E-03 | -2.44E-02 |
| 10824 | Plekha7 | PLEKHA7 | -9.82E-03 | -3.70E-02 |
| 10825 | Mlana | MLANA | 1.15E-02 | 4.04E-02 |
| 10826 | Zswim5 | ZSWIM5 | -7.52E-03 | -8.49E-04 |
| 10827 | Mapk8ip3 | MAPK8IP3 | -5.09E-03 | -5.40E-03 |
| 10828 | Sdr39u1 | SDR39U1 | -6.26E-03 | -2.18E-02 |
| 10829 | Sptssb | SPTSSB | 1.59E-01 | 1.18E-01 |
| 10830 | Cnppd1 | CNPPD1 | 7.03E-03 | -1.44E-02 |
| 10831 | Edrf1 | EDRF1 | -8.03E-03 | -5.45E-03 |
| 10832 | Col1a1 | COL1A1 | 5.07E-02 | 2.57E-01 |
| 10833 | Cdk5r1 | CDK5R1 | -2.00E-02 | 3.26E-02 |
| 10834 | Tbc1d19 | TBC1D19 | -7.34E-03 | -2.03E-02 |
| 10835 | Zdhhc9 | ZDHHC9 | -8.83E-03 | -1.21E-02 |
| 10836 | Art3 | ART3 | -1.81E-02 | -3.32E-03 |
| 10837 | Asb14 | ASB14 | -1.41E-02 | 2.46E-02 |
| 10838 | Tsc2 | TSC2 | -5.92E-03 | -1.96E-02 |
| 10839 | Sppl2a | SPPL2A | 6.70E-03 | -6.43E-03 |
| 10840 | Gata3 | GATA3 | 2.63E-02 | 2.31E-02 |
| 10841 | Gdf9 | GDF9 | -1.32E-02 | 4.62E-03 |
| 10842 | Brwd1 | BRWD1 | 7.09E-03 | -7.81E-03 |
| 10843 | Srrm4 | SRRM4 | 2.53E-02 | -1.44E-02 |
| 10844 | Tmprss13 | TMPRSS13 | 7.80E-02 | 7.56E-02 |
| 10845 | Fam32a | FAM32A | 5.97E-03 | -1.89E-02 |
| 10846 | Ddx46 | DDX46 | 5.70E-03 | -9.76E-03 |
| 10847 | Hectd1 | HECTD1 | 5.68E-03 | -1.35E-02 |
| 10848 | Tmub1 | TMUB1 | -7.32E-03 | -1.28E-03 |
| 10849 | Rasal1 | RASAL1 | 9.64E-02 | -2.33E-02 |
| 10850 | Slc14a2 | SLC14A2 | 1.59E-01 | -5.84E-02 |
| 10851 | Rbfa | RBFA | 5.27E-03 | -1.35E-02 |
| 10852 | Dhx15 | DHX15 | -6.71E-03 | -3.25E-03 |
| 10853 | Pcbd2 | PCBD2 | 9.47E-03 | -2.15E-02 |
| 10854 | Cox4i2 | COX4I2 | -1.94E-02 | -6.76E-02 |
| 10855 | Gipr | GIPR | -8.11E-03 | 1.73E-02 |
| 10856 | Vezf1 | VEZF1 | 1.45E-02 | -1.71E-02 |
| 10857 | Gorab | GORAB | 1.12E-02 | -2.07E-02 |
| 10858 | Armc3 | ARMC3 | -1.09E-02 | -8.01E-03 |
| 10859 | Fastk | FASTK | 6.45E-03 | -1.86E-02 |
| 10860 | Nat9 | NAT9 | -6.83E-03 | -2.87E-02 |
| 10861 | Actn1 | ACTN1 | 9.98E-03 | 5.21E-02 |
| 10862 | Nap1l4 | NAP1L4 | -4.84E-03 | 1.65E-03 |

| | | | | |
|---|---|---|---|---|
| 10863 | Bcam | BCAM | 2.34E-02 | 2.89E-02 |
| 10864 | Derl2 | DERL2 | 7.42E-03 | -4.36E-03 |
| 10865 | Cbx3 | CBX3 | -6.17E-03 | -5.76E-03 |
| 10866 | Sbf2 | SBF2 | 7.66E-03 | -2.57E-02 |
| 10867 | Hic1 | HIC1 | -2.78E-02 | 2.60E-02 |
| 10868 | Ppp1r16a | PPP1R16A | 5.71E-03 | -1.65E-02 |
| 10869 | Pfdn5 | PFDN5 | -5.45E-03 | -8.20E-03 |
| 10870 | Hsp90aa1 | HSP90AA1 | -1.27E-02 | 2.10E-03 |
| 10871 | Slc6a9 | SLC6A9 | 1.27E-02 | -3.73E-02 |
| 10872 | Sostdc1 | SOSTDC1 | 1.47E-02 | 1.40E-02 |
| 10873 | Exog | EXOG | -1.66E-02 | 1.83E-02 |
| 10874 | Ap1m2 | AP1M2 | -1.29E-02 | 1.10E-03 |
| 10875 | Coq5 | COQ5 | -6.51E-03 | -1.89E-02 |
| 10876 | Cacna1c | CACNA1C | 2.96E-02 | -4.36E-02 |
| 10877 | Ndufaf6 | NDUFAF6 | -9.24E-03 | -9.26E-03 |
| 10878 | Rpgr | RPGR | -1.33E-02 | -2.51E-02 |
| 10879 | Prdm16 | PRDM16 | 1.85E-02 | -3.51E-02 |
| 10880 | Ifit2 | IFIT2 | -2.52E-02 | 1.11E-02 |
| 10881 | Ppib | PPIB | 6.16E-03 | 4.17E-03 |
| 10882 | Lin7c | LIN7C | 6.28E-03 | 2.16E-03 |
| 10883 | Cenpl | CENPL | 1.94E-02 | 4.24E-02 |
| 10884 | Calcoco1 | CALCOCO1 | 6.68E-03 | -1.79E-02 |
| 10885 | Spatc1 | SPATC1 | 2.73E-02 | 5.88E-04 |
| 10886 | Prr16 | PRR16 | 1.05E-02 | 9.00E-03 |
| 10887 | Ndufaf5 | NDUFAF5 | 8.52E-03 | -1.29E-02 |
| 10888 | Mks1 | MKS1 | 1.01E-02 | -1.61E-02 |
| 10889 | Nbea | NBEA | -7.07E-03 | 9.88E-03 |
| 10890 | Lmf2 | LMF2 | -6.62E-03 | 5.95E-03 |
| 10891 | Tef | TEF | 1.02E-02 | -5.07E-02 |
| 10892 | Smug1 | SMUG1 | 8.41E-03 | 8.19E-03 |
| 10893 | Phf2 | PHF2 | 4.50E-03 | -2.38E-02 |
| 10894 | Aarsd1 | AARSD1 | 1.87E-02 | 1.10E-02 |
| 10895 | Senp2 | SENP2 | 6.77E-03 | -5.22E-03 |
| 10896 | Vps54 | VPS54 | 4.54E-03 | -7.92E-03 |
| 10897 | Guca1a | GUCA1A | 2.26E-02 | -9.21E-03 |
| 10898 | Pde7a | PDE7A | -9.56E-03 | -1.75E-02 |
| 10899 | Isg20l2 | ISG20L2 | 7.89E-03 | 3.33E-03 |
| 10900 | Pus7 | PUS7 | -9.54E-03 | 5.45E-03 |
| 10901 | Ago1 | AGO1 | 5.84E-03 | -1.79E-02 |
| 10902 | Nefh | NEFH | 1.51E-02 | -9.45E-03 |
| 10903 | 1110032A03Rik | C11orf1 | 5.89E-03 | -1.49E-02 |
| 10904 | Atoh8 | ATOH8 | -3.86E-02 | 6.77E-02 |
| 10905 | Barx2 | BARX2 | -1.25E-02 | 6.06E-02 |
| 10906 | Rnf216 | RNF216 | 5.84E-03 | -4.63E-03 |
| 10907 | Dnaja2 | DNAJA2 | -5.97E-03 | 2.73E-03 |
| 10908 | 9930104L06Rik | C1orf109 | -1.08E-02 | -1.95E-02 |
| 10909 | Spns2 | SPNS2 | 1.12E-02 | 5.24E-03 |
| 10910 | Adh4 | ADH4 | -7.90E-03 | 7.59E-03 |
| 10911 | Trim7 | TRIM7 | -1.32E-02 | -2.77E-02 |
| 10912 | Eif4ebp1 | EIF4EBP1 | 1.55E-02 | 3.34E-02 |
| 10913 | Gadd45a | GADD45A | 1.48E-02 | 9.30E-02 |
| 10914 | Slc2a8 | SLC2A8 | 1.02E-02 | -8.62E-03 |
| 10915 | Atf3 | ATF3 | 7.47E-02 | 1.36E-01 |

| | | | | |
|---|---|---|---|---|
| 10916 | Arhgef10l | ARHGEF10L | 1.04E-02 | 6.64E-03 |
| 10917 | Edn3 | EDN3 | 3.89E-02 | -6.90E-02 |
| 10918 | Taf9b | TAF9B | -8.18E-03 | -3.16E-02 |
| 10919 | Drd3 | DRD3 | -1.93E-02 | 1.01E-02 |
| 10920 | Reep1 | REEP1 | 2.21E-02 | 1.37E-01 |
| 10921 | Pias3 | PIAS3 | 9.50E-03 | -1.90E-02 |
| 10922 | Cnbd2 | CNBD2 | 1.22E-02 | -2.13E-03 |
| 10923 | Taf10 | TAF10 | 5.81E-03 | 8.24E-03 |
| 10924 | Kcnf1 | KCNF1 | 2.86E-02 | -1.99E-02 |
| 10925 | Plek | PLEK | -4.72E-02 | 1.37E-01 |
| 10926 | Pold4 | POLD4 | -1.43E-02 | 1.03E-02 |
| 10927 | Dnajc19 | DNAJC19 | 1.01E-02 | -1.99E-03 |
| 10928 | Cacna1h | CACNA1H | -2.48E-02 | -6.04E-02 |
| 10929 | Gcnt3 | GCNT3 | 1.21E-01 | 8.49E-02 |
| 10930 | Ticam2 | TICAM2 | -1.86E-02 | -3.51E-03 |
| 10931 | Syne3 | SYNE3 | -1.95E-02 | -2.66E-02 |
| 10932 | Phlpp1 | PHLPP1 | 9.08E-03 | -3.40E-02 |
| 10933 | Srp14 | SRP14 | -5.69E-03 | 1.41E-03 |
| 10934 | Tmem30b | TMEM30B | 3.65E-02 | -8.58E-03 |
| 10935 | Fam126a | FAM126A | -1.02E-02 | 5.94E-03 |
| 10936 | Csrnp1 | CSRNP1 | 4.54E-02 | 1.91E-03 |
| 10937 | Twist1 | TWIST1 | 2.07E-02 | 1.61E-02 |
| 10938 | Tal1 | TAL1 | -1.64E-02 | -1.19E-02 |
| 10939 | Ccdc177 | CCDC177 | -8.96E-03 | -5.91E-03 |
| 10940 | Mrps24 | MRPS24 | 7.08E-03 | 1.39E-03 |
| 10941 | Catsper4 | CATSPER4 | -3.50E-02 | -1.04E-02 |
| 10942 | Atpaf2 | ATPAF2 | 5.88E-03 | -1.18E-02 |
| 10943 | Ddx10 | DDX10 | -6.83E-03 | 2.34E-02 |
| 10944 | Fam149a | FAM149A | -8.09E-03 | 3.81E-04 |
| 10945 | Pik3r4 | PIK3R4 | -5.17E-03 | -1.03E-02 |
| 10946 | M1ap | M1AP | 1.05E-02 | -3.47E-03 |
| 10947 | Acadsb | ACADSB | -7.44E-03 | -3.40E-02 |
| 10948 | Map3k13 | MAP3K13 | 2.30E-02 | -4.73E-02 |
| 10949 | Prelid2 | PRELID2 | 2.97E-02 | 4.13E-03 |
| 10950 | Chd3 | CHD3 | -9.92E-03 | -5.68E-03 |
| 10951 | Pias1 | PIAS1 | -5.10E-03 | -1.43E-02 |
| 10952 | Itih2 | ITIH2 | 2.65E-02 | 2.72E-02 |
| 10953 | Dnajc12 | DNAJC12 | -1.55E-02 | -4.38E-02 |
| 10954 | Pomgnt1 | POMGNT1 | 6.67E-03 | -1.09E-02 |
| 10955 | Susd4 | SUSD4 | 2.17E-02 | -3.00E-02 |
| 10956 | Snrpd1 | SNRPD1 | 7.78E-03 | 1.24E-02 |
| 10957 | Tmem207 | TMEM207 | 4.81E-02 | -1.74E-01 |
| 10958 | 1700003E16Rik | C2orf81 | -2.31E-02 | 6.94E-03 |
| 10959 | Tardbp | TARDBP | 5.70E-03 | -6.38E-04 |
| 10960 | Csnk1g2 | CSNK1G2 | 4.36E-03 | -1.29E-02 |
| 10961 | Prps1 | PRPS1 | 1.04E-02 | -3.60E-03 |
| 10962 | Cyp1a2 | CYP1A2 | -9.96E-03 | 2.46E-02 |
| 10963 | Cdc6 | CDC6 | 3.70E-02 | 1.90E-01 |
| 10964 | Ap1g1 | AP1G1 | -6.85E-03 | -2.12E-02 |
| 10965 | Layn | LAYN | 2.79E-02 | 1.07E-02 |
| 10966 | Nmral1 | NMRAL1 | 2.54E-02 | -5.90E-02 |
| 10967 | Gad2 | GAD2 | 7.58E-03 | -1.49E-02 |
| 10968 | Ern2 | ERN2 | -2.67E-02 | 2.56E-02 |

| | | | | |
|---|---|---|---|---|
| 10969 | Il12a | IL12A | 6.67E-03 | -1.81E-03 |
| 10970 | Syce1 | SYCE1 | -7.99E-03 | 1.73E-02 |
| 10971 | Daglb | DAGLB | 1.13E-02 | 7.36E-03 |
| 10972 | Armc5 | ARMC5 | 7.26E-03 | -6.98E-03 |
| 10973 | Mal | MAL | 1.27E-02 | 1.51E-02 |
| 10974 | Klk1 | KLK3 | 9.34E-03 | 2.82E-02 |
| 10975 | Ugt1a9 | UGT1A10 | -3.52E-02 | 3.21E-02 |
| 10976 | Ugt1a9 | UGT1A8 | -3.52E-02 | 3.21E-02 |
| 10977 | Ugt1a9 | UGT1A7 | -3.52E-02 | 3.21E-02 |
| 10978 | Ugt1a9 | UGT1A9 | -3.52E-02 | 3.21E-02 |
| 10979 | Prune2 | PRUNE2 | 3.23E-02 | 4.65E-02 |
| 10980 | Rps16 | RPS16 | 6.57E-03 | 6.94E-03 |
| 10981 | Myl3 | MYL3 | 1.45E-02 | -1.46E-02 |
| 10982 | Olfm3 | OLFM3 | 2.32E-02 | -6.63E-02 |
| 10983 | Ctxn3 | CTXN3 | 4.76E-02 | 4.83E-02 |
| 10984 | Rhobtb1 | RHOBTB1 | -2.50E-02 | 3.80E-02 |
| 10985 | Slco1c1 | SLCO1C1 | -7.72E-03 | 3.90E-03 |
| 10986 | Tnfaip8 | TNFAIP8 | 1.70E-02 | -3.53E-02 |
| 10987 | Slc2a4 | SLC2A4 | 1.04E-02 | -2.69E-02 |
| 10988 | Akap5 | AKAP5 | 2.10E-02 | -6.29E-02 |
| 10989 | Ak3 | AK3 | 8.02E-03 | -2.26E-02 |
| 10990 | Dnah12 | DNAH12 | -2.66E-02 | -6.02E-02 |
| 10991 | Cisd3 | CISD3 | -9.69E-03 | 7.27E-03 |
| 10992 | Igfbp2 | IGFBP2 | 1.08E-01 | 2.33E-01 |
| 10993 | Gp1ba | GP1BA | -6.96E-03 | 9.64E-04 |
| 10994 | Zfp330 | ZNF330 | 6.41E-03 | 3.12E-03 |
| 10995 | Slc23a2 | SLC23A2 | -7.79E-03 | -1.84E-02 |
| 10996 | Cidea | CIDEA | -4.41E-02 | 7.24E-02 |
| 10997 | Cwc25 | CWC25 | 8.21E-03 | -1.32E-03 |
| 10998 | Arhgef11 | ARHGEF11 | 6.87E-03 | -2.59E-02 |
| 10999 | Aip | AIP | -7.76E-03 | 1.04E-02 |
| 11000 | Amigo3 | AMIGO3 | -2.28E-02 | 5.39E-02 |
| 11001 | Rhbdd3 | RHBDD3 | -6.89E-03 | -2.23E-02 |
| 11002 | Cep250 | CEP250 | -8.10E-03 | -1.61E-02 |
| 11003 | Cd14 | CD14 | 3.34E-02 | 1.72E-01 |
| 11004 | Gpr180 | GPR180 | 5.97E-03 | -1.28E-02 |
| 11005 | Mto1 | MTO1 | 8.99E-03 | -1.99E-02 |
| 11006 | Ptar1 | PTAR1 | 1.79E-02 | -2.63E-02 |
| 11007 | Rilpl1 | RILPL1 | 1.79E-02 | -7.09E-03 |
| 11008 | Tecpr2 | TECPR2 | 6.66E-03 | 7.46E-03 |
| 11009 | Srpk2 | SRPK2 | 5.87E-03 | -1.01E-02 |
| 11010 | Pcsk2 | PCSK2 | -1.58E-02 | 3.30E-02 |
| 11011 | Trim38 | TRIM38 | 9.45E-03 | 1.55E-02 |
| 11012 | Cacnb3 | CACNB3 | 1.72E-02 | 3.99E-02 |
| 11013 | Gsdmc2 | GSDMC | 1.51E-01 | 1.65E-01 |
| 11014 | Nodal | NODAL | 8.77E-03 | 1.26E-02 |
| 11015 | Fkbp2 | FKBP2 | -7.00E-03 | -1.54E-02 |
| 11016 | Nfxl1 | NFXL1 | -1.18E-02 | 1.99E-02 |
| 11017 | Naa16 | NAA16 | -1.12E-02 | -1.42E-02 |
| 11018 | Dr1 | DR1 | -6.51E-03 | -1.99E-02 |
| 11019 | Syt8 | SYT8 | 8.36E-02 | 1.69E-01 |
| 11020 | Ctse | CTSE | 9.31E-02 | 9.99E-02 |
| 11021 | Map3k10 | MAP3K10 | 5.74E-03 | 7.99E-03 |

| | | | | |
|---|---|---|---|---|
| 11022 | AA986860 | C1orf116 | -2.20E-02 | 2.99E-04 |
| 11023 | Havcr2 | HAVCR2 | 2.24E-02 | 6.98E-02 |
| 11024 | Ube2n | UBE2N | 6.32E-03 | 1.04E-02 |
| 11025 | Fam110b | FAM110B | 2.61E-02 | 4.79E-02 |
| 11026 | Ascc2 | ASCC2 | -4.66E-03 | -6.46E-03 |
| 11027 | Tmem156 | TMEM156 | 1.01E-02 | 6.25E-03 |
| 11028 | Piwil2 | PIWIL2 | 1.42E-02 | -8.67E-03 |
| 11029 | Gatad1 | GATAD1 | -7.53E-03 | 8.72E-03 |
| 11030 | Entpd3 | ENTPD3 | -3.08E-02 | 1.07E-01 |
| 11031 | Lrrtm2 | LRRTM2 | -1.45E-02 | -1.67E-02 |
| 11032 | Pcdhb13 | PCDHB6 | 1.74E-02 | 2.00E-02 |
| 11033 | Kcnrg | KCNRG | 1.20E-02 | 3.33E-02 |
| 11034 | Creld2 | CRELD2 | 2.42E-02 | 1.90E-02 |
| 11035 | Lrrn2 | LRRN2 | -1.73E-02 | 3.68E-02 |
| 11036 | Gjd3 | GJD3 | 8.22E-03 | -1.10E-02 |
| 11037 | Ccm2 | CCM2 | -9.45E-03 | 3.11E-02 |
| 11038 | Nanos1 | NANOS1 | -1.56E-02 | 3.45E-02 |
| 11039 | Crp | CRP | 1.95E-02 | 1.24E-02 |
| 11040 | Rdh19 | RDH16 | -3.96E-02 | -2.76E-01 |
| 11041 | Acvr2a | ACVR2A | 6.74E-03 | -6.88E-03 |
| 11042 | Dram1 | DRAM1 | 1.66E-02 | 6.69E-03 |
| 11043 | Sidt1 | SIDT1 | -4.93E-02 | 8.14E-02 |
| 11044 | Stt3b | STT3B | 5.55E-03 | 2.17E-03 |
| 11045 | Cxxc1 | CXXC1 | 5.37E-03 | 3.59E-03 |
| 11046 | Prkce | PRKCE | 7.86E-03 | -3.68E-02 |
| 11047 | Ccr10 | CCR10 | -1.03E-02 | 9.17E-03 |
| 11048 | Pink1 | PINK1 | 8.00E-03 | -3.27E-02 |
| 11049 | Cct7 | CCT7 | 5.50E-03 | -1.28E-03 |
| 11050 | Grin2b | GRIN2B | -6.85E-03 | 5.33E-04 |
| 11051 | St6gal1 | ST6GAL1 | -8.42E-03 | 1.08E-02 |
| 11052 | Lrrc6 | LRRC6 | 9.70E-03 | 1.16E-02 |
| 11053 | Tox2 | TOX2 | -1.25E-02 | 1.01E-02 |
| 11054 | Stoml3 | STOML3 | 9.45E-03 | 4.72E-03 |
| 11055 | Bpifc | BPIFC | -1.77E-02 | 4.63E-02 |
| 11056 | Sirt4 | SIRT4 | -7.72E-03 | -5.92E-03 |
| 11057 | Slc22a15 | SLC22A15 | 2.62E-02 | 1.79E-02 |
| 11058 | Gstp1 | GSTP1 | 1.02E-02 | 8.63E-03 |
| 11059 | Stk4 | STK4 | -5.27E-03 | 1.66E-03 |
| 11060 | Ctnnd2 | CTNND2 | 1.34E-02 | 3.27E-02 |
| 11061 | Cpt1b | CPT1B | 2.49E-02 | -6.98E-02 |
| 11062 | Inhba | INHBA | 2.73E-02 | 1.77E-01 |
| 11063 | Pcdhgb2 | PCDHGB2 | -2.72E-02 | -1.03E-01 |
| 11064 | Gjb4 | GJB4 | 7.97E-02 | 1.33E-01 |
| 11065 | Epha7 | EPHA7 | -4.59E-02 | 1.77E-02 |
| 11066 | Cbln3 | CBLN3 | 1.55E-02 | -4.26E-02 |
| 11067 | Aldh8a1 | ALDH8A1 | -9.57E-03 | -1.54E-02 |
| 11068 | Zfp382 | ZNF382 | -2.77E-02 | -3.85E-02 |
| 11069 | Ppp1r12a | PPP1R12A | -4.23E-03 | -2.29E-03 |
| 11070 | Smpdl3b | SMPDL3B | 3.96E-02 | 3.33E-01 |
| 11071 | Zfr2 | ZFR2 | 1.38E-02 | -2.35E-02 |
| 11072 | Ttc26 | TTC26 | -9.94E-03 | 2.54E-03 |
| 11073 | Pard6a | PARD6A | 1.09E-02 | -1.98E-02 |
| 11074 | Ccdc28a | CCDC28A | -7.43E-03 | -3.63E-03 |

| | | | | |
|---|---|---|---|---|
| 11075 | Trem1 | TREM1 | 5.70E-02 | 1.64E-01 |
| 11076 | Nfe2 | NFE2 | 1.74E-02 | 2.78E-02 |
| 11077 | Klf5 | KLF5 | 7.52E-02 | 8.69E-02 |
| 11078 | G2e3 | G2E3 | 1.13E-02 | 1.20E-02 |
| 11079 | Elf2 | ELF2 | -5.83E-03 | -7.72E-03 |
| 11080 | Elmo3 | ELMO3 | -4.97E-03 | -1.22E-02 |
| 11081 | Nipa1 | NIPA1 | 1.13E-02 | -7.67E-02 |
| 11082 | Sgms2 | SGMS2 | 9.39E-03 | -1.09E-02 |
| 11083 | Pfkm | PFKM | -5.99E-03 | -2.89E-02 |
| 11084 | Akap8l | AKAP8L | 6.23E-03 | -2.30E-02 |
| 11085 | Cnrip1 | CNRIP1 | -2.27E-02 | -3.07E-02 |
| 11086 | Fa2h | FA2H | 3.86E-02 | -1.10E-01 |
| 11087 | Glod4 | GLOD4 | 5.47E-03 | -1.50E-02 |
| 11088 | Nrtn | NRTN | 2.03E-02 | 5.11E-02 |
| 11089 | Egr1 | EGR1 | 3.80E-02 | -2.37E-02 |
| 11090 | Hyal3 | HYAL3 | -2.29E-02 | 3.30E-02 |
| 11091 | Dusp4 | DUSP4 | 2.20E-02 | 3.48E-02 |
| 11092 | Khdrbs2 | KHDRBS2 | -9.33E-03 | -4.58E-03 |
| 11093 | Mfsd10 | MFSD10 | -9.94E-03 | 2.98E-02 |
| 11094 | Tmem150a | TMEM150A | -1.02E-02 | 3.24E-02 |
| 11095 | S100a3 | S100A3 | -1.00E-02 | 3.96E-02 |
| 11096 | Tbc1d8b | TBC1D8B | 9.39E-03 | -1.00E-02 |
| 11097 | Hoxd3 | HOXD3 | -2.36E-02 | 5.25E-02 |
| 11098 | Rpp25l | RPP25L | -9.37E-03 | -6.43E-04 |
| 11099 | Nefl | NEFL | -2.87E-02 | 1.16E-01 |
| 11100 | Ppp1r3a | PPP1R3A | 5.46E-03 | 1.44E-02 |
| 11101 | Avpr2 | AVPR2 | 2.99E-02 | -3.84E-02 |
| 11102 | Tfpt | TFPT | -6.71E-03 | 7.41E-03 |
| 11103 | Sos1 | SOS1 | -5.56E-03 | -2.07E-03 |
| 11104 | Trappc5 | TRAPPC5 | 6.50E-03 | -3.13E-03 |
| 11105 | Ythdf3 | YTHDF3 | -5.92E-03 | 8.15E-03 |
| 11106 | Trappc8 | TRAPPC8 | -6.28E-03 | -4.37E-03 |
| 11107 | Mettl7a1 | METTL7A | 1.21E-02 | -6.95E-03 |
| 11108 | Psors1c2 | PSORS1C2 | 1.12E-02 | 2.14E-02 |
| 11109 | Zfp706 | ZNF706 | 6.86E-03 | -3.09E-03 |
| 11110 | Actl9 | ACTL9 | -1.43E-02 | -7.99E-04 |
| 11111 | Hmg20a | HMG20A | 3.96E-03 | -1.17E-02 |
| 11112 | Pigk | PIGK | -5.40E-03 | -2.21E-02 |
| 11113 | Cep85l | CEP85L | -1.51E-02 | -1.87E-03 |
| 11114 | Clcn3 | CLCN3 | 6.17E-03 | -3.27E-03 |
| 11115 | Reep4 | REEP4 | 1.45E-02 | 4.86E-02 |
| 11116 | Bcas2 | BCAS2 | -4.77E-03 | -1.41E-03 |
| 11117 | Pdss2 | PDSS2 | -8.25E-03 | -1.82E-02 |
| 11118 | Lig1 | LIG1 | 2.21E-02 | 1.20E-01 |
| 11119 | Abi3 | ABI3 | -1.27E-02 | -1.31E-02 |
| 11120 | Kif21b | KIF21B | -2.32E-02 | 6.29E-02 |
| 11121 | Lrrc46 | LRRC46 | -1.75E-02 | 1.91E-02 |
| 11122 | Leap2 | LEAP2 | 1.54E-02 | -1.41E-02 |
| 11123 | Aldh5a1 | ALDH5A1 | 9.20E-03 | -3.00E-02 |
| 11124 | Iffo2 | IFFO2 | 2.39E-02 | -6.08E-03 |
| 11125 | Grap | GRAP | -1.81E-02 | -2.54E-02 |
| 11126 | Ccdc159 | CCDC159 | -1.91E-02 | -2.00E-02 |
| 11127 | Hhex | HHEX | -1.56E-02 | 1.17E-02 |

| | | | | |
|---|---|---|---|---|
| 11128 | Snip1 | SNIP1 | -1.04E-02 | -3.24E-04 |
| 11129 | Macc1 | MACC1 | 6.38E-02 | -8.43E-04 |
| 11130 | S100b | S100B | 2.02E-02 | -1.85E-02 |
| 11131 | Myo5b | MYO5B | 6.27E-03 | -3.03E-02 |
| 11132 | Kbtbd4 | KBTBD4 | 1.16E-02 | -1.48E-02 |
| 11133 | Neu2 | NEU2 | 1.15E-02 | -6.43E-02 |
| 11134 | Sema3b | SEMA3B | -6.97E-03 | -1.19E-02 |
| 11135 | Zdhhc17 | ZDHHC17 | 6.16E-03 | -1.87E-02 |
| 11136 | Ammecr1l | AMMECR1L | -6.32E-03 | -2.50E-03 |
| 11137 | Fundc2 | FUNDC2 | 7.72E-03 | -3.57E-03 |
| 11138 | Otos | OTOS | -7.96E-03 | -9.25E-03 |
| 11139 | Lrriq4 | LRRIQ4 | 1.51E-02 | -2.87E-02 |
| 11140 | Mrpl15 | MRPL15 | -5.85E-03 | -7.95E-03 |
| 11141 | Cdhr1 | CDHR1 | 1.35E-02 | 1.34E-02 |
| 11142 | Dcaf11 | DCAF11 | 4.21E-03 | -2.44E-02 |
| 11143 | Tbx1 | TBX1 | -1.19E-02 | -1.28E-02 |
| 11144 | Hk2 | HK2 | 3.70E-02 | 3.08E-01 |
| 11145 | Serpinb12 | SERPINB12 | 9.47E-03 | 2.58E-02 |
| 11146 | Rnf5 | RNF5 | 5.19E-03 | -1.26E-02 |
| 11147 | Gabrb3 | GABRB3 | -3.30E-02 | -1.35E-01 |
| 11148 | Ccl17 | CCL17 | 1.84E-02 | 7.42E-02 |
| 11149 | Eif3j2 | EIF3J | 2.06E-02 | 7.41E-04 |
| 11150 | Slc25a29 | SLC25A29 | -1.66E-02 | -4.90E-02 |
| 11151 | Slc25a28 | SLC25A28 | -7.01E-03 | 1.24E-03 |
| 11152 | A3galt2 | A3GALT2 | -1.67E-02 | -1.07E-02 |
| 11153 | Tada3 | TADA3 | -6.92E-03 | -1.09E-02 |
| 11154 | Sycp3 | SYCP3 | -1.09E-02 | -1.81E-02 |
| 11155 | Gm14137 | C15orf62 | 2.62E-02 | 4.23E-02 |
| 11156 | Mrpl47 | MRPL47 | 7.90E-03 | -7.61E-03 |
| 11157 | Ncapg | NCAPG | 4.77E-02 | 2.29E-01 |
| 11158 | Polr3f | POLR3F | -5.97E-03 | -5.93E-03 |
| 11159 | Bicc1 | BICC1 | 7.40E-03 | -2.09E-02 |
| 11160 | Sptb | SPTB | -1.99E-02 | -3.23E-02 |
| 11161 | Nt5c1a | NT5C1A | -1.87E-02 | 8.48E-02 |
| 11162 | Slc12a3 | SLC12A3 | -1.11E-02 | 4.57E-03 |
| 11163 | Fzd4 | FZD4 | 6.38E-03 | -1.78E-02 |
| 11164 | Iba57 | IBA57 | 1.01E-02 | -3.78E-03 |
| 11165 | Fra10ac1 | FRA10AC1 | -6.78E-03 | 1.80E-03 |
| 11166 | Efcab7 | EFCAB7 | -1.38E-02 | 5.61E-03 |
| 11167 | Amer1 | AMER1 | 1.24E-02 | -1.00E-02 |
| 11168 | Hoxd1 | HOXD1 | 1.58E-02 | -2.60E-02 |
| 11169 | Elovl5 | ELOVL5 | -5.24E-03 | 1.94E-04 |
| 11170 | Triqk | TRIQK | 1.20E-02 | 3.16E-03 |
| 11171 | Mcur1 | MCUR1 | 8.22E-03 | -1.53E-02 |
| 11172 | Lman2 | LMAN2 | -5.38E-03 | -1.59E-02 |
| 11173 | Pcyt2 | PCYT2 | -6.91E-03 | -6.82E-03 |
| 11174 | Mxra8 | MXRA8 | -1.61E-02 | 5.21E-02 |
| 11175 | Amhr2 | AMHR2 | -1.30E-02 | -1.75E-02 |
| 11176 | Nphs1 | NPHS1 | -2.20E-02 | -4.88E-02 |
| 11177 | Arhgap17 | ARHGAP17 | -4.44E-03 | 1.26E-02 |
| 11178 | Coa7 | COA7 | 7.20E-03 | 1.11E-03 |
| 11179 | Zfp689 | ZNF689 | -1.19E-02 | -2.36E-02 |
| 11180 | Lrriq3 | LRRIQ3 | -1.39E-02 | -5.77E-02 |

| | | | | |
|---|---|---|---|---|
| 11181 | Raly | RALY | -5.56E-03 | -5.76E-03 |
| 11182 | Grp | GRP | 6.98E-03 | 1.35E-02 |
| 11183 | Ucp1 | UCP1 | -5.43E-02 | 8.38E-02 |
| 11184 | Lrr1 | LRR1 | 1.80E-02 | 8.65E-02 |
| 11185 | Srm | SRM | 8.71E-03 | 2.55E-02 |
| 11186 | Cdh17 | CDH17 | 1.37E-02 | 6.24E-02 |
| 11187 | Mbd5 | MBD5 | -6.90E-03 | -1.99E-02 |
| 11188 | Gpr183 | GPR183 | -2.53E-02 | 5.10E-02 |
| 11189 | Arid1b | ARID1B | 6.22E-03 | -1.22E-02 |
| 11190 | Slc30a3 | SLC30A3 | 1.11E-02 | 1.32E-02 |
| 11191 | Negr1 | NEGR1 | 3.47E-02 | -6.27E-02 |
| 11192 | Jade1 | JADE1 | -6.38E-03 | -3.98E-02 |
| 11193 | Wipf3 | WIPF3 | 2.09E-02 | -4.05E-02 |
| 11194 | Traf7 | TRAF7 | 4.41E-03 | 7.79E-03 |
| 11195 | Eogt | EOGT | -8.31E-03 | -1.22E-02 |
| 11196 | Ap2m1 | AP2M1 | 6.27E-03 | -1.07E-03 |
| 11197 | Cenpo | CENPO | 1.19E-02 | 1.35E-02 |
| 11198 | Llgl1 | LLGL1 | 7.07E-03 | 2.08E-02 |
| 11199 | Myl7 | MYL7 | -2.03E-02 | -6.25E-02 |
| 11200 | Rps19bp1 | RPS19BP1 | 8.08E-03 | 9.72E-03 |
| 11201 | Pmpcb | PMPCB | 7.66E-03 | -2.74E-02 |
| 11202 | Tead2 | TEAD2 | 1.96E-02 | 4.50E-02 |
| 11203 | Arap1 | ARAP1 | -6.43E-03 | -8.51E-03 |
| 11204 | Rab11fip1 | RAB11FIP1 | -1.75E-02 | -2.25E-02 |
| 11205 | Phf23 | PHF23 | -7.92E-03 | 1.75E-02 |
| 11206 | Scx | SCXB | 5.46E-02 | 6.86E-02 |
| 11207 | Srcap | SRCAP | -1.52E-02 | -3.20E-02 |
| 11208 | Jagn1 | JAGN1 | 6.36E-03 | 1.94E-02 |
| 11209 | Npy | NPY | 6.90E-02 | -5.00E-02 |
| 11210 | Rmdn2 | RMDN2 | -1.37E-02 | 1.73E-02 |
| 11211 | Ppy | PPY | -5.77E-03 | -7.10E-03 |
| 11212 | Rpap3 | RPAP3 | 5.86E-03 | 4.34E-03 |
| 11213 | Ercc8 | ERCC8 | 1.47E-02 | 1.45E-02 |
| 11214 | Rpl9 | RPL9 | -4.86E-03 | 9.02E-03 |
| 11215 | Fmr1 | FMR1 | 6.69E-03 | -6.11E-03 |
| 11216 | Garnl3 | GARNL3 | 3.38E-02 | 4.77E-02 |
| 11217 | Luc7l | LUC7L | -6.07E-03 | 2.11E-02 |
| 11218 | Dynlt1b | DYNLT1 | 2.11E-02 | 1.74E-02 |
| 11219 | Ppp1r15b | PPP1R15B | -5.45E-03 | 1.08E-02 |
| 11220 | Emb | EMB | 7.71E-03 | -3.32E-03 |
| 11221 | Mylk4 | MYLK4 | 1.33E-02 | 1.17E-02 |
| 11222 | Sun1 | SUN1 | 3.28E-03 | -1.58E-02 |
| 11223 | Matr3 | MATR3 | 4.46E-03 | -2.56E-03 |
| 11224 | B3gat3 | B3GAT3 | -5.95E-03 | -8.98E-03 |
| 11225 | Wdr33 | WDR33 | -5.20E-03 | -4.83E-03 |
| 11226 | Tcf12 | TCF12 | 5.21E-03 | -5.21E-03 |
| 11227 | Nprl2 | NPRL2 | 5.25E-03 | -2.88E-02 |
| 11228 | Tln1 | TLN1 | -7.33E-03 | -1.40E-02 |
| 11229 | Hspa1a | HSPA1A | -4.80E-02 | 1.10E-02 |
| 11230 | Hspa1a | HSPA1B | -4.80E-02 | 1.10E-02 |
| 11231 | 4921536K21Rik | C5orf52 | 1.74E-02 | -2.37E-03 |
| 11232 | Dedd | DEDD | -4.82E-03 | 2.99E-03 |
| 11233 | Tmem106a | TMEM106A | -1.10E-02 | -2.04E-02 |

| | | | | |
|---|---|---|---|---|
| 11234 | Fbxl5 | FBXL5 | -5.96E-03 | 1.29E-03 |
| 11235 | Celf3 | CELF3 | 1.05E-01 | 1.59E-01 |
| 11236 | Ccser1 | CCSER1 | 2.09E-02 | -5.20E-02 |
| 11237 | Nf2 | NF2 | 4.37E-03 | -1.52E-02 |
| 11238 | Pdcd2 | PDCD2 | 6.94E-03 | 7.92E-03 |
| 11239 | Jkamp | JKAMP | 3.87E-03 | -7.58E-03 |
| 11240 | Cpsf4 | CPSF4 | 6.59E-03 | 1.20E-02 |
| 11241 | Lamb2 | LAMB2 | -7.45E-03 | 7.67E-03 |
| 11242 | Tbl1xr1 | TBL1XR1 | 5.46E-03 | 8.58E-04 |
| 11243 | Qsox2 | QSOX2 | 4.74E-03 | -8.19E-03 |
| 11244 | Slc5a9 | SLC5A9 | -1.11E-02 | -5.01E-02 |
| 11245 | Hba-a2 | HBA2 | -4.14E-02 | 5.22E-02 |
| 11246 | Hba-a2 | HBA1 | -4.14E-02 | 5.22E-02 |
| 11247 | Pgm5 | PGM5 | 2.47E-02 | 1.80E-02 |
| 11248 | Gemin6 | GEMIN6 | 1.18E-02 | 4.00E-02 |
| 11249 | Igsf23 | IGSF23 | -1.12E-02 | 6.24E-03 |
| 11250 | Tmem125 | TMEM125 | 1.06E-02 | -3.22E-02 |
| 11251 | Rgs20 | RGS20 | 5.38E-03 | 9.99E-03 |
| 11252 | Paqr7 | PAQR7 | 1.87E-02 | 8.50E-03 |
| 11253 | Atmin | ATMIN | -5.76E-03 | -2.41E-02 |
| 11254 | Impg2 | IMPG2 | -1.97E-02 | 3.00E-02 |
| 11255 | Xylb | XYLB | 9.99E-03 | -1.35E-02 |
| 11256 | Atxn1l | ATXN1L | -6.42E-03 | -2.35E-02 |
| 11257 | Ptprf | PTPRF | -6.75E-03 | -1.13E-02 |
| 11258 | Plekha4 | PLEKHA4 | -1.78E-02 | -3.93E-02 |
| 11259 | Plvap | PLVAP | 7.86E-03 | -2.51E-02 |
| 11260 | Trim25 | TRIM25 | -9.77E-03 | -9.29E-03 |
| 11261 | Tubb5 | TUBB | 1.33E-02 | 7.79E-02 |
| 11262 | Src | SRC | 5.38E-03 | 6.74E-03 |
| 11263 | Pcdhgb1 | PCDHGB1 | -1.63E-02 | -3.27E-02 |
| 11264 | Pax2 | PAX2 | 2.33E-02 | -6.51E-03 |
| 11265 | Acat1 | ACAT1 | 6.83E-03 | -1.10E-02 |
| 11266 | Mdfic | MDFIC | -1.49E-02 | 1.88E-03 |
| 11267 | Kdr | KDR | -1.97E-02 | -3.55E-02 |
| 11268 | Syt11 | SYT11 | 1.20E-02 | -2.36E-02 |
| 11269 | Kcnq4 | KCNQ4 | 1.54E-02 | 2.17E-02 |
| 11270 | Tmem120a | TMEM120A | -7.74E-03 | 3.16E-03 |
| 11271 | Cryba2 | CRYBA2 | 7.86E-03 | 9.94E-03 |
| 11272 | Tbx3 | TBX3 | 4.94E-02 | 2.70E-02 |
| 11273 | Ipo8 | IPO8 | -4.82E-03 | -1.35E-02 |
| 11274 | Coq2 | COQ2 | 5.53E-03 | -2.06E-02 |
| 11275 | Tm9sf1 | TM9SF1 | -5.37E-03 | -1.49E-02 |
| 11276 | Neu3 | NEU3 | 1.73E-02 | 3.45E-03 |
| 11277 | Angpt2 | ANGPT2 | -5.36E-02 | -3.95E-02 |
| 11278 | Fhl2 | FHL2 | -2.09E-02 | 3.99E-02 |
| 11279 | Gyg | GYG1 | 6.37E-03 | 1.44E-02 |
| 11280 | Ski | SKI | -4.55E-03 | -2.02E-03 |
| 11281 | Zfp354a | ZNF354A | -8.28E-03 | -1.08E-02 |
| 11282 | Dph1 | DPH1 | 1.08E-02 | 1.34E-03 |
| 11283 | Laptm4a | LAPTM4A | 3.73E-03 | 2.67E-03 |
| 11284 | Rab3ip | RAB3IP | -4.92E-03 | -8.13E-03 |
| 11285 | Cct5 | CCT5 | 5.16E-03 | -1.61E-03 |
| 11286 | 9530068E07Rik | C5orf15 | -4.52E-03 | -1.87E-03 |

| | | | | |
|---|---|---|---|---|
| 11287 | Lrrfip1 | LRRFIP1 | 8.90E-03 | -1.13E-03 |
| 11288 | Fam178b | FAM178B | 8.90E-03 | 9.11E-04 |
| 11289 | Galt | GALT | -4.95E-03 | -1.52E-02 |
| 11290 | Adamts17 | ADAMTS17 | -1.12E-02 | -5.56E-04 |
| 11291 | Pde4a | PDE4A | -2.49E-02 | 5.66E-02 |
| 11292 | 1110059E24Rik | C9orf85 | -7.83E-03 | 6.14E-03 |
| 11293 | Ehmt1 | EHMT1 | -3.52E-03 | -3.81E-04 |
| 11294 | Fxyd5 | FXYD5 | 1.46E-02 | 6.48E-02 |
| 11295 | Znhit6 | ZNHIT6 | 7.32E-03 | 1.25E-02 |
| 11296 | Fancf | FANCF | 1.68E-02 | 3.17E-02 |
| 11297 | Spaca1 | SPACA1 | 1.11E-02 | -6.52E-03 |
| 11298 | Klhl35 | KLHL35 | 9.50E-03 | 4.26E-03 |
| 11299 | Coro2b | CORO2B | 1.26E-02 | -2.75E-02 |
| 11300 | Pnck | PNCK | -1.71E-02 | -7.14E-03 |
| 11301 | Egfr | EGFR | -7.46E-03 | 9.97E-03 |
| 11302 | Cpne9 | CPNE9 | 1.19E-02 | -4.11E-03 |
| 11303 | Sesn2 | SESN2 | 1.79E-02 | 1.60E-02 |
| 11304 | Ikbke | IKBKE | -1.47E-02 | 5.63E-02 |
| 11305 | Fgfr3 | FGFR3 | -8.80E-03 | -3.58E-02 |
| 11306 | Bahcc1 | BAHCC1 | -1.20E-02 | -2.24E-02 |
| 11307 | Rnf113a2 | RNF113A | 7.92E-03 | 1.46E-02 |
| 11308 | Ighmbp2 | IGHMBP2 | 1.21E-02 | -2.69E-03 |
| 11309 | Tcea3 | TCEA3 | 1.15E-02 | -5.24E-02 |
| 11310 | Ikbip | IKBIP | 8.20E-03 | -2.60E-02 |
| 11311 | Gp9 | GP9 | 1.23E-02 | 2.80E-04 |
| 11312 | Fhdc1 | FHDC1 | 1.90E-02 | -8.40E-03 |
| 11313 | Abhd17b | ABHD17B | -7.14E-03 | 4.69E-03 |
| 11314 | Elp2 | ELP2 | -4.86E-03 | -1.35E-02 |
| 11315 | Bcl2l15 | BCL2L15 | 1.60E-02 | -6.01E-03 |
| 11316 | Mrpl18 | MRPL18 | -5.12E-03 | 4.21E-03 |
| 11317 | Spata25 | SPATA25 | -6.31E-03 | 1.09E-02 |
| 11318 | Nelfb | NELFB | 3.85E-03 | 8.50E-05 |
| 11319 | Slc7a11 | SLC7A11 | 3.92E-02 | 1.18E-01 |
| 11320 | Moap1 | MOAP1 | -6.20E-03 | -7.48E-04 |
| 11321 | Prkg1 | PRKG1 | -3.03E-02 | 2.38E-02 |
| 11322 | Ly75 | LY75 | -2.01E-02 | 2.77E-02 |
| 11323 | Rtn2 | RTN2 | -2.00E-02 | 7.02E-02 |
| 11324 | Cops7b | COPS7B | 9.05E-03 | -3.39E-03 |
| 11325 | Slc35c2 | SLC35C2 | 6.50E-03 | -1.68E-02 |
| 11326 | Nav3 | NAV3 | 4.66E-02 | -4.25E-02 |
| 11327 | Vsig10 | VSIG10 | 8.42E-03 | -1.95E-02 |
| 11328 | Rgl3 | RGL3 | 9.86E-03 | -1.24E-02 |
| 11329 | Ccl11 | CCL11 | -1.52E-02 | 3.83E-03 |
| 11330 | Mpzl3 | MPZL3 | 1.21E-02 | 1.63E-02 |
| 11331 | Vsig4 | VSIG4 | 9.44E-03 | 2.75E-02 |
| 11332 | Usp38 | USP38 | 5.74E-03 | 1.76E-03 |
| 11333 | Dusp13 | DUSP13 | 9.85E-03 | -1.69E-02 |
| 11334 | Tor1aip2 | TOR1AIP2 | 4.05E-03 | -7.58E-03 |
| 11335 | Ssrp1 | SSRP1 | 3.31E-03 | -2.23E-03 |
| 11336 | Zbtb46 | ZBTB46 | -1.19E-02 | -1.34E-02 |
| 11337 | Nhlrc3 | NHLRC3 | 7.63E-03 | -1.01E-02 |
| 11338 | Zfp652 | ZNF652 | -1.04E-02 | -4.95E-02 |
| 11339 | Galntl5 | GALNTL5 | 7.49E-03 | 5.43E-03 |

| | | | | |
|---|---|---|---|---|
| 11340 | Suv39h2 | SUV39H2 | -1.88E-02 | 1.02E-01 |
| 11341 | Pik3c2b | PIK3C2B | -2.24E-02 | 4.20E-02 |
| 11342 | Chrnb1 | CHRNB1 | 2.45E-02 | 6.54E-02 |
| 11343 | Ier5 | IER5 | 1.67E-02 | 9.55E-02 |
| 11344 | Rnpep | RNPEP | -6.76E-03 | -8.33E-03 |
| 11345 | Rasgrf2 | RASGRF2 | -2.37E-02 | 2.11E-02 |
| 11346 | Rrad | RRAD | -3.10E-02 | 1.07E-01 |
| 11347 | Kdm1a | KDM1A | 4.11E-03 | 1.07E-02 |
| 11348 | Zbtb20 | ZBTB20 | 5.68E-03 | -1.04E-03 |
| 11349 | Rapgef2 | RAPGEF2 | -8.49E-03 | -1.75E-02 |
| 11350 | Cyp4f39 | CYP4F22 | 1.76E-02 | -3.57E-02 |
| 11351 | Pdpk1 | PDPK1 | 5.33E-03 | -1.56E-02 |
| 11352 | Acer3 | ACER3 | -5.15E-03 | 8.16E-03 |
| 11353 | Ccs | CCS | -7.34E-03 | -2.06E-02 |
| 11354 | Kcne2 | KCNE2 | 1.23E-02 | -1.71E-02 |
| 11355 | Doc2a | DOC2A | 1.95E-02 | 4.88E-02 |
| 11356 | Ndst4 | NDST4 | -1.41E-02 | 1.71E-02 |
| 11357 | Gusb | GUSB | -7.82E-03 | 3.80E-02 |
| 11358 | Pxdn | PXDN | 1.79E-02 | 2.69E-02 |
| 11359 | 4930430F08Rik | C12orf29 | -1.10E-02 | 1.55E-02 |
| 11360 | Fzd9 | FZD9 | 3.42E-02 | -3.67E-02 |
| 11361 | Zfp827 | ZNF827 | -6.45E-03 | 1.28E-03 |
| 11362 | Fos | FOS | 4.68E-02 | 1.67E-02 |
| 11363 | Tcerg1l | TCERG1L | -2.98E-02 | 3.09E-02 |
| 11364 | Hspa2 | HSPA2 | -1.30E-02 | -7.50E-03 |
| 11365 | Ffar2 | FFAR2 | 1.85E-02 | 1.01E-02 |
| 11366 | Spag4 | SPAG4 | 2.18E-02 | 1.65E-02 |
| 11367 | Fbxl15 | FBXL15 | 7.89E-03 | 9.48E-03 |
| 11368 | Fosb | FOSB | 7.40E-02 | -7.35E-02 |
| 11369 | Asns | ASNS | -2.43E-02 | 1.47E-01 |
| 11370 | Mknk2 | MKNK2 | 9.56E-03 | -1.91E-02 |
| 11371 | Crbn | CRBN | -7.63E-03 | -1.96E-02 |
| 11372 | Slc18a1 | SLC18A1 | 1.10E-02 | -3.41E-02 |
| 11373 | Ccdc169 | CCDC169 | -1.55E-02 | -9.06E-02 |
| 11374 | Gdap1l1 | GDAP1L1 | -1.47E-02 | 1.45E-02 |
| 11375 | Them5 | THEM5 | 8.55E-03 | 1.74E-02 |
| 11376 | Slc50a1 | SLC50A1 | 7.97E-03 | -2.09E-03 |
| 11377 | Dock6 | DOCK6 | -8.32E-03 | -2.28E-02 |
| 11378 | Ccdc77 | CCDC77 | -1.09E-02 | 2.57E-02 |
| 11379 | Bloc1s5 | BLOC1S5 | 5.43E-03 | -1.71E-02 |
| 11380 | Dnajb12 | DNAJB12 | 6.39E-03 | -1.90E-02 |
| 11381 | Azgp1 | AZGP1 | 2.68E-02 | 2.68E-02 |
| 11382 | Scube3 | SCUBE3 | -1.18E-02 | -4.46E-02 |
| 11383 | Timmdc1 | TIMMDC1 | -5.87E-03 | 6.46E-04 |
| 11384 | Ncapd2 | NCAPD2 | 1.91E-02 | 7.02E-02 |
| 11385 | Kcne3 | KCNE3 | -1.78E-02 | 3.86E-02 |
| 11386 | Nit1 | NIT1 | 6.90E-03 | -1.03E-02 |
| 11387 | Gabra5 | GABRA5 | 4.55E-03 | -1.36E-02 |
| 11388 | Cmklr1 | CMKLR1 | -1.66E-02 | 3.46E-02 |
| 11389 | Slc7a10 | SLC7A10 | 2.60E-02 | 2.98E-02 |
| 11390 | Il17rc | IL17RC | 1.07E-02 | -4.09E-03 |
| 11391 | Nlrp10 | NLRP10 | -1.39E-02 | 7.70E-02 |
| 11392 | Slco2b1 | SLCO2B1 | -1.74E-02 | -4.26E-02 |

| | | | | |
|---|---|---|---|---|
| 11393 | Pgf | PGF | 2.09E-02 | 6.02E-02 |
| 11394 | Gne | GNE | 4.65E-03 | -1.97E-02 |
| 11395 | Slc52a2 | SLC52A2 | -8.07E-03 | 4.76E-03 |
| 11396 | Med13l | MED13L | 6.21E-03 | -5.94E-03 |
| 11397 | Pvalb | PVALB | -1.77E-02 | -6.82E-03 |
| 11398 | Gm13305 | IL11RA | -1.16E-02 | 1.20E-02 |
| 11399 | Gm13306 | CCL27 | -8.40E-03 | 1.44E-02 |
| 11400 | Dmrta2 | DMRTA2 | 1.15E-02 | 6.73E-04 |
| 11401 | Ift20 | IFT20 | 5.34E-03 | -8.08E-03 |
| 11402 | Psma5 | PSMA5 | -5.16E-03 | 1.19E-02 |
| 11403 | Gja5 | GJA5 | -1.99E-02 | -1.39E-02 |
| 11404 | Slc22a5 | SLC22A5 | -1.12E-02 | -2.22E-02 |
| 11405 | Ankra2 | ANKRA2 | -4.98E-03 | 1.85E-02 |
| 11406 | C1qtnf1 | C1QTNF1 | -2.00E-02 | -6.05E-03 |
| 11407 | Dram2 | DRAM2 | 4.73E-03 | -1.15E-02 |
| 11408 | Cox14 | COX14 | 6.32E-03 | -3.98E-04 |
| 11409 | Stag1 | STAG1 | -6.28E-03 | -1.37E-02 |
| 11410 | Rab40c | RAB40C | -7.23E-03 | 2.25E-03 |
| 11411 | Scamp2 | SCAMP2 | -4.08E-03 | -2.47E-02 |
| 11412 | Dennd2a | DENND2A | 2.46E-02 | 4.79E-02 |
| 11413 | Edc4 | EDC4 | -3.90E-03 | -1.86E-03 |
| 11414 | Gpnmb | GPNMB | 3.38E-02 | 2.67E-01 |
| 11415 | Dbndd2 | DBNDD2 | 2.33E-02 | -2.03E-02 |
| 11416 | Celsr3 | CELSR3 | 6.36E-03 | 1.43E-02 |
| 11417 | Pwwp2b | PWWP2B | -2.00E-02 | 3.54E-02 |
| 11418 | Cep41 | CEP41 | -1.15E-02 | 3.68E-02 |
| 11419 | Vpreb1 | VPREB1 | -3.98E-02 | -2.36E-02 |
| 11420 | Zfyve27 | ZFYVE27 | 4.38E-03 | -1.20E-04 |
| 11421 | Polr2c | POLR2C | 4.65E-03 | -5.16E-03 |
| 11422 | Epas1 | EPAS1 | -9.90E-03 | -2.63E-02 |
| 11423 | Rabl6 | RABL6 | -6.65E-03 | -8.00E-03 |
| 11424 | Dyrk2 | DYRK2 | 6.17E-03 | -2.65E-02 |
| 11425 | Cdca7 | CDCA7 | -2.88E-02 | 7.06E-02 |
| 11426 | Serpina1b | SERPINA1 | 4.43E-02 | 1.63E-01 |
| 11427 | Prrg3 | PRRG3 | -4.50E-02 | -2.30E-02 |
| 11428 | Fam199x | FAM199X | 1.04E-02 | -1.88E-02 |
| 11429 | Adcy5 | ADCY5 | 1.32E-02 | -5.00E-03 |
| 11430 | Rps28 | RPS28 | -5.19E-03 | 1.23E-02 |
| 11431 | Tcaim | TCAIM | 7.56E-03 | -4.42E-02 |
| 11432 | Camkk1 | CAMKK1 | 2.02E-02 | 4.73E-02 |
| 11433 | Oser1 | OSER1 | 7.22E-03 | -1.26E-03 |
| 11434 | Spcs3 | SPCS3 | 5.84E-03 | 1.20E-03 |
| 11435 | Papola | PAPOLA | -4.50E-03 | 3.91E-03 |
| 11436 | Ffar4 | FFAR4 | -2.14E-02 | 7.52E-02 |
| 11437 | Cpsf2 | CPSF2 | 4.89E-03 | 9.40E-03 |
| 11438 | Ddost | DDOST | -4.47E-03 | 1.31E-04 |
| 11439 | Rab27b | RAB27B | -2.42E-02 | 8.26E-02 |
| 11440 | Tsc22d2 | TSC22D2 | -9.00E-03 | 1.75E-02 |
| 11441 | Ldlrad3 | LDLRAD3 | 8.55E-03 | 7.51E-04 |
| 11442 | Morn1 | MORN1 | 1.03E-02 | 1.63E-02 |
| 11443 | Klk1b1 | KLK3 | 1.01E-02 | 3.11E-02 |
| 11444 | Kcne4 | KCNE4 | 3.58E-02 | -1.26E-02 |
| 11445 | Slx4ip | SLX4IP | 9.68E-03 | 4.21E-03 |

| | | | | |
|---|---|---|---|---|
| 11446 | Dxo | DXO | -5.99E-03 | -3.98E-03 |
| 11447 | Cdh24 | CDH24 | 2.62E-02 | 5.20E-02 |
| 11448 | Nudt1 | NUDT1 | -9.37E-03 | -1.42E-02 |
| 11449 | Cplx2 | CPLX2 | -1.28E-02 | -4.28E-03 |
| 11450 | Rbm28 | RBM28 | 5.06E-03 | -1.09E-03 |
| 11451 | Afp | AFP | -2.19E-02 | -2.97E-02 |
| 11452 | Hap1 | HAP1 | 2.04E-02 | 1.16E-02 |
| 11453 | Elavl3 | ELAVL3 | -4.49E-02 | -1.41E-01 |
| 11454 | Cacnb2 | CACNB2 | -2.40E-02 | -3.09E-02 |
| 11455 | Zfp513 | ZNF513 | 5.64E-03 | -2.56E-03 |
| 11456 | Adra2a | ADRA2A | 2.64E-02 | 8.25E-04 |
| 11457 | Itih3 | ITIH3 | 1.76E-02 | 1.45E-01 |
| 11458 | Ptcd3 | PTCD3 | -4.46E-03 | -1.63E-02 |
| 11459 | Diras1 | DIRAS1 | 7.33E-03 | 1.14E-02 |
| 11460 | Cisd2 | CISD2 | 3.85E-03 | -4.80E-03 |
| 11461 | Dapl1 | DAPL1 | 1.49E-02 | 1.02E-02 |
| 11462 | Akr1d1 | AKR1D1 | 1.29E-02 | -6.80E-02 |
| 11463 | Dusp2 | DUSP2 | 2.66E-02 | 4.88E-02 |
| 11464 | Gtdc1 | GTDC1 | 7.69E-03 | -2.00E-02 |
| 11465 | Alox12 | ALOX12 | -1.28E-02 | -5.58E-04 |
| 11466 | Ndufaf1 | NDUFAF1 | -7.69E-03 | -7.61E-03 |
| 11467 | Setx | SETX | 5.49E-03 | -1.05E-02 |
| 11468 | Tex19.2 | TEX19 | 6.68E-03 | -1.89E-02 |
| 11469 | Rgs17 | RGS17 | -6.01E-03 | 1.73E-02 |
| 11470 | Gtf3c1 | GTF3C1 | -5.19E-03 | -1.14E-02 |
| 11471 | Ccl25 | CCL25 | 1.04E-02 | -1.38E-02 |
| 11472 | Mlxipl | MLXIPL | 9.22E-03 | 1.60E-02 |
| 11473 | Zfp362 | ZNF362 | 6.55E-03 | -2.22E-02 |
| 11474 | Depdc5 | DEPDC5 | -5.32E-03 | -1.86E-02 |
| 11475 | Cpe | CPE | 2.08E-02 | 1.17E-01 |
| 11476 | Abcc9 | ABCC9 | -1.75E-02 | -2.99E-02 |
| 11477 | Prss41 | PRSS41 | 1.82E-02 | 6.13E-03 |
| 11478 | Krt23 | KRT23 | -6.28E-02 | 1.32E-01 |
| 11479 | Wnt7a | WNT7A | -7.44E-03 | 3.57E-02 |
| 11480 | Thbs2 | THBS2 | 1.57E-02 | 1.46E-01 |
| 11481 | Lonrf2 | LONRF2 | -1.53E-02 | -3.48E-02 |
| 11482 | Vat1l | VAT1L | -1.24E-02 | 2.82E-02 |
| 11483 | Gabra4 | GABRA4 | 2.36E-02 | -5.74E-02 |
| 11484 | Ccdc80 | CCDC80 | -3.36E-02 | 1.42E-01 |
| 11485 | Lrtm1 | LRTM1 | 9.48E-03 | 2.03E-02 |
| 11486 | Smoc2 | SMOC2 | 4.37E-02 | -3.36E-02 |
| 11487 | Ccar2 | CCAR2 | 4.15E-03 | -2.01E-02 |
| 11488 | Brsk2 | BRSK2 | -1.55E-02 | -4.08E-02 |
| 11489 | Pml | PML | -5.77E-03 | -1.03E-02 |
| 11490 | Eef1a1 | EEF1A1 | -6.04E-03 | 1.39E-02 |
| 11491 | Fam168b | FAM168B | -3.80E-03 | -7.32E-03 |
| 11492 | Itgb5 | ITGB5 | 3.90E-03 | -1.20E-02 |
| 11493 | Gprc5c | GPRC5C | 4.73E-03 | -1.09E-02 |
| 11494 | Tcp11l2 | TCP11L2 | 1.01E-02 | -2.94E-02 |
| 11495 | Hepacam | HEPACAM | 9.13E-03 | 1.32E-03 |
| 11496 | Csf2 | CSF2 | 4.92E-03 | 3.80E-03 |
| 11497 | Wdsub1 | WDSUB1 | 6.19E-03 | -2.53E-02 |
| 11498 | Mrpl23 | LOC102724828 | 5.68E-03 | -1.74E-02 |

| | | | | |
|---|---|---|---|---|
| 11499 | Mrpl23 | MRPL23 | 5.68E-03 | -1.74E-02 |
| 11500 | Ryr2 | RYR2 | 2.33E-02 | -1.01E-01 |
| 11501 | Fmo1 | FMO1 | -9.09E-03 | -4.91E-03 |
| 11502 | Coq6 | COQ6 | -5.35E-03 | -3.07E-02 |
| 11503 | AU040320 | KIAA0319L | -5.18E-03 | -1.36E-02 |
| 11504 | Itgb1bp2 | ITGB1BP2 | 1.64E-02 | 1.82E-02 |
| 11505 | Txk | TXK | -2.44E-02 | 2.43E-02 |
| 11506 | Slc5a11 | SLC5A11 | 1.01E-02 | -2.41E-02 |
| 11507 | Rasgef1b | RASGEF1B | 3.55E-02 | -8.90E-02 |
| 11508 | Plekha2 | PLEKHA2 | 1.31E-02 | 4.22E-03 |
| 11509 | Ccdc18 | CCDC18 | 2.37E-02 | 7.31E-02 |
| 11510 | Lurap1l | LURAP1L | 7.79E-03 | -7.16E-03 |
| 11511 | Tns3 | TNS3 | -7.30E-03 | -1.53E-02 |
| 11512 | Arpc1a | ARPC1A | 4.19E-03 | -3.21E-04 |
| 11513 | Dcx | DCX | 7.37E-03 | -8.96E-03 |
| 11514 | Dcun1d1 | DCUN1D1 | 6.51E-03 | -9.38E-03 |
| 11515 | Tnfsf15 | TNFSF15 | 2.37E-02 | -1.48E-01 |
| 11516 | Onecut2 | ONECUT2 | -7.09E-03 | 2.25E-02 |
| 11517 | Hba-a1 | HBA2 | -2.87E-02 | 2.57E-02 |
| 11518 | Hba-a1 | HBA1 | -2.87E-02 | 2.57E-02 |
| 11519 | Tmem176a | TMEM176A | -6.72E-03 | 3.06E-02 |
| 11520 | Gabbr2 | GABBR2 | 8.29E-03 | -3.18E-02 |
| 11521 | Thbd | THBD | -8.72E-03 | 3.75E-02 |
| 11522 | Apoc4 | APOC4 | 9.64E-03 | 5.59E-02 |
| 11523 | Crygn | CRYGN | -5.98E-03 | -6.89E-03 |
| 11524 | Ccdc154 | CCDC154 | 1.25E-02 | -2.07E-02 |
| 11525 | Dstyk | DSTYK | -4.80E-03 | -1.92E-02 |
| 11526 | Adcy6 | ADCY6 | -6.36E-03 | -1.47E-02 |
| 11527 | Card10 | CARD10 | 8.16E-03 | -3.59E-03 |
| 11528 | Zfp414 | ZNF414 | 8.95E-03 | -1.23E-02 |
| 11529 | Clec18a | CLEC18B | 3.66E-02 | 1.83E-01 |
| 11530 | Sf3b3 | SF3B3 | 3.58E-03 | -4.14E-04 |
| 11531 | Arl6ip5 | ARL6IP5 | -3.92E-03 | 1.67E-02 |
| 11532 | Grpel2 | GRPEL2 | 6.60E-03 | -1.27E-02 |
| 11533 | Ssh3 | SSH3 | 4.77E-03 | 1.77E-02 |
| 11534 | Pdzd3 | PDZD3 | -9.19E-03 | -1.83E-02 |
| 11535 | Fbxo3 | FBXO3 | -4.57E-03 | -1.87E-02 |
| 11536 | Egfl6 | EGFL6 | -3.84E-02 | -9.90E-02 |
| 11537 | Adrb2 | ADRB2 | -1.62E-02 | -7.54E-02 |
| 11538 | Ssc5d | SSC5D | -2.17E-02 | 1.00E-01 |
| 11539 | Arhgef18 | ARHGEF18 | 3.87E-03 | -1.82E-02 |
| 11540 | Snupn | SNUPN | -7.54E-03 | -1.85E-02 |
| 11541 | Chst7 | CHST7 | -2.06E-02 | 3.73E-02 |
| 11542 | Mapkbp1 | MAPKBP1 | 5.38E-03 | -4.94E-05 |
| 11543 | C5ar1 | C5AR1 | 2.74E-02 | 9.43E-02 |
| 11544 | Qrich1 | QRICH1 | 4.04E-03 | 5.81E-03 |
| 11545 | Fam102a | FAM102A | -7.36E-03 | 4.78E-03 |
| 11546 | Med1 | MED1 | -4.32E-03 | -1.03E-02 |
| 11547 | Ints1 | INTS1 | 4.04E-03 | -1.61E-02 |
| 11548 | Ago4 | AGO4 | -9.30E-03 | -1.72E-02 |
| 11549 | Rabl2 | RABL2A | -7.03E-03 | 2.63E-04 |
| 11550 | Samd7 | SAMD7 | 9.13E-03 | 1.23E-03 |
| 11551 | 1700008O03Rik | C19orf81 | 1.60E-02 | 2.70E-02 |

| | | | | |
|---|---|---|---|---|
| 11552 | Fance | FANCE | 6.49E-03 | 5.23E-03 |
| 11553 | Slc39a9 | SLC39A9 | -3.90E-03 | -1.07E-02 |
| 11554 | Rab10 | RAB10 | 3.99E-03 | 7.43E-04 |
| 11555 | Dcps | DCPS | -5.62E-03 | -1.80E-02 |
| 11556 | Sigmar1 | SIGMAR1 | -4.40E-03 | 1.66E-02 |
| 11557 | Ankrd45 | ANKRD45 | 2.72E-02 | 1.66E-02 |
| 11558 | Dhrs9 | DHRS9 | -1.11E-02 | 5.61E-02 |
| 11559 | Axin2 | AXIN2 | 4.17E-02 | -2.00E-02 |
| 11560 | Trim29 | TRIM29 | 6.44E-02 | 5.83E-02 |
| 11561 | Celf2 | CELF2 | -1.05E-02 | 7.32E-02 |
| 11562 | Plekhj1 | PLEKHJ1 | -6.86E-03 | 1.34E-02 |
| 11563 | Dmgdh | DMGDH | 1.08E-02 | -5.12E-02 |
| 11564 | Arhgap19 | ARHGAP19 | 2.08E-02 | 7.30E-02 |
| 11565 | Zfp26 | ZNF778 | 7.28E-03 | 3.44E-04 |
| 11566 | Atp9a | ATP9A | -4.73E-03 | -1.92E-02 |
| 11567 | Clmp | CLMP | -2.52E-02 | -1.01E-03 |
| 11568 | Cbs | CBS | 7.87E-03 | -1.25E-02 |
| 11569 | Cbs | LOC102724560 | 7.87E-03 | -1.25E-02 |
| 11570 | Setd5 | SETD5 | 3.93E-03 | 1.15E-04 |
| 11571 | Ctrl | CTRL | -1.73E-02 | 3.18E-03 |
| 11572 | Scn11a | SCN11A | 1.36E-02 | -1.08E-02 |
| 11573 | Capn15 | CAPN15 | -7.08E-03 | 1.12E-02 |
| 11574 | Sirt6 | SIRT6 | -7.51E-03 | -2.32E-02 |
| 11575 | Wdpcp | WDPCP | -8.06E-03 | -1.35E-03 |
| 11576 | Mrps22 | MRPS22 | 8.40E-03 | -1.07E-02 |
| 11577 | Hsd17b11 | HSD17B11 | -5.43E-03 | -1.02E-02 |
| 11578 | Ldhal6b | LDHAL6B | -1.26E-02 | -2.54E-03 |
| 11579 | Nudt16l1 | NUDT16L1 | 5.77E-03 | -2.47E-02 |
| 11580 | Kdelr3 | KDELR3 | 2.53E-02 | 1.05E-01 |
| 11581 | Pttg1ip | PTTG1IP | 3.75E-03 | -1.26E-02 |
| 11582 | Snapc2 | SNAPC2 | 6.16E-03 | 8.56E-03 |
| 11583 | Pold3 | POLD3 | 4.53E-03 | -3.15E-03 |
| 11584 | Sf3b5 | SF3B5 | -4.78E-03 | 2.09E-03 |
| 11585 | Phf3 | PHF3 | -3.40E-03 | 3.10E-03 |
| 11586 | Usp36 | USP36 | 4.88E-03 | -1.41E-02 |
| 11587 | Prss35 | PRSS35 | -5.24E-02 | 1.62E-01 |
| 11588 | Gphn | GPHN | 7.27E-03 | -2.13E-02 |
| 11589 | Mro | MRO | -2.20E-02 | -8.61E-02 |
| 11590 | Trmt11 | TRMT11 | 8.23E-03 | -5.04E-04 |
| 11591 | 2810004N23Rik | C1orf131 | -3.91E-03 | -4.46E-03 |
| 11592 | Cpeb1 | CPEB1 | 1.01E-02 | -2.28E-02 |
| 11593 | Fzd7 | FZD7 | -1.01E-02 | 1.19E-02 |
| 11594 | Atp2c1 | ATP2C1 | 5.01E-03 | -2.05E-02 |
| 11595 | Gpr31b | GPR31 | -6.71E-03 | -7.06E-03 |
| 11596 | Frat1 | FRAT1 | -1.44E-02 | -2.56E-02 |
| 11597 | Gnpda2 | GNPDA2 | 6.39E-03 | -8.64E-03 |
| 11598 | Snai1 | SNAI1 | 2.38E-02 | -2.90E-02 |
| 11599 | Ptov1 | PTOV1 | 4.37E-03 | 2.98E-03 |
| 11600 | Sgsm2 | SGSM2 | 8.09E-03 | -1.12E-02 |
| 11601 | Bcl2a1b | BCL2A1 | -7.99E-03 | 1.85E-02 |
| 11602 | Sema4f | SEMA4F | -8.04E-03 | 8.57E-03 |
| 11603 | Tnpo2 | TNPO2 | -3.39E-03 | 6.14E-03 |
| 11604 | Trim10 | TRIM10 | -2.57E-02 | 6.98E-02 |

| | | | | |
|---|---|---|---|---|
| 11605 | Amt | AMT | -5.93E-03 | -6.16E-02 |
| 11606 | Dyrk3 | DYRK3 | 1.65E-02 | 3.10E-02 |
| 11607 | Wnt4 | WNT4 | -7.18E-02 | 1.60E-01 |
| 11608 | Tanc1 | TANC1 | -4.33E-03 | -1.13E-02 |
| 11609 | Osbpl1a | OSBPL1A | -6.42E-03 | -4.38E-02 |
| 11610 | F9 | F9 | -1.33E-02 | 3.01E-02 |
| 11611 | Pcsk5 | PCSK5 | -1.15E-02 | -1.10E-02 |
| 11612 | Cplx1 | CPLX1 | 5.08E-03 | -5.57E-03 |
| 11613 | Krt222 | KRT222 | -1.98E-02 | -1.57E-02 |
| 11614 | Zfp169 | ZNF169 | -1.30E-02 | -2.13E-02 |
| 11615 | Patz1 | PATZ1 | -5.78E-03 | -8.38E-03 |
| 11616 | Ddah2 | DDAH2 | -8.43E-03 | 4.80E-02 |
| 11617 | Cda | CDA | -9.92E-03 | 1.61E-02 |
| 11618 | Azi2 | AZI2 | -4.12E-03 | -1.12E-02 |
| 11619 | Naga | NAGA | 4.92E-03 | -1.02E-02 |
| 11620 | Hspa5 | HSPA5 | 1.25E-02 | -5.07E-03 |
| 11621 | Ddx51 | DDX51 | 5.33E-03 | 3.71E-03 |
| 11622 | Tmtc3 | TMTC3 | -5.89E-03 | 5.05E-03 |
| 11623 | Aup1 | AUP1 | 3.99E-03 | -7.53E-03 |
| 11624 | Dnajb6 | DNAJB6 | -5.81E-03 | -2.96E-03 |
| 11625 | Yod1 | YOD1 | -8.74E-03 | -7.21E-03 |
| 11626 | Car14 | CA14 | 9.87E-03 | -2.96E-02 |
| 11627 | Ptpn14 | PTPN14 | -1.90E-02 | -1.07E-02 |
| 11628 | Cdk5rap1 | CDK5RAP1 | 3.05E-02 | -8.05E-02 |
| 11629 | Actn2 | ACTN2 | -1.90E-02 | 6.70E-02 |
| 11630 | Dnajc1 | DNAJC1 | -3.72E-03 | 1.89E-02 |
| 11631 | Rpl5 | RPL5 | -4.37E-03 | 1.37E-02 |
| 11632 | Naa15 | NAA15 | 3.98E-03 | 9.26E-03 |
| 11633 | Ankrd35 | ANKRD35 | -1.50E-02 | 2.45E-02 |
| 11634 | Klhl24 | KLHL24 | -9.24E-03 | -3.82E-02 |
| 11635 | Vps72 | VPS72 | -3.89E-03 | -5.34E-03 |
| 11636 | Gjc2 | GJC2 | 1.40E-02 | -1.37E-02 |
| 11637 | Armc1 | ARMC1 | 5.11E-03 | -9.39E-03 |
| 11638 | Tlcd2 | TLCD2 | -1.23E-02 | 3.82E-02 |
| 11639 | Fgfr1 | FGFR1 | 5.50E-03 | -1.80E-03 |
| 11640 | Mtmr7 | MTMR7 | 1.78E-02 | 8.77E-03 |
| 11641 | Amacr | AMACR | -7.19E-03 | -2.49E-02 |
| 11642 | Psmd5 | PSMD5 | 4.11E-03 | 5.37E-03 |
| 11643 | Clcc1 | CLCC1 | -5.52E-03 | -1.22E-02 |
| 11644 | Aipl1 | AIPL1 | -6.58E-03 | 1.77E-02 |
| 11645 | Mitd1 | MITD1 | 8.47E-03 | 1.72E-02 |
| 11646 | Acbd7 | ACBD7 | 1.03E-02 | -7.68E-03 |
| 11647 | Trim23 | TRIM23 | -6.80E-03 | -2.57E-02 |
| 11648 | Pth2r | PTH2R | -8.60E-03 | 2.68E-02 |
| 11649 | Miox | MIOX | 1.04E-02 | -9.70E-03 |
| 11650 | Fst | FST | 5.48E-02 | -3.23E-02 |
| 11651 | Fat3 | FAT3 | -1.54E-02 | 4.18E-03 |
| 11652 | Erbb3 | ERBB3 | 1.29E-02 | -3.61E-02 |
| 11653 | Prpf6 | PRPF6 | -3.34E-03 | -1.18E-02 |
| 11654 | Alas1 | ALAS1 | -1.07E-02 | -4.66E-02 |
| 11655 | Madd | MADD | -6.26E-03 | -5.80E-03 |
| 11656 | Cacng1 | CACNG1 | -8.64E-03 | -6.21E-03 |
| 11657 | Pias2 | PIAS2 | 4.19E-03 | -4.76E-03 |

| | | | | |
|---|---|---|---|---|
| 11658 | Sim2 | SIM2 | -3.61E-02 | -7.92E-02 |
| 11659 | Mtx1 | MTX1 | 4.41E-03 | -1.86E-02 |
| 11660 | Dcdc2a | DCDC2 | 1.99E-02 | 1.20E-01 |
| 11661 | Chmp5 | CHMP5 | -4.41E-03 | -1.26E-02 |
| 11662 | Cadps | CADPS | 1.79E-02 | 2.89E-02 |
| 11663 | Psmg2 | PSMG2 | 6.80E-03 | -8.25E-04 |
| 11664 | Bicd2 | BICD2 | 6.74E-03 | -1.68E-03 |
| 11665 | Fgf20 | FGF20 | 1.02E-02 | 1.94E-02 |
| 11666 | Lmo1 | LMO1 | 2.72E-02 | -5.98E-02 |
| 11667 | Mettl25 | METTL25 | 8.23E-03 | -5.06E-02 |
| 11668 | Chrd | CHRD | -1.27E-02 | 1.83E-02 |
| 11669 | Ldlrad4 | LDLRAD4 | -8.35E-03 | -4.90E-02 |
| 11670 | Col9a2 | COL9A2 | -5.96E-03 | 6.38E-03 |
| 11671 | Jade3 | JADE3 | -8.25E-03 | 4.02E-02 |
| 11672 | Xxylt1 | XXYLT1 | 6.17E-03 | -1.08E-02 |
| 11673 | Asgr1 | ASGR1 | 2.22E-02 | -4.50E-02 |
| 11674 | Msn | MSN | -7.47E-03 | 1.19E-04 |
| 11675 | Noa1 | NOA1 | -5.91E-03 | -3.64E-04 |
| 11676 | Rtn4rl1 | RTN4RL1 | 7.15E-03 | -3.03E-02 |
| 11677 | C1galt1 | C1GALT1 | 4.90E-03 | -1.05E-02 |
| 11678 | Lonp2 | LONP2 | -6.13E-03 | -1.10E-02 |
| 11679 | Nme4 | NME4 | -1.74E-02 | -1.89E-02 |
| 11680 | Fam189a1 | FAM189A1 | 8.13E-03 | 1.37E-02 |
| 11681 | Map6 | MAP6 | -5.91E-02 | 1.58E-01 |
| 11682 | Zrsr2 | ZRSR2 | 5.10E-03 | -7.45E-03 |
| 11683 | Nol12 | NOL12 | 7.33E-03 | 1.24E-02 |
| 11684 | Myo6 | MYO6 | -4.41E-03 | -1.58E-02 |
| 11685 | Zfp692 | ZNF692 | 9.16E-03 | -7.40E-03 |
| 11686 | Mefv | MEFV | -2.20E-02 | 8.01E-02 |
| 11687 | Adam12 | ADAM12 | 3.58E-02 | 7.98E-02 |
| 11688 | Slitrk5 | SLITRK5 | -6.29E-03 | 2.52E-02 |
| 11689 | Tnfrsf8 | TNFRSF8 | -9.05E-03 | 9.33E-03 |
| 11690 | Lnx1 | LNX1 | -1.42E-02 | -3.33E-02 |
| 11691 | Mob1b | MOB1B | 5.21E-03 | -3.96E-04 |
| 11692 | Cdip1 | CDIP1 | -4.29E-03 | -5.29E-03 |
| 11693 | Polr3b | POLR3B | -4.87E-03 | -2.39E-03 |
| 11694 | Zw10 | ZW10 | 4.85E-03 | 8.95E-03 |
| 11695 | Cdhr5 | CDHR5 | -5.65E-03 | -6.59E-04 |
| 11696 | Gpatch8 | GPATCH8 | 3.33E-03 | -9.91E-03 |
| 11697 | Iws1 | IWS1 | 3.89E-03 | -1.80E-02 |
| 11698 | Hoxb3 | HOXB3 | 8.11E-03 | -3.57E-03 |
| 11699 | Sft2d1 | SFT2D1 | 5.97E-03 | -2.79E-03 |
| 11700 | Atrip | ATRIP | -1.50E-02 | -2.87E-02 |
| 11701 | Mreg | MREG | 2.33E-02 | -5.65E-04 |
| 11702 | Tbcel | TBCEL | 6.23E-03 | -2.28E-02 |
| 11703 | Prkcsh | PRKCSH | 4.30E-03 | 3.46E-04 |
| 11704 | Xrn1 | XRN1 | -4.98E-03 | -1.72E-02 |
| 11705 | Ip6k3 | IP6K3 | 1.29E-02 | 2.10E-03 |
| 11706 | Cdca2 | CDCA2 | 3.78E-02 | 2.44E-01 |
| 11707 | Insl3 | INSL3 | 2.07E-02 | 4.59E-02 |
| 11708 | Rassf10 | RASSF10 | -2.02E-02 | -1.04E-01 |
| 11709 | Npy2r | NPY2R | -1.03E-02 | -2.38E-02 |
| 11710 | Cnot6 | CNOT6 | -5.83E-03 | -8.90E-04 |

| | | | | |
|---|---|---|---|---|
| 11711 | Fat2 | FAT2 | 6.50E-02 | 1.28E-01 |
| 11712 | Dip2c | DIP2C | 5.06E-03 | -3.48E-02 |
| 11713 | Cpsf1 | CPSF1 | 4.56E-03 | -1.52E-02 |
| 11714 | Ssh1 | SSH1 | 5.67E-03 | -4.62E-03 |
| 11715 | Arhgef25 | ARHGEF25 | -1.36E-02 | 8.56E-03 |
| 11716 | Scn3a | SCN3A | 1.52E-02 | 1.46E-01 |
| 11717 | Acot8 | ACOT8 | 7.10E-03 | -7.96E-03 |
| 11718 | Cnot7 | CNOT7 | -4.16E-03 | 3.92E-03 |
| 11719 | Ago2 | AGO2 | 4.92E-03 | -5.96E-03 |
| 11720 | Tcea2 | TCEA2 | 8.22E-03 | 4.89E-03 |
| 11721 | Polr1a | POLR1A | 4.83E-03 | 1.84E-02 |
| 11722 | Zfp516 | ZNF516 | 9.10E-03 | -2.24E-02 |
| 11723 | Neurl1a | NEURL1 | 1.95E-02 | 7.60E-02 |
| 11724 | Abcc6 | ABCC6 | -1.51E-02 | -7.91E-02 |
| 11725 | Saa1 | SAA1 | 2.21E-02 | 3.36E-01 |
| 11726 | Gmppb | GMPPB | -7.04E-03 | 1.43E-02 |
| 11727 | Gsdmc3 | GSDMC | 1.08E-01 | 1.95E-01 |
| 11728 | Nle1 | NLE1 | -9.75E-03 | 2.85E-02 |
| 11729 | Rnf181 | RNF181 | 1.21E-02 | 1.23E-02 |
| 11730 | Pggt1b | PGGT1B | -8.58E-03 | -2.71E-02 |
| 11731 | Lhfpl2 | LHFPL2 | -1.32E-02 | 6.55E-02 |
| 11732 | Ralgds | RALGDS | 8.09E-03 | -5.17E-02 |
| 11733 | Ccdc115 | CCDC115 | 4.88E-03 | -1.03E-02 |
| 11734 | Capn1 | CAPN1 | 9.16E-03 | 1.71E-03 |
| 11735 | Pgap3 | PGAP3 | 5.60E-03 | -1.46E-02 |
| 11736 | U2af2 | U2AF2 | -3.14E-03 | 2.71E-03 |
| 11737 | A530016L24Rik | C14orf180 | 2.64E-02 | 2.96E-02 |
| 11738 | Tlk1 | TLK1 | 4.46E-03 | -1.50E-02 |
| 11739 | Rassf6 | RASSF6 | 1.73E-02 | 8.68E-03 |
| 11740 | Lrrn4cl | LRRN4CL | -1.16E-02 | 6.10E-03 |
| 11741 | 2510009E07Rik | C3orf70 | -6.23E-03 | -4.10E-02 |
| 11742 | Map7 | MAP7 | 7.29E-03 | -2.99E-02 |
| 11743 | Gtpbp6 | GTPBP6 | -6.09E-03 | 9.09E-05 |
| 11744 | Sin3b | SIN3B | 3.59E-03 | -1.28E-03 |
| 11745 | Ptprb | PTPRB | 8.88E-03 | -2.92E-02 |
| 11746 | Gpr87 | GPR87 | -9.75E-03 | 1.15E-02 |
| 11747 | Rpp30 | RPP30 | -9.25E-03 | 2.53E-02 |
| 11748 | Slc23a3 | SLC23A3 | -1.69E-02 | -1.02E-01 |
| 11749 | Rnf2 | RNF2 | 5.34E-03 | 5.54E-03 |
| 11750 | Zdhhc12 | ZDHHC12 | -7.35E-03 | 1.35E-02 |
| 11751 | Lsm2 | LSM2 | -9.38E-03 | 5.26E-02 |
| 11752 | Pip5kl1 | PIP5KL1 | 6.52E-03 | 2.79E-03 |
| 11753 | Slc16a13 | SLC16A13 | 1.13E-02 | 5.80E-03 |
| 11754 | Cfhr1 | CFHR1 | -5.16E-03 | 1.03E-02 |
| 11755 | Nek9 | NEK9 | -3.87E-03 | -1.23E-02 |
| 11756 | Gjb6 | GJB6 | 2.96E-02 | 8.29E-02 |
| 11757 | Postn | POSTN | 5.25E-02 | 1.75E-01 |
| 11758 | Fbxl19 | FBXL19 | 7.30E-03 | 1.69E-02 |
| 11759 | Patl1 | PATL1 | -4.56E-03 | 3.49E-03 |
| 11760 | Nck2 | NCK2 | -6.10E-03 | 7.84E-03 |
| 11761 | Tprn | TPRN | 5.62E-03 | 1.16E-02 |
| 11762 | Ado | ADO | -4.28E-03 | 2.04E-02 |
| 11763 | Duox2 | DUOX2 | 9.51E-03 | -9.22E-03 |

| | | | | |
|---|---|---|---|---|
| 11764 | Car4 | CA4 | 7.28E-03 | -3.33E-02 |
| 11765 | Nkapl | NKAPL | -1.31E-02 | -7.38E-03 |
| 11766 | Esyt1 | ESYT1 | 1.02E-02 | 4.73E-02 |
| 11767 | Hal | HAL | -1.89E-02 | 5.93E-02 |
| 11768 | Agxt | AGXT | 1.30E-02 | 2.05E-02 |
| 11769 | Unc5a | UNC5A | 3.71E-02 | 1.19E-01 |
| 11770 | Axin1 | AXIN1 | 3.46E-03 | -5.19E-03 |
| 11771 | Lrrd1 | LRRD1 | 1.67E-02 | 2.69E-02 |
| 11772 | B3gnt6 | B3GNT6 | -8.65E-03 | -1.17E-02 |
| 11773 | Lyl1 | LYL1 | -1.45E-02 | 4.20E-02 |
| 11774 | Hsd17b3 | HSD17B3 | -5.73E-03 | -9.04E-03 |
| 11775 | Dynlt1c | DYNLT1 | 1.05E-02 | -2.20E-02 |
| 11776 | Erp29 | ERP29 | -5.69E-03 | -1.75E-03 |
| 11777 | Desi1 | DESI1 | -6.37E-03 | 1.38E-02 |
| 11778 | Cldn10 | CLDN10 | 1.13E-02 | -3.41E-02 |
| 11779 | Impa2 | IMPA2 | -8.06E-03 | 6.97E-03 |
| 11780 | Dpy19l1 | DPY19L1 | 8.41E-03 | 1.80E-02 |
| 11781 | Arid3c | ARID3C | -6.37E-03 | 4.56E-03 |
| 11782 | Dnajc15 | DNAJC15 | -5.14E-03 | -1.73E-02 |
| 11783 | Sirt1 | SIRT1 | -7.41E-03 | -1.93E-02 |
| 11784 | Dpm1 | DPM1 | 8.72E-03 | -1.89E-02 |
| 11785 | Gpr141 | GPR141 | 2.50E-02 | 6.84E-02 |
| 11786 | Ormdl3 | ORMDL3 | -5.40E-03 | -5.03E-03 |
| 11787 | Fadd | FADD | 6.17E-03 | -7.03E-03 |
| 11788 | Frat2 | FRAT2 | -1.85E-02 | 1.82E-02 |
| 11789 | Fam151a | FAM151A | -1.04E-02 | -2.96E-02 |
| 11790 | Pigq | PIGQ | 3.87E-03 | -1.86E-02 |
| 11791 | Fdxr | FDXR | -6.15E-03 | -2.09E-02 |
| 11792 | Mars2 | MARS2 | 6.00E-03 | 6.96E-04 |
| 11793 | Poldip3 | POLDIP3 | -3.13E-03 | 2.75E-03 |
| 11794 | Epg5 | EPG5 | -4.83E-03 | -1.71E-02 |
| 11795 | Bbs4 | BBS4 | 5.69E-03 | -1.63E-02 |
| 11796 | Numb | NUMB | -5.56E-03 | -1.89E-04 |
| 11797 | Mphosph10 | MPHOSPH10 | 6.69E-03 | 2.12E-02 |
| 11798 | Tjp2 | TJP2 | 5.50E-03 | 1.02E-02 |
| 11799 | Tssk3 | TSSK3 | -6.53E-03 | 8.03E-03 |
| 11800 | Nudcd1 | NUDCD1 | 8.17E-03 | 1.38E-02 |
| 11801 | Smarcd3 | SMARCD3 | 1.46E-02 | 2.74E-02 |
| 11802 | Mien1 | MIEN1 | -4.48E-03 | -4.01E-03 |
| 11803 | Hbs1l | HBS1L | 3.56E-03 | -1.29E-02 |
| 11804 | Cdh1 | CDH1 | -6.88E-03 | 8.88E-04 |
| 11805 | Dolpp1 | DOLPP1 | 6.64E-03 | -3.89E-03 |
| 11806 | Mrps7 | MRPS7 | -4.36E-03 | -1.25E-02 |
| 11807 | Tmem81 | TMEM81 | 1.14E-02 | -2.62E-02 |
| 11808 | Slx1b | SLX1A | 7.64E-03 | -8.67E-03 |
| 11809 | Slx1b | SLX1B | 7.64E-03 | -8.67E-03 |
| 11810 | Dbn1 | DBN1 | 6.22E-03 | 3.26E-02 |
| 11811 | Baz2a | BAZ2A | 3.39E-03 | -9.17E-03 |
| 11812 | Ugt1a2 | UGT1A3 | 1.70E-02 | -5.90E-02 |
| 11813 | Slc18b1 | SLC18B1 | 7.14E-03 | -6.20E-03 |
| 11814 | Spop | SPOP | -4.04E-03 | -2.14E-02 |
| 11815 | Pth1r | PTH1R | 6.87E-03 | -8.84E-03 |
| 11816 | Klhl7 | KLHL7 | -5.58E-03 | 5.45E-04 |

| | | | | |
|---|---|---|---|---|
| 11817 | Wnt10b | WNT10B | 1.07E-02 | 4.68E-02 |
| 11818 | Hspb2 | HSPB2 | 2.97E-02 | -4.72E-02 |
| 11819 | Gcc2 | GCC2 | -6.05E-03 | -2.50E-02 |
| 11820 | Peli1 | PELI1 | 6.49E-03 | -8.50E-03 |
| 11821 | Rbm18 | RBM18 | -5.66E-03 | -2.21E-03 |
| 11822 | Kmt2b | KMT2B | 4.17E-03 | -4.01E-03 |
| 11823 | Fam209 | FAM209B | 9.27E-03 | 4.74E-03 |
| 11824 | Tmod2 | TMOD2 | 2.04E-02 | 5.97E-03 |
| 11825 | Dgkb | DGKB | 2.13E-02 | -6.57E-02 |
| 11826 | Pigx | PIGX | -4.80E-03 | 1.70E-02 |
| 11827 | Tsks | TSKS | -5.28E-03 | 7.04E-03 |
| 11828 | Cacna1b | CACNA1B | -7.67E-03 | 1.84E-02 |
| 11829 | Ap3m1 | AP3M1 | 3.80E-03 | -5.61E-03 |
| 11830 | Pfn3 | PFN3 | -4.47E-03 | 3.14E-03 |
| 11831 | Cep290 | CEP290 | -8.32E-03 | -8.94E-03 |
| 11832 | Pum1 | PUM1 | -4.97E-03 | -6.21E-03 |
| 11833 | Ier2 | IER2 | 2.85E-02 | -2.34E-02 |
| 11834 | Nxt1 | NXT1 | 9.15E-03 | 3.95E-02 |
| 11835 | Dlg1 | DLG1 | 4.99E-03 | -2.03E-02 |
| 11836 | Lasp1 | LASP1 | -3.44E-03 | -5.29E-03 |
| 11837 | Limd2 | LIMD2 | 8.22E-03 | -5.03E-03 |
| 11838 | Rcn2 | RCN2 | -6.70E-03 | 1.95E-03 |
| 11839 | Greb1l | GREB1L | 1.56E-02 | -1.28E-02 |
| 11840 | Ssbp1 | SSBP1 | 3.88E-03 | -2.86E-03 |
| 11841 | Atr | ATR | -5.35E-03 | -1.20E-02 |
| 11842 | Ppp1r10 | PPP1R10 | 9.69E-03 | -7.77E-03 |
| 11843 | Fam118b | FAM118B | 9.33E-03 | 5.60E-02 |
| 11844 | Sla | SLA | 1.53E-02 | 1.02E-01 |
| 11845 | Atl1 | ATL1 | -1.25E-02 | -4.20E-02 |
| 11846 | Tpmt | TPMT | 5.30E-03 | -1.86E-02 |
| 11847 | Cmtm8 | CMTM8 | 8.48E-03 | 8.29E-04 |
| 11848 | Senp6 | SENP6 | -4.05E-03 | -7.81E-03 |
| 11849 | Ccnt1 | CCNT1 | -6.42E-03 | -6.12E-03 |
| 11850 | Gch1 | GCH1 | 1.62E-02 | 1.23E-01 |
| 11851 | Zc3h3 | ZC3H3 | -7.25E-03 | 4.74E-03 |
| 11852 | Npc1l1 | NPC1L1 | -1.58E-02 | -5.12E-02 |
| 11853 | Gpr62 | GPR62 | -7.43E-03 | 3.95E-03 |
| 11854 | Zkscan1 | ZKSCAN1 | 5.06E-03 | -1.64E-02 |
| 11855 | Pcdhga7 | PCDHGA7 | -1.29E-02 | -3.19E-02 |
| 11856 | Ccdc14 | CCDC14 | 1.50E-02 | 1.43E-02 |
| 11857 | Mid2 | MID2 | -5.91E-03 | -2.15E-02 |
| 11858 | Fgf7 | FGF7 | -1.52E-02 | 4.41E-03 |
| 11859 | Bin1 | BIN1 | -6.64E-03 | 5.61E-03 |
| 11860 | Ap1ar | AP1AR | 4.07E-03 | -5.54E-03 |
| 11861 | Lrrc3 | LRRC3 | -9.24E-03 | -3.15E-02 |
| 11862 | Tex38 | TEX38 | -1.45E-02 | -5.13E-02 |
| 11863 | Rbl2 | RBL2 | 4.29E-03 | -2.73E-02 |
| 11864 | Tceanc | TCEANC | -1.14E-02 | -3.15E-02 |
| 11865 | Oxsm | OXSM | 6.68E-03 | 6.04E-04 |
| 11866 | Tmem202 | TMEM202 | -9.15E-03 | 6.26E-02 |
| 11867 | Scara3 | SCARA3 | 2.16E-02 | 2.26E-02 |
| 11868 | Oc90 | OC90 | 6.68E-03 | 2.76E-02 |
| 11869 | Gtf2h2 | GTF2H2 | 7.76E-03 | -3.13E-02 |

| | | | | |
|---|---|---|---|---|
| 11870 | Serpina7 | SERPINA7 | -1.39E-02 | 5.23E-02 |
| 11871 | Crhbp | CRHBP | 5.81E-03 | 1.14E-02 |
| 11872 | Sarnp | SARNP | -4.35E-03 | 1.07E-02 |
| 11873 | Htatsf1 | HTATSF1 | 3.56E-03 | -1.05E-02 |
| 11874 | Tmem40 | TMEM40 | 2.27E-02 | 1.06E-01 |
| 11875 | Boll | BOLL | -7.72E-03 | 4.44E-03 |
| 11876 | Wdr92 | WDR92 | -5.60E-03 | -1.84E-02 |
| 11877 | Kif14 | KIF14 | 2.96E-02 | 1.87E-01 |
| 11878 | Rps15 | RPS15 | -4.08E-03 | 7.73E-03 |
| 11879 | Nr1h2 | NR1H2 | -4.44E-03 | -1.53E-02 |
| 11880 | Pla2g3 | PLA2G3 | 1.39E-02 | -8.11E-03 |
| 11881 | Efnb3 | EFNB3 | 1.59E-02 | -3.01E-02 |
| 11882 | Tlr4 | TLR4 | -1.84E-02 | 5.29E-02 |
| 11883 | Dgka | DGKA | -4.44E-03 | 8.54E-03 |
| 11884 | Pglyrp1 | PGLYRP1 | 3.11E-02 | 1.21E-01 |
| 11885 | Dcstamp | DCSTAMP | 6.80E-03 | 3.47E-02 |
| 11886 | Apol6 | APOL6 | 4.70E-02 | 8.57E-02 |
| 11887 | 2310057M21Rik | C10orf88 | -8.53E-03 | -2.37E-03 |
| 11888 | Calml4 | CALML4 | -5.42E-03 | -2.14E-02 |
| 11889 | Kdm7a | KDM7A | 5.27E-03 | -1.09E-02 |
| 11890 | Prrc2a | PRRC2A | 3.03E-03 | -1.20E-03 |
| 11891 | Npy5r | NPY5R | -2.14E-02 | -2.38E-02 |
| 11892 | Slamf1 | SLAMF1 | -6.62E-03 | -2.03E-04 |
| 11893 | Bmpr2 | BMPR2 | 4.69E-03 | -8.94E-03 |
| 11894 | Parn | PARN | 4.00E-03 | -1.02E-02 |
| 11895 | Gm10177 | SEC61G | -9.95E-03 | 9.21E-03 |
| 11896 | Cyth2 | CYTH2 | 3.74E-03 | 2.17E-02 |
| 11897 | Fut8 | FUT8 | -8.89E-03 | 6.57E-03 |
| 11898 | Dnase1l2 | DNASE1L2 | -3.72E-02 | 5.83E-02 |
| 11899 | Utp3 | UTP3 | -4.36E-03 | 1.00E-02 |
| 11900 | 4833420G17Rik | C5orf34 | 6.62E-03 | -3.36E-02 |
| 11901 | Pycrl | PYCRL | -5.93E-03 | -5.92E-03 |
| 11902 | Irak1bp1 | IRAK1BP1 | -7.39E-03 | 1.90E-02 |
| 11903 | Eml4 | EML4 | -5.81E-03 | 1.05E-02 |
| 11904 | Klhl23 | PHOSPHO2-KLHL23 | -1.57E-02 | 4.69E-02 |
| 11905 | Klhl23 | KLHL23 | -1.57E-02 | 4.69E-02 |
| 11906 | Parp4 | PARP4 | 3.97E-03 | -1.32E-02 |
| 11907 | Slc26a7 | SLC26A7 | -3.51E-02 | -1.92E-01 |
| 11908 | Tbcc | TBCC | -7.04E-03 | 2.14E-02 |
| 11909 | Ppm1j | PPM1J | -1.68E-02 | 1.29E-01 |
| 11910 | Cadm1 | CADM1 | -8.15E-03 | 5.14E-02 |
| 11911 | Htr1d | HTR1D | 8.07E-03 | 2.03E-02 |
| 11912 | Phka2 | PHKA2 | -7.10E-03 | 5.01E-03 |
| 11913 | Dsp | DSP | 1.74E-02 | 1.24E-02 |
| 11914 | Gmfb | GMFB | 4.91E-03 | -6.33E-03 |
| 11915 | Nckap5 | NCKAP5 | 1.29E-02 | -4.18E-02 |
| 11916 | Bmpr1b | BMPR1B | -2.06E-02 | -3.68E-03 |
| 11917 | Gjc3 | GJC3 | 5.68E-03 | 3.66E-03 |
| 11918 | Sirt3 | SIRT3 | 7.23E-03 | -2.40E-02 |
| 11919 | Itga6 | ITGA6 | -7.20E-03 | 4.20E-02 |
| 11920 | R3hdm2 | R3HDM2 | -4.23E-03 | -8.21E-03 |
| 11921 | Tmem167 | TMEM167A | 5.21E-03 | 4.59E-03 |
| 11922 | Tcf20 | TCF20 | 3.26E-03 | -1.60E-02 |

| | | | | |
|---|---|---|---|---|
| 11923 | Kctd13 | KCTD13 | 6.45E-03 | -1.19E-02 |
| 11924 | Fbl | FBL | 4.73E-03 | -4.91E-03 |
| 11925 | Dus3l | DUS3L | 3.65E-03 | -2.30E-02 |
| 11926 | Ctps | CTPS1 | 9.59E-03 | 5.44E-02 |
| 11927 | Brat1 | BRAT1 | -4.83E-03 | -4.00E-03 |
| 11928 | Rpl12 | RPL12 | 4.74E-03 | 2.44E-02 |
| 11929 | Poc1b | POC1B | -5.09E-03 | -2.05E-03 |
| 11930 | Rps14 | RPS14 | -3.77E-03 | 1.27E-02 |
| 11931 | Mocs3 | MOCS3 | -7.62E-03 | 2.87E-02 |
| 11932 | Cbx4 | CBX4 | -1.15E-02 | -1.86E-02 |
| 11933 | Ift81 | IFT81 | -4.70E-03 | -9.49E-03 |
| 11934 | Sec61b | SEC61B | 5.15E-03 | -7.88E-03 |
| 11935 | Stx18 | STX18 | 4.28E-03 | 1.35E-03 |
| 11936 | Dus2 | DUS2 | 5.89E-03 | 7.54E-03 |
| 11937 | Nr2e3 | NR2E3 | -6.47E-02 | -8.17E-02 |
| 11938 | Itga3 | ITGA3 | 1.12E-02 | 1.91E-02 |
| 11939 | Cldn6 | CLDN6 | 1.52E-02 | 3.77E-02 |
| 11940 | Myzap | MYZAP | -1.53E-02 | 4.05E-03 |
| 11941 | Cyp26b1 | CYP26B1 | -2.62E-02 | -1.67E-01 |
| 11942 | Zxdc | ZXDC | -4.54E-03 | 1.20E-02 |
| 11943 | Tusc2 | TUSC2 | 4.31E-03 | 2.51E-03 |
| 11944 | Ascl1 | ASCL1 | 6.42E-03 | 2.12E-02 |
| 11945 | Cebpd | CEBPD | -2.79E-02 | 4.95E-02 |
| 11946 | Zdhhc5 | ZDHHC5 | 4.32E-03 | -8.21E-03 |
| 11947 | Slc31a2 | SLC31A2 | -8.67E-03 | 1.23E-02 |
| 11948 | Csf2rb | CSF2RB | -2.63E-02 | 8.97E-02 |
| 11949 | C4a | C4B | -1.77E-02 | 1.50E-01 |
| 11950 | C4a | C4A | -1.77E-02 | 1.50E-01 |
| 11951 | Best2 | BEST2 | 5.02E-03 | -3.63E-03 |
| 11952 | Sucnr1 | SUCNR1 | -2.01E-02 | 1.80E-02 |
| 11953 | Rnf44 | RNF44 | -3.73E-03 | -1.12E-02 |
| 11954 | Rpp38 | RPP38 | -9.85E-03 | 3.56E-02 |
| 11955 | Itm2c | ITM2C | 7.61E-03 | 9.29E-03 |
| 11956 | Casz1 | CASZ1 | 3.80E-03 | 5.84E-03 |
| 11957 | Socs5 | SOCS5 | -9.26E-03 | -3.80E-02 |
| 11958 | Soat1 | SOAT1 | -6.44E-03 | -3.14E-03 |
| 11959 | Serpina3m | SERPINA3 | 7.03E-03 | 4.78E-02 |
| 11960 | Mpi | MPI | -5.19E-03 | -1.04E-02 |
| 11961 | Ptpn18 | PTPN18 | -1.01E-02 | -7.22E-04 |
| 11962 | Pcsk4 | PCSK4 | 1.19E-02 | -9.90E-03 |
| 11963 | Trp63 | TP63 | -3.88E-02 | 4.59E-02 |
| 11964 | Pfkfb3 | PFKFB3 | 1.99E-02 | 2.58E-02 |
| 11965 | Tmem251 | TMEM251 | 6.46E-03 | 4.12E-02 |
| 11966 | Gdf7 | GDF7 | -1.24E-02 | -2.02E-02 |
| 11967 | St6galnac6 | ST6GALNAC6 | -9.98E-03 | -1.21E-02 |
| 11968 | Nsmce1 | NSMCE1 | 6.02E-03 | 1.01E-03 |
| 11969 | Pof1b | POF1B | 3.96E-02 | 5.95E-02 |
| 11970 | Zbtb7c | ZBTB7C | 1.21E-02 | 4.96E-02 |
| 11971 | Sdc3 | SDC3 | -9.38E-03 | -2.01E-02 |
| 11972 | Atp7a | ATP7A | -8.14E-03 | -2.79E-02 |
| 11973 | Fabp12 | FABP12 | -6.73E-03 | -8.50E-03 |
| 11974 | 4933427D14Rik | KIAA0753 | 4.68E-03 | 9.06E-03 |
| 11975 | Ptgfrn | PTGFRN | 1.17E-02 | 3.67E-02 |

| | | | | |
|---|---|---|---|---|
| 11976 | Swi5 | SWI5 | 3.82E-03 | -1.40E-03 |
| 11977 | Dpf1 | DPF1 | 1.12E-02 | 4.93E-02 |
| 11978 | Sympk | SYMPK | -3.18E-03 | -1.30E-02 |
| 11979 | Ccl24 | CCL24 | -4.64E-03 | 2.26E-02 |
| 11980 | Mboat7 | MBOAT7 | -4.02E-03 | -7.95E-03 |
| 11981 | Ndst3 | NDST3 | -1.48E-02 | 5.48E-02 |
| 11982 | Kcnk3 | KCNK3 | 1.47E-02 | -4.89E-02 |
| 11983 | Gpa33 | GPA33 | 2.73E-02 | 9.00E-02 |
| 11984 | Auh | AUH | -4.62E-03 | -2.64E-02 |
| 11985 | Hnf4a | HNF4A | -6.63E-03 | -2.49E-02 |
| 11986 | Ppp4r2 | PPP4R2 | 5.28E-03 | 5.23E-03 |
| 11987 | Pcdhgc5 | PCDHGC5 | 8.91E-03 | -1.05E-02 |
| 11988 | Dcaf12 | DCAF12 | 3.62E-03 | 7.25E-03 |
| 11989 | Kndc1 | KNDC1 | -3.62E-03 | 6.05E-03 |
| 11990 | Araf | ARAF | -3.45E-03 | -1.11E-02 |
| 11991 | Rad51b | RAD51B | -1.91E-02 | 5.93E-02 |
| 11992 | Tmem130 | TMEM130 | 8.30E-03 | 2.68E-02 |
| 11993 | Chm | CHM | -4.93E-03 | -1.49E-02 |
| 11994 | Mrps6 | MRPS6 | 1.81E-02 | -9.24E-03 |
| 11995 | Parp1 | PARP1 | -4.27E-03 | -8.83E-03 |
| 11996 | Larp4 | LARP4 | -4.28E-03 | -7.24E-04 |
| 11997 | Cyp2c29 | CYP2C8 | 2.54E-02 | 1.24E-01 |
| 11998 | Abhd16a | ABHD16A | 3.33E-03 | -1.13E-02 |
| 11999 | Aifm1 | AIFM1 | 4.03E-03 | -1.65E-02 |
| 12000 | Mrps16 | MRPS16 | 4.57E-03 | -7.40E-03 |
| 12001 | Commd8 | COMMD8 | -3.85E-03 | -1.09E-02 |
| 12002 | Lrsam1 | LRSAM1 | -4.54E-03 | -1.88E-02 |
| 12003 | Mfap4 | MFAP4 | 2.46E-02 | 1.33E-01 |
| 12004 | Hmces | HMCES | -4.24E-03 | 3.17E-03 |
| 12005 | Tcf7l2 | TCF7L2 | -5.79E-03 | -3.89E-03 |
| 12006 | Zfp746 | ZNF746 | -4.64E-03 | 4.95E-03 |
| 12007 | Vit | VIT | -8.49E-03 | 2.98E-02 |
| 12008 | Atp6v0c | ATP6V0C | -6.01E-03 | -2.24E-02 |
| 12009 | Cby3 | CBY3 | 8.89E-03 | 1.71E-02 |
| 12010 | Hook3 | HOOK3 | 4.42E-03 | -1.01E-02 |
| 12011 | Tle3 | TLE3 | -5.68E-03 | -9.32E-03 |
| 12012 | Ndufaf3 | NDUFAF3 | -4.58E-03 | -1.15E-02 |
| 12013 | Gxylt2 | GXYLT2 | -2.00E-02 | 3.02E-01 |
| 12014 | Mrpl9 | MRPL9 | -4.03E-03 | -3.55E-03 |
| 12015 | Igfbp6 | IGFBP6 | 1.62E-02 | 5.78E-02 |
| 12016 | Tubgcp4 | TUBGCP4 | 4.10E-03 | -9.05E-03 |
| 12017 | Usp45 | USP45 | 4.91E-03 | -1.22E-02 |
| 12018 | Etv5 | ETV5 | 2.07E-02 | -4.20E-02 |
| 12019 | Kcnd2 | KCND2 | 1.54E-02 | -2.53E-02 |
| 12020 | Spire2 | SPIRE2 | 2.09E-02 | 7.10E-02 |
| 12021 | 0610010K14Rik | C17orf49 | 1.22E-02 | 1.12E-02 |
| 12022 | Ttc14 | TTC14 | -4.94E-03 | 1.92E-04 |
| 12023 | Sult1c2 | SULT1C2 | -7.49E-03 | -4.74E-02 |
| 12024 | Tesc | TESC | -1.41E-02 | 1.85E-03 |
| 12025 | Pradc1 | PRADC1 | 6.24E-03 | 2.28E-02 |
| 12026 | Prdx4 | PRDX4 | 8.42E-03 | 3.25E-02 |
| 12027 | Surf6 | SURF6 | 5.00E-03 | 2.67E-03 |
| 12028 | Bri3 | BRI3 | 5.93E-03 | -4.19E-03 |

| | | | | |
|---|---|---|---|---|
| 12029 | Mrpl24 | MRPL24 | -3.91E-03 | -5.94E-03 |
| 12030 | Nudt3 | NUDT3 | -3.70E-03 | -8.43E-03 |
| 12031 | Tmem131 | TMEM131 | -3.42E-03 | -1.03E-02 |
| 12032 | Sync | SYNC | 1.35E-02 | 9.08E-02 |
| 12033 | Dner | DNER | -4.50E-03 | 1.91E-02 |
| 12034 | Nog | NOG | -1.72E-02 | 1.48E-01 |
| 12035 | Hmcn1 | HMCN1 | -2.28E-02 | 2.29E-02 |
| 12036 | Stra6 | STRA6 | -2.23E-02 | -1.04E-01 |
| 12037 | Slc9a5 | SLC9A5 | 1.24E-02 | 4.44E-02 |
| 12038 | Fetub | FETUB | 1.37E-02 | 4.26E-02 |
| 12039 | Ankef1 | ANKEF1 | -1.61E-02 | 2.60E-02 |
| 12040 | Kctd3 | KCTD3 | 4.06E-03 | 1.84E-02 |
| 12041 | Cenph | CENPH | 3.01E-02 | 1.57E-01 |
| 12042 | Hspa4l | HSPA4L | -7.46E-03 | -3.31E-03 |
| 12043 | Aldh1a3 | ALDH1A3 | -1.10E-01 | 3.24E-01 |
| 12044 | Gbf1 | GBF1 | 4.62E-03 | -1.13E-02 |
| 12045 | Hpse | HPSE | 2.43E-02 | 9.26E-02 |
| 12046 | Pcdh11x | PCDH11X | -7.65E-03 | -2.13E-02 |
| 12047 | Dbndd1 | DBNDD1 | 5.30E-03 | 1.35E-02 |
| 12048 | Sar1a | SAR1A | 2.75E-03 | -1.48E-03 |
| 12049 | Dact2 | DACT2 | -7.84E-03 | -1.70E-02 |
| 12050 | Apaf1 | APAF1 | -7.96E-03 | 3.48E-02 |
| 12051 | Pde2a | PDE2A | -7.93E-03 | 3.17E-02 |
| 12052 | Pcdhga6 | PCDHGA6 | -1.23E-02 | -3.08E-02 |
| 12053 | Dmrt2 | DMRT2 | -2.49E-02 | -2.68E-02 |
| 12054 | Icam4 | ICAM4 | 1.18E-02 | -4.97E-02 |
| 12055 | Rpf1 | RPF1 | 4.61E-03 | 2.66E-03 |
| 12056 | Ptgfr | PTGFR | 7.55E-03 | 4.89E-02 |
| 12057 | Apobr | APOBR | 1.85E-02 | 8.44E-02 |
| 12058 | Eri1 | ERI1 | 6.52E-03 | 2.50E-02 |
| 12059 | Vac14 | VAC14 | 7.19E-03 | 3.21E-03 |
| 12060 | Hcfc1 | HCFC1 | 4.11E-03 | -4.97E-03 |
| 12061 | Rps7 | RPS7 | -3.87E-03 | 1.62E-02 |
| 12062 | Snx27 | SNX27 | -4.02E-03 | -1.24E-02 |
| 12063 | Aurkaip1 | AURKAIP1 | 3.37E-03 | -1.60E-02 |
| 12064 | Lrrc17 | LRRC17 | -1.70E-02 | 9.66E-02 |
| 12065 | Usp3 | USP3 | 5.46E-03 | -1.02E-02 |
| 12066 | Zfp707 | ZNF707 | 5.50E-03 | -8.56E-03 |
| 12067 | Samsn1 | SAMSN1 | -2.75E-02 | 8.55E-02 |
| 12068 | Slc17a9 | SLC17A9 | -1.34E-02 | -2.53E-02 |
| 12069 | Ptpn12 | PTPN12 | 4.45E-03 | 2.97E-02 |
| 12070 | Rufy4 | RUFY4 | -1.28E-02 | -8.05E-02 |
| 12071 | Tnpo3 | TNPO3 | -3.98E-03 | -5.90E-03 |
| 12072 | Serpinb5 | SERPINB5 | -2.28E-02 | 9.13E-02 |
| 12073 | Gpr3 | GPR3 | 2.59E-02 | 5.60E-02 |
| 12074 | Naa60 | NAA60 | -3.99E-03 | -1.84E-03 |
| 12075 | Gba2 | GBA2 | -4.92E-03 | -1.98E-03 |
| 12076 | Senp3 | SENP3 | 3.02E-03 | -3.65E-03 |
| 12077 | Lep | LEP | 6.38E-02 | 1.57E-01 |
| 12078 | Crebzf | CREBZF | 3.86E-03 | 7.45E-03 |
| 12079 | Dnajc9 | DNAJC9 | 4.73E-03 | 4.96E-03 |
| 12080 | Isoc2a | ISOC2 | -8.44E-03 | -1.91E-02 |
| 12081 | Prdm2 | PRDM2 | -4.11E-03 | -1.09E-02 |

| | | | | |
|---|---|---|---|---|
| 12082 | Cidec | CIDEC | -5.93E-02 | 7.48E-02 |
| 12083 | Pqbp1 | PQBP1 | -4.05E-03 | 2.78E-03 |
| 12084 | Rnf20 | RNF20 | -3.29E-03 | -3.63E-03 |
| 12085 | Ccnj | CCNJ | -9.97E-03 | 1.40E-02 |
| 12086 | Ubl7 | UBL7 | 3.96E-03 | -1.13E-02 |
| 12087 | Polr2b | POLR2B | -4.91E-03 | -1.27E-02 |
| 12088 | Pef1 | PEF1 | 4.45E-03 | -2.23E-02 |
| 12089 | Med23 | MED23 | -3.64E-03 | -1.80E-02 |
| 12090 | Vcan | VCAN | 5.60E-02 | 7.31E-02 |
| 12091 | B9d1 | B9D1 | 5.77E-03 | 8.16E-03 |
| 12092 | Slc17a7 | SLC17A7 | -6.17E-03 | 1.15E-02 |
| 12093 | Rngtt | RNGTT | 4.94E-03 | -2.19E-03 |
| 12094 | Gstz1 | GSTZ1 | -5.83E-03 | -1.50E-02 |
| 12095 | Orai3 | ORAI3 | -4.09E-03 | 1.11E-02 |
| 12096 | Lrrc1 | LRRC1 | -4.83E-03 | -3.28E-02 |
| 12097 | Rasgrf1 | RASGRF1 | -6.33E-03 | 4.08E-02 |
| 12098 | Camk2n1 | CAMK2N1 | 4.70E-03 | -2.01E-02 |
| 12099 | Rims1 | RIMS1 | 7.59E-03 | 6.39E-03 |
| 12100 | Mthfd1 | MTHFD1 | -5.72E-03 | -2.72E-02 |
| 12101 | Pkp2 | PKP2 | 3.64E-03 | 2.27E-02 |
| 12102 | Alcam | ALCAM | 9.55E-03 | 1.82E-02 |
| 12103 | Slc39a4 | SLC39A4 | 4.69E-02 | 8.92E-02 |
| 12104 | Ndrg4 | NDRG4 | -5.16E-02 | -3.10E-02 |
| 12105 | Fto | FTO | 3.52E-03 | -1.14E-02 |
| 12106 | Tepp | TEPP | -1.02E-02 | 1.83E-02 |
| 12107 | Ap3b1 | AP3B1 | 5.24E-03 | 7.70E-04 |
| 12108 | Rnf25 | RNF25 | -3.99E-03 | -9.61E-03 |
| 12109 | Mrpl22 | MRPL22 | 4.32E-03 | -1.14E-02 |
| 12110 | Dnajc14 | DNAJC14 | 2.38E-03 | -1.43E-02 |
| 12111 | Osbpl5 | OSBPL5 | 6.54E-03 | -3.19E-02 |
| 12112 | Crybb3 | CRYBB3 | -1.34E-02 | 3.36E-02 |
| 12113 | Tbc1d2 | TBC1D2 | 1.31E-02 | 4.00E-02 |
| 12114 | Gramd2 | GRAMD2 | -1.83E-02 | 3.62E-02 |
| 12115 | Lrrc57 | LRRC57 | 4.88E-03 | -1.72E-02 |
| 12116 | Mpl | MPL | -4.08E-03 | 9.94E-03 |
| 12117 | Eme1 | EME1 | -2.34E-02 | 1.47E-01 |
| 12118 | Asic1 | ASIC1 | -1.24E-02 | -2.32E-02 |
| 12119 | S100a16 | S100A16 | -4.39E-03 | -3.77E-03 |
| 12120 | Cox7a1 | COX7A1 | -8.22E-03 | -2.48E-02 |
| 12121 | Agrp | AGRP | 7.39E-03 | 9.91E-03 |
| 12122 | Nlrp1b | NLRP1 | 1.16E-02 | 6.52E-03 |
| 12123 | Cnksr2 | CNKSR2 | 1.29E-02 | -2.52E-02 |
| 12124 | Cilp2 | CILP2 | 1.01E-02 | 6.59E-02 |
| 12125 | Chmp4c | CHMP4C | -7.54E-03 | -2.27E-02 |
| 12126 | Ttll12 | TTLL12 | -5.49E-03 | 3.33E-03 |
| 12127 | Aldh3a1 | ALDH3A1 | 5.76E-02 | 1.80E-01 |
| 12128 | Zc3h7a | ZC3H7A | 3.41E-03 | 1.09E-02 |
| 12129 | 5031439G07Rik | KIAA0930 | -5.12E-03 | -9.08E-03 |
| 12130 | Optc | OPTC | 9.96E-03 | 5.23E-03 |
| 12131 | Pex26 | PEX26 | -4.80E-03 | -1.17E-02 |
| 12132 | Fam53c | FAM53C | -3.45E-03 | 4.09E-03 |
| 12133 | Ilvbl | ILVBL | 3.03E-03 | -1.01E-03 |
| 12134 | Zfyve9 | ZFYVE9 | 3.58E-03 | -1.62E-02 |

| | | | | |
|---|---|---|---|---|
| 12135 | Hcls1 | HCLS1 | -1.21E-02 | 7.49E-02 |
| 12136 | Gadd45gip1 | GADD45GIP1 | -4.13E-03 | -1.38E-02 |
| 12137 | Def6 | DEF6 | 8.98E-03 | 5.31E-02 |
| 12138 | Acp2 | ACP2 | -3.21E-03 | -1.04E-02 |
| 12139 | Uba5 | UBA5 | 5.33E-03 | -1.44E-02 |
| 12140 | Krt25 | KRT25 | 3.67E-03 | 6.45E-03 |
| 12141 | Mycn | MYCN | -1.36E-02 | 2.29E-03 |
| 12142 | Hmgb2 | HMGB2 | -9.75E-03 | 3.96E-02 |
| 12143 | Cd300e | CD300E | -2.30E-02 | 4.19E-02 |
| 12144 | Trpv4 | TRPV4 | -5.35E-03 | -2.61E-03 |
| 12145 | Bhlhe22 | BHLHE22 | -1.42E-02 | -3.62E-02 |
| 12146 | Ankrd39 | ANKRD39 | 7.15E-03 | -3.63E-04 |
| 12147 | Pkn2 | PKN2 | -3.37E-03 | -6.93E-03 |
| 12148 | Tdrd5 | TDRD5 | -1.37E-02 | -6.70E-02 |
| 12149 | Cdk10 | CDK10 | -3.51E-03 | -3.86E-03 |
| 12150 | Rassf3 | RASSF3 | 6.17E-03 | -6.08E-03 |
| 12151 | Adam19 | ADAM19 | -1.53E-02 | 7.79E-02 |
| 12152 | Idh3b | IDH3B | 4.31E-03 | -2.19E-02 |
| 12153 | Rtkn2 | RTKN2 | -7.52E-03 | 4.14E-02 |
| 12154 | Tmem258 | TMEM258 | -4.84E-03 | 3.10E-03 |
| 12155 | Fam131a | FAM131A | 1.43E-02 | 2.57E-02 |
| 12156 | Rpl39 | RPL39 | 4.61E-03 | 2.78E-02 |
| 12157 | Fam72a | FAM72A | 1.40E-02 | -7.69E-03 |
| 12158 | Osbpl7 | OSBPL7 | 4.47E-03 | -1.14E-02 |
| 12159 | Rarres1 | RARRES1 | -1.63E-02 | 1.17E-02 |
| 12160 | D630045J12Rik | KIAA1549 | 8.52E-03 | 6.85E-03 |
| 12161 | Tubgcp5 | TUBGCP5 | -5.85E-03 | 3.10E-03 |
| 12162 | Rev1 | REV1 | -4.76E-03 | 6.76E-03 |
| 12163 | Hs2st1 | HS2ST1 | -3.84E-03 | 1.07E-02 |
| 12164 | Taf5 | TAF5 | -1.05E-02 | -2.20E-02 |
| 12165 | Tubb1 | TUBB1 | 6.61E-03 | 2.56E-02 |
| 12166 | Dmkn | DMKN | 3.17E-02 | 2.36E-02 |
| 12167 | Fcho2 | FCHO2 | 3.73E-03 | -1.49E-02 |
| 12168 | Erc2 | ERC2 | 1.09E-02 | -8.81E-02 |
| 12169 | Mybl1 | MYBL1 | 1.87E-02 | 1.22E-01 |
| 12170 | Pkd1l1 | PKD1L1 | -9.14E-03 | 6.04E-03 |
| 12171 | Agap3 | AGAP3 | 4.04E-03 | -1.03E-02 |
| 12172 | Wls | WLS | -2.84E-03 | -7.25E-03 |
| 12173 | Gnat1 | GNAT1 | 2.32E-02 | 4.38E-02 |
| 12174 | Inhbe | INHBE | -4.21E-03 | 2.14E-03 |
| 12175 | Mlycd | MLYCD | -5.75E-03 | -1.03E-02 |
| 12176 | Gsg1l | GSG1L | -1.95E-02 | -7.35E-02 |
| 12177 | Tmem59 | TMEM59 | 3.21E-03 | -1.02E-02 |
| 12178 | Kirrel | KIRREL | 9.34E-03 | -7.44E-03 |
| 12179 | Ids | IDS | -4.97E-03 | -1.73E-02 |
| 12180 | Amdhd2 | AMDHD2 | 7.38E-03 | -1.64E-02 |
| 12181 | Iqcb1 | IQCB1 | 7.06E-03 | -2.37E-02 |
| 12182 | Mmp14 | MMP14 | 1.26E-02 | 1.05E-01 |
| 12183 | Dennd5b | DENND5B | -5.84E-03 | -4.30E-03 |
| 12184 | Psmg3 | PSMG3 | -8.92E-03 | 2.42E-02 |
| 12185 | Zfp282 | ZNF282 | -5.53E-03 | -1.00E-02 |
| 12186 | Cbx5 | CBX5 | 3.39E-03 | 2.68E-03 |
| 12187 | Fam83a | FAM83A | -1.99E-02 | 6.84E-02 |

| | | | | |
|---|---|---|---|---|
| 12188 | Ass1 | ASS1 | -7.66E-03 | -3.36E-03 |
| 12189 | Cldn1 | CLDN1 | 1.14E-02 | 3.72E-02 |
| 12190 | Hhipl2 | HHIPL2 | -9.57E-03 | 1.32E-02 |
| 12191 | Foxc1 | FOXC1 | 2.55E-02 | -5.34E-02 |
| 12192 | Ubash3b | UBASH3B | -1.69E-02 | -1.43E-02 |
| 12193 | Ttbk2 | TTBK2 | -5.41E-03 | -7.02E-04 |
| 12194 | Fbln2 | FBLN2 | 1.22E-02 | 1.06E-01 |
| 12195 | Tmem86a | TMEM86A | 1.45E-02 | -1.24E-01 |
| 12196 | Dna2 | DNA2 | -8.56E-03 | 2.13E-02 |
| 12197 | Cck | CCK | 7.24E-02 | -1.33E-01 |
| 12198 | Polr2l | POLR2L | -4.33E-03 | 3.03E-03 |
| 12199 | Rin2 | RIN2 | -4.50E-03 | -1.75E-02 |
| 12200 | C1rl | C1RL | 9.06E-03 | -4.66E-02 |
| 12201 | Retn | RETN | -3.73E-02 | 9.78E-02 |
| 12202 | Krt80 | KRT80 | -1.78E-02 | 3.66E-02 |
| 12203 | Cdkn2a | CDKN2A | 6.15E-03 | 3.11E-02 |
| 12204 | Pstpip2 | PSTPIP2 | 8.61E-03 | -1.88E-02 |
| 12205 | Ssr2 | SSR2 | 3.84E-03 | 5.27E-03 |
| 12206 | Prok2 | PROK2 | -2.02E-02 | 1.37E-02 |
| 12207 | Cdc25b | CDC25B | 1.05E-02 | 6.02E-02 |
| 12208 | Hexim1 | HEXIM1 | -4.68E-03 | 2.33E-02 |
| 12209 | Prr11 | PRR11 | 2.91E-02 | 1.97E-01 |
| 12210 | Nhs | NHS | -1.54E-02 | -4.78E-02 |
| 12211 | Tmem177 | TMEM177 | -5.42E-03 | -2.82E-02 |
| 12212 | Cnr1 | CNR1 | -1.02E-02 | 1.83E-02 |
| 12213 | Exoc6 | EXOC6 | -5.96E-03 | -1.16E-02 |
| 12214 | Cxxc5 | CXXC5 | 4.31E-03 | 3.62E-03 |
| 12215 | Cmc2 | CMC2 | -6.08E-03 | 1.54E-02 |
| 12216 | Nif3l1 | NIF3L1 | 5.81E-03 | -2.43E-02 |
| 12217 | Slc25a18 | SLC25A18 | -5.33E-03 | 2.07E-02 |
| 12218 | Cfd | CFD | -9.31E-02 | 1.01E-01 |
| 12219 | Pcdhb2 | PCDHB2 | -4.84E-03 | 1.90E-02 |
| 12220 | Eml5 | EML5 | 7.17E-03 | -3.78E-02 |
| 12221 | Arsj | ARSJ | -8.82E-03 | 7.05E-03 |
| 12222 | Npff | NPFF | -9.25E-03 | 5.58E-02 |
| 12223 | Inip | INIP | 7.57E-03 | 3.52E-04 |
| 12224 | Rbmx2 | RBMX2 | 9.30E-03 | 1.03E-02 |
| 12225 | Fkbpl | FKBPL | 6.55E-03 | -1.25E-02 |
| 12226 | Zdhhc2 | ZDHHC2 | -1.38E-02 | 2.67E-02 |
| 12227 | Plxna2 | PLXNA2 | 6.81E-03 | -3.52E-03 |
| 12228 | Ptprm | PTPRM | 7.48E-03 | -4.85E-02 |
| 12229 | Rdx | RDX | 3.59E-03 | -1.08E-02 |
| 12230 | Cyp3a13 | CYP3A7 | -1.54E-02 | 1.69E-01 |
| 12231 | Usp25 | USP25 | 2.88E-03 | -8.82E-03 |
| 12232 | Efemp2 | EFEMP2 | -1.14E-02 | 9.44E-02 |
| 12233 | Cst8 | CST8 | -4.90E-03 | 1.22E-02 |
| 12234 | Mtfmt | MTFMT | -3.31E-03 | 7.51E-03 |
| 12235 | Hadha | HADHA | 4.40E-03 | -2.87E-02 |
| 12236 | Paip2 | PAIP2 | -3.55E-03 | -4.75E-03 |
| 12237 | Lin52 | LIN52 | -4.91E-03 | -3.29E-02 |
| 12238 | Rgl1 | RGL1 | -8.24E-03 | -2.64E-02 |
| 12239 | Dnm3 | DNM3 | 2.06E-02 | -3.50E-02 |
| 12240 | Cmip | CMIP | 8.42E-03 | -2.13E-02 |

| | | | | |
|---|---|---|---|---|
| 12241 | Crebrf | CREBRF | -8.06E-03 | -4.29E-02 |
| 12242 | Stap2 | STAP2 | 1.62E-02 | 2.40E-02 |
| 12243 | Tmem60 | TMEM60 | 5.47E-03 | -4.25E-04 |
| 12244 | Rpain | RPAIN | -5.05E-03 | -3.30E-02 |
| 12245 | C9 | C9 | -8.27E-03 | 3.36E-02 |
| 12246 | Tgfb1 | TGFB1 | -1.08E-02 | 7.92E-02 |
| 12247 | Mpp1 | MPP1 | 7.98E-03 | 1.77E-02 |
| 12248 | Irak2 | IRAK2 | 5.81E-03 | -2.29E-02 |
| 12249 | Cntn1 | CNTN1 | -3.15E-02 | -7.07E-02 |
| 12250 | Cdc42ep5 | CDC42EP5 | 9.09E-03 | 4.77E-02 |
| 12251 | Scrn1 | SCRN1 | -1.88E-02 | -8.67E-02 |
| 12252 | Rnf180 | RNF180 | 2.22E-02 | 8.93E-02 |
| 12253 | Scel | SCEL | 4.81E-02 | -1.33E-01 |
| 12254 | Zfp51 | ZNF761 | -7.02E-03 | 6.97E-05 |
| 12255 | Fam160b2 | FAM160B2 | 4.20E-03 | 3.98E-04 |
| 12256 | Ush2a | USH2A | -4.23E-03 | -9.15E-04 |
| 12257 | Phb2 | PHB2 | 3.92E-03 | -5.02E-03 |
| 12258 | Spats1 | SPATS1 | -1.43E-02 | -1.53E-02 |
| 12259 | Tfcp2 | TFCP2 | 5.51E-03 | -3.13E-02 |
| 12260 | Ift43 | IFT43 | 5.16E-03 | -2.67E-03 |
| 12261 | Tfam | TFAM | 3.23E-03 | -1.43E-02 |
| 12262 | Armc2 | ARMC2 | -1.56E-02 | 9.14E-03 |
| 12263 | Wnt2b | WNT2B | -8.89E-03 | 6.41E-02 |
| 12264 | Ppl | PPL | 2.98E-02 | 2.93E-02 |
| 12265 | Ift88 | IFT88 | 4.31E-03 | -4.49E-03 |
| 12266 | Ascl4 | ASCL4 | -1.17E-02 | 3.62E-02 |
| 12267 | Wdr37 | WDR37 | -3.78E-03 | -1.65E-03 |
| 12268 | Slc39a11 | SLC39A11 | 5.34E-03 | -2.76E-02 |
| 12269 | Trnau1ap | TRNAU1AP | -4.28E-03 | -1.67E-02 |
| 12270 | Olfml2b | OLFML2B | 1.54E-02 | 4.75E-02 |
| 12271 | Mat2a | MAT2A | 6.01E-03 | -2.15E-03 |
| 12272 | Wfs1 | WFS1 | 1.42E-02 | 2.54E-02 |
| 12273 | Avl9 | AVL9 | 4.32E-03 | -1.25E-02 |
| 12274 | Ocstamp | OCSTAMP | -6.06E-03 | 1.92E-02 |
| 12275 | Enc1 | ENC1 | 9.21E-03 | 6.23E-02 |
| 12276 | Mgat4c | MGAT4C | 1.32E-02 | -2.16E-02 |
| 12277 | Arrdc1 | ARRDC1 | 7.69E-03 | -2.21E-02 |
| 12278 | Pard6g | PARD6G | 1.46E-02 | -2.91E-02 |
| 12279 | Dnmt3a | DNMT3A | 3.70E-03 | 1.49E-03 |
| 12280 | Fzd10 | FZD10 | 7.85E-03 | -1.20E-02 |
| 12281 | Zfyve26 | ZFYVE26 | -3.99E-03 | -2.27E-02 |
| 12282 | Rnf139 | RNF139 | -3.22E-03 | 1.09E-02 |
| 12283 | Slc6a2 | SLC6A2 | 1.06E-02 | 3.14E-02 |
| 12284 | Rars | RARS | 2.94E-03 | 4.29E-03 |
| 12285 | Fam98a | FAM98A | 5.56E-03 | 1.05E-02 |
| 12286 | Hus1 | HUS1 | -4.53E-03 | -2.40E-02 |
| 12287 | Ccdc130 | CCDC130 | -6.41E-03 | -1.95E-03 |
| 12288 | Txn1 | TXN | -4.46E-03 | 1.14E-02 |
| 12289 | Drc1 | DRC1 | -7.97E-03 | -2.43E-02 |
| 12290 | Epo | EPO | -3.11E-02 | 1.67E-01 |
| 12291 | Spint2 | SPINT2 | 4.65E-03 | 8.84E-03 |
| 12292 | Tmx3 | TMX3 | -3.86E-03 | 1.19E-02 |
| 12293 | Cog3 | COG3 | 2.82E-03 | -7.34E-03 |

| | | | | |
|---|---|---|---|---|
| 12294 | Tonsl | TONSL | 2.50E-03 | -3.79E-03 |
| 12295 | Faf2 | FAF2 | 2.67E-03 | -2.72E-03 |
| 12296 | Dpp9 | DPP9 | -2.96E-03 | -1.37E-02 |
| 12297 | Mrps36 | MRPS36 | 4.59E-03 | -8.09E-03 |
| 12298 | Cdc14b | CDC14B | 4.61E-03 | -2.50E-02 |
| 12299 | Chd8 | CHD8 | 2.28E-03 | -1.16E-02 |
| 12300 | Vhl | VHL | -4.60E-03 | 1.29E-02 |
| 12301 | Olr1 | OLR1 | 8.49E-03 | 6.23E-02 |
| 12302 | Fcgrt | FCGRT | -6.55E-03 | 1.76E-03 |
| 12303 | Mmp27 | MMP27 | 6.22E-03 | 3.91E-02 |
| 12304 | Ttc16 | TTC16 | 1.34E-02 | -1.91E-02 |
| 12305 | Ifitm5 | IFITM5 | 6.99E-03 | 8.97E-03 |
| 12306 | Zfp212 | ZNF212 | 4.35E-03 | -2.26E-02 |
| 12307 | Lrif1 | LRIF1 | 6.14E-03 | -4.19E-03 |
| 12308 | Galnt5 | GALNT5 | -6.04E-03 | 3.54E-02 |
| 12309 | Ntm | NTM | 1.41E-02 | 1.17E-01 |
| 12310 | Ap4e1 | AP4E1 | 6.49E-03 | -2.47E-02 |
| 12311 | Alg8 | ALG8 | -6.32E-03 | 3.54E-02 |
| 12312 | Tars2 | TARS2 | 3.31E-03 | -1.73E-02 |
| 12313 | Hps5 | HPS5 | -4.17E-03 | -2.22E-02 |
| 12314 | Afap1 | AFAP1 | 5.80E-03 | -3.55E-02 |
| 12315 | Hps6 | HPS6 | -4.99E-03 | 8.71E-03 |
| 12316 | Itga8 | ITGA8 | 1.07E-02 | -7.27E-03 |
| 12317 | Lrrc56 | LRRC56 | -5.73E-03 | -2.48E-02 |
| 12318 | Smad5 | SMAD5 | -3.45E-03 | -8.20E-03 |
| 12319 | Amd2 | AMD1 | 1.42E-02 | 2.03E-02 |
| 12320 | Ptgds | PTGDS | -1.83E-02 | -5.35E-02 |
| 12321 | Sord | SORD | 7.43E-03 | -1.74E-02 |
| 12322 | Dnaaf2 | DNAAF2 | -9.96E-03 | 3.40E-02 |
| 12323 | Neil2 | NEIL2 | -1.12E-02 | -1.09E-01 |
| 12324 | Sorl1 | SORL1 | 5.24E-03 | -4.00E-02 |
| 12325 | Rrm2b | RRM2B | 3.41E-03 | -2.09E-02 |
| 12326 | Rasa1 | RASA1 | -3.15E-03 | 2.12E-03 |
| 12327 | Taf8 | TAF8 | 3.80E-03 | -2.73E-02 |
| 12328 | Rpl6 | RPL6 | -3.51E-03 | 1.01E-02 |
| 12329 | Bmp7 | BMP7 | 5.06E-03 | -4.11E-02 |
| 12330 | Ascl2 | ASCL2 | 6.69E-03 | 1.40E-02 |
| 12331 | Cir1 | CIR1 | 5.03E-03 | -2.64E-02 |
| 12332 | Tiprl | TIPRL | -2.99E-03 | -4.40E-03 |
| 12333 | Mrps9 | MRPS9 | -3.31E-03 | -1.91E-02 |
| 12334 | Ube2v1 | UBE2V1 | 1.00E-02 | -1.93E-02 |
| 12335 | Samd11 | SAMD11 | -4.63E-03 | 1.71E-03 |
| 12336 | Pus7l | PUS7L | 8.95E-03 | 2.99E-02 |
| 12337 | Tgm5 | TGM5 | 3.71E-03 | -1.14E-02 |
| 12338 | Tmem154 | TMEM154 | -1.63E-02 | 9.70E-02 |
| 12339 | Dffb | DFFB | -6.03E-03 | -3.92E-04 |
| 12340 | Gtpbp10 | GTPBP10 | 4.12E-03 | -2.68E-02 |
| 12341 | Lin37 | LIN37 | -5.53E-03 | 4.67E-03 |
| 12342 | Ppp1ca | PPP1CA | -9.63E-03 | 2.00E-02 |
| 12343 | Blnk | BLNK | -7.29E-03 | 3.36E-02 |
| 12344 | Fam124a | FAM124A | -1.28E-02 | 8.75E-02 |
| 12345 | Trim71 | TRIM71 | -6.49E-03 | 2.69E-02 |
| 12346 | Npr1 | NPR1 | -6.91E-03 | -2.44E-02 |

| | | | | |
|---|---|---|---|---|
| 12347 | Fxyd7 | FXYD7 | 7.48E-03 | 2.54E-03 |
| 12348 | Dixdc1 | DIXDC1 | 1.08E-02 | -1.06E-01 |
| 12349 | Gart | GART | -3.19E-03 | -2.64E-03 |
| 12350 | Parg | PARG | 3.86E-03 | 1.77E-03 |
| 12351 | Gtf3c2 | GTF3C2 | 3.62E-03 | -5.61E-03 |
| 12352 | Lxn | LXN | 9.74E-03 | 7.07E-02 |
| 12353 | Cpb2 | CPB2 | 1.51E-02 | -7.75E-02 |
| 12354 | Plet1 | PLET1 | 2.77E-02 | 7.85E-02 |
| 12355 | Raph1 | RAPH1 | -6.77E-03 | 2.73E-02 |
| 12356 | Il1a | IL1A | 4.80E-03 | 8.83E-03 |
| 12357 | Tuba8 | TUBA8 | 2.30E-02 | -5.76E-02 |
| 12358 | Lamtor5 | LAMTOR5 | -4.46E-03 | -1.31E-02 |
| 12359 | Tubb3 | TUBB3 | -1.06E-02 | 6.22E-02 |
| 12360 | Mrps15 | MRPS15 | -3.38E-03 | -9.31E-03 |
| 12361 | Tmem243 | TMEM243 | 4.06E-03 | -1.81E-02 |
| 12362 | Aoah | AOAH | 9.65E-03 | 1.84E-03 |
| 12363 | Fgf15 | FGF19 | 5.20E-03 | 1.09E-02 |
| 12364 | Tigd2 | TIGD2 | 4.47E-03 | -5.11E-03 |
| 12365 | Vtn | VTN | -2.15E-02 | 7.21E-02 |
| 12366 | Upk3a | UPK3A | 7.87E-02 | 1.72E-01 |
| 12367 | Cuta | CUTA | 3.54E-03 | 4.15E-03 |
| 12368 | Rps12 | RPS12 | -3.49E-03 | 1.09E-02 |
| 12369 | Tbck | TBCK | 3.89E-03 | -4.36E-03 |
| 12370 | Dio2 | DIO2 | 1.62E-02 | 4.12E-02 |
| 12371 | Safb | SAFB | -2.51E-03 | -4.46E-03 |
| 12372 | Ube2c | UBE2C | -2.71E-02 | 9.96E-02 |
| 12373 | Rpl18 | RPL18 | -3.42E-03 | 1.65E-02 |
| 12374 | Sh3d19 | SH3D19 | 4.69E-03 | 8.77E-03 |
| 12375 | Med29 | MED29 | -4.11E-03 | -8.05E-03 |
| 12376 | Prrg2 | PRRG2 | 3.38E-03 | 2.71E-03 |
| 12377 | Spa17 | SPA17 | -1.24E-02 | -4.35E-02 |
| 12378 | Arl2 | ARL2 | 2.88E-03 | -2.01E-02 |
| 12379 | Uqcrc1 | UQCRC1 | 5.27E-03 | -1.72E-02 |
| 12380 | Clpb | CLPB | 3.98E-03 | -2.66E-02 |
| 12381 | Colec11 | COLEC11 | 1.15E-02 | 2.97E-02 |
| 12382 | Pcdhb10 | PCDHB5 | -8.57E-03 | 3.17E-02 |
| 12383 | Ciart | CIART | 2.71E-02 | -2.25E-01 |
| 12384 | Slc1a1 | SLC1A1 | 5.08E-03 | -3.85E-02 |
| 12385 | Jsrp1 | JSRP1 | -3.32E-03 | 2.13E-02 |
| 12386 | Slc16a7 | SLC16A7 | 6.83E-03 | -2.75E-02 |
| 12387 | Rnf208 | RNF208 | -6.22E-03 | -2.64E-02 |
| 12388 | E2f8 | E2F8 | 2.58E-02 | 2.27E-01 |
| 12389 | Snx30 | SNX30 | -7.96E-03 | -3.38E-03 |
| 12390 | Rhpn2 | RHPN2 | -1.26E-02 | -2.37E-02 |
| 12391 | Atn1 | ATN1 | 7.20E-03 | 1.40E-02 |
| 12392 | Tmem50b | TMEM50B | -5.49E-03 | 2.52E-03 |
| 12393 | Fbxo5 | FBXO5 | -1.46E-02 | 2.09E-02 |
| 12394 | Eif3h | EIF3H | -3.05E-03 | 1.02E-02 |
| 12395 | Serf1 | SERF1A | -7.13E-03 | -1.47E-03 |
| 12396 | Serf1 | SERF1B | -7.13E-03 | -1.47E-03 |
| 12397 | Meig1 | MEIG1 | -3.55E-03 | 3.38E-02 |
| 12398 | Esrrg | ESRRG | 5.06E-03 | -3.90E-02 |
| 12399 | Ripk2 | RIPK2 | -7.32E-03 | 2.16E-02 |

| | | | | |
|---|---|---|---|---|
| 12400 | Cul9 | CUL9 | 3.38E-03 | -2.75E-02 |
| 12401 | Nfu1 | NFU1 | 2.77E-03 | -2.56E-02 |
| 12402 | Emc1 | EMC1 | 3.26E-03 | -9.99E-03 |
| 12403 | Lgals3bp | LGALS3BP | -8.07E-03 | 5.89E-02 |
| 12404 | Asic3 | ASIC3 | 7.65E-03 | -1.24E-02 |
| 12405 | Smc2 | SMC2 | 1.26E-02 | 1.00E-01 |
| 12406 | Vipas39 | VIPAS39 | -2.57E-03 | -2.66E-03 |
| 12407 | Krt15 | KRT15 | 7.47E-02 | 1.96E-01 |
| 12408 | Cdk14 | CDK14 | -1.12E-02 | -1.61E-02 |
| 12409 | Cnnm4 | CNNM4 | -5.14E-03 | 9.19E-03 |
| 12410 | Slc25a43 | SLC25A43 | -1.40E-02 | 6.32E-03 |
| 12411 | Rnf13 | RNF13 | 3.60E-03 | -1.83E-02 |
| 12412 | Casp3 | CASP3 | 4.03E-03 | -7.19E-03 |
| 12413 | Rpl37a | RPL37A | -3.13E-03 | 2.10E-03 |
| 12414 | Ranbp6 | RANBP6 | 3.99E-03 | 1.28E-02 |
| 12415 | Foxn2 | FOXN2 | 6.69E-03 | 2.45E-02 |
| 12416 | Tsr3 | TSR3 | 3.84E-03 | 2.75E-03 |
| 12417 | Rab22a | RAB22A | 2.54E-03 | -3.61E-03 |
| 12418 | Dnase1 | DNASE1 | 1.28E-02 | -6.72E-02 |
| 12419 | Tecpr1 | TECPR1 | 2.98E-03 | -2.02E-02 |
| 12420 | Tmem79 | TMEM79 | -7.36E-03 | 1.70E-02 |
| 12421 | Wdr89 | WDR89 | 8.33E-03 | 9.20E-03 |
| 12422 | Kif17 | KIF17 | -1.15E-02 | -1.59E-02 |
| 12423 | Plekha3 | PLEKHA3 | 3.37E-03 | 5.98E-03 |
| 12424 | Ttc7b | TTC7B | 4.62E-03 | -1.18E-02 |
| 12425 | Bnip1 | BNIP1 | -4.33E-03 | -1.08E-02 |
| 12426 | Pcdh17 | PCDH17 | -9.96E-03 | -3.98E-02 |
| 12427 | Gmpr | GMPR | -4.52E-03 | -1.02E-02 |
| 12428 | Enox1 | ENOX1 | 1.32E-02 | -3.82E-02 |
| 12429 | Rfk | RFK | 2.84E-03 | -1.74E-02 |
| 12430 | Ssr1 | SSR1 | -2.19E-03 | -9.56E-03 |
| 12431 | Fdx1 | FDX1 | -3.62E-03 | -5.51E-03 |
| 12432 | Mfrp | MFRP | 4.06E-02 | -1.39E-01 |
| 12433 | Rbm47 | RBM47 | -3.82E-03 | -1.39E-02 |
| 12434 | Cldn14 | CLDN14 | 9.02E-03 | 5.77E-02 |
| 12435 | Tert | TERT | -3.79E-02 | -2.24E-04 |
| 12436 | Pde8b | PDE8B | 1.06E-02 | 1.29E-02 |
| 12437 | Mgat4b | MGAT4B | 3.31E-03 | -1.76E-04 |
| 12438 | Rps24 | RPS24 | -2.93E-03 | 6.57E-03 |
| 12439 | Nelfa | NELFA | -4.15E-03 | 1.49E-02 |
| 12440 | Alpl | ALPL | -6.91E-03 | 9.10E-03 |
| 12441 | Thpo | THPO | -6.84E-03 | 1.33E-02 |
| 12442 | Zfp46 | ZNF436 | -8.29E-03 | 1.66E-04 |
| 12443 | Tmbim6 | TMBIM6 | 3.38E-03 | -7.38E-03 |
| 12444 | Npas3 | NPAS3 | 5.78E-03 | -1.36E-02 |
| 12445 | Mamstr | MAMSTR | -1.31E-02 | 3.29E-02 |
| 12446 | Mapk11 | MAPK11 | 1.55E-02 | 3.22E-02 |
| 12447 | Rab17 | RAB17 | 4.31E-03 | -1.16E-02 |
| 12448 | Gabrq | GABRQ | 5.28E-03 | -9.31E-03 |
| 12449 | Nadk | NADK | 3.04E-03 | 3.70E-03 |
| 12450 | Isca1 | ISCA1 | 2.63E-03 | -1.37E-02 |
| 12451 | Acadl | ACADL | 3.75E-03 | -1.42E-02 |
| 12452 | Rars2 | RARS2 | 3.74E-03 | -2.31E-02 |

| | | | | |
|---|---|---|---|---|
| 12453 | Plekho1 | PLEKHO1 | 7.01E-03 | 7.56E-02 |
| 12454 | Col19a1 | COL19A1 | 7.09E-03 | -3.13E-05 |
| 12455 | Lag3 | LAG3 | -1.16E-02 | 2.97E-02 |
| 12456 | Rs1 | RS1 | -3.33E-03 | 1.50E-02 |
| 12457 | Agtpbp1 | AGTPBP1 | -5.02E-03 | -1.76E-02 |
| 12458 | Eed | EED | -3.27E-03 | 4.49E-03 |
| 12459 | Pmel | PMEL | -9.57E-03 | 7.30E-03 |
| 12460 | Plgrkt | PLGRKT | -3.34E-03 | 2.63E-03 |
| 12461 | Scn1b | SCN1B | -7.87E-03 | 7.56E-02 |
| 12462 | Srsf10 | SRSF10 | -4.24E-03 | 1.35E-02 |
| 12463 | Tlcd1 | TLCD1 | 2.87E-03 | -1.22E-02 |
| 12464 | Rpl24 | RPL24 | 6.45E-03 | 4.00E-02 |
| 12465 | Slc39a12 | SLC39A12 | -5.23E-03 | -1.31E-02 |
| 12466 | Il1rapl1 | IL1RAPL1 | -3.12E-03 | -2.90E-03 |
| 12467 | Cdc42ep1 | CDC42EP1 | 5.49E-03 | 1.11E-02 |
| 12468 | Metrn | METRN | -5.60E-03 | 2.38E-03 |
| 12469 | Xrcc4 | XRCC4 | -4.35E-03 | -5.72E-03 |
| 12470 | Mvp | MVP | -3.63E-03 | 2.12E-02 |
| 12471 | Otop3 | OTOP3 | -6.42E-03 | -2.54E-04 |
| 12472 | Tmem68 | TMEM68 | 4.85E-03 | 1.30E-02 |
| 12473 | Trim33 | TRIM33 | -3.05E-03 | -5.48E-04 |
| 12474 | Clrn3 | CLRN3 | -4.50E-03 | 7.77E-03 |
| 12475 | Kcp | KCP | 6.90E-03 | -2.08E-02 |
| 12476 | Gpatch2l | GPATCH2L | -3.17E-03 | -1.55E-02 |
| 12477 | Slc1a2 | SLC1A2 | -9.91E-03 | 3.03E-02 |
| 12478 | Polk | POLK | 4.33E-03 | -1.62E-02 |
| 12479 | Phex | PHEX | 4.64E-03 | 2.25E-02 |
| 12480 | Mgat1 | MGAT1 | -3.83E-03 | -1.41E-02 |
| 12481 | Mctp1 | MCTP1 | 1.43E-02 | 2.37E-02 |
| 12482 | Coq10b | COQ10B | -7.92E-03 | 7.22E-03 |
| 12483 | Ros1 | ROS1 | 4.88E-02 | 3.17E-02 |
| 12484 | Bcas3 | BCAS3 | -3.62E-03 | -1.59E-02 |
| 12485 | Tsfm | TSFM | 4.39E-03 | -8.95E-03 |
| 12486 | Cox6b2 | COX6B2 | 1.10E-02 | 3.94E-02 |
| 12487 | Tlr6 | TLR6 | 1.13E-02 | 4.52E-02 |
| 12488 | Crtam | CRTAM | 1.35E-02 | -5.65E-02 |
| 12489 | Ipo7 | IPO7 | 3.04E-03 | 1.77E-02 |
| 12490 | Tex10 | TEX10 | 4.56E-03 | -7.73E-03 |
| 12491 | Stradb | STRADB | -2.94E-03 | -1.30E-02 |
| 12492 | Pcbp4 | PCBP4 | -4.58E-03 | -1.97E-02 |
| 12493 | Tmco1 | TMCO1 | -2.93E-03 | -4.35E-03 |
| 12494 | Hivep3 | HIVEP3 | 8.58E-03 | 9.28E-03 |
| 12495 | Eid2 | EID2 | -6.92E-03 | 8.88E-03 |
| 12496 | Klb | KLB | 6.14E-03 | 2.82E-02 |
| 12497 | Lsr | LSR | -3.40E-03 | 2.41E-02 |
| 12498 | Rc3h1 | RC3H1 | 3.59E-03 | -4.70E-03 |
| 12499 | Slc2a7 | SLC2A7 | 7.13E-03 | -3.02E-03 |
| 12500 | Pcdhb19 | PCDHB12 | -8.99E-03 | 2.98E-02 |
| 12501 | Rpl36al | RPL36AL | -7.38E-03 | 6.15E-02 |
| 12502 | Acss1 | ACSS1 | 6.71E-03 | -2.63E-02 |
| 12503 | Heca | HECA | 4.64E-03 | -1.84E-02 |
| 12504 | Shf | SHF | 7.04E-03 | -2.46E-03 |
| 12505 | Fibcd1 | FIBCD1 | -4.63E-03 | -1.25E-03 |

| | | | | |
|---|---|---|---|---|
| 12506 | Cyyr1 | CYYR1 | 1.30E-02 | -2.95E-02 |
| 12507 | Faxc | FAXC | -7.39E-03 | 6.01E-02 |
| 12508 | Gen1 | GEN1 | -1.29E-02 | 6.12E-02 |
| 12509 | Sbk3 | SBK3 | 9.64E-03 | -3.01E-03 |
| 12510 | Clec4e | CLEC4E | 3.66E-02 | 1.30E-01 |
| 12511 | Oasl1 | OASL | -1.16E-02 | 1.33E-01 |
| 12512 | Supv3l1 | SUPV3L1 | 3.29E-03 | -1.07E-02 |
| 12513 | Maf1 | MAF1 | -2.84E-03 | -1.47E-02 |
| 12514 | Trdmt1 | TRDMT1 | -5.53E-03 | -2.30E-02 |
| 12515 | Rab8a | RAB8A | 2.83E-03 | -1.78E-03 |
| 12516 | Iqcd | IQCD | -1.21E-02 | -9.23E-02 |
| 12517 | Cbln1 | CBLN1 | 2.54E-02 | -3.28E-02 |
| 12518 | Npat | NPAT | 5.69E-03 | -1.13E-02 |
| 12519 | Crlf1 | CRLF1 | 3.37E-02 | 3.08E-01 |
| 12520 | Rad54l | RAD54L | -1.70E-02 | 1.81E-01 |
| 12521 | Ptprn | PTPRN | 1.75E-02 | 1.18E-01 |
| 12522 | Ghrhr | GHRHR | 1.81E-02 | -1.30E-01 |
| 12523 | Mycbp2 | MYCBP2 | -2.14E-03 | -1.94E-02 |
| 12524 | Cyth3 | CYTH3 | 3.89E-03 | -2.05E-02 |
| 12525 | Mon1a | MON1A | 3.72E-03 | -5.25E-04 |
| 12526 | Tsr2 | TSR2 | -3.25E-03 | -1.52E-02 |
| 12527 | Hsd3b6 | HSD3B1 | -5.63E-03 | -4.68E-03 |
| 12528 | Enkd1 | ENKD1 | -6.26E-03 | 4.24E-03 |
| 12529 | Llph-ps1 | LLPH | -6.18E-03 | 1.33E-02 |
| 12530 | Gpr37l1 | GPR37L1 | -6.00E-03 | 3.49E-03 |
| 12531 | Gfra4 | GFRA4 | -9.62E-03 | -5.90E-02 |
| 12532 | Kidins220 | KIDINS220 | 3.04E-03 | -2.32E-02 |
| 12533 | Ebpl | EBPL | 4.60E-03 | 1.97E-02 |
| 12534 | Grpr | GRPR | 5.22E-03 | -2.02E-03 |
| 12535 | Fancd2 | FANCD2 | 2.24E-02 | 1.30E-01 |
| 12536 | Gcnt2 | GCNT2 | -4.62E-03 | -1.58E-03 |
| 12537 | Uaca | UACA | 3.96E-03 | -3.49E-02 |
| 12538 | Srsf1 | SRSF1 | 3.06E-03 | 7.37E-03 |
| 12539 | Coil | COIL | 6.65E-03 | 1.65E-02 |
| 12540 | Rpl30 | RPL30 | -2.78E-03 | 2.29E-03 |
| 12541 | Rgs18 | RGS18 | -5.70E-03 | 2.94E-02 |
| 12542 | Sox18 | SOX18 | 1.50E-02 | 7.01E-03 |
| 12543 | Lix1 | LIX1 | -1.47E-02 | 6.15E-02 |
| 12544 | Ankrd17 | ANKRD17 | -3.14E-03 | -2.09E-03 |
| 12545 | Rasl10b | RASL10B | 2.00E-02 | -8.53E-02 |
| 12546 | Pdpr | PDPR | 6.37E-03 | -4.01E-02 |
| 12547 | Tgfbr2 | TGFBR2 | 4.09E-03 | 1.51E-02 |
| 12548 | Tm9sf4 | TM9SF4 | -2.71E-03 | -1.51E-02 |
| 12549 | Myoz2 | MYOZ2 | 5.96E-03 | 2.41E-02 |
| 12550 | Rnf168 | RNF168 | 3.35E-03 | 4.65E-03 |
| 12551 | Limd1 | LIMD1 | -2.35E-03 | 8.89E-04 |
| 12552 | Itgb6 | ITGB6 | 8.68E-03 | -5.54E-02 |
| 12553 | Rnf19b | RNF19B | 4.95E-03 | 1.95E-02 |
| 12554 | Cpne3 | CPNE3 | 3.77E-03 | -1.59E-02 |
| 12555 | Tmem170b | TMEM170B | -4.83E-03 | -5.80E-03 |
| 12556 | Muc5b | MUC5B | -4.56E-03 | 3.32E-02 |
| 12557 | Wdr11 | WDR11 | 3.13E-03 | -1.53E-02 |
| 12558 | Fbxo41 | FBXO41 | 6.19E-03 | 2.16E-02 |

| | | | | |
|---|---|---|---|---|
| 12559 | Ugt2b35 | UGT2B7 | 6.58E-03 | 3.01E-02 |
| 12560 | Cpm | CPM | -9.99E-03 | -5.37E-02 |
| 12561 | Boc | BOC | 1.02E-02 | -2.40E-02 |
| 12562 | Zkscan16 | ZNF483 | -1.14E-02 | -5.59E-03 |
| 12563 | Nptx2 | NPTX2 | 8.42E-03 | 4.03E-02 |
| 12564 | Gria1 | GRIA1 | 4.51E-03 | 2.82E-02 |
| 12565 | Dntt | DNTT | -5.42E-03 | 9.90E-03 |
| 12566 | Nbn | NBN | 3.98E-03 | 9.92E-03 |
| 12567 | Scaf11 | SCAF11 | 2.72E-03 | -1.30E-02 |
| 12568 | Prss22 | PRSS22 | 1.23E-02 | 6.45E-02 |
| 12569 | Ddx59 | DDX59 | -6.01E-03 | 1.30E-02 |
| 12570 | Mmrn2 | MMRN2 | 5.80E-03 | -3.63E-02 |
| 12571 | Irs2 | IRS2 | -1.37E-02 | -2.28E-03 |
| 12572 | Thbs1 | THBS1 | 6.05E-03 | 2.91E-02 |
| 12573 | Bace2 | BACE2 | 1.65E-02 | 3.91E-02 |
| 12574 | Cr2 | CR2 | 9.51E-03 | -1.02E-01 |
| 12575 | Nudt6 | NUDT6 | -5.55E-03 | 2.33E-02 |
| 12576 | Rps29 | RPS29 | 3.16E-03 | -5.57E-03 |
| 12577 | Ddx28 | DDX28 | 5.26E-03 | 1.23E-02 |
| 12578 | Pih1d2 | PIH1D2 | 8.41E-03 | 2.16E-02 |
| 12579 | Polr3g | POLR3G | 5.34E-03 | 6.70E-03 |
| 12580 | Zbtb3 | ZBTB3 | -1.15E-02 | 1.52E-02 |
| 12581 | Hsd17b10 | HSD17B10 | -2.84E-03 | -6.80E-03 |
| 12582 | Homer1 | HOMER1 | -4.82E-03 | 1.59E-02 |
| 12583 | Cela1 | CELA1 | 7.28E-03 | -2.42E-02 |
| 12584 | Rogdi | ROGDI | -3.41E-03 | -1.85E-02 |
| 12585 | Irgq | IRGQ | -2.87E-03 | 1.08E-02 |
| 12586 | Sumo2 | SUMO2 | 2.51E-03 | -4.24E-03 |
| 12587 | Timm10 | TIMM10 | -3.57E-03 | -6.83E-03 |
| 12588 | Ccsap | CCSAP | 1.20E-02 | 7.31E-02 |
| 12589 | 4930404N11Rik | C19orf71 | -6.21E-03 | 1.26E-02 |
| 12590 | Ociad2 | OCIAD2 | 1.68E-02 | 1.03E-01 |
| 12591 | Kirrel2 | KIRREL2 | -6.67E-03 | 4.70E-02 |
| 12592 | Twist2 | TWIST2 | -7.83E-03 | 2.77E-02 |
| 12593 | Rfc1 | RFC1 | -2.63E-03 | -1.01E-02 |
| 12594 | Pik3ip1 | PIK3IP1 | -7.80E-03 | -9.40E-03 |
| 12595 | Rfc2 | RFC2 | -3.34E-03 | -3.61E-04 |
| 12596 | Klhl38 | KLHL38 | 1.21E-02 | -1.83E-02 |
| 12597 | Kctd8 | KCTD8 | -9.11E-03 | -3.37E-02 |
| 12598 | Rbpjl | RBPJL | 4.57E-03 | -1.08E-03 |
| 12599 | Neurl1b | NEURL1B | 1.30E-02 | 9.65E-03 |
| 12600 | Acta1 | ACTA1 | 2.06E-02 | 1.05E-01 |
| 12601 | Stk36 | STK36 | -4.61E-03 | 1.17E-03 |
| 12602 | Fasn | FASN | 5.44E-03 | 3.39E-02 |
| 12603 | Flywch2 | FLYWCH2 | 8.26E-03 | 1.96E-02 |
| 12604 | Fbxl12 | FBXL12 | -3.37E-03 | -1.45E-02 |
| 12605 | Gmnn | GMNN | 5.09E-03 | 1.73E-02 |
| 12606 | Hoxd11 | HOXD11 | -1.28E-02 | 1.15E-02 |
| 12607 | Ocel1 | OCEL1 | 4.42E-03 | 3.04E-03 |
| 12608 | Dnah17 | DNAH17 | 1.78E-02 | -1.20E-01 |
| 12609 | Cybrd1 | CYBRD1 | -1.39E-02 | 9.68E-02 |
| 12610 | Neo1 | NEO1 | 2.44E-03 | -2.39E-02 |
| 12611 | Cyba | CYBA | 4.52E-03 | -9.67E-05 |

| | | | | |
|---|---|---|---|---|
| 12612 | Tshz3 | TSHZ3 | 1.15E-02 | 3.48E-02 |
| 12613 | Dennd1a | DENND1A | 5.79E-03 | -4.61E-02 |
| 12614 | Kdm8 | KDM8 | 6.87E-03 | -1.57E-02 |
| 12615 | Rpl17 | RPL17 | -2.60E-03 | 1.10E-02 |
| 12616 | Mcfd2 | MCFD2 | -2.76E-03 | -9.58E-03 |
| 12617 | Il17c | IL17C | 4.39E-03 | 2.00E-03 |
| 12618 | Otogl | OTOGL | -4.22E-03 | 2.50E-02 |
| 12619 | Spef1 | SPEF1 | 9.40E-03 | 4.04E-02 |
| 12620 | Tbx15 | TBX15 | 8.24E-03 | -1.20E-02 |
| 12621 | Tdp2 | TDP2 | 3.92E-03 | 1.49E-02 |
| 12622 | Tbc1d17 | TBC1D17 | -3.09E-03 | -4.02E-02 |
| 12623 | Midn | MIDN | 6.27E-03 | 1.07E-02 |
| 12624 | Fam118a | FAM118A | -4.02E-03 | -2.30E-02 |
| 12625 | Psma1 | PSMA1 | -2.48E-03 | 4.83E-03 |
| 12626 | Celsr2 | CELSR2 | -3.48E-03 | -2.79E-02 |
| 12627 | Runx1t1 | RUNX1T1 | 1.10E-02 | -2.41E-02 |
| 12628 | Rnaset2a | RNASET2 | 5.47E-03 | 3.95E-02 |
| 12629 | Hapln4 | HAPLN4 | 1.18E-02 | 8.65E-02 |
| 12630 | Pcdhga2 | PCDHGA2 | -1.03E-02 | -3.72E-02 |
| 12631 | Nrd1 | NRD1 | 2.19E-03 | -1.61E-02 |
| 12632 | Fancl | FANCL | 4.48E-03 | 2.69E-02 |
| 12633 | Arhgef39 | ARHGEF39 | 9.87E-03 | -1.77E-02 |
| 12634 | Acd | ACD | 3.44E-03 | -1.11E-03 |
| 12635 | Nanog | NANOG | 4.32E-03 | -1.64E-02 |
| 12636 | Glod5 | GLOD5 | -4.44E-03 | -3.24E-02 |
| 12637 | Cyp2c55 | CYP2C18 | -4.59E-03 | 1.10E-02 |
| 12638 | Caskin1 | CASKIN1 | 1.00E-02 | -2.77E-02 |
| 12639 | Dpp8 | DPP8 | -2.80E-03 | -2.60E-02 |
| 12640 | Tcp11l1 | TCP11L1 | 5.61E-03 | 1.22E-03 |
| 12641 | Igfbp4 | IGFBP4 | 5.84E-03 | 1.59E-02 |
| 12642 | Ppa2 | PPA2 | -3.68E-03 | -1.86E-02 |
| 12643 | Ppef2 | PPEF2 | -7.19E-03 | -3.53E-02 |
| 12644 | Tgif2 | TGIF2 | 1.40E-02 | 4.76E-02 |
| 12645 | Fbxo48 | FBXO48 | 9.29E-03 | 6.09E-02 |
| 12646 | Rapgefl1 | RAPGEFL1 | -1.55E-02 | -3.23E-02 |
| 12647 | Tmem8b | TMEM8B | 5.50E-03 | -6.33E-02 |
| 12648 | Rpl11 | RPL11 | 2.70E-03 | 4.39E-03 |
| 12649 | Tmprss11f | TMPRSS11F | 4.03E-03 | 2.41E-02 |
| 12650 | Pex5l | PEX5L | 1.62E-02 | -1.13E-01 |
| 12651 | Hapln3 | HAPLN3 | 5.75E-03 | -2.21E-02 |
| 12652 | Smchd1 | SMCHD1 | -3.95E-03 | 6.19E-03 |
| 12653 | Syngr4 | SYNGR4 | 8.08E-03 | 4.21E-02 |
| 12654 | Kcng3 | KCNG3 | -5.24E-03 | -7.00E-04 |
| 12655 | Pomc | POMC | -9.18E-03 | -2.94E-02 |
| 12656 | Fzd3 | FZD3 | -6.25E-03 | 3.55E-03 |
| 12657 | Pspc1 | PSPC1 | 4.99E-03 | 1.35E-02 |
| 12658 | Mrpl49 | MRPL49 | -2.69E-03 | -1.63E-02 |
| 12659 | Map1lc3b | MAP1LC3B | 2.76E-03 | -1.07E-02 |
| 12660 | Plcb3 | PLCB3 | 2.62E-03 | -2.41E-03 |
| 12661 | Stk16 | STK16 | 2.71E-03 | -8.57E-03 |
| 12662 | Sertad4 | SERTAD4 | 9.45E-03 | 3.53E-02 |
| 12663 | Rpl10a | RPL10A | 2.93E-03 | 2.82E-02 |
| 12664 | Slc2a3 | SLC2A3 | 1.61E-02 | 5.00E-02 |

| | | | | |
|---|---|---|---|---|
| 12665 | Rap1gap | RAP1GAP | -7.14E-03 | -3.37E-02 |
| 12666 | Recql4 | RECQL4 | -1.26E-02 | 7.72E-02 |
| 12667 | Ano9 | ANO9 | -6.68E-03 | 3.61E-02 |
| 12668 | Gal3st1 | GAL3ST1 | 9.11E-03 | -3.31E-02 |
| 12669 | Zfp536 | ZNF536 | 6.51E-03 | 2.58E-03 |
| 12670 | Adnp2 | ADNP2 | 5.12E-03 | -2.84E-02 |
| 12671 | Mink1 | MINK1 | 2.58E-03 | -2.10E-02 |
| 12672 | Rpl13 | RPL13 | 2.77E-03 | 1.07E-02 |
| 12673 | Mss51 | MSS51 | 8.66E-03 | 5.20E-02 |
| 12674 | Recql | RECQL | 4.51E-03 | 1.89E-02 |
| 12675 | Cspg5 | CSPG5 | 1.17E-02 | 1.98E-02 |
| 12676 | Shroom3 | SHROOM3 | 5.59E-03 | 1.17E-02 |
| 12677 | Fhad1 | FHAD1 | -1.03E-02 | 3.97E-02 |
| 12678 | Trim26 | TRIM26 | 3.03E-03 | 3.06E-03 |
| 12679 | Ccdc174 | CCDC174 | 3.72E-03 | 1.99E-03 |
| 12680 | Mtif3 | MTIF3 | 3.25E-03 | -1.10E-02 |
| 12681 | Anks6 | ANKS6 | 4.00E-03 | 5.89E-03 |
| 12682 | Crebbp | CREBBP | 2.32E-03 | -7.67E-04 |
| 12683 | Exoc2 | EXOC2 | 3.55E-03 | 1.36E-03 |
| 12684 | Mib1 | MIB1 | 3.42E-03 | -1.14E-02 |
| 12685 | Numa1 | NUMA1 | 3.01E-03 | -1.45E-02 |
| 12686 | Dctn1 | DCTN1 | 3.24E-03 | -1.85E-02 |
| 12687 | Mfsd7a | MFSD7 | -7.27E-03 | -3.72E-02 |
| 12688 | Syt16 | SYT16 | 1.56E-02 | 6.63E-02 |
| 12689 | Urod | UROD | 3.10E-03 | -7.76E-03 |
| 12690 | Ubxn11 | UBXN11 | -6.08E-03 | -4.28E-02 |
| 12691 | Has2 | HAS2 | -3.19E-03 | 2.51E-02 |
| 12692 | Tox3 | TOX3 | -6.87E-03 | -1.47E-02 |
| 12693 | Abcc4 | ABCC4 | -6.89E-03 | -2.04E-02 |
| 12694 | Tnfrsf9 | TNFRSF9 | -9.61E-03 | 1.19E-01 |
| 12695 | Sytl2 | SYTL2 | 1.16E-02 | 4.18E-02 |
| 12696 | B3gnt8 | B3GNT8 | -1.27E-02 | 3.91E-02 |
| 12697 | Rpl4 | RPL4 | 2.66E-03 | 7.69E-03 |
| 12698 | Snrk | SNRK | -4.64E-03 | 4.94E-04 |
| 12699 | Soga1 | SOGA1 | -1.07E-02 | -2.71E-03 |
| 12700 | Zscan18 | ZSCAN18 | -1.22E-02 | 6.75E-02 |
| 12701 | Pros1 | PROS1 | 4.86E-03 | 5.71E-02 |
| 12702 | Dtymk | DTYMK | 4.62E-03 | 1.22E-02 |
| 12703 | Plekho2 | PLEKHO2 | -8.68E-03 | 1.44E-02 |
| 12704 | Cdan1 | CDAN1 | -3.78E-03 | 8.28E-04 |
| 12705 | Las1l | LAS1L | 2.60E-03 | 2.32E-03 |
| 12706 | Acss3 | ACSS3 | -6.00E-03 | 4.15E-02 |
| 12707 | Tnfrsf11a | TNFRSF11A | -1.01E-02 | 1.29E-02 |
| 12708 | Ascl3 | ASCL3 | 9.07E-03 | -4.75E-02 |
| 12709 | Kdm5a | KDM5A | 3.55E-03 | -2.51E-02 |
| 12710 | Rps6ka3 | RPS6KA3 | -2.49E-03 | -3.23E-03 |
| 12711 | Zfp786 | ZNF786 | -8.19E-03 | 6.55E-03 |
| 12712 | Pyurf | PYURF | 3.38E-03 | -1.57E-02 |
| 12713 | Xdh | XDH | -8.14E-03 | 9.83E-03 |
| 12714 | Pih1d1 | PIH1D1 | 2.39E-03 | -1.12E-02 |
| 12715 | Hmgcl | HMGCL | 3.22E-03 | -2.99E-02 |
| 12716 | Aldh16a1 | ALDH16A1 | -3.27E-03 | -2.28E-02 |
| 12717 | Diras2 | DIRAS2 | 1.13E-02 | 1.71E-02 |

| | | | | |
|---|---|---|---|---|
| 12718 | Serpina1e | SERPINA1 | 1.57E-02 | -5.45E-02 |
| 12719 | Sos2 | SOS2 | -3.29E-03 | -1.41E-02 |
| 12720 | Cuzd1 | CUZD1 | 1.89E-02 | 5.44E-02 |
| 12721 | Wdr66 | WDR66 | 8.17E-03 | -3.01E-02 |
| 12722 | Lynx1 | LYNX1 | 8.94E-03 | -1.50E-02 |
| 12723 | Dhx38 | DHX38 | -2.16E-03 | -1.87E-02 |
| 12724 | 5730455P16Rik | C17orf75 | 4.47E-03 | -4.11E-02 |
| 12725 | Znrf3 | ZNRF3 | 2.95E-03 | -3.31E-02 |
| 12726 | Ndufb2 | NDUFB2 | -3.63E-03 | -1.23E-02 |
| 12727 | Ftsj1 | FTSJ1 | -4.13E-03 | -2.01E-03 |
| 12728 | Slc36a2 | SLC36A2 | -2.01E-02 | 6.39E-02 |
| 12729 | Fam131c | FAM131C | 1.42E-02 | -4.61E-02 |
| 12730 | Mavs | MAVS | 2.49E-03 | -4.06E-04 |
| 12731 | Tnfaip6 | TNFAIP6 | 4.29E-03 | 1.11E-02 |
| 12732 | Fbxo45 | FBXO45 | 3.77E-03 | 1.32E-02 |
| 12733 | Maged1 | MAGED1 | 2.57E-03 | -1.15E-02 |
| 12734 | Fan1 | FAN1 | 4.59E-03 | -3.27E-02 |
| 12735 | Samd15 | SAMD15 | -9.01E-03 | -1.81E-02 |
| 12736 | Enoph1 | ENOPH1 | -3.70E-03 | 1.21E-03 |
| 12737 | Hspb1 | HSPB1 | -1.69E-02 | 1.15E-01 |
| 12738 | Hrh2 | HRH2 | -9.05E-03 | 7.22E-02 |
| 12739 | Sbspon | SBSPON | -3.72E-02 | 8.80E-02 |
| 12740 | Zfpl1 | ZFPL1 | -3.32E-03 | 5.75E-05 |
| 12741 | Herc1 | HERC1 | 2.05E-03 | -9.17E-03 |
| 12742 | Slc47a1 | SLC47A1 | 4.33E-03 | -6.50E-03 |
| 12743 | Cyp2c65 | CYP2C9 | -2.89E-03 | 1.61E-02 |
| 12744 | Isca2 | ISCA2 | -2.29E-03 | -1.29E-02 |
| 12745 | Cep112 | CEP112 | -6.59E-03 | -2.41E-02 |
| 12746 | Cxcr4 | CXCR4 | 1.04E-02 | 3.66E-02 |
| 12747 | Mcm3ap | MCM3AP | 2.24E-03 | -1.15E-02 |
| 12748 | Snapc5 | SNAPC5 | 3.12E-03 | -5.75E-03 |
| 12749 | Tmem30a | TMEM30A | -2.33E-03 | -1.40E-02 |
| 12750 | Plb1 | PLB1 | -6.39E-03 | 3.24E-02 |
| 12751 | Ndufaf4 | NDUFAF4 | -3.92E-03 | 3.25E-03 |
| 12752 | Smim19 | SMIM19 | 2.56E-03 | -1.90E-02 |
| 12753 | Pla2g1b | PLA2G1B | 4.31E-03 | -9.80E-03 |
| 12754 | Cyp39a1 | CYP39A1 | -7.88E-03 | 9.83E-03 |
| 12755 | Pnpla2 | PNPLA2 | -3.33E-03 | -1.18E-02 |
| 12756 | Naa38 | NAA38 | 3.00E-03 | -1.74E-02 |
| 12757 | Utp15 | UTP15 | 3.08E-03 | -1.26E-02 |
| 12758 | Ubc | UBC | -5.77E-03 | 1.26E-02 |
| 12759 | Ptpru | PTPRU | 6.92E-03 | -4.54E-02 |
| 12760 | Frmpd4 | FRMPD4 | 1.77E-02 | 3.19E-02 |
| 12761 | Cnot1 | CNOT1 | -2.57E-03 | -1.04E-02 |
| 12762 | Senp1 | SENP1 | -2.20E-03 | 5.90E-03 |
| 12763 | Srpx | SRPX | 1.33E-02 | 5.84E-02 |
| 12764 | Wac | WAC | 2.15E-03 | 6.77E-04 |
| 12765 | Cntf | CNTF | 6.51E-03 | 2.64E-02 |
| 12766 | Tcfl5 | TCFL5 | 1.96E-03 | -7.64E-03 |
| 12767 | Rp9 | RP9 | -3.40E-03 | 2.52E-03 |
| 12768 | Prrt2 | PRRT2 | 4.90E-03 | 4.56E-03 |
| 12769 | Pten | PTEN | -3.12E-03 | -4.59E-03 |
| 12770 | Immp2l | IMMP2L | 7.13E-03 | 3.75E-02 |

| | | | | |
|---|---|---|---|---|
| 12771 | Fat4 | FAT4 | -1.18E-02 | -7.63E-03 |
| 12772 | Prpf31 | PRPF31 | 2.41E-03 | 8.66E-03 |
| 12773 | Zc3h12d | ZC3H12D | 1.56E-02 | -2.89E-02 |
| 12774 | Heatr3 | HEATR3 | -3.31E-03 | 2.67E-02 |
| 12775 | Thumpd3 | THUMPD3 | 2.62E-03 | 9.91E-03 |
| 12776 | Stk33 | STK33 | 2.69E-03 | 1.82E-03 |
| 12777 | Tmem47 | TMEM47 | -6.33E-03 | 1.65E-02 |
| 12778 | 6030468B19Rik | C17orf99 | -3.53E-03 | -6.11E-03 |
| 12779 | Cartpt | CARTPT | 4.39E-03 | 6.66E-03 |
| 12780 | Arl8b | ARL8B | -2.41E-03 | 1.94E-03 |
| 12781 | Map3k12 | MAP3K12 | 3.66E-03 | 1.12E-03 |
| 12782 | Ing5 | ING5 | -3.22E-03 | -1.50E-02 |
| 12783 | S1pr2 | S1PR2 | -1.05E-02 | 5.05E-02 |
| 12784 | Arl16 | ARL16 | 3.92E-03 | -1.07E-02 |
| 12785 | Klhdc1 | KLHDC1 | 5.59E-03 | -3.45E-03 |
| 12786 | Ptchd4 | PTCHD4 | -7.74E-03 | -2.29E-03 |
| 12787 | Tmem151a | TMEM151A | -8.03E-03 | 5.63E-02 |
| 12788 | Rora | RORA | -5.04E-03 | 4.94E-03 |
| 12789 | Ung | UNG | -6.64E-03 | 1.02E-01 |
| 12790 | Grb10 | GRB10 | -4.03E-03 | 3.48E-02 |
| 12791 | H1f0 | H1F0 | 3.75E-03 | -3.05E-02 |
| 12792 | Gas6 | GAS6 | -3.92E-03 | -1.83E-02 |
| 12793 | L3mbtl1 | L3MBTL1 | -6.53E-03 | 1.47E-02 |
| 12794 | Iqsec3 | IQSEC3 | -7.18E-03 | 2.78E-02 |
| 12795 | Uvrag | UVRAG | 3.12E-03 | -5.07E-03 |
| 12796 | Pax1 | PAX1 | -3.50E-03 | -7.51E-03 |
| 12797 | Agr2 | AGR2 | -5.38E-03 | 3.14E-03 |
| 12798 | Rrh | RRH | 4.25E-03 | -1.12E-02 |
| 12799 | Ipo11 | IPO11 | -2.33E-03 | -1.75E-03 |
| 12800 | Tpm3 | TPM3 | 2.20E-03 | 2.32E-02 |
| 12801 | Esam | ESAM | 1.00E-02 | -5.67E-02 |
| 12802 | Rnf41 | RNF41 | -2.93E-03 | 2.35E-03 |
| 12803 | Rps8 | RPS8 | -2.34E-03 | 9.94E-03 |
| 12804 | Emx1 | EMX1 | -5.44E-03 | 7.99E-03 |
| 12805 | Perp | PERP | -2.35E-03 | 1.74E-02 |
| 12806 | Prickle2 | PRICKLE2 | 8.44E-03 | -2.25E-02 |
| 12807 | Helq | HELQ | -2.70E-03 | -2.55E-03 |
| 12808 | Dhrs13 | DHRS13 | 5.30E-03 | 2.88E-02 |
| 12809 | Mn1 | MN1 | 9.44E-03 | -1.12E-02 |
| 12810 | Rasgrp1 | RASGRP1 | -1.13E-02 | -8.23E-02 |
| 12811 | Ddx31 | DDX31 | 5.55E-03 | -1.73E-03 |
| 12812 | Pelo | PELO | 5.00E-03 | 3.03E-02 |
| 12813 | Fxyd4 | FXYD4 | 2.01E-02 | -5.58E-02 |
| 12814 | Nphs2 | NPHS2 | -6.68E-03 | 2.79E-04 |
| 12815 | Slc6a19 | SLC6A19 | 7.22E-03 | -5.02E-02 |
| 12816 | Mrpl28 | MRPL28 | 2.39E-03 | -1.14E-02 |
| 12817 | Rasl11b | RASL11B | -1.90E-02 | -1.46E-02 |
| 12818 | E130309D02Rik | C7orf26 | 3.00E-03 | -3.02E-03 |
| 12819 | Etfb | ETFB | -5.24E-03 | -2.38E-03 |
| 12820 | Tgm2 | TGM2 | 4.29E-03 | -3.91E-03 |
| 12821 | Cpn2 | CPN2 | -6.29E-03 | 5.27E-02 |
| 12822 | Pitx2 | PITX2 | 9.25E-03 | -1.84E-02 |
| 12823 | Ccdc88c | CCDC88C | -4.08E-03 | -3.72E-02 |

| | | | | |
|---|---|---|---|---|
| 12824 | Col4a6 | COL4A6 | -2.44E-02 | 7.83E-02 |
| 12825 | Has1 | HAS1 | -2.17E-03 | 1.79E-02 |
| 12826 | Ier3ip1 | IER3IP1 | -4.32E-03 | 1.20E-02 |
| 12827 | Asl | ASL | 5.78E-03 | -1.82E-02 |
| 12828 | Cep170b | CEP170B | 2.15E-03 | -7.43E-03 |
| 12829 | Pls1 | PLS1 | 3.74E-03 | -4.15E-02 |
| 12830 | Tnn | TNN | 5.11E-02 | -1.11E-01 |
| 12831 | Vwa7 | VWA7 | -1.62E-02 | 5.38E-02 |
| 12832 | Duoxa1 | DUOXA1 | 8.28E-03 | 3.58E-02 |
| 12833 | Kcnh8 | KCNH8 | -4.90E-03 | 4.51E-02 |
| 12834 | Tnmd | TNMD | -4.74E-03 | 2.08E-02 |
| 12835 | Itga11 | ITGA11 | -1.20E-02 | 8.77E-02 |
| 12836 | Snrnp25 | SNRNP25 | 6.21E-03 | -2.09E-02 |
| 12837 | Fsd1 | FSD1 | -3.93E-03 | -1.97E-02 |
| 12838 | Scap | SCAP | 2.21E-03 | -2.55E-02 |
| 12839 | Strip2 | STRIP2 | -1.50E-02 | -9.42E-03 |
| 12840 | Yy1 | YY1 | 2.01E-03 | 1.61E-03 |
| 12841 | Mamdc2 | MAMDC2 | -8.44E-03 | 5.95E-02 |
| 12842 | Rpl31 | RPL31 | -2.11E-03 | 6.70E-03 |
| 12843 | Plp2 | PLP2 | 6.26E-03 | 1.03E-01 |
| 12844 | 1700125H20Rik | C17orf64 | 2.99E-03 | 1.09E-02 |
| 12845 | Pnpo | PNPO | 3.33E-03 | -2.39E-02 |
| 12846 | Prpf18 | PRPF18 | 2.17E-03 | -1.26E-02 |
| 12847 | Rbm27 | RBM27 | -2.55E-03 | 6.34E-03 |
| 12848 | Paqr3 | PAQR3 | -6.08E-03 | 2.93E-02 |
| 12849 | Ednrb | EDNRB | 6.84E-03 | 5.63E-03 |
| 12850 | Sorcs2 | SORCS2 | -1.18E-02 | -5.48E-02 |
| 12851 | Dynll1 | DYNLL1 | -4.46E-03 | 2.82E-02 |
| 12852 | Ugp2 | UGP2 | 3.75E-03 | -1.31E-02 |
| 12853 | Atp10b | ATP10B | 3.14E-02 | 1.29E-02 |
| 12854 | Plekhg1 | PLEKHG1 | -5.95E-03 | -5.89E-02 |
| 12855 | Tfip11 | TFIP11 | 1.82E-03 | -5.15E-03 |
| 12856 | Myo1h | MYO1H | 4.59E-03 | 4.96E-02 |
| 12857 | Tmem161a | TMEM161A | 2.40E-03 | -2.27E-02 |
| 12858 | Abcb11 | ABCB11 | -8.15E-03 | 3.57E-02 |
| 12859 | Kif21a | KIF21A | -3.61E-03 | -3.31E-02 |
| 12860 | Pld2 | PLD2 | 4.77E-03 | 2.21E-02 |
| 12861 | Itih5 | ITIH5 | 1.37E-02 | -1.20E-01 |
| 12862 | Krt12 | KRT12 | -4.14E-03 | 5.01E-02 |
| 12863 | Rnf165 | RNF165 | 6.45E-03 | -3.26E-02 |
| 12864 | Cat | CAT | 5.29E-03 | -1.01E-02 |
| 12865 | Itga2 | ITGA2 | 3.79E-02 | 2.21E-01 |
| 12866 | Lrrc61 | LRRC61 | 3.15E-03 | -3.26E-02 |
| 12867 | Nfs1 | NFS1 | -2.83E-03 | -2.86E-02 |
| 12868 | Actg1 | ACTG1 | 2.89E-03 | 1.08E-02 |
| 12869 | Pkp1 | PKP1 | 1.93E-02 | 5.15E-02 |
| 12870 | BC051019 | C11orf16 | 3.47E-03 | 1.27E-02 |
| 12871 | Islr | ISLR | -1.57E-02 | 6.44E-02 |
| 12872 | Klhl25 | KLHL25 | -4.69E-03 | 3.18E-02 |
| 12873 | Upb1 | UPB1 | 3.98E-03 | -1.68E-02 |
| 12874 | Adamdec1 | ADAMDEC1 | 1.10E-02 | 3.72E-02 |
| 12875 | Ttc33 | TTC33 | 2.26E-03 | -2.35E-02 |
| 12876 | Rbms2 | RBMS2 | -2.21E-03 | -5.93E-03 |

| | | | | |
|---|---|---|---|---|
| 12877 | Rpl28 | RPL28 | -2.41E-03 | 4.26E-03 |
| 12878 | Emp3 | EMP3 | -6.89E-03 | 7.93E-02 |
| 12879 | Abcd3 | ABCD3 | 5.68E-03 | 1.96E-02 |
| 12880 | Gramd1a | GRAMD1A | -2.57E-03 | 1.82E-02 |
| 12881 | Cbx8 | CBX8 | 5.27E-03 | -5.48E-03 |
| 12882 | Dtx2 | DTX2 | 3.59E-03 | 2.28E-02 |
| 12883 | Bbs5 | BBS5 | 4.17E-03 | 9.32E-03 |
| 12884 | Krt28 | KRT28 | -6.28E-03 | 6.80E-03 |
| 12885 | Arf5 | ARF5 | -2.64E-03 | 6.68E-04 |
| 12886 | Parp16 | PARP16 | 3.02E-03 | -1.24E-04 |
| 12887 | Adam28 | ADAM28 | 1.57E-02 | 3.12E-02 |
| 12888 | Slc25a41 | SLC25A41 | -2.19E-03 | -4.98E-03 |
| 12889 | Pld1 | PLD1 | 5.84E-03 | -2.99E-02 |
| 12890 | Cd164l2 | CD164L2 | -5.22E-03 | -7.64E-03 |
| 12891 | Nenf | NENF | -3.23E-03 | 2.77E-04 |
| 12892 | Zfp236 | ZNF236 | -3.05E-03 | 8.62E-03 |
| 12893 | Med21 | MED21 | 3.92E-03 | 3.11E-03 |
| 12894 | Golt1b | GOLT1B | -4.81E-03 | 3.23E-02 |
| 12895 | Rgn | RGN | 6.48E-03 | 2.84E-02 |
| 12896 | Prr22 | PRR22 | -7.26E-03 | -3.07E-03 |
| 12897 | Btbd16 | BTBD16 | -4.00E-03 | 5.12E-02 |
| 12898 | Ppargc1a | PPARGC1A | -3.28E-03 | -3.41E-02 |
| 12899 | Tex261 | TEX261 | -1.80E-03 | -1.12E-02 |
| 12900 | Htra3 | HTRA3 | -1.96E-02 | 1.55E-01 |
| 12901 | Fam229b | FAM229B | -5.73E-03 | 6.72E-04 |
| 12902 | Serpina3c | SERPINA3 | -1.05E-02 | 5.39E-02 |
| 12903 | Rnf10 | RNF10 | 1.83E-03 | -1.18E-02 |
| 12904 | Hbegf | HBEGF | -1.41E-02 | 5.53E-02 |
| 12905 | Tmem151b | TMEM151B | -8.69E-03 | 1.05E-01 |
| 12906 | 2300009A05Rik | C15orf61 | -2.80E-03 | 8.94E-03 |
| 12907 | 3110009E18Rik | C2orf76 | 8.79E-03 | -2.59E-02 |
| 12908 | Arl5a | ARL5A | -2.68E-03 | -1.01E-02 |
| 12909 | Hsd17b12 | HSD17B12 | 2.30E-03 | 6.36E-03 |
| 12910 | Ivns1abp | IVNS1ABP | 2.23E-03 | -8.28E-03 |
| 12911 | Gbx2 | GBX2 | 2.71E-03 | 1.58E-02 |
| 12912 | Ftl1 | FTL | -4.62E-03 | -1.42E-03 |
| 12913 | Myl6b | MYL6B | 5.80E-03 | -5.90E-03 |
| 12914 | Clns1a | CLNS1A | 2.07E-03 | 2.72E-03 |
| 12915 | Ltn1 | LTN1 | 2.14E-03 | -1.58E-02 |
| 12916 | Tspan11 | TSPAN11 | -7.80E-03 | -1.25E-02 |
| 12917 | Zbtb21 | ZBTB21 | -3.60E-03 | 4.22E-02 |
| 12918 | Slc38a4 | SLC38A4 | 1.19E-02 | -3.67E-02 |
| 12919 | Elovl4 | ELOVL4 | 1.16E-02 | -5.09E-03 |
| 12920 | Aldh4a1 | ALDH4A1 | 3.61E-03 | -1.32E-02 |
| 12921 | Stard4 | STARD4 | -4.88E-03 | -2.02E-02 |
| 12922 | Bbs10 | BBS10 | 6.91E-03 | -3.40E-02 |
| 12923 | Arl4d | ARL4D | 1.73E-02 | 7.53E-03 |
| 12924 | Strn3 | STRN3 | -2.14E-03 | 1.73E-03 |
| 12925 | Tex11 | TEX11 | 4.59E-03 | 3.58E-03 |
| 12926 | Atp1a3 | ATP1A3 | -6.39E-03 | 6.10E-02 |
| 12927 | Birc6 | BIRC6 | -2.71E-03 | -6.10E-03 |
| 12928 | Slc5a7 | SLC5A7 | 2.74E-03 | 2.02E-02 |
| 12929 | Rmdn1 | RMDN1 | 3.03E-03 | -2.46E-03 |

| | | | | |
|---|---|---|---|---|
| 12930 | Acvr1c | ACVR1C | 1.10E-02 | 3.00E-02 |
| 12931 | Htr3a | HTR3A | 1.02E-02 | 2.14E-01 |
| 12932 | Rpl8 | RPL8 | 2.09E-03 | 7.46E-04 |
| 12933 | Enah | ENAH | -4.78E-03 | -4.20E-02 |
| 12934 | Sox9 | SOX9 | 9.78E-03 | 2.22E-01 |
| 12935 | Klc4 | KLC4 | -2.56E-03 | -1.60E-02 |
| 12936 | Dip2b | DIP2B | 2.79E-03 | 1.20E-02 |
| 12937 | Me1 | ME1 | 3.20E-03 | 4.15E-03 |
| 12938 | Ypel1 | YPEL1 | 8.34E-03 | -2.40E-02 |
| 12939 | Pcbp2 | PCBP2 | 1.78E-03 | -2.49E-04 |
| 12940 | Rab23 | RAB23 | -3.00E-03 | -1.49E-02 |
| 12941 | Aldh1l2 | ALDH1L2 | -1.10E-02 | 1.87E-01 |
| 12942 | Myd88 | MYD88 | -3.95E-03 | 2.86E-02 |
| 12943 | Hdac7 | HDAC7 | -3.66E-03 | 1.57E-02 |
| 12944 | Sema3a | SEMA3A | 9.28E-03 | -3.10E-02 |
| 12945 | Tns4 | TNS4 | 2.05E-02 | 7.85E-02 |
| 12946 | Abcc5 | ABCC5 | 4.38E-03 | -3.10E-03 |
| 12947 | Tfeb | TFEB | -2.69E-03 | 1.94E-02 |
| 12948 | Sorbs1 | SORBS1 | -6.67E-03 | -1.57E-02 |
| 12949 | Bard1 | BARD1 | 1.14E-02 | 1.28E-01 |
| 12950 | Coro7 | CORO7 | 4.03E-03 | -2.25E-03 |
| 12951 | Coro7 | CORO7-PAM16 | 4.03E-03 | -2.25E-03 |
| 12952 | Slc4a1 | SLC4A1 | 9.66E-03 | -7.42E-02 |
| 12953 | Tnni1 | TNNI1 | 2.30E-03 | 1.35E-02 |
| 12954 | Wdr31 | WDR31 | -4.80E-03 | -2.98E-02 |
| 12955 | Prpf39 | PRPF39 | -2.24E-03 | -3.72E-03 |
| 12956 | Tex14 | TEX14 | -7.77E-03 | -8.10E-03 |
| 12957 | Rhebl1 | RHEBL1 | -1.00E-02 | -7.79E-06 |
| 12958 | Lyrm7 | LYRM7 | -3.35E-03 | -1.52E-02 |
| 12959 | Mageb3 | MAGEB1 | 4.11E-03 | -2.01E-03 |
| 12960 | Tnnc2 | TNNC2 | 2.84E-03 | -3.75E-03 |
| 12961 | Fbln5 | FBLN5 | -9.30E-03 | -3.17E-02 |
| 12962 | Zcchc4 | ZCCHC4 | 2.58E-03 | -6.28E-03 |
| 12963 | Ehd2 | EHD2 | -4.55E-03 | 2.37E-02 |
| 12964 | Hnrnpul2 | HNRNPUL2 | -1.66E-03 | -7.39E-03 |
| 12965 | Gemin2 | GEMIN2 | -3.31E-03 | 3.40E-03 |
| 12966 | Fhl1 | FHL1 | -2.74E-03 | 7.29E-04 |
| 12967 | Pde4c | PDE4C | -3.29E-03 | -2.41E-02 |
| 12968 | Nrm | NRM | 1.24E-02 | 1.64E-01 |
| 12969 | Psapl1 | PSAPL1 | 6.44E-03 | 1.13E-02 |
| 12970 | Ccdc33 | CCDC33 | 4.92E-03 | 5.33E-02 |
| 12971 | Exoc1 | EXOC1 | 1.70E-03 | -5.63E-03 |
| 12972 | Gpr4 | GPR4 | -7.94E-03 | 2.01E-02 |
| 12973 | Sdc4 | SDC4 | -1.99E-03 | -4.89E-03 |
| 12974 | Tango6 | TANGO6 | 3.42E-03 | -2.85E-03 |
| 12975 | Wdr75 | WDR75 | 2.17E-03 | 2.75E-02 |
| 12976 | Frem1 | FREM1 | 1.12E-02 | 1.27E-02 |
| 12977 | Sowahb | SOWAHB | -3.58E-03 | 9.25E-04 |
| 12978 | Mical1 | MICAL1 | -6.90E-03 | 2.74E-02 |
| 12979 | Gng3 | GNG3 | -3.78E-03 | 2.07E-04 |
| 12980 | Cdx4 | CDX4 | 3.39E-03 | 2.07E-02 |
| 12981 | Mycbpap | MYCBPAP | -1.42E-02 | -6.63E-03 |
| 12982 | Slc5a5 | SLC5A5 | -5.55E-03 | 2.84E-02 |

| | | | | |
|---|---|---|---|---|
| 12983 | Slc25a31 | SLC25A31 | -3.62E-03 | 4.82E-03 |
| 12984 | Col14a1 | COL14A1 | 1.35E-02 | -5.72E-02 |
| 12985 | Ephb2 | EPHB2 | -1.09E-02 | -2.46E-02 |
| 12986 | Mex3b | MEX3B | -1.00E-02 | -3.08E-02 |
| 12987 | Nrk | NRK | 2.36E-03 | 4.75E-04 |
| 12988 | Tmem88b | TMEM88B | -1.05E-02 | 3.42E-02 |
| 12989 | Sertad1 | SERTAD1 | 5.77E-03 | -4.18E-03 |
| 12990 | Dtx3 | DTX3 | -1.94E-03 | 7.43E-03 |
| 12991 | Larp7 | LARP7 | 2.55E-03 | 3.24E-03 |
| 12992 | Fam228b | FAM228B | 7.38E-03 | -5.60E-02 |
| 12993 | Gtf2e1 | GTF2E1 | 3.85E-03 | 7.27E-03 |
| 12994 | Eml6 | EML6 | -3.50E-03 | -3.93E-02 |
| 12995 | Mrpl39 | MRPL39 | -2.46E-03 | -1.70E-02 |
| 12996 | Pax6 | PAX6 | 2.12E-03 | 1.22E-02 |
| 12997 | Mybpc1 | MYBPC1 | 4.68E-03 | -7.46E-03 |
| 12998 | Mia2 | MIA2 | 2.79E-03 | -1.31E-02 |
| 12999 | Slc25a42 | SLC25A42 | 5.41E-03 | 1.04E-02 |
| 13000 | Flt1 | FLT1 | 5.09E-03 | -1.73E-02 |
| 13001 | Emilin2 | EMILIN2 | 3.11E-02 | 5.57E-02 |
| 13002 | Prr3 | PRR3 | 2.89E-03 | 9.95E-03 |
| 13003 | AI467606 | C16orf54 | -9.72E-03 | -3.82E-02 |
| 13004 | Brd1 | BRD1 | 2.01E-03 | -9.51E-03 |
| 13005 | Stmn3 | STMN3 | 4.46E-03 | 4.36E-02 |
| 13006 | Crnkl1 | CRNKL1 | -2.86E-03 | 8.67E-03 |
| 13007 | Sec31a | SEC31A | 2.99E-03 | -1.25E-02 |
| 13008 | Zeb2 | ZEB2 | -5.94E-03 | 2.60E-02 |
| 13009 | Rtn1 | RTN1 | -2.23E-02 | 5.83E-02 |
| 13010 | Ccr6 | CCR6 | 4.91E-03 | -6.68E-03 |
| 13011 | Prodh2 | PRODH2 | 5.06E-03 | 1.05E-02 |
| 13012 | Hdhd2 | HDHD2 | 1.77E-03 | -2.41E-02 |
| 13013 | Taf1 | TAF1 | 1.73E-03 | -6.36E-03 |
| 13014 | Ecsit | ECSIT | 2.20E-03 | -1.61E-02 |
| 13015 | Mri1 | MRI1 | -2.66E-03 | 2.20E-02 |
| 13016 | Polr2e | POLR2E | -1.79E-03 | -1.26E-02 |
| 13017 | Gpr143 | GPR143 | 8.09E-03 | -3.20E-02 |
| 13018 | Sbno1 | SBNO1 | -1.42E-03 | -1.21E-02 |
| 13019 | Slc9b2 | SLC9B2 | 1.12E-02 | 3.74E-02 |
| 13020 | Kcnj13 | KCNJ13 | 1.06E-02 | -4.13E-02 |
| 13021 | Ddhd1 | DDHD1 | 3.77E-03 | 3.02E-02 |
| 13022 | Maml3 | MAML3 | 3.78E-03 | -1.68E-02 |
| 13023 | Nob1 | NOB1 | 2.63E-03 | 2.26E-02 |
| 13024 | Thap7 | THAP7 | 2.41E-03 | -5.50E-03 |
| 13025 | Phka1 | PHKA1 | 2.60E-03 | -3.07E-02 |
| 13026 | Clk4 | CLK4 | -1.95E-03 | -8.77E-03 |
| 13027 | Phpt1 | PHPT1 | 6.48E-03 | 1.25E-02 |
| 13028 | Wdr4 | WDR4 | -2.20E-03 | 7.34E-03 |
| 13029 | Cenpa | CENPA | 1.49E-02 | 1.82E-01 |
| 13030 | Tmem218 | TMEM218 | 5.04E-03 | -3.56E-02 |
| 13031 | Lancl3 | LANCL3 | -8.15E-03 | -7.30E-02 |
| 13032 | Gapvd1 | GAPVD1 | -1.44E-03 | -3.37E-04 |
| 13033 | Pigw | PIGW | 4.42E-03 | 2.49E-02 |
| 13034 | Ecscr | ECSCR | -6.01E-03 | 5.96E-02 |
| 13035 | Fzd8 | FZD8 | -4.31E-03 | 7.46E-03 |

| | | | | |
|---|---|---|---|---|
| 13036 | Serinc2 | SERINC2 | 6.30E-03 | -3.15E-03 |
| 13037 | Tti1 | TTI1 | -2.11E-03 | -3.54E-03 |
| 13038 | Sphk2 | SPHK2 | -1.74E-03 | -1.33E-02 |
| 13039 | Rab13 | RAB13 | -3.58E-03 | 1.87E-02 |
| 13040 | Pdgfd | PDGFD | 7.00E-03 | -2.55E-02 |
| 13041 | Rcsd1 | RCSD1 | 4.38E-03 | -6.47E-03 |
| 13042 | Samd4b | SAMD4B | -1.99E-03 | -4.18E-03 |
| 13043 | Eya2 | EYA2 | -1.11E-02 | 2.17E-02 |
| 13044 | Map4k5 | MAP4K5 | 3.56E-03 | -1.33E-02 |
| 13045 | Stard13 | STARD13 | 7.89E-03 | -2.13E-02 |
| 13046 | Ptbp3 | PTBP3 | 2.10E-03 | 1.76E-02 |
| 13047 | Morn4 | MORN4 | 3.09E-03 | -3.04E-02 |
| 13048 | Shoc2 | SHOC2 | 2.04E-03 | -4.06E-03 |
| 13049 | Dchs1 | DCHS1 | 7.04E-03 | -1.28E-02 |
| 13050 | Smad4 | SMAD4 | -1.65E-03 | -9.43E-03 |
| 13051 | Slc22a8 | SLC22A8 | -5.46E-03 | -1.44E-02 |
| 13052 | Stx5a | STX5 | 1.50E-03 | -1.59E-02 |
| 13053 | Ano3 | ANO3 | 3.23E-03 | -3.35E-03 |
| 13054 | 4930562C15Rik | C16orf96 | -2.30E-03 | 3.16E-03 |
| 13055 | Zkscan6 | ZNF18 | -3.36E-03 | -4.84E-03 |
| 13056 | Zfp143 | ZNF143 | -3.71E-03 | -6.78E-03 |
| 13057 | Il12rb1 | IL12RB1 | 4.43E-03 | 1.42E-03 |
| 13058 | Tpr | TPR | -1.36E-03 | -5.43E-03 |
| 13059 | Hcfc2 | HCFC2 | 2.26E-03 | -1.22E-02 |
| 13060 | Atrnl1 | ATRNL1 | 2.93E-03 | -2.24E-02 |
| 13061 | Iqcg | IQCG | -3.58E-03 | -6.90E-02 |
| 13062 | Map2k7 | MAP2K7 | 1.58E-03 | -1.08E-02 |
| 13063 | Nr2c2 | NR2C2 | -2.54E-03 | -3.28E-02 |
| 13064 | Ch25h | CH25H | -1.24E-02 | 1.10E-01 |
| 13065 | Ift52 | IFT52 | 1.69E-03 | -1.36E-02 |
| 13066 | Apool | APOOL | -2.36E-03 | -1.75E-02 |
| 13067 | Dcaf8 | DCAF8 | 2.56E-03 | -1.91E-02 |
| 13068 | Lrrcc1 | LRRCC1 | 3.54E-03 | -3.13E-02 |
| 13069 | Gnmt | GNMT | -4.96E-03 | -3.44E-02 |
| 13070 | Sag | SAG | -6.29E-03 | -1.78E-02 |
| 13071 | Igbp1 | IGBP1 | -1.91E-03 | 9.73E-03 |
| 13072 | Cpsf6 | CPSF6 | -2.29E-03 | 1.39E-02 |
| 13073 | Snx18 | SNX18 | 2.30E-03 | 6.27E-04 |
| 13074 | Slc22a28 | SLC22A25 | -7.26E-03 | -2.00E-02 |
| 13075 | Fxyd3 | FXYD3 | -2.50E-02 | 1.67E-01 |
| 13076 | Btbd10 | BTBD10 | 4.73E-03 | 3.94E-02 |
| 13077 | Dync2li1 | DYNC2LI1 | -2.10E-03 | -1.85E-02 |
| 13078 | Topors | TOPORS | 2.16E-03 | -2.27E-03 |
| 13079 | Manf | MANF | 8.08E-03 | 7.18E-03 |
| 13080 | Slc12a1 | SLC12A1 | 5.93E-03 | -7.51E-02 |
| 13081 | Rims4 | RIMS4 | -2.74E-03 | 2.61E-02 |
| 13082 | Parp10 | PARP10 | 5.29E-03 | 2.89E-03 |
| 13083 | Flcn | FLCN | -2.82E-03 | -1.46E-02 |
| 13084 | Wrnip1 | WRNIP1 | -1.62E-03 | -1.01E-02 |
| 13085 | Rab4b | RAB4B | -5.78E-03 | 1.26E-02 |
| 13086 | Cib2 | CIB2 | -6.30E-03 | 1.35E-02 |
| 13087 | Xirp2 | XIRP2 | -2.13E-03 | 2.38E-02 |
| 13088 | Dctn5 | DCTN5 | -1.79E-03 | -3.88E-03 |

| | | | | |
|---|---|---|---|---|
| 13089 | Tekt2 | TEKT2 | -6.38E-03 | 6.19E-02 |
| 13090 | Pum2 | PUM2 | 1.76E-03 | -1.05E-02 |
| 13091 | Smg6 | SMG6 | -2.37E-03 | -1.95E-02 |
| 13092 | Zscan2 | ZSCAN2 | 4.24E-03 | -1.87E-02 |
| 13093 | Zc4h2 | ZC4H2 | 5.13E-03 | 1.99E-03 |
| 13094 | Tbx18 | TBX18 | 1.22E-02 | 6.71E-02 |
| 13095 | Parvb | PARVB | 4.86E-03 | 6.86E-03 |
| 13096 | Iqck | IQCK | -8.09E-03 | 2.30E-02 |
| 13097 | Lrrc40 | LRRC40 | 2.69E-03 | -1.22E-02 |
| 13098 | Hadhb | HADHB | 2.21E-03 | -2.37E-02 |
| 13099 | Hdac8 | HDAC8 | 3.18E-03 | -8.78E-03 |
| 13100 | Fam71d | FAM71D | 9.73E-03 | -5.34E-02 |
| 13101 | B230219D22Rik | C5orf24 | -1.71E-03 | -6.83E-06 |
| 13102 | Pet100 | PET100 | 2.25E-03 | 6.87E-03 |
| 13103 | Nub1 | NUB1 | -1.68E-03 | -1.35E-02 |
| 13104 | Sdf2l1 | SDF2L1 | -1.24E-02 | 1.82E-02 |
| 13105 | Mns1 | MNS1 | -6.86E-03 | 7.20E-02 |
| 13106 | Spdef | SPDEF | 4.73E-03 | 2.34E-02 |
| 13107 | Zfp712 | ZNF726 | -7.14E-03 | -6.55E-02 |
| 13108 | Lrch2 | LRCH2 | 5.93E-03 | -3.14E-02 |
| 13109 | Pitpnm3 | PITPNM3 | -5.07E-03 | 1.54E-02 |
| 13110 | AI597479 | C2orf49 | -2.79E-03 | 1.27E-02 |
| 13111 | Tmem51 | TMEM51 | -2.42E-03 | 1.36E-02 |
| 13112 | Scrn3 | SCRN3 | 1.93E-03 | -2.77E-02 |
| 13113 | Lemd2 | LEMD2 | 2.01E-03 | -9.06E-03 |
| 13114 | Tmem70 | TMEM70 | -2.07E-03 | -1.36E-02 |
| 13115 | Pira1 | LILRB3 | -3.23E-03 | 2.59E-02 |
| 13116 | Abhd2 | ABHD2 | -1.96E-03 | -1.41E-02 |
| 13117 | Cmtm3 | CMTM3 | -6.01E-03 | 1.01E-01 |
| 13118 | Basp1 | BASP1 | 8.25E-03 | 1.11E-01 |
| 13119 | Lgals2 | LGALS2 | -2.47E-03 | -2.18E-03 |
| 13120 | Cytl1 | CYTL1 | -1.23E-02 | 2.44E-02 |
| 13121 | Msantd4 | MSANTD4 | -1.91E-03 | -9.77E-03 |
| 13122 | Zfp184 | ZNF184 | -5.94E-03 | -4.08E-02 |
| 13123 | Gtf2a1 | GTF2A1 | -2.50E-03 | 7.47E-03 |
| 13124 | Palmd | PALMD | 5.28E-03 | 6.49E-02 |
| 13125 | Pcdhb5 | PCDHB4 | 5.06E-03 | 1.26E-02 |
| 13126 | Ceacam19 | CEACAM19 | 1.19E-02 | 2.05E-02 |
| 13127 | Anapc2 | ANAPC2 | 1.20E-03 | -1.35E-02 |
| 13128 | Fbxl6 | FBXL6 | -2.12E-03 | 4.05E-03 |
| 13129 | Mdga2 | MDGA2 | 2.10E-03 | 2.43E-03 |
| 13130 | Syngap1 | SYNGAP1 | -2.44E-03 | 8.23E-03 |
| 13131 | Fiz1 | FIZ1 | 1.67E-03 | -2.99E-03 |
| 13132 | Thoc1 | THOC1 | -1.47E-03 | 5.54E-03 |
| 13133 | Rnaseh2a | RNASEH2A | 2.50E-03 | 1.08E-02 |
| 13134 | Plod3 | PLOD3 | -1.51E-03 | 1.58E-02 |
| 13135 | Hhat | HHAT | -5.21E-03 | -6.83E-02 |
| 13136 | Meis3 | MEIS3 | -5.67E-03 | 8.55E-02 |
| 13137 | Fmo5 | FMO5 | 1.22E-02 | 1.25E-01 |
| 13138 | Aqr | AQR | 1.91E-03 | 1.39E-03 |
| 13139 | Pirt | PIRT | 1.28E-02 | -2.67E-02 |
| 13140 | Fign | FIGN | -5.06E-03 | -1.28E-02 |
| 13141 | Mmrn1 | MMRN1 | -9.80E-03 | -1.08E-01 |

| | | | | |
|---|---|---|---|---|
| 13142 | G6pc2 | G6PC2 | -4.63E-03 | -1.76E-02 |
| 13143 | Stk38l | STK38L | -2.56E-03 | 7.37E-03 |
| 13144 | Uhmk1 | UHMK1 | -1.70E-03 | -1.93E-02 |
| 13145 | D17H6S53E | C6orf47 | -2.78E-03 | -1.05E-02 |
| 13146 | Cwf19l2 | CWF19L2 | 2.61E-03 | -4.09E-02 |
| 13147 | Glyr1 | GLYR1 | 1.57E-03 | -2.12E-02 |
| 13148 | Tssc4 | TSSC4 | 1.98E-03 | -5.79E-04 |
| 13149 | Ophn1 | OPHN1 | 7.65E-03 | 1.26E-02 |
| 13150 | Kcnj9 | KCNJ9 | -4.44E-03 | 1.93E-03 |
| 13151 | Hydin | HYDIN | -3.66E-03 | 1.15E-02 |
| 13152 | Olfr195 | OR5K3 | -3.27E-03 | 2.20E-02 |
| 13153 | Cnot2 | CNOT2 | 1.41E-03 | -6.08E-04 |
| 13154 | Taf11 | TAF11 | 1.97E-03 | 4.71E-04 |
| 13155 | Mpo | MPO | -1.80E-03 | 2.00E-02 |
| 13156 | Rhof | RHOF | -1.23E-02 | -4.59E-02 |
| 13157 | Iah1 | IAH1 | -2.18E-03 | -1.88E-02 |
| 13158 | Cand1 | CAND1 | 1.56E-03 | 5.19E-04 |
| 13159 | Islr2 | ISLR2 | 9.62E-03 | 3.64E-02 |
| 13160 | Kcnh3 | KCNH3 | -7.52E-03 | 1.52E-02 |
| 13161 | Dnajc3 | DNAJC3 | 2.21E-03 | -2.03E-02 |
| 13162 | Hipk2 | HIPK2 | 3.85E-03 | -6.23E-02 |
| 13163 | Tmem184b | TMEM184B | 3.47E-03 | 4.29E-03 |
| 13164 | Ttc21a | TTC21A | 5.03E-03 | 1.38E-02 |
| 13165 | Wdr13 | WDR13 | -1.15E-03 | -8.12E-04 |
| 13166 | Dhx33 | DHX33 | -2.21E-03 | 2.08E-03 |
| 13167 | Gpn3 | GPN3 | -2.15E-03 | -1.07E-02 |
| 13168 | Foxa1 | FOXA1 | 2.91E-02 | 1.66E-01 |
| 13169 | Ckm | CKM | -2.11E-03 | 1.66E-02 |
| 13170 | Degs2 | DEGS2 | 4.48E-03 | 5.07E-02 |
| 13171 | Apoa1 | APOA1 | -1.33E-02 | 8.58E-02 |
| 13172 | Pacs2 | PACS2 | 1.55E-03 | -1.36E-02 |
| 13173 | D630039A03Rik | C9orf152 | 3.63E-03 | 3.98E-03 |
| 13174 | Vipr1 | VIPR1 | 1.07E-02 | -8.33E-02 |
| 13175 | Psmg4 | PSMG4 | 3.26E-03 | 1.91E-02 |
| 13176 | Acot13 | ACOT13 | 2.16E-03 | -8.82E-03 |
| 13177 | Akr7a5 | AKR7A2 | -2.83E-03 | -1.92E-02 |
| 13178 | Ubxn7 | UBXN7 | -1.94E-03 | -1.69E-02 |
| 13179 | Nkiras2 | NKIRAS2 | 1.74E-03 | -1.00E-02 |
| 13180 | Tatdn1 | TATDN1 | 2.18E-03 | 8.21E-03 |
| 13181 | Arhgap31 | ARHGAP31 | -2.48E-03 | -2.78E-02 |
| 13182 | Tc2n | TC2N | -5.43E-03 | 1.89E-03 |
| 13183 | Trmt61a | TRMT61A | 3.52E-03 | 7.46E-02 |
| 13184 | Cd53 | CD53 | -7.22E-03 | 1.26E-01 |
| 13185 | Sgsh | SGSH | -2.28E-03 | -2.66E-02 |
| 13186 | Vim | VIM | -3.77E-03 | 1.19E-01 |
| 13187 | Rln1 | RLN1 | 4.97E-03 | 7.69E-02 |
| 13188 | Pomt2 | POMT2 | -1.86E-03 | -2.06E-03 |
| 13189 | Arhgap42 | ARHGAP42 | -2.85E-03 | -8.55E-03 |
| 13190 | Nup210l | NUP210L | 8.22E-03 | 6.96E-03 |
| 13191 | Il22ra1 | IL22RA1 | -1.01E-02 | -4.53E-02 |
| 13192 | Clta | CLTA | -1.56E-03 | -4.75E-04 |
| 13193 | Bloc1s1 | BLOC1S1 | -6.16E-03 | -3.71E-02 |
| 13194 | Tspan12 | TSPAN12 | -5.00E-03 | -2.46E-02 |

| | | | | |
|---|---|---|---|---|
| 13195 | Slc34a3 | SLC34A3 | -5.88E-03 | -8.06E-02 |
| 13196 | Tet3 | TET3 | 1.89E-03 | -1.82E-02 |
| 13197 | Tmc8 | TMC8 | -6.25E-03 | -7.66E-03 |
| 13198 | Gpr88 | GPR88 | -2.83E-03 | 1.59E-02 |
| 13199 | Gpr85 | GPR85 | 6.92E-03 | -4.44E-02 |
| 13200 | Rdh11 | RDH11 | 2.12E-03 | 1.79E-02 |
| 13201 | Aftph | AFTPH | 1.48E-03 | -1.36E-02 |
| 13202 | Fbxo42 | FBXO42 | 1.96E-03 | 2.12E-03 |
| 13203 | Dhx37 | DHX37 | 1.55E-03 | -2.81E-04 |
| 13204 | Triobp | TRIOBP | -1.60E-03 | 2.27E-02 |
| 13205 | Itgad | ITGAD | 4.55E-03 | 3.38E-02 |
| 13206 | Arg1 | ARG1 | -1.13E-02 | 1.93E-01 |
| 13207 | Bst1 | BST1 | 2.22E-02 | -4.81E-02 |
| 13208 | Polr2m | POLR2M | -1.25E-03 | -1.76E-02 |
| 13209 | Cdk2ap2 | CDK2AP2 | -3.11E-03 | 1.50E-03 |
| 13210 | Fam50a | FAM50A | -1.69E-03 | 9.57E-03 |
| 13211 | Slco1b2 | SLCO1B3 | 6.28E-03 | 5.75E-02 |
| 13212 | Etv1 | ETV1 | 8.49E-03 | -2.08E-03 |
| 13213 | Asphd2 | ASPHD2 | 4.15E-03 | -4.42E-03 |
| 13214 | Ackr4 | ACKR4 | 3.76E-03 | 1.29E-02 |
| 13215 | Qprt | QPRT | -3.25E-03 | -1.15E-02 |
| 13216 | Lifr | LIFR | -1.98E-03 | -2.37E-02 |
| 13217 | Rpl23 | RPL23 | 1.52E-03 | 1.65E-02 |
| 13218 | Fam126b | FAM126B | -3.29E-03 | -2.16E-03 |
| 13219 | Fam180a | FAM180A | -4.15E-03 | 3.89E-02 |
| 13220 | Irak4 | IRAK4 | 3.37E-03 | -2.51E-03 |
| 13221 | Scn1a | SCN1A | 2.47E-03 | -9.60E-03 |
| 13222 | Gpr179 | GPR179 | -5.54E-03 | -3.60E-02 |
| 13223 | Klhl13 | KLHL13 | 4.33E-03 | -5.67E-03 |
| 13224 | Pitx1 | PITX1 | 2.72E-03 | -1.04E-02 |
| 13225 | Ubald1 | UBALD1 | -2.75E-03 | 1.22E-02 |
| 13226 | Brinp2 | BRINP2 | 3.23E-03 | 5.50E-02 |
| 13227 | Cdpf1 | CDPF1 | 4.36E-03 | -6.11E-02 |
| 13228 | Adnp | ADNP | -5.74E-03 | 2.13E-02 |
| 13229 | Dhcr24 | DHCR24 | 3.95E-03 | -1.71E-02 |
| 13230 | Man2b2 | MAN2B2 | 2.73E-03 | -1.13E-02 |
| 13231 | Lyar | LYAR | 1.93E-03 | 1.05E-02 |
| 13232 | C2cd3 | C2CD3 | -1.67E-03 | -5.97E-03 |
| 13233 | Ngfr | NGFR | 7.18E-03 | 1.36E-02 |
| 13234 | Pabpc1l | PABPC1L | -7.35E-03 | -2.40E-03 |
| 13235 | Arv1 | ARV1 | 3.70E-03 | 7.06E-03 |
| 13236 | Ablim2 | ABLIM2 | -4.31E-03 | -2.77E-02 |
| 13237 | Ubr2 | UBR2 | 1.47E-03 | -1.13E-02 |
| 13238 | Ttc17 | TTC17 | 1.49E-03 | 5.92E-03 |
| 13239 | Fcrlb | FCRLB | -2.34E-03 | 8.72E-03 |
| 13240 | Atad2b | ATAD2B | -1.93E-03 | -2.26E-03 |
| 13241 | Cyp3a11 | CYP3A5 | -1.57E-02 | 2.14E-01 |
| 13242 | Cdc7 | CDC7 | 9.62E-03 | 9.53E-02 |
| 13243 | Angpt1 | ANGPT1 | -4.41E-03 | -1.27E-02 |
| 13244 | Cacng7 | CACNG7 | -5.56E-03 | 1.01E-02 |
| 13245 | Megf6 | MEGF6 | 9.83E-03 | 4.47E-02 |
| 13246 | Ppp6r1 | PPP6R1 | -1.58E-03 | -1.14E-02 |
| 13247 | Gpr75 | GPR75 | -3.46E-03 | 1.58E-02 |

| | | | | |
|---|---|---|---|---|
| 13248 | Sdcbp2 | SDCBP2 | 8.63E-03 | 6.07E-02 |
| 13249 | Cyp11a1 | CYP11A1 | 2.18E-03 | 6.15E-03 |
| 13250 | Med11 | MED11 | -2.16E-03 | 1.21E-02 |
| 13251 | Styk1 | STYK1 | 2.26E-02 | 8.59E-02 |
| 13252 | Rspo3 | RSPO3 | 7.10E-03 | 2.85E-02 |
| 13253 | Pak2 | PAK2 | 1.18E-03 | 4.17E-03 |
| 13254 | Vangl1 | VANGL1 | -2.69E-03 | -2.31E-02 |
| 13255 | Pax9 | PAX9 | -2.14E-03 | 9.42E-03 |
| 13256 | Xrcc6 | XRCC6 | -1.53E-03 | -1.63E-02 |
| 13257 | Rrn3 | RRN3 | 1.31E-03 | 1.18E-02 |
| 13258 | Tmod1 | TMOD1 | -5.98E-03 | 4.88E-02 |
| 13259 | Dgcr8 | DGCR8 | 2.66E-03 | 1.11E-02 |
| 13260 | Akr1c6 | AKR1C4 | -5.42E-03 | 5.39E-02 |
| 13261 | C5ar2 | C5AR2 | -6.24E-03 | 3.39E-02 |
| 13262 | Tbx4 | TBX4 | 2.47E-03 | -1.54E-02 |
| 13263 | Lzts1 | LZTS1 | -5.66E-03 | -6.21E-02 |
| 13264 | Tm4sf5 | TM4SF5 | -5.44E-03 | -5.23E-02 |
| 13265 | Aven | AVEN | 2.44E-03 | 2.25E-02 |
| 13266 | Pde1a | PDE1A | 2.42E-03 | -3.24E-02 |
| 13267 | Tead4 | TEAD4 | -5.69E-03 | -3.54E-02 |
| 13268 | Ugt8a | UGT8 | -1.29E-02 | -4.99E-03 |
| 13269 | Pcdh18 | PCDH18 | -9.48E-03 | -1.20E-02 |
| 13270 | Mzt2 | MZT2A | -2.92E-03 | -1.78E-02 |
| 13271 | Klf15 | KLF15 | 3.61E-03 | -4.92E-02 |
| 13272 | Map3k1 | MAP3K1 | 2.02E-03 | 2.03E-02 |
| 13273 | Dclre1c | DCLRE1C | -2.57E-03 | 1.23E-02 |
| 13274 | Polr2d | POLR2D | -2.14E-03 | 2.28E-02 |
| 13275 | Tmed4 | TMED4 | 1.18E-03 | -2.78E-03 |
| 13276 | Elmod3 | ELMOD3 | -3.23E-03 | -1.54E-02 |
| 13277 | Rgs7bp | RGS7BP | -5.93E-03 | 4.89E-02 |
| 13278 | Mfsd8 | MFSD8 | -2.21E-03 | -8.34E-03 |
| 13279 | Echdc1 | ECHDC1 | 2.13E-03 | -1.35E-02 |
| 13280 | Ccdc142 | CCDC142 | 2.11E-03 | -3.26E-02 |
| 13281 | Smc4 | SMC4 | 3.10E-03 | 4.21E-02 |
| 13282 | Fam107b | FAM107B | -1.96E-03 | -1.14E-02 |
| 13283 | Crlf2 | CRLF2 | 4.57E-03 | 8.26E-02 |
| 13284 | Vamp3 | VAMP3 | -1.51E-03 | 1.74E-02 |
| 13285 | Hars2 | HARS2 | -1.49E-03 | -1.26E-02 |
| 13286 | Abcd2 | ABCD2 | -9.45E-03 | 3.95E-02 |
| 13287 | Rpl35 | RPL35 | 1.54E-03 | 9.03E-03 |
| 13288 | Golga4 | GOLGA4 | -1.56E-03 | -1.60E-02 |
| 13289 | Myc | MYC | 6.50E-03 | 1.74E-01 |
| 13290 | Fam43a | FAM43A | -9.10E-03 | -5.57E-02 |
| 13291 | Ppp1r3f | PPP1R3F | -4.78E-03 | 4.95E-02 |
| 13292 | Pde4b | PDE4B | -3.60E-03 | 1.17E-03 |
| 13293 | Hnrnpf | HNRNPF | -1.36E-03 | 1.28E-02 |
| 13294 | Cgrrf1 | CGRRF1 | -1.63E-03 | -3.48E-03 |
| 13295 | Ube2h | UBE2H | 1.52E-03 | -4.54E-03 |
| 13296 | Secisbp2 | SECISBP2 | 1.65E-03 | -1.51E-02 |
| 13297 | Pbld1 | PBLD | 2.86E-03 | -3.11E-02 |
| 13298 | Ddx17 | DDX17 | -1.11E-03 | -3.51E-03 |
| 13299 | Eef1e1 | EEF1E1 | 2.25E-03 | 3.46E-02 |
| 13300 | Rwdd2b | RWDD2B | 2.03E-03 | -7.41E-03 |

| | | | | |
|---|---|---|---|---|
| 13301 | Fpr1 | FPR1 | 1.26E-02 | 6.64E-02 |
| 13302 | 1700016K19Rik | C17orf97 | 2.87E-03 | 1.61E-02 |
| 13303 | Myh3 | MYH3 | -2.50E-03 | 1.54E-02 |
| 13304 | Luc7l2 | LUC7L2 | 1.32E-03 | -1.23E-02 |
| 13305 | Defb19 | DEFB119 | 5.30E-03 | 7.12E-02 |
| 13306 | Palb2 | PALB2 | 5.14E-03 | 2.11E-02 |
| 13307 | Omp | OMP | 5.98E-03 | 6.08E-02 |
| 13308 | Phf6 | PHF6 | -4.39E-03 | 2.80E-03 |
| 13309 | St14 | ST14 | -3.41E-03 | -4.68E-03 |
| 13310 | Zfp384 | ZNF384 | 1.11E-03 | -4.08E-03 |
| 13311 | Rps6kb1 | RPS6KB1 | -1.21E-03 | 3.31E-03 |
| 13312 | Ncf1 | NCF1 | -6.69E-03 | 1.63E-01 |
| 13313 | Hsd17b13 | HSD17B13 | 4.84E-03 | 4.06E-02 |
| 13314 | Dlx5 | DLX5 | 4.00E-03 | 2.80E-02 |
| 13315 | Slc7a6os | SLC7A6OS | -1.85E-03 | 2.23E-03 |
| 13316 | Slc10a7 | SLC10A7 | -1.63E-03 | 4.46E-03 |
| 13317 | Trpm1 | TRPM1 | -1.29E-02 | -4.64E-02 |
| 13318 | Slc34a2 | SLC34A2 | -8.03E-03 | 1.01E-01 |
| 13319 | Rbbp6 | RBBP6 | -1.35E-03 | 7.34E-04 |
| 13320 | Syncrip | SYNCRIP | -1.28E-03 | 9.80E-03 |
| 13321 | Golph3l | GOLPH3L | 1.58E-03 | -1.76E-02 |
| 13322 | Nfkbid | NFKBID | -6.61E-03 | 5.94E-02 |
| 13323 | Cxcl2 | CXCL3 | 1.32E-02 | 4.11E-01 |
| 13324 | Gpbp1l1 | GPBP1L1 | -9.93E-04 | -1.36E-02 |
| 13325 | Bcap31 | BCAP31 | -1.24E-03 | -5.67E-03 |
| 13326 | Rnf6 | RNF6 | -1.58E-03 | -2.00E-03 |
| 13327 | Vezt | VEZT | -1.43E-03 | 1.66E-02 |
| 13328 | Mbd6 | MBD6 | 1.44E-03 | 8.15E-04 |
| 13329 | Clasp2 | CLASP2 | 1.51E-03 | -8.42E-03 |
| 13330 | Yipf6 | YIPF6 | 1.24E-03 | -6.75E-03 |
| 13331 | Taf15 | TAF15 | 1.14E-03 | 1.20E-03 |
| 13332 | Cxcl5 | CXCL6 | -4.19E-03 | 1.63E-01 |
| 13333 | Bud13 | BUD13 | 1.77E-03 | -4.30E-03 |
| 13334 | Zbtb2 | ZBTB2 | 2.45E-03 | 1.30E-02 |
| 13335 | Kng1 | KNG1 | 6.77E-03 | 1.03E-01 |
| 13336 | Utp23 | UTP23 | 3.16E-03 | 2.86E-02 |
| 13337 | Dohh | DOHH | -3.04E-03 | 2.73E-02 |
| 13338 | Mrpl34 | MRPL34 | -1.94E-03 | -2.77E-03 |
| 13339 | Rab25 | RAB25 | -4.40E-03 | 8.40E-03 |
| 13340 | Eddm3b | EDDM3B | 1.05E-02 | -9.62E-03 |
| 13341 | Cby1 | CBY1 | -1.51E-03 | -5.83E-03 |
| 13342 | Hoxc9 | HOXC9 | -2.64E-03 | 3.82E-02 |
| 13343 | Med19 | MED19 | 1.65E-03 | 1.32E-02 |
| 13344 | Crtc3 | CRTC3 | 1.96E-03 | -4.64E-06 |
| 13345 | Cd63 | CD63 | 1.66E-03 | 1.02E-02 |
| 13346 | Nxt2 | NXT2 | -2.54E-03 | -1.54E-03 |
| 13347 | Trafd1 | TRAFD1 | -1.71E-03 | 6.56E-03 |
| 13348 | Lrrc32 | LRRC32 | -4.91E-03 | 3.26E-02 |
| 13349 | Cct8l1 | CCT8L2 | 1.65E-03 | -1.04E-02 |
| 13350 | Ubap1 | UBAP1 | -1.20E-03 | 9.32E-03 |
| 13351 | Rgs9 | RGS9 | -1.12E-02 | -9.42E-02 |
| 13352 | Taf3 | TAF3 | -1.66E-03 | -4.39E-03 |
| 13353 | Xrcc5 | XRCC5 | -1.80E-03 | -2.39E-02 |

| | | | | |
|---|---|---|---|---|
| 13354 | Htra4 | HTRA4 | 5.96E-03 | -6.01E-02 |
| 13355 | Maf | MAF | 4.64E-03 | -8.04E-03 |
| 13356 | Insig1 | INSIG1 | 5.22E-03 | -3.77E-02 |
| 13357 | Fn1 | FN1 | -5.35E-03 | 7.86E-02 |
| 13358 | Dctn4 | DCTN4 | 9.33E-04 | 2.33E-03 |
| 13359 | Spic | SPIC | -4.55E-03 | 4.40E-02 |
| 13360 | Cep152 | CEP152 | -3.26E-03 | 1.37E-02 |
| 13361 | Ablim1 | ABLIM1 | 1.16E-03 | -1.48E-02 |
| 13362 | Pkig | PKIG | -1.41E-03 | 1.30E-02 |
| 13363 | Sbk2 | SBK2 | 2.50E-03 | 1.73E-02 |
| 13364 | Alx1 | ALX1 | 1.63E-02 | -5.52E-02 |
| 13365 | Ciao1 | CIAO1 | 1.22E-03 | -1.78E-02 |
| 13366 | Mapk15 | MAPK15 | -2.66E-03 | -6.08E-03 |
| 13367 | Acsf3 | ACSF3 | -1.75E-03 | -2.26E-02 |
| 13368 | BC016579 | C3orf52 | -7.43E-03 | 4.63E-02 |
| 13369 | Aldh1l1 | ALDH1L1 | -1.99E-03 | -1.82E-02 |
| 13370 | Pira2 | LILRB3 | -2.69E-03 | 3.23E-02 |
| 13371 | Slc7a7 | SLC7A7 | 3.27E-03 | 2.59E-03 |
| 13372 | Capn10 | CAPN10 | 1.40E-03 | -4.26E-03 |
| 13373 | Rras2 | RRAS2 | 1.58E-03 | 3.02E-02 |
| 13374 | Tmigd1 | TMIGD1 | 3.36E-03 | -3.67E-02 |
| 13375 | Mmp24 | MMP24 | -2.18E-03 | 2.26E-02 |
| 13376 | Barhl1 | BARHL1 | 2.50E-03 | -1.40E-02 |
| 13377 | Lrrc36 | LRRC36 | -3.44E-03 | 2.23E-02 |
| 13378 | Dolk | DOLK | 1.53E-03 | -1.66E-02 |
| 13379 | Accsl | ACCSL | -2.61E-03 | -6.83E-03 |
| 13380 | Rptn | RPTN | -3.13E-03 | -3.34E-03 |
| 13381 | Rps19 | RPS19 | 1.67E-03 | 1.46E-02 |
| 13382 | Atf1 | ATF1 | 1.30E-03 | -4.70E-04 |
| 13383 | Katnal2 | KATNAL2 | 5.29E-03 | 2.42E-02 |
| 13384 | Nudt22 | NUDT22 | 1.77E-03 | -7.69E-04 |
| 13385 | Asb1 | ASB1 | -1.68E-03 | -2.06E-02 |
| 13386 | Slc1a7 | SLC1A7 | 2.92E-03 | -2.22E-02 |
| 13387 | Alox5ap | ALOX5AP | -5.76E-03 | 1.55E-01 |
| 13388 | Grn | GRN | -1.19E-03 | 1.54E-02 |
| 13389 | Trim11 | TRIM11 | 1.18E-03 | 5.09E-04 |
| 13390 | Agap1 | AGAP1 | 1.58E-03 | -2.93E-03 |
| 13391 | Smarcc2 | SMARCC2 | -1.25E-03 | -1.29E-02 |
| 13392 | Gng8 | GNG8 | 4.31E-03 | 1.27E-02 |
| 13393 | Bola1 | BOLA1 | 1.91E-03 | 8.54E-04 |
| 13394 | Dscc1 | DSCC1 | 5.88E-03 | 8.76E-02 |
| 13395 | Sdr9c7 | SDR9C7 | -2.10E-03 | 2.03E-02 |
| 13396 | Emp2 | EMP2 | -3.57E-03 | 5.85E-02 |
| 13397 | Rsad1 | RSAD1 | 1.54E-03 | -3.08E-02 |
| 13398 | Clstn3 | CLSTN3 | 4.53E-03 | 1.51E-03 |
| 13399 | Ep400 | EP400 | -1.08E-03 | -3.39E-03 |
| 13400 | Itm2a | ITM2A | 7.92E-03 | -6.79E-03 |
| 13401 | Hmbox1 | HMBOX1 | 9.17E-04 | -1.71E-02 |
| 13402 | Nrip3 | NRIP3 | 2.36E-02 | 1.18E-01 |
| 13403 | Hbq1b | HBQ1 | -1.96E-03 | -2.12E-03 |
| 13404 | Asgr2 | ASGR2 | -4.66E-03 | 2.87E-02 |
| 13405 | Arl11 | ARL11 | 4.94E-03 | 5.03E-02 |
| 13406 | Vps51 | VPS51 | -1.26E-03 | -7.29E-03 |

| | | | | |
|---|---|---|---|---|
| 13407 | Krt5 | KRT5 | 3.10E-02 | 2.06E-01 |
| 13408 | Pon1 | PON1 | -4.61E-03 | 7.50E-02 |
| 13409 | Ncam2 | NCAM2 | -5.51E-03 | -3.00E-02 |
| 13410 | Parm1 | PARM1 | -5.08E-03 | 3.41E-03 |
| 13411 | Nrip1 | NRIP1 | 2.22E-03 | -6.84E-05 |
| 13412 | Flt4 | FLT4 | -3.08E-03 | -9.86E-03 |
| 13413 | Abl2 | ABL2 | -1.54E-03 | -1.55E-02 |
| 13414 | Dgkh | DGKH | 3.93E-03 | 1.17E-02 |
| 13415 | Smyd3 | SMYD3 | 2.20E-03 | -4.49E-02 |
| 13416 | Actc1 | ACTC1 | -1.95E-03 | 5.01E-03 |
| 13417 | 1700020L24Rik | C17orf50 | 1.25E-03 | 6.01E-03 |
| 13418 | Cstf1 | CSTF1 | 1.23E-03 | -4.83E-03 |
| 13419 | Klk1b4 | KLK3 | -3.49E-03 | 1.27E-01 |
| 13420 | Mob4 | MOB4 | -1.47E-03 | 3.03E-03 |
| 13421 | Ctu1 | CTU1 | -2.73E-03 | -7.87E-03 |
| 13422 | Cyp2a5 | CYP2A6 | 1.23E-02 | -2.21E-02 |
| 13423 | Cyp2a5 | CYP2A7 | 1.23E-02 | -2.21E-02 |
| 13424 | Cyp2a5 | CYP2A13 | 1.23E-02 | -2.21E-02 |
| 13425 | Zfyve21 | ZFYVE21 | 1.39E-03 | -2.56E-02 |
| 13426 | Cass4 | CASS4 | 3.82E-03 | 1.12E-02 |
| 13427 | Zdhhc7 | ZDHHC7 | -1.09E-03 | 7.19E-03 |
| 13428 | Igdcc4 | IGDCC4 | 6.54E-03 | 7.54E-02 |
| 13429 | Cd5l | CD5L | 1.55E-03 | 2.20E-02 |
| 13430 | Rnase1 | RNASE1 | -2.27E-03 | -3.29E-03 |
| 13431 | Mcrs1 | MCRS1 | -1.23E-03 | -6.99E-03 |
| 13432 | Krt1 | KRT1 | -1.57E-03 | 5.25E-04 |
| 13433 | Gfpt1 | GFPT1 | -1.30E-03 | -1.79E-02 |
| 13434 | Syt12 | SYT12 | 5.72E-03 | 2.21E-01 |
| 13435 | Nucb1 | NUCB1 | -9.16E-04 | -7.12E-03 |
| 13436 | Gramd1b | GRAMD1B | -2.19E-03 | -4.31E-02 |
| 13437 | Matn4 | MATN4 | 5.75E-03 | -5.97E-02 |
| 13438 | Sowahd | SOWAHD | -2.11E-03 | -1.50E-02 |
| 13439 | Ube2ql1 | UBE2QL1 | -6.96E-03 | -2.23E-02 |
| 13440 | Fam120a | FAM120A | 1.21E-03 | -2.49E-03 |
| 13441 | Moxd1 | MOXD1 | 3.21E-03 | 8.13E-02 |
| 13442 | Tmem219 | TMEM219 | 1.50E-03 | 8.04E-03 |
| 13443 | Slc16a2 | SLC16A2 | 2.82E-03 | -3.17E-03 |
| 13444 | Abcb6 | ABCB6 | -4.46E-03 | 3.09E-02 |
| 13445 | Adamts7 | ADAMTS7 | -4.78E-03 | -3.81E-02 |
| 13446 | Zfp821 | ZNF821 | -1.27E-03 | -2.35E-03 |
| 13447 | Cbfa2t2 | CBFA2T2 | -1.12E-03 | -1.57E-02 |
| 13448 | Oaz3 | OAZ3 | 1.72E-03 | -4.66E-03 |
| 13449 | Lta4h | LTA4H | -1.33E-03 | -3.00E-03 |
| 13450 | Hr | HR | 7.78E-03 | 1.23E-01 |
| 13451 | Pabpn1l | PABPN1L | -4.89E-03 | -3.63E-02 |
| 13452 | Arfip1 | ARFIP1 | -2.94E-03 | 1.40E-02 |
| 13453 | Mier3 | MIER3 | 4.04E-03 | 3.45E-03 |
| 13454 | Bet1 | BET1 | -1.40E-03 | -5.47E-03 |
| 13455 | Slc36a1 | SLC36A1 | -2.38E-03 | -1.76E-02 |
| 13456 | Gmpr2 | GMPR2 | 1.53E-03 | -1.23E-02 |
| 13457 | St7l | ST7L | 1.60E-03 | -1.64E-02 |
| 13458 | 6820408C15Rik | C20orf96 | -6.92E-03 | -7.04E-02 |
| 13459 | Rexo4 | REXO4 | -1.11E-03 | 1.17E-02 |

| | | | | |
|---|---|---|---|---|
| 13460 | Rabggta | RABGGTA | 1.51E-03 | 8.37E-03 |
| 13461 | Exo1 | EXO1 | 6.58E-03 | 1.98E-01 |
| 13462 | Dusp28 | DUSP28 | 1.98E-03 | -8.33E-03 |
| 13463 | Nup37 | NUP37 | 2.34E-03 | -9.52E-03 |
| 13464 | Sox15 | SOX15 | 4.34E-03 | 4.59E-02 |
| 13465 | Tvp23a | TVP23A | 3.18E-03 | 3.98E-02 |
| 13466 | Slc35a3 | SLC35A3 | 1.07E-03 | -1.26E-02 |
| 13467 | Tmem158 | TMEM158 | 1.13E-02 | 1.26E-01 |
| 13468 | Pcdh10 | PCDH10 | 1.56E-03 | 8.56E-03 |
| 13469 | Zkscan14 | ZNF394 | 2.93E-03 | -2.55E-02 |
| 13470 | Apof | APOF | -2.89E-03 | 3.22E-02 |
| 13471 | Reps2 | REPS2 | -1.70E-03 | -1.23E-02 |
| 13472 | Smyd1 | SMYD1 | 4.34E-03 | -1.83E-03 |
| 13473 | Atrx | ATRX | 1.11E-03 | -1.74E-02 |
| 13474 | Blcap | BLCAP | 1.15E-03 | 6.41E-03 |
| 13475 | Vsig2 | VSIG2 | 9.54E-03 | -9.64E-03 |
| 13476 | Zfp37 | ZFP37 | -3.98E-03 | -2.88E-02 |
| 13477 | Slc35e3 | SLC35E3 | 2.06E-03 | 1.77E-02 |
| 13478 | Tas2r108 | TAS2R4 | 1.64E-03 | -1.52E-02 |
| 13479 | Zfp575 | ZNF575 | 3.15E-03 | 5.91E-03 |
| 13480 | Gckr | GCKR | 3.02E-03 | -5.42E-03 |
| 13481 | Gfpt2 | GFPT2 | -5.34E-03 | -2.97E-02 |
| 13482 | Acbd6 | ACBD6 | 1.16E-03 | -2.07E-03 |
| 13483 | Syne1 | SYNE1 | 1.24E-03 | -3.14E-02 |
| 13484 | Mrgbp | MRGBP | -1.76E-03 | -6.34E-03 |
| 13485 | Ttpal | TTPAL | -1.45E-03 | -2.13E-03 |
| 13486 | Otop1 | OTOP1 | -2.67E-03 | 6.39E-03 |
| 13487 | Gemin7 | GEMIN7 | 9.56E-04 | -7.88E-03 |
| 13488 | 9430015G10Rik | C1orf159 | 3.03E-03 | 2.37E-02 |
| 13489 | Thsd7b | THSD7B | -2.06E-03 | 5.91E-03 |
| 13490 | Rps10 | RPS10 | -1.13E-03 | 2.85E-03 |
| 13491 | Desi2 | DESI2 | -3.56E-03 | -2.62E-02 |
| 13492 | Nit2 | NIT2 | 1.65E-03 | -1.08E-02 |
| 13493 | Stoml1 | STOML1 | -1.34E-03 | -5.41E-03 |
| 13494 | Cldn8 | CLDN8 | -2.99E-03 | 4.51E-03 |
| 13495 | Tmem240 | TMEM240 | 3.82E-03 | 1.68E-02 |
| 13496 | Abcc10 | ABCC10 | 1.66E-03 | -2.17E-02 |
| 13497 | Klc3 | KLC3 | -3.89E-03 | 3.65E-02 |
| 13498 | Snrpd2 | SNRPD2 | -1.10E-03 | 2.02E-03 |
| 13499 | Cdh16 | CDH16 | -8.66E-04 | -1.44E-02 |
| 13500 | Sez6l2 | SEZ6L2 | -5.02E-03 | 6.09E-03 |
| 13501 | Fcgbp | FCGBP | -2.92E-02 | 1.44E-01 |
| 13502 | P2ry12 | P2RY12 | 3.63E-03 | 6.26E-02 |
| 13503 | Ccdc89 | CCDC89 | -3.44E-03 | -1.61E-03 |
| 13504 | Nfkb2 | NFKB2 | -2.01E-03 | 6.24E-02 |
| 13505 | Chga | CHGA | 1.91E-03 | 3.06E-02 |
| 13506 | Inpp5k | INPP5K | 1.24E-03 | -1.83E-02 |
| 13507 | Sardh | SARDH | -2.33E-03 | -1.54E-02 |
| 13508 | Slco1a5 | SLCO1A2 | 1.06E-03 | 4.26E-03 |
| 13509 | Bend3 | BEND3 | 1.54E-03 | 7.44E-03 |
| 13510 | Vgf | VGF | -1.52E-03 | 4.04E-02 |
| 13511 | Abcc1 | ABCC1 | 1.93E-03 | -2.22E-02 |
| 13512 | Noxo1 | NOXO1 | -4.66E-03 | 1.82E-02 |

| | | | | |
|---|---|---|---|---|
| 13513 | Rps23 | RPS23 | -1.15E-03 | 5.92E-03 |
| 13514 | Rgmb | RGMB | -3.25E-03 | -3.51E-02 |
| 13515 | Nedd1 | NEDD1 | -1.37E-03 | 8.83E-03 |
| 13516 | Cpne5 | CPNE5 | -7.87E-03 | 3.39E-02 |
| 13517 | Pgap1 | PGAP1 | 2.16E-03 | -8.86E-03 |
| 13518 | Pkmyt1 | PKMYT1 | -6.25E-03 | 1.90E-01 |
| 13519 | Stoml2 | STOML2 | 1.17E-03 | -6.32E-03 |
| 13520 | Hnrnpr | HNRNPR | 1.70E-03 | -1.44E-02 |
| 13521 | Ptpre | PTPRE | 2.92E-03 | 2.98E-02 |
| 13522 | Chtf18 | CHTF18 | 6.90E-03 | 1.59E-01 |
| 13523 | Pelp1 | PELP1 | 1.08E-03 | -9.26E-04 |
| 13524 | Aloxe3 | ALOXE3 | -2.38E-03 | 8.97E-03 |
| 13525 | Esr2 | ESR2 | 4.99E-03 | -3.71E-02 |
| 13526 | Defb29 | DEFB116 | -2.88E-03 | -6.83E-02 |
| 13527 | Fam71f2 | FAM71F2 | 1.84E-03 | 4.96E-04 |
| 13528 | Sbk1 | SBK1 | -2.02E-03 | -6.42E-03 |
| 13529 | Sirt7 | SIRT7 | -9.31E-04 | -1.16E-03 |
| 13530 | Mpv17 | MPV17 | 1.14E-03 | -4.13E-03 |
| 13531 | Usp42 | USP42 | 1.35E-03 | 1.36E-02 |
| 13532 | Tufm | TUFM | 1.26E-03 | -2.20E-02 |
| 13533 | Saa2 | SAA1 | 1.05E-03 | 2.93E-02 |
| 13534 | Rxfp2 | RXFP2 | -1.76E-03 | -2.14E-02 |
| 13535 | Camp | CAMP | 2.51E-03 | 1.86E-02 |
| 13536 | Musk | MUSK | 2.00E-03 | -3.48E-03 |
| 13537 | Scin | SCIN | 4.74E-03 | -7.64E-03 |
| 13538 | Fut1 | FUT1 | -1.64E-03 | -9.54E-04 |
| 13539 | Fuca2 | FUCA2 | -1.10E-03 | -2.01E-02 |
| 13540 | Atf7 | ATF7 | -9.63E-04 | 2.75E-03 |
| 13541 | Zfc3h1 | ZFC3H1 | -1.01E-03 | 1.80E-02 |
| 13542 | Myh10 | MYH10 | 1.33E-03 | -1.88E-02 |
| 13543 | Myocd | MYOCD | 3.32E-03 | 7.31E-02 |
| 13544 | Gcdh | GCDH | -1.75E-03 | -2.35E-02 |
| 13545 | Sestd1 | SESTD1 | -1.00E-03 | -1.34E-02 |
| 13546 | Esyt3 | ESYT3 | 5.15E-03 | -2.85E-03 |
| 13547 | Kcnt1 | KCNT1 | -1.12E-02 | -1.16E-01 |
| 13548 | Stam | STAM | 9.53E-04 | 1.93E-02 |
| 13549 | Dsel | DSEL | 2.29E-03 | 3.91E-03 |
| 13550 | Txn2 | TXN2 | -1.05E-03 | -1.27E-02 |
| 13551 | Plcd1 | PLCD1 | -2.32E-03 | 1.75E-02 |
| 13552 | Gpaa1 | GPAA1 | 9.49E-04 | 6.51E-03 |
| 13553 | Zfr | ZFR | -7.96E-04 | 6.21E-03 |
| 13554 | Anks3 | ANKS3 | -8.38E-04 | -1.95E-02 |
| 13555 | Ddx39b | DDX39B | 8.03E-04 | -5.09E-03 |
| 13556 | Sppl2b | SPPL2B | 8.98E-04 | 1.18E-02 |
| 13557 | Mif4gd | MIF4GD | -1.43E-03 | 1.89E-02 |
| 13558 | Nin | NIN | 1.88E-03 | -1.66E-03 |
| 13559 | Mthfd2 | MTHFD2 | -4.58E-03 | 1.40E-01 |
| 13560 | Gpr45 | GPR45 | -1.33E-03 | 1.85E-02 |
| 13561 | Usp9x | USP9X | 1.15E-03 | -1.47E-02 |
| 13562 | Far1 | FAR1 | -1.27E-03 | -1.74E-02 |
| 13563 | Asic4 | ASIC4 | -1.61E-03 | -2.00E-02 |
| 13564 | Smyd4 | SMYD4 | -1.42E-03 | -2.70E-02 |
| 13565 | Cldn23 | CLDN23 | -7.91E-03 | 5.93E-02 |

| | | | | |
|---|---|---|---|---|
| 13566 | Lamtor2 | LAMTOR2 | -9.14E-04 | -1.66E-02 |
| 13567 | Kdm6b | KDM6B | 1.49E-03 | -5.54E-03 |
| 13568 | Slc8a2 | SLC8A2 | 3.14E-03 | 4.81E-02 |
| 13569 | Zfp41 | ZFP41 | -3.37E-03 | -1.70E-02 |
| 13570 | Pla2g2e | PLA2G2E | 1.54E-03 | 1.42E-02 |
| 13571 | Npc2 | NPC2 | -1.04E-03 | 1.72E-02 |
| 13572 | Srr | SRR | 1.30E-03 | -1.30E-02 |
| 13573 | Ppm1b | PPM1B | -8.69E-04 | 2.69E-03 |
| 13574 | Dhx30 | DHX30 | 7.76E-04 | -1.13E-02 |
| 13575 | Uncx | UNCX | 1.08E-02 | -9.54E-02 |
| 13576 | Cryaa | CRYAA | 1.45E-03 | -2.09E-02 |
| 13577 | Cryaa | LOC102724652 | 1.45E-03 | -2.09E-02 |
| 13578 | Capn13 | CAPN13 | -1.44E-02 | 1.99E-01 |
| 13579 | Prtn3 | PRTN3 | -2.29E-03 | 3.25E-02 |
| 13580 | Gml | GML | 1.25E-03 | 4.13E-02 |
| 13581 | Zfp619 | ZNF208 | 2.30E-03 | -2.74E-02 |
| 13582 | Mycbp | MYCBP | 1.19E-03 | -3.36E-03 |
| 13583 | Foxf1 | FOXF1 | -3.05E-03 | 2.44E-02 |
| 13584 | Gpld1 | GPLD1 | -4.05E-03 | 6.77E-03 |
| 13585 | Ms4a8a | MS4A8 | 4.18E-03 | 9.25E-02 |
| 13586 | Tigit | TIGIT | 1.77E-03 | -1.38E-02 |
| 13587 | Rad9b | RAD9B | 1.87E-03 | -1.35E-02 |
| 13588 | Dkkl1 | DKKL1 | -1.61E-03 | 8.62E-04 |
| 13589 | Amy1 | AMY2A | -3.40E-03 | -2.00E-02 |
| 13590 | Recql5 | RECQL5 | 9.07E-04 | -1.49E-02 |
| 13591 | Igfbp7 | IGFBP7 | -8.62E-04 | -1.16E-02 |
| 13592 | Plekhg3 | PLEKHG3 | 1.03E-03 | 2.32E-03 |
| 13593 | Tm6sf1 | TM6SF1 | -4.22E-03 | 7.69E-02 |
| 13594 | Dio3 | DIO3 | -3.04E-03 | 3.24E-02 |
| 13595 | Samd8 | SAMD8 | -1.34E-03 | -1.60E-02 |
| 13596 | Mex3c | MEX3C | 1.17E-03 | 1.95E-02 |
| 13597 | Vwa5b2 | VWA5B2 | -1.67E-03 | 1.75E-02 |
| 13598 | Mdga1 | MDGA1 | 3.90E-03 | 9.69E-02 |
| 13599 | Thada | THADA | 1.14E-03 | -1.06E-03 |
| 13600 | Camkmt | CAMKMT | 1.85E-03 | -2.90E-02 |
| 13601 | Efhb | EFHB | -1.68E-03 | 2.06E-02 |
| 13602 | Sp7 | SP7 | -1.14E-03 | 7.62E-03 |
| 13603 | Nfic | NFIC | -1.93E-03 | -3.76E-03 |
| 13604 | Atg4d | ATG4D | -9.09E-04 | -1.49E-02 |
| 13605 | Cdon | CDON | 3.18E-03 | 9.74E-03 |
| 13606 | Rgs16 | RGS16 | -4.78E-03 | 1.79E-01 |
| 13607 | Tat | TAT | 7.08E-03 | 1.50E-01 |
| 13608 | Arf6 | ARF6 | 7.54E-04 | 1.02E-02 |
| 13609 | 2700062C07Rik | C18orf21 | 1.18E-03 | 2.33E-03 |
| 13610 | Atp8b1 | ATP8B1 | 4.46E-03 | 1.16E-02 |
| 13611 | Tgm4 | TGM4 | -2.02E-03 | -2.42E-02 |
| 13612 | Fem1b | FEM1B | 1.01E-03 | 1.16E-02 |
| 13613 | Clcf1 | CLCF1 | -3.41E-03 | 1.23E-01 |
| 13614 | Tnf | TNF | -3.70E-03 | 7.00E-02 |
| 13615 | Nuak1 | NUAK1 | 2.04E-03 | -4.72E-03 |
| 13616 | Timm8a1 | TIMM8A | -1.70E-03 | 3.23E-02 |
| 13617 | Bloc1s4 | BLOC1S4 | 1.11E-03 | -1.09E-02 |
| 13618 | Prrx1 | PRRX1 | -4.72E-03 | 4.14E-02 |

| | | | | |
|---|---|---|---|---|
| 13619 | Gpx7 | GPX7 | -3.47E-03 | 5.85E-02 |
| 13620 | Psma8 | PSMA8 | -2.56E-03 | 3.17E-02 |
| 13621 | Fmod | FMOD | -3.62E-03 | -8.78E-03 |
| 13622 | Mylpf | MYLPF | -2.34E-03 | -8.34E-03 |
| 13623 | Nr4a2 | NR4A2 | -1.14E-02 | 9.56E-02 |
| 13624 | Ints4 | INTS4 | -7.22E-04 | -1.49E-02 |
| 13625 | Myo5c | MYO5C | 3.81E-03 | -2.40E-02 |
| 13626 | Kars | KARS | -7.54E-04 | -6.94E-03 |
| 13627 | Trpv2 | TRPV2 | 3.51E-03 | 1.23E-01 |
| 13628 | Ddt | DDT | -1.85E-03 | -3.50E-02 |
| 13629 | Ap3b2 | AP3B2 | 2.01E-03 | 9.51E-04 |
| 13630 | Ttc38 | TTC38 | 1.66E-03 | -2.73E-02 |
| 13631 | Gstm2 | GSTM1 | -4.07E-03 | 9.81E-03 |
| 13632 | Slc41a2 | SLC41A2 | -1.92E-03 | 5.47E-03 |
| 13633 | Ccnl1 | CCNL1 | -1.29E-03 | 2.81E-02 |
| 13634 | Nans | NANS | -1.51E-03 | 2.26E-02 |
| 13635 | Ints9 | INTS9 | -9.19E-04 | -2.66E-03 |
| 13636 | Il18r1 | IL18R1 | -7.79E-03 | 8.70E-02 |
| 13637 | Cldn11 | CLDN11 | 1.15E-02 | 2.58E-02 |
| 13638 | Rbpms2 | RBPMS2 | -9.77E-04 | 2.27E-03 |
| 13639 | Rps26 | RPS26 | 8.12E-04 | 1.27E-02 |
| 13640 | Gucd1 | GUCD1 | -1.14E-03 | -1.56E-02 |
| 13641 | Nup50 | NUP50 | -7.69E-04 | -4.51E-03 |
| 13642 | Ppat | PPAT | -1.08E-03 | -1.81E-02 |
| 13643 | Lap3 | LAP3 | 1.04E-03 | -3.05E-02 |
| 13644 | Aspa | ASPA | -1.55E-03 | -3.62E-02 |
| 13645 | Supt4a | SUPT4H1 | 7.44E-04 | -1.95E-02 |
| 13646 | Myadml2 | MYADML2 | 1.13E-03 | -7.53E-03 |
| 13647 | Surf1 | SURF1 | -8.30E-04 | -1.56E-02 |
| 13648 | Ahdc1 | AHDC1 | -1.36E-03 | 2.04E-02 |
| 13649 | Kcnh4 | KCNH4 | 1.23E-03 | -1.75E-03 |
| 13650 | Pdzd9 | PDZD9 | -1.37E-03 | -6.03E-03 |
| 13651 | E4f1 | E4F1 | -8.92E-04 | -1.53E-02 |
| 13652 | Mgp | MGP | -2.96E-03 | 1.37E-01 |
| 13653 | Mcm8 | MCM8 | -2.05E-03 | 3.93E-02 |
| 13654 | Pcdh12 | PCDH12 | -2.91E-03 | -1.08E-02 |
| 13655 | Cep192 | CEP192 | -1.53E-03 | 2.43E-02 |
| 13656 | Wdr73 | WDR73 | 7.08E-04 | -1.68E-03 |
| 13657 | Gls2 | GLS2 | -2.68E-03 | 8.01E-02 |
| 13658 | Hps3 | HPS3 | 9.72E-04 | -1.85E-02 |
| 13659 | Selenbp2 | SELENBP1 | 2.22E-03 | -1.86E-02 |
| 13660 | Fam102b | FAM102B | 1.22E-03 | -3.88E-02 |
| 13661 | Mfap3 | MFAP3 | 7.98E-04 | 1.16E-02 |
| 13662 | Srpk3 | SRPK3 | -2.39E-03 | 1.15E-02 |
| 13663 | Hsp90b1 | HSP90B1 | -1.38E-03 | -6.62E-03 |
| 13664 | Akirin2 | AKIRIN2 | 1.10E-03 | 7.78E-03 |
| 13665 | Trmt2b | TRMT2B | 7.90E-04 | -1.16E-02 |
| 13666 | Osgin2 | OSGIN2 | -1.62E-03 | 1.90E-02 |
| 13667 | Ahsg | AHSG | 7.88E-03 | 2.16E-01 |
| 13668 | Vsig8 | VSIG8 | -2.27E-03 | -1.49E-02 |
| 13669 | Pigs | PIGS | 6.02E-04 | -9.33E-03 |
| 13670 | Heatr1 | HEATR1 | 9.55E-04 | 2.36E-02 |
| 13671 | Fcgr3 | FCGR2A | -2.84E-03 | 1.86E-01 |

| | | | | |
|---|---|---|---|---|
| 13672 | Lrwd1 | LRWD1 | -8.60E-04 | 6.92E-03 |
| 13673 | Dpf2 | DPF2 | 6.39E-04 | -6.40E-03 |
| 13674 | Mfap3l | MFAP3L | 1.03E-03 | -2.57E-02 |
| 13675 | Gpr39 | GPR39 | -4.02E-03 | 9.81E-02 |
| 13676 | Epcam | EPCAM | -3.16E-03 | -1.44E-02 |
| 13677 | Apoa5 | APOA5 | -3.39E-03 | 7.72E-02 |
| 13678 | Mis18a | MIS18A | -1.97E-03 | 6.67E-02 |
| 13679 | Spen | SPEN | -1.05E-03 | -1.33E-02 |
| 13680 | Rpl26 | RPL26 | -6.65E-04 | 3.80E-03 |
| 13681 | Col3a1 | COL3A1 | -5.57E-03 | 1.69E-01 |
| 13682 | Mettl21a | METTL21A | 1.10E-03 | 3.88E-04 |
| 13683 | Dysf | DYSF | -1.26E-03 | 1.33E-02 |
| 13684 | 4933411K16Rik | C10orf62 | 2.58E-03 | -1.61E-02 |
| 13685 | Acpp | ACPP | 2.67E-03 | -9.38E-03 |
| 13686 | Spg11 | SPG11 | -8.53E-04 | -3.33E-02 |
| 13687 | Dapk3 | DAPK3 | -7.61E-04 | -1.56E-03 |
| 13688 | Tdo2 | TDO2 | 5.42E-03 | 1.20E-01 |
| 13689 | S100a13 | S100A13 | 9.01E-04 | 1.01E-02 |
| 13690 | Scn5a | SCN5A | 3.49E-03 | -1.41E-01 |
| 13691 | Alkbh6 | ALKBH6 | -1.36E-03 | -1.11E-02 |
| 13692 | Zdbf2 | ZDBF2 | -1.31E-03 | -7.14E-03 |
| 13693 | Csf3r | CSF3R | -3.77E-03 | 1.20E-01 |
| 13694 | Smarcad1 | SMARCAD1 | -7.14E-04 | 1.89E-02 |
| 13695 | Slc9a1 | SLC9A1 | 4.88E-04 | -3.84E-03 |
| 13696 | Fsip1 | FSIP1 | -1.98E-03 | 1.84E-02 |
| 13697 | Icam5 | ICAM5 | -2.64E-03 | 2.11E-02 |
| 13698 | Cav3 | CAV3 | 1.95E-03 | 2.75E-02 |
| 13699 | Dhx8 | DHX8 | 5.65E-04 | -5.83E-03 |
| 13700 | Cst7 | CST7 | -1.06E-03 | 3.19E-02 |
| 13701 | Arhgap28 | ARHGAP28 | 2.17E-03 | -2.65E-02 |
| 13702 | Ly6g6d | LY6G6D | 2.69E-03 | -3.30E-02 |
| 13703 | Rnf141 | RNF141 | -8.19E-04 | -2.04E-02 |
| 13704 | Inpp5f | INPP5F | -6.61E-04 | -2.55E-02 |
| 13705 | Cpa2 | CPA2 | -7.69E-04 | 1.32E-04 |
| 13706 | Brd9 | BRD9 | 6.89E-04 | -7.92E-03 |
| 13707 | Popdc3 | POPDC3 | -1.45E-03 | 6.78E-02 |
| 13708 | Ube2r2 | UBE2R2 | -5.92E-04 | -3.37E-03 |
| 13709 | Lsm8 | LSM8 | 8.79E-04 | 9.00E-03 |
| 13710 | Tmem39b | TMEM39B | 1.02E-03 | -1.76E-02 |
| 13711 | Itpripl1 | ITPRIPL1 | 1.38E-03 | -2.98E-02 |
| 13712 | Sccpdh | SCCPDH | -6.32E-04 | -2.94E-02 |
| 13713 | Dhfr | DHFR | 1.35E-03 | -3.88E-03 |
| 13714 | Pcdhb16 | PCDHB8 | 2.45E-03 | -4.10E-02 |
| 13715 | Gata4 | GATA4 | -8.40E-04 | 1.48E-02 |
| 13716 | Rhcg | RHCG | -8.77E-04 | 9.76E-03 |
| 13717 | Trib2 | TRIB2 | -1.25E-03 | 1.91E-02 |
| 13718 | F11r | F11R | -5.91E-04 | 1.13E-02 |
| 13719 | Myrf | MYRF | 2.43E-03 | -9.50E-03 |
| 13720 | Mphosph8 | MPHOSPH8 | -8.20E-04 | 4.28E-03 |
| 13721 | Pgm1 | PGM2 | -1.10E-03 | -1.46E-02 |
| 13722 | Atp8b3 | ATP8B3 | 1.04E-03 | 1.88E-02 |
| 13723 | Btaf1 | BTAF1 | -6.59E-04 | 1.69E-03 |
| 13724 | Shc2 | SHC2 | -2.53E-03 | 2.82E-02 |

| | | | | |
|---|---|---|---|---|
| 13725 | Tspan17 | TSPAN17 | 1.72E-03 | 8.85E-03 |
| 13726 | Mcm7 | MCM7 | 1.64E-03 | 5.28E-02 |
| 13727 | Csnk1g1 | CSNK1G1 | -8.76E-04 | -3.18E-03 |
| 13728 | Snx31 | SNX31 | 4.70E-03 | 1.76E-02 |
| 13729 | H2-Ke6 | HSD17B8 | 9.61E-04 | 5.10E-03 |
| 13730 | Ttll10 | TTLL10 | 1.20E-03 | -6.13E-02 |
| 13731 | Itgb4 | ITGB4 | -2.73E-03 | 5.83E-02 |
| 13732 | Prss27 | PRSS27 | 1.57E-03 | 3.96E-02 |
| 13733 | Chad | CHAD | -9.93E-04 | 1.56E-02 |
| 13734 | Brsk1 | BRSK1 | -2.11E-03 | 1.75E-02 |
| 13735 | Gsk3b | GSK3B | -6.06E-04 | -1.02E-02 |
| 13736 | Tmed1 | TMED1 | -7.88E-04 | -1.83E-04 |
| 13737 | Hao2 | HAO2 | -1.01E-03 | -4.73E-02 |
| 13738 | Xrcc3 | XRCC3 | -2.20E-03 | -3.57E-03 |
| 13739 | Faim2 | FAIM2 | -4.56E-03 | -1.02E-02 |
| 13740 | Ppt2 | PPT2 | 8.95E-04 | 1.14E-02 |
| 13741 | Gigyf1 | GIGYF1 | 6.28E-04 | -2.45E-02 |
| 13742 | Mxd1 | MXD1 | -1.26E-03 | -4.76E-02 |
| 13743 | Mak | MAK | 9.49E-04 | 2.57E-02 |
| 13744 | Ak2 | AK2 | 1.03E-03 | -1.08E-02 |
| 13745 | Gpr84 | GPR84 | 3.46E-03 | 5.94E-02 |
| 13746 | Kansl3 | KANSL3 | 6.05E-04 | -1.20E-02 |
| 13747 | Pkdrej | PKDREJ | 1.07E-03 | 8.01E-03 |
| 13748 | Hes7 | HES7 | -1.49E-03 | 1.93E-02 |
| 13749 | Dis3l2 | DIS3L2 | -6.62E-04 | -2.75E-02 |
| 13750 | Tubgcp6 | TUBGCP6 | -6.76E-04 | 7.82E-03 |
| 13751 | Cpxm2 | CPXM2 | -3.90E-03 | 1.18E-01 |
| 13752 | Tenm3 | TENM3 | 3.50E-03 | -1.97E-02 |
| 13753 | Cdcp1 | CDCP1 | 7.62E-04 | -8.71E-03 |
| 13754 | Rtn4ip1 | RTN4IP1 | -8.29E-04 | -3.44E-02 |
| 13755 | Hsd17b7 | HSD17B7 | -1.07E-03 | 1.15E-02 |
| 13756 | Ptcd1 | ATP5J2-PTCD1 | 6.93E-04 | 1.20E-04 |
| 13757 | Taok2 | TAOK2 | -5.53E-04 | -1.33E-02 |
| 13758 | Klhdc2 | KLHDC2 | 6.03E-04 | -9.49E-03 |
| 13759 | Sft2d3 | SFT2D3 | -1.00E-03 | -1.17E-02 |
| 13760 | Haao | HAAO | 1.18E-03 | -1.89E-02 |
| 13761 | Fsbp | FSBP | 1.67E-03 | 1.52E-02 |
| 13762 | Ninl | NINL | 1.44E-03 | -2.94E-02 |
| 13763 | Nsmce2 | NSMCE2 | 8.13E-04 | -6.02E-03 |
| 13764 | Sobp | SOBP | -2.32E-03 | 3.40E-02 |
| 13765 | Lims2 | LIMS2 | 1.43E-03 | -1.04E-02 |
| 13766 | Pttg1 | PTTG1 | -6.71E-04 | 8.27E-03 |
| 13767 | Mesp2 | MESP2 | -1.77E-03 | 2.29E-02 |
| 13768 | Bnipl | BNIPL | -3.23E-03 | 4.79E-02 |
| 13769 | Ctdsp1 | CTDSP1 | -5.10E-04 | -1.51E-02 |
| 13770 | Med26 | MED26 | -1.10E-03 | -2.97E-03 |
| 13771 | Sptssa | SPTSSA | 6.00E-04 | -1.45E-02 |
| 13772 | Cdh11 | CDH11 | -3.40E-03 | 1.73E-02 |
| 13773 | Plin1 | PLIN1 | 1.09E-02 | 1.24E-01 |
| 13774 | Dcakd | DCAKD | 7.15E-04 | 4.39E-03 |
| 13775 | Prss23 | PRSS23 | 7.86E-04 | -8.27E-03 |
| 13776 | Azin1 | AZIN1 | 6.95E-04 | 6.48E-03 |
| 13777 | Nkiras1 | NKIRAS1 | -7.62E-04 | -7.83E-03 |

| | | | | |
|---|---|---|---|---|
| 13778 | Aldh1a2 | ALDH1A2 | -3.76E-03 | 9.09E-02 |
| 13779 | Ap4m1 | AP4M1 | 6.23E-04 | -3.37E-03 |
| 13780 | Tnfrsf19 | TNFRSF19 | 8.18E-03 | 1.10E-01 |
| 13781 | Timp2 | TIMP2 | 1.46E-03 | 1.29E-02 |
| 13782 | Clp1 | CLP1 | 9.28E-04 | -6.30E-03 |
| 13783 | Pcdhga1 | PCDHGA1 | 1.59E-03 | -2.90E-02 |
| 13784 | Prr14 | PRR14 | -3.68E-04 | -2.36E-03 |
| 13785 | Fyco1 | FYCO1 | 7.71E-04 | -2.73E-02 |
| 13786 | Paqr8 | PAQR8 | -2.60E-03 | 1.56E-02 |
| 13787 | Zfp36l1 | ZFP36L1 | 9.91E-04 | 2.71E-02 |
| 13788 | Gltp | GLTP | 5.28E-04 | -5.67E-03 |
| 13789 | Dlk2 | DLK2 | -1.88E-03 | 7.10E-02 |
| 13790 | Creb5 | CREB5 | 2.61E-03 | 1.64E-01 |
| 13791 | Itih4 | ITIH4 | -4.20E-03 | 1.65E-01 |
| 13792 | Phykpl | PHYKPL | 7.03E-04 | -4.63E-03 |
| 13793 | Ccdc167 | CCDC167 | 1.34E-03 | -6.29E-02 |
| 13794 | Adamtsl5 | ADAMTSL5 | -2.47E-03 | 1.14E-01 |
| 13795 | Apip | APIP | -7.48E-04 | -7.67E-03 |
| 13796 | Txlnb | TXLNB | -1.53E-03 | 4.16E-02 |
| 13797 | Rps13 | RPS13 | -5.32E-04 | 7.57E-03 |
| 13798 | Vpreb3 | VPREB3 | 7.07E-04 | -9.81E-03 |
| 13799 | Grm4 | GRM4 | -5.99E-03 | -1.15E-02 |
| 13800 | Casp8ap2 | CASP8AP2 | 1.17E-03 | -1.18E-02 |
| 13801 | Atp5g2 | ATP5G2 | 5.26E-04 | -5.60E-03 |
| 13802 | Ift122 | IFT122 | -5.75E-04 | -1.87E-02 |
| 13803 | Hc | C5 | -2.18E-03 | 7.72E-02 |
| 13804 | Rad50 | RAD50 | 8.00E-04 | 8.80E-03 |
| 13805 | Afmid | AFMID | 1.70E-03 | -6.34E-03 |
| 13806 | Dlx1 | DLX1 | 1.27E-03 | -1.09E-02 |
| 13807 | Nr1i2 | NR1I2 | 1.80E-03 | 4.04E-04 |
| 13808 | Pak4 | PAK4 | 5.95E-04 | 8.94E-03 |
| 13809 | Lama3 | LAMA3 | 1.56E-03 | -6.96E-03 |
| 13810 | Ttll3 | TTLL3 | -1.58E-03 | -6.43E-03 |
| 13811 | Ubac1 | UBAC1 | 4.76E-04 | 1.05E-02 |
| 13812 | Gsdmc4 | GSDMC | -4.44E-03 | 1.19E-01 |
| 13813 | Kcnj12 | KCNJ12 | -1.96E-03 | -6.82E-02 |
| 13814 | Stag3 | STAG3 | 1.32E-03 | 3.25E-02 |
| 13815 | Ebf2 | EBF2 | -1.25E-03 | 1.58E-02 |
| 13816 | Whrn | DFNB31 | 9.24E-04 | 5.77E-03 |
| 13817 | Gm7694 | C1orf226 | -8.58E-04 | -5.09E-02 |
| 13818 | Ogfod1 | OGFOD1 | -6.12E-04 | -1.68E-02 |
| 13819 | Zfp661 | ZNF2 | -1.24E-03 | -2.00E-02 |
| 13820 | Akt2 | AKT2 | -4.22E-04 | -1.46E-02 |
| 13821 | Ccl19 | CCL19 | 6.29E-04 | -1.09E-02 |
| 13822 | Kctd14 | KCTD14 | 2.07E-03 | -4.62E-02 |
| 13823 | Il1rl2 | IL1RL2 | -1.97E-03 | 3.88E-02 |
| 13824 | Ubqln4 | UBQLN4 | 3.99E-04 | 3.94E-03 |
| 13825 | Lgals1 | LGALS1 | -1.61E-03 | 4.74E-02 |
| 13826 | Eid3 | EID3 | 1.23E-03 | 3.34E-02 |
| 13827 | Treml1 | TREML1 | -8.27E-04 | 1.93E-02 |
| 13828 | Star | STAR | 1.62E-03 | -4.47E-03 |
| 13829 | Osm | OSM | -1.85E-03 | 5.91E-02 |
| 13830 | Epm2a | EPM2A | -7.16E-04 | -4.18E-02 |

| | | | | |
|---|---|---|---|---|
| 13831 | Tmem44 | TMEM44 | 1.18E-03 | -5.45E-02 |
| 13832 | Mlf1 | MLF1 | -1.14E-03 | 3.99E-02 |
| 13833 | Col28a1 | COL28A1 | 1.48E-03 | 2.85E-02 |
| 13834 | Col10a1 | COL10A1 | 1.10E-03 | 9.56E-02 |
| 13835 | Kcnj2 | KCNJ2 | -1.63E-03 | -7.97E-02 |
| 13836 | Clvs2 | CLVS2 | 1.01E-03 | 4.91E-02 |
| 13837 | Sri | SRI | -3.62E-04 | -1.44E-02 |
| 13838 | Commd10 | COMMD10 | 7.46E-04 | 1.48E-03 |
| 13839 | Adra2c | ADRA2C | -2.40E-03 | 1.23E-02 |
| 13840 | Adcy8 | ADCY8 | 8.38E-04 | -3.51E-04 |
| 13841 | Exph5 | EXPH5 | -1.34E-03 | -2.33E-02 |
| 13842 | Mrgprf | MRGPRF | -1.87E-03 | 2.12E-02 |
| 13843 | Tcf7l1 | TCF7L1 | 1.46E-03 | -1.58E-02 |
| 13844 | Awat2 | AWAT2 | -1.77E-03 | 4.84E-03 |
| 13845 | Gm11992 | C7orf57 | 9.84E-04 | -5.26E-02 |
| 13846 | Tex15 | TEX15 | 1.44E-03 | 8.71E-02 |
| 13847 | Ppp1r14d | PPP1R14D | -1.76E-03 | 2.19E-02 |
| 13848 | Foxn1 | FOXN1 | 9.61E-04 | 1.71E-02 |
| 13849 | Medag | MEDAG | 1.14E-03 | 7.81E-02 |
| 13850 | Slc12a8 | SLC12A8 | -1.78E-03 | 3.06E-02 |
| 13851 | Inpp4a | INPP4A | 5.62E-04 | -2.04E-02 |
| 13852 | Tdrkh | TDRKH | -1.32E-03 | -4.31E-02 |
| 13853 | Tab3 | TAB3 | -5.28E-04 | 7.00E-04 |
| 13854 | Rpl13a | RPL13A | -8.42E-04 | 4.79E-02 |
| 13855 | 1110051M20Rik | C11orf49 | 5.68E-04 | -2.84E-02 |
| 13856 | Ppp1r1a | PPP1R1A | -1.25E-03 | -5.07E-02 |
| 13857 | Ripk3 | RIPK3 | -1.62E-03 | 1.09E-01 |
| 13858 | Ccndbp1 | CCNDBP1 | -3.14E-04 | -7.88E-03 |
| 13859 | Col5a1 | COL5A1 | 2.32E-03 | 3.15E-02 |
| 13860 | Schip1 | SCHIP1 | 1.28E-03 | -3.07E-02 |
| 13861 | Ppie | PPIE | -6.02E-04 | -1.55E-02 |
| 13862 | Fbxo33 | FBXO33 | -8.29E-04 | 3.35E-03 |
| 13863 | 4933430I17Rik | C9orf43 | -1.25E-03 | -4.83E-02 |
| 13864 | Ap1g2 | AP1G2 | -4.59E-04 | 1.04E-04 |
| 13865 | Tomt | LRTOMT | 1.01E-03 | 2.72E-02 |
| 13866 | Kcnj1 | KCNJ1 | 8.96E-04 | -2.87E-02 |
| 13867 | Psenen | PSENEN | -4.01E-04 | -9.64E-03 |
| 13868 | Rdh16 | RDH16 | -8.49E-04 | 1.18E-05 |
| 13869 | Cdhr3 | CDHR3 | 4.20E-04 | -1.04E-03 |
| 13870 | Mthfsd | MTHFSD | 3.48E-04 | -2.37E-02 |
| 13871 | Adipoq | ADIPOQ | -8.10E-03 | 1.35E-01 |
| 13872 | Ugt2b36 | UGT2B7 | 1.20E-03 | 4.13E-02 |
| 13873 | Crip1 | CRIP1 | 8.44E-04 | 1.01E-01 |
| 13874 | Trip10 | TRIP10 | 8.05E-04 | -9.81E-03 |
| 13875 | Ramp1 | RAMP1 | -1.78E-03 | 1.09E-01 |
| 13876 | Cps1 | CPS1 | -3.48E-03 | 1.45E-01 |
| 13877 | Polr2j | POLR2J | 3.81E-04 | 5.63E-03 |
| 13878 | Plin2 | PLIN2 | 9.45E-04 | 4.32E-02 |
| 13879 | Abcg4 | ABCG4 | 1.17E-03 | -7.66E-03 |
| 13880 | Alb | ALB | -3.73E-03 | 3.64E-02 |
| 13881 | Impa1 | IMPA1 | -4.36E-04 | -1.15E-03 |
| 13882 | Slc35a5 | SLC35A5 | -3.82E-04 | -1.44E-02 |
| 13883 | Itpkb | ITPKB | -6.01E-04 | 7.78E-03 |

| | | | | |
|---|---|---|---|---|
| 13884 | Zfp462 | ZNF462 | -8.11E-04 | -7.40E-03 |
| 13885 | Kif19a | KIF19 | 1.11E-03 | 1.77E-02 |
| 13886 | Abcb5 | ABCB5 | -2.73E-04 | 7.62E-03 |
| 13887 | Adamtsl1 | ADAMTSL1 | -1.65E-03 | -6.19E-03 |
| 13888 | Tekt1 | TEKT1 | 5.42E-04 | 1.47E-02 |
| 13889 | Tmem189 | TMEM189 | -6.41E-04 | -2.85E-03 |
| 13890 | Rsbn1 | RSBN1 | -6.11E-04 | -2.26E-02 |
| 13891 | Hoxa2 | HOXA2 | 1.53E-03 | -1.88E-02 |
| 13892 | Znfx1 | ZNFX1 | -3.40E-04 | -1.63E-03 |
| 13893 | Mtmr3 | MTMR3 | -3.92E-04 | -2.61E-02 |
| 13894 | Tmem150b | TMEM150B | 1.51E-03 | 1.07E-02 |
| 13895 | Kcnj14 | KCNJ14 | 7.19E-04 | 4.05E-02 |
| 13896 | Ap5m1 | AP5M1 | -4.70E-04 | -1.79E-02 |
| 13897 | Marco | MARCO | 5.46E-03 | 1.50E-01 |
| 13898 | Acsl1 | ACSL1 | 6.15E-04 | -1.28E-02 |
| 13899 | Cacna2d4 | CACNA2D4 | -1.05E-03 | 1.28E-02 |
| 13900 | Pam | PAM | 2.85E-04 | -1.01E-02 |
| 13901 | Brinp1 | BRINP1 | 6.23E-04 | 1.31E-02 |
| 13902 | Mybpc2 | MYBPC2 | 1.31E-03 | 1.58E-01 |
| 13903 | Nuggc | NUGGC | 1.11E-03 | -7.46E-02 |
| 13904 | Slc25a13 | SLC25A13 | 6.88E-04 | -1.35E-02 |
| 13905 | Tcp11 | TCP11 | -1.49E-03 | 8.36E-03 |
| 13906 | Zscan25 | ZSCAN25 | -6.40E-04 | -1.14E-02 |
| 13907 | Pde3a | PDE3A | -1.13E-03 | -1.58E-02 |
| 13908 | Gnao1 | GNAO1 | -2.59E-03 | 5.60E-02 |
| 13909 | Otc | OTC | 1.22E-03 | 3.59E-02 |
| 13910 | Raf1 | RAF1 | 2.37E-04 | 1.47E-03 |
| 13911 | Vopp1 | VOPP1 | -1.63E-03 | 7.32E-02 |
| 13912 | Simc1 | SIMC1 | -6.42E-04 | 3.40E-02 |
| 13913 | Adap2 | ADAP2 | 1.05E-03 | -5.30E-03 |
| 13914 | Anxa6 | ANXA6 | 2.32E-04 | 8.61E-03 |
| 13915 | Palm | PALM | 4.69E-04 | -1.61E-02 |
| 13916 | Tnfrsf10b | TNFRSF10A | 9.05E-04 | 4.94E-02 |
| 13917 | Hnf1a | HNF1A | 7.08E-04 | -2.10E-02 |
| 13918 | Tdrd6 | TDRD6 | -6.89E-04 | -1.07E-02 |
| 13919 | Ccl2 | CCL13 | -2.03E-03 | 4.13E-01 |
| 13920 | Tmem252 | TMEM252 | -1.73E-03 | 1.20E-01 |
| 13921 | Rhbg | RHBG | -9.36E-04 | -3.68E-02 |
| 13922 | Fam174b | FAM174B | 2.34E-03 | 1.44E-02 |
| 13923 | Bbip1 | BBIP1 | 4.11E-04 | -6.00E-03 |
| 13924 | Rps11 | RPS11 | 2.73E-04 | 1.37E-02 |
| 13925 | Dad1 | DAD1 | -2.69E-04 | -2.50E-03 |
| 13926 | Frrs1 | FRRS1 | -5.78E-04 | -4.40E-04 |
| 13927 | Arhgdib | ARHGDIB | 1.65E-03 | 1.01E-02 |
| 13928 | Rnf223 | RNF223 | -1.11E-03 | 1.68E-03 |
| 13929 | Hid1 | HID1 | -1.66E-03 | 3.13E-02 |
| 13930 | Axl | AXL | 8.14E-04 | 3.95E-02 |
| 13931 | Tmod3 | TMOD3 | 3.33E-04 | 1.30E-02 |
| 13932 | Csnk1g3 | CSNK1G3 | 4.00E-04 | -3.84E-03 |
| 13933 | Wdr83 | WDR83 | 2.94E-04 | -4.27E-03 |
| 13934 | Msx2 | MSX2 | 2.02E-03 | 7.88E-02 |
| 13935 | Tspan6 | TSPAN6 | -2.00E-03 | 4.73E-02 |
| 13936 | Lsmem1 | LSMEM1 | 5.87E-04 | 1.36E-02 |

| | | | | |
|---|---|---|---|---|
| 13937 | Mrpl46 | MRPL46 | -2.92E-04 | -2.09E-02 |
| 13938 | Aldh2 | ALDH2 | -5.33E-04 | -2.22E-02 |
| 13939 | Rps17 | RPS17 | -2.71E-04 | 5.31E-03 |
| 13940 | Wdr76 | WDR76 | 5.73E-04 | 2.94E-02 |
| 13941 | Six1 | SIX1 | -8.55E-04 | 3.66E-02 |
| 13942 | Cntfr | CNTFR | -1.11E-03 | -5.99E-02 |
| 13943 | Zfp668 | ZNF668 | -5.16E-04 | 1.33E-02 |
| 13944 | Use1 | USE1 | -3.06E-04 | 4.48E-03 |
| 13945 | Asxl3 | ASXL3 | -8.21E-04 | 5.74E-03 |
| 13946 | Tlr8 | TLR8 | -1.27E-03 | 1.11E-01 |
| 13947 | Pgrmc2 | PGRMC2 | -2.22E-04 | -6.85E-03 |
| 13948 | Paqr6 | PAQR6 | -8.93E-04 | 5.38E-02 |
| 13949 | Fkbp8 | FKBP8 | -2.67E-04 | -1.47E-02 |
| 13950 | Gm14325 | ZNF442 | -5.82E-04 | -2.01E-02 |
| 13951 | Tymp | TYMP | -7.53E-04 | -4.47E-02 |
| 13952 | Gfap | GFAP | -6.53E-04 | 4.14E-03 |
| 13953 | Mfsd12 | MFSD12 | 5.18E-04 | 2.26E-02 |
| 13954 | Gm15753 | SP110 | 8.74E-04 | 1.56E-02 |
| 13955 | Svep1 | SVEP1 | 1.35E-03 | -5.54E-02 |
| 13956 | Appl1 | APPL1 | -2.59E-04 | -1.56E-02 |
| 13957 | Fabp7 | FABP7 | 1.82E-03 | -3.36E-02 |
| 13958 | C2cd4b | C2CD4B | -1.54E-03 | 1.01E-01 |
| 13959 | Rasgrp2 | RASGRP2 | -7.14E-04 | -1.00E-02 |
| 13960 | Thap4 | THAP4 | 2.46E-04 | -2.22E-02 |
| 13961 | Mycl | MYCL | -2.01E-03 | 1.46E-01 |
| 13962 | Ccnc | CCNC | -3.19E-04 | -2.20E-02 |
| 13963 | Cideb | CIDEB | 3.31E-04 | -1.70E-02 |
| 13964 | Vwce | VWCE | 6.87E-04 | -2.46E-03 |
| 13965 | Dnajc30 | DNAJC30 | -3.00E-04 | -1.89E-02 |
| 13966 | Armc7 | ARMC7 | -5.77E-04 | -2.21E-02 |
| 13967 | Rwdd4a | RWDD4 | -3.04E-04 | -6.89E-03 |
| 13968 | Fezf2 | FEZF2 | -2.73E-04 | 1.96E-02 |
| 13969 | Gzmk | GZMK | 6.25E-04 | 2.16E-02 |
| 13970 | Ptger1 | PTGER1 | 1.90E-03 | -1.02E-01 |
| 13971 | Hdac1 | HDAC1 | -1.65E-04 | 1.22E-02 |
| 13972 | Spcs1 | SPCS1 | -1.96E-04 | -2.76E-03 |
| 13973 | Snrpa1 | SNRPA1 | 3.67E-04 | 1.21E-03 |
| 13974 | Car6 | CA6 | -3.78E-04 | 8.12E-03 |
| 13975 | Tiam2 | TIAM2 | -8.40E-04 | 4.32E-02 |
| 13976 | F2 | F2 | 1.30E-03 | 6.48E-02 |
| 13977 | Mpdz | MPDZ | -3.99E-04 | -6.12E-02 |
| 13978 | Elmod2 | ELMOD2 | -2.76E-04 | -3.56E-03 |
| 13979 | Snapc3 | SNAPC3 | -3.92E-04 | 7.61E-03 |
| 13980 | Nbl1 | NBL1 | -1.38E-03 | 6.33E-02 |
| 13981 | Nbl1 | MINOS1-NBL1 | -1.38E-03 | 6.33E-02 |
| 13982 | Oxsr1 | OXSR1 | -2.02E-04 | 8.56E-03 |
| 13983 | Erg | ERG | -4.87E-04 | -1.18E-02 |
| 13984 | Foxh1 | FOXH1 | 3.40E-04 | 1.29E-02 |
| 13985 | Dscaml1 | DSCAML1 | -6.32E-04 | -1.94E-02 |
| 13986 | Rtn4rl2 | RTN4RL2 | -6.68E-04 | -9.47E-03 |
| 13987 | Myo18b | MYO18B | -6.85E-04 | 4.99E-03 |
| 13988 | Akap11 | AKAP11 | -2.28E-04 | -1.13E-02 |
| 13989 | Slco5a1 | SLCO5A1 | -4.50E-04 | 6.92E-02 |

| | | | | |
|---|---|---|---|---|
| 13990 | Myo7b | MYO7B | -5.60E-04 | -6.82E-03 |
| 13991 | Cpne1 | CPNE1 | -4.91E-04 | -2.52E-02 |
| 13992 | Cog8 | COG8 | -1.79E-04 | 3.84E-03 |
| 13993 | Tnks2 | TNKS2 | -1.78E-04 | -1.74E-03 |
| 13994 | Son | SON | -1.48E-04 | -1.50E-02 |
| 13995 | Yeats4 | YEATS4 | 1.86E-04 | 1.10E-02 |
| 13996 | Serpina3k | SERPINA3 | -1.81E-03 | 1.77E-01 |
| 13997 | Gtf3c3 | GTF3C3 | 2.20E-04 | -1.23E-02 |
| 13998 | Muc5ac | MUC5AC | -2.73E-04 | 2.97E-02 |
| 13999 | Sugct | SUGCT | 3.51E-04 | -2.11E-02 |
| 14000 | Capsl | CAPSL | -7.43E-04 | 2.19E-02 |
| 14001 | Rgs19 | RGS19 | 5.61E-04 | 9.39E-02 |
| 14002 | Trim36 | TRIM36 | -5.75E-04 | -6.12E-03 |
| 14003 | Slc7a8 | SLC7A8 | -5.10E-04 | -4.27E-02 |
| 14004 | Ctsz | CTSZ | -2.26E-04 | 1.42E-02 |
| 14005 | Gnb1l | GNB1L | 3.78E-04 | -1.35E-02 |
| 14006 | Zbtb8os | ZBTB8OS | -1.76E-04 | -1.89E-02 |
| 14007 | Crip3 | CRIP3 | -6.76E-04 | -2.54E-02 |
| 14008 | Socs6 | SOCS6 | 1.61E-04 | -4.21E-03 |
| 14009 | Gss | GSS | -2.08E-04 | -3.12E-02 |
| 14010 | Tmem175 | TMEM175 | -1.57E-04 | -2.08E-02 |
| 14011 | Elf1 | ELF1 | 1.97E-04 | 5.94E-03 |
| 14012 | Xpr1 | XPR1 | -2.16E-04 | -1.53E-02 |
| 14013 | Spcs2 | SPCS2 | 1.44E-04 | -6.27E-03 |
| 14014 | Tbk1 | TBK1 | -1.48E-04 | -1.99E-03 |
| 14015 | Ankrd23 | ANKRD23 | 4.24E-04 | 1.28E-02 |
| 14016 | Terf2ip | TERF2IP | 1.21E-04 | -1.68E-03 |
| 14017 | Sox11 | SOX11 | 6.05E-04 | 3.24E-02 |
| 14018 | 4933405L10Rik | C16orf86 | 2.72E-04 | 9.98E-03 |
| 14019 | Piezo2 | PIEZO2 | -3.89E-04 | 7.81E-02 |
| 14020 | Spn | SPN | 8.66E-04 | 2.19E-02 |
| 14021 | Pnma1 | PNMA1 | -3.00E-04 | 4.97E-03 |
| 14022 | Top3a | TOP3A | 2.45E-04 | -2.19E-02 |
| 14023 | Fut2 | FUT2 | 1.06E-03 | 2.11E-01 |
| 14024 | Rnf215 | RNF215 | -2.05E-04 | -1.25E-03 |
| 14025 | Cdh2 | CDH2 | 7.83E-04 | -4.74E-02 |
| 14026 | Tmem41b | TMEM41B | -1.45E-04 | 1.81E-02 |
| 14027 | Tbc1d4 | TBC1D4 | -3.70E-04 | -3.28E-02 |
| 14028 | Pgm2l1 | PGM2L1 | 5.02E-04 | 1.74E-02 |
| 14029 | Fam217a | FAM217A | 2.95E-04 | -4.58E-02 |
| 14030 | Gfod1 | GFOD1 | 3.73E-04 | -8.52E-03 |
| 14031 | Rfx7 | RFX7 | 1.50E-04 | -6.78E-03 |
| 14032 | Lbr | LBR | 1.96E-04 | -5.51E-03 |
| 14033 | Lgi4 | LGI4 | -4.56E-04 | -3.34E-02 |
| 14034 | Ube2s | UBE2S | -1.43E-04 | 9.97E-03 |
| 14035 | Zbtb49 | ZBTB49 | -2.45E-04 | -2.15E-02 |
| 14036 | Spsb2 | SPSB2 | 1.85E-04 | -2.59E-02 |
| 14037 | Rnaset2b | RNASET2 | -3.17E-04 | -1.21E-02 |
| 14038 | Ugt2b1 | UGT2B17 | -2.73E-04 | 3.48E-02 |
| 14039 | Jmy | JMY | -1.04E-04 | -3.33E-03 |
| 14040 | Usp35 | USP35 | 3.09E-04 | 8.37E-03 |
| 14041 | Rarres2 | RARRES2 | 2.47E-04 | 3.38E-02 |
| 14042 | Dnm1 | DNM1 | -2.89E-04 | -5.03E-02 |

| | | | | |
|---|---|---|---|---|
| 14043 | Crabp2 | CRABP2 | -9.18E-04 | 7.01E-02 |
| 14044 | Upk3bl | UPK3BL | -6.14E-04 | 1.05E-01 |
| 14045 | Cxxc4 | CXXC4 | -3.87E-04 | -3.24E-03 |
| 14046 | Rps18 | RPS18 | -1.10E-04 | 1.92E-02 |
| 14047 | Prg2 | PRG2 | 8.69E-05 | -9.73E-03 |
| 14048 | Sds | SDS | -2.73E-04 | 4.88E-02 |
| 14049 | Slc35g2 | SLC35G2 | -3.54E-04 | 2.54E-02 |
| 14050 | Eef2 | EEF2 | 1.08E-04 | 7.84E-03 |
| 14051 | Sipa1l1 | SIPA1L1 | 1.27E-04 | -1.62E-02 |
| 14052 | Zc3h12b | ZC3H12B | 3.20E-04 | -2.97E-02 |
| 14053 | Ripply3 | RIPPLY3 | -2.70E-04 | -2.37E-02 |
| 14054 | Zdhhc1 | ZDHHC1 | 5.24E-05 | -8.76E-03 |
| 14055 | Smarce1 | SMARCE1 | 4.81E-05 | 1.50E-02 |
| 14056 | Tle6 | TLE6 | -3.09E-04 | 3.66E-02 |
| 14057 | Orc4 | ORC4 | -9.49E-05 | -9.18E-03 |
| 14058 | Ncor1 | NCOR1 | -6.87E-05 | -3.45E-03 |
| 14059 | Lysmd3 | LYSMD3 | -8.91E-05 | 1.39E-02 |
| 14060 | Kif5a | KIF5A | 1.05E-04 | 2.49E-02 |
| 14061 | Irf2bp2 | IRF2BP2 | -1.41E-04 | 3.98E-02 |
| 14062 | Prom2 | PROM2 | -1.96E-04 | 4.69E-03 |
| 14063 | Sec24c | SEC24C | 5.57E-05 | -3.17E-03 |
| 14064 | Sgce | SGCE | -1.61E-04 | -1.81E-02 |
| 14065 | Yes1 | YES1 | -6.61E-05 | -1.51E-03 |
| 14066 | Opn4 | OPN4 | -7.86E-05 | -5.92E-03 |
| 14067 | Hnrnpu | HNRNPU | 5.16E-05 | 1.80E-02 |
| 14068 | Ttc5 | TTC5 | 4.36E-05 | -2.79E-02 |
| 14069 | Ntrk3 | NTRK3 | 1.72E-04 | 1.08E-02 |
| 14070 | Fam220a | FAM220A | 6.60E-05 | 2.80E-03 |
| 14071 | Atf6 | ATF6 | 3.05E-05 | -1.08E-02 |
| 14072 | Ifitm2 | IFITM2 | 4.91E-05 | 1.69E-02 |
| 14073 | Abca6 | ABCA6 | -2.14E-04 | 5.41E-02 |
| 14074 | Timm17b | TIMM17B | 3.55E-05 | -1.19E-02 |
| 14075 | Accs | ACCS | -7.36E-05 | -4.11E-02 |
| 14076 | Ube2g2 | UBE2G2 | 4.45E-05 | 2.46E-02 |
| 14077 | Pamr1 | PAMR1 | -1.00E-04 | 7.00E-02 |
| 14078 | Lnx2 | LNX2 | 3.50E-05 | 1.26E-02 |
| 14079 | Cend1 | CEND1 | -1.25E-04 | 5.87E-02 |
| 14080 | Daam1 | DAAM1 | 2.34E-05 | -1.32E-03 |
| 14081 | Gmeb1 | GMEB1 | 1.90E-05 | -2.43E-02 |
| 14082 | Pdp1 | PDP1 | 4.93E-05 | 2.56E-02 |
| 14083 | Tmem236 | TMEM236 | 3.28E-05 | 1.81E-02 |
| 14084 | Cyp4f18 | CYP4F2 | -8.22E-05 | 8.97E-02 |
| 14085 | Hoxb9 | HOXB9 | 4.18E-05 | 3.50E-02 |
| 14086 | Med25 | MED25 | 8.92E-06 | -4.99E-03 |
| 14087 | Phactr4 | PHACTR4 | 1.23E-05 | -1.74E-03 |
| 14088 | A430005L14Rik | C1orf174 | 1.29E-05 | 1.75E-02 |
| 14089 | Filip1 | FILIP1 | -1.61E-05 | -1.60E-02 |
| 14090 | Mat1a | MAT1A | 6.09E-05 | 2.44E-01 |
| 14091 | Ccdc141 | CCDC141 | 6.38E-06 | 3.40E-04 |
| 14092 | Pcca | PCCA | -6.29E-06 | -2.06E-02 |
| 14093 | Scmh1 | SCMH1 | 4.70E-06 | -2.15E-02 |
| 14094 | Med18 | MED18 | 8.18E-06 | 5.83E-03 |
| 14095 | Il6 | IL6 | -6.56E-06 | 8.41E-02 |

| | | | | |
|---|---|---|---|---|
| 14096 | Cacfd1 | CACFD1 | 4.45E-07 | -1.72E-03 |
